# Alterations in the plasma metabolome associated with maternal smoking during the first trimester and fetal growth

**DOI:** 10.1101/2024.11.28.24318123

**Authors:** Wadzanai Masvosva, Taija Voutilainen, Marko Lehtonen, Retu Haikonen, Seppo Auriola, Leea Keski-Nisula, Jaana Rysä, Olli Kärkkäinen

## Abstract

Tobacco smoking during pregnancy has been associated with an increased risk of adverse outcomes like low birth weight. This study determined changes in the circulating metabolome linked to maternal smoking in the first trimester and correlated these changes to the growth of the fetus. The circulating metabolome was examined from first trimester plasma samples by non-targeted (global) liquid chromatography mass spectrometry-based metabolite profiling of 227 pregnant women (99 smokers and 117 non-smokers) from the Kuopio Birth Cohort Study. Tobacco smoking was self-reported through a questionnaire and verified with cotinine measurements from plasma samples. In summary, 64 significant differences were observed between the groups after correction for multiple testing e.g. in metabolites indicating endocrine disruption (e.g., dehydroepiandrosterone sulfate [DHEA-S], VIP=2.70, d=0.68, p<0.0001), metabolites associated with oxidative stress (e.g., bilirubin, VIP=2.00, d=0.50, p<0.0001) and lipid metabolism (e.g., LysoPC 16:1, VIP=2.07, d=0.51, p<0.0001). Some of these metabolites, e.g., DHEA-S and bilirubin, correlated with low birth weight, and some, e.g. LysoPC 16:1, correlated with small head circumference at birth. In conclusion, maternal smoking during the first trimester of pregnancy was associated with an altered metabolite profile linked to endocrine disruption and increased oxidative stress.

**Plain language summary:** Tobacco smoking during pregnancy has been associated with an increased risk of adverse outcomes like low birth weight. In this study, we measured changes in the circulating metabolome linked to maternal smoking in the first trimester and correlated these changes to the growth of the fetus. Maternal smoking during the first trimester of pregnancy was associated with an altered metabolite profile linked to endocrine disruption and increased oxidative stress. Furthermore, some of these changes were associated with reduced fetal growth.

## Introduction

Smoking has been recognized as the most important preventable cause of mortality in the world^1^. Approximately 10% of women worldwide smoke during pregnancy, although the rate varies greatly between regions and countries^2^. Smoking during pregnancy exposes the fetus to the harmful effects of tobacco.

Nicotine is the main psychoactive ingredient in tobacco, and it can easily cross the placental barrier. Nicotine concentrations in fetal circulation have been reported to be even 15% higher than those detected in the maternal circulation^3^. Nicotine causes a general constriction of blood vessels, decreasing the delivery of nutrients to the developing fetus. Carbon monoxide is another harmful chemical present in tobacco smoking; it binds avidly to hemoglobin and subsequently reduces the amount of oxygen being carried to the fetus thus increasing the risk for hypoxia^4^. During pregnancy, even low levels of tobacco smoking have been associated with an increased risk of adverse outcomes like low birth weight^5^, sudden infant death syndrome^6^, behavioral and neurodevelopmental impairments^7^ and asthma and obesity^8^. The effects of maternal tobacco smoking during pregnancy may be permanent and not just be evident from childhood but also last into adulthood, for example, leading to elevated blood pressure, endothelial dysfunction, and even changes in renal structure^9^.

Only a limited number of studies have investigated changes in the metabolite profile associated with smoking during pregnancy. Gray et al. (2010) observed metabolic differences in the umbilical venous samples taken at birth between smoking exposed and non-exposed neonates^10^. For example, they reported lower concentrations of lysophosphatidylcholine (LysoPC) 16:0, LysoPC18:0, and LysoPC 18:1 in the umbilical venous samples from smoking exposed newborns. Indeed, low lysophosphatidylcholine levels have been associated with a lowered birth weight in the newborns^11^. Furthermore, Rolle-Kampczyk et al. (2016) examined the impact of maternal smoking on endogenous metabolites in 40 maternal and umbilical blood samples. They revealed significant alterations in several metabolites, including sphingomyelin SM 26:0 and several phosphatidylcholines (PCs), in both mothers and children within the smoking-exposed group^12^. In their evaluation of urine samples from the first and second antenatal visit of 105 pregnant Afro-American women, Tan et al. (2022) revealed that 47 metabolites that have been linked with increased inflammation and decreased oxidation of amino acids were associated with high cotinine levels. These detected metabolites also have potential health-related effects during gestation with the possibility to influence several pathophysiological pathways causing adverse birth outcomes^13^. However, to our knowledge, there have been no studies investigating tobacco smoking associated alterations in the metabolite profiles during the first trimester, which is a crucial time during pregnancy since it is the period of organogenesis ^14^.

Therefore, here we wanted to examine the changes in the maternal metabolome associated with smoking in the first trimester to increase our understanding of the metabolic consequences of gestational smoking. In this nested case-control study, our aim was to identify cigarette smoking-induced changes in the circulating metabolome by applying a non-targeted, or global, metabolomics approach in the evaluation of first trimester plasma samples of women in the Kuopio Birth Cohort (KuBiCo). In addition, we aimed to investigate how these changes in the metabolome could be associated with fetal growth.

## Methods

### Study Cohort

This study is part of Kuopio Birth Cohort (KuBiCo), which is a joint research effort between the University of Eastern Finland (UEF), Kuopio University Hospital (KUH), and National Institute for Health and Welfare (THL)^15^. The KuBiCo study was approved by the Research Ethics Committee, Hospital District of Central Finland in 15.11.2011 and all participants have given their written electronic consent. The study was conducted in accordance with the Basic & Clinical Pharmacology & Toxicology policy for experimental and clinical studies^16^.

The cohort consists of women from Finnish Northern Savo county expected to have delivery at KUH. The participants were recruited during their first trimester routine antenatal visit by a health care nurse, during weeks 6-9 of gestation mainly from the outpatient maternity clinics. Additionally, these women filled in several questionnaires which evaluated their background risk for ongoing pregnancy in the Pikku-Haikara system designed in the Kuopio University Hospital for pregnant women. The information on gestational smoking/tobacco exposure was gathered via standardized questionnaires. Between the years 2012 and 2020, approximately 5000 first trimester plasma samples were collected routinely for the KuBiCo study at the same time with the first trimester fetal screening samples. This study utilizes ethylenediaminetetraacetic acid (EDTA) plasma samples, which were kept at -80°C until the metabolomics analysis, collected during the first trimester between years 2013 and 2018^15^.

To allow the samples to be used for this study, the subjects of interest should be pregnant with a parity of six or less, with the preference to use samples from their first pregnancies. Participants’ self-reported tobacco use gathered in an online questionnaire was used to evaluate tobacco smoking during pregnancy ^17^. Tobacco smoking was defined as taking any tobacco products after conception. There was no limitation on the number of cigarettes smoked per day. The smokers were compared against controls who recorded not having smoked at all during pregnancy. Since it is evident that self-reported data is not 100% reliable, we initially selected more controls than tobacco smokers. After liquid chromatography mass spectrometry (LC-MS) analysis, we excluded seven subjects from the control group as they had elevated levels of nicotine or cotinine in their blood sample. This led to the selection of two study groups: a control group with 117 samples of women who had not smoked during pregnancy and a study group with 99 samples of women who had smoked during their pregnancy but did not report any alcohol use during pregnancy. We excluded alcohol users, since alcohol use during pregnancy is more common in those who smoke and is known to influence the metabolome during pregnancy^18^. The analysis order of the samples was randomized using a random number generation algorithm.

This study used the following descriptive characteristics collected from the KUH birth registry: maternal age, height and weight, body mass index (BMI; kg/m^2^), parity, gravidity, duration of pregnancy at birth, length of maternal hospital stay after birth, relationship status, and in the smoking women, the daily amount of cigarettes before and during pregnancy to allow the estimation of the smoking index. Additionally, in the newborns, data was collected on gender, birthweight, head circumference, placental weight, APGAR scores (1 and 5 minutes), need for special or neonatal intensive care, ventilation assistance or intubation, need for postnatal antibiotic treatment or phototherapy, and the presence of any congenital malformations.

### Non-targeted LC-MS based metabolite profiling

The metabolomics analysis was performed according to previously released guidelines with some modifications^19^. In brief, sample preparation was done by thawing plasma samples in ice-cold water; the samples were maintained in ice-cold water during all waiting periods before they were transferred for sample preparation into 96 well plates (Thermo Scientific Nunc) embedded on wet ice and covered with a filter plate (Captiva ND filter plate 0.2 um PP 5/pk, Part no. A5969002, 732-2669). Then, 320 µL of cold acetonitrile (HPLC LC-MS Grade 83640.320, VWR) were dispensed into the filter plate well. The plasma samples were then vortexed for 10 seconds at maximum speed and subsequently 80 µL of the plasma samples were added to the same well with acetonitrile. The Andrew Alliance robot with a Microplate shaker (519.4004.14HAK, Vacuum + 518.6093.160UY, 8-ch 1200) was used for sample preparation. Preparation of the quality control sample (QC) was done by adding 10 µL of each sample to the same clean microcentrifuge tube and mixing. These QC samples were injected at the beginning of the analysis to stabilize the liquid chromatography (LC) column. QC samples were also tested after every 12^th^ sample in the analysis runs to assess batch and drift correction and data quality analysis, as well as in the tandem mass spectrometry analyses. The samples were precipitated by centrifuging the plate for 5 minutes at 700xg at 4 °C after which, the filter plate was sealed tightly to avoid sample evaporation prior to the following metabolomics analysis

The metabolomics analysis used ultrahigh performance liquid chromatography (UPLC) with a Thermo Q Exactive Hybrid quadrupole-orbitrap mass spectrometer (MS). Samples were analyzed using two chromatographic techniques, reversed phase (RP) and hydrophilic interaction (HILIC), in both positive and negative polarities. The RP chromatography column was Zorbax Eclipse XDB-C18. The mobile phase A was H2O + 0.1 % HCOOH, mobile phase B was MeOH + 0.1 % HCOOH with a 16.5 min gradient and a flow rate of 0.4 ml/min. The HILIC chromatographic set-up used an Acquity BEH amide column, Mobile phase A: 50% acetonitrile + 20 mM ammonium formate buffer, B: 90 % acetonitrile + 20 mM ammonium formate buffer, 12.5 min gradient, 0.6 ml/min.

Both negative and positive electrospray ionization (ESI) were used in both analytical modes. ESI settings were ray voltage 3.5 kV for positive and 3.0 kV for negative, max spray current 100, flow rates 40 for sheath gas, 10 for auxiliary gas, and 2 for spare gas (as arbitrary units for ion source), S-lens RF level 50 V, capillary and probe heater temperature 300 °C. A full scan range from m/z 60 to 700 in the HILIC modes and 120 to 1200 in the RP modes with a resolution of 70 000 (m/Δm, full width at half maximum at 200 u) and an automated injection time and gain control targeted at 1 000 000 ions. The m/z ranges were limited to ions of interest in each mode to increase the sensitivity of the measurement. In the tandem mass spectrometry (MS/MS), three peaks with apex triggers ranging from 0.2 up to 3 s were selected for MS/MS fragmentation with 15 s dynamic exclusion. A normalized collision energy at 20, 30, and 40, was used in MS/MS with a mass resolution of 17 500 (m/Δm, full width at half maximum at 200 u), an automated gain targeted at 50 000, and an isolation window of 1.5 m/z.

Data analysis was done using MS-dial software 4.90 version for peak picking, peak alignment, and metabolite identification. Data collection had an MS1 tolerance of 0.005 and an MS2 tolerance of 0.025. The settings for the retention time parameters started at 0.5 minutes and stopped at the end of the analytical run, collecting all the features within the mass ranges set for each mode. Peak detection parameters had a minimum peak height of 300,000 with an amplitude smoothing level of 3 scans, minimum peak width of 8 scans, and a mass slice width of 0.1 Da. Deconvolution parameters had a sigma window value of 0.5 with 0 as the amplitude cut-off. The identification was performed against the University of Eastern Finland in-house database with accurate mass tolerance of MS1 0.008 Da and MS2 0.025 Da. Automatic identifications were verified and corrected manually. Adduct ion settings of [M+H]+, [M+NH4]+, [M+Na]+, [M+K]+, [M+Li]+, [M+ACN+H]+, [M+H-H2O]+, [M+H-2H2O]+, [2M+H]+ were utilized in positive polarity and [M-H]-, [M-H2O-H]-, [M+Na-2H]-, [M+Cl]-, [M+FA-H], [M+Hac-H]-, [2M-H]- when we applied negative polarity. The alignment parameters settings had a retention time of 0.05 mins with MS1 tolerance 0.015 for molecular features detected in at least 30% of the samples in one of the groups with a gap-filling by compulsion.

Preprocessing of the data (data quality analysis, drift and batch effect correction, imputation of missing values), and statistical analyses were done using the R-package notame^19^.

The identification of metabolites focused on those molecular features with p-values <0.05. The molecular features were grouped into 4 levels according to the community guidelines^20^. Level 1 identification included those compounds confirmed by matching to exact mass, isotope pattern, retention time, and MS2 fragmentation with chemical standards from the in-house library. Level 2 identification comprised putatively annotated compounds, identified by matching to exact mass and MS2 spectra against public libraries. Level 3 identification referred to the compounds identified at the chemical group level but lacking an exact compound identification, while level 4 consisted of unknown compounds.

### Statistical analysis

Differences between background characteristics were evaluated with Welch’s t-test and Chi^2^ test. With respect to the metabolomics data, univariate analysis was done with Cohen’s d effect sizes with p-values being calculated using Welch’s t-test. Since molecular features in metabolomics analysis are not independent, the number of principal components needed to explain 95% of the variance in the metabolomics data in the principal component analysis (PCA) was used and to account for multiple testing the Bonferroni method was applied. Here, 184 PCs were needed to explain 95% of the variation in the metabolomics data, therefore the α level was adjusted to 0.0003. Findings with p-values between 0.05 and 0.0003 were considered to be trends. Additionally, we used partial-least-squares discriminant analysis (PLS-DA) to identify the variables that contributed most extensively to the variation between smoking and nonsmoking pregnant women in a multivariate analysis. Cross-validation was done by splitting the dataset into 7 subsets. Variable importance to the projection (VIP) values from PLS-DA are reported for each variable. Correlations between molecular features and birth weight or head circumference were calculated using linear regression. We used SIMCA 17.0 for multivariate data analysis solution, whereas we utilized the notame (version 0.3.0) R-package for the calculation of univariate statistics for metabolomics data^19^. JASP (version 0.17) software was used to calculate statistics related to the background characteristics between the non-smoking and smoking groups. Figures were made using Prism (GraphPad software Inc. version 10.0) and SIMCA version 17.0.

## Results

The characteristics of the pregnant women and their newborns women are shown in Table 1. Smokers had a significantly lower parity, higher BMI values, pre-pregnancy weights and were less likely to be in a relationship when compared to the nonsmoking controls. On average, the newborns of smokers had a lower birth weight and head circumference when compared to control newborns. Furthermore, they needed more antibiotic therapy and treatment in the intensive care unit than the controls.

**Table 1:** Background characteristics of the study women.

| **Maternal characteristics** |  |  |  |  |  |
| --- | --- | --- | --- | --- | --- |
|  | **Controls n=117** |  | **Cases n=99** |  | **p value** |
| Maternal age (years, mean; ±SD) | 28.4 | $\pm 5.7$ | 27.4 | $\pm 5.7$ | 0.192<sup>a</sup> |
| Maternal height (cm, mean; ±SD) | 163.9 | $\pm 5.6$ | 165.3 | $\pm 5.5$ | 0.077<sup>a</sup> |
| **Maternal weight, kg (pre-pregnancy) (mean; ±SD)** | **64.2** | **$\pm 14.0$** | **69.5** | **$\pm 13.8$** | **0.006<sup>a</sup>** |
| **Maternal BMI kg/m<sup>2</sup> (mean; ±SD)** | **23.8** | **$\pm 4.9$** | **25.5** | **$\pm 5.1$** | **0.020<sup>a</sup>** |
| **Parity (N, mean; SD)** | **1.7** | **$\pm 2.2$** | **1.2** | **$\pm 1.2$** | **0.026<sup>a</sup>** |
| Gravidity (N, mean; SD) | 3.3 | $\pm 2.8$ | 3.1 | $\pm 1.9$ | 0.522<sup>a</sup> |
| Duration of pregnancy at birth (days, mean; SD) | 278 | $\pm 11$ | 277 | $\pm 15$ | 0.566<sup>a</sup> |
| Hospital stay after birth, days (mean; SD) | 3.3 | $\pm 1.7$ | 3.7 | $\pm 2.7$ | 0.152<sup>a</sup> |
| **In a relationship (n, %)** | **108** | **92%** | **62** | **63%** | **<0.0001<sup>b</sup>** |
| **Cigarettes per day before pregnancy (mean; ±SD)** | **0** | **$\pm 0$** | **15.2** | **$\pm 6.3$** | **<0.0001<sup>a</sup>** |
| **Cigarettes per day during pregnancy (mean; ±SD)** | **0** | **$\pm 0$** | **6.3** | **$\pm 4.0$** | **<0.0001<sup>a</sup>** |
| **Smoking Index (mean; SD)** | **0** | **$\pm 0$** | **2.4** | **$\pm 0.9$** | **<0.0001<sup>a</sup>** |
| **Characteristics of the newborns** |  |  |  |  |  |
| Female (n, %) | 57 | 49% | 43 | 43% | 0.438<sup>b</sup> |
| **Birth weight (g, mean ±SD)** | **3552** | **$\pm 513$** | **3397** | **$\pm 534$** | **0.032** |
| **Head circumference (cm, mean; ±SD)** | **35.2** | **$\pm 1.4$** | **34.7** | **$\pm 1.6$** | **0.020<sup>a</sup>** |
| Placenta weight (grams, mean; ±SD) | 618.6 | $\pm 141.4$ | 605.5 | $\pm 135.0$ | 0.489<sup>a</sup> |
| APGAR scores 1 min (mean; ±SD) | 8.8 | $\pm 1.0$ | 8. | $\pm 1.1$ | 0.615<sup>a</sup> |
| APGAR scores 5 min (mean; ±SD) | 8.9 | $\pm 1.3$ | 9.0 | $\pm 0.8$ | 0.611<sup>a</sup> |
| **Child's special care after birth (n, %)** | **13** | **11%** | **21** | **21%** | **0.042<sup>b</sup>** |
| **Need for treatment in Neonatal Intensive Care Unit (n, %)** | **10** | **8.5%** | **21** | **21%** | **0.008<sup>b</sup>** |
| Ventilation assistance (n, %) | 0 | 0% | 1 | 1% | 0.276<sup>b</sup> |
| Intubation (%) | 0 | 0% | 1 | 1% | 0.276<sup>b</sup> |
| **Postnatal antibiotic treatment (n, %)** | **4** | **3.4%** | **13** | **13.1%** | **0.008<sup>b</sup>** |
| Phototherapy (%) | 5 | 4.3% | 5 | 5% | 0.787<sup>b</sup> |
| Malformations (%) | 1% | 0.9% | 0 | 0% | 0.357<sup>b</sup> |

Legend: <sup>a</sup> Welch's t test; <sup>b</sup> Chi<sup>2</sup> test.

A total of 9600 molecular features were observed in the non-targeted metabolomics analysis (Supplementary table 1). The PCA analysis revealed that 184 principal components were required to explain 95% of the overall variance in the metabolomics data. These results were used to adjust the α level to 0.0003 to account for multiple testing using Bonferroni correction after which a total of 1109 molecular features had a p-value < 0.05, and 193 molecular features had a p-value < 0.0003 (Figure 1). The PLS-DA model also revealed a difference between the first trimester plasma samples of nonsmokers and smokers (6 components, cum R2Y = 0.99, Q2 = 0.70) (Figure 1, Supplementary table 1).

**Figure 1:**
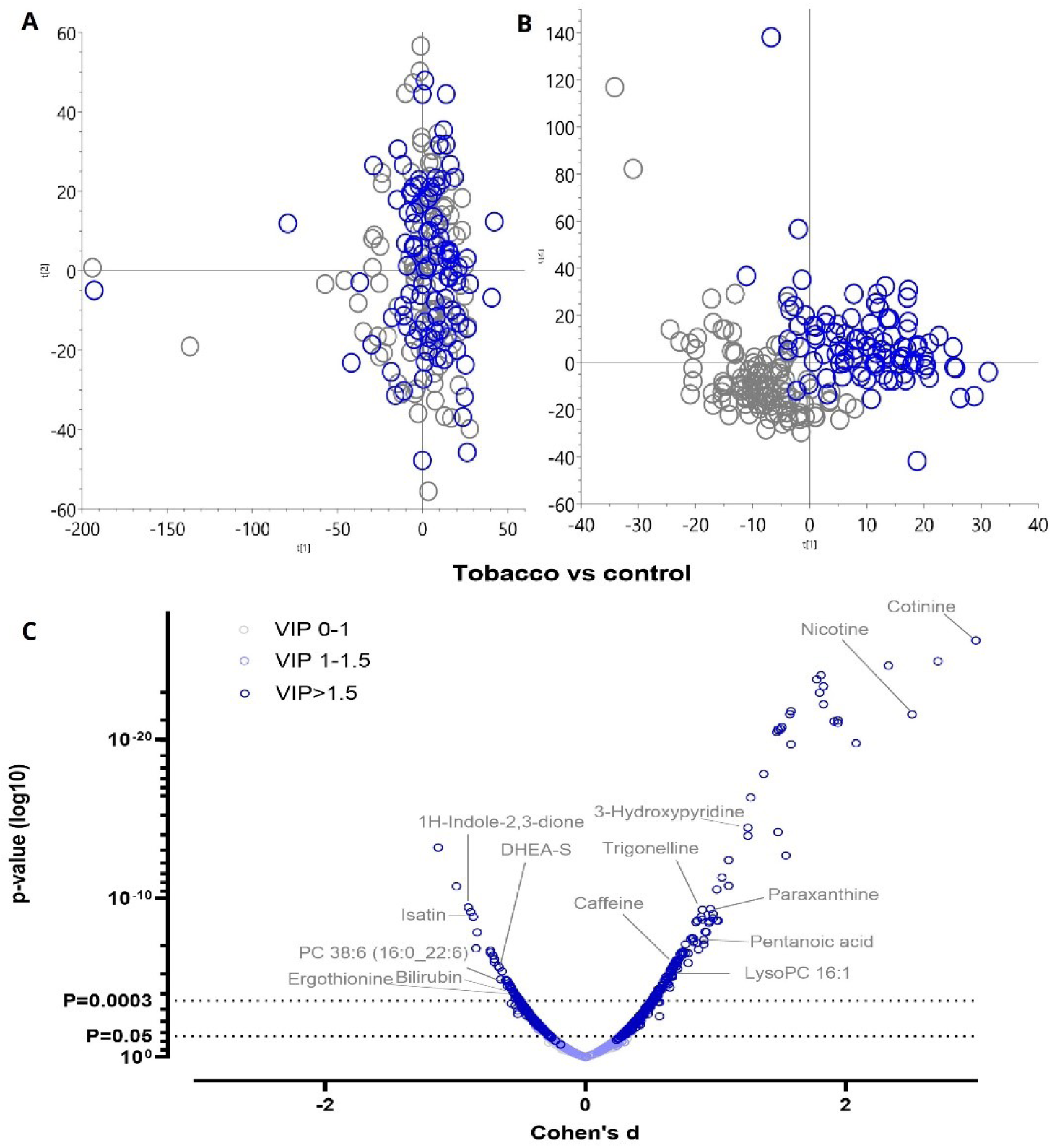
Overview of metabolite profiling from the first trimester maternal plasma samples. A total of 9600 molecular features were observed from the chosen samples. Panel **A** shows the outcomes of the principal component analysis (PCA). A total of 184 components were needed to account for 95% of the variance in the data. PC1 (t1) explained 4.5 % of the variation in the data, and PC2 (t2) explained 4.3% of the data. Panel **B** shows partial least squares discriminant analysis (PLS-DA) which illustrates the good separation between the study groups (6 components, cum R2Y = 0.99, Q2 = 0.70). Panel **C** shows the p values (Welch’s t-test), Cohen’s d effect sizes, and VIP values from the PLS-DA) of all of the relevant molecular features. A total of 193 molecular features were significantly altered between the smokers’ and nonsmokers’ samples (adjusted α level= 0.0003, Bonferroni’s correction).

Our results showed that the circulating metabolite profiles from the first trimester plasma samples of smoking pregnant women were different from the nonsmoking controls (Figure 1). The identified metabolites were grouped according to the human metabolome database (HMDB)^21^. For example, smokers had significantly lower levels of bilirubin and biliverdin, and significantly higher levels of dehydroepiandrosterone sulfate (DHEA-S) (Figure 2). High levels were also evident for several exogenous compounds e.g. cotinine, nicotine, and trigonelline along with xanthines like caffeine, paraxanthine, and theophylline in the smokers when compared to the controls.

**Figure 2:**
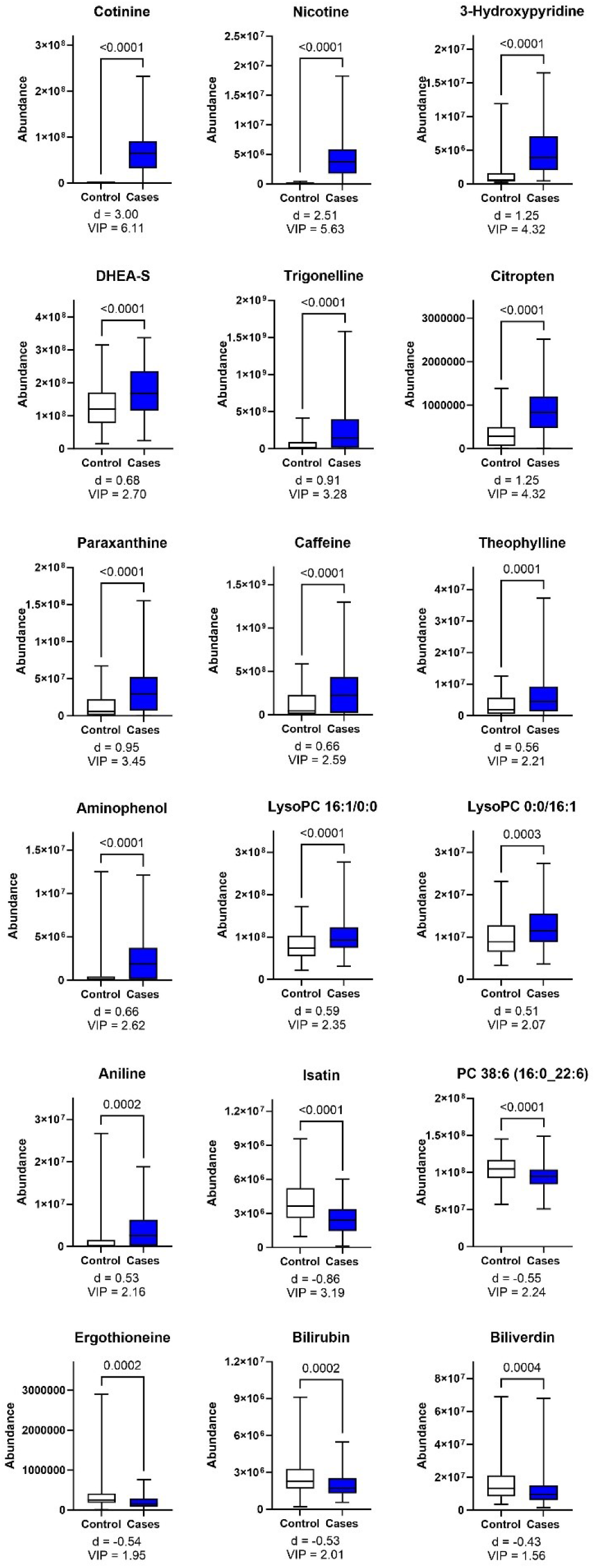
Smokers show altered metabolite profile in the first-trimester plasma samples when compared to nonsmoking controls. Box plots with whiskers (min and max) are shown with p-value from a Welch’s t-test (multiple testing adjusted α level was 0.0003). Biliverdin result should be considered a trend. Abbreviations: DHEA-S, dehydroepiandrosterone sulfate; PC, phosphatidylcholine.

The correlation analysis indicated that birth weight was also positively associated with the levels of biliverdin (r = 0.20, p = 0.0029), creatine (r = 0.15, p = 0.0029) and PC38:6 (16:0_22:6) (r = 0.14, p = 0.0035) and negatively associated with those of cotinine (r = -0.18, p = 0.00096), Asp-Phe (r-0.15, p = 0.0026), DHEA-S (r = -0.15, p = 0.0265) and AC14:1 (r =-0,13, p = 0.0049). Head circumference was negatively associated with the levels of 7a-hydroxy-3-oxo-4-cholestenoic acid (r = -0.18, p=0.001), cotinine (r = -0.17,p=0.0013), LysoPC 16:1 (16:1/0:0) (r = -0.16, p = 0.0019), LysoPC 16:1 (0:0/16:1) (r = -0.15, p = 0.0028), AC 06:0 (r = -0.15, p = 0.0037) and lenticin (r = -0.14, p = 0.0043). These results are shown graphically in Figure 3 and Figure 4. Furthermore, levels of nicotine, cotinine and 3-hydroxypyridine correlated (r values between 0.56-0.76 and p values < 0.0001) with the self-reported number of cigarettes smoked per day both before and during pregnancy (Supplementary Figure 1).

**Figure 3:**
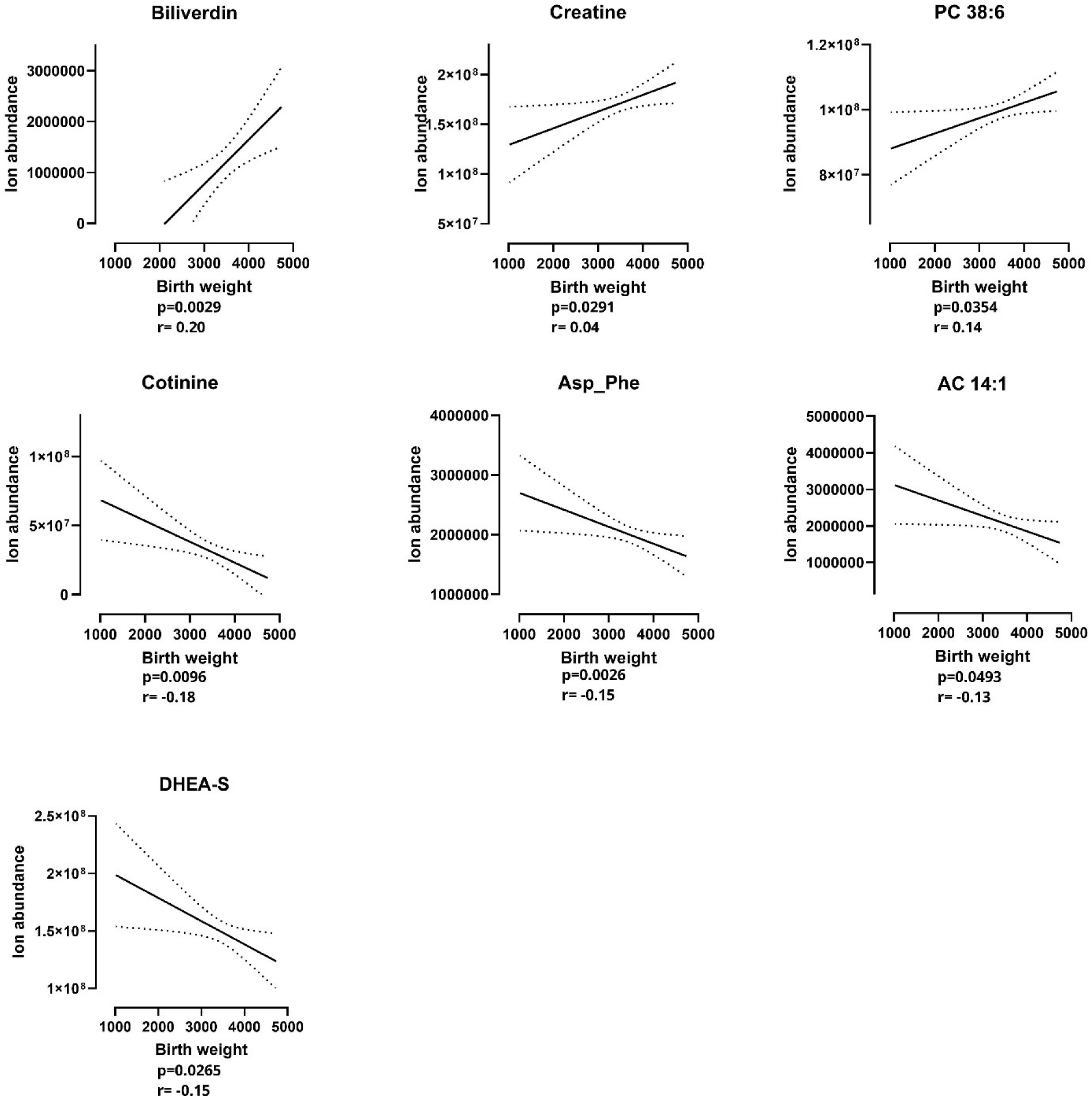
Association between the birth weight of the newborn and metabolite levels in the first trimester of pregnancy. Three metabolites were positively associated with birth weight and four were negatively associated with birth weight. Abbreviations: AC 14.1: tetradecenoylcarnitine; Asp_Phe: Aspartyl-phenylalanine; PC: phosphatidylcholine; p: p-value, r: Pearson correlation coefficient

**Figure 4:**
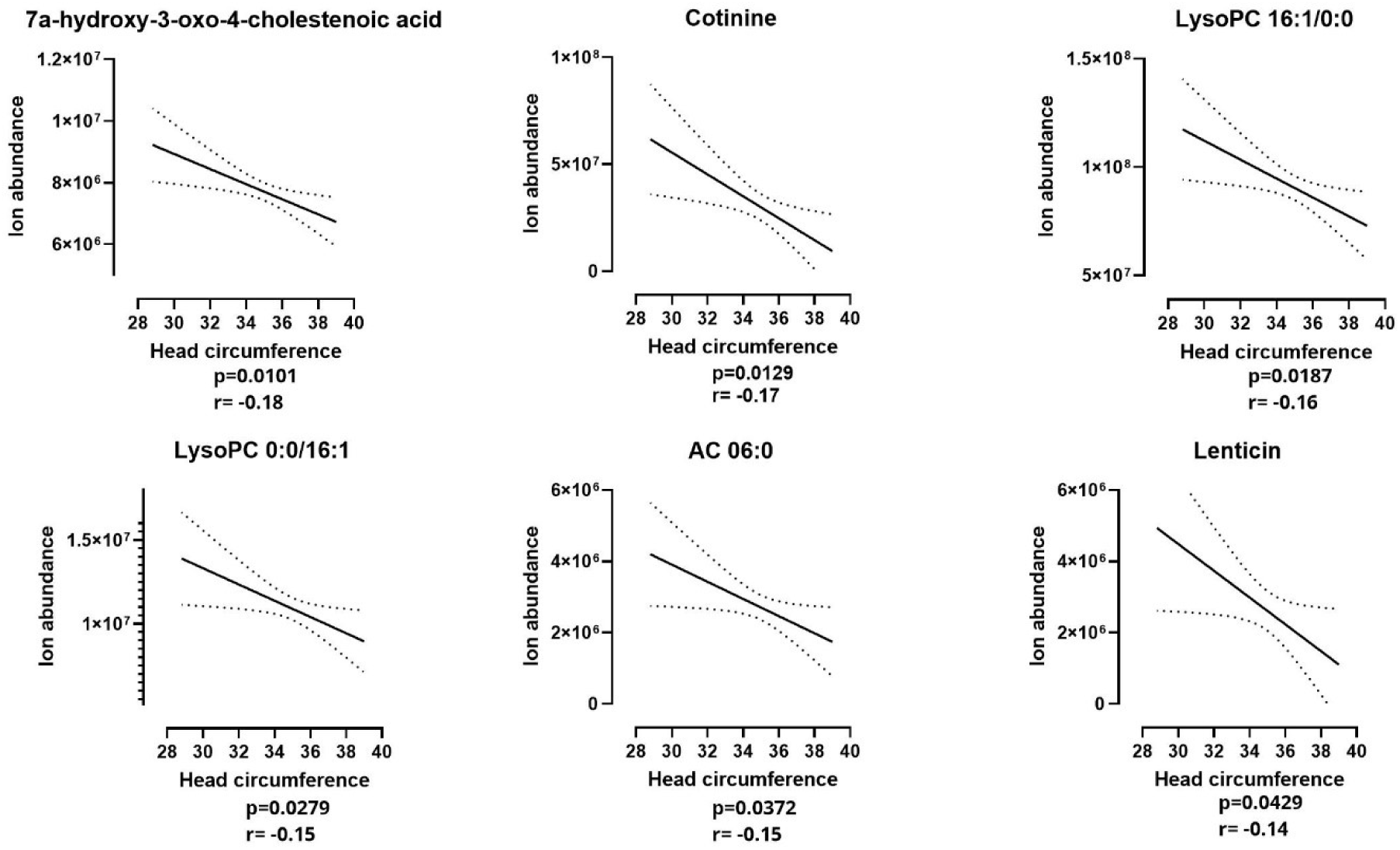
Correlations between the head circumference of the newborn and first trimester levels of metabolites. Six metabolites were negatively associated with differences in the head circumference between the groups. Abbreviations: AC 06:0, Hexanoylcarnitine; LysoPC: lysophosphatidylcholine, p: p-value, r: Pearson correlation coefficient

## Discussion

We demonstrated that the profiles of the first trimester circulating metabolites of smoking pregnant women were different from those of non-smoking controls. Furthermore, some of these differences were associated with the known unfavorable effects of smoking such as low birth weight and smaller head circumference. Key differences in the metabolite profiles of smoking pregnant women were detected in several factors e.g., elevated levels of DHEA-S and reduced levels of bilirubin and ergothioneine, as well as in certain specific small molecules which are linked directly to the tobacco smoking (nicotine, cotinine, 3-hydroxypyridine) and a higher coffee intake (caffeine, paraxanthine, trigonelline). This finding is in line with the published literature that there is an association between higher nicotine/ cotinine levels and unfavorable fetal/ neonatal effects like low birth weight ^22^. However, this study expands these findings by linking also other metabolites such as DHEA-S and bilirubin to fetal and neonatal growth.

We found that pregnant women who were smokers had high levels of DHEA-S in the first trimester plasma samples, and that these were further associated with the lower birth weight. DHEA-S is as a metabolic intermediate in the biosynthesis of sex steroids ^23^. Smoking has been shown to influence adrenal hormone production by upregulating the adrenal β-arrestin 1 enzyme, which is involved in the development of hyperaldosteronism and consequently to higher basal levels of DHEA-S^24,25^. The elevated DHEA-S levels are evidence of an endocrine disorder attributable to maternal smoking and it has been reported that endocrine disturbances can be linked to growth retardation and neurological impairments in the newborn^26^.

Moreover, smokers displayed lowered plasma levels of ergothioneine, which is a microbiota-associated antioxidant. Earlier studies have shown that low levels of ergothioneine during pregnancy are associated with negative pregnancy outcomes like pre-eclampsia^27^. Conversely, high ergothioneine levels are associated with health benefits like protection against cardiovascular diseases^28^, and this seems to be the case for biliverdin and bilirubin^29^, implying that this protective mechanism has become impaired in smokers. Smokers had low levels of two endogenous antioxidant biomarkers i.e., bilirubin and biliverdin, when compared to the controls, a finding which is in line with previous reports^30,31^. Biliverdin is a metabolite formed from the breakdown of hemoproteins; it is further reduced to bilirubin by biliverdin reductase ^32^. Low levels of bilirubin have been associated not only with smoking-related respiratory diseases^30^, but also with autoimmune diseases like lupus and rheumatoid arthrtis^33^. Thus, the low levels ergothioneine, biliverdin and bilirubin indicate that the pregnant women who smoke have a reduced antioxidative capacity while being subjected to increased oxidative stress; these types of changes could well trigger unfavorable outcomes like growth restrictions ^5^. In line, we observed that the presence of low biliverdin levels in the first trimester plasma samples was associated with low birth weights in the newborns.

Additionally, our study showed a lower abundance of isatin (1H-indole-2,3-dione) in the smoking group when compared to the control group. The levels of isatin have been reported to be lower in smokers due to the impact of monoamine oxidase activity and oxidative stress since monoamine oxidase is the enzyme that metabolizes isatin^34^. An increased level of oxidative stress has been reported to alter the regulation of isatin in the body, where it acts as a neuroprotective agent^35^. It would be enlightening to clarify if low isatin levels would be associated with neurological impairments in newborns.

Moreover, the metabolomics analysis also showed that the levels of three metabolites linked to tobacco smoking, namely nicotine, cotinine, 3-hydroxypyridine, were significantly increased in blood samples of the tobacco-exposed mothers. Positive correlations were shown between the self-reported number of cigarettes smoked per day both before and during pregnancy with these 3 mentioned metabolites (Supplementary Figure 1). Here, cotinine exhibited a significant negative correlation to birth weight and head circumference. Interestingly, previous studies have shown that the metabolism of nicotine is accelerated during pregnancy; this was assessed by measuring the nicotine metabolite ratio at different gestational weeks^36^.

Furthermore, the observed high levels of metabolites associated with caffeinated products (caffeine, theophylline, paraxanthine and trigonelline) are in line with previous studies that found that smoking pregnant women consume more caffeinated products compared to their nonsmoking pregnant counterparts^37^. It is recognized that the metabolism of compounds present in coffee is reduced during pregnancy^38^, and additionally, after crossing the placental barrier, caffeine’s metabolism is decreased possibly leading to prolonged effects in the fetus ^39^. Previously, an increased coffee intake has been associated with a low birth weight ^40^, although here we did not observe correlations between birth weight and the levels of coffee metabolites.

Additionally, we detected significant alterations in the concentrations of lipids; higher levels (e.g., LysoPC 16:1), and lower levels (e.g., PC 38:6) in the tobacco smoking group. Disturbances in lipid levels have been associated with negative developmental effects in the newborns’ brains and retinas ^41^. Also here, the high levels of LysoPC 16:1 were associated with a small head circumference whereas low levels of PC 38:6 were linked with a reduced body weight at birth.

The limitations of the current study include the self-reported use of tobacco smoking by the pregnant women. Although we had access to nicotine and cotinine levels which made it possible to detect current users, there might be some cases of pregnant women who were infrequent smokers as the blood samples were taken as a part of routine visits during the first trimester. We also observed some limitations of self-reporting, i.e., there were 7 controls that had to be excluded from the study because of high cotinine levels in these samples. Another limitation is the sample size, which was relatively small and limited the statistical power. For example, with a higher sample size, differences in the levels of sterols and lipids could have reached statistical significance (Supplementary table 1). In addition, the population used for this study was mostly from the Northern Savo area of Finland, likely with similar lifestyle factors such as their diet. This could explain the increased abundance of metabolites like paraxanthine which might not be evident in other populations that do not consume as many caffeinated products. Therefore, a similar study would need to be done with a different target population. Additionally, for the correlation analysis, the time between the 1^st^ trimester samples collection and its linkage to the health of the newborns, means that many positive and negative changes could have taken place in the remaining months of the pregnancy. The maternal metabolite profile may have significantly altered during this time, and this would be reflected in the attained results^42^. Moreover, the plasma samples were stored in -80°C for between 5-10 years before analysis. Although these are the recommended conditions for plasma sample storage^43^, measuring metabolomes is highly sensitive and there could be a possibility of it being altered during the storage from example in case of lipids. However, the storage times were similar between the study groups making the results comparable between the groups.

In conclusion, the current study successfully compared the differences between smoking and nonsmoking pregnant women in the circulating metabolome as analyzed from plasma samples taken in the first trimester. Tobacco smoking during early pregnancy was associated with alterations in metabolites associated with a disruption of endocrine functions and increased oxidative stress and linked with a restricted growth of the fetus. Further studies will need to be conducted in several different populations in order to widen the generalizability of these results.

## Acknowledgements

This work was supported by the Finnish Ministry of Social Affairs and Health for the VAURAS project, and the Finnish Foundation for Alcohol Studies. This paper belongs to the studies carried out by the Kuopio Birth Cohort consortium (www.KuBiCo.fi) and we thank our colleagues who are responsible for the design and conduct of the KuBiCo. We want to thank Essi Järvelä for assistance with sample collection, Miia Reponen and Juulia Kuparinen for technical assistance with the mass spectrometry analyses, Ewen MacDonald for proof-reading, and the staff of the Department of Obstetrics and Gynecology in Kuopio University Hospital. The authors also want to thank Biocenter Finland and Biocenter Kuopio for supporting the core LC-MS laboratory facility.

## Data Sharing Statement

The data that support the findings of this study are available on request from the corresponding author, OK. The study plan approved by the ethical committee and the participant consent terms preclude public sharing of these sensitive data, even in anonymized form.

## References

1. Reitan T, Callinan S. Changes in Smoking Rates Among Pregnant Women and the General Female Population in Australia, Finland, Norway, and Sweden. Nicotine Tob Res Off J Soc Res Nicotine Tob. 2017;19(3):282–289. doi:10.1093/ntr/ntw188

2. Havard A, Chandran JJ, Oei JL. Tobacco use during pregnancy. Addict Abingdon Engl. 2022;117(6):1801–1810. doi:10.1111/add.15792

3. Wickström R. Effects of nicotine during pregnancy: human and experimental evidence. Curr Neuropharmacol. 2007;5(3):213–222. doi:10.2174/157015907781695955

4. Kubista E. [Smoking in pregnancy]. Wien Med Wochenschr 1946. 1994;144(22-23):529-531.

5. Rumrich I, Vähäkangas K, Viluksela M, Gissler M, de Ruyter H, Hänninen O. Effects of maternal smoking on body size and proportions at birth: a register-based cohort study of 1.4 million births. BMJ Open. 2020;10(2):e033465. doi:10.1136/bmjopen-2019-033465

6. Anderson TM, Lavista Ferres JM, Ren SY, et al. Maternal Smoking Before and During Pregnancy and the Risk of Sudden Unexpected Infant Death. Pediatrics. 2019;143(4):e20183325. doi:10.1542/peds.2018-3325

7. Wehby GL, Prater K, McCarthy AM, Castilla EE, Murray JC. The Impact of Maternal Smoking during Pregnancy on Early Child Neurodevelopment. J Hum Cap. 2011;5(2):207–254. doi:10.1086/660885

8. Ino T, Shibuya T, Saito K, Ohtani T. Effects of maternal smoking during pregnancy on body composition in offspring. Pediatr Int Off J Jpn Pediatr Soc. 2011;53(6):851–857. doi:10.1111/j.1442-200X.2011.03383.x

9. Banderali G, Martelli A, Landi M, et al. Short and long term health effects of parental tobacco smoking during pregnancy and lactation: a descriptive review. J Transl Med. 2015;13:327. doi:10.1186/s12967-015-0690-y

10. Peacock JL, Palys TJ, Halchenko Y, et al. Assessing tobacco smoke exposure in pregnancy from self-report, urinary cotinine and NNAL: a validation study using the New Hampshire Birth Cohort Study. BMJ Open. 2022;12(2):e054535. doi:10.1136/bmjopen-2021-054535

11. LaBarre JL, Puttabyatappa M, Song PXK, et al. Maternal lipid levels across pregnancy impact the umbilical cord blood lipidome and infant birth weight. Sci Rep. 2020;10(1):14209. doi:10.1038/s41598-020-71081-z

12. Rolle-Kampczyk UE, Krumsiek J, Otto W, et al. Metabolomics reveals effects of maternal smoking on endogenous metabolites from lipid metabolism in cord blood of newborns. Metabolomics Off J Metabolomic Soc. 2016;12:76. doi:10.1007/s11306-016-0983-z

13. Tan Y, Barr DB, Ryan PB, et al. High-resolution metabolomics of exposure to tobacco smoke during pregnancy and adverse birth outcomes in the Atlanta African American maternal-child cohort. Environ Pollut Barking Essex 1987. 2022;292(Pt A):118361. doi:10.1016/j.envpol.2021.118361

14. Donovan MF, Cascella M. Embryology, Weeks 6-8. In: StatPearls. StatPearls Publishing; 2024. Accessed October 14, 2024. http://www.ncbi.nlm.nih.gov/books/NBK563181/

15. Huuskonen P, Keski-Nisula L, Heinonen S, et al. Kuopio birth cohort - design of a Finnish joint research effort for identification of environmental and lifestyle risk factors for the wellbeing of the mother and the newborn child. BMC Pregnancy Childbirth. 2018;18(1):381. doi:10.1186/s12884-018-2013-9

16. Tveden-Nyborg P, Bergmann TK, Jessen N, Simonsen U, Lykkesfeldt J. BCPT 2023 policy for experimental and clinical studies. Basic Clin Pharmacol Toxicol. 2023;133(4):391-396. doi:10.1111/bcpt.13944

17. Voutilainen T, Keski-Nisula L, Rysä J, Kärkkäinen O. Parental cigarette smoking before and during pregnancy in a cohort of 21 472 pregnancies. Basic Clin Pharmacol Toxicol. 2024;134(4):543–555. doi:10.1111/bcpt.13987

18. Lehikoinen AI, Kärkkäinen OK, Lehtonen MAS, Auriola SOK, Hanhineva KJ, Heinonen ST. Alcohol and substance use are associated with altered metabolome in the first trimester serum samples of pregnant mothers. Eur J Obstet Gynecol Reprod Biol. 2018;223:79–84. doi:10.1016/j.ejogrb.2018.02.004

19. Klåvus A, Kokla M, Noerman S, et al. “notame”: Workflow for Non-Targeted LC-MS Metabolic Profiling. Metabolites. 2020;10(4):135. doi:10.3390/metabo10040135

20. Sumner LW, Amberg A, Barrett D, et al. Proposed minimum reporting standards for chemical analysis Chemical Analysis Working Group (CAWG) Metabolomics Standards Initiative (MSI). Metabolomics Off J Metabolomic Soc. 2007;3(3):211–221. doi:10.1007/s11306-007-0082-2

21. Wishart DS, Guo A, Oler E, et al. HMDB 5.0: the Human Metabolome Database for 2022. Nucleic Acids Res. 2022;50(D1):D622–D631. doi:10.1093/nar/gkab1062

22. Tappin DM, Ford RP, Schluter PJ. Smoking during pregnancy measured by population cotinine testing. N Z Med J. 1997;110(1050):311-314.

23. Nenezic N, Kostic S, Strac DS, et al. Dehydroepiandrosterone (DHEA): Pharmacological Effects and Potential Therapeutic Application. Mini Rev Med Chem. 2023;23(8):941–952. doi:10.2174/1389557522666220919125817

24. Baron JA, Comi RJ, Cryns V, Brinck-Johnsen T, Mercer NG. The effect of cigarette smoking on adrenal cortical hormones. J Pharmacol Exp Ther. 1995;272(1):151–155.

25. Cora N, Ghandour J, Pollard CM, et al. Nicotine-induced adrenal beta-arrestin1 upregulation mediates tobacco-related hyperaldosteronism leading to cardiac dysfunction. World J Cardiol. 2020;12(5):192–202. doi:10.4330/wjc.v12.i5.192

26. Yan Y, Guo F, Liu K, Ding R, Wang Y. The effect of endocrine-disrupting chemicals on placental development. Front Endocrinol. 2023;14:1059854. doi:10.3389/fendo.2023.1059854

27. Kenny LC, SCOPE Consortium, Brown LW, Ortea P, Tuytten R, Kell DB. Relationship between the concentration of ergothioneine in plasma and the likelihood of developing pre-eclampsia. Biosci Rep. 2023;43(7):BSR20230160. doi:10.1042/BSR20230160

28. Smith E, Ottosson F, Hellstrand S, et al. Ergothioneine is associated with reduced mortality and decreased risk of cardiovascular disease. Heart Br Card Soc. 2020;106(9):691–697. doi:10.1136/heartjnl-2019-315485

29. Hinds TD, Stec DE. Bilirubin, a Cardiometabolic Signaling Molecule. Hypertens Dallas Tex 1979. 2018;72(4):788-795. doi:10.1161/HYPERTENSIONAHA.118.11130

30. Kodal JB, Çolak Y, Kobylecki CJ, Vedel-Krogh S, Nordestgaard BG, Afzal S. Smoking Reduces Plasma Bilirubin: Observational and Genetic Analyses in the Copenhagen General Population Study. Nicotine Tob Res Off J Soc Res Nicotine Tob. 2020;22(1):104–110. doi:10.1093/ntr/nty188

31. Kim NH, Kim HC, Lee JY, Lee JM, Suh I. Active and Passive Smoking and Serum Total Bilirubin in a Rural Korean Population. Nicotine Tob Res Off J Soc Res Nicotine Tob. 2016;18(5):572–579. doi:10.1093/ntr/ntv251

32. Weaver L, Hamoud AR, Stec DE, Hinds TD. Biliverdin reductase and bilirubin in hepatic disease. Am J Physiol Gastrointest Liver Physiol. 2018;314(6):G668–G676. doi:10.1152/ajpgi.00026.2018

33. Koh M, Ahmad I, Ko Y, et al. A short ORF-encoded transcriptional regulator. Proc Natl Acad Sci U S A. 2021;118(4):e2021943118. doi:10.1073/pnas.2021943118

34. Hamaue N. [Pharmacological role of isatin, an endogenous MAO inhibitor]. Yakugaku Zasshi. 2000;120(4):352–362. doi:10.1248/yakushi1947.120.4_352

35. Buneeva OA, Kapitsa IG, Kazieva LS, Vavilov NE, Zgoda VG. Quantitative changes of brain isatin-binding proteins of rats with the rotenone-induced experimental parkinsonism. Biomeditsinskaia Khimiia. 2023;69(3):188–192. doi:10.18097/PBMC20236903188

36. Bowker K, Lewis S, Coleman T, Cooper S. Changes in the rate of nicotine metabolism across pregnancy: a longitudinal study. Addict Abingdon Engl. 2015;110(11):1827–1832. doi:10.1111/add.13029

37. Qian J, Chen Q, Ward SM, Duan E, Zhang Y. Impacts of Caffeine during Pregnancy. Trends Endocrinol Metab TEM. 2020;31(3):218–227. doi:10.1016/j.tem.2019.11.004

38. Yu T, Campbell SC, Stockmann C, et al. Pregnancy-induced changes in the pharmacokinetics of caffeine and its metabolites. J Clin Pharmacol. 2016;56(5):590–596. doi:10.1002/jcph.632

39. Hey E. Coffee and pregnancy. BMJ. 2007;334(7590):377. doi:10.1136/bmj.39122.395058.80

40. Jin F, Qiao C. Association of maternal caffeine intake during pregnancy with low birth weight, childhood overweight, and obesity: a meta-analysis of cohort studies. Int J Obes 2005. 2021;45(2):279-287. doi:10.1038/s41366-020-0617-4

41. Huffman SL, Harika RK, Eilander A, Osendarp SJM. Essential fats: how do they affect growth and development of infants and young children in developing countries? A literature review. Matern Child Nutr. 2011;7 Suppl 3(Suppl 3):44-65. doi:10.1111/j.1740-8709.2011.00356.x

42. Jääskeläinen T, Kärkkäinen O, Jokkala J, et al. A non-targeted LC-MS metabolic profiling of pregnancy: longitudinal evidence from healthy and pre-eclamptic pregnancies. Metabolomics Off J Metabolomic Soc. 2021;17(2):20. doi:10.1007/s11306-020-01752-5

43. Kitchen S, Adcock DM, Dauer R, et al. International Council for Standardization in Haematology (ICSH) recommendations for processing of blood samples for coagulation testing. Int J Lab Hematol. 2021;43(6):1272–1283. doi:10.1111/ijlh.13702

