## Supplementary Table 1 for "Alterations in the plasma metabolome associated with maternal smoking during the first trimester and fetal growth" (from another version of this preprint: version 1, 2024-11-29)

**Supplemantary table 1**: The non-targeted metabolomics results ordered by the VIP value (p value < 0.0003 are shaded dark green, p value < 0.05 are shaded light green, d values below 0 are blue and above 0 are red)

| | | | | | | | | | Cases | | Controls | | Welch test | Cohen's | |
|---|---|---|---|---|---|---|---|---|---|---|---|---|---|---|---|
| Feature_ID | Metabolite_name | Average_Mz | Average_RT_min_ | HMDB ID | RP_HILIC | POS_NEG | Cureted_MSMS | ID_level | Mean | SD | Mean2 | SD3 | p | d | VIP |
| HILIC_POS_177_1023a0_533 | Cotinine | 177.1023 | 0.53 | HMDB0001046 | HILIC | POS | 177.1013 (100), 80.0466 (10) | 1 | 28845.7 | 22850.5 | 6.8E+07 | 4.5E+07 | 5.45E-27 | 3.00 | 6.11 |
| RP_POS_177_10233a0_682 | Cotinine and serotonin | 177.1023 | 0.68 | | RP | POS | 177.103(100), 80.0506(10) | 2 | 831121 | 394534 | 1.1E+07 | 6995510 | 1.13E-25 | 2.71 | 5.98 |
| HILIC_POS_141_10228a0_547 | | 141.1023 | 0.55 | | HILIC | POS | | | 221214 | 116450 | 995606 | 547561 | 2.14E-25 | 2.33 | 5.85 |
| RP_NEG_183_84825a0_587 | | 183.8483 | 0.59 | | RP | NEG | | | 958361 | 466159 | 2133135 | 835238 | 8.73E-25 | 1.81 | 5.43 |
| RP_NEG_185_84526a0_587 | | 185.8453 | 0.59 | | RP | NEG | | | 629944 | 303724 | 1379763 | 536709 | 1.60E-24 | 1.78 | 5.4 |
| RP_NEG_241_82365a0_586 | | 241.8237 | 0.59 | | RP | NEG | | | 1809358 | 1063442 | 4775203 | 2173767 | 4.32E-24 | 1.83 | 5.44 |
| RP_NEG_243_8203a0_586 | | 243.8203 | 0.59 | | RP | NEG | | | 1252450 | 752695 | 3334526 | 1525631 | 4.38E-24 | 1.83 | 5.43 |
| RP_NEG_245_8172a0_587 | | 245.8172 | 0.59 | | RP | NEG | | | 251140 | 154610 | 673291 | 313207 | 1.08E-23 | 1.8 | 5.39 |
| RP_NEG_199_88109a0_585 | | 199.8811 | 0.59 | | RP | NEG | | | 1297128 | 719814 | 3486266 | 1673892 | 5.80E-23 | 1.83 | 5.38 |
| RP_NEG_230_84334a0_606 | | 230.8433 | 0.61 | | RP | NEG | | | 365480 | 116392 | 583319 | 159709 | 1.57E-22 | 1.58 | 5.08 |
| RP_NEG_228_84586a0_602 | | 228.8459 | 0.60 | | RP | NEG | | | 544021 | 173529 | 868800 | 239932 | 2.45E-22 | 1.57 | 5.06 |
| HILIC_POS_163_12306a1_271 | Nicotine | 163.1231 | 1.27 | HMDB0243542 | HILIC | POS | 163.1221(100), 132.0811(65), 106.0656(20) | 2 | 9157.76 | 43997.4 | 4214689 | 3308104 | 2.55E-22 | 2.51 | 5.63 |
| RP_NEG_264_82974a0_585 | | 264.8297 | 0.59 | | RP | NEG | | | 410852 | 376281 | 2031993 | 1292281 | 5.73E-22 | 1.94 | 5.4 |
| RP_NEG_206_85451a0_586 | | 206.8545 | 0.59 | | RP | NEG | | | 201617 | 165625 | 868244 | 532534 | 6.80E-22 | 1.91 | 5.38 |
| RP_NEG_266_82639a0_586 | | 266.8264 | 0.59 | | RP | NEG | | | 167706 | 155320 | 850866 | 548314 | 8.74E-22 | 1.94 | 5.39 |
| RP_NEG_222_81129a0_589 | | 222.8113 | 0.59 | | RP | NEG | | | 2133612 | 620583 | 3195154 | 788915 | 1.59E-21 | 1.51 | 4.94 |
| RP_NEG_220_81432a0_589 | | 220.8143 | 0.59 | | RP | NEG | | | 6406744 | 1874235 | 9579631 | 2361779 | 2.11E-21 | 1.5 | 4.92 |
| RP_NEG_218_81734a0_589 | | 218.8173 | 0.59 | | RP | NEG | | | 6421254 | 1873992 | 9603161 | 2371872 | 2.16E-21 | 1.5 | 4.93 |
| RP_NEG_176_87492a0_588 | | 176.8749 | 0.59 | | RP | NEG | | | 2279376 | 621981 | 3276987 | 727817 | 2.32E-21 | 1.48 | 4.91 |
| RP_NEG_178_87201a0_588 | | 178.8720 | 0.59 | | RP | NEG | | | 736733 | 203457 | 1064422 | 241182 | 3.29E-21 | 1.47 | 4.89 |
| RP_POS_163_12302a0_68 | | 163.1230 | 0.68 | | RP | POS | | | 45745.6 | 78321.5 | 706085 | 557267 | 1.68E-20 | 2.08 | 5.37 |
| RP_NEG_209_85283a0_589 | | 209.8528 | 0.59 | | RP | NEG | | | 345893 | 248879 | 912330 | 465868 | 1.95E-20 | 1.58 | 5.02 |
| RP_NEG_157_90083a0_585 | | 157.9008 | 0.59 | | RP | NEG | | | 1021564 | 429213 | 1700608 | 562048 | 1.45E-18 | 1.37 | 4.66 |
| HILIC_POS_193_09486a0_651 | | 193.0949 | 0.65 | | HILIC | POS | | | 358308 | 3703037 | 5350940 | 4172014 | 4.40E-17 | 1.27 | 4.41 |
| RP_POS_207_06529a5_931 | Citropten | 207.0653 | 5.93 | HMDB0032952 | RP | POS | 207.0644(100) 179.0618(10) | 2 | 334831 | 325976 | 874327 | 536301 | 3.55E-15 | 1.25 | 4.32 |
| RP_NEG_232_85913a0_587 | | 232.8591 | 0.59 | | RP | NEG | | | 120982 | 112435 | 526778 | 434486 | 6.53E-15 | 1.48 | 4.52 |
| HILIC_POS_96_04498a0_784 | 3-Hydroxypyridine | 96.0450 | 0.78 | HMDB0240652 | HILIC | POS | 96.0449(100), 78.0425(20) | 2 | 1414796 | 1884236 | 4734104 | 3428614 | 1.14E-14 | 1.25 | 4.32 |
| RP_NEG_127_8704a0_624 | | 127.8704 | 0.62 | | RP | NEG | | | 800925 | 161296 | 597201 | 200787 | 6.14E-14 | -1.13 | 4.03 |
| RP_NEG_287_836a0_585 | | 287.8360 | 0.59 | | RP | NEG | | | 70110.2 | 61506.5 | 522592 | 527388 | 1.97E-13 | 1.54 | 4.34 |
| RP_POS_221_08063a6_651 | 3,5-dimethyl-7-oxo-6-oxabicyclo[3.2.1]octane-2-carboxylic acid | 221.0806 | 6.65 | CID: 2827403 | RP | POS | 221.082(100) | 3 | 254180 | 250523 | 577337 | 338793 | 3.84E-13 | 1.10 | 3.98 |
| RP_NEG_188_84357a0_589 | | 188.8436 | 0.59 | | RP | NEG | | | 710260 | 291055 | 1083032 | 420446 | 4.74E-12 | 1.05 | 3.86 |
| RP_POS_181_07207a0_677 | | 181.0721 | 0.68 | | RP | POS | | | 178798 | 196932 | 519181 | 422246 | 1.59E-11 | 1.1 | 3.85 |
| RP_POS_549_40906a11_497 | | 549.4091 | 11.50 | | RP | POS | | | 871689 | 437761 | 530918 | 249362 | 1.75E-11 | -0.99 | 3.46 |
| RP_POS_180_10175a0_682 | | 180.1018 | 0.68 | | RP | POS | | | 530073 | 64454.1 | 610970 | 95923.2 | 2.78E-11 | 1.01 | 3.72 |
| RP_POS_568_42737a11_42 | | 568.4274 | 11.42 | | RP | POS | | | 8090201 | 2841882 | 5865258 | 2119798 | 3.71E-10 | -0.9 | 3.29 |
| RP_POS_269_1243a0_579 | | 269.1243 | 0.58 | | RP | POS | | | 258937 | 319488 | 669265 | 537895 | 4.57E-10 | 0.96 | 3.53 |
| RP_NEG_215_00214a4_053 | | 215.0021 | 4.05 | | RP | NEG | | | 849287 | 2287590 | 3093138 | 2683257 | 5.13E-10 | 0.9 | 3.43 |
| HILIC_POS_143_1178a1_508 | | 143.1178 | 1.51 | | HILIC | POS | | | 3011180 | 785595 | 2413589 | 569895 | 7.18E-10 | -0.88 | 3.2 |
| RP_NEG_181_03693a0_687 | | 181.0369 | 0.69 | | RP | NEG | | | 160164 | 137367 | 365752 | 284180 | 9.48E-10 | 0.98 | 3.54 |
| RP_POS_148_03944a3_886 | 1H-Indole-2,3-dione (Isatin) | 148.0394 | 3.89 | HMDB0061933 | RP | POS | 148.0391(100) | 2 | 3953911 | 1816521 | 2554355 | 1429094 | 1.40E-09 | -0.86 | 3.19 |
| RP_POS_176_12816a0_594 | | 176.1282 | 0.59 | | RP | POS | | | 521010 | 277125 | 800983 | 354664 | 1.41E-09 | 0.89 | 3.33 |
| RP_NEG_225_0629a0_681 | | 225.0629 | 0.68 | | RP | NEG | | | 282691 | 154623 | 530280 | 352789 | 1.78E-09 | 0.98 | 3.48 |
| RP_NEG_203_00221a3_498 | | 203.0022 | 3.50 | | RP | NEG | | | 568580 | 929539 | 1639864 | 1447561 | 2.20E-09 | 0.9 | 3.39 |
| RP_POS_193_09723a0_679 | | 193.0972 | 0.68 | | RP | POS | | | 1201748 | 477283 | 2148747 | 1389966 | 2.42E-09 | 1.01 | 3.53 |
| RP_POS_182_11765a7_507 | (S)-4-methyl-2-(1H-pyrrol-1-yl)pentanoic acid | 182.1177 | 7.51 | CID: 15629024 | RP | POS | 136.112(100),182.1167(60) | 2 | 702261 | 339134 | 1016720 | 390737 | 2.42E-09 | 0.86 | 3.25 |
| RP_POS_177_10226a1_334 | | 177.1023 | 1.33 | | RP | POS | | | 1339147 | 553365 | 2465028 | 1657781 | 2.53E-09 | 1.02 | 3.58 |
| RP_POS_181_07204a3_06 | Paraxanthine | 181.0720 | 3.06 | HMDB0001860 | RP | POS | 181.0722(100), 124.0508(30) | 1 | 1.3E+07 | 1.5E+07 | 3.5E+07 | 3.2E+07 | 2.75E-09 | 0.95 | 3.45 |
| RP_NEG_180_10289a7_506 | | 180.1029 | 7.51 | | RP | NEG | | | 219308 | 108619 | 318916 | 124389 | 3.07E-09 | 0.85 | 3.22 |
| RP_POS_181_07205a2_721 | | 181.0721 | 2.72 | | RP | POS | 181.0721(100), 124.0508(30) | | 1.2E+07 | 1.4E+07 | 3.2E+07 | 2.9E+07 | 3.43E-09 | 0.95 | 3.42 |
| HILIC_POS_199_10768a0_862 | | 199.1077 | 0.86 | | HILIC | POS | | | 231846 | 253214 | 612617 | 577548 | 1.23E-08 | 0.92 | 3.35 |
| RP_POS_138_05505a0_564 | | 138.0551 | 0.56 | | RP | POS | | | 1.1E+07 | 1.1E+07 | 2.9E+07 | 2.7E+07 | 1.28E-08 | 0.93 | 3.34 |
| RP_POS_551_42474a11_42 | | 551.4247 | 11.42 | | RP | POS | | | 1984689 | 1053647 | 1321428 | 537324 | 1.35E-08 | -0.83 | 2.96 |
| RP_NEG_369_17413a6_866 | | 369.1741 | 6.87 | | RP | NEG | | | 3137635 | 1710510 | 4834788 | 2446639 | 3.06E-08 | 0.82 | 3.11 |
| RP_NEG_191_05629a0_581 | | 191.0563 | 0.58 | | RP | NEG | | | 458983 | 668844 | 1157121 | 1025283 | 3.19E-08 | 0.82 | 3.15 |
| RP_NEG_197_06792a0_679 | | 197.0679 | 0.68 | | RP | NEG | | | 312265 | 247096 | 578099 | 394561 | 3.33E-08 | 0.83 | 3.13 |
| RP_NEG_186_84702a0_589 | | 186.8470 | 0.59 | | RP | NEG | | | 1104731 | 466533 | 1550147 | 638118 | 3.62E-08 | 0.81 | 3.12 |
| HILIC_POS_138_05505a4_206 | Trigonelline | 138.0551 | 4.21 | HMDB0000875 | HILIC | POS | 138.0549(100), 94.0656(3) | 1 | 7.2E+07 | 1.1E+08 | 2.5E+08 | 2.9E+08 | 4.07E-08 | 0.91 | 3.28 |
| RP_POS_160_03694a0_567 | | 160.0369 | 0.57 | | RP | POS | | | 796697 | 368743 | 1227398 | 666443 | 5.47E-08 | 0.83 | 3.15 |
| HILIC_POS_181_07201a0_588 | | 181.0720 | 0.59 | | HILIC | POS | | | 670517 | 622913 | 1508715 | 1336750 | 6.24E-08 | 0.86 | 3.15 |
| RP_NEG_217_01775a4_488 | | 217.0178 | 4.49 | | RP | NEG | | | 441708 | 697255 | 1036028 | 840260 | 7.61E-08 | 0.77 | 3 |
| HILIC_POS_160_03696a4_217 | | 160.0370 | 4.22 | | HILIC | POS | | | 984358 | 1482224 | 3686100 | 4482379 | 7.77E-08 | 0.91 | 3.2 |
| RP_POS_445_2948a10_167 | | 445.2948 | 10.17 | | RP | POS | | | 843011 | 1124242 | 241365 | 300741 | 1.40E-07 | -0.84 | 2.71 |
| HILIC_NEG_225_06316a0_757 | | 225.0632 | 0.76 | | HILIC | NEG | | | 75152.7 | 78594.1 | 207802 | 225561 | 1.61E-07 | 0.87 | 3.11 |
| RP_NEG_207_83688a0_609 | | 207.8369 | 0.61 | | RP | NEG | | | 1953195 | 777099 | 1413250 | 696108 | 1.91E-07 | -0.73 | 2.92 |
| RP_POS_255_21049a7_353 | | 255.2105 | 7.35 | | RP | POS | | | 629926 | 343670 | 918715 | 430737 | 2.24E-07 | 0.75 | 2.88 |
| RP_NEG_413_20065a7_351 | | 413.2007 | 7.35 | | RP | NEG | | | 7096577 | 3735024 | 1E+07 | 4540223 | 2.53E-07 | 0.74 | 2.87 |
| RP_NEG_160_84206a0_588 | | 160.8421 | 0.59 | | RP | NEG | | | 4463818 | 1068255 | 5074729 | 568216 | 2.55E-07 | 0.75 | 2.73 |
| RP_NEG_201_80208a0_544 | | 201.8021 | 0.54 | | RP | NEG | | | 9965276 | 1393953 | 8985456 | 1308586 | 2.62E-07 | -0.73 | 2.78 |
| RP_POS_191_07031a5_819 | | 191.0703 | 5.82 | | RP | POS | 191.0712(100), 148.0527(15) | 4 | 326978 | 609322 | 944563 | 997280 | 2.78E-07 | 0.77 | 2.98 |
| HILIC_POS_166_08604a4_41 | | 166.0860 | 4.41 | | HILIC | POS | 217.1044(100) | | 1270848 | 1469164 | 2922876 | 2737329 | 2.88E-07 | 0.79 | 3.12 |
| RP_POS_281_22623a7_345 | | 281.2262 | 7.35 | | RP | POS | | | 352331 | 194590 | 510267 | 235864 | 3.08E-07 | 0.73 | 2.85 |
| RP_POS_195_11276a2_912 | | 195.1128 | 2.91 | | RP | POS | | | 1036963 | 524572 | 1452127 | 616255 | 3.49E-07 | 0.73 | 2.8 |
| RP_NEG_445_33289a11_122 | | 445.3329 | 11.12 | | RP | NEG | | | 428984 | 230520 | 651109 | 358183 | 3.57E-07 | 0.75 | 2.9 |
| RP_POS_565_40326a11_535 | | 565.4033 | 11.54 | | RP | POS | | | 787191 | 322534 | 587739 | 235933 | 3.99E-07 | -0.71 | 2.71 |
| RP_NEG_229_01776a4_529 | | 229.0178 | 4.53 | | RP | NEG | | | 263967 | 483952 | 658913 | 618339 | 6.43E-07 | 0.72 | 2.81 |
| RP_POS_568_42712a12_032 | | 568.4271 | 12.03 | | RP | POS | | | 975777 | 532672 | 654723 | 384923 | 6.70E-07 | -0.7 | 2.66 |
| RP_NEG_205_84006a0_611 | | 205.8401 | 0.61 | | RP | NEG | | | 1942040 | 825297 | 1395107 | 746366 | 7.09E-07 | -0.7 | 2.79 |
| RP_NEG_467_08374a7_254 | | 467.0837 | 7.25 | | RP | NEG | | | 280405 | 137882 | 384866 | 159254 | 7.81E-07 | 0.7 | 2.77 |
| RP_NEG_463_1777a8_188 | | 463.1777 | 8.19 | | RP | NEG | | | 262266 | 129119 | 363246 | 157109 | 8.14E-07 | 0.71 | 2.77 |
| RP_NEG_399_20132a7_901 | | 399.2013 | 7.90 | | RP | NEG | | | 839887 | 360997 | 1138731 | 478685 | 8.41E-07 | 0.71 | 2.77 |
| RP_POS_399_32556a10_738 | | 399.3256 | 10.74 | | RP | POS | | | 271528 | 113778 | 363541 | 145972 | 8.53E-07 | 0.71 | 2.95 |
| RP_NEG_516_08673a7_262 | | 516.0867 | 7.26 | | RP | NEG | | | 547585 | 244236 | 730936 | 280221 | 8.79E-07 | 0.7 | 2.76 |
| RP_NEG_395_18985a8_188 | | 395.1899 | 8.19 | | RP | NEG | | | 7384438 | 3357342 | 1E+07 | 4276086 | 8.91E-07 | 0.71 | 2.75 |
| RP_NEG_465_19421a7_9 | | 465.1942 | 7.90 | | RP | NEG | | | 713326 | 311417 | 960918 | 391775 | 9.31E-07 | 0.7 | 2.74 |
| HILIC_POS_72_08162a1_383 | | 72.0816 | 1.38 | | HILIC | POS | | | 3096903 | 1467787 | 2298630 | 802033 | 1.02E-06 | -0.7 | 2.56 |
| RP_NEG_397_20578a7_898 | | 397.2058 | 7.90 | | RP | NEG | | | 2.3E+07 | 9990746 | 3.1E+07 | 1.3E+07 | 1.08E-06 | 0.7 | 2.73 |
| RP_NEG_515_08917a7_265 | | 515.0892 | 7.27 | | RP | NEG | | | 512511 | 229360 | 682659 | 261823 | 1.08E-06 | 0.69 | 2.77 |
| HILIC_POS_176_01099a4_238 | | 176.0110 | 4.24 | | HILIC | POS | | | 457953 | 763826 | 1533947 | 1973069 | 1.19E-06 | 0.79 | 2.89 |
| RP_NEG_735_3252a7_259 | | 735.3252 | 7.26 | | RP | NEG | | | 431705 | 364028 | 743377 | 517559 | 1.22E-06 | 0.71 | 2.78 |
| RP_NEG_481_18811a7_36 | | 481.1881 | 7.36 | | RP | NEG | | | 219227 | 117059 | 311184 | 148330 | 1.38E-06 | 0.69 | 2.72 |
| RP_NEG_310_83868a6_949 | | 310.8387 | 6.95 | | RP | NEG | | | 180767 | 132590 | 254536 | 84519.3 | 1.60E-06 | 0.68 | 2.64 |
| RP_NEG_425_11646a7_252 | | 425.1165 | 7.25 | | RP | NEG | | | 273215 | 101502 | 345528 | 111798 | 1.65E-06 | 0.68 | 2.68 |
| RP_POS_271_20569a7_257 | | 271.2057 | 7.26 | | RP | POS | | | 3877276 | 2006446 | 5355776 | 2335492 | 1.66E-06 | 0.68 | 2.7 |
| RP_NEG_367_15857a7_264 | Dehydroepiandrosterone sulfate (DHEA-S) | 367.1586 | 7.26 | HMDB0001032 | RP | NEG | 96.9597(100), 367.1557(95) | 2 | 1.3E+08 | 6.2E+07 | 1.7E+08 | 7.2E+07 | 1.74E-06 | 0.68 | 2.7 |
| RP_NEG_125_87311a0_633 | | 125.8731 | 0.63 | | RP | NEG | | | 660547 | 237975 | 520234 | 181455 | 1.82E-06 | -0.67 | 2.58 |
| HILIC_NEG_224_06226a1_221 | | 224.0623 | 1.22 | | HILIC | NEG | | | 251346 | 148675 | 386345 | 235126 | 1.97E-06 | 0.7 | 2.73 |
| RP_NEG_435_14606a7_264 | | 435.1461 | 7.26 | | RP | NEG | | | 3635163 | 1508733 | 4692607 | 1639334 | 1.98E-06 | 0.67 | 2.67 |
| RP_NEG_369_15408a7_264 | | 369.1541 | 7.26 | | RP | NEG | | | 5006159 | 2444983 | 6782038 | 2825162 | 2.07E-06 | 0.67 | 2.69 |
| RP_NEG_203_00218a2_809 | | 203.0022 | 2.81 | | RP | NEG | | | 470616 | 386413 | 751844 | 450135 | 2.22E-06 | 0.67 | 2.65 |
| RP_NEG_757_30652a7_264 | | 757.3065 | 7.26 | | RP | NEG | | | 553227 | 406566 | 868764 | 523458 | 2.30E-06 | 0.68 | 2.71 |
| RP_NEG_836_54565a12_7 | | 836.5457 | 12.70 | | RP | NEG | | | 2064940 | 670773 | 1676128 | 503766 | 2.32E-06 | -0.66 | 2.63 |
| RP_NEG_224_06216a5_45 | | 224.0622 | 5.45 | | RP | NEG | | | 3821496 | 2249448 | 6024703 | 3979786 | 2.62E-06 | 0.71 | 2.71 |
| RP_NEG_513_08881a7_276 | | 513.0888 | 7.28 | | RP | NEG | | | 266774 | 132664 | 359528 | 147064 | 2.72E-06 | 0.66 | 2.67 |
| RP_POS_293_18796a7_265 | | 293.1880 | 7.27 | | RP | POS | | | 569192 | 300667 | 779173 | 341005 | 3.76E-06 | 0.65 | 2.64 |
| RP_POS_486_13672a7_264 | | 486.1367 | 7.26 | | RP | POS | | | 411912 | 197005 | 546144 | 215583 | 4.00E-06 | 0.65 | 2.6 |
| HILIC_POS_287_24426a5_973 | | 287.2443 | 5.97 | | HILIC | POS | | | 600352 | 405230 | 363021 | 331537 | 4.01E-06 | -0.64 | 2.49 |
| HILIC_NEG_413_20108a0_611 | | 413.2011 | 0.61 | | HILIC | NEG | | | 120008 | 54712.5 | 159095 | 65255.9 | 4.55E-06 | 0.65 | 2.59 |
| HILIC_POS_110_06046a1_459 | Aminophenol | 110.0605 | 1.46 | HMDB0001169 | HILIC | POS | 110.0607(100) | 2 | 944370 | 1974695 | 2474837 | 2663726 | 4.70E-06 | 0.66 | 2.62 |
| RP_POS_301_25208a7_906 | | 301.2521 | 7.91 | | RP | POS | | | 619895 | 358842 | 870693 | 425480 | 6.60E-06 | 0.64 | 2.56 |
| RP_POS_408_16953a3_804 | | 408.1695 | 3.80 | | RP | POS | | | 643497 | 600040 | 1168079 | 974391 | 6.67E-06 | 0.67 | 2.61 |
| RP_POS_138_06618a3_805 | | 138.0662 | 3.81 | | RP | POS | | | 390138 | 462473 | 802259 | 775156 | 7.48E-06 | 0.67 | 2.6 |
| RP_POS_195_08768a3_805 | Caffeine | 195.0877 | 3.81 | HMDB0001847 | RP | POS | 195.0868(100), 138.0668(40) | 1 | 1.3E+08 | 1.6E+08 | 2.7E+08 | 2.6E+08 | 7.93E-06 | 0.66 | 2.59 |
| RP_NEG_503_13358a7_263 | | 503.1336 | 7.26 | | RP | NEG | | | 356083 | 141141 | 445107 | 144398 | 8.70E-06 | 0.62 | 2.52 |
| RP_POS_195_08762a3_696 | | 195.0876 | 3.70 | | RP | POS | 195.087(100), 138.0669(35) | | 7.7E+07 | 9.7E+07 | 1.7E+08 | 1.7E+08 | 8.72E-06 | 0.67 | 2.59 |
| RP_POS_469_32907a11_133 | | 469.3291 | 11.13 | | RP | POS | | | 966636 | 511060 | 1380480 | 774588 | 1.06E-05 | 0.64 | 2.54 |
| RP_POS_447_31049a10_236 | | 447.3105 | 10.24 | | RP | POS | | | 963435 | 1367283 | 340742 | 554095 | 1.27E-05 | -0.65 | 2.33 |
| RP_POS_503_19843a7_266 | | 503.1984 | 7.27 | | RP | POS | | | 304230 | 172535 | 409364 | 172481 | 1.32E-05 | 0.61 | 2.46 |
| RP_NEG_304_73672a0_641 | | 304.7367 | 0.64 | | RP | NEG | | | 300074 | 70943.7 | 263311 | 49698.4 | 1.35E-05 | -0.61 | 2.39 |
| RP_NEG_383_15402a6_438 | | 383.1540 | 6.44 | | RP | NEG | | | 418014 | 193030 | 553549 | 243995 | 1.37E-05 | 0.62 | 2.41 |
| RP_NEG_325_89981a0_588 | | 325.8998 | 0.59 | | RP | NEG | | | 726496 | 305008 | 539424 | 309937 | 1.39E-05 | -0.61 | 2.4 |
| RP_NEG_173_98631a0_677 | | 173.9863 | 0.68 | | RP | NEG | | | 1159853 | 2395753 | 2905716 | 3227168 | 1.54E-05 | 0.62 | 2.47 |
| RP_NEG_705_52124a11_992 | | 705.5212 | 11.99 | | RP | NEG | | | 521327 | 146602 | 442610 | 114746 | 1.55E-05 | -0.6 | 2.32 |
| RP_POS_350_2066a4_164 | | 350.2066 | 4.16 | | RP | POS | | | 365385 | 251935 | 242231 | 150977 | 1.57E-05 | -0.61 | 2.29 |
| HILIC_POS_256_11777a1_803 | | 256.1178 | 1.80 | | HILIC | POS | | | 269165 | 583217 | 744807 | 935890 | 2.09E-05 | 0.63 | 2.51 |
| RP_NEG_323_9028a0_589 | | 323.9028 | 0.59 | | RP | NEG | | | 753412 | 328324 | 558021 | 330882 | 2.23E-05 | -0.59 | 2.34 |
| RP_POS_299_23615a8_209 | | 299.2362 | 8.21 | | RP | POS | | | 375895 | 178457 | 544456 | 348434 | 2.55E-05 | 0.64 | 2.46 |
| RP_NEG_397_18542a8_191 | | 397.1854 | 8.19 | | RP | NEG | | | 258498 | 138345 | 352413 | 175745 | 2.69E-05 | 0.6 | 2.4 |
| RP_POS_132_04446a3_235 | | 132.0445 | 3.24 | | RP | POS | | | 371578 | 206612 | 262583 | 167659 | 2.83E-05 | -0.58 | 2.29 |
| RP_NEG_427_1796a6_89 | | 427.1796 | 6.89 | | RP | NEG | | | 180852 | 63910.4 | 220991 | 72292.7 | 2.84E-05 | 0.59 | 2.38 |
| RP_NEG_209_83409a0_631 | | 209.8341 | 0.63 | | RP | NEG | | | 545800 | 248575 | 420368 | 183784 | 3.16E-05 | -0.58 | 2.25 |
| RP_POS_494_32425a10_108 | LysoPC 16:1 (16:1/0:0) | 494.3243 | 10.11 | HMDB0010383 | RP | POS | 184.0745(100), 104.1078(70) | 1 | 8E+07 | 3.4E+07 | 1E+08 | 4.3E+07 | 3.24E-05 | 0.59 | 2.35 |
| RP_POS_230_09575a0_588 | | 230.0958 | 0.59 | | RP | POS | | | 985184 | 889499 | 589026 | 429727 | 3.28E-05 | -0.6 | 2.18 |
| HILIC_NEG_225_06189a6_263 | | 225.0619 | 6.26 | | HILIC | NEG | | | 149196 | 38249.4 | 131039 | 23982.9 | 3.39E-05 | -0.58 | 2.2 |
| HILIC_POS_130_09769a1_533 | | 130.0977 | 1.53 | | HILIC | POS | | | 1642578 | 410279 | 2100108 | 1000693 | 4.03E-05 | 0.65 | 2.45 |
| HILIC_POS_167_09274a1_147 | | 167.0927 | 1.15 | | HILIC | POS | | | 719953 | 247741 | 590101 | 207424 | 4.03E-05 | -0.57 | 2.22 |
| HILIC_POS_86_0607a0_836 | | 86.0607 | 0.84 | | HILIC | POS | | | 1847610 | 462013 | 1619640 | 334568 | 4.05E-05 | -0.57 | 2.18 |
| RP_NEG_310_9325a0_588 | | 310.9325 | 0.59 | | RP | NEG | | | 321616 | 131235 | 407001 | 163371 | 4.43E-05 | 0.58 | 2.31 |
| RP_NEG_537_41699a11_031 | | 537.4170 | 11.03 | | RP | NEG | | | 594824 | 330096 | 804286 | 395672 | 4.46E-05 | 0.58 | 2.33 |
| RP_NEG_606_30304a10_109 | | 606.3030 | 10.11 | | RP | NEG | | | 1902022 | 817085 | 2382838 | 872651 | 4.81E-05 | 0.57 | 2.31 |
| RP_NEG_674_29047a10_109 | | 674.2905 | 10.11 | | RP | NEG | | | 501590 | 182311 | 611521 | 202971 | 4.84E-05 | 0.57 | 2.3 |
| HILIC_NEG_255_08789a0_614 | | 255.0879 | 0.61 | | HILIC | NEG | | | 209647 | 195781 | 116308 | 133283 | 4.97E-05 | -0.57 | 2.32 |
| RP_POS_447_34662a11_131 | | 447.3466 | 11.13 | | RP | POS | | | 595696 | 228729 | 756930 | 324153 | 5.19E-05 | 0.58 | 2.37 |
| RP_NEG_627_37604a9_067 | | 627.3760 | 9.07 | | RP | NEG | | | 609622 | 299224 | 765894 | 257670 | 5.33E-05 | 0.56 | 2.21 |
| RP_NEG_188_00214a0_681 | | 188.0021 | 0.68 | | RP | NEG | | | 286323 | 472826 | 622579 | 688234 | 6.17E-05 | 0.58 | 2.3 |
| HILIC_NEG_173_98669a0_569 | | 173.9867 | 0.57 | | HILIC | NEG | | | 410800 | 900589 | 1001576 | 1174599 | 6.48E-05 | 0.57 | 2.3 |
| HILIC_NEG_211_9774a1_218 | | 211.9774 | 1.22 | | HILIC | NEG | | | 292593 | 65880.1 | 261380 | 46516 | 6.81E-05 | -0.56 | 2.13 |
| RP_NEG_478_29437a10_108 | | 478.2944 | 10.11 | | RP | NEG | | | 834893 | 338232 | 1040060 | 397758 | 7.63E-05 | 0.56 | 2.26 |
| RP_NEG_538_31531a10_109 | LysoPC 16:1 (16:1/0:0) | 538.3153 | 10.11 | HMDB0010383 | RP | NEG | 253.2189(100), 478.296(15) | 1 | 2.3E+07 | 1E+07 | 3E+07 | 1.2E+07 | 7.71E-05 | 0.56 | 2.26 |
| RP_NEG_822_53101a12_425 | | 822.5310 | 12.43 | | RP | NEG | | | 3297651 | 1357528 | 2634554 | 1058348 | 7.75E-05 | -0.55 | 2.15 |
| HILIC_POS_84_91185a2_802 | | 84.9119 | 2.80 | | HILIC | POS | | | 6348480 | 1389674 | 7163174 | 1550266 | 7.81E-05 | 0.55 | 2.23 |
| RP_NEG_850_56201a13_005 | PC 38:6 (16:0_22:6) | 850.5620 | 13.01 | HMDB0013409 | RP | NEG | 255.2328(100), 327.2315(65), 790.5338(10) | 2 | 1E+08 | 1.7E+07 | 9.5E+07 | 1.6E+07 | 8.01E-05 | -0.55 | 2.24 |
| HILIC_POS_150_05843a0_837 | | 150.0584 | 0.84 | | HILIC | POS | | | 3825920 | 1245374 | 3245693 | 869538 | 8.34E-05 | -0.55 | 2.09 |
| RP_POS_719_42383a10_583 | | 719.4238 | 10.58 | | RP | POS | | | 1231804 | 1054255 | 767282 | 628557 | 9.03E-05 | -0.55 | 2.14 |
| RP_NEG_211_09767a5_549 | | 211.0977 | 5.55 | | RP | NEG | | | 369180 | 327785 | 216749 | 231206 | 9.10E-05 | -0.55 | 2.27 |
| RP_NEG_517_24261a10_04 | | 517.2426 | 10.04 | | RP | NEG | | | 406191 | 220630 | 561055 | 329497 | 1.02E-04 | 0.56 | 2.24 |
| RP_NEG_302_73978a0_64 | | 302.7398 | 0.64 | | RP | NEG | | | 235977 | 61053.5 | 208653 | 39557.6 | 1.05E-04 | -0.54 | 2.17 |
| RP_POS_792_5531a12_713 | | 792.5531 | 12.71 | | RP | POS | | | 1.4E+07 | 5505465 | 1.2E+07 | 4040911 | 1.06E-04 | -0.54 | 2.18 |
| RP_POS_303_23169a7_268 | | 303.2317 | 7.27 | | RP | POS | | | 321075 | 195816 | 435612 | 225387 | 1.09E-04 | 0.54 | 2.24 |
| RP_NEG_224_06201a6_03 | | 224.0620 | 6.03 | | RP | NEG | | | 7489931 | 2765823 | 9173621 | 3395830 | 1.11E-04 | 0.55 | 2.26 |
| RP_POS_714_46844a10_586 | | 714.4684 | 10.59 | | RP | POS | | | 932111 | 828205 | 570119 | 505560 | 1.14E-04 | -0.54 | 2.14 |
| HILIC_POS_86_90932a2_802 | | 86.9093 | 2.80 | | HILIC | POS | | | 2365626 | 518019 | 2660722 | 577166 | 1.20E-04 | 0.54 | 2.17 |

**Supplemantary table 1**: The non-targeted metabolomics results ordered by the VIP value (p value < 0.0003 are shaded dark green, p value < 0.05 are shaded light green, d values below 0 are blue and above 0 are red)

| | | | | | | | | | Cases | | Controls | | Welch test | Cohen's | |
|---|---|---|---|---|---|---|---|---|---|---|---|---|---|---|---|
| Feature_ID | Metabolite_name | Average_Mz | Average_RT_min_ | HMDB ID | RP_HILIC | POS_NEG | Cureted_MSMS | ID_level | Mean | SD | Mean2 | SD3 | p | d | VIP |
| RP_NEG_289_92322a0_589 | | 289.9232 | 0.59 | | RP | NEG | | | 959694 | 361993 | 758853 | 386262 | 1.22E-04 | -0.54 | 2.14 |
| HILIC_POS_186_11256a3_975 | | 186.1126 | 3.98 | | HILIC | POS | | | 2924966 | 1087455 | 3486604 | 1030191 | 1.33E-04 | 0.53 | 2.16 |
| RP_POS_516_30603a10_108 | | 516.3060 | 10.11 | | RP | POS | | | 9408374 | 4259356 | 1.2E+07 | 5745345 | 1.34E-04 | 0.55 | 2.2 |
| RP_NEG_255_08754a5_548 | | 255.0875 | 5.55 | | RP | NEG | | | 3236005 | 3086106 | 1825872 | 2231060 | 1.37E-04 | -0.53 | 2.23 |
| RP_POS_181_07205a3_195 | Theophylline | 181.0721 | 3.20 | HMDB0001889 | RP | POS | 181.0723(100), 124.0509(25) | 2 | 3295919 | 3474960 | 5797991 | 5514959 | 1.38E-04 | 0.56 | 2.21 |
| RP_POS_239_16408a9_668 | | 239.1641 | 9.67 | | RP | POS | | | 5098644 | 1870096 | 4256407 | 1303577 | 1.39E-04 | -0.53 | 2.1 |
| RP_POS_415_32043a9_835 | | 415.3204 | 9.84 | | RP | POS | | | 837355 | 295943 | 1011070 | 353326 | 1.46E-04 | 0.54 | 2.21 |
| HILIC_POS_387_21417a4_281 | | 387.2142 | 4.28 | | HILIC | POS | | | 1149400 | 276481 | 994374 | 307261 | 1.49E-04 | -0.53 | 2.11 |
| HILIC_POS_585_27075a0_621 | Bilirubin | 585.2708 | 0.62 | HMDB0000054 | HILIC | POS | 299.1379(100),297.1262(5) | 2 | 2737898 | 1680200 | 2012317 | 1061473 | 1.59E-04 | -0.53 | 2.01 |
| RP_NEG_287_92621a0_589 | | 287.9262 | 0.59 | | RP | NEG | | | 1574608 | 599403 | 1251387 | 636548 | 1.78E-04 | -0.52 | 2.09 |
| HILIC_NEG_179_05762a0_561 | | 179.0576 | 0.56 | | HILIC | NEG | | | 1370719 | 1403209 | 2325576 | 2118292 | 1.82E-04 | 0.54 | 2.18 |
| HILIC_POS_94_06578a1_194 | Aniline | 94.0658 | 1.19 | HMDB0003012 | HILIC | POS | 94.0655(100) | 2 | 1759634 | 3875588 | 3997175 | 4629757 | 1.87E-04 | 0.53 | 2.16 |
| RP_NEG_431_31699a9_833 | | 431.3170 | 9.83 | | RP | NEG | | | 744927 | 262173 | 894463 | 307807 | 1.90E-04 | 0.52 | 2.18 |
| HILIC_NEG_224_06203a1_331 | | 224.0620 | 1.33 | | HILIC | NEG | | | 415695 | 154805 | 505214 | 185977 | 1.93E-04 | 0.53 | 2.2 |
| RP_POS_239_09131a5_549 | | 239.0913 | 5.55 | | RP | POS | | | 7077900 | 6706221 | 4076862 | 4906819 | 1.98E-04 | -0.52 | 2.19 |
| RP_NEG_610_24603a10_108 | | 610.2460 | 10.11 | | RP | NEG | | | 439370 | 189279 | 546665 | 221408 | 2.01E-04 | 0.52 | 2.15 |
| HILIC_POS_230_09567a5_411 | Ergothioneine | 230.0957 | 5.41 | HMDB0003045 | HILIC | POS | 127.0326(100), 186.1073(40) | 1 | 342027 | 351057 | 204817 | 157346 | 2.02E-04 | -0.54 | 1.95 |
| RP_NEG_413_20078a7_784 | | 413.2008 | 7.78 | | RP | NEG | | | 811745 | 433993 | 617828 | 320172 | 2.12E-04 | -0.51 | 2.02 |
| RP_POS_521_4198a11_032 | | 521.4198 | 11.03 | | RP | POS | | | 918924 | 515389 | 1203616 | 581988 | 2.13E-04 | 0.52 | 2.13 |
| RP_NEG_179_0576a2_851 | | 179.0576 | 2.85 | | RP | NEG | | | 729085 | 795739 | 1266110 | 1217874 | 2.36E-04 | 0.53 | 2.13 |
| RP_NEG_179_05762a3_197 | Theophylline | 179.0576 | 3.20 | HMDB0001889 | RP | NEG | 179.0578(100), 134.9884(20) | 2 | 876526 | 954670 | 1536294 | 1508052 | 2.36E-04 | 0.54 | 2.14 |
| RP_NEG_449_25531a10_04 | | 449.2553 | 10.04 | | RP | NEG | | | 4472938 | 2414782 | 6060682 | 3580795 | 2.44E-04 | 0.53 | 2.13 |
| RP_POS_279_08383a5_549 | | 279.0838 | 5.55 | | RP | POS | | | 1000089 | 990002 | 557981 | 748795 | 2.45E-04 | -0.51 | 2.17 |
| RP_POS_817_5589a13_008 | | 817.5589 | 13.01 | | RP | POS | | | 1.1E+07 | 3596527 | 8992374 | 3218536 | 2.47E-04 | -0.51 | 2.01 |
| HILIC_NEG_115_92093a0_559 | | 115.9209 | 0.56 | | HILIC | NEG | | | 6482022 | 1314423 | 5871036 | 1097271 | 2.51E-04 | -0.51 | 2.05 |
| RP_POS_257_2262a10_043 | | 257.2262 | 10.04 | | RP | POS | | | 2282004 | 1214535 | 3073879 | 1787729 | 2.55E-04 | 0.53 | 2.12 |
| RP_POS_494_3241a9_964 | LysoPC 16:1 (0:0/16:1) | 494.3241 | 9.96 | HMDB0010383 | RP | POS | 184.0744(100) | 1 | 9833737 | 4312197 | 1.2E+07 | 4887017 | 2.62E-04 | 0.51 | 2.07 |
| RP_NEG_538_31543a9_963 | | 538.3154 | 9.96 | | RP | NEG | | | 2852949 | 1179496 | 3519067 | 1415359 | 2.65E-04 | 0.51 | 2.12 |
| RP_POS_130_08675a0_676 | Pipecolic acid | 130.0868 | 0.68 | HMDB0000070 | RP | POS | 84.0814(100), 130.0867(45) | 2 | 8832345 | 2365828 | 7735700 | 1988261 | 2.73E-04 | -0.50 | 2.03 |
| RP_NEG_583_2569a11_845 | Bilirubin | 583.2569 | 11.85 | HMDB0000054 | RP | NEG | 285.1228(100), 241.1345(10), 213.1042(10) | 2 | 4598136 | 2864070 | 3324538 | 2190412 | 2.76E-04 | -0.50 | 2 |
| RP_POS_566_41132a11_521 | | 566.4113 | 11.52 | | RP | POS | | | 1052236 | 611284 | 786658 | 442801 | 2.84E-04 | -0.5 | 2 |
| RP_POS_607_25256a11_849 | | 607.2526 | 11.85 | | RP | POS | | | 7955252 | 4202002 | 6051480 | 3388490 | 2.90E-04 | -0.5 | 1.98 |
| RP_NEG_528_28711a10_109 | | 528.2871 | 10.11 | | RP | NEG | | | 719975 | 294078 | 877912 | 331254 | 3.07E-04 | 0.51 | 2.07 |
| RP_POS_494_22861a11_851 | | 494.2286 | 11.85 | | RP | POS | | | 1237539 | 665052 | 933354 | 553240 | 3.07E-04 | -0.5 | 1.99 |
| RP_POS_283_24246a7_894 | | 283.2425 | 7.89 | | RP | POS | | | 1405281 | 681919 | 2057027 | 1639021 | 3.26E-04 | 0.56 | 2.14 |
| RP_POS_814_53595a12_704 | | 814.5360 | 12.70 | | RP | POS | | | 3629568 | 1536558 | 2974090 | 1089632 | 3.26E-04 | -0.5 | 2.02 |
| RP_NEG_415_21652a8_066 | | 415.2165 | 8.07 | | RP | NEG | | | 434902 | 301485 | 302798 | 229396 | 3.28E-04 | -0.5 | 1.97 |
| RP_POS_516_30621a9_947 | | 516.3062 | 9.95 | | RP | POS | | | 941834 | 451319 | 1183850 | 512791 | 3.36E-04 | 0.5 | 2.07 |
| RP_POS_299_13831a11_849 | | 299.1383 | 11.85 | | RP | POS | | | 5729155 | 3972651 | 4036588 | 2825019 | 3.37E-04 | -0.5 | 1.96 |
| RP_POS_473_25098a10_044 | | 473.2510 | 10.04 | | RP | POS | | | 1765599 | 939051 | 2377016 | 1419370 | 3.38E-04 | 0.52 | 2.1 |
| HILIC_POS_220_11917a1_594 | Zeatin | 220.1192 | 1.59 | HMDB0012204 | HILIC | POS | 220.1182(100), 202.1074(20) | 2 | 198544 | 262961 | 466005 | 680187 | 3.40E-04 | 0.57 | 2.22 |
| HILIC_NEG_246_07494a1_595 | | 246.0749 | 1.60 | | HILIC | NEG | | | 177160 | 115224 | 244071 | 148974 | 3.53E-04 | 0.51 | 2.07 |
| RP_NEG_133_01431a0_64 | | 133.0143 | 0.64 | | RP | NEG | | | 202333 | 71721.7 | 171753 | 51669 | 3.55E-04 | -0.5 | 2.03 |
| RP_NEG_450_2634a10_105 | | 450.2634 | 10.11 | | RP | NEG | | | 798902 | 483625 | 1135613 | 802716 | 3.57E-04 | 0.52 | 2.12 |
| RP_POS_484_2905a8_444 | | 484.2905 | 8.44 | | RP | POS | | | 523103 | 240303 | 671796 | 342983 | 3.80E-04 | 0.51 | 2.01 |
| RP_NEG_658_36115a9_587 | | 658.3612 | 9.59 | | RP | NEG | | | 818982 | 881362 | 478091 | 471952 | 3.87E-04 | -0.5 | 1.87 |
| RP_NEG_367_34381a7_265 | | 367.3438 | 7.27 | | RP | NEG | | | 325476 | 205751 | 442796 | 263841 | 4.17E-04 | 0.5 | 2.03 |
| HILIC_POS_583_255a0_621 | | 583.2550 | 0.62 | | HILIC | POS | | | 133644 | 88627.7 | 98259.9 | 54537.9 | 4.19E-04 | -0.49 | 1.9 |
| RP_POS_585_27063a9_274 | | 585.2706 | 9.27 | | RP | POS | | | 2385264 | 1746846 | 1663757 | 1195776 | 4.22E-04 | -0.49 | 1.93 |
| RP_POS_658_3584a8_923 | | 658.3584 | 8.92 | | RP | POS | | | 896490 | 955717 | 533709 | 486177 | 4.23E-04 | -0.5 | 1.86 |
| RP_POS_445_29465a9_728 | | 445.2947 | 9.73 | | RP | POS | | | 962238 | 1928769 | 299488 | 415204 | 4.24E-04 | -0.57 | 1.83 |
| RP_POS_583_25507a11_849 | | 583.2551 | 11.85 | | RP | POS | 297.1218(100), 583.2565(65) | | 1.7E+07 | 1.3E+07 | 1.2E+07 | 9127762 | 4.27E-04 | -0.49 | 1.92 |
| RP_NEG_605_2392a11_846 | | 605.2392 | 11.85 | | RP | NEG | | | 1463798 | 1163125 | 982338 | 804914 | 4.32E-04 | -0.49 | 1.92 |
| RP_POS_303_19522a7_299 | | 303.1952 | 7.30 | | RP | POS | | | 229965 | 107062 | 294859 | 151817 | 4.66E-04 | 0.5 | 1.99 |
| RP_POS_585_27057a11_849 | Bilirubin | 585.2706 | 11.85 | HMDB0240584 | RP | POS | 229.1381(100), 271.1465(100) | 2 | 4.1E+07 | 2.4E+07 | 3.1E+07 | 1.8E+07 | 4.66E-04 | -0.48 | 1.92 |
| RP_NEG_313_11972a7_246 | | 313.1197 | 7.25 | | RP | NEG | | | 346819 | 225688 | 255287 | 150189 | 4.71E-04 | -0.49 | 1.88 |
| RP_POS_247_13316a7_214 | | 247.1332 | 7.21 | | RP | POS | | | 798044 | 289896 | 674675 | 221628 | 4.92E-04 | -0.48 | 1.91 |
| RP_NEG_844_53333a13_031 | | 844.5333 | 13.03 | | RP | NEG | | | 1281112 | 578328 | 1608031 | 746598 | 4.93E-04 | 0.49 | 1.97 |
| RP_POS_383_3298a10_613 | | 383.3298 | 10.61 | | RP | POS | | | 1082813 | 316766 | 926135 | 331258 | 5.07E-04 | -0.48 | 1.97 |
| RP_NEG_595_49567a11_477 | | 595.4957 | 11.48 | | RP | NEG | | | 1204748 | 822460 | 1689926 | 1136707 | 5.21E-04 | 0.5 | 2.01 |
| RP_POS_350_17999a5_568 | | 350.1800 | 5.57 | | RP | POS | | | 1354644 | 544004 | 1670862 | 736308 | 5.22E-04 | 0.49 | 2 |
| RP_NEG_279_6142a7_355 | | 279.6142 | 7.36 | | RP | NEG | | | 4438349 | 2325893 | 5850341 | 3349351 | 5.25E-04 | 0.5 | 2.05 |
| RP_POS_365_10516a0_556 | | 365.1052 | 0.56 | | RP | POS | | | 1541747 | 1938759 | 885038 | 476200 | 5.53E-04 | -0.54 | 1.88 |
| RP_POS_178_08649a0_627 | | 178.0865 | 0.63 | | RP | POS | | | 974474 | 344658 | 1179087 | 484241 | 5.56E-04 | 0.49 | 2 |
| RP_NEG_465_24976a8_443 | | 465.2498 | 8.44 | | RP | NEG | | | 1183103 | 536732 | 1511300 | 787861 | 5.71E-04 | 0.5 | 1.97 |
| RP_NEG_874_56146a12_684 | | 874.5615 | 12.68 | | RP | NEG | | | 2554541 | 516194 | 2323520 | 459576 | 6.11E-04 | -0.47 | 1.94 |
| RP_POS_431_31558a9_951 | 7a-hydroxy-3-oxo-4-cholestenoic acid | 431.3156 | 9.95 | HMDB0012458 | RP | POS | 97.0654(100),177.1279(60) | 2 | 7273328 | 2023381 | 8230204 | 2014879 | 6.30E-04 | 0.47 | 1.89 |
| RP_POS_581_2395a11_851 | | 581.2395 | 11.85 | | RP | POS | | | 1776160 | 1545672 | 1163352 | 1036318 | 6.45E-04 | -0.47 | 1.87 |
| RP_POS_617_2605a11_85 | | 617.2605 | 11.85 | | RP | POS | | | 1258000 | 864191 | 898396 | 661080 | 6.51E-04 | -0.47 | 1.89 |
| RP_NEG_222_82977a15_501 | | 222.8298 | 15.50 | | RP | NEG | | | 263658 | 58718.7 | 237173 | 53810.2 | 6.61E-04 | -0.47 | 2.07 |
| RP_POS_288_23206a7_242 | | 288.2321 | 7.24 | | RP | POS | | | 157230 | 131901 | 222092 | 142317 | 6.80E-04 | 0.47 | 1.95 |
| RP_POS_166_08389a0_588 | | 166.0839 | 0.59 | | RP | POS | | | 3519847 | 3071821 | 2260064 | 2288794 | 6.84E-04 | -0.47 | 1.83 |
| RP_NEG_605_23859a7_233 | | 605.2386 | 7.23 | | RP | NEG | | | 278964 | 231485 | 191493 | 135077 | 6.86E-04 | -0.48 | 1.82 |
| RP_NEG_404_18915a7_245 | | 404.1892 | 7.25 | | RP | NEG | | | 839870 | 76035.8 | 806399 | 67015.2 | 7.04E-04 | -0.47 | 1.9 |
| RP_POS_660_37408a9_59 | | 660.3741 | 9.59 | | RP | POS | | | 1398835 | 1586842 | 819229 | 817380 | 7.06E-04 | -0.48 | 1.79 |
| RP_NEG_165_05574a4_682 | 3-(3-Hydroxyphenyl)propionic acid | 165.0557 | 4.68 | HMDB0000375 | RP | NEG | 121.0662(100), 164.8955(20), 119.0502(49) | 2 | 472687 | 827035 | 1041685 | 1450695 | 7.13E-04 | 0.50 | 2 |
| HILIC_NEG_294_69153a1_408 | | 294.6915 | 1.41 | | HILIC | NEG | | | 146951 | 79620.4 | 118732 | 36275.2 | 7.46E-04 | -0.49 | 1.8 |
| RP_NEG_469_22659a7_787 | | 469.2266 | 7.79 | | RP | NEG | | | 282074 | 128899 | 230062 | 93955.3 | 7.50E-04 | -0.47 | 1.93 |
| RP_POS_585_27063a9_466 | | 585.2706 | 9.47 | | RP | POS | | | 1224310 | 958657 | 834327 | 713435 | 7.50E-04 | -0.47 | 1.85 |
| RP_NEG_162_8392a0_588 | | 162.8392 | 0.59 | | RP | NEG | | | 4525208 | 1293188 | 5010477 | 758897 | 7.60E-04 | 0.47 | 1.84 |
| RP_NEG_429_3017a9_95 | 7a-hydroxy-3-oxo-4-cholestenoic acid | 429.3017 | 9.95 | HMDB0012458 | RP | NEG | 429.3044(100), 411.2892(30) | 2 | 1308506 | 372854 | 1477184 | 352035 | 7.65E-04 | 0.47 | 1.86 |
| HILIC_POS_494_32492a1_277 | LysoPC 16:1 | 494.3249 | 1.28 | HMDB0010383 | HILIC | POS | 184.0743(100), 104.1076(75) | 2 | 1.1E+07 | 3783725 | 1.3E+07 | 4065708 | 7.81E-04 | 0.47 | 1.93 |
| RP_NEG_246_0751a5_137 | | 246.0751 | 5.14 | | RP | NEG | | | 1812908 | 1227131 | 2515750 | 1710104 | 7.99E-04 | 0.48 | 1.97 |
| RP_POS_452_27704a10_106 | | 452.2770 | 10.11 | | RP | POS | | | 1843482 | 1208405 | 2603218 | 1912677 | 8.03E-04 | 0.49 | 2.01 |
| RP_POS_790_57385a13_298 | | 790.5739 | 13.30 | | RP | POS | | | 9608471 | 3058861 | 8336785 | 2439460 | 8.11E-04 | -0.46 | 1.92 |
| RP_NEG_475_30746a9_95 | | 475.3075 | 9.95 | | RP | NEG | | | 965854 | 269338 | 1090268 | 267253 | 8.16E-04 | 0.46 | 1.86 |
| RP_POS_645_29175a11_85 | | 645.2918 | 11.85 | | RP | POS | | | 5759143 | 4692258 | 3934906 | 3154692 | 8.24E-04 | -0.46 | 1.83 |
| RP_NEG_312_90292a15_371 | | 312.9029 | 15.37 | | RP | NEG | | | 787174 | 215986 | 680242 | 242163 | 8.24E-04 | -0.47 | 2.06 |
| RP_NEG_581_24146a11_845 | | 581.2415 | 11.85 | | RP | NEG | | | 1096676 | 1015235 | 702153 | 683348 | 8.32E-04 | -0.46 | 1.82 |
| RP_POS_249_10684a0_541 | | 249.1068 | 0.54 | | RP | POS | | | 1482068 | 238832 | 1378686 | 209454 | 8.36E-04 | -0.46 | 1.85 |
| RP_POS_401_34082a10_614 | | 401.3408 | 10.61 | | RP | POS | | | 969900 | 294393 | 833243 | 298183 | 8.79E-04 | -0.46 | 1.88 |
| RP_POS_613_26544a11_85 | | 613.2654 | 11.85 | | RP | POS | | | 2102873 | 1757528 | 1421364 | 1205481 | 9.21E-04 | -0.46 | 1.82 |
| RP_NEG_656_3454a8_921 | | 656.3454 | 8.92 | | RP | NEG | | | 565680 | 632291 | 342024 | 313613 | 9.31E-04 | -0.47 | 1.79 |
| RP_NEG_283_10904a11_846 | | 283.1090 | 11.85 | | RP | NEG | | | 774116 | 785566 | 475254 | 514948 | 9.60E-04 | -0.46 | 1.8 |
| RP_NEG_335_11151a8_825 | | 335.1115 | 8.83 | | RP | NEG | | | 191559 | 78583.8 | 159710 | 61388.4 | 9.84E-04 | -0.46 | 1.83 |
| RP_POS_800_5199a12_424 | | 800.5199 | 12.42 | | RP | POS | | | 3848135 | 1935799 | 3078725 | 1449409 | 1.01E-03 | -0.45 | 1.83 |
| RP_POS_605_23657a11_849 | | 605.2366 | 11.85 | | RP | POS | | | 1946915 | 1642950 | 1313972 | 1136035 | 1.03E-03 | -0.46 | 1.82 |
| RP_NEG_465_25a8_311 | | 465.2500 | 8.31 | | RP | NEG | | | 653821 | 346601 | 828621 | 413349 | 1.04E-03 | 0.46 | 1.84 |
| RP_NEG_193_81563a0_544 | | 193.8156 | 0.54 | | RP | NEG | | | 4368655 | 701867 | 4060477 | 659021 | 1.05E-03 | -0.45 | 1.86 |
| RP_NEG_481_24487a7_189 | | 481.2449 | 7.19 | | RP | NEG | | | 348209 | 178418 | 437216 | 209775 | 1.06E-03 | 0.46 | 1.83 |
| HILIC_POS_86_07198a1_218 | | 86.0720 | 1.22 | | HILIC | POS | | | 4602479 | 568916 | 4346168 | 564361 | 1.09E-03 | -0.45 | 1.87 |
| RP_NEG_273_12469a5_982 | | 273.1247 | 5.98 | | RP | NEG | | | 447642 | 389346 | 309581 | 209840 | 1.13E-03 | -0.46 | 1.72 |
| RP_POS_778_53754a12_426 | | 778.5375 | 12.43 | | RP | POS | | | 1.7E+07 | 7876674 | 1.4E+07 | 5935060 | 1.15E-03 | -0.45 | 1.8 |
| RP_NEG_583_25671a7_233 | | 583.2567 | 7.23 | | RP | NEG | | | 4544043 | 3753448 | 3199864 | 2137832 | 1.18E-03 | -0.46 | 1.74 |
| HILIC_POS_159_14919a6_118 | | 159.1492 | 6.12 | | HILIC | POS | | | 970687 | 1466573 | 507601 | 331507 | 1.18E-03 | -0.52 | 1.7 |
| RP_NEG_495_2243a6_378 | | 495.2243 | 6.38 | | RP | NEG | | | 985124 | 762774 | 1506965 | 1405903 | 1.20E-03 | 0.48 | 1.91 |
| RP_POS_661_52924a11_993 | | 661.5292 | 11.99 | | RP | POS | | | 2002656 | 636636 | 1743153 | 528032 | 1.23E-03 | -0.45 | 1.79 |
| RP_POS_806_56995a13_008 | PC 38:6 (16:0_22:6) | 806.5700 | 13.01 | HMDB0013409 | RP | POS | 184.0742(100) | 1 | 8.8E+08 | 2.2E+08 | 7.9E+08 | 1.9E+08 | 1.27E-03 | -0.44 | 1.84 |
| RP_POS_355_22653a10_078 | | 355.2265 | 10.08 | | RP | POS | | | 3299033 | 1542128 | 4090674 | 1949153 | 1.29E-03 | 0.45 | 1.92 |
| HILIC_POS_249_10652a1_218 | | 249.1065 | 1.22 | | HILIC | POS | | | 3088046 | 746834 | 2772020 | 676261 | 1.29E-03 | -0.44 | 1.83 |
| RP_NEG_583_25726a9_274 | | 583.2573 | 9.27 | | RP | NEG | | | 628110 | 559982 | 416695 | 387894 | 1.29E-03 | -0.45 | 1.77 |
| RP_POS_236_07149a9_267 | | 236.0715 | 9.27 | | RP | POS | | | 9.5E+07 | 3.6E+07 | 8.1E+07 | 2.9E+07 | 1.32E-03 | -0.44 | 1.78 |
| HILIC_NEG_279_61462a1_781 | | 279.6146 | 1.78 | | HILIC | NEG | | | 148167 | 81883.6 | 194816 | 120760 | 1.34E-03 | 0.46 | 1.93 |
| RP_POS_377_20856a10_078 | | 377.2086 | 10.08 | | RP | POS | | | 662385 | 299276 | 811899 | 364872 | 1.34E-03 | 0.45 | 1.91 |
| RP_NEG_631_242a8_454 | | 631.2420 | 8.45 | | RP | NEG | | | 698070 | 513190 | 512772 | 314191 | 1.35E-03 | -0.45 | 1.73 |
| HILIC_POS_114_0665a1_218 | Creatinine | 114.0665 | 1.22 | HMDB0000562 | HILIC | POS | 114.067(100), 86.0718(15) | 1 | 1.1E+09 | 1.3E+08 | 1E+09 | 1.3E+08 | 1.36E-03 | -0.44 | 1.85 |
| RP_NEG_273_12473a5_853 | | 273.1247 | 5.85 | | RP | NEG | | | 385490 | 315561 | 274776 | 175188 | 1.37E-03 | -0.45 | 1.7 |
| RP_POS_278_21136a9_347 | | 278.2114 | 9.35 | | RP | POS | | | 573076 | 281301 | 714877 | 350554 | 1.42E-03 | 0.45 | 1.85 |
| RP_POS_619_4906a11_598 | | 619.4906 | 11.60 | | RP | POS | | | 3656029 | 2391626 | 4944233 | 3284958 | 1.42E-03 | 0.45 | 1.89 |
| RP_NEG_317_11435a5_862 | | 317.1144 | 5.86 | | RP | NEG | | | 934503 | 773728 | 663619 | 431649 | 1.42E-03 | -0.45 | 1.7 |
| RP_NEG_255_11403a5_982 | | 255.1140 | 5.98 | | RP | NEG | | | 313781 | 263737 | 221623 | 146584 | 1.44E-03 | -0.45 | 1.69 |
| RP_NEG_583_2569a5_966 | | 583.2569 | 5.97 | | RP | NEG | | | 2822915 | 2382013 | 1988483 | 1341167 | 1.45E-03 | -0.45 | 1.71 |
| RP_POS_167_03378a11_408 | | 167.0338 | 11.41 | | RP | POS | | | 690497 | 128483 | 639932 | 101977 | 1.47E-03 | -0.44 | 1.8 |
| RP_NEG_238_07773a5_781 | | 238.0777 | 5.78 | | RP | NEG | | | 1.2E+07 | 7125359 | 1.6E+07 | 9427034 | 1.48E-03 | 0.45 | 1.82 |
| RP_POS_830_56873a12_688 | | 830.5687 | 12.69 | | RP | POS | | | 1.5E+07 | 3954592 | 1.4E+07 | 3224351 | 1.52E-03 | -0.44 | 1.79 |
| HILIC_NEG_538_31537a1_277 | | 538.3154 | 1.28 | | HILIC | NEG | | | 298575 | 99550.1 | 344479 | 109015 | 1.56E-03 | 0.44 | 1.83 |
| RP_NEG_261_07437a7_79 | | 261.0744 | 7.79 | | RP | NEG | | | 427745 | 400569 | 275297 | 298084 | 1.58E-03 | -0.44 | 1.86 |
| RP_NEG_239_09258a7_79 | 3-Carboxy-4-methyl-5-propyl-2-furanpropionic acid (CMPF) | 239.0926 | 7.79 | HMDB0061112 | RP | NEG | 195.1023(100), 239.0939(25) | 2 | 9494612 | 9369301 | 6018586 | 6584386 | 1.65E-03 | -0.44 | 1.84 |
| RP_POS_287_20041a7_997 | | 287.2004 | 8.00 | | RP | POS | | | 496408 | 203543 | 606741 | 288456 | 1.68E-03 | 0.45 | 1.81 |
| HILIC_NEG_206_10356a4_81 | | 206.1036 | 4.81 | | HILIC | NEG | | | 857657 | 163419 | 937205 | 198060 | 1.70E-03 | 0.44 | 1.96 |
| RP_POS_261_18481a9_345 | | 261.1848 | 9.35 | | RP | POS | | | 2367176 | 753206 | 2744924 | 956984 | 1.72E-03 | 0.44 | 1.83 |
| RP_NEG_289_10568a8_824 | | 289.1057 | 8.82 | | RP | NEG | | | 280539 | 103754 | 239469 | 86463.9 | 1.73E-03 | -0.43 | 1.75 |
| RP_POS_629_23322a11_85 | | 629.2332 | 11.85 | | RP | POS | | | 1000973 | 579706 | 773347 | 474174 | 1.73E-03 | -0.43 | 1.76 |
| RP_POS_292_63528a5_967 | | 292.6353 | 5.97 | | RP | POS | | | 207878 | 215711 | 132741 | 126772 | 1.75E-03 | -0.44 | 1.71 |
| RP_NEG_353_21198a10_077 | | 353.2120 | 10.08 | | RP | NEG | | | 4100440 | 2033455 | 5082470 | 2458384 | 1.82E-03 | 0.44 | 1.88 |
| HILIC_POS_682_32794a1_214 | | 682.3279 | 1.21 | | HILIC | POS | | | 174757 | 121425 | 238310 | 165703 | 1.83E-03 | 0.44 | 1.82 |
| HILIC_POS_585_27094a1_253 | | 585.2709 | 1.25 | | HILIC | POS | | | 379887 | 341571 | 254329 | 240505 | 1.83E-03 | -0.43 | 1.7 |
| RP_POS_189_71233a12_996 | | 189.7123 | 13.00 | | RP | POS | | | 353220 | 278184 | 255394 | 171703 | 1.84E-03 | -0.43 | 1.67 |
| RP_POS_642_6524a11_434 | | 642.6524 | 11.43 | | RP | POS | | | 119384 | 62398.8 | 155755 | 98819.9 | 1.85E-03 | 0.45 | 1.79 |
| RP_POS_854_56854a12_589 | | 854.5685 | 12.59 | | RP | POS | | | 3831704 | 1261070 | 3326005 | 1097308 | 1.86E-03 | -0.43 | 1.8 |
| RP_NEG_130_94427a11_593 | | 130.9443 | 11.59 | | RP | NEG | | | 7100849 | 1999093 | 7979592 | 2081759 | 1.89E-03 | 0.43 | 1.86 |
| HILIC_POS_156_04218a3_582 | | 156.0422 | 3.58 | | HILIC | POS | | | 8376709 | 2E+07 | 2548558 | 2770935 | 1.89E-03 | -0.52 | 1.6 |
| RP_POS_172_13306a0_682 | | 172.1331 | 0.68 | | RP | POS | | | 333780 | 65959.8 | 361978 | 65350.1 | 1.90E-03 | 0.43 | 1.73 |
| RP_NEG_195_10265a7_79 | | 195.1027 | 7.79 | | RP | NEG | | | 1315010 | 1293912 | 841462 | 911146 | 1.91E-03 | -0.43 | 1.82 |
| RP_NEG_311_22308a8_972 | | 311.2231 | 8.97 | | RP | NEG | | | 660437 | 341022 | 530556 | 266403 | 1.93E-03 | -0.43 | 1.69 |
| RP_POS_326_37778a9_12 | | 326.3778 | 9.12 | | RP | POS | | | 960470 | 200274 | 1124892 | 482990 | 1.94E-03 | 0.48 | 2.04 |
| RP_NEG_191_01947a0_678 | Citrate | 191.0195 | 0.68 | HMDB0000094 | RP | NEG | 111.0084(100), 87.0085(40),85.0292(28) | 2 | 2592170 | 1260091 | 2131799 | 889800 | 1.97E-03 | -0.43 | 1.69 |
| RP_NEG_220_9416a6_425 | | 220.9416 | 6.43 | | RP | NEG | | | 265847 | 145059 | 322970 | 123162 | 1.99E-03 | 0.43 | 1.79 |
| RP_POS_223_09639a7_791 | CMPF | 223.0964 | 7.79 | HMDB0061112 | RP | POS | 181.086(100), 139.0382(2) | 1 | 6913156 | 6684558 | 4482524 | 4676711 | 2.00E-03 | -0.43 | 1.81 |
| RP_POS_181_08595a7_791 | | 181.0860 | 7.79 | | RP | POS | 181.0859(100),139.0398(15) | | 1.2E+07 | 1.2E+07 | 7987129 | 8509835 | 2.12E-03 | -0.43 | 1.8 |
| RP_POS_263_20047a9_345 | | 263.2005 | 9.35 | | RP | POS | | | 4019489 | 1797631 | 4833716 | 2028758 | 2.25E-03 | 0.43 | 1.79 |
| RP_POS_229_15466a0_679 | | 229.1547 | 0.68 | | RP | POS | | | 1.2E+07 | 2928546 | 1.1E+07 | 1912102 | 2.26E-03 | -0.42 | 1.71 |
| RP_POS_793_54285a12_804 | | 793.5429 | 12.80 | | RP | POS | | | 2421321 | 775332 | 2740799 | 741408 | 2.27E-03 | 0.42 | 1.81 |
| RP_NEG_199_00711a4_421 | | 199.0071 | 4.42 | | RP | NEG | | | 1059227 | 3221953 | 2051643 | 1155400 | 2.28E-03 | 0.45 | 1.68 |
| HILIC_POS_162_11249a4_796 | Carnitine | 162.1125 | 4.80 | HMDB0000062 | HILIC | POS | 162.1118 (100), 103.0386 (15), 60.0803 (15) | 1 | 1.8E+09 | 2.8E+08 | 1.9E+09 | 3.2E+08 | 2.30E-03 | 0.43 | 1.92 |
| RP_NEG_305_24899a11_182 | | 305.2490 | 11.18 | | RP | NEG | | | 456073 | 292395 | 583878 | 312087 | 2.31E-03 | 0.42 | 1.93 |
| HILIC_NEG_198_93422a0_983 | | 198.9342 | 0.98 | | HILIC | NEG | | | 114690 | 46100.1 | 135799 | 53228.7 | 2.32E-03 | 0.43 | 1.78 |
| RP_NEG_383_31705a11_13 | | 383.3171 | 11.13 | | RP | NEG | | | 428728 | 49869.6 | 406857 | 53738.1 | 2.36E-03 | -0.42 | 1.79 |
| RP_NEG_746_513a13_223 | | 746.5130 | 13.22 | | RP | NEG | | | 1481077 | 414721 | 1323521 | 337653 | 2.36E-03 | -0.42 | 1.71 |
| HILIC_POS_200_08939a4_453 | | 200.0894 | 4.45 | | HILIC | POS | | | 251597 | 174459 | 191615 | 108903 | 2.39E-03 | -0.42 | 1.65 |
| RP_POS_563_42841a11_14 | | 563.4284 | 11.14 | | RP | POS | | | 544819 | 370483 | 724551 | 471020 | 2.42E-03 | 0.43 | 1.77 |

**Supplemantary table 1**: The non-targeted metabolomics results ordered by the VIP value (p value < 0.0003 are shaded dark green, p value < 0.05 are shaded light green, d values below 0 are blue and above 0 are red)

| | | | | | | | | | Cases | | Controls | | Welch test | Cohen's | |
|---|---|---|---|---|---|---|---|---|---|---|---|---|---|---|---|
| Feature_ID | Metabolite_name | Average_Mz | Average_RT_min_ | HMDB ID | RP_HILIC | POS_NEG | Cureted_MSMS | ID_level | Mean | SD | Mean2 | SD3 | p | d | VIP |
| RP_POS_665_24524a8_686 | | 665.2452 | 8.69 | | RP | POS | | | 306134 | 292458 | 206193 | 180005 | 2.43E-03 | -0.42 | 1.68 |
| RP_NEG_413_20093a7_176 | | 413.2009 | 7.18 | | RP | NEG | | | 602694 | 280678 | 721909 | 288147 | 2.46E-03 | 0.42 | 1.7 |
| RP_POS_576_49854a12_233 | | 576.4985 | 12.23 | | RP | POS | | | 834668 | 742179 | 1258384 | 1189959 | 2.49E-03 | 0.44 | 1.81 |
| HILIC_NEG_379_60132a1_412 | | 379.6013 | 1.41 | | HILIC | NEG | | | 383788 | 164436 | 325440 | 114254 | 2.49E-03 | -0.42 | 1.62 |
| RP_NEG_377_08569a0_556 | | 377.0857 | 0.56 | | RP | NEG | | | 1198285 | 1170268 | 841775 | 419614 | 2.54E-03 | -0.45 | 1.71 |
| HILIC_POS_132_0657a5_655 | | 132.0657 | 5.66 | | HILIC | POS | | | 5528260 | 2291630 | 6724675 | 3266838 | 2.55E-03 | 0.43 | 1.77 |
| RP_POS_263_08896a7_791 | Methyl 2-((4-methyl-2-oxo-2H-chromen-7-yl)oxy)propanoate | 263.0890 | 7.79 | CID: 3782756 | RP | POS | 263.0891(100) | 2 | 3868146 | 4104526 | 2434327 | 2746350 | 2.55E-03 | -0.42 | 1.76 |
| RP_POS_484_2905a8_312 | | 484.2905 | 8.31 | | RP | POS | | | 536468 | 286114 | 667287 | 335345 | 2.58E-03 | 0.42 | 1.7 |
| RP_NEG_399_22131a8_089 | | 399.2213 | 8.09 | | RP | NEG | | | 3.7E+07 | 1.5E+07 | 3.1E+07 | 1.3E+07 | 2.58E-03 | -0.42 | 1.64 |
| RP_NEG_691_50543a11_774 | | 691.5054 | 11.77 | | RP | NEG | | | 1647225 | 489974 | 1455784 | 432424 | 2.58E-03 | -0.42 | 1.69 |
| RP_POS_144_10204a0_588 | | 144.1020 | 0.59 | | RP | POS | | | 1.2E+08 | 1.2E+08 | 7.5E+07 | 8.9E+07 | 2.63E-03 | -0.42 | 1.62 |
| HILIC_NEG_373_60724a1_41 | | 373.6072 | 1.41 | | HILIC | NEG | | | 1798748 | 787356 | 1525894 | 521175 | 2.66E-03 | -0.42 | 1.61 |
| HILIC_POS_166_08609a3_062 | | 166.0861 | 3.06 | | HILIC | POS | 166.0871(100) | | 107020 | 151114 | 608538 | 1615001 | 2.69E-03 | 0.57 | 1.89 |
| RP_POS_255_12259a7_791 | | 255.1226 | 7.79 | | RP | POS | | | 2781734 | 2576432 | 1863880 | 1853700 | 2.71E-03 | -0.41 | 1.78 |
| HILIC_POS_176_04057a5_585 | | 176.0406 | 5.59 | | HILIC | POS | | | 306153 | 93020.5 | 342140 | 81386.3 | 2.73E-03 | 0.41 | 1.76 |
| RP_POS_398_32629a9_096 | | 398.3263 | 9.10 | | RP | POS | | | 2781150 | 1390535 | 3556823 | 2189845 | 2.74E-03 | 0.43 | 1.92 |
| RP_POS_535_40045a10_573 | | 535.4005 | 10.57 | | RP | POS | | | 1027615 | 431803 | 850645 | 424140 | 2.75E-03 | -0.41 | 1.73 |
| RP_POS_491_37122a11_139 | | 491.3712 | 11.14 | | RP | POS | | | 413918 | 231086 | 511590 | 242200 | 2.89E-03 | 0.41 | 1.76 |
| HILIC_NEG_299_68423a1_411 | | 299.6842 | 1.41 | | HILIC | NEG | | | 699340 | 166162 | 646153 | 86154.1 | 2.93E-03 | -0.42 | 1.59 |
| HILIC_NEG_146_08247a4_803 | | 146.0825 | 4.80 | | HILIC | NEG | | | 391362 | 78059 | 427246 | 94362.5 | 2.96E-03 | 0.42 | 1.85 |
| RP_POS_241_10686a7_792 | | 241.1069 | 7.79 | | RP | POS | | | 934028 | 853486 | 633399 | 612091 | 2.98E-03 | -0.41 | 1.77 |
| RP_POS_361_20331a11_148 | | 361.2033 | 11.15 | | RP | POS | | | 522981 | 102627 | 478805 | 111720 | 2.99E-03 | -0.41 | 1.67 |
| HILIC_NEG_375_60498a1_41 | | 375.6050 | 1.41 | | HILIC | NEG | | | 2560699 | 1128316 | 2175399 | 746355 | 3.05E-03 | -0.41 | 1.58 |
| RP_NEG_147_02966a0_679 | | 147.0297 | 0.68 | | RP | NEG | | | 208714 | 48518.7 | 191394 | 36255.1 | 3.05E-03 | -0.41 | 1.65 |
| RP_POS_583_25476a8_251 | Biliverdin | 583.2548 | 8.25 | HMDB0001008 | RP | POS | 297.1216(100), 583.2557(100) | 2 | 1592050 | 2850306 | 719215 | 1212305 | 3.06E-03 | -0.43 | 1.56 |
| HILIC_NEG_371_60843a1_411 | | 371.6084 | 1.41 | | HILIC | NEG | | | 493327 | 213484 | 420276 | 142185 | 3.06E-03 | -0.41 | 1.59 |
| RP_NEG_331_13037a6_351 | | 331.1304 | 6.35 | | RP | NEG | | | 154092 | 167918 | 100326 | 89185.2 | 3.08E-03 | -0.42 | 1.59 |
| RP_POS_251_12775a8_827 | Furan fatty acid | 251.1278 | 8.83 | HMDB0031005 | RP | POS | 209.1172(100) | 3 | 5537852 | 1856431 | 4843321 | 1560226 | 3.14E-03 | -0.41 | 1.67 |
| RP_NEG_267_12372a8_826 | | 267.1237 | 8.83 | | RP | NEG | | | 6439308 | 2534967 | 5511594 | 2044787 | 3.25E-03 | -0.41 | 1.66 |
| HILIC_NEG_80_91636a1_41 | | 80.9164 | 1.41 | | HILIC | NEG | | | 3048225 | 1182043 | 2675487 | 610776 | 3.35E-03 | -0.42 | 1.57 |
| RP_POS_420_25424a11_362 | | 420.2542 | 11.36 | | RP | POS | | | 758996 | 281077 | 669230 | 153581 | 3.37E-03 | -0.41 | 1.71 |
| HILIC_NEG_78_91837a1_41 | | 78.9184 | 1.41 | | HILIC | NEG | | | 2998047 | 1147977 | 2634292 | 609922 | 3.40E-03 | -0.41 | 1.56 |
| RP_NEG_188_02094a0_56 | | 188.0209 | 0.56 | | RP | NEG | | | 359873 | 130888 | 414928 | 140342 | 3.42E-03 | 0.41 | 1.63 |
| RP_NEG_282_99167a4_771 | | 282.9917 | 4.77 | | RP | NEG | | | 370192 | 84096.4 | 333077 | 98315 | 3.55E-03 | -0.41 | 1.89 |
| HILIC_POS_398_32639a0_612 | | 398.3264 | 0.61 | | HILIC | POS | | | 299345 | 149026 | 380607 | 236374 | 3.56E-03 | 0.42 | 1.89 |
| HILIC_NEG_377_60306a1_41 | | 377.6031 | 1.41 | | HILIC | NEG | | | 1614372 | 709818 | 1375946 | 469806 | 3.56E-03 | -0.4 | 1.56 |
| RP_POS_211_1442a4_793 | | 211.1442 | 4.79 | | RP | POS | 211.1456(100), 70.066(40) | | 1946184 | 1326550 | 2717658 | 2295085 | 3.65E-03 | 0.43 | 1.8 |
| RP_POS_301_17734a9_345 | | 301.1773 | 9.35 | | RP | POS | | | 4178540 | 1738877 | 4949333 | 2062304 | 3.70E-03 | 0.41 | 1.7 |
| RP_POS_455_31314a9_833 | | 455.3131 | 9.83 | | RP | POS | | | 550927 | 197196 | 641624 | 247616 | 3.70E-03 | 0.41 | 1.68 |
| HILIC_POS_176_12808a4_058 | | 176.1281 | 4.06 | | HILIC | POS | | | 740625 | 613801 | 1025588 | 782487 | 3.72E-03 | 0.41 | 1.74 |
| RP_POS_241_15465a0_68 | | 241.1547 | 0.68 | | RP | POS | | | 1598410 | 284948 | 1492204 | 247212 | 3.72E-03 | -0.4 | 1.75 |
| HILIC_NEG_296_6886a1_41 | | 296.6886 | 1.41 | | HILIC | NEG | | | 381755 | 217131 | 315103 | 105196 | 3.75E-03 | -0.41 | 1.54 |
| RP_POS_357_20847a11_362 | | 357.2085 | 11.36 | | RP | POS | | | 870774 | 385580 | 754349 | 176710 | 3.94E-03 | -0.41 | 1.59 |
| RP_POS_292_63428a7_239 | | 292.6343 | 7.24 | | RP | POS | | | 440089 | 391523 | 314669 | 230758 | 3.95E-03 | -0.4 | 1.61 |
| RP_NEG_369_17444a8_46 | | 369.1744 | 8.46 | | RP | NEG | | | 3663012 | 2207469 | 4609922 | 2516013 | 3.98E-03 | 0.4 | 1.71 |
| RP_NEG_840_53296a13_006 | | 840.5330 | 13.01 | | RP | NEG | | | 2608914 | 579165 | 2397634 | 487702 | 3.99E-03 | -0.4 | 1.64 |
| RP_POS_302_21432a8_385 | | 302.2143 | 8.39 | | RP | POS | | | 3499273 | 1396806 | 4210371 | 2056315 | 4.01E-03 | 0.41 | 1.72 |
| RP_POS_339_1412a6_285 | Columbamine | 339.1412 | 6.29 | CID: 72310 | RP | POS | 339.1753(100) | 2 | 195162 | 104031 | 161994 | 60739.8 | 4.02E-03 | -0.40 | 1.77 |
| RP_POS_209_11748a8_827 | | 209.1175 | 8.83 | | RP | POS | | | 6602317 | 2413092 | 5735575 | 1975165 | 4.09E-03 | -0.4 | 1.62 |
| RP_NEG_527_21387a7_139 | | 527.2139 | 7.14 | | RP | NEG | | | 283131 | 103910 | 327339 | 117486 | 4.10E-03 | 0.4 | 1.66 |
| RP_POS_302_10211a5_856 | | 302.1021 | 5.86 | | RP | POS | | | 373142 | 369933 | 252636 | 234203 | 4.12E-03 | -0.4 | 1.57 |
| RP_POS_585_27118a5_969 | | 585.2712 | 5.97 | | RP | POS | | | 1.1E+07 | 9979806 | 8221252 | 5853979 | 4.17E-03 | -0.4 | 1.56 |
| HILIC_POS_132_07684a5_587 | Creatine | 132.0768 | 5.59 | HMDB0000064 | HILIC | POS | 90.0557(100), 132.0762(70) | 1 | 1.6E+08 | 5.7E+07 | 1.8E+08 | 6.1E+07 | 4.22E-03 | 0.40 | 1.62 |
| RP_POS_231_17429a9_344 | | 231.1743 | 9.34 | | RP | POS | | | 3465023 | 1475098 | 4102343 | 1726661 | 4.32E-03 | 0.4 | 1.67 |
| RP_POS_323_25552a10_625 | | 323.2555 | 10.63 | | RP | POS | | | 2469450 | 416448 | 2308188 | 403230 | 4.32E-03 | -0.39 | 1.76 |
| RP_POS_676_67035a5_81 | | 676.6704 | 5.81 | | RP | POS | | | 763374 | 1197931 | 410209 | 510287 | 4.33E-03 | -0.41 | 1.63 |
| HILIC_POS_130_04985a6_383 | adduct of glutamate (Pyroglutamic acid) | 130.0499 | 6.38 | | HILIC | POS | 84.0451(100), 130.1588(40) | | 711025 | 219725 | 814478 | 293468 | 4.35E-03 | 0.4 | 1.68 |
| RP_NEG_223_13416a8_825 | | 223.1342 | 8.83 | | RP | NEG | | | 421796 | 161167 | 364457 | 131889 | 4.45E-03 | -0.39 | 1.6 |
| RP_NEG_467_20853a8_09 | | 467.2085 | 8.09 | | RP | NEG | | | 1152326 | 471704 | 979450 | 412320 | 4.47E-03 | -0.39 | 1.56 |
| RP_POS_282_11954a0_678 | | 282.1195 | 0.68 | | RP | POS | | | 1180519 | 126915 | 1129898 | 130992 | 4.53E-03 | -0.39 | 1.58 |
| RP_NEG_188_98648a1_868 | Pyrocatechol sulfate | 188.9865 | 1.87 | HMDB0059724 | RP | NEG | 109.0296(100), 188.9875(45) | 2 | 2354012 | 1714645 | 3191975 | 2433242 | 4.55E-03 | 0.40 | 1.72 |
| RP_POS_450_35794a11_408 | | 450.3579 | 11.41 | | RP | POS | | | 1.8E+07 | 3238750 | 1.7E+07 | 2688304 | 4.67E-03 | -0.39 | 1.77 |
| RP_NEG_613_36023a8_789 | | 613.3602 | 8.79 | | RP | NEG | | | 1253406 | 562638 | 1457430 | 486984 | 4.70E-03 | 0.39 | 1.57 |
| RP_POS_283_15396a8_826 | | 283.1540 | 8.83 | | RP | POS | | | 2336524 | 840531 | 2038550 | 699503 | 4.89E-03 | -0.39 | 1.61 |
| RP_POS_329_2684a10_504 | | 329.2684 | 10.50 | | RP | POS | | | 755302 | 828614 | 1321810 | 1817247 | 4.92E-03 | 0.43 | 1.72 |
| HILIC_NEG_242_95975a5_121 | | 242.9598 | 5.12 | | HILIC | NEG | | | 275123 | 140796 | 348715 | 222292 | 5.00E-03 | 0.41 | 1.95 |
| RP_NEG_401_21674a8_09 | | 401.2167 | 8.09 | | RP | NEG | | | 1337087 | 563757 | 1134624 | 485647 | 5.02E-03 | -0.39 | 1.53 |
| RP_NEG_470_34897a9_537 | | 470.3490 | 9.54 | | RP | NEG | | | 237891 | 88499.5 | 272113 | 88611.3 | 5.11E-03 | 0.39 | 1.75 |
| RP_NEG_130_06219a0_576 | | 130.0622 | 0.58 | | RP | NEG | | | 344427 | 132759 | 398831 | 147857 | 5.24E-03 | 0.39 | 1.56 |
| HILIC_POS_144_1019a3_42 | Proline betaine | 144.1019 | 3.42 | HMDB0004827 | HILIC | POS | 144.1028(100) | 1 | 1.3E+09 | 1.8E+09 | 7.3E+08 | 1.1E+09 | 5.26E-03 | -0.39 | 1.49 |
| RP_NEG_382_14557a10_81 | | 382.1456 | 10.81 | | RP | NEG | | | 246809 | 191535 | 320527 | 191431 | 5.28E-03 | 0.38 | 1.75 |
| RP_POS_389_23492a11_363 | | 389.2349 | 11.36 | | RP | POS | | | 610961 | 219258 | 543797 | 124061 | 5.28E-03 | -0.39 | 1.58 |
| HILIC_POS_131_11806a1_354 | | 131.1181 | 1.35 | | HILIC | POS | | | 1271188 | 616277 | 1526335 | 701622 | 5.38E-03 | 0.39 | 1.6 |
| RP_NEG_467_3739a11_076 | | 467.3739 | 11.08 | | RP | NEG | | | 236613 | 118721 | 284235 | 128372 | 5.42E-03 | 0.39 | 1.65 |
| RP_POS_203_1066a7_846 | | 203.1066 | 7.85 | | RP | POS | | | 1220288 | 168178 | 1290618 | 195128 | 5.46E-03 | 0.39 | 1.65 |
| RP_NEG_557_45807a12_23 | | 557.4581 | 12.23 | | RP | NEG | | | 939753 | 616982 | 1196437 | 710422 | 5.48E-03 | 0.39 | 1.6 |
| RP_NEG_245_06712a5_784 | | 245.0671 | 5.78 | | RP | NEG | | | 1005702 | 475353 | 1185773 | 465248 | 5.49E-03 | 0.38 | 1.7 |
| HILIC_POS_585_37408a1_012 | | 585.3741 | 1.01 | | HILIC | POS | | | 211571 | 91848.4 | 178829 | 79703.8 | 5.50E-03 | -0.38 | 1.55 |
| RP_POS_583_25494a9_272 | | 583.2549 | 9.27 | | RP | POS | | | 993589 | 939986 | 686837 | 665659 | 5.64E-03 | -0.38 | 1.64 |
| HILIC_POS_284_1062a6_198 | | 284.1062 | 6.20 | | HILIC | POS | | | 121606 | 58723.5 | 100459 | 52479.9 | 5.68E-03 | -0.38 | 1.74 |
| RP_POS_453_29623a9_953 | Ruscogenin | 453.2962 | 9.95 | HMDB0257364 | RP | POS | 453.2987(100) | 2 | 1112319 | 342567 | 1239593 | 325818 | 5.69E-03 | 0.38 | 1.55 |
| RP_NEG_213_02303a5_294 | | 213.0230 | 5.29 | | RP | NEG | | | 1036952 | 1183858 | 669609 | 723176 | 5.69E-03 | -0.39 | 1.48 |
| RP_POS_413_13586a7_265 | | 413.1359 | 7.27 | | RP | POS | | | 544094 | 193087 | 621179 | 209371 | 5.72E-03 | 0.38 | 1.74 |
| RP_POS_359_20837a11_362 | | 359.2084 | 11.36 | | RP | POS | | | 785795 | 223042 | 714373 | 151569 | 5.84E-03 | -0.38 | 1.67 |
| HILIC_NEG_130_06221a5_591 | Creatine | 130.0622 | 5.59 | HMDB0000064 | HILIC | NEG | 88.0401(100),130.0402(4) | 1 | 383834 | 151609 | 443375 | 160598 | 5.85E-03 | 0.38 | 1.56 |
| HILIC_NEG_88_04a5_59 | Alanine | 88.0400 | 5.59 | HMDB0000161 | HILIC | NEG | 88.0401(100) | 1 | 445716 | 177613 | 515371 | 187832 | 5.87E-03 | 0.38 | 1.55 |
| HILIC_NEG_495_29636a2_532 | | 495.2964 | 2.53 | | HILIC | NEG | | | 171425 | 72688.6 | 141296 | 84444.2 | 5.91E-03 | -0.38 | 1.55 |
| RP_POS_253_14088a7_972 | | 253.1409 | 7.97 | | RP | POS | | | 1340721 | 183653 | 1271894 | 179166 | 5.91E-03 | -0.38 | 1.6 |
| RP_POS_604_0752a10_389 | | 604.0752 | 10.39 | | RP | POS | | | 413052 | 251081 | 531006 | 352146 | 5.92E-03 | 0.39 | 1.71 |
| RP_POS_161_0961a9_345 | | 161.0961 | 9.35 | | RP | POS | | | 1605514 | 587685 | 1845940 | 668443 | 5.92E-03 | 0.38 | 1.6 |
| RP_POS_287_12524a7_216 | | 287.1252 | 7.22 | | RP | POS | | | 456046 | 204809 | 379418 | 199290 | 5.93E-03 | -0.38 | 1.51 |
| RP_NEG_165_05574a4_768 | 3-Phenyllactic acid | 165.0557 | 4.77 | HMDB0000748 | RP | NEG | 147.0452(100), 119.05(30), 72.9928(20) | 2 | 957878 | 552967 | 756524 | 511196 | 5.96E-03 | -0.38 | 1.52 |
| HILIC_NEG_389_66876a1_409 | | 389.6688 | 1.41 | | HILIC | NEG | | | 299100 | 97505.1 | 266974 | 72089.6 | 5.97E-03 | -0.38 | 1.49 |
| RP_POS_637_48029a13_606 | | 637.4803 | 13.61 | | RP | POS | | | 777048 | 648347 | 566561 | 461588 | 5.99E-03 | -0.38 | 1.56 |
| RP_POS_595_49323a11_349 | | 595.4932 | 11.35 | | RP | POS | | | 869229 | 623041 | 1151087 | 830031 | 6.01E-03 | 0.39 | 1.64 |
| RP_POS_189_07857a6_031 | | 189.0786 | 6.03 | | RP | POS | | | 829805 | 475223 | 559275 | 860956 | 6.01E-03 | -0.4 | 1.69 |
| HILIC_NEG_302_10101a6_216 | | 302.1010 | 6.22 | | HILIC | NEG | | | 290120 | 103165 | 251004 | 103505 | 6.08E-03 | -0.38 | 1.87 |
| HILIC_POS_106_05034a6_196 | Serine | 106.0503 | 6.20 | HMDB0000187 | HILIC | POS | 60.0454(100), 88.0763(55) | 1 | 2764974 | 747849 | 3021300 | 613171 | 6.14E-03 | 0.38 | 1.54 |
| RP_POS_426_3569a9_541 | AC 18:1 (Oleoylcarnitine) | 426.3569 | 9.54 | HMDB0005065 | RP | POS | 85.0292(100), 426.3578(60) | 2 | 1.8E+07 | 5913145 | 2E+07 | 6151607 | 6.15E-03 | 0.38 | 1.75 |
| RP_POS_444_40521a12_309 | | 444.4052 | 12.31 | | RP | POS | | | 421219 | 65868.4 | 449774 | 82788.7 | 6.18E-03 | 0.38 | 1.59 |
| HILIC_POS_178_08604a3_548 | | 178.0860 | 3.55 | | HILIC | POS | | | 2186034 | 1282094 | 2722990 | 1527931 | 6.19E-03 | 0.38 | 1.57 |
| RP_POS_626_30182a5_523 | | 626.3018 | 5.52 | | RP | POS | | | 410325 | 105116 | 452938 | 119307 | 6.31E-03 | 0.38 | 1.57 |
| RP_POS_272_25793a10_123 | | 272.2579 | 10.12 | | RP | POS | | | 607926 | 69320.3 | 581980 | 68518.2 | 6.33E-03 | -0.38 | 1.54 |
| HILIC_POS_229_1546a4_136 | | 229.1546 | 4.14 | | HILIC | POS | | | 9424315 | 2563426 | 8619339 | 1695816 | 6.35E-03 | -0.38 | 1.56 |
| RP_POS_247_14403a2_752 | | 247.1440 | 2.75 | | RP | POS | 188.0708(100), 146.0601(55) | | 1953451 | 2965444 | 1109918 | 1333285 | 6.35E-03 | -0.39 | 1.52 |
| HILIC_NEG_139_94464a5_925 | | 139.9446 | 5.93 | | HILIC | NEG | | | 270023 | 40776.6 | 287335 | 49899.7 | 6.36E-03 | 0.38 | 1.64 |
| HILIC_NEG_232_90669a6_176 | | 232.9067 | 6.18 | | HILIC | NEG | | | 151487 | 28077.4 | 161764 | 26656 | 6.38E-03 | 0.38 | 1.52 |
| HILIC_POS_196_07365a2_969 | | 196.0737 | 2.97 | | HILIC | POS | | | 888954 | 596048 | 702799 | 388450 | 6.38E-03 | -0.38 | 1.48 |
| HILIC_NEG_295_68845a1_408 | | 295.6885 | 1.41 | | HILIC | NEG | | | 240021 | 59116.4 | 222696 | 30750 | 6.39E-03 | -0.39 | 1.48 |
| RP_NEG_264_99118a8_771 | | 264.9912 | 8.77 | | RP | NEG | | | 921005 | 103029 | 883168 | 98536.6 | 6.41E-03 | -0.38 | 1.52 |
| RP_POS_319_12857a5_86 | | 319.1286 | 5.86 | | RP | POS | | | 529632 | 510075 | 369254 | 340180 | 6.43E-03 | -0.38 | 1.51 |
| RP_NEG_399_22159a7_754 | | 399.2216 | 7.75 | | RP | NEG | | | 709952 | 570146 | 541362 | 309708 | 6.47E-03 | -0.38 | 1.48 |
| RP_POS_566_32129a10_292 | | 566.3213 | 10.29 | | RP | POS | | | 2.9E+07 | 5477035 | 3.1E+07 | 6679759 | 6.50E-03 | 0.38 | 1.6 |
| RP_NEG_188_98642a2_048 | | 188.9864 | 2.05 | | RP | NEG | 109.0297(100), 188.9853(45) | | 3268936 | 2353263 | 4360016 | 3293940 | 6.51E-03 | 0.39 | 1.66 |
| HILIC_NEG_243_76511a1_407 | | 243.7651 | 1.41 | | HILIC | NEG | | | 182691 | 118744 | 148276 | 59452.3 | 6.51E-03 | -0.39 | 1.44 |
| HILIC_NEG_146_04604a6_383 | Glutamic acid | 146.0460 | 6.38 | HMDB0000148 | HILIC | NEG | 102.0559(100), 128.0353(35), 146.0454(20) | 1 | 850750 | 343711 | 1008245 | 473496 | 6.54E-03 | 0.39 | 1.62 |
| RP_POS_333_16669a8_246 | N-Methylnuciferine | 333.1667 | 8.25 | HMDB0242040 | RP | POS | 333.1701(100) | 2 | 2300357 | 109482 | 2258225 | 114692 | 6.56E-03 | -0.38 | 1.6 |
| RP_POS_669_49347a11_776 | | 669.4935 | 11.78 | | RP | POS | | | 939611 | 389022 | 809214 | 308690 | 6.56E-03 | -0.37 | 1.59 |
| HILIC_POS_166_08366a3_422 | Phenylalanine | 166.0837 | 3.42 | HMDB0250791 | HILIC | POS | 166.0833(100), 120.0825(2) | 2 | 5607017 | 7730916 | 3215844 | 4955459 | 6.60E-03 | -0.38 | 1.45 |
| RP_NEG_740_29083a5_322 | | 740.2908 | 5.32 | | RP | NEG | | | 302994 | 92102.7 | 265235 | 107513 | 6.62E-03 | -0.38 | 1.54 |
| HILIC_POS_183_11285a0_745 | | 183.1129 | 0.75 | | HILIC | POS | | | 1082218 | 641272 | 892260 | 355522 | 6.66E-03 | -0.38 | 1.44 |
| RP_POS_413_28696a10_656 | | 413.2870 | 10.66 | | RP | POS | | | 576130 | 220726 | 707835 | 430535 | 6.69E-03 | 0.4 | 1.67 |
| RP_NEG_300_98969a6_63 | | 300.9897 | 6.63 | | RP | NEG | | | 2542015 | 156368 | 2482797 | 159894 | 6.69E-03 | -0.37 | 1.57 |
| HILIC_POS_227_12489a1_218 | | 227.1249 | 1.22 | | HILIC | POS | | | 1.2E+08 | 2.3E+07 | 1.2E+08 | 2.2E+07 | 6.70E-03 | -0.37 | 1.61 |
| RP_NEG_130_94437a11_238 | | 130.9444 | 11.24 | | RP | NEG | | | 4859089 | 1312346 | 5348473 | 1306570 | 6.73E-03 | 0.37 | 1.72 |
| HILIC_POS_148_06049a6_385 | Glutamic acid | 148.0605 | 6.39 | HMDB0000148 | HILIC | POS | 84.045(100), 102.059(40) | 2 | 2794795 | 1140174 | 3323116 | 1609499 | 6.86E-03 | 0.38 | 1.62 |
| RP_NEG_733_54999a12_511 | | 733.5500 | 12.51 | | RP | NEG | | | 4165651 | 815188 | 3889283 | 673816 | 6.91E-03 | -0.37 | 1.55 |
| RP_POS_302_10211a5_983 | | 302.1021 | 5.98 | | RP | POS | | | 402086 | 411002 | 275081 | 267405 | 6.92E-03 | -0.37 | 1.51 |
| HILIC_NEG_360_69406a1_41 | | 360.6941 | 1.41 | | HILIC | NEG | | | 138485 | 55611.8 | 121399 | 35632.8 | 6.96E-03 | -0.37 | 1.44 |
| RP_POS_276_1991a7_944 | | 276.1991 | 7.94 | | RP | POS | | | 304452 | 147868 | 371350 | 202406 | 6.99E-03 | 0.38 | 1.68 |
| RP_POS_299_13895a6_887 | | 299.1390 | 6.89 | | RP | POS | | | 657343 | 613858 | 462691 | 432440 | 7.02E-03 | -0.37 | 1.5 |
| RP_NEG_367_15909a8_007 | | 367.1591 | 8.01 | | RP | NEG | 367.0563(100), 96.9601(70) | | 2025949 | 1106859 | 2452595 | 1182613 | 7.09E-03 | 0.37 | 1.77 |
| RP_POS_266_63901a10_104 | | 266.6390 | 10.10 | | RP | POS | | | 409177 | 275263 | 532496 | 373134 | 7.16E-03 | 0.38 | 1.74 |
| RP_POS_319_12866a5_671 | | 319.1287 | 5.67 | | RP | POS | | | 359083 | 360054 | 247753 | 238083 | 7.18E-03 | -0.37 | 1.48 |
| HILIC_NEG_119_03487a5_119 | | 119.0349 | 5.12 | | HILIC | NEG | | | 6106378 | 1916618 | 6944076 | 2508348 | 7.19E-03 | 0.38 | 1.58 |
| HILIC_POS_136_04813a5_587 | | 136.0481 | 5.59 | | HILIC | POS | | | 2663952 | 822033 | 2973504 | 846555 | 7.22E-03 | 0.37 | 1.53 |
| HILIC_POS_516_30737a1_282 | | 516.3074 | 1.28 | | HILIC | POS | | | 261403 | 116959 | 308009 | 132867 | 7.27E-03 | 0.37 | 1.53 |
| RP_NEG_195_811a0_566 | | 195.8110 | 0.57 | | RP | NEG | | | 7.3E+07 | 2.6E+07 | 6.2E+07 | 2.9E+07 | 7.29E-03 | -0.37 | 1.58 |
| RP_POS_265_14078a7_212 | 9-Hydroxyparthenolide | 265.1408 | 7.21 | CID: 5477074 | RP | POS | 265.1409(100) | 2 | 1216449 | 379856 | 1088352 | 315415 | 7.32E-03 | -0.37 | 1.47 |
| RP_POS_132_07693a0_559 | | 132.0769 | 0.56 | | RP | POS | | | 5.2E+07 | 1.7E+07 | 5.9E+07 | 1.8E+07 | 7.38E-03 | 0.37 | 1.5 |
| RP_POS_313_27322a11_601 | | 313.2732 | 11.60 | | RP | POS | | | 1145408 | 671082 | 1421561 | 806088 | 7.42E-03 | 0.37 | 1.57 |
| HILIC_POS_263_14621a5_588 | | 263.1462 | 5.59 | | HILIC | POS | | | 1022136 | 697916 | 1315710 | 870190 | 7.54E-03 | 0.37 | 1.52 |
| RP_POS_672_46454a10_403 | | 672.4645 | 10.40 | | RP | POS | | | 506099 | 276689 | 415527 | 216374 | 7.54E-03 | -0.37 | 1.61 |
| RP_POS_396_31064a10_649 | | 396.3106 | 10.65 | | RP | POS | | | 245935 | 188920 | 349255 | 339241 | 7.81E-03 | 0.39 | 1.76 |
| RP_POS_379_28406a10_651 | | 379.2841 | 10.65 | | RP | POS | | | 623453 | 378026 | 833697 | 693506 | 7.83E-03 | 0.39 | 1.73 |
| RP_POS_342_26373a7_85 | AC 12:0 (Dodecanoylcarnitine) | 342.2637 | 7.85 | HMDB0002250 | RP | POS | 85.0292(100), 342.2648(40) | 2 | 2715085 | 2149388 | 3704021 | 3084684 | 7.96E-03 | 0.38 | 1.62 |
| HILIC_NEG_391_6662a1_409 | | 391.6662 | 1.41 | | HILIC | NEG | | | 291857 | 99700.7 | 260478 | 72097.5 | 8.03E-03 | -0.37 | 1.44 |
| RP_POS_632_52484a13_602 | | 632.5248 | 13.60 | | RP | POS | | | 544714 | 850845 | 305865 | 420704 | 8.19E-03 | -0.38 | 1.48 |
| RP_NEG_744_29388a10_525 | | 744.2939 | 10.53 | | RP | NEG | | | 943996 | 54166.8 | 922606 | 62195.8 | 8.21E-03 | -0.37 | 1.51 |
| HILIC_POS_260_14914a2_298 | | 260.1491 | 2.30 | | HILIC | POS | | | 215201 | 256126 | 393497 | 618043 | 8.29E-03 | 0.41 | 1.61 |
| RP_NEG_413_20065a6_407 | | 413.2007 | 6.41 | | RP | NEG | | | 277419 | 107787 | 324454 | 144591 | 8.30E-03 | 0.37 | 1.56 |
| RP_POS_647_51312a11_774 | | 647.5131 | 11.77 | | RP | POS | | | 5705639 | 1939249 | 5079455 | 1515223 | 8.36E-03 | -0.36 | 1.48 |
| HILIC_NEG_115_92094a0_933 | | 115.9209 | 0.93 | | HILIC | NEG | | | 4E+07 | 3873802 | 3.9E+07 | 4033945 | 8.45E-03 | -0.36 | 1.64 |
| HILIC_POS_154_05878a5_587 | | 154.0588 | 5.59 | | HILIC | POS | | | 6167482 | 1950883 | 6882366 | 1990710 | 8.57E-03 | 0.36 | 1.5 |
| RP_NEG_255_11398a6_255 | | 255.1140 | 6.26 | | RP | NEG | | | 481046 | 383359 | 367013 | 241544 | 8.59E-03 | -0.36 | 1.42 |
| RP_NEG_495_29694a8_971 | | 495.2969 | 8.97 | | RP | NEG | | | 1.3E+07 | 4924590 | 1.1E+07 | 5730951 | 8.63E-03 | -0.37 | 1.49 |
| HILIC_POS_426_35754a0_6 | | 426.3575 | 0.60 | | HILIC | POS | | | 582733 | 206314 | 659837 | 218457 | 8.65E-03 | 0.36 | 1.67 |
| RP_POS_754_53485a13_039 | | 754.5349 | 13.04 | | RP | POS | | | 2E+07 | 9374946 | 2.4E+07 | 1.4E+07 | 8.66E-03 | 0.38 | 1.6 |
| RP_NEG_490_16574a10_926 | | 490.1657 | 10.93 | | RP | NEG | | | 236207 | 73253.7 | 264285 | 80997.1 | 8.66E-03 | 0.36 | 1.64 |
| RP_NEG_946_57996a13_769 | | 946.5800 | 13.77 | | RP | NEG | | | 1661910 | 994963 | 1335176 | 818764 | 8.72E-03 | -0.36 | 1.5 |
| RP_POS_301_11813a5_859 | | 301.1181 | 5.86 | | RP | POS | | | 790735 | 728569 | 571509 | 478782 | 8.73E-03 | -0.36 | 1.45 |

**Supplemantary table 1**: The non-targeted metabolomics results ordered by the VIP value (p value < 0.0003 are shaded dark green, p value < 0.05 are shaded light green, d values below 0 are blue and above 0 are red)

| | | | | | | | | | Cases | | Controls | | Welch test | Cohen's | |
|---|---|---|---|---|---|---|---|---|---|---|---|---|---|---|---|
| Feature_ID | Metabolite_name | Average_Mz | Average_RT_min_ | HMDB ID | RP_HILIC | POS_NEG | Cureted_MSMS | ID_level | Mean | SD | Mean2 | SD3 | p | d | VIP |
| RP_NEG_285_20694a9_555 | | 285.2069 | 9.56 | | RP | NEG | | | 1140304 | 125405 | 1098336 | 107797 | 8.78E-03 | -0.36 | 1.54 |
| RP_NEG_185_05661a0_679 | | 185.0566 | 0.68 | | RP | NEG | | | 551096 | 291171 | 686919 | 435531 | 9.00E-03 | 0.37 | 1.7 |
| HILIC_POS_148_00369a5_351 | | 148.0037 | 5.35 | | HILIC | POS | | | 533864 | 142162 | 581145 | 121545 | 9.03E-03 | 0.36 | 1.44 |
| RP_POS_175_11185a9_345 | | 175.1119 | 9.35 | | RP | POS | | | 1317914 | 433885 | 1492993 | 526623 | 9.07E-03 | 0.36 | 1.54 |
| RP_POS_497_39627a13_292 | | 497.3963 | 13.29 | | RP | POS | | | 767286 | 400178 | 959060 | 621838 | 9.09E-03 | 0.38 | 1.77 |
| RP_POS_291_12067a8_826 | | 291.1207 | 8.83 | | RP | POS | | | 2286753 | 912041 | 1989518 | 746959 | 9.09E-03 | -0.36 | 1.51 |
| RP_NEG_271_22833a10_641 | Hydroxyhexadecanoic acid | 271.2283 | 10.64 | HMDB0006294 | RP | NEG | 271.228(100), 225.223(65) | 2 | 6174889 | 710535 | 5919437 | 711464 | 9.15E-03 | -0.36 | 1.55 |
| RP_NEG_344_97974a8_173 | | 344.9797 | 8.17 | | RP | NEG | | | 1393336 | 217379 | 1313723 | 225040 | 9.15E-03 | -0.36 | 1.79 |
| RP_POS_413_36279a11_138 | | 413.3628 | 11.14 | | RP | POS | | | 527741 | 96562.7 | 492702 | 98393.1 | 9.18E-03 | -0.36 | 1.49 |
| RP_NEG_440_14996a10_807 | | 440.1500 | 10.81 | | RP | NEG | | | 184069 | 148712 | 239607 | 159636 | 9.23E-03 | 0.36 | 1.66 |
| HILIC_NEG_104_03534a6_177 | Serine | 104.0353 | 6.18 | HMDB0000187 | HILIC | NEG | 74.0239(100), 104.0353(50) | 1 | 816761 | 181118 | 882254 | 183769 | 9.26E-03 | 0.36 | 1.47 |
| RP_POS_149_13263a10_655 | | 149.1326 | 10.66 | | RP | POS | | | 578777 | 256654 | 677772 | 291219 | 9.28E-03 | 0.36 | 1.68 |
| HILIC_NEG_180_84236a1_41 | | 180.8424 | 1.41 | | HILIC | NEG | | | 1.5E+07 | 3488344 | 1.4E+07 | 1928585 | 9.29E-03 | -0.37 | 1.4 |
| RP_POS_360_36246a9_417 | | 360.3625 | 9.42 | | RP | POS | | | 5433706 | 2238584 | 6587073 | 3841069 | 9.33E-03 | 0.38 | 1.67 |
| RP_POS_315_13416a7_247 | | 315.1342 | 7.25 | | RP | POS | | | 398146 | 301675 | 302938 | 231302 | 9.37E-03 | -0.36 | 1.47 |
| RP_POS_351_25034a10_455 | | 351.2503 | 10.46 | | RP | POS | | | 325815 | 595924 | 847768 | 1887772 | 9.40E-03 | 0.42 | 1.6 |
| RP_NEG_313_11948a7_625 | | 313.1195 | 7.63 | | RP | NEG | | | 126876 | 80745.9 | 103252 | 50147.3 | 9.40E-03 | -0.36 | 1.48 |
| RP_NEG_876_57678a13_133 | | 876.5768 | 13.13 | | RP | NEG | | | 4776456 | 1505530 | 4261929 | 1377608 | 9.40E-03 | -0.36 | 1.52 |
| HILIC_NEG_178_84428a1_409 | | 178.8443 | 1.41 | | HILIC | NEG | | | 1.6E+07 | 3509727 | 1.5E+07 | 1932604 | 9.44E-03 | -0.36 | 1.4 |
| RP_NEG_460_18192a9_97 | | 460.1819 | 9.97 | | RP | NEG | | | 252596 | 120293 | 215292 | 88624.5 | 9.48E-03 | -0.36 | 1.45 |
| RP_POS_433_33237a9_021 | | 433.3324 | 9.02 | | RP | POS | | | 365941 | 92830.1 | 335002 | 80846.9 | 9.48E-03 | -0.36 | 1.44 |
| RP_POS_308_12552a8_397 | | 308.1255 | 8.40 | | RP | POS | 308.1252(100) | | 1492314 | 1836525 | 951675 | 1178949 | 9.70E-03 | -0.36 | 1.41 |
| RP_POS_388_30588a8_503 | | 388.3059 | 8.50 | | RP | POS | | | 412431 | 178895 | 489734 | 244134 | 9.76E-03 | 0.37 | 1.64 |
| HILIC_POS_172_13313a3_104 | | 172.1331 | 3.10 | | HILIC | POS | | | 488455 | 210484 | 571274 | 249474 | 9.78E-03 | 0.36 | 1.69 |
| RP_POS_329_15674a5_969 | | 329.1567 | 5.97 | | RP | POS | | | 486119 | 257163 | 412742 | 149600 | 9.81E-03 | -0.36 | 1.41 |
| HILIC_POS_58_06567a4_795 | | 58.0657 | 4.80 | | HILIC | POS | | | 3046402 | 473208 | 3212860 | 463227 | 9.83E-03 | 0.36 | 1.79 |
| HILIC_NEG_176_84959a1_408 | | 176.8496 | 1.41 | | HILIC | NEG | | | 713573 | 167746 | 666305 | 93571.9 | 9.89E-03 | -0.36 | 1.4 |
| RP_NEG_563_28448a8_971 | | 563.2845 | 8.97 | | RP | NEG | | | 948068 | 363068 | 806744 | 425833 | 1.01E-02 | -0.36 | 1.45 |
| RP_POS_337_21353a9_041 | | 337.2135 | 9.04 | | RP | POS | | | 1324546 | 451696 | 1114872 | 685942 | 1.01E-02 | -0.37 | 1.5 |
| RP_POS_151_07545a7_214 | | 151.0755 | 7.21 | | RP | POS | | | 1357374 | 450588 | 1211522 | 377842 | 1.03E-02 | -0.35 | 1.41 |
| HILIC_POS_323_21732a4_773 | | 323.2173 | 4.77 | | HILIC | POS | | | 380149 | 180062 | 466843 | 288572 | 1.04E-02 | 0.37 | 1.67 |
| RP_POS_324_6452a5_518 | | 324.6452 | 5.52 | | RP | POS | | | 145603 | 69361 | 173117 | 84385.5 | 1.04E-02 | 0.36 | 1.54 |
| RP_NEG_435_25052a8_798 | | 435.2505 | 8.80 | | RP | NEG | | | 228404 | 127345 | 184979 | 119159 | 1.04E-02 | -0.35 | 1.43 |
| RP_POS_372_08737a8_842 | | 372.0874 | 8.84 | | RP | POS | | | 210093 | 138844 | 169171 | 92091 | 1.04E-02 | -0.35 | 1.38 |
| RP_POS_427_37772a12_31 | | 427.3777 | 12.31 | | RP | POS | | | 1174750 | 138996 | 1227954 | 160086 | 1.05E-02 | 0.36 | 1.52 |
| HILIC_POS_282_1196a4_435 | 1-Methyladenosine | 282.1196 | 4.44 | HMDB0011599 | HILIC | POS | 150.0778(100), 282.12(5) | 2 | 2612336 | 291359 | 2519167 | 239046 | 1.05E-02 | -0.35 | 1.41 |
| HILIC_POS_553_40833a5_716 | | 553.4083 | 5.72 | | HILIC | POS | | | 107424 | 107240 | 71282.2 | 98323.3 | 1.05E-02 | -0.35 | 1.48 |
| RP_POS_633_25519a8_455 | | 633.2552 | 8.46 | | RP | POS | | | 1139335 | 1121098 | 812165 | 723574 | 1.05E-02 | -0.35 | 1.52 |
| RP_POS_459_24631a8_8 | | 459.2463 | 8.80 | | RP | POS | | | 541782 | 309463 | 434290 | 301976 | 1.06E-02 | -0.35 | 1.45 |
| HILIC_NEG_100_9336a0_934 | | 100.9336 | 0.93 | | HILIC | NEG | | | 3.2E+07 | 2811283 | 3.1E+07 | 2749543 | 1.07E-02 | -0.35 | 1.48 |
| HILIC_NEG_415_21646a0_638 | | 415.2165 | 0.64 | | HILIC | NEG | | | 133552 | 90696.7 | 105084 | 71750 | 1.07E-02 | -0.35 | 1.43 |
| RP_POS_430_25037a13 | | 430.2504 | 13.00 | | RP | POS | | | 216803 | 187116 | 162547 | 119384 | 1.07E-02 | -0.35 | 1.42 |
| RP_POS_829_55371a13_004 | | 829.5537 | 13.00 | | RP | POS | | | 9.4E+07 | 3.3E+07 | 8.1E+07 | 3.5E+07 | 1.07E-02 | -0.35 | 1.54 |
| RP_NEG_281_24805a9_537 | | 281.2481 | 9.54 | | RP | NEG | | | 266074 | 98355.5 | 300274 | 96563 | 1.08E-02 | 0.35 | 1.65 |
| RP_NEG_656_31836a10_292 | | 656.3184 | 10.29 | | RP | NEG | | | 2197095 | 402119 | 2349749 | 460338 | 1.08E-02 | 0.35 | 1.51 |
| RP_POS_211_14452a4_634 | Cyclo(Leu-Pro) | 211.1445 | 4.63 | HMDB0094673 | RP | POS | 211.1415(100), 70.0659(30) | 2 | 4108792 | 1395795 | 4835559 | 2487852 | 1.08E-02 | 0.37 | 1.59 |
| RP_NEG_392_19348a9_957 | | 392.1935 | 9.96 | | RP | NEG | | | 1134217 | 427062 | 1003359 | 319192 | 1.08E-02 | -0.35 | 1.39 |
| RP_NEG_286_97638a7_589 | | 286.9764 | 7.59 | | RP | NEG | | | 962141 | 162911 | 907391 | 150058 | 1.09E-02 | -0.35 | 1.47 |
| RP_NEG_918_54865a13_005 | | 918.5487 | 13.01 | | RP | NEG | | | 1.1E+07 | 1842802 | 1E+07 | 1513901 | 1.09E-02 | -0.35 | 1.57 |
| RP_POS_349_27115a10_626 | | 349.2712 | 10.63 | | RP | POS | | | 964740 | 203412 | 896086 | 188900 | 1.09E-02 | -0.35 | 1.68 |
| HILIC_POS_330_2272a0_974 | | 330.2272 | 0.97 | | HILIC | POS | | | 173114 | 159709 | 127089 | 101179 | 1.10E-02 | -0.35 | 1.37 |
| RP_NEG_350_15186a4_76 | | 350.1519 | 4.76 | | RP | NEG | | | 786341 | 267037 | 895584 | 345432 | 1.11E-02 | 0.36 | 1.82 |
| RP_NEG_432_16068a10_925 | | 432.1607 | 10.93 | | RP | NEG | | | 194957 | 63381.8 | 217360 | 64546.4 | 1.11E-02 | 0.35 | 1.57 |
| HILIC_POS_186_11247a1_826 | | 186.1125 | 1.83 | | HILIC | POS | | | 403156 | 881038 | 179092 | 310148 | 1.12E-02 | -0.38 | 1.33 |
| RP_POS_508_37518a11_106 | LysoPC O-18:1 (O-18:1/0:0) | 508.3752 | 11.11 | | RP | POS | 104.1077(100), 508.374(90) | 1 | 2049822 | 820753 | 1792685 | 659076 | 1.14E-02 | -0.35 | 1.41 |
| HILIC_POS_302_19562a1_512 | | 302.1956 | 1.51 | | HILIC | POS | | | 2679886 | 1266793 | 2267217 | 1109242 | 1.14E-02 | -0.35 | 1.46 |
| RP_NEG_583_25696a12_084 | | 583.2570 | 12.08 | | RP | NEG | | | 380550 | 803559 | 169977 | 351655 | 1.14E-02 | -0.36 | 1.35 |
| RP_POS_597_42596a10_646 | | 597.4260 | 10.65 | | RP | POS | | | 224370 | 146143 | 289685 | 216205 | 1.16E-02 | 0.36 | 1.48 |
| HILIC_POS_182_05766a3_413 | | 182.0577 | 3.41 | | HILIC | POS | | | 1605954 | 2202603 | 963659 | 1482175 | 1.16E-02 | -0.35 | 1.37 |
| RP_POS_609_43646a10_134 | | 609.4365 | 10.13 | | RP | POS | | | 8941626 | 2957610 | 7841403 | 3332749 | 1.17E-02 | -0.35 | 1.58 |
| RP_NEG_397_20541a8_432 | | 397.2054 | 8.43 | | RP | NEG | | | 6627808 | 4545648 | 5330067 | 2886362 | 1.18E-02 | -0.35 | 1.36 |
| RP_NEG_786_56671a13_577 | | 786.5667 | 13.58 | | RP | NEG | | | 5366265 | 1417142 | 4917690 | 1178527 | 1.18E-02 | -0.35 | 1.48 |
| RP_POS_803_54303a11_408 | | 803.5430 | 11.41 | | RP | POS | | | 6271773 | 1291958 | 5888991 | 914108 | 1.18E-02 | -0.35 | 1.49 |
| RP_POS_524_2735a10_293 | | 524.2735 | 10.29 | | RP | POS | | | 4015791 | 767325 | 4312164 | 922445 | 1.19E-02 | 0.35 | 1.61 |
| HILIC_NEG_263_10349a1_895 | Phenylacetylglutamine | 263.1035 | 1.90 | | HILIC | NEG | | | 728929 | 471309 | 579087 | 396667 | 1.19E-02 | -0.35 | 1.41 |
| RP_POS_202_18036a0_679 | | 202.1804 | 0.68 | | RP | POS | | | 3115562 | 1655169 | 2564660 | 1534767 | 1.19E-02 | -0.35 | 1.46 |
| RP_POS_312_25253a10_251 | | 312.2525 | 10.25 | | RP | POS | | | 403317 | 85201.1 | 435534 | 99103 | 1.20E-02 | 0.35 | 1.56 |
| RP_POS_519_29279a8_971 | | 519.2928 | 8.97 | | RP | POS | | | 3329331 | 1237471 | 2851702 | 1491623 | 1.21E-02 | -0.35 | 1.43 |
| RP_POS_160_09686a0_545 | | 160.0969 | 0.55 | | RP | POS | | | 2308188 | 4845514 | 1044241 | 2171212 | 1.21E-02 | -0.36 | 1.33 |
| RP_POS_302_19556a2_085 | | 302.1956 | 2.09 | | RP | POS | | | 437672 | 231744 | 361491 | 210889 | 1.22E-02 | -0.34 | 1.46 |
| RP_POS_645_49603a11_477 | | 645.4960 | 11.48 | | RP | POS | | | 938639 | 636283 | 758926 | 396501 | 1.22E-02 | -0.35 | 1.51 |
| RP_POS_343_19266a11_144 | | 343.1927 | 11.14 | | RP | POS | | | 5822555 | 918548 | 5514423 | 871637 | 1.23E-02 | -0.34 | 1.46 |
| HILIC_NEG_278_76974a1_406 | | 278.7697 | 1.41 | | HILIC | NEG | | | 189146 | 39684 | 177552 | 27416.5 | 1.23E-02 | -0.35 | 1.4 |
| RP_NEG_411_18481a6_703 | | 411.1848 | 6.70 | | RP | NEG | | | 406058 | 212922 | 481093 | 221947 | 1.24E-02 | 0.35 | 1.45 |
| HILIC_POS_158_11771a3_021 | Homostachydrine (Pipecolic acid betaine) | 158.1177 | 3.02 | HMDB0033433 | HILIC | POS | 158.1182(100) | 1 | 3735765 | 2326293 | 3063326 | 1565971 | 1.24E-02 | -0.35 | 1.35 |
| RP_POS_524_70874a10_337 | | 524.7087 | 10.34 | | RP | POS | | | 663261 | 433201 | 841950 | 580690 | 1.25E-02 | 0.35 | 1.48 |
| RP_POS_585_27112a7_237 | | 585.2711 | 7.24 | | RP | POS | | | 2.6E+07 | 2.2E+07 | 2E+07 | 1.3E+07 | 1.26E-02 | -0.35 | 1.37 |
| HILIC_POS_285_12863a5_581 | | 285.1286 | 5.58 | | HILIC | POS | | | 484769 | 327098 | 609636 | 391525 | 1.27E-02 | 0.35 | 1.43 |
| HILIC_POS_182_07886a4_569 | Tyrosine | 182.0789 | 4.57 | HMDB0011741 | HILIC | POS | 182.0794(100) | 2 | 1444398 | 3419327 | 564516 | 1474905 | 1.27E-02 | -0.36 | 1.32 |
| RP_POS_639_49567a14_21 | | 639.4957 | 14.21 | | RP | POS | | | 3580325 | 2331881 | 2885945 | 1716562 | 1.27E-02 | -0.34 | 1.41 |
| RP_POS_352_16489a4_763 | | 352.1649 | 4.76 | | RP | POS | | | 4983806 | 1679909 | 5662713 | 2197673 | 1.28E-02 | 0.35 | 1.81 |
| RP_NEG_602_30188a10_278 | | 602.3019 | 10.28 | | RP | NEG | | | 348920 | 91516.2 | 316956 | 94622.2 | 1.28E-02 | -0.34 | 1.55 |
| RP_POS_587_42426a10_153 | | 587.4243 | 10.15 | | RP | POS | | | 9580077 | 3710781 | 8373975 | 3351821 | 1.29E-02 | -0.34 | 1.5 |
| RP_NEG_279_09897a3_038 | | 279.0990 | 3.04 | | RP | NEG | | | 225952 | 104448 | 267110 | 131947 | 1.30E-02 | 0.35 | 1.57 |
| HILIC_NEG_387_67175a1_41 | | 387.6718 | 1.41 | | HILIC | NEG | | | 105196 | 36184.5 | 94239.1 | 28075.4 | 1.30E-02 | -0.34 | 1.38 |
| RP_POS_285_25772a8_971 | | 285.2577 | 8.97 | | RP | POS | | | 5073878 | 1982914 | 4344946 | 2247448 | 1.30E-02 | -0.34 | 1.42 |
| RP_POS_282_27905a9_053 | | 282.2791 | 9.05 | | RP | POS | | | 368805 | 652148 | 866288 | 1871999 | 1.31E-02 | 0.39 | 1.63 |
| RP_NEG_383_18924a11_592 | | 383.1892 | 11.59 | | RP | NEG | | | 4.4E+07 | 2544140 | 4.3E+07 | 3029974 | 1.31E-02 | -0.35 | 1.62 |
| HILIC_NEG_162_9827a0_934 | | 162.9827 | 0.93 | | HILIC | NEG | 118.9923(100), 162.9984(30) | | 1437809 | 151612 | 1391374 | 121410 | 1.32E-02 | -0.34 | 1.36 |
| RP_NEG_197_808a0_573 | | 197.8080 | 0.57 | | RP | NEG | | | 8.9E+07 | 3.5E+07 | 7.7E+07 | 3.5E+07 | 1.32E-02 | -0.34 | 1.38 |
| HILIC_NEG_182_84053a1_408 | | 182.8405 | 1.41 | | HILIC | NEG | | | 363006 | 85825.7 | 339604 | 49295 | 1.32E-02 | -0.35 | 1.33 |
| RP_POS_756_59088a13_831 | | 756.5909 | 13.83 | | RP | POS | | | 538845 | 551072 | 390145 | 304940 | 1.32E-02 | -0.35 | 1.38 |
| HILIC_POS_258_11035a6_217 | Glycerophosphocholine | 258.1104 | 6.22 | | HILIC | POS | 104.1077(100), 124.999(15) | 2 | 3.5E+07 | 1.4E+07 | 3E+07 | 1.5E+07 | 1.33E-02 | -0.34 | 1.77 |
| RP_POS_240_10136a2_053 | | 240.1014 | 2.05 | | RP | POS | | | 280556 | 120895 | 326490 | 145309 | 1.33E-02 | 0.35 | 1.42 |
| RP_POS_214_00487a0_74 | | 214.0049 | 0.74 | | RP | POS | | | 999017 | 370274 | 1153236 | 510754 | 1.34E-02 | 0.35 | 1.47 |
| RP_NEG_401_14868a10_807 | | 401.1487 | 10.81 | | RP | NEG | | | 336281 | 300574 | 445443 | 336024 | 1.34E-02 | 0.34 | 1.6 |
| RP_POS_720_98962a10_523 | | 720.9896 | 10.52 | | RP | POS | | | 532340 | 303038 | 437455 | 256151 | 1.34E-02 | -0.34 | 1.46 |
| RP_NEG_415_32248a10_72 | | 415.3225 | 10.72 | | RP | NEG | | | 176390 | 57792.6 | 198779 | 71760.9 | 1.35E-02 | 0.35 | 1.44 |
| RP_NEG_344_97916a7_279 | | 344.9792 | 7.28 | | RP | NEG | | | 2042777 | 57380.3 | 2061229 | 51391.9 | 1.35E-02 | 0.34 | 1.41 |
| RP_POS_302_23282a6_464 | | 302.2328 | 6.46 | | RP | POS | | | 2046669 | 1556502 | 1574915 | 1223451 | 1.35E-02 | -0.34 | 1.37 |
| RP_POS_323_25537a10_924 | | 323.2554 | 10.92 | | RP | POS | | | 4529533 | 843385 | 4257223 | 762238 | 1.35E-02 | -0.34 | 1.65 |
| HILIC_NEG_311_14017a1_723 | | 311.1402 | 1.72 | | HILIC | NEG | | | 2447659 | 873449 | 2793555 | 1123603 | 1.36E-02 | 0.35 | 1.63 |
| RP_POS_223_06355a8_97 | | 223.0636 | 8.97 | | RP | POS | | | 995670 | 164524 | 1058868 | 202053 | 1.36E-02 | 0.34 | 1.46 |
| HILIC_NEG_280_76706a1_406 | | 280.7671 | 1.41 | | HILIC | NEG | | | 197137 | 40155.8 | 185981 | 24972.7 | 1.36E-02 | -0.34 | 1.37 |
| RP_NEG_325_27539a10_624 | | 325.2754 | 10.62 | | RP | NEG | | | 474718 | 99763.9 | 443026 | 87380 | 1.36E-02 | -0.34 | 1.76 |
| RP_POS_138_98338a15_488 | | 138.9834 | 15.49 | | RP | POS | | | 316768 | 42277.5 | 303217 | 37730.5 | 1.36E-02 | -0.34 | 1.43 |
| RP_POS_337_2337a10_635 | | 337.2337 | 10.64 | | RP | POS | | | 1614010 | 213807 | 1543622 | 201135 | 1.36E-02 | -0.34 | 1.42 |
| RP_POS_1198_35645a13_462 | | 1198.3565 | 13.46 | | RP | POS | | | 1252582 | 948913 | 1683526 | 1483308 | 1.37E-02 | 0.35 | 1.49 |
| HILIC_POS_170_03242a5_588 | | 170.0324 | 5.59 | | HILIC | POS | | | 1681379 | 451492 | 1837012 | 464267 | 1.37E-02 | 0.34 | 1.43 |
| HILIC_POS_265_11853a1_892 | Phenylacetylglutamine | 265.1185 | 1.89 | HMDB0006344 | HILIC | POS | 130.0501(100), 84.045(20) | 2 | 4398792 | 3023747 | 3458847 | 2536088 | 1.37E-02 | -0.34 | 1.37 |
| RP_NEG_280_99301a4_771 | | 280.9930 | 4.77 | | RP | NEG | | | 368395 | 86865.1 | 334746 | 108392 | 1.38E-02 | -0.34 | 1.44 |
| RP_POS_315_23172a9_041 | Progesterone | 315.2317 | 9.04 | HMDB0001830 | RP | POS | 97.0656(100), 315.2334(75) | 2 | 9732983 | 3365835 | 8246252 | 5079839 | 1.39E-02 | -0.35 | 1.45 |
| RP_POS_822_63623a14_887 | | 822.6362 | 14.89 | | RP | POS | | | 1111618 | 698501 | 1423360 | 1070754 | 1.40E-02 | 0.35 | 1.72 |
| HILIC_NEG_196_93024a6_175 | | 196.9302 | 6.18 | | HILIC | NEG | | | 122563 | 21452.5 | 129493 | 19626 | 1.40E-02 | 0.34 | 1.41 |
| RP_NEG_417_18045a11_361 | | 417.1805 | 11.36 | | RP | NEG | | | 698458 | 75608.5 | 671413 | 83282.9 | 1.40E-02 | -0.34 | 1.73 |
| HILIC_POS_136_04825a1_217 | | 136.0483 | 1.22 | | HILIC | POS | | | 1.1E+07 | 1991348 | 1.1E+07 | 1924382 | 1.41E-02 | -0.34 | 1.5 |
| RP_NEG_606_32373a13_141 | | 606.3237 | 13.14 | | RP | NEG | | | 316602 | 48364.4 | 299880 | 50402.9 | 1.41E-02 | -0.34 | 1.44 |
| HILIC_POS_128_07076a1_826 | | 128.0708 | 1.83 | | HILIC | POS | | | 402806 | 712391 | 223680 | 296094 | 1.42E-02 | -0.36 | 1.29 |
| RP_NEG_414_06406a4_816 | | 414.0641 | 4.82 | | RP | NEG | | | 685714 | 233645 | 775491 | 290565 | 1.43E-02 | 0.34 | 1.67 |
| HILIC_POS_280_09155a6_205 | | 280.0916 | 6.21 | | HILIC | POS | | | 445757 | 192526 | 379013 | 202200 | 1.43E-02 | -0.34 | 1.73 |
| RP_POS_273_15308a9_209 | | 273.1531 | 9.21 | | RP | POS | | | 309763 | 362431 | 213138 | 200254 | 1.43E-02 | -0.34 | 1.41 |
| RP_POS_236_07162a3_804 | | 236.0716 | 3.80 | | RP | POS | | | 2.9E+07 | 1.5E+07 | 3.4E+07 | 1.7E+07 | 1.44E-02 | 0.34 | 1.4 |
| HILIC_POS_130_08646a3_7 | | 130.0865 | 3.70 | | HILIC | POS | 84.0816(100), 130.0869(80) | | 3.7E+07 | 6E+07 | 2.2E+07 | 2.6E+07 | 1.45E-02 | -0.35 | 1.3 |
| RP_NEG_158_97878a14_348 | | 158.9788 | 14.35 | | RP | NEG | | | 2518465 | 1273762 | 2967047 | 1381750 | 1.46E-02 | 0.34 | 1.44 |
| RP_POS_1162_35547a13_8 | | 1162.3555 | 13.80 | | RP | POS | | | 3949490 | 1788075 | 4604373 | 2070921 | 1.46E-02 | 0.34 | 1.51 |
| HILIC_POS_229_15472a4_938 | | 229.1547 | 4.94 | | HILIC | POS | | | 4423230 | 1137748 | 4048205 | 1096667 | 1.46E-02 | -0.34 | 1.42 |
| RP_POS_285_00531a4_771 | | 285.0053 | 4.77 | | RP | POS | | | 1062306 | 321338 | 955698 | 313126 | 1.46E-02 | -0.34 | 1.44 |
| RP_POS_137_0961a10_653 | | 137.0961 | 10.65 | | RP | POS | | | 1343410 | 443988 | 1510329 | 537081 | 1.47E-02 | 0.34 | 1.56 |
| HILIC_NEG_195_81099a0_557 | | 195.8110 | 0.56 | | HILIC | NEG | | | 616539 | 284744 | 538113 | 178345 | 1.47E-02 | -0.34 | 1.35 |
| HILIC_POS_293_19095a1_024 | | 293.1910 | 1.02 | | HILIC | POS | | | 319283 | 156761 | 271516 | 128779 | 1.48E-02 | -0.33 | 1.39 |
| RP_NEG_773_58228a12_963 | | 773.5823 | 12.96 | | RP | NEG | | | 4643929 | 1143124 | 5021784 | 1114164 | 1.49E-02 | 0.33 | 1.39 |
| RP_POS_391_28412a11_408 | Dioctyl phthalate | 391.2841 | 11.41 | HMDB0251427 | RP | POS | 149.0232(100), 71.0855(10) | 1 | 1.2E+08 | 2.1E+07 | 1.1E+08 | 1.4E+07 | 1.49E-02 | -0.34 | 1.43 |
| RP_POS_796_07672a14_057 | | 796.0767 | 14.06 | | RP | POS | | | 724309 | 714562 | 986531 | 835366 | 1.50E-02 | 0.34 | 1.78 |
| RP_POS_247_14404a3_002 | Lenticin (Tryptophan betaine) | 247.1440 | 3.00 | HMDB0061115 | RP | POS | 188.0707(100), 146.0603(45) | 2 | 3088653 | 4838722 | 1844268 | 2375939 | 1.51E-02 | -0.34 | 1.32 |
| RP_NEG_295_22818a9_668 | | 295.2282 | 9.67 | | RP | NEG | | | 750356 | 852810 | 487773 | 723025 | 1.51E-02 | -0.33 | 1.4 |
| RP_POS_230_96657a15_356 | | 230.9666 | 15.36 | | RP | POS | | | 1539529 | 996749 | 1241461 | 790097 | 1.51E-02 | -0.33 | 1.55 |
| HILIC_POS_189_07307a5_928 | | 189.0731 | 5.93 | | HILIC | POS | | | 2543414 | 378174 | 2677458 | 419017 | 1.52E-02 | 0.34 | 1.48 |
| HILIC_POS_370_29535a0_62 | AC 14:1 (Tetradecenoylcarnitine) | 370.2954 | 0.62 | HMDB0258884 | HILIC | POS | 85.029(100), 370.2932(55) | 1 | 1807295 | 1188248 | 2382550 | 2059751 | 1.52E-02 | 0.35 | 1.6 |
| HILIC_POS_130_08652a4_976 | Pipecolinic acid | 130.0865 | 4.98 | HMDB0000138 | HILIC | POS | 84.0814(100), 130.0867(55) | 1 | 6968059 | 2555043 | 6140846 | 2404923 | 1.52E-02 | -0.33 | 1.34 |
| RP_POS_388_3938a9_797 | | 388.3938 | 9.80 | | RP | POS | | | 6723974 | 2761162 | 7973991 | 4394647 | 1.54E-02 | 0.35 | 1.58 |
| RP_NEG_263_10385a3_647 | | 263.1039 | 3.65 | | RP | NEG | | | 4784638 | 3023467 | 3853061 | 2579927 | 1.54E-02 | -0.33 | 1.36 |
| RP_POS_218_91017a15_387 | | 218.9102 | 15.39 | | RP | POS | | | 498557 | 323514 | 408608 | 213509 | 1.54E-02 | -0.33 | 1.59 |
| RP_NEG_578_30206a10_293 | | 578.3021 | 10.29 | | RP | NEG | | | 873877 | 175144 | 936083 | 195847 | 1.55E-02 | 0.34 | 1.42 |
| RP_POS_197_1286a3_677 | | 197.1286 | 3.68 | | RP | POS | | | 3655187 | 2033454 | 4504012 | 2905460 | 1.55E-02 | 0.34 | 1.38 |
| RP_POS_413_29703a8_583 | | 413.2970 | 8.58 | | RP | POS | | | 190829 | 182120 | 143571 | 94841.3 | 1.55E-02 | -0.34 | 1.29 |
| RP_POS_379_2818a10_851 | | 379.2818 | 10.85 | | RP | POS | | | 2502512 | 1730907 | 3405929 | 3307631 | 1.56E-02 | 0.36 | 1.61 |
| RP_POS_289_88095a15_386 | | 289.8810 | 15.39 | | RP | POS | | | 154127 | 86998.1 | 188872 | 116817 | 1.56E-02 | 0.34 | 1.8 |
| HILIC_POS_378_17648a1_326 | | 378.1765 | 1.33 | | HILIC | POS | | | 163217 | 37847 | 177370 | 46076.7 | 1.57E-02 | 0.34 | 1.44 |
| RP_NEG_648_46313a11_59 | | 648.4631 | 11.59 | | RP | NEG | | | 390051 | 46719.8 | 375784 | 39411.5 | 1.57E-02 | -0.33 | 1.55 |
| RP_POS_634_53815a10_838 | | 634.5382 | 10.84 | | RP | POS | | | 314418 | 247474 | 450687 | 506713 | 1.59E-02 | 0.36 | 1.52 |
| RP_NEG_379_12753a4_811 | | 379.1275 | 4.81 | | RP | NEG | | | 750827 | 257378 | 848651 | 322424 | 1.59E-02 | 0.34 | 1.66 |
| RP_POS_828_55084a13_008 | | 828.5508 | 13.01 | | RP | POS | | | 1.9E+08 | 5.9E+07 | 1.7E+08 | 5.3E+07 | 1.60E-02 | -0.33 | 1.44 |
| RP_POS_281_11356a2_646 | | 281.1136 | 2.65 | | RP | POS | 120.0808(100), 166.0876(45) | | 1650334 | 900076 | 1983005 | 1084535 | 1.62E-02 | 0.34 | 1.49 |
| RP_POS_771_57434a13_8 | | 771.5743 | 13.80 | | RP | POS | | | 1.3E+07 | 4753650 | 1.5E+07 | 5237569 | 1.62E-02 | 0.33 | 1.55 |
| RP_POS_339_28925a10_851 | | 339.2893 | 10.85 | | RP | POS | | | 1009322 | 698030 | 1367896 | 1320006 | 1.63E-02 | 0.36 | 1.59 |
| HILIC_POS_302_23187a0_751 | | 302.2319 | 0.75 | | HILIC | POS | | | 3002695 | 1953093 | 2446204 | 1414101 | 1.63E-02 | -0.33 | 1.32 |
| RP_POS_401_26599a10_647 | | 401.2660 | 10.65 | | RP | POS | | | 818729 | 508150 | 1090162 | 1009114 | 1.64E-02 | 0.36 | 1.56 |
| HILIC_NEG_191_99522a5_353 | | 191.9952 | 5.35 | | HILIC | NEG | | | 160604 | 33772 | 171525 | 32435.4 | 1.64E-02 | 0.33 | 1.41 |
| RP_NEG_387_02289a0_671 | | 387.0229 | 0.67 | | RP | NEG | | | 261691 | 39191.2 | 274113 | 36195.9 | 1.64E-02 | 0.33 | 1.4 |
| RP_POS_170_11754a4_714 | | 170.1175 | 4.71 | | RP | POS | | | 864135 | 276535 | 956740 | 283250 | 1.64E-02 | 0.33 | 1.43 |
| RP_POS_357_29984a10_852 | Monoolein | 357.2998 | 10.85 | HMDB0011567 | RP | POS | 121.1019(100), 247.2426(60), 339.2883(40) | 2 | 1393333 | 985128 | 1910077 | 1912425 | 1.64E-02 | 0.36 | 1.6 |
| RP_POS_181_12247a6_84 | | 181.1225 | 6.84 | | RP | POS | | | 1187840 | 190630 | 1122295 | 204782 | 1.64E-02 | -0.33 | 1.43 |

**Supplemantary table 1**: The non-targeted metabolomics results ordered by the VIP value (p value < 0.0003 are shaded dark green, p value < 0.05 are shaded light green, d values below 0 are blue and above 0 are red)

| | | | | | | | | | Cases | | Controls | | Welch test | Cohen's | |
|---|---|---|---|---|---|---|---|---|---|---|---|---|---|---|---|
| Feature_ID | Metabolite_name | Average_Mz | Average_RT_min_ | HMDB ID | RP_HILIC | POS_NEG | Cureted_MSMS | ID_level | Mean | SD | Mean2 | SD3 | p | d | VIP |
| RP_POS_416_22137a11_141 | | 416.2214 | 11.14 | | RP | POS | | | 631460 | 93647 | 597497 | 109910 | 1.64E-02 | -0.33 | 1.53 |
| HILIC_POS_396_31143a0_612 | | 396.3114 | 0.61 | | HILIC | POS | | | 85029.2 | 48402.1 | 107980 | 83049.2 | 1.66E-02 | 0.35 | 1.62 |
| HILIC_POS_352_16504a1_779 | | 352.1650 | 1.78 | | HILIC | POS | | | 356571 | 125794 | 405166 | 163042 | 1.66E-02 | 0.34 | 1.75 |
| RP_NEG_413_06552a4_81 | | 413.0655 | 4.81 | | RP | NEG | | | 705828 | 242083 | 797439 | 304348 | 1.66E-02 | 0.34 | 1.65 |
| RP_POS_370_29507a8_737 | AC 14:1 (Tetradecenoylcarnitine) | 370.2951 | 8.74 | HMDB0258884 | RP | POS | 85.0289(100), 370.2971(40) | 2 | 895476 | 517332 | 1082072 | 602794 | 1.66E-02 | 0.33 | 1.58 |
| RP_POS_160_13321a4_326 | | 160.1332 | 4.33 | | RP | POS | | | 666011 | 517916 | 525581 | 328213 | 1.66E-02 | -0.33 | 1.31 |
| RP_POS_313_15482a4_813 | | 313.1548 | 4.81 | | RP | POS | | | 1.7E+08 | 6E+07 | 1.9E+08 | 7.6E+07 | 1.67E-02 | 0.34 | 1.63 |
| RP_POS_120_08042a4_81 | Indoline | 120.0804 | 4.81 | HMDB0253472 | RP | POS | 120.0807(100), 103.0548(20) | 2 | 1095924 | 389925 | 1242039 | 483797 | 1.67E-02 | 0.33 | 1.63 |
| RP_POS_286_14368a8_392 | Piperine | 286.1437 | 8.39 | HMDB0029377 | RP | POS | 201.0543(100),286.1451(60) | 2 | 2.4E+07 | 3.3E+07 | 1.5E+07 | 2E+07 | 1.67E-02 | -0.33 | 1.31 |
| RP_POS_742_57526a13_576 | | 742.5753 | 13.58 | | RP | POS | | | 3.7E+07 | 1.1E+07 | 3.4E+07 | 8123663 | 1.68E-02 | -0.33 | 1.39 |
| RP_POS_239_23677a10_838 | | 239.2368 | 10.84 | | RP | POS | | | 474747 | 208758 | 570810 | 346733 | 1.71E-02 | 0.35 | 1.46 |
| RP_POS_335_13635a4_813 | | 335.1364 | 4.81 | | RP | POS | | | 1239992 | 520512 | 1443097 | 689723 | 1.71E-02 | 0.34 | 1.62 |
| RP_POS_438_29779a8_987 | | 438.2978 | 8.99 | | RP | POS | | | 765723 | 664221 | 567556 | 547165 | 1.71E-02 | -0.33 | 1.47 |
| RP_POS_700_28149a6_05 | | 700.2815 | 6.05 | | RP | POS | | | 573145 | 342176 | 477161 | 242591 | 1.71E-02 | -0.33 | 1.36 |
| RP_POS_504_30811a10_489 | | 504.3081 | 10.49 | | RP | POS | | | 3572813 | 1109243 | 4010730 | 1498451 | 1.73E-02 | 0.34 | 1.45 |
| RP_NEG_411_06595a4_811 | | 411.0660 | 4.81 | | RP | NEG | | | 422385 | 157964 | 486772 | 223686 | 1.74E-02 | 0.34 | 1.63 |
| RP_NEG_776_54633a13_031 | | 776.5463 | 13.03 | | RP | NEG | | | 1.4E+07 | 5420734 | 1.6E+07 | 8052990 | 1.75E-02 | 0.34 | 1.37 |
| HILIC_POS_331_24756a3_211 | | 331.2476 | 3.21 | | HILIC | POS | | | 527524 | 135066 | 574685 | 151392 | 1.75E-02 | 0.33 | 1.51 |
| RP_POS_348_31079a10_839 | 2,3-dihydroxypropyl hexadecanoate | 348.3108 | 10.84 | | RP | POS | 313.2729(100), 239.2379(40) | 3 | 973934 | 548325 | 1215142 | 864508 | 1.76E-02 | 0.34 | 1.47 |
| HILIC_POS_76_04005a5_926 | | 76.0401 | 5.93 | | HILIC | POS | | | 1124494 | 250062 | 1223466 | 340621 | 1.76E-02 | 0.34 | 1.39 |
| RP_NEG_593_47986a11_335 | | 593.4799 | 11.34 | | RP | NEG | | | 1589376 | 1051206 | 1945897 | 1122464 | 1.76E-02 | 0.33 | 1.42 |
| RP_POS_293_19055a2_787 | | 293.1906 | 2.79 | | RP | POS | | | 484594 | 326176 | 390773 | 249123 | 1.76E-02 | -0.33 | 1.34 |
| RP_POS_776_55853a14_064 | | 776.5585 | 14.06 | | RP | POS | | | 1894784 | 922113 | 1636608 | 658668 | 1.76E-02 | -0.33 | 1.6 |
| RP_POS_363_19586a11_145 | | 363.1959 | 11.15 | | RP | POS | | | 305853 | 653031 | 149929 | 238902 | 1.76E-02 | -0.35 | 1.31 |
| RP_POS_677_00446a5_812 | | 677.0045 | 5.81 | | RP | POS | | | 950610 | 1905754 | 500224 | 641016 | 1.76E-02 | -0.35 | 1.32 |
| RP_POS_339_2894a10_937 | | 339.2894 | 10.94 | | RP | POS | | | 3976965 | 2787979 | 5376343 | 5204093 | 1.77E-02 | 0.35 | 1.58 |
| RP_POS_413_26544a11_408 | 2-acetoxy-4-pentadecylbenzoic acid | 413.2654 | 11.41 | | RP | POS | 413.2647(100), 301.1394(20) | 2 | 7.7E+07 | 1.3E+07 | 7.3E+07 | 8920964 | 1.77E-02 | -0.33 | 1.41 |
| RP_POS_432_23792a8_636 | | 432.2379 | 8.64 | | RP | POS | | | 438421 | 595217 | 280413 | 363732 | 1.77E-02 | -0.33 | 1.26 |
| RP_POS_198_05284a5_287 | | 198.0528 | 5.29 | | RP | POS | | | 573116 | 292641 | 489883 | 219193 | 1.79E-02 | -0.33 | 1.35 |
| HILIC_POS_118_08641a3_177 | | 118.0864 | 3.18 | | HILIC | POS | 118.0865(100) | | 9350385 | 3129372 | 1E+07 | 3543501 | 1.80E-02 | 0.33 | 1.37 |
| RP_NEG_311_13986a4_81 | | 311.1399 | 4.81 | | RP | NEG | | | 2.4E+07 | 8394503 | 2.7E+07 | 1.1E+07 | 1.80E-02 | 0.33 | 1.64 |
| RP_POS_786_60205a13_953 | PC 36:2 (18:1/18:1) | 786.6021 | 13.95 | HMDB0000593 | RP | POS | 184.0745(100), 86.095(10) | 2 | 5.1E+07 | 2.6E+07 | 6.6E+07 | 5.6E+07 | 1.80E-02 | 0.36 | 1.55 |
| RP_NEG_698_37659a7_973 | | 698.3766 | 7.97 | | RP | NEG | | | 398605 | 325854 | 312875 | 195026 | 1.80E-02 | -0.33 | 1.29 |
| RP_POS_250_9366a15_387 | | 250.9366 | 15.39 | | RP | POS | | | 473112 | 223772 | 552714 | 261527 | 1.83E-02 | 0.33 | 1.37 |
| RP_POS_177_12733a10_241 | | 177.1273 | 10.24 | | RP | POS | | | 656089 | 249925 | 746917 | 302640 | 1.84E-02 | 0.33 | 1.59 |
| RP_POS_837_52521a12_999 | | 837.5252 | 13.00 | | RP | POS | | | 941737 | 692357 | 752999 | 468821 | 1.85E-02 | -0.33 | 1.38 |
| HILIC_NEG_273_0177a1_295 | | 273.0177 | 1.30 | | HILIC | NEG | | | 301927 | 144363 | 349941 | 151413 | 1.86E-02 | 0.32 | 1.37 |
| RP_NEG_239_08548a5_958 | | 239.0855 | 5.96 | | RP | NEG | | | 2.3E+07 | 1.4E+07 | 2E+07 | 9288906 | 1.86E-02 | -0.32 | 1.29 |
| RP_POS_385_75345a13_027 | | 385.7535 | 13.03 | | RP | POS | | | 918043 | 679682 | 1270718 | 1336318 | 1.87E-02 | 0.35 | 1.43 |
| RP_POS_803_55945a13_722 | | 803.5595 | 13.72 | | RP | POS | | | 247618 | 268279 | 357097 | 386947 | 1.87E-02 | 0.33 | 1.51 |
| RP_POS_309_22729a10_808 | | 309.2273 | 10.81 | | RP | POS | | | 373297 | 409640 | 509631 | 430669 | 1.87E-02 | 0.32 | 1.53 |
| RP_NEG_854_59259a13_624 | PC 38:4 (18:1_20:3) | 854.5926 | 13.62 | HMDB0008113 | RP | NEG | 305.2475(100), 794.5718(10), 281.2481(45) | 1 | 7993045 | 2112778 | 8663325 | 2036038 | 1.87E-02 | 0.32 | 1.33 |
| RP_NEG_329_26956a10_024 | | 329.2696 | 10.02 | | RP | NEG | | | 423315 | 26812.3 | 414772 | 26035.3 | 1.87E-02 | -0.32 | 1.55 |
| HILIC_NEG_150_88918a3_174 | | 150.8892 | 3.17 | | HILIC | NEG | | | 758036 | 360585 | 643447 | 349757 | 1.89E-02 | -0.32 | 1.34 |
| HILIC_NEG_335_65009a1_409 | | 335.6501 | 1.41 | | HILIC | NEG | | | 130233 | 29709.7 | 121442 | 24930.3 | 1.89E-02 | -0.32 | 1.28 |
| HILIC_NEG_301_68332a1_408 | | 301.6833 | 1.41 | | HILIC | NEG | | | 265615 | 62850.9 | 249786 | 32786.6 | 1.89E-02 | -0.33 | 1.24 |
| RP_POS_163_03905a11_408 | | 163.0391 | 11.41 | | RP | POS | 163.0394(100) | | 3144666 | 425544 | 3021564 | 339817 | 1.90E-02 | -0.32 | 1.37 |
| RP_POS_271_11307a8_842 | | 271.1131 | 8.84 | | RP | POS | | | 458700 | 186642 | 406559 | 136641 | 1.90E-02 | -0.32 | 1.5 |
| RP_POS_833_52893a12_999 | | 833.5289 | 13.00 | | RP | POS | | | 1447703 | 1028424 | 1153036 | 802807 | 1.90E-02 | -0.32 | 1.55 |
| RP_NEG_236_97903a7_367 | | 236.9790 | 7.37 | | RP | NEG | | | 1903068 | 424629 | 2019189 | 294699 | 1.91E-02 | 0.32 | 1.3 |
| RP_POS_1183_33276a13_167 | | 1183.3328 | 13.17 | | RP | POS | | | 1508962 | 892344 | 1842730 | 1141751 | 1.92E-02 | 0.33 | 1.55 |
| RP_NEG_331_26462a11_262 | FA 22:4 (Adrenic acid, docosatetraenoic acid) | 331.2646 | 11.26 | HMDB0002226 | RP | NEG | 331.2623(100), 59.013(2) | 2 | 1345050 | 875855 | 1615131 | 805420 | 1.92E-02 | 0.32 | 1.61 |
| RP_POS_751_57184a12_965 | | 751.5718 | 12.97 | | RP | POS | | | 6411619 | 1817963 | 6968719 | 1653594 | 1.93E-02 | 0.32 | 1.41 |
| RP_POS_345_20578a6_999 | | 345.2058 | 7.00 | | RP | POS | | | 270834 | 139292 | 329285 | 210281 | 1.94E-02 | 0.33 | 1.39 |
| RP_NEG_253_10823a7_215 | | 253.1082 | 7.22 | | RP | NEG | | | 1054515 | 86501 | 1026829 | 85776.4 | 1.95E-02 | -0.32 | 1.41 |
| HILIC_POS_188_07079a1_397 | | 188.0708 | 1.40 | | HILIC | POS | | | 1391093 | 2174375 | 847223 | 1130656 | 1.96E-02 | -0.33 | 1.29 |
| RP_POS_309_15106a10_808 | | 309.1511 | 10.81 | | RP | POS | | | 456458 | 500707 | 623060 | 534107 | 1.97E-02 | 0.32 | 1.52 |
| RP_NEG_465_1929a8_709 | | 465.1929 | 8.71 | | RP | NEG | | | 519412 | 803308 | 324474 | 363341 | 1.97E-02 | -0.33 | 1.27 |
| RP_POS_1172_33691a13_282 | | 1172.3369 | 13.28 | | RP | POS | | | 1235506 | 668556 | 1489320 | 880337 | 1.98E-02 | 0.33 | 1.44 |
| HILIC_NEG_160_93578a0_934 | | 160.9358 | 0.93 | | HILIC | NEG | | | 209212 | 44325.6 | 195314 | 42510 | 1.98E-02 | -0.32 | 1.39 |
| RP_NEG_197_80798a3_68 | | 197.8080 | 3.68 | | RP | NEG | | | 992997 | 434996 | 864062 | 372142 | 1.98E-02 | -0.32 | 1.34 |
| RP_POS_732_55402a13_027 | | 732.5540 | 13.03 | | RP | POS | | | 1.1E+08 | 4.7E+07 | 1.3E+08 | 7.1E+07 | 1.99E-02 | 0.33 | 1.38 |
| RP_NEG_599_25348a5_5 | | 599.2535 | 5.50 | | RP | NEG | | | 438465 | 491610 | 312264 | 285844 | 1.99E-02 | -0.32 | 1.31 |
| HILIC_NEG_331_65567a1_41 | | 331.6557 | 1.41 | | HILIC | NEG | | | 1749401 | 376680 | 1637330 | 327070 | 2.02E-02 | -0.32 | 1.29 |
| RP_NEG_251_12904a7_961 | | 251.1290 | 7.96 | | RP | NEG | | | 243380 | 116722 | 213058 | 71138.2 | 2.02E-02 | -0.32 | 1.33 |
| RP_NEG_488_22653a10_492 | | 488.2265 | 10.49 | | RP | NEG | | | 1272723 | 1044968 | 1624525 | 1147108 | 2.03E-02 | 0.32 | 1.45 |
| RP_POS_544_33978a10_295 | LysoPC 20:4 (0:0/20:4) | 544.3398 | 10.30 | HMDB0061699 | RP | POS | 184.0747(100), 86.0963(70) | 1 | 2.2E+08 | 5E+07 | 2.3E+08 | 5.6E+07 | 2.04E-02 | 0.32 | 1.35 |
| RP_NEG_556_21802a13_3 | | 556.2180 | 13.30 | | RP | NEG | | | 269369 | 90761 | 299771 | 99027.9 | 2.05E-02 | 0.32 | 1.41 |
| RP_POS_265_11829a3_648 | Phenylacetylglutamine | 265.1183 | 3.65 | | RP | POS | 130.0503(100), 84.045(25) | 2 | 1.1E+07 | 9171492 | 8128139 | 7332183 | 2.05E-02 | -0.32 | 1.32 |
| RP_POS_176_07068a5_288 | 3-Indoleacetic acid | 176.0707 | 5.29 | HMDB0000197 | RP | POS | 130.0656(100), 176.0703(20) | 1 | 9094931 | 4345676 | 7872330 | 3350569 | 2.06E-02 | -0.32 | 1.33 |
| RP_POS_130_06532a5_288 | Quinoline | 130.0653 | 5.29 | HMDB0033731 | RP | POS | 130.0657(100) | 1 | 4567681 | 2086434 | 3977064 | 1632126 | 2.06E-02 | -0.32 | 1.33 |
| RP_NEG_397_2059a7_519 | | 397.2059 | 7.52 | | RP | NEG | | | 513492 | 222272 | 588104 | 244031 | 2.07E-02 | 0.32 | 1.38 |
| HILIC_NEG_180_98921a0_934 | | 180.9892 | 0.93 | | HILIC | NEG | | | 1100364 | 115293 | 1065388 | 105219 | 2.08E-02 | -0.32 | 1.31 |
| RP_NEG_136_89157a0_618 | | 136.8916 | 0.62 | | RP | NEG | | | 1166529 | 732031 | 936281 | 718159 | 2.09E-02 | -0.32 | 1.32 |
| RP_NEG_680_31885a10_281 | | 680.3189 | 10.28 | | RP | NEG | | | 963986 | 232868 | 887010 | 250182 | 2.10E-02 | -0.32 | 1.47 |
| RP_POS_211_09689a6_463 | | 211.0969 | 6.46 | | RP | POS | | | 5781491 | 1.8E+07 | 1748567 | 5012630 | 2.10E-02 | -0.35 | 1.22 |
| RP_NEG_316_94788a12_945 | | 316.9479 | 12.95 | | RP | NEG | | | 1419804 | 624065 | 1642335 | 759890 | 2.11E-02 | 0.32 | 1.35 |
| RP_POS_586_30914a11_603 | | 586.3091 | 11.60 | | RP | POS | | | 344920 | 144763 | 394405 | 164735 | 2.11E-02 | 0.32 | 1.41 |
| RP_NEG_402_16373a11_592 | | 402.1637 | 11.59 | | RP | NEG | | | 5445345 | 244192 | 5368526 | 240449 | 2.11E-02 | -0.32 | 1.38 |
| RP_POS_286_16437a6_735 | | 286.1644 | 6.74 | | RP | POS | | | 637553 | 68256.1 | 659041 | 67376.4 | 2.12E-02 | 0.32 | 1.44 |
| RP_POS_354_28391a9_045 | | 354.2839 | 9.05 | | RP | POS | | | 1410832 | 102400 | 1375807 | 116699 | 2.12E-02 | -0.32 | 1.48 |
| RP_NEG_532_28918a8_734 | | 532.2892 | 8.73 | | RP | NEG | | | 547921 | 1156921 | 287214 | 325634 | 2.12E-02 | -0.35 | 1.25 |
| RP_POS_1167_51953a11_852 | | 1167.5195 | 11.85 | | RP | POS | | | 2456469 | 4778363 | 1245011 | 2764127 | 2.13E-02 | -0.32 | 1.29 |
| HILIC_POS_335_13672a1_719 | | 335.1367 | 1.72 | | HILIC | POS | | | 258205 | 140784 | 319455 | 228067 | 2.14E-02 | 0.33 | 1.58 |
| RP_POS_271_09229a0_679 | | 271.0923 | 0.68 | | RP | POS | | | 358204 | 121583 | 322095 | 107258 | 2.14E-02 | -0.32 | 1.28 |
| HILIC_POS_281_11273a5_9 | | 281.1127 | 5.90 | | HILIC | POS | | | 191219 | 87991.9 | 222806 | 108710 | 2.15E-02 | 0.32 | 1.46 |
| RP_POS_374_3266a10_938 | | 374.3266 | 10.94 | | RP | POS | | | 3542970 | 2598199 | 4774970 | 4706116 | 2.16E-02 | 0.34 | 1.54 |
| RP_POS_742_5376a12_533 | | 742.5376 | 12.53 | | RP | POS | | | 1417593 | 1108644 | 1079264 | 1037410 | 2.16E-02 | -0.32 | 1.36 |
| RP_POS_494_36209a10_841 | | 494.3621 | 10.84 | | RP | POS | | | 914688 | 407915 | 797124 | 339267 | 2.17E-02 | -0.31 | 1.31 |
| RP_POS_415_21127a8_635 | | 415.2113 | 8.64 | | RP | POS | 119.0859(100) | | 575284 | 1263067 | 256874 | 723782 | 2.17E-02 | -0.32 | 1.25 |
| RP_POS_357_29987a10_938 | MG 18:1 | 357.2999 | 10.94 | HMDB0011536 | RP | POS | 121.1021(100), 265.252(70),0339.288(60) | 1 | 1.1E+07 | 7670932 | 1.4E+07 | 1.4E+07 | 2.18E-02 | 0.34 | 1.55 |
| RP_NEG_440_22421a12_105 | | 440.2242 | 12.11 | | RP | NEG | | | 331533 | 50176 | 316082 | 47877.7 | 2.18E-02 | -0.32 | 1.51 |
| RP_POS_722_51111a12_933 | | 722.5111 | 12.93 | | RP | POS | | | 1763896 | 1470254 | 1366550 | 1047982 | 2.18E-02 | -0.32 | 1.25 |
| RP_POS_257_13861a4_8 | | 257.1386 | 4.80 | | RP | POS | | | 305027 | 243572 | 449469 | 577622 | 2.19E-02 | 0.35 | 1.37 |
| RP_POS_605_24756a10_091 | | 605.2476 | 10.09 | | RP | POS | | | 548999 | 320409 | 657147 | 360568 | 2.19E-02 | 0.32 | 1.58 |
| RP_POS_281_11368a3_038 | Asp-Phe | 281.1137 | 3.04 | HMDB0000706 | RP | POS | 120.0808(100), 166.0871(55) | 2 | 1852099 | 886837 | 2165802 | 1076954 | 2.19E-02 | 0.32 | 1.49 |
| RP_POS_274_62454a10_108 | | 274.6245 | 10.11 | | RP | POS | | | 464565 | 330653 | 587638 | 434546 | 2.21E-02 | 0.32 | 1.57 |
| RP_POS_716_22815a11_574 | | 716.2282 | 11.57 | | RP | POS | | | 362604 | 110957 | 398328 | 115504 | 2.21E-02 | 0.32 | 1.33 |
| RP_NEG_850_5614a12_783 | PC 38:6 (18:2_20:4) | 850.5614 | 12.78 | | RP | NEG | 255.2328(100), 327.2315(65), 790.5338(15) | 1 | 1.1E+07 | 3683632 | 1E+07 | 3466780 | 2.21E-02 | -0.31 | 1.36 |
| HILIC_NEG_194_98892a0_935 | | 194.9889 | 0.94 | | HILIC | NEG | | | 1769466 | 164645 | 1718558 | 159439 | 2.22E-02 | -0.31 | 1.28 |
| HILIC_NEG_329_65689a1_41 | | 329.6569 | 1.41 | | HILIC | NEG | | | 1542522 | 328610 | 1445919 | 287744 | 2.22E-02 | -0.31 | 1.26 |
| RP_NEG_537_3797a11_996 | | 537.3797 | 12.00 | | RP | NEG | | | 128870 | 201821 | 245059 | 464204 | 2.23E-02 | 0.35 | 1.52 |
| HILIC_NEG_278_98178a5_121 | | 278.9818 | 5.12 | | HILIC | NEG | | | 3.5E+07 | 1.7E+07 | 4.1E+07 | 1.9E+07 | 2.23E-02 | 0.32 | 1.47 |
| HILIC_POS_313_15515a1_722 | | 313.1552 | 1.72 | | HILIC | POS | | | 1.5E+07 | 5964041 | 1.7E+07 | 8062589 | 2.24E-02 | 0.32 | 1.6 |
| HILIC_POS_202_17995a1_32 | | 202.1800 | 1.32 | | HILIC | POS | | | 3.3E+07 | 1.8E+07 | 2.8E+07 | 1.5E+07 | 2.24E-02 | -0.31 | 1.39 |
| RP_POS_367_21594a11_236 | | 367.2159 | 11.24 | | RP | POS | | | 366542 | 207011 | 439394 | 251049 | 2.25E-02 | 0.32 | 1.45 |
| RP_POS_573_40527a10_424 | | 573.4053 | 10.42 | | RP | POS | | | 587785 | 257640 | 687283 | 358658 | 2.25E-02 | 0.32 | 1.41 |
| RP_POS_530_36957a10_873 | | 530.3696 | 10.87 | | RP | POS | | | 2310277 | 1261075 | 2888983 | 2213169 | 2.26E-02 | 0.33 | 1.53 |
| RP_POS_279_23178a9_656 | | 279.2318 | 9.66 | | RP | POS | | | 2491579 | 2135352 | 1895501 | 1677336 | 2.26E-02 | -0.31 | 1.33 |
| RP_POS_335_21967a8_989 | | 335.2197 | 8.99 | | RP | POS | | | 1310275 | 478235 | 1171328 | 411154 | 2.26E-02 | -0.31 | 1.29 |
| HILIC_NEG_161_04546a5_119 | | 161.0455 | 5.12 | | HILIC | NEG | | | 4973367 | 1876988 | 5755059 | 2907855 | 2.27E-02 | 0.33 | 1.43 |
| RP_POS_343_17758a10_809 | | 343.1776 | 10.81 | | RP | POS | | | 798610 | 868104 | 1085842 | 954998 | 2.27E-02 | 0.32 | 1.48 |
| RP_POS_634_54034a14_205 | | 634.5403 | 14.21 | | RP | POS | | | 1791428 | 1236615 | 1450202 | 944762 | 2.27E-02 | -0.31 | 1.27 |
| RP_POS_159_7048a4_81 | | 159.7048 | 4.81 | | RP | POS | | | 993446 | 369978 | 1129196 | 479787 | 2.28E-02 | 0.32 | 1.6 |
| RP_POS_260_18558a4_277 | AC 06:0 (Hexanoylcarnitine) | 260.1856 | 4.28 | HMDB0241848 | RP | POS | 85.0291(100), 260.1861(40) | 1 | 2303369 | 1278095 | 3119213 | 3317032 | 2.28E-02 | 0.36 | 1.47 |
| HILIC_POS_186_18539a0_953 | | 186.1854 | 0.95 | | HILIC | POS | | | 37135.8 | 20134.7 | 31622 | 15194.9 | 2.30E-02 | -0.31 | 1.3 |
| HILIC_POS_120_08117a1_711 | | 120.0812 | 1.71 | | HILIC | POS | | | 541985 | 216433 | 621964 | 284436 | 2.31E-02 | 0.32 | 1.62 |
| HILIC_POS_247_14421a1_398 | Lenticin (Tryptophan betaine) | 247.1442 | 1.40 | HMDB0061115 | HILIC | POS | 188.0708(100), 146.0599(45) | 2 | 1.2E+07 | 1.9E+07 | 7418823 | 9834673 | 2.32E-02 | -0.32 | 1.25 |
| RP_NEG_195_81058a3_679 | | 195.8106 | 3.68 | | RP | NEG | | | 737557 | 382715 | 631376 | 299357 | 2.32E-02 | -0.31 | 1.27 |
| RP_POS_240_15665a11_145 | | 240.1567 | 11.15 | | RP | POS | | | 2821261 | 593565 | 2634837 | 600015 | 2.32E-02 | -0.31 | 1.27 |
| RP_POS_168_00806a15_54 | | 168.0081 | 15.54 | | RP | POS | | | 1.5E+07 | 2215484 | 1.4E+07 | 3194297 | 2.32E-02 | -0.32 | 1.34 |
| RP_NEG_333_13519a9_648 | | 333.1352 | 9.65 | | RP | NEG | | | 51648.9 | 163714 | 16536.5 | 19886.9 | 2.32E-02 | -0.38 | 1.22 |
| RP_POS_815_56732a11_641 | | 815.5673 | 11.64 | | RP | POS | | | 157223 | 89662.9 | 194718 | 140532 | 2.33E-02 | 0.33 | 1.43 |
| RP_POS_326_30481a10_791 | | 326.3048 | 10.79 | | RP | POS | | | 1166881 | 444208 | 1310486 | 473009 | 2.33E-02 | 0.31 | 1.36 |
| RP_POS_792_55115a13_175 | | 792.5512 | 13.18 | | RP | POS | | | 5646293 | 1000385 | 5943738 | 912965 | 2.34E-02 | 0.31 | 1.38 |
| RP_POS_521_20825a8_391 | | 521.2083 | 8.39 | | RP | POS | | | 464278 | 563261 | 317329 | 375670 | 2.34E-02 | -0.31 | 1.3 |
| RP_POS_615_49548a14_691 | | 615.4955 | 14.69 | | RP | POS | | | 3647395 | 2085433 | 4395012 | 2635135 | 2.36E-02 | 0.32 | 1.48 |
| RP_NEG_425_36407a11_698 | | 425.3641 | 11.70 | | RP | NEG | | | 373809 | 63389.8 | 354181 | 62722.7 | 2.36E-02 | -0.31 | 1.59 |
| RP_POS_340_23947a10_157 | | 340.2395 | 10.16 | | RP | POS | | | 221900 | 184501 | 304964 | 319606 | 2.38E-02 | 0.33 | 1.48 |
| RP_POS_287_10025a3_635 | | 287.1003 | 3.64 | | RP | POS | | | 990700 | 766464 | 780548 | 588148 | 2.38E-02 | -0.31 | 1.26 |
| RP_POS_193_0862a6_463 | | 193.0862 | 6.46 | | RP | POS | | | 2343981 | 7042325 | 787059 | 1991582 | 2.38E-02 | -0.34 | 1.2 |
| RP_NEG_568_26764a10_291 | | 568.2676 | 10.29 | | RP | NEG | | | 899749 | 165858 | 955137 | 188043 | 2.39E-02 | 0.31 | 1.6 |
| RP_POS_681_49463a10_705 | | 681.4946 | 10.71 | | RP | POS | | | 694239 | 349429 | 584680 | 355146 | 2.39E-02 | -0.31 | 1.43 |
| HILIC_NEG_226_99461a3_058 | | 226.9946 | 3.06 | | HILIC | NEG | | | 449125 | 38483.6 | 437454 | 36830.3 | 2.40E-02 | -0.31 | 1.49 |
| HILIC_NEG_118_9241a4_246 | | 118.9241 | 4.25 | | HILIC | NEG | | | 2883423 | 548613 | 2681011 | 726817 | 2.40E-02 | -0.32 | 1.44 |
| RP_NEG_212_07509a15_486 | | 212.0751 | 15.49 | | RP | NEG | | | 8529153 | 4431879 | 9967771 | 4798305 | 2.41E-02 | 0.31 | 1.29 |
| HILIC_POS_203_14993a6_944 | | 203.1499 | 6.94 | | HILIC | POS | | | 411386 | 156953 | 371557 | 97693 | 2.41E-02 | -0.31 | 1.29 |
| RP_NEG_287_70544a15_495 | | 287.7054 | 15.50 | | RP | NEG | | | 3282766 | 128216 | 3244342 | 119970 | 2.41E-02 | -0.31 | 1.29 |
| RP_NEG_399_25201a11_261 | | 399.2520 | 11.26 | | RP | NEG | | | 108909 | 78798.7 | 132029 | 70870.1 | 2.43E-02 | 0.31 | 1.59 |
| RP_POS_351_12589a0_587 | | 351.1259 | 0.59 | | RP | POS | | | 607737 | 309701 | 708571 | 338149 | 2.43E-02 | 0.31 | 1.31 |
| HILIC_POS_512_33447a1_706 | | 512.3345 | 1.71 | | HILIC | POS | | | 2217122 | 5764657 | 902979 | 2224293 | 2.43E-02 | -0.33 | 1.23 |
| RP_POS_312_36221a9_675 | | 312.3622 | 9.68 | | RP | POS | | | 304528 | 258147 | 428683 | 488183 | 2.44E-02 | 0.33 | 1.4 |
| RP_POS_687_30798a11_387 | | 687.3080 | 11.39 | | RP | POS | | | 101918 | 54226.8 | 121859 | 71789.2 | 2.44E-02 | 0.32 | 1.34 |
| HILIC_POS_287_10025a1_883 | | 287.1003 | 1.88 | | HILIC | POS | | | 682330 | 514584 | 537342 | 426019 | 2.45E-02 | -0.31 | 1.27 |
| RP_POS_345_03314a15_586 | | 345.0331 | 15.59 | | RP | POS | | | 1.1E+07 | 5867198 | 9236068 | 5201668 | 2.45E-02 | -0.31 | 1.27 |
| RP_POS_599_50928a11_144 | | 599.5093 | 11.14 | | RP | POS | | | 1270086 | 246590 | 1194872 | 240179 | 2.45E-02 | -0.31 | 1.43 |
| RP_POS_141_11343a10_578 | | 141.1134 | 10.58 | | RP | POS | | | 105479 | 71205.9 | 88133.4 | 38727.1 | 2.45E-02 | -0.32 | 1.21 |
| RP_POS_848_53802a13_218 | | 848.5380 | 13.22 | | RP | POS | | | 4249405 | 3091593 | 5525120 | 4817692 | 2.46E-02 | 0.32 | 1.32 |
| HILIC_POS_242_0789a2_967 | | 242.0789 | 2.97 | | HILIC | POS | | | 1416116 | 455070 | 1578493 | 577973 | 2.47E-02 | 0.31 | 1.47 |
| RP_POS_224_12802a0_596 | | 224.1280 | 0.60 | | RP | POS | | | 1711563 | 532747 | 1870447 | 498333 | 2.47E-02 | 0.31 | 1.25 |
| RP_POS_699_27808a6_084 | | 699.2781 | 6.08 | | RP | POS | | | 1812589 | 1135577 | 1525457 | 709073 | 2.47E-02 | -0.31 | 1.26 |
| RP_NEG_150_00195a10_387 | | 150.0020 | 10.39 | | RP | NEG | 150.0011(100) | | 1251782 | 857862 | 1528032 | 924219 | 2.48E-02 | 0.31 | 1.57 |
| RP_POS_331_2843a10_838 | 2-Palmitoylglycerol | 331.2843 | 10.84 | HMDB0011533 | RP | POS | 57.071(100), 71.0862(85) | 2 | 5022728 | 2300916 | 6103836 | 4243213 | 2.48E-02 | 0.33 | 1.44 |
| RP_POS_766_5379a12_458 | | 766.5379 | 12.46 | | RP | POS | | | 2683638 | 1930103 | 2192754 | 1229274 | 2.48E-02 | -0.31 | 1.27 |
| RP_POS_265_98105a4_765 | | 265.9811 | 4.77 | | RP | POS | | | 480898 | 153090 | 431058 | 168199 | 2.49E-02 | -0.31 | 1.36 |
| RP_POS_283_00769a4_77 | | 283.0077 | 4.77 | | RP | POS | | | 1080836 | 288322 | 985284 | 326383 | 2.49E-02 | -0.31 | 1.33 |
| RP_POS_408_13919a10_088 | | 408.1392 | 10.09 | | RP | POS | | | 508107 | 319391 | 608004 | 327811 | 2.50E-02 | 0.31 | 1.54 |
| RP_POS_402_7728a12_882 | | 402.7728 | 12.88 | | RP | POS | | | 591895 | 443864 | 466213 | 374571 | 2.50E-02 | -0.31 | 1.33 |
| RP_POS_525_034a9_877 | | 525.0340 | 9.88 | | RP | POS | | | 464092 | 293645 | 556158 | 303225 | 2.51E-02 | 0.31 | 1.34 |
| RP_NEG_787_52246a12_239 | | 787.5225 | 12.24 | | RP | NEG | | | 1826343 | 366808 | 1719707 | 328090 | 2.52E-02 | -0.31 | 1.27 |
| RP_NEG_650_45709a11_592 | | 650.4571 | 11.59 | | RP | NEG | | | 7865096 | 811901 | 7608121 | 854302 | 2.53E-02 | -0.31 | 1.61 |
| HILIC_NEG_130_92921a3_142 | | 130.9292 | 3.14 | | HILIC | NEG | | | 715099 | 112250 | 745681 | 87298.6 | 2.54E-02 | 0.31 | 1.27 |

**Supplemantary table 1**: The non-targeted metabolomics results ordered by the VIP value (p value < 0.0003 are shaded dark green, p value < 0.05 are shaded light green, d values below 0 are blue and above 0 are red)

| | | | | | | | | | Cases | | Controls | | Welch test | Cohen's | |
|---|---|---|---|---|---|---|---|---|---|---|---|---|---|---|---|
| Feature_ID | Metabolite_name | Average_Mz | Average_RT_min_ | HMDB ID | RP_HILIC | POS_NEG | Cureted_MSMS | ID_level | Mean | SD | Mean2 | SD3 | p | d | VIP |
| HILIC_POS_60_05653a2_156 | | 60.0565 | 2.16 | | HILIC | POS | | | 4056936 | 1.4E+07 | 1033408 | 4045477 | 2.54E-02 | -0.34 | 1.16 |
| HILIC_NEG_157_90088a0_709 | | 157.9009 | 0.71 | | HILIC | NEG | | | 1615126 | 428279 | 1780172 | 613303 | 2.55E-02 | 0.32 | 1.41 |
| RP_POS_340_16946a10_808 | | 340.1695 | 10.81 | | RP | POS | | | 726854 | 773630 | 969277 | 801760 | 2.55E-02 | 0.31 | 1.47 |
| RP_POS_898_62946a11_144 | | 898.6295 | 11.14 | | RP | POS | | | 832209 | 116710 | 795663 | 120852 | 2.55E-02 | -0.31 | 1.32 |
| RP_NEG_407_22052a10_65 | | 407.2205 | 10.65 | | RP | NEG | | | 1104516 | 568442 | 1288508 | 624257 | 2.57E-02 | 0.31 | 1.55 |
| RP_POS_821_08276a14_018 | | 821.0828 | 14.02 | | RP | POS | | | 1488433 | 962089 | 1836397 | 1261001 | 2.58E-02 | 0.31 | 1.38 |
| RP_NEG_719_5368a12_24 | | 719.5368 | 12.24 | | RP | NEG | | | 1.3E+07 | 2700678 | 1.3E+07 | 2347981 | 2.58E-02 | -0.31 | 1.26 |
| RP_POS_227_12502a0_541 | | 227.1250 | 0.54 | | RP | POS | | | 1.2E+07 | 1891964 | 1.2E+07 | 2313398 | 2.58E-02 | -0.31 | 1.34 |
| RP_POS_371_1535a8_842 | | 371.1535 | 8.84 | | RP | POS | | | 192051 | 118065 | 160627 | 87095.7 | 2.58E-02 | -0.31 | 1.3 |
| RP_POS_266_15961a4_063 | | 266.1596 | 4.06 | | RP | POS | | | 1902178 | 231456 | 1966971 | 192971 | 2.59E-02 | 0.31 | 1.29 |
| RP_POS_406_18103a10_389 | | 406.1810 | 10.39 | | RP | POS | | | 2797341 | 1765021 | 3367261 | 1939062 | 2.60E-02 | 0.31 | 1.54 |
| RP_NEG_426_1723a11_239 | | 426.1723 | 11.24 | | RP | NEG | | | 303356 | 181367 | 361210 | 195397 | 2.61E-02 | 0.31 | 1.5 |
| RP_POS_395_25531a10_939 | | 395.2553 | 10.94 | | RP | POS | | | 427745 | 348521 | 587447 | 631246 | 2.63E-02 | 0.33 | 1.36 |
| HILIC_NEG_118_92439a2_272 | | 118.9244 | 2.27 | | HILIC | NEG | | | 3412864 | 536599 | 3596911 | 652280 | 2.63E-02 | 0.31 | 1.34 |
| RP_POS_229_11826a0_594 | | 229.1183 | 0.59 | | RP | POS | | | 2514461 | 1320222 | 2919400 | 1331840 | 2.65E-02 | 0.31 | 1.42 |
| RP_POS_568_29358a6_693 | | 568.2936 | 6.69 | | RP | POS | | | 277510 | 173814 | 327788 | 157094 | 2.67E-02 | 0.3 | 1.24 |
| RP_POS_399_35748a10_331 | | 399.3575 | 10.33 | | RP | POS | | | 290435 | 298027 | 397765 | 392014 | 2.68E-02 | 0.31 | 1.3 |
| RP_POS_619_43781a11_239 | | 619.4378 | 11.24 | | RP | POS | | | 1604838 | 891790 | 1900647 | 1033945 | 2.69E-02 | 0.31 | 1.44 |
| HILIC_NEG_333_6525a1_41 | | 333.6525 | 1.41 | | HILIC | NEG | | | 816200 | 175121 | 766646 | 151592 | 2.69E-02 | -0.3 | 1.22 |
| RP_NEG_369_21259a11_144 | | 369.2126 | 11.14 | | RP | NEG | | | 468060 | 41474.7 | 453779 | 51023.2 | 2.69E-02 | -0.31 | 1.43 |
| RP_POS_177_10219a1_641 | | 177.1022 | 1.64 | | RP | POS | 160.0767(100) | | 464792 | 221177 | 535794 | 243109 | 2.70E-02 | 0.31 | 1.38 |
| RP_NEG_353_30643a11_02 | | 353.3064 | 11.02 | | RP | NEG | | | 393360 | 60524.7 | 374606 | 62685.4 | 2.71E-02 | -0.3 | 1.58 |
| RP_POS_491_02078a10_217 | | 491.0208 | 10.22 | | RP | POS | | | 272475 | 180916 | 224150 | 138063 | 2.72E-02 | -0.3 | 1.42 |
| RP_NEG_854_59308a13_956 | PC 38:4 (18:0_20:4) | 854.5931 | 13.96 | HMDB0008431 | RP | NEG | 303.2345(100), 283.2641(62) | 2 | 4.7E+07 | 9431085 | 5E+07 | 9625258 | 2.74E-02 | 0.30 | 1.35 |
| RP_POS_183_10143a2_726 | | 183.1014 | 2.73 | | RP | POS | | | 179314 | 83183.3 | 154692 | 79455.3 | 2.74E-02 | -0.3 | 1.37 |
| RP_NEG_150_00194a10_088 | | 150.0019 | 10.09 | | RP | NEG | | | 4006900 | 2564248 | 4803847 | 2681070 | 2.75E-02 | 0.3 | 1.53 |
| HILIC_NEG_276_99142a1_226 | | 276.9914 | 1.23 | | HILIC | NEG | | | 1545926 | 665704 | 1363023 | 545763 | 2.76E-02 | -0.3 | 1.4 |
| RP_NEG_328_75253a15_359 | | 328.7525 | 15.36 | | RP | NEG | | | 3613339 | 218342 | 3681245 | 229271 | 2.77E-02 | 0.3 | 1.41 |
| RP_POS_363_04282a15_581 | | 363.0428 | 15.58 | | RP | POS | | | 2321919 | 1361892 | 1966727 | 985318 | 2.77E-02 | -0.3 | 1.39 |
| RP_POS_160_13322a0_595 | | 160.1332 | 0.60 | | RP | POS | | | 2.1E+07 | 8136865 | 2.4E+07 | 9003664 | 2.78E-02 | 0.3 | 1.29 |
| RP_NEG_488_79739a15_396 | | 488.7974 | 15.40 | | RP | NEG | | | 344137 | 144744 | 301636 | 136696 | 2.78E-02 | -0.3 | 1.58 |
| RP_POS_460_14969a8_385 | | 460.1497 | 8.39 | | RP | POS | | | 204297 | 35555.9 | 215195 | 36461.1 | 2.79E-02 | 0.3 | 1.29 |
| HILIC_POS_160_09686a4_562 | 2-(4-aminotetrahydro-2H-pyran-4-yl)acetic acid | 160.0969 | 4.56 | | HILIC | POS | 160.0961(100) | 2 | 3.3E+07 | 1.3E+08 | 6473377 | 2.5E+07 | 2.79E-02 | -0.35 | 1.16 |
| RP_POS_267_97879a4_765 | | 267.9788 | 4.77 | | RP | POS | | | 453310 | 159434 | 401711 | 179763 | 2.80E-02 | -0.3 | 1.29 |
| RP_NEG_321_20505a10_808 | | 321.2051 | 10.81 | | RP | NEG | | | 208546 | 281744 | 301012 | 325368 | 2.81E-02 | 0.3 | 1.47 |
| HILIC_NEG_355_15823a0_933 | | 355.1582 | 0.93 | | HILIC | NEG | | | 211968 | 46451.2 | 198156 | 45155.3 | 2.81E-02 | -0.3 | 1.26 |
| RP_POS_713_99915a11_489 | | 713.9992 | 11.49 | | RP | POS | | | 144713 | 98354.8 | 188107 | 172607 | 2.82E-02 | 0.32 | 1.55 |
| HILIC_POS_152_02197a1_217 | | 152.0220 | 1.22 | | HILIC | POS | | | 2641711 | 363309 | 2531371 | 368154 | 2.83E-02 | -0.3 | 1.4 |
| RP_POS_742_30267a5_325 | | 742.3027 | 5.33 | | RP | POS | | | 908460 | 304743 | 807692 | 356759 | 2.83E-02 | -0.3 | 1.25 |
| RP_POS_318_30042a10_928 | | 318.3004 | 10.93 | | RP | POS | | | 507886 | 138840 | 468109 | 125846 | 2.84E-02 | -0.3 | 1.48 |
| RP_POS_377_76492a13_026 | | 377.7649 | 13.03 | | RP | POS | | | 177905 | 164354 | 285885 | 460342 | 2.85E-02 | 0.35 | 1.32 |
| RP_NEG_433_18958a11_612 | | 433.1896 | 11.61 | | RP | NEG | | | 526322 | 97510.5 | 497106 | 96577.5 | 2.85E-02 | -0.3 | 1.38 |
| HILIC_NEG_118_92417a0_934 | | 118.9242 | 0.93 | | HILIC | NEG | | | 4125893 | 355370 | 4018452 | 359055 | 2.88E-02 | -0.3 | 1.21 |
| RP_NEG_355_22839a10_244 | | 355.2284 | 10.24 | | RP | NEG | | | 337510 | 182634 | 400960 | 232211 | 2.89E-02 | 0.31 | 1.6 |
| RP_NEG_398_17596a11_364 | | 398.1760 | 11.36 | | RP | NEG | | | 429211 | 41812.6 | 416598 | 42181 | 2.90E-02 | -0.3 | 1.47 |
| RP_NEG_458_19846a11_237 | | 458.1985 | 11.24 | | RP | NEG | | | 366598 | 141041 | 409344 | 143613 | 2.91E-02 | 0.3 | 1.45 |
| RP_NEG_197_80803a15_687 | | 197.8080 | 15.69 | | RP | NEG | | | 735057 | 368744 | 635700 | 295781 | 2.91E-02 | -0.3 | 1.3 |
| RP_POS_339_23172a10_646 | | 339.2317 | 10.65 | | RP | POS | | | 711419 | 395062 | 850096 | 512015 | 2.92E-02 | 0.31 | 1.5 |
| RP_POS_186_14893a10_571 | | 186.1489 | 10.57 | | RP | POS | | | 768725 | 323663 | 678911 | 277802 | 2.93E-02 | -0.3 | 1.29 |
| RP_POS_622_42847a10_11 | | 622.4285 | 10.11 | | RP | POS | | | 273015 | 159642 | 325722 | 188538 | 2.94E-02 | 0.3 | 1.5 |
| HILIC_POS_260_1857a1_051 | AC 06:0 (Hexanoylcarnitine) | 260.1857 | 1.05 | HMDB0241848 | HILIC | POS | 85.0292(100), 260.1864(45) | 1 | 2953227 | 1551297 | 3947099 | 4251965 | 2.94E-02 | 0.34 | 1.43 |
| RP_POS_301_25241a8_867 | | 301.2524 | 8.87 | | RP | POS | | | 463177 | 290420 | 378298 | 277234 | 2.94E-02 | -0.3 | 1.22 |
| RP_POS_379_28174a10_94 | | 379.2817 | 10.94 | | RP | POS | | | 1.6E+07 | 1.2E+07 | 2.1E+07 | 1.9E+07 | 2.95E-02 | 0.32 | 1.45 |
| RP_POS_579_07135a10_144 | | 579.0714 | 10.14 | | RP | POS | | | 248803 | 157179 | 305788 | 214152 | 2.95E-02 | 0.31 | 1.48 |
| RP_NEG_369_17371a11_361 | | 369.1737 | 11.36 | | RP | NEG | | | 980014 | 161565 | 930597 | 167931 | 2.95E-02 | -0.3 | 1.53 |
| RP_NEG_653_47491a11_237 | | 653.4749 | 11.24 | | RP | NEG | | | 259669 | 170554 | 311579 | 175936 | 2.96E-02 | 0.3 | 1.5 |
| RP_POS_341_19913a11_141 | | 341.1991 | 11.14 | | RP | POS | | | 891090 | 149217 | 845793 | 153197 | 2.96E-02 | -0.3 | 1.3 |
| RP_POS_514_33765a8_971 | | 514.3377 | 8.97 | | RP | POS | | | 7320050 | 2734647 | 6416592 | 3240725 | 2.96E-02 | -0.3 | 1.27 |
| RP_POS_626_34955a11_411 | | 626.3496 | 11.41 | | RP | POS | | | 844202 | 215473 | 789068 | 153380 | 2.97E-02 | -0.3 | 1.23 |
| RP_NEG_413_20084a7_619 | | 413.2008 | 7.62 | | RP | NEG | | | 463454 | 309289 | 379925 | 252279 | 2.99E-02 | -0.3 | 1.25 |
| RP_NEG_467_20834a8_688 | | 467.2083 | 8.69 | | RP | NEG | | | 1382460 | 562099 | 1197899 | 661106 | 2.99E-02 | -0.3 | 1.31 |
| RP_NEG_397_20547a8_679 | | 397.2055 | 8.68 | | RP | NEG | | | 7296094 | 4731000 | 5963707 | 4226404 | 3.00E-02 | -0.3 | 1.23 |
| RP_POS_361_28085a11_144 | | 361.2809 | 11.14 | | RP | POS | | | 1576140 | 296027 | 1487984 | 295107 | 3.00E-02 | -0.3 | 1.47 |
| RP_NEG_168_83606a7_263 | | 168.8361 | 7.26 | | RP | NEG | | | 1472123 | 445400 | 1608568 | 467324 | 3.01E-02 | 0.3 | 1.27 |
| RP_POS_285_12842a9_207 | | 285.1284 | 9.21 | | RP | POS | | | 831981 | 426687 | 718734 | 335116 | 3.01E-02 | -0.3 | 1.35 |
| RP_NEG_382_18524a10_386 | | 382.1852 | 10.39 | | RP | NEG | | | 229866 | 158439 | 280835 | 181089 | 3.03E-02 | 0.3 | 1.55 |
| HILIC_NEG_124_00736a5_355 | Taurine | 124.0074 | 5.36 | HMDB0000251 | HILIC | NEG | 124.0069(100), 79.9571(55) | 2 | 5642087 | 1581443 | 6142441 | 1757783 | 3.03E-02 | 0.30 | 1.29 |
| RP_POS_524_27502a10_186 | | 524.2750 | 10.19 | | RP | POS | | | 713302 | 190310 | 774889 | 219856 | 3.04E-02 | 0.3 | 1.44 |
| RP_POS_502_29279a10_294 | | 502.2928 | 10.29 | | RP | POS | | | 4.5E+07 | 9735822 | 4.8E+07 | 1.2E+07 | 3.05E-02 | 0.3 | 1.46 |
| RP_POS_677_95215a10_088 | | 677.9522 | 10.09 | | RP | POS | | | 1377170 | 617496 | 1195863 | 604159 | 3.07E-02 | -0.3 | 1.25 |
| RP_NEG_299_25952a10_925 | | 299.2595 | 10.93 | | RP | NEG | | | 1.1E+07 | 1454335 | 1.1E+07 | 1395836 | 3.08E-02 | -0.3 | 1.58 |
| RP_NEG_249_7888a15_493 | | 249.7888 | 15.49 | | RP | NEG | | | 393514 | 45317.2 | 408065 | 51916.4 | 3.09E-02 | 0.3 | 1.35 |
| HILIC_NEG_194_98837a5_111 | | 194.9884 | 5.11 | | HILIC | NEG | | | 1372767 | 655169 | 1619841 | 956982 | 3.11E-02 | 0.31 | 1.66 |
| RP_POS_820_58435a13_374 | | 820.5844 | 13.37 | | RP | POS | | | 7693436 | 4022409 | 6704090 | 2626454 | 3.12E-02 | -0.3 | 1.28 |
| RP_POS_181_07205a2_609 | Theobromine | 181.0721 | 2.61 | HMDB0002825 | RP | POS | 181.0721(100), 138.0665(10) | 2 | 1.9E+07 | 1.9E+07 | 2.5E+07 | 1.9E+07 | 3.13E-02 | 0.30 | 1.23 |
| RP_NEG_226_966a15_315 | | 226.9660 | 15.32 | | RP | NEG | | | 1034407 | 132228 | 995026 | 133812 | 3.14E-02 | -0.3 | 1.35 |
| RP_POS_299_13889a5_75 | | 299.1389 | 5.75 | | RP | POS | | | 551058 | 635760 | 388924 | 461665 | 3.15E-02 | -0.3 | 1.3 |
| RP_POS_345_21707a8_435 | | 345.2171 | 8.44 | | RP | POS | | | 404726 | 582367 | 271744 | 292535 | 3.15E-02 | -0.3 | 1.18 |
| HILIC_NEG_457_23615a1_492 | | 457.2362 | 1.49 | | HILIC | NEG | | | 208840 | 159551 | 259805 | 182475 | 3.16E-02 | 0.3 | 1.31 |
| RP_POS_437_1936a8_633 | | 437.1936 | 8.63 | | RP | POS | | | 154004 | 275774 | 88078.3 | 165940 | 3.17E-02 | -0.3 | 1.2 |
| HILIC_POS_331_24719a0_653 | | 331.2472 | 0.65 | | HILIC | POS | | | 639751 | 233258 | 575538 | 204094 | 3.21E-02 | -0.29 | 1.19 |
| HILIC_POS_371_24008a4_197 | | 371.2401 | 4.20 | | HILIC | POS | | | 6756664 | 1364023 | 6411601 | 978956 | 3.21E-02 | -0.29 | 1.25 |
| RP_POS_416_22784a3_691 | | 416.2278 | 3.69 | | RP | POS | | | 913037 | 471702 | 779114 | 439396 | 3.21E-02 | -0.29 | 1.27 |
| RP_POS_733_31464a11_587 | | 733.3146 | 11.59 | | RP | POS | | | 141090 | 88074.5 | 117740 | 70964.2 | 3.21E-02 | -0.29 | 1.48 |
| RP_NEG_144_99179a15_514 | | 144.9918 | 15.51 | | RP | NEG | | | 1850620 | 397218 | 1744732 | 324844 | 3.23E-02 | -0.29 | 1.21 |
| RP_POS_810_6015a13_956 | PC 38:4 (18:0_20:4) | 810.6015 | 13.96 | HMDB0008431 | RP | POS | 184.0743(100) | 1 | 3E+08 | 8.2E+07 | 3.2E+08 | 8.1E+07 | 3.24E-02 | 0.29 | 1.46 |
| HILIC_POS_64_01667a0_933 | | 64.0167 | 0.93 | | HILIC | POS | | | 6263222 | 752463 | 6044163 | 738660 | 3.24E-02 | -0.29 | 1.25 |
| RP_POS_418_80048a9_39 | | 418.8005 | 9.39 | | RP | POS | | | 480149 | 268377 | 572086 | 345484 | 3.25E-02 | 0.3 | 1.24 |
| RP_NEG_509_27557a8_722 | | 509.2756 | 8.72 | | RP | NEG | | | 808373 | 431782 | 962050 | 588681 | 3.27E-02 | 0.3 | 1.25 |
| RP_POS_350_30511a11_264 | | 350.3051 | 11.26 | | RP | POS | | | 1068395 | 752074 | 1281203 | 700775 | 3.27E-02 | 0.29 | 1.42 |
| RP_POS_370_18359a10_09 | | 370.1836 | 10.09 | | RP | POS | | | 1.9E+07 | 1.2E+07 | 2.3E+07 | 1.3E+07 | 3.29E-02 | 0.29 | 1.5 |
| HILIC_POS_370_99896a7_481 | | 370.9990 | 7.48 | | HILIC | POS | | | 117102 | 9264.31 | 119954 | 10102.7 | 3.30E-02 | 0.29 | 1.21 |
| RP_POS_581_45404a12_232 | | 581.4540 | 12.23 | | RP | POS | | | 735464 | 918220 | 1002322 | 903516 | 3.30E-02 | 0.29 | 1.37 |
| HILIC_POS_342_26419a0_652 | AC 12:0 (Dodecanoylcarnitine) | 342.2642 | 0.65 | HMDB0002250 | HILIC | POS | 85.0292(100), 342.2654(40) | 1 | 2546846 | 1695132 | 3113660 | 2113256 | 3.30E-02 | 0.30 | 1.42 |
| RP_NEG_788_54846a12_89 | | 788.5485 | 12.89 | | RP | NEG | | | 5431426 | 1113406 | 5102319 | 1131731 | 3.31E-02 | -0.29 | 1.28 |
| RP_POS_384_19907a10_389 | | 384.1991 | 10.39 | | RP | POS | | | 6906496 | 4627575 | 8320669 | 4996178 | 3.32E-02 | 0.29 | 1.51 |
| RP_NEG_627_37585a8_824 | | 627.3759 | 8.82 | | RP | NEG | | | 285593 | 117420 | 317405 | 100817 | 3.33E-02 | 0.29 | 1.21 |
| RP_POS_309_20297a9_555 | Hexadecanedioic acid | 309.2030 | 9.56 | HMDB0304377 | RP | POS | 309.2051(100) | 2 | 1147121 | 134330 | 1110356 | 118032 | 3.34E-02 | -0.29 | 1.35 |
| RP_NEG_397_00388a3_68 | | 397.0039 | 3.68 | | RP | NEG | | | 1764037 | 1527546 | 1370115 | 1173583 | 3.34E-02 | -0.29 | 1.23 |
| RP_NEG_134_89488a11_238 | | 134.8949 | 11.24 | | RP | NEG | | | 567728 | 259277 | 645711 | 273083 | 3.35E-02 | 0.29 | 1.4 |
| RP_POS_296_22241a9_345 | | 296.2224 | 9.35 | | RP | POS | | | 872143 | 258664 | 949735 | 271440 | 3.36E-02 | 0.29 | 1.3 |
| HILIC_POS_126_02229a5_347 | Taurine | 126.0223 | 5.35 | HMDB0000251 | HILIC | POS | 126.0229(100), 108.012(55) | 2 | 1.1E+07 | 3212736 | 1.2E+07 | 3569443 | 3.36E-02 | 0.29 | 1.26 |
| RP_POS_372_17905a10_09 | | 372.1791 | 10.09 | | RP | POS | | | 697612 | 472494 | 837846 | 486689 | 3.37E-02 | 0.29 | 1.5 |
| HILIC_POS_467_34784a0_989 | | 467.3478 | 0.99 | | HILIC | POS | | | 168156 | 251041 | 114673 | 92572.5 | 3.40E-02 | -0.31 | 1.2 |
| HILIC_NEG_127_00152a0_934 | | 127.0015 | 0.93 | | HILIC | NEG | | | 410791 | 63662.2 | 391506 | 68299.7 | 3.41E-02 | -0.29 | 1.24 |
| HILIC_POS_461_35721a4_266 | | 461.3572 | 4.27 | | HILIC | POS | | | 88046.4 | 59617.7 | 74089.1 | 35248.8 | 3.44E-02 | -0.29 | 1.21 |
| RP_NEG_464_30157a8_734 | Glycocholic acid | 464.3016 | 8.73 | HMDB0000138 | RP | NEG | 464.3042(100), 74.0239(85) | 1 | 8502657 | 2E+07 | 4338519 | 4986351 | 3.46E-02 | -0.33 | 1.16 |
| RP_NEG_800_54596a12_815 | | 800.5460 | 12.82 | | RP | NEG | | | 2.7E+07 | 7716326 | 2.5E+07 | 7715323 | 3.47E-02 | -0.29 | 1.25 |
| RP_POS_129_06607a0_552 | | 129.0661 | 0.55 | | RP | POS | | | 2346058 | 1564494 | 1996700 | 772674 | 3.47E-02 | -0.3 | 1.17 |
| RP_NEG_227_03867a6_166 | | 227.0387 | 6.17 | | RP | NEG | | | 135007 | 338586 | 241244 | 388091 | 3.49E-02 | 0.29 | 1.24 |
| RP_NEG_285_24359a10_711 | | 285.2436 | 10.71 | | RP | NEG | | | 951911 | 123335 | 916920 | 118345 | 3.49E-02 | -0.29 | 1.48 |
| RP_POS_472_30353a9_224 | | 472.3035 | 9.22 | HMDB0000631 | RP | POS | 472.3047(100), 397.2722(2) | | 4499990 | 5469065 | 3308133 | 2403367 | 3.49E-02 | -0.3 | 1.2 |
| RP_POS_579_42511a10_52 | | 579.4251 | 10.52 | | RP | POS | | | 1960174 | 904123 | 1710971 | 822048 | 3.51E-02 | -0.29 | 1.28 |
| RP_POS_212_0202a7 | | 212.0202 | 7.00 | | RP | POS | | | 1786208 | 2952061 | 2793000 | 3860904 | 3.52E-02 | 0.3 | 1.33 |
| RP_NEG_368_16931a10_088 | | 368.1693 | 10.09 | | RP | NEG | | | 1113429 | 707834 | 1323272 | 739500 | 3.53E-02 | 0.29 | 1.5 |
| RP_NEG_189_04045a0_68 | | 189.0405 | 0.68 | | RP | NEG | | | 363138 | 104111 | 335145 | 90085.8 | 3.53E-02 | -0.29 | 1.19 |
| RP_POS_385_34628a13_3 | | 385.3463 | 13.30 | | RP | POS | | | 1082818 | 398105 | 959597 | 447735 | 3.53E-02 | -0.29 | 1.23 |
| HILIC_POS_74_0608a5_792 | | 74.0608 | 5.79 | | HILIC | POS | | | 1655058 | 417323 | 1784370 | 470644 | 3.54E-02 | 0.29 | 1.28 |
| RP_POS_357_27869a9_824 | Chenodeoxycholic acid | 357.2787 | 9.82 | HMDB0000518 | RP | POS | 357.2766(100) | 1 | 2801881 | 4051270 | 4492542 | 6996294 | 3.55E-02 | 0.31 | 1.26 |
| RP_POS_300_28958a10_668 | | 300.2896 | 10.67 | | RP | POS | | | 1558818 | 327011 | 1654423 | 334379 | 3.56E-02 | 0.29 | 1.22 |
| RP_NEG_437_2912a9_823 | Ursodeoxycholic acid | 437.2912 | 9.82 | HMDB0000946 | RP | NEG | 391.2835(100) | 2 | 898419 | 1324493 | 1436389 | 2211276 | 3.56E-02 | 0.30 | 1.27 |
| RP_POS_450_32104a9_224 | Glycochenodeoxycholic acid | 450.3210 | 9.22 | HMDB0002497 | RP | POS | 414.302(100), 339.2696(30) | 1 | 8730580 | 1.1E+07 | 6321572 | 4514532 | 3.56E-02 | -0.31 | 1.2 |
| RP_POS_285_25763a8_709 | | 285.2576 | 8.71 | | RP | POS | | | 740813 | 466198 | 612655 | 424810 | 3.58E-02 | -0.29 | 1.36 |
| RP_POS_496_27481a6_861 | | 496.2748 | 6.86 | | RP | POS | | | 523214 | 45572.4 | 510355 | 43703 | 3.58E-02 | -0.29 | 1.31 |
| RP_NEG_397_13837a10_075 | | 397.1384 | 10.08 | | RP | NEG | | | 27830.6 | 24859.8 | 140897 | 528338 | 3.59E-02 | 0.41 | 1.46 |
| RP_POS_296_1488a7_665 | | 296.1488 | 7.67 | | RP | POS | | | 458524 | 62587.1 | 476694 | 63327.9 | 3.59E-02 | 0.29 | 1.44 |
| HILIC_POS_689_5611a0_854 | | 689.5611 | 0.85 | | HILIC | POS | | | 637911 | 145963 | 597514 | 135002 | 3.59E-02 | -0.29 | 1.23 |
| RP_NEG_516_29419a9_222 | | 516.2942 | 9.22 | | RP | NEG | | | 1513626 | 1732547 | 1134676 | 800618 | 3.60E-02 | -0.3 | 1.2 |
| RP_POS_265_25247a10_935 | | 265.2525 | 10.94 | | RP | POS | | | 4694908 | 1798269 | 5547936 | 3653070 | 3.61E-02 | 0.31 | 1.42 |
| RP_POS_432_31067a9_224 | | 432.3107 | 9.22 | | RP | POS | 414.305(100), 339.2693(40) | | 1.8E+07 | 2.3E+07 | 1.3E+07 | 9164847 | 3.61E-02 | -0.3 | 1.2 |
| HILIC_POS_414_29938a1_001 | | 414.2994 | 1.00 | | HILIC | POS | | | 228231 | 322377 | 158422 | 141995 | 3.61E-02 | -0.3 | 1.19 |
| HILIC_POS_240_10162a1_715 | | 240.1016 | 1.72 | | HILIC | POS | 240.1031(100) | | 1969861 | 801367 | 2214983 | 891129 | 3.62E-02 | 0.29 | 1.23 |
| RP_POS_689_56091a12_511 | | 689.5609 | 12.51 | | RP | POS | | | 1.9E+07 | 4496139 | 1.8E+07 | 3753852 | 3.62E-02 | -0.29 | 1.36 |
| RP_NEG_588_33081a10_295 | | 588.3308 | 10.30 | | RP | NEG | | | 4.3E+07 | 9360841 | 4.5E+07 | 9834200 | 3.63E-02 | 0.29 | 1.27 |
| RP_NEG_674_27631a10_293 | | 674.2763 | 10.29 | | RP | NEG | | | 435802 | 102766 | 467188 | 114039 | 3.63E-02 | 0.29 | 1.35 |
| RP_POS_628_77618a11_541 | | 628.7762 | 11.54 | | RP | POS | | | 141957 | 58809 | 165049 | 94471.4 | 3.64E-02 | 0.3 | 1.3 |
| RP_NEG_402_14444a11_238 | | 402.1444 | 11.24 | | RP | NEG | | | 220412 | 257504 | 298058 | 280115 | 3.64E-02 | 0.29 | 1.42 |
| RP_NEG_411_20203a9_095 | | 411.2020 | 9.10 | | RP | NEG | | | 362656 | 145463 | 401806 | 127684 | 3.64E-02 | 0.29 | 1.21 |
| RP_NEG_448_30701a9_222 | Glycohyodeoxycholic acid | 448.3070 | 9.22 | | RP | NEG | 448.3041(100), 74.0248(65) | 1 | 2.1E+07 | 2.7E+07 | 1.5E+07 | 1.1E+07 | 3.64E-02 | -0.30 | 1.2 |
| RP_NEG_339_23297a10_156 | | 339.2330 | 10.16 | | RP | NEG | | | 552110 | 380560 | 693565 | 568439 | 3.65E-02 | 0.3 | 1.41 |
| RP_NEG_216_85385a11_591 | | 216.8539 | 11.59 | | RP | NEG | | | 591936 | 139128 | 550707 | 147137 | 3.66E-02 | -0.29 | 1.49 |
| RP_POS_414_30005a9_223 | | 414.3001 | 9.22 | | RP | POS | | | 1.5E+07 | 2E+07 | 1.1E+07 | 7811879 | 3.66E-02 | -0.3 | 1.2 |
| RP_POS_477_32095a10_99 | | 477.3210 | 10.99 | | RP | POS | | | 637441 | 298042 | 766168 | 542114 | 3.67E-02 | 0.31 | 1.54 |
| HILIC_POS_360_27472a0_988 | | 360.2747 | 0.99 | | HILIC | POS | | | 173223 | 93184.3 | 204005 | 117643 | 3.67E-02 | 0.29 | 1.41 |
| RP_NEG_257_81973a0_639 | | 257.8197 | 0.64 | | RP | NEG | | | 687880 | 208509 | 633302 | 173117 | 3.67E-02 | -0.29 | 1.19 |
| RP_POS_183_10161a7_361 | | 183.1016 | 7.36 | | RP | POS | | | 1810630 | 592010 | 2036263 | 916515 | 3.68E-02 | 0.3 | 1.53 |
| RP_NEG_433_2374a10_732 | | 433.2374 | 10.73 | | RP | NEG | | | 613635 | 420053 | 751514 | 525409 | 3.68E-02 | 0.29 | 1.43 |
| HILIC_POS_184_06056a0_821 | 4-Pyridoxate | 184.0606 | 0.82 | | HILIC | POS | 166.0506(100), 184.0602(30) | 2 | 1062922 | 1844316 | 658920 | 877803 | 3.68E-02 | -0.30 | 1.12 |
| RP_POS_711_53833a12_509 | | 711.5383 | 12.51 | | RP | POS | | | 4014708 | 1167343 | 3715044 | 928003 | 3.68E-02 | -0.29 | 1.4 |
| RP_NEG_665_43042a11_237 | | 665.4304 | 11.24 | | RP | NEG | | | 382992 | 206001 | 442689 | 209827 | 3.69E-02 | 0.29 | 1.41 |
| HILIC_NEG_327_65918a1_411 | | 327.6592 | 1.41 | | HILIC | NEG | | | 497593 | 105458 | 468871 | 95558.5 | 3.70E-02 | -0.29 | 1.17 |
| RP_POS_370_29523a8_595 | AC 14:1 (Tetradecenoylcarnitine) | 370.2952 | 8.60 | HMDB0240588 | RP | POS | 85.0292(100),370.2929(40) | 1 | 5210413 | 3609996 | 6617897 | 5776290 | 3.71E-02 | 0.30 | 1.38 |
| RP_POS_425_36273a12_428 | | 425.3627 | 12.43 | | RP | POS | | | 4921389 | 2408258 | 4239110 | 2367296 | 3.75E-02 | -0.29 | 1.24 |
| RP_NEG_195_81064a7_233 | | 195.8106 | 7.23 | | RP | NEG | | | 181586 | 152110 | 238767 | 232801 | 3.77E-02 | 0.3 | 1.25 |
| HILIC_NEG_183_93456a7_092 | | 183.9346 | 7.09 | | HILIC | NEG | | | 1397706 | 204683 | 1463758 | 251236 | 3.77E-02 | 0.29 | 1.41 |
| RP_POS_636_27557a11_276 | | 636.2756 | 11.28 | | RP | POS | | | 122602 | 68423.3 | 144606 | 83528.3 | 3.77E-02 | 0.29 | 1.24 |
| RP_POS_860_49341a12_732 | | 860.4934 | 12.73 | | RP | POS | | | 1329871 | 701579 | 1528967 | 693044 | 3.77E-02 | 0.29 | 1.25 |
| RP_POS_313_27371a10_838 | | 313.2737 | 10.84 | | RP | POS | | | 4311727 | 1773482 | 5058993 | 3149833 | 3.78E-02 | 0.3 | 1.36 |
| RP_POS_476_67136a9_743 | | 476.6714 | 9.74 | | RP | POS | | | 132442 | 67417 | 156574 | 96484.2 | 3.78E-02 | 0.29 | 1.24 |
| RP_NEG_212_00259a3_038 | | 212.0026 | 3.04 | | RP | NEG | | | 645230 | 342172 | 550942 | 320107 | 3.78E-02 | -0.28 | 1.26 |
| RP_NEG_842_51782a12_583 | | 842.5178 | 12.58 | | RP | NEG | | | 1452884 | 524523 | 1305588 | 508819 | 3.78E-02 | -0.29 | 1.23 |

**Supplemantary table 1**: The non-targeted metabolomics results ordered by the VIP value (p value < 0.0003 are shaded dark green, p value < 0.05 are shaded light green, d values below 0 are blue and above 0 are red)

| | | | | | | | | | Cases | | Controls | | Welch test | Cohen's | |
|---|---|---|---|---|---|---|---|---|---|---|---|---|---|---|---|
| Feature_ID | Metabolite_name | Average_Mz | Average_RT_min_ | HMDB ID | RP_HILIC | POS_NEG | Cureted_MSMS | ID_level | Mean | SD | Mean2 | SD3 | p | d | VIP |
| RP_NEG_826_59827a10_927 | | 826.5983 | 10.93 | | RP | NEG | | | 1362096 | 570741 | 1536353 | 642420 | 3.79E-02 | 0.29 | 1.49 |
| RP_POS_325_20483a10_805 | | 325.2048 | 10.81 | | RP | POS | | | 205879 | 208466 | 269468 | 234244 | 3.79E-02 | 0.29 | 1.27 |
| RP_POS_184_05821a15_241 | | 184.0582 | 15.24 | | RP | POS | | | 5053665 | 3283021 | 4211864 | 2638293 | 3.79E-02 | -0.28 | 1.36 |
| RP_POS_545_8761a10_194 | | 545.8761 | 10.19 | | RP | POS | | | 3184472 | 1569950 | 2716930 | 1694742 | 3.79E-02 | -0.29 | 1.36 |
| RP_POS_353_26617a10_838 | | 353.2662 | 10.84 | | RP | POS | | | 1.5E+07 | 6375984 | 1.8E+07 | 1.1E+07 | 3.80E-02 | 0.3 | 1.36 |
| RP_POS_508_34a10_374 | | 508.3400 | 10.37 | | RP | POS | | | 9868705 | 3141741 | 1.1E+07 | 3534832 | 3.80E-02 | 0.29 | 1.27 |
| RP_POS_554_16663a10_547 | | 554.1666 | 10.55 | | RP | POS | | | 1348523 | 601852 | 1533628 | 686354 | 3.80E-02 | 0.29 | 1.47 |
| RP_POS_646_27545a10_933 | | 646.2755 | 10.93 | | RP | POS | | | 2064766 | 382321 | 1952378 | 403709 | 3.80E-02 | -0.29 | 1.23 |
| RP_NEG_367_24698a11_072 | | 367.2470 | 11.07 | | RP | NEG | | | 650424 | 87280.1 | 625123 | 90047.7 | 3.81E-02 | -0.29 | 1.56 |
| RP_POS_488_34302a6_691 | | 488.3430 | 6.69 | | RP | POS | | | 822833 | 519042 | 686714 | 440527 | 3.83E-02 | -0.28 | 1.25 |
| RP_POS_425_19974a10_648 | | 425.1997 | 10.65 | | RP | POS | | | 274652 | 205347 | 336780 | 228557 | 3.84E-02 | 0.29 | 1.3 |
| RP_POS_375_22012a11_595 | | 375.2201 | 11.60 | | RP | POS | | | 1936843 | 333454 | 1839979 | 346151 | 3.84E-02 | -0.29 | 1.36 |
| RP_NEG_470_19742a11_592 | | 470.1974 | 11.59 | | RP | NEG | | | 2127103 | 181727 | 2076991 | 171364 | 3.85E-02 | -0.28 | 1.44 |
| RP_NEG_381_33762a11_401 | | 381.3376 | 11.40 | | RP | NEG | | | 269404 | 45600.9 | 256647 | 44304.7 | 3.87E-02 | -0.28 | 1.57 |
| HILIC_POS_317_05618a5_463 | | 317.0562 | 5.46 | | HILIC | POS | | | 3943334 | 685270 | 4139063 | 692928 | 3.88E-02 | 0.28 | 1.26 |
| RP_POS_766_57349a13_176 | | 766.5735 | 13.18 | | RP | POS | | | 5606112 | 4086390 | 4630642 | 2766968 | 3.88E-02 | -0.28 | 1.13 |
| RP_NEG_151_99771a10_037 | | 151.9977 | 10.04 | | RP | NEG | | | 152059 | 121293 | 190321 | 145230 | 3.90E-02 | 0.29 | 1.37 |
| RP_POS_581_90387a10_524 | | 581.9039 | 10.52 | | RP | POS | | | 719368 | 402254 | 618093 | 313883 | 3.90E-02 | -0.28 | 1.16 |
| HILIC_NEG_182_04605a0_821 | 4-Pyridoxic acid | 182.0461 | 0.82 | | HILIC | NEG | 138.0564(100), 182.0466(30) | 2 | 816163 | 1381552 | 515512 | 677438 | 3.92E-02 | -0.29 | 1.1 |
| RP_POS_856_5824a13_448 | | 856.5824 | 13.45 | | RP | POS | | | 1744574 | 1304793 | 1407539 | 1082437 | 3.92E-02 | -0.28 | 1.13 |
| HILIC_POS_228_19543a3_079 | | 228.1954 | 3.08 | | HILIC | POS | | | 590303 | 161650 | 539365 | 193562 | 3.92E-02 | -0.29 | 1.17 |
| RP_POS_159_0132a0_647 | | 159.0132 | 0.65 | | RP | POS | | | 5180596 | 1096996 | 5470033 | 954525 | 3.93E-02 | 0.28 | 1.17 |
| HILIC_NEG_115_92085a6_158 | | 115.9209 | 6.16 | | HILIC | NEG | | | 3.6E+07 | 1E+07 | 3.3E+07 | 9082910 | 3.93E-02 | -0.28 | 1.29 |
| RP_NEG_239_08568a6_492 | | 239.0857 | 6.49 | | RP | NEG | | | 9200002 | 7560896 | 7292846 | 5943244 | 3.93E-02 | -0.28 | 1.28 |
| RP_NEG_493_28119a8_865 | | 493.2812 | 8.87 | | RP | NEG | | | 1274300 | 709246 | 1078449 | 677724 | 3.95E-02 | -0.28 | 1.16 |
| RP_POS_321_2399a10_639 | | 321.2399 | 10.64 | | RP | POS | | | 3601169 | 600944 | 3437940 | 556019 | 3.95E-02 | -0.28 | 1.38 |
| RP_NEG_389_19284a10_806 | | 389.1928 | 10.81 | | RP | NEG | | | 98975.9 | 134287 | 139824 | 152445 | 3.96E-02 | 0.28 | 1.39 |
| RP_NEG_299_25989a11_071 | | 299.2599 | 11.07 | | RP | NEG | | | 6530341 | 835830 | 6284024 | 899133 | 3.96E-02 | -0.28 | 1.54 |
| RP_POS_454_29092a8_8 | | 454.2909 | 8.80 | | RP | POS | | | 289159 | 165776 | 241088 | 173518 | 3.97E-02 | -0.28 | 1.17 |
| RP_POS_585_27051a8_181 | | 585.2705 | 8.18 | | RP | POS | | | 290166 | 536644 | 170697 | 292657 | 3.97E-02 | -0.29 | 1.14 |
| RP_NEG_401_21652a8_689 | | 401.2165 | 8.69 | | RP | NEG | | | 1181015 | 508874 | 1012919 | 658209 | 3.98E-02 | -0.29 | 1.26 |
| RP_POS_699_48926a11_518 | | 699.4893 | 11.52 | | RP | POS | | | 165293 | 74509.8 | 190854 | 102081 | 4.01E-02 | 0.29 | 1.18 |
| RP_POS_235_1804a3_684 | | 235.1804 | 3.68 | HMDB0014426 | RP | POS | 86.0974(100), 235.1803(25) | | 1.8E+08 | 1.1E+07 | 1.8E+08 | 8278381 | 4.01E-02 | 0.28 | 1.2 |
| RP_POS_638_61157a11_349 | | 638.6116 | 11.35 | | RP | POS | | | 148472 | 91647.8 | 176700 | 106637 | 4.01E-02 | 0.28 | 1.27 |
| RP_NEG_185_11829a7_945 | | 185.1183 | 7.95 | | RP | NEG | | | 1128849 | 976033 | 863617 | 909407 | 4.01E-02 | -0.28 | 1.2 |
| RP_NEG_461_22266a12_107 | | 461.2227 | 12.11 | | RP | NEG | | | 252511 | 41657.6 | 240690 | 42126.5 | 4.01E-02 | -0.28 | 1.52 |
| RP_NEG_367_24738a10_925 | | 367.2474 | 10.93 | | RP | NEG | | | 1167516 | 163466 | 1120756 | 167786 | 4.02E-02 | -0.28 | 1.63 |
| RP_POS_425_78516a14_436 | | 425.7852 | 14.44 | | RP | POS | | | 575263 | 479321 | 735050 | 630781 | 4.03E-02 | 0.29 | 1.27 |
| RP_POS_582_29529a10_289 | | 582.2953 | 10.29 | | RP | POS | | | 1031262 | 323491 | 1125121 | 340929 | 4.03E-02 | 0.28 | 1.37 |
| RP_NEG_168_83656a14_342 | | 168.8366 | 14.34 | | RP | NEG | | | 1583123 | 653557 | 1405963 | 606869 | 4.03E-02 | -0.28 | 1.23 |
| RP_POS_255_05862a15_26 | | 255.0586 | 15.26 | | RP | POS | 255.0601(100) | | 2118683 | 1083351 | 2442196 | 1200885 | 4.04E-02 | 0.28 | 1.18 |
| HILIC_POS_626_30139a0_991 | | 626.3014 | 0.99 | | HILIC | POS | | | 325128 | 69976.5 | 347063 | 84102.1 | 4.07E-02 | 0.28 | 1.25 |
| RP_POS_266_01126a1_41 | | 266.0113 | 1.41 | | RP | POS | | | 333016 | 163292 | 384560 | 198497 | 4.07E-02 | 0.28 | 1.17 |
| HILIC_NEG_132_03015a6_759 | Aspartate | 132.0302 | 6.76 | HMDB0006483 | HILIC | NEG | 88.04(60), 132.0205(35) | 1 | 350523 | 111517 | 386771 | 141740 | 4.07E-02 | 0.29 | 1.44 |
| RP_NEG_349_11212a6_635 | Estrone-3-sulfate | 349.1121 | 6.64 | | RP | NEG | 349.113(100), 269.1549(40) | 2 | 806490 | 775382 | 623381 | 523148 | 4.07E-02 | -0.28 | 1.35 |
| RP_POS_149_02341a11_408 | | 149.0234 | 11.41 | | RP | POS | 149.0231(100), 121.0287(20) | | 2.1E+07 | 3369929 | 2E+07 | 2360121 | 4.08E-02 | -0.28 | 1.17 |
| RP_POS_848_57568a10_959 | | 848.5757 | 10.96 | | RP | POS | | | 195501 | 144260 | 238960 | 163040 | 4.10E-02 | 0.28 | 1.29 |
| HILIC_NEG_272_93658a5_353 | | 272.9366 | 5.35 | | HILIC | NEG | | | 381301 | 128982 | 418645 | 136350 | 4.11E-02 | 0.28 | 1.2 |
| RP_POS_729_59106a12_967 | | 729.5911 | 12.97 | | RP | POS | | | 2.9E+07 | 7820022 | 3.1E+07 | 7240536 | 4.13E-02 | 0.28 | 1.3 |
| RP_POS_728_98785a10_217 | | 728.9879 | 10.22 | | RP | POS | | | 678948 | 429961 | 573767 | 321384 | 4.13E-02 | -0.28 | 1.29 |
| RP_POS_488_29825a8_735 | | 488.2983 | 8.74 | | RP | POS | 412.2869(100), 337.2515(80) | | 1479069 | 4007035 | 695045 | 858646 | 4.13E-02 | -0.32 | 1.12 |
| RP_NEG_704_43347a11_235 | | 704.4335 | 11.24 | | RP | NEG | | | 144846 | 103900 | 174878 | 109828 | 4.14E-02 | 0.28 | 1.35 |
| HILIC_POS_434_78018a0_821 | | 434.7802 | 0.82 | | HILIC | POS | | | 102105 | 86174.2 | 81616.9 | 59917.9 | 4.15E-02 | -0.28 | 1.19 |
| RP_NEG_868_53339a12_813 | | 868.5334 | 12.81 | | RP | NEG | | | 3144348 | 1372283 | 2778430 | 1249511 | 4.16E-02 | -0.28 | 1.22 |
| RP_POS_213_18509a8_12 | | 213.1851 | 8.12 | | RP | POS | | | 719025 | 58512.7 | 701901 | 63336.8 | 4.17E-02 | -0.28 | 1.24 |
| RP_POS_421_35202a10_699 | | 421.3520 | 10.70 | | RP | POS | | | 2084322 | 210204 | 2027951 | 193863 | 4.17E-02 | -0.28 | 1.23 |
| HILIC_POS_432_31076a1 | Glycohyodeoxcholic acid | 432.3108 | 1.00 | | HILIC | POS | 414.3026(100), 76.0401(40) | 1 | 472729 | 655961 | 335392 | 281997 | 4.18E-02 | -0.29 | 1.16 |
| RP_POS_475_41531a11_14 | | 475.4153 | 11.14 | | RP | POS | | | 1125403 | 285647 | 1048394 | 266401 | 4.18E-02 | -0.28 | 1.23 |
| RP_NEG_548_23218a9_232 | | 548.2322 | 9.23 | | RP | NEG | | | 494831 | 736181 | 344713 | 266323 | 4.18E-02 | -0.3 | 1.16 |
| RP_POS_219_17409a10_09 | | 219.1741 | 10.09 | | RP | POS | | | 2829794 | 1054571 | 3128050 | 1077253 | 4.19E-02 | 0.28 | 1.41 |
| RP_POS_184_05818a9_339 | | 184.0582 | 9.34 | | RP | POS | | | 1875876 | 1002525 | 1623445 | 809331 | 4.19E-02 | -0.28 | 1.24 |
| RP_POS_612_76147a10_255 | | 612.7615 | 10.26 | | RP | POS | | | 465321 | 299426 | 554198 | 333457 | 4.22E-02 | 0.28 | 1.18 |
| RP_NEG_474_12741a10_651 | | 474.1274 | 10.65 | | RP | NEG | | | 269999 | 81214.2 | 293586 | 87242.6 | 4.23E-02 | 0.28 | 1.34 |
| RP_POS_392_16537a10_089 | | 392.1654 | 10.09 | | RP | POS | | | 7366430 | 4433627 | 8631096 | 4614438 | 4.23E-02 | 0.28 | 1.46 |
| RP_NEG_253_21761a10_808 | FA 16:1 | 253.2176 | 10.81 | HMDB0012328 | RP | NEG | 253.2186(100) | 2 | 1191507 | 1477474 | 1633731 | 1669250 | 4.23E-02 | 0.28 | 1.41 |
| RP_POS_592_92657a10_489 | | 592.9266 | 10.49 | | RP | POS | | | 646548 | 370076 | 541106 | 384686 | 4.24E-02 | -0.28 | 1.16 |
| RP_NEG_662_42224a11_238 | | 662.4222 | 11.24 | | RP | NEG | | | 514952 | 591097 | 688260 | 646078 | 4.25E-02 | 0.28 | 1.39 |
| HILIC_POS_203_13907a4_669 | | 203.1391 | 4.67 | | HILIC | POS | | | 3278453 | 813151 | 3075743 | 645789 | 4.25E-02 | -0.28 | 1.15 |
| RP_POS_818_60486a13_652 | | 818.6049 | 13.65 | | RP | POS | | | 1.4E+07 | 4506668 | 1.3E+07 | 4695584 | 4.27E-02 | -0.28 | 1.25 |
| RP_NEG_500_2785a10_293 | | 500.2785 | 10.29 | | RP | NEG | | | 1.8E+07 | 3945140 | 1.9E+07 | 4719631 | 4.28E-02 | 0.28 | 1.42 |
| RP_POS_544_40656a8_799 | | 544.4066 | 8.80 | | RP | POS | | | 868945 | 626921 | 724801 | 403224 | 4.28E-02 | -0.28 | 1.19 |
| RP_NEG_453_39496a12_16 | | 453.3950 | 12.16 | | RP | NEG | | | 219739 | 57970.8 | 203691 | 57454.3 | 4.29E-02 | -0.28 | 1.52 |
| RP_NEG_339_21545a10_473 | | 339.2155 | 10.47 | | RP | NEG | | | 474405 | 72628.9 | 454468 | 70881.6 | 4.30E-02 | -0.28 | 1.41 |
| HILIC_NEG_180_98933a3_051 | | 180.9893 | 3.05 | | HILIC | NEG | | | 359625 | 39530.2 | 371203 | 43402.7 | 4.32E-02 | 0.28 | 1.25 |
| RP_NEG_397_16833a11_238 | | 397.1683 | 11.24 | | RP | NEG | | | 439513 | 343162 | 535580 | 348355 | 4.33E-02 | 0.28 | 1.39 |
| RP_POS_127_03915a7_361 | | 127.0392 | 7.36 | | RP | POS | | | 799335 | 322182 | 914676 | 479976 | 4.35E-02 | 0.29 | 1.53 |
| RP_POS_788_61597a14_924 | PC 36:1 (18:0/18:1) | 788.6160 | 14.92 | HMDB0013427 | RP | POS | 184.0744(100) | 1 | 2.1E+07 | 1.3E+07 | 2.5E+07 | 1.9E+07 | 4.35E-02 | 0.29 | 1.41 |
| HILIC_POS_368_22a5_019 | | 368.2200 | 5.02 | | HILIC | POS | | | 233032 | 116179 | 204093 | 93191.1 | 4.35E-02 | -0.28 | 1.34 |
| RP_POS_300_28961a9_054 | | 300.2896 | 9.05 | | RP | POS | | | 519612 | 1447422 | 1228590 | 3195439 | 4.36E-02 | 0.31 | 1.3 |
| RP_NEG_246_90527a6_688 | | 246.9053 | 6.69 | | RP | NEG | | | 428641 | 354344 | 525148 | 342590 | 4.36E-02 | 0.28 | 1.29 |
| RP_POS_138_09126a3_765 | | 138.0913 | 3.77 | | RP | POS | | | 84345.1 | 64455.7 | 69255.2 | 44243.2 | 4.37E-02 | -0.28 | 1.22 |
| RP_NEG_468_1828a11_237 | | 468.1828 | 11.24 | | RP | NEG | | | 658016 | 188452 | 713220 | 207912 | 4.38E-02 | 0.28 | 1.36 |
| RP_NEG_600_28546a6_498 | | 600.2855 | 6.50 | | RP | NEG | | | 453257 | 439250 | 566489 | 381275 | 4.38E-02 | 0.28 | 1.2 |
| HILIC_POS_386_2897a0_886 | | 386.2897 | 0.89 | | HILIC | POS | | | 218027 | 133969 | 257481 | 149307 | 4.39E-02 | 0.28 | 1.38 |
| HILIC_NEG_244_84848a5_323 | | 244.8485 | 5.32 | | HILIC | NEG | | | 837468 | 371123 | 741554 | 324345 | 4.39E-02 | -0.28 | 1.23 |
| RP_POS_611_91284a10_136 | | 611.9128 | 10.14 | | RP | POS | | | 3153823 | 1588325 | 2731451 | 1470665 | 4.39E-02 | -0.28 | 1.38 |
| RP_POS_154_05875a0_559 | | 154.0588 | 0.56 | | RP | POS | | | 964671 | 336789 | 1061355 | 359473 | 4.40E-02 | 0.28 | 1.15 |
| RP_POS_297_0824a9_614 | | 297.0824 | 9.61 | | RP | POS | | | 850966 | 176229 | 898279 | 166526 | 4.41E-02 | 0.28 | 1.2 |
| RP_NEG_429_19522a7_753 | | 429.1952 | 7.75 | | RP | NEG | | | 188914 | 313918 | 116800 | 205271 | 4.42E-02 | -0.28 | 1.13 |
| RP_NEG_459_22659a12_107 | | 459.2266 | 12.11 | | RP | NEG | | | 718397 | 108702 | 687591 | 113654 | 4.42E-02 | -0.28 | 1.55 |
| RP_POS_253_19493a7_258 | | 253.1949 | 7.26 | | RP | POS | | | 1711488 | 1468194 | 2380478 | 2987401 | 4.43E-02 | 0.3 | 1.26 |
| RP_NEG_353_14307a10_809 | | 353.1431 | 10.81 | | RP | NEG | | | 564696 | 804897 | 802359 | 904443 | 4.44E-02 | 0.28 | 1.37 |
| RP_POS_236_07149a10_877 | | 236.0715 | 10.88 | | RP | POS | | | 1.1E+08 | 3.4E+07 | 1E+08 | 2.6E+07 | 4.44E-02 | -0.28 | 1.11 |
| RP_NEG_351_16339a10_363 | | 351.1634 | 10.36 | | RP | NEG | | | 106577 | 126937 | 166293 | 268674 | 4.45E-02 | 0.3 | 1.24 |
| RP_POS_782_56854a11_145 | | 782.5685 | 11.15 | | RP | POS | | | 1687811 | 347266 | 1594402 | 330684 | 4.45E-02 | -0.28 | 1.21 |
| RP_NEG_552_33136a10_376 | | 552.3314 | 10.38 | | RP | NEG | | | 2425058 | 796255 | 2653218 | 851461 | 4.46E-02 | 0.28 | 1.24 |
| HILIC_NEG_448_30725a1_005 | Glycohyodeoxycholic acid | 448.3073 | 1.01 | | HILIC | NEG | 448.3047(100), 74.0239(90) | 1 | 711498 | 913681 | 518531 | 441255 | 4.46E-02 | -0.28 | 1.14 |
| RP_NEG_646_42792a11_238 | | 646.4279 | 11.24 | | RP | NEG | | | 1043824 | 1186007 | 1380194 | 1249662 | 4.49E-02 | 0.28 | 1.39 |
| RP_POS_670_46851a11_354 | | 670.4685 | 11.35 | | RP | POS | | | 188764 | 116415 | 229566 | 169967 | 4.49E-02 | 0.28 | 1.18 |
| RP_POS_589_53381a11_144 | | 589.5338 | 11.14 | | RP | POS | | | 353029 | 165303 | 311855 | 134601 | 4.49E-02 | -0.27 | 1.45 |
| RP_POS_782_56769a11_594 | | 782.5677 | 11.59 | | RP | POS | | | 2342310 | 480489 | 2219604 | 413670 | 4.50E-02 | -0.27 | 1.3 |
| RP_POS_856_58154a13_785 | | 856.5815 | 13.79 | | RP | POS | | | 3.1E+07 | 1.2E+07 | 2.8E+07 | 1E+07 | 4.50E-02 | -0.27 | 1.19 |
| RP_POS_691_96838a10_397 | | 691.9684 | 10.40 | | RP | POS | | | 1293043 | 599573 | 1127715 | 600983 | 4.50E-02 | -0.28 | 1.26 |
| RP_NEG_619_28943a11_347 | | 619.2894 | 11.35 | | RP | NEG | | | 159044 | 199742 | 251296 | 415052 | 4.51E-02 | 0.3 | 1.22 |
| RP_POS_538_72467a10_722 | | 538.7247 | 10.72 | | RP | POS | | | 124507 | 74829.1 | 150900 | 110456 | 4.52E-02 | 0.28 | 1.4 |
| RP_POS_302_93671a15_398 | | 302.9367 | 15.40 | | RP | POS | | | 8008757 | 1998152 | 7470303 | 1922516 | 4.53E-02 | -0.27 | 1.36 |
| RP_NEG_283_3898a11_591 | | 283.3898 | 11.59 | | RP | NEG | | | 175143 | 63761.7 | 157931 | 61638.4 | 4.54E-02 | -0.27 | 1.47 |
| RP_POS_403_28156a10_829 | | 403.2816 | 10.83 | | RP | POS | | | 337438 | 238109 | 442487 | 469251 | 4.55E-02 | 0.3 | 1.45 |
| RP_NEG_339_39728a10_65 | | 339.3973 | 10.65 | | RP | NEG | | | 586094 | 325086 | 681644 | 365595 | 4.55E-02 | 0.28 | 1.46 |
| RP_NEG_631_10425a10_548 | | 631.1043 | 10.55 | | RP | NEG | | | 619310 | 317000 | 709451 | 336997 | 4.55E-02 | 0.28 | 1.45 |
| RP_POS_244_07898a0_578 | | 244.0790 | 0.58 | | RP | POS | | | 911400 | 271900 | 989260 | 292674 | 4.55E-02 | 0.28 | 1.14 |
| RP_NEG_299_25946a10_621 | | 299.2595 | 10.62 | | RP | NEG | | | 3580829 | 515601 | 3445970 | 469149 | 4.55E-02 | -0.27 | 1.47 |
| RP_NEG_381_17429a11_238 | | 381.1743 | 11.24 | | RP | NEG | | | 7034834 | 6112814 | 8781485 | 6557598 | 4.56E-02 | 0.28 | 1.39 |
| RP_NEG_400_1478a11_238 | | 400.1478 | 11.24 | | RP | NEG | | | 717755 | 802417 | 948265 | 869255 | 4.56E-02 | 0.28 | 1.4 |
| RP_POS_163_11182a9_255 | | 163.1118 | 9.26 | | RP | POS | | | 6513922 | 2121224 | 7095495 | 2116637 | 4.57E-02 | 0.27 | 1.38 |
| RP_NEG_612_33124a10_283 | | 612.3312 | 10.28 | | RP | NEG | | | 1.6E+07 | 4542625 | 1.4E+07 | 4284423 | 4.57E-02 | -0.27 | 1.27 |
| HILIC_POS_181_05254a1_503 | | 181.0525 | 1.50 | | HILIC | POS | | | 616109 | 136656 | 585344 | 85885.5 | 4.57E-02 | -0.28 | 1.17 |
| RP_NEG_539_25043a8_457 | | 539.2504 | 8.46 | | RP | NEG | | | 561029 | 572091 | 427328 | 401643 | 4.58E-02 | -0.27 | 1.18 |
| RP_NEG_743_16388a8_847 | | 743.1639 | 8.85 | | RP | NEG | | | 767354 | 50570.5 | 754352 | 44510 | 4.58E-02 | -0.27 | 1.17 |
| RP_POS_580_29248a10_476 | | 580.2925 | 10.48 | | RP | POS | | | 291328 | 131318 | 258067 | 112006 | 4.58E-02 | -0.27 | 1.12 |
| HILIC_NEG_168_9894a1_352 | | 168.9894 | 1.35 | | HILIC | NEG | | | 641311 | 213475 | 704809 | 245698 | 4.59E-02 | 0.28 | 1.23 |
| HILIC_NEG_550_7641a2_992 | | 550.7641 | 2.99 | | HILIC | NEG | | | 217984 | 37446.7 | 228712 | 40455 | 4.59E-02 | 0.28 | 1.13 |
| RP_POS_201_1122a7_362 | | 201.1122 | 7.36 | | RP | POS | | | 1314646 | 624270 | 1529425 | 894493 | 4.60E-02 | 0.28 | 1.54 |
| RP_POS_546_35303a10_813 | | 546.3530 | 10.81 | | RP | POS | | | 1.9E+07 | 5463861 | 2.1E+07 | 6738666 | 4.61E-02 | 0.28 | 1.34 |
| RP_POS_670_99518a6_429 | | 670.9952 | 6.43 | | RP | POS | | | 554526 | 579082 | 412989 | 457313 | 4.61E-02 | -0.27 | 1.64 |
| RP_NEG_199_80502a0_563 | | 199.8050 | 0.56 | | RP | NEG | | | 4.5E+07 | 1.3E+07 | 4.1E+07 | 1.2E+07 | 4.62E-02 | -0.27 | 1.1 |
| RP_POS_371_22461a11_595 | | 371.2246 | 11.60 | | RP | POS | | | 4645089 | 1060273 | 4388323 | 819910 | 4.62E-02 | -0.27 | 1.31 |
| HILIC_NEG_144_92355a4_867 | | 144.9236 | 4.87 | | HILIC | NEG | | | 4.6E+07 | 1.7E+07 | 5.1E+07 | 1.9E+07 | 4.63E-02 | 0.28 | 1.43 |
| RP_POS_290_23221a9_215 | | 290.2322 | 9.22 | | RP | POS | | | 1396309 | 421641 | 1288822 | 366460 | 4.63E-02 | -0.27 | 1.11 |
| RP_POS_298_87204a15_412 | | 298.8720 | 15.41 | | RP | POS | | | 2604007 | 862760 | 2351232 | 971353 | 4.63E-02 | -0.28 | 1.38 |
| RP_POS_573_74158a10_42 | | 573.7416 | 10.42 | | RP | POS | | | 280188 | 141030 | 323965 | 174213 | 4.64E-02 | 0.28 | 1.57 |
| RP_NEG_407_20816a6_482 | | 407.2082 | 6.48 | | RP | NEG | | | 207472 | 83383.3 | 228318 | 69512.8 | 4.64E-02 | 0.27 | 1.15 |
| RP_POS_490_42538a14_232 | alpha-Tocopherol acetate (vitamin E acetate) | 490.4254 | 14.23 | HMDB0034227 | RP | POS | 207.1014(100) 165.091(30) | 2 | 868250 | 488977 | 1063124 | 854254 | 4.64E-02 | 0.29 | 1.41 |
| RP_POS_448_30576a8_735 | | 448.3058 | 8.74 | HMDB0000138 | RP | POS | 412.2868(100), 337.2515(75) | | 3166972 | 8383347 | 1565380 | 1825120 | 4.64E-02 | -0.31 | 1.1 |
| RP_NEG_427_16486a10_961 | | 427.1649 | 10.96 | | RP | NEG | | | 966282 | 343310 | 1059202 | 337373 | 4.67E-02 | 0.27 | 1.37 |
| RP_POS_417_33621a10_072 | | 417.3362 | 10.07 | | RP | POS | | | 1348533 | 354056 | 1448695 | 376605 | 4.67E-02 | 0.27 | 1.14 |
| HILIC_POS_127_12313a1_63 | | 127.1231 | 1.63 | | HILIC | POS | | | 478788 | 448479 | 383203 | 236605 | 4.70E-02 | -0.28 | 1.27 |
| RP_NEG_172_82964a7_267 | | 172.8296 | 7.27 | | RP | NEG | | | 999158 | 337532 | 1087556 | 312373 | 4.71E-02 | 0.27 | 1.23 |
| HILIC_NEG_186_03836a5_795 | | 186.0384 | 5.80 | | HILIC | NEG | | | 130151 | 34054.6 | 139234 | 32713.9 | 4.73E-02 | 0.27 | 1.16 |
| RP_POS_565_56555a15_293 | | 565.5656 | 15.29 | | RP | POS | | | 2.9E+07 | 7995367 | 3.1E+07 | 1.1E+07 | 4.74E-02 | 0.28 | 1.36 |
| RP_POS_143_07307a2_882 | | 143.0731 | 2.88 | | RP | POS | | | 826084 | 173111 | 872340 | 167056 | 4.74E-02 | 0.27 | 1.41 |
| RP_POS_525_36798a9_875 | | 525.3680 | 9.88 | | RP | POS | | | 316274 | 154141 | 373752 | 248838 | 4.76E-02 | 0.29 | 1.37 |
| RP_POS_610_62549a15_287 | | 610.6255 | 15.29 | | RP | POS | | | 852029 | 422064 | 986835 | 548985 | 4.76E-02 | 0.28 | 1.23 |
| HILIC_NEG_126_00328a5_35 | | 126.0033 | 5.35 | | HILIC | NEG | | | 224717 | 63878.7 | 243729 | 74479.3 | 4.76E-02 | 0.27 | 1.18 |
| HILIC_POS_102_05543a5_795 | 1-Aminocyclopropane-1-carboxylic acid | 102.0554 | 5.80 | | HILIC | POS | 102.0921(100), 56.0503(80) | 1 | 1328084 | 389311 | 1438335 | 418431 | 4.77E-02 | 0.27 | 1.21 |
| RP_POS_129_03047a15_416 | | 129.0305 | 15.42 | | RP | POS | | | 1163836 | 694564 | 1005795 | 463507 | 4.77E-02 | -0.27 | 1.21 |
| HILIC_POS_128_01797a5_348 | | 128.0180 | 5.35 | | HILIC | POS | | | 440303 | 158339 | 484650 | 166968 | 4.78E-02 | 0.27 | 1.28 |
| RP_NEG_383_15359a5_646 | | 383.1536 | 5.65 | | RP | NEG | | | 434121 | 280960 | 511359 | 286838 | 4.79E-02 | 0.27 | 1.18 |
| RP_POS_233_18985a10_39 | | 233.1899 | 10.39 | | RP | POS | | | 904604 | 473902 | 1039110 | 511950 | 4.79E-02 | 0.27 | 1.47 |
| RP_NEG_616_36249a10_672 | | 616.3625 | 10.67 | | RP | NEG | | | 995581 | 287332 | 1084486 | 357390 | 4.80E-02 | 0.28 | 1.23 |
| RP_POS_447_16138a8_081 | | 447.1614 | 8.08 | | RP | POS | | | 218729 | 42696.1 | 232011 | 53537 | 4.80E-02 | 0.28 | 1.19 |
| RP_POS_626_77631a10_61 | | 626.7763 | 10.61 | | RP | POS | | | 184597 | 131160 | 225927 | 167737 | 4.80E-02 | 0.28 | 1.33 |
| RP_POS_287_19644a0_586 | | 287.1964 | 0.59 | | RP | POS | | | 7803772 | 1.5E+07 | 4628277 | 8226317 | 4.80E-02 | -0.28 | 1.05 |
| RP_NEG_199_80527a3_675 | | 199.8053 | 3.68 | | RP | NEG | | | 461718 | 228719 | 405703 | 185325 | 4.81E-02 | -0.27 | 1.16 |
| RP_NEG_498_22916a12_099 | | 498.2292 | 12.10 | | RP | NEG | | | 334408 | 49962.5 | 319396 | 59375.8 | 4.81E-02 | -0.27 | 1.47 |
| RP_NEG_512_26917a8_962 | | 512.2692 | 8.96 | | RP | NEG | | | 9071926 | 7556555 | 7173835 | 6482374 | 4.82E-02 | -0.27 | 1.27 |
| RP_NEG_302_8739a15_336 | | 302.8739 | 15.34 | | RP | NEG | | | 337165 | 82825.6 | 312709 | 95826.7 | 4.83E-02 | -0.27 | 1.16 |
| RP_NEG_528_30957a10_295 | | 528.3096 | 10.30 | | RP | NEG | | | 1084213 | 253784 | 1154330 | 262476 | 4.84E-02 | 0.27 | 1.22 |
| RP_NEG_588_89856a11_11 | | 588.8986 | 11.11 | | RP | NEG | | | 410859 | 124494 | 377734 | 120210 | 4.84E-02 | -0.27 | 1.17 |
| RP_POS_752_52191a12_202 | | 752.5219 | 12.20 | | RP | POS | | | 3353514 | 2676557 | 2720288 | 2001422 | 4.84E-02 | -0.27 | 1.12 |
| RP_NEG_246_07498a6_013 | | 246.0750 | 6.01 | | RP | NEG | | | 905918 | 1997044 | 511092 | 722430 | 4.84E-02 | -0.29 | 1.12 |
| RP_NEG_379_1799a11_237 | | 379.1799 | 11.24 | | RP | NEG | | | 337736 | 306095 | 424819 | 333614 | 4.85E-02 | 0.27 | 1.38 |

**Supplemantary table 1**: The non-targeted metabolomics results ordered by the VIP value (p value < 0.0003 are shaded dark green, p value < 0.05 are shaded light green, d values below 0 are blue and above 0 are red)

| | | | | | | | | | Cases | | Controls | | Welch test | Cohen's | |
|---|---|---|---|---|---|---|---|---|---|---|---|---|---|---|---|
| Feature_ID | Metabolite_name | Average_Mz | Average_RT_min_ | HMDB ID | RP_HILIC | POS_NEG | Cureted_MSMS | ID_level | Mean | SD | Mean2 | SD3 | p | d | VIP |
| RP_POS_565_71606a10_149 | | 565.7161 | 10.15 | | RP | POS | | | 459185 | 195809 | 530989 | 311214 | 4.86E-02 | 0.28 | 1.35 |
| RP_POS_293_17502a8_67 | | 293.1750 | 8.67 | | RP | POS | | | 1.1E+07 | 658201 | 1.1E+07 | 710823 | 4.87E-02 | 0.27 | 1.45 |
| RP_NEG_663_13013a10_548 | | 663.1301 | 10.55 | | RP | NEG | | | 456680 | 264424 | 532372 | 291814 | 4.88E-02 | 0.27 | 1.47 |
| RP_NEG_294_98212a7_586 | | 294.9821 | 7.59 | | RP | NEG | | | 1158494 | 408003 | 1048891 | 403064 | 4.90E-02 | -0.27 | 1.18 |
| RP_NEG_502_29401a10_487 | | 502.2940 | 10.49 | | RP | NEG | | | 1736967 | 542076 | 1908499 | 703055 | 4.92E-02 | 0.28 | 1.23 |
| HILIC_NEG_308_99844a1_23 | | 308.9984 | 1.23 | | HILIC | NEG | | | 1326545 | 477913 | 1202667 | 441439 | 4.92E-02 | -0.27 | 1.22 |
| RP_POS_430_29517a8_735 | | 430.2952 | 8.74 | | RP | POS | | | 2532949 | 6884683 | 1234510 | 1492086 | 4.92E-02 | -0.31 | 1.09 |
| RP_POS_194_1153a10_363 | | 194.1153 | 10.36 | | RP | POS | 194.1544(100) | | 2140066 | 447976 | 2014282 | 480143 | 4.93E-02 | -0.27 | 1.17 |
| RP_POS_790_58844a10_154 | | 790.5884 | 10.15 | | RP | POS | | | 1847005 | 1049843 | 1596374 | 810761 | 4.93E-02 | -0.27 | 1.22 |
| HILIC_NEG_239_08559a0_945 | | 239.0856 | 0.95 | | HILIC | NEG | | | 1802746 | 1783821 | 1428378 | 921099 | 4.93E-02 | -0.28 | 1.07 |
| RP_POS_468_84433a10_268 | | 468.8443 | 10.27 | | RP | POS | | | 1020068 | 535141 | 877659 | 521108 | 4.94E-02 | -0.27 | 1.14 |
| HILIC_POS_241_15448a0_788 | | 241.1545 | 0.79 | | HILIC | POS | | | 136078 | 290244 | 220166 | 328459 | 4.95E-02 | 0.27 | 1.23 |
| RP_NEG_397_20599a8_731 | | 397.2060 | 8.73 | | RP | NEG | | | 4750648 | 6027920 | 3310854 | 4680052 | 4.96E-02 | -0.27 | 1.13 |
| HILIC_POS_129_06598a5_715 | | 129.0660 | 5.72 | | HILIC | POS | | | 6379042 | 4970015 | 5381686 | 2063229 | 4.96E-02 | -0.28 | 1.07 |
| RP_POS_672_80298a11_423 | | 672.8030 | 11.42 | | RP | POS | | | 162706 | 80364.2 | 190025 | 115967 | 4.97E-02 | 0.28 | 1.19 |
| RP_POS_329_15271a0_573 | | 329.1527 | 0.57 | | RP | POS | | | 1307437 | 293272 | 1382591 | 266025 | 4.97E-02 | 0.27 | 1.14 |
| RP_POS_623_29041a11_312 | | 623.2904 | 11.31 | | RP | POS | | | 111172 | 50936.5 | 127192 | 65681.1 | 4.97E-02 | 0.27 | 1.17 |
| RP_POS_141_95885a0_951 | | 141.9589 | 0.95 | | RP | POS | | | 1.6E+07 | 1.1E+07 | 2E+07 | 1.4E+07 | 4.98E-02 | 0.27 | 1.24 |
| RP_POS_402_2438a11_146 | | 402.2438 | 11.15 | | RP | POS | | | 542077 | 77795.2 | 522144 | 70675.8 | 4.99E-02 | -0.27 | 1.14 |
| RP_POS_412_28442a8_735 | | 412.2844 | 8.74 | | RP | POS | | | 1203730 | 3264311 | 587703 | 756511 | 4.99E-02 | -0.31 | 1.09 |
| RP_NEG_349_23648a11_238 | | 349.2365 | 11.24 | | RP | NEG | | | 2148045 | 1081275 | 2444868 | 1119893 | 5.00E-02 | 0.27 | 1.45 |
| RP_POS_257_22623a8_762 | | 257.2262 | 8.76 | | RP | POS | | | 2475855 | 778928 | 2781604 | 1363464 | 5.01E-02 | 0.29 | 1.21 |
| RP_POS_578_737a10_143 | | 578.7370 | 10.14 | | RP | POS | | | 593512 | 295850 | 690418 | 406123 | 5.01E-02 | 0.28 | 1.26 |
| HILIC_POS_394_31708a6_697 | | 394.3171 | 6.70 | | HILIC | POS | | | 1043901 | 356860 | 1150714 | 427499 | 5.01E-02 | 0.27 | 1.12 |
| HILIC_NEG_131_07138a0_619 | Leucic acid | 131.0714 | 0.62 | | HILIC | NEG | 85.0653(100), 131.0718(60) | 2 | 1384636 | 560096 | 1242109 | 503046 | 5.02E-02 | -0.27 | 1.14 |
| RP_POS_466_3165a8_735 | Glycocholic acid | 466.3165 | 8.74 | | RP | POS | 412.2863(100),337.2516(60) | 1 | 9625564 | 2.6E+07 | 4759539 | 5537026 | 5.02E-02 | -0.31 | 1.09 |
| HILIC_POS_675_54364a0_852 | | 675.5436 | 0.85 | | HILIC | POS | | | 1555341 | 421944 | 1450122 | 363388 | 5.02E-02 | -0.27 | 1.24 |
| RP_POS_447_34549a12_335 | | 447.3455 | 12.34 | | RP | POS | | | 4190991 | 1027689 | 4601861 | 1842950 | 5.03E-02 | 0.29 | 1.21 |
| HILIC_POS_118_08641a5_358 | | 118.0864 | 5.36 | | HILIC | POS | 140.069(100), 74.0974(40) | | 1.2E+07 | 5424614 | 1.4E+07 | 5529422 | 5.03E-02 | 0.27 | 1.1 |
| RP_POS_804_55109a13_293 | | 804.5511 | 13.29 | | RP | POS | | | 2.2E+07 | 1.2E+07 | 2.5E+07 | 1.1E+07 | 5.03E-02 | 0.27 | 1.13 |
| RP_POS_486_32736a10_041 | | 486.3274 | 10.04 | | RP | POS | | | 325271 | 192878 | 273045 | 195362 | 5.03E-02 | -0.27 | 1.41 |
| HILIC_NEG_199_03809a2_271 | | 199.0381 | 2.27 | | HILIC | NEG | | | 553922 | 172860 | 607501 | 219070 | 5.04E-02 | 0.27 | 1.22 |
| RP_NEG_130_94415a12_937 | | 130.9442 | 12.94 | | RP | NEG | | | 2314424 | 861471 | 2574734 | 1050069 | 5.04E-02 | 0.27 | 1.14 |
| RP_POS_407_31323a11_137 | | 407.3132 | 11.14 | | RP | POS | | | 912099 | 178503 | 862976 | 186334 | 5.04E-02 | -0.27 | 1.24 |
| RP_POS_555_17474a10_548 | | 555.1747 | 10.55 | | RP | POS | | | 2.1E+07 | 1.2E+07 | 2.4E+07 | 1.3E+07 | 5.05E-02 | 0.27 | 1.47 |
| RP_POS_460_33218a10_277 | | 460.3322 | 10.28 | | RP | POS | | | 423153 | 256173 | 363973 | 184505 | 5.05E-02 | -0.27 | 1.1 |
| HILIC_NEG_99_92572a6_29 | | 99.9257 | 6.29 | | HILIC | NEG | | | 6712643 | 4186514 | 5624302 | 3936962 | 5.06E-02 | -0.27 | 1.17 |
| RP_POS_172_07571a6_03 | | 172.0757 | 6.03 | | RP | POS | | | 916081 | 663725 | 632585 | 1294032 | 5.06E-02 | -0.29 | 1.31 |
| RP_POS_285_18539a10_652 | | 285.1854 | 10.65 | | RP | POS | | | 2857069 | 1485894 | 3278700 | 1639492 | 5.08E-02 | 0.27 | 1.42 |
| RP_POS_341_2475a10_657 | | 341.2475 | 10.66 | | RP | POS | | | 949487 | 524206 | 1095509 | 560886 | 5.09E-02 | 0.27 | 1.47 |
| HILIC_POS_156_0768a7_094 | | 156.0768 | 7.09 | | HILIC | POS | 110.0716(1009, 156.0768(40) | | 1656279 | 505246 | 1542643 | 339788 | 5.09E-02 | -0.27 | 1.1 |
| HILIC_POS_105_0428a0_927 | | 105.0428 | 0.93 | | HILIC | POS | | | 1158204 | 126315 | 1123528 | 131773 | 5.09E-02 | -0.27 | 1.16 |
| RP_POS_158_11763a0_677 | | 158.1176 | 0.68 | | RP | POS | | | 582051 | 344978 | 501642 | 255552 | 5.09E-02 | -0.27 | 1.09 |
| RP_POS_311_30554a10_68 | | 311.3055 | 10.68 | | RP | POS | | | 1463346 | 2695669 | 2158148 | 2501303 | 5.10E-02 | 0.27 | 1.13 |
| RP_NEG_243_75537a15_494 | | 243.7554 | 15.49 | | RP | NEG | | | 1.4E+07 | 290070 | 1.4E+07 | 274523 | 5.10E-02 | -0.27 | 1.17 |
| RP_POS_290_66806a10_928 | | 290.6681 | 10.93 | | RP | POS | | | 609403 | 393268 | 512179 | 334853 | 5.10E-02 | -0.27 | 1.18 |
| RP_NEG_403_16431a11_147 | | 403.1643 | 11.15 | | RP | NEG | | | 1461015 | 310261 | 1381619 | 284147 | 5.11E-02 | -0.27 | 1.11 |
| RP_NEG_182_04597a0_682 | | 182.0460 | 0.68 | | RP | NEG | | | 377953 | 693348 | 236923 | 321625 | 5.11E-02 | -0.28 | 1.06 |
| RP_POS_223_09398a7_361 | | 223.0940 | 7.36 | | RP | POS | | | 2699058 | 1312407 | 3146752 | 1922635 | 5.14E-02 | 0.28 | 1.54 |
| RP_NEG_687_53821a12_799 | | 687.5382 | 12.80 | | RP | NEG | | | 235304 | 147950 | 275318 | 150901 | 5.14E-02 | 0.27 | 1.39 |
| HILIC_POS_120_06583a5_795 | Threonine | 120.0658 | 5.80 | | HILIC | POS | 74.0611(100), 56.0504(50) | 2 | 1.2E+07 | 3597544 | 1.3E+07 | 3826805 | 5.15E-02 | 0.27 | 1.19 |
| RP_NEG_567_15497a10_547 | | 567.1550 | 10.55 | | RP | NEG | | | 461148 | 307662 | 546944 | 331913 | 5.17E-02 | 0.27 | 1.5 |
| RP_POS_671_32953a6_43 | | 671.3295 | 6.43 | | RP | POS | | | 612549 | 534984 | 473827 | 505441 | 5.17E-02 | -0.27 | 1.7 |
| RP_POS_523_86133a10_217 | | 523.8613 | 10.22 | | RP | POS | | | 2779000 | 1366332 | 2460142 | 1025883 | 5.18E-02 | -0.27 | 1.21 |
| HILIC_NEG_253_73727a1_407 | | 253.7373 | 1.41 | | HILIC | NEG | | | 191528 | 93864 | 214892 | 81735.7 | 5.19E-02 | 0.27 | 1.21 |
| RP_POS_141_05464a7_363 | | 141.0546 | 7.36 | | RP | POS | | | 706228 | 283316 | 806940 | 440465 | 5.20E-02 | 0.28 | 1.51 |
| RP_POS_362_88617a15_374 | | 362.8862 | 15.37 | | RP | POS | | | 824275 | 333956 | 935517 | 474885 | 5.20E-02 | 0.28 | 1.19 |
| RP_NEG_397_33334a11_309 | | 397.3333 | 11.31 | | RP | NEG | | | 459278 | 51557.6 | 444703 | 57044.5 | 5.20E-02 | -0.27 | 1.35 |
| RP_POS_601_26398a5_502 | | 601.2640 | 5.50 | | RP | POS | | | 964801 | 1099824 | 720241 | 725239 | 5.20E-02 | -0.27 | 1.21 |
| RP_NEG_469_22662a7_973 | | 469.2266 | 7.97 | | RP | NEG | | | 359554 | 426949 | 268043 | 250548 | 5.21E-02 | -0.27 | 1.09 |
| RP_POS_293_20865a10_095 | | 293.2087 | 10.10 | | RP | POS | | | 2283960 | 258039 | 2218310 | 235616 | 5.21E-02 | -0.27 | 1.34 |
| RP_NEG_201_02277a4_88 | | 201.0228 | 4.88 | | RP | NEG | | | 1020885 | 1459014 | 1314892 | 660342 | 5.22E-02 | 0.28 | 1.2 |
| HILIC_NEG_262_98627a2_269 | | 262.9863 | 2.27 | | HILIC | NEG | | | 542471 | 261632 | 625985 | 350442 | 5.22E-02 | 0.27 | 1.25 |
| RP_NEG_486_80011a15_385 | | 486.8001 | 15.39 | | RP | NEG | | | 961658 | 193782 | 905644 | 222740 | 5.22E-02 | -0.27 | 1.31 |
| RP_POS_587_41504a11_481 | | 587.4150 | 11.48 | | RP | POS | | | 209644 | 78141.8 | 237107 | 119889 | 5.23E-02 | 0.28 | 1.19 |
| RP_POS_648_93323a9_928 | | 648.9332 | 9.93 | | RP | POS | | | 1158662 | 598325 | 1002474 | 575553 | 5.23E-02 | -0.27 | 1.26 |
| RP_NEG_500_27859a10_185 | | 500.2786 | 10.19 | | RP | NEG | | | 2718841 | 643997 | 2911906 | 785922 | 5.24E-02 | 0.27 | 1.36 |
| HILIC_POS_302_0199a6_307 | | 302.0199 | 6.31 | | HILIC | POS | | | 528866 | 234939 | 473669 | 180525 | 5.24E-02 | -0.27 | 1.29 |
| RP_NEG_244_90823a6_688 | | 244.9082 | 6.69 | | RP | NEG | | | 447364 | 376760 | 546972 | 371583 | 5.25E-02 | 0.27 | 1.24 |
| RP_POS_303_92227a15_452 | | 303.9223 | 15.45 | | RP | POS | | | 742988 | 567635 | 960115 | 974741 | 5.26E-02 | 0.28 | 1.19 |
| RP_NEG_832_60913a14_915 | PC 36:1 (18:0_18:1) | 832.6091 | 14.92 | | RP | NEG | 281.2493(100), 772.5884(10) | 2 | 4652535 | 3260818 | 5505933 | 3162458 | 5.27E-02 | 0.27 | 1.2 |
| HILIC_NEG_209_06699a2_275 | | 209.0670 | 2.28 | | HILIC | NEG | | | 4301334 | 1276434 | 4683946 | 1562681 | 5.29E-02 | 0.27 | 1.2 |
| RP_NEG_150_00197a10_546 | | 150.0020 | 10.55 | | RP | NEG | 150.0026(100) | | 721629 | 498874 | 860544 | 542082 | 5.30E-02 | 0.27 | 1.5 |
| RP_NEG_590_34674a10_39 | | 590.3467 | 10.39 | | RP | NEG | | | 2552197 | 715835 | 2758045 | 820430 | 5.30E-02 | 0.27 | 1.19 |
| RP_NEG_381_19455a11_592 | | 381.1946 | 11.59 | | RP | NEG | | | 2166697 | 134290 | 2128189 | 153445 | 5.32E-02 | -0.27 | 1.38 |
| RP_NEG_525_27081a7_119 | | 525.2708 | 7.12 | | RP | NEG | | | 575690 | 278682 | 662199 | 360735 | 5.33E-02 | 0.27 | 1.1 |
| RP_POS_576_27917a10_356 | | 576.2792 | 10.36 | | RP | POS | | | 362306 | 156664 | 410648 | 201080 | 5.34E-02 | 0.27 | 1.18 |
| HILIC_POS_86_09708a3_962 | Piperidine | 86.0971 | 3.96 | | HILIC | POS | 86.0968(100), 69.0711(2) | 2 | 5.3E+07 | 8.9E+07 | 3.3E+07 | 6.2E+07 | 5.34E-02 | -0.27 | 1.14 |
| RP_POS_383_35229a11_208 | | 383.3523 | 11.21 | | RP | POS | | | 669031 | 170014 | 626848 | 149090 | 5.34E-02 | -0.26 | 1.2 |
| HILIC_POS_170_05994a4_092 | | 170.0599 | 4.09 | | HILIC | POS | | | 176663 | 142491 | 215882 | 152283 | 5.35E-02 | 0.27 | 1.15 |
| RP_POS_358_27399a10_651 | | 358.2740 | 10.65 | | RP | POS | | | 2.8E+07 | 1.5E+07 | 3.3E+07 | 1.7E+07 | 5.35E-02 | 0.27 | 1.41 |
| RP_NEG_766_56128a10_475 | | 766.5613 | 10.48 | | RP | NEG | | | 515163 | 78898.3 | 495339 | 71104.9 | 5.35E-02 | -0.26 | 1.28 |
| RP_POS_236_07159a6_199 | | 236.0716 | 6.20 | | RP | POS | | | 3.7E+07 | 1.6E+07 | 3.3E+07 | 1.4E+07 | 5.37E-02 | -0.26 | 1.5 |
| RP_POS_629_29767a11_527 | | 629.2977 | 11.53 | | RP | POS | | | 119067 | 65463.4 | 138033 | 76438.4 | 5.39E-02 | 0.27 | 1.12 |
| RP_POS_649_40094a12_173 | | 649.4009 | 12.17 | | RP | POS | | | 108332 | 35526.6 | 118578 | 41158.5 | 5.39E-02 | 0.27 | 1.21 |
| HILIC_POS_371_23996a3_95 | | 371.2400 | 3.95 | | HILIC | POS | | | 9916356 | 2249497 | 1.1E+07 | 2441325 | 5.41E-02 | 0.27 | 1.19 |
| RP_POS_416_31589a8_96 | Glycolithocholic acid | 416.3159 | 8.96 | | RP | POS | 76.0399(100), 323.2731(60) | 2 | 6300653 | 5239850 | 5016207 | 4506143 | 5.41E-02 | -0.26 | 1.24 |
| RP_POS_182_10246a0_586 | | 182.1025 | 0.59 | | RP | POS | | | 4305087 | 1072114 | 4626002 | 1320269 | 5.42E-02 | 0.27 | 1.15 |
| HILIC_POS_162_11237a5_46 | | 162.1124 | 5.46 | | HILIC | POS | 162.1133(100) | | 1810933 | 495637 | 1936802 | 459112 | 5.42E-02 | 0.26 | 1.29 |
| RP_NEG_409_25812a11_51 | | 409.2581 | 11.51 | | RP | NEG | | | 244336 | 29577.2 | 235350 | 37309.5 | 5.43E-02 | -0.27 | 1.25 |
| RP_POS_189_12341a0_676 | N-epsilon-Acetyllysine | 189.1234 | 0.68 | | RP | POS | 126.0916(100), 84.0815(60) | 2 | 1030748 | 195164 | 981968 | 175355 | 5.44E-02 | -0.26 | 1.12 |
| RP_NEG_474_23331a10_853 | | 474.2333 | 10.85 | | RP | NEG | | | 927548 | 705920 | 751183 | 633636 | 5.44E-02 | -0.26 | 1.16 |
| RP_POS_405_17224a10_392 | | 405.1722 | 10.39 | | RP | POS | | | 255324 | 195550 | 307086 | 196449 | 5.45E-02 | 0.26 | 1.22 |
| HILIC_POS_565_56549a10_678 | | 565.5655 | 10.68 | | HILIC | POS | | | 366229 | 75207.6 | 344634 | 86900.3 | 5.45E-02 | -0.27 | 1.16 |
| RP_NEG_352_08621a6_086 | | 352.0862 | 6.09 | | RP | NEG | | | 244265 | 226062 | 189763 | 188457 | 5.46E-02 | -0.26 | 1.07 |
| RP_POS_344_2796a8_259 | AC 12:0 (Dodecanoylcarnitine) | 344.2796 | 8.26 | | RP | POS | 85.0292(100), 344.2801(40) | 2 | 5479687 | 3448178 | 6772639 | 5846743 | 5.50E-02 | 0.28 | 1.27 |
| RP_POS_291_05295a6_5 | | 291.0530 | 6.50 | | RP | POS | | | 315129 | 99739.5 | 292466 | 72474.1 | 5.51E-02 | -0.26 | 1.12 |
| RP_POS_427_28409a11_313 | | 427.2841 | 11.31 | | RP | POS | | | 656484 | 798260 | 499461 | 338608 | 5.51E-02 | -0.28 | 1.18 |
| RP_NEG_351_16373a9_945 | | 351.1637 | 9.95 | | RP | NEG | | | 277111 | 252484 | 388525 | 523964 | 5.52E-02 | 0.29 | 1.15 |
| RP_POS_177_1273a10_651 | | 177.1273 | 10.65 | | RP | POS | | | 3.8E+07 | 2.1E+07 | 4.4E+07 | 2.3E+07 | 5.54E-02 | 0.26 | 1.41 |
| RP_NEG_385_16968a6_434 | | 385.1697 | 6.43 | | RP | NEG | | | 427212 | 243436 | 505031 | 333508 | 5.55E-02 | 0.27 | 1.12 |
| HILIC_NEG_358_99597a0_589 | | 358.9960 | 0.59 | | HILIC | NEG | | | 868853 | 323698 | 956444 | 340677 | 5.55E-02 | 0.26 | 1.12 |
| RP_POS_667_48029a10_42 | | 667.4803 | 10.42 | | RP | POS | | | 3942011 | 1587300 | 3496306 | 1780016 | 5.55E-02 | -0.26 | 1.28 |
| RP_POS_483_34265a8_736 | | 483.3427 | 8.74 | | RP | POS | | | 501907 | 1283205 | 260937 | 386425 | 5.56E-02 | -0.29 | 1.06 |
| HILIC_NEG_340_00003a5_589 | | 340.0000 | 5.59 | | HILIC | NEG | | | 349217 | 85849.4 | 374541 | 104361 | 5.57E-02 | 0.27 | 1.3 |
| HILIC_NEG_241_01053a5_11 | | 241.0105 | 5.11 | | HILIC | NEG | | | 416671 | 199230 | 472011 | 219631 | 5.57E-02 | 0.26 | 1.12 |
| RP_NEG_666_45258a11_592 | | 666.4526 | 11.59 | | RP | NEG | | | 4330898 | 292110 | 4244252 | 358128 | 5.57E-02 | -0.27 | 1.33 |
| HILIC_NEG_183_93456a5_928 | | 183.9346 | 5.93 | | HILIC | NEG | | | 2260229 | 413438 | 2373800 | 447764 | 5.58E-02 | 0.26 | 1.2 |
| RP_POS_860_49597a13_206 | | 860.4960 | 13.21 | | RP | POS | | | 895065 | 444989 | 1016457 | 477369 | 5.62E-02 | 0.26 | 1.22 |
| HILIC_NEG_115_92082a3_107 | | 115.9208 | 3.11 | | HILIC | NEG | | | 2.2E+07 | 6897009 | 2.4E+07 | 8128231 | 5.63E-02 | 0.26 | 1.16 |
| HILIC_NEG_220_97423a5_793 | | 220.9742 | 5.79 | | HILIC | NEG | | | 106575 | 22103.7 | 112415 | 22424.2 | 5.63E-02 | 0.26 | 1.13 |
| RP_POS_466_31638a7_884 | | 466.3164 | 7.88 | | RP | POS | | | 776885 | 1529114 | 474925 | 684019 | 5.65E-02 | -0.27 | 1.04 |
| RP_POS_530_32037a10_373 | | 530.3204 | 10.37 | | RP | POS | | | 1691794 | 546719 | 1841724 | 594724 | 5.69E-02 | 0.26 | 1.21 |
| RP_POS_441_17209a10_648 | | 441.1721 | 10.65 | | RP | POS | | | 339834 | 222512 | 403817 | 262019 | 5.70E-02 | 0.26 | 1.28 |
| RP_POS_242_17513a7_989 | | 242.1751 | 7.99 | | RP | POS | | | 269265 | 79845.1 | 250622 | 63239.4 | 5.70E-02 | -0.26 | 1.16 |
| RP_POS_316_32123a8_911 | | 316.3212 | 8.91 | | RP | POS | | | 883813 | 2369371 | 444188 | 664997 | 5.70E-02 | -0.29 | 1.16 |
| RP_NEG_721_46204a11_236 | | 721.4620 | 11.24 | | RP | NEG | | | 148321 | 106115 | 175752 | 104218 | 5.73E-02 | 0.26 | 1.4 |
| RP_NEG_285_20746a10_185 | | 285.2075 | 10.19 | | RP | NEG | | | 253034 | 116423 | 285266 | 129141 | 5.74E-02 | 0.26 | 1.26 |
| RP_POS_345_21692a11_591 | | 345.2169 | 11.59 | | RP | POS | | | 408207 | 103263 | 382256 | 96226.9 | 5.75E-02 | -0.26 | 1.18 |
| RP_POS_401_73816a12_932 | | 401.7382 | 12.93 | | RP | POS | | | 686466 | 250679 | 614434 | 295798 | 5.75E-02 | -0.26 | 1.12 |
| RP_POS_392_73242a12_586 | | 392.7324 | 12.59 | | RP | POS | | | 199668 | 175628 | 158039 | 144858 | 5.76E-02 | -0.26 | 1.24 |
| RP_POS_499_37131a10_241 | | 499.3713 | 10.24 | | RP | POS | | | 4734464 | 1741869 | 4288240 | 1685179 | 5.76E-02 | -0.26 | 1.14 |
| RP_NEG_165_97893a10_641 | 2-Mercaptobenzothiazole | 165.9789 | 10.64 | | RP | NEG | 165.9798100), 134.0069(15) | 2 | 231915 | 141984 | 274272 | 177854 | 5.77E-02 | 0.26 | 1.4 |
| HILIC_POS_590_32251a1_129 | | 590.3225 | 1.13 | | HILIC | POS | | | 210951 | 100524 | 184991 | 98830.3 | 5.77E-02 | -0.26 | 1.18 |
| HILIC_POS_396_03873a4_83 | | 396.0387 | 4.83 | | HILIC | POS | | | 108203 | 54279.6 | 95730.7 | 41700.1 | 5.78E-02 | -0.26 | 1.04 |
| RP_POS_223_13237a7_802 | | 223.1324 | 7.80 | | RP | POS | | | 4384660 | 298797 | 4459157 | 274941 | 5.79E-02 | 0.26 | 1.14 |
| RP_POS_424_26334a10_672 | | 424.2633 | 10.67 | | RP | POS | | | 234926 | 191348 | 290381 | 229410 | 5.79E-02 | 0.26 | 1.32 |
| RP_NEG_451_16437a10_926 | | 451.1644 | 10.93 | | RP | NEG | | | 415821 | 136656 | 451567 | 138033 | 5.82E-02 | 0.26 | 1.35 |
| RP_POS_236_07159a10_569 | | 236.0716 | 10.57 | | RP | POS | | | 1E+08 | 8013480 | 1E+08 | 9026440 | 5.82E-02 | -0.26 | 1.26 |
| RP_POS_360_32327a11_409 | | 360.3233 | 11.41 | | RP | POS | | | 654160 | 526955 | 822133 | 731338 | 5.84E-02 | 0.27 | 1.23 |
| RP_POS_533_19305a10_547 | | 533.1931 | 10.55 | | RP | POS | | | 2.9E+07 | 2.1E+07 | 3.5E+07 | 2.3E+07 | 5.84E-02 | 0.26 | 1.47 |
| RP_POS_617_47534a11_278 | | 617.4753 | 11.28 | | RP | POS | | | 899418 | 1038141 | 1211989 | 1324291 | 5.84E-02 | 0.26 | 1.26 |
| RP_POS_553_46075a11_143 | | 553.4608 | 11.14 | | RP | POS | | | 2800740 | 528419 | 2662996 | 531285 | 5.84E-02 | -0.26 | 1.39 |
| RP_POS_185_11511a14_244 | | 185.1151 | 14.24 | | RP | POS | | | 2327911 | 1117543 | 2681966 | 1538318 | 5.85E-02 | 0.27 | 1.43 |
| RP_POS_315_16193a11_143 | | 315.1619 | 11.14 | | RP | POS | | | 1259458 | 217607 | 1199888 | 238998 | 5.87E-02 | -0.26 | 1.17 |
| RP_POS_832_58185a13_957 | | 832.5819 | 13.96 | | RP | POS | | | 9.2E+07 | 2.5E+07 | 9.8E+07 | 2.5E+07 | 5.88E-02 | 0.26 | 1.29 |
| RP_NEG_460_21222a11_595 | | 460.2122 | 11.60 | | RP | NEG | | | 891385 | 89244.3 | 863955 | 117691 | 5.88E-02 | -0.27 | 1.25 |
| RP_POS_259_13068a8_419 | | 259.1307 | 8.42 | | RP | POS | | | 1209586 | 139123 | 1246052 | 141867 | 5.89E-02 | 0.26 | 1.13 |
| RP_POS_177_09088a7_07 | | 177.0909 | 7.07 | | RP | POS | | | 1132477 | 146152 | 1169873 | 142613 | 5.90E-02 | 0.26 | 1.21 |
| RP_POS_187_07112a2_227 | | 187.0711 | 2.23 | | RP | POS | | | 3429920 | 912196 | 3682976 | 1026161 | 5.90E-02 | 0.26 | 1.14 |
| RP_POS_282_11823a0_588 | | 282.1182 | 0.59 | | RP | POS | | | 550612 | 230908 | 612339 | 244031 | 5.91E-02 | 0.26 | 1.22 |
| RP_POS_332_24298a6_579 | | 332.2430 | 6.58 | | RP | POS | | | 536913 | 344259 | 632155 | 385932 | 5.91E-02 | 0.26 | 1.2 |
| RP_POS_416_18881a9_968 | | 416.1888 | 9.97 | | RP | POS | | | 203915 | 156188 | 170045 | 104265 | 5.91E-02 | -0.26 | 1.17 |
| RP_POS_341_07458a8_842 | | 341.0746 | 8.84 | | RP | POS | | | 263793 | 136567 | 230734 | 119589 | 5.92E-02 | -0.26 | 1.13 |
| HILIC_NEG_99_9257a1_331 | | 99.9257 | 1.33 | | HILIC | NEG | | | 2225926 | 759607 | 2420068 | 741164 | 5.93E-02 | 0.26 | 1.16 |
| HILIC_POS_450_3212a1_008 | Glycodeoxycholic acid | 450.3212 | 1.01 | HMDB0000631 | HILIC | POS | 414.3026(100), 76.0399(20) | 1 | 931442 | 1211513 | 683772 | 667077 | 5.93E-02 | -0.26 | 1.08 |
| HILIC_NEG_276_98718a5_116 | | 276.9872 | 5.12 | | HILIC | NEG | | | 513102 | 260393 | 591227 | 332478 | 5.94E-02 | 0.26 | 1.23 |
| RP_NEG_615_44916a14_036 | | 615.4492 | 14.04 | | RP | NEG | | | 594761 | 83961.5 | 567930 | 117620 | 5.95E-02 | -0.27 | 1.17 |
| HILIC_NEG_194_90591a0_934 | | 194.9059 | 0.93 | | HILIC | NEG | | | 193241 | 39699.2 | 183136 | 38530.6 | 5.96E-02 | -0.26 | 1.16 |
| RP_NEG_268_98499a15_477 | | 268.9850 | 15.48 | | RP | NEG | | | 1513489 | 111182 | 1483015 | 123202 | 5.97E-02 | -0.26 | 1.45 |
| RP_POS_617_75873a9_981 | | 617.7587 | 9.98 | | RP | POS | | | 140151 | 80358.3 | 165545 | 111014 | 5.98E-02 | 0.27 | 1.35 |
| RP_POS_311_22183a7_402 | | 311.2218 | 7.40 | | RP | POS | | | 101781 | 42332.7 | 114081 | 51606.4 | 5.99E-02 | 0.26 | 1.38 |
| RP_POS_674_87415a11_531 | | 674.8742 | 11.53 | | RP | POS | | | 115932 | 64595.5 | 135135 | 81606.5 | 5.99E-02 | 0.26 | 1.2 |
| RP_POS_575_11676a11_283 | | 575.1168 | 11.28 | | RP | POS | | | 74833.9 | 35997 | 90109.8 | 73004.4 | 6.00E-02 | 0.28 | 1.13 |
| RP_NEG_429_15994a10_961 | | 429.1599 | 10.96 | | RP | NEG | | | 343558 | 132852 | 377529 | 130459 | 6.00E-02 | 0.26 | 1.32 |
| RP_NEG_599_16638a10_547 | | 599.1664 | 10.55 | | RP | NEG | | | 420066 | 296752 | 500836 | 325669 | 6.00E-02 | 0.26 | 1.47 |
| RP_POS_127_07565a4_303 | | 127.0757 | 4.30 | | RP | POS | | | 1012186 | 263526 | 1087666 | 314264 | 6.00E-02 | 0.26 | 1.26 |
| HILIC_NEG_132_90033a0_942 | | 132.9003 | 0.94 | | HILIC | NEG | | | 1661113 | 360747 | 1747966 | 314440 | 6.01E-02 | 0.26 | 1.11 |
| RP_NEG_163_11208a10_65 | | 163.1121 | 10.65 | | RP | NEG | | | 659468 | 349345 | 750390 | 354844 | 6.02E-02 | 0.26 | 1.44 |
| RP_NEG_186_11388a11_144 | | 186.1139 | 11.14 | | RP | NEG | | | 403112 | 74010 | 421449 | 68479 | 6.02E-02 | 0.26 | 1.17 |
| RP_POS_203_13934a0_682 | | 203.1393 | 0.68 | | RP | POS | | | 2957604 | 752874 | 2766649 | 729639 | 6.03E-02 | -0.26 | 1.14 |
| RP_POS_642_01953a11_492 | | 642.0195 | 11.49 | | RP | POS | | | 123730 | 54938.8 | 140285 | 71082.9 | 6.05E-02 | 0.26 | 1.24 |

**Supplemantary table 1**: The non-targeted metabolomics results ordered by the VIP value (p value < 0.0003 are shaded dark green, p value < 0.05 are shaded light green, d values below 0 are blue and above 0 are red)

| | | | | | | | | | Cases | | Controls | | Welch test | Cohen's | |
|---|---|---|---|---|---|---|---|---|---|---|---|---|---|---|---|
| Feature_ID | Metabolite_name | Average_Mz | Average_RT_min_ | HMDB ID | RP_HILIC | POS_NEG | Cureted_MSMS | ID_level | Mean | SD | Mean2 | SD3 | p | d | VIP |
| RP_NEG_212_00256a2_608 | 3-Indoxyl sulfate | 212.0026 | 2.61 | | RP | NEG | 79.9569(100), 212.002(90) | 2 | 1.7E+07 | 9239487 | 1.5E+07 | 8549737 | 6.06E-02 | -0.26 | 1.17 |
| RP_NEG_464_30222a7_883 | | 464.3022 | 7.88 | | RP | NEG | 464.3029(100), 74.0244(70) | | 785711 | 1540869 | 486626 | 688314 | 6.07E-02 | -0.27 | 1.03 |
| RP_NEG_447_27039a12_107 | | 447.2704 | 12.11 | | RP | NEG | | | 260446 | 80135.5 | 239743 | 80641.5 | 6.08E-02 | -0.26 | 1.53 |
| RP_POS_287_12842a10_311 | | 287.1284 | 10.31 | | RP | POS | | | 914819 | 513935 | 800186 | 377536 | 6.08E-02 | -0.26 | 1.33 |
| RP_POS_826_53448a12_539 | | 826.5345 | 12.54 | | RP | POS | | | 4244425 | 1957033 | 3769489 | 1746334 | 6.09E-02 | -0.26 | 1.19 |
| RP_NEG_417_2247a11_237 | | 417.2247 | 11.24 | | RP | NEG | | | 1165530 | 572948 | 1312640 | 571411 | 6.11E-02 | 0.26 | 1.39 |
| HILIC_NEG_118_05107a5_796 | Threonine | 118.0511 | 5.80 | | HILIC | NEG | 74.0244(100), 118.0512(30) | 2 | 2136630 | 604745 | 2292885 | 610112 | 6.11E-02 | 0.26 | 1.12 |
| RP_POS_123_08065a6_041 | | 123.0807 | 6.04 | | RP | POS | | | 2109889 | 466210 | 1993947 | 437568 | 6.11E-02 | -0.26 | 1.23 |
| HILIC_POS_304_2114a1_503 | | 304.2114 | 1.50 | | HILIC | POS | | | 1716864 | 937369 | 1987883 | 1144271 | 6.14E-02 | 0.26 | 1.24 |
| RP_POS_279_95514a15_361 | | 279.9551 | 15.36 | | RP | POS | | | 1179137 | 596922 | 1344717 | 682351 | 6.14E-02 | 0.26 | 1.27 |
| HILIC_NEG_262_90735a5_352 | | 262.9074 | 5.35 | | HILIC | NEG | | | 351113 | 108624 | 379797 | 114290 | 6.15E-02 | 0.26 | 1.15 |
| RP_NEG_370_70547a15_361 | | 370.7055 | 15.36 | | RP | NEG | | | 1410310 | 160526 | 1374784 | 116377 | 6.15E-02 | -0.26 | 1.4 |
| RP_NEG_464_85629a15_357 | | 464.8563 | 15.36 | | RP | NEG | | | 988459 | 80218 | 961119 | 124229 | 6.15E-02 | -0.27 | 1.29 |
| RP_POS_782_56732a13_803 | | 782.5673 | 13.80 | | RP | POS | | | 2.2E+08 | 8E+07 | 2.4E+08 | 6.8E+07 | 6.18E-02 | 0.26 | 1.19 |
| HILIC_NEG_85_00391a5_135 | | 85.0039 | 5.14 | | HILIC | NEG | | | 5342411 | 2780632 | 6030554 | 2603112 | 6.20E-02 | 0.26 | 1.1 |
| HILIC_NEG_244_84854a4_827 | | 244.8485 | 4.83 | | HILIC | NEG | | | 182023 | 47829.8 | 171782 | 31819.7 | 6.20E-02 | -0.26 | 1.15 |
| RP_NEG_860_58234a10_647 | | 860.5823 | 10.65 | | RP | NEG | | | 2397266 | 1392502 | 2759190 | 1431393 | 6.22E-02 | 0.26 | 1.47 |
| RP_NEG_432_31232a9_776 | Glycolithocholic acid | 432.3123 | 9.78 | HMDB0002639 | RP | NEG | 74.0238(100), 432.3152(80) | 2 | 744890 | 886383 | 530301 | 796566 | 6.24E-02 | -0.26 | 1.16 |
| RP_NEG_339_23276a10_649 | | 339.2328 | 10.65 | | RP | NEG | | | 1.9E+08 | 1E+08 | 2.2E+08 | 1.1E+08 | 6.25E-02 | 0.26 | 1.41 |
| RP_POS_558_29071a8_458 | | 558.2907 | 8.46 | | RP | POS | | | 413259 | 455101 | 315264 | 309385 | 6.25E-02 | -0.26 | 1.1 |
| RP_POS_382_18314a10_547 | | 382.1831 | 10.55 | | RP | POS | | | 756417 | 601116 | 916976 | 649915 | 6.26E-02 | 0.26 | 1.44 |
| RP_NEG_248_96088a12_95 | | 248.9609 | 12.95 | | RP | NEG | | | 1632380 | 651034 | 1813067 | 750598 | 6.27E-02 | 0.26 | 1.08 |
| HILIC_POS_359_09106a6_137 | | 359.0911 | 6.14 | | HILIC | POS | | | 185951 | 31112.4 | 192820 | 22717.8 | 6.28E-02 | 0.26 | 1.1 |
| RP_POS_658_12543a11_46 | | 658.1254 | 11.46 | | RP | POS | | | 175799 | 95032.3 | 207485 | 143949 | 6.30E-02 | 0.27 | 1.2 |
| RP_POS_766_53754a13_316 | | 766.5375 | 13.32 | | RP | POS | | | 1631668 | 3246049 | 2801720 | 5452616 | 6.30E-02 | 0.27 | 1.33 |
| HILIC_NEG_219_84525a3_101 | | 219.8453 | 3.10 | | HILIC | NEG | | | 296437 | 70728.9 | 314629 | 71701.8 | 6.30E-02 | 0.26 | 1.22 |
| RP_NEG_605_40717a11_804 | | 605.4072 | 11.80 | | RP | NEG | | | 1025236 | 500072 | 911269 | 395714 | 6.30E-02 | -0.25 | 1.09 |
| RP_POS_155_97446a1_678 | | 155.9745 | 1.68 | | RP | POS | | | 2188058 | 196406 | 2241476 | 219566 | 6.31E-02 | 0.26 | 1.1 |
| RP_POS_999_63715a10_488 | | 999.6372 | 10.49 | | RP | POS | | | 491492 | 331427 | 592673 | 443880 | 6.32E-02 | 0.26 | 1.19 |
| RP_NEG_528_30969a10_656 | | 528.3097 | 10.66 | | RP | NEG | | | 474423 | 211431 | 538878 | 282786 | 6.33E-02 | 0.26 | 1.16 |
| RP_NEG_526_26074a12_734 | | 526.2607 | 12.73 | | RP | NEG | | | 171081 | 53690.2 | 158191 | 47771.1 | 6.33E-02 | -0.25 | 1.35 |
| RP_NEG_429_15396a11_593 | | 429.1540 | 11.59 | | RP | NEG | | | 353792 | 40591.8 | 342868 | 44655.4 | 6.33E-02 | -0.26 | 1.32 |
| RP_NEG_709_07507a9_099 | | 709.0751 | 9.10 | | RP | NEG | | | 360068 | 23999.1 | 353622 | 26317.6 | 6.33E-02 | -0.26 | 1.11 |
| RP_POS_333_27866a11_263 | | 333.2787 | 11.26 | | RP | POS | | | 542746 | 381580 | 637398 | 362509 | 6.34E-02 | 0.25 | 1.34 |
| RP_POS_130_06532a6_03 | Indole-3-carbinol | 130.0653 | 6.03 | | RP | POS | 130.0658(100), 103.0561(2) | 1 | 4714922 | 3450941 | 3316468 | 6726904 | 6.35E-02 | -0.27 | 1.28 |
| RP_POS_866_54175a14_087 | | 866.5418 | 14.09 | | RP | POS | | | 2072228 | 2097225 | 2665813 | 2510182 | 6.36E-02 | 0.26 | 1.31 |
| RP_POS_587_48853a11_235 | | 587.4885 | 11.24 | | RP | POS | | | 216197 | 185741 | 264955 | 196254 | 6.37E-02 | 0.26 | 1.22 |
| RP_POS_657_14154a8_642 | | 657.1415 | 8.64 | | RP | POS | | | 646152 | 54765.1 | 660235 | 55791.4 | 6.37E-02 | 0.25 | 1.12 |
| HILIC_POS_145_04976a2_877 | | 145.0498 | 2.88 | | HILIC | POS | | | 637600 | 277324 | 719026 | 351690 | 6.38E-02 | 0.26 | 1.19 |
| HILIC_NEG_210_94534a5_794 | | 210.9453 | 5.79 | | HILIC | NEG | | | 161542 | 41173.2 | 172119 | 41892.4 | 6.38E-02 | 0.25 | 1.08 |
| RP_NEG_531_17853a10_546 | | 531.1785 | 10.55 | | RP | NEG | | | 1.2E+07 | 8011580 | 1.4E+07 | 8680710 | 6.39E-02 | 0.26 | 1.47 |
| RP_NEG_350_80936a15_404 | | 350.8094 | 15.40 | | RP | NEG | | | 361892 | 148643 | 326011 | 134288 | 6.39E-02 | -0.25 | 1.26 |
| HILIC_NEG_222_90057a3_188 | | 222.9006 | 3.19 | | HILIC | NEG | | | 295425 | 34483.7 | 303053 | 25594 | 6.40E-02 | 0.25 | 1.09 |
| HILIC_NEG_248_96017a5_464 | | 248.9602 | 5.46 | | HILIC | NEG | | | 189639 | 14394.7 | 186174 | 12943.7 | 6.40E-02 | -0.25 | 1.06 |
| RP_POS_391_15726a10_089 | | 391.1573 | 10.09 | | RP | POS | | | 1136449 | 580337 | 1290896 | 630470 | 6.43E-02 | 0.26 | 1.4 |
| HILIC_POS_105_0428a4_338 | | 105.0428 | 4.34 | | HILIC | POS | | | 3.3E+07 | 1.2E+07 | 3.6E+07 | 1.1E+07 | 6.43E-02 | 0.25 | 1.34 |
| RP_NEG_415_21634a7_64 | | 415.2163 | 7.64 | | RP | NEG | | | 4754574 | 5303974 | 3606881 | 3726151 | 6.43E-02 | -0.25 | 1.06 |
| RP_POS_187_05779a0_587 | | 187.0578 | 0.59 | | RP | POS | | | 1.4E+07 | 3806903 | 1.5E+07 | 4301389 | 6.44E-02 | 0.26 | 1.08 |
| RP_POS_278_24518a11_145 | | 278.2452 | 11.15 | | RP | POS | | | 763827 | 124006 | 730521 | 137009 | 6.45E-02 | -0.26 | 1.13 |
| RP_POS_278_96979a15_367 | | 278.9698 | 15.37 | | RP | POS | | | 5181637 | 1311953 | 5505603 | 1247220 | 6.47E-02 | 0.25 | 1.15 |
| RP_POS_798_58722a11_409 | | 798.5872 | 11.41 | | RP | POS | | | 374606 | 186626 | 335850 | 116733 | 6.47E-02 | -0.26 | 1.28 |
| RP_POS_546_35522a10_5 | | 546.3552 | 10.50 | | RP | POS | | | 6.2E+07 | 1.5E+07 | 6.6E+07 | 1.7E+07 | 6.48E-02 | 0.26 | 1.17 |
| HILIC_NEG_212_00246a0_557 | | 212.0025 | 0.56 | | HILIC | NEG | | | 2041957 | 1117983 | 1771475 | 1023823 | 6.50E-02 | -0.25 | 1.15 |
| RP_NEG_612_33124a10_176 | | 612.3312 | 10.18 | | RP | NEG | | | 2304537 | 721409 | 2129667 | 662862 | 6.50E-02 | -0.25 | 1.17 |
| RP_POS_651_45746a10_465 | | 651.4575 | 10.47 | | RP | POS | | | 481333 | 214482 | 546986 | 291363 | 6.51E-02 | 0.26 | 1.51 |
| RP_NEG_135_9966a10_649 | | 135.9966 | 10.65 | | RP | NEG | | | 1552744 | 825550 | 1765203 | 851241 | 6.53E-02 | 0.25 | 1.41 |
| RP_POS_780_55121a13_298 | | 780.5512 | 13.30 | | RP | POS | | | 2.1E+08 | 4.2E+07 | 2.2E+08 | 4E+07 | 6.54E-02 | 0.25 | 1.15 |
| RP_POS_357_27875a9_903 | Deoxycholic acid [M+H-2H2O]+ | 357.2788 | 9.90 | | RP | POS | 357.2765(100), 215.1808(30) | 1 | 9030053 | 7486777 | 1.1E+07 | 8875300 | 6.56E-02 | 0.26 | 1.11 |
| RP_NEG_585_48706a11_236 | | 585.4871 | 11.24 | | RP | NEG | | | 265713 | 188606 | 313819 | 192076 | 6.58E-02 | 0.25 | 1.42 |
| RP_NEG_267_23337a11_026 | | 267.2334 | 11.03 | | RP | NEG | | | 114553 | 124468 | 148134 | 139781 | 6.59E-02 | 0.25 | 1.31 |
| RP_NEG_725_23193a6_031 | | 725.2319 | 6.03 | | RP | NEG | | | 472505 | 207881 | 419639 | 210619 | 6.59E-02 | -0.25 | 1.34 |
| RP_NEG_774_5304a12_589 | | 774.5304 | 12.59 | | RP | NEG | | | 1.1E+07 | 3535486 | 1E+07 | 3465473 | 6.59E-02 | -0.25 | 1.05 |
| RP_NEG_339_06821a10_649 | | 339.0682 | 10.65 | | RP | NEG | | | 629994 | 366566 | 741379 | 495212 | 6.60E-02 | 0.26 | 1.42 |
| RP_POS_344_87476a15_373 | | 344.8748 | 15.37 | | RP | POS | | | 2437020 | 1215911 | 2869966 | 2040726 | 6.61E-02 | 0.27 | 1.15 |
| RP_POS_347_22144a8_457 | | 347.2214 | 8.46 | | RP | POS | | | 592033 | 533111 | 477382 | 375627 | 6.62E-02 | -0.25 | 1.1 |
| HILIC_NEG_130_99278a0_649 | | 130.9928 | 0.65 | | HILIC | NEG | | | 1960967 | 512717 | 1817779 | 610646 | 6.64E-02 | -0.25 | 1.48 |
| RP_POS_279_23178a9_761 | | 279.2318 | 9.76 | | RP | POS | | | 2356951 | 1338949 | 2020311 | 1333022 | 6.64E-02 | -0.25 | 1.09 |
| RP_POS_792_04449a13_001 | | 792.0445 | 13.00 | | RP | POS | | | 2770941 | 3068793 | 3724645 | 4292857 | 6.65E-02 | 0.26 | 1.35 |
| RP_POS_502_29279a10_186 | LysoPE 20:4 (0:0/20:4) | 502.2928 | 10.19 | | RP | POS | 361.2749(100) | 1 | 6584430 | 1581606 | 7031150 | 1918748 | 6.65E-02 | 0.26 | 1.31 |
| RP_POS_311_16705a11_144 | | 311.1671 | 11.14 | | RP | POS | | | 1115438 | 216402 | 1059715 | 225599 | 6.68E-02 | -0.25 | 1.1 |
| RP_POS_458_34744a11_16 | | 458.3474 | 11.16 | | RP | POS | | | 451536 | 238011 | 522140 | 311898 | 6.69E-02 | 0.26 | 1.44 |
| RP_POS_278_17493a9_77 | | 278.1749 | 9.77 | | RP | POS | | | 782089 | 59214.9 | 798002 | 66475 | 6.69E-02 | 0.25 | 1.27 |
| RP_NEG_135_23697a10_649 | | 135.2370 | 10.65 | | RP | NEG | | | 352368 | 191961 | 401836 | 200597 | 6.70E-02 | 0.25 | 1.42 |
| RP_POS_223_0631a10_222 | | 223.0631 | 10.22 | | RP | POS | | | 162872 | 84262.8 | 144545 | 61652.8 | 6.70E-02 | -0.25 | 1.26 |
| RP_NEG_391_28589a9_901 | Deoxycholic acid | 391.2859 | 9.90 | | RP | NEG | 391.2846(100), 345.2823(5) | 1 | 4373437 | 3582764 | 5382328 | 4342471 | 6.71E-02 | 0.25 | 1.09 |
| HILIC_NEG_391_00159a1_228 | | 391.0016 | 1.23 | | HILIC | NEG | | | 727358 | 303799 | 654199 | 279991 | 6.71E-02 | -0.25 | 1.09 |
| RP_POS_832_58405a13_134 | | 832.5841 | 13.13 | | RP | POS | | | 4.3E+07 | 1.5E+07 | 3.9E+07 | 1.2E+07 | 6.71E-02 | -0.25 | 1.23 |
| RP_NEG_828_57794a13_447 | | 828.5779 | 13.45 | | RP | NEG | | | 8.3E+07 | 2.7E+07 | 8.9E+07 | 2.5E+07 | 6.73E-02 | 0.25 | 1.14 |
| RP_NEG_212_00256a2_449 | Indoxyl sulfate | 212.0026 | 2.45 | | RP | NEG | 212.0023(100), 79.9599(80) | 1 | 1.1E+07 | 6146107 | 9356437 | 5430844 | 6.73E-02 | -0.25 | 1.13 |
| RP_NEG_595_75397a5_532 | | 595.7540 | 5.53 | | RP | NEG | | | 312875 | 147417 | 275100 | 152953 | 6.74E-02 | -0.25 | 1.16 |
| RP_POS_574_39954a10_005 | | 574.3995 | 10.01 | | RP | POS | | | 136585 | 91234.4 | 162969 | 115450 | 6.75E-02 | 0.26 | 1.2 |
| RP_POS_582_57666a11_31 | | 582.5767 | 11.31 | | RP | POS | | | 104560 | 55338.8 | 123273 | 87507.9 | 6.77E-02 | 0.26 | 1.25 |
| RP_NEG_281_24908a11_238 | FA 18:1 (Oleic acid) | 281.2491 | 11.24 | HMDB0000207 | RP | NEG | 281.2492(100) | 1 | 1.7E+07 | 9913292 | 1.9E+07 | 1E+07 | 6.77E-02 | 0.25 | 1.38 |
| RP_POS_229_12228a10_651 | | 229.1223 | 10.65 | | RP | POS | | | 4673811 | 2537223 | 5341339 | 2762847 | 6.78E-02 | 0.25 | 1.37 |
| RP_NEG_696_77283a6_032 | | 696.7728 | 6.03 | | RP | NEG | | | 1.4E+07 | 5999388 | 1.2E+07 | 6178861 | 6.78E-02 | -0.25 | 1.34 |
| HILIC_NEG_343_99521a6_313 | | 343.9952 | 6.31 | | HILIC | NEG | | | 2E+07 | 4094422 | 1.8E+07 | 5572614 | 6.78E-02 | -0.26 | 1.06 |
| RP_POS_513_34204a10_873 | | 513.3420 | 10.87 | | RP | POS | | | 644298 | 359360 | 754725 | 498452 | 6.79E-02 | 0.26 | 1.47 |
| RP_NEG_207_96742a13_29 | | 207.9674 | 13.29 | | RP | NEG | | | 886537 | 307336 | 803960 | 347131 | 6.80E-02 | -0.25 | 1.33 |
| RP_NEG_762_50873a12_933 | | 762.5087 | 12.93 | | RP | NEG | | | 9987937 | 3787517 | 9075096 | 3521254 | 6.81E-02 | -0.25 | 1.05 |
| RP_POS_353_26599a10_745 | | 353.2660 | 10.75 | | RP | POS | | | 1671321 | 665598 | 1921144 | 1206558 | 6.82E-02 | 0.27 | 1.26 |
| RP_NEG_851_1145a9_277 | | 851.1145 | 9.28 | | RP | NEG | | | 4733568 | 386594 | 4642163 | 345895 | 6.82E-02 | -0.25 | 1.39 |
| HILIC_NEG_277_15646a1_62 | | 277.1565 | 1.62 | | HILIC | NEG | | | 174571 | 87700.2 | 199920 | 111413 | 6.83E-02 | 0.25 | 1.3 |
| RP_NEG_411_22214a12_107 | | 411.2221 | 12.11 | | RP | NEG | | | 513288 | 99518.3 | 488214 | 100737 | 6.83E-02 | -0.25 | 1.4 |
| RP_NEG_232_05948a5_407 | | 232.0595 | 5.41 | | RP | NEG | | | 616502 | 385036 | 723004 | 457547 | 6.86E-02 | 0.25 | 1.16 |
| RP_NEG_366_97443a7_266 | | 366.9744 | 7.27 | | RP | NEG | | | 252118 | 356819 | 344522 | 379985 | 6.86E-02 | 0.25 | 1.14 |
| RP_POS_530_32391a10_656 | | 530.3239 | 10.66 | | RP | POS | | | 1154402 | 512815 | 1305279 | 670274 | 6.87E-02 | 0.26 | 1.14 |
| RP_POS_410_32632a9_903 | | 410.3263 | 9.90 | | RP | POS | 357.277(100) | | 2908423 | 2447588 | 3578010 | 2859097 | 6.87E-02 | 0.25 | 1.09 |
| RP_POS_191_97206a0_566 | | 191.9721 | 0.57 | | RP | POS | | | 1165196 | 239884 | 1227284 | 255393 | 6.87E-02 | 0.25 | 1.26 |
| HILIC_NEG_183_93439a3_064 | | 183.9344 | 3.06 | | HILIC | NEG | | | 233586 | 54478.6 | 220263 | 52363 | 6.88E-02 | -0.25 | 1.22 |
| RP_NEG_830_49634a12_933 | | 830.4963 | 12.93 | | RP | NEG | | | 1315459 | 482302 | 1198198 | 459723 | 6.92E-02 | -0.25 | 1.05 |
| RP_POS_551_39471a9_786 | | 551.3947 | 9.79 | | RP | POS | | | 6364151 | 2496435 | 7007664 | 2650700 | 6.94E-02 | 0.25 | 1.28 |
| HILIC_POS_394_20764a0_898 | | 394.2076 | 0.90 | | HILIC | POS | | | 182022 | 69651 | 166404 | 56168.1 | 6.96E-02 | -0.25 | 1.1 |
| RP_NEG_326_98889a15_471 | | 326.9889 | 15.47 | | RP | NEG | | | 1559261 | 137165 | 1528005 | 114756 | 6.96E-02 | -0.25 | 1.08 |
| HILIC_NEG_192_97945a5_65 | | 192.9795 | 5.65 | | HILIC | NEG | | | 123963 | 26302.1 | 118362 | 18670 | 6.97E-02 | -0.25 | 1.09 |
| RP_POS_579_43402a11_197 | | 579.4340 | 11.20 | | RP | POS | | | 1133867 | 336828 | 1047139 | 357927 | 6.98E-02 | -0.25 | 1.27 |
| HILIC_NEG_141_92397a4_399 | | 141.9240 | 4.40 | | HILIC | NEG | | | 920610 | 420781 | 1026180 | 427822 | 7.01E-02 | 0.25 | 1.32 |
| RP_POS_594_35291a10_677 | | 594.3529 | 10.68 | | RP | POS | | | 754119 | 265697 | 830577 | 338507 | 7.01E-02 | 0.25 | 1.12 |
| RP_POS_776_57953a12_74 | | 776.5795 | 12.74 | | RP | POS | | | 2099823 | 500554 | 2210383 | 391227 | 7.01E-02 | 0.25 | 1.25 |
| HILIC_POS_368_21997a2_278 | | 368.2200 | 2.28 | | HILIC | POS | | | 95376.8 | 79725 | 119906 | 112090 | 7.02E-02 | 0.26 | 1.14 |
| HILIC_NEG_117_0196a1_134 | Succinic acid | 117.0196 | 1.13 | | HILIC | NEG | 73.0288(100), 116.9292(50) | 2 | 936508 | 183546 | 892160 | 174063 | 7.02E-02 | -0.25 | 1.11 |
| RP_POS_146_08124a0_68 | | 146.0812 | 0.68 | | RP | POS | | | 1298997 | 1194205 | 1042750 | 869096 | 7.02E-02 | -0.25 | 1.02 |
| RP_POS_571_14832a10_547 | | 571.1483 | 10.55 | | RP | POS | | | 1611383 | 917754 | 1835632 | 889905 | 7.03E-02 | 0.25 | 1.38 |
| HILIC_POS_84_08081a3_422 | | 84.0808 | 3.42 | | HILIC | POS | | | 1666974 | 6508816 | 509229 | 2012154 | 7.03E-02 | -0.27 | 0.95 |
| RP_POS_313_23636a8_996 | | 313.2364 | 9.00 | | RP | POS | | | 663268 | 212345 | 613919 | 186400 | 7.04E-02 | -0.25 | 1.03 |
| RP_POS_536_27185a3_969 | | 536.2719 | 3.97 | | RP | POS | | | 373831 | 205616 | 326688 | 175462 | 7.04E-02 | -0.25 | 1.04 |
| RP_NEG_437_29135a9_902 | | 437.2914 | 9.90 | | RP | NEG | | | 956918 | 786512 | 1173897 | 941060 | 7.05E-02 | 0.25 | 1.06 |
| RP_POS_378_21182a8_257 | | 378.2118 | 8.26 | | RP | POS | | | 190128 | 61510 | 214265 | 119072 | 7.06E-02 | 0.27 | 1.1 |
| RP_POS_257_19003a10_796 | | 257.1900 | 10.80 | | RP | POS | | | 1247969 | 221049 | 1200586 | 161185 | 7.06E-02 | -0.25 | 1.11 |
| RP_NEG_371_22058a10_926 | | 371.2206 | 10.93 | | RP | NEG | | | 1709394 | 719167 | 1885295 | 700562 | 7.07E-02 | 0.25 | 1.37 |
| RP_POS_574_39771a8_932 | | 574.3977 | 8.93 | | RP | POS | | | 391743 | 250658 | 338848 | 175494 | 7.08E-02 | -0.25 | 1.19 |
| RP_NEG_305_24902a11_114 | | 305.2490 | 11.11 | | RP | NEG | | | 1348042 | 658480 | 1514861 | 684627 | 7.09E-02 | 0.25 | 1.53 |
| RP_POS_161_09604a10_653 | | 161.0960 | 10.65 | | RP | POS | | | 792024 | 365690 | 887989 | 404133 | 7.09E-02 | 0.25 | 1.4 |
| RP_NEG_280_98328a7_291 | | 280.9833 | 7.29 | | RP | NEG | | | 2902727 | 153177 | 2937996 | 132613 | 7.11E-02 | 0.25 | 1 |
| RP_NEG_527_22125a6_799 | | 527.2213 | 6.80 | | RP | NEG | | | 590887 | 657729 | 457233 | 413546 | 7.11E-02 | -0.25 | 1.11 |
| RP_POS_812_55627a13_298 | | 812.5563 | 13.30 | | RP | POS | | | 3849848 | 1276317 | 3547819 | 1169021 | 7.11E-02 | -0.25 | 1.33 |
| RP_POS_403_24973a11_595 | | 403.2497 | 11.60 | | RP | POS | | | 3080797 | 600178 | 2950551 | 453792 | 7.12E-02 | -0.25 | 1.3 |
| HILIC_NEG_238_07799a0_937 | | 238.0780 | 0.94 | | HILIC | NEG | | | 2906589 | 2069138 | 3443099 | 2248205 | 7.14E-02 | 0.25 | 1.05 |
| RP_POS_587_54938a15_29 | | 587.5494 | 15.29 | | RP | POS | | | 3.1E+07 | 1.4E+07 | 3.5E+07 | 1.9E+07 | 7.14E-02 | 0.25 | 1.32 |
| HILIC_NEG_234_81949a7_751 | | 234.8195 | 7.75 | | HILIC | NEG | | | 2991048 | 444180 | 3108428 | 498482 | 7.15E-02 | 0.25 | 1.05 |
| RP_POS_432_31085a9_374 | | 432.3109 | 9.37 | | RP | POS | | | 5318441 | 6834484 | 3838929 | 5152073 | 7.15E-02 | -0.25 | 1.06 |
| HILIC_POS_210_10979a5_66 | | 210.1098 | 5.66 | | HILIC | POS | | | 79529.5 | 127536 | 121868 | 200446 | 7.16E-02 | 0.26 | 1.15 |
| RP_NEG_563_50507a11_237 | | 563.5051 | 11.24 | | RP | NEG | | | 380133 | 307764 | 459746 | 333372 | 7.16E-02 | 0.25 | 1.37 |
| HILIC_NEG_179_88254a7_734 | | 179.8825 | 7.73 | | HILIC | NEG | | | 1594096 | 122486 | 1565601 | 108760 | 7.16E-02 | -0.25 | 1.04 |
| RP_NEG_277_15536a4_543 | | 277.1554 | 4.54 | | RP | NEG | | | 1881221 | 921412 | 2143398 | 1164976 | 7.18E-02 | 0.25 | 1.37 |
| RP_NEG_485_2121a11_237 | | 485.2121 | 11.24 | | RP | NEG | | | 537147 | 258985 | 601664 | 262893 | 7.18E-02 | 0.25 | 1.36 |
| RP_POS_468_38986a10_531 | | 468.3899 | 10.53 | | RP | POS | | | 4545778 | 879563 | 4353900 | 677251 | 7.18E-02 | -0.25 | 1.2 |
| RP_POS_636_40503a10_115 | | 636.4050 | 10.12 | | RP | POS | | | 307670 | 161820 | 269821 | 145448 | 7.18E-02 | -0.25 | 1.15 |
| RP_POS_462_29953a12_108 | | 462.2995 | 12.11 | | RP | POS | | | 517517 | 221269 | 466681 | 191979 | 7.20E-02 | -0.25 | 1.4 |
| RP_POS_279_17023a4_546 | | 279.1702 | 4.55 | | RP | POS | 120.0808(100),279.1715(10) | | 1.3E+07 | 6286797 | 1.5E+07 | 8208742 | 7.21E-02 | 0.25 | 1.29 |
| HILIC_POS_372_31055a0_616 | | 372.3106 | 0.62 | | HILIC | POS | | | 604051 | 212880 | 673183 | 325870 | 7.22E-02 | 0.26 | 1.43 |
| HILIC_NEG_308_99857a5_116 | | 308.9986 | 5.12 | | HILIC | NEG | | | 634815 | 330930 | 727879 | 411908 | 7.22E-02 | 0.25 | 1.18 |
| RP_NEG_187_00699a3_681 | p-Cresol sulfate | 187.0070 | 3.68 | | RP | NEG | 107.0498(100), 187.0077(65), 79.957(35) | 2 | 1.3E+08 | 1E+08 | 1.1E+08 | 8E+07 | 7.23E-02 | -0.25 | 1.1 |
| RP_POS_569_43427a14_227 | | 569.4343 | 14.23 | | RP | POS | | | 404507 | 294312 | 343739 | 196848 | 7.23E-02 | -0.25 | 1.24 |
| RP_POS_240_15688a6_27 | | 240.1569 | 6.27 | | RP | POS | | | 5221856 | 605955 | 5066415 | 650070 | 7.24E-02 | -0.25 | 1.1 |
| HILIC_POS_217_12956a5_674 | N-Acetylarginine | 217.1296 | 5.67 | | HILIC | POS | 217.1291(100) | 2 | 196620 | 92469.1 | 220024 | 96976 | 7.25E-02 | 0.25 | 1.13 |
| RP_NEG_878_59296a13_783 | | 878.5930 | 13.78 | | RP | NEG | | | 1.7E+07 | 4157895 | 1.5E+07 | 4027190 | 7.25E-02 | -0.25 | 1.24 |
| RP_NEG_307_26465a11_366 | | 307.2647 | 11.37 | | RP | NEG | | | 423803 | 296833 | 496154 | 290949 | 7.26E-02 | 0.25 | 1.44 |
| RP_NEG_236_97987a15_481 | | 236.9799 | 15.48 | | RP | NEG | | | 1079244 | 68430 | 1062001 | 71262.1 | 7.26E-02 | -0.25 | 1.17 |
| HILIC_POS_140_06818a5_317 | | 140.0682 | 5.32 | | HILIC | POS | 140.069(100), 74.0974(40) | | 3460717 | 2772110 | 4685614 | 6233329 | 7.27E-02 | 0.27 | 1.12 |
| RP_NEG_568_76361a15_406 | | 568.7636 | 15.41 | | RP | NEG | | | 242153 | 80696.1 | 224223 | 65400.4 | 7.27E-02 | -0.25 | 1.12 |
| RP_POS_281_18723a10_064 | | 281.1872 | 10.06 | | RP | POS | | | 1092400 | 275599 | 1029486 | 237185 | 7.28E-02 | -0.25 | 1.38 |
| RP_POS_518_32886a9_874 | | 518.3289 | 9.87 | | RP | POS | | | 3316346 | 1583395 | 2903449 | 1752068 | 7.29E-02 | -0.25 | 1.1 |
| RP_POS_363_21637a6_482 | | 363.2164 | 6.48 | | RP | POS | 363.2173(100) | | 762265 | 311776 | 830265 | 242366 | 7.30E-02 | 0.25 | 1.05 |
| HILIC_NEG_246_92216a5_795 | | 246.9222 | 5.80 | | HILIC | NEG | | | 134659 | 35193 | 143162 | 34011.4 | 7.30E-02 | 0.25 | 1.05 |
| RP_POS_352_65906a5_567 | | 352.6591 | 5.57 | | RP | POS | | | 331499 | 235898 | 394521 | 271895 | 7.30E-02 | 0.25 | 1.24 |
| HILIC_POS_325_03836a4_256 | | 325.0384 | 4.26 | | HILIC | POS | | | 118614 | 34030.6 | 108493 | 46248.6 | 7.30E-02 | -0.25 | 1.07 |
| RP_POS_236_07156a15_187 | | 236.0716 | 15.19 | | RP | POS | | | 1.2E+08 | 7.9E+07 | 1.4E+08 | 1.2E+08 | 7.31E-02 | 0.25 | 1.26 |
| RP_POS_379_21814a11_591 | | 379.2181 | 11.59 | | RP | POS | | | 408034 | 153241 | 373745 | 126719 | 7.33E-02 | -0.24 | 1.28 |
| RP_POS_165_12772a7_924 | | 165.1277 | 7.92 | | RP | POS | | | 1668607 | 160515 | 1707991 | 160236 | 7.36E-02 | 0.25 | 1.02 |
| RP_NEG_411_18491a7_505 | | 411.1849 | 7.51 | | RP | NEG | | | 256985 | 288393 | 541929 | 1547200 | 7.38E-02 | 0.31 | 1.12 |
| HILIC_POS_214_05869a6_382 | | 214.0587 | 6.38 | | HILIC | POS | | | 330166 | 130626 | 364778 | 149222 | 7.38E-02 | 0.25 | 1.06 |
| RP_POS_399_28253a7_97 | | 399.2825 | 7.97 | | RP | POS | | | 183926 | 191459 | 146298 | 111755 | 7.42E-02 | -0.25 | 0.99 |

**Supplemantary table 1**: The non-targeted metabolomics results ordered by the VIP value (p value < 0.0003 are shaded dark green, p value < 0.05 are shaded light green, d values below 0 are blue and above 0 are red)

| | | | | | | | | | Cases | | Controls | | Welch test | Cohen's | |
|---|---|---|---|---|---|---|---|---|---|---|---|---|---|---|---|
| Feature_ID | Metabolite_name | Average_Mz | Average_RT_min_ | HMDB ID | RP_HILIC | POS_NEG | Cureted_MSMS | ID_level | Mean | SD | Mean2 | SD3 | p | d | VIP |
| RP_POS_322_27396a10_927 | | 322.2740 | 10.93 | | RP | POS | | | 3622357 | 1627618 | 4021163 | 1629100 | 7.44E-02 | 0.24 | 1.26 |
| HILIC_NEG_84_0086a0_933 | | 84.0086 | 0.93 | | HILIC | NEG | | | 547893 | 97779.9 | 523014 | 104685 | 7.44E-02 | -0.25 | 1.1 |
| RP_POS_748_11993a11_459 | | 748.1199 | 11.46 | | RP | POS | | | 181042 | 85313.7 | 206423 | 116839 | 7.45E-02 | 0.25 | 1.1 |
| HILIC_NEG_112_9857a0_573 | | 112.9857 | 0.57 | | HILIC | NEG | | | 1.1E+07 | 2381518 | 1.2E+07 | 2290151 | 7.47E-02 | 0.24 | 1.04 |
| RP_NEG_439_38004a11_917 | | 439.3800 | 11.92 | | RP | NEG | | | 293503 | 61291.5 | 278485 | 61477.3 | 7.47E-02 | -0.24 | 1.45 |
| RP_NEG_523_72705a5_483 | | 523.7271 | 5.48 | | RP | NEG | | | 450184 | 178266 | 406064 | 182099 | 7.47E-02 | -0.24 | 1.14 |
| RP_POS_389_11191a9_614 | | 389.1119 | 9.61 | | RP | POS | | | 501958 | 86562.6 | 523352 | 88272 | 7.48E-02 | 0.24 | 1.11 |
| RP_POS_299_16141a9_072 | 6-Shogaol | 299.1614 | 9.07 | | RP | POS | 299.1622(100) | 2 | 3301282 | 461060 | 3191845 | 436010 | 7.49E-02 | -0.24 | 1.07 |
| RP_NEG_352_08624a6_192 | | 352.0862 | 6.19 | | RP | NEG | | | 197998 | 182416 | 157252 | 152131 | 7.49E-02 | -0.24 | 0.98 |
| HILIC_POS_230_18636a2_894 | | 230.1864 | 2.89 | | HILIC | POS | | | 706772 | 1664301 | 396270 | 792920 | 7.49E-02 | -0.25 | 1.04 |
| RP_POS_190_08633a6_03 | Indole-3-methyl acetate | 190.0863 | 6.03 | | RP | POS | 130.0657(100), 172.0755(20) | 1 | 1.1E+07 | 8068595 | 7637817 | 1.6E+07 | 7.53E-02 | -0.26 | 1.25 |
| RP_POS_452_35669a10_653 | | 452.3567 | 10.65 | | RP | POS | | | 822203 | 137166 | 789683 | 129768 | 7.53E-02 | -0.24 | 1.13 |
| RP_NEG_511_29141a8_761 | | 511.2914 | 8.76 | | RP | NEG | | | 3108684 | 949274 | 3453472 | 1706040 | 7.55E-02 | 0.26 | 1.11 |
| RP_POS_480_1579a10_127 | | 480.1579 | 10.13 | | RP | POS | | | 213356 | 61789.5 | 229171 | 67291.5 | 7.55E-02 | 0.25 | 1.07 |
| RP_POS_588_43127a8_815 | | 588.4313 | 8.82 | | RP | POS | | | 635080 | 428942 | 549308 | 269222 | 7.55E-02 | -0.25 | 1 |
| RP_NEG_197_80806a7_247 | | 197.8081 | 7.25 | | RP | NEG | | | 330269 | 319739 | 426002 | 444356 | 7.56E-02 | 0.25 | 1.05 |
| RP_POS_351_28915a11_337 | | 351.2892 | 11.34 | | RP | POS | | | 805435 | 340340 | 738141 | 206172 | 7.56E-02 | -0.25 | 1.06 |
| RP_POS_668_61835a11_017 | | 668.6184 | 11.02 | | RP | POS | | | 325769 | 1089729 | 143973 | 116597 | 7.56E-02 | -0.3 | 1.01 |
| RP_POS_303_25278a10_36 | | 303.2528 | 10.36 | | RP | POS | | | 538290 | 534328 | 745088 | 1040105 | 7.58E-02 | 0.26 | 1.09 |
| HILIC_NEG_215_03278a5_121 | 5-Methoxypsoralen | 215.0328 | 5.12 | | HILIC | NEG | 89.0235(100), 59.0131(90) | 1 | 3.3E+07 | 1.2E+07 | 3.6E+07 | 1.7E+07 | 7.58E-02 | 0.25 | 1.14 |
| RP_NEG_213_99791a2_606 | | 213.9979 | 2.61 | | RP | NEG | | | 727051 | 413742 | 630745 | 380188 | 7.63E-02 | -0.24 | 1.11 |
| RP_POS_330_24588a8_872 | | 330.2459 | 8.87 | | RP | POS | | | 1134460 | 524833 | 1286037 | 695103 | 7.64E-02 | 0.25 | 1.05 |
| HILIC_POS_305_96069a5_387 | | 305.9607 | 5.39 | | HILIC | POS | | | 97270.9 | 32693.2 | 89852.8 | 28524 | 7.64E-02 | -0.24 | 0.99 |
| RP_NEG_588_33099a10_182 | | 588.3310 | 10.18 | | RP | NEG | | | 6598023 | 1725619 | 7024353 | 1776494 | 7.65E-02 | 0.24 | 1.1 |
| RP_NEG_423_1467a11_141 | | 423.1467 | 11.14 | | RP | NEG | | | 462975 | 58701 | 448938 | 56906.7 | 7.65E-02 | -0.24 | 1.28 |
| RP_POS_486_19064a5_262 | | 486.1906 | 5.26 | | RP | POS | | | 733767 | 661735 | 601546 | 418545 | 7.65E-02 | -0.24 | 1.16 |
| RP_POS_715_8974a11_359 | | 715.8974 | 11.36 | | RP | POS | | | 155374 | 81391 | 177901 | 101254 | 7.67E-02 | 0.25 | 1.02 |
| RP_POS_277_17978a9_073 | | 277.1798 | 9.07 | | RP | POS | | | 8920246 | 1258887 | 8623895 | 1186054 | 7.67E-02 | -0.24 | 1.02 |
| RP_NEG_271_22791a10_471 | | 271.2279 | 10.47 | | RP | NEG | 271.2275(45), 225.224(2) | | 4627478 | 613735 | 4485437 | 559845 | 7.69E-02 | -0.24 | 1.3 |
| RP_POS_181_0493a11_408 | | 181.0493 | 11.41 | | RP | POS | | | 3027355 | 411989 | 2938865 | 318921 | 7.69E-02 | -0.24 | 1.07 |
| RP_NEG_667_47412a14_035 | | 667.4741 | 14.04 | | RP | NEG | | | 2409013 | 259174 | 2347787 | 246323 | 7.70E-02 | -0.24 | 1.18 |
| RP_NEG_462_29858a10_771 | | 462.2986 | 10.77 | | RP | NEG | | | 1027277 | 450057 | 1144536 | 509444 | 7.71E-02 | 0.24 | 1.21 |
| RP_POS_426_18521a10_648 | | 426.1852 | 10.65 | | RP | POS | | | 395094 | 242718 | 456637 | 262476 | 7.71E-02 | 0.24 | 1.25 |
| RP_NEG_445_21173a11_84 | | 445.2117 | 11.84 | | RP | NEG | | | 135228 | 24583 | 129288 | 24399.4 | 7.71E-02 | -0.24 | 1.41 |
| RP_POS_329_19571a8_993 | | 329.1957 | 8.99 | | RP | POS | | | 463332 | 272870 | 550139 | 415724 | 7.72E-02 | 0.25 | 1.42 |
| RP_POS_755_52997a10_351 | | 755.5300 | 10.35 | | RP | POS | | | 1579077 | 740551 | 1809015 | 1091642 | 7.72E-02 | 0.25 | 1.17 |
| RP_POS_355_2605a11_263 | | 355.2605 | 11.26 | | RP | POS | | | 1567450 | 1187713 | 1847228 | 1124814 | 7.73E-02 | 0.24 | 1.27 |
| RP_NEG_534_25134a8_97 | | 534.2513 | 8.97 | | RP | NEG | | | 615305 | 569171 | 491845 | 452391 | 7.73E-02 | -0.24 | 1.15 |
| HILIC_POS_269_12601a1_378 | | 269.1260 | 1.38 | | HILIC | POS | | | 162421 | 232735 | 119167 | 113287 | 7.73E-02 | -0.25 | 1.05 |
| RP_NEG_293_78357a15_484 | | 293.7836 | 15.48 | | RP | NEG | | | 295925 | 46975.5 | 285531 | 39132.3 | 7.75E-02 | -0.24 | 1.08 |
| RP_POS_512_32184a8_719 | | 512.3218 | 8.72 | | RP | POS | | | 1537420 | 653972 | 1382514 | 626777 | 7.75E-02 | -0.24 | 1.06 |
| RP_POS_764_55597a13_577 | | 764.5560 | 13.58 | | RP | POS | | | 9787982 | 3158353 | 9120315 | 2362411 | 7.75E-02 | -0.24 | 1.07 |
| RP_POS_544_34021a10_182 | LysoPC 20:4 (0:0/20:4) | 544.3402 | 10.18 | | RP | POS | 184.0743(100) | 1 | 2.6E+07 | 7489498 | 2.8E+07 | 7491061 | 7.76E-02 | 0.24 | 1.06 |
| RP_NEG_367_32254a11_207 | | 367.3225 | 11.21 | | RP | NEG | | | 238555 | 40168.1 | 228935 | 39338.2 | 7.76E-02 | -0.24 | 1.46 |
| HILIC_NEG_92_9278a3_192 | | 92.9278 | 3.19 | | HILIC | NEG | | | 2416064 | 162474 | 2360820 | 270920 | 7.76E-02 | -0.25 | 1.22 |
| HILIC_POS_466_31598a2_181 | | 466.3160 | 2.18 | | HILIC | POS | | | 199804 | 557995 | 104103 | 154621 | 7.79E-02 | -0.27 | 1 |
| RP_NEG_848_54565a12_539 | | 848.5457 | 12.54 | | RP | NEG | | | 3319710 | 1063318 | 3061916 | 1068281 | 7.80E-02 | -0.24 | 1.33 |
| RP_POS_444_25372a11_591 | | 444.2537 | 11.59 | | RP | POS | | | 658302 | 131771 | 627360 | 124601 | 7.80E-02 | -0.24 | 1.23 |
| RP_POS_537_37842a9_384 | | 537.3784 | 9.38 | | RP | POS | | | 738088 | 364024 | 854063 | 558691 | 7.83E-02 | 0.25 | 1.16 |
| RP_NEG_622_29028a10_316 | | 622.2903 | 10.32 | | RP | NEG | | | 528077 | 64254.5 | 511839 | 69589.5 | 7.83E-02 | -0.24 | 1.02 |
| RP_POS_464_31345a10_772 | | 464.3135 | 10.77 | | RP | POS | | | 2684531 | 1190019 | 2997497 | 1378549 | 7.85E-02 | 0.24 | 1.18 |
| RP_POS_304_21228a4_676 | | 304.2123 | 4.68 | | RP | POS | | | 2226585 | 1301984 | 2567908 | 1501819 | 7.86E-02 | 0.24 | 1.15 |
| RP_POS_132_9585a0_636 | | 132.9585 | 0.64 | | RP | POS | | | 7116593 | 2059119 | 6600249 | 2203849 | 7.86E-02 | -0.24 | 1.07 |
| HILIC_NEG_157_90086a0_953 | | 157.9009 | 0.95 | | HILIC | NEG | | | 1560489 | 553742 | 1693309 | 548740 | 7.90E-02 | 0.24 | 1.09 |
| RP_NEG_330_98026a7_819 | | 330.9803 | 7.82 | | RP | NEG | | | 783544 | 314616 | 861281 | 329189 | 7.91E-02 | 0.24 | 0.99 |
| RP_NEG_187_07368a3_679 | | 187.0737 | 3.68 | | RP | NEG | | | 756150 | 591086 | 628815 | 469788 | 7.93E-02 | -0.24 | 1.14 |
| RP_POS_627_43597a10_243 | | 627.4360 | 10.24 | | RP | POS | | | 365515 | 215572 | 422385 | 252226 | 7.94E-02 | 0.24 | 1.09 |
| HILIC_POS_104_10759a1_433 | Choline | 104.1076 | 1.43 | | HILIC | POS | 104.1077(100), 60.0815(14) | 2 | 7.4E+08 | 1.4E+08 | 7E+08 | 1.4E+08 | 7.94E-02 | -0.24 | 1.37 |
| RP_NEG_211_9182a15_527 | | 211.9182 | 15.53 | | RP | NEG | | | 3547713 | 1230788 | 3882073 | 1509385 | 7.95E-02 | 0.24 | 1.02 |
| RP_POS_227_12473a7_272 | | 227.1247 | 7.27 | | RP | POS | | | 2414522 | 417068 | 2318018 | 386938 | 7.95E-02 | -0.24 | 1.4 |
| RP_POS_560_39996a7_687 | | 560.4000 | 7.69 | | RP | POS | | | 604782 | 395601 | 524038 | 274644 | 7.95E-02 | -0.24 | 0.99 |
| HILIC_NEG_276_99124a0_936 | | 276.9912 | 0.94 | | HILIC | NEG | | | 1491681 | 152045 | 1455938 | 145604 | 7.96E-02 | -0.24 | 1.05 |
| RP_POS_379_31766a11_162 | | 379.3177 | 11.16 | | RP | POS | | | 610266 | 155843 | 573314 | 151847 | 7.98E-02 | -0.24 | 1.17 |
| RP_POS_372_31055a8_915 | Myristoylcarnitine | 372.3106 | 8.92 | | RP | POS | 85.0292(100), 372.3084(45) | 1 | 2833231 | 1058951 | 3159469 | 1563723 | 7.99E-02 | 0.25 | 1.43 |
| HILIC_POS_422_30295a3_09 | | 422.3030 | 3.09 | | HILIC | POS | | | 40743.1 | 16926.6 | 45398.7 | 21202.5 | 7.99E-02 | 0.24 | 1.04 |
| RP_POS_334_22223a8_797 | | 334.2222 | 8.80 | | RP | POS | | | 385795 | 251597 | 449908 | 278924 | 7.99E-02 | 0.24 | 1.41 |
| RP_POS_270_96661a0_549 | | 270.9666 | 0.55 | | RP | POS | | | 1659751 | 221239 | 1711904 | 213484 | 8.00E-02 | 0.24 | 1.1 |
| RP_NEG_339_29013a10_826 | | 339.2901 | 10.83 | | RP | NEG | | | 297960 | 47691.6 | 286390 | 48532.2 | 8.00E-02 | -0.24 | 1.47 |
| RP_POS_887_15533a9_406 | | 887.1553 | 9.41 | | RP | POS | | | 761606 | 91920.4 | 740230 | 86428.9 | 8.00E-02 | -0.24 | 1.07 |
| RP_POS_312_12051a0_691 | | 312.1205 | 0.69 | | RP | POS | | | 307153 | 183387 | 265723 | 162884 | 8.02E-02 | -0.24 | 1.03 |
| RP_POS_660_48944a9_417 | | 660.4894 | 9.42 | | RP | POS | | | 3722226 | 1915714 | 3295167 | 1657592 | 8.05E-02 | -0.24 | 1.33 |
| RP_NEG_188_89391a15_401 | | 188.8939 | 15.40 | | RP | NEG | | | 2704469 | 972245 | 2950119 | 1066336 | 8.06E-02 | 0.24 | 1.01 |
| RP_NEG_410_17764a11_239 | | 410.1776 | 11.24 | | RP | NEG | | | 845191 | 230526 | 898516 | 215268 | 8.06E-02 | 0.24 | 1.3 |
| HILIC_NEG_255_73523a1_41 | | 255.7352 | 1.41 | | HILIC | NEG | | | 131539 | 64914.8 | 146857 | 63043.3 | 8.07E-02 | 0.24 | 1.08 |
| RP_POS_568_33972a10_176 | LysoPC(0:0/22:6) | 568.3397 | 10.18 | | RP | POS | 184.0744(100).568.3422(2) | 1 | 8992888 | 3123739 | 8286972 | 2786681 | 8.07E-02 | -0.24 | 1.08 |
| RP_POS_217_15858a10_267 | | 217.1586 | 10.27 | | RP | POS | | | 945932 | 112155 | 919548 | 108304 | 8.07E-02 | -0.24 | 1.29 |
| RP_POS_151_00542a15_413 | | 151.0054 | 15.41 | | RP | POS | | | 6798930 | 3400141 | 7688167 | 3952612 | 8.08E-02 | 0.24 | 1.04 |
| RP_POS_560_47064a14_356 | | 560.4706 | 14.36 | | RP | POS | | | 1.6E+07 | 5113862 | 1.7E+07 | 4610796 | 8.08E-02 | 0.24 | 0.98 |
| RP_POS_573_73169a10_001 | | 573.7317 | 10.00 | | RP | POS | | | 434174 | 228380 | 504873 | 340992 | 8.09E-02 | 0.25 | 1.16 |
| RP_NEG_159_10289a5_537 | | 159.1029 | 5.54 | | RP | NEG | | | 222284 | 73097.1 | 240767 | 80450.5 | 8.10E-02 | 0.24 | 1.23 |
| RP_NEG_280_98303a15_463 | | 280.9830 | 15.46 | | RP | NEG | | | 1012059 | 108304 | 1039659 | 120800 | 8.10E-02 | 0.24 | 1.04 |
| RP_POS_267_05856a0_676 | | 267.0586 | 0.68 | | RP | POS | | | 716896 | 136787 | 687314 | 111117 | 8.10E-02 | -0.24 | 1.08 |
| RP_POS_261_14581a8_911 | | 261.1458 | 8.91 | | RP | POS | | | 1924145 | 180627 | 1884659 | 150766 | 8.14E-02 | -0.24 | 1.33 |
| HILIC_POS_344_27972a0_64 | AC 12:0 (Dodecanoylcarnitine) | 344.2797 | 0.64 | | HILIC | POS | 85.0292(100), 344.28(40) | 1 | 2236853 | 1392824 | 2709290 | 2354539 | 8.15E-02 | 0.25 | 1.2 |
| HILIC_POS_448_30573a0_826 | | 448.3057 | 0.83 | | HILIC | POS | | | 143007 | 170406 | 101322 | 177685 | 8.15E-02 | -0.24 | 1.1 |
| RP_POS_619_52539a13_288 | | 619.5254 | 13.29 | | RP | POS | | | 212978 | 229009 | 274663 | 280224 | 8.16E-02 | 0.24 | 1.07 |
| RP_POS_416_1412a15_378 | | 416.1412 | 15.38 | | RP | POS | | | 306869 | 183032 | 266515 | 155982 | 8.16E-02 | -0.24 | 1.14 |
| RP_POS_121_06505a10_651 | | 121.0651 | 10.65 | | RP | POS | | | 2345628 | 1290686 | 2666194 | 1383877 | 8.18E-02 | 0.24 | 1.34 |
| RP_NEG_267_07346a0_682 | | 267.0735 | 0.68 | | RP | NEG | 135.0315(100), 267.0721(40) | | 3362977 | 3685104 | 2651243 | 2216251 | 8.19E-02 | -0.24 | 1.09 |
| HILIC_POS_542_73236a2_978 | | 542.7324 | 2.98 | | HILIC | POS | | | 300242 | 46619.3 | 311617 | 48509.1 | 8.20E-02 | 0.24 | 1.01 |
| RP_NEG_600_20331a10_289 | | 600.2033 | 10.29 | | RP | NEG | | | 315391 | 66351.1 | 332414 | 75231.5 | 8.20E-02 | 0.24 | 1.56 |
| RP_NEG_466_16669a10_958 | | 466.1667 | 10.96 | | RP | NEG | | | 515871 | 154164 | 552435 | 152530 | 8.21E-02 | 0.24 | 1.28 |
| RP_POS_272_25836a10_617 | | 272.2584 | 10.62 | | RP | POS | | | 176291 | 113631 | 153521 | 76794.6 | 8.22E-02 | -0.24 | 1.04 |
| HILIC_POS_279_1701a1_609 | | 279.1701 | 1.61 | | HILIC | POS | 120.0806(100) | | 2432862 | 6774926 | 1250015 | 2534778 | 8.22E-02 | -0.25 | 1.08 |
| RP_POS_535_28772a8_761 | | 535.2877 | 8.76 | | RP | POS | | | 793863 | 237262 | 876999 | 419703 | 8.24E-02 | 0.25 | 1.1 |
| RP_POS_572_37091a10_671 | | 572.3709 | 10.67 | | RP | POS | | | 4040064 | 1229808 | 4367005 | 1479903 | 8.24E-02 | 0.24 | 1.11 |
| HILIC_POS_128_06842a5_24 | | 128.0684 | 5.24 | | HILIC | POS | | | 278433 | 153096 | 243027 | 144590 | 8.24E-02 | -0.24 | 1.35 |
| RP_POS_599_43311a11_144 | | 599.4331 | 11.14 | | RP | POS | | | 924712 | 205763 | 874658 | 213618 | 8.25E-02 | -0.24 | 1.15 |
| HILIC_NEG_115_92093a7_378 | | 115.9209 | 7.38 | | HILIC | NEG | | | 3.2E+07 | 7254560 | 3.3E+07 | 8056048 | 8.26E-02 | 0.24 | 1.32 |
| RP_POS_702_21417a11_045 | | 702.2142 | 11.05 | | RP | POS | | | 604935 | 131289 | 639436 | 155334 | 8.26E-02 | 0.24 | 1.03 |
| HILIC_POS_162_07616a1_079 | | 162.0762 | 1.08 | | HILIC | POS | | | 1469121 | 2345365 | 1056063 | 937515 | 8.26E-02 | -0.25 | 0.91 |
| HILIC_NEG_343_99533a7_095 | | 343.9953 | 7.10 | | HILIC | NEG | | | 750195 | 123372 | 724677 | 91344.5 | 8.28E-02 | -0.24 | 0.96 |
| HILIC_NEG_308_99771a3_049 | | 308.9977 | 3.05 | | HILIC | NEG | | | 482806 | 52444.7 | 470999 | 47141.1 | 8.30E-02 | -0.24 | 1.13 |
| RP_POS_237_1489a7_009 | | 237.1489 | 7.01 | | RP | POS | | | 683379 | 270251 | 624299 | 228358 | 8.30E-02 | -0.24 | 1.17 |
| RP_POS_370_68869a9_207 | | 370.6887 | 9.21 | | RP | POS | | | 300597 | 227280 | 251868 | 183757 | 8.30E-02 | -0.24 | 1 |
| RP_POS_533_33936a10_625 | | 533.3394 | 10.63 | | RP | POS | | | 1993899 | 594112 | 2143050 | 655332 | 8.35E-02 | 0.24 | 1.19 |
| RP_POS_416_31577a9_779 | | 416.3158 | 9.78 | | RP | POS | | | 1045792 | 1234583 | 769451 | 1100532 | 8.35E-02 | -0.24 | 1.08 |
| RP_POS_315_18005a8_838 | | 315.1801 | 8.84 | | RP | POS | | | 993139 | 612476 | 1205324 | 1073399 | 8.36E-02 | 0.25 | 1.47 |
| RP_NEG_411_34851a11_496 | | 411.3485 | 11.50 | | RP | NEG | | | 434828 | 61424 | 420029 | 63154.7 | 8.38E-02 | -0.24 | 1.4 |
| RP_NEG_615_1413a8_764 | | 615.1413 | 8.76 | | RP | NEG | | | 763809 | 42941.7 | 754236 | 38095 | 8.41E-02 | -0.24 | 1.32 |
| RP_POS_137_0459a1_701 | | 137.0459 | 1.70 | | RP | POS | 137.04628100) | | 1578680 | 1756347 | 1243358 | 1041909 | 8.42E-02 | -0.24 | 1.07 |
| RP_POS_825_03772a13_104 | | 825.0377 | 13.10 | | RP | POS | | | 702574 | 472748 | 846901 | 703268 | 8.43E-02 | 0.25 | 1.18 |
| RP_POS_776_57263a9_911 | | 776.5726 | 9.91 | | RP | POS | | | 3603337 | 1438787 | 3248280 | 1547860 | 8.43E-02 | -0.24 | 1.28 |
| RP_POS_588_74103a9_993 | | 588.7410 | 9.99 | | RP | POS | | | 273590 | 154142 | 315268 | 192383 | 8.44E-02 | 0.24 | 1.04 |
| RP_POS_439_27579a12_109 | | 439.2758 | 12.11 | | RP | POS | | | 153058 | 54308.3 | 140303 | 53483.3 | 8.44E-02 | -0.24 | 1.16 |
| HILIC_POS_274_87317a2_981 | | 274.8732 | 2.98 | | HILIC | POS | | | 343225 | 57593.9 | 356541 | 55104.9 | 8.45E-02 | 0.24 | 1.22 |
| RP_POS_327_2276a11_239 | | 327.2276 | 11.24 | | RP | POS | | | 4770462 | 4196105 | 5829627 | 4697808 | 8.46E-02 | 0.24 | 1.22 |
| RP_POS_612_55591a11_532 | | 612.5559 | 11.53 | | RP | POS | | | 887352 | 2068938 | 521076 | 888387 | 8.46E-02 | -0.25 | 1.05 |
| RP_POS_730_5401a12_59 | PC(18:2/14:0) | 730.5401 | 12.59 | | RP | POS | 184.0744(100), 86.0972(20) | 1 | 6E+07 | 2.1E+07 | 5.5E+07 | 2.1E+07 | 8.48E-02 | -0.24 | 1.05 |
| RP_POS_568_3371a10_509 | | 568.3371 | 10.51 | | RP | POS | | | 7048171 | 1481143 | 7427291 | 1699732 | 8.49E-02 | 0.24 | 1.15 |
| HILIC_POS_566_32233a1_152 | | 566.3223 | 1.15 | | HILIC | POS | | | 574324 | 209972 | 622361 | 197418 | 8.50E-02 | 0.24 | 1 |
| RP_POS_185_11465a13_183 | | 185.1147 | 13.18 | | RP | POS | | | 1897318 | 893349 | 2130674 | 1059717 | 8.50E-02 | 0.24 | 1.09 |
| RP_NEG_601_16028a9_418 | | 601.1603 | 9.42 | | RP | NEG | | | 73025.7 | 11162 | 70633.1 | 9158.21 | 8.50E-02 | -0.24 | 1.06 |
| RP_POS_550_40485a10_373 | | 550.4049 | 10.37 | | RP | POS | | | 4083809 | 1708227 | 3644597 | 1975684 | 8.50E-02 | -0.24 | 1.35 |
| HILIC_POS_291_06937a2_49 | Inosine | 291.0694 | 2.49 | | HILIC | POS | 159.0274(100) | 2 | 950735 | 1032845 | 754501 | 608195 | 8.51E-02 | -0.24 | 1.04 |
| RP_POS_148_97713a0_629 | | 148.9771 | 0.63 | | RP | POS | | | 318403 | 114576 | 290304 | 122441 | 8.51E-02 | -0.24 | 1.04 |
| RP_NEG_1085_6792a10_498 | | 1085.6792 | 10.50 | | RP | NEG | | | 1856971 | 492573 | 1981137 | 552016 | 8.52E-02 | 0.24 | 1.12 |
| RP_POS_179_14287a6_923 | | 179.1429 | 6.92 | | RP | POS | | | 445732 | 283370 | 388866 | 197993 | 8.54E-02 | -0.24 | 1.16 |
| RP_NEG_288_61984a7_546 | | 288.6198 | 7.55 | | RP | NEG | | | 4036824 | 4238185 | 5043344 | 4289977 | 8.55E-02 | 0.24 | 1.08 |
| RP_POS_668_13257a11_286 | | 668.1326 | 11.29 | | RP | POS | | | 204557 | 98904.9 | 231067 | 122516 | 8.55E-02 | 0.24 | 1.15 |
| RP_POS_386_10272a9_208 | | 386.1027 | 9.21 | | RP | POS | | | 443092 | 339628 | 371306 | 270850 | 8.55E-02 | -0.24 | 1.01 |
| RP_POS_357_08975a9_207 | | 357.0898 | 9.21 | | RP | POS | | | 342700 | 271709 | 287983 | 192162 | 8.57E-02 | -0.24 | 1.03 |
| RP_POS_361_20074a6_738 | Cortisone | 361.2007 | 6.74 | | RP | POS | 361.2034(100),163.1125(60) | 1 | 4039423 | 946713 | 4261087 | 934981 | 8.58E-02 | 0.24 | 1 |
| RP_NEG_388_87329a15_376 | | 388.8733 | 15.38 | | RP | NEG | | | 654039 | 319687 | 583282 | 282899 | 8.59E-02 | -0.23 | 0.96 |
| RP_NEG_309_28073a11_673 | | 309.2807 | 11.67 | | RP | NEG | | | 411072 | 305866 | 485649 | 325339 | 8.60E-02 | 0.24 | 1.34 |
| HILIC_POS_184_09427a4_818 | | 184.0943 | 4.82 | | HILIC | POS | | | 4004506 | 1428274 | 4309653 | 1171633 | 8.60E-02 | 0.23 | 1.07 |
| RP_POS_375_29572a11_144 | | 375.2957 | 11.14 | | RP | POS | | | 2809549 | 543846 | 2687450 | 495818 | 8.60E-02 | -0.23 | 1.26 |
| RP_POS_342_26419a8_021 | | 342.2642 | 8.02 | | RP | POS | 85.0291(100), 342.2649(35) | | 1452179 | 834766 | 1661520 | 932847 | 8.63E-02 | 0.24 | 1.23 |
| RP_POS_641_77692a10_235 | | 641.7769 | 10.24 | | RP | POS | | | 243555 | 161349 | 283980 | 180031 | 8.63E-02 | 0.24 | 1.09 |
| RP_POS_257_11267a0_68 | | 257.1127 | 0.68 | | RP | POS | | | 386422 | 74230.5 | 371304 | 54469.2 | 8.65E-02 | -0.23 | 1.01 |
| RP_POS_741_91846a11_561 | | 741.9185 | 11.56 | | RP | POS | | | 124116 | 58155.3 | 142169 | 89420.4 | 8.66E-02 | 0.24 | 1.29 |
| RP_POS_304_21115a4_936 | | 304.2112 | 4.94 | | RP | POS | | | 755515 | 413487 | 860480 | 472836 | 8.68E-02 | 0.24 | 1.24 |
| RP_POS_477_12991a10_006 | | 477.1299 | 10.01 | | RP | POS | | | 895423 | 771667 | 1089576 | 870608 | 8.70E-02 | 0.24 | 1.23 |
| RP_NEG_552_30994a10_28 | | 552.3099 | 10.28 | | RP | NEG | | | 344765 | 107489 | 320340 | 101002 | 8.70E-02 | -0.23 | 1.14 |
| RP_NEG_707_76495a6_032 | | 707.7650 | 6.03 | | RP | NEG | | | 792693 | 339713 | 710927 | 355192 | 8.70E-02 | -0.24 | 1.28 |
| RP_POS_644_94916a10_105 | | 644.9492 | 10.11 | | RP | POS | | | 1598005 | 703482 | 1429241 | 731374 | 8.70E-02 | -0.24 | 1.09 |
| RP_POS_375_28912a9_901 | | 375.2891 | 9.90 | | RP | POS | | | 815474 | 597276 | 962447 | 649309 | 8.71E-02 | 0.24 | 1 |
| HILIC_POS_175_10771a5_84 | | 175.1077 | 5.84 | | HILIC | POS | | | 623223 | 287863 | 563348 | 223588 | 8.71E-02 | -0.23 | 1.01 |
| RP_POS_509_50348a13_145 | | 509.5035 | 13.15 | | RP | POS | | | 965792 | 415447 | 1071071 | 474948 | 8.73E-02 | 0.24 | 0.99 |
| RP_POS_482_84683a10_237 | | 482.8468 | 10.24 | | RP | POS | | | 284345 | 147747 | 250124 | 144564 | 8.76E-02 | -0.23 | 1.17 |
| RP_POS_441_3208a11_158 | | 441.3208 | 11.16 | | RP | POS | | | 770475 | 387989 | 880848 | 530778 | 8.77E-02 | 0.24 | 1.35 |
| HILIC_NEG_267_07358a2_486 | Inosine | 267.0736 | 2.49 | | HILIC | NEG | 135.0317(100), 264.0731(40) | 2 | 659726 | 738046 | 521104 | 429676 | 8.77E-02 | -0.24 | 1.04 |
| RP_POS_724_90289a11_333 | | 724.9029 | 11.33 | | RP | POS | | | 153850 | 79518.5 | 178136 | 120183 | 8.78E-02 | 0.24 | 1.05 |
| RP_POS_564_23456a10_549 | | 564.2346 | 10.55 | | RP | POS | | | 1094739 | 759211 | 1276426 | 789890 | 8.79E-02 | 0.23 | 1.39 |
| RP_POS_400_34195a9_422 | AC 16:0 (Palmitoylcarnitine) | 400.3420 | 9.42 | | RP | POS | 85.0293(100), 400.3419(50) | 1 | 1.2E+07 | 2934352 | 1.2E+07 | 3031263 | 8.80E-02 | 0.23 | 1.43 |
| RP_POS_627_32642a14_339 | | 627.3264 | 14.34 | | RP | POS | | | 1300990 | 750658 | 1155231 | 488760 | 8.80E-02 | -0.24 | 0.92 |
| HILIC_POS_144_98209a7_431 | | 144.9821 | 7.43 | | HILIC | POS | | | 2040734 | 197503 | 1996524 | 181401 | 8.81E-02 | -0.23 | 1.11 |
| RP_NEG_187_00717a3_63 | | 187.0072 | 3.63 | | RP | NEG | | | 5.8E+07 | 4.3E+07 | 4.8E+07 | 3.6E+07 | 8.81E-02 | -0.23 | 1.04 |
| RP_NEG_787_10144a9_416 | | 787.1014 | 9.42 | | RP | NEG | | | 5345827 | 418040 | 5254771 | 363844 | 8.85E-02 | -0.23 | 1.15 |
| RP_POS_514_37567a10_022 | | 514.3757 | 10.02 | | RP | POS | | | 9695711 | 4438922 | 8777242 | 3443024 | 8.85E-02 | -0.23 | 1 |
| HILIC_POS_140_06825a5_455 | | 140.0683 | 5.46 | | HILIC | POS | | | 3213472 | 319847 | 3131235 | 376681 | 8.85E-02 | -0.24 | 0.98 |
| HILIC_POS_288_21689a0_833 | | 288.2169 | 0.83 | | HILIC | POS | 85.0293(100), 288.218(40) | | 6749088 | 5728154 | 9029663 | 1.2E+07 | 8.87E-02 | 0.26 | 1.08 |

**Supplemantary table 1**: The non-targeted metabolomics results ordered by the VIP value (p value < 0.0003 are shaded dark green, p value < 0.05 are shaded light green, d values below 0 are blue and above 0 are red)

| | | | | | | | | | Cases | | Controls | | Welch test | Cohen's | |
|---|---|---|---|---|---|---|---|---|---|---|---|---|---|---|---|
| Feature_ID | Metabolite_name | Average_Mz | Average_RT_min_ | HMDB ID | RP_HILIC | POS_NEG | Cureted_MSMS | ID_level | Mean | SD | Mean2 | SD3 | p | d | VIP |
| RP_NEG_267_07364a1_704 | Inosine | 267.0736 | 1.70 | | RP | NEG | 135.0316(100), 267.0729(45) | 2 | 919720 | 1013183 | 728260 | 609205 | 8.87E-02 | -0.24 | 1.06 |
| RP_POS_377_30194a11_021 | | 377.3019 | 11.02 | | RP | POS | | | 931127 | 144433 | 897747 | 141631 | 8.87E-02 | -0.23 | 1.46 |
| RP_POS_134_08125a0_628 | N-Methylthreonine | 134.0813 | 0.63 | | RP | POS | 88.0762(100), 70.066(38), 116.0713(30) | 2 | 5171092 | 1038791 | 5403425 | 956571 | 8.88E-02 | 0.23 | 1.14 |
| HILIC_NEG_209_00459a5_118 | | 209.0046 | 5.12 | | HILIC | NEG | | | 621147 | 343933 | 551143 | 256705 | 8.88E-02 | -0.23 | 0.95 |
| RP_POS_459_16687a8_842 | | 459.1669 | 8.84 | | RP | POS | | | 132116 | 88568.5 | 114720 | 60077.9 | 8.88E-02 | -0.23 | 1.05 |
| HILIC_NEG_180_98929a1_355 | | 180.9893 | 1.36 | | HILIC | NEG | | | 1159400 | 369894 | 1074598 | 357752 | 8.89E-02 | -0.23 | 1.16 |
| RP_POS_760_5849a13_799 | | 760.5849 | 13.80 | | RP | POS | | | 8.8E+08 | 2.5E+08 | 9.4E+08 | 2.6E+08 | 8.91E-02 | 0.23 | 1.21 |
| HILIC_POS_161_01859a1_012 | | 161.0186 | 1.01 | | HILIC | POS | | | 226819 | 64929.4 | 242980 | 72848.4 | 8.94E-02 | 0.23 | 1.19 |
| RP_POS_158_15402a6_704 | | 158.1540 | 6.70 | | RP | POS | | | 1277692 | 1970221 | 1799363 | 2442079 | 8.95E-02 | 0.24 | 0.96 |
| RP_NEG_530_3031a10_491 | | 530.3031 | 10.49 | | RP | NEG | | | 8858018 | 875505 | 8651592 | 894631 | 8.95E-02 | -0.23 | 1 |
| RP_POS_495_02307a10_366 | | 495.0231 | 10.37 | | RP | POS | | | 664439 | 405196 | 574547 | 368805 | 8.95E-02 | -0.23 | 1.06 |
| RP_NEG_415_21671a6_986 | | 415.2167 | 6.99 | | RP | NEG | | | 691688 | 569476 | 543467 | 688335 | 8.98E-02 | -0.24 | 1.08 |
| RP_NEG_590_34668a10_502 | | 590.3467 | 10.50 | | RP | NEG | | | 1.1E+07 | 2471527 | 1.2E+07 | 2769149 | 9.00E-02 | 0.23 | 1.08 |
| HILIC_POS_204_12337a2_891 | AC 02:0 (Acetylcarnitine) | 204.1234 | 2.89 | | HILIC | POS | 85.0292(100), 204.1229(50) | 1 | 6.7E+08 | 2E+08 | 7.2E+08 | 2.3E+08 | 9.02E-02 | 0.23 | 1.25 |
| RP_POS_399_25443a12_111 | | 399.2544 | 12.11 | | RP | POS | | | 581374 | 211547 | 534408 | 193625 | 9.02E-02 | -0.23 | 1.4 |
| RP_NEG_552_36676a11_102 | | 552.3668 | 11.10 | | RP | NEG | | | 510789 | 155345 | 474850 | 154274 | 9.05E-02 | -0.23 | 1.04 |
| RP_POS_167_55321a15_441 | | 167.5532 | 15.44 | | RP | POS | | | 367140 | 183268 | 417595 | 242207 | 9.06E-02 | 0.24 | 1.09 |
| RP_POS_361_20398a11_367 | | 361.2040 | 11.37 | | RP | POS | | | 262443 | 103647 | 241381 | 78158.3 | 9.06E-02 | -0.23 | 1.29 |
| RP_POS_508_35965a8_904 | | 508.3597 | 8.90 | | RP | POS | | | 454589 | 299789 | 394267 | 220566 | 9.07E-02 | -0.23 | 1 |
| RP_POS_687_54156a12_13 | | 687.5416 | 12.13 | | RP | POS | | | 1262001 | 469450 | 1148009 | 508715 | 9.07E-02 | -0.23 | 1.1 |
| HILIC_NEG_500_27954a1_756 | | 500.2795 | 1.76 | | HILIC | NEG | | | 305071 | 83204.6 | 326466 | 99119.4 | 9.08E-02 | 0.23 | 1.22 |
| RP_POS_379_20316a10_651 | | 379.2032 | 10.65 | | RP | POS | | | 1696468 | 734729 | 1858522 | 666486 | 9.08E-02 | 0.23 | 1.2 |
| RP_POS_339_19806a11_594 | | 339.1981 | 11.59 | | RP | POS | | | 821154 | 262362 | 764493 | 227750 | 9.08E-02 | -0.23 | 1.24 |
| RP_POS_780_2121a8_892 | | 780.2121 | 8.89 | | RP | POS | | | 607209 | 73139.8 | 623832 | 70380.8 | 9.09E-02 | 0.23 | 1.3 |
| RP_POS_337_27359a10_685 | | 337.2736 | 10.69 | | RP | POS | | | 2207345 | 1204633 | 2613868 | 2103444 | 9.10E-02 | 0.25 | 1.17 |
| HILIC_NEG_312_83643a1_31 | | 312.8364 | 1.31 | | HILIC | NEG | | | 150815 | 47788.8 | 140901 | 37985.1 | 9.10E-02 | -0.23 | 1.09 |
| RP_NEG_453_28644a9_285 | Muricholic acid | 453.2864 | 9.29 | | RP | NEG | 407.2766(100) | 2 | 341272 | 570210 | 593444 | 1377234 | 9.11E-02 | 0.26 | 1 |
| RP_NEG_289_70258a15_495 | | 289.7026 | 15.50 | | RP | NEG | | | 1157298 | 51145.2 | 1144843 | 55815.7 | 9.11E-02 | -0.23 | 1.11 |
| RP_POS_394_1972a5_296 | | 394.1972 | 5.30 | | RP | POS | | | 729878 | 569406 | 871483 | 644243 | 9.13E-02 | 0.23 | 1.32 |
| RP_POS_307_11118a4_875 | Fluconazole | 307.1112 | 4.88 | | RP | POS | 220.0684(100), 238.0798(80) | 1 | 184091 | 383400 | 459717 | 1570948 | 9.14E-02 | 0.28 | 1.13 |
| RP_POS_697_52545a12_24 | | 697.5255 | 12.24 | | RP | POS | | | 1.2E+07 | 3299661 | 1.2E+07 | 2620002 | 9.14E-02 | -0.23 | 1.11 |
| RP_POS_579_2644a11_415 | | 579.2644 | 11.42 | | RP | POS | | | 84538.2 | 45632.3 | 96129.9 | 53503.5 | 9.15E-02 | 0.23 | 1.01 |
| RP_POS_718_90381a11_429 | | 718.9038 | 11.43 | | RP | POS | | | 142351 | 74106.9 | 161788 | 91389 | 9.15E-02 | 0.23 | 1.01 |
| HILIC_NEG_194_10355a1_437 | | 194.1036 | 1.44 | | HILIC | NEG | | | 475102 | 110794 | 449975 | 106594 | 9.15E-02 | -0.23 | 1.32 |
| HILIC_NEG_134_89474a6_478 | | 134.8947 | 6.48 | | HILIC | NEG | | | 1.3E+07 | 8692766 | 1.5E+07 | 9301800 | 9.16E-02 | 0.23 | 1.04 |
| RP_NEG_407_28067a9_285 | Cholic acid | 407.2807 | 9.29 | | RP | NEG | 407.2766(100), 343.2675(100) | 2 | 878196 | 1468905 | 1536804 | 3609553 | 9.16E-02 | 0.26 | 1 |
| RP_POS_269_08795a0_68 | | 269.0880 | 0.68 | | RP | POS | | | 2244579 | 2400155 | 1797223 | 1421747 | 9.17E-02 | -0.23 | 1.03 |
| HILIC_NEG_191_81664a3_169 | | 191.8166 | 3.17 | | HILIC | NEG | | | 709148 | 103291 | 730220 | 79370.2 | 9.18E-02 | 0.23 | 1.02 |
| HILIC_POS_144_10181a4_578 | | 144.1018 | 4.58 | | HILIC | POS | 144.1026(100), 98.0969(5) | | 696979 | 952920 | 514001 | 621937 | 9.18E-02 | -0.23 | 0.91 |
| RP_NEG_573_12915a9_386 | | 573.1292 | 9.39 | | RP | NEG | | | 2751950 | 574497 | 2616407 | 595588 | 9.18E-02 | -0.23 | 1.21 |
| RP_POS_429_32095a11_057 | | 429.3210 | 11.06 | | RP | POS | | | 1977924 | 696884 | 2183950 | 1025518 | 9.19E-02 | 0.24 | 1.39 |
| RP_NEG_772_52936a13_362 | | 772.5294 | 13.36 | | RP | NEG | | | 1298303 | 377235 | 1217051 | 328005 | 9.19E-02 | -0.23 | 1.13 |
| RP_NEG_553_19855a11_237 | | 553.1986 | 11.24 | | RP | NEG | | | 398372 | 183013 | 441415 | 189073 | 9.22E-02 | 0.23 | 1.36 |
| RP_NEG_160_97676a15_345 | | 160.9768 | 15.35 | | RP | NEG | | | 5594919 | 1750139 | 5231211 | 1411493 | 9.24E-02 | -0.23 | 1.07 |
| RP_POS_546_35541a10_381 | | 546.3554 | 10.38 | | RP | POS | | | 1E+07 | 3039565 | 1.1E+07 | 3350234 | 9.25E-02 | 0.23 | 1.12 |
| RP_POS_674_50403a9_699 | | 674.5040 | 9.70 | | RP | POS | | | 4828678 | 2022366 | 4394374 | 1754136 | 9.25E-02 | -0.23 | 1.21 |
| RP_POS_355_26324a9_287 | | 355.2632 | 9.29 | | RP | POS | | | 912071 | 1329010 | 1525119 | 3384917 | 9.26E-02 | 0.26 | 1 |
| HILIC_POS_158_09229a6_958 | | 158.0923 | 6.96 | | HILIC | POS | | | 304730 | 58637.7 | 318672 | 61901 | 9.26E-02 | 0.23 | 1.02 |
| HILIC_NEG_115_04032a1_014 | | 115.0403 | 1.01 | | HILIC | NEG | | | 6616223 | 1606864 | 6993152 | 1656442 | 9.27E-02 | 0.23 | 1.2 |
| RP_POS_444_33185a11_049 | | 444.3319 | 11.05 | | RP | POS | | | 751017 | 394869 | 847807 | 439442 | 9.27E-02 | 0.23 | 1.42 |
| RP_NEG_376_98605a7_2 | | 376.9861 | 7.20 | | RP | NEG | | | 1910911 | 465350 | 1812477 | 391253 | 9.27E-02 | -0.23 | 1.13 |
| RP_NEG_248_96085a13_289 | | 248.9609 | 13.29 | | RP | NEG | | | 1646679 | 322698 | 1715843 | 279406 | 9.28E-02 | 0.23 | 1.14 |
| HILIC_NEG_136_89169a7_743 | | 136.8917 | 7.74 | | HILIC | NEG | | | 6822812 | 1499625 | 6489009 | 1402015 | 9.28E-02 | -0.23 | 1.04 |
| RP_POS_1005_98859a6_429 | | 1005.9886 | 6.43 | | RP | POS | | | 1098302 | 948235 | 881765 | 931155 | 9.28E-02 | -0.23 | 1.62 |
| RP_POS_369_19839a11_144 | | 369.1984 | 11.14 | | RP | POS | | | 1739563 | 202773 | 1690435 | 221300 | 9.28E-02 | -0.23 | 0.99 |
| RP_POS_510_24377a11_141 | | 510.2438 | 11.14 | | RP | POS | | | 857829 | 522302 | 989766 | 611492 | 9.30E-02 | 0.23 | 1.36 |
| RP_POS_667_79968a11_52 | | 667.7997 | 11.52 | | RP | POS | | | 150278 | 69078.3 | 170024 | 97513.4 | 9.32E-02 | 0.24 | 1.06 |
| RP_NEG_303_23306a10_926 | FA 20:4 (Arachidonic acid) | 303.2331 | 10.93 | | RP | NEG | 303.2345(100), 259.2434(15) | 2 | 2E+07 | 8996373 | 2.2E+07 | 8848021 | 9.32E-02 | 0.23 | 1.3 |
| RP_POS_288_21686a6_109 | AC 08:0 (Octanoylcarnitine) | 288.2169 | 6.11 | | RP | POS | 85.0291(100), 288.2174(40) | 2 | 9125769 | 7799016 | 1.2E+07 | 1.8E+07 | 9.32E-02 | 0.25 | 1.08 |
| RP_POS_420_40329a11_974 | | 420.4033 | 11.97 | | RP | POS | | | 157199 | 70053.7 | 175571 | 87210.1 | 9.35E-02 | 0.23 | 1.13 |
| RP_POS_427_15918a8_06 | | 427.1592 | 8.06 | | RP | POS | | | 759441 | 421150 | 669247 | 366161 | 9.37E-02 | -0.23 | 0.98 |
| RP_POS_377_32635a10_694 | | 377.3264 | 10.69 | | RP | POS | | | 4033485 | 486235 | 3939568 | 328841 | 9.39E-02 | -0.23 | 0.94 |
| RP_POS_526_87231a10_194 | | 526.8723 | 10.19 | | RP | POS | | | 605318 | 356042 | 530550 | 297166 | 9.40E-02 | -0.23 | 1.06 |
| RP_NEG_609_04846a8_98 | | 609.0485 | 8.98 | | RP | NEG | | | 294471 | 33373.2 | 302348 | 35064.3 | 9.41E-02 | 0.23 | 1.39 |
| RP_POS_429_2395a15_413 | | 429.2395 | 15.41 | | RP | POS | | | 1713529 | 363622 | 1791476 | 317439 | 9.41E-02 | 0.23 | 0.96 |
| HILIC_NEG_187_12025a6_842 | | 187.1203 | 6.84 | | HILIC | NEG | | | 100173 | 48885.5 | 89749.5 | 42205.9 | 9.41E-02 | -0.23 | 0.94 |
| RP_POS_374_21036a11_143 | | 374.2104 | 11.14 | | RP | POS | | | 1795582 | 317687 | 1723512 | 310570 | 9.41E-02 | -0.23 | 1.39 |
| RP_POS_381_70935a6_78 | | 381.7094 | 6.78 | | RP | POS | | | 394953 | 53182.8 | 406868 | 50784.1 | 9.42E-02 | 0.23 | 1.03 |
| HILIC_NEG_383_18997a0_933 | | 383.1900 | 0.93 | | HILIC | NEG | | | 100954 | 23176.8 | 95710.3 | 22588 | 9.45E-02 | -0.23 | 0.98 |
| HILIC_NEG_241_75854a1_404 | | 241.7585 | 1.40 | | HILIC | NEG | | | 128290 | 33210 | 135161 | 26940.3 | 9.47E-02 | 0.23 | 0.94 |
| RP_POS_304_26147a15_264 | | 304.2615 | 15.26 | | RP | POS | | | 1014566 | 503763 | 904720 | 457503 | 9.47E-02 | -0.23 | 1.19 |
| RP_POS_373_2738a9_287 | | 373.2738 | 9.29 | | RP | POS | 355.261(100) | | 1070879 | 1692270 | 1825533 | 4180616 | 9.48E-02 | 0.26 | 0.99 |
| HILIC_POS_387_21393a2_236 | | 387.2139 | 2.24 | | HILIC | POS | | | 460714 | 656718 | 639666 | 871493 | 9.49E-02 | 0.23 | 1.13 |
| HILIC_NEG_136_89172a0_69 | | 136.8917 | 0.69 | | HILIC | NEG | | | 5550215 | 1669685 | 5139017 | 1893266 | 9.49E-02 | -0.23 | 1.12 |
| RP_NEG_485_27228a13_294 | | 485.2723 | 13.29 | | RP | NEG | | | 262737 | 61936.4 | 277247 | 64609.5 | 9.52E-02 | 0.23 | 1.28 |
| RP_POS_204_1235a0_671 | Acetylcarnitine | 204.1235 | 0.67 | | RP | POS | 85.02938100), 204.1232(50) | 2 | 1.4E+08 | 4.2E+07 | 1.5E+08 | 4.4E+07 | 9.52E-02 | 0.23 | 1.29 |
| RP_POS_316_24814a7_373 | AC 10:0 (Decanoylcarnitine) | 316.2481 | 7.37 | | RP | POS | 85.0292(100), 316.2497(40) | 1 | 1.9E+07 | 1.6E+07 | 2.5E+07 | 3.2E+07 | 9.52E-02 | 0.25 | 1.07 |
| RP_POS_327_33624a9_768 | | 327.3362 | 9.77 | | RP | POS | | | 115949 | 147884 | 190940 | 422473 | 9.54E-02 | 0.26 | 1.01 |
| RP_POS_530_44299a12_422 | | 530.4430 | 12.42 | | RP | POS | | | 679588 | 783670 | 550720 | 252327 | 9.54E-02 | -0.25 | 0.94 |
| RP_POS_797_08539a13_923 | | 797.0854 | 13.92 | | RP | POS | | | 1464189 | 1689061 | 2060257 | 3174830 | 9.55E-02 | 0.25 | 1.28 |
| RP_POS_332_24298a6_404 | | 332.2430 | 6.40 | | RP | POS | | | 1445902 | 1019763 | 1704582 | 1216944 | 9.55E-02 | 0.23 | 1.06 |
| RP_POS_421_79465a9_883 | | 421.7947 | 9.88 | | RP | POS | | | 1084826 | 645726 | 958122 | 462877 | 9.56E-02 | -0.23 | 1.05 |
| RP_POS_465_43076a11_285 | | 465.4308 | 11.29 | | RP | POS | | | 185876 | 61705.2 | 172087 | 59106 | 9.56E-02 | -0.23 | 0.96 |
| RP_POS_404_78741a14_105 | | 404.7874 | 14.11 | | RP | POS | | | 1748361 | 2195087 | 2426437 | 3483390 | 9.58E-02 | 0.24 | 1.15 |
| RP_NEG_431_11288a11_141 | | 431.1129 | 11.14 | | RP | NEG | | | 431078 | 40381 | 421327 | 44527.4 | 9.58E-02 | -0.23 | 1.16 |
| HILIC_POS_211_13278a1 | | 211.1328 | 1.00 | | HILIC | POS | | | 845354 | 427648 | 762038 | 301488 | 9.59E-02 | -0.23 | 0.96 |
| HILIC_NEG_212_9111a3_45 | | 212.9111 | 3.45 | | HILIC | NEG | | | 2748014 | 1283274 | 2470046 | 1158966 | 9.60E-02 | -0.23 | 0.95 |
| RP_POS_655_46954a14_203 | | 655.4695 | 14.20 | | RP | POS | | | 430391 | 599015 | 325468 | 292537 | 9.60E-02 | -0.24 | 0.95 |
| HILIC_POS_269_08786a2_482 | Inosine | 269.0879 | 2.48 | | HILIC | POS | 137.0462(100) | 1 | 977338 | 1102441 | 776282 | 634705 | 9.61E-02 | -0.23 | 1.04 |
| HILIC_NEG_178_85938a3_143 | | 178.8594 | 3.14 | | HILIC | NEG | | | 770530 | 161691 | 801519 | 109175 | 9.62E-02 | 0.23 | 0.93 |
| RP_NEG_826_56183a12_878 | PC 36:4 (18:2_18:2) | 826.5618 | 12.88 | | RP | NEG | 279.2337(100), 766.5322(5) | 2 | 3.2E+07 | 9645172 | 3E+07 | 7966097 | 9.63E-02 | -0.23 | 0.96 |
| RP_POS_589_90039a10_153 | | 589.9004 | 10.15 | | RP | POS | | | 3753660 | 1836933 | 3344804 | 1753243 | 9.63E-02 | -0.23 | 1.21 |
| HILIC_POS_90_05558a5_649 | Alanine | 90.0556 | 5.65 | | HILIC | POS | 90.0557(100) | 2 | 2.6E+07 | 1E+07 | 2.4E+07 | 9444974 | 9.64E-02 | -0.23 | 1.03 |
| RP_NEG_747_37018a4_856 | | 747.3702 | 4.86 | | RP | NEG | | | 974211 | 500104 | 855415 | 537809 | 9.64E-02 | -0.23 | 1.41 |
| RP_POS_279_17026a4_648 | | 279.1703 | 4.65 | | RP | POS | 86.0973(100) | | 2253074 | 2314321 | 1745459 | 2149523 | 9.65E-02 | -0.23 | 1.15 |
| RP_POS_634_1095a11_206 | | 634.1095 | 11.21 | | RP | POS | | | 165949 | 65797.6 | 186952 | 109397 | 9.66E-02 | 0.24 | 1.08 |
| RP_POS_415_28174a9_902 | | 415.2817 | 9.90 | | RP | POS | 415.2807(100) | | 2043492 | 1556002 | 2438901 | 1871739 | 9.66E-02 | 0.23 | 0.99 |
| RP_POS_191_12782a6_5 | | 191.1278 | 6.50 | | RP | POS | | | 9.1E+07 | 8248049 | 9.3E+07 | 7869750 | 9.66E-02 | 0.23 | 1.37 |
| HILIC_NEG_367_10599a5_121 | | 367.1060 | 5.12 | | HILIC | NEG | | | 472769 | 271715 | 577872 | 573472 | 9.69E-02 | 0.25 | 1.04 |
| RP_POS_569_43439a14_534 | | 569.4344 | 14.53 | | RP | POS | | | 1017327 | 702651 | 1196444 | 850675 | 9.69E-02 | 0.23 | 1.13 |
| RP_NEG_381_02902a5_567 | | 381.0290 | 5.57 | | RP | NEG | | | 441618 | 1154871 | 233285 | 644784 | 9.70E-02 | -0.23 | 0.97 |
| RP_POS_805_02594a10_926 | | 805.0259 | 10.93 | | RP | POS | | | 246834 | 197624 | 293550 | 211626 | 9.72E-02 | 0.23 | 1.06 |
| RP_POS_258_1088a0_677 | | 258.1088 | 0.68 | | RP | POS | | | 297165 | 74870.3 | 316186 | 90416.9 | 9.75E-02 | 0.23 | 1.38 |
| RP_NEG_201_12442a0_682 | | 201.1244 | 0.68 | | RP | NEG | | | 274732 | 100970 | 251382 | 104143 | 9.75E-02 | -0.23 | 1.14 |
| RP_NEG_448_30719a9_374 | | 448.3072 | 9.37 | | RP | NEG | 448.3039(100), 74.0243(55) | | 1.1E+07 | 1.3E+07 | 7968933 | 1E+07 | 9.76E-02 | -0.23 | 0.96 |
| HILIC_POS_274_20117a0_93 | | 274.2012 | 0.93 | | HILIC | POS | | | 321057 | 159393 | 357589 | 162002 | 9.77E-02 | 0.23 | 1.07 |
| HILIC_POS_137_04584a2_476 | | 137.0458 | 2.48 | | HILIC | POS | 137.0462(100), 119.0318(1) | | 3923341 | 4576113 | 3090179 | 2662127 | 9.77E-02 | -0.23 | 1.03 |
| RP_POS_275_22168a9_73 | | 275.2217 | 9.73 | | RP | POS | | | 169747 | 104193 | 255861 | 503816 | 9.78E-02 | 0.28 | 1.49 |
| RP_POS_273_20575a8_449 | | 273.2058 | 8.45 | | RP | POS | | | 948445 | 96601.3 | 926604 | 95783.8 | 9.78E-02 | -0.23 | 1.29 |
| RP_NEG_567_53595a11_592 | | 567.5360 | 11.59 | | RP | NEG | | | 2603077 | 1010651 | 2353809 | 1166014 | 9.79E-02 | -0.23 | 1.45 |
| RP_NEG_461_14584a11_592 | | 461.1458 | 11.59 | | RP | NEG | | | 2136904 | 143029 | 2103701 | 148978 | 9.80E-02 | -0.23 | 1.24 |
| RP_NEG_385_16931a5_67 | | 385.1693 | 5.67 | | RP | NEG | | | 2060406 | 1020691 | 2293830 | 1035703 | 9.81E-02 | 0.23 | 1.06 |
| RP_NEG_333_9823a0_672 | | 333.9823 | 0.67 | | RP | NEG | | | 153604 | 51465.2 | 143822 | 34478.7 | 9.81E-02 | -0.23 | 0.94 |
| RP_POS_331_2265a8_363 | | 331.2265 | 8.36 | | RP | POS | | | 498918 | 166669 | 584600 | 488154 | 9.83E-02 | 0.26 | 1.07 |
| RP_POS_516_35315a10_772 | | 516.3532 | 10.77 | | RP | POS | | | 2348104 | 1289961 | 2709123 | 1806957 | 9.84E-02 | 0.23 | 1.46 |
| HILIC_POS_227_17522a1_084 | | 227.1752 | 1.08 | | HILIC | POS | | | 386453 | 293072 | 322512 | 272408 | 9.84E-02 | -0.23 | 1.14 |
| HILIC_POS_146_08125a4_867 | 4-Acetamidobutanoic acid | 146.0813 | 4.87 | | HILIC | POS | 146.0821(100) | 1 | 725659 | 1008979 | 533856 | 679317 | 9.86E-02 | -0.23 | 0.92 |
| HILIC_POS_380_30112a0_995 | | 380.3011 | 1.00 | | HILIC | POS | | | 259035 | 93766.9 | 239151 | 82392.7 | 9.87E-02 | -0.23 | 0.92 |
| RP_POS_136_98961a15_536 | | 136.9896 | 15.54 | | RP | POS | | | 7381237 | 2470078 | 6805406 | 2601968 | 9.87E-02 | -0.23 | 0.98 |
| RP_POS_563_4032a10_558 | | 563.4032 | 10.56 | | RP | POS | | | 538191 | 298175 | 615022 | 370238 | 9.88E-02 | 0.23 | 1.28 |
| RP_POS_791_52838a12_358 | | 791.5284 | 12.36 | | RP | POS | | | 1696600 | 463935 | 1797497 | 429540 | 9.88E-02 | 0.23 | 1.04 |
| HILIC_NEG_117_91898a4_855 | | 117.9190 | 4.86 | | HILIC | NEG | | | 1422070 | 584836 | 1292794 | 558933 | 9.88E-02 | -0.23 | 1.09 |
| HILIC_NEG_262_99289a0_935 | | 262.9929 | 0.94 | | HILIC | NEG | | | 443482 | 48184.3 | 455887 | 59814.5 | 9.90E-02 | 0.23 | 1.11 |
| HILIC_NEG_276_99136a2_266 | | 276.9914 | 2.27 | | HILIC | NEG | | | 897261 | 294599 | 960845 | 268900 | 9.90E-02 | 0.23 | 0.99 |
| RP_POS_426_32156a9_287 | Cholic Acid | 426.3216 | 9.29 | | RP | POS | 355.2613(100), 159.1164(12), 145.1025(11) | 1 | 969906 | 1628815 | 1671808 | 3927928 | 9.91E-02 | 0.25 | 0.98 |
| RP_NEG_285_70822a15_495 | | 285.7082 | 15.50 | | RP | NEG | | | 3790281 | 136031 | 3759332 | 137523 | 9.92E-02 | -0.23 | 1.05 |
| RP_NEG_254_99461a3_677 | | 254.9946 | 3.68 | | RP | NEG | | | 323522 | 239580 | 274716 | 193772 | 9.94E-02 | -0.23 | 0.98 |
| RP_NEG_552_36682a10_872 | | 552.3668 | 10.87 | | RP | NEG | | | 4120155 | 1039686 | 4364656 | 1116317 | 9.95E-02 | 0.23 | 1.09 |
| RP_NEG_215_16539a9_119 | | 215.1654 | 9.12 | | RP | NEG | | | 646590 | 437103 | 755144 | 514145 | 9.96E-02 | 0.23 | 1.25 |
| RP_POS_386_1882a10_339 | | 386.1882 | 10.34 | | RP | POS | | | 239486 | 471730 | 376951 | 702822 | 9.96E-02 | 0.23 | 1.16 |
| RP_NEG_314_71124a15_496 | | 314.7112 | 15.50 | | RP | NEG | | | 141335 | 37824.8 | 132653 | 38933.9 | 9.96E-02 | -0.23 | 0.98 |
| RP_POS_770_50995a13_053 | | 770.5100 | 13.05 | | RP | POS | | | 1204165 | 970169 | 1429054 | 1017134 | 9.97E-02 | 0.23 | 1.19 |
| RP_POS_544_05292a10_45 | | 544.0529 | 10.45 | | RP | POS | | | 670874 | 424017 | 768116 | 436286 | 9.98E-02 | 0.23 | 1.01 |
| RP_POS_157_12224a7_323 | | 157.1222 | 7.32 | | RP | POS | | | 782720 | 116922 | 756240 | 117703 | 9.99E-02 | -0.23 | 0.97 |
| RP_POS_682_30847a11_482 | | 682.3085 | 11.48 | | RP | POS | | | 110356 | 68242.8 | 128392 | 88638 | 1.00E-01 | 0.23 | 0.99 |
| HILIC_POS_84_08146a3_652 | | 84.0815 | 3.65 | | HILIC | POS | | | 809836 | 2922182 | 324449 | 1136554 | 1.00E-01 | -0.24 | 0.89 |
| RP_POS_441_23871a10_638 | | 441.2387 | 10.64 | | RP | POS | | | 528844 | 731610 | 905555 | 2168710 | 1.01E-01 | 0.26 | 1.24 |
| HILIC_NEG_315_04178a6_427 | | 315.0418 | 6.43 | | HILIC | NEG | | | 660752 | 379782 | 765468 | 526120 | 1.01E-01 | 0.23 | 0.98 |
| RP_NEG_556_31873a10_61 | | 556.3187 | 10.61 | | RP | NEG | | | 4509284 | 672438 | 4667049 | 723113 | 1.01E-01 | 0.23 | 1.05 |
| RP_POS_449_28741a11_047 | | 449.2874 | 11.05 | | RP | POS | | | 757737 | 366290 | 852684 | 463221 | 1.01E-01 | 0.23 | 1.42 |
| RP_POS_282_2785a11_586 | | 282.2785 | 11.59 | | RP | POS | 282.2813(100), 69.0709(70) | | 3228791 | 1091826 | 3006441 | 889751 | 1.01E-01 | -0.22 | 1.06 |
| HILIC_POS_495_34213a3_955 | | 495.3421 | 3.96 | | HILIC | POS | | | 43446 | 18543.4 | 39711.5 | 14776.4 | 1.01E-01 | -0.22 | 1.25 |
| RP_NEG_311_25955a10_392 | | 311.2596 | 10.39 | | RP | NEG | | | 266572 | 42812.7 | 256715 | 44643.1 | 1.01E-01 | -0.23 | 1.39 |
| RP_POS_239_08894a5_996 | | 239.0889 | 6.00 | | RP | POS | | | 514155 | 178101 | 569139 | 289569 | 1.02E-01 | 0.24 | 1.32 |
| RP_POS_337_1619a8_835 | | 337.1619 | 8.84 | | RP | POS | | | 980418 | 532913 | 1149641 | 897162 | 1.02E-01 | 0.24 | 1.42 |
| HILIC_POS_175_10777a3_046 | | 175.1078 | 3.05 | | HILIC | POS | | | 191898 | 224776 | 256404 | 330603 | 1.02E-01 | 0.23 | 1.08 |
| RP_POS_368_27933a8_222 | | 368.2793 | 8.22 | | RP | POS | | | 1753376 | 1186425 | 2080339 | 1653825 | 1.02E-01 | 0.23 | 1.08 |
| RP_POS_460_14984a8_016 | | 460.1498 | 8.02 | | RP | POS | | | 398218 | 42668.5 | 409091 | 52855.9 | 1.02E-01 | 0.23 | 1.13 |
| RP_POS_460_36334a10_593 | | 460.3633 | 10.59 | | RP | POS | | | 324247 | 263323 | 402582 | 407645 | 1.02E-01 | 0.23 | 1.3 |
| RP_POS_741_51532a10_054 | | 741.5153 | 10.05 | | RP | POS | | | 1273191 | 593868 | 1431762 | 788321 | 1.02E-01 | 0.23 | 1.16 |
| RP_POS_522_35425a10_608 | LysoPC 18:1 (18:1/0:0) | 522.3543 | 10.61 | | RP | POS | 184.0742(100), 522.355(20) | 1 | 9.7E+08 | 2.2E+08 | 1E+09 | 2.4E+08 | 1.02E-01 | 0.22 | 1.05 |
| RP_POS_187_97118a15_499 | | 187.9712 | 15.50 | | RP | POS | | | 372148 | 36612.8 | 364143 | 34838.5 | 1.02E-01 | -0.22 | 1.18 |
| RP_POS_728_52209a12_259 | | 728.5221 | 12.26 | | RP | POS | | | 2965925 | 1943892 | 2541908 | 1846050 | 1.02E-01 | -0.22 | 0.95 |
| RP_NEG_329_26917a11_592 | | 329.2692 | 11.59 | | RP | NEG | | | 1438278 | 416901 | 1341087 | 446939 | 1.02E-01 | -0.23 | 1.45 |
| RP_POS_255_21037a10_844 | | 255.2104 | 10.84 | | RP | POS | | | 1327350 | 264524 | 1266578 | 275536 | 1.02E-01 | -0.23 | 1.11 |
| RP_NEG_150_00189a9_82 | | 150.0019 | 9.82 | | RP | NEG | | | 250964 | 215565 | 302949 | 245658 | 1.03E-01 | 0.23 | 1.12 |
| RP_POS_193_15907a7_044 | | 193.1591 | 7.04 | | RP | POS | | | 5108037 | 370073 | 5200408 | 446850 | 1.03E-01 | 0.23 | 1.06 |
| RP_POS_365_2771a8_963 | | 365.2771 | 8.96 | | RP | POS | | | 329412 | 65064.6 | 347509 | 92226.4 | 1.03E-01 | 0.23 | 1.14 |
| RP_POS_653_289a11_314 | | 653.2890 | 11.31 | | RP | POS | | | 147308 | 90378.7 | 171181 | 118529 | 1.03E-01 | 0.23 | 1.02 |
| HILIC_NEG_100_9335a4_089 | | 100.9335 | 4.09 | | HILIC | NEG | | | 2.5E+07 | 4896692 | 2.4E+07 | 3914995 | 1.03E-01 | -0.22 | 1.36 |
| HILIC_NEG_251_86726a7_735 | | 251.8673 | 7.74 | | HILIC | NEG | | | 786568 | 82840.9 | 768644 | 77612.5 | 1.03E-01 | -0.22 | 0.97 |
| RP_POS_181_00832a0_548 | | 181.0083 | 0.55 | | RP | POS | | | 605778 | 226460 | 553267 | 241552 | 1.03E-01 | -0.22 | 1.09 |

**Supplemantary table 1**: The non-targeted metabolomics results ordered by the VIP value (p value < 0.0003 are shaded dark green, p value < 0.05 are shaded light green, d values below 0 are blue and above 0 are red)

| | | | | | | | | | Cases | | Controls | | Welch test | Cohen's | |
|---|---|---|---|---|---|---|---|---|---|---|---|---|---|---|---|
| Feature_ID | Metabolite_name | Average_Mz | Average_RT_min_ | HMDB ID | RP_HILIC | POS_NEG | Cureted_MSMS | ID_level | Mean | SD | Mean2 | SD3 | p | d | VIP |
| RP_POS_684_96313a10_249 | | 684.9631 | 10.25 | | RP | POS | | | 1608027 | 824735 | 1445647 | 632429 | 1.03E-01 | -0.22 | 1.11 |
| HILIC_POS_640_31854a1_313 | | 640.3185 | 1.31 | | HILIC | POS | | | 273237 | 59881.4 | 262312 | 37092.7 | 1.03E-01 | -0.23 | 0.97 |
| RP_POS_317_24747a8_96 | | 317.2475 | 8.96 | | RP | POS | | | 2681536 | 859038 | 2432495 | 1289875 | 1.03E-01 | -0.23 | 1.01 |
| HILIC_POS_338_04959a3_142 | | 338.0496 | 3.14 | | HILIC | POS | | | 1247936 | 245696 | 1286877 | 71472.2 | 1.04E-01 | 0.25 | 0.91 |
| RP_NEG_330_74994a15_44 | | 330.7499 | 15.44 | | RP | NEG | | | 2098635 | 239540 | 2148675 | 210522 | 1.04E-01 | 0.22 | 1.04 |
| RP_NEG_384_93668a12_945 | | 384.9367 | 12.95 | | RP | NEG | | | 914088 | 461694 | 1015494 | 450058 | 1.04E-01 | 0.22 | 0.92 |
| HILIC_POS_358_19653a5_917 | | 358.1965 | 5.92 | | HILIC | POS | | | 145613 | 34741.7 | 138539 | 28899.8 | 1.04E-01 | -0.22 | 0.99 |
| RP_POS_211_09444a5_973 | | 211.0944 | 5.97 | | RP | POS | | | 847557 | 139803 | 817146 | 133697 | 1.04E-01 | -0.22 | 1.07 |
| HILIC_POS_150_01381a6_196 | | 150.0138 | 6.20 | | HILIC | POS | | | 189737 | 32472.6 | 197712 | 38405.6 | 1.05E-01 | 0.23 | 1.08 |
| RP_POS_554_71973a9_865 | | 554.7197 | 9.87 | | RP | POS | | | 216598 | 109370 | 247753 | 161668 | 1.05E-01 | 0.23 | 1.04 |
| RP_POS_656_78735a10_227 | | 656.7874 | 10.23 | | RP | POS | | | 151245 | 84943.1 | 174087 | 115358 | 1.05E-01 | 0.23 | 1 |
| RP_POS_742_51392a11_301 | | 742.5139 | 11.30 | | RP | POS | | | 144296 | 67042.5 | 161361 | 83941.1 | 1.05E-01 | 0.23 | 1 |
| RP_POS_221_18996a9_629 | | 221.1900 | 9.63 | | RP | POS | 221.1903(100) | | 4328388 | 275545 | 4382402 | 211437 | 1.05E-01 | 0.22 | 1.08 |
| RP_POS_650_45038a11_403 | | 650.4504 | 11.40 | | RP | POS | | | 64917.6 | 41135 | 75047.8 | 48987.1 | 1.05E-01 | 0.22 | 1.03 |
| RP_NEG_257_17566a8_862 | | 257.1757 | 8.86 | | RP | NEG | | | 1460561 | 201780 | 1420141 | 162591 | 1.05E-01 | -0.22 | 1.01 |
| RP_POS_677_55933a12_487 | | 677.5593 | 12.49 | | RP | POS | | | 1125252 | 581872 | 1005167 | 501596 | 1.05E-01 | -0.22 | 1.11 |
| RP_POS_247_24228a10_855 | | 247.2423 | 10.86 | | RP | POS | | | 1752841 | 939864 | 2073699 | 1763012 | 1.06E-01 | 0.24 | 1.14 |
| RP_POS_347_22433a14_353 | | 347.2243 | 14.35 | | RP | POS | | | 557917 | 453361 | 707542 | 813661 | 1.06E-01 | 0.24 | 1.16 |
| RP_POS_411_76981a13_458 | | 411.7698 | 13.46 | | RP | POS | | | 3168861 | 2422254 | 3893355 | 3829575 | 1.06E-01 | 0.23 | 1.33 |
| RP_POS_593_84424a11_464 | | 593.8442 | 11.46 | | RP | POS | | | 109942 | 44513.6 | 122835 | 67418.1 | 1.06E-01 | 0.23 | 1.16 |
| RP_POS_283_26309a11_239 | | 283.2631 | 11.24 | | RP | POS | | | 2121768 | 1411000 | 2458671 | 1606513 | 1.06E-01 | 0.22 | 1.21 |
| RP_POS_382_18344a9_821 | | 382.1834 | 9.82 | | RP | POS | | | 2519822 | 2168661 | 3040622 | 2490248 | 1.06E-01 | 0.22 | 1.12 |
| HILIC_NEG_612_33087a1_125 | | 612.3309 | 1.13 | | HILIC | NEG | | | 147095 | 86537 | 129774 | 70097.1 | 1.06E-01 | -0.22 | 1.09 |
| HILIC_POS_387_21414a4_164 | | 387.2141 | 4.16 | | HILIC | POS | | | 1072051 | 213618 | 1028992 | 176590 | 1.06E-01 | -0.22 | 0.96 |
| RP_POS_379_38434a8_846 | | 379.3843 | 8.85 | | RP | POS | | | 298283 | 149640 | 269897 | 105973 | 1.06E-01 | -0.22 | 0.98 |
| RP_POS_581_40381a11_308 | | 581.4038 | 11.31 | | RP | POS | | | 209756 | 174270 | 267660 | 317213 | 1.07E-01 | 0.24 | 1.02 |
| RP_POS_207_13797a8_362 | | 207.1380 | 8.36 | | RP | POS | | | 1398427 | 153120 | 1437524 | 194357 | 1.07E-01 | 0.23 | 1 |
| RP_NEG_373_23724a11_112 | | 373.2372 | 11.11 | | RP | NEG | | | 109566 | 59298.5 | 122927 | 61342.8 | 1.07E-01 | 0.22 | 1.47 |
| HILIC_POS_316_24829a0_707 | AC 10:0 (Decanoylcarnitine) | 316.2483 | 0.71 | | HILIC | POS | 85.0292(100), 316.2497(45) | 1 | 9432806 | 7823044 | 1.3E+07 | 1.8E+07 | 1.07E-01 | 0.24 | 1.06 |
| HILIC_POS_146_08124a0_791 | Allysine (not validated) | 146.0812 | 0.79 | | HILIC | POS | 146.082(100) | 2 | 1.4E+07 | 2.5E+07 | 9454928 | 1.8E+07 | 1.07E-01 | -0.22 | 0.9 |
| HILIC_NEG_241_0105a0_934 | | 241.0105 | 0.93 | | HILIC | NEG | | | 738482 | 74386.2 | 721871 | 75887.4 | 1.07E-01 | -0.22 | 0.99 |
| HILIC_POS_338_34183a3_119 | | 338.3418 | 3.12 | | HILIC | POS | | | 337797 | 35074.2 | 329586 | 38710.4 | 1.07E-01 | -0.22 | 1.05 |
| RP_NEG_242_92719a0_572 | | 242.9272 | 0.57 | | RP | NEG | | | 1024492 | 221530 | 976270 | 215106 | 1.07E-01 | -0.22 | 1.06 |
| RP_NEG_456_18292a11_359 | | 456.1829 | 11.36 | | RP | NEG | | | 346403 | 37533.8 | 337757 | 40359 | 1.07E-01 | -0.22 | 1.29 |
| RP_POS_401_25537a12_114 | | 401.2554 | 12.11 | | RP | POS | | | 515249 | 151823 | 481559 | 152924 | 1.07E-01 | -0.22 | 1.44 |
| RP_POS_686_28369a11_615 | | 686.2837 | 11.62 | | RP | POS | | | 88432.6 | 50874.8 | 102479 | 72714.4 | 1.08E-01 | 0.23 | 1.12 |
| RP_POS_1041_68738a10_493 | | 1041.6874 | 10.49 | | RP | POS | | | 1.3E+07 | 4236362 | 1.4E+07 | 4593209 | 1.08E-01 | 0.22 | 1.11 |
| RP_POS_364_34161a10_878 | | 364.3416 | 10.88 | | RP | POS | | | 2404536 | 313059 | 2471461 | 295787 | 1.08E-01 | 0.22 | 1.21 |
| RP_POS_480_34744a10_387 | | 480.3474 | 10.39 | | RP | POS | | | 529860 | 409647 | 627268 | 468135 | 1.08E-01 | 0.22 | 1.02 |
| RP_POS_548_5022a14_534 | | 548.5022 | 14.53 | | RP | POS | | | 4583016 | 1976280 | 5077102 | 2441314 | 1.08E-01 | 0.22 | 1.08 |
| RP_POS_597_75189a10_265 | | 597.7519 | 10.27 | | RP | POS | | | 668370 | 353396 | 758878 | 452500 | 1.08E-01 | 0.22 | 1.07 |
| RP_POS_522_35529a10_484 | LysoPC 18:1 (0:0/18:1) | 522.3553 | 10.48 | | RP | POS | 184.0746(100) | 1 | 8.1E+07 | 1.6E+07 | 8.5E+07 | 1.8E+07 | 1.08E-01 | 0.22 | 1.01 |
| RP_NEG_516_29443a9_374 | | 516.2944 | 9.37 | | RP | NEG | | | 805267 | 959345 | 616833 | 755227 | 1.08E-01 | -0.22 | 0.95 |
| RP_POS_281_91125a15_544 | | 281.9113 | 15.54 | | RP | POS | | | 1144570 | 563041 | 1029915 | 479870 | 1.08E-01 | -0.22 | 1.21 |
| RP_POS_532_39105a11_463 | | 532.3911 | 11.46 | | RP | POS | | | 837314 | 270901 | 781614 | 236458 | 1.08E-01 | -0.22 | 1.24 |
| RP_POS_538_38666a11_138 | | 538.3867 | 11.14 | | RP | POS | | | 2769834 | 862542 | 2572957 | 918907 | 1.08E-01 | -0.22 | 1.11 |
| RP_POS_795_57465a13_476 | | 795.5747 | 13.48 | | RP | POS | | | 9396061 | 7688883 | 1.1E+07 | 1E+07 | 1.09E-01 | 0.23 | 1 |
| HILIC_POS_106_08676a1_051 | | 106.0868 | 1.05 | | HILIC | POS | 88.0762(100), 106.0874(90) | | 1.1E+07 | 5716944 | 1.3E+07 | 6782325 | 1.09E-01 | 0.22 | 1.2 |
| RP_NEG_452_92371a14_35 | | 452.9237 | 14.35 | | RP | NEG | | | 2278141 | 1264137 | 2573623 | 1404913 | 1.09E-01 | 0.22 | 0.96 |
| RP_NEG_459_2735a9_9 | | 459.2735 | 9.90 | | RP | NEG | | | 535582 | 432895 | 639891 | 507256 | 1.09E-01 | 0.22 | 0.93 |
| RP_POS_192_97949a0_562 | | 192.9795 | 0.56 | | RP | POS | | | 1211161 | 572424 | 1330674 | 518746 | 1.09E-01 | 0.22 | 1.16 |
| RP_POS_265_25247a11_239 | | 265.2525 | 11.24 | | RP | POS | | | 1131743 | 725781 | 1305795 | 841695 | 1.09E-01 | 0.22 | 1.17 |
| RP_POS_296_87381a15_413 | | 296.8738 | 15.41 | | RP | POS | | | 3607228 | 1013093 | 3849455 | 1173646 | 1.09E-01 | 0.22 | 1.3 |
| RP_POS_419_75613a13_467 | | 419.7561 | 13.47 | | RP | POS | | | 2332687 | 1939775 | 2815855 | 2392264 | 1.09E-01 | 0.22 | 1.12 |
| RP_POS_551_31262a10_297 | | 551.3126 | 10.30 | | RP | POS | | | 332988 | 182486 | 379430 | 232873 | 1.09E-01 | 0.22 | 1.35 |
| HILIC_NEG_112_98565a0_934 | | 112.9857 | 0.93 | | HILIC | NEG | 68.995(100), 112.9858(3) | | 2.2E+07 | 2585669 | 2.1E+07 | 2269593 | 1.09E-01 | -0.22 | 0.95 |
| RP_NEG_212_97948a15_486 | | 212.9795 | 15.49 | | RP | NEG | | | 536418 | 92967.5 | 512903 | 117265 | 1.09E-01 | -0.22 | 0.92 |
| RP_NEG_720_49841a12_926 | | 720.4984 | 12.93 | | RP | NEG | | | 523211 | 331541 | 460155 | 243180 | 1.09E-01 | -0.22 | 1.1 |
| RP_POS_171_09938a4_075 | | 171.0994 | 4.08 | | RP | POS | | | 1579943 | 827017 | 1427073 | 562556 | 1.09E-01 | -0.22 | 1.02 |
| RP_POS_276_65234a10_487 | | 276.6523 | 10.49 | | RP | POS | | | 2652394 | 1414092 | 2382828 | 1041123 | 1.09E-01 | -0.22 | 1.26 |
| RP_POS_373_22534a11_595 | | 373.2253 | 11.60 | | RP | POS | | | 5218821 | 853611 | 5040053 | 779773 | 1.09E-01 | -0.22 | 1.18 |
| RP_POS_424_28247a11_024 | | 424.2825 | 11.02 | | RP | POS | | | 613012 | 909470 | 449989 | 560882 | 1.09E-01 | -0.22 | 0.95 |
| RP_POS_424_30347a9_861 | | 424.3035 | 9.86 | | RP | POS | | | 557642 | 278217 | 496233 | 280934 | 1.09E-01 | -0.22 | 1.15 |
| RP_POS_464_31625a10_071 | | 464.3163 | 10.07 | | RP | POS | | | 231580 | 180869 | 197418 | 130313 | 1.09E-01 | -0.22 | 1.08 |
| RP_POS_149_01205a10_508 | | 149.0121 | 10.51 | | RP | POS | | | 714122 | 1656001 | 461561 | 323786 | 1.09E-01 | -0.26 | 1 |
| RP_POS_567_38483a14_979 | | 567.3848 | 14.98 | | RP | POS | | | 688177 | 559652 | 908733 | 1264812 | 1.10E-01 | 0.24 | 1.21 |
| RP_POS_352_24564a13_293 | | 352.2456 | 13.29 | | RP | POS | | | 542190 | 405390 | 657766 | 610337 | 1.10E-01 | 0.23 | 1.23 |
| HILIC_NEG_136_89172a4_753 | | 136.8917 | 4.75 | | HILIC | NEG | | | 6395989 | 3314208 | 7127767 | 3357418 | 1.10E-01 | 0.22 | 0.94 |
| HILIC_POS_130_15915a0_57 | | 130.1592 | 0.57 | | HILIC | POS | | | 1.1E+07 | 1456477 | 1.1E+07 | 1524331 | 1.10E-01 | 0.22 | 1.07 |
| HILIC_POS_351_01254a7_483 | | 351.0125 | 7.48 | | HILIC | POS | | | 720746 | 37256.4 | 712832 | 35200.5 | 1.10E-01 | -0.22 | 1.02 |
| RP_POS_342_24863a5_529 | | 342.2486 | 5.53 | | RP | POS | | | 769433 | 363073 | 695165 | 317139 | 1.10E-01 | -0.22 | 1.11 |
| RP_POS_470_68057a10_553 | | 470.6806 | 10.55 | | RP | POS | | | 182849 | 108077 | 161498 | 87191.3 | 1.10E-01 | -0.22 | 0.92 |
| RP_POS_586_30609a6_406 | | 586.3061 | 6.41 | | RP | POS | | | 369267 | 52232 | 358470 | 46723.3 | 1.10E-01 | -0.22 | 1.02 |
| RP_POS_173_09618a9_069 | | 173.0962 | 9.07 | | RP | POS | | | 726734 | 377870 | 893181 | 971017 | 1.11E-01 | 0.25 | 1.39 |
| RP_NEG_134_89468a11_593 | | 134.8947 | 11.59 | | RP | NEG | | | 1188835 | 376407 | 1265206 | 325446 | 1.11E-01 | 0.22 | 0.93 |
| RP_NEG_327_18149a8_99 | | 327.1815 | 8.99 | | RP | NEG | | | 213185 | 130507 | 246445 | 168545 | 1.11E-01 | 0.22 | 1.41 |
| RP_POS_317_19568a8_796 | | 317.1957 | 8.80 | | RP | POS | | | 1255129 | 815673 | 1447390 | 930952 | 1.11E-01 | 0.22 | 1.35 |
| RP_POS_379_16916a1_422 | | 379.1692 | 1.42 | | RP | POS | | | 1122979 | 655412 | 1258124 | 586296 | 1.11E-01 | 0.22 | 1.11 |
| RP_POS_450_23349a7_243 | | 450.2335 | 7.24 | | RP | POS | | | 820743 | 70151.4 | 837733 | 83608.4 | 1.11E-01 | 0.22 | 1.07 |
| RP_POS_461_66083a9_745 | | 461.6608 | 9.75 | | RP | POS | | | 171689 | 107954 | 195491 | 109705 | 1.11E-01 | 0.22 | 0.98 |
| RP_NEG_249_15028a10_204 | | 249.1503 | 10.20 | | RP | NEG | 249.1483(100), 205.1595(25) | | 855559 | 40307.6 | 846720 | 40630.2 | 1.11E-01 | -0.22 | 1.01 |
| RP_POS_236_07188a13_609 | | 236.0719 | 13.61 | | RP | POS | | | 1.1E+08 | 7.3E+07 | 9.4E+07 | 5.2E+07 | 1.11E-01 | -0.22 | 1.15 |
| RP_POS_499_21786a5_456 | | 499.2179 | 5.46 | | RP | POS | | | 571069 | 224687 | 523463 | 211328 | 1.11E-01 | -0.22 | 0.92 |
| RP_POS_201_09108a9_069 | | 201.0911 | 9.07 | | RP | POS | | | 565267 | 401213 | 733015 | 975168 | 1.12E-01 | 0.24 | 1.4 |
| HILIC_NEG_199_80559a10_94 | | 199.8056 | 10.94 | | HILIC | NEG | | | 8938237 | 4681972 | 1E+07 | 5211026 | 1.12E-01 | 0.22 | 1 |
| HILIC_POS_104_08235a2_45 | | 104.0824 | 2.45 | | HILIC | POS | | | 717114 | 359002 | 795528 | 359635 | 1.12E-01 | 0.22 | 1.1 |
| RP_POS_416_30035a10_39 | | 416.3004 | 10.39 | | RP | POS | | | 1262089 | 598655 | 1405449 | 704519 | 1.12E-01 | 0.22 | 1.32 |
| RP_POS_463_30273a11_157 | | 463.3027 | 11.16 | | RP | POS | | | 1010781 | 492039 | 1131576 | 601029 | 1.12E-01 | 0.22 | 1.32 |
| RP_POS_807_57318a13_927 | | 807.5732 | 13.93 | | RP | POS | | | 5359874 | 3805155 | 6225879 | 4103692 | 1.12E-01 | 0.22 | 1.13 |
| HILIC_NEG_141_924a0_592 | | 141.9240 | 0.59 | | HILIC | NEG | | | 245117 | 49106 | 233930 | 53251.7 | 1.12E-01 | -0.22 | 0.94 |
| HILIC_NEG_236_81679a5_524 | | 236.8168 | 5.52 | | HILIC | NEG | | | 351850 | 156627 | 320632 | 131249 | 1.12E-01 | -0.22 | 1.07 |
| HILIC_NEG_696_77448a6_365 | | 696.7745 | 6.37 | | HILIC | NEG | | | 166099 | 85296.3 | 148014 | 81073 | 1.12E-01 | -0.22 | 1.33 |
| HILIC_POS_356_2424a3_338 | | 356.2424 | 3.34 | | HILIC | POS | | | 46766.7 | 26563.3 | 41548.6 | 21499.4 | 1.12E-01 | -0.22 | 1.06 |
| RP_NEG_140_00243a15_485 | | 140.0024 | 15.49 | | RP | NEG | | | 111655 | 26209.6 | 117777 | 29760 | 1.13E-01 | 0.22 | 1.02 |
| RP_POS_254_12482a0_675 | | 254.1248 | 0.68 | | RP | POS | | | 1143161 | 471239 | 1251148 | 518162 | 1.13E-01 | 0.22 | 0.93 |
| RP_POS_363_2294a10_651 | | 363.2294 | 10.65 | | RP | POS | | | 1.1E+07 | 6180418 | 1.3E+07 | 5922485 | 1.13E-01 | 0.22 | 1.26 |
| RP_POS_499_32648a10_774 | | 499.3265 | 10.77 | | RP | POS | | | 832972 | 467081 | 956560 | 642002 | 1.13E-01 | 0.22 | 1.41 |
| RP_POS_521_30853a10_772 | | 521.3085 | 10.77 | | RP | POS | | | 2085626 | 1071474 | 2373533 | 1506375 | 1.13E-01 | 0.22 | 1.41 |
| HILIC_POS_131_04926a3_905 | trans-Cinnamic acid | 131.0493 | 3.91 | | HILIC | POS | 103.0549(100) | 1 | 1100329 | 1463196 | 799283 | 1318277 | 1.13E-01 | -0.22 | 1.17 |
| HILIC_NEG_326_98892a0_95 | | 326.9889 | 0.95 | | HILIC | NEG | 242.9875(100), 326.988(10) | | 956068 | 105657 | 933975 | 97912.8 | 1.13E-01 | -0.22 | 0.97 |
| HILIC_POS_249_10805a7_751 | | 249.1081 | 7.75 | | HILIC | POS | | | 285549 | 33064.2 | 277463 | 40336.1 | 1.13E-01 | -0.22 | 1.15 |
| HILIC_POS_384_34637a1_354 | | 384.3464 | 1.35 | | HILIC | POS | | | 122730 | 59390.4 | 111397 | 45101.8 | 1.13E-01 | -0.22 | 1.02 |
| RP_NEG_612_26105a10_496 | | 612.2611 | 10.50 | | RP | NEG | | | 4885937 | 977698 | 4651810 | 1151592 | 1.13E-01 | -0.22 | 0.91 |
| RP_POS_458_8114a9_612 | | 458.8114 | 9.61 | | RP | POS | | | 1428896 | 1072080 | 1242410 | 622235 | 1.13E-01 | -0.22 | 0.91 |
| HILIC_POS_167_04849a5_333 | | 167.0485 | 5.33 | | HILIC | POS | | | 402810 | 150183 | 434583 | 143554 | 1.14E-01 | 0.22 | 1.07 |
| RP_POS_197_15388a7_527 | | 197.1539 | 7.53 | | RP | POS | | | 1929932 | 294728 | 2001057 | 354526 | 1.14E-01 | 0.22 | 1.08 |
| RP_POS_687_14996a11_635 | | 687.1500 | 11.64 | | RP | POS | | | 125784 | 63766.2 | 139899 | 66080.2 | 1.14E-01 | 0.22 | 0.91 |
| HILIC_NEG_204_9894a0_933 | | 204.9894 | 0.93 | | HILIC | NEG | | | 291663 | 36229.7 | 284115 | 33653.9 | 1.14E-01 | -0.22 | 0.96 |
| RP_POS_284_16971a3_344 | | 284.1697 | 3.34 | | RP | POS | | | 336415 | 78479.5 | 318865 | 83065 | 1.14E-01 | -0.22 | 0.99 |
| RP_POS_519_86676a10_001 | | 519.8668 | 10.00 | | RP | POS | | | 784983 | 487121 | 693000 | 363509 | 1.14E-01 | -0.22 | 0.98 |
| RP_NEG_239_16599a10_082 | | 239.1660 | 10.08 | | RP | NEG | | | 241279 | 122006 | 268953 | 132867 | 1.15E-01 | 0.22 | 1.01 |
| RP_NEG_329_95459a6_808 | | 329.9546 | 6.81 | | RP | NEG | | | 613238 | 99636.6 | 633650 | 89964.4 | 1.15E-01 | 0.22 | 1.01 |
| RP_NEG_398_17984a9_819 | | 398.1798 | 9.82 | | RP | NEG | | | 704035 | 610439 | 846028 | 694224 | 1.15E-01 | 0.22 | 1.1 |
| RP_NEG_624_33972a8_777 | | 624.3397 | 8.78 | | RP | NEG | | | 964379 | 1027362 | 1203965 | 1171518 | 1.15E-01 | 0.22 | 0.9 |
| HILIC_POS_170_11755a1_361 | | 170.1176 | 1.36 | | HILIC | POS | | | 510961 | 274637 | 458219 | 214346 | 1.15E-01 | -0.22 | 1.09 |
| HILIC_POS_189_07356a4_94 | | 189.0736 | 4.94 | | HILIC | POS | | | 3058507 | 1131227 | 2808995 | 1175526 | 1.15E-01 | -0.22 | 1.03 |
| RP_NEG_420_60336a15_484 | | 420.6034 | 15.48 | | RP | NEG | | | 481619 | 74054.5 | 467118 | 60610.4 | 1.15E-01 | -0.22 | 0.98 |
| RP_POS_461_83643a10_064 | | 461.8364 | 10.06 | | RP | POS | | | 2323708 | 1150013 | 2090462 | 1013751 | 1.15E-01 | -0.22 | 0.97 |
| RP_POS_552_88354a10_375 | | 552.8835 | 10.38 | | RP | POS | | | 1816271 | 968456 | 1624456 | 810958 | 1.15E-01 | -0.22 | 1.15 |
| RP_POS_568_34003a10_284 | | 568.3400 | 10.28 | | RP | POS | | | 7.4E+07 | 2.4E+07 | 6.9E+07 | 2.3E+07 | 1.15E-01 | -0.22 | 1.06 |
| RP_POS_297_20355a9_73 | | 297.2036 | 9.73 | | RP | POS | | | 923969 | 342395 | 1215225 | 1799586 | 1.16E-01 | 0.27 | 1.38 |
| RP_NEG_283_26389a11_592 | Methyl Heptadecanoic acid | 283.2639 | 11.59 | | RP | NEG | 283.2645(100),239.1939(1) | 2 | 5.1E+07 | 1.7E+07 | 4.7E+07 | 1.9E+07 | 1.16E-01 | -0.22 | 1.45 |
| HILIC_POS_164_02945a5_8 | | 164.0295 | 5.80 | | HILIC | POS | | | 1051489 | 172293 | 1090236 | 185675 | 1.16E-01 | 0.22 | 0.93 |
| RP_POS_389_26843a9_006 | | 389.2684 | 9.01 | | RP | POS | | | 860527 | 356671 | 939334 | 373886 | 1.16E-01 | 0.22 | 0.9 |
| RP_POS_427_30533a11_047 | | 427.3053 | 11.05 | | RP | POS | | | 677974 | 304849 | 754744 | 394864 | 1.16E-01 | 0.22 | 1.4 |
| RP_POS_499_11188a10_006 | | 499.1119 | 10.01 | | RP | POS | | | 650617 | 569253 | 776088 | 592350 | 1.16E-01 | 0.22 | 1.16 |
| RP_NEG_243_19675a9_884 | beta-Hydroxymyristic acid | 243.1968 | 9.88 | | RP | NEG | 59.0132(100), 243.1956(35) | 2 | 765765 | 273718 | 827448 | 295607 | 1.16E-01 | 0.22 | 1.15 |
| HILIC_POS_189_13464a6_845 | Homoarginine | 189.1346 | 6.85 | | HILIC | POS | 189.1346(100), 144.1141(60) | 2 | 3183784 | 1627988 | 2861513 | 1374746 | 1.16E-01 | -0.21 | 0.89 |
| HILIC_POS_59_06139a0_931 | | 59.0614 | 0.93 | | HILIC | POS | | | 1.1E+07 | 4624131 | 1E+07 | 4218230 | 1.16E-01 | -0.21 | 1.04 |
| RP_NEG_139_90825a0_622 | | 139.9083 | 0.62 | | RP | NEG | | | 567060 | 142855 | 538334 | 124605 | 1.16E-01 | -0.21 | 0.9 |
| RP_POS_321_23993a10_236 | | 321.2399 | 10.24 | | RP | POS | | | 7494600 | 1022190 | 7279527 | 976533 | 1.16E-01 | -0.22 | 1.31 |
| RP_POS_380_33755a10_522 | | 380.3376 | 10.52 | | RP | POS | | | 2266811 | 442189 | 2177215 | 390968 | 1.16E-01 | -0.22 | 1.13 |
| RP_POS_545_40173a10_874 | | 545.4017 | 10.87 | | RP | POS | | | 1430123 | 409110 | 1344961 | 381942 | 1.16E-01 | -0.22 | 1.2 |
| RP_POS_560_36963a11_14 | | 560.3696 | 11.14 | | RP | POS | | | 1111281 | 324277 | 1037959 | 353457 | 1.16E-01 | -0.22 | 1.1 |
| RP_POS_610_4306a11_635 | | 610.4306 | 11.64 | | RP | POS | | | 649054 | 539432 | 556242 | 310920 | 1.16E-01 | -0.22 | 0.92 |
| RP_POS_764_52228a12_942 | | 764.5223 | 12.94 | | RP | POS | | | 3.2E+07 | 1.3E+07 | 2.9E+07 | 1.2E+07 | 1.16E-01 | -0.22 | 0.96 |
| HILIC_POS_176_12827a3_935 | | 176.1283 | 3.94 | | HILIC | POS | | | 298649 | 654650 | 2484328 | 1.4E+07 | 1.17E-01 | 0.3 | 1.15 |
| RP_POS_744_59009a14_213 | | 744.5901 | 14.21 | | RP | POS | | | 4758795 | 9618363 | 7646060 | 1.6E+07 | 1.17E-01 | 0.23 | 1.16 |
| HILIC_POS_246_10995a5_474 | | 246.1100 | 5.47 | | HILIC | POS | | | 99577.7 | 82511.1 | 121379 | 114740 | 1.17E-01 | 0.22 | 0.95 |
| HILIC_POS_332_24332a1_16 | | 332.2433 | 1.16 | | HILIC | POS | | | 1009329 | 677584 | 1172942 | 823291 | 1.17E-01 | 0.22 | 1.04 |
| RP_POS_283_22672a11_203 | | 283.2267 | 11.20 | | RP | POS | | | 961192 | 477472 | 1094385 | 716233 | 1.17E-01 | 0.22 | 1.36 |
| RP_NEG_408_1604a10_964 | | 408.1604 | 10.96 | | RP | NEG | | | 558469 | 177979 | 595422 | 167035 | 1.17E-01 | 0.21 | 1.24 |
| HILIC_POS_137_07101a2_14 | 1-Methylnicotinamide | 137.0710 | 2.14 | | HILIC | POS | 137.0707(100), 94.0654(2) | 2 | 3.6E+07 | 2.7E+07 | 3.1E+07 | 1.5E+07 | 1.17E-01 | -0.22 | 0.89 |
| RP_NEG_425_27716a12_11 | | 425.2772 | 12.11 | | RP | NEG | | | 154570 | 29799.9 | 148237 | 29241.5 | 1.17E-01 | -0.21 | 1.43 |
| RP_NEG_556_32477a8_872 | | 556.3248 | 8.87 | | RP | NEG | | | 4075050 | 1475894 | 3789436 | 1190431 | 1.17E-01 | -0.21 | 0.97 |
| RP_POS_786_50372a12_941 | | 786.5037 | 12.94 | | RP | POS | | | 8252601 | 3254890 | 7582598 | 2999125 | 1.17E-01 | -0.21 | 0.95 |
| HILIC_NEG_259_0011a1_23 | | 259.0011 | 1.23 | | HILIC | NEG | | | 1467637 | 495892 | 1358981 | 513791 | 1.17E-01 | -0.22 | 0.91 |
| RP_POS_311_63306a10_281 | | 311.6331 | 10.28 | | RP | POS | | | 363045 | 141498 | 331036 | 154860 | 1.17E-01 | -0.22 | 1.19 |
| HILIC_POS_352_24625a2_277 | | 352.2463 | 2.28 | | HILIC | POS | | | 62095.3 | 25288.7 | 80419.1 | 113382 | 1.18E-01 | 0.26 | 0.96 |
| RP_POS_525_04272a10_334 | | 525.0427 | 10.33 | | RP | POS | | | 289830 | 181353 | 344914 | 305848 | 1.18E-01 | 0.23 | 1.39 |
| HILIC_NEG_214_96561a1_013 | | 214.9656 | 1.01 | | HILIC | NEG | | | 1173576 | 327424 | 1247003 | 355164 | 1.18E-01 | 0.22 | 1.16 |
| HILIC_POS_189_05203a2_989 | | 189.0520 | 2.99 | | HILIC | POS | | | 661331 | 730271 | 845701 | 956106 | 1.18E-01 | 0.22 | 1.04 |
| RP_NEG_329_2489a11_116 | | 329.2489 | 11.12 | | RP | NEG | | | 1646883 | 978073 | 1852300 | 942530 | 1.18E-01 | 0.21 | 1.28 |
| RP_POS_341_24377a6_751 | | 341.2438 | 6.75 | | RP | POS | | | 577368 | 76767 | 594116 | 79388.4 | 1.18E-01 | 0.21 | 1.17 |
| RP_NEG_391_28625a9_261 | Hyodeoxycholic acid | 391.2863 | 9.26 | | RP | NEG | 391.2843(100), 345.2823(5) | 1 | 977980 | 837125 | 822301 | 618311 | 1.18E-01 | -0.21 | 0.96 |
| RP_NEG_172_83014a15_467 | | 172.8301 | 15.47 | | RP | NEG | | | 5618636 | 731102 | 5461801 | 733785 | 1.18E-01 | -0.21 | 1.09 |
| RP_NEG_232_99475a4_407 | | 232.9948 | 4.41 | | RP | NEG | | | 238909 | 440049 | 158477 | 310977 | 1.18E-01 | -0.21 | 0.91 |
| RP_POS_206_1385a5_972 | | 206.1385 | 5.97 | | RP | POS | | | 255382 | 48347.2 | 245673 | 42623.4 | 1.18E-01 | -0.21 | 0.99 |
| RP_POS_299_25793a10_778 | | 299.2579 | 10.78 | | RP | POS | | | 2143530 | 298543 | 2079944 | 295316 | 1.18E-01 | -0.21 | 1.23 |
| RP_POS_450_32156a9_375 | | 450.3216 | 9.38 | | RP | POS | | | 1.2E+07 | 1.6E+07 | 9236751 | 1.2E+07 | 1.18E-01 | -0.21 | 0.92 |
| HILIC_NEG_226_94044a5_654 | | 226.9404 | 5.65 | | HILIC | NEG | | | 365403 | 80128.1 | 351089 | 52712.6 | 1.18E-01 | -0.22 | 1.03 |
| HILIC_POS_368_27914a0_628 | | 368.2791 | 0.63 | | HILIC | POS | | | 903827 | 564879 | 1046304 | 742355 | 1.19E-01 | 0.22 | 1.08 |

**Supplemantary table 1**: The non-targeted metabolomics results ordered by the VIP value (p value < 0.0003 are shaded dark green, p value < 0.05 are shaded light green, d values below 0 are blue and above 0 are red)

| | | | | | | | | | Cases | | Controls | | Welch test | Cohen's | |
|---|---|---|---|---|---|---|---|---|---|---|---|---|---|---|---|
| Feature_ID | Metabolite_name | Average_Mz | Average_RT_min_ | HMDB ID | RP_HILIC | POS_NEG | Cureted_MSMS | ID_level | Mean | SD | Mean2 | SD3 | p | d | VIP |
| RP_POS_1151_36548a13_784 | | 1151.3655 | 13.78 | | RP | POS | | | 1194613 | 781089 | 1406431 | 1136977 | 1.19E-01 | 0.22 | 1.06 |
| RP_POS_418_31598a10_597 | | 418.3160 | 10.60 | | RP | POS | | | 1003681 | 586375 | 1159870 | 832179 | 1.19E-01 | 0.22 | 1.31 |
| RP_POS_481_67624a9_906 | | 481.6762 | 9.91 | | RP | POS | | | 115327 | 82257.9 | 134240 | 93570.8 | 1.19E-01 | 0.22 | 1.09 |
| RP_NEG_329_17596a6_737 | | 329.1760 | 6.74 | | RP | NEG | | | 301032 | 73355.5 | 316629 | 72704.4 | 1.19E-01 | 0.21 | 0.91 |
| RP_POS_191_10654a7_509 | | 191.1065 | 7.51 | | RP | POS | | | 3671268 | 254097 | 3725494 | 253293 | 1.19E-01 | 0.21 | 1.03 |
| RP_POS_340_24039a10_653 | | 340.2404 | 10.65 | | RP | POS | | | 1014972 | 521370 | 1126879 | 525275 | 1.19E-01 | 0.21 | 1.23 |
| RP_POS_386_28995a8_114 | | 386.2900 | 8.11 | | RP | POS | | | 621013 | 384218 | 706043 | 409155 | 1.19E-01 | 0.21 | 1.09 |
| RP_NEG_181_05106a3_031 | 3-(4-Hydroxyphenyl)lactic acid | 181.0511 | 3.03 | | RP | NEG | 163.0397(100), 135.0452(60), 119.0507(45) | 2 | 913370 | 263760 | 979444 | 342356 | 1.19E-01 | 0.22 | 0.95 |
| RP_POS_241_14085a8_075 | | 241.1409 | 8.08 | | RP | POS | 241.1403(100) | | 4704984 | 589203 | 4586316 | 524972 | 1.19E-01 | -0.21 | 1.34 |
| HILIC_NEG_341_00375a1_23 | | 341.0038 | 1.23 | | HILIC | NEG | | | 865409 | 338749 | 793520 | 334299 | 1.19E-01 | -0.21 | 1 |
| RP_NEG_295_22833a9_77 | | 295.2283 | 9.77 | | RP | NEG | | | 588501 | 448981 | 488516 | 482390 | 1.19E-01 | -0.21 | 0.99 |
| RP_POS_692_95679a9_906 | | 692.9568 | 9.91 | | RP | POS | | | 430834 | 250287 | 381541 | 212633 | 1.19E-01 | -0.21 | 1.01 |
| RP_POS_431_27676a9_286 | | 431.2768 | 9.29 | | RP | POS | | | 330795 | 553814 | 539578 | 1224662 | 1.20E-01 | 0.23 | 0.92 |
| RP_NEG_136_87204a13_284 | | 136.8720 | 13.28 | | RP | NEG | | | 241547 | 88579.3 | 259927 | 84022.6 | 1.20E-01 | 0.21 | 0.94 |
| RP_POS_235_1694a7_893 | | 235.1694 | 7.89 | | RP | POS | | | 5446719 | 388965 | 5531788 | 408160 | 1.20E-01 | 0.21 | 1.08 |
| RP_POS_252_04553a12_668 | | 252.0455 | 12.67 | | RP | POS | | | 2698847 | 1348275 | 3006563 | 1519976 | 1.20E-01 | 0.21 | 1.26 |
| HILIC_NEG_209_00455a0_934 | | 209.0046 | 0.93 | | HILIC | NEG | | | 855749 | 80626.7 | 837509 | 89659.4 | 1.20E-01 | -0.21 | 1.03 |
| RP_NEG_179_10809a7_719 | | 179.1081 | 7.72 | | RP | NEG | | | 2296001 | 71523 | 2280046 | 77663.4 | 1.20E-01 | -0.21 | 1.09 |
| RP_NEG_370_74969a15_416 | | 370.7497 | 15.42 | | RP | NEG | | | 321375 | 137972 | 293371 | 125561 | 1.20E-01 | -0.21 | 1.06 |
| RP_POS_355_32019a10_828 | | 355.3202 | 10.83 | | RP | POS | | | 895816 | 183585 | 856608 | 183731 | 1.20E-01 | -0.21 | 1.33 |
| HILIC_NEG_424_03265a5_122 | | 424.0327 | 5.12 | | HILIC | NEG | | | 127068 | 91157.1 | 160063 | 193067 | 1.21E-01 | 0.23 | 0.98 |
| RP_POS_794_56885a13_236 | | 794.5689 | 13.24 | | RP | POS | | | 4440396 | 4001933 | 5474681 | 5488480 | 1.21E-01 | 0.22 | 1.2 |
| RP_POS_383_1835a6_738 | | 383.1835 | 6.74 | | RP | POS | | | 406766 | 105372 | 427549 | 91076.2 | 1.21E-01 | 0.21 | 0.89 |
| RP_POS_397_16724a10_651 | | 397.1672 | 10.65 | | RP | POS | | | 469585 | 324688 | 539566 | 332360 | 1.21E-01 | 0.21 | 1.23 |
| HILIC_NEG_212_91119a3_912 | | 212.9112 | 3.91 | | HILIC | NEG | | | 2437343 | 627132 | 2314051 | 535467 | 1.21E-01 | -0.21 | 0.88 |
| HILIC_NEG_308_99854a0_936 | | 308.9985 | 0.94 | | HILIC | NEG | | | 1228050 | 111484 | 1204561 | 109693 | 1.21E-01 | -0.21 | 1.01 |
| HILIC_NEG_98_93915a0_934 | | 98.9392 | 0.93 | | HILIC | NEG | | | 1436343 | 135365 | 1409881 | 114551 | 1.21E-01 | -0.21 | 1.02 |
| RP_NEG_157_08711a6_66 | | 157.0871 | 6.66 | | RP | NEG | | | 727349 | 516945 | 638018 | 314708 | 1.21E-01 | -0.21 | 0.91 |
| RP_POS_1058_26501a9_767 | | 1058.2650 | 9.77 | | RP | POS | | | 2052956 | 279569 | 1996652 | 251980 | 1.21E-01 | -0.21 | 1.16 |
| RP_POS_275_08823a7_334 | | 275.0882 | 7.33 | | RP | POS | | | 370359 | 37683.6 | 362814 | 33566.3 | 1.21E-01 | -0.21 | 0.98 |
| RP_POS_303_64709a10_278 | | 303.6471 | 10.28 | | RP | POS | | | 391111 | 168265 | 356034 | 162144 | 1.21E-01 | -0.21 | 1.08 |
| RP_POS_323_25552a10_744 | | 323.2555 | 10.74 | | RP | POS | | | 2516759 | 314794 | 2450899 | 306046 | 1.21E-01 | -0.21 | 1.31 |
| RP_POS_327_28967a10_393 | | 327.2897 | 10.39 | | RP | POS | | | 1173039 | 172154 | 1136449 | 171612 | 1.21E-01 | -0.21 | 1.32 |
| RP_POS_472_30344a9_375 | | 472.3034 | 9.38 | | RP | POS | | | 2351788 | 3242337 | 1761236 | 2317226 | 1.21E-01 | -0.21 | 0.92 |
| RP_POS_749_38306a4_704 | | 749.3831 | 4.70 | | RP | POS | | | 2787595 | 1476522 | 2462049 | 1574431 | 1.21E-01 | -0.21 | 1.36 |
| HILIC_POS_307_0437a2_502 | | 307.0437 | 2.50 | | HILIC | POS | | | 272616 | 326502 | 216960 | 190658 | 1.21E-01 | -0.22 | 0.94 |
| RP_POS_204_97922a1_626 | | 204.9792 | 1.63 | | RP | POS | | | 499446 | 237161 | 556744 | 294811 | 1.22E-01 | 0.22 | 1.13 |
| RP_POS_796_61938a14_341 | | 796.6194 | 14.34 | | RP | POS | | | 2181137 | 1435489 | 2537935 | 1865106 | 1.22E-01 | 0.22 | 1.18 |
| RP_NEG_359_29596a11_648 | | 359.2960 | 11.65 | | RP | NEG | | | 118204 | 60377.1 | 131091 | 61229.3 | 1.22E-01 | 0.21 | 1.23 |
| RP_POS_1017_68732a10_486 | | 1017.6873 | 10.49 | | RP | POS | | | 1.9E+07 | 5468650 | 2E+07 | 5962396 | 1.22E-01 | 0.21 | 1.04 |
| RP_POS_245_13206a0_634 | | 245.1321 | 0.63 | | RP | POS | | | 278685 | 67645.8 | 292977 | 67124 | 1.22E-01 | 0.21 | 1.05 |
| RP_POS_263_23676a10_96 | | 263.2368 | 10.96 | | RP | POS | | | 1275868 | 679198 | 1417721 | 659152 | 1.22E-01 | 0.21 | 1.3 |
| RP_POS_744_55182a12_889 | PC 33:2 (18:2_15:0) | 744.5518 | 12.89 | | RP | POS | 184.0744(100), 86.0966(10) | 1 | 3.7E+07 | 9419302 | 3.5E+07 | 9625930 | 1.22E-01 | -0.21 | 1.03 |
| HILIC_NEG_177_85999a3_151 | | 177.8600 | 3.15 | | HILIC | NEG | | | 801413 | 149617 | 768068 | 163390 | 1.22E-01 | -0.21 | 0.89 |
| HILIC_POS_142_1225a3_104 | | 142.1225 | 3.10 | | HILIC | POS | | | 113302 | 44930.5 | 103812 | 44544 | 1.22E-01 | -0.21 | 0.94 |
| HILIC_POS_371_23975a7_1 | | 371.2398 | 7.10 | | HILIC | POS | | | 2290438 | 216262 | 2245485 | 208740 | 1.22E-01 | -0.21 | 1.12 |
| RP_POS_334_186a6_267 | | 334.1860 | 6.27 | | RP | POS | | | 479785 | 593358 | 378774 | 348231 | 1.22E-01 | -0.21 | 1.03 |
| RP_POS_601_43799a10_474 | | 601.4380 | 10.47 | | RP | POS | | | 1653221 | 754791 | 1502831 | 667830 | 1.22E-01 | -0.21 | 0.94 |
| RP_POS_721_57654a15_253 | | 721.5765 | 15.25 | | RP | POS | | | 379221 | 336761 | 460653 | 420672 | 1.23E-01 | 0.22 | 0.93 |
| RP_POS_451_22934a9_172 | | 451.2293 | 9.17 | | RP | POS | | | 1486728 | 683084 | 1645292 | 801266 | 1.23E-01 | 0.21 | 1.14 |
| RP_POS_832_58197a13_628 | | 832.5820 | 13.63 | | RP | POS | | | 1.5E+07 | 5999801 | 1.6E+07 | 5192473 | 1.23E-01 | 0.21 | 1.01 |
| HILIC_NEG_103_03989a0_948 | 2-hydroxy-butanoic acid | 103.0399 | 0.95 | | HILIC | NEG | 103.0398(100), 57.0339(90) | 1 | 1.2E+07 | 4999713 | 1.3E+07 | 6073028 | 1.23E-01 | 0.21 | 1.04 |
| HILIC_POS_130_08649a3_566 | Pipecolic acid | 130.0865 | 3.57 | | HILIC | POS | 84.0816(100), 130.1588(85) | 2 | 6.9E+07 | 4.7E+08 | 1580870 | 2089789 | 1.23E-01 | -0.29 | 0.85 |
| RP_NEG_372_99112a8_516 | | 372.9911 | 8.52 | | RP | NEG | | | 1760110 | 239566 | 1710741 | 227631 | 1.23E-01 | -0.21 | 0.93 |
| RP_NEG_562_31549a9_875 | | 562.3155 | 9.88 | | RP | NEG | | | 1004851 | 444062 | 908266 | 467605 | 1.23E-01 | -0.21 | 0.97 |
| RP_POS_304_28448a8_418 | | 304.2845 | 8.42 | | RP | POS | | | 1289109 | 2401726 | 865039 | 1591104 | 1.23E-01 | -0.21 | 0.84 |
| RP_POS_336_31052a10_424 | | 336.3105 | 10.42 | | RP | POS | | | 980332 | 181989 | 942443 | 176950 | 1.23E-01 | -0.21 | 1.12 |
| RP_POS_286_2012a5_646 | | 286.2012 | 5.65 | | RP | POS | 85.0292(100)286.2(40) | | 604363 | 361953 | 811542 | 1288579 | 1.24E-01 | 0.25 | 1.02 |
| RP_POS_697_15045a11_449 | | 697.1505 | 11.45 | | RP | POS | | | 322662 | 164623 | 366156 | 235524 | 1.24E-01 | 0.22 | 1.07 |
| RP_POS_429_76974a10_957 | | 429.7697 | 10.96 | | RP | POS | | | 255466 | 136601 | 288200 | 169033 | 1.24E-01 | 0.21 | 1.21 |
| HILIC_NEG_102_93201a0_935 | | 102.9320 | 0.94 | | HILIC | NEG | | | 808747 | 86360.8 | 791097 | 81436.5 | 1.24E-01 | -0.21 | 0.95 |
| RP_NEG_299_25952a10_744 | | 299.2595 | 10.74 | | RP | NEG | | | 3168444 | 416504 | 3081331 | 410564 | 1.24E-01 | -0.21 | 1.27 |
| RP_POS_130_92375a15_48 | | 130.9238 | 15.48 | | RP | POS | | | 905574 | 354654 | 834735 | 318557 | 1.24E-01 | -0.21 | 1.17 |
| RP_POS_271_18759a8_215 | | 271.1876 | 8.22 | | RP | POS | | | 812776 | 333125 | 745243 | 308140 | 1.24E-01 | -0.21 | 1.06 |
| RP_POS_914_48444a13_002 | | 914.4844 | 13.00 | | RP | POS | | | 585347 | 321555 | 518244 | 314621 | 1.24E-01 | -0.21 | 1.01 |
| RP_POS_801_54114a13_107 | | 801.5411 | 13.11 | | RP | POS | | | 1526308 | 1368205 | 1986057 | 2687069 | 1.25E-01 | 0.23 | 1.1 |
| RP_POS_203_14297a8_856 | | 203.1430 | 8.86 | | RP | POS | | | 8106499 | 1154222 | 8345072 | 1117604 | 1.25E-01 | 0.21 | 1.03 |
| RP_NEG_471_15033a11_145 | | 471.1503 | 11.15 | | RP | NEG | | | 344972 | 79321.5 | 327705 | 84412.2 | 1.25E-01 | -0.21 | 1.07 |
| RP_POS_393_29745a11_456 | | 393.2975 | 11.46 | | RP | POS | | | 6765539 | 977538 | 6569719 | 890816 | 1.25E-01 | -0.21 | 1.11 |
| RP_POS_414_30008a9_375 | | 414.3001 | 9.38 | | RP | POS | | | 3694877 | 4748252 | 2805893 | 3739070 | 1.25E-01 | -0.21 | 0.93 |
| RP_POS_472_32861a8_377 | | 472.3286 | 8.38 | | RP | POS | | | 482378 | 284376 | 430062 | 214192 | 1.25E-01 | -0.21 | 1.02 |
| RP_POS_479_39337a10_88 | | 479.3934 | 10.88 | | RP | POS | | | 721344 | 170298 | 685221 | 173135 | 1.25E-01 | -0.21 | 1.09 |
| RP_POS_741_05438a12_8 | | 741.0544 | 12.80 | | RP | POS | | | 440997 | 387116 | 366168 | 327051 | 1.25E-01 | -0.21 | 0.98 |
| RP_POS_499_30292a10_847 | | 499.3029 | 10.85 | | RP | POS | | | 764815 | 452525 | 884793 | 655781 | 1.26E-01 | 0.22 | 1.4 |
| RP_POS_805_57562a14_081 | | 805.5756 | 14.08 | | RP | POS | | | 904341 | 588360 | 1059224 | 844622 | 1.26E-01 | 0.22 | 1.03 |
| RP_POS_170_11751a4_605 | | 170.1175 | 4.61 | | RP | POS | | | 821839 | 136789 | 855050 | 174012 | 1.26E-01 | 0.21 | 1.1 |
| RP_POS_445_27957a9_742 | | 445.2796 | 9.74 | | RP | POS | | | 637869 | 408273 | 724293 | 415121 | 1.26E-01 | 0.21 | 1.24 |
| RP_POS_446_34744a11_058 | | 446.3474 | 11.06 | | RP | POS | | | 1212089 | 641579 | 1359489 | 749892 | 1.26E-01 | 0.21 | 1.39 |
| HILIC_NEG_343_72153a1_407 | | 343.7215 | 1.41 | | HILIC | NEG | | | 96658 | 19599.5 | 92782.6 | 17470.2 | 1.26E-01 | -0.21 | 0.9 |
| RP_NEG_189_00266a3_68 | | 189.0027 | 3.68 | | RP | NEG | | | 5888106 | 4474387 | 5038509 | 3651292 | 1.26E-01 | -0.21 | 0.98 |
| RP_POS_276_14398a0_68 | | 276.1440 | 0.68 | | RP | POS | | | 343555 | 134131 | 317872 | 111482 | 1.26E-01 | -0.21 | 0.88 |
| RP_POS_392_77332a9_654 | | 392.7733 | 9.65 | | RP | POS | | | 453979 | 245598 | 405513 | 218201 | 1.26E-01 | -0.21 | 1 |
| RP_POS_675_54376a12_24 | | 675.5438 | 12.24 | | RP | POS | | | 5.7E+07 | 1.4E+07 | 5.4E+07 | 1.1E+07 | 1.26E-01 | -0.21 | 1.03 |
| RP_POS_778_53516a12_819 | | 778.5352 | 12.82 | | RP | POS | | | 5.1E+07 | 1.9E+07 | 4.7E+07 | 1.8E+07 | 1.26E-01 | -0.21 | 0.96 |
| RP_POS_499_30304a10_986 | | 499.3030 | 10.99 | | RP | POS | | | 1128615 | 617748 | 1288160 | 866800 | 1.27E-01 | 0.21 | 1.34 |
| RP_POS_343_29483a8_959 | Cocamidopropyl betaine | 343.2948 | 8.96 | | RP | POS | 240.2336(100) | 2 | 2429092 | 307990 | 2506325 | 413117 | 1.27E-01 | 0.21 | 1.19 |
| HILIC_POS_200_98346a0_93 | | 200.9835 | 0.93 | | HILIC | POS | | | 162162 | 57157.9 | 150562 | 53983.1 | 1.27E-01 | -0.21 | 1.05 |
| RP_POS_134_08125a10_408 | | 134.0813 | 10.41 | | RP | POS | | | 5767122 | 2787599 | 5253640 | 2127166 | 1.27E-01 | -0.21 | 1.02 |
| RP_POS_219_1745a9_611 | | 219.1745 | 9.61 | | RP | POS | | | 3202056 | 525066 | 3095842 | 493713 | 1.27E-01 | -0.21 | 1.08 |
| RP_POS_453_24564a3_762 | | 453.2456 | 3.76 | | RP | POS | | | 5128663 | 2669180 | 4552891 | 2821311 | 1.27E-01 | -0.21 | 1.14 |
| RP_POS_548_50232a11_591 | | 548.5023 | 11.59 | | RP | POS | | | 2140375 | 615019 | 2024677 | 494284 | 1.27E-01 | -0.21 | 0.97 |
| HILIC_NEG_143_0349a5_12 | | 143.0349 | 5.12 | | HILIC | NEG | | | 2524433 | 1104972 | 2861609 | 1940847 | 1.28E-01 | 0.22 | 0.99 |
| HILIC_POS_213_12341a4_865 | | 213.1234 | 4.87 | | HILIC | POS | | | 31760.9 | 17655.2 | 36455.5 | 25842.6 | 1.28E-01 | 0.22 | 0.99 |
| HILIC_POS_626_3006a1_661 | | 626.3006 | 1.66 | | HILIC | POS | | | 130403 | 39612.7 | 139284 | 44850.1 | 1.28E-01 | 0.21 | 0.96 |
| RP_NEG_439_20816a10_923 | | 439.2082 | 10.92 | | RP | NEG | | | 266173 | 128648 | 292007 | 119870 | 1.28E-01 | 0.21 | 1.15 |
| RP_POS_1043_70337a10_607 | | 1043.7034 | 10.61 | | RP | POS | | | 2.6E+07 | 1.5E+07 | 2.9E+07 | 1.7E+07 | 1.28E-01 | 0.21 | 1.01 |
| RP_POS_451_30304a11_057 | | 451.3030 | 11.06 | | RP | POS | | | 2545653 | 1212372 | 2832202 | 1493358 | 1.28E-01 | 0.21 | 1.35 |
| HILIC_POS_189_16003a6_769 | Ne,Ne,Ne-Trimethyllysine | 189.1600 | 6.77 | | HILIC | POS | 84.0814(100), 130.0867(95) | 1 | 6673655 | 1959894 | 7081639 | 1955608 | 1.28E-01 | 0.21 | 1.05 |
| HILIC_POS_59_06134a1_347 | | 59.0613 | 1.35 | | HILIC | POS | | | 7398638 | 2993639 | 6754676 | 3166621 | 1.28E-01 | -0.21 | 0.9 |
| RP_POS_258_08136a15_416 | | 258.0814 | 15.42 | | RP | POS | | | 240043 | 143234 | 212930 | 117763 | 1.28E-01 | -0.21 | 1.03 |
| RP_POS_352_22479a9_358 | | 352.2248 | 9.36 | | RP | POS | | | 918535 | 312592 | 861390 | 235693 | 1.28E-01 | -0.21 | 0.9 |
| RP_POS_416_2858a5_434 | | 416.2858 | 5.43 | | RP | POS | | | 577970 | 318389 | 511622 | 318120 | 1.28E-01 | -0.21 | 1.05 |
| RP_POS_465_32153a8_101 | | 465.3215 | 8.10 | | RP | POS | | | 300081 | 244955 | 258623 | 148304 | 1.28E-01 | -0.21 | 1.06 |
| HILIC_POS_177_04347a3_146 | | 177.0435 | 3.15 | | HILIC | POS | | | 409811 | 47026.1 | 395198 | 84474 | 1.28E-01 | -0.22 | 0.92 |
| RP_POS_287_14697a8_501 | | 287.1470 | 8.50 | | RP | POS | | | 1098260 | 3604413 | 555472 | 1215614 | 1.28E-01 | -0.23 | 0.82 |
| HILIC_NEG_139_94464a0_939 | | 139.9446 | 0.94 | | HILIC | NEG | | | 546508 | 90637.9 | 561754 | 54115.7 | 1.29E-01 | 0.21 | 0.9 |
| HILIC_POS_544_34076a1_161 | | 544.3408 | 1.16 | | HILIC | POS | | | 1.9E+07 | 5421246 | 2E+07 | 5560425 | 1.29E-01 | 0.21 | 1.01 |
| RP_NEG_558_31464a10_61 | | 558.3146 | 10.61 | | RP | NEG | | | 1334660 | 215434 | 1382093 | 237489 | 1.29E-01 | 0.21 | 0.97 |
| RP_POS_382_05542a0_585 | | 382.0554 | 0.59 | | RP | POS | | | 1827832 | 786450 | 2000039 | 858492 | 1.29E-01 | 0.21 | 0.98 |
| RP_POS_225_14873a7_874 | Methyl jasmonate | 225.1487 | 7.87 | | RP | POS | 225.1495(100) | 2 | 2878204 | 113351 | 2902316 | 118067 | 1.29E-01 | 0.21 | 0.91 |
| HILIC_NEG_226_99515a0_935 | | 226.9952 | 0.94 | | HILIC | NEG | | | 1656336 | 154885 | 1623385 | 160977 | 1.29E-01 | -0.21 | 0.97 |
| HILIC_POS_198_12364a6_136 | | 198.1236 | 6.14 | | HILIC | POS | | | 627515 | 488496 | 515990 | 573547 | 1.29E-01 | -0.21 | 1.04 |
| RP_NEG_335_93561a3_68 | | 335.9356 | 3.68 | | RP | NEG | | | 979899 | 720218 | 840512 | 622485 | 1.29E-01 | -0.21 | 1.04 |
| RP_NEG_798_53076a12_509 | | 798.5308 | 12.51 | | RP | NEG | | | 5049458 | 2073459 | 4654769 | 1737428 | 1.29E-01 | -0.21 | 0.88 |
| RP_POS_736_54193a13_695 | | 736.5419 | 13.70 | | RP | POS | | | 3104426 | 880853 | 2936270 | 741705 | 1.29E-01 | -0.21 | 1.02 |
| RP_POS_840_58728a13_641 | | 840.5873 | 13.64 | | RP | POS | | | 3218138 | 1486404 | 2920977 | 1372995 | 1.29E-01 | -0.21 | 0.95 |
| HILIC_POS_394_31525a6_303 | | 394.3153 | 6.30 | | HILIC | POS | | | 926918 | 435272 | 1018971 | 450133 | 1.30E-01 | 0.21 | 1.23 |
| RP_POS_560_3017a10_304 | | 560.3017 | 10.30 | | RP | POS | | | 1309699 | 348071 | 1386192 | 384922 | 1.30E-01 | 0.21 | 1 |
| RP_POS_342_26416a7_959 | Dodecenoylcarnitine | 342.2642 | 7.96 | | RP | POS | 85.0298(100), 342.2652(40) | 1 | 1691680 | 902677 | 1884747 | 951748 | 1.30E-01 | 0.21 | 1.13 |
| RP_NEG_737_0719a9_194 | | 737.0719 | 9.19 | | RP | NEG | | | 404560 | 34284.9 | 397551 | 33257 | 1.30E-01 | -0.21 | 1.04 |
| RP_POS_130_0865a0_584 | | 130.0865 | 0.58 | | RP | POS | | | 3182864 | 3976621 | 2441147 | 3198954 | 1.30E-01 | -0.21 | 0.87 |
| RP_POS_408_31058a9_616 | | 408.3106 | 9.62 | | RP | POS | | | 496253 | 451273 | 413541 | 347049 | 1.30E-01 | -0.21 | 0.88 |
| RP_POS_596_37006a10_62 | | 596.3701 | 10.62 | | RP | POS | | | 693768 | 340463 | 617570 | 388068 | 1.30E-01 | -0.21 | 0.98 |
| RP_POS_606_29584a10_281 | | 606.2958 | 10.28 | | RP | POS | | | 350680 | 151357 | 319253 | 151148 | 1.30E-01 | -0.21 | 1.02 |
| RP_NEG_352_85312a15_427 | | 352.8531 | 15.43 | | RP | NEG | | | 1020656 | 4540946 | 3128222 | 1.3E+07 | 1.31E-01 | 0.24 | 1.23 |
| HILIC_POS_357_00732a7_361 | | 357.0073 | 7.36 | | HILIC | POS | | | 1824463 | 87428.5 | 1843998 | 99741 | 1.31E-01 | 0.21 | 0.88 |
| RP_POS_311_18521a10_389 | | 311.1852 | 10.39 | | RP | POS | | | 4811367 | 2325384 | 5336601 | 2696977 | 1.31E-01 | 0.21 | 1.32 |
| RP_POS_462_30563a9_744 | | 462.3056 | 9.74 | | RP | POS | | | 688220 | 413633 | 776698 | 438512 | 1.31E-01 | 0.21 | 1.24 |
| HILIC_NEG_316_94833a5_464 | | 316.9483 | 5.46 | | HILIC | NEG | | | 120518 | 8427.42 | 118610 | 9821.24 | 1.31E-01 | -0.21 | 0.93 |
| RP_NEG_487_25827a12_746 | | 487.2583 | 12.75 | | RP | NEG | | | 189283 | 76947.9 | 174468 | 66812.1 | 1.31E-01 | -0.21 | 1.36 |
| RP_POS_311_1459a6_229 | | 311.1459 | 6.23 | | RP | POS | | | 302097 | 500220 | 228914 | 133410 | 1.31E-01 | -0.23 | 0.82 |
| RP_POS_459_25043a10_837 | | 459.2504 | 10.84 | | RP | POS | | | 974553 | 2622966 | 3188642 | 1.4E+07 | 1.32E-01 | 0.26 | 0.96 |
| RP_POS_212_20032a10_832 | | 212.2003 | 10.83 | | RP | POS | | | 183788 | 98385.1 | 211787 | 160382 | 1.32E-01 | 0.22 | 1.17 |
| RP_POS_1085_65356a10_301 | | 1085.6536 | 10.30 | | RP | POS | | | 2319541 | 674170 | 2462601 | 708694 | 1.32E-01 | 0.21 | 0.97 |
| RP_POS_255_1225a10_39 | | 255.1225 | 10.39 | | RP | POS | | | 988922 | 461436 | 1092626 | 534154 | 1.32E-01 | 0.21 | 1.3 |
| RP_POS_256_13089a15_426 | | 256.1309 | 15.43 | | RP | POS | | | 838487 | 350168 | 912389 | 364462 | 1.32E-01 | 0.21 | 0.86 |
| HILIC_POS_174_0135a4_257 | | 174.0135 | 4.26 | | HILIC | POS | | | 354139 | 151146 | 321957 | 160022 | 1.32E-01 | -0.21 | 0.89 |
| HILIC_POS_59_06134a3_043 | | 59.0613 | 3.04 | | HILIC | POS | | | 9757874 | 3472255 | 9057528 | 3317066 | 1.32E-01 | -0.21 | 1 |
| RP_NEG_179_88275a3_678 | | 179.8828 | 3.68 | | RP | NEG | | | 637560 | 544177 | 534934 | 452667 | 1.32E-01 | -0.21 | 0.96 |
| RP_NEG_227_9604a0_681 | | 227.9604 | 0.68 | | RP | NEG | | | 1156196 | 210509 | 1111502 | 221363 | 1.32E-01 | -0.21 | 0.98 |
| RP_POS_347_18912a11_143 | | 347.1891 | 11.14 | | RP | POS | | | 2374605 | 412090 | 2291813 | 391297 | 1.32E-01 | -0.21 | 1.1 |
| RP_POS_648_45398a7_825 | | 648.4540 | 7.83 | | RP | POS | | | 541828 | 356836 | 473369 | 307998 | 1.32E-01 | -0.21 | 1.03 |
| RP_POS_774_54327a13_368 | | 774.5433 | 13.37 | | RP | POS | | | 4209048 | 1469514 | 3943260 | 1106730 | 1.32E-01 | -0.21 | 1.12 |
| RP_POS_854_56616a13_143 | | 854.5662 | 13.14 | | RP | POS | | | 6879754 | 2665489 | 6343612 | 2540139 | 1.32E-01 | -0.21 | 1.33 |
| RP_POS_624_4469a11_799 | | 624.4469 | 11.80 | | RP | POS | | | 906631 | 1562145 | 672897 | 545787 | 1.32E-01 | -0.22 | 0.83 |
| RP_POS_372_31097a10_669 | | 372.3110 | 10.67 | | RP | POS | | | 3266716 | 2350839 | 3971784 | 4107314 | 1.33E-01 | 0.22 | 1.15 |
| RP_POS_192_15944a0_676 | | 192.1594 | 0.68 | | RP | POS | | | 1196304 | 200386 | 1238889 | 211554 | 1.33E-01 | 0.21 | 1.09 |
| RP_POS_263_23691a10_191 | | 263.2369 | 10.19 | | RP | POS | | | 1802692 | 1314069 | 2120221 | 1711643 | 1.33E-01 | 0.21 | 0.99 |
| RP_POS_332_3306a8_93 | | 332.3306 | 8.93 | | RP | POS | | | 1479834 | 768017 | 1664352 | 991576 | 1.33E-01 | 0.21 | 1.16 |
| RP_POS_523_00598a10_189 | | 523.0060 | 10.19 | | RP | POS | | | 175731 | 95392.8 | 201657 | 146510 | 1.33E-01 | 0.21 | 1.07 |
| RP_POS_535_32416a10_872 | | 535.3242 | 10.87 | | RP | POS | | | 1873872 | 968966 | 2119394 | 1352696 | 1.33E-01 | 0.21 | 1.38 |
| RP_POS_389_16269a7_243 | Cetirizine | 389.1627 | 7.24 | | RP | POS | 201.046(100),166.0787(10) | 2 | 2354641 | 9794094 | 945181 | 2195771 | 1.33E-01 | -0.24 | 0.85 |
| RP_NEG_324_98917a7_822 | | 324.9892 | 7.82 | | RP | NEG | | | 2463444 | 319009 | 2402729 | 273016 | 1.33E-01 | -0.21 | 0.87 |
| RP_NEG_708_46375a11_592 | | 708.4638 | 11.59 | | RP | NEG | | | 735529 | 72346.4 | 720013 | 77788 | 1.33E-01 | -0.21 | 1.09 |
| RP_POS_421_3291a11_597 | | 421.3291 | 11.60 | | RP | POS | | | 732297 | 279538 | 682510 | 204132 | 1.33E-01 | -0.21 | 0.83 |
| RP_POS_512_87091a10_216 | | 512.8709 | 10.22 | | RP | POS | | | 2515057 | 1135448 | 2285423 | 1098923 | 1.33E-01 | -0.21 | 0.92 |
| RP_POS_523_36359a8_673 | | 523.3636 | 8.67 | | RP | POS | | | 441889 | 270642 | 386216 | 269904 | 1.33E-01 | -0.21 | 1.04 |
| RP_POS_529_38287a9_797 | | 529.3829 | 9.80 | | RP | POS | | | 7491712 | 2891912 | 6908898 | 2781979 | 1.33E-01 | -0.21 | 1.01 |
| RP_POS_661_46075a9_744 | | 661.4608 | 9.74 | | RP | POS | | | 1571882 | 716265 | 1437588 | 591959 | 1.33E-01 | -0.21 | 0.99 |
| RP_POS_874_55377a13_473 | | 874.5538 | 13.47 | | RP | POS | | | 9486700 | 5608237 | 1.1E+07 | 5838118 | 1.34E-01 | 0.21 | 1.04 |
| HILIC_NEG_231_93163a6_139 | | 231.9316 | 6.14 | | HILIC | NEG | | | 1255551 | 194238 | 1217823 | 174133 | 1.34E-01 | -0.2 | 0.96 |
| RP_POS_269_11438a8_844 | | 269.1144 | 8.84 | | RP | POS | | | 5664844 | 2155550 | 5254086 | 1855702 | 1.34E-01 | -0.2 | 1.25 |

**Supplemantary table 1**: The non-targeted metabolomics results ordered by the VIP value (p value < 0.0003 are shaded dark green, p value < 0.05 are shaded light green, d values below 0 are blue and above 0 are red)

| | | | | | | | | | Cases | | Controls | | Welch test | Cohen's | |
|---|---|---|---|---|---|---|---|---|---|---|---|---|---|---|---|
| Feature_ID | Metabolite_name | Average_Mz | Average_RT_min_ | HMDB ID | RP_HILIC | POS_NEG | Cureted_MSMS | ID_level | Mean | SD | Mean2 | SD3 | p | d | VIP |
| HILIC_POS_387_21399a7_1 | | 387.2140 | 7.10 | | HILIC | POS | | | 597183 | 60854.3 | 584402 | 63399 | 1.34E-01 | -0.21 | 1.01 |
| RP_POS_203_10646a9_074 | | 203.1065 | 9.07 | | RP | POS | | | 1634883 | 208556 | 1592608 | 203624 | 1.34E-01 | -0.21 | 0.9 |
| RP_POS_385_25854a10_389 | | 385.2585 | 10.39 | | RP | POS | 385.2928(100) | | 1E+07 | 5042946 | 1.1E+07 | 5761924 | 1.35E-01 | 0.21 | 1.32 |
| HILIC_NEG_325_05609a5_109 | | 325.0561 | 5.11 | | HILIC | NEG | | | 329077 | 175428 | 366675 | 189608 | 1.35E-01 | 0.21 | 0.94 |
| RP_NEG_475_11575a10_004 | | 475.1158 | 10.00 | | RP | NEG | | | 228637 | 196655 | 270958 | 214222 | 1.35E-01 | 0.21 | 1.16 |
| RP_POS_549_06189a10_581 | | 549.0619 | 10.58 | | RP | POS | | | 155334 | 68812.1 | 172020 | 90629.6 | 1.35E-01 | 0.21 | 1.12 |
| RP_NEG_357_30167a12_11 | | 357.3017 | 12.11 | | RP | NEG | | | 79525.2 | 44227 | 70637.3 | 42598.1 | 1.35E-01 | -0.2 | 1.36 |
| RP_NEG_358_99542a15_466 | | 358.9954 | 15.47 | | RP | NEG | | | 1557538 | 186390 | 1520824 | 172524 | 1.35E-01 | -0.2 | 0.87 |
| RP_POS_171_14899a5_896 | | 171.1490 | 5.90 | | RP | POS | | | 6112347 | 1145424 | 5878211 | 1139170 | 1.35E-01 | -0.2 | 1.13 |
| RP_POS_182_90227a15_434 | | 182.9023 | 15.43 | | RP | POS | | | 1.3E+07 | 1757597 | 1.3E+07 | 1649469 | 1.35E-01 | -0.2 | 0.9 |
| RP_POS_350_21249a10_634 | | 350.2125 | 10.63 | | RP | POS | | | 1.1E+07 | 3217349 | 1E+07 | 2792912 | 1.35E-01 | -0.2 | 0.94 |
| HILIC_NEG_307_90732a7_414 | | 307.9073 | 7.41 | | HILIC | NEG | | | 118901 | 44475.1 | 109609 | 46124.7 | 1.35E-01 | -0.21 | 0.95 |
| RP_POS_316_88315a15_412 | | 316.8832 | 15.41 | | RP | POS | | | 1877540 | 2040100 | 1531557 | 1320418 | 1.35E-01 | -0.21 | 1.06 |
| RP_POS_616_46454a9_435 | | 616.4645 | 9.44 | | RP | POS | | | 4286058 | 1842690 | 3883282 | 2063779 | 1.35E-01 | -0.21 | 1 |
| RP_POS_355_28427a10_675 | | 355.2843 | 10.68 | | RP | POS | | | 1.4E+07 | 5198766 | 1.6E+07 | 9543874 | 1.36E-01 | 0.22 | 1.18 |
| HILIC_POS_203_05264a5_16 | | 203.0526 | 5.16 | | HILIC | POS | 203.0539(100) | | 3.3E+07 | 1.3E+07 | 3.6E+07 | 1.3E+07 | 1.36E-01 | 0.2 | 0.99 |
| HILIC_NEG_194_98898a2_268 | | 194.9890 | 2.27 | | HILIC | NEG | | | 1184870 | 292226 | 1243892 | 285695 | 1.36E-01 | 0.2 | 0.95 |
| RP_NEG_735_52386a12_354 | | 735.5239 | 12.35 | | RP | NEG | | | 672201 | 135345 | 699200 | 129125 | 1.36E-01 | 0.2 | 0.89 |
| RP_POS_277_17981a9_178 | | 277.1798 | 9.18 | | RP | POS | | | 3479568 | 323550 | 3539340 | 263913 | 1.36E-01 | 0.2 | 0.98 |
| HILIC_POS_130_04987a1_882 | | 130.0499 | 1.88 | | HILIC | POS | 130.1586(100), 84.0813(20) | | 294002 | 337471 | 232839 | 263296 | 1.36E-01 | -0.2 | 0.92 |
| RP_NEG_455_24802a9_29 | | 455.2480 | 9.29 | | RP | NEG | | | 510127 | 505678 | 416334 | 415584 | 1.36E-01 | -0.2 | 0.84 |
| RP_POS_652_47137a10_594 | | 652.4714 | 10.59 | | RP | POS | | | 1610118 | 806205 | 1458963 | 678006 | 1.36E-01 | -0.2 | 0.94 |
| RP_POS_583_73499a9_851 | | 583.7350 | 9.85 | | RP | POS | | | 154424 | 93392.2 | 175173 | 108551 | 1.37E-01 | 0.21 | 0.95 |
| RP_POS_496_01727a9_892 | | 496.0173 | 9.89 | | RP | POS | | | 422090 | 269450 | 478232 | 280673 | 1.37E-01 | 0.2 | 0.91 |
| RP_POS_583_55109a11_334 | | 583.5511 | 11.33 | | RP | POS | | | 85783.4 | 53505.1 | 96742.5 | 53868.4 | 1.37E-01 | 0.2 | 0.86 |
| HILIC_POS_522_3551a1_18 | LysoPC 18:1 | 522.3551 | 1.18 | | HILIC | POS | 184.0744(100), 104.1075(75) | 2 | 5.8E+07 | 1.7E+07 | 6.1E+07 | 1.8E+07 | 1.37E-01 | 0.20 | 1.01 |
| HILIC_NEG_126_90514a0_932 | | 126.9051 | 0.93 | | HILIC | NEG | | | 196339 | 38057 | 188649 | 37518.6 | 1.37E-01 | -0.2 | 0.98 |
| HILIC_POS_118_08659a5_651 | | 118.0866 | 5.65 | | HILIC | POS | | | 1.3E+07 | 3639222 | 1.2E+07 | 3988557 | 1.37E-01 | -0.2 | 0.89 |
| RP_POS_147_06508a8_844 | | 147.0651 | 8.84 | | RP | POS | | | 914458 | 315098 | 854724 | 273241 | 1.37E-01 | -0.2 | 0.98 |
| RP_POS_410_7612a12_872 | | 410.7612 | 12.87 | | RP | POS | | | 1362498 | 746697 | 1214149 | 711107 | 1.37E-01 | -0.2 | 1.03 |
| RP_POS_248_14908a0_678 | | 248.1491 | 0.68 | | RP | POS | | | 499080 | 300442 | 565218 | 344348 | 1.38E-01 | 0.21 | 1.22 |
| RP_POS_181_10123a8_342 | Prednisolone | 181.1012 | 8.34 | | RP | POS | 181.1021(100), 165.0705(65) | 2 | 102936 | 413133 | 483870 | 2509945 | 1.38E-01 | 0.26 | 0.98 |
| RP_NEG_134_87509a15_379 | | 134.8751 | 15.38 | | RP | NEG | | | 1156671 | 507175 | 1045469 | 579113 | 1.38E-01 | -0.2 | 1.07 |
| RP_NEG_144_92355a11_591 | | 144.9236 | 11.59 | | RP | NEG | | | 1513247 | 156979 | 1482101 | 150174 | 1.38E-01 | -0.2 | 1.03 |
| RP_POS_486_34506a8_891 | | 486.3451 | 8.89 | | RP | POS | | | 513192 | 296699 | 448883 | 332451 | 1.38E-01 | -0.2 | 1.1 |
| HILIC_NEG_234_81947a3_094 | | 234.8195 | 3.09 | | HILIC | NEG | | | 501322 | 30932 | 491659 | 57810.2 | 1.38E-01 | -0.22 | 1.33 |
| RP_POS_518_36896a10_789 | | 518.3690 | 10.79 | | RP | POS | | | 6587330 | 3966246 | 7574680 | 5519507 | 1.39E-01 | 0.21 | 1.39 |
| RP_POS_796_62103a14_737 | | 796.6210 | 14.74 | | RP | POS | | | 5178524 | 3073083 | 5865503 | 3624778 | 1.39E-01 | 0.21 | 1.07 |
| HILIC_NEG_158_97874a7_403 | | 158.9787 | 7.40 | | HILIC | NEG | | | 5300059 | 358674 | 5369962 | 332607 | 1.39E-01 | 0.2 | 1.16 |
| RP_POS_256_19061a8_912 | | 256.1906 | 8.91 | | RP | POS | | | 359017 | 43479 | 367778 | 43061 | 1.39E-01 | 0.2 | 1.29 |
| HILIC_NEG_179_05646a5_122 | Glucose | 179.0565 | 5.12 | | HILIC | NEG | 59.0132(100), 71.013(100) | 2 | 4.1E+07 | 9549613 | 4.3E+07 | 1.4E+07 | 1.39E-01 | 0.21 | 0.97 |
| RP_POS_196_00287a0_637 | | 196.0029 | 0.64 | | RP | POS | | | 525727 | 224453 | 479616 | 230049 | 1.39E-01 | -0.2 | 0.97 |
| RP_POS_590_32135a10_284 | | 590.3214 | 10.28 | | RP | POS | | | 1.1E+07 | 3154591 | 1E+07 | 3376830 | 1.39E-01 | -0.2 | 1.06 |
| RP_POS_599_43433a10_96 | | 599.4343 | 10.96 | | RP | POS | | | 1064220 | 2299707 | 676092 | 1508120 | 1.39E-01 | -0.2 | 0.88 |
| RP_POS_709_77631a6_032 | | 709.7763 | 6.03 | | RP | POS | | | 2236345 | 1139146 | 2012291 | 1074369 | 1.39E-01 | -0.2 | 1.25 |
| RP_POS_749_3822a4_858 | | 749.3822 | 4.86 | | RP | POS | | | 4188086 | 2270511 | 3710861 | 2415149 | 1.39E-01 | -0.2 | 1.29 |
| RP_POS_334_89471a15_406 | | 334.8947 | 15.41 | | RP | POS | | | 466330 | 192909 | 517742 | 295682 | 1.40E-01 | 0.21 | 1.36 |
| RP_POS_221_15306a7_236 | | 221.1531 | 7.24 | | RP | POS | 221.1531(100) | | 1786488 | 145226 | 1815100 | 138348 | 1.40E-01 | 0.2 | 0.85 |
| RP_NEG_474_7724a15_394 | | 474.7724 | 15.39 | | RP | NEG | | | 408240 | 147954 | 376691 | 162247 | 1.40E-01 | -0.2 | 1.08 |
| RP_POS_1002_6593a10_488 | | 1002.6593 | 10.49 | | RP | POS | | | 6041074 | 838638 | 5869593 | 856236 | 1.40E-01 | -0.2 | 0.96 |
| RP_POS_336_31079a10_519 | | 336.3108 | 10.52 | | RP | POS | | | 811300 | 204854 | 772054 | 183872 | 1.40E-01 | -0.2 | 1.13 |
| RP_NEG_367_3576a13_556 | Lignoceric acid | 367.3576 | 13.56 | | RP | NEG | 367.3565(100) | 2 | 332839 | 152749 | 301726 | 155659 | 1.41E-01 | -0.20 | 1.41 |
| RP_NEG_396_86832a15_362 | | 396.8683 | 15.36 | | RP | NEG | | | 2180412 | 203709 | 2210674 | 79268 | 1.41E-01 | 0.21 | 0.98 |
| RP_NEG_595_28973a10_807 | | 595.2897 | 10.81 | | RP | NEG | | | 441112 | 386302 | 549186 | 634555 | 1.41E-01 | 0.21 | 0.99 |
| HILIC_POS_388_31635a3_08 | | 388.3164 | 3.08 | | HILIC | POS | | | 339224 | 101291 | 359910 | 103538 | 1.41E-01 | 0.2 | 0.94 |
| HILIC_POS_582_29663a1_151 | | 582.2966 | 1.15 | | HILIC | POS | | | 159941 | 67427.7 | 173714 | 69077.5 | 1.41E-01 | 0.2 | 0.88 |
| RP_NEG_530_26959a13_289 | | 530.2696 | 13.29 | | RP | NEG | | | 750910 | 141958 | 780571 | 151390 | 1.41E-01 | 0.2 | 1.21 |
| RP_POS_187_08134a15_33 | | 187.0813 | 15.33 | | RP | POS | | | 1452628 | 343660 | 1523901 | 361824 | 1.41E-01 | 0.2 | 0.94 |
| RP_POS_568_73322a10_291 | | 568.7332 | 10.29 | | RP | POS | | | 686048 | 376100 | 768818 | 435901 | 1.41E-01 | 0.2 | 1.08 |
| RP_POS_161_05974a6_702 | 6-Methylcoumarin | 161.0597 | 6.70 | | RP | POS | 161.0592(100), 105.0705(30) | 2 | 1299158 | 239660 | 1500647 | 1332527 | 1.41E-01 | 0.26 | 1.31 |
| RP_POS_339_1777a8_795 | Picrinine | 339.1777 | 8.80 | | RP | POS | 339.1809(100) | 2 | 1817685 | 1088929 | 2051816 | 1219340 | 1.41E-01 | 0.20 | 1.29 |
| HILIC_NEG_125_87312a3_241 | | 125.8731 | 3.24 | | HILIC | NEG | | | 216650 | 149390 | 189122 | 124079 | 1.41E-01 | -0.2 | 1.05 |
| RP_NEG_131_0714a4_509 | | 131.0714 | 4.51 | | RP | NEG | | | 603125 | 237008 | 556610 | 224342 | 1.41E-01 | -0.2 | 0.88 |
| RP_NEG_162_89352a14_335 | | 162.8935 | 14.34 | | RP | NEG | | | 2046823 | 592401 | 1921871 | 639616 | 1.41E-01 | -0.2 | 0.9 |
| RP_NEG_387_63165a15_5 | | 387.6317 | 15.50 | | RP | NEG | | | 210543 | 33003.5 | 204205 | 29957.6 | 1.41E-01 | -0.2 | 0.99 |
| RP_NEG_801_08124a9_197 | | 801.0812 | 9.20 | | RP | NEG | | | 1039443 | 77948.5 | 1023756 | 77473.8 | 1.41E-01 | -0.2 | 1.01 |
| RP_POS_405_25122a11_596 | | 405.2512 | 11.60 | | RP | POS | | | 4271021 | 591145 | 4153844 | 570888 | 1.41E-01 | -0.2 | 1.23 |
| RP_POS_406_32889a11_049 | | 406.3289 | 11.05 | | RP | POS | | | 8484196 | 472021 | 8384110 | 514384 | 1.41E-01 | -0.2 | 0.99 |
| RP_POS_665_44482a9_424 | | 665.4448 | 9.42 | | RP | POS | | | 387092 | 286701 | 336352 | 216867 | 1.41E-01 | -0.2 | 1.12 |
| HILIC_NEG_201_8591a5_41 | | 201.8591 | 5.41 | | HILIC | NEG | | | 970535 | 355315 | 1053485 | 453726 | 1.42E-01 | 0.21 | 0.93 |
| RP_POS_250_10332a2_503 | | 250.1033 | 2.50 | | RP | POS | | | 335036 | 153057 | 373270 | 216418 | 1.42E-01 | 0.21 | 0.86 |
| RP_POS_561_70038a10_43 | | 561.7004 | 10.43 | | RP | POS | | | 193916 | 89842.2 | 218575 | 144208 | 1.42E-01 | 0.21 | 0.97 |
| RP_NEG_241_18132a9_588 | | 241.1813 | 9.59 | | RP | NEG | | | 656460 | 546262 | 767491 | 557007 | 1.42E-01 | 0.2 | 1.16 |
| RP_POS_243_12251a8_795 | | 243.1225 | 8.80 | | RP | POS | | | 691500 | 400240 | 778592 | 459075 | 1.42E-01 | 0.2 | 1.3 |
| RP_POS_723_83551a11_54 | | 723.8355 | 11.54 | | RP | POS | | | 191897 | 80865.5 | 209842 | 95747.9 | 1.42E-01 | 0.2 | 0.95 |
| RP_POS_265_17926a8_979 | | 265.1793 | 8.98 | | RP | POS | | | 1481665 | 341802 | 1414361 | 327456 | 1.42E-01 | -0.2 | 0.83 |
| RP_POS_431_28055a12_107 | | 431.2806 | 12.11 | | RP | POS | | | 337498 | 137707 | 311319 | 123262 | 1.42E-01 | -0.2 | 1.37 |
| RP_POS_529_30341a13_007 | | 529.3034 | 13.01 | | RP | POS | | | 217778 | 141295 | 190694 | 128393 | 1.42E-01 | -0.2 | 1.16 |
| RP_POS_971_60632a10_477 | | 971.6063 | 10.48 | | RP | POS | | | 861442 | 295952 | 800562 | 307191 | 1.42E-01 | -0.2 | 1.12 |
| HILIC_POS_336_18387a3_023 | | 336.1839 | 3.02 | | HILIC | POS | | | 2553604 | 5295619 | 1782675 | 1791341 | 1.42E-01 | -0.22 | 0.91 |
| RP_NEG_367_15848a7_627 | | 367.1585 | 7.63 | | RP | NEG | 96.9599(100), 367.1552(90) | | 859603 | 725265 | 1004891 | 721852 | 1.43E-01 | 0.2 | 0.89 |
| HILIC_NEG_61_9876a5_118 | | 61.9876 | 5.12 | | HILIC | NEG | | | 6331681 | 2175811 | 6812422 | 2561420 | 1.43E-01 | 0.2 | 1.02 |
| RP_POS_388_2037a10_935 | | 388.2037 | 10.94 | | RP | POS | | | 100323 | 70680.8 | 116418 | 87245.5 | 1.43E-01 | 0.2 | 1.2 |
| RP_POS_413_7937a10_956 | | 413.7937 | 10.96 | | RP | POS | | | 278897 | 135201 | 306353 | 138089 | 1.43E-01 | 0.2 | 1.14 |
| RP_POS_718_57501a14_035 | | 718.5750 | 14.04 | | RP | POS | | | 862390 | 923896 | 1049727 | 942599 | 1.43E-01 | 0.2 | 1.16 |
| HILIC_POS_279_17026a2_275 | | 279.1703 | 2.28 | | HILIC | POS | | | 190691 | 239204 | 146573 | 202345 | 1.43E-01 | -0.2 | 1.05 |
| RP_NEG_192_98875a0_551 | | 192.9888 | 0.55 | | RP | NEG | | | 1186782 | 395341 | 1103678 | 429627 | 1.43E-01 | -0.2 | 0.99 |
| RP_POS_510_39197a11_199 | | 510.3920 | 11.20 | | RP | POS | | | 3608252 | 1717887 | 3280909 | 1553702 | 1.43E-01 | -0.2 | 0.86 |
| RP_NEG_311_16901a10_075 | | 311.1690 | 10.08 | | RP | NEG | 311.1699(100) | | 2310891 | 975185 | 2530462 | 1190782 | 1.44E-01 | 0.2 | 0.88 |
| HILIC_NEG_169_96793a0_941 | | 169.9679 | 0.94 | | HILIC | NEG | | | 47646.6 | 30669.6 | 54920.9 | 40435.6 | 1.44E-01 | 0.2 | 0.95 |
| RP_POS_294_91995a15_368 | | 294.9200 | 15.37 | | RP | POS | | | 6553329 | 3774584 | 7324047 | 3901920 | 1.44E-01 | 0.2 | 0.99 |
| RP_POS_361_16641a10_926 | | 361.1664 | 10.93 | | RP | POS | | | 1312296 | 641166 | 1434716 | 583492 | 1.44E-01 | 0.2 | 1.12 |
| RP_NEG_461_59424a15_493 | | 461.5942 | 15.49 | | RP | NEG | | | 406289 | 37660.9 | 398724 | 37878.9 | 1.44E-01 | -0.2 | 0.91 |
| RP_POS_177_0546a8_615 | | 177.0546 | 8.62 | | RP | POS | | | 779841 | 86990.8 | 761855 | 92310.6 | 1.44E-01 | -0.2 | 1.03 |
| RP_POS_610_92377a10_641 | | 610.9238 | 10.64 | | RP | POS | | | 380856 | 233580 | 332506 | 247658 | 1.44E-01 | -0.2 | 1.15 |
| RP_POS_263_23694a10_666 | | 263.2369 | 10.67 | | RP | POS | | | 4250936 | 2066798 | 4851937 | 3617239 | 1.45E-01 | 0.21 | 1.11 |
| HILIC_NEG_174_83932a3_226 | | 174.8393 | 3.23 | | HILIC | NEG | | | 1998470 | 780342 | 2143689 | 679630 | 1.45E-01 | 0.2 | 0.81 |
| RP_NEG_661_39917a10_96 | | 661.3992 | 10.96 | | RP | NEG | | | 202073 | 157336 | 233026 | 152572 | 1.45E-01 | 0.2 | 1.2 |
| RP_POS_249_13303a4_056 | | 249.1330 | 4.06 | | RP | POS | | | 679930 | 152291 | 708206 | 132040 | 1.45E-01 | 0.2 | 0.95 |
| RP_POS_566_32141a10_181 | | 566.3214 | 10.18 | | RP | POS | | | 3808964 | 1355388 | 4077036 | 1332253 | 1.45E-01 | 0.2 | 0.93 |
| HILIC_POS_244_1545a1_434 | Tiglylcarnitine | 244.1545 | 1.43 | | HILIC | POS | 85.0291(100), 185.0811(20) | 1 | 842660 | 304987 | 922434 | 463474 | 1.45E-01 | 0.21 | 1.19 |
| RP_NEG_405_19235a6_738 | Cortisone | 405.1924 | 6.74 | | RP | NEG | 329.1781(100), 137.0615(15), 301.1803(25) | 2 | 1028006 | 243263 | 1076686 | 244382 | 1.45E-01 | 0.20 | 0.87 |
| RP_NEG_251_09267a7_33 | | 251.0927 | 7.33 | | RP | NEG | | | 2322442 | 99906.3 | 2300686 | 116140 | 1.45E-01 | -0.2 | 0.91 |
| RP_NEG_281_03598a0_679 | | 281.0360 | 0.68 | | RP | NEG | | | 292142 | 95085.8 | 274664 | 80648.1 | 1.45E-01 | -0.2 | 1.01 |
| RP_NEG_296_99783a7_321 | | 296.9978 | 7.32 | | RP | NEG | | | 1423531 | 729758 | 1275855 | 747908 | 1.45E-01 | -0.2 | 0.94 |
| RP_POS_163_07542a8_394 | | 163.0754 | 8.39 | | RP | POS | | | 1622487 | 637034 | 1500377 | 588472 | 1.45E-01 | -0.2 | 1.06 |
| RP_POS_285_13074a4_788 | | 285.1307 | 4.79 | | RP | POS | | | 242431 | 231730 | 203060 | 162169 | 1.45E-01 | -0.2 | 1.11 |
| RP_POS_424_2822a11_112 | | 424.2822 | 11.11 | | RP | POS | | | 403616 | 542566 | 313597 | 354051 | 1.45E-01 | -0.2 | 0.88 |
| RP_POS_619_91272a9_755 | | 619.9127 | 9.76 | | RP | POS | | | 790762 | 460486 | 707168 | 379486 | 1.45E-01 | -0.2 | 0.98 |
| RP_NEG_620_35547a10_87 | | 620.3555 | 10.87 | | RP | NEG | | | 350521 | 94830 | 369689 | 97406 | 1.46E-01 | 0.2 | 0.97 |
| RP_NEG_243_06197a0_678 | Pseudouridine | 243.0620 | 0.68 | | RP | NEG | 153.0311(95), 183.0405(15)243.0623(12) | 2 | 3662511 | 515882 | 3567521 | 441209 | 1.46E-01 | -0.20 | 0.98 |
| HILIC_POS_112_01827a0_927 | | 112.0183 | 0.93 | | HILIC | POS | | | 423846 | 280873 | 373694 | 224526 | 1.46E-01 | -0.2 | 0.99 |
| RP_NEG_166_96404a0_679 | | 166.9640 | 0.68 | | RP | NEG | | | 834516 | 112375 | 810197 | 129598 | 1.46E-01 | -0.2 | 0.89 |
| RP_POS_241_25261a11_996 | | 241.2526 | 12.00 | | RP | POS | | | 1345220 | 447213 | 1430753 | 414681 | 1.47E-01 | 0.2 | 1.12 |
| RP_POS_402_28497a10_389 | | 402.2850 | 10.39 | | RP | POS | | | 5138337 | 2594291 | 5697859 | 2986718 | 1.47E-01 | 0.2 | 1.32 |
| RP_POS_598_08459a10_266 | | 598.0846 | 10.27 | | RP | POS | | | 553785 | 277974 | 612980 | 313243 | 1.47E-01 | 0.2 | 1.02 |
| RP_POS_627_77075a10_243 | | 627.7708 | 10.24 | | RP | POS | | | 154956 | 83938.8 | 173689 | 102189 | 1.47E-01 | 0.2 | 0.92 |
| RP_POS_804_55066a12_549 | PC 38:7 (20:5_18:2) | 804.5507 | 12.55 | | RP | POS | 184.0743(100), 86.0954(20) | 1 | 1.8E+07 | 6835394 | 1.7E+07 | 6123666 | 1.47E-01 | -0.20 | 1.03 |
| RP_POS_255_05876a2_229 | | 255.0588 | 2.23 | | RP | POS | 255.0567(100) | | 3058514 | 546599 | 2956473 | 484057 | 1.47E-01 | -0.2 | 1.15 |
| HILIC_NEG_216_91141a5_652 | | 216.9114 | 5.65 | | HILIC | NEG | | | 181898 | 39540.4 | 175343 | 26172.9 | 1.47E-01 | -0.2 | 0.92 |
| HILIC_POS_265_99426a0_927 | | 265.9943 | 0.93 | | HILIC | POS | | | 236469 | 108788 | 215834 | 99206.7 | 1.47E-01 | -0.2 | 1.02 |
| RP_NEG_135_02991a0_548 | | 135.0299 | 0.55 | | RP | NEG | | | 4880684 | 1613821 | 4555130 | 1657846 | 1.47E-01 | -0.2 | 1.03 |
| RP_NEG_297_24335a10_158 | | 297.2434 | 10.16 | | RP | NEG | | | 8056519 | 1173058 | 7822195 | 1182796 | 1.47E-01 | -0.2 | 1.19 |
| RP_POS_235_24173a10_156 | | 235.2417 | 10.16 | | RP | POS | | | 373537 | 79383.8 | 358074 | 76539 | 1.47E-01 | -0.2 | 1.06 |
| RP_POS_392_22177a11_143 | | 392.2218 | 11.14 | | RP | POS | | | 987292 | 365694 | 918262 | 331518 | 1.47E-01 | -0.2 | 1.14 |
| RP_POS_698_78638a6_032 | | 698.7864 | 6.03 | | RP | POS | | | 2.3E+07 | 1.3E+07 | 2.1E+07 | 1.2E+07 | 1.47E-01 | -0.2 | 1.24 |
| RP_POS_834_60028a13_788 | | 834.6003 | 13.79 | | RP | POS | | | 1.2E+08 | 3.6E+07 | 1.1E+08 | 3.2E+07 | 1.47E-01 | -0.2 | 1.11 |
| RP_POS_955_56879a11_433 | | 955.5688 | 11.43 | | RP | POS | | | 138212 | 96952.8 | 163315 | 146796 | 1.48E-01 | 0.21 | 0.92 |
| HILIC_POS_153_03281a7_419 | | 153.0328 | 7.42 | | HILIC | POS | | | 1661243 | 228075 | 1707042 | 232889 | 1.48E-01 | 0.2 | 1.01 |
| RP_POS_422_17578a9_821 | | 422.1758 | 9.82 | | RP | POS | | | 1127399 | 979409 | 1333797 | 1091795 | 1.48E-01 | 0.2 | 1.06 |
| RP_POS_812_61584a14_443 | | 812.6158 | 14.44 | | RP | POS | | | 6.8E+07 | 3.5E+07 | 7.5E+07 | 3.4E+07 | 1.48E-01 | 0.2 | 1.03 |
| RP_POS_868_50562a13_797 | | 868.5056 | 13.80 | | RP | POS | | | 850209 | 497057 | 963673 | 628080 | 1.48E-01 | 0.2 | 0.89 |
| RP_NEG_515_25732a12_106 | | 515.2573 | 12.11 | | RP | NEG | | | 148864 | 54468 | 137474 | 59763.5 | 1.48E-01 | -0.2 | 1.39 |
| RP_NEG_822_62421a10_926 | | 822.6242 | 10.93 | | RP | NEG | | | 871182 | 150961 | 840294 | 159466 | 1.48E-01 | -0.2 | 1.05 |
| RP_POS_231_11642a15_327 | | 231.1164 | 15.33 | | RP | POS | | | 4234098 | 2325942 | 3788333 | 2178062 | 1.48E-01 | -0.2 | 1.02 |
| RP_POS_512_32123a8_872 | | 512.3212 | 8.87 | | RP | POS | | | 1.3E+07 | 4619100 | 1.2E+07 | 3820333 | 1.48E-01 | -0.2 | 0.9 |
| RP_POS_622_26569a10_315 | | 622.2657 | 10.32 | | RP | POS | | | 463539 | 199143 | 424148 | 198209 | 1.48E-01 | -0.2 | 0.9 |
| RP_POS_781_0531a12_928 | | 781.0531 | 12.93 | | RP | POS | | | 533187 | 910965 | 792536 | 1567043 | 1.49E-01 | 0.21 | 0.94 |
| RP_NEG_658_33441a10_513 | | 658.3344 | 10.51 | | RP | NEG | | | 406691 | 105192 | 429480 | 122862 | 1.49E-01 | 0.2 | 1.03 |
| RP_POS_270_31516a9_658 | | 270.3152 | 9.66 | | RP | POS | | | 1267684 | 243690 | 1319265 | 274252 | 1.49E-01 | 0.2 | 0.95 |
| RP_POS_608_71033a11_277 | | 608.7103 | 11.28 | | RP | POS | | | 115894 | 56739.9 | 128080 | 65386.6 | 1.49E-01 | 0.2 | 1.05 |
| RP_POS_806_56635a13_382 | | 806.5664 | 13.38 | | RP | POS | | | 1.2E+08 | 5.1E+07 | 1.3E+08 | 6.5E+07 | 1.49E-01 | 0.2 | 0.93 |
| HILIC_POS_388_31613a1_359 | | 388.3161 | 1.36 | | HILIC | POS | | | 268895 | 121638 | 244489 | 124960 | 1.49E-01 | -0.2 | 1.07 |
| RP_NEG_368_97745a8_239 | | 368.9775 | 8.24 | | RP | NEG | | | 1314085 | 687772 | 1180826 | 662071 | 1.49E-01 | -0.2 | 0.91 |
| RP_NEG_383_31754a11_396 | | 383.3175 | 11.40 | | RP | NEG | | | 299149 | 24480.2 | 294118 | 26194.8 | 1.49E-01 | -0.2 | 0.88 |
| RP_POS_313_16248a5_068 | | 313.1625 | 5.07 | | RP | POS | | | 187437 | 112972 | 168097 | 82911.5 | 1.49E-01 | -0.2 | 1.23 |
| RP_POS_406_29102a8_289 | | 406.2910 | 8.29 | | RP | POS | | | 217239 | 152551 | 189237 | 131436 | 1.49E-01 | -0.2 | 1.02 |
| RP_POS_600_92426a10_136 | | 600.9243 | 10.14 | | RP | POS | | | 2364420 | 1088690 | 2165137 | 935481 | 1.49E-01 | -0.2 | 0.99 |
| RP_POS_725_74438a6_032 | | 725.7444 | 6.03 | | RP | POS | | | 752263 | 502524 | 663518 | 397833 | 1.49E-01 | -0.2 | 1.2 |
| HILIC_POS_206_10194a3_116 | | 206.1019 | 3.12 | | HILIC | POS | | | 101815 | 36433 | 111421 | 56988.4 | 1.50E-01 | 0.21 | 0.97 |
| HILIC_POS_400_34222a0_609 | AC 16:0 (Palmitoylcarnitine) | 400.3422 | 0.61 | | HILIC | POS | 85.0291(100), 400.3391(60) | 1 | 831557 | 229312 | 877620 | 237058 | 1.50E-01 | 0.20 | 1.31 |
| RP_NEG_385_11819a10_693 | | 385.1182 | 10.69 | | RP | NEG | | | 95375.1 | 56468.8 | 108220 | 71635.6 | 1.50E-01 | 0.2 | 0.97 |
| RP_POS_324_28973a11_117 | | 324.2897 | 11.12 | | RP | POS | | | 461278 | 274212 | 516549 | 284837 | 1.50E-01 | 0.2 | 1.32 |
| RP_POS_534_29608a10_495 | | 534.2961 | 10.50 | | RP | POS | | | 7491125 | 1788699 | 7862797 | 1962827 | 1.50E-01 | 0.2 | 0.96 |
| RP_POS_577_74481a10_543 | | 577.7448 | 10.54 | | RP | POS | | | 457712 | 285598 | 520181 | 340551 | 1.50E-01 | 0.2 | 1.01 |
| HILIC_POS_553_409a4_227 | | 553.4090 | 4.23 | | HILIC | POS | | | 204215 | 179123 | 169838 | 169615 | 1.50E-01 | -0.2 | 0.85 |
| RP_POS_625_92969a10_459 | | 625.9297 | 10.46 | | RP | POS | | | 1283610 | 527115 | 1183750 | 487449 | 1.50E-01 | -0.2 | 0.97 |
| RP_POS_842_56586a13_382 | | 842.5659 | 13.38 | | RP | POS | | | 1611435 | 1404898 | 1357341 | 1178351 | 1.50E-01 | -0.2 | 1.02 |
| RP_NEG_457_2356a11_056 | | 457.2356 | 11.06 | | RP | NEG | | | 1158217 | 4402118 | 2434016 | 7814229 | 1.51E-01 | 0.21 | 0.87 |
| HILIC_NEG_464_80487a2_998 | | 464.8049 | 3.00 | | HILIC | NEG | | | 795078 | 149721 | 825076 | 154972 | 1.51E-01 | 0.2 | 0.84 |
| RP_NEG_309_1712a8_245 | | 309.1712 | 8.25 | | RP | NEG | | | 1983454 | 87083.1 | 2000851 | 89275.9 | 1.51E-01 | 0.2 | 1.23 |
| RP_POS_1065_6853a10_608 | | 1065.6853 | 10.61 | | RP | POS | | | 9436091 | 3997017 | 1E+07 | 4141508 | 1.51E-01 | 0.2 | 0.97 |

**Supplemantary table 1**: The non-targeted metabolomics results ordered by the VIP value (p value < 0.0003 are shaded dark green, p value < 0.05 are shaded light green, d values below 0 are blue and above 0 are red)

| | | | | | | | | | Cases | | Controls | | Welch test | Cohen's | |
|---|---|---|---|---|---|---|---|---|---|---|---|---|---|---|---|
| Feature_ID | Metabolite_name | Average_Mz | Average_RT_min_ | HMDB ID | RP_HILIC | POS_NEG | Cureted_MSMS | ID_level | Mean | SD | Mean2 | SD3 | p | d | VIP |
| RP_POS_373_06064a0_585 | | 373.0606 | 0.59 | | RP | POS | | | 445733 | 176877 | 482716 | 196512 | 1.51E-01 | 0.2 | 0.88 |
| RP_POS_312_63351a10_284 | | 312.6335 | 10.28 | | RP | POS | | | 494672 | 183721 | 457324 | 195046 | 1.51E-01 | -0.2 | 1.03 |
| RP_POS_345_19373a11_143 | | 345.1937 | 11.14 | | RP | POS | | | 6395434 | 1035249 | 6193448 | 1019688 | 1.51E-01 | -0.2 | 1.06 |
| RP_POS_506_38055a10_43 | | 506.3806 | 10.43 | | RP | POS | | | 1641573 | 688842 | 1505624 | 691000 | 1.51E-01 | -0.2 | 0.92 |
| HILIC_NEG_180_98926a2_266 | | 180.9893 | 2.27 | | HILIC | NEG | | | 641159 | 181308 | 677751 | 190711 | 1.52E-01 | 0.2 | 1.01 |
| HILIC_NEG_135_02998a5_654 | | 135.0300 | 5.65 | | HILIC | NEG | | | 912139 | 367649 | 840798 | 359651 | 1.52E-01 | -0.2 | 0.97 |
| HILIC_NEG_475_52985a1_41 | | 475.5299 | 1.41 | | HILIC | NEG | | | 108436 | 27274.1 | 103428 | 23918.9 | 1.52E-01 | -0.2 | 0.87 |
| RP_POS_383_29529a11_145 | | 383.2953 | 11.15 | | RP | POS | | | 1415098 | 287580 | 1361387 | 261756 | 1.52E-01 | -0.2 | 0.9 |
| RP_POS_483_29236a8_844 | | 483.2924 | 8.84 | | RP | POS | | | 3.2E+07 | 2.2E+07 | 2.8E+07 | 1.7E+07 | 1.52E-01 | -0.2 | 1.21 |
| RP_POS_517_33063a9_117 | | 517.3306 | 9.12 | | RP | POS | | | 1851745 | 5628706 | 929053 | 3725145 | 1.52E-01 | -0.2 | 0.84 |
| HILIC_NEG_217_02988a5_121 | | 217.0299 | 5.12 | | HILIC | NEG | | | 9755292 | 3765502 | 1.1E+07 | 6879417 | 1.53E-01 | 0.21 | 0.92 |
| RP_POS_238_06763a13_298 | | 238.0676 | 13.30 | | RP | POS | | | 4336085 | 1667687 | 4696348 | 1970020 | 1.53E-01 | 0.2 | 0.99 |
| RP_POS_592_26581a10_547 | | 592.2658 | 10.55 | | RP | POS | | | 728648 | 576075 | 839843 | 560614 | 1.53E-01 | 0.2 | 1.32 |
| RP_NEG_239_76106a15_494 | | 239.7611 | 15.49 | | RP | NEG | | | 1.1E+07 | 238079 | 1.1E+07 | 245084 | 1.53E-01 | -0.2 | 0.91 |
| RP_POS_225_22095a10_095 | | 225.2210 | 10.10 | | RP | POS | | | 1100712 | 138848 | 1073369 | 140398 | 1.53E-01 | -0.2 | 1.05 |
| RP_POS_275_08914a7_72 | | 275.0891 | 7.72 | | RP | POS | | | 1588889 | 99449.2 | 1568883 | 104219 | 1.53E-01 | -0.2 | 1.23 |
| RP_POS_398_21558a9_589 | | 398.2156 | 9.59 | | RP | POS | | | 1395213 | 746429 | 1256626 | 673533 | 1.53E-01 | -0.2 | 1.26 |
| RP_POS_411_38275a11_608 | | 411.3828 | 11.61 | | RP | POS | | | 520910 | 142510 | 494085 | 132256 | 1.53E-01 | -0.2 | 1.14 |
| RP_POS_419_16687a7_783 | | 419.1669 | 7.78 | | RP | POS | | | 432606 | 43644 | 424496 | 39447.6 | 1.53E-01 | -0.2 | 0.97 |
| RP_POS_430_24353a4_317 | | 430.2435 | 4.32 | | RP | POS | | | 31441.9 | 46937.6 | 24241.5 | 25147 | 1.53E-01 | -0.2 | 0.94 |
| RP_POS_377_26608a10_67 | | 377.2661 | 10.67 | | RP | POS | | | 1.5E+07 | 9913299 | 1.8E+07 | 1.7E+07 | 1.54E-01 | 0.21 | 1.1 |
| HILIC_POS_194_02223a4_632 | | 194.0222 | 4.63 | | HILIC | POS | | | 684803 | 326076 | 743157 | 273228 | 1.54E-01 | 0.19 | 0.88 |
| RP_POS_288_25354a9_325 | Lauryl diethanolamide | 288.2535 | 9.33 | | RP | POS | 106.0872(100), 227.2014(30) | 2 | 5227978 | 322678 | 5162531 | 345013 | 1.54E-01 | -0.20 | 1.25 |
| HILIC_NEG_61_98755a0_931 | | 61.9876 | 0.93 | | HILIC | NEG | | | 8653723 | 2451407 | 8183341 | 2366420 | 1.54E-01 | -0.2 | 1 |
| RP_NEG_551_12756a8_596 | | 551.1276 | 8.60 | | RP | NEG | | | 618491 | 28731.4 | 612768 | 29809.4 | 1.54E-01 | -0.2 | 0.91 |
| RP_POS_269_24734a10_531 | | 269.2473 | 10.53 | | RP | POS | | | 926487 | 149884 | 897992 | 142336 | 1.54E-01 | -0.2 | 1.03 |
| RP_POS_534_31665a10_062 | | 534.3167 | 10.06 | | RP | POS | | | 981181 | 336353 | 915735 | 333052 | 1.54E-01 | -0.2 | 0.97 |
| HILIC_POS_546_3559a1_165 | | 546.3559 | 1.17 | | HILIC | POS | | | 7953513 | 2352829 | 8421638 | 2437199 | 1.55E-01 | 0.2 | 0.95 |
| RP_NEG_159_89404a0_633 | | 159.8940 | 0.63 | | RP | NEG | | | 462900 | 239726 | 518563 | 319324 | 1.55E-01 | 0.2 | 0.96 |
| RP_POS_402_28497a10_242 | | 402.2850 | 10.24 | | RP | POS | | | 1116592 | 508515 | 1218806 | 536429 | 1.55E-01 | 0.2 | 1.25 |
| RP_POS_555_71051a10_302 | | 555.7105 | 10.30 | | RP | POS | | | 400037 | 237091 | 457434 | 334922 | 1.55E-01 | 0.2 | 1.18 |
| RP_POS_591_98816a11_398 | | 591.9882 | 11.40 | | RP | POS | | | 130829 | 67544.4 | 146011 | 85654 | 1.55E-01 | 0.2 | 0.96 |
| RP_POS_626_35333a8_777 | | 626.3533 | 8.78 | | RP | POS | | | 1440346 | 1587176 | 1768757 | 1759668 | 1.55E-01 | 0.2 | 0.81 |
| RP_NEG_480_52722a15_496 | | 480.5272 | 15.50 | | RP | NEG | | | 585351 | 38210.7 | 592829 | 38543.9 | 1.55E-01 | 0.19 | 0.9 |
| RP_POS_369_16547a5_596 | | 369.1655 | 5.60 | | RP | POS | | | 463000 | 192366 | 429569 | 151569 | 1.55E-01 | -0.19 | 0.84 |
| HILIC_POS_697_33777a1_889 | | 697.3378 | 1.89 | | HILIC | POS | | | 164039 | 28799.4 | 158063 | 32076.4 | 1.55E-01 | -0.2 | 0.92 |
| RP_NEG_315_25473a9_073 | | 315.2547 | 9.07 | | RP | NEG | | | 481683 | 38152.5 | 474103 | 39462 | 1.55E-01 | -0.2 | 0.97 |
| RP_POS_622_86218a11_536 | | 622.8622 | 11.54 | | RP | POS | | | 154306 | 73513.3 | 169638 | 83242.6 | 1.56E-01 | 0.2 | 0.91 |
| RP_POS_667_63177a11_517 | | 667.6318 | 11.52 | | RP | POS | | | 134138 | 72640 | 151508 | 101088 | 1.56E-01 | 0.2 | 0.91 |
| RP_POS_820_58142a13_97 | | 820.5814 | 13.97 | | RP | POS | | | 2340292 | 1601238 | 2658871 | 1666103 | 1.56E-01 | 0.2 | 0.98 |
| RP_NEG_315_18188a8_796 | | 315.1819 | 8.80 | | RP | NEG | | | 490967 | 316365 | 554645 | 336781 | 1.56E-01 | 0.19 | 1.28 |
| RP_POS_701_42993a12_03 | | 701.4299 | 12.03 | | RP | POS | | | 499046 | 92941.7 | 515896 | 81073.7 | 1.56E-01 | 0.19 | 0.93 |
| RP_POS_810_60028a13_632 | PC(18:0/20:4) | 810.6003 | 13.63 | | RP | POS | 184.0745(100) | 1 | 4.8E+07 | 1.9E+07 | 5.1E+07 | 1.7E+07 | 1.56E-01 | 0.19 | 0.98 |
| HILIC_POS_367_99097a6_308 | | 367.9910 | 6.31 | | HILIC | POS | | | 557804 | 240713 | 511394 | 236791 | 1.56E-01 | -0.19 | 1.02 |
| RP_POS_169_08607a3_892 | | 169.0861 | 3.89 | | RP | POS | | | 1315209 | 652506 | 1195211 | 584406 | 1.56E-01 | -0.19 | 0.97 |
| RP_POS_375_32129a10_264 | | 375.3213 | 10.26 | | RP | POS | | | 459590 | 94471.5 | 441242 | 94103.7 | 1.56E-01 | -0.19 | 1.1 |
| RP_POS_422_32513a11_408 | | 422.3251 | 11.41 | | RP | POS | | | 5681035 | 955704 | 5515797 | 749351 | 1.56E-01 | -0.19 | 0.97 |
| RP_POS_441_32858a9_862 | | 441.3286 | 9.86 | | RP | POS | | | 4114733 | 2017385 | 3745042 | 1794212 | 1.56E-01 | -0.19 | 0.95 |
| RP_POS_447_82111a9_615 | | 447.8211 | 9.62 | | RP | POS | | | 1634977 | 951970 | 1470979 | 739408 | 1.56E-01 | -0.19 | 0.94 |
| RP_POS_347_25266a10_645 | | 347.2527 | 10.65 | | RP | POS | | | 458861 | 396898 | 558665 | 594964 | 1.57E-01 | 0.2 | 0.94 |
| RP_NEG_800_5462a12_708 | | 800.5462 | 12.71 | | RP | NEG | | | 9442422 | 2890614 | 1E+07 | 3075652 | 1.57E-01 | 0.19 | 0.89 |
| HILIC_POS_406_10413a5_674 | | 406.1041 | 5.67 | | HILIC | POS | | | 74105.3 | 23802.3 | 69748.9 | 21290.3 | 1.57E-01 | -0.19 | 0.87 |
| RP_NEG_244_92404a0_572 | | 244.9240 | 0.57 | | RP | NEG | | | 646947 | 139424 | 620432 | 134532 | 1.57E-01 | -0.19 | 0.95 |
| RP_NEG_435_21094a10_916 | | 435.2109 | 10.92 | | RP | NEG | | | 42226.6 | 116812 | 25989.1 | 36527.9 | 1.57E-01 | -0.21 | 0.76 |
| RP_POS_476_01114a10_233 | | 476.0111 | 10.23 | | RP | POS | | | 414419 | 213922 | 475060 | 376520 | 1.58E-01 | 0.21 | 0.89 |
| RP_POS_286_88208a15_437 | | 286.8821 | 15.44 | | RP | POS | | | 1872824 | 514957 | 1990588 | 678153 | 1.58E-01 | 0.2 | 1.15 |
| HILIC_POS_192_06563a4_114 | | 192.0656 | 4.11 | | HILIC | POS | | | 346985 | 210990 | 387580 | 208704 | 1.58E-01 | 0.19 | 0.88 |
| RP_POS_234_09698a0_683 | | 234.0970 | 0.68 | | RP | POS | | | 192325 | 174805 | 228520 | 197087 | 1.58E-01 | 0.19 | 1.02 |
| RP_NEG_556_32471a10_062 | | 556.3247 | 10.06 | | RP | NEG | | | 2425588 | 760600 | 2279012 | 753845 | 1.58E-01 | -0.19 | 0.99 |
| RP_POS_231_15919a8_844 | | 231.1592 | 8.84 | | RP | POS | | | 2.4E+08 | 8.4E+07 | 2.2E+08 | 7.3E+07 | 1.58E-01 | -0.19 | 0.94 |
| RP_POS_590_32135a10_177 | | 590.3214 | 10.18 | | RP | POS | | | 1330643 | 578112 | 1226336 | 503339 | 1.58E-01 | -0.19 | 0.9 |
| RP_POS_732_54803a9_94 | | 732.5480 | 9.94 | | RP | POS | | | 3617493 | 1533384 | 3346456 | 1278206 | 1.58E-01 | -0.19 | 1.1 |
| RP_POS_253_11809a0_694 | | 253.1181 | 0.69 | | RP | POS | | | 769732 | 377538 | 855978 | 497777 | 1.59E-01 | 0.2 | 1.04 |
| RP_POS_357_29977a11_202 | | 357.2998 | 11.20 | | RP | POS | | | 2049723 | 1069958 | 2305943 | 1509604 | 1.59E-01 | 0.2 | 1.35 |
| RP_POS_379_28183a11_202 | | 379.2818 | 11.20 | | RP | POS | | | 3144409 | 1474820 | 3495728 | 2062724 | 1.59E-01 | 0.2 | 1.32 |
| RP_POS_144_99747a15_746 | | 144.9975 | 15.75 | | RP | POS | | | 4991433 | 3153840 | 5637586 | 3501226 | 1.59E-01 | 0.19 | 0.96 |
| RP_NEG_334_78638a15_438 | | 334.7864 | 15.44 | | RP | NEG | | | 5989753 | 1780680 | 5678736 | 1451263 | 1.59E-01 | -0.19 | 1.03 |
| RP_NEG_452_92413a11_196 | | 452.9241 | 11.20 | | RP | NEG | | | 1067694 | 234562 | 1025276 | 206356 | 1.59E-01 | -0.19 | 0.8 |
| RP_POS_752_51953a12_588 | | 752.5195 | 12.59 | | RP | POS | | | 1.3E+07 | 5951244 | 1.2E+07 | 5743755 | 1.59E-01 | -0.19 | 0.89 |
| RP_NEG_309_10159a6_768 | | 309.1016 | 6.77 | | RP | NEG | | | 397562 | 273155 | 462366 | 381416 | 1.60E-01 | 0.2 | 0.89 |
| HILIC_NEG_170_97543a1_014 | | 170.9754 | 1.01 | | HILIC | NEG | | | 110906 | 33095.7 | 117280 | 33107.1 | 1.60E-01 | 0.19 | 1.08 |
| RP_POS_179_14296a8_633 | | 179.1430 | 8.63 | | RP | POS | | | 6816145 | 835641 | 6971294 | 780155 | 1.60E-01 | 0.19 | 1.14 |
| RP_POS_564_06262a10_155 | | 564.0626 | 10.16 | | RP | POS | | | 725924 | 385189 | 808085 | 458368 | 1.60E-01 | 0.19 | 0.92 |
| RP_POS_305_24744a10_927 | FA 20:4 (Arachidonic acid) | 305.2474 | 10.93 | | RP | POS | 305.2467(100), 121.102(40) | 1 | 7535773 | 3272066 | 8137559 | 2997802 | 1.60E-01 | 0.19 | 1.13 |
| RP_POS_179_1066a5_348 | Methyleugenol | 179.1066 | 5.35 | | RP | POS | 179.0641(100) | 2 | 1181590 | 155060 | 1149500 | 175497 | 1.60E-01 | -0.19 | 0.88 |
| HILIC_NEG_132_92361a0_933 | | 132.9236 | 0.93 | | HILIC | NEG | | | 863570 | 149418 | 834745 | 149925 | 1.60E-01 | -0.19 | 0.91 |
| HILIC_POS_172_09419a4_562 | | 172.0942 | 4.56 | | HILIC | POS | | | 2727668 | 2240039 | 2372064 | 1430936 | 1.60E-01 | -0.19 | 1.23 |
| RP_POS_302_86758a15_435 | | 302.8676 | 15.44 | | RP | POS | | | 287647 | 181141 | 257347 | 133951 | 1.60E-01 | -0.19 | 1 |
| RP_POS_315_25351a9_557 | | 315.2535 | 9.56 | | RP | POS | | | 1080880 | 119417 | 1059019 | 108352 | 1.60E-01 | -0.19 | 1 |
| RP_POS_635_49615a11_596 | | 635.4962 | 11.60 | | RP | POS | | | 947671 | 191855 | 910976 | 189751 | 1.60E-01 | -0.19 | 0.97 |
| RP_POS_765_37732a13_704 | | 765.3773 | 13.70 | | RP | POS | | | 509869 | 264519 | 458307 | 270977 | 1.60E-01 | -0.19 | 1.24 |
| RP_POS_573_29596a9_209 | | 573.2960 | 9.21 | | RP | POS | | | 1032870 | 1692248 | 776268 | 919288 | 1.60E-01 | -0.2 | 0.87 |
| RP_NEG_280_62234a8_05 | | 280.6223 | 8.05 | | RP | NEG | | | 9132005 | 7224099 | 1.1E+07 | 1.1E+07 | 1.61E-01 | 0.2 | 1.47 |
| RP_POS_304_29941a8_355 | | 304.2994 | 8.36 | | RP | POS | | | 1076257 | 304202 | 1143474 | 383459 | 1.61E-01 | 0.2 | 1.04 |
| HILIC_NEG_134_89478a1_35 | | 134.8948 | 1.35 | | HILIC | NEG | | | 2.3E+07 | 4986245 | 2.4E+07 | 3829412 | 1.61E-01 | 0.19 | 1.02 |
| RP_POS_297_08246a10_126 | | 297.0825 | 10.13 | | RP | POS | | | 356554 | 75654.6 | 371995 | 84312.3 | 1.61E-01 | 0.19 | 0.92 |
| RP_POS_548_37128a10_632 | | 548.3713 | 10.63 | | RP | POS | | | 1417906 | 439447 | 1505977 | 474321 | 1.61E-01 | 0.19 | 0.94 |
| HILIC_POS_136_11226a0_628 | C9H14N | 136.1123 | 0.63 | | HILIC | POS | 136.1122(100) | 2 | 3401940 | 182604 | 3438944 | 201026 | 1.61E-01 | 0.19 | 1.17 |
| HILIC_NEG_180_93452a5_651 | | 180.9345 | 5.65 | | HILIC | NEG | | | 118329 | 27053.8 | 114016 | 17570.9 | 1.61E-01 | -0.19 | 0.88 |
| RP_POS_290_23233a8_844 | | 290.2323 | 8.84 | | RP | POS | | | 504334 | 205435 | 467702 | 176867 | 1.61E-01 | -0.19 | 1.26 |
| RP_POS_471_22662a9_207 | | 471.2266 | 9.21 | | RP | POS | | | 190894 | 170040 | 162296 | 128526 | 1.61E-01 | -0.19 | 1.07 |
| HILIC_POS_287_1965a3_421 | | 287.1965 | 3.42 | | HILIC | POS | | | 4.9E+07 | 2.4E+08 | 1.4E+07 | 9.6E+07 | 1.61E-01 | -0.2 | 0.78 |
| RP_NEG_180_86206a15_449 | | 180.8621 | 15.45 | | RP | NEG | | | 3078772 | 975658 | 3289424 | 1192157 | 1.62E-01 | 0.19 | 1.01 |
| RP_POS_665_46558a11_434 | | 665.4656 | 11.43 | | RP | POS | | | 160338 | 88158 | 177681 | 92452.2 | 1.62E-01 | 0.19 | 0.94 |
| RP_POS_675_46906a11_266 | | 675.4691 | 11.27 | | RP | POS | | | 191844 | 132294 | 221166 | 168697 | 1.62E-01 | 0.19 | 0.83 |
| RP_POS_876_56903a14_103 | | 876.5690 | 14.10 | | RP | POS | | | 1.4E+07 | 5955231 | 1.5E+07 | 6351392 | 1.62E-01 | 0.19 | 0.84 |
| HILIC_POS_308_15259a4_402 | Tebuconazole | 308.1526 | 4.40 | | HILIC | POS | 308.1508(100) | 2 | 5501351 | 7731988 | 4386757 | 3409839 | 1.62E-01 | -0.20 | 0.83 |
| HILIC_NEG_114_94923a4_404 | | 114.9492 | 4.40 | | HILIC | NEG | | | 937823 | 212484 | 894367 | 237751 | 1.62E-01 | -0.19 | 0.87 |
| HILIC_NEG_173_92621a0_933 | | 173.9262 | 0.93 | | HILIC | NEG | | | 294831 | 64447.2 | 281044 | 77592.6 | 1.62E-01 | -0.19 | 0.94 |
| HILIC_POS_102_0667a0_935 | | 102.0667 | 0.94 | | HILIC | POS | | | 2084636 | 519072 | 1988202 | 488674 | 1.62E-01 | -0.19 | 0.99 |
| RP_POS_178_90157a15_407 | | 178.9016 | 15.41 | | RP | POS | | | 162925 | 131424 | 139991 | 108546 | 1.62E-01 | -0.19 | 0.9 |
| RP_POS_181_02609a0_69 | | 181.0261 | 0.69 | | RP | POS | | | 887495 | 532526 | 792692 | 460223 | 1.62E-01 | -0.19 | 0.84 |
| RP_POS_501_34213a10_79 | | 501.3421 | 10.79 | | RP | POS | | | 2638802 | 1488914 | 2988717 | 2070204 | 1.63E-01 | 0.2 | 1.36 |
| HILIC_NEG_273_01639a0_935 | | 273.0164 | 0.94 | | HILIC | NEG | | | 420612 | 45211.3 | 411686 | 47823.7 | 1.63E-01 | -0.19 | 0.93 |
| RP_NEG_419_17532a11_361 | | 419.1753 | 11.36 | | RP | NEG | | | 248477 | 32137.2 | 242103 | 34373.8 | 1.63E-01 | -0.19 | 1.5 |
| RP_NEG_459_14471a11_592 | | 459.1447 | 11.59 | | RP | NEG | | | 1487951 | 93152.8 | 1469276 | 101525 | 1.63E-01 | -0.19 | 1.16 |
| RP_POS_209_05573a0_676 | | 209.0557 | 0.68 | | RP | POS | | | 736424 | 181377 | 708088 | 113032 | 1.63E-01 | -0.19 | 0.9 |
| RP_POS_695_50787a11_875 | | 695.5079 | 11.88 | | RP | POS | | | 2390218 | 749646 | 2259526 | 621374 | 1.63E-01 | -0.19 | 0.93 |
| RP_POS_327_3374a10_837 | | 327.3374 | 10.84 | | RP | POS | | | 159052 | 93946 | 205457 | 318075 | 1.64E-01 | 0.23 | 0.96 |
| RP_NEG_525_22388a10_712 | | 525.2239 | 10.71 | | RP | NEG | | | 177160 | 157267 | 226047 | 316453 | 1.64E-01 | 0.21 | 1.01 |
| RP_POS_351_22675a11_025 | | 351.2268 | 11.03 | | RP | POS | | | 526868 | 1416453 | 882815 | 2170678 | 1.64E-01 | 0.2 | 0.91 |
| RP_POS_635_44312a11_489 | | 635.4431 | 11.49 | | RP | POS | | | 143233 | 76438.9 | 161426 | 108710 | 1.64E-01 | 0.2 | 0.96 |
| RP_POS_684_81018a11_552 | | 684.8102 | 11.55 | | RP | POS | | | 155673 | 70585.8 | 172531 | 100848 | 1.64E-01 | 0.2 | 1.08 |
| HILIC_NEG_200_85924a6_003 | | 200.8592 | 6.00 | | HILIC | NEG | | | 1809738 | 835662 | 1978717 | 926834 | 1.64E-01 | 0.19 | 0.82 |
| RP_NEG_506_32614a10_609 | | 506.3261 | 10.61 | | RP | NEG | | | 5689751 | 1155157 | 5922113 | 1268821 | 1.64E-01 | 0.19 | 1 |
| RP_POS_241_10698a10_252 | | 241.1070 | 10.25 | | RP | POS | | | 433273 | 212473 | 474473 | 218751 | 1.64E-01 | 0.19 | 1.25 |
| HILIC_POS_64_01667a4_85 | | 64.0167 | 4.85 | | HILIC | POS | | | 4481212 | 2813587 | 3985193 | 2404840 | 1.64E-01 | -0.19 | 1.01 |
| RP_NEG_653_30316a12_29 | | 653.3032 | 12.29 | | RP | NEG | | | 188359 | 114056 | 167450 | 105717 | 1.64E-01 | -0.19 | 0.8 |
| RP_NEG_676_30713a10_517 | | 676.3071 | 10.52 | | RP | NEG | | | 2808707 | 130050 | 2783138 | 137582 | 1.64E-01 | -0.19 | 0.99 |
| RP_POS_253_14111a8_844 | | 253.1411 | 8.84 | | RP | POS | | | 2.2E+08 | 8.5E+07 | 2E+08 | 7.5E+07 | 1.64E-01 | -0.19 | 1.23 |
| RP_POS_331_17831a11_148 | | 331.1783 | 11.15 | | RP | POS | | | 684026 | 157453 | 654202 | 155668 | 1.64E-01 | -0.19 | 1.06 |
| RP_POS_269_13623a4_114 | | 269.1362 | 4.11 | | RP | POS | | | 1003254 | 2268338 | 1820019 | 5432468 | 1.65E-01 | 0.21 | 0.98 |
| HILIC_POS_262_16458a3_319 | | 262.1646 | 3.32 | | HILIC | POS | | | 312185 | 332385 | 399190 | 539570 | 1.65E-01 | 0.2 | 0.89 |
| RP_POS_491_01013a9_727 | | 491.0101 | 9.73 | | RP | POS | | | 228454 | 134222 | 259382 | 182740 | 1.65E-01 | 0.2 | 0.89 |
| RP_POS_554_06018a10_305 | | 554.0602 | 10.31 | | RP | POS | | | 639598 | 419230 | 736384 | 573403 | 1.65E-01 | 0.2 | 1.02 |
| RP_POS_651_12201a10_463 | | 651.1220 | 10.46 | | RP | POS | | | 210203 | 129856 | 235754 | 137732 | 1.65E-01 | 0.19 | 0.84 |
| HILIC_NEG_259_83936a7_735 | | 259.8394 | 7.74 | | HILIC | NEG | | | 1.9E+07 | 1803568 | 1.9E+07 | 1584314 | 1.65E-01 | -0.19 | 0.96 |
| HILIC_POS_537_53503a10_652 | | 537.5350 | 10.65 | | HILIC | POS | | | 413867 | 53387.1 | 402110 | 68044.5 | 1.65E-01 | -0.19 | 0.96 |
| RP_NEG_315_25427a9_574 | | 315.2543 | 9.57 | | RP | NEG | | | 627067 | 220968 | 587146 | 199733 | 1.65E-01 | -0.19 | 0.89 |
| RP_NEG_327_25409a10_368 | | 327.2541 | 10.37 | | RP | NEG | | | 470376 | 52215.3 | 460205 | 54425.5 | 1.65E-01 | -0.19 | 1.04 |
| RP_POS_236_07176a13_165 | | 236.0718 | 13.17 | | RP | POS | | | 7.4E+07 | 2.3E+07 | 8.1E+07 | 4.1E+07 | 1.66E-01 | 0.2 | 0.93 |
| RP_POS_506_35938a14_093 | | 506.3594 | 14.09 | | RP | POS | | | 456474 | 472855 | 561536 | 612488 | 1.66E-01 | 0.19 | 1.1 |
| RP_POS_381_08408a9_522 | | 381.0841 | 9.52 | | RP | POS | | | 165670 | 68741.9 | 192530 | 181402 | 1.67E-01 | 0.21 | 0.92 |
| RP_POS_131_08568a6_382 | | 131.0857 | 6.38 | | RP | POS | | | 701511 | 88217.6 | 719312 | 98531.2 | 1.67E-01 | 0.19 | 0.95 |
| RP_POS_695_68719a11_315 | | 695.6872 | 11.32 | | RP | POS | | | 189280 | 90674.4 | 209411 | 117644 | 1.67E-01 | 0.19 | 0.99 |
| HILIC_POS_443_32275a1_209 | | 443.3228 | 1.21 | | HILIC | POS | | | 1446119 | 1287706 | 1240749 | 877837 | 1.67E-01 | -0.19 | 1.09 |
| RP_NEG_178_86525a14_342 | | 178.8653 | 14.34 | | RP | NEG | | | 712623 | 344823 | 652336 | 293733 | 1.67E-01 | -0.19 | 0.8 |
| RP_NEG_554_38214a11_201 | | 554.3821 | 11.20 | | RP | NEG | | | 970252 | 328185 | 907782 | 330744 | 1.67E-01 | -0.19 | 0.81 |
| RP_POS_221_117a8_486 | | 221.1170 | 8.49 | | RP | POS | | | 1156799 | 512973 | 1061992 | 489273 | 1.67E-01 | -0.19 | 1.05 |
| RP_POS_425_8074a9_624 | | 425.8074 | 9.62 | | RP | POS | | | 1394982 | 758356 | 1255451 | 717091 | 1.67E-01 | -0.19 | 1.1 |
| RP_POS_512_33459a10_062 | | 512.3346 | 10.06 | | RP | POS | | | 8891560 | 2773467 | 8367523 | 2756284 | 1.67E-01 | -0.19 | 0.98 |
| RP_POS_518_32135a10_487 | | 518.3214 | 10.49 | | RP | POS | | | 2.6E+08 | 6E+07 | 2.5E+08 | 5.7E+07 | 1.67E-01 | -0.19 | 0.98 |
| RP_POS_265_25208a10_809 | | 265.2521 | 10.81 | | RP | POS | | | 5590328 | 1746074 | 6026954 | 2697875 | 1.68E-01 | 0.2 | 1.3 |
| RP_NEG_349_08636a7_087 | | 349.0864 | 7.09 | | RP | NEG | | | 4857485 | 2267058 | 5318497 | 2578606 | 1.68E-01 | 0.19 | 0.86 |
| RP_POS_331_24716a9_635 | | 331.2472 | 9.64 | | RP | POS | | | 263478 | 57510.8 | 275757 | 70658.1 | 1.68E-01 | 0.19 | 0.89 |
| RP_POS_373_20432a11_239 | | 373.2043 | 11.24 | | RP | POS | | | 1191210 | 609501 | 1301210 | 558021 | 1.68E-01 | 0.19 | 1.09 |
| RP_POS_563_72949a10_154 | | 563.7295 | 10.15 | | RP | POS | | | 936063 | 542112 | 1044409 | 598793 | 1.68E-01 | 0.19 | 0.98 |
| HILIC_NEG_261_83615a7_735 | | 261.8362 | 7.74 | | HILIC | NEG | | | 1558922 | 149826 | 1532357 | 132103 | 1.68E-01 | -0.19 | 0.9 |
| HILIC_POS_249_13333a0_77 | | 249.1333 | 0.77 | | HILIC | POS | | | 239919 | 21852.3 | 235623 | 23488.7 | 1.68E-01 | -0.19 | 0.9 |
| HILIC_POS_74_09721a1_032 | | 74.0972 | 1.03 | | HILIC | POS | | | 2891608 | 1002836 | 2720637 | 814891 | 1.68E-01 | -0.19 | 0.82 |
| RP_POS_236_07149a8_785 | | 236.0715 | 8.79 | | RP | POS | | | 5.9E+07 | 2.4E+07 | 5.5E+07 | 2E+07 | 1.68E-01 | -0.19 | 0.87 |
| RP_POS_368_21893a10_253 | | 368.2189 | 10.25 | | RP | POS | | | 2210432 | 278893 | 2157636 | 279324 | 1.68E-01 | -0.19 | 1.08 |
| RP_POS_385_33173a10_776 | | 385.3317 | 10.78 | | RP | POS | | | 436557 | 58559.3 | 424625 | 66894.8 | 1.68E-01 | -0.19 | 0.92 |
| RP_POS_396_20276a9_959 | | 396.2028 | 9.96 | | RP | POS | | | 425528 | 331896 | 363162 | 328274 | 1.68E-01 | -0.19 | 1.05 |
| HILIC_NEG_214_9192a2_988 | | 214.9192 | 2.99 | | HILIC | NEG | | | 258005 | 43202 | 266271 | 44325.9 | 1.69E-01 | 0.19 | 0.84 |
| RP_NEG_231_0515a4_621 | | 231.0515 | 4.62 | | RP | NEG | | | 166683 | 154169 | 203602 | 224819 | 1.69E-01 | 0.19 | 1.1 |
| RP_POS_445_121a10_701 | | 445.1210 | 10.70 | | RP | POS | | | 716496 | 124866 | 741949 | 143073 | 1.69E-01 | 0.19 | 1.04 |
| RP_NEG_365_34229a12_827 | Nervonic acid | 365.3423 | 12.83 | | RP | NEG | 365.3406(100) | 2 | 91658.5 | 55445.3 | 102259 | 56960.6 | 1.69E-01 | 0.19 | 0.84 |
| HILIC_POS_302_1962a1_226 | | 302.1962 | 1.23 | | HILIC | POS | | | 1392020 | 813664 | 1251778 | 679525 | 1.69E-01 | -0.19 | 0.94 |
| HILIC_NEG_142_99284a0_933 | | 142.9928 | 0.93 | | HILIC | NEG | | | 484188 | 52642.5 | 474452 | 50825.3 | 1.69E-01 | -0.19 | 0.92 |
| RP_NEG_250_99463a15_483 | | 250.9946 | 15.48 | | RP | NEG | | | 1019860 | 225631 | 982238 | 174376 | 1.69E-01 | -0.19 | 0.77 |
| RP_POS_175_09666a8_844 | | 175.0967 | 8.84 | | RP | POS | | | 4E+07 | 1.4E+07 | 3.8E+07 | 1.3E+07 | 1.69E-01 | -0.19 | 0.93 |

**Supplemantary table 1**: The non-targeted metabolomics results ordered by the VIP value (p value < 0.0003 are shaded dark green, p value < 0.05 are shaded light green, d values below 0 are blue and above 0 are red)

| | | | | | | | | | Cases | | Controls | | Welch test | Cohen's | |
|---|---|---|---|---|---|---|---|---|---|---|---|---|---|---|---|
| Feature_ID | Metabolite_name | Average_Mz | Average_RT_min_ | HMDB ID | RP_HILIC | POS_NEG | Cureted_MSMS | ID_level | Mean | SD | Mean2 | SD3 | p | d | VIP |
| RP_POS_177_09084a6_31 | | 177.0908 | 6.31 | | RP | POS | | | 1986684 | 350078 | 1923703 | 319648 | 1.69E-01 | -0.19 | 0.94 |
| RP_POS_269_08792a1_701 | | 269.0879 | 1.70 | | RP | POS | | | 430699 | 508452 | 353449 | 302866 | 1.69E-01 | -0.19 | 0.89 |
| RP_POS_448_30579a8_597 | | 448.3058 | 8.60 | | RP | POS | | | 2295717 | 2738741 | 1708654 | 3396945 | 1.69E-01 | -0.19 | 0.98 |
| RP_POS_540_04401a9_871 | | 540.0440 | 9.87 | | RP | POS | | | 265632 | 141180 | 295495 | 172437 | 1.70E-01 | 0.19 | 0.93 |
| RP_POS_802_21411a9_539 | | 802.2141 | 9.54 | | RP | POS | | | 1532033 | 136975 | 1558347 | 142193 | 1.70E-01 | 0.19 | 1.07 |
| RP_POS_508_37628a10_87 | LysoPC O-18:1 | 508.3763 | 10.87 | | RP | POS | 104.1078(100) | 1 | 1.8E+07 | 4553069 | 1.9E+07 | 5218403 | 1.70E-01 | 0.19 | 0.98 |
| HILIC_POS_132_10211a3_951 | Leucine | 132.1021 | 3.95 | | HILIC | POS | 86.0974(100), 132.1028(2) | 2 | 1.3E+09 | 5.3E+09 | 5.3E+08 | 3.3E+09 | 1.70E-01 | -0.19 | 0.85 |
| RP_NEG_192_8894a15_4 | | 192.8894 | 15.40 | | RP | NEG | | | 893971 | 333297 | 823280 | 407984 | 1.70E-01 | -0.19 | 0.96 |
| RP_NEG_334_93805a3_677 | | 334.9381 | 3.68 | | RP | NEG | | | 540840 | 403283 | 471164 | 340949 | 1.70E-01 | -0.19 | 0.92 |
| RP_NEG_350_98721a7_957 | | 350.9872 | 7.96 | | RP | NEG | | | 1177248 | 753836 | 1050762 | 596565 | 1.70E-01 | -0.19 | 0.81 |
| RP_NEG_788_5238a13_058 | | 788.5238 | 13.06 | | RP | NEG | | | 755988 | 619820 | 661532 | 374289 | 1.70E-01 | -0.19 | 0.73 |
| RP_POS_534_37787a10_183 | | 534.3779 | 10.18 | | RP | POS | | | 1156943 | 606247 | 1405902 | 1708172 | 1.71E-01 | 0.22 | 1.13 |
| HILIC_NEG_186_95554a1_349 | | 186.9555 | 1.35 | | HILIC | NEG | | | 867346 | 177641 | 900079 | 171453 | 1.71E-01 | 0.19 | 0.84 |
| RP_POS_327_22934a10_927 | | 327.2293 | 10.93 | | RP | POS | | | 8507928 | 3778445 | 9176972 | 3384297 | 1.71E-01 | 0.19 | 1.1 |
| RP_POS_810_59778a14_928 | | 810.5978 | 14.93 | | RP | POS | | | 5117598 | 4561081 | 5933169 | 4158405 | 1.71E-01 | 0.19 | 0.89 |
| HILIC_NEG_212_91116a2_278 | | 212.9112 | 2.28 | | HILIC | NEG | | | 4797866 | 1077983 | 4580330 | 1222116 | 1.71E-01 | -0.19 | 1.07 |
| RP_NEG_637_58813a12_955 | | 637.5881 | 12.96 | | RP | NEG | | | 792852 | 502819 | 698899 | 498897 | 1.71E-01 | -0.19 | 0.84 |
| RP_POS_479_83707a10_264 | | 479.8371 | 10.26 | | RP | POS | | | 1075040 | 541434 | 976347 | 512436 | 1.71E-01 | -0.19 | 0.91 |
| HILIC_POS_258_10806a1_463 | | 258.1081 | 1.46 | | HILIC | POS | | | 244342 | 52839.8 | 254803 | 58456.9 | 1.72E-01 | 0.19 | 0.93 |
| HILIC_POS_64_01667a5_642 | | 64.0167 | 5.64 | | HILIC | POS | | | 2535109 | 1259624 | 2751825 | 1066777 | 1.72E-01 | 0.19 | 0.75 |
| RP_POS_426_23364a6_941 | | 426.2336 | 6.94 | | RP | POS | | | 178428 | 48366.6 | 187311 | 46599.6 | 1.72E-01 | 0.19 | 0.96 |
| RP_POS_357_27869a9_261 | Hyodeoxycholic acid | 357.2787 | 9.26 | | RP | POS | 357.2769(100), 95.086(40) | 1 | 2075863 | 1622612 | 1816627 | 1149971 | 1.72E-01 | -0.19 | 0.9 |
| RP_NEG_448_30759a8_243 | | 448.3076 | 8.24 | | RP | NEG | 448.3044(100), 74.0244(60) | | 2277923 | 3088153 | 1825844 | 1639880 | 1.72E-01 | -0.19 | 1 |
| RP_NEG_652_40063a11_142 | | 652.4006 | 11.14 | | RP | NEG | | | 630414 | 65784.5 | 618626 | 60636.7 | 1.72E-01 | -0.19 | 0.9 |
| RP_POS_352_24554a10_366 | | 352.2455 | 10.37 | | RP | POS | | | 1.3E+07 | 1832927 | 1.3E+07 | 1896971 | 1.72E-01 | -0.19 | 1.01 |
| RP_POS_375_21884a11_144 | | 375.2188 | 11.14 | | RP | POS | | | 4484294 | 854926 | 4335128 | 746401 | 1.72E-01 | -0.19 | 0.91 |
| RP_POS_535_39868a11_324 | | 535.3987 | 11.32 | | RP | POS | | | 277148 | 246908 | 240124 | 143653 | 1.72E-01 | -0.19 | 0.77 |
| RP_POS_759_12946a9_278 | | 759.1295 | 9.28 | | RP | POS | | | 683861 | 83038.5 | 668668 | 79489.8 | 1.72E-01 | -0.19 | 1.15 |
| RP_NEG_324_75824a15_356 | | 324.7582 | 15.36 | | RP | NEG | | | 4536576 | 315192 | 4595108 | 312663 | 1.73E-01 | 0.19 | 1.03 |
| RP_NEG_348_81119a15_4 | | 348.8112 | 15.40 | | RP | NEG | | | 1182902 | 363123 | 1252043 | 375470 | 1.73E-01 | 0.19 | 1.02 |
| RP_POS_237_14597a7_529 | | 237.1460 | 7.53 | | RP | POS | | | 901234 | 224199 | 954978 | 331623 | 1.73E-01 | 0.19 | 1.13 |
| RP_POS_332_29276a11_235 | | 332.2928 | 11.24 | | RP | POS | | | 1436670 | 2092354 | 1913031 | 2884813 | 1.73E-01 | 0.19 | 1.2 |
| HILIC_POS_241_03139a7_692 | Cystine | 241.0314 | 7.69 | | HILIC | POS | 120.0119(100), 151.9834(90) | 2 | 337680 | 178808 | 369005 | 157851 | 1.73E-01 | 0.19 | 0.93 |
| RP_NEG_370_18167a11_144 | | 370.1817 | 11.14 | | RP | NEG | | | 352885 | 30779.7 | 346741 | 34630.3 | 1.73E-01 | -0.19 | 1.09 |
| RP_NEG_465_43243a11_591 | | 465.4324 | 11.59 | | RP | NEG | | | 214076 | 121485 | 194470 | 88721.8 | 1.73E-01 | -0.19 | 0.85 |
| HILIC_POS_371_24008a5_795 | | 371.2401 | 5.80 | | HILIC | POS | | | 7438816 | 960314 | 7614208 | 925580 | 1.74E-01 | 0.19 | 0.97 |
| RP_NEG_1087_69568a10_609 | | 1087.6957 | 10.61 | | RP | NEG | | | 7748929 | 2843267 | 8305015 | 3100069 | 1.74E-01 | 0.19 | 0.97 |
| RP_NEG_174_82866a15_475 | | 174.8287 | 15.48 | | RP | NEG | | | 1870106 | 1091407 | 2091635 | 1267963 | 1.74E-01 | 0.19 | 0.85 |
| RP_POS_282_99548a0_547 | | 282.9955 | 0.55 | | RP | POS | | | 1054974 | 118446 | 1079553 | 142199 | 1.74E-01 | 0.19 | 0.98 |
| RP_POS_368_20038a11_239 | | 368.2004 | 11.24 | | RP | POS | | | 2891430 | 1271073 | 3120978 | 1200019 | 1.74E-01 | 0.19 | 1.13 |
| RP_POS_399_34653a12 | | 399.3465 | 12.00 | | RP | POS | | | 622876 | 182558 | 657345 | 186722 | 1.74E-01 | 0.19 | 0.95 |
| RP_POS_488_26169a11_142 | | 488.2617 | 11.14 | | RP | POS | | | 668547 | 407283 | 756122 | 516294 | 1.74E-01 | 0.19 | 1.27 |
| RP_POS_682_80566a11_258 | | 682.8057 | 11.26 | | RP | POS | | | 137889 | 57489.3 | 151007 | 79643.3 | 1.74E-01 | 0.19 | 0.94 |
| HILIC_NEG_183_06635a0_933 | | 183.0664 | 0.93 | | HILIC | NEG | | | 717390 | 118573 | 695873 | 112960 | 1.74E-01 | -0.19 | 0.84 |
| HILIC_NEG_92_99541a0_934 | | 92.9954 | 0.93 | | HILIC | NEG | | | 263687 | 32886.8 | 257807 | 30392.5 | 1.74E-01 | -0.19 | 0.85 |
| HILIC_POS_453_24585a6_248 | | 453.2459 | 6.25 | | HILIC | POS | | | 307156 | 176262 | 273617 | 183064 | 1.74E-01 | -0.19 | 1.07 |
| HILIC_POS_64_01669a3_943 | | 64.0167 | 3.94 | | HILIC | POS | | | 1910595 | 1287326 | 1688967 | 1101807 | 1.74E-01 | -0.19 | 0.87 |
| RP_NEG_269_21265a9_933 | | 269.2127 | 9.93 | | RP | NEG | | | 700059 | 89720 | 683940 | 83883.3 | 1.74E-01 | -0.19 | 1.23 |
| RP_POS_238_06734a11_144 | | 238.0673 | 11.14 | | RP | POS | | | 7285628 | 2237415 | 6888843 | 2031345 | 1.74E-01 | -0.19 | 0.89 |
| RP_POS_597_89703a9_765 | | 597.8970 | 9.77 | | RP | POS | | | 1169473 | 618634 | 1059625 | 563212 | 1.74E-01 | -0.19 | 1.12 |
| RP_POS_653_45557a11_317 | | 653.4556 | 11.32 | | RP | POS | | | 218220 | 221453 | 185500 | 124412 | 1.74E-01 | -0.19 | 0.74 |
| HILIC_POS_170_02457a5_176 | | 170.0246 | 5.18 | | HILIC | POS | | | 462513 | 475237 | 545970 | 425410 | 1.75E-01 | 0.19 | 0.93 |
| HILIC_POS_371_24005a6_173 | | 371.2401 | 6.17 | | HILIC | POS | | | 1858878 | 348874 | 1918949 | 299419 | 1.75E-01 | 0.19 | 0.78 |
| RP_POS_803_05182a13_229 | | 803.0518 | 13.23 | | RP | POS | | | 1062510 | 558630 | 1167704 | 572470 | 1.75E-01 | 0.19 | 0.94 |
| RP_POS_930_23956a9_461 | | 930.2396 | 9.46 | | RP | POS | | | 753774 | 426426 | 841621 | 508797 | 1.75E-01 | 0.19 | 1.05 |
| RP_NEG_566_34521a10_485 | LysoPC 18:1 | 566.3452 | 10.49 | | RP | NEG | 281.2495(100), 506.3267(10) | 1 | 1.4E+07 | 2785252 | 1.5E+07 | 2816124 | 1.75E-01 | 0.19 | 0.91 |
| RP_NEG_176_99783a15_508 | | 176.9978 | 15.51 | | RP | NEG | | | 1238786 | 209305 | 1204319 | 162310 | 1.75E-01 | -0.19 | 1.08 |
| RP_NEG_223_13412a7_871 | | 223.1341 | 7.87 | | RP | NEG | | | 340107 | 23278.3 | 335847 | 22661.2 | 1.75E-01 | -0.19 | 0.96 |
| RP_POS_157_08607a8_845 | | 157.0861 | 8.85 | | RP | POS | | | 8858514 | 2910815 | 8348093 | 2596097 | 1.75E-01 | -0.19 | 0.93 |
| RP_POS_608_44623a10_642 | | 608.4462 | 10.64 | | RP | POS | | | 867574 | 559249 | 779390 | 389195 | 1.75E-01 | -0.19 | 0.92 |
| HILIC_NEG_380_84418a2_995 | | 380.8442 | 3.00 | | HILIC | NEG | | | 1785412 | 342602 | 1849598 | 349698 | 1.76E-01 | 0.19 | 0.83 |
| RP_POS_613_09528a10_258 | | 613.0953 | 10.26 | | RP | POS | | | 196183 | 113856 | 221955 | 156831 | 1.76E-01 | 0.19 | 0.91 |
| RP_POS_135_06525a6_5 | | 135.0653 | 6.50 | | RP | POS | | | 3544927 | 301120 | 3597950 | 272343 | 1.76E-01 | 0.18 | 1.21 |
| RP_POS_200_03224a15_419 | | 200.0322 | 15.42 | | RP | POS | | | 1446381 | 419400 | 1518588 | 361987 | 1.76E-01 | 0.18 | 0.9 |
| RP_NEG_207_0119a4_215 | 2-Naphthalenesulfonic acid | 207.0119 | 4.22 | | RP | NEG | 207.0123(100), 143.0502(10) | 2 | 190442 | 110942 | 173971 | 64462.4 | 1.76E-01 | -0.19 | 0.73 |
| HILIC_POS_246_20625a2_991 | | 246.2063 | 2.99 | | HILIC | POS | | | 253286 | 180437 | 222990 | 147290 | 1.76E-01 | -0.18 | 0.88 |
| RP_NEG_315_25446a10_229 | | 315.2545 | 10.23 | | RP | NEG | | | 484583 | 85961.5 | 469244 | 79893.4 | 1.76E-01 | -0.18 | 1.26 |
| RP_POS_271_11545a4_97 | | 271.1155 | 4.97 | | RP | POS | | | 238327 | 150321 | 212081 | 133512 | 1.76E-01 | -0.18 | 1.15 |
| RP_POS_808_58197a11_594 | | 808.5820 | 11.59 | | RP | POS | | | 1152526 | 246889 | 1109541 | 218507 | 1.76E-01 | -0.18 | 0.89 |
| RP_NEG_221_11836a7_575 | | 221.1184 | 7.58 | | RP | NEG | | | 1179415 | 99148.3 | 1161512 | 94168.7 | 1.76E-01 | -0.19 | 1.35 |
| HILIC_POS_276_1803a2_289 | | 276.1803 | 2.29 | | HILIC | POS | | | 500575 | 1226273 | 786377 | 1770921 | 1.77E-01 | 0.19 | 1.07 |
| RP_POS_252_19542a9_828 | | 252.1954 | 9.83 | | RP | POS | | | 2639777 | 280170 | 2694645 | 309307 | 1.77E-01 | 0.19 | 1.27 |
| RP_POS_298_27365a10_964 | | 298.2737 | 10.96 | | RP | POS | | | 1091639 | 621459 | 1208370 | 638347 | 1.77E-01 | 0.19 | 1.09 |
| RP_POS_477_80658a12_805 | | 477.8066 | 12.81 | | RP | POS | | | 371031 | 209951 | 416109 | 268884 | 1.77E-01 | 0.19 | 1.12 |
| RP_POS_523_32434a10_789 | | 523.3243 | 10.79 | | RP | POS | | | 7775279 | 4353070 | 8777702 | 6170021 | 1.77E-01 | 0.19 | 1.34 |
| RP_POS_207_17439a9_41 | | 207.1744 | 9.41 | | RP | POS | | | 928983 | 307791 | 985988 | 308710 | 1.77E-01 | 0.18 | 0.78 |
| RP_POS_413_09647a0_585 | | 413.0965 | 0.59 | | RP | POS | | | 197181 | 90459.5 | 213474 | 86168.1 | 1.77E-01 | 0.18 | 1 |
| RP_POS_586_3067a6_323 | | 586.3067 | 6.32 | | RP | POS | | | 951892 | 79307.1 | 965734 | 70676.2 | 1.77E-01 | 0.18 | 0.83 |
| RP_POS_276_83188a8_842 | | 276.8319 | 8.84 | | RP | POS | | | 496004 | 231759 | 455586 | 206809 | 1.77E-01 | -0.18 | 1.24 |
| HILIC_NEG_134_89474a4_834 | | 134.8947 | 4.83 | | HILIC | NEG | | | 1.3E+07 | 2035317 | 1.2E+07 | 2417434 | 1.77E-01 | -0.19 | 0.84 |
| RP_NEG_669_46198a11_591 | | 669.4620 | 11.59 | | RP | NEG | | | 962450 | 69591.1 | 948942 | 75686.4 | 1.77E-01 | -0.19 | 1.21 |
| RP_POS_135_08049a10_261 | | 135.0805 | 10.26 | | RP | POS | | | 1051525 | 156851 | 1020648 | 174620 | 1.77E-01 | -0.19 | 0.9 |
| RP_POS_676_96533a10_566 | | 676.9653 | 10.57 | | RP | POS | | | 621082 | 284848 | 566790 | 300027 | 1.77E-01 | -0.19 | 0.91 |
| RP_NEG_421_22644a10_48 | | 421.2264 | 10.48 | | RP | NEG | | | 139936 | 111240 | 281865 | 1035609 | 1.78E-01 | 0.25 | 0.91 |
| HILIC_POS_458_76929a2_98 | | 458.7693 | 2.98 | | HILIC | POS | | | 449530 | 83197.4 | 465300 | 87124 | 1.78E-01 | 0.19 | 0.79 |
| RP_POS_1043_60681a10_3 | | 1043.6068 | 10.30 | | RP | POS | | | 777227 | 266163 | 829944 | 300859 | 1.78E-01 | 0.19 | 1.03 |
| RP_POS_355_19205a11_239 | | 355.1921 | 11.24 | | RP | POS | | | 460648 | 218766 | 502548 | 233760 | 1.78E-01 | 0.19 | 1.1 |
| RP_POS_546_45496a14_994 | | 546.4550 | 14.99 | | RP | POS | | | 4705251 | 1.1E+07 | 7272200 | 1.6E+07 | 1.78E-01 | 0.19 | 0.97 |
| RP_POS_716_55646a13_349 | | 716.5565 | 13.35 | | RP | POS | | | 745383 | 477230 | 834090 | 483279 | 1.78E-01 | 0.18 | 0.99 |
| HILIC_POS_131_53362a0_926 | | 131.5336 | 0.93 | | HILIC | POS | | | 375676 | 242755 | 334964 | 199818 | 1.78E-01 | -0.18 | 1 |
| HILIC_POS_304_10184a4_415 | | 304.1018 | 4.42 | | HILIC | POS | | | 602824 | 102581 | 585437 | 86553.1 | 1.78E-01 | -0.18 | 0.78 |
| RP_POS_141_0909a5_637 | | 141.0909 | 5.64 | | RP | POS | | | 1809113 | 786962 | 1667796 | 747332 | 1.78E-01 | -0.18 | 0.91 |
| RP_POS_235_20543a9_936 | | 235.2054 | 9.94 | | RP | POS | | | 975278 | 139006 | 950761 | 127413 | 1.78E-01 | -0.18 | 1.01 |
| RP_POS_382_28027a10_833 | | 382.2803 | 10.83 | | RP | POS | | | 93062.8 | 53928.3 | 84255.8 | 41687.2 | 1.78E-01 | -0.18 | 1.1 |
| RP_POS_566_91382a10_178 | | 566.9138 | 10.18 | | RP | POS | | | 186900 | 119490 | 166940 | 97308.7 | 1.78E-01 | -0.18 | 1.03 |
| RP_POS_546_38446a7_271 | | 546.3845 | 7.27 | | RP | POS | | | 784523 | 763024 | 667684 | 496527 | 1.78E-01 | -0.19 | 0.78 |
| HILIC_NEG_212_92068a2_996 | | 212.9207 | 3.00 | | HILIC | NEG | | | 1840184 | 320309 | 1898586 | 314540 | 1.79E-01 | 0.18 | 0.84 |
| RP_POS_548_50208a13_292 | | 548.5021 | 13.29 | | RP | POS | | | 1630918 | 694821 | 1757647 | 681721 | 1.79E-01 | 0.18 | 0.99 |
| RP_POS_805_01135a10_608 | | 805.0114 | 10.61 | | RP | POS | | | 3381954 | 1143013 | 3596494 | 1185633 | 1.79E-01 | 0.18 | 0.87 |
| RP_POS_243_09891a8_392 | | 243.0989 | 8.39 | | RP | POS | 243.0987(100) | | 418115 | 368973 | 355682 | 312313 | 1.79E-01 | -0.18 | 1.24 |
| RP_NEG_257_81973a0_731 | | 257.8197 | 0.73 | | RP | NEG | | | 571606 | 65202.7 | 560269 | 58269.7 | 1.79E-01 | -0.18 | 1.01 |
| RP_NEG_369_30188a11_743 | | 369.3019 | 11.74 | | RP | NEG | | | 1365340 | 120068 | 1344412 | 108013 | 1.79E-01 | -0.18 | 1.33 |
| RP_POS_302_24515a11_14 | | 302.2452 | 11.14 | | RP | POS | | | 662250 | 207322 | 627614 | 170225 | 1.79E-01 | -0.18 | 1.02 |
| RP_POS_492_36234a10_042 | | 492.3623 | 10.04 | | RP | POS | | | 7464879 | 3504119 | 6819541 | 3499363 | 1.79E-01 | -0.18 | 1.07 |
| HILIC_POS_357_68604a1_881 | | 357.6860 | 1.88 | | HILIC | POS | | | 575255 | 95168.3 | 553698 | 132671 | 1.79E-01 | -0.19 | 0.91 |
| RP_NEG_595_06293a8_833 | | 595.0629 | 8.83 | | RP | NEG | | | 526108 | 32382.5 | 519715 | 36621.7 | 1.79E-01 | -0.19 | 0.97 |
| RP_POS_461_21893a5_458 | | 461.2189 | 5.46 | | RP | POS | | | 778498 | 374614 | 693228 | 526361 | 1.79E-01 | -0.19 | 1.03 |
| RP_NEG_457_23663a10_86 | | 457.2366 | 10.86 | | RP | NEG | | | 1094525 | 3952878 | 2340101 | 8453615 | 1.80E-01 | 0.2 | 0.8 |
| HILIC_NEG_373_01007a5_123 | | 373.0101 | 5.12 | | HILIC | NEG | | | 287597 | 119214 | 312739 | 149933 | 1.80E-01 | 0.19 | 1.1 |
| RP_POS_377_26611a10_578 | | 377.2661 | 10.58 | | RP | POS | | | 1284994 | 826238 | 1491501 | 1322096 | 1.80E-01 | 0.19 | 1.08 |
| RP_POS_683_87817a11_255 | | 683.8782 | 11.26 | | RP | POS | | | 160618 | 93576.6 | 181397 | 127194 | 1.80E-01 | 0.19 | 0.92 |
| HILIC_POS_140_06828a4_382 | | 140.0683 | 4.38 | | HILIC | POS | 140.0692(100), 74.0975(100) | | 2731719 | 1465620 | 3012999 | 1583243 | 1.80E-01 | 0.18 | 0.8 |
| HILIC_NEG_383_15396a0_606 | | 383.1540 | 0.61 | | HILIC | NEG | | | 179784 | 150793 | 206462 | 140316 | 1.80E-01 | 0.18 | 0.86 |
| RP_POS_362_22064a10_653 | | 362.2206 | 10.65 | | RP | POS | | | 438562 | 161948 | 467896 | 157512 | 1.80E-01 | 0.18 | 0.99 |
| HILIC_NEG_186_99892a0_935 | | 186.9989 | 0.94 | | HILIC | NEG | | | 219048 | 28233.3 | 214060 | 26191.1 | 1.80E-01 | -0.18 | 0.82 |
| HILIC_NEG_259_00116a0_935 | | 259.0012 | 0.94 | | HILIC | NEG | | | 1401354 | 143974 | 1375088 | 142138 | 1.80E-01 | -0.18 | 0.83 |
| RP_POS_852_55072a12_689 | | 852.5507 | 12.69 | | RP | POS | | | 3965245 | 1708066 | 3663163 | 1592129 | 1.80E-01 | -0.18 | 1.05 |
| RP_POS_779_55725a13_782 | | 779.5573 | 13.78 | | RP | POS | | | 1348390 | 637745 | 1478065 | 762053 | 1.81E-01 | 0.19 | 0.8 |
| RP_NEG_508_87668a15_332 | | 508.8767 | 15.33 | | RP | NEG | | | 977944 | 44645.1 | 986231 | 45650.5 | 1.81E-01 | 0.18 | 0.83 |
| RP_POS_129_00351a15_335 | | 129.0035 | 15.34 | | RP | POS | | | 290448 | 122947 | 314373 | 136767 | 1.81E-01 | 0.18 | 0.89 |
| RP_POS_395_215a11_239 | | 395.2150 | 11.24 | | RP | POS | | | 522682 | 313228 | 581126 | 323588 | 1.81E-01 | 0.18 | 1.02 |
| RP_POS_407_3371a10_49 | | 407.3371 | 10.49 | | RP | POS | | | 708534 | 243504 | 751800 | 230099 | 1.81E-01 | 0.18 | 0.92 |
| HILIC_POS_358_29523a0_617 | | 358.2952 | 0.62 | | HILIC | POS | | | 504493 | 310679 | 450233 | 282657 | 1.81E-01 | -0.18 | 0.83 |
| RP_NEG_653_26746a5_64 | | 653.2675 | 5.64 | | RP | NEG | | | 325821 | 185227 | 294761 | 155187 | 1.81E-01 | -0.18 | 1.04 |
| RP_POS_495_30057a11_465 | | 495.3006 | 11.47 | | RP | POS | | | 338579 | 150767 | 313476 | 124251 | 1.81E-01 | -0.18 | 1.15 |
| HILIC_POS_532_33545a1_158 | | 532.3355 | 1.16 | | HILIC | POS | | | 39915.1 | 53420.4 | 32317.8 | 27535.6 | 1.81E-01 | -0.19 | 0.88 |
| RP_POS_792_55286a13_687 | | 792.5529 | 13.69 | | RP | POS | | | 5816935 | 5557099 | 5013784 | 3061183 | 1.81E-01 | -0.19 | 0.93 |
| HILIC_NEG_200_85883a1_012 | | 200.8588 | 1.01 | | HILIC | NEG | | | 608615 | 251739 | 656290 | 267888 | 1.82E-01 | 0.18 | 0.87 |
| RP_NEG_566_34576a10_609 | | 566.3458 | 10.61 | | RP | NEG | | | 1.9E+08 | 3.8E+07 | 2E+08 | 4.1E+07 | 1.82E-01 | 0.18 | 0.97 |
| RP_NEG_717_51825a11_878 | | 717.5183 | 11.88 | | RP | NEG | | | 3320816 | 764912 | 3187077 | 700898 | 1.82E-01 | -0.18 | 0.79 |
| RP_POS_245_13141a15_328 | | 245.1314 | 15.33 | | RP | POS | | | 1258448 | 682478 | 1143526 | 580318 | 1.82E-01 | -0.18 | 1.04 |
| RP_POS_257_17459a9_496 | | 257.1746 | 9.50 | | RP | POS | | | 818052 | 893788 | 677630 | 642454 | 1.82E-01 | -0.18 | 0.83 |
| RP_POS_366_18613a10_633 | | 366.1861 | 10.63 | | RP | POS | | | 731435 | 260267 | 687799 | 218345 | 1.82E-01 | -0.18 | 0.93 |
| RP_POS_424_83466a15_412 | | 424.8347 | 15.41 | | RP | POS | | | 630806 | 345226 | 569546 | 326320 | 1.82E-01 | -0.18 | 1.01 |
| RP_POS_552_40302a11_335 | | 552.4030 | 11.34 | | RP | POS | | | 1493906 | 801893 | 1375057 | 484159 | 1.82E-01 | -0.18 | 0.88 |
| HILIC_POS_173_07849a0_672 | | 173.0785 | 0.67 | | HILIC | POS | | | 304601 | 631087 | 215333 | 321147 | 1.82E-01 | -0.19 | 0.72 |
| RP_NEG_397_22668a10_682 | | 397.2267 | 10.68 | | RP | NEG | | | 59014.3 | 75108.3 | 48637.2 | 34066.1 | 1.82E-01 | -0.19 | 0.76 |
| RP_POS_186_99461a15_384 | | 186.9946 | 15.38 | | RP | POS | | | 328285 | 210518 | 373169 | 271942 | 1.83E-01 | 0.19 | 1.15 |
| HILIC_NEG_548_76831a2_994 | | 548.7683 | 2.99 | | HILIC | NEG | | | 508304 | 87738.7 | 524649 | 91200.2 | 1.83E-01 | 0.18 | 0.77 |
| RP_NEG_526_29395a10_524 | | 526.2940 | 10.52 | | RP | NEG | | | 853896 | 392069 | 928207 | 419193 | 1.83E-01 | 0.18 | 0.87 |
| RP_POS_524_1626a8_274 | | 524.1626 | 8.27 | | RP | POS | | | 750637 | 76724.6 | 765237 | 82565.4 | 1.83E-01 | 0.18 | 1.2 |
| RP_NEG_268_98557a6_03 | | 268.9856 | 6.03 | | RP | NEG | | | 1646520 | 166467 | 1613750 | 190210 | 1.83E-01 | -0.18 | 0.78 |
| RP_POS_458_33163a7_049 | | 458.3316 | 7.05 | | RP | POS | | | 871674 | 599513 | 771454 | 503726 | 1.83E-01 | -0.18 | 0.89 |
| RP_POS_713_97998a10_378 | | 713.9800 | 10.38 | | RP | POS | | | 927412 | 513430 | 840318 | 445622 | 1.83E-01 | -0.18 | 1.05 |
| RP_POS_701_8241a11_594 | | 701.8241 | 11.59 | | RP | POS | | | 184856 | 74056.2 | 203420 | 120603 | 1.84E-01 | 0.19 | 1.14 |
| RP_POS_350_32593a10_685 | | 350.3259 | 10.69 | | RP | POS | | | 3245832 | 331415 | 3312115 | 390215 | 1.84E-01 | 0.18 | 0.94 |
| RP_POS_494_31955a11_853 | | 494.3196 | 11.85 | | RP | POS | | | 1988660 | 240399 | 2034891 | 265071 | 1.84E-01 | 0.18 | 0.96 |
| HILIC_NEG_144_92355a1_301 | | 144.9236 | 1.30 | | HILIC | NEG | | | 9904122 | 1746625 | 9605055 | 1547670 | 1.84E-01 | -0.18 | 0.91 |
| RP_POS_311_29401a11_33 | | 311.2940 | 11.33 | | RP | POS | | | 1013057 | 286639 | 970588 | 175766 | 1.84E-01 | -0.18 | 0.79 |
| RP_POS_351_18704a11_14 | | 351.1870 | 11.14 | | RP | POS | | | 295629 | 129597 | 276892 | 73201.6 | 1.84E-01 | -0.18 | 0.87 |
| RP_POS_524_04883a10_733 | | 524.0488 | 10.73 | | RP | POS | | | 130168 | 142430 | 166111 | 234615 | 1.85E-01 | 0.19 | 0.93 |
| RP_POS_441_29755a11_838 | | 441.2976 | 11.84 | | RP | POS | | | 4731053 | 3041402 | 5347977 | 3673495 | 1.85E-01 | 0.18 | 1.02 |
| HILIC_NEG_144_92355a0_933 | | 144.9236 | 0.93 | | HILIC | NEG | | | 2.9E+07 | 2833122 | 2.8E+07 | 3089627 | 1.85E-01 | -0.18 | 0.83 |
| RP_NEG_376_98572a7_788 | | 376.9857 | 7.79 | | RP | NEG | | | 2071907 | 486694 | 1988982 | 429240 | 1.85E-01 | -0.18 | 0.86 |
| RP_POS_406_23779a11_144 | | 406.2378 | 11.14 | | RP | POS | | | 4816530 | 819892 | 4665928 | 835534 | 1.85E-01 | -0.18 | 1.29 |
| RP_POS_748_52728a13_055 | | 748.5273 | 13.06 | | RP | POS | | | 1828011 | 2431516 | 1443057 | 1814619 | 1.85E-01 | -0.18 | 0.89 |
| RP_POS_754_5379a12_51 | | 754.5379 | 12.51 | | RP | POS | | | 2.7E+07 | 1.2E+07 | 2.5E+07 | 9833762 | 1.85E-01 | -0.18 | 0.83 |
| RP_NEG_313_16507a10_668 | | 313.1651 | 10.67 | | RP | NEG | | | 247563 | 892863 | 136775 | 100769 | 1.85E-01 | -0.22 | 0.69 |
| RP_POS_1001_65631a10_601 | | 1001.6563 | 10.60 | | RP | POS | | | 1959737 | 1129568 | 2180515 | 1287354 | 1.86E-01 | 0.18 | 0.93 |
| RP_POS_329_24493a11_115 | | 329.2449 | 11.12 | | RP | POS | | | 1588860 | 856282 | 1746883 | 885018 | 1.86E-01 | 0.18 | 1.27 |
| RP_POS_610_71057a11_34 | | 610.7106 | 11.34 | | RP | POS | | | 126370 | 74475.4 | 140486 | 80582.4 | 1.86E-01 | 0.18 | 0.79 |
| HILIC_POS_140_06841a3_616 | | 140.0684 | 3.62 | | HILIC | POS | 140.069(100) | | 1.8E+07 | 4836339 | 1.7E+07 | 5502985 | 1.86E-01 | -0.18 | 0.89 |

**Supplemantary table 1**: The non-targeted metabolomics results ordered by the VIP value (p value < 0.0003 are shaded dark green, p value < 0.05 are shaded light green, d values below 0 are blue and above 0 are red)

| Feature_ID | Metabolite_name | Average_Mz | Average_RT_min_ | HMDB ID | RP_HILIC | POS_NEG | Cureted_MSMS | ID_level | Cases Mean | Cases SD | Controls Mean2 | Controls SD3 | Welch test p | Cohen's d | VIP |
|---|---|---|---|---|---|---|---|---|---|---|---|---|---|---|---|
| RP_POS_301_211a11_144 | | 301.2110 | 11.14 | | RP | POS | 301.2098(100) | | 5583247 | 784081 | 5432853 | 866302 | 1.86E-01 | -0.18 | 0.97 |
| HILIC_POS_219_98882a0_931 | | 219.9888 | 0.93 | | HILIC | POS | | | 829246 | 411672 | 759040 | 365137 | 1.86E-01 | -0.18 | 1 |
| HILIC_POS_312_21692a0_8 | | 312.2169 | 0.80 | | HILIC | POS | | | 1355385 | 1243136 | 1178130 | 674450 | 1.86E-01 | -0.18 | 0.89 |
| RP_NEG_337_23724a11_361 | | 337.2372 | 11.36 | | RP | NEG | | | 428559 | 126415 | 404296 | 139756 | 1.86E-01 | -0.18 | 1.41 |
| RP_NEG_365_23135a10_155 | | 365.2314 | 10.16 | | RP | NEG | | | 392740 | 71264.7 | 380495 | 64280.2 | 1.86E-01 | -0.18 | 1.2 |
| RP_POS_154_0499a0_682 | | 154.0499 | 0.68 | | RP | POS | | | 692744 | 420197 | 623223 | 350190 | 1.86E-01 | -0.18 | 0.76 |
| RP_POS_215_1274a6_056 | | 215.1274 | 6.06 | | RP | POS | | | 1498530 | 313846 | 1444079 | 289004 | 1.86E-01 | -0.18 | 1.06 |
| RP_POS_324_25955a10_944 | | 324.2596 | 10.94 | | RP | POS | | | 749102 | 321580 | 689982 | 330732 | 1.86E-01 | -0.18 | 1.03 |
| RP_POS_526_39807a14_356 | | 526.3981 | 14.36 | | RP | POS | | | 2700717 | 2861158 | 2255860 | 2054416 | 1.86E-01 | -0.18 | 0.93 |
| RP_POS_589_73413a10_405 | | 589.7341 | 10.41 | | RP | POS | | | 538273 | 336597 | 483615 | 268910 | 1.86E-01 | -0.18 | 0.89 |
| RP_NEG_409_2366a11_418 | | 409.2366 | 11.42 | | RP | NEG | | | 21735.8 | 37362 | 38197.7 | 118364 | 1.87E-01 | 0.21 | 1.03 |
| HILIC_POS_176_9995a1_388 | | 176.9995 | 1.39 | | HILIC | POS | | | 298671 | 133980 | 322920 | 134527 | 1.87E-01 | 0.18 | 0.88 |
| RP_NEG_286_84601a15_402 | | 286.8460 | 15.40 | | RP | NEG | | | 1065157 | 369215 | 1129958 | 349391 | 1.87E-01 | 0.18 | 1.03 |
| RP_NEG_487_32837a9_789 | | 487.3284 | 9.79 | | RP | NEG | | | 268860 | 159175 | 295547 | 137378 | 1.87E-01 | 0.18 | 0.9 |
| RP_NEG_834_56708a10_649 | | 834.5671 | 10.65 | | RP | NEG | | | 394664 | 234300 | 437185 | 235809 | 1.87E-01 | 0.18 | 1.4 |
| RP_POS_1070_66003a10_607 | | 1070.6600 | 10.61 | | RP | POS | | | 443532 | 428644 | 517530 | 392287 | 1.87E-01 | 0.18 | 0.96 |
| RP_NEG_792_53412a13_212 | PC 34:2 (16:0_18:2) | 792.5341 | 13.21 | | RP | NEG | 279.2337(100), 255.2329(40) | 2 | 3414400 | 1107011 | 3623189 | 1195928 | 1.87E-01 | 0.18 | 0.97 |
| HILIC_NEG_130_99268a0_934 | | 130.9927 | 0.93 | | HILIC | NEG | | | 2650587 | 237735 | 2609898 | 213923 | 1.87E-01 | -0.18 | 0.82 |
| HILIC_NEG_153_86879a6_307 | | 153.8688 | 6.31 | | HILIC | NEG | | | 2335049 | 1236832 | 2130754 | 1031805 | 1.87E-01 | -0.18 | 1.02 |
| RP_NEG_355_2858a10_776 | | 355.2858 | 10.78 | | RP | NEG | | | 475970 | 48239.4 | 466261 | 57884.5 | 1.87E-01 | -0.18 | 0.88 |
| RP_POS_442_33716a8_371 | | 442.3372 | 8.37 | | RP | POS | | | 642776 | 256836 | 596666 | 253170 | 1.87E-01 | -0.18 | 1.11 |
| RP_POS_282_27853a14_248 | | 282.2785 | 14.25 | | RP | POS | 282.2776(100), 83.0863(70) | | 2818430 | 2022662 | 3172932 | 1911849 | 1.88E-01 | 0.18 | 1.03 |
| HILIC_NEG_158_84665a3_175 | | 158.8467 | 3.18 | | HILIC | NEG | | | 626463 | 248747 | 667086 | 202894 | 1.88E-01 | 0.18 | 0.97 |
| RP_NEG_1061_68005a10_487 | | 1061.6801 | 10.49 | | RP | NEG | | | 2479122 | 564266 | 2586007 | 615533 | 1.88E-01 | 0.18 | 0.94 |
| RP_NEG_986_53638a13_006 | | 986.5364 | 13.01 | | RP | NEG | | | 2505698 | 888192 | 2666958 | 898825 | 1.88E-01 | 0.18 | 0.8 |
| RP_POS_782_56989a12_876 | PC 36:4 (18:2/18:2) | 782.5699 | 12.88 | | RP | POS | 184.0746(100), 782.5822(2) | 1 | 2E+08 | 7.4E+07 | 1.9E+08 | 6.1E+07 | 1.88E-01 | -0.18 | 0.8 |
| RP_NEG_478_53024a15_493 | | 478.5302 | 15.49 | | RP | NEG | | | 775917 | 43213.2 | 768671 | 37435.8 | 1.88E-01 | -0.18 | 0.88 |
| RP_POS_236_07161a9_394 | | 236.0716 | 9.39 | | RP | POS | | | 6.8E+07 | 2.4E+07 | 6.4E+07 | 2.5E+07 | 1.88E-01 | -0.18 | 0.79 |
| RP_POS_643_44489a10_268 | | 643.4449 | 10.27 | | RP | POS | | | 725635 | 354817 | 654917 | 421587 | 1.88E-01 | -0.18 | 1.09 |
| HILIC_POS_192_06184a7_419 | | 192.0618 | 7.42 | | HILIC | POS | | | 2323329 | 362990 | 2386749 | 342748 | 1.89E-01 | 0.18 | 0.9 |
| RP_NEG_134_89476a0_578 | | 134.8948 | 0.58 | | RP | NEG | | | 4620510 | 4443449 | 5494135 | 5170296 | 1.89E-01 | 0.18 | 0.88 |
| RP_POS_356_03119a15_431 | | 356.0312 | 15.43 | | RP | POS | | | 4143634 | 1366012 | 4442196 | 1869870 | 1.89E-01 | 0.18 | 0.92 |
| HILIC_POS_111_02062a0_931 | | 111.0206 | 0.93 | | HILIC | POS | | | 1114585 | 757481 | 991310 | 616323 | 1.89E-01 | -0.18 | 0.99 |
| RP_NEG_418_89453a15_344 | | 418.8945 | 15.34 | | RP | NEG | | | 468641 | 121702 | 446874 | 120280 | 1.89E-01 | -0.18 | 1.14 |
| RP_NEG_814_51709a12_735 | | 814.5171 | 12.74 | | RP | NEG | | | 1184487 | 483313 | 1104501 | 409501 | 1.89E-01 | -0.18 | 0.8 |
| RP_POS_501_37561a10_928 | | 501.3756 | 10.93 | | RP | POS | | | 2795696 | 598766 | 2695822 | 515179 | 1.89E-01 | -0.18 | 1.24 |
| RP_POS_659_48157a11_677 | | 659.4816 | 11.68 | | RP | POS | | | 376124 | 438999 | 313779 | 241155 | 1.89E-01 | -0.18 | 0.74 |
| HILIC_NEG_128_95969a2_995 | | 128.9597 | 3.00 | | HILIC | NEG | | | 5.8E+07 | 6309546 | 5.9E+07 | 6144312 | 1.90E-01 | 0.18 | 0.83 |
| RP_POS_182_95061a0_562 | | 182.9506 | 0.56 | | RP | POS | | | 486292 | 218160 | 523344 | 196041 | 1.90E-01 | 0.18 | 1.06 |
| RP_NEG_297_24341a10_219 | | 297.2434 | 10.22 | | RP | NEG | | | 2885941 | 530294 | 2789538 | 541835 | 1.90E-01 | -0.18 | 0.96 |
| RP_POS_136_11208a2_955 | | 136.1121 | 2.96 | | RP | POS | | | 476298 | 393061 | 408162 | 367426 | 1.90E-01 | -0.18 | 0.83 |
| RP_POS_195_09901a5_556 | | 195.0990 | 5.56 | | RP | POS | | | 578183 | 285364 | 530427 | 248367 | 1.90E-01 | -0.18 | 0.94 |
| RP_POS_293_20862a9_939 | | 293.2086 | 9.94 | | RP | POS | | | 2364219 | 312805 | 2308854 | 304826 | 1.90E-01 | -0.18 | 0.88 |
| RP_POS_374_19455a8_842 | | 374.1946 | 8.84 | | RP | POS | | | 444481 | 596868 | 355576 | 388790 | 1.90E-01 | -0.18 | 1.1 |
| RP_POS_415_16437a9_207 | | 415.1644 | 9.21 | | RP | POS | | | 264467 | 204359 | 229689 | 183884 | 1.90E-01 | -0.18 | 0.8 |
| RP_POS_135_04401a4_155 | | 135.0440 | 4.16 | | RP | POS | | | 4031199 | 992101 | 4213282 | 1036317 | 1.91E-01 | 0.18 | 0.91 |
| RP_POS_308_09076a15_536 | | 308.0908 | 15.54 | | RP | POS | | | 428567 | 201753 | 470081 | 254063 | 1.91E-01 | 0.18 | 0.94 |
| RP_POS_407_24054a10_389 | | 407.2405 | 10.39 | | RP | POS | | | 1.2E+07 | 5961075 | 1.3E+07 | 6314524 | 1.91E-01 | 0.18 | 1.22 |
| RP_POS_595_35083a14_347 | | 595.3508 | 14.35 | | RP | POS | | | 616495 | 416921 | 687607 | 379713 | 1.91E-01 | 0.18 | 0.83 |
| RP_NEG_824_54626a12_74 | | 824.5463 | 12.74 | | RP | NEG | | | 4.5E+07 | 1.9E+07 | 4.2E+07 | 1.5E+07 | 1.91E-01 | -0.18 | 0.79 |
| RP_POS_576_27966a10_528 | | 576.2797 | 10.53 | | RP | POS | | | 472846 | 181004 | 442401 | 159785 | 1.91E-01 | -0.18 | 0.9 |
| RP_POS_1194_32361a13_017 | | 1194.3236 | 13.02 | | RP | POS | | | 889354 | 632246 | 1008383 | 692258 | 1.92E-01 | 0.18 | 0.84 |
| RP_POS_572_41541a10_869 | | 572.4154 | 10.87 | | RP | POS | | | 416180 | 236181 | 466149 | 311472 | 1.92E-01 | 0.18 | 1.33 |
| RP_POS_641_45032a10_605 | | 641.4503 | 10.61 | | RP | POS | | | 193124 | 123866 | 215663 | 127934 | 1.92E-01 | 0.18 | 0.85 |
| RP_POS_666_13312a10_454 | | 666.1331 | 10.45 | | RP | POS | | | 170437 | 100003 | 190532 | 121722 | 1.92E-01 | 0.18 | 0.81 |
| RP_POS_742_11963a11_562 | | 742.1196 | 11.56 | | RP | POS | | | 154291 | 75665 | 169401 | 91219.5 | 1.92E-01 | 0.18 | 0.86 |
| HILIC_NEG_144_99213a0_935 | | 144.9921 | 0.94 | | HILIC | NEG | | | 2534940 | 269796 | 2489025 | 245316 | 1.92E-01 | -0.18 | 0.94 |
| HILIC_NEG_176_99847a0_934 | | 176.9985 | 0.93 | | HILIC | NEG | | | 1215495 | 126074 | 1193140 | 124383 | 1.92E-01 | -0.18 | 0.83 |
| RP_POS_363_21442a7_932 | | 363.2144 | 7.93 | | RP | POS | | | 139289 | 94716 | 124979 | 65210.5 | 1.92E-01 | -0.18 | 0.88 |
| RP_POS_1017_68738a10_606 | | 1017.6874 | 10.61 | | RP | POS | | | 2484105 | 1220404 | 2707189 | 1274525 | 1.93E-01 | 0.18 | 1 |
| RP_POS_192_1228a6_085 | | 192.1228 | 6.09 | | RP | POS | | | 59256 | 27503.7 | 64792.6 | 33711 | 1.93E-01 | 0.18 | 0.9 |
| RP_POS_297_16953a10_25 | | 297.1695 | 10.25 | | RP | POS | | | 2111394 | 1045925 | 2299596 | 1064683 | 1.93E-01 | 0.18 | 1.24 |
| RP_POS_440_81403a9_382 | | 440.8140 | 9.38 | | RP | POS | | | 540509 | 260034 | 597011 | 357712 | 1.93E-01 | 0.18 | 0.88 |
| RP_POS_463_12933a10_125 | | 463.1293 | 10.13 | | RP | POS | | | 567743 | 111837 | 588158 | 116798 | 1.93E-01 | 0.18 | 0.88 |
| HILIC_NEG_236_96899a5_655 | | 236.9690 | 5.66 | | HILIC | NEG | | | 357801 | 73672.6 | 346635 | 51357.1 | 1.93E-01 | -0.18 | 0.85 |
| RP_POS_278_16391a9_456 | | 278.1639 | 9.46 | | RP | POS | | | 226796 | 298230 | 297646 | 466344 | 1.94E-01 | 0.19 | 0.91 |
| RP_NEG_295_26425a11_45 | | 295.2643 | 11.45 | | RP | NEG | | | 85137.3 | 88696.7 | 102526 | 104597 | 1.94E-01 | 0.18 | 1.17 |
| RP_POS_417_79745a14_46 | | 417.7975 | 14.46 | | RP | POS | | | 140065 | 119825 | 163196 | 138141 | 1.94E-01 | 0.18 | 0.84 |
| HILIC_POS_160_13315a2_019 | 5-amino valeric acid betaine (5-AVAB) | 160.1332 | 2.02 | HMDB0240732 | HILIC | POS | 160.1326(100), 101.06(35) | 1 | 1.2E+08 | 5.3E+07 | 1.3E+08 | 4.8E+07 | 1.94E-01 | 0.18 | 0.91 |
| RP_POS_185_0807a3_35 | | 185.0807 | 3.35 | | RP | POS | | | 5374023 | 1889189 | 5029585 | 1972752 | 1.94E-01 | -0.18 | 1 |
| RP_POS_369_33655a11_021 | | 369.3366 | 11.02 | | RP | POS | | | 1149449 | 171757 | 1118775 | 173014 | 1.94E-01 | -0.18 | 1.33 |
| HILIC_NEG_118_92437a10_858 | | 118.9244 | 10.86 | | HILIC | NEG | | | 2462091 | 1859453 | 2821289 | 2154995 | 1.95E-01 | 0.18 | 0.81 |
| RP_NEG_412_84183a15_407 | | 412.8418 | 15.41 | | RP | NEG | | | 375938 | 75030.3 | 388042 | 61849.2 | 1.95E-01 | 0.18 | 0.93 |
| RP_POS_1063_66882a10_504 | | 1063.6688 | 10.50 | | RP | POS | | | 981100 | 372147 | 1049428 | 395669 | 1.95E-01 | 0.18 | 0.9 |
| RP_POS_266_08337a0_585 | | 266.0834 | 0.59 | | RP | POS | | | 180923 | 122984 | 205364 | 148627 | 1.95E-01 | 0.18 | 0.99 |
| RP_POS_287_6507a0_678 | | 287.6507 | 0.68 | | RP | POS | | | 476648 | 244958 | 526796 | 310018 | 1.95E-01 | 0.18 | 1.22 |
| RP_POS_369_31161a8_644 | | 369.3116 | 8.64 | | RP | POS | | | 2270915 | 88771.1 | 2286381 | 85775.9 | 1.95E-01 | 0.18 | 0.86 |
| RP_POS_423_21414a10_387 | | 423.2141 | 10.39 | | RP | POS | | | 675208 | 317264 | 732961 | 331643 | 1.95E-01 | 0.18 | 1.14 |
| RP_POS_500_35532a10_053 | | 500.3553 | 10.05 | | RP | POS | | | 1078271 | 536001 | 1175940 | 561426 | 1.95E-01 | 0.18 | 0.87 |
| RP_POS_738_52264a12_028 | | 738.5226 | 12.03 | | RP | POS | | | 189672 | 92339.6 | 205212 | 83312.2 | 1.95E-01 | 0.18 | 0.87 |
| HILIC_NEG_566_34705a1_188 | LysoPC 18:1 | 566.3471 | 1.19 | | HILIC | NEG | 281.2491(100), 506.3268(20) | 2 | 2159232 | 530327 | 2256588 | 562548 | 1.95E-01 | 0.18 | 0.88 |
| RP_POS_279_15872a11_408 | | 279.1587 | 11.41 | | RP | POS | 149.023(100), 71.0862(2) | | 4641746 | 820706 | 4511620 | 648294 | 1.95E-01 | -0.18 | 0.8 |
| HILIC_NEG_186_10365a3_045 | | 186.1037 | 3.05 | | HILIC | NEG | | | 3271386 | 606432 | 3160530 | 638678 | 1.95E-01 | -0.18 | 0.89 |
| HILIC_NEG_207_83649a3_248 | | 207.8365 | 3.25 | | HILIC | NEG | | | 1230869 | 296293 | 1181090 | 266549 | 1.95E-01 | -0.18 | 0.81 |
| HILIC_POS_61_04055a0_934 | | 61.0406 | 0.93 | | HILIC | POS | | | 5.3E+07 | 1.4E+07 | 5.1E+07 | 1.3E+07 | 1.95E-01 | -0.18 | 0.94 |
| RP_POS_231_11572a9_326 | | 231.1157 | 9.33 | | RP | POS | | | 4475792 | 2674876 | 4037231 | 2280859 | 1.95E-01 | -0.18 | 0.89 |
| RP_POS_744_58997a13_786 | | 744.5900 | 13.79 | | RP | POS | | | 1.7E+07 | 6464630 | 1.6E+07 | 6179462 | 1.95E-01 | -0.18 | 0.74 |
| HILIC_NEG_298_88095a2_997 | | 298.8810 | 3.00 | | HILIC | NEG | | | 789195 | 147127 | 815228 | 146670 | 1.96E-01 | 0.18 | 0.78 |
| HILIC_POS_200_05424a1_378 | | 200.0542 | 1.38 | | HILIC | POS | | | 1324767 | 456979 | 1406131 | 460688 | 1.96E-01 | 0.18 | 0.86 |
| RP_POS_262_91568a15_388 | | 262.9157 | 15.39 | | RP | POS | | | 3254149 | 1444061 | 3490123 | 1227367 | 1.96E-01 | 0.18 | 0.84 |
| RP_POS_337_18173a11_238 | | 337.1817 | 11.24 | | RP | POS | | | 2172589 | 1005819 | 2350558 | 1004916 | 1.96E-01 | 0.18 | 1.09 |
| RP_POS_369_20901a11_241 | | 369.2090 | 11.24 | | RP | POS | | | 2993806 | 1252946 | 3222533 | 1324567 | 1.96E-01 | 0.18 | 1.05 |
| HILIC_POS_154_04974a0_734 | C7H7NO3 | 154.0497 | 0.73 | | HILIC | POS | 154.0493(100),136.0317(30) | 4 | 687438 | 416232 | 618516 | 364374 | 1.96E-01 | -0.18 | 0.75 |
| HILIC_NEG_113_9264a0_933 | | 113.9264 | 0.93 | | HILIC | NEG | | | 1782878 | 194689 | 1747903 | 200117 | 1.96E-01 | -0.18 | 0.84 |
| HILIC_NEG_68_99506a0_934 | | 68.9951 | 0.93 | | HILIC | NEG | | | 2595723 | 269961 | 2549382 | 254165 | 1.96E-01 | -0.18 | 0.76 |
| RP_NEG_709_5061a12_23 | | 709.5061 | 12.23 | | RP | NEG | | | 390312 | 102842 | 374116 | 80428.2 | 1.96E-01 | -0.18 | 0.83 |
| RP_NEG_828_57751a13_384 | | 828.5775 | 13.38 | | RP | NEG | | | 4.8E+07 | 1.8E+07 | 4.5E+07 | 1.4E+07 | 1.96E-01 | -0.18 | 0.76 |
| RP_POS_142_12267a10_359 | | 142.1227 | 10.36 | | RP | POS | | | 569188 | 157499 | 542150 | 148582 | 1.96E-01 | -0.18 | 0.76 |
| RP_POS_690_48004a9_913 | | 690.4800 | 9.91 | | RP | POS | | | 2163684 | 1071464 | 1984340 | 959903 | 1.96E-01 | -0.18 | 0.79 |
| HILIC_POS_140_06818a5_901 | | 140.0682 | 5.90 | | HILIC | POS | 140.069(100), 74.0974(40) | | 504623 | 449099 | 590319 | 513779 | 1.97E-01 | 0.18 | 0.88 |
| HILIC_NEG_192_98953a5_113 | | 192.9895 | 5.11 | | HILIC | NEG | | | 302378 | 138704 | 329285 | 162673 | 1.97E-01 | 0.18 | 1.35 |
| RP_POS_175_07521a6_385 | | 175.0752 | 6.39 | | RP | POS | | | 1348479 | 153125 | 1376085 | 158790 | 1.97E-01 | 0.18 | 0.97 |
| RP_POS_380_20901a11_239 | | 380.2090 | 11.24 | | RP | POS | | | 136057 | 106664 | 154150 | 98390.2 | 1.97E-01 | 0.18 | 0.96 |
| RP_POS_522_79602a10_356 | | 522.7960 | 10.36 | | RP | POS | | | 710664 | 229872 | 755020 | 267110 | 1.97E-01 | 0.18 | 1.12 |
| HILIC_POS_198_08493a6_383 | N-Acetylhistidine | 198.0849 | 6.38 | | HILIC | POS | 198.1861(100) | 2 | 1159270 | 400102 | 1232934 | 429962 | 1.97E-01 | 0.18 | 0.8 |
| HILIC_POS_118_08642a0_749 | | 118.0864 | 0.75 | | HILIC | POS | 118.0864(100), 95.0497(10) | | 2871727 | 1648739 | 2613965 | 1275212 | 1.97E-01 | -0.18 | 1.06 |
| RP_NEG_241_21716a10_919 | | 241.2172 | 10.92 | | RP | NEG | | | 154731 | 68191.8 | 142848 | 66531.3 | 1.97E-01 | -0.18 | 1.13 |
| RP_NEG_294_87848a15_388 | | 294.8785 | 15.39 | | RP | NEG | | | 813215 | 1068504 | 659358 | 656349 | 1.97E-01 | -0.18 | 0.93 |
| RP_POS_358_29492a8_507 | | 358.2949 | 8.51 | | RP | POS | | | 1702759 | 1034696 | 1529182 | 933621 | 1.97E-01 | -0.18 | 0.85 |
| RP_POS_402_76508a13_426 | | 402.7651 | 13.43 | | RP | POS | | | 682194 | 604572 | 589078 | 450061 | 1.97E-01 | -0.18 | 0.83 |
| RP_POS_516_35626a8_424 | | 516.3563 | 8.42 | | RP | POS | | | 353405 | 224960 | 318431 | 171995 | 1.97E-01 | -0.18 | 0.88 |
| RP_POS_684_55573a14_593 | | 684.5557 | 14.59 | | RP | POS | | | 343677 | 530070 | 267456 | 325056 | 1.97E-01 | -0.18 | 0.78 |
| RP_POS_707_29968a6_034 | | 707.2997 | 6.03 | | RP | POS | | | 1008947 | 558955 | 913626 | 521928 | 1.97E-01 | -0.18 | 1.18 |
| HILIC_POS_362_92593a5_463 | | 362.9259 | 5.46 | | HILIC | POS | | | 175827 | 304715 | 262368 | 603257 | 1.98E-01 | 0.19 | 0.8 |
| RP_POS_245_22627a10_637 | | 245.2263 | 10.64 | | RP | POS | | | 1432609 | 700226 | 1605237 | 1162704 | 1.98E-01 | 0.19 | 0.92 |
| HILIC_POS_371_24014a3_288 | | 371.2401 | 3.29 | | HILIC | POS | | | 526157 | 211881 | 566663 | 244027 | 1.98E-01 | 0.18 | 0.92 |
| RP_NEG_331_17648a8_052 | | 331.1765 | 8.05 | | RP | NEG | | | 1509667 | 2308622 | 2003867 | 3153641 | 1.98E-01 | 0.18 | 1.11 |
| RP_POS_163_13264a5_731 | | 163.1326 | 5.73 | | RP | POS | | | 6.5E+07 | 4005265 | 6.6E+07 | 4316010 | 1.98E-01 | 0.18 | 1.09 |
| RP_POS_279_09283a7_8 | Triphenylphosphine oxide | 279.0928 | 7.80 | | RP | POS | 279.0922(100), 219.0564(30) | 2 | 1872245 | 145585 | 1903076 | 196088 | 1.98E-01 | 0.18 | 0.91 |
| RP_POS_151_07547a8_532 | 1-(4-methoxyphenyl)ethanone | 151.0755 | 8.53 | | RP | POS | 151.0748(100) | 2 | 1131109 | 99034.2 | 1148832 | 101540 | 1.98E-01 | 0.18 | 0.8 |
| HILIC_POS_419_31122a5_661 | | 419.3112 | 5.66 | | HILIC | POS | | | 33612.4 | 34332.3 | 27891.2 | 30796.5 | 1.98E-01 | -0.18 | 1.21 |
| RP_NEG_1121_60974a10_49 | | 1121.6097 | 10.49 | | RP | NEG | | | 696035 | 106054 | 675552 | 123917 | 1.98E-01 | -0.18 | 1.02 |
| RP_NEG_243_19676a10_1 | | 243.1968 | 10.10 | | RP | NEG | | | 370191 | 45475.2 | 362741 | 39391.5 | 1.98E-01 | -0.18 | 0.78 |
| RP_POS_283_20294a10_589 | | 283.2029 | 10.59 | | RP | POS | | | 1418424 | 239403 | 1376459 | 237206 | 1.98E-01 | -0.18 | 0.95 |
| RP_POS_497_86646a10_424 | | 497.8665 | 10.42 | | RP | POS | | | 690986 | 357662 | 632825 | 304741 | 1.98E-01 | -0.18 | 0.96 |
| RP_POS_534_31665a8_873 | | 534.3167 | 8.87 | | RP | POS | | | 970318 | 388359 | 908597 | 313625 | 1.98E-01 | -0.18 | 0.83 |
| RP_POS_181_04962a9_44 | | 181.0496 | 9.44 | | RP | POS | | | 1995400 | 169611 | 2024618 | 162925 | 1.99E-01 | 0.18 | 1 |
| RP_POS_418_24954a13_11 | | 418.2495 | 13.11 | | RP | POS | | | 245676 | 193581 | 285973 | 255009 | 1.99E-01 | 0.18 | 1.15 |
| RP_POS_467_2616a9_742 | | 467.2616 | 9.74 | | RP | POS | | | 813803 | 481570 | 898751 | 484021 | 1.99E-01 | 0.18 | 1.17 |
| RP_POS_186_10262a0_675 | Ecgonine | 186.1026 | 0.68 | | RP | POS | 186.1119(100), 168.103(10) | 2 | 536419 | 445359 | 619705 | 494764 | 1.99E-01 | 0.18 | 0.77 |
| RP_NEG_279_03488a0_68 | | 279.0349 | 0.68 | | RP | NEG | | | 974446 | 168233 | 942300 | 193700 | 1.99E-01 | -0.18 | 0.92 |
| RP_NEG_355_15891a11_142 | | 355.1589 | 11.14 | | RP | NEG | | | 2.3E+07 | 2022749 | 2.3E+07 | 1981223 | 1.99E-01 | -0.18 | 1.11 |
| RP_NEG_654_30298a10_015 | | 654.3030 | 10.02 | | RP | NEG | | | 620493 | 313053 | 573403 | 222620 | 1.99E-01 | -0.18 | 0.78 |
| RP_POS_247_09229a0_674 | | 247.0923 | 0.67 | | RP | POS | | | 327403 | 78117.8 | 315130 | 61954.3 | 1.99E-01 | -0.18 | 0.92 |
| RP_POS_286_229a10_844 | | 286.2290 | 10.84 | | RP | POS | | | 3949583 | 655875 | 3826651 | 733462 | 1.99E-01 | -0.18 | 0.95 |
| RP_POS_607_41406a9_942 | | 607.4141 | 9.94 | | RP | POS | | | 1022599 | 473507 | 938311 | 483872 | 1.99E-01 | -0.18 | 1.04 |
| RP_POS_388_26938a10_251 | | 388.2694 | 10.25 | | RP | POS | | | 8442167 | 4286679 | 9204497 | 4392986 | 2.00E-01 | 0.18 | 1.24 |
| RP_POS_494_34766a10_99 | | 494.3477 | 10.99 | | RP | POS | | | 740458 | 486501 | 851663 | 733757 | 2.00E-01 | 0.18 | 1.17 |
| RP_POS_762_50366a13_046 | | 762.5037 | 13.05 | | RP | POS | | | 2295613 | 992073 | 2492724 | 1221443 | 2.00E-01 | 0.18 | 1.16 |
| RP_POS_561_39722a10_509 | | 561.3972 | 10.51 | | RP | POS | | | 500887 | 369204 | 443014 | 292654 | 2.00E-01 | -0.17 | 0.82 |
| RP_POS_186_14919a10_36 | | 186.1492 | 10.36 | | RP | POS | | | 1906450 | 406008 | 1835152 | 406259 | 2.00E-01 | -0.18 | 0.84 |
| RP_POS_295_2243a10_469 | | 295.2243 | 10.47 | | RP | POS | | | 944201 | 200633 | 908376 | 207290 | 2.00E-01 | -0.18 | 1.26 |
| RP_POS_304_24762a9_207 | | 304.2476 | 9.21 | | RP | POS | | | 918721 | 427522 | 841343 | 451524 | 2.00E-01 | -0.18 | 1.15 |
| RP_POS_417_36841a11_026 | | 417.3684 | 11.03 | | RP | POS | | | 426362 | 806004 | 306906 | 553888 | 2.00E-01 | -0.18 | 0.86 |
| RP_POS_485_35522a9_824 | | 485.3552 | 9.82 | | RP | POS | | | 7762864 | 3321458 | 7198124 | 3131128 | 2.00E-01 | -0.18 | 0.94 |
| RP_POS_609_3078a10_928 | | 609.3078 | 10.93 | | RP | POS | | | 804256 | 297924 | 753287 | 283743 | 2.00E-01 | -0.18 | 1.02 |
| HILIC_POS_482_32419a1_792 | | 482.3242 | 1.79 | | HILIC | POS | | | 1148195 | 2936676 | 1784959 | 4128021 | 2.01E-01 | 0.18 | 0.81 |
| RP_NEG_486_28046a13_292 | | 486.2805 | 13.29 | | RP | NEG | | | 415633 | 81801.3 | 430641 | 88668.3 | 2.01E-01 | 0.18 | 1.09 |
| RP_POS_205_12234a7_812 | | 205.1223 | 7.81 | | RP | POS | | | 1411591 | 151961 | 1441216 | 182600 | 2.01E-01 | 0.18 | 0.75 |
| RP_NEG_403_15857a10_929 | | 403.1586 | 10.93 | | RP | NEG | | | 422649 | 216796 | 459619 | 205861 | 2.01E-01 | 0.17 | 1.13 |
| RP_POS_287_20053a6_255 | | 287.2005 | 6.26 | | RP | POS | | | 372836 | 359383 | 433604 | 336107 | 2.01E-01 | 0.17 | 0.82 |
| RP_POS_295_11566a3_985 | | 295.1157 | 3.99 | | RP | POS | 295.1154(100) | | 177929 | 133585 | 157209 | 103574 | 2.01E-01 | -0.17 | 1.22 |
| RP_NEG_257_21277a10_201 | | 257.2128 | 10.20 | | RP | NEG | | | 444802 | 56668.7 | 435443 | 50431.9 | 2.01E-01 | -0.17 | 1.18 |
| RP_POS_120_03733a8_846 | | 120.0373 | 8.85 | | RP | POS | | | 171602 | 82112.8 | 158085 | 72621.2 | 2.01E-01 | -0.17 | 0.84 |
| RP_POS_147_06532a9_21 | | 147.0653 | 9.21 | | RP | POS | | | 771130 | 416694 | 698679 | 411753 | 2.01E-01 | -0.17 | 0.87 |
| RP_POS_471_34702a10_452 | | 471.3470 | 10.45 | | RP | POS | | | 1420434 | 3507950 | 935153 | 1935224 | 2.01E-01 | -0.18 | 0.76 |
| RP_POS_567_74078a10_677 | | 567.7408 | 10.68 | | RP | POS | | | 217210 | 125840 | 241449 | 148862 | 2.02E-01 | 0.18 | 0.9 |
| RP_POS_236_07161a13_308 | | 236.0716 | 13.31 | | RP | POS | | | 1.4E+08 | 3.3E+07 | 1.4E+08 | 3.2E+07 | 2.02E-01 | 0.17 | 0.98 |
| RP_POS_341_1778a11_239 | | 341.1778 | 11.24 | | RP | POS | | | 1554673 | 822541 | 1692353 | 756784 | 2.02E-01 | 0.17 | 1.06 |
| RP_POS_371_20871a11_24 | | 371.2087 | 11.24 | | RP | POS | | | 3314852 | 1588407 | 3583796 | 1498487 | 2.02E-01 | 0.17 | 1.06 |
| HILIC_NEG_151_02641a1_701 | Xanthine | 151.0264 | 1.70 | | HILIC | NEG | 151.0258(100), 108.0208(40) | 2 | 891189 | 203962 | 928944 | 225674 | 2.02E-01 | 0.18 | 1.02 |
| RP_POS_348_12222a8_844 | | 348.1222 | 8.84 | | RP | POS | | | 1012894 | 562801 | 921752 | 483624 | 2.02E-01 | -0.17 | 1.21 |

**Supplemantary table 1**: The non-targeted metabolomics results ordered by the VIP value (p value < 0.0003 are shaded dark green, p value < 0.05 are shaded light green, d values below 0 are blue and above 0 are red)

| | | | | | | | | | Cases | | Controls | | Welch test | Cohen's | |
|---|---|---|---|---|---|---|---|---|---|---|---|---|---|---|---|
| Feature_ID | Metabolite_name | Average_Mz | Average_RT_min_ | HMDB ID | RP_HILIC | POS_NEG | Cureted_MSMS | ID_level | Mean | SD | Mean2 | SD3 | p | d | VIP |
| RP_POS_175_98534a15_393 | | 175.9853 | 15.39 | | RP | POS | | | 845980 | 384280 | 1415066 | 4399812 | 2.03E-01 | 0.24 | 1.05 |
| RP_NEG_260_10703a7_289 | | 260.1070 | 7.29 | | RP | NEG | | | 12907 | 57203.4 | 33826.5 | 153988 | 2.03E-01 | 0.2 | 0.84 |
| RP_POS_745_31885a11_38 | | 745.3189 | 11.38 | | RP | POS | | | 137257 | 71204.1 | 150760 | 82380 | 2.03E-01 | 0.18 | 0.85 |
| HILIC_NEG_296_88211a2_995 | | 296.8821 | 3.00 | | HILIC | NEG | | | 3854801 | 726417 | 3980884 | 720479 | 2.03E-01 | 0.17 | 0.79 |
| RP_POS_371_10153a11_008 | | 371.1015 | 11.01 | | RP | POS | | | 393711 | 173849 | 423208 | 165059 | 2.03E-01 | 0.17 | 0.9 |
| RP_POS_544_33698a10_609 | | 544.3370 | 10.61 | | RP | POS | | | 1.6E+08 | 2.8E+07 | 1.7E+08 | 2.9E+07 | 2.03E-01 | 0.17 | 0.88 |
| RP_POS_248_14886a6_06 | | 248.1489 | 6.06 | | RP | POS | | | 389308 | 107356 | 373534 | 72986.3 | 2.03E-01 | -0.17 | 0.79 |
| RP_POS_453_24609a3_525 | | 453.2461 | 3.53 | | RP | POS | | | 7553893 | 4256816 | 6788293 | 4507321 | 2.03E-01 | -0.17 | 1.05 |
| RP_NEG_321_17471a10_178 | | 321.1747 | 10.18 | | RP | NEG | | | 70119 | 143019 | 123543 | 394475 | 2.04E-01 | 0.2 | 0.77 |
| RP_POS_232_09183a11_599 | | 232.0918 | 11.60 | | RP | POS | | | 283190 | 111203 | 305038 | 136269 | 2.04E-01 | 0.18 | 1.15 |
| RP_POS_498_80075a15_413 | | 498.8008 | 15.41 | | RP | POS | | | 355482 | 420260 | 433444 | 470430 | 2.04E-01 | 0.18 | 0.93 |
| RP_POS_539_72131a10_317 | | 539.7213 | 10.32 | | RP | POS | | | 235711 | 164697 | 266228 | 183791 | 2.04E-01 | 0.18 | 0.8 |
| HILIC_NEG_99_92575a1_456 | | 99.9258 | 1.46 | | HILIC | NEG | | | 1443338 | 538557 | 1540615 | 576704 | 2.04E-01 | 0.17 | 0.85 |
| HILIC_POS_175_00185a1_376 | | 175.0019 | 1.38 | | HILIC | POS | | | 4543199 | 2047301 | 4895420 | 2005533 | 2.04E-01 | 0.17 | 0.84 |
| HILIC_POS_392_27921a0_621 | | 392.2792 | 0.62 | | HILIC | POS | | | 181912 | 108328 | 201507 | 115864 | 2.04E-01 | 0.17 | 0.91 |
| RP_NEG_366_9808a9_019 | | 366.9808 | 9.02 | | RP | NEG | | | 748641 | 69193.4 | 761347 | 76128.9 | 2.04E-01 | 0.17 | 0.73 |
| RP_NEG_482_80569a15_388 | | 482.8057 | 15.39 | | RP | NEG | | | 848866 | 197560 | 880716 | 170192 | 2.04E-01 | 0.17 | 1.01 |
| HILIC_POS_136_03931a0_734 | | 136.0393 | 0.73 | | HILIC | POS | | | 729032 | 468978 | 654673 | 389540 | 2.04E-01 | -0.17 | 0.73 |
| RP_NEG_894_54895a12_878 | | 894.5490 | 12.88 | | RP | NEG | | | 4236007 | 1276878 | 4033671 | 1058513 | 2.04E-01 | -0.17 | 0.75 |
| RP_POS_390_13245a9_458 | | 390.1325 | 9.46 | | RP | POS | | | 402304 | 477092 | 329598 | 361074 | 2.04E-01 | -0.17 | 0.94 |
| HILIC_NEG_226_99532a1_355 | | 226.9953 | 1.36 | | HILIC | NEG | | | 1785348 | 488298 | 1873979 | 527694 | 2.05E-01 | 0.17 | 0.74 |
| RP_NEG_292_98306a6_705 | | 292.9831 | 6.71 | | RP | NEG | | | 643462 | 80856 | 656075 | 65041.5 | 2.05E-01 | 0.17 | 0.95 |
| RP_POS_807_56982a13_461 | | 807.5698 | 13.46 | | RP | POS | | | 8.6E+07 | 3.3E+07 | 9.2E+07 | 3E+07 | 2.05E-01 | 0.17 | 1.04 |
| RP_NEG_165_00574a0_677 | | 165.0057 | 0.68 | | RP | NEG | | | 287309 | 46808.1 | 280059 | 36962.1 | 2.05E-01 | -0.17 | 0.73 |
| RP_NEG_471_24289a8_678 | | 471.2429 | 8.68 | | RP | NEG | | | 885363 | 997418 | 722874 | 880084 | 2.05E-01 | -0.17 | 0.84 |
| RP_NEG_613_39862a11_143 | | 613.3986 | 11.14 | | RP | NEG | | | 840462 | 73541.9 | 827977 | 70570 | 2.05E-01 | -0.17 | 1.24 |
| RP_POS_227_23663a10_871 | | 227.2366 | 10.87 | | RP | POS | | | 411220 | 81581.2 | 397552 | 76248 | 2.05E-01 | -0.17 | 1.02 |
| RP_POS_245_13116a10_564 | | 245.1312 | 10.56 | | RP | POS | | | 886903 | 339378 | 835912 | 248965 | 2.05E-01 | -0.17 | 0.89 |
| RP_POS_325_27151a10_86 | | 325.2715 | 10.86 | | RP | POS | | | 2147494 | 417300 | 2070879 | 460971 | 2.05E-01 | -0.17 | 1.09 |
| RP_POS_352_20798a10_635 | | 352.2080 | 10.64 | | RP | POS | | | 384390 | 139199 | 359637 | 145444 | 2.05E-01 | -0.17 | 0.89 |
| RP_POS_643_46106a9_42 | | 643.4611 | 9.42 | | RP | POS | | | 525159 | 316352 | 477356 | 234525 | 2.05E-01 | -0.17 | 0.82 |
| RP_POS_401_28964a10_602 | | 401.2896 | 10.60 | | RP | POS | | | 1226547 | 687835 | 1360822 | 842769 | 2.06E-01 | 0.18 | 1.17 |
| RP_POS_747_92157a11_46 | | 747.9216 | 11.46 | | RP | POS | | | 124923 | 54586 | 136269 | 73453.3 | 2.06E-01 | 0.18 | 0.8 |
| RP_POS_750_92719a11_541 | | 750.9272 | 11.54 | | RP | POS | | | 146302 | 73346.8 | 162483 | 107485 | 2.06E-01 | 0.18 | 0.91 |
| HILIC_NEG_356_99329a7_352 | | 356.9933 | 7.35 | | HILIC | NEG | | | 478388 | 25796.6 | 483235 | 29686.8 | 2.06E-01 | 0.17 | 0.87 |
| RP_NEG_292_9838a7_793 | | 292.9838 | 7.79 | | RP | NEG | | | 1227754 | 500456 | 1321959 | 577196 | 2.06E-01 | 0.17 | 0.93 |
| RP_POS_325_21158a10_964 | | 325.2116 | 10.96 | | RP | POS | | | 3138231 | 2082119 | 3500727 | 2104084 | 2.06E-01 | 0.17 | 1.1 |
| HILIC_POS_237_99966a0_927 | | 237.9997 | 0.93 | | HILIC | POS | | | 347919 | 148191 | 322987 | 140102 | 2.06E-01 | -0.17 | 1.01 |
| RP_POS_320_23495a11_139 | | 320.2350 | 11.14 | | RP | POS | | | 193274 | 109314 | 175952 | 91154.4 | 2.06E-01 | -0.17 | 1.28 |
| RP_POS_434_26917a11_594 | | 434.2692 | 11.59 | | RP | POS | | | 3995174 | 849795 | 3852799 | 797949 | 2.06E-01 | -0.17 | 1.31 |
| RP_POS_506_67865a10_206 | | 506.6787 | 10.21 | | RP | POS | | | 861758 | 474715 | 782202 | 446499 | 2.06E-01 | -0.17 | 0.91 |
| RP_POS_564_30579a10_015 | | 564.3058 | 10.02 | | RP | POS | | | 3211989 | 1911741 | 2934954 | 1272187 | 2.06E-01 | -0.17 | 0.78 |
| RP_POS_302_19568a2_451 | | 302.1957 | 2.45 | | RP | POS | | | 349613 | 176904 | 315476 | 212186 | 2.06E-01 | -0.18 | 0.97 |
| RP_NEG_826_56183a13_116 | PC 36:4 (16:0_20:4) | 826.5618 | 13.12 | | RP | NEG | 303.2345(100), 255.2329(75) | 1 | 1.3E+08 | 2E+07 | 1.4E+08 | 1.7E+07 | 2.07E-01 | 0.17 | 0.79 |
| RP_NEG_311_16925a15_459 | | 311.1693 | 15.46 | | RP | NEG | 311.1699(100) | | 767917 | 106509 | 785556 | 98484.7 | 2.08E-01 | 0.17 | 0.81 |
| RP_POS_291_64661a10_291 | | 291.6466 | 10.29 | | RP | POS | | | 797950 | 295761 | 855513 | 363187 | 2.08E-01 | 0.17 | 1.17 |
| RP_NEG_379_1553a10_219 | | 379.1553 | 10.22 | | RP | NEG | | | 221161 | 107664 | 203100 | 102271 | 2.08E-01 | -0.17 | 0.75 |
| RP_POS_191_10645a6_555 | | 191.1065 | 6.56 | | RP | POS | | | 533204 | 179730 | 502645 | 174709 | 2.08E-01 | -0.17 | 0.83 |
| RP_POS_421_29611a8_018 | | 421.2961 | 8.02 | | RP | POS | | | 317504 | 326714 | 272769 | 183380 | 2.08E-01 | -0.18 | 0.8 |
| RP_NEG_435_25186a11_115 | | 435.2519 | 11.12 | | RP | NEG | | | 177868 | 170117 | 213564 | 234473 | 2.09E-01 | 0.18 | 1.1 |
| RP_POS_535_37854a14_771 | | 535.3785 | 14.77 | | RP | POS | | | 452368 | 375495 | 548732 | 677621 | 2.09E-01 | 0.18 | 1.02 |
| RP_NEG_1155_68335a10_61 | | 1155.6834 | 10.61 | | RP | NEG | | | 1177326 | 345524 | 1238045 | 358440 | 2.09E-01 | 0.17 | 0.94 |
| RP_POS_277_1098a8_842 | | 277.1098 | 8.84 | | RP | POS | | | 372130 | 160282 | 400654 | 170139 | 2.09E-01 | 0.17 | 0.77 |
| RP_POS_380_89529a15_401 | | 380.8953 | 15.40 | | RP | POS | | | 168706 | 157762 | 198694 | 186679 | 2.09E-01 | 0.17 | 1.01 |
| RP_POS_383_28665a9_407 | | 383.2867 | 9.41 | | RP | POS | | | 311795 | 182089 | 347509 | 226413 | 2.09E-01 | 0.17 | 0.8 |
| HILIC_NEG_132_90034a6_134 | | 132.9003 | 6.13 | | HILIC | NEG | | | 831496 | 228667 | 792283 | 227364 | 2.09E-01 | -0.17 | 0.83 |
| RP_NEG_353_16467a11_142 | | 353.1647 | 11.14 | | RP | NEG | | | 1116713 | 103360 | 1099698 | 95117.5 | 2.09E-01 | -0.17 | 1.1 |
| RP_NEG_405_22421a11_361 | | 405.2242 | 11.36 | | RP | NEG | | | 245565 | 75929.8 | 231841 | 82922 | 2.09E-01 | -0.17 | 1.4 |
| RP_NEG_596_3941a11_333 | | 596.3941 | 11.33 | | RP | NEG | | | 485477 | 142098 | 459826 | 154739 | 2.09E-01 | -0.17 | 0.87 |
| RP_NEG_790_54047a13_679 | | 790.5405 | 13.68 | | RP | NEG | | | 1831706 | 959131 | 1685529 | 743388 | 2.09E-01 | -0.17 | 0.74 |
| RP_POS_355_2515a15_4 | | 355.2515 | 15.40 | | RP | POS | | | 229766 | 125901 | 209431 | 111301 | 2.09E-01 | -0.17 | 1.07 |
| RP_POS_395_79718a10_149 | | 395.7972 | 10.15 | | RP | POS | | | 144937 | 105630 | 128858 | 81517.1 | 2.09E-01 | -0.17 | 1.04 |
| RP_POS_567_8877a10_172 | | 567.8877 | 10.17 | | RP | POS | | | 3469379 | 1780293 | 3163289 | 1776208 | 2.09E-01 | -0.17 | 0.94 |
| RP_POS_585_53296a10_835 | | 585.5330 | 10.84 | | RP | POS | | | 5725445 | 4497416 | 1E+07 | 3.4E+07 | 2.10E-01 | 0.23 | 0.94 |
| RP_NEG_285_06839a10_245 | | 285.0684 | 10.25 | | RP | NEG | | | 417860 | 615875 | 516243 | 533221 | 2.10E-01 | 0.17 | 0.72 |
| RP_NEG_427_21625a7_451 | | 427.2163 | 7.45 | | RP | NEG | | | 476840 | 415111 | 542792 | 355923 | 2.10E-01 | 0.17 | 1.07 |
| RP_POS_366_26367a7_832 | | 366.2637 | 7.83 | | RP | POS | | | 372848 | 334034 | 439331 | 427220 | 2.10E-01 | 0.17 | 0.78 |
| HILIC_NEG_100_93352a7_742 | | 100.9335 | 7.74 | | HILIC | NEG | | | 2.2E+07 | 4019394 | 2.1E+07 | 4055799 | 2.10E-01 | -0.17 | 0.72 |
| RP_NEG_313_16376a10_328 | | 313.1638 | 10.33 | | RP | NEG | | | 241268 | 186511 | 213452 | 137995 | 2.10E-01 | -0.17 | 0.93 |
| RP_NEG_457_1452a11_592 | | 457.1452 | 11.59 | | RP | NEG | | | 755833 | 56525.3 | 745906 | 58795.9 | 2.10E-01 | -0.17 | 1.01 |
| RP_NEG_586_31561a9_889 | | 586.3156 | 9.89 | | RP | NEG | | | 1054924 | 541275 | 975869 | 377561 | 2.10E-01 | -0.17 | 0.79 |
| RP_NEG_665_17358a9_738 | | 665.1736 | 9.74 | | RP | NEG | | | 537136 | 28143 | 532401 | 27116 | 2.10E-01 | -0.17 | 0.93 |
| RP_POS_381_29788a11_601 | | 381.2979 | 11.60 | | RP | POS | | | 492975 | 125528 | 470331 | 136967 | 2.10E-01 | -0.17 | 0.81 |
| HILIC_POS_118_08645a1_379 | | 118.0865 | 1.38 | | HILIC | POS | 118.0866(100) | | 4381347 | 1634495 | 4660436 | 1625354 | 2.11E-01 | 0.17 | 0.84 |
| HILIC_POS_130_15935a1_369 | | 130.1594 | 1.37 | | HILIC | POS | | | 1921694 | 947742 | 2091440 | 1025388 | 2.11E-01 | 0.17 | 0.95 |
| RP_POS_127_11197a5_561 | | 127.1120 | 5.56 | | RP | POS | | | 159335 | 78301 | 172870 | 79622.1 | 2.11E-01 | 0.17 | 1.13 |
| RP_POS_600_13623a11_444 | | 600.1362 | 11.44 | | RP | POS | | | 115104 | 84653.3 | 129014 | 77999.7 | 2.11E-01 | 0.17 | 0.72 |
| RP_POS_701_29279a11_476 | | 701.2928 | 11.48 | | RP | POS | | | 164667 | 73989.3 | 179177 | 92575.7 | 2.11E-01 | 0.17 | 0.82 |
| RP_NEG_269_24918a11_361 | | 269.2492 | 11.36 | | RP | NEG | | | 1762062 | 638894 | 1647821 | 688350 | 2.11E-01 | -0.17 | 1.39 |
| RP_NEG_282_99454a3_584 | | 282.9945 | 3.58 | | RP | NEG | | | 405486 | 131301 | 382527 | 136085 | 2.11E-01 | -0.17 | 0.87 |
| RP_NEG_586_31549a10_016 | | 586.3155 | 10.02 | | RP | NEG | | | 8040193 | 4141950 | 7435564 | 2902638 | 2.11E-01 | -0.17 | 0.78 |
| RP_POS_584_91479a11_392 | | 584.9148 | 11.39 | | RP | POS | | | 119750 | 76866.7 | 108370 | 55965.2 | 2.11E-01 | -0.17 | 1.03 |
| RP_POS_617_43475a9_759 | | 617.4348 | 9.76 | | RP | POS | | | 3286069 | 1605230 | 3010903 | 1605061 | 2.11E-01 | -0.17 | 0.98 |
| RP_POS_335_24283a10_963 | | 335.2428 | 10.96 | | RP | POS | | | 1184429 | 608707 | 1286449 | 587179 | 2.12E-01 | 0.17 | 1.1 |
| RP_POS_359_1666a10_925 | | 359.1666 | 10.93 | | RP | POS | | | 874432 | 413160 | 941433 | 373297 | 2.12E-01 | 0.17 | 0.94 |
| RP_POS_641_11823a10_596 | | 641.1182 | 10.60 | | RP | POS | | | 189946 | 132761 | 212814 | 134553 | 2.12E-01 | 0.17 | 0.84 |
| RP_POS_780_55438a12_742 | PC 36:5 (16:1_20:4) | 780.5544 | 12.74 | | RP | POS | 184.0743(100), 780.5491(2) | 1 | 3.2E+08 | 1.6E+08 | 2.9E+08 | 1.2E+08 | 2.12E-01 | -0.17 | 0.74 |
| HILIC_POS_150_11234a1_377 | | 150.1123 | 1.38 | | HILIC | POS | 150.1126(100), 132.1046(45) | | 2636818 | 1730867 | 2395702 | 1069358 | 2.12E-01 | -0.17 | 0.99 |
| HILIC_POS_178_10744a3_112 | | 178.1074 | 3.11 | | HILIC | POS | | | 1059723 | 525975 | 985269 | 340327 | 2.12E-01 | -0.17 | 1.04 |
| RP_POS_473_31784a11_464 | | 473.3178 | 11.46 | | RP | POS | | | 1298171 | 334997 | 1246757 | 268924 | 2.12E-01 | -0.17 | 0.85 |
| RP_POS_752_55963a13_695 | | 752.5596 | 13.70 | | RP | POS | | | 916940 | 728487 | 794981 | 699889 | 2.12E-01 | -0.17 | 1.07 |
| RP_POS_850_55383a13_822 | | 850.5538 | 13.82 | | RP | POS | | | 1.1E+07 | 5078906 | 9851929 | 4685303 | 2.12E-01 | -0.17 | 0.97 |
| RP_POS_384_20416a10_835 | | 384.2042 | 10.84 | | RP | POS | | | 947049 | 358473 | 1104167 | 1203617 | 2.13E-01 | 0.2 | 0.92 |
| RP_POS_435_24991a10_752 | | 435.2499 | 10.75 | | RP | POS | | | 148801 | 137844 | 193015 | 328035 | 2.13E-01 | 0.19 | 0.95 |
| HILIC_NEG_166_83524a3_199 | | 166.8352 | 3.20 | | HILIC | NEG | | | 7523303 | 307255 | 7571646 | 261605 | 2.13E-01 | 0.17 | 0.94 |
| RP_NEG_241_88251a15_642 | | 241.8825 | 15.64 | | RP | NEG | | | 2697578 | 1170940 | 2896831 | 1164238 | 2.13E-01 | 0.17 | 0.82 |
| RP_POS_133_03473a15_326 | | 133.0347 | 15.33 | | RP | POS | | | 9.2E+07 | 1.1E+07 | 9.4E+07 | 1.1E+07 | 2.13E-01 | 0.17 | 0.8 |
| HILIC_POS_172_09418a7_089 | | 172.0942 | 7.09 | | HILIC | POS | | | 697840 | 280951 | 652054 | 257493 | 2.13E-01 | -0.17 | 1.02 |
| RP_NEG_364_75693a15_432 | | 364.7569 | 15.43 | | RP | NEG | | | 882147 | 320580 | 825521 | 340954 | 2.13E-01 | -0.17 | 0.81 |
| RP_POS_245_13138a14_233 | | 245.1314 | 14.23 | | RP | POS | | | 979580 | 481623 | 901144 | 439930 | 2.13E-01 | -0.17 | 0.84 |
| RP_POS_267_15686a9_21 | | 267.1569 | 9.21 | | RP | POS | | | 3.7E+08 | 1.8E+08 | 3.4E+08 | 1.7E+08 | 2.13E-01 | -0.17 | 1.08 |
| RP_POS_283_13065a9_209 | | 283.1307 | 9.21 | | RP | POS | | | 1.1E+07 | 4928291 | 9701650 | 4846889 | 2.13E-01 | -0.17 | 1.08 |
| RP_POS_404_31085a10_661 | | 404.3109 | 10.66 | | RP | POS | | | 204142 | 112913 | 187123 | 87218.2 | 2.13E-01 | -0.17 | 0.81 |
| RP_POS_433_13574a9_207 | | 433.1357 | 9.21 | | RP | POS | | | 275921 | 286852 | 230702 | 244984 | 2.13E-01 | -0.17 | 0.78 |
| RP_POS_445_39978a11_025 | | 445.3998 | 11.03 | | RP | POS | | | 311614 | 532576 | 236546 | 341686 | 2.13E-01 | -0.17 | 0.79 |
| RP_POS_493_34875a11_903 | | 493.3488 | 11.90 | | RP | POS | | | 5814423 | 1.4E+07 | 3738368 | 1E+07 | 2.13E-01 | -0.17 | 0.79 |
| RP_POS_563_46637a13_648 | | 563.4664 | 13.65 | | RP | POS | | | 928296 | 494311 | 845712 | 476211 | 2.13E-01 | -0.17 | 1.15 |
| RP_NEG_345_15552a6_298 | | 345.1555 | 6.30 | | RP | NEG | | | 390616 | 1061583 | 252291 | 507695 | 2.13E-01 | -0.18 | 0.77 |
| RP_POS_1023_63776a10_6 | | 1023.6378 | 10.60 | | RP | POS | | | 787191 | 393683 | 861477 | 469343 | 2.14E-01 | 0.17 | 0.87 |
| HILIC_POS_106_08679a0_879 | | 106.0868 | 0.88 | | HILIC | POS | 88.0761(100), 70.0659(75) | | 9995023 | 4334429 | 9309831 | 3747469 | 2.14E-01 | -0.17 | 1.25 |
| HILIC_NEG_226_99538a0_623 | | 226.9954 | 0.62 | | HILIC | NEG | | | 1366346 | 380715 | 1307875 | 309379 | 2.14E-01 | -0.17 | 0.75 |
| HILIC_POS_153_06587a0_729 | | 153.0659 | 0.73 | | HILIC | POS | | | 1.9E+07 | 1.2E+07 | 1.7E+07 | 1E+07 | 2.14E-01 | -0.17 | 0.73 |
| RP_POS_1051_80054a14_356 | | 1051.8005 | 14.36 | | RP | POS | | | 2E+08 | 5.3E+07 | 1.9E+08 | 4.2E+07 | 2.14E-01 | -0.17 | 1.04 |
| RP_POS_236_07153a11_254 | | 236.0715 | 11.25 | | RP | POS | | | 1E+08 | 5.2E+07 | 9.5E+07 | 2.7E+07 | 2.14E-01 | -0.17 | 0.78 |
| RP_POS_309_17841a0_584 | | 309.1784 | 0.58 | | RP | POS | | | 1468259 | 2585908 | 1106649 | 1638946 | 2.14E-01 | -0.17 | 0.73 |
| RP_POS_338_23242a6_854 | | 338.2324 | 6.85 | | RP | POS | | | 444962 | 256872 | 403078 | 236618 | 2.14E-01 | -0.17 | 0.76 |
| RP_POS_368_27942a8_412 | | 368.2794 | 8.41 | | RP | POS | | | 264254 | 354909 | 213206 | 243203 | 2.14E-01 | -0.17 | 1.14 |
| RP_POS_494_25656a9_459 | | 494.2566 | 9.46 | | RP | POS | | | 408103 | 398863 | 346941 | 321411 | 2.14E-01 | -0.17 | 0.91 |
| RP_POS_539_26971a4_476 | | 539.2697 | 4.48 | | RP | POS | | | 1325333 | 845154 | 1186380 | 790054 | 2.14E-01 | -0.17 | 1.27 |
| RP_POS_860_61572a13_905 | | 860.6157 | 13.91 | | RP | POS | | | 1527122 | 1563806 | 1308343 | 987811 | 2.14E-01 | -0.17 | 0.97 |
| HILIC_NEG_130_95764a2_995 | | 130.9576 | 3.00 | | HILIC | NEG | | | 4293853 | 483622 | 4375067 | 473137 | 2.15E-01 | 0.17 | 0.8 |
| RP_NEG_443_26572a9_742 | | 443.2657 | 9.74 | | RP | NEG | | | 415124 | 246138 | 457491 | 252373 | 2.15E-01 | 0.17 | 1.13 |
| RP_POS_181_04951a10_39 | | 181.0495 | 10.39 | | RP | POS | | | 817479 | 309411 | 873550 | 346012 | 2.15E-01 | 0.17 | 1.11 |
| RP_POS_335_16602a10_964 | | 335.1660 | 10.96 | | RP | POS | | | 3665033 | 1718330 | 3955976 | 1709366 | 2.15E-01 | 0.17 | 1.14 |
| RP_POS_427_28168a10_788 | | 427.2817 | 10.79 | | RP | POS | | | 1263868 | 1111617 | 1476645 | 1362098 | 2.15E-01 | 0.17 | 1.41 |
| RP_POS_612_10382a10_626 | | 612.1038 | 10.63 | | RP | POS | | | 247018 | 172793 | 279863 | 208954 | 2.15E-01 | 0.17 | 0.76 |
| HILIC_POS_229_1185a6_135 | Proline-hydroxyproline | 229.1185 | 6.14 | | HILIC | POS | 70.066(100), 132.0655(30), 229.1175(35) | 2 | 1095007 | 541437 | 1184816 | 517870 | 2.15E-01 | 0.17 | 1 |
| RP_POS_283_26306a10_538 | | 283.2631 | 10.54 | | RP | POS | | | 3375563 | 458371 | 3295804 | 478830 | 2.15E-01 | -0.17 | 1.06 |
| RP_POS_365_20825a8_842 | | 365.2083 | 8.84 | | RP | POS | | | 410941 | 672379 | 322485 | 342415 | 2.15E-01 | -0.17 | 0.85 |
| RP_POS_377_21994a11_144 | | 377.2199 | 11.14 | | RP | POS | | | 6192531 | 984240 | 6031177 | 920513 | 2.15E-01 | -0.17 | 1.07 |
| RP_POS_542_32404a9_889 | | 542.3240 | 9.89 | | RP | POS | | | 3562380 | 1858014 | 3295883 | 1270628 | 2.15E-01 | -0.17 | 0.76 |
| RP_POS_717_75958a6_032 | | 717.7596 | 6.03 | | RP | POS | | | 861809 | 592082 | 768485 | 510952 | 2.15E-01 | -0.17 | 1.25 |
| RP_POS_1179_87a13_801 | | 1179.8700 | 13.80 | | RP | POS | | | 702799 | 603987 | 837232 | 921551 | 2.16E-01 | 0.18 | 0.82 |
| HILIC_NEG_271_99924a0_944 | | 271.9992 | 0.94 | | HILIC | NEG | | | 1497362 | 900693 | 1656066 | 964194 | 2.16E-01 | 0.17 | 0.89 |
| HILIC_POS_79_99043a2_981 | | 79.9904 | 2.98 | | HILIC | POS | | | 5781711 | 468263 | 5860785 | 464470 | 2.16E-01 | 0.17 | 0.78 |
| RP_POS_580_05627a10_144 | | 580.0563 | 10.14 | | RP | POS | | | 397295 | 207450 | 439250 | 276486 | 2.16E-01 | 0.17 | 1.02 |
| HILIC_POS_568_34137a1_142 | LysoPC 22:6 (22:6/0:0) | 568.3414 | 1.14 | | HILIC | POS | 184.0744(100), 104.1071(95) | 2 | 4895974 | 1824829 | 4600274 | 1674651 | 2.16E-01 | -0.17 | 0.83 |
| HILIC_POS_159_02792a1_378 | | 159.0279 | 1.38 | | HILIC | POS | 159.0275(100) | | 2.1E+07 | 7769818 | 2.3E+07 | 7771730 | 2.17E-01 | 0.17 | 0.83 |
| RP_NEG_263_16537a8_501 | | 263.1654 | 8.50 | | RP | NEG | | | 1073525 | 61107 | 1085051 | 73661.9 | 2.17E-01 | 0.17 | 0.77 |
| RP_POS_339_16217a10_963 | | 339.1622 | 10.96 | | RP | POS | | | 2716201 | 1572030 | 2978533 | 1533325 | 2.17E-01 | 0.17 | 1.14 |
| RP_POS_441_23975a10_846 | | 441.2398 | 10.85 | | RP | POS | | | 434727 | 1575647 | 762948 | 2201401 | 2.17E-01 | 0.17 | 1.19 |
| HILIC_POS_357_25021a6_684 | | 357.2502 | 6.68 | | HILIC | POS | | | 130222 | 66501.9 | 120124 | 53362.3 | 2.17E-01 | -0.17 | 0.85 |
| RP_POS_266_12473a0_677 | | 266.1247 | 0.68 | | RP | POS | | | 263369 | 139838 | 240891 | 126835 | 2.17E-01 | -0.17 | 0.81 |
| RP_POS_266_84534a15_426 | | 266.8453 | 15.43 | | RP | POS | | | 216029 | 145284 | 194323 | 112039 | 2.17E-01 | -0.17 | 0.77 |
| HILIC_NEG_101_0244a5_118 | | 101.0244 | 5.12 | | HILIC | NEG | | | 2740129 | 1385617 | 2963342 | 1267035 | 2.18E-01 | 0.17 | 0.97 |
| HILIC_POS_374_80835a2_98 | | 374.8084 | 2.98 | | HILIC | POS | | | 402294 | 84234.9 | 416905 | 88607.4 | 2.18E-01 | 0.17 | 0.77 |
| HILIC_POS_458_09683a5_813 | | 458.0968 | 5.81 | | HILIC | POS | | | 57473.7 | 19465.6 | 61649.6 | 28456.9 | 2.18E-01 | 0.17 | 0.81 |
| RP_NEG_418_1593a11_596 | | 418.1593 | 11.60 | | RP | NEG | | | 288798 | 119838 | 308506 | 114349 | 2.18E-01 | 0.17 | 0.92 |
| RP_POS_160_95982a15_417 | | 160.9598 | 15.42 | | RP | POS | | | 3481764 | 1339317 | 3703030 | 1289487 | 2.18E-01 | 0.17 | 0.84 |
| RP_POS_490_026a10_653 | | 490.0260 | 10.65 | | RP | POS | | | 183561 | 100865 | 202955 | 125752 | 2.18E-01 | 0.17 | 0.78 |
| RP_POS_617_7663a10_377 | | 617.7663 | 10.38 | | RP | POS | | | 189797 | 109514 | 210133 | 129186 | 2.18E-01 | 0.17 | 0.83 |
| HILIC_POS_177_06602a0_733 | 2-(hydroxymethyl)-4(3H)-quinazolinone | 177.0660 | 0.73 | | HILIC | POS | 154.0493(100), 177.1056(45) | 2 | 357603 | 213963 | 323855 | 187342 | 2.18E-01 | -0.17 | 0.71 |
| RP_NEG_299_22266a9_854 | | 299.2227 | 9.85 | | RP | NEG | | | 677244 | 53379.8 | 667906 | 56956.1 | 2.18E-01 | -0.17 | 0.99 |
| RP_NEG_452_92398a10_862 | | 452.9240 | 10.86 | | RP | NEG | | | 974049 | 261383 | 931131 | 248425 | 2.18E-01 | -0.17 | 0.72 |
| RP_POS_287_2215a8_864 | | 287.2215 | 8.86 | | RP | POS | | | 852770 | 114243 | 834260 | 105766 | 2.18E-01 | -0.17 | 0.85 |
| RP_POS_466_22437a8_844 | | 466.2244 | 8.84 | | RP | POS | | | 2112490 | 458447 | 2037241 | 435792 | 2.18E-01 | -0.17 | 0.9 |
| RP_NEG_255_23299a10_487 | | 255.2330 | 10.49 | | RP | NEG | | | 846958 | 108718 | 866962 | 126724 | 2.19E-01 | 0.17 | 0.95 |
| RP_NEG_379_15857a10_962 | | 379.1586 | 10.96 | | RP | NEG | | | 2089568 | 1656924 | 2366996 | 1639797 | 2.19E-01 | 0.17 | 1.1 |
| RP_POS_198_18518a13_297 | | 198.1852 | 13.30 | | RP | POS | | | 2.2E+07 | 8481345 | 2.3E+07 | 8658311 | 2.19E-01 | 0.17 | 0.74 |
| RP_POS_542_88177a9_786 | | 542.8818 | 9.79 | | RP | POS | | | 1860297 | 889384 | 2021863 | 1014656 | 2.19E-01 | 0.17 | 0.91 |
| RP_POS_642_11285a10_232 | | 642.1129 | 10.23 | | RP | POS | | | 256064 | 163797 | 287430 | 203221 | 2.19E-01 | 0.17 | 0.78 |
| RP_NEG_907_72925a11_59 | | 907.7293 | 11.59 | | RP | NEG | | | 355910 | 66101.2 | 344368 | 70440.5 | 2.19E-01 | -0.17 | 1.27 |

**Supplemantary table 1**: The non-targeted metabolomics results ordered by the VIP value (p value < 0.0003 are shaded dark green, p value < 0.05 are shaded light green, d values below 0 are blue and above 0 are red)

| | | | | | | | | | Cases | | Controls | | Welch test | Cohen's | |
|---|---|---|---|---|---|---|---|---|---|---|---|---|---|---|---|
| Feature_ID | Metabolite_name | Average_Mz | Average_RT_min_ | HMDB ID | RP_HILIC | POS_NEG | Cureted_MSMS | ID_level | Mean | SD | Mean2 | SD3 | p | d | VIP |
| RP_POS_299_25793a9_843 | | 299.2579 | 9.84 | | RP | POS | | | 2139866 | 352761 | 2079418 | 363933 | 2.19E-01 | -0.17 | 0.94 |
| RP_POS_512_35657a9_795 | | 512.3566 | 9.80 | | RP | POS | | | 1158198 | 642265 | 1061506 | 510767 | 2.19E-01 | -0.17 | 0.92 |
| RP_POS_516_42633a12_426 | | 516.4263 | 12.43 | | RP | POS | | | 2.3E+07 | 6E+07 | 1.4E+07 | 4.3E+07 | 2.19E-01 | -0.17 | 0.78 |
| RP_POS_549_41687a11_743 | | 549.4169 | 11.74 | | RP | POS | | | 647487 | 533510 | 574217 | 328520 | 2.19E-01 | -0.17 | 0.69 |
| RP_POS_698_69061a11_39 | | 698.6906 | 11.39 | | RP | POS | | | 195714 | 92756.9 | 212583 | 106543 | 2.20E-01 | 0.17 | 0.89 |
| RP_POS_701_40674a15_414 | | 701.4067 | 15.41 | | RP | POS | | | 517610 | 232092 | 558185 | 249486 | 2.20E-01 | 0.17 | 0.9 |
| RP_POS_432_31064a8_248 | Glycohyodeoxcholic acid | 432.3106 | 8.25 | | RP | POS | 414.3018(100),339.2697(45) | 1 | 1707698 | 2307574 | 1403215 | 1245289 | 2.20E-01 | -0.17 | 0.93 |
| RP_NEG_514_2843a8_447 | Taurocholic acid | 514.2843 | 8.45 | | RP | NEG | 514.2859(100) | 2 | 4530240 | 1.7E+07 | 2588855 | 3666046 | 2.20E-01 | -0.19 | 0.73 |
| HILIC_NEG_158_95038a0_933 | | 158.9504 | 0.93 | | HILIC | NEG | | | 1646775 | 730494 | 1533268 | 625981 | 2.20E-01 | -0.17 | 0.91 |
| RP_POS_250_17752a11_025 | | 250.1775 | 11.03 | | RP | POS | | | 496267 | 874055 | 376437 | 539591 | 2.20E-01 | -0.17 | 0.79 |
| RP_POS_429_23972a11_408 | | 429.2397 | 11.41 | | RP | POS | | | 5168303 | 858774 | 5042212 | 646729 | 2.20E-01 | -0.17 | 0.93 |
| RP_POS_439_82404a10_088 | | 439.8240 | 10.09 | | RP | POS | | | 1375117 | 876588 | 1243794 | 690704 | 2.20E-01 | -0.17 | 0.81 |
| RP_POS_655_57141a11_594 | | 655.5714 | 11.59 | | RP | POS | | | 1014361 | 298160 | 967855 | 257850 | 2.20E-01 | -0.17 | 1.17 |
| RP_NEG_362_15756a8_804 | | 362.1576 | 8.80 | | RP | NEG | | | 312530 | 446044 | 416913 | 738692 | 2.21E-01 | 0.18 | 0.8 |
| HILIC_NEG_131_0827a6_955 | | 131.0827 | 6.96 | | HILIC | NEG | | | 248042 | 47759 | 256440 | 52025.3 | 2.21E-01 | 0.17 | 0.78 |
| RP_NEG_836_58209a13_575 | | 836.5821 | 13.58 | | RP | NEG | | | 3633545 | 1322265 | 3843530 | 1188394 | 2.21E-01 | 0.17 | 1 |
| RP_POS_355_28409a11_02 | | 355.2841 | 11.02 | | RP | POS | | | 1675648 | 664615 | 1820604 | 1003134 | 2.21E-01 | 0.17 | 0.85 |
| RP_POS_824_55591a14_1 | | 824.5559 | 14.10 | | RP | POS | | | 1.1E+07 | 5172641 | 1.2E+07 | 4537705 | 2.21E-01 | 0.17 | 1.12 |
| HILIC_POS_331_24698a3_126 | | 331.2470 | 3.13 | | HILIC | POS | | | 864072 | 104049 | 845804 | 113047 | 2.21E-01 | -0.17 | 0.84 |
| RP_NEG_432_18188a11_143 | | 432.1819 | 11.14 | | RP | NEG | | | 740507 | 61274.7 | 730797 | 55035.7 | 2.21E-01 | -0.17 | 0.87 |
| RP_POS_270_20657a6_596 | | 270.2066 | 6.60 | | RP | POS | | | 340420 | 71402.6 | 328967 | 65605.5 | 2.21E-01 | -0.17 | 1.21 |
| RP_POS_409_27069a11_451 | | 409.2707 | 11.45 | | RP | POS | | | 451868 | 150893 | 428979 | 122976 | 2.21E-01 | -0.17 | 0.87 |
| RP_POS_511_32394a9_21 | | 511.3239 | 9.21 | | RP | POS | | | 7.2E+07 | 5.1E+07 | 6.3E+07 | 4.7E+07 | 2.21E-01 | -0.17 | 1.06 |
| RP_POS_531_85913a9_795 | | 531.8591 | 9.80 | | RP | POS | | | 2308371 | 1199319 | 2102251 | 1254687 | 2.21E-01 | -0.17 | 1.03 |
| RP_POS_221_15321a8_033 | | 221.1532 | 8.03 | | RP | POS | | | 1706492 | 177115 | 1737011 | 187054 | 2.22E-01 | 0.17 | 0.84 |
| RP_POS_348_28952a11_119 | | 348.2895 | 11.12 | | RP | POS | | | 1034159 | 657954 | 1141972 | 634094 | 2.22E-01 | 0.17 | 1.11 |
| RP_POS_393_23981a10_668 | | 393.2398 | 10.67 | | RP | POS | | | 609404 | 386656 | 691574 | 564239 | 2.22E-01 | 0.17 | 1.06 |
| RP_POS_626_27216a11_455 | | 626.2722 | 11.46 | | RP | POS | | | 143821 | 77688.9 | 159689 | 107307 | 2.22E-01 | 0.17 | 0.87 |
| RP_POS_632_3772a14_346 | | 632.3772 | 14.35 | | RP | POS | | | 918876 | 597020 | 1037197 | 788157 | 2.22E-01 | 0.17 | 0.96 |
| HILIC_POS_150_11266a4_124 | | 150.1127 | 4.12 | | HILIC | POS | 150.1127(100), 132.1046(40) | | 1987336 | 1218579 | 1794524 | 1091599 | 2.22E-01 | -0.17 | 1.18 |
| HILIC_NEG_197_80769a3_089 | | 197.8077 | 3.09 | | HILIC | NEG | | | 2.3E+07 | 2015171 | 2.2E+07 | 2195564 | 2.22E-01 | -0.17 | 0.97 |
| RP_NEG_350_2106a9_358 | | 350.2106 | 9.36 | | RP | NEG | | | 435637 | 140272 | 414818 | 109059 | 2.22E-01 | -0.17 | 0.75 |
| RP_POS_143_07022a8_101 | | 143.0702 | 8.10 | | RP | POS | | | 579527 | 145697 | 555647 | 140074 | 2.22E-01 | -0.17 | 0.8 |
| RP_POS_159_1169a6_709 | | 159.1169 | 6.71 | | RP | POS | | | 1494905 | 1615751 | 1309294 | 241505 | 2.22E-01 | -0.2 | 0.69 |
| HILIC_POS_460_76782a2_978 | | 460.7678 | 2.98 | | HILIC | POS | | | 183485 | 40600.2 | 189930 | 36807.9 | 2.23E-01 | 0.17 | 0.69 |
| RP_POS_173_09613a10_513 | | 173.0961 | 10.51 | | RP | POS | | | 3776351 | 1817945 | 4080547 | 1827394 | 2.23E-01 | 0.17 | 1.17 |
| HILIC_NEG_253_79387a7_048 | | 253.7939 | 7.05 | | HILIC | NEG | | | 161053 | 76171.5 | 149179 | 66449.7 | 2.23E-01 | -0.17 | 0.78 |
| RP_NEG_1043_15234a9_708 | | 1043.1523 | 9.71 | | RP | NEG | | | 4645754 | 342866 | 4589688 | 329613 | 2.23E-01 | -0.17 | 1.19 |
| RP_NEG_269_21243a10_269 | | 269.2124 | 10.27 | | RP | NEG | | | 1164234 | 135375 | 1142108 | 129913 | 2.23E-01 | -0.17 | 1.06 |
| RP_POS_271_22662a10_098 | | 271.2266 | 10.10 | | RP | POS | | | 1276471 | 201279 | 1244734 | 179850 | 2.23E-01 | -0.17 | 0.99 |
| RP_POS_491_37366a11_342 | | 491.3737 | 11.34 | | RP | POS | | | 281142 | 162751 | 255422 | 146236 | 2.23E-01 | -0.17 | 0.76 |
| RP_POS_585_40356a9_955 | | 585.4036 | 9.96 | | RP | POS | | | 1135514 | 540974 | 1051340 | 471279 | 2.23E-01 | -0.17 | 0.7 |
| RP_POS_660_46997a10_265 | | 660.4700 | 10.27 | | RP | POS | | | 5559010 | 2220131 | 5178850 | 2321567 | 2.23E-01 | -0.17 | 0.79 |
| RP_NEG_229_05391a6_458 | | 229.0539 | 6.46 | | RP | NEG | | | 687331 | 2843907 | 358911 | 517184 | 2.23E-01 | -0.2 | 0.64 |
| HILIC_NEG_215_0675a6_58 | | 215.0675 | 6.58 | | HILIC | NEG | | | 1719600 | 332585 | 1769980 | 274660 | 2.24E-01 | 0.17 | 1.05 |
| RP_POS_309_12915a0_548 | | 309.1292 | 0.55 | | RP | POS | | | 2000940 | 493544 | 2108806 | 752378 | 2.24E-01 | 0.17 | 0.9 |
| RP_POS_538_29449a10_967 | | 538.2945 | 10.97 | | RP | POS | | | 280362 | 161239 | 306988 | 158940 | 2.24E-01 | 0.17 | 1.11 |
| RP_POS_657_96118a11_458 | | 657.9612 | 11.46 | | RP | POS | | | 192704 | 101588 | 212103 | 127604 | 2.24E-01 | 0.17 | 0.94 |
| RP_NEG_265_14807a9_691 | | 265.1481 | 9.69 | | RP | NEG | 265.1464(100), 96.9601(70) | | 2119239 | 1256531 | 1932741 | 988509 | 2.24E-01 | -0.17 | 0.83 |
| HILIC_POS_205_96178a0_931 | | 205.9618 | 0.93 | | HILIC | POS | | | 295732 | 57493.7 | 286259 | 56459.8 | 2.24E-01 | -0.17 | 0.92 |
| RP_POS_341_19916a11_595 | | 341.1992 | 11.60 | | RP | POS | | | 2733117 | 527830 | 2647333 | 503502 | 2.24E-01 | -0.17 | 0.91 |
| RP_POS_379_21548a11_144 | | 379.2155 | 11.14 | | RP | POS | | | 2279066 | 379132 | 2217522 | 360591 | 2.24E-01 | -0.17 | 0.98 |
| RP_POS_483_85141a10_039 | | 483.8514 | 10.04 | | RP | POS | | | 3013506 | 1216632 | 2787940 | 1461353 | 2.24E-01 | -0.17 | 0.88 |
| RP_POS_563_55145a10_835 | | 563.5515 | 10.84 | | RP | POS | | | 3397276 | 2928791 | 6346482 | 2.4E+07 | 2.25E-01 | 0.22 | 0.9 |
| HILIC_POS_445_32639a4_869 | | 445.3264 | 4.87 | | HILIC | POS | | | 111638 | 49593.3 | 121721 | 68706.2 | 2.25E-01 | 0.17 | 0.99 |
| RP_POS_537_39301a13_671 | | 537.3930 | 13.67 | | RP | POS | | | 510954 | 350435 | 587544 | 536395 | 2.25E-01 | 0.17 | 1.13 |
| RP_POS_840_52124a13_814 | | 840.5212 | 13.81 | | RP | POS | | | 1326876 | 702860 | 1466934 | 943609 | 2.25E-01 | 0.17 | 0.83 |
| HILIC_NEG_96_95981a7_735 | | 96.9598 | 7.74 | | HILIC | NEG | | | 9323594 | 558658 | 9231766 | 546771 | 2.25E-01 | -0.17 | 0.81 |
| RP_POS_455_34464a10_3 | | 455.3446 | 10.30 | | RP | POS | | | 818321 | 583119 | 726877 | 521029 | 2.25E-01 | -0.17 | 0.86 |
| RP_POS_551_43488a11_142 | | 551.4349 | 11.14 | | RP | POS | | | 1192172 | 433586 | 1119057 | 445961 | 2.25E-01 | -0.17 | 1.29 |
| RP_POS_246_17024a3_14 | | 246.1702 | 3.14 | | RP | POS | 85.0291(100), 246.1692(45) | | 3067931 | 1639493 | 3342093 | 1665225 | 2.26E-01 | 0.17 | 0.75 |
| RP_NEG_388_76495a15_406 | | 388.7650 | 15.41 | | RP | NEG | | | 168545 | 83764.9 | 182954 | 89497.1 | 2.26E-01 | 0.17 | 0.91 |
| RP_POS_260_0264a15_438 | | 260.0264 | 15.44 | | RP | POS | | | 366486 | 149057 | 391887 | 156356 | 2.26E-01 | 0.17 | 0.76 |
| RP_NEG_183_10262a7_309 | | 183.1026 | 7.31 | | RP | NEG | | | 325428 | 164404 | 301515 | 124602 | 2.26E-01 | -0.17 | 0.73 |
| RP_NEG_274_97675a7_696 | | 274.9768 | 7.70 | | RP | NEG | | | 1E+07 | 266058 | 9964808 | 247427 | 2.26E-01 | -0.17 | 0.74 |
| RP_NEG_428_18753a11_592 | | 428.1875 | 11.59 | | RP | NEG | | | 1312954 | 91894.5 | 1297230 | 97214.9 | 2.26E-01 | -0.17 | 0.94 |
| RP_POS_359_24698a11_141 | | 359.2470 | 11.14 | | RP | POS | | | 408078 | 218146 | 373477 | 200700 | 2.26E-01 | -0.17 | 0.98 |
| RP_POS_486_36365a8_409 | | 486.3637 | 8.41 | | RP | POS | | | 502822 | 309782 | 453178 | 289771 | 2.26E-01 | -0.17 | 0.9 |
| HILIC_POS_210_10973a5_9 | | 210.1097 | 5.90 | | HILIC | POS | | | 54088.1 | 53973.2 | 64824.4 | 72699.5 | 2.27E-01 | 0.17 | 0.82 |
| HILIC_POS_383_11627a5_088 | | 383.1163 | 5.09 | | HILIC | POS | | | 186268 | 93432.4 | 205024 | 127658 | 2.27E-01 | 0.17 | 0.73 |
| HILIC_POS_387_21423a4_004 | | 387.2142 | 4.00 | | HILIC | POS | | | 1251582 | 398586 | 1315571 | 376819 | 2.27E-01 | 0.17 | 0.75 |
| RP_POS_293_93411a15_373 | | 293.9341 | 15.37 | | RP | POS | | | 380949 | 243513 | 421292 | 244295 | 2.27E-01 | 0.17 | 1 |
| RP_POS_354_86276a15_425 | | 354.8628 | 15.43 | | RP | POS | | | 361188 | 218861 | 407761 | 324354 | 2.27E-01 | 0.17 | 0.79 |
| RP_POS_553_45868a13_293 | | 553.4587 | 13.29 | | RP | POS | | | 1354528 | 877632 | 1514697 | 1036841 | 2.27E-01 | 0.17 | 1.21 |
| RP_POS_571_90155a9_968 | | 571.9016 | 9.97 | | RP | POS | | | 2411477 | 1114148 | 2626266 | 1435422 | 2.27E-01 | 0.17 | 1.18 |
| RP_NEG_417_18015a11_291 | | 417.1802 | 11.29 | | RP | NEG | | | 365410 | 50541.5 | 357473 | 45678.1 | 2.27E-01 | -0.16 | 1.33 |
| RP_POS_418_74716a12_885 | | 418.7472 | 12.89 | | RP | POS | | | 678236 | 531104 | 595929 | 467835 | 2.27E-01 | -0.16 | 0.84 |
| RP_POS_273_10928a8_169 | | 273.1093 | 8.17 | | RP | POS | 273.1089(100) | | 1385574 | 144119 | 1361611 | 145219 | 2.27E-01 | -0.17 | 0.93 |
| RP_POS_266_20892a9_403 | | 266.2089 | 9.40 | | RP | POS | | | 379929 | 27984.8 | 375308 | 27845.9 | 2.27E-01 | -0.17 | 1.01 |
| RP_POS_659_52124a10_36 | | 659.5212 | 10.36 | | RP | POS | | | 721347 | 326906 | 673879 | 248299 | 2.27E-01 | -0.17 | 0.8 |
| RP_POS_752_04651a12_806 | | 752.0465 | 12.81 | | RP | POS | | | 2282307 | 829924 | 2148623 | 790471 | 2.27E-01 | -0.17 | 0.84 |
| HILIC_NEG_202_92911a5_122 | | 202.9291 | 5.12 | | HILIC | NEG | | | 2594287 | 1030102 | 2765558 | 1043799 | 2.28E-01 | 0.17 | 0.8 |
| RP_POS_331_26294a11_118 | | 331.2629 | 11.12 | | RP | POS | | | 601239 | 344025 | 657495 | 337851 | 2.28E-01 | 0.17 | 1.12 |
| RP_POS_472_65497a10_075 | | 472.6550 | 10.08 | | RP | POS | | | 557157 | 236188 | 605829 | 336031 | 2.28E-01 | 0.17 | 1.05 |
| HILIC_POS_292_84448a2_98 | | 292.8445 | 2.98 | | HILIC | POS | | | 1622593 | 333402 | 1677250 | 329133 | 2.28E-01 | 0.16 | 0.77 |
| RP_POS_622_28076a10_607 | | 622.2808 | 10.61 | | RP | POS | | | 467061 | 203735 | 437266 | 158240 | 2.28E-01 | -0.16 | 1.04 |
| RP_NEG_369_3017a10_958 | | 369.3017 | 10.96 | | RP | NEG | | | 465928 | 51123.7 | 457006 | 56356.4 | 2.28E-01 | -0.17 | 0.98 |
| RP_POS_243_19533a11_026 | | 243.1953 | 11.03 | | RP | POS | | | 319758 | 578756 | 241363 | 363608 | 2.28E-01 | -0.17 | 0.81 |
| RP_POS_492_8439a15_396 | | 492.8439 | 15.40 | | RP | POS | | | 198354 | 175396 | 174157 | 116532 | 2.28E-01 | -0.17 | 0.86 |
| RP_POS_379_0827a9_521 | | 379.0827 | 9.52 | | RP | POS | | | 177946 | 95000.5 | 200053 | 159899 | 2.29E-01 | 0.17 | 0.93 |
| RP_NEG_473_40039a13_28 | | 473.4004 | 13.28 | | RP | NEG | | | 675121 | 149741 | 698784 | 138251 | 2.29E-01 | 0.16 | 1.21 |
| HILIC_POS_508_37601a1_242 | LysoPC O-18:1 | 508.3760 | 1.24 | | HILIC | POS | 104.1079(100), 184.0738(45) | 2 | 1312651 | 429733 | 1381904 | 411679 | 2.29E-01 | 0.16 | 0.74 |
| RP_POS_297_14594a9_459 | | 297.1459 | 9.46 | | RP | POS | | | 3618791 | 3361048 | 3104342 | 2910836 | 2.29E-01 | -0.16 | 1.01 |
| RP_NEG_233_15517a9_091 | | 233.1552 | 9.09 | | RP | NEG | 233.1555(100) | | 4811076 | 237719 | 4770277 | 256024 | 2.29E-01 | -0.17 | 0.83 |
| HILIC_POS_406_32968a2_297 | | 406.3297 | 2.30 | | HILIC | POS | | | 35497.8 | 44075.9 | 43932.6 | 56653.9 | 2.30E-01 | 0.17 | 0.73 |
| HILIC_NEG_466_80234a2_992 | | 466.8023 | 2.99 | | HILIC | NEG | | | 283420 | 54975.6 | 292559 | 56206.1 | 2.30E-01 | 0.16 | 0.73 |
| RP_NEG_385_1489a6_26 | | 385.1489 | 6.26 | | RP | NEG | | | 415177 | 371296 | 474793 | 355640 | 2.30E-01 | 0.16 | 0.78 |
| RP_NEG_580_36188a10_82 | | 580.3619 | 10.82 | | RP | NEG | | | 618351 | 185541 | 648351 | 179568 | 2.30E-01 | 0.16 | 0.82 |
| RP_NEG_259_00131a7_319 | | 259.0013 | 7.32 | | RP | NEG | | | 1386171 | 206068 | 1351995 | 209439 | 2.30E-01 | -0.16 | 1.02 |
| RP_POS_295_15152a5_768 | | 295.1515 | 5.77 | | RP | POS | | | 166283 | 103806 | 150502 | 88940.7 | 2.30E-01 | -0.16 | 1.17 |
| RP_POS_475_3248a9_163 | | 475.3248 | 9.16 | | RP | POS | | | 264752 | 162022 | 240243 | 137080 | 2.30E-01 | -0.16 | 0.91 |
| HILIC_POS_125_09631a3_099 | | 125.0963 | 3.10 | | HILIC | POS | | | 45875.6 | 43316.6 | 40008.9 | 27682.4 | 2.30E-01 | -0.17 | 1.04 |
| RP_POS_574_89661a10_348 | | 574.8966 | 10.35 | | RP | POS | | | 1936670 | 1095604 | 2158102 | 1530397 | 2.31E-01 | 0.17 | 1.25 |
| RP_POS_603_40289a10_138 | | 603.4029 | 10.14 | | RP | POS | | | 212802 | 98174.9 | 233960 | 150105 | 2.31E-01 | 0.17 | 1.14 |
| RP_POS_866_52069a13_103 | | 866.5207 | 13.10 | | RP | POS | | | 652165 | 260884 | 706045 | 375486 | 2.31E-01 | 0.17 | 1.06 |
| HILIC_POS_161_12865a1_07 | | 161.1287 | 1.07 | | HILIC | POS | | | 608270 | 250662 | 651922 | 278911 | 2.31E-01 | 0.16 | 0.78 |
| HILIC_POS_290_84769a2_982 | | 290.8477 | 2.98 | | HILIC | POS | | | 5913010 | 1258469 | 6117886 | 1242038 | 2.31E-01 | 0.16 | 0.77 |
| RP_NEG_1149_24243a9_935 | | 1149.2424 | 9.94 | | RP | NEG | | | 2610145 | 168533 | 2637221 | 161922 | 2.31E-01 | 0.16 | 1.05 |
| RP_POS_368_30011a8_786 | | 368.3001 | 8.79 | | RP | POS | | | 184821 | 126528 | 206198 | 133380 | 2.31E-01 | 0.16 | 1 |
| RP_POS_733_50916a11_306 | | 733.5092 | 11.31 | | RP | POS | | | 206762 | 98252.1 | 223910 | 109707 | 2.31E-01 | 0.16 | 0.95 |
| RP_POS_257_5162a9_46 | | 257.5162 | 9.46 | | RP | POS | | | 314747 | 393364 | 258321 | 296071 | 2.31E-01 | -0.16 | 0.83 |
| RP_POS_542_32458a10_015 | | 542.3246 | 10.02 | | RP | POS | | | 2.9E+07 | 1.6E+07 | 2.7E+07 | 1.1E+07 | 2.31E-01 | -0.16 | 0.75 |
| RP_POS_645_27191a7_235 | | 645.2719 | 7.24 | | RP | POS | | | 2794811 | 611417 | 2699770 | 550193 | 2.31E-01 | -0.16 | 1.31 |
| HILIC_POS_663_45544a10_662 | | 663.4554 | 10.66 | | HILIC | POS | | | 349717 | 52757.4 | 339857 | 65606.1 | 2.31E-01 | -0.17 | 0.83 |
| RP_POS_247_13019a7_984 | | 247.1302 | 7.98 | | RP | POS | | | 742355 | 145906 | 771251 | 198481 | 2.32E-01 | 0.17 | 1.09 |
| HILIC_POS_349_23083a6_96 | | 349.2308 | 6.96 | | HILIC | POS | | | 159730 | 78393 | 174230 | 96234.9 | 2.32E-01 | 0.17 | 0.77 |
| RP_NEG_305_82166a15_559 | | 305.8217 | 15.56 | | RP | NEG | | | 334757 | 105966 | 352281 | 107926 | 2.32E-01 | 0.16 | 0.9 |
| RP_POS_326_90244a15_41 | | 326.9024 | 15.41 | | RP | POS | | | 5136286 | 1595178 | 5363755 | 1190361 | 2.32E-01 | 0.16 | 1.17 |
| RP_POS_393_19781a10_97 | | 393.1978 | 10.97 | | RP | POS | | | 165072 | 96499.7 | 181986 | 108916 | 2.32E-01 | 0.16 | 0.91 |
| HILIC_POS_150_1127a2_993 | | 150.1127 | 2.99 | | HILIC | POS | 150.1126(100), 132.1026(30) | | 1639041 | 1488862 | 1416841 | 1238367 | 2.32E-01 | -0.16 | 1.14 |
| HILIC_NEG_328_81024a5_43 | | 328.8102 | 5.43 | | HILIC | NEG | | | 378809 | 125254 | 358548 | 122560 | 2.32E-01 | -0.16 | 0.69 |
| RP_POS_284_09888a0_679 | | 284.0989 | 0.68 | | RP | POS | | | 620942 | 819233 | 504837 | 601749 | 2.32E-01 | -0.16 | 0.8 |
| HILIC_NEG_382_84143a2_996 | | 382.8414 | 3.00 | | HILIC | NEG | | | 500448 | 96842.5 | 516181 | 95780.5 | 2.33E-01 | 0.16 | 0.74 |
| HILIC_POS_536_37329a1_144 | | 536.3733 | 1.14 | | HILIC | POS | | | 162952 | 67889.4 | 174233 | 70005.9 | 2.33E-01 | 0.16 | 0.8 |
| RP_NEG_371_18997a7_673 | | 371.1900 | 7.67 | | RP | NEG | | | 516812 | 412457 | 584237 | 413355 | 2.33E-01 | 0.16 | 0.75 |
| HILIC_NEG_236_81674a3_074 | | 236.8167 | 3.07 | | HILIC | NEG | | | 155253 | 12264.7 | 153332 | 11286.2 | 2.33E-01 | -0.16 | 1.07 |
| RP_NEG_794_54681a13_798 | | 794.5468 | 13.80 | | RP | NEG | | | 2956174 | 1017942 | 2778178 | 1144956 | 2.33E-01 | -0.16 | 0.93 |
| RP_POS_357_24911a12_235 | | 357.2491 | 12.24 | | RP | POS | | | 956178 | 314017 | 906729 | 292832 | 2.33E-01 | -0.16 | 0.74 |
| RP_POS_776_51971a12_51 | | 776.5197 | 12.51 | | RP | POS | | | 6139710 | 3076079 | 5675660 | 2631157 | 2.33E-01 | -0.16 | 0.84 |
| HILIC_POS_178_05875a6_785 | | 178.0588 | 6.79 | | HILIC | POS | | | 2970895 | 941594 | 2846748 | 561867 | 2.33E-01 | -0.17 | 0.8 |
| RP_POS_203_14316a8_964 | | 203.1432 | 8.96 | | RP | POS | | | 8423016 | 499594 | 8503636 | 490622 | 2.34E-01 | 0.16 | 0.98 |
| RP_POS_274_08365a6_02 | | 274.0837 | 6.02 | | RP | POS | | | 632884 | 237236 | 671721 | 239519 | 2.34E-01 | 0.16 | 0.72 |
| RP_POS_757_62323a13_745 | | 757.6232 | 13.75 | | RP | POS | | | 6241161 | 3918803 | 6833640 | 3383650 | 2.34E-01 | 0.16 | 1.18 |
| HILIC_POS_118_08648a3_615 | Glycine betaine | 118.0865 | 3.62 | | HILIC | POS | 118.0866(100), 59.074(10), 58.0659(7) | 1 | 1.1E+09 | 2.8E+08 | 1E+09 | 3.5E+08 | 2.34E-01 | -0.17 | 0.83 |
| RP_POS_182_98544a0_727 | | 182.9854 | 0.73 | | RP | POS | | | 8566891 | 4899646 | 7750858 | 5103907 | 2.34E-01 | -0.16 | 0.81 |
| RP_POS_345_33545a11_145 | | 345.3355 | 11.15 | | RP | POS | | | 850590 | 185255 | 819156 | 198888 | 2.34E-01 | -0.16 | 1.01 |
| RP_POS_594_43188a10_325 | | 594.4319 | 10.33 | | RP | POS | | | 5990127 | 2827752 | 5552699 | 2552477 | 2.34E-01 | -0.16 | 0.89 |
| HILIC_POS_266_13904a1_386 | | 266.1390 | 1.39 | | HILIC | POS | | | 622349 | 318596 | 682879 | 412308 | 2.35E-01 | 0.17 | 0.77 |
| RP_POS_719_5033a10_064 | | 719.5033 | 10.06 | | RP | POS | | | 2063437 | 916944 | 2244439 | 1254623 | 2.35E-01 | 0.17 | 1.25 |
| RP_NEG_296_87558a15_407 | | 296.8756 | 15.41 | | RP | NEG | | | 456646 | 164351 | 486242 | 195836 | 2.35E-01 | 0.16 | 0.98 |
| RP_NEG_437_29166a8_92 | | 437.2917 | 8.92 | | RP | NEG | | | 396167 | 603114 | 487709 | 525801 | 2.35E-01 | 0.16 | 1.08 |
| RP_POS_219_15865a8_073 | | 219.1587 | 8.07 | | RP | POS | | | 4499193 | 369600 | 4557030 | 343709 | 2.35E-01 | 0.16 | 1.19 |
| RP_NEG_313_23923a10_126 | Octadecanedioic acid | 313.2392 | 10.13 | | RP | NEG | 313.2364(100), 251.238(98) | 2 | 938215 | 75721.7 | 924551 | 90321.7 | 2.35E-01 | -0.16 | 0.88 |
| RP_NEG_201_11345a6_757 | | 201.1135 | 6.76 | | RP | NEG | | | 1610850 | 155561 | 1586833 | 140476 | 2.35E-01 | -0.16 | 0.95 |
| RP_NEG_584_3576a10_573 | | 584.3576 | 10.57 | | RP | NEG | | | 519569 | 185505 | 489264 | 187119 | 2.35E-01 | -0.16 | 1.01 |
| RP_POS_361_15335a9_209 | | 361.1534 | 9.21 | | RP | POS | | | 998967 | 504922 | 921502 | 449721 | 2.35E-01 | -0.16 | 0.74 |
| RP_POS_280_26352a9_583 | | 280.2635 | 9.58 | | RP | POS | | | 442878 | 552743 | 551551 | 753152 | 2.36E-01 | 0.17 | 1.08 |
| RP_NEG_262_72867a15_476 | | 262.7287 | 15.48 | | RP | NEG | | | 248479 | 158851 | 277359 | 192730 | 2.36E-01 | 0.16 | 0.87 |
| RP_POS_163_11182a15_291 | | 163.1118 | 15.29 | | RP | POS | | | 8491732 | 4188329 | 9181397 | 4302256 | 2.36E-01 | 0.16 | 0.75 |
| RP_POS_200_08925a14_219 | | 200.0893 | 14.22 | | RP | POS | | | 274745 | 138778 | 297690 | 143480 | 2.36E-01 | 0.16 | 0.77 |
| RP_POS_255_15892a7_695 | | 255.1589 | 7.70 | | RP | POS | | | 4E+07 | 2229652 | 4.1E+07 | 2242203 | 2.36E-01 | 0.16 | 0.87 |
| RP_POS_316_05276a15_383 | | 316.0528 | 15.38 | | RP | POS | | | 1.2E+07 | 4783721 | 1.3E+07 | 4949558 | 2.36E-01 | 0.16 | 0.72 |
| RP_POS_394_29501a8_615 | | 394.2950 | 8.62 | | RP | POS | | | 198748 | 141271 | 223058 | 156485 | 2.36E-01 | 0.16 | 0.97 |
| RP_POS_409_19846a10_246 | | 409.1985 | 10.25 | | RP | POS | | | 527645 | 291917 | 571448 | 249979 | 2.36E-01 | 0.16 | 1.08 |
| RP_POS_494_17288a10_7 | | 494.1729 | 10.70 | | RP | POS | | | 449045 | 106229 | 467539 | 120075 | 2.36E-01 | 0.16 | 0.84 |
| RP_POS_153_06589a0_682 | | 153.0659 | 0.68 | | RP | POS | 153.0656(100), 110.0607(3) | | 5853313 | 3745793 | 5289647 | 3231740 | 2.36E-01 | -0.16 | 0.69 |
| RP_POS_203_12782a9_21 | | 203.1278 | 9.21 | | RP | POS | | | 8162236 | 3881978 | 7528984 | 3917809 | 2.36E-01 | -0.16 | 0.83 |
| RP_POS_436_34286a11_408 | | 436.3429 | 11.41 | | RP | POS | | | 3695878 | 528618 | 3616919 | 448099 | 2.36E-01 | -0.16 | 0.94 |
| RP_POS_271_11484a4_736 | | 271.1148 | 4.74 | | RP | POS | | | 310342 | 621265 | 232103 | 318171 | 2.36E-01 | -0.17 | 0.81 |
| HILIC_POS_141_06573a4_272 | | 141.0657 | 4.27 | | HILIC | POS | | | 626227 | 1625133 | 958202 | 2347518 | 2.37E-01 | 0.17 | 0.73 |
| HILIC_POS_168_08807a1_373 | | 168.0881 | 1.37 | | HILIC | POS | | | 527751 | 167688 | 554748 | 166043 | 2.37E-01 | 0.16 | 0.78 |
| HILIC_POS_191_0403a6_138 | | 191.0403 | 6.14 | | HILIC | POS | | | 1199873 | 57826.5 | 1210680 | 73446.5 | 2.37E-01 | 0.16 | 0.78 |

**Supplemantary table 1**: The non-targeted metabolomics results ordered by the VIP value (p value < 0.0003 are shaded dark green, p value < 0.05 are shaded light green, d values below 0 are blue and above 0 are red)

| | | | | | | | | | Cases | | Controls | | Welch test | Cohen's | |
|---|---|---|---|---|---|---|---|---|---|---|---|---|---|---|---|
| Feature_ID | Metabolite_name | Average_Mz | Average_RT_min_ | HMDB ID | RP_HILIC | POS_NEG | Cureted_MSMS | ID_level | Mean | SD | Mean2 | SD3 | p | d | VIP |
| HILIC_POS_531_40881a1_35 | | 531.4088 | 1.35 | | HILIC | POS | | | 34733.3 | 15340.8 | 37180.9 | 14936.4 | 2.37E-01 | 0.16 | 0.81 |
| RP_NEG_838_59784a13_788 | | 838.5978 | 13.79 | | RP | NEG | | | 7837622 | 1381591 | 8087540 | 1665495 | 2.37E-01 | 0.16 | 0.85 |
| RP_POS_262_08606a7_985 | | 262.0861 | 7.99 | | RP | POS | | | 364154 | 177757 | 392677 | 174732 | 2.37E-01 | 0.16 | 0.89 |
| RP_POS_353_34146a10_767 | | 353.3415 | 10.77 | | RP | POS | | | 348033 | 122189 | 368607 | 130891 | 2.37E-01 | 0.16 | 0.8 |
| RP_POS_548_80969a10_608 | | 548.8097 | 10.61 | | RP | POS | | | 1502945 | 482073 | 1586490 | 542697 | 2.37E-01 | 0.16 | 1 |
| RP_NEG_416_60858a15_493 | | 416.6086 | 15.49 | | RP | NEG | | | 708253 | 122448 | 688642 | 119965 | 2.37E-01 | -0.16 | 0.72 |
| RP_NEG_459_19601a11_142 | | 459.1960 | 11.14 | | RP | NEG | | | 845585 | 163208 | 820547 | 146827 | 2.37E-01 | -0.16 | 1.34 |
| RP_POS_654_95404a10_594 | | 654.9540 | 10.59 | | RP | POS | | | 567407 | 312991 | 519025 | 286490 | 2.37E-01 | -0.16 | 0.83 |
| RP_POS_762_55701a9_657 | | 762.5570 | 9.66 | | RP | POS | | | 3870849 | 1781369 | 3591095 | 1679837 | 2.37E-01 | -0.16 | 0.8 |
| RP_NEG_415_20737a10_962 | | 415.2074 | 10.96 | | RP | NEG | | | 468257 | 285609 | 513149 | 270904 | 2.38E-01 | 0.16 | 1.14 |
| RP_POS_236_07137a10_44 | | 236.0714 | 10.44 | | RP | POS | | | 9E+07 | 4E+07 | 9.7E+07 | 3.8E+07 | 2.38E-01 | 0.16 | 0.75 |
| RP_POS_336_01852a15_493 | | 336.0185 | 15.49 | | RP | POS | | | 216767 | 96436.1 | 232545 | 98455.6 | 2.38E-01 | 0.16 | 0.93 |
| RP_POS_366_18466a10_963 | | 366.1847 | 10.96 | | RP | POS | | | 2110129 | 1085698 | 2280930 | 1033201 | 2.38E-01 | 0.16 | 1.15 |
| HILIC_NEG_183_9341a4_637 | | 183.9341 | 4.64 | | HILIC | NEG | | | 3302912 | 1039824 | 3123001 | 1171353 | 2.38E-01 | -0.16 | 0.73 |
| RP_NEG_390_12744a11_144 | | 390.1274 | 11.14 | | RP | NEG | | | 205828 | 27571.2 | 201227 | 29197.6 | 2.38E-01 | -0.16 | 1.04 |
| RP_NEG_724_53125a13_595 | | 724.5313 | 13.60 | | RP | NEG | | | 749839 | 332363 | 695994 | 333879 | 2.38E-01 | -0.16 | 0.73 |
| RP_POS_318_19092a5_571 | | 318.1909 | 5.57 | | RP | POS | | | 230140 | 221726 | 201548 | 126787 | 2.38E-01 | -0.16 | 0.68 |
| RP_POS_394_80554a15_418 | | 394.8055 | 15.42 | | RP | POS | | | 585009 | 332537 | 531650 | 327982 | 2.38E-01 | -0.16 | 0.89 |
| RP_POS_666_26819a11_292 | | 666.2682 | 11.29 | | RP | POS | | | 187444 | 150610 | 164850 | 129757 | 2.38E-01 | -0.16 | 0.75 |
| RP_POS_349_63678a5_554 | | 349.6368 | 5.55 | | RP | POS | | | 175601 | 84793.2 | 193706 | 131113 | 2.39E-01 | 0.17 | 0.85 |
| HILIC_NEG_100_93348a5_175 | | 100.9335 | 5.18 | | HILIC | NEG | | | 2.9E+07 | 1.3E+07 | 3.2E+07 | 1.5E+07 | 2.39E-01 | 0.16 | 0.81 |
| RP_POS_520_33405a5_084 | | 520.3341 | 5.08 | | RP | POS | | | 577527 | 124992 | 601081 | 161770 | 2.39E-01 | 0.16 | 0.87 |
| HILIC_POS_252_18076a3_11 | | 252.1808 | 3.11 | | HILIC | POS | | | 102803 | 54862.9 | 94454.9 | 49007.8 | 2.39E-01 | -0.16 | 0.88 |
| RP_NEG_311_29593a12_108 | | 311.2959 | 12.11 | | RP | NEG | | | 2704955 | 923252 | 2551758 | 972833 | 2.39E-01 | -0.16 | 1.38 |
| RP_NEG_550_79419a15_381 | | 550.7942 | 15.38 | | RP | NEG | | | 667313 | 103688 | 648333 | 128284 | 2.39E-01 | -0.16 | 0.79 |
| RP_POS_245_27605a9_211 | | 245.2761 | 9.21 | | RP | POS | | | 301164 | 236571 | 267576 | 181269 | 2.39E-01 | -0.16 | 0.92 |
| RP_POS_322_24319a10_394 | | 322.2432 | 10.39 | | RP | POS | | | 1361998 | 307608 | 1314256 | 285712 | 2.39E-01 | -0.16 | 1.14 |
| RP_POS_343_11533a6_619 | | 343.1153 | 6.62 | | RP | POS | | | 107605 | 51858.1 | 99975.4 | 43052 | 2.39E-01 | -0.16 | 1.09 |
| RP_POS_512_41559a10_536 | | 512.4156 | 10.54 | | RP | POS | | | 4304434 | 796313 | 4184001 | 702742 | 2.39E-01 | -0.16 | 0.89 |
| RP_POS_695_98602a10_235 | | 695.9860 | 10.24 | | RP | POS | | | 866297 | 391616 | 802837 | 394962 | 2.39E-01 | -0.16 | 0.83 |
| RP_POS_705_59021a13_108 | | 705.5902 | 13.11 | | RP | POS | | | 2985890 | 1253228 | 2790001 | 1180904 | 2.39E-01 | -0.16 | 1.03 |
| RP_NEG_570_84546a15_342 | | 570.8455 | 15.34 | | RP | NEG | | | 1073438 | 61518.1 | 1060741 | 90816.5 | 2.39E-01 | -0.17 | 1.03 |
| RP_POS_471_73779a5_158 | | 471.7378 | 5.16 | | RP | POS | | | 5112303 | 3665792 | 4352364 | 5431140 | 2.39E-01 | -0.17 | 0.7 |
| RP_POS_355_28412a10_586 | | 355.2841 | 10.59 | | RP | POS | | | 1336962 | 1138915 | 1563037 | 1593127 | 2.40E-01 | 0.17 | 0.68 |
| RP_POS_275_16403a10_513 | | 275.1640 | 10.51 | | RP | POS | | | 3862344 | 1932688 | 4172125 | 1915287 | 2.40E-01 | 0.16 | 1.19 |
| RP_POS_428_37238a9_835 | Octadecanoylcarnitine | 428.3724 | 9.84 | | RP | POS | 85.0293(100), 428.3777(40) | 1 | 3537348 | 1081283 | 3711736 | 1087577 | 2.40E-01 | 0.16 | 0.95 |
| HILIC_NEG_170_98744a0_934 | | 170.9874 | 0.93 | | HILIC | NEG | | | 387050 | 44394.6 | 379959 | 43818.5 | 2.40E-01 | -0.16 | 0.82 |
| HILIC_NEG_200_85919a4_905 | | 200.8592 | 4.91 | | HILIC | NEG | | | 4550496 | 1494299 | 4315046 | 1438232 | 2.40E-01 | -0.16 | 0.98 |
| HILIC_POS_609_39825a1_233 | | 609.3983 | 1.23 | | HILIC | POS | | | 3589294 | 1047343 | 3434513 | 882249 | 2.40E-01 | -0.16 | 0.95 |
| RP_NEG_217_86566a0_638 | | 217.8657 | 0.64 | | RP | NEG | | | 231782 | 95088.5 | 219142 | 61046.5 | 2.40E-01 | -0.16 | 0.8 |
| RP_NEG_287_18622a7_961 | | 287.1862 | 7.96 | | RP | NEG | | | 246832 | 35923.1 | 241351 | 32341.4 | 2.40E-01 | -0.16 | 1.06 |
| RP_POS_313_16687a11_144 | | 313.1669 | 11.14 | | RP | POS | | | 3519124 | 606085 | 3417907 | 647677 | 2.40E-01 | -0.16 | 0.93 |
| RP_POS_448_33859a10_09 | | 448.3386 | 10.09 | | RP | POS | | | 3117631 | 1583530 | 2880934 | 1369083 | 2.40E-01 | -0.16 | 0.84 |
| HILIC_POS_208_13319a1_32 | | 208.1332 | 1.32 | | HILIC | POS | 208.1314(100) | | 504680 | 430185 | 453247 | 177570 | 2.40E-01 | -0.17 | 0.69 |
| HILIC_POS_429_0062a5_481 | | 429.0062 | 5.48 | | HILIC | POS | | | 99671.3 | 50788.1 | 108424 | 57456 | 2.41E-01 | 0.16 | 0.74 |
| RP_NEG_178_86478a15_439 | | 178.8648 | 15.44 | | RP | NEG | | | 2442723 | 498849 | 2535823 | 640157 | 2.41E-01 | 0.16 | 0.93 |
| RP_NEG_462_83398a15_373 | | 462.8340 | 15.37 | | RP | NEG | | | 635351 | 115187 | 655504 | 133508 | 2.41E-01 | 0.16 | 0.8 |
| RP_POS_376_32083a11_402 | | 376.3208 | 11.40 | | RP | POS | | | 271575 | 220815 | 311117 | 265743 | 2.41E-01 | 0.16 | 0.98 |
| RP_POS_481_01767a10_386 | | 481.0177 | 10.39 | | RP | POS | | | 140873 | 75826.5 | 154623 | 93184 | 2.41E-01 | 0.16 | 1.05 |
| RP_POS_422_30209a11_047 | | 422.3021 | 11.05 | | RP | POS | 422.3042(100) | | 484356 | 178968 | 459231 | 134463 | 2.41E-01 | -0.16 | 0.8 |
| HILIC_NEG_315_04187a5_468 | | 315.0419 | 5.47 | | HILIC | NEG | | | 4202382 | 649140 | 4095500 | 680426 | 2.41E-01 | -0.16 | 0.86 |
| RP_POS_623_45197a10_462 | | 623.4520 | 10.46 | | RP | POS | | | 3143977 | 1112806 | 2943505 | 1350835 | 2.41E-01 | -0.16 | 0.97 |
| HILIC_POS_197_10065a6_964 | | 197.1007 | 6.96 | | HILIC | POS | | | 649334 | 100836 | 666506 | 112321 | 2.42E-01 | 0.16 | 0.73 |
| HILIC_POS_85_02923a2_858 | | 85.0292 | 2.86 | | HILIC | POS | | | 818702 | 388317 | 887453 | 460406 | 2.42E-01 | 0.16 | 0.88 |
| RP_POS_371_24292a10_251 | | 371.2429 | 10.25 | | RP | POS | | | 1.1E+07 | 5975617 | 1.2E+07 | 5816219 | 2.42E-01 | 0.16 | 1.19 |
| RP_POS_321_2403a10_159 | | 321.2403 | 10.16 | | RP | POS | 321.2424(100) | | 2.7E+07 | 4301792 | 2.6E+07 | 4169065 | 2.42E-01 | -0.16 | 1.16 |
| HILIC_POS_64_01669a5_462 | | 64.0167 | 5.46 | | HILIC | POS | | | 6454928 | 967518 | 6309419 | 855772 | 2.42E-01 | -0.16 | 0.69 |
| RP_NEG_446_29184a8_595 | | 446.2918 | 8.60 | | RP | NEG | | | 1043506 | 1286621 | 805043 | 1640551 | 2.42E-01 | -0.16 | 0.92 |
| RP_NEG_452_92407a10_434 | | 452.9241 | 10.43 | | RP | NEG | | | 336324 | 146505 | 314209 | 130594 | 2.42E-01 | -0.16 | 0.72 |
| RP_POS_123_08061a9_702 | | 123.0806 | 9.70 | | RP | POS | | | 6879921 | 370735 | 6823964 | 329610 | 2.42E-01 | -0.16 | 0.83 |
| RP_POS_197_15347a7_957 | | 197.1535 | 7.96 | | RP | POS | | | 2367913 | 264648 | 2321531 | 308326 | 2.42E-01 | -0.16 | 0.81 |
| RP_POS_358_10812a8_992 | | 358.1081 | 8.99 | | RP | POS | | | 497810 | 257021 | 455508 | 270106 | 2.42E-01 | -0.16 | 0.76 |
| RP_POS_224_16443a8_942 | | 224.1644 | 8.94 | | RP | POS | | | 856287 | 5310092 | 271345 | 814704 | 2.42E-01 | -0.19 | 0.69 |
| HILIC_NEG_100_9335a4_257 | | 100.9335 | 4.26 | | HILIC | NEG | | | 3.7E+07 | 6626284 | 3.8E+07 | 7207672 | 2.43E-01 | 0.16 | 1.18 |
| HILIC_POS_480_30811a1_796 | | 480.3081 | 1.80 | | HILIC | POS | | | 703889 | 282698 | 756387 | 362831 | 2.43E-01 | 0.16 | 0.8 |
| RP_NEG_253_10838a6_387 | | 253.1084 | 6.39 | | RP | NEG | | | 819833 | 35759.5 | 825796 | 38601.6 | 2.43E-01 | 0.16 | 1.04 |
| RP_POS_433_28259a12_117 | | 433.2826 | 12.12 | | RP | POS | | | 361634 | 95572.8 | 345349 | 106697 | 2.43E-01 | -0.16 | 1.23 |
| RP_POS_617_51135a11_585 | | 617.5114 | 11.59 | | RP | POS | | | 4112372 | 9974968 | 2778897 | 6649700 | 2.43E-01 | -0.16 | 0.75 |
| RP_NEG_652_29395a10_61 | | 652.2940 | 10.61 | | RP | NEG | | | 2964863 | 499943 | 3048041 | 538115 | 2.44E-01 | 0.16 | 0.99 |
| RP_POS_505_6955a10_203 | | 505.6955 | 10.20 | | RP | POS | | | 294760 | 179477 | 322652 | 171032 | 2.44E-01 | 0.16 | 0.76 |
| RP_NEG_187_09753a5_974 | Azelaic acid | 187.0975 | 5.97 | | RP | NEG | 125.097(100), 187.0971(40) | 1 | 2087079 | 204055 | 2047586 | 278590 | 2.44E-01 | -0.16 | 1.14 |
| HILIC_NEG_221_90726a6_316 | | 221.9073 | 6.32 | | HILIC | NEG | | | 480629 | 126063 | 459827 | 134134 | 2.44E-01 | -0.16 | 0.82 |
| HILIC_POS_178_10744a3_241 | | 178.1074 | 3.24 | | HILIC | POS | | | 730677 | 385674 | 668269 | 395605 | 2.44E-01 | -0.16 | 0.83 |
| RP_NEG_601_77753a4_57 | | 601.7775 | 4.57 | | RP | NEG | | | 449525 | 298053 | 405016 | 262043 | 2.44E-01 | -0.16 | 1.33 |
| RP_POS_369_23206a11_592 | | 369.2321 | 11.59 | | RP | POS | | | 632026 | 495407 | 567961 | 299523 | 2.44E-01 | -0.16 | 0.91 |
| RP_POS_796_52429a13_223 | | 796.5243 | 13.22 | | RP | POS | | | 7748883 | 3257543 | 8315469 | 3788166 | 2.45E-01 | 0.16 | 0.86 |
| RP_NEG_231_93182a13_78 | | 231.9318 | 13.78 | | RP | NEG | | | 1796370 | 1202452 | 1618272 | 1042534 | 2.45E-01 | -0.16 | 0.83 |
| RP_POS_408_30856a11_025 | | 408.3086 | 11.03 | | RP | POS | | | 4613487 | 6955318 | 3685310 | 4682394 | 2.45E-01 | -0.16 | 0.76 |
| RP_POS_550_70477a10_161 | | 550.7048 | 10.16 | | RP | POS | | | 692809 | 368367 | 637666 | 326084 | 2.45E-01 | -0.16 | 0.84 |
| RP_POS_820_62061a13_939 | | 820.6206 | 13.94 | | RP | POS | | | 2928096 | 1926782 | 2628900 | 1839520 | 2.45E-01 | -0.16 | 0.9 |
| HILIC_POS_145_13359a2_597 | | 145.1336 | 2.60 | | HILIC | POS | | | 231139 | 701549 | 507463 | 2267296 | 2.46E-01 | 0.19 | 0.86 |
| HILIC_NEG_85_00388a0_934 | | 85.0039 | 0.93 | | HILIC | NEG | | | 1.2E+07 | 2575374 | 1.2E+07 | 3031962 | 2.46E-01 | 0.16 | 0.85 |
| RP_NEG_550_35272a10_792 | | 550.3527 | 10.79 | | RP | NEG | | | 629654 | 189210 | 658588 | 175678 | 2.46E-01 | 0.16 | 0.88 |
| RP_NEG_220_77654a15_476 | | 220.7765 | 15.48 | | RP | NEG | | | 382819 | 162395 | 357062 | 161705 | 2.46E-01 | -0.16 | 0.75 |
| RP_NEG_264_99084a8_124 | | 264.9908 | 8.12 | | RP | NEG | | | 1662591 | 201961 | 1631535 | 189534 | 2.46E-01 | -0.16 | 0.78 |
| RP_POS_158_15404a7_627 | | 158.1540 | 7.63 | | RP | POS | | | 3.9E+07 | 2656061 | 3.9E+07 | 2000346 | 2.46E-01 | -0.16 | 0.83 |
| RP_POS_159_96922a0_631 | | 159.9692 | 0.63 | | RP | POS | | | 2132547 | 866835 | 2006087 | 730127 | 2.46E-01 | -0.16 | 0.85 |
| RP_POS_275_63336a10_245 | | 275.6334 | 10.25 | | RP | POS | | | 1344870 | 1327112 | 1144353 | 1203531 | 2.46E-01 | -0.16 | 0.81 |
| RP_POS_399_30865a10_644 | | 399.3087 | 10.64 | | RP | POS | | | 6068168 | 1194633 | 5867744 | 1313351 | 2.46E-01 | -0.16 | 1.25 |
| RP_POS_683_47363a9_737 | | 683.4736 | 9.74 | | RP | POS | | | 904919 | 443504 | 839377 | 383833 | 2.46E-01 | -0.16 | 0.96 |
| RP_NEG_250_14549a9_647 | | 250.1455 | 9.65 | | RP | NEG | | | 883886 | 115749 | 901790 | 110381 | 2.47E-01 | 0.16 | 0.84 |
| RP_POS_622_76312a10_113 | | 622.7631 | 10.11 | | RP | POS | | | 241366 | 139477 | 266615 | 173999 | 2.47E-01 | 0.16 | 0.84 |
| RP_POS_844_22455a9_275 | | 844.2246 | 9.28 | | RP | POS | | | 825839 | 239071 | 862981 | 230081 | 2.47E-01 | 0.16 | 0.8 |
| RP_POS_786_60144a14_099 | PC 36:2 (18:1/18:1) | 786.6014 | 14.10 | | RP | POS | 184.0743(100), 86.095(10) | 1 | 5.5E+08 | 1.3E+08 | 5.7E+08 | 1.3E+08 | 2.47E-01 | 0.16 | 0.85 |
| RP_NEG_818_55035a13_459 | | 818.5504 | 13.46 | | RP | NEG | | | 2349681 | 1530332 | 2142188 | 1087549 | 2.47E-01 | -0.16 | 0.86 |
| HILIC_NEG_259_00128a3_052 | | 259.0013 | 3.05 | | HILIC | NEG | | | 440661 | 42864.2 | 431235 | 70389.7 | 2.47E-01 | -0.17 | 0.82 |
| RP_POS_193_09488a3_083 | | 193.0949 | 3.08 | | RP | POS | | | 1.3E+07 | 2.6E+07 | 9748701 | 1E+07 | 2.47E-01 | -0.17 | 0.85 |
| RP_POS_592_4198a10_519 | | 592.4198 | 10.52 | | RP | POS | | | 385030 | 256175 | 910516 | 4494734 | 2.48E-01 | 0.22 | 0.73 |
| RP_NEG_383_15347a6_259 | | 383.1535 | 6.26 | | RP | NEG | | | 1.1E+07 | 9949780 | 1.3E+07 | 9349164 | 2.48E-01 | 0.16 | 0.76 |
| RP_POS_288_14346a5_405 | | 288.1435 | 5.41 | | RP | POS | | | 395215 | 86347 | 408197 | 78212.8 | 2.48E-01 | 0.16 | 0.69 |
| RP_POS_336_2507a9_544 | | 336.2507 | 9.54 | | RP | POS | | | 308388 | 64862.5 | 321641 | 96859.9 | 2.48E-01 | 0.16 | 0.99 |
| RP_POS_514_28711a6_047 | | 514.2871 | 6.05 | | RP | POS | | | 1.6E+07 | 912061 | 1.6E+07 | 980179 | 2.48E-01 | 0.16 | 0.75 |
| RP_POS_560_30646a6_892 | | 560.3065 | 6.89 | | RP | POS | | | 466813 | 259230 | 512886 | 315225 | 2.48E-01 | 0.16 | 0.74 |
| RP_POS_578_40442a10_141 | | 578.4044 | 10.14 | | RP | POS | | | 791861 | 420486 | 858169 | 418313 | 2.48E-01 | 0.16 | 0.83 |
| RP_POS_652_18823a8_642 | | 652.1882 | 8.64 | | RP | POS | | | 1215459 | 110025 | 1233127 | 112942 | 2.48E-01 | 0.16 | 1.05 |
| RP_POS_203_13892a1_758 | Ala-Ile | 203.1389 | 1.76 | | RP | POS | 86.0974(100) | 2 | 514411 | 212765 | 480274 | 218237 | 2.48E-01 | -0.16 | 0.89 |
| HILIC_POS_220_11798a1_015 | Pantothenic acid | 220.1180 | 1.02 | | HILIC | POS | 90.0558(100), 202.1076(40) | 2 | 696641 | 259652 | 656859 | 244404 | 2.48E-01 | -0.16 | 0.75 |
| RP_NEG_521_38507a12_05 | | 521.3851 | 12.05 | | RP | NEG | | | 1351321 | 112946 | 1333993 | 106488 | 2.48E-01 | -0.16 | 1.25 |
| RP_POS_145_10133a10_841 | | 145.1013 | 10.84 | | RP | POS | | | 443723 | 98445.1 | 427666 | 104189 | 2.48E-01 | -0.16 | 0.68 |
| RP_POS_290_23221a9_461 | | 290.2322 | 9.46 | | RP | POS | | | 2973027 | 2163786 | 2643798 | 2005118 | 2.48E-01 | -0.16 | 0.8 |
| RP_POS_332_89532a15_413 | | 332.8953 | 15.41 | | RP | POS | | | 1711963 | 608263 | 1624321 | 504581 | 2.48E-01 | -0.16 | 1.04 |
| RP_POS_394_20746a9_961 | | 394.2075 | 9.96 | | RP | POS | | | 5798709 | 3631583 | 5154614 | 4407072 | 2.48E-01 | -0.16 | 1.15 |
| RP_POS_600_4671a10_458 | | 600.4671 | 10.46 | | RP | POS | | | 4245493 | 448388 | 4177028 | 419440 | 2.48E-01 | -0.16 | 0.96 |
| RP_NEG_283_07043a10_245 | | 283.0704 | 10.25 | | RP | NEG | | | 431845 | 644870 | 525025 | 538991 | 2.49E-01 | 0.16 | 0.66 |
| RP_NEG_312_25443a10_609 | | 312.2544 | 10.61 | | RP | NEG | | | 275483 | 45610 | 282972 | 48905.2 | 2.49E-01 | 0.16 | 0.91 |
| RP_POS_337_16669a10_963 | | 337.1667 | 10.96 | | RP | POS | | | 7286341 | 4108477 | 7915195 | 3876917 | 2.49E-01 | 0.16 | 1.12 |
| RP_POS_584_72748a10_266 | | 584.7275 | 10.27 | | RP | POS | | | 463044 | 242150 | 507915 | 315252 | 2.49E-01 | 0.16 | 0.89 |
| RP_POS_712_89856a11_538 | | 712.8986 | 11.54 | | RP | POS | | | 165394 | 118976 | 185878 | 138199 | 2.49E-01 | 0.16 | 0.78 |
| RP_POS_313_27353a10_235 | | 313.2735 | 10.24 | | RP | POS | | | 8905651 | 1235369 | 8714628 | 1189135 | 2.49E-01 | -0.16 | 1.11 |
| RP_POS_375_25049a10_412 | | 375.2505 | 10.41 | | RP | POS | | | 1536199 | 1082897 | 1784682 | 1893400 | 2.50E-01 | 0.17 | 0.92 |
| RP_POS_420_31067a10_629 | | 420.3107 | 10.63 | | RP | POS | | | 214169 | 220445 | 262685 | 365399 | 2.50E-01 | 0.17 | 1 |
| HILIC_POS_189_07312a5_223 | | 189.0731 | 5.22 | | HILIC | POS | | | 1890542 | 850057 | 2053933 | 1172099 | 2.50E-01 | 0.16 | 0.7 |
| RP_POS_276_63342a10_369 | | 276.6334 | 10.37 | | RP | POS | | | 813382 | 248032 | 855734 | 285217 | 2.50E-01 | 0.16 | 1.02 |
| RP_POS_296_2225a9_113 | | 296.2225 | 9.11 | | RP | POS | | | 1.2E+07 | 813695 | 1.2E+07 | 921869 | 2.50E-01 | 0.16 | 1.26 |
| HILIC_POS_158_15379a1_22 | | 158.1538 | 1.22 | | HILIC | POS | | | 3168296 | 1759684 | 2927084 | 1309886 | 2.50E-01 | -0.16 | 0.86 |
| RP_NEG_365_23135a10_773 | | 365.2314 | 10.77 | | RP | NEG | | | 318188 | 45257.1 | 311232 | 43228.5 | 2.50E-01 | -0.16 | 1.19 |
| RP_POS_220_1181a2_572 | | 220.1181 | 2.57 | | RP | POS | | | 421330 | 167029 | 396984 | 143274 | 2.50E-01 | -0.16 | 0.74 |
| RP_POS_669_95721a10_42 | | 669.9572 | 10.42 | | RP | POS | | | 1406276 | 564045 | 1307378 | 677127 | 2.50E-01 | -0.16 | 1.14 |
| RP_POS_691_98877a11_546 | | 691.9888 | 11.55 | | RP | POS | | | 98055.9 | 54818.3 | 113991 | 127971 | 2.51E-01 | 0.17 | 0.75 |
| HILIC_NEG_276_99133a1_362 | | 276.9913 | 1.36 | | HILIC | NEG | | | 1485057 | 475174 | 1553992 | 405848 | 2.51E-01 | 0.16 | 0.77 |
| HILIC_POS_206_88597a2_982 | | 206.8860 | 2.98 | | HILIC | POS | | | 7936103 | 1593670 | 8183371 | 1554528 | 2.51E-01 | 0.16 | 0.74 |
| RP_POS_324_25305a9_79 | | 324.2531 | 9.79 | | RP | POS | | | 438087 | 108510 | 456013 | 118585 | 2.51E-01 | 0.16 | 0.93 |
| RP_POS_830_56604a13_293 | | 830.5660 | 13.29 | | RP | POS | | | 5.5E+07 | 1.5E+07 | 5.7E+07 | 1.7E+07 | 2.51E-01 | 0.16 | 1.01 |
| RP_NEG_244_94289a0_667 | | 244.9429 | 0.67 | | RP | NEG | | | 587451 | 175721 | 563568 | 128275 | 2.51E-01 | -0.16 | 0.73 |
| RP_NEG_405_15781a11_152 | | 405.1578 | 11.15 | | RP | NEG | | | 532782 | 93755.1 | 519420 | 76819.2 | 2.51E-01 | -0.16 | 0.72 |
| RP_NEG_694_48517a11_591 | | 694.4852 | 11.59 | | RP | NEG | | | 358598 | 68091 | 348437 | 61595.9 | 2.51E-01 | -0.16 | 1.15 |
| RP_POS_213_14851a5_914 | | 213.1485 | 5.91 | | RP | POS | | | 83707.1 | 105617 | 133644 | 420778 | 2.52E-01 | 0.19 | 0.85 |
| HILIC_POS_151_03551a7_405 | | 151.0355 | 7.41 | | HILIC | POS | | | 1.4E+07 | 1137551 | 1.4E+07 | 1202570 | 2.52E-01 | 0.16 | 1.2 |
| RP_POS_347_22153a10_39 | | 347.2215 | 10.39 | | RP | POS | | | 1002024 | 474446 | 1079579 | 510432 | 2.52E-01 | 0.16 | 1.28 |
| RP_POS_386_32602a10_67 | | 386.3260 | 10.67 | | RP | POS | | | 119035 | 101939 | 138313 | 137951 | 2.52E-01 | 0.16 | 0.93 |
| RP_POS_593_41187a10_133 | | 593.4119 | 10.13 | | RP | POS | | | 1072582 | 424808 | 1154662 | 593951 | 2.52E-01 | 0.16 | 0.92 |
| RP_POS_872_5379a13_13 | | 872.5379 | 13.13 | | RP | POS | | | 5112545 | 1596806 | 5370492 | 1683519 | 2.52E-01 | 0.16 | 0.67 |
| RP_NEG_221_15446a8_304 | | 221.1545 | 8.30 | | RP | NEG | | | 213049 | 35956.8 | 207657 | 33002.1 | 2.52E-01 | -0.16 | 1.01 |
| RP_POS_347_09979a9_209 | | 347.0998 | 9.21 | | RP | POS | | | 3225356 | 1625642 | 2967189 | 1660350 | 2.52E-01 | -0.16 | 1.03 |
| RP_POS_378_95132a15_454 | | 378.9513 | 15.45 | | RP | POS | | | 369404 | 121514 | 353080 | 86659.5 | 2.52E-01 | -0.16 | 0.78 |
| RP_POS_680_46265a10_07 | | 680.4627 | 10.07 | | RP | POS | | | 606724 | 308089 | 552898 | 369743 | 2.52E-01 | -0.16 | 0.93 |
| HILIC_NEG_187_94228a6_181 | | 187.9423 | 6.18 | | HILIC | NEG | | | 205044 | 91155.8 | 219660 | 95076.1 | 2.53E-01 | 0.16 | 0.73 |
| HILIC_NEG_222_90079a3_106 | | 222.9008 | 3.11 | | HILIC | NEG | | | 349845 | 23795.2 | 353408 | 21891 | 2.53E-01 | 0.16 | 0.67 |
| RP_POS_163_11188a8_171 | | 163.1119 | 8.17 | | RP | POS | | | 5478452 | 477970 | 5552532 | 468669 | 2.53E-01 | 0.16 | 1.2 |
| RP_POS_621_44556a15_162 | | 621.4456 | 15.16 | | RP | POS | | | 398200 | 375347 | 467944 | 497074 | 2.53E-01 | 0.16 | 0.83 |
| RP_NEG_415_19794a7_569 | | 415.1979 | 7.57 | | RP | NEG | 415.2095(100), 113.0244(40) | | 509964 | 293855 | 467319 | 252459 | 2.53E-01 | -0.16 | 0.7 |
| RP_NEG_130_94434a15_485 | | 130.9443 | 15.49 | | RP | NEG | | | 3744898 | 840497 | 3614570 | 825562 | 2.53E-01 | -0.16 | 0.9 |
| RP_NEG_816_57727a13_632 | | 816.5773 | 13.63 | | RP | NEG | | | 4884234 | 1095480 | 4706834 | 1164579 | 2.53E-01 | -0.16 | 0.93 |
| RP_POS_327_22076a11_142 | | 327.2208 | 11.14 | | RP | POS | | | 1307212 | 498293 | 1233231 | 450376 | 2.53E-01 | -0.16 | 1.22 |
| RP_POS_301_10477a7_666 | | 301.1048 | 7.67 | | RP | POS | | | 1272552 | 83943.5 | 1286129 | 89414.8 | 2.54E-01 | 0.16 | 0.8 |
| RP_POS_560_41559a10_789 | | 560.4156 | 10.79 | | RP | POS | | | 1227432 | 744528 | 1366303 | 995411 | 2.54E-01 | 0.16 | 1.27 |
| RP_POS_610_42584a9_572 | | 610.4258 | 9.57 | | RP | POS | | | 1116819 | 584924 | 1217691 | 691742 | 2.54E-01 | 0.16 | 1.13 |
| RP_POS_717_59052a13_137 | | 717.5905 | 13.14 | | RP | POS | | | 1976291 | 2690986 | 2493107 | 3747885 | 2.54E-01 | 0.16 | 1.04 |
| RP_NEG_584_35669a9_499 | | 584.3567 | 9.50 | | RP | NEG | | | 559157 | 254678 | 521534 | 228976 | 2.54E-01 | -0.16 | 0.77 |
| RP_POS_482_00116a10_386 | | 482.0012 | 10.39 | | RP | POS | | | 517480 | 277312 | 475384 | 263043 | 2.54E-01 | -0.16 | 0.75 |
| RP_POS_504_35666a10_218 | | 504.3567 | 10.22 | | RP | POS | | | 1009630 | 606417 | 924800 | 483997 | 2.54E-01 | -0.16 | 0.77 |
| RP_NEG_222_99176a0_68 | | 222.9918 | 0.68 | | RP | NEG | | | 781116 | 842678 | 941658 | 1162678 | 2.55E-01 | 0.16 | 0.96 |

**Supplemantary table 1**: The non-targeted metabolomics results ordered by the VIP value (p value < 0.0003 are shaded dark green, p value < 0.05 are shaded light green, d values below 0 are blue and above 0 are red)

| | | | | | | | | | Cases | | Controls | | Welch test | Cohen's | |
|---|---|---|---|---|---|---|---|---|---|---|---|---|---|---|---|
| Feature_ID | Metabolite_name | Average_Mz | Average_RT_min_ | HMDB ID | RP_HILIC | POS_NEG | Cureted_MSMS | ID_level | Mean | SD | Mean2 | SD3 | p | d | VIP |
| RP_NEG_557_32092a10_487 | | 557.3209 | 10.49 | | RP | NEG | | | 399930 | 119575 | 417490 | 106370 | 2.55E-01 | 0.16 | 0.77 |
| RP_POS_374_18829a10_494 | | 374.1883 | 10.49 | | RP | POS | | | 193701 | 134471 | 215066 | 139312 | 2.55E-01 | 0.16 | 0.87 |
| RP_POS_387_21405a9_526 | | 387.2141 | 9.53 | | RP | POS | | | 547678 | 216079 | 586421 | 272873 | 2.55E-01 | 0.16 | 1.06 |
| RP_POS_470_42178a11_709 | | 470.4218 | 11.71 | | RP | POS | | | 581940 | 72194.3 | 593222 | 72663.2 | 2.55E-01 | 0.16 | 0.88 |
| RP_POS_515_36475a10_041 | | 515.3648 | 10.04 | | RP | POS | | | 567936 | 367062 | 624166 | 355762 | 2.55E-01 | 0.16 | 0.69 |
| RP_NEG_389_27032a9_612 | | 389.2703 | 9.61 | | RP | NEG | | | 373071 | 362363 | 323676 | 272199 | 2.55E-01 | -0.16 | 0.68 |
| RP_POS_262_20074a9_21 | | 262.2007 | 9.21 | | RP | POS | | | 6670294 | 3023279 | 6192189 | 3100211 | 2.55E-01 | -0.16 | 0.72 |
| RP_POS_516_37396a7_609 | | 516.3740 | 7.61 | | RP | POS | | | 648593 | 407266 | 590774 | 337521 | 2.55E-01 | -0.16 | 0.95 |
| RP_NEG_180_97351a15_487 | | 180.9735 | 15.49 | | RP | NEG | | | 1011453 | 210913 | 1059309 | 369530 | 2.56E-01 | 0.16 | 0.93 |
| RP_NEG_457_23691a11_294 | | 457.2369 | 11.29 | | RP | NEG | | | 591236 | 394718 | 656971 | 444143 | 2.56E-01 | 0.16 | 0.87 |
| RP_NEG_464_09863a10_651 | | 464.0986 | 10.65 | | RP | NEG | | | 117925 | 49021.9 | 126487 | 59547.1 | 2.56E-01 | 0.16 | 0.75 |
| RP_NEG_922_58026a13_956 | | 922.5803 | 13.96 | | RP | NEG | | | 6206489 | 1180887 | 6415721 | 1468556 | 2.56E-01 | 0.16 | 0.74 |
| RP_NEG_297_24371a10_775 | FA 18:1+1O | 297.2437 | 10.78 | | RP | NEG | 297.2449(100), 279.2342(10) | 2 | 3086402 | 361483 | 3029917 | 365122 | 2.56E-01 | -0.16 | 1.16 |
| RP_POS_135_11697a8_874 | Carveol | 135.1170 | 8.87 | | RP | POS | 135.1174(100), 119.0856(70) | 2 | 628061 | 377915 | 690975 | 425723 | 2.56E-01 | 0.16 | 1.03 |
| HILIC_NEG_323_01434a1_26 | | 323.0143 | 1.26 | | HILIC | NEG | | | 483653 | 195074 | 455218 | 171831 | 2.56E-01 | -0.15 | 0.81 |
| RP_NEG_197_04295a3_585 | | 197.0430 | 3.59 | | RP | NEG | | | 624522 | 177700 | 596703 | 180092 | 2.56E-01 | -0.16 | 0.93 |
| RP_NEG_528_2641a8_395 | | 528.2641 | 8.40 | | RP | NEG | | | 2312832 | 2863203 | 1960771 | 1580450 | 2.56E-01 | -0.16 | 0.96 |
| RP_POS_484_38461a9_527 | | 484.3846 | 9.53 | | RP | POS | | | 1456877 | 770378 | 1337507 | 766289 | 2.56E-01 | -0.16 | 0.86 |
| RP_POS_534_3689a9_784 | | 534.3689 | 9.78 | | RP | POS | | | 1096840 | 583727 | 1013739 | 488535 | 2.56E-01 | -0.16 | 0.81 |
| RP_POS_544_38818a9_585 | | 544.3882 | 9.59 | | RP | POS | | | 2771064 | 1481201 | 3012500 | 1612721 | 2.57E-01 | 0.16 | 0.99 |
| RP_POS_685_14282a11_325 | | 685.1428 | 11.33 | | RP | POS | | | 173456 | 88739 | 188162 | 99695.8 | 2.57E-01 | 0.16 | 0.93 |
| RP_NEG_288_61935a7_436 | | 288.6194 | 7.44 | | RP | NEG | | | 1857512 | 2413785 | 2187273 | 1842051 | 2.57E-01 | 0.15 | 0.65 |
| RP_POS_129_05463a9_21 | 3-Methylglutaric acid | 129.0546 | 9.21 | | RP | POS | 101.06(100),59.0503(90) | 2 | 2.1E+07 | 1.1E+07 | 1.9E+07 | 1.1E+07 | 2.57E-01 | -0.16 | 0.84 |
| RP_NEG_339_19937a15_453 | | 339.1994 | 15.45 | | RP | NEG | | | 685699 | 84910 | 672084 | 89984.6 | 2.57E-01 | -0.16 | 0.67 |
| RP_POS_322_25037a11_147 | | 322.2504 | 11.15 | | RP | POS | | | 156137 | 96576 | 175279 | 142227 | 2.58E-01 | 0.16 | 0.88 |
| RP_POS_347_22076a9_529 | | 347.2208 | 9.53 | | RP | POS | | | 603465 | 211577 | 639676 | 251096 | 2.58E-01 | 0.16 | 1 |
| RP_POS_535_87415a9_586 | | 535.8742 | 9.59 | | RP | POS | | | 735689 | 347691 | 794948 | 409057 | 2.58E-01 | 0.16 | 1.09 |
| RP_POS_687_47827a11_39 | | 687.4783 | 11.39 | | RP | POS | | | 146919 | 75102.9 | 163484 | 127681 | 2.58E-01 | 0.16 | 0.77 |
| RP_POS_230_88991a15_393 | | 230.8899 | 15.39 | | RP | POS | | | 2310722 | 1661622 | 2550356 | 1440650 | 2.58E-01 | 0.15 | 0.81 |
| HILIC_POS_663_4552a1_348 | | 663.4552 | 1.35 | | HILIC | POS | | | 140482 | 52720 | 132336 | 52528.6 | 2.58E-01 | -0.15 | 0.69 |
| RP_POS_452_7373a5_095 | | 452.7373 | 5.10 | | RP | POS | | | 2713020 | 5547741 | 1989448 | 3781266 | 2.58E-01 | -0.16 | 0.96 |
| RP_POS_433_33121a11_962 | | 433.3312 | 11.96 | | RP | POS | | | 380006 | 483400 | 504113 | 994659 | 2.59E-01 | 0.17 | 0.79 |
| HILIC_POS_178_10738a1_054 | | 178.1074 | 1.05 | | HILIC | POS | | | 532762 | 253232 | 578739 | 330467 | 2.59E-01 | 0.16 | 0.87 |
| RP_NEG_329_16095a8_211 | | 329.1610 | 8.21 | | RP | NEG | | | 410733 | 268795 | 454313 | 292310 | 2.59E-01 | 0.16 | 1.25 |
| RP_POS_529_04541a10_458 | | 529.0454 | 10.46 | | RP | POS | | | 621940 | 250086 | 671638 | 370682 | 2.59E-01 | 0.16 | 0.8 |
| RP_NEG_221_01018a0_671 | | 221.0102 | 0.67 | | RP | NEG | | | 157927 | 34897.8 | 163251 | 34101.1 | 2.59E-01 | 0.15 | 0.7 |
| RP_POS_615_40656a10_962 | | 615.4066 | 10.96 | | RP | POS | | | 453720 | 427860 | 520942 | 441528 | 2.59E-01 | 0.15 | 1.08 |
| HILIC_POS_110_07156a6_783 | | 110.0716 | 6.78 | | HILIC | POS | | | 4533868 | 1222854 | 4374958 | 829230 | 2.59E-01 | -0.15 | 0.74 |
| RP_POS_307_10312a9_21 | | 307.1031 | 9.21 | | RP | POS | | | 1036975 | 589084 | 945580 | 592334 | 2.59E-01 | -0.15 | 0.8 |
| RP_POS_476_2774a10_608 | | 476.2774 | 10.61 | | RP | POS | | | 2784383 | 7085286 | 1871814 | 4685438 | 2.59E-01 | -0.16 | 0.89 |
| HILIC_NEG_212_91115a4_743 | | 212.9112 | 4.74 | | HILIC | NEG | | | 1368257 | 644690 | 1475032 | 729509 | 2.60E-01 | 0.16 | 0.84 |
| HILIC_NEG_236_81767a6_908 | | 236.8177 | 6.91 | | HILIC | NEG | | | 240628 | 101189 | 257438 | 115305 | 2.60E-01 | 0.16 | 0.81 |
| RP_NEG_191_86034a15_473 | | 191.8603 | 15.47 | | RP | NEG | | | 499353 | 54976.6 | 508859 | 66758.8 | 2.60E-01 | 0.16 | 0.79 |
| RP_NEG_307_1149a0_548 | | 307.1149 | 0.55 | | RP | NEG | | | 982577 | 213780 | 1024534 | 313075 | 2.60E-01 | 0.16 | 0.67 |
| RP_NEG_462_14264a9_583 | | 462.1426 | 9.58 | | RP | NEG | | | 274787 | 165607 | 302312 | 188864 | 2.60E-01 | 0.16 | 0.94 |
| RP_POS_1058_7196a10_928 | | 1058.7196 | 10.93 | | RP | POS | | | 539311 | 423499 | 609939 | 484568 | 2.60E-01 | 0.16 | 0.94 |
| RP_POS_365_26602a10_67 | | 365.2660 | 10.67 | | RP | POS | | | 385061 | 496502 | 479316 | 692536 | 2.60E-01 | 0.16 | 0.98 |
| HILIC_POS_114_06663a5_589 | | 114.0666 | 5.59 | | HILIC | POS | 114.0671(100), 86.0719(20) | | 414599 | 165295 | 441186 | 178153 | 2.60E-01 | 0.15 | 0.68 |
| RP_NEG_207_10254a7_719 | | 207.1025 | 7.72 | | RP | NEG | | | 7655200 | 242446 | 7690831 | 221046 | 2.60E-01 | 0.15 | 0.83 |
| RP_NEG_802_59796a10_957 | | 802.5980 | 10.96 | | RP | NEG | | | 349866 | 191674 | 380332 | 202646 | 2.60E-01 | 0.15 | 1.28 |
| RP_POS_589_47992a14_448 | | 589.4799 | 14.45 | | RP | POS | | | 2488074 | 1672804 | 2756540 | 1797830 | 2.60E-01 | 0.15 | 0.8 |
| RP_POS_814_57214a13_575 | | 814.5721 | 13.58 | | RP | POS | | | 6197010 | 2975267 | 6675277 | 3197487 | 2.60E-01 | 0.15 | 0.8 |
| HILIC_POS_118_12292a1_112 | | 118.1229 | 1.11 | | HILIC | POS | | | 568524 | 112776 | 553216 | 86328.8 | 2.60E-01 | -0.15 | 0.93 |
| RP_NEG_865_0932a9_278 | | 865.0932 | 9.28 | | RP | NEG | | | 1483218 | 102120 | 1467999 | 95802.2 | 2.60E-01 | -0.15 | 1.1 |
| RP_POS_411_43051a11_896 | | 411.4305 | 11.90 | | RP | POS | | | 559848 | 293611 | 515907 | 277292 | 2.60E-01 | -0.15 | 1.19 |
| RP_POS_517_01782a10_034 | | 517.0178 | 10.03 | | RP | POS | | | 457591 | 223468 | 422430 | 231503 | 2.60E-01 | -0.15 | 0.97 |
| RP_POS_422_76221a12_996 | | 422.7622 | 13.00 | | RP | POS | | | 4251244 | 4571981 | 3667496 | 2957303 | 2.60E-01 | -0.16 | 1.19 |
| RP_POS_791_58856a14_342 | | 791.5886 | 14.34 | | RP | POS | | | 820887 | 968557 | 703750 | 518898 | 2.60E-01 | -0.16 | 0.82 |
| RP_POS_203_50584a10_835 | | 203.5058 | 10.84 | | RP | POS | | | 556322 | 341495 | 747430 | 1651324 | 2.61E-01 | 0.19 | 0.84 |
| RP_POS_299_08914a4_465 | | 299.0891 | 4.47 | | RP | POS | 299.0891(100 | | 6.6E+09 | 1.3E+10 | 9E+09 | 1.7E+10 | 2.61E-01 | 0.16 | 1.04 |
| RP_NEG_430_29663a9_061 | | 430.2966 | 9.06 | | RP | NEG | | | 218335 | 111540 | 240736 | 168763 | 2.61E-01 | 0.16 | 0.73 |
| RP_POS_319_1947a10_962 | | 319.1947 | 10.96 | | RP | POS | | | 447321 | 275321 | 494264 | 328011 | 2.61E-01 | 0.16 | 1.01 |
| HILIC_NEG_144_92352a6_375 | | 144.9235 | 6.38 | | HILIC | NEG | | | 2.4E+07 | 5030794 | 2.5E+07 | 4886427 | 2.61E-01 | 0.15 | 0.86 |
| RP_NEG_654_13068a9_099 | | 654.1307 | 9.10 | | RP | NEG | | | 643416 | 38798.1 | 649481 | 39875 | 2.61E-01 | 0.15 | 0.87 |
| RP_NEG_660_26172a10_289 | | 660.2617 | 10.29 | | RP | NEG | | | 521636 | 152533 | 545613 | 158228 | 2.61E-01 | 0.15 | 0.94 |
| RP_POS_509_40469a10_682 | | 509.4047 | 10.68 | | RP | POS | | | 1092912 | 361357 | 1143681 | 300481 | 2.61E-01 | 0.15 | 1.03 |
| RP_POS_665_45685a10_251 | | 665.4569 | 10.25 | | RP | POS | | | 811237 | 385420 | 867147 | 342856 | 2.61E-01 | 0.15 | 0.84 |
| RP_NEG_820_56451a14_089 | PC 36:2 (18:0_18:2) | 820.5645 | 14.09 | | RP | NEG | 279.2332(100) | 2 | 2215858 | 1499616 | 2441904 | 1440030 | 2.61E-01 | 0.15 | 0.95 |
| HILIC_POS_144_10196a1_945 | | 144.1020 | 1.95 | | HILIC | POS | 144.1025(100), 98.096(5) | | 713058 | 614708 | 625068 | 533362 | 2.61E-01 | -0.15 | 0.72 |
| RP_NEG_288_93646a15_331 | | 288.9365 | 15.33 | | RP | NEG | | | 3406459 | 323285 | 3356534 | 325063 | 2.61E-01 | -0.15 | 1.06 |
| RP_POS_434_74432a13_001 | | 434.7443 | 13.00 | | RP | POS | | | 1293133 | 592342 | 1209568 | 496881 | 2.61E-01 | -0.15 | 1.02 |
| RP_POS_603_52167a15_389 | | 603.5217 | 15.39 | | RP | POS | | | 2965837 | 2246880 | 2648576 | 1888569 | 2.61E-01 | -0.15 | 0.69 |
| HILIC_POS_495_06305a3_146 | | 495.0631 | 3.15 | | HILIC | POS | | | 196201 | 13818 | 193159 | 23615 | 2.61E-01 | -0.16 | 0.72 |
| RP_NEG_297_15353a10_194 | | 297.1535 | 10.19 | | RP | NEG | | | 1033323 | 522939 | 1123188 | 632551 | 2.62E-01 | 0.16 | 1.04 |
| RP_POS_218_13799a1_429 | | 218.1380 | 1.43 | | RP | POS | 85.0291(100), 218.139(40) | | 2628065 | 998234 | 2778660 | 964956 | 2.62E-01 | 0.15 | 0.85 |
| RP_NEG_347_22119a10_961 | | 347.2212 | 10.96 | | RP | NEG | | | 1141275 | 714619 | 1247976 | 677068 | 2.62E-01 | 0.15 | 1.18 |
| RP_POS_305_02206a2_885 | | 305.0221 | 2.89 | | RP | POS | | | 760871 | 134201 | 781754 | 137330 | 2.62E-01 | 0.15 | 0.89 |
| HILIC_NEG_144_99216a3_055 | | 144.9922 | 3.06 | | HILIC | NEG | | | 810989 | 129802 | 793463 | 99002.5 | 2.62E-01 | -0.15 | 0.94 |
| RP_POS_245_17485a9_21 | | 245.1749 | 9.21 | | RP | POS | | | 5.1E+08 | 2.7E+08 | 4.7E+08 | 2.7E+08 | 2.62E-01 | -0.15 | 0.81 |
| RP_POS_386_88846a15_419 | | 386.8885 | 15.42 | | RP | POS | | | 280156 | 213328 | 248523 | 199679 | 2.62E-01 | -0.15 | 0.85 |
| RP_NEG_130_94437a13_287 | | 130.9444 | 13.29 | | RP | NEG | | | 2737151 | 300963 | 2785108 | 323001 | 2.63E-01 | 0.15 | 0.68 |
| RP_POS_339_18265a11_239 | | 339.1827 | 11.24 | | RP | POS | | | 4276725 | 2176732 | 4595054 | 1988007 | 2.63E-01 | 0.15 | 1 |
| RP_POS_282_27911a10_835 | Oleamide | 282.2791 | 10.84 | | RP | POS | 282.2814(100), 83.0862(70) | 1 | 2.1E+08 | 7.3E+07 | 2.3E+08 | 1.5E+08 | 2.63E-01 | 0.17 | 1.03 |
| RP_POS_626_29639a10_931 | | 626.2964 | 10.93 | | RP | POS | | | 916613 | 234913 | 877165 | 274733 | 2.63E-01 | -0.15 | 0.86 |
| RP_POS_668_46686a9_92 | | 668.4669 | 9.92 | | RP | POS | | | 3223458 | 1526048 | 2976945 | 1677090 | 2.63E-01 | -0.15 | 1.07 |
| RP_POS_414_3002a8_249 | | 414.3002 | 8.25 | | RP | POS | | | 1923078 | 2336987 | 1638717 | 1312997 | 2.63E-01 | -0.16 | 0.9 |
| RP_NEG_134_06116a3_535 | | 134.0612 | 3.54 | | RP | NEG | | | 319811 | 299298 | 376805 | 424671 | 2.64E-01 | 0.16 | 0.83 |
| RP_POS_433_17392a12_372 | | 433.1739 | 12.37 | | RP | POS | | | 342204 | 136699 | 367706 | 188039 | 2.64E-01 | 0.16 | 0.94 |
| HILIC_POS_122_92461a2_982 | | 122.9246 | 2.98 | | HILIC | POS | | | 3.3E+07 | 4939019 | 3.4E+07 | 4765604 | 2.64E-01 | 0.15 | 0.73 |
| RP_NEG_406_1459a10_717 | | 406.1459 | 10.72 | | RP | NEG | | | 139001 | 83662.6 | 151585 | 81095 | 2.64E-01 | 0.15 | 1.06 |
| RP_POS_1054_68872a10_607 | | 1054.6887 | 10.61 | | RP | POS | | | 1170689 | 676285 | 1276964 | 709818 | 2.64E-01 | 0.15 | 0.88 |
| RP_POS_174_97585a15_408 | | 174.9759 | 15.41 | | RP | POS | | | 5.2E+07 | 1.1E+07 | 5.4E+07 | 1.3E+07 | 2.64E-01 | 0.15 | 0.97 |
| RP_POS_675_47638a10_089 | | 675.4764 | 10.09 | | RP | POS | | | 4408038 | 2009759 | 4738510 | 2276681 | 2.64E-01 | 0.15 | 0.79 |
| RP_POS_271_11554a4_06 | 7-prenyl-theophylline | 271.1155 | 4.06 | | RP | POS | 271.1162(100) | 2 | 957964 | 689659 | 879834 | 280701 | 2.64E-01 | -0.16 | 0.85 |
| HILIC_POS_191_07661a7_099 | | 191.0766 | 7.10 | | HILIC | POS | 191.0768(100) | | 421880 | 69227.1 | 411203 | 70327.5 | 2.64E-01 | -0.15 | 0.72 |
| RP_NEG_372_10513a0_68 | | 372.1051 | 0.68 | | RP | NEG | | | 384763 | 153117 | 363105 | 130950 | 2.64E-01 | -0.15 | 0.84 |
| RP_NEG_915_1275a9_347 | | 915.1275 | 9.35 | | RP | NEG | | | 3216356 | 348455 | 3165672 | 315715 | 2.64E-01 | -0.15 | 1.11 |
| RP_POS_817_06635a13_774 | | 817.0664 | 13.77 | | RP | POS | | | 592791 | 410883 | 536588 | 325632 | 2.64E-01 | -0.15 | 0.91 |
| RP_NEG_381_15558a7_967 | | 381.1556 | 7.97 | | RP | NEG | | | 890364 | 4274615 | 436337 | 861234 | 2.64E-01 | -0.18 | 0.63 |
| HILIC_NEG_264_07257a5_117 | | 264.0726 | 5.12 | | HILIC | NEG | | | 3503130 | 1391186 | 3736095 | 1631061 | 2.65E-01 | 0.15 | 0.88 |
| RP_NEG_755_13916a9_482 | | 755.1392 | 9.48 | | RP | NEG | | | 2174705 | 160110 | 2198113 | 147561 | 2.65E-01 | 0.15 | 0.73 |
| RP_POS_294_8765a15_437 | | 294.8765 | 15.44 | | RP | POS | | | 3528941 | 2723591 | 3990023 | 3252048 | 2.65E-01 | 0.15 | 0.69 |
| RP_NEG_1021_21814a9_736 | | 1021.2181 | 9.74 | | RP | NEG | | | 5880661 | 552700 | 5803286 | 465040 | 2.65E-01 | -0.15 | 1.05 |
| RP_NEG_378_98206a9_038 | | 378.9821 | 9.04 | | RP | NEG | | | 851918 | 109915 | 836034 | 98709.1 | 2.65E-01 | -0.15 | 0.8 |
| RP_POS_352_2453a11_596 | | 352.2453 | 11.60 | | RP | POS | | | 2534144 | 575209 | 2442424 | 622158 | 2.65E-01 | -0.15 | 0.76 |
| RP_POS_692_48267a11_307 | | 692.4827 | 11.31 | | RP | POS | | | 222453 | 95464.4 | 239784 | 127327 | 2.66E-01 | 0.16 | 1.03 |
| RP_NEG_326_75458a15_354 | | 326.7546 | 15.35 | | RP | NEG | | | 4937297 | 312505 | 4982549 | 283033 | 2.66E-01 | 0.15 | 0.96 |
| RP_POS_419_0401a0_585 | | 419.0401 | 0.59 | | RP | POS | | | 664896 | 258600 | 705449 | 272489 | 2.66E-01 | 0.15 | 0.76 |
| RP_POS_812_50897a10_927 | | 812.5090 | 10.93 | | RP | POS | | | 280920 | 189623 | 309689 | 188172 | 2.66E-01 | 0.15 | 0.82 |
| RP_NEG_326_08838a0_706 | Acetaminophen glucuronide | 326.0884 | 0.71 | | RP | NEG | 150.0567(100), 113.0233(80), 85.0283(60) | 2 | 2388588 | 1E+07 | 4573466 | 1.7E+07 | 2.66E-01 | 0.16 | 0.96 |
| HILIC_NEG_183_93459a3_982 | | 183.9346 | 3.98 | | HILIC | NEG | | | 2985897 | 978691 | 2838007 | 964482 | 2.66E-01 | -0.15 | 0.76 |
| HILIC_POS_177_08667a6_327 | | 177.0867 | 6.33 | | HILIC | POS | | | 1370622 | 430849 | 1307938 | 394484 | 2.66E-01 | -0.15 | 1.08 |
| RP_NEG_203_00217a3_263 | | 203.0022 | 3.26 | | RP | NEG | | | 2046745 | 1962917 | 1756746 | 1852535 | 2.66E-01 | -0.15 | 0.63 |
| RP_POS_341_30487a10_625 | | 341.3049 | 10.63 | | RP | POS | | | 1731145 | 295068 | 1687939 | 273184 | 2.66E-01 | -0.15 | 1.27 |
| RP_POS_479_33804a8_642 | | 479.3380 | 8.64 | | RP | POS | | | 542027 | 319700 | 494725 | 303060 | 2.66E-01 | -0.15 | 0.89 |
| RP_POS_766_53369a12_98 | | 766.5337 | 12.98 | | RP | POS | | | 6960125 | 6590203 | 6020327 | 5792686 | 2.66E-01 | -0.15 | 0.71 |
| HILIC_NEG_144_92355a0_579 | | 144.9236 | 0.58 | | HILIC | NEG | | | 3457182 | 1809156 | 3717665 | 1629563 | 2.67E-01 | 0.15 | 0.72 |
| HILIC_POS_502_29327a1_725 | | 502.2933 | 1.73 | | HILIC | POS | | | 887195 | 334744 | 946435 | 430345 | 2.67E-01 | 0.15 | 0.88 |
| RP_NEG_790_56158a13_386 | | 790.5616 | 13.39 | | RP | NEG | | | 2818690 | 916129 | 2961899 | 962665 | 2.67E-01 | 0.15 | 0.88 |
| RP_NEG_999_64349a10_599 | | 999.6435 | 10.60 | | RP | NEG | | | 1021964 | 465265 | 1098690 | 535865 | 2.67E-01 | 0.15 | 0.84 |
| RP_POS_371_02048a15_492 | | 371.0205 | 15.49 | | RP | POS | | | 529306 | 103948 | 546457 | 120019 | 2.67E-01 | 0.15 | 0.99 |
| HILIC_POS_156_07681a6_784 | L-Histidine | 156.0768 | 6.78 | | HILIC | POS | 110.0714(100), 156.0766(30) | 1 | 1.1E+08 | 2.9E+07 | 1.1E+08 | 1.9E+07 | 2.67E-01 | -0.15 | 0.73 |
| HILIC_POS_140_06825a7_092 | | 140.0683 | 7.09 | | HILIC | POS | 140.069(100), 74.0974(40) | | 424561 | 63758.6 | 415193 | 59746.2 | 2.67E-01 | -0.15 | 0.94 |
| HILIC_POS_106_08676a5_919 | | 106.0868 | 5.92 | | HILIC | POS | 88.0761(100), 70.066(60) | | 1.5E+07 | 7085381 | 1.4E+07 | 5800068 | 2.67E-01 | -0.15 | 0.85 |
| HILIC_POS_513_81677a0_935 | | 513.8168 | 0.94 | | HILIC | POS | | | 254589 | 70594.2 | 243977 | 69051.1 | 2.67E-01 | -0.15 | 0.66 |
| RP_NEG_341_6828a15_496 | | 341.6828 | 15.50 | | RP | NEG | | | 930521 | 62465.4 | 920621 | 67289.5 | 2.67E-01 | -0.15 | 0.99 |
| RP_POS_290_15976a3_986 | | 290.1598 | 3.99 | | RP | POS | | | 356411 | 182558 | 331146 | 151296 | 2.67E-01 | -0.15 | 0.79 |
| RP_POS_333_0842a8_845 | | 333.0842 | 8.85 | | RP | POS | | | 1426161 | 666606 | 1328586 | 621157 | 2.67E-01 | -0.15 | 1.18 |
| RP_POS_383_16742a8_076 | | 383.1674 | 8.08 | | RP | POS | | | 380101 | 137469 | 360448 | 121744 | 2.67E-01 | -0.15 | 0.95 |
| RP_POS_385_21045a11_141 | | 385.2105 | 11.14 | | RP | POS | | | 350924 | 85560.9 | 338100 | 83395.6 | 2.67E-01 | -0.15 | 1.02 |
| HILIC_POS_236_07179a4_386 | | 236.0718 | 4.39 | | HILIC | POS | | | 1034791 | 2794777 | 1638938 | 4762158 | 2.68E-01 | 0.16 | 0.8 |
| RP_POS_425_26599a10_642 | | 425.2660 | 10.64 | | RP | POS | | | 459979 | 460112 | 550986 | 697030 | 2.68E-01 | 0.16 | 1.04 |
| HILIC_POS_140_06834a5_158 | | 140.0683 | 5.16 | | HILIC | POS | 140.069(100), 74.0974(40) | | 6120351 | 1920483 | 6440733 | 2259789 | 2.68E-01 | 0.15 | 0.9 |
| HILIC_POS_295_06668a1_378 | | 295.0667 | 1.38 | | HILIC | POS | | | 1576026 | 1044013 | 1732037 | 1015513 | 2.68E-01 | 0.15 | 0.75 |
| RP_NEG_755_1391a9_278 | | 755.1391 | 9.28 | | RP | NEG | | | 2033522 | 158955 | 2057892 | 162237 | 2.68E-01 | 0.15 | 1.08 |
| RP_POS_171_10146a9_21 | | 171.1015 | 9.21 | | RP | POS | | | 6.8E+07 | 3.7E+07 | 6.3E+07 | 3.7E+07 | 2.68E-01 | -0.15 | 0.81 |
| RP_POS_355_284a12_429 | | 355.2840 | 12.43 | | RP | POS | | | 1084407 | 841551 | 971991 | 643356 | 2.68E-01 | -0.15 | 0.76 |
| RP_POS_557_41168a11_315 | | 557.4117 | 11.32 | | RP | POS | | | 252258 | 263032 | 220063 | 156611 | 2.68E-01 | -0.15 | 0.71 |
| RP_POS_192_13832a7_327 | | 192.1383 | 7.33 | | RP | POS | | | 805357 | 170674 | 883619 | 682662 | 2.69E-01 | 0.18 | 0.74 |
| RP_POS_221_06969a15_372 | | 221.0697 | 15.37 | | RP | POS | | | 3464380 | 1347139 | 3668756 | 1354268 | 2.69E-01 | 0.15 | 0.68 |
| RP_POS_718_53784a12_711 | | 718.5378 | 12.71 | | RP | POS | | | 1662444 | 1302968 | 1894680 | 1702367 | 2.69E-01 | 0.15 | 0.86 |
| RP_NEG_480_82962a15_399 | | 480.8296 | 15.40 | | RP | NEG | | | 795449 | 148078 | 773511 | 142288 | 2.69E-01 | -0.15 | 0.64 |
| RP_POS_227_12733a4_945 | | 227.1273 | 4.95 | | RP | POS | | | 258636 | 119831 | 240181 | 123823 | 2.69E-01 | -0.15 | 0.99 |
| RP_POS_293_20862a10_269 | | 293.2086 | 10.27 | | RP | POS | | | 1368365 | 173991 | 1342769 | 164692 | 2.69E-01 | -0.15 | 1.16 |
| RP_POS_507_32507a10_488 | | 507.3251 | 10.49 | | RP | POS | | | 5698754 | 697744 | 5588406 | 754177 | 2.69E-01 | -0.15 | 1.22 |
| RP_POS_802_5354a12_742 | | 802.5354 | 12.74 | | RP | POS | | | 8.9E+07 | 4.1E+07 | 8.4E+07 | 3E+07 | 2.69E-01 | -0.15 | 0.65 |
| RP_NEG_294_61957a8_812 | | 294.6196 | 8.81 | | RP | NEG | | | 218744 | 602748 | 153692 | 181746 | 2.69E-01 | -0.17 | 0.75 |
| HILIC_POS_192_15977a1_015 | | 192.1598 | 1.02 | | HILIC | POS | | | 6774660 | 1380968 | 6986015 | 1412680 | 2.70E-01 | 0.15 | 1.06 |
| RP_POS_279_19327a8_751 | | 279.1933 | 8.75 | | RP | POS | 279.1942(100) | | 732788 | 287833 | 698873 | 151299 | 2.70E-01 | -0.15 | 0.73 |
| RP_NEG_787_10187a9_747 | | 787.1019 | 9.75 | | RP | NEG | | | 2416056 | 115849 | 2399281 | 106859 | 2.70E-01 | -0.15 | 1.04 |
| RP_NEG_892_53296a12_74 | | 892.5330 | 12.74 | | RP | NEG | | | 5539388 | 2135603 | 5246703 | 1751171 | 2.70E-01 | -0.15 | 0.7 |
| RP_POS_183_1019a4_179 | | 183.1019 | 4.18 | | RP | POS | | | 531789 | 280458 | 490693 | 264604 | 2.70E-01 | -0.15 | 0.9 |
| RP_POS_213_10999a6_5 | | 213.1100 | 6.50 | | RP | POS | | | 3.6E+07 | 5228627 | 3.5E+07 | 4324155 | 2.70E-01 | -0.15 | 1.23 |
| RP_POS_241_14359a7_174 | | 241.1436 | 7.17 | | RP | POS | | | 894212 | 405191 | 835647 | 371966 | 2.70E-01 | -0.15 | 1.01 |
| RP_POS_347_17044a7_071 | | 347.1704 | 7.07 | | RP | POS | | | 1522404 | 170269 | 1498961 | 140985 | 2.70E-01 | -0.15 | 1.09 |
| RP_POS_268_14014a0_681 | | 268.1401 | 0.68 | | RP | POS | | | 2278615 | 948306 | 2427275 | 1015545 | 2.71E-01 | 0.15 | 0.78 |
| RP_POS_351_25037a10_66 | | 351.2504 | 10.66 | | RP | POS | | | 1301730 | 879171 | 1450774 | 1071799 | 2.71E-01 | 0.15 | 0.81 |
| RP_POS_745_12067a11_383 | | 745.1207 | 11.38 | | RP | POS | | | 169556 | 88858.4 | 184972 | 112221 | 2.71E-01 | 0.15 | 0.9 |
| RP_POS_303_22986a10_963 | Linoleic acid | 303.2299 | 10.96 | | RP | POS | 303.2293(100) | 2 | 8187852 | 4593042 | 8856803 | 4310233 | 2.71E-01 | 0.15 | 1.1 |
| HILIC_POS_368_21994a2_956 | | 368.2199 | 2.96 | | HILIC | POS | | | 370948 | 134933 | 351918 | 118400 | 2.71E-01 | -0.15 | 1 |
| RP_POS_262_16461a3_647 | | 262.1646 | 3.65 | | RP | POS | | | 127625 | 65845.8 | 118202 | 59616.1 | 2.71E-01 | -0.15 | 1.22 |
| RP_POS_372_25919a5_177 | | 372.2592 | 5.18 | | RP | POS | | | 739548 | 436846 | 680428 | 350557 | 2.71E-01 | -0.15 | 0.84 |

**Supplemantary table 1**: The non-targeted metabolomics results ordered by the VIP value (p value < 0.0003 are shaded dark green, p value < 0.05 are shaded light green, d values below 0 are blue and above 0 are red)

| | | | | | | | | | Cases | | Controls | | Welch test | Cohen's | |
|---|---|---|---|---|---|---|---|---|---|---|---|---|---|---|---|
| Feature_ID | Metabolite_name | Average_Mz | Average_RT_min_ | HMDB ID | RP_HILIC | POS_NEG | Cureted_MSMS | ID_level | Mean | SD | Mean2 | SD3 | p | d | VIP |
| RP_POS_727_50659a11_414 | | 727.5066 | 11.41 | | RP | POS | | | 202783 | 131355 | 186470 | 83817.1 | 2.71E-01 | -0.15 | 1 |
| RP_POS_466_29163a10_413 | | 466.2916 | 10.41 | | RP | POS | | | 232926 | 127688 | 298612 | 580484 | 2.72E-01 | 0.19 | 0.81 |
| HILIC_NEG_84_0087a5_117 | | 84.0087 | 5.12 | | HILIC | NEG | | | 2530275 | 1061974 | 2714074 | 1343332 | 2.72E-01 | 0.15 | 0.82 |
| RP_POS_120_97929a15_413 | | 120.9793 | 15.41 | | RP | POS | | | 3278322 | 1269853 | 3484896 | 1454800 | 2.72E-01 | 0.15 | 0.68 |
| RP_POS_266_01123a2_159 | | 266.0112 | 2.16 | | RP | POS | | | 398660 | 92826.1 | 412383 | 89783 | 2.72E-01 | 0.15 | 0.78 |
| RP_POS_647_42377a12_076 | | 647.4238 | 12.08 | | RP | POS | | | 124026 | 45739.4 | 131571 | 53564.9 | 2.72E-01 | 0.15 | 0.7 |
| RP_POS_702_15411a11_355 | | 702.1541 | 11.36 | | RP | POS | | | 326825 | 137947 | 350062 | 166900 | 2.72E-01 | 0.15 | 0.71 |
| RP_POS_280_99545a0_547 | | 280.9955 | 0.55 | | RP | POS | | | 3769288 | 486853 | 3872202 | 816022 | 2.73E-01 | 0.16 | 0.73 |
| RP_NEG_397_23627a11_014 | | 397.2363 | 11.01 | | RP | NEG | | | 260567 | 175975 | 286505 | 170402 | 2.73E-01 | 0.15 | 1.12 |
| RP_NEG_455_14719a11_592 | | 455.1472 | 11.59 | | RP | NEG | | | 1232070 | 72499.5 | 1219709 | 89693.1 | 2.73E-01 | -0.15 | 0.95 |
| RP_POS_131_95061a15_603 | | 131.9506 | 15.60 | | RP | POS | | | 2155289 | 1248961 | 1985721 | 1018124 | 2.73E-01 | -0.15 | 0.61 |
| RP_POS_189_1123a9_21 | | 189.1123 | 9.21 | | RP | POS | | | 1.8E+07 | 9765931 | 1.7E+07 | 9824824 | 2.73E-01 | -0.15 | 0.81 |
| RP_POS_299_13412a8_843 | | 299.1341 | 8.84 | | RP | POS | | | 739125 | 409430 | 681457 | 361582 | 2.73E-01 | -0.15 | 1.22 |
| RP_POS_430_74792a13_007 | | 430.7479 | 13.01 | | RP | POS | | | 4258422 | 4261969 | 3721924 | 2869761 | 2.73E-01 | -0.15 | 0.73 |
| RP_POS_442_31927a9_15 | | 442.3193 | 9.15 | | RP | POS | | | 506082 | 258579 | 467532 | 255214 | 2.73E-01 | -0.15 | 0.8 |
| RP_NEG_451_14078a6_26 | | 451.1408 | 6.26 | | RP | NEG | | | 290270 | 255514 | 326917 | 235541 | 2.74E-01 | 0.15 | 0.75 |
| RP_POS_236_91486a15_402 | | 236.9149 | 15.40 | | RP | POS | | | 237913 | 119984 | 259535 | 161983 | 2.74E-01 | 0.15 | 0.77 |
| RP_POS_260_89062a15_406 | | 260.8906 | 15.41 | | RP | POS | | | 2158604 | 612006 | 2253031 | 646531 | 2.74E-01 | 0.15 | 0.74 |
| RP_POS_266_13904a6_693 | | 266.1390 | 6.69 | | RP | POS | | | 860971 | 269770 | 899844 | 250512 | 2.74E-01 | 0.15 | 0.73 |
| RP_POS_300_6329a10_295 | | 300.6329 | 10.30 | | RP | POS | | | 1227661 | 352358 | 1279327 | 339237 | 2.74E-01 | 0.15 | 0.93 |
| RP_NEG_455_24789a9_427 | | 455.2479 | 9.43 | | RP | NEG | | | 610306 | 767819 | 511288 | 555084 | 2.74E-01 | -0.15 | 0.69 |
| RP_POS_750_59991a11_411 | | 750.5999 | 11.41 | | RP | POS | | | 497399 | 93462.9 | 486166 | 54617.4 | 2.74E-01 | -0.15 | 0.86 |
| HILIC_POS_399_23532a2_28 | | 399.2353 | 2.28 | | HILIC | POS | | | 58939.7 | 24834.2 | 70280.7 | 100236 | 2.75E-01 | 0.18 | 0.74 |
| HILIC_POS_128_95078a1_349 | | 128.9508 | 1.35 | | HILIC | POS | | | 321560 | 174913 | 348142 | 180141 | 2.75E-01 | 0.15 | 0.84 |
| RP_POS_405_2998a11_293 | | 405.2998 | 11.29 | | RP | POS | | | 425678 | 238482 | 465574 | 289141 | 2.75E-01 | 0.15 | 1.16 |
| RP_POS_604_79193a15_389 | | 604.7919 | 15.39 | | RP | POS | | | 208355 | 176831 | 237929 | 214102 | 2.75E-01 | 0.15 | 0.89 |
| RP_NEG_297_15338a9_712 | | 297.1534 | 9.71 | | RP | NEG | 297.1525(100), 183.0123(20) | | 2331233 | 1354792 | 2153869 | 1020616 | 2.75E-01 | -0.15 | 0.62 |
| HILIC_POS_553_40631a1_081 | | 553.4063 | 1.08 | | HILIC | POS | | | 161273 | 84107.2 | 150308 | 62802.7 | 2.75E-01 | -0.15 | 0.92 |
| RP_NEG_299_2598a10_192 | | 299.2598 | 10.19 | | RP | NEG | | | 9384000 | 1423254 | 9179366 | 1323521 | 2.75E-01 | -0.15 | 1.15 |
| RP_NEG_659_07782a8_98 | | 659.0778 | 8.98 | | RP | NEG | | | 3050607 | 155388 | 3027645 | 152032 | 2.75E-01 | -0.15 | 1.03 |
| RP_POS_235_85146a9_209 | | 235.8515 | 9.21 | | RP | POS | | | 1120411 | 596894 | 1032961 | 575320 | 2.75E-01 | -0.15 | 1.06 |
| RP_POS_299_10999a6_514 | | 299.1100 | 6.51 | | RP | POS | | | 1438182 | 163123 | 1414816 | 150522 | 2.75E-01 | -0.15 | 1.1 |
| RP_POS_348_9895a8_09 | | 348.9895 | 8.09 | | RP | POS | | | 1170324 | 275142 | 1127856 | 291937 | 2.75E-01 | -0.15 | 0.76 |
| RP_POS_472_828a10_065 | | 472.8280 | 10.07 | | RP | POS | | | 2503279 | 1250971 | 2323103 | 1167757 | 2.75E-01 | -0.15 | 0.94 |
| RP_POS_236_07143a14_468 | | 236.0714 | 14.47 | | RP | POS | | | 1.3E+08 | 6.5E+07 | 1.4E+08 | 6.5E+07 | 2.76E-01 | 0.15 | 0.86 |
| RP_POS_404_30032a10_252 | | 404.3003 | 10.25 | | RP | POS | | | 2414186 | 1012387 | 2564267 | 1000572 | 2.76E-01 | 0.15 | 1.09 |
| RP_POS_529_71484a10_46 | | 529.7148 | 10.46 | | RP | POS | | | 212733 | 118502 | 230711 | 122358 | 2.76E-01 | 0.15 | 0.8 |
| HILIC_NEG_234_81949a5_498 | | 234.8195 | 5.50 | | HILIC | NEG | | | 1089226 | 413587 | 1033187 | 341125 | 2.76E-01 | -0.15 | 0.74 |
| RP_POS_332_99936a8_095 | | 332.9994 | 8.10 | | RP | POS | | | 179472 | 83160.8 | 168820 | 59862.4 | 2.76E-01 | -0.15 | 0.84 |
| HILIC_NEG_195_81087a10_938 | | 195.8109 | 10.94 | | HILIC | NEG | | | 1.6E+07 | 8386055 | 1.7E+07 | 8736800 | 2.77E-01 | 0.15 | 0.67 |
| RP_NEG_262_99231a6_569 | | 262.9923 | 6.57 | | RP | NEG | | | 2787623 | 108825 | 2806806 | 143787 | 2.77E-01 | 0.15 | 0.69 |
| RP_NEG_313_11938a6_796 | | 313.1194 | 6.80 | | RP | NEG | | | 89089.8 | 111817 | 104680 | 98425.2 | 2.77E-01 | 0.15 | 0.68 |
| RP_NEG_522_76794a15_416 | | 522.7679 | 15.42 | | RP | NEG | | | 268565 | 95333.6 | 286408 | 137317 | 2.77E-01 | 0.15 | 0.99 |
| RP_POS_334_15848a10_961 | | 334.1585 | 10.96 | | RP | POS | | | 282594 | 184994 | 309453 | 176253 | 2.77E-01 | 0.15 | 1.05 |
| RP_POS_410_12967a5_909 | | 410.1297 | 5.91 | | RP | POS | | | 67464.2 | 25938.2 | 71753.4 | 31001 | 2.77E-01 | 0.15 | 0.93 |
| HILIC_POS_349_2587a3_131 | | 349.2587 | 3.13 | | HILIC | POS | | | 371711 | 54442.1 | 363533 | 55445.6 | 2.77E-01 | -0.15 | 0.82 |
| RP_NEG_312_9736a6_959 | | 312.9736 | 6.96 | | RP | NEG | | | 482913 | 22423.4 | 479471 | 23698.1 | 2.77E-01 | -0.15 | 0.67 |
| RP_POS_413_18149a11_144 | | 413.1815 | 11.14 | | RP | POS | | | 554101 | 115605 | 538026 | 101314 | 2.77E-01 | -0.15 | 0.76 |
| RP_POS_499_6947a10_52 | | 499.6947 | 10.52 | | RP | POS | | | 290561 | 178645 | 264519 | 171818 | 2.77E-01 | -0.15 | 0.84 |
| RP_POS_557_31598a9_208 | | 557.3160 | 9.21 | | RP | POS | | | 556851 | 600064 | 481880 | 404340 | 2.77E-01 | -0.15 | 0.78 |
| HILIC_NEG_117_91919a5_652 | | 117.9192 | 5.65 | | HILIC | NEG | | | 1148007 | 139840 | 1171138 | 168144 | 2.78E-01 | 0.15 | 0.83 |
| HILIC_NEG_132_90036a2_276 | | 132.9004 | 2.28 | | HILIC | NEG | | | 1988351 | 319225 | 2039021 | 358955 | 2.78E-01 | 0.15 | 0.8 |
| HILIC_NEG_132_92361a5_122 | | 132.9236 | 5.12 | | HILIC | NEG | | | 7067614 | 2850420 | 7482038 | 2736963 | 2.78E-01 | 0.15 | 0.72 |
| RP_NEG_367_24734a10_198 | | 367.2473 | 10.20 | | RP | NEG | | | 745829 | 116024 | 729032 | 110729 | 2.78E-01 | -0.15 | 1.19 |
| RP_POS_574_72406a10_417 | | 574.7241 | 10.42 | | RP | POS | | | 634597 | 319985 | 590001 | 282773 | 2.78E-01 | -0.15 | 0.8 |
| RP_POS_684_64502a11_55 | | 684.6450 | 11.55 | | RP | POS | | | 134307 | 140743 | 118204 | 69927.9 | 2.78E-01 | -0.15 | 0.67 |
| RP_POS_754_99005a10_488 | | 754.9901 | 10.49 | | RP | POS | | | 2164761 | 412830 | 2100149 | 452099 | 2.78E-01 | -0.15 | 0.85 |
| RP_NEG_459_25241a10_938 | | 459.2524 | 10.94 | | RP | NEG | | | 81997.5 | 74786.5 | 94859.5 | 95649.3 | 2.79E-01 | 0.15 | 0.74 |
| RP_NEG_554_78729a15_384 | | 554.7873 | 15.38 | | RP | NEG | | | 700859 | 163071 | 724351 | 154403 | 2.79E-01 | 0.15 | 0.79 |
| RP_POS_480_30893a10_48 | | 480.3089 | 10.48 | | RP | POS | | | 3149844 | 1115440 | 3332627 | 1325222 | 2.79E-01 | 0.15 | 0.76 |
| RP_POS_519_34906a9_135 | | 519.3491 | 9.14 | | RP | POS | | | 252939 | 183293 | 227316 | 163601 | 2.79E-01 | -0.15 | 1 |
| RP_POS_531_40839a14_361 | | 531.4084 | 14.36 | | RP | POS | | | 2755043 | 1590482 | 2536473 | 1367059 | 2.79E-01 | -0.15 | 0.84 |
| RP_NEG_621_30475a11_224 | | 621.3048 | 11.22 | | RP | NEG | | | 167325 | 153734 | 188202 | 129300 | 2.80E-01 | 0.15 | 0.75 |
| RP_POS_582_31335a7_071 | | 582.3134 | 7.07 | | RP | POS | | | 513475 | 60609.7 | 522926 | 66619.9 | 2.80E-01 | 0.15 | 0.86 |
| RP_POS_738_20386a9_484 | | 738.2039 | 9.48 | | RP | POS | | | 955452 | 119273 | 973913 | 129434 | 2.80E-01 | 0.15 | 0.99 |
| RP_POS_140_06842a11_144 | | 140.0684 | 11.14 | | RP | POS | | | 783432 | 81926.4 | 770216 | 95086.6 | 2.80E-01 | -0.15 | 0.65 |
| RP_POS_299_2579a10_638 | | 299.2579 | 10.64 | | RP | POS | | | 2754473 | 374382 | 2700881 | 351484 | 2.80E-01 | -0.15 | 0.88 |
| RP_POS_356_19217a4_684 | | 356.1922 | 4.68 | | RP | POS | | | 136508 | 60080.4 | 128088 | 54143.1 | 2.80E-01 | -0.15 | 0.72 |
| RP_POS_689_49121a10_401 | | 689.4912 | 10.40 | | RP | POS | | | 3607571 | 1495946 | 3391121 | 1434299 | 2.80E-01 | -0.15 | 0.8 |
| RP_POS_392_27921a8_259 | | 392.2792 | 8.26 | | RP | POS | | | 418936 | 309252 | 466316 | 330393 | 2.81E-01 | 0.15 | 0.79 |
| RP_POS_726_50208a11_384 | | 726.5021 | 11.38 | | RP | POS | | | 167461 | 73408.1 | 179903 | 92373.2 | 2.81E-01 | 0.15 | 0.73 |
| RP_POS_784_58582a13_383 | | 784.5858 | 13.38 | | RP | POS | | | 3.8E+08 | 1.7E+08 | 4.1E+08 | 2.2E+08 | 2.81E-01 | 0.15 | 0.76 |
| HILIC_POS_349_01718a7_483 | | 349.0172 | 7.48 | | HILIC | POS | | | 1968597 | 82938.1 | 1955425 | 94080.9 | 2.81E-01 | -0.15 | 0.78 |
| RP_POS_532_36926a6_818 | | 532.3693 | 6.82 | | RP | POS | | | 587611 | 429752 | 528368 | 376243 | 2.81E-01 | -0.15 | 0.72 |
| HILIC_NEG_87_00847a5_115 | | 87.0085 | 5.12 | | HILIC | NEG | | | 365234 | 177498 | 396466 | 237112 | 2.82E-01 | 0.15 | 0.69 |
| RP_POS_349_23715a10_513 | | 349.2372 | 10.51 | | RP | POS | | | 7281664 | 3502614 | 7800306 | 3541158 | 2.82E-01 | 0.15 | 1.16 |
| RP_NEG_151_02628a0_681 | Xanthine | 151.0263 | 0.68 | | RP | NEG | 151.0256(100), 108.0203(30) | 2 | 1564231 | 365442 | 1621545 | 408598 | 2.82E-01 | 0.15 | 1.05 |
| RP_POS_299_25839a10_159 | | 299.2584 | 10.16 | | RP | POS | | | 1.4E+07 | 2010894 | 1.4E+07 | 2007491 | 2.82E-01 | -0.15 | 1.08 |
| RP_NEG_253_78468a15_493 | | 253.7847 | 15.49 | | RP | NEG | | | 287142 | 49094 | 294651 | 52783.8 | 2.83E-01 | 0.15 | 0.67 |
| RP_POS_809_98938a10_609 | | 809.9894 | 10.61 | | RP | POS | | | 769264 | 396183 | 828639 | 411065 | 2.83E-01 | 0.15 | 0.88 |
| HILIC_POS_211_10509a5_678 | | 211.1051 | 5.68 | | HILIC | POS | | | 241023 | 131386 | 223051 | 114089 | 2.83E-01 | -0.15 | 0.95 |
| HILIC_POS_310_20099a0_838 | | 310.2010 | 0.84 | | HILIC | POS | | | 5032312 | 2082812 | 4713082 | 2243913 | 2.83E-01 | -0.15 | 0.61 |
| RP_NEG_226_99532a15_481 | | 226.9953 | 15.48 | | RP | NEG | | | 2110537 | 522035 | 2039918 | 442347 | 2.83E-01 | -0.15 | 0.69 |
| RP_NEG_339_1994a13_449 | | 339.1994 | 13.45 | | RP | NEG | | | 657852 | 384234 | 603137 | 361886 | 2.83E-01 | -0.15 | 1.01 |
| RP_POS_413_32336a10_93 | | 413.3234 | 10.93 | | RP | POS | | | 4575678 | 817220 | 4462245 | 731013 | 2.83E-01 | -0.15 | 1.11 |
| RP_POS_193_09491a2_811 | | 193.0949 | 2.81 | | RP | POS | | | 3434058 | 8817651 | 2519268 | 2341402 | 2.83E-01 | -0.16 | 0.68 |
| RP_POS_728_55878a14_393 | | 728.5588 | 14.39 | | RP | POS | | | 420476 | 409860 | 513329 | 771798 | 2.84E-01 | 0.16 | 0.95 |
| HILIC_POS_124_92272a2_982 | | 124.9227 | 2.98 | | HILIC | POS | | | 4738019 | 745854 | 4845343 | 718798 | 2.84E-01 | 0.15 | 0.71 |
| RP_POS_344_22763a3_793 | | 344.2276 | 3.79 | | RP | POS | | | 475169 | 233725 | 515661 | 307214 | 2.84E-01 | 0.15 | 0.77 |
| HILIC_POS_170_11749a3_108 | | 170.1175 | 3.11 | | HILIC | POS | | | 395100 | 171953 | 372156 | 141883 | 2.84E-01 | -0.15 | 0.69 |
| RP_NEG_636_26202a10_312 | | 636.2620 | 10.31 | | RP | NEG | | | 2739407 | 518946 | 2662704 | 526460 | 2.84E-01 | -0.15 | 0.91 |
| RP_POS_242_21138a11_026 | | 242.2114 | 11.03 | | RP | POS | | | 1061362 | 1689045 | 848466 | 1216090 | 2.84E-01 | -0.15 | 0.75 |
| RP_POS_386_98215a15_449 | | 386.9822 | 15.45 | | RP | POS | | | 403483 | 101269 | 387666 | 113204 | 2.84E-01 | -0.15 | 0.83 |
| RP_POS_445_39978a11_112 | | 445.3998 | 11.11 | | RP | POS | | | 185483 | 295675 | 149496 | 192525 | 2.84E-01 | -0.15 | 0.7 |
| RP_POS_497_39554a11_591 | | 497.3955 | 11.59 | | RP | POS | | | 686113 | 178367 | 657375 | 209350 | 2.84E-01 | -0.15 | 0.85 |
| RP_POS_657_47852a9_702 | | 657.4785 | 9.70 | | RP | POS | | | 807810 | 589754 | 729159 | 485271 | 2.84E-01 | -0.15 | 0.81 |
| RP_POS_382_20383a10_834 | | 382.2038 | 10.83 | | RP | POS | | | 9.6E+08 | 3.4E+09 | 3.4E+09 | 2.3E+10 | 2.85E-01 | 0.19 | 0.81 |
| HILIC_POS_168_12289a0_623 | | 168.1229 | 0.62 | | HILIC | POS | | | 869316 | 171323 | 897829 | 212247 | 2.85E-01 | 0.15 | 0.77 |
| HILIC_POS_476_30664a0_61 | | 476.3066 | 0.61 | | HILIC | POS | | | 83727.8 | 57397.4 | 95055.4 | 90830.4 | 2.85E-01 | 0.15 | 0.67 |
| RP_POS_296_25833a10_716 | | 296.2583 | 10.72 | | RP | POS | | | 1809748 | 1278687 | 1998908 | 1304588 | 2.85E-01 | 0.15 | 0.95 |
| RP_POS_547_35529a10_928 | | 547.3553 | 10.93 | | RP | POS | | | 5.8E+07 | 9280436 | 5.9E+07 | 1.1E+07 | 2.85E-01 | 0.15 | 0.9 |
| RP_POS_636_44489a11_278 | | 636.4449 | 11.28 | | RP | POS | | | 269381 | 130220 | 291785 | 169692 | 2.85E-01 | 0.15 | 0.69 |
| RP_NEG_351_25223a11_592 | | 351.2522 | 11.59 | | RP | NEG | | | 4303960 | 1159046 | 4137689 | 1117074 | 2.85E-01 | -0.15 | 1.34 |
| RP_NEG_379_28357a12_106 | | 379.2836 | 12.11 | | RP | NEG | | | 377719 | 124380 | 358396 | 138052 | 2.85E-01 | -0.15 | 1.34 |
| RP_NEG_960_52039a12_737 | | 960.5204 | 12.74 | | RP | NEG | | | 2014614 | 671872 | 1923692 | 576096 | 2.85E-01 | -0.15 | 0.67 |
| RP_POS_122_92978a15_489 | | 122.9298 | 15.49 | | RP | POS | | | 1022956 | 94728.3 | 1009688 | 87225.3 | 2.85E-01 | -0.15 | 1.14 |
| RP_POS_335_25604a10_395 | | 335.2560 | 10.40 | | RP | POS | | | 927591 | 125576 | 908254 | 137422 | 2.85E-01 | -0.15 | 0.98 |
| RP_POS_386_27432a5_779 | | 386.2743 | 5.78 | | RP | POS | | | 1285356 | 723767 | 1191454 | 561797 | 2.85E-01 | -0.15 | 0.79 |
| RP_POS_644_49408a10_541 | | 644.4941 | 10.54 | | RP | POS | | | 1384618 | 292230 | 1342404 | 284746 | 2.85E-01 | -0.15 | 0.84 |
| RP_POS_785_65259a14_706 | | 785.6526 | 14.71 | | RP | POS | | | 1888723 | 1575420 | 1676426 | 1337808 | 2.85E-01 | -0.15 | 0.79 |
| HILIC_POS_106_08658a4_719 | | 106.0866 | 4.72 | | HILIC | POS | 88.0761(100), 70.066(60) | | 8198952 | 5698414 | 9020586 | 5557016 | 2.86E-01 | 0.15 | 1.19 |
| HILIC_NEG_118_92409a1_179 | | 118.9241 | 1.18 | | HILIC | NEG | | | 863495 | 911421 | 1006884 | 1037214 | 2.86E-01 | 0.15 | 0.65 |
| RP_NEG_1019_61188a10_299 | | 1019.6119 | 10.30 | | RP | NEG | | | 1408659 | 470966 | 1483222 | 540803 | 2.86E-01 | 0.15 | 0.99 |
| RP_NEG_413_20062a8_371 | | 413.2006 | 8.37 | | RP | NEG | | | 408787 | 468751 | 471842 | 397542 | 2.86E-01 | 0.15 | 0.79 |
| HILIC_NEG_130_05093a1_584 | N-Acetyl-L-alanine | 130.0509 | 1.58 | | HILIC | NEG | 88.04(100), 130.0415(5) | 1 | 617598 | 93291.2 | 606102 | 63613.6 | 2.86E-01 | -0.15 | 0.76 |
| HILIC_POS_371_2399a1_141 | | 371.2399 | 1.14 | | HILIC | POS | | | 3493702 | 1214511 | 3302904 | 1377154 | 2.86E-01 | -0.15 | 0.74 |
| RP_POS_417_30576a11_592 | | 417.3058 | 11.59 | | RP | POS | | | 651108 | 151406 | 627478 | 169919 | 2.86E-01 | -0.15 | 0.7 |
| RP_NEG_474_23157a10_728 | | 474.2316 | 10.73 | | RP | NEG | | | 340394 | 390423 | 423288 | 683563 | 2.87E-01 | 0.15 | 0.69 |
| RP_NEG_638_27594a10_61 | | 638.2759 | 10.61 | | RP | NEG | | | 3027597 | 453009 | 3096107 | 484041 | 2.87E-01 | 0.15 | 0.94 |
| RP_POS_492_44067a13_175 | | 492.4407 | 13.18 | | RP | POS | | | 5263332 | 945193 | 5413360 | 1093132 | 2.87E-01 | 0.15 | 0.65 |
| RP_POS_529_46136a11_826 | | 529.4614 | 11.83 | | RP | POS | | | 530766 | 421944 | 592745 | 427222 | 2.87E-01 | 0.15 | 0.89 |
| RP_POS_568_19244a11_017 | | 568.1924 | 11.02 | | RP | POS | | | 1060591 | 211974 | 1093792 | 240131 | 2.87E-01 | 0.15 | 0.77 |
| RP_POS_621_76935a10_504 | | 621.7694 | 10.50 | | RP | POS | | | 241495 | 164579 | 269562 | 213475 | 2.87E-01 | 0.15 | 0.91 |
| HILIC_NEG_154_06223a6_776 | Histidine | 154.0622 | 6.78 | | HILIC | NEG | 154.0616(100), 930.459(50) | 2 | 4307655 | 909971 | 4188987 | 724930 | 2.87E-01 | -0.15 | 0.82 |
| RP_POS_228_19579a11_025 | | 228.1958 | 11.03 | | RP | POS | 95.0862(100), 175.1479(50) | | 4354724 | 6722893 | 3515551 | 4792894 | 2.87E-01 | -0.15 | 0.73 |
| RP_NEG_1107_16516a9_736 | | 1107.1652 | 9.74 | | RP | NEG | | | 4252783 | 314459 | 4211862 | 248428 | 2.87E-01 | -0.15 | 1.13 |
| RP_NEG_852_57715a13_438 | | 852.5772 | 13.44 | | RP | NEG | | | 1.7E+07 | 4979692 | 1.6E+07 | 3788786 | 2.87E-01 | -0.15 | 0.81 |
| RP_POS_196_0482a0_723 | | 196.0482 | 0.72 | | RP | POS | | | 697003 | 327511 | 651247 | 302477 | 2.87E-01 | -0.15 | 0.89 |
| RP_POS_230_17477a6_026 | | 230.1748 | 6.03 | | RP | POS | | | 360675 | 95439.9 | 347604 | 84335.7 | 2.87E-01 | -0.15 | 0.81 |
| RP_POS_343_33875a11_144 | | 343.3388 | 11.14 | | RP | POS | | | 3.6E+07 | 7757249 | 3.5E+07 | 7805575 | 2.87E-01 | -0.15 | 0.92 |
| RP_POS_349_09482a9_209 | | 349.0948 | 9.21 | | RP | POS | | | 1197127 | 556411 | 1117178 | 541040 | 2.87E-01 | -0.15 | 0.88 |
| RP_POS_596_92145a11_539 | | 596.9215 | 11.54 | | RP | POS | | | 134406 | 146745 | 117641 | 78287.8 | 2.87E-01 | -0.15 | 0.71 |
| RP_POS_614_39374a10_132 | | 614.3937 | 10.13 | | RP | POS | | | 369083 | 225807 | 339102 | 187216 | 2.87E-01 | -0.15 | 0.96 |
| RP_POS_830_55688a13_006 | | 830.5569 | 13.01 | | RP | POS | | | 2.6E+07 | 1.8E+07 | 2.3E+07 | 1.6E+07 | 2.87E-01 | -0.15 | 0.74 |
| HILIC_NEG_236_81747a5_877 | | 236.8175 | 5.88 | | HILIC | NEG | | | 332682 | 133989 | 352889 | 142768 | 2.88E-01 | 0.15 | 0.62 |
| RP_POS_313_14044a0_683 | | 313.1404 | 0.68 | | RP | POS | | | 294264 | 165055 | 320097 | 187310 | 2.88E-01 | 0.15 | 0.74 |
| RP_POS_527_87793a10 | | 527.8779 | 10.00 | | RP | POS | | | 3209457 | 1413761 | 3424138 | 1526068 | 2.88E-01 | 0.15 | 1.05 |
| HILIC_NEG_234_82028a6_735 | | 234.8203 | 6.74 | | HILIC | NEG | | | 650694 | 87971.5 | 662665 | 77247.4 | 2.88E-01 | 0.14 | 0.8 |
| HILIC_NEG_218_10361a1_025 | | 218.1036 | 1.03 | | HILIC | NEG | | | 124738 | 49101.7 | 118029 | 43477.2 | 2.88E-01 | -0.14 | 0.69 |
| RP_POS_259_19025a7_972 | | 259.1903 | 7.97 | | RP | POS | | | 721746 | 79343.6 | 710747 | 72437.9 | 2.88E-01 | -0.14 | 1.1 |
| RP_POS_636_5575a15_049 | | 636.5575 | 15.05 | | RP | POS | | | 6197025 | 2741738 | 5813971 | 2542113 | 2.88E-01 | -0.14 | 0.86 |
| RP_NEG_209_08197a7_207 | | 209.0820 | 7.21 | | RP | NEG | | | 368505 | 24324.8 | 365287 | 20031.5 | 2.88E-01 | -0.15 | 0.68 |
| RP_POS_277_14145a7_009 | | 277.1415 | 7.01 | | RP | POS | | | 203759 | 154176 | 184335 | 113413 | 2.88E-01 | -0.15 | 0.86 |
| RP_POS_314_23236a7_693 | | 314.2324 | 7.69 | | RP | POS | | | 235779 | 86112.2 | 223135 | 87543.8 | 2.88E-01 | -0.15 | 1.22 |
| RP_NEG_339_32721a12_739 | Behenic acid | 339.3272 | 12.74 | | RP | NEG | 339.3662(100) | 2 | 665283 | 279603 | 625023 | 275886 | 2.89E-01 | -0.14 | 1.33 |
| HILIC_NEG_206_97307a5_469 | | 206.9731 | 5.47 | | HILIC | NEG | | | 327108 | 36022.6 | 332662 | 40052.3 | 2.89E-01 | 0.15 | 0.79 |
| RP_POS_570_35266a10_749 | | 570.3527 | 10.75 | | RP | POS | | | 1682035 | 524837 | 1763268 | 586630 | 2.89E-01 | 0.15 | 0.83 |
| RP_POS_170_97806a15_417 | | 170.9781 | 15.42 | | RP | POS | | | 705937 | 389061 | 757720 | 326467 | 2.89E-01 | 0.14 | 0.59 |
| RP_POS_306_23999a10_253 | | 306.2400 | 10.25 | | RP | POS | | | 893251 | 123537 | 875576 | 120422 | 2.89E-01 | -0.14 | 0.73 |
| RP_POS_313_27328a10_159 | | 313.2733 | 10.16 | | RP | POS | | | 3.7E+07 | 5282502 | 3.6E+07 | 5015579 | 2.89E-01 | -0.14 | 1.06 |
| RP_POS_341_23618a11_36 | | 341.2362 | 11.36 | | RP | POS | | | 158992 | 79119.8 | 148423 | 67094.6 | 2.89E-01 | -0.14 | 1.17 |
| RP_POS_161_0806a9_21 | | 161.0806 | 9.21 | | RP | POS | | | 4001415 | 1913956 | 3720287 | 1953700 | 2.89E-01 | -0.15 | 0.79 |
| RP_POS_389_27466a11_142 | | 389.2747 | 11.14 | | RP | POS | | | 445927 | 194401 | 422051 | 134366 | 2.89E-01 | -0.15 | 1.06 |
| HILIC_POS_371_23999a2_263 | | 371.2400 | 2.26 | | HILIC | POS | | | 8820221 | 1959731 | 9132054 | 2299160 | 2.90E-01 | 0.15 | 0.68 |
| RP_POS_121_01733a15_492 | | 121.0173 | 15.49 | | RP | POS | | | 132211 | 232760 | 176393 | 354176 | 2.90E-01 | 0.15 | 0.75 |
| RP_POS_137_04584a0_974 | | 137.0458 | 0.97 | | RP | POS | 137.0464(100), 119.0355(2) | | 6225339 | 3254388 | 6684401 | 3097428 | 2.90E-01 | 0.14 | 0.71 |
| RP_POS_594_16003a9_644 | | 594.1600 | 9.64 | | RP | POS | | | 2520679 | 232154 | 2552856 | 213060 | 2.90E-01 | 0.14 | 1.05 |
| RP_POS_794_60492a13_792 | | 794.6049 | 13.79 | | RP | POS | | | 6.7E+07 | 1.6E+07 | 6.9E+07 | 1.5E+07 | 2.90E-01 | 0.14 | 0.81 |
| RP_NEG_778_56152a13_668 | PC(16:0/16:0) | 778.5615 | 13.67 | | RP | NEG | 255.2325(100), 718.531(10) | 2 | 2798584 | 990213 | 2936961 | 923766 | 2.90E-01 | 0.14 | 0.77 |
| HILIC_NEG_100_93359a3_202 | | 100.9336 | 3.20 | | HILIC | NEG | | | 1.7E+07 | 879229 | 1.7E+07 | 627438 | 2.90E-01 | -0.14 | 0.66 |
| RP_POS_331_15979a8_842 | | 331.1598 | 8.84 | | RP | POS | | | 465249 | 183962 | 440006 | 165863 | 2.90E-01 | -0.14 | 0.82 |
| HILIC_POS_678_50513a0_612 | | 678.5051 | 0.61 | | HILIC | POS | | | 244014 | 158673 | 220168 | 169661 | 2.90E-01 | -0.15 | 0.62 |

**Supplemantary table 1**: The non-targeted metabolomics results ordered by the VIP value (p value < 0.0003 are shaded dark green, p value < 0.05 are shaded light green, d values below 0 are blue and above 0 are red)

| | | | | | | | | | Cases | | Controls | | Welch test | Cohen's | |
|---|---|---|---|---|---|---|---|---|---|---|---|---|---|---|---|
| Feature_ID | Metabolite_name | Average_Mz | Average_RT_min_ | HMDB ID | RP_HILIC | POS_NEG | Cureted_MSMS | ID_level | Mean | SD | Mean2 | SD3 | p | d | VIP |
| RP_POS_390_98831a15_493 | | 390.9883 | 15.49 | | RP | POS | | | 132740 | 36923.3 | 127226 | 39051.1 | 2.90E-01 | -0.15 | 1.07 |
| HILIC_NEG_241_9138a6_302 | | 241.9138 | 6.30 | | HILIC | NEG | | | 393659 | 176515 | 421307 | 203084 | 2.91E-01 | 0.15 | 0.72 |
| RP_POS_353_26852a10_412 | | 353.2685 | 10.41 | | RP | POS | | | 1350751 | 759298 | 1514990 | 1374940 | 2.91E-01 | 0.15 | 0.88 |
| RP_POS_658_50934a10_652 | | 658.5093 | 10.65 | | RP | POS | | | 4019931 | 397462 | 4084343 | 482857 | 2.91E-01 | 0.15 | 0.99 |
| HILIC_NEG_241_97366a1_346 | | 241.9737 | 1.35 | | HILIC | NEG | | | 588467 | 160701 | 565902 | 152277 | 2.91E-01 | -0.14 | 1.01 |
| HILIC_NEG_391_00156a0_935 | | 391.0016 | 0.94 | | HILIC | NEG | | | 704394 | 78234.9 | 693584 | 71602.7 | 2.91E-01 | -0.14 | 0.77 |
| RP_NEG_219_84579a0_644 | | 219.8458 | 0.64 | | RP | NEG | | | 238357 | 84990.9 | 228312 | 52780.7 | 2.91E-01 | -0.15 | 0.64 |
| RP_POS_236_07164a10_689 | | 236.0716 | 10.69 | | RP | POS | | | 1.4E+08 | 2.4E+07 | 1.3E+08 | 2.9E+07 | 2.91E-01 | -0.15 | 0.77 |
| RP_POS_163_03903a9_523 | | 163.0390 | 9.52 | | RP | POS | 163.0394(100) | | 5036152 | 1052005 | 5217593 | 1407921 | 2.92E-01 | 0.15 | 1 |
| RP_POS_236_1852a8_078 | | 236.1852 | 8.08 | | RP | POS | | | 935876 | 90002.5 | 949602 | 99175.6 | 2.92E-01 | 0.15 | 1.05 |
| RP_POS_694_48615a11_592 | | 694.4862 | 11.59 | | RP | POS | | | 153798 | 122732 | 175165 | 166658 | 2.92E-01 | 0.15 | 0.9 |
| HILIC_NEG_226_99535a2_267 | | 226.9954 | 2.27 | | HILIC | NEG | | | 904086 | 220827 | 935451 | 214353 | 2.92E-01 | 0.14 | 0.82 |
| RP_NEG_135_03123a0_974 | | 135.0312 | 0.97 | | RP | NEG | | | 309618 | 171922 | 333661 | 162225 | 2.92E-01 | 0.14 | 0.73 |
| RP_POS_173_02908a15_235 | | 173.0291 | 15.24 | | RP | POS | | | 4142369 | 1953007 | 4437540 | 2120943 | 2.92E-01 | 0.14 | 0.81 |
| RP_POS_272_18579a7_695 | | 272.1858 | 7.70 | | RP | POS | | | 9729929 | 839506 | 9847166 | 790399 | 2.92E-01 | 0.14 | 1.08 |
| RP_POS_784_58575a13_476 | | 784.5858 | 13.48 | | RP | POS | | | 6.6E+08 | 1.7E+08 | 6.9E+08 | 1.5E+08 | 2.92E-01 | 0.14 | 0.89 |
| HILIC_POS_102_09175a4_765 | | 102.0918 | 4.77 | | HILIC | POS | | | 477406 | 417901 | 418040 | 405561 | 2.92E-01 | -0.14 | 0.73 |
| RP_POS_545_26337a8_842 | | 545.2634 | 8.84 | | RP | POS | | | 292011 | 312181 | 254161 | 211674 | 2.92E-01 | -0.14 | 1.04 |
| RP_POS_323_1955a10_715 | | 323.1955 | 10.72 | | RP | POS | | | 790041 | 620612 | 887027 | 714942 | 2.93E-01 | 0.15 | 0.96 |
| RP_POS_253_14334a6_885 | | 253.1433 | 6.89 | | RP | POS | | | 1382985 | 262276 | 1422810 | 288357 | 2.93E-01 | 0.14 | 0.82 |
| RP_POS_257_05704a6_02 | | 257.0570 | 6.02 | | RP | POS | | | 1419663 | 515432 | 1491095 | 478441 | 2.93E-01 | 0.14 | 0.66 |
| RP_POS_153_09099a9_21 | | 153.0910 | 9.21 | | RP | POS | | | 1443152 | 549588 | 1360546 | 594260 | 2.93E-01 | -0.14 | 0.76 |
| RP_POS_236_07172a12_656 | | 236.0717 | 12.66 | | RP | POS | | | 1.2E+08 | 6E+07 | 1.1E+08 | 4.9E+07 | 2.93E-01 | -0.14 | 0.62 |
| RP_POS_379_24994a12_807 | | 379.2499 | 12.81 | | RP | POS | | | 1193487 | 892591 | 1079760 | 692330 | 2.93E-01 | -0.14 | 0.86 |
| RP_POS_443_19427a8_842 | | 443.1943 | 8.84 | | RP | POS | | | 171908 | 119164 | 155795 | 105422 | 2.93E-01 | -0.14 | 1.18 |
| RP_POS_613_51379a11_6 | | 613.5138 | 11.60 | | RP | POS | | | 419722 | 103748 | 405304 | 97118.1 | 2.93E-01 | -0.14 | 0.68 |
| RP_NEG_552_78833a15_374 | | 552.7883 | 15.37 | | RP | NEG | | | 767405 | 196763 | 799592 | 244310 | 2.94E-01 | 0.15 | 0.97 |
| RP_POS_285_24234a10_359 | | 285.2423 | 10.36 | | RP | POS | | | 913029 | 609361 | 1011256 | 739476 | 2.94E-01 | 0.15 | 0.76 |
| RP_POS_558_39996a10_771 | | 558.4000 | 10.77 | | RP | POS | | | 381429 | 215713 | 418542 | 289716 | 2.94E-01 | 0.15 | 1.3 |
| RP_POS_718_53778a13_669 | | 718.5378 | 13.67 | | RP | POS | | | 1234185 | 1058869 | 1442278 | 1708156 | 2.94E-01 | 0.15 | 0.93 |
| HILIC_NEG_158_93909a4_412 | | 158.9391 | 4.41 | | HILIC | NEG | | | 4158038 | 426941 | 4212760 | 336914 | 2.94E-01 | 0.14 | 1.08 |
| RP_NEG_848_58179a10_653 | | 848.5818 | 10.65 | | RP | NEG | | | 426518 | 276595 | 465394 | 265908 | 2.94E-01 | 0.14 | 1.07 |
| RP_POS_313_06601a2_887 | | 313.0660 | 2.89 | | RP | POS | | | 214207 | 63715.3 | 222826 | 56545.8 | 2.94E-01 | 0.14 | 0.85 |
| HILIC_POS_133_09723a7_154 | Ornithine | 133.0972 | 7.15 | | HILIC | POS | 70.0661(100). 116.0713(55) | 1 | 5488760 | 2408298 | 5843046 | 2513424 | 2.94E-01 | 0.14 | 0.72 |
| HILIC_POS_184_07359a1_102 | | 184.0736 | 1.10 | | HILIC | POS | 184.0745(100), 86.0975(40) | | 2434614 | 2477552 | 2102830 | 2161706 | 2.94E-01 | -0.14 | 0.87 |
| RP_NEG_337_23712a11_291 | | 337.2371 | 11.29 | | RP | NEG | | | 191540 | 67004.3 | 182176 | 63574.9 | 2.94E-01 | -0.14 | 1.32 |
| RP_POS_299_1441a9_442 | | 299.1441 | 9.44 | | RP | POS | | | 381529 | 433606 | 331440 | 254536 | 2.94E-01 | -0.15 | 0.93 |
| RP_POS_310_8764a15_422 | | 310.8764 | 15.42 | | RP | POS | | | 215918 | 107131 | 198480 | 132239 | 2.94E-01 | -0.15 | 0.68 |
| HILIC_NEG_157_9008a4_546 | | 157.9008 | 4.55 | | HILIC | NEG | | | 1456267 | 1144486 | 1602396 | 898578 | 2.95E-01 | 0.14 | 0.6 |
| HILIC_POS_182_10342a1_388 | | 182.1034 | 1.39 | | HILIC | POS | | | 796710 | 270136 | 835470 | 270359 | 2.95E-01 | 0.14 | 0.71 |
| RP_NEG_294_98282a15_47 | | 294.9828 | 15.47 | | RP | NEG | | | 949604 | 90036 | 962791 | 93651.9 | 2.95E-01 | 0.14 | 0.64 |
| RP_POS_286_20142a7_696 | | 286.2014 | 7.70 | | RP | POS | | | 715052 | 92476 | 729670 | 109412 | 2.95E-01 | 0.14 | 1.04 |
| RP_POS_624_10303a11_38 | | 624.1030 | 11.38 | | RP | POS | | | 188433 | 105522 | 204836 | 121502 | 2.95E-01 | 0.14 | 0.74 |
| RP_POS_718_5755a13_557 | | 718.5755 | 13.56 | | RP | POS | | | 1179580 | 779296 | 1301509 | 904667 | 2.95E-01 | 0.14 | 0.7 |
| RP_POS_838_63123a14_832 | | 838.6312 | 14.83 | | RP | POS | | | 1327298 | 1385550 | 1536521 | 1521127 | 2.95E-01 | 0.14 | 1.05 |
| RP_POS_281_24747a10_656 | | 281.2475 | 10.66 | | RP | POS | | | 3471901 | 444208 | 3407619 | 452112 | 2.95E-01 | -0.14 | 0.96 |
| RP_POS_352_24564a10_563 | | 352.2456 | 10.56 | | RP | POS | | | 1838027 | 620317 | 1750209 | 604779 | 2.95E-01 | -0.14 | 0.66 |
| RP_POS_540_36566a10_573 | | 540.3657 | 10.57 | | RP | POS | | | 1942769 | 745954 | 1835825 | 746828 | 2.95E-01 | -0.14 | 0.98 |
| RP_POS_673_52911a11_878 | | 673.5291 | 11.88 | | RP | POS | | | 1.3E+07 | 3488003 | 1.3E+07 | 2985766 | 2.95E-01 | -0.14 | 0.75 |
| HILIC_POS_375_32159a1_397 | | 375.3216 | 1.40 | | HILIC | POS | | | 173897 | 93129.4 | 190999 | 137785 | 2.96E-01 | 0.15 | 0.76 |
| RP_NEG_221_94725a13_284 | | 221.9473 | 13.28 | | RP | NEG | | | 5821421 | 1081062 | 5996570 | 1332373 | 2.96E-01 | 0.15 | 0.6 |
| RP_POS_299_63324a10_293 | | 299.6332 | 10.29 | | RP | POS | | | 914191 | 285868 | 961853 | 368288 | 2.96E-01 | 0.15 | 1.03 |
| RP_POS_280_94055a15_332 | | 280.9406 | 15.33 | | RP | POS | | | 2985688 | 2376533 | 3328246 | 2411491 | 2.96E-01 | 0.14 | 0.8 |
| HILIC_POS_118_08662a10_54 | | 118.0866 | 10.54 | | HILIC | POS | 140.069(100), 74.0974(40) | | 2.1E+07 | 1.8E+07 | 1.8E+07 | 1.6E+07 | 2.96E-01 | -0.14 | 0.86 |
| HILIC_NEG_132_90034a7_741 | | 132.9003 | 7.74 | | HILIC | NEG | | | 962387 | 335763 | 917200 | 297768 | 2.96E-01 | -0.14 | 0.73 |
| HILIC_NEG_155_86555a7_034 | | 155.8656 | 7.03 | | HILIC | NEG | | | 625792 | 311236 | 583114 | 286657 | 2.96E-01 | -0.14 | 0.9 |
| RP_POS_332_0246a0_643 | | 332.0246 | 0.64 | | RP | POS | | | 3018113 | 1212148 | 2844137 | 1221212 | 2.96E-01 | -0.14 | 0.91 |
| RP_NEG_325_18472a11_727 | | 325.1847 | 11.73 | | RP | NEG | 325.1825(100), 183.0121(10) | | 3156469 | 6664706 | 4455792 | 1.1E+07 | 2.97E-01 | 0.15 | 0.76 |
| RP_POS_302_2453a10_641 | | 302.2453 | 10.64 | | RP | POS | | | 1375261 | 3578848 | 1980751 | 4733244 | 2.97E-01 | 0.15 | 1.33 |
| HILIC_NEG_497_04507a7_662 | | 497.0451 | 7.66 | | HILIC | NEG | | | 258626 | 12678.4 | 260561 | 14242.9 | 2.97E-01 | 0.14 | 0.81 |
| RP_NEG_134_89478a11_143 | | 134.8948 | 11.14 | | RP | NEG | | | 1173049 | 108008 | 1188489 | 108252 | 2.97E-01 | 0.14 | 1.08 |
| RP_POS_1159_33228a13_216 | | 1159.3323 | 13.22 | | RP | POS | | | 8628352 | 3697929 | 9146200 | 3568826 | 2.97E-01 | 0.14 | 0.85 |
| RP_POS_225_10951a7_653 | Sebacic acid | 225.1095 | 7.65 | | RP | POS | 225.112(100) | 2 | 899515 | 204641 | 873457 | 161806 | 2.97E-01 | -0.14 | 0.95 |
| HILIC_POS_201_99084a0_937 | | 201.9908 | 0.94 | | HILIC | POS | | | 241849 | 74476.9 | 232063 | 63198.2 | 2.97E-01 | -0.14 | 0.86 |
| HILIC_POS_84_08141a7_098 | | 84.0814 | 7.10 | | HILIC | POS | | | 1128997 | 400661 | 1076042 | 343106 | 2.97E-01 | -0.14 | 0.76 |
| RP_NEG_979_14008a9_406 | | 979.1401 | 9.41 | | RP | NEG | | | 2257887 | 314469 | 2213188 | 311639 | 2.97E-01 | -0.14 | 1.01 |
| RP_POS_217_14288a8_099 | | 217.1429 | 8.10 | | RP | POS | | | 2505974 | 776160 | 2396574 | 758200 | 2.97E-01 | -0.14 | 0.86 |
| RP_POS_229_084a15_416 | | 229.0840 | 15.42 | | RP | POS | | | 579723 | 210869 | 549516 | 212024 | 2.97E-01 | -0.14 | 0.79 |
| HILIC_POS_393_05023a3_146 | | 393.0502 | 3.15 | | HILIC | POS | | | 325504 | 19824.2 | 321040 | 38269.4 | 2.97E-01 | -0.15 | 0.72 |
| HILIC_NEG_259_00119a2_265 | | 259.0012 | 2.27 | | HILIC | NEG | | | 706441 | 238588 | 738499 | 212675 | 2.98E-01 | 0.14 | 0.84 |
| RP_NEG_187_13411a8_085 | | 187.1341 | 8.09 | | RP | NEG | | | 724789 | 385523 | 787031 | 475763 | 2.98E-01 | 0.14 | 1.14 |
| RP_NEG_248_9608a13_892 | | 248.9608 | 13.89 | | RP | NEG | | | 1951517 | 967841 | 2088478 | 954085 | 2.98E-01 | 0.14 | 0.73 |
| RP_POS_603_08167a9_988 | | 603.0817 | 9.99 | | RP | POS | | | 231529 | 159486 | 254864 | 167627 | 2.98E-01 | 0.14 | 0.63 |
| HILIC_POS_478_32794a1_246 | | 478.3279 | 1.25 | | HILIC | POS | 478.3312(100) | | 207006 | 120993 | 189743 | 121508 | 2.98E-01 | -0.14 | 0.73 |
| HILIC_NEG_257_84503a7_735 | | 257.8450 | 7.74 | | HILIC | NEG | | | 900052 | 88091.9 | 888088 | 80474.4 | 2.98E-01 | -0.14 | 0.87 |
| RP_NEG_277_18146a9_112 | | 277.1815 | 9.11 | | RP | NEG | | | 2434792 | 207358 | 2406125 | 195909 | 2.98E-01 | -0.14 | 1.18 |
| RP_NEG_698_47748a11_593 | | 698.4775 | 11.59 | | RP | NEG | | | 573105 | 67435.2 | 562945 | 74353.4 | 2.98E-01 | -0.14 | 1.09 |
| RP_POS_180_13577a7_626 | | 180.1358 | 7.63 | | RP | POS | | | 1039735 | 250794 | 1006473 | 218195 | 2.98E-01 | -0.14 | 1.28 |
| RP_POS_189_11186a6_714 | | 189.1119 | 6.71 | | RP | POS | | | 352008 | 223844 | 320345 | 220734 | 2.98E-01 | -0.14 | 0.82 |
| RP_POS_448_3421a9_285 | | 448.3421 | 9.29 | | RP | POS | | | 700948 | 851432 | 586823 | 757229 | 2.98E-01 | -0.14 | 0.95 |
| RP_POS_302_24582a11_59 | | 302.2458 | 11.59 | | RP | POS | | | 1.1E+08 | 3.9E+08 | 1.8E+08 | 6.6E+08 | 2.99E-01 | 0.15 | 0.82 |
| HILIC_POS_359_00711a7_362 | | 359.0071 | 7.36 | | HILIC | POS | | | 1202308 | 54567.8 | 1210996 | 66070.9 | 2.99E-01 | 0.14 | 0.66 |
| RP_NEG_775_59796a13_55 | | 775.5980 | 13.55 | | RP | NEG | | | 1679293 | 709363 | 1786946 | 794997 | 2.99E-01 | 0.14 | 0.75 |
| RP_POS_564_43738a11_709 | | 564.4374 | 11.71 | | RP | POS | | | 377006 | 263437 | 416699 | 292205 | 2.99E-01 | 0.14 | 0.76 |
| RP_POS_382_36777a11_422 | | 382.3678 | 11.42 | | RP | POS | | | 407375 | 255226 | 373890 | 217205 | 2.99E-01 | -0.14 | 0.98 |
| RP_POS_563_90558a10_347 | | 563.9056 | 10.35 | | RP | POS | | | 1783144 | 783443 | 1666213 | 854642 | 2.99E-01 | -0.14 | 0.79 |
| RP_NEG_185_96259a1_39 | | 185.9626 | 1.39 | | RP | NEG | | | 328716 | 277682 | 375853 | 371941 | 3.00E-01 | 0.15 | 0.98 |
| RP_NEG_337_20609a11_336 | | 337.2061 | 11.34 | | RP | NEG | | | 15926.1 | 12676.1 | 18749.8 | 24379 | 3.00E-01 | 0.15 | 0.74 |
| RP_NEG_221_94682a15_55 | | 221.9468 | 15.55 | | RP | NEG | | | 3.1E+07 | 723292 | 3.1E+07 | 736363 | 3.00E-01 | 0.14 | 0.83 |
| RP_NEG_284_22293a10_066 | | 284.2229 | 10.07 | | RP | NEG | | | 412431 | 118126 | 428314 | 106565 | 3.00E-01 | 0.14 | 0.88 |
| RP_POS_233_15311a9_163 | | 233.1531 | 9.16 | | RP | POS | | | 2224827 | 172748 | 2247488 | 148100 | 3.00E-01 | 0.14 | 1.03 |
| RP_POS_181_07205a2_084 | | 181.0721 | 2.08 | | RP | POS | 181.0722(100),138.0668(5) | | 3.1E+07 | 3.7E+07 | 2.7E+07 | 2.6E+07 | 3.00E-01 | -0.14 | 0.62 |
| RP_NEG_1107_16504a9_506 | | 1107.1650 | 9.51 | | RP | NEG | | | 810555 | 110849 | 796040 | 94660.8 | 3.00E-01 | -0.14 | 1.08 |
| RP_POS_329_24332a11_595 | | 329.2433 | 11.60 | | RP | POS | | | 1.1E+07 | 1600095 | 1.1E+07 | 1610799 | 3.00E-01 | -0.14 | 0.86 |
| RP_POS_232_8905a15_461 | | 232.8905 | 15.46 | | RP | POS | | | 56004.7 | 34061.5 | 61083.3 | 37261.1 | 3.01E-01 | 0.14 | 0.74 |
| HILIC_POS_106_08662a3_882 | | 106.0866 | 3.88 | | HILIC | POS | 88.0761(100) | | 6526325 | 3535255 | 6031761 | 3459817 | 3.01E-01 | -0.14 | 0.95 |
| HILIC_NEG_151_87422a3_212 | | 151.8742 | 3.21 | | HILIC | NEG | | | 1226509 | 179913 | 1196465 | 236196 | 3.01E-01 | -0.14 | 0.74 |
| RP_POS_310_20071a8_98 | | 310.2007 | 8.98 | | RP | POS | | | 586215 | 194964 | 557754 | 205859 | 3.01E-01 | -0.14 | 0.65 |
| RP_POS_402_35721a11_449 | | 402.3572 | 11.45 | | RP | POS | | | 613288 | 133600 | 596108 | 109866 | 3.01E-01 | -0.14 | 0.68 |
| RP_POS_569_43292a15_407 | | 569.4329 | 15.41 | | RP | POS | | | 1.1E+07 | 1710179 | 1E+07 | 1963940 | 3.01E-01 | -0.14 | 0.75 |
| HILIC_POS_336_05243a3_146 | | 336.0524 | 3.15 | | HILIC | POS | | | 660300 | 37548 | 651670 | 75133.6 | 3.01E-01 | -0.15 | 0.63 |
| RP_POS_161_0295a15_328 | | 161.0295 | 15.33 | | RP | POS | | | 2285127 | 500583 | 2360919 | 565271 | 3.02E-01 | 0.14 | 0.67 |
| RP_POS_486_01431a10_065 | | 486.0143 | 10.07 | | RP | POS | | | 607900 | 288357 | 651649 | 326968 | 3.02E-01 | 0.14 | 0.82 |
| RP_POS_528_30859a10_438 | | 528.3086 | 10.44 | | RP | POS | | | 4612280 | 1959856 | 4909617 | 2219465 | 3.02E-01 | 0.14 | 0.79 |
| RP_POS_822_54218a13_468 | | 822.5422 | 13.47 | | RP | POS | | | 9713058 | 6866105 | 1.1E+07 | 4912828 | 3.02E-01 | 0.14 | 0.89 |
| HILIC_POS_103_12338a6_169 | Cadaverine | 103.1234 | 6.17 | | HILIC | POS | 86.0968(100) | 1 | 1149776 | 619064 | 1231708 | 543393 | 3.02E-01 | 0.14 | 0.67 |
| HILIC_POS_361_30652a1_335 | | 361.3065 | 1.34 | | HILIC | POS | | | 1478359 | 2523935 | 1153294 | 2096377 | 3.02E-01 | -0.14 | 0.91 |
| RP_NEG_788_5827a13_776 | | 788.5827 | 13.78 | | RP | NEG | | | 1934615 | 1723446 | 1685755 | 1795132 | 3.02E-01 | -0.14 | 0.88 |
| RP_POS_280_15424a3_41 | | 280.1542 | 3.41 | | RP | POS | | | 1341455 | 1339724 | 1181187 | 925497 | 3.02E-01 | -0.14 | 0.83 |
| RP_POS_393_26117a8_552 | | 393.2612 | 8.55 | | RP | POS | | | 264573 | 110112 | 249260 | 106991 | 3.02E-01 | -0.14 | 1.11 |
| RP_POS_597_76709a5_536 | | 597.7671 | 5.54 | | RP | POS | | | 745517 | 562203 | 666234 | 560727 | 3.02E-01 | -0.14 | 0.7 |
| RP_NEG_255_6312a8_994 | | 255.6312 | 8.99 | | RP | NEG | | | 939327 | 2747682 | 1717584 | 7042600 | 3.03E-01 | 0.16 | 0.9 |
| RP_POS_387_1929a10_55 | | 387.1929 | 10.55 | | RP | POS | | | 262898 | 179292 | 290543 | 209340 | 3.03E-01 | 0.14 | 0.74 |
| RP_POS_593_07977a10_132 | | 593.0798 | 10.13 | | RP | POS | | | 548743 | 269337 | 594092 | 359323 | 3.03E-01 | 0.14 | 0.67 |
| RP_POS_688_97583a10_078 | | 688.9758 | 10.08 | | RP | POS | | | 715806 | 324619 | 770817 | 438100 | 3.03E-01 | 0.14 | 1.19 |
| HILIC_NEG_173_92604a6_304 | | 173.9260 | 6.30 | | HILIC | NEG | | | 1E+07 | 2919628 | 9581971 | 3099601 | 3.03E-01 | -0.14 | 0.87 |
| HILIC_POS_167_01288a7_989 | | 167.0129 | 7.99 | | HILIC | POS | | | 1.1E+07 | 5055561 | 9844172 | 4404001 | 3.03E-01 | -0.14 | 0.82 |
| RP_POS_866_22699a9_832 | | 866.2270 | 9.83 | | RP | POS | | | 926801 | 163402 | 907553 | 108332 | 3.03E-01 | -0.14 | 0.72 |
| RP_POS_202_04753a3_536 | | 202.0475 | 3.54 | | RP | POS | | | 1252135 | 1272254 | 1498948 | 2074878 | 3.04E-01 | 0.15 | 0.81 |
| RP_POS_596_60913a15_298 | | 596.6091 | 15.30 | | RP | POS | | | 1560775 | 900917 | 1698964 | 1045186 | 3.04E-01 | 0.14 | 0.81 |
| RP_NEG_241_75838a15_494 | | 241.7584 | 15.49 | | RP | NEG | | | 2.2E+07 | 439185 | 2.2E+07 | 431619 | 3.04E-01 | -0.14 | 0.64 |
| RP_POS_414_30576a6_916 | | 414.3058 | 6.92 | | RP | POS | | | 697315 | 381476 | 642263 | 399215 | 3.04E-01 | -0.14 | 0.97 |
| RP_POS_573_40894a9_775 | | 573.4089 | 9.78 | | RP | POS | | | 5551739 | 2334720 | 5240616 | 2098158 | 3.04E-01 | -0.14 | 1.12 |
| RP_POS_256_01514a4_409 | | 256.0151 | 4.41 | | RP | POS | 256.0171(100) | | 1.9E+08 | 3.8E+07 | 2E+08 | 6.7E+07 | 3.05E-01 | 0.15 | 0.94 |
| RP_POS_401_34109a11_642 | | 401.3411 | 11.64 | | RP | POS | | | 402371 | 164175 | 436398 | 292006 | 3.05E-01 | 0.15 | 0.86 |
| RP_POS_709_89764a11_456 | | 709.8976 | 11.46 | | RP | POS | | | 148430 | 73923.4 | 162882 | 122205 | 3.05E-01 | 0.15 | 0.66 |
| HILIC_POS_398_21539a4_375 | | 398.2154 | 4.38 | | HILIC | POS | | | 169632 | 77675.8 | 181268 | 87068.6 | 3.05E-01 | 0.14 | 0.92 |
| RP_POS_410_76123a13_107 | | 410.7612 | 13.11 | | RP | POS | | | 5701663 | 3348278 | 6258133 | 4420741 | 3.05E-01 | 0.14 | 0.85 |
| RP_POS_588_41504a10_407 | | 588.4150 | 10.41 | | RP | POS | | | 457699 | 253715 | 499599 | 331343 | 3.05E-01 | 0.14 | 0.75 |
| RP_POS_756_55182a12_816 | PC 34:3 (16:1/18:2) | 756.5518 | 12.82 | | RP | POS | 184.0744(100), 86.0947(15) | 1 | 1.8E+08 | 8.1E+07 | 1.7E+08 | 7.6E+07 | 3.05E-01 | -0.14 | 0.7 |
| HILIC_POS_364_19659a2_453 | | 364.1966 | 2.45 | | HILIC | POS | | | 335305 | 173143 | 309214 | 195575 | 3.05E-01 | -0.14 | 0.82 |
| RP_POS_235_13048a5_972 | | 235.1305 | 5.97 | | RP | POS | | | 114271 | 72008.1 | 105333 | 55679.6 | 3.05E-01 | -0.14 | 0.93 |
| RP_POS_283_26309a10_196 | | 283.2631 | 10.20 | | RP | POS | | | 1E+07 | 1403277 | 9893868 | 1286441 | 3.05E-01 | -0.14 | 1.02 |
| RP_POS_555_32831a10_926 | | 555.3283 | 10.93 | | RP | POS | | | 290005 | 202068 | 266135 | 137190 | 3.05E-01 | -0.14 | 0.77 |
| RP_NEG_236_09625a6_456 | | 236.0963 | 6.46 | | RP | NEG | | | 189800 | 37948.4 | 202654 | 119294 | 3.06E-01 | 0.16 | 0.81 |
| RP_POS_304_22705a6_245 | | 304.2271 | 6.25 | | RP | POS | | | 205306 | 220380 | 235076 | 205623 | 3.06E-01 | 0.14 | 0.75 |
| RP_POS_281_17194a9_452 | | 281.1719 | 9.45 | | RP | POS | 281.174(100) | | 1E+08 | 9.4E+07 | 8.8E+07 | 8.5E+07 | 3.06E-01 | -0.14 | 0.96 |
| RP_NEG_718_14661a9_196 | | 718.1466 | 9.20 | | RP | NEG | | | 564273 | 41632.1 | 558811 | 36629.7 | 3.06E-01 | -0.14 | 0.81 |
| RP_POS_204_06332a0_679 | | 204.0633 | 0.68 | | RP | POS | | | 490245 | 235579 | 461376 | 176821 | 3.06E-01 | -0.14 | 0.66 |
| RP_POS_262_20068a8_845 | | 262.2007 | 8.85 | | RP | POS | | | 2733736 | 580377 | 2657225 | 514761 | 3.06E-01 | -0.14 | 0.64 |
| RP_POS_283_15381a7_658 | | 283.1538 | 7.66 | | RP | POS | | | 1092939 | 120617 | 1074952 | 134299 | 3.06E-01 | -0.14 | 0.72 |
| RP_NEG_327_65228a7_655 | | 327.6523 | 7.66 | | RP | NEG | | | 419551 | 1083740 | 697362 | 2506143 | 3.07E-01 | 0.15 | 0.7 |
| HILIC_NEG_243_9679a1_343 | | 243.9679 | 1.34 | | HILIC | NEG | | | 1.3E+07 | 2893549 | 1.4E+07 | 2452256 | 3.07E-01 | 0.14 | 0.9 |
| RP_NEG_182_85936a15_445 | | 182.8594 | 15.45 | | RP | NEG | | | 955323 | 227581 | 988712 | 247551 | 3.07E-01 | 0.14 | 0.69 |
| RP_POS_239_16385a8_911 | | 239.1639 | 8.91 | | RP | POS | | | 6108313 | 402602 | 6166187 | 422340 | 3.07E-01 | 0.14 | 0.99 |
| RP_POS_263_20004a9_747 | | 263.2000 | 9.75 | | RP | POS | | | 1680285 | 213155 | 1709306 | 202750 | 3.07E-01 | 0.14 | 0.72 |
| RP_POS_539_38361a10_317 | | 539.3836 | 10.32 | | RP | POS | | | 644400 | 514202 | 719674 | 557119 | 3.07E-01 | 0.14 | 0.77 |
| RP_NEG_229_14432a7_972 | | 229.1443 | 7.97 | | RP | NEG | | | 1612771 | 181184 | 1587605 | 178603 | 3.07E-01 | -0.14 | 1.12 |
| RP_POS_281_17197a8_864 | | 281.1720 | 8.86 | | RP | POS | | | 909689 | 138473 | 891841 | 117623 | 3.07E-01 | -0.14 | 0.65 |
| RP_POS_640_315a5_598 | | 640.3150 | 5.60 | | RP | POS | | | 399434 | 124718 | 383710 | 100925 | 3.07E-01 | -0.14 | 0.79 |
| RP_NEG_183_06613a3_303 | | 183.0661 | 3.30 | | RP | NEG | | | 757209 | 389201 | 812062 | 396545 | 3.08E-01 | 0.14 | 0.78 |
| RP_POS_252_04553a14_227 | | 252.0455 | 14.23 | | RP | POS | | | 3080067 | 1629139 | 3311933 | 1690343 | 3.08E-01 | 0.14 | 0.99 |
| HILIC_POS_129_10242a7_098 | | 129.1024 | 7.10 | | HILIC | POS | | | 1292797 | 390314 | 1241129 | 351787 | 3.08E-01 | -0.14 | 0.73 |
| RP_NEG_737_41907a11_143 | | 737.4191 | 11.14 | | RP | NEG | | | 222989 | 59492.5 | 214778 | 58212.3 | 3.08E-01 | -0.14 | 1.23 |
| RP_POS_248_18581a8_846 | | 248.1858 | 8.85 | | RP | POS | | | 718781 | 243677 | 686933 | 214036 | 3.08E-01 | -0.14 | 0.65 |
| RP_POS_427_31458a9_387 | | 427.3146 | 9.39 | | RP | POS | | | 1430237 | 804011 | 1323606 | 728746 | 3.08E-01 | -0.14 | 0.67 |
| RP_POS_443_30927a8_063 | | 443.3093 | 8.06 | | RP | POS | | | 206599 | 153165 | 186435 | 136665 | 3.08E-01 | -0.14 | 0.66 |
| RP_POS_445_36743a11_985 | | 445.3674 | 11.99 | | RP | POS | | | 1040479 | 1617495 | 869080 | 755556 | 3.08E-01 | -0.14 | 0.97 |
| HILIC_NEG_348_00067a7_398 | | 348.0007 | 7.40 | | HILIC | NEG | | | 547521 | 78649.8 | 558806 | 83034.3 | 3.09E-01 | 0.14 | 0.75 |
| RP_POS_132_9585a15_38 | | 132.9585 | 15.38 | | RP | POS | | | 2E+07 | 1.1E+07 | 2.1E+07 | 1.3E+07 | 3.09E-01 | 0.14 | 0.62 |
| RP_POS_723_48828a9_679 | | 723.4883 | 9.68 | | RP | POS | | | 445676 | 288437 | 484461 | 270185 | 3.09E-01 | 0.14 | 1.03 |
| RP_NEG_288_84473a15_405 | | 288.8447 | 15.41 | | RP | NEG | | | 919946 | 245284 | 887159 | 226847 | 3.09E-01 | -0.14 | 0.71 |

**Supplemantary table 1**: The non-targeted metabolomics results ordered by the VIP value (p value < 0.0003 are shaded dark green, p value < 0.05 are shaded light green, d values below 0 are blue and above 0 are red)

| | | | | | | | | | Cases | | Controls | | Welch test | Cohen's | |
|---|---|---|---|---|---|---|---|---|---|---|---|---|---|---|---|
| Feature_ID | Metabolite_name | Average_Mz | Average_RT_min_ | HMDB ID | RP_HILIC | POS_NEG | Cureted_MSMS | ID_level | Mean | SD | Mean2 | SD3 | p | d | VIP |
| RP_POS_147_11665a4_643 | | 147.1167 | 4.64 | | RP | POS | | | 58056.5 | 43722.7 | 52501.8 | 36289.1 | 3.09E-01 | -0.14 | 0.97 |
| HILIC_POS_497_05991a3_146 | | 497.0599 | 3.15 | | HILIC | POS | | | 559696 | 64980.2 | 547485 | 102899 | 3.09E-01 | -0.15 | 0.61 |
| RP_POS_437_25089a9_901 | | 437.2509 | 9.90 | | RP | POS | 437.2534(100) | | 726991 | 356304 | 779055 | 389567 | 3.10E-01 | 0.14 | 1.14 |
| RP_NEG_339_19934a12_881 | | 339.1993 | 12.88 | | RP | NEG | | | 1178159 | 985914 | 1347753 | 1385747 | 3.10E-01 | 0.14 | 0.67 |
| RP_POS_281_6712a10_609 | | 281.6712 | 10.61 | | RP | POS | | | 1089400 | 383514 | 1147207 | 441275 | 3.10E-01 | 0.14 | 1.12 |
| RP_POS_505_02707a10_204 | | 505.0271 | 10.20 | | RP | POS | | | 1117173 | 556204 | 1209635 | 743954 | 3.10E-01 | 0.14 | 0.77 |
| RP_POS_530_46881a15_159 | | 530.4688 | 15.16 | | RP | POS | | | 335983 | 228888 | 367372 | 223251 | 3.10E-01 | 0.14 | 0.71 |
| RP_POS_633_94238a11_213 | | 633.9424 | 11.21 | | RP | POS | | | 293223 | 125053 | 316106 | 191604 | 3.10E-01 | 0.14 | 0.71 |
| RP_NEG_279_2334a10_962 | Linoleic acid | 279.2334 | 10.96 | | RP | NEG | 279.2336(100) | 2 | 9371515 | 6536181 | 1E+07 | 6254710 | 3.10E-01 | 0.14 | 1.12 |
| RP_NEG_629_1109a9_337 | | 629.1109 | 9.34 | | RP | NEG | | | 668025 | 71376.6 | 658535 | 65623.1 | 3.10E-01 | -0.14 | 0.73 |
| RP_POS_210_10985a11_144 | | 210.1099 | 11.14 | | RP | POS | | | 2542954 | 337255 | 2491746 | 392601 | 3.10E-01 | -0.14 | 0.68 |
| RP_POS_470_28772a10_057 | | 470.2877 | 10.06 | | RP | POS | | | 608316 | 236349 | 572290 | 277031 | 3.10E-01 | -0.14 | 0.92 |
| HILIC_POS_560_31146a1_176 | | 560.3115 | 1.18 | | HILIC | POS | | | 436410 | 148638 | 457712 | 157725 | 3.11E-01 | 0.14 | 0.84 |
| RP_NEG_125_87308a15_487 | | 125.8731 | 15.49 | | RP | NEG | | | 997691 | 120426 | 1018652 | 172582 | 3.11E-01 | 0.14 | 0.76 |
| RP_NEG_441_11041a8_228 | | 441.1104 | 8.23 | | RP | NEG | | | 221537 | 18847.9 | 223871 | 14920.4 | 3.11E-01 | 0.14 | 0.75 |
| RP_POS_671_87299a11_45 | | 671.8730 | 11.45 | | RP | POS | | | 155692 | 92207.9 | 169101 | 100342 | 3.11E-01 | 0.14 | 0.62 |
| HILIC_NEG_234_81953a7_038 | | 234.8195 | 7.04 | | HILIC | NEG | | | 945774 | 391687 | 892466 | 378425 | 3.11E-01 | -0.14 | 0.58 |
| RP_NEG_353_20044a10_327 | | 353.2004 | 10.33 | | RP | NEG | | | 66664 | 51097.9 | 60189.3 | 42523 | 3.11E-01 | -0.14 | 0.64 |
| RP_POS_247_24171a10_196 | | 247.2417 | 10.20 | | RP | POS | | | 852015 | 134358 | 831760 | 155266 | 3.11E-01 | -0.14 | 0.93 |
| RP_NEG_205_8396a15_477 | | 205.8396 | 15.48 | | RP | NEG | | | 2228402 | 130077 | 2247097 | 139200 | 3.12E-01 | 0.14 | 0.91 |
| RP_POS_287_20071a8_977 | | 287.2007 | 8.98 | | RP | POS | | | 2865980 | 163237 | 2890504 | 188129 | 3.12E-01 | 0.14 | 0.79 |
| RP_POS_348_28967a11_038 | | 348.2897 | 11.04 | | RP | POS | | | 4432568 | 3110106 | 4844131 | 2856341 | 3.12E-01 | 0.14 | 1.1 |
| RP_POS_452_39545a10_874 | | 452.3955 | 10.87 | | RP | POS | | | 8144277 | 717225 | 8244268 | 727504 | 3.12E-01 | 0.14 | 0.92 |
| RP_POS_519_32501a10_359 | | 519.3250 | 10.36 | | RP | POS | | | 2.3E+07 | 6594819 | 2.4E+07 | 8964369 | 3.12E-01 | 0.14 | 0.63 |
| RP_POS_546_35455a10_929 | | 546.3546 | 10.93 | | RP | POS | | | 2.1E+08 | 3.3E+07 | 2.1E+08 | 3.8E+07 | 3.12E-01 | 0.14 | 0.88 |
| RP_POS_798_03015a13_167 | | 798.0302 | 13.17 | | RP | POS | | | 392803 | 292885 | 436684 | 335995 | 3.12E-01 | 0.14 | 0.85 |
| RP_POS_367_22757a10_878 | | 367.2276 | 10.88 | | RP | POS | 367.1959(100) | | 1513909 | 462155 | 1454369 | 402075 | 3.12E-01 | -0.14 | 0.75 |
| RP_NEG_175_07237a0_544 | | 175.0724 | 0.54 | | RP | NEG | | | 659464 | 188666 | 632893 | 194435 | 3.12E-01 | -0.14 | 0.85 |
| RP_POS_241_21591a9_087 | | 241.2159 | 9.09 | | RP | POS | | | 734673 | 79401.6 | 722932 | 89148.7 | 3.12E-01 | -0.14 | 0.6 |
| RP_NEG_506_79346a15_409 | | 506.7935 | 15.41 | | RP | NEG | | | 303928 | 71579.9 | 314522 | 80726.6 | 3.13E-01 | 0.14 | 0.96 |
| RP_POS_318_19095a5_839 | | 318.1910 | 5.84 | | RP | POS | | | 215017 | 104477 | 231046 | 125038 | 3.13E-01 | 0.14 | 1.03 |
| HILIC_POS_147_11285a7_098 | L-Lysine | 147.1129 | 7.10 | | HILIC | POS | 84.0815(100), 130.0868(40) | 1 | 7E+07 | 2.5E+07 | 6.7E+07 | 2.2E+07 | 3.13E-01 | -0.14 | 0.75 |
| HILIC_POS_175_11874a6_96 | Arginine | 175.1187 | 6.96 | | HILIC | POS | 175.1193(100), 70.0661(60), 116.0714(55) | 1 | 6.4E+07 | 1.3E+07 | 6.6E+07 | 1.4E+07 | 3.13E-01 | 0.14 | 0.69 |
| HILIC_POS_150_11269a2_248 | | 150.1127 | 2.25 | | HILIC | POS | 150.1126(100), 132.1046(40) | | 3220720 | 2657656 | 2869462 | 2442055 | 3.13E-01 | -0.14 | 1.22 |
| HILIC_POS_184_05774a5_461 | | 184.0577 | 5.46 | | HILIC | POS | | | 314686 | 158916 | 293105 | 154099 | 3.13E-01 | -0.14 | 0.66 |
| RP_NEG_787_10175a9_196 | | 787.1018 | 9.20 | | RP | NEG | | | 5573534 | 373986 | 5527453 | 294560 | 3.13E-01 | -0.14 | 1.08 |
| RP_POS_177_95583a0_607 | | 177.9558 | 0.61 | | RP | POS | | | 1243228 | 269576 | 1204215 | 292851 | 3.13E-01 | -0.14 | 0.68 |
| RP_POS_368_21906a10_365 | | 368.2191 | 10.37 | | RP | POS | | | 1970443 | 689532 | 1881295 | 605023 | 3.13E-01 | -0.14 | 0.86 |
| HILIC_NEG_141_9239a2_995 | | 141.9239 | 3.00 | | HILIC | NEG | | | 1070657 | 114610 | 1086459 | 114502 | 3.14E-01 | 0.14 | 0.65 |
| RP_POS_322_95129a15_384 | | 322.9513 | 15.38 | | RP | POS | | | 1045722 | 722048 | 1137835 | 618278 | 3.14E-01 | 0.14 | 0.8 |
| RP_POS_779_60309a13_753 | | 779.6031 | 13.75 | | RP | POS | | | 1386535 | 2225118 | 1690951 | 2194176 | 3.14E-01 | 0.14 | 0.66 |
| RP_POS_795_57422a13_401 | | 795.5742 | 13.40 | | RP | POS | | | 5012474 | 4878350 | 5745118 | 5666494 | 3.14E-01 | 0.14 | 0.79 |
| RP_NEG_204_79132a15_497 | | 204.7913 | 15.50 | | RP | NEG | | | 601258 | 38274.7 | 596182 | 35551 | 3.14E-01 | -0.14 | 0.82 |
| RP_NEG_245_7518a15_494 | | 245.7518 | 15.49 | | RP | NEG | | | 3680529 | 101806 | 3666619 | 100164 | 3.14E-01 | -0.14 | 0.72 |
| RP_NEG_246_9402a0_667 | | 246.9402 | 0.67 | | RP | NEG | | | 421982 | 115901 | 407894 | 88839.5 | 3.14E-01 | -0.14 | 0.65 |
| RP_POS_484_36728a10_454 | | 484.3673 | 10.45 | | RP | POS | | | 750125 | 389503 | 703176 | 292421 | 3.14E-01 | -0.14 | 0.77 |
| RP_POS_574_25677a10_313 | | 574.2568 | 10.31 | | RP | POS | | | 549823 | 218066 | 522278 | 183213 | 3.14E-01 | -0.14 | 0.93 |
| RP_POS_370_29501a10_413 | | 370.2950 | 10.41 | | RP | POS | | | 339519 | 269671 | 393893 | 476430 | 3.15E-01 | 0.15 | 0.88 |
| HILIC_NEG_183_93456a5_353 | | 183.9346 | 5.35 | | HILIC | NEG | | | 2568591 | 370478 | 2620573 | 384601 | 3.15E-01 | 0.14 | 0.67 |
| HILIC_NEG_188_94989a1_346 | | 188.9499 | 1.35 | | HILIC | NEG | | | 2E+07 | 2886093 | 2.1E+07 | 2467626 | 3.15E-01 | 0.14 | 0.87 |
| RP_POS_640_78845a11_679 | | 640.7885 | 11.68 | | RP | POS | | | 121723 | 66702.5 | 132782 | 90248.8 | 3.15E-01 | 0.14 | 0.73 |
| RP_NEG_467_80038a15_386 | | 467.8004 | 15.39 | | RP | NEG | | | 525585 | 246343 | 496083 | 183642 | 3.15E-01 | -0.14 | 0.71 |
| RP_POS_239_12495a5_603 | | 239.1250 | 5.60 | | RP | POS | | | 307975 | 128581 | 289818 | 134679 | 3.15E-01 | -0.14 | 0.83 |
| RP_POS_371_31561a11_455 | | 371.3156 | 11.46 | | RP | POS | | | 7829410 | 891919 | 7708562 | 865909 | 3.15E-01 | -0.14 | 0.79 |
| RP_POS_530_87122a10_4 | | 530.8712 | 10.40 | | RP | POS | | | 1405726 | 694746 | 1309153 | 709053 | 3.15E-01 | -0.14 | 0.81 |
| RP_POS_615_38696a11_672 | | 615.3870 | 11.67 | | RP | POS | | | 407275 | 541913 | 346313 | 337193 | 3.15E-01 | -0.14 | 0.59 |
| RP_POS_806_06018a13_264 | | 806.0602 | 13.26 | | RP | POS | | | 5479133 | 2683924 | 5147823 | 2144738 | 3.15E-01 | -0.14 | 0.71 |
| HILIC_NEG_259_00119a3_977 | | 259.0012 | 3.98 | | HILIC | NEG | | | 599752 | 282026 | 639679 | 298247 | 3.16E-01 | 0.14 | 0.78 |
| RP_POS_195_13806a8_784 | | 195.1381 | 8.78 | | RP | POS | | | 1936407 | 589726 | 2032222 | 776975 | 3.16E-01 | 0.14 | 0.67 |
| RP_POS_444_33179a10_389 | | 444.3318 | 10.39 | | RP | POS | | | 1718692 | 838851 | 1832876 | 826348 | 3.16E-01 | 0.14 | 1.12 |
| RP_POS_550_29034a10_43 | | 550.2903 | 10.43 | | RP | POS | | | 579443 | 236976 | 614535 | 270019 | 3.16E-01 | 0.14 | 0.83 |
| RP_POS_632_43719a9_576 | | 632.4372 | 9.58 | | RP | POS | | | 722535 | 353749 | 777462 | 435514 | 3.16E-01 | 0.14 | 0.63 |
| RP_NEG_122_89406a15_504 | | 122.8941 | 15.50 | | RP | NEG | | | 339484 | 82594.4 | 328036 | 84091.5 | 3.16E-01 | -0.14 | 0.8 |
| RP_NEG_155_86565a15_495 | | 155.8657 | 15.50 | | RP | NEG | | | 722546 | 56583.9 | 715029 | 53214.2 | 3.16E-01 | -0.14 | 0.69 |
| RP_NEG_589_51892a11_591 | | 589.5189 | 11.59 | | RP | NEG | | | 733667 | 293879 | 694216 | 281789 | 3.16E-01 | -0.14 | 1.3 |
| RP_POS_381_78204a9_656 | | 381.7820 | 9.66 | | RP | POS | | | 460135 | 290711 | 424860 | 224585 | 3.16E-01 | -0.14 | 0.64 |
| HILIC_NEG_480_30899a1_785 | | 480.3090 | 1.79 | | HILIC | NEG | | | 121305 | 34213.3 | 126158 | 36395.4 | 3.17E-01 | 0.14 | 0.92 |
| RP_NEG_173_0817a5_052 | | 173.0817 | 5.05 | | RP | NEG | | | 748815 | 134441 | 766574 | 125613 | 3.17E-01 | 0.14 | 1.15 |
| RP_NEG_239_16562a9_187 | | 239.1656 | 9.19 | | RP | NEG | | | 262579 | 192135 | 289663 | 202377 | 3.17E-01 | 0.14 | 1.05 |
| RP_NEG_736_49347a12_665 | | 736.4935 | 12.67 | | RP | NEG | | | 530689 | 381044 | 484890 | 288662 | 3.17E-01 | -0.14 | 0.61 |
| RP_POS_352_28244a9_669 | | 352.2824 | 9.67 | | RP | POS | | | 859131 | 138505 | 835196 | 200267 | 3.17E-01 | -0.14 | 0.89 |
| RP_POS_363_30978a10_52 | | 363.3098 | 10.52 | | RP | POS | | | 1508985 | 301369 | 1463335 | 358209 | 3.17E-01 | -0.14 | 0.84 |
| RP_POS_126_09157a2_802 | | 126.0916 | 2.80 | | RP | POS | | | 2878990 | 2.9E+07 | 144464 | 99856.7 | 3.17E-01 | -0.19 | 0.61 |
| RP_POS_535_04626a10_179 | | 535.0463 | 10.18 | | RP | POS | | | 329284 | 158188 | 356377 | 226468 | 3.18E-01 | 0.14 | 0.67 |
| RP_POS_590_55029a15_163 | | 590.5503 | 15.16 | | RP | POS | | | 1866548 | 1406541 | 2052267 | 1318291 | 3.18E-01 | 0.14 | 0.95 |
| HILIC_POS_170_08124a0_871 | Pyridoxine (Vitamin B6) | 170.0812 | 0.87 | | HILIC | POS | 152.0716(100), 170.0821(30) | 1 | 1.4E+07 | 1.5E+08 | 27211.8 | 39000 | 3.18E-01 | -0.19 | 0.61 |
| RP_NEG_216_78082a15_467 | | 216.7808 | 15.47 | | RP | NEG | | | 2728682 | 156652 | 2709147 | 130041 | 3.18E-01 | -0.14 | 0.79 |
| RP_NEG_318_85229a15_378 | | 318.8523 | 15.38 | | RP | NEG | | | 2397092 | 682383 | 2303956 | 681756 | 3.18E-01 | -0.14 | 0.63 |
| RP_POS_362_10165a8_844 | | 362.1017 | 8.84 | | RP | POS | | | 1242506 | 540647 | 1169322 | 531116 | 3.18E-01 | -0.14 | 0.97 |
| RP_POS_521_38177a12_426 | | 521.3818 | 12.43 | | RP | POS | | | 7866766 | 1.8E+07 | 5650180 | 1.4E+07 | 3.18E-01 | -0.14 | 0.69 |
| RP_POS_591_38959a11_147 | | 591.3896 | 11.15 | | RP | POS | | | 343969 | 96290.3 | 330892 | 95067.1 | 3.18E-01 | -0.14 | 0.63 |
| RP_NEG_325_18472a10_889 | | 325.1847 | 10.89 | | RP | NEG | 325.1825(100), 183.0122(5) | | 8345586 | 5687354 | 9341575 | 8430152 | 3.19E-01 | 0.14 | 0.64 |
| HILIC_POS_388_31622a2_278 | | 388.3162 | 2.28 | | HILIC | POS | | | 97341.7 | 32324.5 | 101954 | 35019.3 | 3.19E-01 | 0.14 | 0.7 |
| RP_POS_1067_69299a10_607 | | 1067.6930 | 10.61 | | RP | POS | | | 1725111 | 732723 | 1826341 | 749613 | 3.19E-01 | 0.14 | 0.81 |
| RP_POS_203_14296a8_063 | | 203.1430 | 8.06 | | RP | POS | | | 1555520 | 159061 | 1579008 | 182228 | 3.19E-01 | 0.14 | 0.91 |
| RP_POS_435_23459a9_866 | | 435.2346 | 9.87 | | RP | POS | | | 510512 | 1469125 | 703313 | 1362415 | 3.19E-01 | 0.14 | 0.66 |
| RP_POS_538_39154a10_728 | | 538.3915 | 10.73 | | RP | POS | | | 154181 | 93360.6 | 167824 | 105442 | 3.19E-01 | 0.14 | 0.77 |
| RP_POS_828_55035a12_785 | | 828.5504 | 12.79 | | RP | POS | | | 2.2E+07 | 8325983 | 2.3E+07 | 8263123 | 3.19E-01 | 0.14 | 0.73 |
| HILIC_NEG_134_89478a5_65 | | 134.8948 | 5.65 | | HILIC | NEG | | | 1.7E+07 | 4438376 | 1.6E+07 | 4215380 | 3.19E-01 | -0.14 | 0.66 |
| HILIC_POS_210_08891a3_028 | | 210.0889 | 3.03 | | HILIC | POS | | | 70470.6 | 32480 | 66354.6 | 28085 | 3.19E-01 | -0.14 | 0.93 |
| RP_POS_735_99664a10_362 | | 735.9966 | 10.36 | | RP | POS | | | 668476 | 410764 | 611212 | 427143 | 3.19E-01 | -0.14 | 0.96 |
| HILIC_POS_141_11354a2_439 | | 141.1135 | 2.44 | | HILIC | POS | | | 7346045 | 7.9E+07 | 63727.3 | 29545.4 | 3.19E-01 | -0.18 | 0.58 |
| RP_NEG_331_03931a5_308 | | 331.0393 | 5.31 | | RP | NEG | | | 567075 | 6120779 | 1104.88 | 706.01 | 3.19E-01 | -0.18 | 0.61 |
| HILIC_POS_371_2403a2_853 | | 371.2403 | 2.85 | | HILIC | POS | | | 770129 | 264482 | 807549 | 283389 | 3.20E-01 | 0.14 | 1.04 |
| RP_POS_193_15855a8_916 | | 193.1586 | 8.92 | | RP | POS | | | 7810996 | 495741 | 7878733 | 498425 | 3.20E-01 | 0.14 | 0.79 |
| RP_POS_236_07172a13_763 | | 236.0717 | 13.76 | | RP | POS | | | 1.2E+08 | 4.9E+07 | 1.3E+08 | 4.2E+07 | 3.20E-01 | 0.14 | 0.69 |
| HILIC_NEG_168_83192a3_113 | | 168.8319 | 3.11 | | HILIC | NEG | | | 2734685 | 132871 | 2717856 | 115333 | 3.20E-01 | -0.14 | 1.38 |
| RP_NEG_553_49487a13_134 | | 553.4949 | 13.13 | | RP | NEG | | | 772719 | 62154.9 | 764643 | 56771.1 | 3.20E-01 | -0.14 | 0.87 |
| RP_POS_223_12975a7_137 | | 223.1298 | 7.14 | | RP | POS | | | 599427 | 123410 | 582916 | 119291 | 3.20E-01 | -0.14 | 0.75 |
| RP_POS_154_08629a3_083 | | 154.0863 | 3.08 | | RP | POS | | | 3766553 | 4E+07 | 39221.5 | 51191.1 | 3.20E-01 | -0.18 | 0.61 |
| RP_POS_154_08632a2_808 | | 154.0863 | 2.81 | | RP | POS | | | 2101185 | 2.2E+07 | 25419.4 | 23435.8 | 3.20E-01 | -0.18 | 0.61 |
| HILIC_POS_167_07364a2_534 | | 167.0736 | 2.53 | | HILIC | POS | | | 1621512 | 6159977 | 4063949 | 2.4E+07 | 3.21E-01 | 0.16 | 1.03 |
| RP_POS_304_26141a10_836 | | 304.2614 | 10.84 | | RP | POS | | | 6.1E+07 | 2.3E+07 | 6.6E+07 | 4.6E+07 | 3.21E-01 | 0.15 | 1 |
| HILIC_NEG_182_86192a3_148 | | 182.8619 | 3.15 | | HILIC | NEG | | | 500846 | 192894 | 528252 | 209256 | 3.21E-01 | 0.14 | 0.58 |
| HILIC_NEG_277_97641a0_986 | | 277.9764 | 0.99 | | HILIC | NEG | | | 146924 | 141931 | 171155 | 203832 | 3.21E-01 | 0.14 | 0.79 |
| RP_NEG_177_092a7_703 | | 177.0920 | 7.70 | | RP | NEG | | | 786320 | 49298.5 | 792602 | 43454.9 | 3.21E-01 | 0.14 | 1.19 |
| RP_POS_1087_36011a10_548 | | 1087.3601 | 10.55 | | RP | POS | | | 732841 | 1210498 | 884119 | 1023250 | 3.21E-01 | 0.14 | 1.16 |
| RP_POS_267_00052a0_587 | | 267.0005 | 0.59 | | RP | POS | | | 1218204 | 247029 | 1254010 | 276730 | 3.21E-01 | 0.14 | 0.73 |
| RP_POS_289_6409a10_609 | | 289.6409 | 10.61 | | RP | POS | | | 5050200 | 1066740 | 5193845 | 1048257 | 3.21E-01 | 0.14 | 1 |
| RP_POS_310_31055a11_234 | | 310.3106 | 11.23 | | RP | POS | | | 3048019 | 3499934 | 3646745 | 5048090 | 3.21E-01 | 0.14 | 1.06 |
| RP_POS_534_38666a14_344 | | 534.3867 | 14.34 | | RP | POS | | | 1555386 | 867128 | 1692362 | 1112710 | 3.21E-01 | 0.14 | 0.89 |
| HILIC_POS_371_01895a6_246 | | 371.0190 | 6.25 | | HILIC | POS | | | 1638618 | 409662 | 1577221 | 484365 | 3.21E-01 | -0.14 | 0.81 |
| RP_POS_281_20859a10_387 | | 281.2086 | 10.39 | | RP | POS | | | 2364691 | 620856 | 2288458 | 504234 | 3.21E-01 | -0.14 | 0.67 |
| RP_POS_430_30051a5_99 | | 430.3005 | 5.99 | | RP | POS | | | 977533 | 771318 | 884180 | 606950 | 3.21E-01 | -0.14 | 0.83 |
| RP_POS_591_26617a9_209 | | 591.2662 | 9.21 | | RP | POS | | | 467388 | 796721 | 384389 | 388939 | 3.21E-01 | -0.14 | 0.68 |
| HILIC_NEG_134_89478a2_272 | | 134.8948 | 2.27 | | HILIC | NEG | | | 4.4E+07 | 6305340 | 4.5E+07 | 7063713 | 3.22E-01 | 0.14 | 0.65 |
| RP_POS_314_23233a7_054 | | 314.2323 | 7.05 | | RP | POS | | | 2051560 | 1078978 | 2203052 | 1149584 | 3.22E-01 | 0.14 | 0.79 |
| RP_POS_376_27893a9_133 | | 376.2789 | 9.13 | | RP | POS | | | 126828 | 71992.1 | 139043 | 103004 | 3.22E-01 | 0.14 | 1.08 |
| RP_POS_481_00754a9_9 | | 481.0075 | 9.90 | | RP | POS | | | 560290 | 376933 | 610086 | 359504 | 3.22E-01 | 0.14 | 0.66 |
| RP_POS_512_26758a8_215 | | 512.2676 | 8.22 | | RP | POS | | | 2969671 | 1349731 | 3172500 | 1609895 | 3.22E-01 | 0.14 | 0.76 |
| HILIC_POS_115_0869a4_016 | | 115.0869 | 4.02 | | HILIC | POS | | | 665705 | 1510950 | 484645 | 1168459 | 3.22E-01 | -0.14 | 0.61 |
| HILIC_POS_209_15366a3_109 | | 209.1537 | 3.11 | | HILIC | POS | | | 119149 | 51683.6 | 112790 | 42454.9 | 3.22E-01 | -0.14 | 0.77 |
| RP_POS_136_95334a0_627 | | 136.9533 | 0.63 | | RP | POS | | | 471353 | 180582 | 447187 | 176607 | 3.22E-01 | -0.14 | 1.16 |
| RP_POS_433_33292a10_37 | | 433.3329 | 10.37 | | RP | POS | | | 291412 | 339843 | 252587 | 231826 | 3.22E-01 | -0.14 | 0.62 |
| RP_POS_618_24701a10_523 | | 618.2470 | 10.52 | | RP | POS | | | 1815272 | 169700 | 1793154 | 157475 | 3.22E-01 | -0.14 | 0.72 |
| RP_POS_710_49042a12_026 | | 710.4904 | 12.03 | | RP | POS | | | 108488 | 65716.2 | 100491 | 52692.8 | 3.22E-01 | -0.14 | 0.93 |
| RP_POS_982_72235a11_593 | | 982.7224 | 11.59 | | RP | POS | | | 1008138 | 189012 | 985119 | 151629 | 3.22E-01 | -0.14 | 0.73 |
| HILIC_POS_171_1129a0_544 | | 171.1129 | 0.54 | | HILIC | POS | | | 3659134 | 3.9E+07 | 55144.5 | 38659.7 | 3.22E-01 | -0.18 | 0.6 |
| HILIC_POS_314_23282a0_741 | | 314.2328 | 0.74 | | HILIC | POS | | | 4957545 | 2756116 | 5382248 | 3424563 | 3.23E-01 | 0.14 | 0.76 |
| RP_NEG_1107_66357a10_302 | | 1107.6636 | 10.30 | | RP | NEG | | | 2749934 | 942754 | 2878286 | 953723 | 3.23E-01 | 0.14 | 0.79 |
| RP_NEG_185_96275a0_718 | | 185.9628 | 0.72 | | RP | NEG | | | 368665 | 149129 | 391138 | 179005 | 3.23E-01 | 0.14 | 0.65 |
| RP_POS_393_22476a10_25 | | 393.2248 | 10.25 | | RP | POS | | | 8453925 | 4289889 | 9015058 | 4022306 | 3.23E-01 | 0.14 | 1.11 |
| RP_POS_808_58197a14_1 | | 808.5820 | 14.10 | | RP | POS | | | 1.7E+08 | 5E+07 | 1.8E+08 | 5.3E+07 | 3.23E-01 | 0.14 | 0.92 |
| HILIC_NEG_178_0511a0_987 | Hippuric acid | 178.0511 | 0.99 | | HILIC | NEG | 134.0607(100), 178.0509(65) | 1 | 2930873 | 2745394 | 3397477 | 3939273 | 3.23E-01 | 0.14 | 0.78 |
| HILIC_POS_134_01881a5_651 | | 134.0188 | 5.65 | | HILIC | POS | | | 490111 | 135146 | 472828 | 121343 | 3.23E-01 | -0.13 | 0.74 |
| RP_NEG_653_30365a11_217 | | 653.3037 | 11.22 | | RP | NEG | | | 584879 | 222596 | 556448 | 198771 | 3.23E-01 | -0.13 | 0.93 |
| RP_NEG_508_50256a15_5 | | 508.5026 | 15.50 | | RP | NEG | | | 330088 | 35361 | 325312 | 35298.5 | 3.23E-01 | -0.14 | 0.9 |
| RP_POS_303_23175a9_946 | | 303.2318 | 9.95 | | RP | POS | 303.233(100) | | 6212007 | 6510342 | 7806766 | 1.5E+07 | 3.24E-01 | 0.15 | 0.78 |
| RP_NEG_391_21313a7_554 | | 391.2131 | 7.55 | | RP | NEG | | | 182316 | 159926 | 207618 | 207452 | 3.24E-01 | 0.14 | 0.72 |
| RP_POS_309_20346a10_17 | | 309.2035 | 10.17 | | RP | POS | | | 1087811 | 373484 | 1144567 | 455839 | 3.24E-01 | 0.14 | 0.82 |
| RP_POS_431_75491a13_231 | | 431.7549 | 13.23 | | RP | POS | | | 439929 | 567279 | 544630 | 913590 | 3.24E-01 | 0.14 | 1 |
| HILIC_POS_130_08652a7_098 | | 130.0865 | 7.10 | | HILIC | POS | 84.0815(100), 130.0868(50) | | 2.4E+07 | 8569205 | 2.3E+07 | 7420700 | 3.24E-01 | -0.13 | 0.74 |
| RP_NEG_442_15143a11_59 | | 442.1514 | 11.59 | | RP | NEG | | | 490334 | 66765.9 | 481162 | 68944.1 | 3.24E-01 | -0.14 | 0.64 |
| RP_POS_169_97719a0_734 | | 169.9772 | 0.73 | | RP | POS | | | 589322 | 246722 | 555271 | 256567 | 3.24E-01 | -0.14 | 0.88 |
| RP_POS_250_17758a11_114 | | 250.1776 | 11.11 | | RP | POS | | | 465912 | 702366 | 384928 | 495626 | 3.24E-01 | -0.14 | 0.68 |
| HILIC_POS_126_09158a0_602 | | 126.0916 | 0.60 | | HILIC | POS | | | 1.2E+07 | 1.2E+08 | 1158114 | 1163341 | 3.24E-01 | -0.18 | 0.6 |
| RP_POS_420_30716a9_141 | | 420.3072 | 9.14 | | RP | POS | | | 378691 | 228342 | 412751 | 271273 | 3.25E-01 | 0.14 | 0.79 |
| RP_POS_203_14314a8_77 | | 203.1431 | 8.77 | | RP | POS | | | 3539191 | 170813 | 3562371 | 173195 | 3.25E-01 | 0.13 | 1.06 |
| HILIC_POS_130_10898a1_461 | Metformin | 130.1090 | 1.46 | | HILIC | POS | 60.0568(100), 130.1094(98) | 2 | 3565542 | 3.9E+07 | 46515.2 | 418690 | 3.25E-01 | -0.18 | 0.63 |
| RP_POS_171_11281a3_083 | Levetiracetam | 171.1128 | 3.08 | | RP | POS | 126.0913(100), 171.15(35) | 2 | 4030396 | 4.1E+07 | 278821 | 1882479 | 3.25E-01 | -0.18 | 0.61 |
| HILIC_NEG_656_88562a4_352 | | 656.8856 | 4.35 | | HILIC | NEG | | | 3201953 | 212410 | 3174717 | 192816 | 3.25E-01 | -0.13 | 0.82 |
| RP_NEG_306_99918a7_248 | | 306.9992 | 7.25 | | RP | NEG | | | 1340335 | 453386 | 1288020 | 323536 | 3.25E-01 | -0.13 | 0.6 |
| RP_POS_408_30835a11_114 | | 408.3084 | 11.11 | | RP | POS | | | 2698088 | 3801497 | 2258236 | 2731169 | 3.25E-01 | -0.13 | 0.67 |
| RP_POS_792_58875a13_299 | | 792.5888 | 13.30 | | RP | POS | | | 7711556 | 4055673 | 7247792 | 2820644 | 3.25E-01 | -0.13 | 0.86 |
| RP_NEG_200_8587a11_144 | | 200.8587 | 11.14 | | RP | NEG | | | 556086 | 108044 | 541056 | 114311 | 3.25E-01 | -0.14 | 0.64 |
| RP_POS_548_38531a11_494 | | 548.3853 | 11.49 | | RP | POS | | | 454968 | 275216 | 424823 | 168073 | 3.25E-01 | -0.14 | 0.66 |
| HILIC_POS_143_08165a5_308 | Ectoine | 143.0817 | 5.31 | | HILIC | POS | 143.0811(100), 97.0766(55) | 2 | 785518 | 1917733 | 584111 | 1009546 | 3.25E-01 | -0.14 | 0.56 |
| RP_NEG_207_83644a14_35 | | 207.8364 | 14.35 | | RP | NEG | | | 237578 | 143971 | 259841 | 181725 | 3.26E-01 | 0.14 | 0.63 |
| RP_NEG_264_99191a6_681 | | 264.9919 | 6.68 | | RP | NEG | | | 8427530 | 273550 | 8468626 | 330789 | 3.26E-01 | 0.14 | 0.65 |
| RP_POS_569_06055a9_867 | | 569.0606 | 9.87 | | RP | POS | | | 207278 | 134862 | 226659 | 151665 | 3.26E-01 | 0.14 | 0.76 |
| HILIC_NEG_632_73138a2_991 | | 632.7314 | 2.99 | | HILIC | NEG | | | 255060 | 40164.6 | 260352 | 38729.7 | 3.26E-01 | 0.13 | 0.58 |
| RP_NEG_451_17587a11_594 | | 451.1759 | 11.59 | | RP | NEG | | | 548273 | 129143 | 566436 | 140151 | 3.26E-01 | 0.13 | 0.67 |

**Supplemantary table 1**: The non-targeted metabolomics results ordered by the VIP value (p value < 0.0003 are shaded dark green, p value < 0.05 are shaded light green, d values below 0 are blue and above 0 are red)

| | | | | | | | | | Cases | | Controls | | Welch test | Cohen's | |
|---|---|---|---|---|---|---|---|---|---|---|---|---|---|---|---|
| Feature_ID | Metabolite_name | Average_Mz | Average_RT_min_ | HMDB ID | RP_HILIC | POS_NEG | Cureted_MSMS | ID_level | Mean | SD | Mean2 | SD3 | p | d | VIP |
| RP_POS_337_2579a11_239 | | 337.2579 | 11.24 | | RP | POS | | | 1555354 | 786794 | 1653658 | 679216 | 3.26E-01 | 0.13 | 1.01 |
| RP_POS_496_42102a10_878 | | 496.4210 | 10.88 | | RP | POS | | | 9012174 | 786917 | 9119363 | 807394 | 3.26E-01 | 0.13 | 0.96 |
| RP_POS_772_58472a13_644 | PC 35:2 (18:2_17:0) | 772.5847 | 13.64 | | RP | POS | 184.0744(100), 86.0959(20) | 1 | 2.8E+07 | 8042659 | 2.7E+07 | 6780812 | 3.26E-01 | -0.13 | 0.88 |
| RP_POS_218_13863a0_681 | Propionylcarnitine | 218.1386 | 0.68 | | RP | POS | 85.0291(100), 218.1393(30) | 2 | 8943673 | 3222710 | 9367236 | 3085595 | 3.26E-01 | 0.13 | 0.86 |
| HILIC_NEG_347_71609a1_406 | | 347.7161 | 1.41 | | HILIC | NEG | | | 149249 | 30382.3 | 145454 | 26324.6 | 3.26E-01 | -0.13 | 0.59 |
| HILIC_POS_257_11319a1_268 | | 257.1132 | 1.27 | | HILIC | POS | | | 211464 | 57286.7 | 204960 | 39366.1 | 3.26E-01 | -0.13 | 0.8 |
| RP_NEG_241_92944a0_572 | | 241.9294 | 0.57 | | RP | NEG | | | 546710 | 88071.8 | 535270 | 82612.3 | 3.26E-01 | -0.13 | 0.8 |
| RP_NEG_476_53152a15_496 | | 476.5315 | 15.50 | | RP | NEG | | | 590436 | 42060.3 | 584638 | 44054.8 | 3.26E-01 | -0.13 | 0.75 |
| RP_POS_242_92738a15_45 | | 242.9274 | 15.45 | | RP | POS | | | 394800 | 61414.8 | 386844 | 57322 | 3.26E-01 | -0.13 | 0.79 |
| RP_POS_376_11838a9_209 | | 376.1184 | 9.21 | | RP | POS | | | 2056509 | 1312489 | 1879525 | 1318964 | 3.26E-01 | -0.13 | 1.07 |
| RP_NEG_339_19965a11_399 | | 339.1997 | 11.40 | | RP | NEG | | | 2447180 | 2915934 | 2158810 | 1135050 | 3.26E-01 | -0.14 | 0.73 |
| RP_POS_586_45282a9_769 | | 586.4528 | 9.77 | | RP | POS | | | 2338812 | 988232 | 2194969 | 1134298 | 3.26E-01 | -0.14 | 1.18 |
| RP_POS_609_26288a11_417 | | 609.2629 | 11.42 | | RP | POS | | | 161552 | 103691 | 177882 | 135205 | 3.27E-01 | 0.14 | 0.64 |
| RP_POS_496_3403a15_477 | | 496.3403 | 15.48 | | RP | POS | 184.0745(100) | | 772820 | 666384 | 860066 | 637266 | 3.27E-01 | 0.13 | 0.82 |
| HILIC_NEG_295_73575a3_325 | | 295.7358 | 3.33 | | HILIC | NEG | | | 1666997 | 112390 | 1682798 | 122057 | 3.27E-01 | 0.13 | 0.66 |
| HILIC_POS_115_08695a7_152 | | 115.0870 | 7.15 | | HILIC | POS | | | 841316 | 328114 | 886915 | 348937 | 3.27E-01 | 0.13 | 0.68 |
| RP_POS_870_55237a14_076 | | 870.5524 | 14.08 | | RP | POS | | | 562831 | 308371 | 606821 | 343571 | 3.27E-01 | 0.13 | 0.65 |
| HILIC_NEG_118_92419a0_781 | | 118.9242 | 0.78 | | HILIC | NEG | | | 758599 | 379797 | 713315 | 297641 | 3.27E-01 | -0.13 | 0.59 |
| RP_POS_171_03333a15_675 | | 171.0333 | 15.68 | | RP | POS | | | 5035050 | 2587388 | 4669718 | 2835399 | 3.27E-01 | -0.13 | 0.78 |
| RP_POS_253_25194a10_159 | | 253.2519 | 10.16 | | RP | POS | | | 2834880 | 466807 | 2772917 | 457366 | 3.27E-01 | -0.13 | 0.93 |
| RP_POS_310_20111a6_04 | | 310.2011 | 6.04 | | RP | POS | | | 4006701 | 1752602 | 3766791 | 1820326 | 3.27E-01 | -0.13 | 0.59 |
| RP_POS_780_55066a11_595 | | 780.5507 | 11.60 | | RP | POS | | | 3005810 | 515666 | 2939174 | 479610 | 3.27E-01 | -0.13 | 0.59 |
| RP_POS_834_59985a13_424 | | 834.5999 | 13.42 | | RP | POS | | | 8621180 | 4030992 | 8100349 | 3758434 | 3.27E-01 | -0.13 | 0.59 |
| RP_POS_199_06903a0_617 | | 199.0690 | 0.62 | | RP | POS | | | 2140273 | 6509607 | 1450607 | 3585887 | 3.27E-01 | -0.14 | 0.98 |
| RP_POS_343_28445a10_128 | | 343.2845 | 10.13 | | RP | POS | | | 801597 | 77379.9 | 790392 | 88221.6 | 3.27E-01 | -0.14 | 0.87 |
| RP_NEG_327_29102a11_229 | | 327.2910 | 11.23 | | RP | NEG | | | 255186 | 212260 | 290641 | 301749 | 3.28E-01 | 0.14 | 0.78 |
| RP_POS_383_72479a14_336 | | 383.7248 | 14.34 | | RP | POS | | | 288234 | 147380 | 309869 | 172747 | 3.28E-01 | 0.14 | 0.64 |
| RP_POS_515_04016a10_473 | | 515.0402 | 10.47 | | RP | POS | | | 150761 | 86506.3 | 165145 | 122408 | 3.28E-01 | 0.14 | 0.74 |
| HILIC_POS_358_02155a6_478 | | 358.0216 | 6.48 | | HILIC | POS | | | 152318 | 63059 | 160870 | 64471.4 | 3.28E-01 | 0.13 | 0.61 |
| RP_POS_369_29739a11_147 | | 369.2974 | 11.15 | | RP | POS | | | 1178286 | 770531 | 1287165 | 846106 | 3.28E-01 | 0.13 | 0.93 |
| RP_POS_154_95587a15_49 | | 154.9559 | 15.49 | | RP | POS | | | 2691583 | 217632 | 2663458 | 203274 | 3.28E-01 | -0.13 | 1.15 |
| RP_POS_678_50653a12_323 | | 678.5065 | 12.32 | | RP | POS | | | 3704111 | 2972898 | 3304400 | 2998498 | 3.28E-01 | -0.13 | 0.59 |
| RP_POS_698_26862a10_626 | | 698.2686 | 10.63 | | RP | POS | | | 523856 | 237624 | 494354 | 204894 | 3.28E-01 | -0.13 | 0.7 |
| RP_POS_439_35007a11_813 | | 439.3501 | 11.81 | | RP | POS | | | 296956 | 225080 | 274002 | 106599 | 3.28E-01 | -0.14 | 0.67 |
| RP_NEG_230_98631a5_601 | | 230.9863 | 5.60 | | RP | NEG | | | 807707 | 126109 | 834433 | 245560 | 3.29E-01 | 0.14 | 0.79 |
| RP_POS_440_27682a9_21 | | 440.2768 | 9.21 | | RP | POS | | | 648634 | 703871 | 761916 | 953354 | 3.29E-01 | 0.14 | 0.95 |
| RP_POS_541_32843a10_606 | | 541.3284 | 10.61 | | RP | POS | | | 1327705 | 563883 | 1410492 | 662330 | 3.29E-01 | 0.14 | 1.1 |
| RP_POS_608_37067a12_612 | | 608.3707 | 12.61 | | RP | POS | | | 197269 | 170108 | 223001 | 209356 | 3.29E-01 | 0.14 | 0.85 |
| HILIC_POS_210_10971a2_277 | | 210.1097 | 2.28 | | HILIC | POS | | | 57906 | 65081.7 | 66368.7 | 61837.9 | 3.29E-01 | 0.13 | 0.88 |
| HILIC_POS_387_21408a6_171 | | 387.2141 | 6.17 | | HILIC | POS | | | 424315 | 118478 | 438813 | 99313 | 3.29E-01 | 0.13 | 0.6 |
| RP_POS_173_02864a0_637 | | 173.0286 | 0.64 | | RP | POS | | | 3329228 | 728815 | 3431875 | 799286 | 3.29E-01 | 0.13 | 0.69 |
| RP_POS_583_40979a10_279 | | 583.4098 | 10.28 | | RP | POS | | | 659701 | 355796 | 708892 | 377660 | 3.29E-01 | 0.13 | 0.88 |
| HILIC_NEG_142_92894a0_933 | | 142.9289 | 0.93 | | HILIC | NEG | | | 1225107 | 123524 | 1208406 | 126349 | 3.29E-01 | -0.13 | 0.86 |
| HILIC_NEG_175_07237a6_325 | | 175.0724 | 6.33 | | HILIC | NEG | | | 236900 | 71019.9 | 227483 | 70127.6 | 3.29E-01 | -0.13 | 1.03 |
| HILIC_POS_358_36758a3_043 | | 358.3676 | 3.04 | | HILIC | POS | | | 941486 | 413495 | 885796 | 418839 | 3.29E-01 | -0.13 | 0.64 |
| RP_POS_199_0114a15_442 | | 199.0114 | 15.44 | | RP | POS | | | 1688038 | 625018 | 1603717 | 636965 | 3.29E-01 | -0.13 | 0.72 |
| RP_POS_240_15939a6_775 | | 240.1594 | 6.78 | | RP | POS | | | 1923116 | 1.2E+07 | 824958 | 1368102 | 3.29E-01 | -0.16 | 0.55 |
| RP_POS_320_23492a15_41 | | 320.2349 | 15.41 | | RP | POS | | | 1.5E+10 | 4.1E+10 | 2.5E+10 | 9.6E+10 | 3.30E-01 | 0.15 | 0.76 |
| RP_NEG_212_78589a15_469 | | 212.7859 | 15.47 | | RP | NEG | | | 1494285 | 86116.7 | 1508794 | 124610 | 3.30E-01 | 0.14 | 1.09 |
| RP_POS_369_20352a10_389 | | 369.2035 | 10.39 | | RP | POS | | | 815679 | 383946 | 868505 | 406955 | 3.30E-01 | 0.13 | 1.24 |
| RP_POS_636_11609a11_518 | | 636.1161 | 11.52 | | RP | POS | | | 180659 | 93850.6 | 193776 | 102154 | 3.30E-01 | 0.13 | 0.75 |
| HILIC_POS_178_10742a1_373 | | 178.1074 | 1.37 | | HILIC | POS | | | 639280 | 348729 | 593923 | 332606 | 3.30E-01 | -0.13 | 0.83 |
| HILIC_POS_394_31564a3_286 | | 394.3156 | 3.29 | | HILIC | POS | | | 1577253 | 849220 | 1475487 | 682874 | 3.30E-01 | -0.13 | 0.64 |
| RP_NEG_168_83659a11_592 | | 168.8366 | 11.59 | | RP | NEG | | | 333464 | 85366.7 | 322705 | 76643.6 | 3.30E-01 | -0.13 | 0.94 |
| RP_POS_346_00952a15_499 | | 346.0095 | 15.50 | | RP | POS | | | 677887 | 121120 | 660446 | 138637 | 3.30E-01 | -0.13 | 0.93 |
| RP_POS_403_32758a11_594 | | 403.3276 | 11.59 | | RP | POS | | | 2573487 | 516691 | 2508659 | 458166 | 3.30E-01 | -0.13 | 1.23 |
| RP_NEG_243_19701a9_432 | | 243.1970 | 9.43 | | RP | NEG | | | 217526 | 40424.4 | 222607 | 36100.2 | 3.31E-01 | 0.13 | 0.67 |
| RP_POS_628_19293a10_803 | | 628.1929 | 10.80 | | RP | POS | | | 1165325 | 245467 | 1198617 | 254536 | 3.31E-01 | 0.13 | 0.74 |
| HILIC_POS_188_17584a6_206 | | 188.1758 | 6.21 | | HILIC | POS | | | 1849657 | 632202 | 1772732 | 527780 | 3.31E-01 | -0.13 | 0.59 |
| HILIC_POS_402_21188a0_959 | | 402.2119 | 0.96 | | HILIC | POS | | | 121254 | 70145.6 | 112576 | 60822.3 | 3.31E-01 | -0.13 | 0.94 |
| RP_NEG_170_83342a14_342 | | 170.8334 | 14.34 | | RP | NEG | | | 2515106 | 629529 | 2445290 | 416317 | 3.31E-01 | -0.13 | 0.68 |
| RP_NEG_254_97128a0_627 | | 254.9713 | 0.63 | | RP | NEG | | | 280812 | 159564 | 261415 | 132909 | 3.31E-01 | -0.13 | 0.61 |
| RP_NEG_276_99222a15_475 | | 276.9922 | 15.48 | | RP | NEG | | | 2085310 | 154871 | 2062276 | 187376 | 3.31E-01 | -0.13 | 0.67 |
| RP_NEG_610_78772a15_397 | | 610.7877 | 15.40 | | RP | NEG | | | 508165 | 98223.8 | 494538 | 105902 | 3.31E-01 | -0.13 | 0.69 |
| RP_POS_215_13913a0_677 | | 215.1391 | 0.68 | | RP | POS | | | 367387 | 77745.2 | 357558 | 70292.8 | 3.31E-01 | -0.13 | 0.72 |
| RP_POS_298_6246a10_015 | | 298.6246 | 10.02 | | RP | POS | | | 201934 | 142820 | 185085 | 110861 | 3.31E-01 | -0.13 | 0.7 |
| RP_POS_589_42792a10_872 | | 589.4279 | 10.87 | | RP | POS | | | 918640 | 367739 | 874377 | 299389 | 3.31E-01 | -0.13 | 1.16 |
| HILIC_POS_448_0509a3_148 | | 448.0509 | 3.15 | | HILIC | POS | | | 617416 | 290585 | 781767 | 1655758 | 3.32E-01 | 0.17 | 0.79 |
| RP_POS_396_18185a10_614 | | 396.1819 | 10.61 | | RP | POS | | | 379256 | 167370 | 405374 | 218295 | 3.32E-01 | 0.14 | 0.94 |
| RP_NEG_449_21457a9_167 | | 449.2146 | 9.17 | | RP | NEG | | | 710586 | 286069 | 748221 | 281703 | 3.32E-01 | 0.13 | 0.72 |
| RP_POS_267_17181a9_412 | | 267.1718 | 9.41 | | RP | POS | | | 853894 | 181932 | 878856 | 192933 | 3.32E-01 | 0.13 | 1.1 |
| RP_NEG_242_98637a7_221 | | 242.9864 | 7.22 | | RP | NEG | | | 1273851 | 684194 | 1187997 | 612127 | 3.32E-01 | -0.13 | 0.89 |
| RP_NEG_396_89075a11_195 | | 396.8908 | 11.20 | | RP | NEG | | | 441060 | 79619.7 | 431055 | 71580.4 | 3.32E-01 | -0.13 | 0.88 |
| RP_POS_419_34991a11_605 | | 419.3499 | 11.61 | | RP | POS | | | 1544076 | 363520 | 1498221 | 328528 | 3.32E-01 | -0.13 | 1.22 |
| RP_POS_450_81424a10_087 | | 450.8142 | 10.09 | | RP | POS | | | 1425877 | 637793 | 1331210 | 771118 | 3.32E-01 | -0.13 | 0.68 |
| RP_POS_126_09156a3_083 | | 126.0916 | 3.08 | | RP | POS | | | 5039489 | 5.4E+07 | 181781 | 1004810 | 3.32E-01 | -0.18 | 0.6 |
| RP_POS_474_83292a15_377 | | 474.8329 | 15.38 | | RP | POS | | | 860305 | 588453 | 955947 | 817164 | 3.33E-01 | 0.14 | 0.72 |
| RP_POS_589_56445a11_159 | | 589.5645 | 11.16 | | RP | POS | | | 153699 | 135126 | 176847 | 201868 | 3.33E-01 | 0.14 | 1.14 |
| HILIC_POS_575_49231a2_955 | | 575.4923 | 2.96 | | HILIC | POS | | | 64687.5 | 27307.1 | 68578.2 | 31039.6 | 3.33E-01 | 0.13 | 0.88 |
| RP_POS_332_10068a15_326 | | 332.1007 | 15.33 | | RP | POS | | | 3030612 | 644820 | 3110814 | 570802 | 3.33E-01 | 0.13 | 0.62 |
| RP_POS_896_53809a13_004 | | 896.5381 | 13.00 | | RP | POS | | | 5613491 | 2014148 | 5896664 | 2231612 | 3.33E-01 | 0.13 | 0.6 |
| RP_POS_319_22406a10_096 | | 319.2241 | 10.10 | | RP | POS | | | 1455159 | 550602 | 1377319 | 617102 | 3.33E-01 | -0.13 | 0.62 |
| RP_NEG_319_22784a9_938 | | 319.2278 | 9.94 | | RP | NEG | 319.2262(75), 275.2398(10) | | 2245380 | 2673712 | 2888488 | 6127333 | 3.34E-01 | 0.15 | 0.78 |
| RP_POS_654_65942a11_238 | | 654.6594 | 11.24 | | RP | POS | | | 166024 | 79781.7 | 180354 | 127406 | 3.34E-01 | 0.14 | 0.74 |
| RP_POS_249_18436a9_189 | | 249.1844 | 9.19 | | RP | POS | | | 9940014 | 1428047 | 1E+07 | 1519248 | 3.34E-01 | 0.13 | 0.94 |
| RP_POS_252_04549a10_432 | | 252.0455 | 10.43 | | RP | POS | | | 1445185 | 551925 | 1522044 | 604913 | 3.34E-01 | 0.13 | 0.92 |
| RP_POS_391_3421a10_885 | | 391.3421 | 10.89 | | RP | POS | | | 2049152 | 289567 | 2083440 | 230268 | 3.34E-01 | 0.13 | 0.61 |
| RP_POS_253_09729a6_13 | Trileptal | 253.0973 | 6.13 | | RP | POS | 236.0715(100),208.0756(60) | 2 | 1.6E+08 | 7.6E+07 | 1.5E+08 | 4.3E+07 | 3.34E-01 | -0.13 | 0.7 |
| RP_POS_152_05675a0_679 | | 152.0568 | 0.68 | | RP | POS | | | 1818298 | 788792 | 1719680 | 708808 | 3.34E-01 | -0.13 | 0.74 |
| RP_POS_343_22433a9_939 | | 343.2243 | 9.94 | | RP | POS | 343.222(100) | | 2465830 | 2773499 | 3122316 | 6253117 | 3.35E-01 | 0.15 | 0.77 |
| RP_POS_716_10101a11_363 | | 716.1010 | 11.36 | | RP | POS | | | 184416 | 98100.1 | 200913 | 143689 | 3.35E-01 | 0.14 | 0.59 |
| RP_POS_238_06683a15_244 | | 238.0668 | 15.24 | | RP | POS | 238.0656(100) | | 4339756 | 2489452 | 4714325 | 3097794 | 3.35E-01 | 0.13 | 0.68 |
| RP_NEG_275_82419a15_51 | | 275.8242 | 15.51 | | RP | NEG | | | 520907 | 46964.2 | 527323 | 50053.7 | 3.35E-01 | 0.13 | 0.75 |
| RP_POS_1021_62506a10_301 | | 1021.6251 | 10.30 | | RP | POS | | | 3169089 | 1457071 | 3374912 | 1643260 | 3.35E-01 | 0.13 | 0.82 |
| RP_POS_272_25824a8_815 | | 272.2582 | 8.82 | | RP | POS | | | 765595 | 146829 | 784058 | 133973 | 3.35E-01 | 0.13 | 0.79 |
| RP_POS_273_22159a10_435 | | 273.2216 | 10.44 | | RP | POS | | | 867859 | 73980.9 | 877906 | 77822.7 | 3.35E-01 | 0.13 | 0.89 |
| RP_POS_409_18771a2_884 | | 409.1877 | 2.88 | | RP | POS | | | 819030 | 367310 | 868152 | 376475 | 3.35E-01 | 0.13 | 0.72 |
| RP_NEG_419_2403a11_592 | | 419.2403 | 11.59 | | RP | NEG | | | 2422890 | 564274 | 2349076 | 555068 | 3.35E-01 | -0.13 | 1.31 |
| RP_POS_292_92435a15_41 | | 292.9244 | 15.41 | | RP | POS | | | 1645982 | 1186192 | 1491279 | 1158459 | 3.35E-01 | -0.13 | 0.95 |
| RP_POS_384_745a12_583 | | 384.7450 | 12.58 | | RP | POS | | | 1024239 | 777433 | 927559 | 693815 | 3.35E-01 | -0.13 | 0.88 |
| RP_POS_502_35989a7_166 | | 502.3599 | 7.17 | | RP | POS | | | 762984 | 593343 | 691286 | 497164 | 3.35E-01 | -0.13 | 0.67 |
| RP_POS_582_89313a9_967 | | 582.8931 | 9.97 | | RP | POS | | | 2771798 | 1304786 | 2594343 | 1377116 | 3.35E-01 | -0.13 | 0.73 |
| RP_POS_694_81268a11_379 | | 694.8127 | 11.38 | | RP | POS | | | 283288 | 122611 | 305359 | 197448 | 3.36E-01 | 0.14 | 0.59 |
| HILIC_NEG_157_90082a6_136 | | 157.9008 | 6.14 | | HILIC | NEG | | | 1902870 | 613712 | 1993989 | 750918 | 3.36E-01 | 0.13 | 0.8 |
| HILIC_NEG_280_62186a2_071 | | 280.6219 | 2.07 | | HILIC | NEG | | | 159168 | 177326 | 185038 | 211014 | 3.36E-01 | 0.13 | 0.93 |
| RP_NEG_883_54211a12_802 | | 883.5421 | 12.80 | | RP | NEG | | | 1708267 | 303455 | 1748464 | 306489 | 3.36E-01 | 0.13 | 1.04 |
| RP_POS_1074_69312a10_929 | | 1074.6931 | 10.93 | | RP | POS | | | 323606 | 356320 | 371279 | 366822 | 3.36E-01 | 0.13 | 0.9 |
| RP_POS_177_12724a7_769 | | 177.1272 | 7.77 | | RP | POS | | | 1395456 | 76458.8 | 1406065 | 83878.7 | 3.36E-01 | 0.13 | 0.64 |
| RP_POS_418_19308a0_684 | | 418.1931 | 0.68 | | RP | POS | | | 1330787 | 871489 | 1458605 | 1048059 | 3.36E-01 | 0.13 | 1.15 |
| RP_POS_655_78992a11_387 | | 655.7899 | 11.39 | | RP | POS | | | 193392 | 90599.9 | 206798 | 110215 | 3.36E-01 | 0.13 | 0.7 |
| HILIC_POS_422_31085a1_348 | | 422.3109 | 1.35 | | HILIC | POS | | | 185076 | 74779.2 | 175515 | 70698.8 | 3.36E-01 | -0.13 | 0.74 |
| HILIC_POS_59_06136a4_458 | | 59.0614 | 4.46 | | HILIC | POS | | | 1.5E+07 | 7138516 | 1.4E+07 | 7832954 | 3.36E-01 | -0.13 | 0.72 |
| RP_NEG_174_95609a15_354 | | 174.9561 | 15.35 | | RP | NEG | | | 4E+07 | 4502950 | 3.9E+07 | 5661474 | 3.36E-01 | -0.13 | 0.62 |
| RP_NEG_308_93793a0_68 | | 308.9379 | 0.68 | | RP | NEG | | | 340027 | 72475.7 | 331550 | 56537.3 | 3.36E-01 | -0.13 | 0.88 |
| RP_NEG_617_4447a14_035 | | 617.4447 | 14.04 | | RP | NEG | | | 580687 | 79628.1 | 571261 | 64087.4 | 3.36E-01 | -0.13 | 0.85 |
| RP_NEG_829_18048a9_685 | | 829.1805 | 9.69 | | RP | NEG | | | 3085704 | 394205 | 3038171 | 329512 | 3.36E-01 | -0.13 | 0.82 |
| RP_POS_264_14835a9_208 | | 264.1484 | 9.21 | | RP | POS | | | 734309 | 875997 | 627901 | 747467 | 3.36E-01 | -0.13 | 1.09 |
| RP_POS_327_14099a6_622 | | 327.1410 | 6.62 | | RP | POS | | | 291654 | 149385 | 273387 | 128837 | 3.36E-01 | -0.13 | 0.94 |
| HILIC_POS_194_03259a6_786 | | 194.0326 | 6.79 | | HILIC | POS | | | 295299 | 147829 | 280530 | 68386.6 | 3.36E-01 | -0.14 | 0.61 |
| RP_POS_413_1217a6_552 | | 413.1217 | 6.55 | | RP | POS | | | 139736 | 66932 | 179219 | 402677 | 3.37E-01 | 0.17 | 0.87 |
| HILIC_POS_202_17978a0_727 | | 202.1798 | 0.73 | | HILIC | POS | | | 384570 | 485051 | 451655 | 530144 | 3.37E-01 | 0.13 | 0.74 |
| RP_NEG_310_93109a15_324 | | 310.9311 | 15.32 | | RP | NEG | | | 1684924 | 147837 | 1704339 | 147881 | 3.37E-01 | 0.13 | 0.7 |
| RP_POS_1047_73486a10_928 | | 1047.7349 | 10.93 | | RP | POS | | | 2E+07 | 1.2E+07 | 2.1E+07 | 1.3E+07 | 3.37E-01 | 0.13 | 0.83 |
| RP_POS_420_7634a14_108 | | 420.7634 | 14.11 | | RP | POS | | | 2221681 | 1498678 | 2454359 | 1970926 | 3.37E-01 | 0.13 | 1.02 |
| HILIC_POS_105_04275a5_435 | | 105.0428 | 5.44 | | HILIC | POS | | | 667315 | 416035 | 620138 | 302517 | 3.37E-01 | -0.13 | 0.91 |
| RP_NEG_297_24356a10_281 | | 297.2436 | 10.28 | | RP | NEG | | | 2675138 | 386001 | 2624746 | 381411 | 3.37E-01 | -0.13 | 1.16 |
| RP_POS_163_07539a6_127 | | 163.0754 | 6.13 | | RP | POS | | | 7361447 | 1002080 | 7498447 | 1079022 | 3.38E-01 | 0.13 | 0.84 |
| RP_POS_548_37097a10_748 | | 548.3710 | 10.75 | | RP | POS | | | 1E+07 | 2824299 | 1.1E+07 | 3172643 | 3.38E-01 | 0.13 | 0.79 |
| RP_POS_688_52191a10_518 | | 688.5219 | 10.52 | | RP | POS | | | 799243 | 344615 | 853453 | 463323 | 3.38E-01 | 0.13 | 0.9 |
| RP_POS_813_98737a10_304 | | 813.9874 | 10.30 | | RP | POS | | | 726738 | 212702 | 757350 | 249964 | 3.38E-01 | 0.13 | 0.71 |
| RP_NEG_440_9902a0_679 | | 440.9902 | 0.68 | | RP | NEG | | | 197480 | 55650 | 190490 | 51215.6 | 3.38E-01 | -0.13 | 0.7 |
| RP_NEG_743_16327a9_09 | | 743.1633 | 9.09 | | RP | NEG | | | 695933 | 304158 | 655789 | 307607 | 3.38E-01 | -0.13 | 0.76 |
| RP_POS_304_24771a9_461 | | 304.2477 | 9.46 | | RP | POS | | | 757047 | 514151 | 691722 | 484767 | 3.38E-01 | -0.13 | 0.72 |
| RP_POS_397_14703a6_563 | | 397.1470 | 6.56 | | RP | POS | | | 2843392 | 611073 | 2769512 | 519710 | 3.38E-01 | -0.13 | 1.24 |
| RP_POS_431_13544a9_207 | | 431.1354 | 9.21 | | RP | POS | | | 409057 | 465675 | 354820 | 364343 | 3.38E-01 | -0.13 | 0.88 |
| RP_POS_488_39441a11_904 | | 488.3944 | 11.90 | | RP | POS | | | 8826451 | 2.1E+07 | 6526197 | 1.5E+07 | 3.38E-01 | -0.13 | 0.72 |
| RP_POS_279_9906a3_532 | | 279.9906 | 3.53 | | RP | POS | | | 256038 | 262205 | 299723 | 383594 | 3.39E-01 | 0.14 | 0.78 |
| RP_POS_154_12262a6_27 | | 154.1226 | 6.27 | | RP | POS | | | 270223 | 98754.6 | 284249 | 113684 | 3.39E-01 | 0.13 | 0.67 |
| RP_POS_211_13367a6_597 | | 211.1337 | 6.60 | | RP | POS | | | 1.2E+08 | 4496776 | 1.2E+08 | 4910529 | 3.39E-01 | 0.13 | 1.1 |
| RP_POS_500_36029a9_597 | | 500.3603 | 9.60 | | RP | POS | | | 3954362 | 1959651 | 4270668 | 2741385 | 3.39E-01 | 0.13 | 0.83 |
| RP_POS_232_15408a1_737 | isomer of butanoylcarnitine | 232.1541 | 1.74 | | RP | POS | 85.0291(100), 232.1542(40) | 2 | 5799574 | 3193900 | 5430061 | 2472869 | 3.39E-01 | -0.13 | 0.82 |
| HILIC_NEG_135_03137a1_38 | Hypoxanthine | 135.0314 | 1.38 | | HILIC | NEG | 134.8951(100), 92.0254(35) | 1 | 4736639 | 2444113 | 5047288 | 2310775 | 3.39E-01 | 0.13 | 0.68 |
| HILIC_POS_682_36481a0_682 | | 682.3648 | 0.68 | | HILIC | POS | | | 195582 | 65843.6 | 187487 | 58209.3 | 3.39E-01 | -0.13 | 0.78 |
| RP_NEG_473_21164a11_36 | | 473.2116 | 11.36 | | RP | NEG | | | 130710 | 43425.1 | 124872 | 45654.5 | 3.39E-01 | -0.13 | 1.22 |
| RP_POS_329_17523a5_591 | | 329.1752 | 5.59 | | RP | POS | | | 387653 | 95024.1 | 375528 | 90554.9 | 3.39E-01 | -0.13 | 0.65 |
| RP_POS_637_46942a11_69 | | 637.4694 | 11.69 | | RP | POS | | | 478064 | 648658 | 407116 | 432206 | 3.39E-01 | -0.13 | 0.59 |
| HILIC_NEG_89_02399a5_122 | | 89.0240 | 5.12 | | HILIC | NEG | 89.0237(100),71.0128(9) | | 2E+07 | 5245540 | 2.1E+07 | 9099816 | 3.40E-01 | 0.14 | 0.63 |
| RP_NEG_180_98949a4_819 | | 180.9895 | 4.82 | | RP | NEG | | | 2980260 | 860265 | 3095837 | 904250 | 3.40E-01 | 0.13 | 0.87 |
| RP_NEG_186_11391a4_61 | | 186.1139 | 4.61 | | RP | NEG | | | 350602 | 25026.3 | 354263 | 30321.3 | 3.40E-01 | 0.13 | 1.01 |
| RP_POS_191_1069a7_695 | | 191.1069 | 7.70 | | RP | POS | | | 2363817 | 194800 | 2393687 | 253498 | 3.40E-01 | 0.13 | 0.68 |
| RP_POS_314_23251a6_901 | | 314.2325 | 6.90 | | RP | POS | | | 4790262 | 3209861 | 5277280 | 4120551 | 3.40E-01 | 0.13 | 0.73 |
| RP_POS_428_17096a11_21 | | 428.1710 | 11.21 | | RP | POS | | | 405683 | 352674 | 453744 | 380095 | 3.40E-01 | 0.13 | 1.05 |
| RP_POS_506_36172a10_792 | | 506.3617 | 10.79 | | RP | POS | | | 2745033 | 803689 | 2852230 | 836417 | 3.40E-01 | 0.13 | 0.72 |
| RP_POS_162_07611a10_563 | | 162.0761 | 10.56 | | RP | POS | | | 4983118 | 2059742 | 4727742 | 1860004 | 3.40E-01 | -0.13 | 0.67 |
| RP_POS_229_1198a7_3 | | 229.1198 | 7.30 | | RP | POS | | | 572366 | 160657 | 551748 | 155444 | 3.40E-01 | -0.13 | 0.77 |
| HILIC_POS_257_1358a4_432 | | 257.1358 | 4.43 | | HILIC | POS | | | 65245 | 78640.5 | 57703.9 | 30259.9 | 3.40E-01 | -0.14 | 0.96 |
| RP_POS_466_29858a8_059 | | 466.2986 | 8.06 | | RP | POS | | | 490458 | 549131 | 602585 | 1052904 | 3.41E-01 | 0.14 | 1.12 |
| RP_NEG_277_21762a10_715 | | 277.2176 | 10.72 | | RP | NEG | | | 1900547 | 1484323 | 2097338 | 1532317 | 3.41E-01 | 0.13 | 0.99 |
| RP_POS_127_03476a15_507 | | 127.0348 | 15.51 | | RP | POS | | | 3124247 | 1034162 | 3267863 | 1155999 | 3.41E-01 | 0.13 | 0.83 |
| RP_POS_514_28448a5_973 | | 514.2845 | 5.97 | | RP | POS | | | 1397992 | 73996.5 | 1407747 | 75702.4 | 3.41E-01 | 0.13 | 0.79 |
| RP_POS_834_59802a14_445 | | 834.5980 | 14.45 | | RP | POS | | | 1.6E+07 | 1E+07 | 1.8E+07 | 9516443 | 3.41E-01 | 0.13 | 0.94 |
| RP_POS_804_55322a12_462 | PC 38:7 (20:5_18:2) | 804.5532 | 12.46 | | RP | POS | 184.0754(100), 86.0954(20) | 1 | 8741600 | 5840000 | 7995621 | 5614179 | 3.41E-01 | -0.13 | 0.74 |

**Supplemantary table 1**: The non-targeted metabolomics results ordered by the VIP value (p value < 0.0003 are shaded dark green, p value < 0.05 are shaded light green, d values below 0 are blue and above 0 are red)

| | | | | | | | | | Cases | | Controls | | Welch test | Cohen's | |
|---|---|---|---|---|---|---|---|---|---|---|---|---|---|---|---|
| Feature_ID | Metabolite_name | Average_Mz | Average_RT_min_ | HMDB ID | RP_HILIC | POS_NEG | Cureted_MSMS | ID_level | Mean | SD | Mean2 | SD3 | p | d | VIP |
| HILIC_NEG_216_9808a0_829 | | 216.9808 | 0.83 | | HILIC | NEG | | | 172609 | 55166 | 165947 | 47331.8 | 3.41E-01 | -0.13 | 0.82 |
| RP_NEG_725_49487a11_592 | | 725.4949 | 11.59 | | RP | NEG | | | 469372 | 138217 | 450500 | 150024 | 3.41E-01 | -0.13 | 1.25 |
| RP_NEG_844_56366a13_945 | | 844.5637 | 13.95 | | RP | NEG | | | 1346634 | 658539 | 1260416 | 665248 | 3.41E-01 | -0.13 | 0.69 |
| RP_POS_539_35559a9_459 | | 539.3556 | 9.46 | | RP | POS | | | 1.2E+07 | 2E+07 | 1E+07 | 1.5E+07 | 3.41E-01 | -0.13 | 0.88 |
| HILIC_NEG_114_98908a7_409 | | 114.9891 | 7.41 | | HILIC | NEG | | | 420464 | 84605.8 | 431027 | 78294.3 | 3.42E-01 | 0.13 | 0.69 |
| RP_NEG_248_96094a11_549 | | 248.9609 | 11.55 | | RP | NEG | | | 1598192 | 543737 | 1670224 | 561340 | 3.42E-01 | 0.13 | 0.59 |
| RP_POS_304_2605a13_296 | | 304.2605 | 13.30 | | RP | POS | | | 1388391 | 1115076 | 1558267 | 1448516 | 3.42E-01 | 0.13 | 0.96 |
| RP_POS_390_28503a10_253 | | 390.2850 | 10.25 | | RP | POS | | | 1.3E+07 | 6443257 | 1.4E+07 | 6323567 | 3.42E-01 | 0.13 | 1.12 |
| RP_POS_405_33444a10_787 | | 405.3344 | 10.79 | | RP | POS | | | 220240 | 120813 | 235463 | 113618 | 3.42E-01 | 0.13 | 0.74 |
| RP_POS_631_27399a11_366 | | 631.2740 | 11.37 | | RP | POS | | | 138398 | 75868 | 150898 | 110275 | 3.42E-01 | 0.13 | 0.71 |
| HILIC_NEG_103_04003a0_706 | 3R-hydroxy-butanoic acid | 103.0400 | 0.71 | | HILIC | NEG | 59.0132(100), 103.0399(15) | 1 | 1925437 | 1815835 | 2204221 | 2381843 | 3.42E-01 | 0.13 | 0.9 |
| HILIC_POS_421_29129a3_294 | | 421.2913 | 3.29 | | HILIC | POS | | | 369628 | 533773 | 307824 | 419501 | 3.42E-01 | -0.13 | 0.86 |
| RP_NEG_282_98187a8_9 | | 282.9819 | 8.90 | | RP | NEG | | | 835611 | 78591.9 | 825707 | 74087.2 | 3.42E-01 | -0.13 | 0.65 |
| RP_NEG_374_69998a15_458 | | 374.7000 | 15.46 | | RP | NEG | | | 917502 | 138318 | 901305 | 111323 | 3.42E-01 | -0.13 | 0.78 |
| RP_NEG_298_8735a15_409 | | 298.8735 | 15.41 | | RP | NEG | | | 357558 | 147288 | 378251 | 169027 | 3.43E-01 | 0.13 | 0.76 |
| RP_POS_252_99699a8_09 | | 252.9970 | 8.09 | | RP | POS | | | 502294 | 211099 | 529006 | 201535 | 3.43E-01 | 0.13 | 0.67 |
| RP_POS_286_23773a10_069 | | 286.2377 | 10.07 | | RP | POS | | | 642183 | 181965 | 665645 | 180071 | 3.43E-01 | 0.13 | 0.8 |
| RP_NEG_178_05095a3_537 | Hippuric acid | 178.0510 | 3.54 | | RP | NEG | 134.0607(100), 178.0508(60) | 2 | 6187801 | 5567667 | 7081862 | 7838975 | 3.43E-01 | 0.13 | 0.75 |
| HILIC_POS_118_08641a3_939 | Valine | 118.0864 | 3.94 | | HILIC | POS | 118.0865(100) | 2 | 1.1E+07 | 1.3E+07 | 9985970 | 5185372 | 3.43E-01 | -0.14 | 0.63 |
| HILIC_POS_184_0732a0_593 | | 184.0732 | 0.59 | | HILIC | POS | 184.0743(100) | | 967248 | 77272.6 | 957270 | 76455.3 | 3.43E-01 | -0.13 | 0.8 |
| HILIC_POS_185_12842a0_612 | | 185.1284 | 0.61 | | HILIC | POS | | | 1E+07 | 3616066 | 9761844 | 3294754 | 3.43E-01 | -0.13 | 0.6 |
| HILIC_POS_454_29337a1_901 | | 454.2934 | 1.90 | | HILIC | POS | | | 758440 | 651757 | 677431 | 600348 | 3.43E-01 | -0.13 | 0.87 |
| RP_POS_217_08836a0_63 | | 217.0884 | 0.63 | | RP | POS | | | 2361811 | 890435 | 2245745 | 896948 | 3.43E-01 | -0.13 | 0.84 |
| RP_POS_331_08307a8_844 | | 331.0831 | 8.84 | | RP | POS | | | 2477084 | 1547268 | 2288060 | 1373523 | 3.43E-01 | -0.13 | 1.22 |
| RP_POS_376_34241a11_248 | | 376.3424 | 11.25 | | RP | POS | | | 718471 | 237296 | 690466 | 195703 | 3.43E-01 | -0.13 | 0.68 |
| HILIC_NEG_181_83771a3_233 | | 181.8377 | 3.23 | | HILIC | NEG | | | 426221 | 85503.5 | 436901 | 79721.2 | 3.44E-01 | 0.13 | 0.74 |
| RP_NEG_508_34155a10_815 | | 508.3416 | 10.82 | | RP | NEG | | | 640341 | 161096 | 662419 | 177959 | 3.44E-01 | 0.13 | 0.85 |
| RP_POS_194_00793a15_566 | | 194.0079 | 15.57 | | RP | POS | | | 1267882 | 625991 | 1355483 | 716094 | 3.44E-01 | 0.13 | 0.72 |
| RP_POS_314_93115a15_351 | | 314.9312 | 15.35 | | RP | POS | | | 223674 | 126934 | 239196 | 113318 | 3.44E-01 | 0.13 | 0.74 |
| HILIC_POS_496_34052a1_125 | LysoPC 16:0 | 496.3405 | 1.13 | | HILIC | POS | 184.0746(100) | 2 | 4.6E+07 | 1.2E+07 | 4.7E+07 | 1.2E+07 | 3.44E-01 | 0.13 | 0.86 |
| RP_POS_1183_86365a13_687 | | 1183.8637 | 13.69 | | RP | POS | | | 398551 | 315031 | 361016 | 266163 | 3.44E-01 | -0.13 | 0.86 |
| RP_POS_712_69989a11_539 | | 712.6999 | 11.54 | | RP | POS | | | 183097 | 95174.9 | 171682 | 81810.3 | 3.44E-01 | -0.13 | 0.85 |
| RP_POS_270_17496a9_456 | | 270.1750 | 9.46 | | RP | POS | | | 64641 | 71943.7 | 77508.6 | 117971 | 3.45E-01 | 0.14 | 0.9 |
| RP_POS_513_3692a15_399 | | 513.3692 | 15.40 | | RP | POS | | | 360232 | 133791 | 383889 | 215698 | 3.45E-01 | 0.14 | 0.89 |
| RP_NEG_134_89474a0_679 | | 134.8947 | 0.68 | | RP | NEG | | | 4388706 | 453818 | 4440914 | 356222 | 3.45E-01 | 0.13 | 0.56 |
| RP_POS_558_42181a9_123 | | 558.4218 | 9.12 | | RP | POS | | | 2278284 | 1374821 | 2479798 | 1700667 | 3.45E-01 | 0.13 | 0.98 |
| RP_POS_846_54004a13_289 | | 846.5400 | 13.29 | | RP | POS | | | 3091054 | 1732849 | 3330071 | 1939143 | 3.45E-01 | 0.13 | 1.03 |
| RP_POS_150_11252a10_491 | | 150.1125 | 10.49 | | RP | POS | | | 1016210 | 458167 | 961479 | 392454 | 3.45E-01 | -0.13 | 0.64 |
| RP_POS_632_93622a10_618 | | 632.9362 | 10.62 | | RP | POS | | | 452769 | 300944 | 412804 | 315878 | 3.45E-01 | -0.13 | 0.81 |
| RP_POS_718_5296a9_676 | | 718.5296 | 9.68 | | RP | POS | | | 4692627 | 1799611 | 4463517 | 1746070 | 3.45E-01 | -0.13 | 0.66 |
| RP_POS_271_11554a4_536 | | 271.1155 | 4.54 | | RP | POS | | | 1418640 | 1095074 | 1314243 | 434816 | 3.45E-01 | -0.14 | 0.78 |
| RP_POS_355_18231a10_683 | | 355.1823 | 10.68 | | RP | POS | | | 106276 | 43780.5 | 114074 | 71488.5 | 3.46E-01 | 0.14 | 0.75 |
| HILIC_NEG_158_93909a3_166 | | 158.9391 | 3.17 | | HILIC | NEG | | | 2542489 | 265928 | 2579048 | 297426 | 3.46E-01 | 0.13 | 0.8 |
| HILIC_POS_153_09093a1_352 | | 153.0909 | 1.35 | | HILIC | POS | | | 171374 | 91584.2 | 183777 | 99842.2 | 3.46E-01 | 0.13 | 0.82 |
| HILIC_POS_366_26425a0_643 | | 366.2643 | 0.64 | | HILIC | POS | | | 320195 | 190947 | 346501 | 214478 | 3.46E-01 | 0.13 | 0.74 |
| RP_POS_250_11061a5_001 | | 250.1106 | 5.00 | | RP | POS | | | 1136693 | 974539 | 1263290 | 985715 | 3.46E-01 | 0.13 | 0.76 |
| RP_POS_487_27994a10_931 | | 487.2799 | 10.93 | | RP | POS | | | 428589 | 126850 | 446856 | 152771 | 3.46E-01 | 0.13 | 0.64 |
| HILIC_POS_265_01532a7_421 | | 265.0153 | 7.42 | | HILIC | POS | | | 49187.8 | 35602.4 | 45221.8 | 25979.6 | 3.46E-01 | -0.13 | 0.97 |
| RP_NEG_215_12865a7_416 | | 215.1287 | 7.42 | | RP | NEG | | | 1473628 | 145930 | 1454941 | 144061 | 3.46E-01 | -0.13 | 1.2 |
| RP_NEG_566_27771a9_168 | | 566.2777 | 9.17 | | RP | NEG | | | 359950 | 529206 | 305909 | 294906 | 3.46E-01 | -0.13 | 0.61 |
| RP_POS_281_24744a10_158 | | 281.2474 | 10.16 | | RP | POS | | | 4741626 | 656488 | 4657126 | 654381 | 3.46E-01 | -0.13 | 1.03 |
| RP_POS_299_20053a10_676 | | 299.2005 | 10.68 | | RP | POS | | | 1813876 | 362403 | 1769635 | 325759 | 3.46E-01 | -0.13 | 0.81 |
| RP_POS_596_91028a10_329 | | 596.9103 | 10.33 | | RP | POS | | | 2130851 | 947992 | 2012334 | 893344 | 3.46E-01 | -0.13 | 0.6 |
| RP_POS_768_58966a13_678 | | 768.5897 | 13.68 | | RP | POS | | | 6.9E+07 | 2.1E+07 | 6.7E+07 | 1.9E+07 | 3.46E-01 | -0.13 | 0.72 |
| HILIC_POS_154_08636a0_544 | | 154.0864 | 0.54 | | HILIC | POS | | | 3380186 | 3.4E+07 | 418065 | 188459 | 3.46E-01 | -0.17 | 0.58 |
| RP_NEG_541_26642a6_889 | | 541.2664 | 6.89 | | RP | NEG | | | 929821 | 508895 | 1002323 | 605189 | 3.47E-01 | 0.13 | 0.61 |
| RP_NEG_597_30615a11_588 | | 597.3062 | 11.59 | | RP | NEG | | | 506534 | 491986 | 570857 | 505717 | 3.47E-01 | 0.13 | 0.74 |
| HILIC_POS_164_14349a0_702 | | 164.1435 | 0.70 | | HILIC | POS | | | 1149265 | 566088 | 1080312 | 509552 | 3.47E-01 | -0.13 | 0.92 |
| RP_NEG_251_09268a7_719 | | 251.0927 | 7.72 | | RP | NEG | | | 5320964 | 187875 | 5297289 | 180362 | 3.47E-01 | -0.13 | 0.67 |
| RP_POS_308_65952a10_937 | | 308.6595 | 10.94 | | RP | POS | | | 424461 | 235277 | 395073 | 222724 | 3.47E-01 | -0.13 | 0.88 |
| RP_POS_385_29269a10_523 | | 385.2927 | 10.52 | | RP | POS | | | 1872704 | 454206 | 1814618 | 448579 | 3.47E-01 | -0.13 | 1.1 |
| RP_POS_563_23395a8_842 | | 563.2340 | 8.84 | | RP | POS | | | 158896 | 115402 | 144759 | 104772 | 3.47E-01 | -0.13 | 0.73 |
| RP_POS_675_6759a11_656 | | 675.6759 | 11.66 | | RP | POS | | | 7129103 | 1.6E+07 | 5335829 | 1.2E+07 | 3.47E-01 | -0.13 | 0.65 |
| RP_POS_171_11284a2_808 | | 171.1128 | 2.81 | | RP | POS | 126.0913(100), 171.1498(30) | | 2236583 | 2.2E+07 | 318640 | 1382321 | 3.47E-01 | -0.16 | 0.57 |
| RP_POS_301_21375a10_716 | | 301.2138 | 10.72 | | RP | POS | 301.2046(100) | | 5709644 | 3905161 | 6227149 | 4128932 | 3.48E-01 | 0.13 | 0.96 |
| RP_POS_184_05824a14_251 | | 184.0582 | 14.25 | | RP | POS | | | 3843197 | 2150663 | 4100848 | 1874351 | 3.48E-01 | 0.13 | 0.73 |
| RP_POS_240_23196a8_964 | | 240.2320 | 8.96 | | RP | POS | | | 315054 | 49853.7 | 323095 | 71631.4 | 3.48E-01 | 0.13 | 0.74 |
| RP_POS_797_58746a13_365 | | 797.5875 | 13.37 | | RP | POS | | | 6096894 | 5916771 | 6941198 | 7075252 | 3.48E-01 | 0.13 | 0.84 |
| RP_NEG_478_29401a10_48 | LysoPE 18:1 | 478.2940 | 10.48 | | RP | NEG | 281(100), 214.049(2) | 2 | 1711071 | 616996 | 1798252 | 727107 | 3.48E-01 | 0.13 | 0.73 |
| HILIC_NEG_243_06262a3_465 | | 243.0626 | 3.47 | | HILIC | NEG | | | 428321 | 84225.9 | 419803 | 45929.6 | 3.48E-01 | -0.13 | 0.63 |
| HILIC_POS_427_26648a7_098 | | 427.2665 | 7.10 | | HILIC | POS | | | 303241 | 48780.8 | 297575 | 39806.3 | 3.48E-01 | -0.13 | 0.63 |
| RP_NEG_511_47327a11_142 | | 511.4733 | 11.14 | | RP | NEG | | | 613790 | 226365 | 584709 | 226663 | 3.48E-01 | -0.13 | 1.29 |
| RP_NEG_624_20459a10_279 | | 624.2046 | 10.28 | | RP | NEG | | | 489242 | 125156 | 472310 | 137053 | 3.48E-01 | -0.13 | 0.73 |
| RP_NEG_657_50775a11_592 | | 657.5078 | 11.59 | | RP | NEG | | | 647258 | 231681 | 617443 | 232478 | 3.48E-01 | -0.13 | 1.29 |
| RP_POS_646_50739a13_541 | | 646.5074 | 13.54 | | RP | POS | | | 587670 | 342113 | 544032 | 337626 | 3.48E-01 | -0.13 | 0.86 |
| RP_POS_762_53693a10_487 | | 762.5369 | 10.49 | | RP | POS | | | 843222 | 435880 | 776669 | 578491 | 3.48E-01 | -0.13 | 0.83 |
| RP_POS_194_0965a5_759 | 1-(pyrimidin-2-yl)pyrrolidine-2-carboxylic acid | 194.0965 | 5.76 | | RP | POS | 194.0956(100) | 2 | 3685040 | 1.6E+07 | 2250281 | 3200587 | 3.48E-01 | -0.15 | 0.6 |
| RP_POS_254_13857a6_77 | | 254.1386 | 6.77 | | RP | POS | | | 1011730 | 8431135 | 265857 | 1411450 | 3.48E-01 | -0.15 | 0.61 |
| RP_POS_325_12549a6_227 | Demethyleneberberine | 325.1255 | 6.23 | | RP | POS | 325.1281(100) | 2 | 6831601 | 1469939 | 6655097 | 1291047 | 3.49E-01 | -0.13 | 1.25 |
| RP_NEG_362_98615a7_786 | | 362.9862 | 7.79 | | RP | NEG | | | 2036931 | 208029 | 2062050 | 185539 | 3.49E-01 | 0.13 | 0.68 |
| RP_POS_161_13242a8_481 | | 161.1324 | 8.48 | | RP | POS | | | 4079571 | 524386 | 4150281 | 573213 | 3.49E-01 | 0.13 | 0.71 |
| RP_POS_477_66287a10_233 | | 477.6629 | 10.23 | | RP | POS | | | 427385 | 236201 | 459401 | 260918 | 3.49E-01 | 0.13 | 0.72 |
| RP_POS_481_4736a12_468 | | 481.4736 | 12.47 | | RP | POS | | | 315630 | 81502.2 | 326410 | 86343.6 | 3.49E-01 | 0.13 | 0.69 |
| RP_POS_133_04971a9_211 | Glutaric acid | 133.0497 | 9.21 | | RP | POS | 87.0448(100),115.0392(20) | 2 | 920261 | 506367 | 856346 | 491575 | 3.49E-01 | -0.13 | 0.76 |
| HILIC_POS_175_10779a3_204 | | 175.1078 | 3.20 | | HILIC | POS | | | 706181 | 405383 | 659741 | 321860 | 3.49E-01 | -0.13 | 0.59 |
| HILIC_POS_573_84851a0_936 | | 573.8485 | 0.94 | | HILIC | POS | | | 404528 | 115654 | 389739 | 115304 | 3.49E-01 | -0.13 | 0.57 |
| RP_POS_159_02647a0_552 | | 159.0265 | 0.55 | | RP | POS | | | 2461346 | 511961 | 2388964 | 606466 | 3.49E-01 | -0.13 | 0.61 |
| RP_POS_242_21135a11_113 | | 242.2114 | 11.11 | | RP | POS | | | 756995 | 1141828 | 632189 | 804074 | 3.49E-01 | -0.13 | 0.71 |
| RP_POS_323_25552a10_195 | | 323.2555 | 10.20 | | RP | POS | | | 9354253 | 1352381 | 9187795 | 1253587 | 3.49E-01 | -0.13 | 1.07 |
| RP_POS_453_96225a15_396 | | 453.9623 | 15.40 | | RP | POS | | | 1003021 | 355625 | 959628 | 323076 | 3.49E-01 | -0.13 | 0.58 |
| RP_POS_559_3241a9_449 | | 559.3241 | 9.45 | | RP | POS | | | 1437566 | 1255580 | 1286976 | 1102455 | 3.49E-01 | -0.13 | 0.93 |
| RP_POS_692_28693a11_502 | | 692.2869 | 11.50 | | RP | POS | | | 254739 | 185208 | 233779 | 142851 | 3.49E-01 | -0.13 | 0.76 |
| RP_POS_246_17026a3_026 | | 246.1703 | 3.03 | | RP | POS | 85.0291(100), 246.1695(45) | | 2602403 | 1808261 | 2813707 | 1508908 | 3.50E-01 | 0.13 | 0.63 |
| HILIC_NEG_478_29337a1_814 | | 478.2934 | 1.81 | | HILIC | NEG | | | 259785 | 97928.5 | 273490 | 114332 | 3.50E-01 | 0.13 | 0.77 |
| HILIC_POS_169_0457a7_418 | | 169.0457 | 7.42 | | HILIC | POS | | | 658417 | 155872 | 679976 | 178785 | 3.50E-01 | 0.13 | 0.71 |
| RP_NEG_266_94604a0_916 | | 266.9460 | 0.92 | | RP | NEG | | | 500727 | 96305.8 | 513411 | 101353 | 3.50E-01 | 0.13 | 0.54 |
| RP_POS_253_14098a4_008 | | 253.1410 | 4.01 | | RP | POS | | | 331236 | 121131 | 347760 | 135534 | 3.50E-01 | 0.13 | 0.89 |
| HILIC_POS_170_11749a3_248 | | 170.1175 | 3.25 | | HILIC | POS | | | 334691 | 181090 | 312323 | 169387 | 3.50E-01 | -0.13 | 0.58 |
| RP_NEG_499_0896a8_833 | | 499.0896 | 8.83 | | RP | NEG | | | 1386662 | 79788.2 | 1376025 | 86038.5 | 3.50E-01 | -0.13 | 0.93 |
| RP_POS_436_27219a14_117 | | 436.2722 | 14.12 | | RP | POS | | | 133387 | 122275 | 120202 | 83289 | 3.50E-01 | -0.13 | 0.79 |
| RP_NEG_209_06667a0_587 | | 209.0667 | 0.59 | | RP | NEG | | | 6426064 | 1634407 | 6655775 | 1930609 | 3.51E-01 | 0.13 | 0.6 |
| RP_POS_180_15919a5_732 | | 180.1592 | 5.73 | | RP | POS | | | 2667672 | 450610 | 2729134 | 505649 | 3.51E-01 | 0.13 | 0.58 |
| RP_POS_268_15405a7_658 | | 268.1541 | 7.66 | | RP | POS | | | 8770702 | 959727 | 8895908 | 999400 | 3.51E-01 | 0.13 | 1.18 |
| RP_POS_315_19275a9_788 | | 315.1928 | 9.79 | | RP | POS | | | 4814447 | 357042 | 4859958 | 355800 | 3.51E-01 | 0.13 | 1.06 |
| RP_POS_343_16086a10_961 | | 343.1609 | 10.96 | | RP | POS | | | 446722 | 268776 | 481455 | 275071 | 3.51E-01 | 0.13 | 1.09 |
| RP_POS_746_56964a13_387 | | 746.5696 | 13.39 | | RP | POS | | | 2.2E+07 | 7505880 | 2.3E+07 | 8515001 | 3.51E-01 | 0.13 | 0.82 |
| RP_POS_293_20847a9_558 | 8-[3-oxo-2-[(E)-pent-2-enyl]cyclopenten-1-yl]octanoic acid | 293.2085 | 9.56 | | RP | POS | 293.2105(100) | 2 | 6441374 | 763289 | 6346663 | 722250 | 3.51E-01 | -0.13 | 1.07 |
| RP_NEG_370_70599a15_456 | | 370.7060 | 15.46 | | RP | NEG | | | 1972032 | 260750 | 1944608 | 166293 | 3.51E-01 | -0.13 | 0.81 |
| RP_NEG_433_11292a11_142 | | 433.1129 | 11.14 | | RP | NEG | | | 628228 | 59419.9 | 620632 | 59660.2 | 3.51E-01 | -0.13 | 0.8 |
| RP_POS_285_22104a10_883 | | 285.2210 | 10.88 | | RP | POS | | | 1163580 | 340477 | 1124096 | 279930 | 3.51E-01 | -0.13 | 0.9 |
| RP_POS_395_21875a9_209 | | 395.2188 | 9.21 | | RP | POS | | | 998133 | 1780858 | 805306 | 1236825 | 3.51E-01 | -0.13 | 0.92 |
| RP_POS_429_37305a13_151 | | 429.3731 | 13.15 | | RP | POS | | | 7147812 | 1235371 | 6981372 | 1358669 | 3.51E-01 | -0.13 | 0.63 |
| RP_POS_558_50897a14_241 | | 558.5090 | 14.24 | | RP | POS | | | 244983 | 288725 | 213361 | 207109 | 3.51E-01 | -0.13 | 0.6 |
| RP_NEG_437_16177a7_949 | | 437.1618 | 7.95 | | RP | NEG | | | 1488626 | 1061677 | 1615149 | 929993 | 3.52E-01 | 0.13 | 0.66 |
| RP_NEG_520_91101a12_188 | | 520.9110 | 12.19 | | RP | NEG | | | 1376415 | 642609 | 1460652 | 676334 | 3.52E-01 | 0.13 | 0.79 |
| RP_POS_216_01846a2_737 | | 216.0185 | 2.74 | | RP | POS | | | 733960 | 230823 | 762202 | 214098 | 3.52E-01 | 0.13 | 0.79 |
| RP_POS_333_15103a10_715 | | 333.1510 | 10.72 | | RP | POS | | | 1168565 | 801191 | 1277305 | 894893 | 3.52E-01 | 0.13 | 0.96 |
| RP_POS_378_21136a7_605 | | 378.2114 | 7.61 | | RP | POS | | | 255454 | 61808 | 264795 | 81810.9 | 3.52E-01 | 0.13 | 0.67 |
| RP_POS_521_68768a10_186 | | 521.6877 | 10.19 | | RP | POS | | | 585265 | 328785 | 626475 | 319399 | 3.52E-01 | 0.13 | 0.64 |
| HILIC_POS_391_20187a4_858 | | 391.2019 | 4.86 | | HILIC | POS | | | 235023 | 162567 | 215973 | 137888 | 3.52E-01 | -0.13 | 0.78 |
| RP_NEG_341_27008a11_51 | | 341.2701 | 11.51 | | RP | NEG | | | 1972519 | 179048 | 1948741 | 192603 | 3.52E-01 | -0.13 | 0.94 |
| RP_NEG_571_49756a14_037 | | 571.4976 | 14.04 | | RP | NEG | | | 3972324 | 174639 | 3951281 | 156461 | 3.52E-01 | -0.13 | 0.86 |
| RP_NEG_669_42932a11_142 | | 669.4293 | 11.14 | | RP | NEG | | | 269525 | 77325.2 | 259904 | 74065.9 | 3.52E-01 | -0.13 | 1.22 |
| RP_POS_148_09711a0_677 | | 148.0971 | 0.68 | | RP | POS | | | 2002802 | 397782 | 1959392 | 282966 | 3.52E-01 | -0.13 | 0.77 |
| RP_POS_162_07614a11_201 | | 162.0761 | 11.20 | | RP | POS | | | 2670645 | 1032258 | 2554975 | 789005 | 3.52E-01 | -0.13 | 0.61 |
| RP_POS_205_55859a9_458 | | 205.5586 | 9.46 | | RP | POS | | | 375887 | 382232 | 329801 | 343867 | 3.52E-01 | -0.13 | 0.71 |
| RP_NEG_425_14813a10_714 | | 425.1481 | 10.71 | | RP | NEG | | | 245734 | 148682 | 264611 | 148491 | 3.53E-01 | 0.13 | 1 |
| RP_POS_205_08569a9_524 | | 205.0857 | 9.52 | | RP | POS | | | 7999295 | 1696800 | 8260813 | 2320098 | 3.53E-01 | 0.13 | 0.94 |
| RP_POS_293_11447a7_694 | | 293.1145 | 7.69 | | RP | POS | | | 1048206 | 103095 | 1061311 | 102927 | 3.53E-01 | 0.13 | 0.77 |
| RP_POS_337_14636a10_715 | | 337.1464 | 10.72 | | RP | POS | | | 628019 | 468672 | 689169 | 492076 | 3.53E-01 | 0.13 | 0.96 |
| RP_POS_388_25391a4_152 | | 388.2539 | 4.15 | | RP | POS | | | 573158 | 259758 | 615368 | 382590 | 3.53E-01 | 0.13 | 0.64 |
| HILIC_NEG_328_81046a3_064 | | 328.8105 | 3.06 | | HILIC | NEG | | | 78525.6 | 11516.5 | 77102.2 | 10918.3 | 3.53E-01 | -0.13 | 0.95 |
| RP_NEG_466_29526a10_231 | | 466.2953 | 10.23 | | RP | NEG | | | 462274 | 128944 | 446109 | 125535 | 3.53E-01 | -0.13 | 0.81 |
| RP_NEG_638_83374a15_345 | | 638.8337 | 15.35 | | RP | NEG | | | 430923 | 25939.7 | 427423 | 28761.7 | 3.53E-01 | -0.13 | 0.77 |
| RP_NEG_701_15448a9_196 | | 701.1545 | 9.20 | | RP | NEG | | | 1.6E+07 | 1021728 | 1.6E+07 | 924273 | 3.53E-01 | -0.13 | 0.98 |
| RP_POS_312_2168a5_765 | | 312.2168 | 5.77 | | RP | POS | | | 475523 | 1304553 | 339925 | 812750 | 3.53E-01 | -0.13 | 0.74 |
| HILIC_POS_388_25397a0_61 | | 388.2540 | 0.61 | | HILIC | POS | | | 217605 | 197145 | 255941 | 367980 | 3.54E-01 | 0.14 | 0.57 |
| RP_POS_319_22656a8_886 | | 319.2266 | 8.89 | | RP | POS | | | 395091 | 347693 | 480544 | 856616 | 3.54E-01 | 0.14 | 0.63 |
| RP_POS_348_04065a15_462 | | 348.0407 | 15.46 | | RP | POS | | | 2380609 | 531275 | 2485451 | 1008774 | 3.54E-01 | 0.14 | 0.69 |
| HILIC_NEG_183_93465a2_879 | | 183.9347 | 2.88 | | HILIC | NEG | | | 1272099 | 527813 | 1345787 | 622910 | 3.54E-01 | 0.13 | 0.58 |
| HILIC_POS_208_88391a2_98 | | 208.8839 | 2.98 | | HILIC | POS | | | 1634044 | 320809 | 1673982 | 309437 | 3.54E-01 | 0.13 | 0.67 |
| RP_POS_228_00027a15_432 | | 228.0003 | 15.43 | | RP | POS | | | 382113 | 186995 | 406399 | 194869 | 3.54E-01 | 0.13 | 0.56 |
| HILIC_POS_137_04565a1_38 | Hypoxanthine | 137.0457 | 1.38 | | HILIC | POS | 137.0464(100) | 1 | 8.8E+07 | 3.5E+07 | 9.2E+07 | 3.3E+07 | 3.54E-01 | 0.13 | 0.67 |
| HILIC_POS_387_21399a3_035 | | 387.2140 | 3.04 | | HILIC | POS | | | 866411 | 287146 | 826835 | 331719 | 3.54E-01 | -0.13 | 0.77 |
| RP_NEG_415_21637a7_338 | | 415.2164 | 7.34 | | RP | NEG | | | 1875268 | 3101385 | 1510971 | 2658576 | 3.54E-01 | -0.13 | 0.72 |
| RP_POS_519_8797a10_397 | | 519.8797 | 10.40 | | RP | POS | | | 1059985 | 456382 | 999156 | 498876 | 3.54E-01 | -0.13 | 0.77 |
| RP_POS_364_24792a10_444 | | 364.2479 | 10.44 | | RP | POS | | | 242362 | 142658 | 260154 | 138807 | 3.55E-01 | 0.13 | 1.24 |
| RP_POS_505_36069a10_204 | | 505.3607 | 10.20 | | RP | POS | | | 1297316 | 566929 | 1379543 | 712315 | 3.55E-01 | 0.13 | 0.7 |
| HILIC_NEG_183_93459a3_625 | | 183.9346 | 3.63 | | HILIC | NEG | | | 2503420 | 930233 | 2392598 | 827812 | 3.55E-01 | -0.13 | 0.57 |
| HILIC_POS_428_08469a5_674 | | 428.0847 | 5.67 | | HILIC | POS | | | 60287.8 | 21177.2 | 57507.1 | 22626.6 | 3.55E-01 | -0.13 | 0.58 |
| RP_POS_329_26907a9_855 | | 329.2691 | 9.86 | | RP | POS | | | 689418 | 66840.3 | 681144 | 64213.3 | 3.55E-01 | -0.13 | 0.96 |
| RP_POS_529_22443a6_968 | | 529.2244 | 6.97 | | RP | POS | | | 514487 | 147160 | 496690 | 134645 | 3.55E-01 | -0.13 | 1.27 |
| HILIC_POS_204_04713a2_858 | | 204.0471 | 2.86 | | HILIC | POS | | | 410504 | 209479 | 434905 | 178020 | 3.56E-01 | 0.13 | 0.94 |
| RP_NEG_345_20505a10_714 | | 345.2051 | 10.71 | | RP | NEG | | | 201034 | 168706 | 222138 | 165489 | 3.56E-01 | 0.13 | 0.99 |
| RP_POS_136_95419a15_448 | | 136.9542 | 15.45 | | RP | POS | | | 2618978 | 1360371 | 2800856 | 1505268 | 3.56E-01 | 0.13 | 0.9 |
| RP_POS_172_13312a6_269 | | 172.1331 | 6.27 | | RP | POS | | | 4322443 | 596491 | 4397823 | 596121 | 3.56E-01 | 0.13 | 0.89 |
| HILIC_POS_260_19666a6_56 | | 260.1967 | 6.56 | | HILIC | POS | | | 135012 | 99979.4 | 123210 | 87453.1 | 3.56E-01 | -0.13 | 0.9 |
| RP_NEG_368_70697a15_456 | | 368.7070 | 15.46 | | RP | NEG | | | 1537616 | 261134 | 1503662 | 275210 | 3.56E-01 | -0.13 | 0.7 |
| RP_NEG_824_54651a12_55 | | 824.5465 | 12.55 | | RP | NEG | | | 2678983 | 1525321 | 2471467 | 1734176 | 3.56E-01 | -0.13 | 0.6 |
| RP_POS_259_19031a8_45 | | 259.1903 | 8.45 | | RP | POS | | | 720300 | 80403.5 | 710306 | 77955.8 | 3.56E-01 | -0.13 | 0.88 |
| RP_POS_461_33606a10_25 | | 461.3361 | 10.25 | | RP | POS | | | 354175 | 218842 | 329942 | 165662 | 3.56E-01 | -0.13 | 0.76 |
| RP_POS_528_41095a9_483 | | 528.4110 | 9.48 | | RP | POS | | | 2660853 | 1297447 | 2485254 | 1464845 | 3.56E-01 | -0.13 | 0.85 |

**Supplemantary table 1**: The non-targeted metabolomics results ordered by the VIP value (p value < 0.0003 are shaded dark green, p value < 0.05 are shaded light green, d values below 0 are blue and above 0 are red)

| | | | | | | | | | Cases | | Controls | | Welch test | Cohen's | |
|---|---|---|---|---|---|---|---|---|---|---|---|---|---|---|---|
| Feature_ID | Metabolite_name | Average_Mz | Average_RT_min_ | HMDB ID | RP_HILIC | POS_NEG | Cureted_MSMS | ID_level | Mean | SD | Mean2 | SD3 | p | d | VIP |
| RP_NEG_457_23782a10_568 | | 457.2378 | 10.57 | | RP | NEG | | | 62665.2 | 144470 | 91912.7 | 285446 | 3.57E-01 | 0.14 | 0.77 |
| HILIC_NEG_134_06128a0_986 | | 134.0613 | 0.99 | | HILIC | NEG | | | 92046.9 | 89354 | 106189 | 128272 | 3.57E-01 | 0.13 | 0.75 |
| HILIC_POS_169_9857a5_36 | | 169.9857 | 5.36 | | HILIC | POS | | | 1669307 | 1859741 | 1917537 | 2058093 | 3.57E-01 | 0.13 | 0.79 |
| RP_NEG_460_83502a15_373 | | 460.8350 | 15.37 | | RP | NEG | | | 934014 | 131708 | 949991 | 122536 | 3.57E-01 | 0.13 | 0.64 |
| RP_POS_151_09663a10_658 | | 151.0966 | 10.66 | | RP | POS | | | 828788 | 247314 | 859664 | 243108 | 3.57E-01 | 0.13 | 0.8 |
| RP_POS_250_10329a2_226 | | 250.1033 | 2.23 | | RP | POS | | | 1095533 | 424342 | 1146549 | 386566 | 3.57E-01 | 0.13 | 0.68 |
| RP_POS_316_17557a7_909 | | 316.1756 | 7.91 | | RP | POS | | | 1988881 | 202757 | 2012595 | 174855 | 3.57E-01 | 0.13 | 1.09 |
| RP_POS_433_7717a14_462 | | 433.7717 | 14.46 | | RP | POS | | | 237086 | 192487 | 263136 | 218230 | 3.57E-01 | 0.13 | 0.77 |
| RP_POS_606_42255a11_424 | | 606.4226 | 11.42 | | RP | POS | | | 111074 | 63774.7 | 121549 | 96318.4 | 3.57E-01 | 0.13 | 0.7 |
| HILIC_NEG_221_8425a3_201 | | 221.8425 | 3.20 | | HILIC | NEG | | | 193324 | 56056.3 | 185823 | 62334.6 | 3.57E-01 | -0.13 | 0.58 |
| RP_POS_298_04181a15_373 | | 298.0418 | 15.37 | | RP | POS | | | 2E+07 | 6655174 | 1.9E+07 | 6948800 | 3.57E-01 | -0.13 | 0.65 |
| HILIC_NEG_181_07199a4_878 | Mannitol | 181.0720 | 4.88 | | HILIC | NEG | 180.9888(100), 101.0245(45), 71.015(30) | 2 | 477063 | 671911 | 392365 | 672016 | 3.57E-01 | -0.13 | 0.57 |
| HILIC_POS_398_21509a2_281 | | 398.2151 | 2.28 | | HILIC | POS | | | 56132.9 | 18213.1 | 59114.8 | 27521.4 | 3.58E-01 | 0.13 | 0.55 |
| RP_POS_638_28107a11_612 | | 638.2811 | 11.61 | | RP | POS | | | 94590 | 50604.5 | 101870 | 63360.5 | 3.58E-01 | 0.13 | 0.93 |
| RP_NEG_311_16934a10_727 | | 311.1693 | 10.73 | | RP | NEG | 311.1701(100) | | 9015926 | 6451640 | 8305313 | 4856918 | 3.58E-01 | -0.13 | 0.81 |
| RP_NEG_180_06668a0_684 | | 180.0667 | 0.68 | | RP | NEG | 180.0665(100), 163.0399(70), 119.0502(60) | | 1840670 | 917714 | 1744904 | 596876 | 3.58E-01 | -0.13 | 0.67 |
| HILIC_NEG_153_86879a7_021 | | 153.8688 | 7.02 | | HILIC | NEG | | | 1049596 | 481100 | 994814 | 393633 | 3.58E-01 | -0.13 | 0.57 |
| RP_POS_156_06329a10_498 | | 156.0633 | 10.50 | | RP | POS | | | 1079679 | 527628 | 1016362 | 481772 | 3.58E-01 | -0.13 | 0.57 |
| HILIC_NEG_230_98582a0_616 | | 230.9858 | 0.62 | | HILIC | NEG | | | 380381 | 104839 | 394985 | 125173 | 3.59E-01 | 0.13 | 0.54 |
| HILIC_POS_138_09151a3_105 | | 138.0915 | 3.11 | | HILIC | POS | | | 591871 | 214897 | 622904 | 271239 | 3.59E-01 | 0.13 | 0.69 |
| RP_NEG_147_92136a0_647 | | 147.9214 | 0.65 | | RP | NEG | | | 343876 | 91005.6 | 356358 | 106031 | 3.59E-01 | 0.13 | 0.56 |
| HILIC_POS_231_16992a2_696 | | 231.1699 | 2.70 | | HILIC | POS | | | 312146 | 1029279 | 202121 | 721550 | 3.59E-01 | -0.13 | 0.62 |
| RP_NEG_498_93106a8_857 | | 498.9311 | 8.86 | | RP | NEG | | | 496027 | 360417 | 452129 | 340974 | 3.59E-01 | -0.13 | 0.65 |
| RP_POS_769_54626a10_613 | | 769.5463 | 10.61 | | RP | POS | | | 482797 | 324367 | 445022 | 279875 | 3.59E-01 | -0.13 | 0.74 |
| RP_NEG_221_15514a9_372 | | 221.1551 | 9.37 | | RP | NEG | | | 1935842 | 86936.6 | 1948311 | 109127 | 3.60E-01 | 0.13 | 0.62 |
| RP_NEG_221_94678a15_364 | | 221.9468 | 15.36 | | RP | NEG | | | 1734135 | 806621 | 1850060 | 1015447 | 3.60E-01 | 0.13 | 0.58 |
| RP_POS_242_175a7_683 | | 242.1750 | 7.68 | | RP | POS | | | 907325 | 111250 | 922360 | 126719 | 3.60E-01 | 0.13 | 1.05 |
| RP_POS_456_66293a10_091 | | 456.6629 | 10.09 | | RP | POS | | | 229873 | 152841 | 249681 | 162409 | 3.60E-01 | 0.13 | 0.72 |
| RP_POS_466_00717a10_401 | | 466.0072 | 10.40 | | RP | POS | | | 199619 | 120945 | 216655 | 147593 | 3.60E-01 | 0.13 | 0.96 |
| RP_POS_641_11426a11_416 | | 641.1143 | 11.42 | | RP | POS | | | 231466 | 120947 | 249863 | 165758 | 3.60E-01 | 0.13 | 0.81 |
| RP_POS_777_53906a11_485 | | 777.5391 | 11.49 | | RP | POS | | | 205310 | 91135.1 | 218100 | 110473 | 3.60E-01 | 0.13 | 0.64 |
| HILIC_POS_562_32666a1_164 | | 562.3267 | 1.16 | | HILIC | POS | | | 435654 | 153728 | 454035 | 140789 | 3.60E-01 | 0.12 | 0.66 |
| RP_POS_850_60876a9_628 | | 850.6088 | 9.63 | | RP | POS | | | 1599697 | 866645 | 1499013 | 747994 | 3.60E-01 | -0.12 | 0.86 |
| RP_NEG_591_39075a11_649 | | 591.3908 | 11.65 | | RP | NEG | | | 781200 | 425010 | 734501 | 321462 | 3.60E-01 | -0.13 | 0.56 |
| RP_NEG_433_23727a11_108 | | 433.2373 | 11.11 | | RP | NEG | | | 269733 | 273971 | 331009 | 614802 | 3.61E-01 | 0.14 | 0.77 |
| RP_POS_279_15924a9_523 | | 279.1592 | 9.52 | | RP | POS | 149.0231(100), 57.0707(5) | | 2.6E+07 | 6838314 | 2.7E+07 | 1E+07 | 3.61E-01 | 0.13 | 0.79 |
| RP_NEG_311_16925a10_513 | | 311.1693 | 10.51 | | RP | NEG | 311.1698(100) | | 1E+07 | 5338553 | 1.1E+07 | 9334792 | 3.61E-01 | 0.13 | 0.65 |
| RP_POS_321_90741a15_384 | | 321.9074 | 15.38 | | RP | POS | | | 114894 | 92045.9 | 126685 | 96172.1 | 3.61E-01 | 0.13 | 0.83 |
| RP_POS_684_08002a11_253 | | 684.0800 | 11.25 | | RP | POS | | | 165256 | 96829.6 | 178041 | 106729 | 3.61E-01 | 0.13 | 0.74 |
| RP_POS_203_08127a5_367 | Vasicinone | 203.0813 | 5.37 | | RP | POS | 203.0812(100) | 2 | 516142 | 182165 | 539974 | 197294 | 3.61E-01 | 0.13 | 0.74 |
| RP_POS_194_11774a0_68 | | 194.1177 | 0.68 | | RP | POS | 194.1188(100) | | 2930696 | 195939 | 2907523 | 175647 | 3.61E-01 | -0.12 | 0.86 |
| HILIC_NEG_617_737a6_134 | | 617.7370 | 6.13 | | HILIC | NEG | | | 268195 | 99246.8 | 255770 | 99374.4 | 3.61E-01 | -0.13 | 0.58 |
| RP_POS_299_20044a10_602 | | 299.2004 | 10.60 | | RP | POS | | | 985234 | 167644 | 1018391 | 326100 | 3.62E-01 | 0.13 | 0.74 |
| RP_POS_991_67212a10_357 | | 991.6721 | 10.36 | | RP | POS | | | 9586080 | 4995011 | 1E+07 | 5944764 | 3.62E-01 | 0.13 | 1.02 |
| HILIC_NEG_201_80206a3_091 | | 201.8021 | 3.09 | | HILIC | NEG | | | 1919061 | 227443 | 1890284 | 233228 | 3.62E-01 | -0.12 | 0.74 |
| HILIC_POS_175_14404a6_692 | | 175.1440 | 6.69 | | HILIC | POS | | | 2210749 | 920759 | 2098459 | 881958 | 3.62E-01 | -0.12 | 0.6 |
| RP_POS_130_06537a4_824 | | 130.0654 | 4.82 | | RP | POS | | | 483826 | 130552 | 468523 | 115789 | 3.62E-01 | -0.12 | 0.74 |
| RP_POS_253_10675a7_721 | | 253.1068 | 7.72 | | RP | POS | | | 2704087 | 212201 | 2679028 | 190935 | 3.62E-01 | -0.12 | 0.81 |
| RP_POS_347_13712a8_843 | | 347.1371 | 8.84 | | RP | POS | | | 858457 | 396970 | 810850 | 368744 | 3.62E-01 | -0.12 | 0.89 |
| RP_POS_304_26141a11_143 | | 304.2614 | 11.14 | | RP | POS | | | 6623516 | 1195227 | 6455123 | 1465870 | 3.62E-01 | -0.13 | 1.14 |
| HILIC_NEG_297_73315a3_324 | | 297.7332 | 3.32 | | HILIC | NEG | | | 2099183 | 136083 | 2117537 | 156509 | 3.63E-01 | 0.13 | 0.71 |
| RP_NEG_738_50873a13_042 | | 738.5087 | 13.04 | | RP | NEG | | | 2825527 | 1017872 | 2958804 | 1111619 | 3.63E-01 | 0.13 | 0.78 |
| HILIC_POS_375_32172a10_78 | | 375.3217 | 10.78 | | HILIC | POS | | | 180114 | 140327 | 195768 | 111956 | 3.63E-01 | 0.12 | 0.8 |
| RP_POS_603_46564a11_237 | | 603.4656 | 11.24 | | RP | POS | | | 532791 | 529963 | 592001 | 424902 | 3.63E-01 | 0.12 | 0.86 |
| HILIC_NEG_100_93343a6_145 | | 100.9334 | 6.15 | | HILIC | NEG | | | 2.8E+07 | 7993114 | 2.7E+07 | 8031143 | 3.63E-01 | -0.12 | 0.6 |
| HILIC_POS_418_20633a0_917 | | 418.2063 | 0.92 | | HILIC | POS | | | 420676 | 114359 | 406935 | 106950 | 3.63E-01 | -0.12 | 0.88 |
| RP_POS_804_55096a12_872 | | 804.5510 | 12.87 | | RP | POS | | | 6.5E+07 | 2.6E+07 | 6.2E+07 | 2.4E+07 | 3.63E-01 | -0.12 | 0.82 |
| RP_POS_387_20792a11_153 | | 387.2079 | 11.15 | | RP | POS | | | 235316 | 185055 | 217948 | 82883.3 | 3.63E-01 | -0.13 | 0.65 |
| HILIC_POS_138_05504a2_044 | | 138.0550 | 2.04 | | HILIC | POS | 138.0547(100), 94.0656(2) | | 370245 | 2171914 | 1387982 | 1.1E+07 | 3.64E-01 | 0.16 | 0.79 |
| RP_NEG_245_12953a6_225 | | 245.1295 | 6.23 | | RP | NEG | | | 90022.9 | 72122.3 | 99680.4 | 82101.1 | 3.64E-01 | 0.13 | 0.73 |
| RP_POS_512_41656a10_453 | | 512.4166 | 10.45 | | RP | POS | | | 7842714 | 706790 | 7934915 | 771963 | 3.64E-01 | 0.12 | 1.04 |
| RP_NEG_829_18036a9_347 | | 829.1804 | 9.35 | | RP | NEG | | | 6253742 | 644809 | 6175159 | 621391 | 3.64E-01 | -0.12 | 0.85 |
| RP_POS_245_1315a12_968 | | 245.1315 | 12.97 | | RP | POS | | | 920868 | 758480 | 839267 | 555553 | 3.64E-01 | -0.12 | 0.62 |
| HILIC_POS_371_23947a5_34 | | 371.2395 | 5.34 | | HILIC | POS | | | 4591952 | 1102443 | 4439721 | 1318354 | 3.64E-01 | -0.13 | 0.86 |
| HILIC_NEG_315_0419a7_661 | | 315.0419 | 7.66 | | HILIC | NEG | | | 895739 | 221140 | 922997 | 218716 | 3.65E-01 | 0.12 | 0.65 |
| HILIC_POS_228_23213a0_548 | | 228.2321 | 0.55 | | HILIC | POS | | | 901209 | 526448 | 966760 | 531114 | 3.65E-01 | 0.12 | 0.8 |
| RP_NEG_565_09833a8_98 | | 565.0983 | 8.98 | | RP | NEG | | | 846347 | 50110.1 | 852676 | 51891.8 | 3.65E-01 | 0.12 | 0.89 |
| RP_POS_172_16965a8_493 | | 172.1697 | 8.49 | | RP | POS | | | 952540 | 200740 | 978717 | 219618 | 3.65E-01 | 0.12 | 1 |
| RP_POS_636_8512a11_531 | | 636.8512 | 11.53 | | RP | POS | | | 95401.8 | 59889.2 | 102879 | 60625.2 | 3.65E-01 | 0.12 | 0.63 |
| RP_POS_431_38931a13_146 | Alpha-tocopherol | 431.3893 | 13.15 | | RP | POS | 165.0907(100), 55.054(10) | 1 | 5345312 | 1063140 | 5202408 | 1222251 | 3.65E-01 | -0.13 | 0.9 |
| HILIC_NEG_173_92603a4_408 | | 173.9260 | 4.41 | | HILIC | NEG | | | 824341 | 182121 | 800725 | 197371 | 3.65E-01 | -0.12 | 0.71 |
| RP_NEG_465_28641a10_407 | | 465.2864 | 10.41 | | RP | NEG | | | 188478 | 182172 | 166476 | 173131 | 3.65E-01 | -0.12 | 0.64 |
| RP_POS_246_02821a15_351 | | 246.0282 | 15.35 | | RP | POS | | | 884040 | 416761 | 833503 | 399585 | 3.65E-01 | -0.12 | 0.68 |
| RP_POS_283_26306a10_719 | | 283.2631 | 10.72 | | RP | POS | | | 4334900 | 1361165 | 4181638 | 1121450 | 3.65E-01 | -0.12 | 0.56 |
| RP_POS_476_34317a10_233 | | 476.3432 | 10.23 | | RP | POS | | | 182793 | 158202 | 166639 | 100727 | 3.65E-01 | -0.12 | 0.72 |
| RP_POS_229_11981a8_938 | Bisphenol A | 229.1198 | 8.94 | | RP | POS | 229.1206(100) | 2 | 717873 | 6982053 | 130167 | 300395 | 3.65E-01 | -0.16 | 0.53 |
| RP_POS_733_51355a11_596 | | 733.5136 | 11.60 | | RP | POS | | | 245396 | 239325 | 278475 | 288954 | 3.66E-01 | 0.13 | 0.79 |
| RP_POS_243_12256a10_253 | | 243.1226 | 10.25 | | RP | POS | | | 1984112 | 983474 | 2102358 | 931668 | 3.66E-01 | 0.12 | 1.11 |
| RP_POS_527_29663a10_485 | | 527.2966 | 10.49 | | RP | POS | | | 905154 | 523458 | 966601 | 472624 | 3.66E-01 | 0.12 | 0.94 |
| RP_POS_172_0717a2_457 | Metronidazole | 172.0717 | 2.46 | | RP | POS | 128.0453(100), 172.0712(45) | 2 | 1395682 | 2945606 | 1817017 | 3747946 | 3.66E-01 | 0.13 | 0.81 |
| HILIC_NEG_98_93897a5_647 | | 98.9390 | 5.65 | | HILIC | NEG | | | 1617727 | 212251 | 1593236 | 184888 | 3.66E-01 | -0.12 | 0.65 |
| HILIC_POS_158_96428a4_336 | | 158.9643 | 4.34 | | HILIC | POS | | | 1.5E+08 | 7859346 | 1.5E+08 | 8137051 | 3.66E-01 | -0.12 | 0.89 |
| HILIC_POS_200_04074a6_794 | | 200.0407 | 6.79 | | HILIC | POS | | | 642317 | 138628 | 626439 | 119030 | 3.66E-01 | -0.12 | 0.66 |
| RP_POS_252_04552a8_759 | | 252.0455 | 8.76 | | RP | POS | | | 1647525 | 737220 | 1562223 | 646781 | 3.66E-01 | -0.12 | 0.56 |
| RP_POS_523_37903a13_815 | | 523.3790 | 13.82 | | RP | POS | | | 3037875 | 1224742 | 2872191 | 1427076 | 3.66E-01 | -0.12 | 0.72 |
| RP_POS_624_44299a9_94 | | 624.4430 | 9.94 | | RP | POS | | | 5935949 | 2479054 | 5630933 | 2454072 | 3.66E-01 | -0.12 | 0.93 |
| RP_POS_286_20139a5_289 | | 286.2014 | 5.29 | | RP | POS | 85.0293(100), 286.2003(40) | | 8572572 | 4162715 | 9143137 | 4975218 | 3.67E-01 | 0.12 | 0.54 |
| RP_NEG_172_99138a2_4 | | 172.9914 | 2.40 | | RP | NEG | | | 7445001 | 6656403 | 6618864 | 6712671 | 3.67E-01 | -0.12 | 0.6 |
| RP_POS_261_91895a15_384 | | 261.9190 | 15.38 | | RP | POS | | | 759902 | 304158 | 723284 | 290446 | 3.67E-01 | -0.12 | 0.59 |
| RP_POS_271_65842a10_31 | | 271.6584 | 10.31 | | RP | POS | | | 2877106 | 989065 | 2764361 | 844362 | 3.67E-01 | -0.12 | 1.06 |
| RP_POS_323_22559a11_594 | | 323.2256 | 11.59 | | RP | POS | | | 409441 | 163284 | 389314 | 162537 | 3.67E-01 | -0.12 | 1.08 |
| RP_POS_345_18439a0_683 | | 345.1844 | 0.68 | | RP | POS | | | 2854317 | 526142 | 2794470 | 447467 | 3.67E-01 | -0.12 | 0.64 |
| RP_POS_417_74048a12_742 | | 417.7405 | 12.74 | | RP | POS | | | 1389227 | 971190 | 1274170 | 897712 | 3.67E-01 | -0.12 | 0.82 |
| RP_POS_467_44678a11_6 | | 467.4468 | 11.60 | | RP | POS | | | 243330 | 80276.7 | 232677 | 91003.3 | 3.67E-01 | -0.12 | 0.64 |
| RP_POS_495_69174a10_368 | | 495.6917 | 10.37 | | RP | POS | | | 187067 | 114415 | 205445 | 173221 | 3.68E-01 | 0.13 | 0.94 |
| RP_POS_423_25076a9_952 | | 423.2508 | 9.95 | | RP | POS | | | 843718 | 75648.5 | 852731 | 71109.6 | 3.68E-01 | 0.12 | 0.63 |
| RP_POS_243_08751a6_001 | 7,8-dimethylalloxazine (lumichrome) | 243.0875 | 6.00 | | RP | POS | 243.0879(100),172.0875(10) | 2 | 379872 | 742733 | 303556 | 492874 | 3.68E-01 | -0.12 | 0.8 |
| HILIC_NEG_194_92767a7_735 | | 194.9277 | 7.74 | | HILIC | NEG | | | 6320877 | 715698 | 6235376 | 675850 | 3.68E-01 | -0.12 | 0.75 |
| RP_POS_398_76114a13_201 | | 398.7611 | 13.20 | | RP | POS | | | 6929817 | 6886887 | 6166467 | 5554102 | 3.68E-01 | -0.12 | 0.99 |
| HILIC_POS_250_1801a0_89 | | 250.1801 | 0.89 | | HILIC | POS | | | 378215 | 3554954 | 79778.2 | 334515 | 3.68E-01 | -0.15 | 0.53 |
| RP_POS_610_54028a14_703 | | 610.5403 | 14.70 | | RP | POS | | | 1754777 | 1789001 | 1970036 | 1718836 | 3.69E-01 | 0.12 | 0.81 |
| RP_NEG_187_13412a8_431 | 10-Hydroxydecanoic acid | 187.1341 | 8.43 | | RP | NEG | 141.1283(100), 187.1336(85) | 2 | 1938105 | 1186118 | 1756153 | 1685633 | 3.69E-01 | -0.13 | 0.84 |
| HILIC_POS_296_0087a4_847 | | 296.0087 | 4.85 | | HILIC | POS | | | 580902 | 290252 | 547695 | 251337 | 3.69E-01 | -0.12 | 0.93 |
| RP_NEG_271_22842a10_253 | | 271.2284 | 10.25 | | RP | NEG | | | 1333116 | 158074 | 1313664 | 158354 | 3.69E-01 | -0.12 | 1.03 |
| RP_POS_193_15851a7_352 | | 193.1585 | 7.35 | | RP | POS | | | 3025972 | 199177 | 3002008 | 191600 | 3.69E-01 | -0.12 | 0.92 |
| RP_POS_572_41724a10_349 | | 572.4172 | 10.35 | | RP | POS | | | 5170210 | 2059125 | 4904224 | 2250869 | 3.69E-01 | -0.12 | 0.71 |
| RP_POS_313_32056a10_835 | | 313.3206 | 10.84 | | RP | POS | | | 1307482 | 392943 | 1397121 | 924186 | 3.70E-01 | 0.14 | 0.88 |
| HILIC_POS_146_09238a3_74 | | 146.0924 | 3.74 | | HILIC | POS | | | 286185 | 780564 | 438796 | 1529550 | 3.70E-01 | 0.13 | 0.74 |
| RP_POS_603_53448a11_957 | | 603.5345 | 11.96 | | RP | POS | | | 735235 | 1005136 | 889016 | 1428271 | 3.70E-01 | 0.13 | 0.8 |
| RP_POS_615_42853a15_162 | | 615.4285 | 15.16 | | RP | POS | | | 741354 | 433919 | 816911 | 733939 | 3.70E-01 | 0.13 | 0.67 |
| RP_NEG_530_30286a10_359 | | 530.3029 | 10.36 | | RP | NEG | | | 3171765 | 435985 | 3227733 | 473152 | 3.70E-01 | 0.12 | 0.97 |
| RP_POS_244_19057a7_673 | | 244.1906 | 7.67 | | RP | POS | | | 761169 | 488665 | 817881 | 438863 | 3.70E-01 | 0.12 | 0.74 |
| RP_POS_753_58801a13_544 | | 753.5880 | 13.54 | | RP | POS | | | 2192845 | 1709296 | 2392927 | 1561467 | 3.70E-01 | 0.12 | 0.74 |
| HILIC_NEG_132_90028a3_144 | | 132.9003 | 3.14 | | HILIC | NEG | | | 1.7E+07 | 1282770 | 1.7E+07 | 1410358 | 3.70E-01 | -0.12 | 0.73 |
| HILIC_NEG_276_99243a0_589 | | 276.9924 | 0.59 | | HILIC | NEG | | | 1467024 | 417923 | 1420044 | 350664 | 3.70E-01 | -0.12 | 0.55 |
| HILIC_POS_216_92213a5_432 | | 216.9221 | 5.43 | | HILIC | POS | | | 306912 | 107991 | 293027 | 117562 | 3.70E-01 | -0.12 | 0.73 |
| HILIC_POS_246_98834a0_927 | | 246.9883 | 0.93 | | HILIC | POS | | | 213688 | 40078.3 | 208717 | 40861.8 | 3.70E-01 | -0.12 | 0.93 |
| RP_POS_481_23245a10_898 | | 481.2325 | 10.90 | | RP | POS | | | 922167 | 922021 | 822863 | 701938 | 3.70E-01 | -0.12 | 0.52 |
| RP_POS_489_31125a11_498 | | 489.3113 | 11.50 | | RP | POS | | | 554814 | 87252 | 543789 | 92099.4 | 3.70E-01 | -0.12 | 0.7 |
| RP_POS_615_42804a15_343 | | 615.4280 | 15.34 | | RP | POS | | | 523747 | 353748 | 480878 | 345983 | 3.70E-01 | -0.12 | 0.71 |
| HILIC_NEG_219_84576a3_199 | | 219.8458 | 3.20 | | HILIC | NEG | | | 311861 | 90657.8 | 322883 | 89487.4 | 3.71E-01 | 0.12 | 0.64 |
| RP_NEG_297_27991a11_755 | | 297.2799 | 11.76 | | RP | NEG | | | 143971 | 86208.3 | 132796 | 95460 | 3.71E-01 | -0.12 | 1.24 |
| RP_NEG_851_11481a9_482 | | 851.1148 | 9.48 | | RP | NEG | | | 5594556 | 349945 | 5549813 | 377857 | 3.71E-01 | -0.12 | 0.82 |
| RP_NEG_214_78349a15_326 | | 214.7835 | 15.33 | | RP | NEG | | | 1334018 | 126437 | 1349993 | 134371 | 3.72E-01 | 0.12 | 0.73 |
| RP_NEG_737_5379a12_802 | | 737.5379 | 12.80 | | RP | NEG | | | 1088853 | 200465 | 1115014 | 225372 | 3.72E-01 | 0.12 | 1.05 |
| RP_NEG_962_53589a13_134 | | 962.5359 | 13.13 | | RP | NEG | | | 2331708 | 767478 | 2423918 | 745304 | 3.72E-01 | 0.12 | 0.83 |
| RP_POS_213_10966a13_31 | | 213.1097 | 13.31 | | RP | POS | | | 2122554 | 1083969 | 2256077 | 1100184 | 3.72E-01 | 0.12 | 0.96 |
| RP_POS_267_98926a0_675 | | 267.9893 | 0.68 | | RP | POS | | | 280231 | 56778.9 | 286943 | 53406.4 | 3.72E-01 | 0.12 | 0.63 |
| RP_POS_798_53857a13_801 | | 798.5386 | 13.80 | | RP | POS | | | 1.2E+07 | 8253418 | 1.3E+07 | 9548490 | 3.72E-01 | 0.12 | 0.86 |
| RP_POS_786_60046a14_353 | PC 36:2 (18:1/18:1) | 786.6005 | 14.35 | | RP | POS | 184.0743(100), 86.095(10) | 1 | 5.3E+07 | 2.5E+07 | 5E+07 | 2.2E+07 | 3.72E-01 | -0.12 | 0.67 |
| HILIC_NEG_162_98273a3_05 | | 162.9827 | 3.05 | | HILIC | NEG | | | 498744 | 67451.3 | 491239 | 55942.2 | 3.72E-01 | -0.12 | 0.7 |
| HILIC_NEG_216_88025a0_904 | | 216.8803 | 0.90 | | HILIC | NEG | | | 260525 | 37647.7 | 255872 | 38471.3 | 3.72E-01 | -0.12 | 0.61 |
| HILIC_POS_338_23221a0_73 | | 338.2322 | 0.73 | | HILIC | POS | | | 348492 | 150638 | 329672 | 156742 | 3.72E-01 | -0.12 | 0.62 |
| RP_NEG_386_83932a15_362 | | 386.8393 | 15.36 | | RP | NEG | | | 547709 | 121554 | 531686 | 138709 | 3.72E-01 | -0.12 | 0.76 |
| RP_NEG_526_82642a15_363 | | 526.8264 | 15.36 | | RP | NEG | | | 1095968 | 48073.8 | 1090107 | 47828.4 | 3.72E-01 | -0.12 | 0.65 |
| RP_POS_150_12759a4_058 | | 150.1276 | 4.06 | | RP | POS | | | 1225142 | 604228 | 1150555 | 615067 | 3.72E-01 | -0.12 | 0.73 |
| RP_POS_225_22089a9_56 | | 225.2209 | 9.56 | | RP | POS | | | 1123228 | 140845 | 1106688 | 130829 | 3.72E-01 | -0.12 | 0.78 |
| RP_POS_353_13052a10_151 | | 353.1305 | 10.15 | | RP | POS | | | 469265 | 205961 | 445763 | 180095 | 3.72E-01 | -0.12 | 0.7 |
| RP_POS_423_74249a13_115 | | 423.7425 | 13.12 | | RP | POS | | | 946660 | 592150 | 876482 | 557927 | 3.72E-01 | -0.12 | 0.64 |
| RP_POS_507_36871a9_81 | | 507.3687 | 9.81 | | RP | POS | | | 8102811 | 3514577 | 7671415 | 3549969 | 3.72E-01 | -0.12 | 0.71 |
| RP_POS_566_42682a5_753 | | 566.4268 | 5.75 | | RP | POS | | | 2031744 | 1233954 | 1893119 | 1042951 | 3.72E-01 | -0.12 | 0.81 |
| RP_POS_549_31415a10_56 | | 549.3142 | 10.56 | | RP | POS | | | 2802714 | 2463800 | 3177195 | 3506386 | 3.73E-01 | 0.13 | 0.66 |
| HILIC_POS_352_24625a5_659 | | 352.2463 | 5.66 | | HILIC | POS | | | 201742 | 206734 | 224953 | 175420 | 3.73E-01 | 0.12 | 1.12 |
| RP_POS_203_10667a7_207 | | 203.1067 | 7.21 | | RP | POS | | | 1829731 | 125001 | 1843600 | 103275 | 3.73E-01 | 0.12 | 0.78 |
| RP_POS_278_2478a9_27 | | 278.2478 | 9.27 | | RP | POS | | | 532055 | 483618 | 601711 | 634925 | 3.73E-01 | 0.12 | 1.2 |
| RP_POS_269_13623a4_354 | | 269.1362 | 4.35 | | RP | POS | 269.1372(100) | | 210682 | 145037 | 194696 | 117966 | 3.73E-01 | -0.12 | 1.16 |
| RP_POS_317_24738a10_843 | | 317.2474 | 10.84 | | RP | POS | | | 1728225 | 301017 | 1691678 | 298740 | 3.73E-01 | -0.12 | 0.55 |
| RP_POS_396_78732a9_396 | | 396.7873 | 9.40 | | RP | POS | | | 514338 | 314050 | 477479 | 291839 | 3.73E-01 | -0.12 | 0.76 |
| RP_POS_639_39691a15_162 | | 639.3969 | 15.16 | | RP | POS | | | 521455 | 259253 | 491385 | 235327 | 3.73E-01 | -0.12 | 1.2 |
| RP_NEG_1171_17737a9_917 | | 1171.1774 | 9.92 | | RP | NEG | | | 2534362 | 220491 | 2560385 | 208052 | 3.74E-01 | 0.12 | 1.06 |
| RP_NEG_135_03125a0_68 | | 135.0313 | 0.68 | | RP | NEG | | | 2874514 | 1442082 | 3047450 | 1403105 | 3.74E-01 | 0.12 | 0.69 |
| RP_NEG_346_17712a11_64 | | 346.1771 | 11.64 | | RP | NEG | | | 251029 | 69472.8 | 259832 | 74630.1 | 3.74E-01 | 0.12 | 0.52 |
| RP_NEG_479_78198a15_385 | | 479.7820 | 15.39 | | RP | NEG | | | 421414 | 232747 | 450655 | 246694 | 3.74E-01 | 0.12 | 1.06 |
| RP_POS_436_98096a9_933 | | 436.9810 | 9.93 | | RP | POS | | | 125038 | 62071.6 | 134023 | 82404.5 | 3.74E-01 | 0.12 | 0.59 |
| RP_NEG_437_19943a11_142 | | 437.1994 | 11.14 | | RP | NEG | | | 325047 | 62697.5 | 317172 | 66311.8 | 3.74E-01 | -0.12 | 0.86 |
| RP_POS_345_09882a9_209 | | 345.0988 | 9.21 | | RP | POS | | | 4770731 | 2549949 | 4451167 | 2688903 | 3.74E-01 | -0.12 | 0.97 |
| RP_POS_271_16626a9_195 | | 271.1663 | 9.20 | | RP | POS | 271.1718(100) | | 668462 | 138239 | 685143 | 136830 | 3.75E-01 | 0.12 | 0.68 |
| HILIC_POS_169_97734a1_347 | | 169.9773 | 1.35 | | HILIC | POS | | | 520649 | 195128 | 544722 | 200933 | 3.75E-01 | 0.12 | 0.69 |
| HILIC_POS_300_21704a0_809 | | 300.2170 | 0.81 | | HILIC | POS | | | 742795 | 378724 | 697271 | 371166 | 3.75E-01 | -0.12 | 0.58 |
| RP_NEG_358_97479a9_012 | | 358.9748 | 9.01 | | RP | NEG | | | 982941 | 83524.6 | 972229 | 92177.8 | 3.75E-01 | -0.12 | 0.6 |
| RP_POS_663_46759a10_683 | | 663.4676 | 10.68 | | RP | POS | | | 613445 | 231023 | 584813 | 239570 | 3.75E-01 | -0.12 | 0.84 |
| RP_POS_887_15576a9_586 | | 887.1558 | 9.59 | | RP | POS | | | 979773 | 91508.4 | 969217 | 82920.6 | 3.75E-01 | -0.12 | 0.7 |

**Supplemantary table 1**: The non-targeted metabolomics results ordered by the VIP value (p value < 0.0003 are shaded dark green, p value < 0.05 are shaded light green, d values below 0 are blue and above 0 are red)

| Feature_ID | Metabolite_name | Average_Mz | Average_RT_min_ | HMDB ID | RP_HILIC | POS_NEG | Cureted_MSMS | ID_level | Cases Mean | SD | Controls Mean2 | SD3 | Welch test p | Cohen's d | VIP |
|---|---|---|---|---|---|---|---|---|---|---|---|---|---|---|---|
| HILIC_POS_158_1541a0_748 | | 158.1541 | 0.75 | | HILIC | POS | | | 832682 | 135383 | 848607 | 127878 | 3.76E-01 | 0.12 | 0.68 |
| RP_POS_265_9393a15_537 | | 265.9393 | 15.54 | | RP | POS | | | 966936 | 295752 | 1009982 | 398406 | 3.76E-01 | 0.12 | 0.73 |
| RP_POS_390_29477a9_66 | | 390.2948 | 9.66 | | RP | POS | | | 747563 | 379234 | 797231 | 434729 | 3.76E-01 | 0.12 | 0.78 |
| RP_POS_480_6825a10_384 | | 480.6825 | 10.38 | | RP | POS | | | 292909 | 190734 | 316429 | 196928 | 3.76E-01 | 0.12 | 0.82 |
| RP_POS_580_41492a9_967 | | 580.4149 | 9.97 | | RP | POS | | | 8354336 | 3373378 | 8781945 | 3653115 | 3.76E-01 | 0.12 | 0.9 |
| HILIC_NEG_165_01927a0_935 | C8H8O5 | 165.0193 | 0.94 | | HILIC | NEG | 121.0297(100), 165.0181(5) | 3 | 962840 | 264451 | 930189 | 273225 | 3.76E-01 | -0.12 | 0.73 |
| RP_POS_257_15109a9_092 | | 257.1511 | 9.09 | | RP | POS | 257.1532(100) | | 1821250 | 195300 | 1797691 | 193681 | 3.76E-01 | -0.12 | 1.18 |
| HILIC_NEG_373_01004a1_256 | | 373.0100 | 1.26 | | HILIC | NEG | | | 624705 | 248441 | 592713 | 276589 | 3.76E-01 | -0.12 | 0.66 |
| HILIC_POS_133_09737a1_342 | | 133.0974 | 1.34 | | HILIC | POS | | | 74070.8 | 67895.5 | 66205.4 | 62373.3 | 3.76E-01 | -0.12 | 0.59 |
| RP_NEG_183_06612a3_46 | | 183.0661 | 3.46 | | RP | NEG | | | 931136 | 134683 | 913336 | 156475 | 3.76E-01 | -0.12 | 0.67 |
| HILIC_POS_432_28a0_61 | | 432.2800 | 0.61 | | HILIC | POS | | | 144783 | 111517 | 164247 | 193206 | 3.77E-01 | 0.13 | 0.54 |
| RP_POS_240_23235a10_834 | | 240.2324 | 10.83 | | RP | POS | | | 135095 | 74643.2 | 150570 | 159407 | 3.77E-01 | 0.13 | 0.81 |
| RP_NEG_371_16971a7_949 | | 371.1697 | 7.95 | | RP | NEG | | | 1390250 | 1087104 | 1512354 | 938279 | 3.77E-01 | 0.12 | 0.61 |
| RP_POS_397_41565a11_409 | | 397.4157 | 11.41 | | RP | POS | | | 7.4E+08 | 8.5E+08 | 8.4E+08 | 8.1E+08 | 3.77E-01 | 0.12 | 1.17 |
| RP_NEG_509_11673a8_834 | | 509.1167 | 8.83 | | RP | NEG | | | 1.3E+07 | 778320 | 1.3E+07 | 737491 | 3.77E-01 | -0.12 | 1.1 |
| RP_NEG_527_44769a11_831 | | 527.4477 | 11.83 | | RP | NEG | | | 750065 | 456452 | 693380 | 479389 | 3.77E-01 | -0.12 | 0.58 |
| RP_NEG_594_39594a11_142 | | 594.3959 | 11.14 | | RP | NEG | | | 5752219 | 660257 | 5673768 | 639800 | 3.77E-01 | -0.12 | 1.03 |
| RP_POS_146_93773a0_629 | | 146.9377 | 0.63 | | RP | POS | | | 445782 | 183472 | 423985 | 177408 | 3.77E-01 | -0.12 | 0.54 |
| RP_POS_272_13406a9_208 | | 272.1341 | 9.21 | | RP | POS | | | 553539 | 728674 | 473061 | 608711 | 3.77E-01 | -0.12 | 1.05 |
| RP_POS_353_09317a9_209 | | 353.0932 | 9.21 | | RP | POS | | | 1027002 | 656146 | 947581 | 658666 | 3.77E-01 | -0.12 | 0.64 |
| RP_POS_836_61517a14_376 | | 836.6152 | 14.38 | | RP | POS | | | 4070108 | 3070538 | 4559564 | 4727893 | 3.78E-01 | 0.13 | 1.14 |
| HILIC_NEG_165_83717a3_197 | | 165.8372 | 3.20 | | HILIC | NEG | | | 2.4E+07 | 964364 | 2.4E+07 | 856807 | 3.78E-01 | 0.12 | 0.56 |
| HILIC_NEG_236_81673a7_748 | | 236.8167 | 7.75 | | HILIC | NEG | | | 965012 | 163488 | 985618 | 176962 | 3.78E-01 | 0.12 | 0.78 |
| RP_POS_224_1279a0_821 | 4-Hydroxybenzoylcholine | 224.1279 | 0.82 | | RP | POS | 224.1277(100), 165.0542(60) | 2 | 2544169 | 784644 | 2640754 | 813008 | 3.78E-01 | 0.12 | 0.76 |
| RP_POS_276_21716a9_459 | | 276.2172 | 9.46 | | RP | POS | | | 6720069 | 5856947 | 6018988 | 5779069 | 3.78E-01 | -0.12 | 0.67 |
| RP_POS_397_22968a11_594 | | 397.2297 | 11.59 | | RP | POS | | | 1940081 | 222313 | 1912387 | 235804 | 3.78E-01 | -0.12 | 0.73 |
| RP_POS_590_08173a11_3 | | 590.0817 | 11.30 | | RP | POS | | | 118813 | 76439.3 | 110998 | 52804.8 | 3.78E-01 | -0.12 | 0.83 |
| RP_NEG_269_24911a11_292 | Heptadecanoic acid | 269.2491 | 11.29 | | RP | NEG | 269.2475(100) | 2 | 834303 | 312793 | 797238 | 303300 | 3.79E-01 | -0.12 | 1.31 |
| RP_NEG_214_78342a15_466 | | 214.7834 | 15.47 | | RP | NEG | | | 2992995 | 127944 | 3010579 | 159688 | 3.79E-01 | 0.12 | 0.92 |
| RP_NEG_592_36346a10_75 | | 592.3635 | 10.75 | | RP | NEG | | | 2785961 | 752152 | 2881108 | 822515 | 3.79E-01 | 0.12 | 0.81 |
| RP_POS_358_27713a9_332 | | 358.2771 | 9.33 | | RP | POS | | | 1042712 | 1392201 | 1242542 | 1852739 | 3.79E-01 | 0.12 | 0.76 |
| RP_POS_672_52594a10_867 | | 672.5259 | 10.87 | | RP | POS | | | 2025092 | 229526 | 2052487 | 226196 | 3.79E-01 | 0.12 | 0.7 |
| RP_POS_254_24779a10_344 | Palmitoleamide | 254.2478 | 10.34 | | RP | POS | 254.2472(100) | 2 | 457626 | 1316067 | 338651 | 578577 | 3.79E-01 | -0.13 | 0.87 |
| RP_NEG_213_91531a13_28 | | 213.9153 | 13.28 | | RP | NEG | | | 350281 | 189231 | 328615 | 171404 | 3.79E-01 | -0.12 | 0.8 |
| RP_NEG_269_21243a9_558 | | 269.2124 | 9.56 | | RP | NEG | | | 1476861 | 206598 | 1452834 | 193741 | 3.79E-01 | -0.12 | 1.09 |
| RP_POS_252_04561a12_921 | | 252.0456 | 12.92 | | RP | POS | | | 2423932 | 1272748 | 2280843 | 1111955 | 3.79E-01 | -0.12 | 0.6 |
| RP_POS_329_15677a5_445 | | 329.1568 | 5.45 | | RP | POS | | | 379240 | 149120 | 361085 | 152130 | 3.79E-01 | -0.12 | 0.98 |
| RP_POS_718_513a10_522 | | 718.5130 | 10.52 | | RP | POS | | | 1374488 | 648421 | 1295667 | 659203 | 3.79E-01 | -0.12 | 0.82 |
| RP_NEG_415_64685a15_493 | | 415.6469 | 15.49 | | RP | NEG | | | 592886 | 55099.8 | 599821 | 59899.2 | 3.80E-01 | 0.12 | 0.64 |
| RP_POS_679_64209a11_667 | | 679.6421 | 11.67 | | RP | POS | | | 109002 | 52557 | 116312 | 67138 | 3.80E-01 | 0.12 | 0.66 |
| RP_NEG_384_16043a11_142 | | 384.1604 | 11.14 | | RP | NEG | | | 1652952 | 73030.2 | 1643661 | 80798 | 3.80E-01 | -0.12 | 0.71 |
| RP_POS_173_0287a15_682 | | 173.0287 | 15.68 | | RP | POS | | | 9.2E+07 | 4.7E+07 | 8.6E+07 | 4.8E+07 | 3.80E-01 | -0.12 | 0.54 |
| RP_POS_253_21608a10_096 | | 253.2161 | 10.10 | | RP | POS | | | 1753553 | 218541 | 1727837 | 210592 | 3.80E-01 | -0.12 | 0.93 |
| RP_POS_277_6691a10_491 | | 277.6691 | 10.49 | | RP | POS | | | 395468 | 334418 | 360978 | 240218 | 3.80E-01 | -0.12 | 0.6 |
| RP_POS_298_64655a10_615 | | 298.6466 | 10.62 | | RP | POS | | | 350964 | 233747 | 326092 | 180990 | 3.80E-01 | -0.12 | 0.84 |
| RP_POS_321_13031a7_909 | | 321.1303 | 7.91 | | RP | POS | | | 612859 | 49350.7 | 606161 | 60553.7 | 3.80E-01 | -0.12 | 0.67 |
| RP_POS_451_80548a9_394 | | 451.8055 | 9.39 | | RP | POS | | | 382703 | 206418 | 358254 | 200782 | 3.80E-01 | -0.12 | 0.73 |
| RP_POS_655_28052a5_639 | | 655.2805 | 5.64 | | RP | POS | | | 607480 | 810010 | 530872 | 441682 | 3.80E-01 | -0.12 | 0.73 |
| RP_POS_356_673a8_842 | | 356.6730 | 8.84 | | RP | POS | | | 504675 | 505928 | 459371 | 213488 | 3.80E-01 | -0.13 | 0.65 |
| HILIC_POS_414_16714a2_807 | | 414.1671 | 2.81 | | HILIC | POS | | | 73740.2 | 24459.8 | 76909.5 | 27990.7 | 3.81E-01 | 0.12 | 0.76 |
| RP_POS_633_61206a11_437 | | 633.6121 | 11.44 | | RP | POS | | | 178170 | 98199.5 | 192184 | 130472 | 3.81E-01 | 0.12 | 0.75 |
| RP_POS_866_48743a13_223 | | 866.4874 | 13.22 | | RP | POS | | | 775948 | 457118 | 833743 | 502370 | 3.81E-01 | 0.12 | 0.83 |
| HILIC_NEG_315_04187a5_656 | | 315.0419 | 5.66 | | HILIC | NEG | | | 3760359 | 527797 | 3700101 | 480968 | 3.81E-01 | -0.12 | 0.64 |
| HILIC_POS_106_08676a5_643 | | 106.0868 | 5.64 | | HILIC | POS | | | 2.1E+07 | 4749496 | 2E+07 | 4470202 | 3.81E-01 | -0.12 | 1.04 |
| RP_NEG_722_49866a12_324 | | 722.4987 | 12.32 | | RP | NEG | | | 754390 | 586543 | 680434 | 641793 | 3.81E-01 | -0.12 | 0.52 |
| RP_POS_324_20175a5_729 | | 324.2018 | 5.73 | | RP | POS | | | 892087 | 74727.9 | 883910 | 62117.3 | 3.81E-01 | -0.12 | 0.85 |
| RP_POS_461_66962a10_245 | | 461.6696 | 10.25 | | RP | POS | | | 117376 | 55009.7 | 110540 | 58646.6 | 3.81E-01 | -0.12 | 0.76 |
| HILIC_POS_204_12337a3_178 | | 204.1234 | 3.18 | | HILIC | POS | 85.0291(100), 204.1231(40), 145.049(20) | | 1864307 | 1465949 | 2047195 | 1580370 | 3.82E-01 | 0.12 | 1 |
| HILIC_POS_189_07304a4_363 | | 189.0730 | 4.36 | | HILIC | POS | | | 2321805 | 1115192 | 2454367 | 1102182 | 3.82E-01 | 0.12 | 0.66 |
| RP_POS_608_09045a10_122 | | 608.0905 | 10.12 | | RP | POS | | | 392191 | 308881 | 429822 | 319853 | 3.82E-01 | 0.12 | 0.63 |
| RP_POS_695_09021a11_583 | | 695.0902 | 11.58 | | RP | POS | | | 143138 | 63208.7 | 151091 | 69226.7 | 3.82E-01 | 0.12 | 0.96 |
| HILIC_POS_146_1176a3_182 | 4-Trimethylammoniobutanoic acid (butyrobetaine, deoxycarnitine) | 146.1176 | 3.18 | HMDB0001161 | HILIC | POS | 146.1172(100), 87.0449(55) | 1 | 2.5E+07 | 1.1E+07 | 2.7E+07 | 1.4E+07 | 3.82E-01 | 0.12 | 0.74 |
| HILIC_NEG_198_86473a5_501 | | 198.8647 | 5.50 | | HILIC | NEG | | | 583732 | 225572 | 556982 | 222090 | 3.82E-01 | -0.12 | 0.67 |
| HILIC_NEG_94_99484a0_933 | | 94.9948 | 0.93 | | HILIC | NEG | | | 731932 | 128794 | 717129 | 119337 | 3.82E-01 | -0.12 | 0.83 |
| RP_NEG_168_98904a15_481 | | 168.9890 | 15.48 | | RP | NEG | | | 1120653 | 253388 | 1090975 | 243789 | 3.82E-01 | -0.12 | 0.53 |
| RP_POS_530_69781a10_452 | | 530.6978 | 10.45 | | RP | POS | | | 541095 | 201536 | 516584 | 207461 | 3.82E-01 | -0.12 | 0.86 |
| HILIC_NEG_343_99548a7_352 | | 343.9955 | 7.35 | | HILIC | NEG | | | 826218 | 73653 | 836130 | 90164.6 | 3.83E-01 | 0.12 | 0.64 |
| RP_POS_182_19022a4_685 | | 182.1902 | 4.69 | | RP | POS | | | 2883641 | 365068 | 2921841 | 276700 | 3.83E-01 | 0.12 | 0.56 |
| RP_POS_730_51239a11_5 | | 730.5124 | 11.50 | | RP | POS | | | 244658 | 154145 | 264619 | 177579 | 3.83E-01 | 0.12 | 0.72 |
| RP_NEG_311_29599a11_662 | Arachidic acid | 311.2960 | 11.66 | | RP | NEG | 311.2961(100) | 1 | 272413 | 147835 | 253825 | 162062 | 3.83E-01 | -0.12 | 0.95 |
| RP_NEG_209_83376a15_476 | | 209.8338 | 15.48 | | RP | NEG | | | 713048 | 49084.7 | 706813 | 54675.8 | 3.83E-01 | -0.12 | 0.82 |
| RP_POS_262_87854a15_438 | | 262.8785 | 15.44 | | RP | POS | | | 1032727 | 520077 | 969941 | 531035 | 3.83E-01 | -0.12 | 0.65 |
| RP_POS_276_13113a9_207 | | 276.1311 | 9.21 | | RP | POS | | | 267894 | 227861 | 241123 | 221364 | 3.83E-01 | -0.12 | 0.74 |
| RP_NEG_130_94438a10_559 | | 130.9444 | 10.56 | | RP | NEG | | | 4345988 | 788242 | 4434093 | 696038 | 3.84E-01 | 0.12 | 0.58 |
| RP_POS_353_15729a4_431 | | 353.1573 | 4.43 | | RP | POS | | | 1.14E+16 | 3.70E+16 | 1.71E+16 | 5.43E+16 | 3.84E-01 | 0.12 | 0.72 |
| RP_POS_396_80365a15_432 | | 396.8037 | 15.43 | | RP | POS | | | 454130 | 349482 | 496989 | 367731 | 3.84E-01 | 0.12 | 0.61 |
| HILIC_NEG_234_81956a5_754 | | 234.8196 | 5.75 | | HILIC | NEG | | | 910785 | 220302 | 885384 | 206654 | 3.84E-01 | -0.12 | 0.53 |
| HILIC_POS_398_21512a0_94 | | 398.2151 | 0.94 | | HILIC | POS | | | 218622 | 151660 | 201514 | 136686 | 3.84E-01 | -0.12 | 0.82 |
| RP_NEG_134_87541a14_343 | | 134.8754 | 14.34 | | RP | NEG | | | 641562 | 299577 | 608370 | 259456 | 3.84E-01 | -0.12 | 0.53 |
| RP_NEG_526_31445a10_085 | | 526.3145 | 10.09 | | RP | NEG | | | 1556725 | 425445 | 1507573 | 402032 | 3.84E-01 | -0.12 | 0.79 |
| RP_NEG_731_31854a4_891 | | 731.3185 | 4.89 | | RP | NEG | | | 253362 | 249534 | 227599 | 183686 | 3.84E-01 | -0.12 | 1.2 |
| RP_POS_160_0757a0_682 | | 160.0757 | 0.68 | | RP | POS | | | 4246754 | 1938498 | 4018162 | 1901858 | 3.84E-01 | -0.12 | 0.68 |
| HILIC_POS_553_40607a4_002 | | 553.4061 | 4.00 | | HILIC | POS | | | 434798 | 134069 | 450629 | 132362 | 3.85E-01 | 0.12 | 0.65 |
| RP_NEG_1061_68103a10_605 | | 1061.6810 | 10.61 | | RP | NEG | | | 921950 | 261615 | 954380 | 281738 | 3.85E-01 | 0.12 | 0.9 |
| RP_NEG_634_33325a10_61 | | 634.3333 | 10.61 | | RP | NEG | | | 1.3E+07 | 1860489 | 1.3E+07 | 1933639 | 3.85E-01 | 0.12 | 0.81 |
| RP_POS_200_97244a0_946 | | 200.9724 | 0.95 | | RP | POS | | | 1014890 | 516807 | 1082670 | 611380 | 3.85E-01 | 0.12 | 0.66 |
| RP_POS_264_95441a15_369 | | 264.9544 | 15.37 | | RP | POS | | | 4104113 | 1096747 | 4236030 | 1119819 | 3.85E-01 | 0.12 | 1.03 |
| RP_POS_279_23187a10_715 | | 279.2319 | 10.72 | | RP | POS | | | 5198749 | 3318412 | 5594588 | 3339613 | 3.85E-01 | 0.12 | 0.93 |
| RP_POS_379_16885a2_162 | | 379.1689 | 2.16 | | RP | POS | | | 1004524 | 215896 | 1030158 | 215144 | 3.85E-01 | 0.12 | 0.66 |
| HILIC_NEG_315_85449a3_246 | | 315.8545 | 3.25 | | HILIC | NEG | | | 720587 | 90335.1 | 709825 | 90793.1 | 3.85E-01 | -0.12 | 0.6 |
| RP_POS_137_06604a15_333 | | 137.0660 | 15.33 | | RP | POS | | | 616919 | 185679 | 594903 | 184625 | 3.85E-01 | -0.12 | 0.56 |
| RP_POS_211_0941a2_409 | | 211.0941 | 2.41 | | RP | POS | | | 190097 | 80776.7 | 180207 | 85108.7 | 3.85E-01 | -0.12 | 0.89 |
| RP_POS_359_20822a11_598 | | 359.2082 | 11.60 | | RP | POS | | | 409089 | 138061 | 394310 | 111244 | 3.85E-01 | -0.12 | 0.72 |
| HILIC_NEG_278_98166a5_227 | | 278.9817 | 5.23 | | HILIC | NEG | | | 8859383 | 3182296 | 9302022 | 4142849 | 3.86E-01 | 0.12 | 0.51 |
| HILIC_NEG_297_87823a7_417 | | 297.8782 | 7.42 | | HILIC | NEG | | | 101879 | 38834.4 | 106439 | 38161.9 | 3.86E-01 | 0.12 | 0.51 |
| HILIC_POS_148_04257a5_172 | | 148.0426 | 5.17 | | HILIC | POS | | | 398834 | 529677 | 457146 | 456618 | 3.86E-01 | 0.12 | 0.88 |
| RP_NEG_452_92389a12_94 | | 452.9239 | 12.94 | | RP | NEG | | | 846792 | 459007 | 904452 | 507938 | 3.86E-01 | 0.12 | 0.89 |
| RP_POS_739_11658a11_482 | | 739.1166 | 11.48 | | RP | POS | | | 212716 | 93198.6 | 227788 | 149586 | 3.86E-01 | 0.12 | 0.7 |
| RP_POS_243_09895a8_486 | | 243.0990 | 8.49 | | RP | POS | 243.0987(100) | | 928092 | 882717 | 829690 | 782966 | 3.86E-01 | -0.12 | 1.2 |
| RP_NEG_257_21274a10_391 | | 257.2127 | 10.39 | | RP | NEG | | | 285185 | 33261.2 | 281099 | 35402.4 | 3.86E-01 | -0.12 | 0.64 |
| RP_NEG_594_30322a10_231 | | 594.3032 | 10.23 | | RP | NEG | | | 964790 | 257859 | 934764 | 249064 | 3.86E-01 | -0.12 | 0.79 |
| RP_NEG_213_11336a6_827 | | 213.1134 | 6.83 | | RP | NEG | | | 277931 | 140241 | 302163 | 246349 | 3.87E-01 | 0.13 | 0.67 |
| HILIC_POS_391_31647a3_794 | | 391.3165 | 3.79 | | HILIC | POS | | | 49465.3 | 17034.8 | 51912.7 | 23269.3 | 3.87E-01 | 0.12 | 0.65 |
| RP_POS_592_75458a10_528 | | 592.7546 | 10.53 | | RP | POS | | | 376022 | 239416 | 404751 | 245003 | 3.87E-01 | 0.12 | 0.59 |
| RP_POS_692_48212a13_711 | | 692.4821 | 13.71 | | RP | POS | | | 721292 | 293952 | 757820 | 320427 | 3.87E-01 | 0.12 | 1 |
| RP_POS_797_5882a13_506 | | 797.5882 | 13.51 | | RP | POS | | | 6601486 | 3281651 | 6987823 | 3247983 | 3.87E-01 | 0.12 | 1.06 |
| RP_POS_448_33835a10_659 | | 448.3384 | 10.66 | | RP | POS | | | 127408 | 66540.7 | 119986 | 59299.6 | 3.87E-01 | -0.12 | 0.65 |
| RP_POS_697_65765a11_656 | | 697.6577 | 11.66 | | RP | POS | | | 2470967 | 4892169 | 1959197 | 3783881 | 3.87E-01 | -0.12 | 0.64 |
| RP_NEG_452_92377a13_713 | | 452.9238 | 13.71 | | RP | NEG | | | 1012932 | 499180 | 1069776 | 466419 | 3.88E-01 | 0.12 | 0.58 |
| RP_POS_288_29004a8_289 | | 288.2900 | 8.29 | | RP | POS | | | 1316701 | 2006622 | 1635564 | 3164707 | 3.88E-01 | 0.12 | 0.79 |
| RP_POS_541_03552a10_319 | | 541.0355 | 10.32 | | RP | POS | | | 489147 | 307025 | 527057 | 332377 | 3.88E-01 | 0.12 | 0.66 |
| RP_POS_541_87616a14_35 | | 541.8762 | 14.35 | | RP | POS | | | 799199 | 666063 | 905299 | 1053176 | 3.88E-01 | 0.12 | 0.9 |
| HILIC_POS_430_91461a5_472 | | 430.9146 | 5.47 | | HILIC | POS | | | 154010 | 360650 | 117164 | 263347 | 3.88E-01 | -0.12 | 0.52 |
| RP_POS_245_07646a0_677 | | 245.0765 | 0.68 | | RP | POS | | | 1897461 | 408060 | 1856690 | 280264 | 3.88E-01 | -0.12 | 0.74 |
| RP_POS_951_16833a9_704 | | 951.1683 | 9.70 | | RP | POS | | | 1213345 | 116121 | 1200701 | 98599.4 | 3.88E-01 | -0.12 | 0.54 |
| RP_POS_500_30389a9_292 | | 500.3039 | 9.29 | | RP | POS | 464.2795(100), 126.0214(40) | | 1718790 | 2168846 | 1986233 | 2347214 | 3.89E-01 | 0.12 | 0.85 |
| RP_POS_226_18004a7_657 | | 226.1800 | 7.66 | | RP | POS | | | 1123300 | 441942 | 1178355 | 487046 | 3.89E-01 | 0.12 | 0.92 |
| RP_POS_287_07574a7_983 | | 287.0757 | 7.98 | | RP | POS | | | 1075781 | 1438219 | 1263162 | 1708327 | 3.89E-01 | 0.12 | 1.05 |
| RP_POS_323_25549a10_437 | | 323.2555 | 10.44 | | RP | POS | | | 2148137 | 328251 | 2183895 | 280428 | 3.89E-01 | 0.12 | 1.03 |
| HILIC_POS_504_30688a1_311 | | 504.3069 | 1.31 | | HILIC | POS | | | 106156 | 63568.6 | 98418.2 | 67446.9 | 3.89E-01 | -0.12 | 0.72 |
| RP_NEG_301_23853a11_141 | | 301.2385 | 11.14 | | RP | NEG | | | 467371 | 123694 | 452868 | 122496 | 3.89E-01 | -0.12 | 1.26 |
| RP_POS_1050_66016a10_311 | | 1050.6602 | 10.31 | | RP | POS | | | 1218359 | 701289 | 1136939 | 682974 | 3.89E-01 | -0.12 | 0.8 |
| RP_POS_362_13843a9_209 | | 362.1384 | 9.21 | | RP | POS | | | 2523988 | 2744008 | 2214801 | 2521131 | 3.89E-01 | -0.12 | 1.17 |
| HILIC_NEG_355_01947a1_243 | | 355.0195 | 1.24 | | HILIC | NEG | | | 357600 | 148196 | 374752 | 143574 | 3.90E-01 | 0.12 | 0.57 |
| HILIC_POS_223_10738a5_15 | | 223.1074 | 5.15 | | HILIC | POS | | | 231842 | 121294 | 245274 | 107851 | 3.90E-01 | 0.12 | 0.61 |
| RP_POS_231_11594a13_295 | | 231.1159 | 13.30 | | RP | POS | | | 6493657 | 2208661 | 6770744 | 2472059 | 3.90E-01 | 0.12 | 0.54 |
| RP_POS_301_14053a14_246 | | 301.1405 | 14.25 | | RP | POS | | | 1212213 | 576501 | 1283717 | 633193 | 3.90E-01 | 0.12 | 0.86 |
| RP_POS_446_65991a10_264 | | 446.6599 | 10.26 | | RP | POS | | | 209854 | 123041 | 223988 | 117814 | 3.90E-01 | 0.12 | 0.73 |
| RP_POS_615_57983a12_943 | | 615.5798 | 12.94 | | RP | POS | | | 2316076 | 4879990 | 2950745 | 5793643 | 3.90E-01 | 0.12 | 0.7 |
| RP_POS_713_10254a11_537 | | 713.1025 | 11.54 | | RP | POS | | | 158853 | 90085.1 | 172522 | 134390 | 3.90E-01 | 0.12 | 0.93 |
| HILIC_POS_160_07567a1_598 | | 160.0757 | 1.60 | | HILIC | POS | 160.0751(100) | | 7699341 | 3860561 | 7263669 | 3570942 | 3.90E-01 | -0.12 | 0.65 |
| HILIC_NEG_244_98553a3_052 | | 244.9855 | 3.05 | | HILIC | NEG | | | 421541 | 39936.5 | 415578 | 58308.3 | 3.90E-01 | -0.12 | 0.65 |
| RP_POS_538_86786a10_001 | | 538.8679 | 10.00 | | RP | POS | | | 3293730 | 1463591 | 3098252 | 1811900 | 3.90E-01 | -0.12 | 0.78 |
| RP_POS_574_48523a14_355 | | 574.4852 | 14.36 | | RP | POS | | | 5.7E+07 | 1.5E+07 | 5.6E+07 | 1.2E+07 | 3.90E-01 | -0.12 | 1.13 |
| RP_NEG_321_17426a10_379 | | 321.1743 | 10.38 | | RP | NEG | | | 741917 | 2356887 | 1315410 | 6271111 | 3.91E-01 | 0.13 | 0.58 |
| RP_POS_157_04955a6_515 | | 157.0496 | 6.52 | | RP | POS | | | 216376 | 50478.8 | 222196 | 48825.8 | 3.91E-01 | 0.12 | 0.83 |
| RP_POS_798_60028a13_906 | | 798.6003 | 13.91 | | RP | POS | | | 3923471 | 3276030 | 4312958 | 3354266 | 3.91E-01 | 0.12 | 1.16 |
| RP_NEG_283_14069a11_59 | | 283.1407 | 11.59 | | RP | NEG | | | 168086 | 86637 | 156792 | 103718 | 3.91E-01 | -0.12 | 1.21 |
| RP_NEG_441_2533a10_58 | | 441.2533 | 10.58 | | RP | NEG | | | 36546.9 | 59722.5 | 30253.9 | 47753.2 | 3.91E-01 | -0.12 | 0.57 |
| RP_POS_166_0865a1_411 | | 166.0865 | 1.41 | | RP | POS | 120.0809(100), 166.0871(3) | | 2.3E+08 | 4.5E+07 | 2.4E+08 | 4.3E+07 | 3.92E-01 | 0.12 | 0.74 |
| HILIC_NEG_202_96533a0_942 | | 202.9653 | 0.94 | | HILIC | NEG | | | 1808812 | 773924 | 1906467 | 881208 | 3.92E-01 | 0.12 | 0.84 |
| RP_NEG_560_33105a10_93 | | 560.3311 | 10.93 | | RP | NEG | | | 1068564 | 166707 | 1089815 | 193100 | 3.92E-01 | 0.12 | 0.76 |
| RP_POS_203_10664a6_703 | | 203.1066 | 6.70 | | RP | POS | | | 1078565 | 225780 | 1105106 | 227680 | 3.92E-01 | 0.12 | 0.62 |
| RP_POS_369_04089a15_491 | | 369.0409 | 15.49 | | RP | POS | | | 490137 | 129354 | 507062 | 155927 | 3.92E-01 | 0.12 | 0.75 |
| RP_POS_553_36859a15_443 | | 553.3686 | 15.44 | | RP | POS | | | 694783 | 424635 | 747411 | 469255 | 3.92E-01 | 0.12 | 0.6 |
| RP_NEG_297_27994a11_838 | | 297.2799 | 11.84 | | RP | NEG | | | 344791 | 140870 | 327861 | 147763 | 3.92E-01 | -0.12 | 1.27 |
| RP_NEG_412_98355a8_254 | | 412.9836 | 8.25 | | RP | NEG | | | 1036249 | 727332 | 958699 | 600411 | 3.92E-01 | -0.12 | 0.67 |
| RP_POS_379_24243a12_365 | | 379.2424 | 12.37 | | RP | POS | | | 937776 | 689057 | 856405 | 698031 | 3.92E-01 | -0.12 | 0.73 |
| RP_POS_457_34982a10_93 | | 457.3498 | 10.93 | | RP | POS | | | 5592226 | 790666 | 5504611 | 710953 | 3.92E-01 | -0.12 | 1.06 |
| HILIC_NEG_115_92082a4_862 | | 115.9208 | 4.86 | | HILIC | NEG | | | 6.9E+07 | 2.3E+07 | 7.1E+07 | 2.4E+07 | 3.93E-01 | 0.12 | 0.87 |
| HILIC_POS_680_48083a10_66 | | 680.4808 | 10.66 | | HILIC | POS | | | 777221 | 92433.9 | 787396 | 82351 | 3.93E-01 | 0.12 | 0.64 |
| RP_NEG_563_97736a10_309 | | 563.9774 | 10.31 | | RP | NEG | | | 224354 | 135498 | 241738 | 158891 | 3.93E-01 | 0.12 | 0.68 |
| RP_POS_219_17426a11_641 | | 219.1743 | 11.64 | | RP | POS | | | 1659912 | 476555 | 1729850 | 684939 | 3.93E-01 | 0.12 | 0.78 |
| RP_POS_240_09979a10_607 | | 240.0998 | 10.61 | | RP | POS | | | 673826 | 258593 | 703540 | 250141 | 3.93E-01 | 0.12 | 0.99 |
| RP_POS_278_00836a0_586 | | 278.0084 | 0.59 | | RP | POS | | | 485481 | 122974 | 500521 | 133280 | 3.93E-01 | 0.12 | 0.79 |
| RP_POS_311_164a10_796 | | 311.1640 | 10.80 | | RP | POS | | | 2267874 | 7216219 | 3170678 | 8139954 | 3.93E-01 | 0.12 | 0.84 |
| RP_POS_558_39636a10_458 | | 558.3964 | 10.46 | | RP | POS | | | 950902 | 870486 | 1077599 | 1234409 | 3.93E-01 | 0.12 | 0.66 |
| HILIC_NEG_199_80489a4_207 | | 199.8049 | 4.21 | | HILIC | NEG | | | 135618 | 217415 | 110820 | 207927 | 3.93E-01 | -0.12 | 0.6 |
| RP_NEG_487_22763a11_593 | | 487.2276 | 11.59 | | RP | NEG | | | 1325304 | 262388 | 1295657 | 246141 | 3.93E-01 | -0.12 | 1.29 |
| RP_POS_274_90555a15_397 | | 274.9056 | 15.40 | | RP | POS | | | 1E+07 | 2045135 | 1E+07 | 1809592 | 3.93E-01 | -0.12 | 0.76 |
| RP_POS_455_33469a11_144 | | 455.3347 | 11.14 | | RP | POS | | | 461254 | 191345 | 442417 | 129939 | 3.93E-01 | -0.12 | 0.5 |
| RP_POS_830_56622a13_437 | | 830.5662 | 13.44 | | RP | POS | | | 3.2E+07 | 1.1E+07 | 3E+07 | 8156453 | 3.93E-01 | -0.12 | 0.77 |

**Supplemantary table 1**: The non-targeted metabolomics results ordered by the VIP value (p value < 0.0003 are shaded dark green, p value < 0.05 are shaded light green, d values below 0 are blue and above 0 are red)

| | | | | | | | | | Cases | | Controls | | Welch test | Cohen's | |
|---|---|---|---|---|---|---|---|---|---|---|---|---|---|---|---|
| Feature_ID | Metabolite_name | Average_Mz | Average_RT_min_ | HMDB ID | RP_HILIC | POS_NEG | Cureted_MSMS | ID_level | Mean | SD | Mean2 | SD3 | p | d | VIP |
| RP_POS_255_05852a9_357 | | 255.0585 | 9.36 | | RP | POS | 255.0601(100) | | 931535 | 308288 | 976116 | 435344 | 3.94E-01 | 0.12 | 0.5 |
| RP_NEG_242_17642a7_655 | | 242.1764 | 7.66 | | RP | NEG | | | 718707 | 328047 | 758670 | 354977 | 3.94E-01 | 0.12 | 0.81 |
| RP_POS_231_11588a0_636 | | 231.1159 | 0.64 | | RP | POS | | | 6539671 | 1005414 | 6653231 | 945099 | 3.94E-01 | 0.12 | 0.84 |
| RP_POS_354_03403a15_419 | | 354.0340 | 15.42 | | RP | POS | | | 7808459 | 2514846 | 8085601 | 2251218 | 3.94E-01 | 0.12 | 0.64 |
| RP_POS_537_53516a14_05 | | 537.5352 | 14.05 | | RP | POS | | | 8.6E+07 | 7069230 | 8.7E+07 | 7939793 | 3.94E-01 | 0.12 | 0.67 |
| HILIC_POS_218_13901a2_04 | Propionylcarnitine | 218.1390 | 2.04 | | HILIC | POS | 85.0291(100), 218.1393(40) | 1 | 5.2E+07 | 2E+07 | 5.4E+07 | 1.8E+07 | 3.94E-01 | 0.12 | 0.78 |
| HILIC_POS_106_08665a6_12 | | 106.0867 | 6.12 | | HILIC | POS | 88.0761(100), 70.066(60) | | 1.4E+07 | 5770058 | 1.3E+07 | 5007006 | 3.94E-01 | -0.12 | 0.68 |
| HILIC_NEG_194_98839a3_056 | | 194.9884 | 3.06 | | HILIC | NEG | | | 628617 | 82673.7 | 619758 | 69928.3 | 3.94E-01 | -0.12 | 0.78 |
| HILIC_POS_124_08727a6_265 | | 124.0873 | 6.27 | | HILIC | POS | | | 1986505 | 602094 | 1916545 | 598949 | 3.94E-01 | -0.12 | 0.55 |
| RP_POS_335_25601a10_876 | | 335.2560 | 10.88 | | RP | POS | | | 662290 | 380742 | 617881 | 380340 | 3.94E-01 | -0.12 | 0.73 |
| RP_POS_570_3125a7_199 | | 570.3125 | 7.20 | | RP | POS | | | 427051 | 46193 | 421419 | 50069.4 | 3.94E-01 | -0.12 | 0.8 |
| RP_POS_612_32562a10_612 | | 612.3256 | 10.61 | | RP | POS | | | 2632756 | 371728 | 2590052 | 360800 | 3.94E-01 | -0.12 | 0.67 |
| RP_POS_814_53552a13_684 | | 814.5355 | 13.68 | | RP | POS | | | 1781789 | 2541524 | 1544949 | 1461389 | 3.94E-01 | -0.12 | 0.69 |
| RP_NEG_179_07155a5_835 | | 179.0716 | 5.84 | | RP | NEG | | | 347806 | 160973 | 367819 | 180646 | 3.95E-01 | 0.12 | 1.15 |
| RP_POS_387_18268a11_039 | | 387.1827 | 11.04 | | RP | POS | | | 336997 | 279541 | 367670 | 248983 | 3.95E-01 | 0.12 | 1 |
| RP_POS_490_68506a10_22 | | 490.6851 | 10.22 | | RP | POS | | | 654440 | 293113 | 694224 | 377918 | 3.95E-01 | 0.12 | 0.83 |
| RP_POS_545_04907a10_023 | | 545.0491 | 10.02 | | RP | POS | | | 208047 | 128700 | 226524 | 180198 | 3.95E-01 | 0.12 | 0.61 |
| HILIC_NEG_212_91061a0_958 | | 212.9106 | 0.96 | | HILIC | NEG | | | 1588044 | 186819 | 1563721 | 226285 | 3.95E-01 | -0.12 | 0.63 |
| RP_POS_302_19583a1_964 | | 302.1958 | 1.96 | | RP | POS | | | 629179 | 430058 | 579870 | 418527 | 3.95E-01 | -0.12 | 0.86 |
| HILIC_NEG_138_89015a0_941 | | 138.8902 | 0.94 | | HILIC | NEG | | | 263580 | 58971.3 | 270173 | 54901.9 | 3.96E-01 | 0.12 | 1.06 |
| RP_NEG_293_17978a10_695 | | 293.1798 | 10.70 | | RP | NEG | | | 167090 | 114577 | 180659 | 118707 | 3.96E-01 | 0.12 | 0.53 |
| RP_POS_549_05328a10_166 | | 549.0533 | 10.17 | | RP | POS | | | 1025742 | 508768 | 1095831 | 672510 | 3.96E-01 | 0.12 | 0.62 |
| HILIC_NEG_526_31561a1_295 | | 526.3156 | 1.30 | | HILIC | NEG | | | 144264 | 41218.5 | 139390 | 42556.5 | 3.96E-01 | -0.12 | 0.68 |
| RP_NEG_146_04611a0_66 | | 146.0461 | 0.66 | | RP | NEG | | | 248638 | 41729.7 | 244265 | 33782.2 | 3.96E-01 | -0.12 | 0.69 |
| RP_POS_363_28677a10_828 | | 363.2868 | 10.83 | | RP | POS | | | 800170 | 146848 | 783847 | 134697 | 3.96E-01 | -0.12 | 0.91 |
| RP_POS_604_42798a7_766 | | 604.4280 | 7.77 | | RP | POS | | | 604919 | 484748 | 549841 | 465149 | 3.96E-01 | -0.12 | 0.66 |
| HILIC_POS_292_19806a6_958 | | 292.1981 | 6.96 | | HILIC | POS | | | 271150 | 94946.7 | 282676 | 103075 | 3.97E-01 | 0.12 | 0.62 |
| RP_POS_282_16968a8_412 | | 282.1697 | 8.41 | | RP | POS | | | 1082386 | 135951 | 1097617 | 127660 | 3.97E-01 | 0.12 | 1.02 |
| RP_POS_336_12814a6_701 | | 336.1281 | 6.70 | | RP | POS | | | 113209 | 34338.5 | 117546 | 39816.3 | 3.97E-01 | 0.12 | 0.69 |
| RP_POS_461_00015a10_247 | | 461.0002 | 10.25 | | RP | POS | | | 306866 | 179854 | 329066 | 200812 | 3.97E-01 | 0.12 | 0.72 |
| RP_POS_711_82654a11_407 | | 711.8265 | 11.41 | | RP | POS | | | 397640 | 476289 | 463004 | 628693 | 3.97E-01 | 0.12 | 0.67 |
| RP_POS_750_54303a13_506 | | 750.5430 | 13.51 | | RP | POS | | | 9929636 | 3778011 | 1E+07 | 3328297 | 3.97E-01 | 0.12 | 0.92 |
| RP_NEG_329_24884a11_036 | FA 22:5 (Docosapentaenoic acid) | 329.2488 | 11.04 | | RP | NEG | 329.2475(100), 285.2609(12) | 2 | 2799190 | 1986039 | 3016760 | 1783678 | 3.97E-01 | 0.12 | 1.12 |
| RP_POS_480_30884a10_595 | LysoPE 18:1 (18:1/0:0) | 480.3088 | 10.60 | | RP | POS | 339.2874(100) | 1 | 4.5E+07 | 1.5E+07 | 4.7E+07 | 1.7E+07 | 3.97E-01 | 0.12 | 0.7 |
| RP_NEG_693_12463a9_196 | | 693.1246 | 9.20 | | RP | NEG | | | 830708 | 59147.4 | 823923 | 58058.3 | 3.97E-01 | -0.12 | 0.64 |
| RP_POS_780_55115a11_142 | | 780.5512 | 11.14 | | RP | POS | | | 1103389 | 223198 | 1079215 | 195583 | 3.97E-01 | -0.12 | 0.68 |
| RP_POS_159_06532a4_825 | | 159.0653 | 4.83 | | RP | POS | | | 6.6E+07 | 3.6E+07 | 7.3E+07 | 7.6E+07 | 3.98E-01 | 0.13 | 0.73 |
| HILIC_NEG_274_7648a3_273 | | 274.7648 | 3.27 | | HILIC | NEG | | | 378145 | 166180 | 398369 | 181863 | 3.98E-01 | 0.12 | 0.54 |
| RP_POS_410_1019a9_524 | | 410.1019 | 9.52 | | RP | POS | | | 83764.6 | 37599.6 | 88740.8 | 47082.8 | 3.98E-01 | 0.12 | 0.68 |
| RP_POS_468_35684a11_224 | | 468.3568 | 11.22 | | RP | POS | | | 1422746 | 500515 | 1484609 | 561263 | 3.98E-01 | 0.12 | 0.66 |
| RP_NEG_207_0119a4_353 | | 207.0119 | 4.35 | | RP | NEG | | | 232392 | 163732 | 217035 | 99003.3 | 3.98E-01 | -0.12 | 0.62 |
| RP_POS_469_83582a9_604 | | 469.8358 | 9.60 | | RP | POS | | | 1563447 | 832037 | 1464622 | 872500 | 3.98E-01 | -0.12 | 0.89 |
| RP_POS_476_2742a10_473 | | 476.2742 | 10.47 | | RP | POS | | | 2438839 | 528611 | 2372970 | 602897 | 3.98E-01 | -0.12 | 0.69 |
| HILIC_NEG_202_96539a0_705 | | 202.9654 | 0.71 | | HILIC | NEG | | | 1231876 | 1128823 | 1378360 | 1376142 | 3.99E-01 | 0.12 | 0.93 |
| RP_POS_235_13261a8_067 | | 235.1326 | 8.07 | | RP | POS | | | 2337481 | 202983 | 2360669 | 199079 | 3.99E-01 | 0.12 | 0.89 |
| RP_POS_240_15681a11_581 | | 240.1568 | 11.58 | | RP | POS | | | 2005569 | 918805 | 2112483 | 932643 | 3.99E-01 | 0.12 | 0.51 |
| RP_POS_250_14091a9_996 | | 250.1409 | 10.00 | | RP | POS | | | 628507 | 663076 | 710576 | 749885 | 3.99E-01 | 0.12 | 1.13 |
| RP_POS_477_29236a11_854 | | 477.2924 | 11.85 | | RP | POS | | | 760426 | 117945 | 773668 | 111738 | 3.99E-01 | 0.12 | 0.72 |
| RP_POS_886_50781a13_01 | | 886.5078 | 13.01 | | RP | POS | | | 975942 | 421390 | 1027004 | 459067 | 3.99E-01 | 0.12 | 0.57 |
| HILIC_POS_371_23969a3_13 | | 371.2397 | 3.13 | | HILIC | POS | | | 399996 | 58768.1 | 394084 | 43737.5 | 3.99E-01 | -0.12 | 0.82 |
| HILIC_POS_461_3573a4_029 | | 461.3573 | 4.03 | | HILIC | POS | | | 1016698 | 1954272 | 805116 | 1723352 | 3.99E-01 | -0.12 | 0.85 |
| RP_POS_232_09174a11_147 | | 232.0917 | 11.15 | | RP | POS | | | 485720 | 160449 | 469168 | 127031 | 3.99E-01 | -0.12 | 0.76 |
| RP_POS_428_80167a10_119 | | 428.8017 | 10.12 | | RP | POS | | | 632193 | 430968 | 588703 | 323200 | 3.99E-01 | -0.12 | 0.64 |
| RP_NEG_1011_19a9_736 | | 1011.1900 | 9.74 | | RP | NEG | | | 1569551 | 229234 | 1599078 | 276789 | 4.00E-01 | 0.12 | 0.79 |
| RP_NEG_508_34131a10_931 | | 508.3413 | 10.93 | | RP | NEG | | | 4656967 | 939936 | 4770250 | 1020895 | 4.00E-01 | 0.12 | 0.81 |
| RP_POS_131_95065a0_624 | | 131.9507 | 0.62 | | RP | POS | | | 1042931 | 452951 | 1095867 | 464913 | 4.00E-01 | 0.12 | 0.7 |
| RP_POS_262_16461a5_053 | | 262.1646 | 5.05 | | RP | POS | | | 178239 | 80452.5 | 188512 | 96029.6 | 4.00E-01 | 0.12 | 0.83 |
| RP_POS_626_43896a11_438 | | 626.4390 | 11.44 | | RP | POS | | | 167354 | 86961.7 | 178735 | 107867 | 4.00E-01 | 0.12 | 0.71 |
| RP_POS_667_96783a11_522 | | 667.9678 | 11.52 | | RP | POS | | | 232152 | 136800 | 248373 | 144252 | 4.00E-01 | 0.12 | 0.68 |
| RP_POS_768_55042a13_381 | | 768.5504 | 13.38 | | RP | POS | | | 6432878 | 1.1E+07 | 7674012 | 1.1E+07 | 4.00E-01 | 0.12 | 0.87 |
| RP_NEG_211_91821a13_296 | | 211.9182 | 13.30 | | RP | NEG | | | 1303246 | 421931 | 1349204 | 378022 | 4.00E-01 | 0.11 | 0.73 |
| RP_POS_838_50714a13_288 | | 838.5071 | 13.29 | | RP | POS | | | 1349259 | 1144261 | 1476795 | 1077508 | 4.00E-01 | 0.11 | 0.84 |
| HILIC_NEG_540_33173a1_12 | LysoPC 16:0 | 540.3317 | 1.12 | | HILIC | NEG | 225.2324(100) | 2 | 1484434 | 417685 | 1531825 | 406148 | 4.00E-01 | 0.12 | 0.75 |
| RP_NEG_554_34796a10_662 | | 554.3480 | 10.66 | | RP | NEG | | | 7619000 | 2263770 | 7372247 | 2036848 | 4.00E-01 | -0.11 | 0.7 |
| RP_POS_454_2926a8_519 | | 454.2926 | 8.52 | | RP | POS | | | 458502 | 410254 | 413671 | 370804 | 4.00E-01 | -0.11 | 0.6 |
| HILIC_NEG_136_89172a3_108 | | 136.8917 | 3.11 | | HILIC | NEG | | | 9.7E+07 | 2457418 | 9.7E+07 | 2426688 | 4.00E-01 | -0.12 | 0.9 |
| RP_NEG_190_92865a15_532 | | 190.9287 | 15.53 | | RP | NEG | | | 3316017 | 942062 | 3197434 | 1099362 | 4.00E-01 | -0.12 | 0.73 |
| RP_NEG_198_92297a15_397 | | 198.9230 | 15.40 | | RP | NEG | | | 5386526 | 1103965 | 5258620 | 1115094 | 4.00E-01 | -0.12 | 0.78 |
| HILIC_NEG_200_85922a7_731 | | 200.8592 | 7.73 | | HILIC | NEG | | | 1463303 | 286650 | 1497789 | 311498 | 4.01E-01 | 0.12 | 0.59 |
| RP_POS_537_68256a10_178 | | 537.6826 | 10.18 | | RP | POS | | | 176590 | 86917.2 | 187393 | 99731.4 | 4.01E-01 | 0.12 | 0.64 |
| RP_NEG_614_34662a10_534 | | 614.3466 | 10.53 | | RP | NEG | | | 903384 | 370758 | 944554 | 346948 | 4.01E-01 | 0.11 | 0.69 |
| RP_NEG_526_3147a10_231 | LysoPC 15:0 | 526.3147 | 10.23 | | RP | NEG | 241.2188(100) | 1 | 1.3E+07 | 3415149 | 1.2E+07 | 3275554 | 4.01E-01 | -0.11 | 0.79 |
| HILIC_POS_249_08839a2_941 | | 249.0884 | 2.94 | | HILIC | POS | | | 64319 | 32263.6 | 60634.5 | 31912.3 | 4.01E-01 | -0.11 | 0.71 |
| HILIC_POS_261_07294a2_965 | | 261.0729 | 2.97 | | HILIC | POS | | | 204470 | 61764.4 | 197584 | 58342 | 4.01E-01 | -0.11 | 0.64 |
| RP_POS_269_13556a4_182 | | 269.1356 | 4.18 | | RP | POS | | | 184703 | 131681 | 170666 | 113262 | 4.01E-01 | -0.11 | 1.2 |
| RP_POS_378_20615a10_686 | | 378.2062 | 10.69 | | RP | POS | | | 1255007 | 285005 | 1224481 | 247731 | 4.01E-01 | -0.11 | 0.96 |
| RP_POS_397_75467a12_811 | | 397.7547 | 12.81 | | RP | POS | | | 1115416 | 600903 | 1047414 | 584009 | 4.01E-01 | -0.11 | 0.57 |
| RP_POS_746_56378a10_186 | | 746.5638 | 10.19 | | RP | POS | | | 1246191 | 642793 | 1173694 | 619508 | 4.01E-01 | -0.11 | 0.87 |
| RP_POS_756_55188a12_723 | PC 34:3 | 756.5519 | 12.72 | | RP | POS | 184.0743(100), 756.5604(2) | 1 | 6.6E+07 | 3.6E+07 | 7.1E+07 | 5E+07 | 4.01E-01 | 0.12 | 0.56 |
| RP_NEG_255_23285a11_142 | Palmitic Acid | 255.2329 | 11.14 | | RP | NEG | 255.2335(100) | 1 | 1.8E+07 | 5262705 | 1.7E+07 | 5116691 | 4.02E-01 | -0.11 | 1.29 |
| RP_NEG_311_16815a10_196 | | 311.1682 | 10.20 | | RP | NEG | 311.1699(100) | | 5878442 | 3321924 | 6411398 | 5527411 | 4.02E-01 | 0.12 | 0.69 |
| RP_NEG_355_26495a11_192 | | 355.2650 | 11.19 | | RP | NEG | | | 441606 | 275409 | 478350 | 353346 | 4.02E-01 | 0.12 | 0.89 |
| RP_NEG_581_52704a14_038 | | 581.5270 | 14.04 | | RP | NEG | | | 1.9E+07 | 657830 | 1.9E+07 | 708348 | 4.02E-01 | 0.12 | 1.05 |
| RP_NEG_640_29163a10_931 | | 640.2916 | 10.93 | | RP | NEG | | | 2344932 | 352733 | 2388177 | 396784 | 4.02E-01 | 0.12 | 0.8 |
| RP_POS_646_11713a11_333 | | 646.1171 | 11.33 | | RP | POS | | | 157526 | 82371.3 | 168841 | 110582 | 4.02E-01 | 0.12 | 0.69 |
| RP_POS_789_52051a13_205 | | 789.5205 | 13.21 | | RP | POS | | | 623088 | 518368 | 702647 | 811631 | 4.02E-01 | 0.12 | 0.82 |
| RP_NEG_192_99344a15_472 | | 192.9934 | 15.47 | | RP | NEG | | | 3299588 | 232210 | 3325613 | 222804 | 4.02E-01 | 0.11 | 0.89 |
| HILIC_POS_172_13319a0_816 | | 172.1332 | 0.82 | | HILIC | POS | 172.1335(100) | | 1930111 | 729994 | 1848777 | 691136 | 4.02E-01 | -0.11 | 0.79 |
| RP_POS_539_31299a10_309 | | 539.3130 | 10.31 | | RP | POS | | | 1603155 | 846675 | 1510844 | 769066 | 4.02E-01 | -0.11 | 1.02 |
| RP_POS_679_44897a12_035 | | 679.4490 | 12.04 | | RP | POS | | | 321388 | 172459 | 302889 | 150961 | 4.02E-01 | -0.11 | 0.73 |
| RP_POS_681_49707a11_665 | | 681.4971 | 11.67 | | RP | POS | | | 221835 | 164679 | 204708 | 134906 | 4.02E-01 | -0.11 | 0.61 |
| HILIC_POS_156_04207a2_995 | | 156.0421 | 3.00 | | HILIC | POS | | | 2320342 | 336421 | 2270840 | 498435 | 4.02E-01 | -0.12 | 0.62 |
| RP_NEG_249_11293a7_657 | | 249.1129 | 7.66 | | RP | NEG | | | 1924644 | 128743 | 1908748 | 146542 | 4.02E-01 | -0.12 | 0.72 |
| RP_POS_417_28235a8_762 | | 417.2824 | 8.76 | | RP | POS | | | 175315 | 143220 | 193291 | 167651 | 4.03E-01 | 0.12 | 0.79 |
| RP_POS_724_31079a11_595 | | 724.3108 | 11.60 | | RP | POS | | | 100920 | 63452.8 | 110755 | 100956 | 4.03E-01 | 0.12 | 0.67 |
| RP_POS_255_05867a14_24 | | 255.0587 | 14.24 | | RP | POS | 255.0599(100) | | 1625569 | 768167 | 1709027 | 693953 | 4.03E-01 | 0.11 | 0.66 |
| RP_POS_332_14325a10_712 | | 332.1433 | 10.71 | | RP | POS | | | 191587 | 176522 | 210409 | 153705 | 4.03E-01 | 0.11 | 0.88 |
| RP_POS_353_245a11_038 | | 353.2450 | 11.04 | | RP | POS | | | 4947135 | 3273626 | 5300945 | 2935457 | 4.03E-01 | 0.11 | 1.06 |
| RP_POS_418_17981a2_885 | | 418.1798 | 2.89 | | RP | POS | | | 649565 | 128483 | 664445 | 131486 | 4.03E-01 | 0.11 | 0.64 |
| RP_POS_232_15428a2_183 | Butanoylcarnitine | 232.1543 | 2.18 | | RP | POS | 85.0291(100), 232.1538(40) | 1 | 3322842 | 1820991 | 3141055 | 1362235 | 4.03E-01 | -0.11 | 0.79 |
| RP_POS_329_12738a9_208 | | 329.1274 | 9.21 | | RP | POS | | | 3254626 | 2051647 | 3010326 | 2198872 | 4.03E-01 | -0.11 | 1.1 |
| RP_POS_345_66931a5_944 | | 345.6693 | 5.94 | | RP | POS | | | 204196 | 121925 | 191555 | 99698 | 4.03E-01 | -0.11 | 0.8 |
| RP_POS_382_98792a15_449 | | 382.9879 | 15.45 | | RP | POS | | | 290084 | 89913.3 | 279544 | 94045.1 | 4.03E-01 | -0.11 | 1.16 |
| RP_NEG_832_51764a10_608 | | 832.5176 | 10.61 | | RP | NEG | | | 159265 | 71299.2 | 168316 | 85338.1 | 4.04E-01 | 0.12 | 0.51 |
| RP_POS_510_6936a9_883 | | 510.6936 | 9.88 | | RP | POS | | | 375361 | 207456 | 401865 | 251086 | 4.04E-01 | 0.12 | 0.91 |
| RP_POS_602_42297a10_394 | | 602.4230 | 10.39 | | RP | POS | | | 647922 | 311839 | 686871 | 363918 | 4.04E-01 | 0.12 | 0.66 |
| RP_NEG_813_5387a12_361 | | 813.5387 | 12.36 | | RP | NEG | | | 2953970 | 451775 | 3006168 | 461752 | 4.04E-01 | 0.11 | 0.62 |
| RP_POS_314_09a15_323 | | 314.0900 | 15.32 | | RP | POS | | | 8824991 | 1688350 | 9012178 | 1594028 | 4.04E-01 | 0.11 | 0.63 |
| RP_POS_408_22333a6_586 | | 408.2233 | 6.59 | | RP | POS | | | 1162960 | 81743.1 | 1172264 | 81268.2 | 4.04E-01 | 0.11 | 0.83 |
| RP_NEG_426_96817a8_312 | 6:2 Fluorotelomer sulfonic acid | 426.9682 | 8.31 | | RP | NEG | 429.9683(65), 406.9645(45) | 2 | 1.1E+07 | 558035 | 1.1E+07 | 533530 | 4.04E-01 | -0.11 | 0.78 |
| RP_NEG_149_93709a0_647 | | 149.9371 | 0.65 | | RP | NEG | | | 1099903 | 416495 | 1052273 | 418018 | 4.04E-01 | -0.11 | 0.77 |
| RP_NEG_194_98888a15_484 | | 194.9889 | 15.48 | | RP | NEG | | | 2240101 | 538119 | 2184884 | 432836 | 4.04E-01 | -0.11 | 0.56 |
| RP_POS_164_98476a15_44 | | 164.9848 | 15.44 | | RP | POS | | | 3E+07 | 1.1E+07 | 2.9E+07 | 1.1E+07 | 4.04E-01 | -0.11 | 0.56 |
| RP_POS_352_24603a11_144 | | 352.2460 | 11.14 | | RP | POS | | | 2790178 | 494769 | 2733075 | 504494 | 4.04E-01 | -0.11 | 0.88 |
| RP_POS_803_53821a12_741 | | 803.5382 | 12.74 | | RP | POS | | | 4.4E+07 | 2E+07 | 4.2E+07 | 1.5E+07 | 4.04E-01 | -0.11 | 0.53 |
| RP_POS_808_48535a12_937 | | 808.4854 | 12.94 | | RP | POS | | | 1017411 | 368446 | 974756 | 377058 | 4.04E-01 | -0.11 | 0.58 |
| RP_NEG_128_03545a0_681 | Pyroglutamic acid | 128.0355 | 0.68 | | RP | NEG | 128.0356(100), 82.0295(5) | 2 | 4909284 | 871903 | 4806768 | 918649 | 4.04E-01 | -0.11 | 0.57 |
| HILIC_POS_195_12276a3_059 | | 195.1228 | 3.06 | | HILIC | POS | | | 806340 | 250906 | 838055 | 299135 | 4.05E-01 | 0.12 | 1.01 |
| RP_POS_373_25854a10_253 | | 373.2585 | 10.25 | | RP | POS | | | 2.1E+07 | 1E+07 | 2.2E+07 | 9559158 | 4.05E-01 | 0.11 | 1.09 |
| RP_POS_496_33942a10_357 | LysoPC 16:0 (0:0/16:0) | 496.3394 | 10.36 | | RP | POS | 184.0743(100), 104.1078(70) | 1 | 5.8E+08 | 1.1E+08 | 6E+08 | 1.3E+08 | 4.05E-01 | 0.12 | 0.97 |
| HILIC_NEG_236_81674a3_343 | | 236.8167 | 3.34 | | HILIC | NEG | | | 1046258 | 674959 | 977300 | 539961 | 4.05E-01 | -0.11 | 0.72 |
| HILIC_POS_90_97723a4_336 | | 90.9772 | 4.34 | | HILIC | POS | | | 3.8E+08 | 1.8E+07 | 3.8E+08 | 1.9E+07 | 4.05E-01 | -0.11 | 0.89 |
| RP_NEG_807_18091a9_065 | | 807.1809 | 9.07 | | RP | NEG | | | 137490 | 302940 | 109902 | 174774 | 4.05E-01 | -0.12 | 0.55 |
| RP_POS_451_68188a5_62 | | 451.6819 | 5.62 | | RP | POS | | | 168808 | 116197 | 184954 | 160747 | 4.06E-01 | 0.12 | 0.73 |
| RP_POS_181_08565a9_79 | | 181.0857 | 9.79 | | RP | POS | | | 1261865 | 155931 | 1281275 | 182123 | 4.06E-01 | 0.11 | 0.74 |
| RP_POS_294_20627a9_061 | | 294.2063 | 9.06 | | RP | POS | | | 557131 | 81462.2 | 566271 | 79419.8 | 4.06E-01 | 0.11 | 0.91 |
| RP_POS_456_33453a9_615 | | 456.3345 | 9.62 | | RP | POS | | | 4165439 | 1998324 | 4404537 | 2188602 | 4.06E-01 | 0.11 | 0.83 |
| HILIC_POS_482_32434a1_294 | LysoPC 15:0 | 482.3243 | 1.29 | | HILIC | POS | 104.1078(100), 184.0745(55) | 2 | 5261743 | 1660776 | 5082671 | 1498646 | 4.06E-01 | -0.11 | 0.82 |
| RP_NEG_391_20834a11_142 | | 391.2083 | 11.14 | | RP | NEG | | | 1460844 | 342960 | 1423117 | 322528 | 4.06E-01 | -0.11 | 1.3 |
| RP_POS_218_03317a15_383 | | 218.0332 | 15.38 | | RP | POS | | | 603447 | 299343 | 571544 | 263274 | 4.06E-01 | -0.11 | 0.55 |
| RP_POS_669_96997a11_616 | | 669.9700 | 11.62 | | RP | POS | | | 138075 | 61886.9 | 130830 | 65233.3 | 4.06E-01 | -0.11 | 0.63 |
| RP_POS_818_50861a12_74 | | 818.5086 | 12.74 | | RP | POS | | | 6403433 | 3680471 | 6013911 | 3196710 | 4.06E-01 | -0.11 | 0.72 |
| HILIC_NEG_130_99274a5_113 | | 130.9927 | 5.11 | | HILIC | NEG | | | 1726636 | 834231 | 1841919 | 1148109 | 4.07E-01 | 0.12 | 0.61 |
| RP_NEG_748_51495a12_435 | | 748.5150 | 12.44 | | RP | NEG | | | 1320160 | 859478 | 1433990 | 1109530 | 4.07E-01 | 0.12 | 0.53 |
| RP_POS_388_25378a4_324 | | 388.2538 | 4.32 | | RP | POS | | | 849833 | 262940 | 891152 | 431353 | 4.07E-01 | 0.12 | 0.62 |
| RP_POS_483_38049a13_138 | | 483.3805 | 13.14 | | RP | POS | | | 291537 | 310619 | 333308 | 409757 | 4.07E-01 | 0.12 | 0.83 |
| RP_POS_670_94513a9_918 | | 670.9451 | 9.92 | | RP | POS | | | 680713 | 369846 | 731574 | 505532 | 4.07E-01 | 0.12 | 1.05 |
| HILIC_NEG_588_33051a1_154 | | 588.3305 | 1.15 | | HILIC | NEG | | | 505499 | 157091 | 524201 | 171354 | 4.07E-01 | 0.11 | 0.69 |
| HILIC_POS_103_03934a4_797 | | 103.0393 | 4.80 | | HILIC | POS | | | 157245 | 118462 | 171027 | 124160 | 4.07E-01 | 0.11 | 0.5 |
| RP_POS_259_08389a6_806 | | 259.0839 | 6.81 | | RP | POS | 259.0832(100) | | 1889560 | 4996864 | 1450065 | 2563144 | 4.07E-01 | -0.12 | 0.62 |
| RP_POS_511_51978a10_716 | | 511.5198 | 10.72 | | RP | POS | | | 257716 | 256266 | 297908 | 419870 | 4.08E-01 | 0.12 | 0.96 |
| HILIC_POS_155_10641a3_101 | | 155.1064 | 3.10 | | HILIC | POS | | | 108702 | 78051.1 | 116892 | 67276.1 | 4.08E-01 | 0.11 | 0.66 |
| HILIC_POS_346_02063a7_827 | | 346.0206 | 7.83 | | HILIC | POS | | | 637297 | 630442 | 705726 | 582523 | 4.08E-01 | 0.11 | 0.58 |
| RP_POS_125_98641a15_387 | | 125.9864 | 15.39 | | RP | POS | | | 157811 | 103187 | 169273 | 99657.7 | 4.08E-01 | 0.11 | 0.62 |
| RP_POS_398_76056a15_394 | | 398.7606 | 15.39 | | RP | POS | | | 315952 | 150891 | 335287 | 185668 | 4.08E-01 | 0.11 | 0.81 |
| RP_NEG_478_2941a10_593 | LysoPE 18:1 (18:1/0:0) | 478.2941 | 10.59 | | RP | NEG | 281.2493(100), 478.2952(10) | 1 | 1.7E+07 | 5647198 | 1.8E+07 | 6553202 | 4.08E-01 | 0.11 | 0.69 |
| RP_NEG_456_86285a15_386 | | 456.8629 | 15.39 | | RP | NEG | | | 754735 | 126828 | 738431 | 157106 | 4.08E-01 | -0.11 | 0.61 |
| RP_NEG_516_28821a10_232 | | 516.2882 | 10.23 | | RP | NEG | | | 370938 | 101851 | 359749 | 96013.8 | 4.08E-01 | -0.11 | 0.76 |
| RP_NEG_632_31848a10_314 | | 632.3185 | 10.31 | | RP | NEG | | | 1.1E+07 | 1677590 | 1.1E+07 | 1793730 | 4.08E-01 | -0.11 | 0.78 |
| RP_NEG_723_0874a9_099 | | 723.0874 | 9.10 | | RP | NEG | | | 5280346 | 240642 | 5252633 | 248195 | 4.08E-01 | -0.11 | 0.96 |
| RP_POS_784_55048a10_479 | | 784.5505 | 10.48 | | RP | POS | | | 725620 | 376011 | 682341 | 388100 | 4.08E-01 | -0.11 | 0.7 |
| HILIC_POS_534_29688a1_268 | | 534.2969 | 1.27 | | HILIC | POS | | | 1925236 | 757580 | 2009050 | 727219 | 4.09E-01 | 0.11 | 0.49 |
| RP_NEG_271_2283a9_643 | | 271.2283 | 9.64 | | RP | NEG | 271.2277(100), 225.224(1) | | 3468736 | 448721 | 3419541 | 424176 | 4.09E-01 | -0.11 | 0.93 |
| HILIC_POS_375_32169a3_281 | | 375.3217 | 3.28 | | HILIC | POS | | | 225616 | 120833 | 241978 | 162972 | 4.10E-01 | 0.12 | 0.86 |
| RP_POS_258_01196a4_409 | | 258.0120 | 4.41 | | RP | POS | | | 8.4E+07 | 2.4E+07 | 8.9E+07 | 4.8E+07 | 4.10E-01 | 0.12 | 1.12 |
| RP_POS_348_10114a3_82 | | 348.1011 | 3.82 | | RP | POS | | | 4457029 | 5097389 | 5150709 | 6927507 | 4.10E-01 | 0.12 | 0.93 |
| HILIC_POS_125_98651a7_99 | | 125.9865 | 7.99 | | HILIC | POS | | | 3824024 | 1589979 | 4012750 | 1738775 | 4.10E-01 | 0.11 | 0.67 |
| RP_NEG_377_1431a10_714 | | 377.1431 | 10.71 | | RP | NEG | | | 258048 | 233519 | 284920 | 242217 | 4.10E-01 | 0.11 | 0.88 |
| RP_NEG_676_30676a10_36 | | 676.3068 | 10.36 | | RP | NEG | | | 1961731 | 191581 | 1984937 | 216751 | 4.10E-01 | 0.11 | 0.92 |
| RP_POS_193_15854a9_083 | | 193.1585 | 9.08 | | RP | POS | | | 1.5E+07 | 947226 | 1.5E+07 | 1005665 | 4.10E-01 | 0.11 | 0.79 |
| RP_POS_635_58752a11_503 | | 635.5875 | 11.50 | | RP | POS | | | 98708.6 | 57458.4 | 105339 | 60047.1 | 4.10E-01 | 0.11 | 0.63 |
| RP_POS_953_54065a12_617 | | 953.5407 | 12.62 | | RP | POS | | | 424558 | 626095 | 497848 | 670122 | 4.10E-01 | 0.11 | 0.81 |

**Supplemantary table 1**: The non-targeted metabolomics results ordered by the VIP value (p value < 0.0003 are shaded dark green, p value < 0.05 are shaded light green, d values below 0 are blue and above 0 are red)

| | | | | | | | | | Cases | | Controls | | Welch test | Cohen's | |
|---|---|---|---|---|---|---|---|---|---|---|---|---|---|---|---|
| Feature_ID | Metabolite_name | Average_Mz | Average_RT_min_ | HMDB ID | RP_HILIC | POS_NEG | Cureted_MSMS | ID_level | Mean | SD | Mean2 | SD3 | p | d | VIP |
| RP_POS_280_26343a10_546 | Linoleamide | 280.2634 | 10.55 | | RP | POS | 263.2318(100) | 1 | 1.4E+07 | 5524086 | 1.4E+07 | 9004926 | 4.10E-01 | 0.12 | 1.09 |
| RP_NEG_260_02298a0_68 | | 260.0230 | 0.68 | | RP | NEG | | | 747644 | 143348 | 732798 | 120801 | 4.10E-01 | -0.11 | 0.64 |
| HILIC_NEG_308_99869a2_267 | | 308.9987 | 2.27 | | HILIC | NEG | | | 736217 | 201225 | 758798 | 200595 | 4.11E-01 | 0.11 | 0.86 |
| RP_NEG_157_03673a0_548 | | 157.0367 | 0.55 | | RP | NEG | | | 479317 | 233357 | 504162 | 209516 | 4.11E-01 | 0.11 | 0.64 |
| RP_NEG_174_95621a14_343 | | 174.9562 | 14.34 | | RP | NEG | | | 2974452 | 1535321 | 3145207 | 1504419 | 4.11E-01 | 0.11 | 0.77 |
| RP_NEG_190_94969a0_86 | | 190.9497 | 0.86 | | RP | NEG | | | 1271710 | 355583 | 1309672 | 320931 | 4.11E-01 | 0.11 | 0.83 |
| RP_POS_223_1118a8_732 | | 223.1118 | 8.73 | | RP | POS | | | 1870598 | 683060 | 1955596 | 811594 | 4.11E-01 | 0.11 | 0.68 |
| RP_POS_394_75955a12_807 | | 394.7596 | 12.81 | | RP | POS | | | 159547 | 119605 | 173231 | 123424 | 4.11E-01 | 0.11 | 0.69 |
| RP_POS_417_23969a10_943 | | 417.2397 | 10.94 | | RP | POS | | | 120399 | 116829 | 133781 | 120912 | 4.11E-01 | 0.11 | 0.91 |
| RP_POS_765_98737a10_357 | | 765.9874 | 10.36 | | RP | POS | | | 1658892 | 618272 | 1736326 | 743121 | 4.11E-01 | 0.11 | 1.07 |
| RP_NEG_187_0416a0_68 | Psoralen | 187.0416 | 0.68 | | RP | NEG | 187.0418(100), 115.0211(40) | 2 | 1651775 | 359713 | 1695189 | 406484 | 4.11E-01 | 0.11 | 0.84 |
| RP_NEG_312_76935a0_644 | | 312.7694 | 0.64 | | RP | NEG | | | 456084 | 146031 | 441574 | 112357 | 4.11E-01 | -0.11 | 0.6 |
| RP_NEG_354_06845a0_684 | | 354.0685 | 0.68 | | RP | NEG | | | 663191 | 205884 | 641989 | 172502 | 4.11E-01 | -0.11 | 0.64 |
| RP_NEG_898_58008a14_099 | | 898.5801 | 14.10 | | RP | NEG | | | 1.3E+07 | 3166499 | 1.3E+07 | 3256003 | 4.11E-01 | -0.11 | 0.69 |
| RP_POS_171_03342a15_395 | | 171.0334 | 15.40 | | RP | POS | | | 2329822 | 1178090 | 2209334 | 971334 | 4.11E-01 | -0.11 | 0.98 |
| RP_POS_358_36771a9_558 | | 358.3677 | 9.56 | | RP | POS | | | 1064407 | 3275674 | 741901 | 2462398 | 4.11E-01 | -0.11 | 0.49 |
| RP_POS_268_94067a15_394 | | 268.9407 | 15.39 | | RP | POS | | | 187221 | 131243 | 201903 | 130465 | 4.12E-01 | 0.11 | 0.71 |
| RP_POS_589_39813a10_401 | | 589.3981 | 10.40 | | RP | POS | | | 558883 | 294362 | 594939 | 342499 | 4.12E-01 | 0.11 | 0.55 |
| HILIC_POS_245_09972a4_221 | | 245.0997 | 4.22 | | HILIC | POS | 245.0993(100) | | 342648 | 171208 | 325093 | 142339 | 4.12E-01 | -0.11 | 0.81 |
| RP_POS_239_12506a8_097 | | 239.1251 | 8.10 | | RP | POS | 239.125(100) | | 1891713 | 802708 | 1806818 | 714941 | 4.12E-01 | -0.11 | 1.04 |
| HILIC_POS_403_01102a4_819 | | 403.0110 | 4.82 | | HILIC | POS | | | 210263 | 54296 | 203485 | 65053.3 | 4.12E-01 | -0.11 | 0.6 |
| HILIC_POS_498_90176a4_338 | | 498.9018 | 4.34 | | HILIC | POS | | | 1.2E+07 | 711689 | 1.2E+07 | 746762 | 4.12E-01 | -0.11 | 0.8 |
| RP_NEG_285_03168a0_687 | | 285.0317 | 0.69 | | RP | NEG | | | 220718 | 82165 | 212811 | 58814.3 | 4.12E-01 | -0.11 | 0.56 |
| RP_NEG_880_60803a14_116 | | 880.6080 | 14.12 | | RP | NEG | | | 3500472 | 1475398 | 3335733 | 1461361 | 4.12E-01 | -0.11 | 0.54 |
| RP_POS_217_10403a2_069 | | 217.1040 | 2.07 | | RP | POS | | | 1036929 | 540539 | 979585 | 483374 | 4.12E-01 | -0.11 | 0.88 |
| RP_POS_206_13841a3_568 | | 206.1384 | 3.57 | | RP | POS | | | 6.10E+15 | 1.37E+16 | 9.37E+15 | 3.75E+16 | 4.13E-01 | 0.13 | 0.88 |
| HILIC_POS_203_10254a7_746 | | 203.1025 | 7.75 | | HILIC | POS | | | 462651 | 39965.6 | 467396 | 44289.4 | 4.13E-01 | 0.11 | 0.71 |
| HILIC_POS_384_77188a1_151 | | 384.7719 | 1.15 | | HILIC | POS | | | 381162 | 531063 | 441742 | 549466 | 4.13E-01 | 0.11 | 0.6 |
| RP_POS_141_05458a10_254 | | 141.0546 | 10.25 | | RP | POS | | | 944450 | 436464 | 992475 | 422141 | 4.13E-01 | 0.11 | 1.1 |
| RP_POS_207_96619a15_538 | | 207.9662 | 15.54 | | RP | POS | | | 617289 | 297570 | 649770 | 283103 | 4.13E-01 | 0.11 | 0.55 |
| RP_POS_245_07848a7_47 | Beta-uridine | 245.0785 | 7.47 | | RP | POS | 245.077(100), | 2 | 3842171 | 674743 | 3773559 | 554239 | 4.13E-01 | -0.11 | 1.03 |
| HILIC_POS_694_4964a10_662 | | 694.4964 | 10.66 | | HILIC | POS | | | 179829 | 42852.6 | 174748 | 47398.4 | 4.13E-01 | -0.11 | 0.51 |
| RP_POS_198_18513a13_04 | | 198.1851 | 13.04 | | RP | POS | | | 1.7E+07 | 6919905 | 1.6E+07 | 7047033 | 4.13E-01 | -0.11 | 0.63 |
| RP_POS_357_27872a8_852 | | 357.2787 | 8.85 | | RP | POS | | | 1279766 | 3081389 | 987757 | 2122860 | 4.13E-01 | -0.11 | 0.81 |
| RP_POS_738_50671a12_669 | | 738.5067 | 12.67 | | RP | POS | | | 1879938 | 2024890 | 1675375 | 1638952 | 4.13E-01 | -0.11 | 0.61 |
| RP_NEG_815_55408a12_805 | | 815.5541 | 12.81 | | RP | NEG | | | 4558583 | 711669 | 4640605 | 751352 | 4.14E-01 | 0.11 | 0.96 |
| RP_POS_297_9285a15_383 | | 297.9285 | 15.38 | | RP | POS | | | 939896 | 612788 | 1004447 | 545377 | 4.14E-01 | 0.11 | 0.85 |
| RP_POS_345_13052a5_445 | | 345.1305 | 5.45 | | RP | POS | | | 507185 | 491390 | 562771 | 501937 | 4.14E-01 | 0.11 | 0.85 |
| RP_POS_833_58539a13_952 | | 833.5854 | 13.95 | | RP | POS | | | 4.8E+07 | 2.7E+07 | 5.1E+07 | 2.6E+07 | 4.14E-01 | 0.11 | 1.13 |
| RP_POS_291_64691a10_487 | | 291.6469 | 10.49 | | RP | POS | | | 601982 | 318911 | 566877 | 309438 | 4.14E-01 | -0.11 | 0.75 |
| RP_POS_341_26608a10_51 | | 341.2661 | 10.51 | | RP | POS | | | 1476482 | 365713 | 1436747 | 346602 | 4.14E-01 | -0.11 | 1 |
| RP_POS_603_79065a4_572 | | 603.7907 | 4.57 | | RP | POS | | | 2589896 | 2797262 | 2329878 | 1839650 | 4.14E-01 | -0.11 | 1.08 |
| RP_POS_816_5896a13_421 | | 816.5896 | 13.42 | | RP | POS | | | 2611354 | 1734212 | 2423951 | 1627512 | 4.14E-01 | -0.11 | 0.65 |
| RP_NEG_192_98964a5_443 | | 192.9896 | 5.44 | | RP | NEG | | | 2632102 | 510974 | 2685105 | 442483 | 4.15E-01 | 0.11 | 0.5 |
| RP_NEG_510_25357a8_22 | | 510.2536 | 8.22 | | RP | NEG | | | 4857962 | 2188937 | 5127713 | 2595450 | 4.15E-01 | 0.11 | 0.72 |
| RP_POS_307_09662a7_585 | | 307.0966 | 7.59 | | RP | POS | | | 936419 | 193924 | 957260 | 180835 | 4.15E-01 | 0.11 | 0.83 |
| RP_POS_400_22632a11_237 | | 400.2263 | 11.24 | | RP | POS | | | 667636 | 366631 | 704690 | 299950 | 4.15E-01 | 0.11 | 0.96 |
| HILIC_NEG_149_04543a5_116 | | 149.0454 | 5.12 | | HILIC | NEG | | | 538373 | 324392 | 503580 | 301338 | 4.15E-01 | -0.11 | 0.65 |
| HILIC_POS_139_98801a0_93 | | 139.9880 | 0.93 | | HILIC | POS | | | 324879 | 82516 | 315768 | 80905.6 | 4.15E-01 | -0.11 | 0.87 |
| RP_POS_248_03452a3_575 | | 248.0345 | 3.58 | | RP | POS | | | 533618 | 257804 | 506109 | 237029 | 4.15E-01 | -0.11 | 0.84 |
| RP_POS_573_2533a7_071 | | 573.2533 | 7.07 | | RP | POS | | | 5.3E+07 | 1.1E+07 | 5.2E+07 | 1E+07 | 4.15E-01 | -0.11 | 1.24 |
| HILIC_NEG_118_92409a3_989 | | 118.9241 | 3.99 | | HILIC | NEG | | | 2671946 | 1217811 | 2832888 | 1611300 | 4.16E-01 | 0.11 | 0.59 |
| RP_POS_338_18085a5_05 | | 338.1809 | 5.05 | | RP | POS | | | 313982 | 25764.7 | 317040 | 28842.2 | 4.16E-01 | 0.11 | 0.53 |
| RP_POS_608_52472a13_907 | | 608.5247 | 13.91 | | RP | POS | | | 916507 | 495435 | 974157 | 537085 | 4.16E-01 | 0.11 | 0.69 |
| HILIC_NEG_134_89474a4_273 | | 134.8947 | 4.27 | | HILIC | NEG | | | 1.8E+07 | 6356901 | 1.8E+07 | 5788265 | 4.16E-01 | -0.11 | 0.9 |
| HILIC_NEG_155_95219a1_35 | | 155.9522 | 1.35 | | HILIC | NEG | | | 701732 | 193248 | 681729 | 167291 | 4.16E-01 | -0.11 | 0.77 |
| HILIC_NEG_221_84203a3_099 | | 221.8420 | 3.10 | | HILIC | NEG | | | 157954 | 60636.2 | 150963 | 64591.8 | 4.16E-01 | -0.11 | 0.52 |
| RP_POS_265_25275a10_196 | | 265.2528 | 10.20 | | RP | POS | | | 3657762 | 543158 | 3599858 | 499184 | 4.16E-01 | -0.11 | 1.03 |
| RP_NEG_338_87427a15_551 | | 338.8743 | 15.55 | | RP | NEG | | | 3618696 | 207172 | 3587433 | 330058 | 4.16E-01 | -0.12 | 1.24 |
| RP_POS_310_20056a9_451 | | 310.2006 | 9.45 | | RP | POS | | | 925585 | 313091 | 963081 | 356880 | 4.17E-01 | 0.11 | 0.91 |
| RP_POS_524_29962a10_361 | | 524.2996 | 10.36 | | RP | POS | | | 1526113 | 480197 | 1582910 | 535608 | 4.17E-01 | 0.11 | 1.01 |
| HILIC_POS_144_10213a5_115 | | 144.1021 | 5.12 | | HILIC | POS | 144.1026(100) 58.0663(30) | | 1362954 | 2152977 | 1161671 | 1462814 | 4.17E-01 | -0.11 | 0.5 |
| RP_POS_1046_84387a14_355 | | 1046.8439 | 14.36 | | RP | POS | | | 4290462 | 1631714 | 4133998 | 1191289 | 4.17E-01 | -0.11 | 0.56 |
| RP_POS_158_96313a15_409 | | 158.9631 | 15.41 | | RP | POS | | | 7721036 | 3250959 | 7352561 | 3370647 | 4.17E-01 | -0.11 | 1.09 |
| RP_POS_165_09082a13_148 | | 165.0908 | 13.15 | | RP | POS | | | 5826364 | 1486857 | 5653048 | 1618290 | 4.17E-01 | -0.11 | 0.76 |
| RP_POS_309_20306a9_927 | | 309.2031 | 9.93 | | RP | POS | | | 1.5E+07 | 7749503 | 1.5E+07 | 7908623 | 4.17E-01 | -0.11 | 0.74 |
| HILIC_NEG_157_90088a7_094 | | 157.9009 | 7.09 | | HILIC | NEG | | | 1043045 | 454147 | 1092633 | 442236 | 4.18E-01 | 0.11 | 0.55 |
| RP_NEG_413_19281a10_714 | | 413.1928 | 10.71 | | RP | NEG | | | 99279.4 | 89059.6 | 108863 | 84274.3 | 4.18E-01 | 0.11 | 0.9 |
| RP_NEG_480_31018a10_358 | | 480.3102 | 10.36 | | RP | NEG | | | 3930454 | 696572 | 4011914 | 767345 | 4.18E-01 | 0.11 | 1 |
| RP_NEG_819_15216a9_629 | | 819.1522 | 9.63 | | RP | NEG | | | 2262074 | 144975 | 2277750 | 138249 | 4.18E-01 | 0.11 | 0.66 |
| RP_POS_162_07614a10_426 | | 162.0761 | 10.43 | | RP | POS | | | 4726853 | 2026513 | 4939539 | 1826446 | 4.18E-01 | 0.11 | 0.49 |
| RP_POS_577_48273a11_141 | | 577.4827 | 11.14 | | RP | POS | | | 831807 | 790577 | 916237 | 735960 | 4.18E-01 | 0.11 | 0.52 |
| HILIC_POS_177_10194a1_597 | Serotonin | 177.1019 | 1.60 | HMDB0000259 | HILIC | POS | 160.0751(100), 115.0545(2) | 2 | 2541040 | 1267546 | 2406517 | 1164232 | 4.18E-01 | -0.11 | 0.64 |
| HILIC_NEG_234_8195a4_185 | | 234.8195 | 4.19 | | HILIC | NEG | | | 702314 | 341030 | 667149 | 295155 | 4.18E-01 | -0.11 | 0.5 |
| HILIC_NEG_255_82196a3_098 | | 255.8220 | 3.10 | | HILIC | NEG | | | 2516327 | 162338 | 2497674 | 173205 | 4.18E-01 | -0.11 | 0.73 |
| RP_NEG_340_86481a15_395 | | 340.8648 | 15.40 | | RP | NEG | | | 3007324 | 554172 | 2950713 | 470862 | 4.18E-01 | -0.11 | 0.96 |
| RP_POS_278_16a4_547 | | 278.1600 | 4.55 | | RP | POS | | | 132381 | 50741.7 | 127240 | 42371.1 | 4.18E-01 | -0.11 | 0.9 |
| RP_POS_513_3866a11_333 | | 513.3866 | 11.33 | | RP | POS | | | 195944 | 190151 | 178379 | 125828 | 4.18E-01 | -0.11 | 0.59 |
| HILIC_NEG_162_98267a5_112 | | 162.9827 | 5.11 | | HILIC | NEG | 118.9923(100), 162.9998(8) | | 1529008 | 919405 | 1627972 | 872541 | 4.19E-01 | 0.11 | 0.95 |
| HILIC_POS_316_21179a0_584 | | 316.2118 | 0.58 | | HILIC | POS | | | 382038 | 826075 | 483897 | 993411 | 4.19E-01 | 0.11 | 0.64 |
| RP_NEG_248_96114a10_425 | | 248.9611 | 10.43 | | RP | NEG | | | 566788 | 201814 | 591556 | 240727 | 4.19E-01 | 0.11 | 0.75 |
| RP_NEG_559_47369a10_961 | | 559.4737 | 10.96 | | RP | NEG | | | 118848 | 139764 | 133600 | 127955 | 4.19E-01 | 0.11 | 1.05 |
| RP_POS_205_00497a15_407 | | 205.0050 | 15.41 | | RP | POS | | | 1168107 | 1558452 | 1350877 | 1729646 | 4.19E-01 | 0.11 | 0.85 |
| RP_POS_716_20117a9_017 | | 716.2012 | 9.02 | | RP | POS | | | 882942 | 106400 | 895341 | 116822 | 4.19E-01 | 0.11 | 0.89 |
| RP_POS_792_58954a13_568 | | 792.5895 | 13.57 | | RP | POS | | | 2.1E+07 | 8250764 | 2.2E+07 | 8527974 | 4.19E-01 | 0.11 | 0.56 |
| RP_POS_253_09714a5_825 | Carbamazepine-10,11-epoxide | 253.0971 | 5.83 | | RP | POS | 180.0802(100), 236.0968(15) | 2 | 2.9E+07 | 1.5E+07 | 3.1E+07 | 1.5E+07 | 4.19E-01 | 0.11 | 1.12 |
| RP_NEG_174_987a2_4 | | 174.9870 | 2.40 | | RP | NEG | | | 304001 | 284924 | 271030 | 308865 | 4.19E-01 | -0.11 | 0.59 |
| RP_POS_141_95882a0_642 | | 141.9588 | 0.64 | | RP | POS | | | 6840431 | 5526277 | 6304036 | 4187631 | 4.19E-01 | -0.11 | 0.57 |
| RP_POS_265_15848a10_75 | | 265.1585 | 10.75 | | RP | POS | | | 564629 | 52734.8 | 558777 | 53060.6 | 4.19E-01 | -0.11 | 0.79 |
| HILIC_POS_371_23981a1_684 | | 371.2398 | 1.68 | | HILIC | POS | | | 2464344 | 2739451 | 2807130 | 3383583 | 4.20E-01 | 0.11 | 0.75 |
| RP_NEG_184_85762a15_405 | | 184.8576 | 15.41 | | RP | NEG | | | 441746 | 216240 | 466005 | 223175 | 4.20E-01 | 0.11 | 0.5 |
| RP_POS_240_15698a14_22 | | 240.1570 | 14.22 | | RP | POS | | | 1453569 | 806915 | 1545344 | 852934 | 4.20E-01 | 0.11 | 1 |
| RP_POS_263_99258a0_587 | | 263.9926 | 0.59 | | RP | POS | | | 2222807 | 446046 | 2275580 | 503306 | 4.20E-01 | 0.11 | 0.71 |
| RP_POS_432_76462a13_961 | | 432.7646 | 13.96 | | RP | POS | | | 1438893 | 1067735 | 1569973 | 1280785 | 4.20E-01 | 0.11 | 0.71 |
| RP_POS_724_49677a10_249 | | 724.4968 | 10.25 | | RP | POS | | | 416964 | 178398 | 437459 | 191933 | 4.20E-01 | 0.11 | 0.89 |
| RP_POS_755_60498a13_067 | | 755.6050 | 13.07 | | RP | POS | | | 638279 | 564505 | 694840 | 463368 | 4.20E-01 | 0.11 | 0.77 |
| HILIC_NEG_199_97838a0_734 | | 199.9784 | 0.73 | | HILIC | NEG | | | 175235 | 43960 | 170179 | 47270.2 | 4.20E-01 | -0.11 | 0.56 |
| RP_NEG_390_98102a9_194 | | 390.9810 | 9.19 | | RP | NEG | | | 998205 | 51434.5 | 992730 | 48080.1 | 4.20E-01 | -0.11 | 0.71 |
| RP_POS_187_13293a6_27 | | 187.1329 | 6.27 | | RP | POS | | | 561187 | 132774 | 544988 | 157487 | 4.20E-01 | -0.11 | 0.68 |
| RP_POS_332_31595a9_012 | | 332.3160 | 9.01 | | RP | POS | | | 1973583 | 5038405 | 1486056 | 3808788 | 4.20E-01 | -0.11 | 0.46 |
| RP_POS_361_11584a9_458 | | 361.1158 | 9.46 | | RP | POS | | | 768839 | 788540 | 685643 | 724706 | 4.20E-01 | -0.11 | 0.95 |
| RP_POS_631_11212a11_632 | | 631.1121 | 11.63 | | RP | POS | | | 143821 | 98605.6 | 134478 | 70680.2 | 4.20E-01 | -0.11 | 0.81 |
| HILIC_NEG_158_93909a5_119 | | 158.9391 | 5.12 | | HILIC | NEG | | | 1218171 | 554279 | 1276512 | 508133 | 4.21E-01 | 0.11 | 0.54 |
| RP_POS_466_66641a9_913 | | 466.6664 | 9.91 | | RP | POS | | | 250395 | 143128 | 269700 | 198681 | 4.21E-01 | 0.11 | 0.64 |
| RP_POS_612_42676a10_221 | | 612.4268 | 10.22 | | RP | POS | | | 1060458 | 4525134 | 1684437 | 6472326 | 4.21E-01 | 0.11 | 0.53 |
| RP_NEG_1025_63501a10_49 | | 1025.6350 | 10.49 | | RP | NEG | | | 1296667 | 139466 | 1280390 | 154268 | 4.21E-01 | -0.11 | 0.87 |
| RP_POS_221_11719a8_394 | | 221.1172 | 8.39 | | RP | POS | | | 784781 | 301320 | 750457 | 320621 | 4.21E-01 | -0.11 | 0.82 |
| RP_POS_259_1904a9_459 | | 259.1904 | 9.46 | | RP | POS | | | 1.2E+08 | 1.1E+08 | 1.1E+08 | 1E+08 | 4.21E-01 | -0.11 | 0.7 |
| RP_POS_592_52979a15_163 | | 592.5298 | 15.16 | | RP | POS | | | 868150 | 434079 | 821888 | 407250 | 4.21E-01 | -0.11 | 0.77 |
| RP_POS_632_42242a10_297 | | 632.4224 | 10.30 | | RP | POS | | | 171247 | 109443 | 159846 | 98453.2 | 4.21E-01 | -0.11 | 0.47 |
| HILIC_NEG_102_93199a3_115 | | 102.9320 | 3.12 | | HILIC | NEG | | | 499131 | 90592.3 | 508284 | 76607 | 4.22E-01 | 0.11 | 0.63 |
| HILIC_POS_339_03775a5_465 | | 339.0378 | 5.47 | | HILIC | POS | | | 452903 | 210822 | 476940 | 225420 | 4.22E-01 | 0.11 | 0.56 |
| HILIC_POS_406_32834a4_402 | | 406.3283 | 4.40 | | HILIC | POS | | | 43223.8 | 23128.8 | 46175.9 | 29661.6 | 4.22E-01 | 0.11 | 0.52 |
| HILIC_POS_554_39246a1_252 | | 554.3925 | 1.25 | | HILIC | POS | | | 164260 | 120545 | 178571 | 138070 | 4.22E-01 | 0.11 | 0.73 |
| RP_POS_185_97414a15_495 | | 185.9741 | 15.50 | | RP | POS | | | 1756353 | 163145 | 1773739 | 154059 | 4.22E-01 | 0.11 | 0.5 |
| RP_POS_614_42145a10_119 | | 614.4215 | 10.12 | | RP | POS | | | 1167206 | 636322 | 1244593 | 756161 | 4.22E-01 | 0.11 | 0.94 |
| RP_POS_295_2261a9_026 | 9-Hydroxyoctadecadienoic acid | 295.2261 | 9.03 | | RP | POS | 277.2176(100) | 2 | 1842375 | 255808 | 1874347 | 317205 | 4.22E-01 | 0.11 | 1.05 |
| RP_NEG_309_17429a10_568 | | 309.1743 | 10.57 | | RP | NEG | 309.1754(100), 96.96(35) | | 289296 | 143688 | 273378 | 146114 | 4.22E-01 | -0.11 | 0.93 |
| HILIC_POS_128_06841a4_375 | | 128.0684 | 4.38 | | HILIC | POS | | | 287586 | 248682 | 261612 | 225165 | 4.22E-01 | -0.11 | 0.79 |
| RP_POS_407_27625a11_143 | | 407.2763 | 11.14 | | RP | POS | | | 1160104 | 150751 | 1142594 | 165995 | 4.22E-01 | -0.11 | 0.96 |
| RP_POS_617_27869a7_165 | | 617.2787 | 7.17 | | RP | POS | | | 268415 | 95542.5 | 257730 | 98555.3 | 4.22E-01 | -0.11 | 1.11 |
| RP_POS_176_07404a2_552 | | 176.0740 | 2.55 | | RP | POS | | | 4624432 | 4349093 | 5330176 | 7766202 | 4.23E-01 | 0.12 | 0.89 |
| HILIC_NEG_112_98567a1_347 | | 112.9857 | 1.35 | | HILIC | NEG | 68.9949(100), 112.9856(2) | | 2.2E+07 | 5500279 | 2.3E+07 | 5440800 | 4.23E-01 | 0.11 | 0.62 |
| HILIC_NEG_170_82907a3_184 | | 170.8291 | 3.18 | | HILIC | NEG | | | 607707 | 134520 | 621946 | 125773 | 4.23E-01 | 0.11 | 0.68 |
| RP_POS_195_13789a7_693 | | 195.1379 | 7.69 | | RP | POS | | | 8103829 | 488003 | 8159138 | 517254 | 4.23E-01 | 0.11 | 0.66 |
| RP_POS_394_35217a10_695 | | 394.3522 | 10.70 | | RP | POS | | | 1E+07 | 1053885 | 1E+07 | 1111691 | 4.23E-01 | 0.11 | 0.77 |
| RP_POS_552_26459a11_239 | | 552.2646 | 11.24 | | RP | POS | | | 707048 | 394561 | 748019 | 355044 | 4.23E-01 | 0.11 | 0.87 |
| RP_POS_584_52448a14_445 | | 584.5245 | 14.45 | | RP | POS | | | 1079255 | 937798 | 1175776 | 828273 | 4.23E-01 | 0.11 | 0.61 |
| RP_POS_153_06592a1_659 | | 153.0659 | 1.66 | | RP | POS | 153.0655(100), 111.0241(2) | | 4325715 | 2906979 | 4023537 | 2627449 | 4.23E-01 | -0.11 | 0.49 |
| HILIC_NEG_128_92867a0_935 | | 128.9287 | 0.94 | | HILIC | NEG | | | 701207 | 78751.5 | 692634 | 77619.1 | 4.23E-01 | -0.11 | 0.81 |
| HILIC_POS_280_15433a1_276 | | 280.1543 | 1.28 | | HILIC | POS | | | 1648384 | 1268545 | 1519185 | 1096532 | 4.23E-01 | -0.11 | 0.93 |
| RP_NEG_462_55502a15_495 | | 462.5550 | 15.50 | | RP | NEG | | | 807928 | 63623.7 | 801277 | 58021.2 | 4.23E-01 | -0.11 | 0.78 |
| RP_NEG_812_54706a12_794 | | 812.5471 | 12.79 | | RP | NEG | | | 2387417 | 693677 | 2314074 | 648124 | 4.23E-01 | -0.11 | 0.69 |
| RP_POS_414_78549a9_639 | | 414.7855 | 9.64 | | RP | POS | | | 922694 | 590735 | 862293 | 514546 | 4.23E-01 | -0.11 | 0.88 |
| HILIC_POS_160_09691a0_754 | | 160.0969 | 0.75 | | HILIC | POS | 160.0979(100) | | 2000883 | 1844528 | 2246252 | 2531710 | 4.24E-01 | 0.11 | 0.65 |
| HILIC_NEG_191_94792a1_351 | | 191.9479 | 1.35 | | HILIC | NEG | | | 555252 | 139775 | 569755 | 126235 | 4.24E-01 | 0.11 | 0.71 |
| RP_POS_294_91986a15_553 | | 294.9199 | 15.55 | | RP | POS | | | 1.1E+07 | 3599611 | 1.1E+07 | 3198016 | 4.24E-01 | 0.11 | 1.02 |
| RP_POS_303_23169a10_209 | | 303.2317 | 10.21 | | RP | POS | | | 1724038 | 283508 | 1754812 | 279884 | 4.24E-01 | 0.11 | 0.64 |
| RP_POS_408_92844a15_385 | | 408.9284 | 15.39 | | RP | POS | | | 229032 | 104793 | 240262 | 100656 | 4.24E-01 | 0.11 | 0.56 |
| RP_POS_435_36755a10_898 | | 435.3676 | 10.90 | | RP | POS | | | 912238 | 242093 | 939109 | 248776 | 4.24E-01 | 0.11 | 0.59 |
| RP_POS_501_84918a10_239 | | 501.8492 | 10.24 | | RP | POS | | | 1821054 | 674382 | 1917743 | 1026344 | 4.24E-01 | 0.11 | 0.91 |
| HILIC_NEG_180_8596a3_163 | | 180.8596 | 3.16 | | HILIC | NEG | | | 1359918 | 142556 | 1341128 | 192851 | 4.24E-01 | -0.11 | 0.73 |
| HILIC_POS_151_09659a0_623 | | 151.0966 | 0.62 | | HILIC | POS | | | 4348811 | 836441 | 4452215 | 1032489 | 4.25E-01 | 0.11 | 0.63 |
| RP_NEG_201_88931a15_455 | | 201.8893 | 15.46 | | RP | NEG | | | 760838 | 513234 | 821274 | 584872 | 4.25E-01 | 0.11 | 0.87 |
| RP_POS_235_09618a7_213 | | 235.0962 | 7.21 | | RP | POS | | | 450341 | 92670.3 | 459992 | 84616.2 | 4.25E-01 | 0.11 | 0.59 |
| RP_POS_283_65826a10_294 | | 283.6583 | 10.29 | | RP | POS | | | 491015 | 171206 | 511113 | 194055 | 4.25E-01 | 0.11 | 0.97 |
| RP_POS_312_98605a0_676 | | 312.9861 | 0.68 | | RP | POS | | | 349559 | 68077.5 | 357002 | 68264.3 | 4.25E-01 | 0.11 | 0.6 |
| RP_POS_467_34763a9_225 | | 467.3476 | 9.23 | | RP | POS | | | 657381 | 908963 | 571594 | 664060 | 4.25E-01 | -0.11 | 0.7 |
| RP_POS_286_09671a0_678 | | 286.0967 | 0.68 | | RP | POS | | | 319673 | 152318 | 335423 | 138005 | 4.26E-01 | 0.11 | 0.73 |
| RP_POS_407_77875a9_386 | | 407.7788 | 9.39 | | RP | POS | | | 353608 | 220986 | 332638 | 164783 | 4.26E-01 | -0.11 | 0.72 |
| RP_POS_626_9184a9_94 | | 626.9184 | 9.94 | | RP | POS | | | 1634478 | 753616 | 1552454 | 753438 | 4.26E-01 | -0.11 | 0.52 |
| RP_POS_638_45782a10_286 | | 638.4578 | 10.29 | | RP | POS | | | 6125925 | 2682026 | 5828564 | 2773109 | 4.26E-01 | -0.11 | 0.71 |
| RP_POS_347_22159a7_555 | Corticosterone | 347.2216 | 7.56 | | RP | POS | 347.2192(100), 121.0655(40) | 1 | 1252618 | 1016101 | 1379254 | 1271757 | 4.26E-01 | 0.11 | 0.59 |
| RP_NEG_321_17453a10_702 | | 321.1745 | 10.70 | | RP | NEG | | | 1465561 | 5708884 | 2931578 | 1.8E+07 | 4.27E-01 | 0.13 | 1.12 |
| RP_POS_439_20319a10_892 | | 439.2032 | 10.89 | | RP | POS | | | 616787 | 1207053 | 895204 | 3291172 | 4.27E-01 | 0.12 | 0.5 |
| RP_POS_1013_65363a10_357 | | 1013.6536 | 10.36 | | RP | POS | | | 3981791 | 1752033 | 4185113 | 1962032 | 4.27E-01 | 0.11 | 0.97 |
| HILIC_POS_221_09161a1_592 | | 221.0916 | 1.59 | | HILIC | POS | | | 299134 | 147318 | 283686 | 137725 | 4.27E-01 | -0.11 | 0.64 |
| HILIC_POS_229_15488a4_793 | | 229.1549 | 4.79 | | HILIC | POS | | | 1658001 | 1074297 | 1550252 | 914226 | 4.27E-01 | -0.11 | 0.59 |
| HILIC_POS_374_30112a2_281 | | 374.3011 | 2.28 | | HILIC | POS | | | 756045 | 1426983 | 609738 | 1276386 | 4.27E-01 | -0.11 | 0.85 |
| RP_NEG_358_84036a15_403 | | 358.8404 | 15.40 | | RP | NEG | | | 295660 | 108148 | 283936 | 107633 | 4.27E-01 | -0.11 | 0.51 |
| RP_NEG_637_14276a9_515 | | 637.1428 | 9.52 | | RP | NEG | | | 3060857 | 226199 | 3036018 | 230137 | 4.27E-01 | -0.11 | 0.9 |
| RP_POS_161_09601a9_075 | | 161.0960 | 9.08 | | RP | POS | | | 2480052 | 411072 | 2435500 | 409394 | 4.27E-01 | -0.11 | 0.52 |

**Supplemantary table 1**: The non-targeted metabolomics results ordered by the VIP value (p value < 0.0003 are shaded dark green, p value < 0.05 are shaded light green, d values below 0 are blue and above 0 are red)

| | | | | | | | | | Cases | | Controls | | Welch test | Cohen's | |
|---|---|---|---|---|---|---|---|---|---|---|---|---|---|---|---|
| Feature_ID | Metabolite_name | Average_Mz | Average_RT_min_ | HMDB ID | RP_HILIC | POS_NEG | Cureted_MSMS | ID_level | Mean | SD | Mean2 | SD3 | p | d | VIP |
| RP_POS_194_99907a15_403 | | 194.9991 | 15.40 | | RP | POS | | | 599939 | 866970 | 527636 | 424067 | 4.27E-01 | -0.11 | 0.74 |
| RP_POS_318_88034a15_454 | | 318.8803 | 15.45 | | RP | POS | | | 581499 | 435810 | 534370 | 432607 | 4.27E-01 | -0.11 | 0.74 |
| RP_POS_336_20132a5_157 | | 336.2013 | 5.16 | | RP | POS | | | 280674 | 98228.4 | 269815 | 101169 | 4.27E-01 | -0.11 | 0.76 |
| RP_POS_489_19888a9_207 | | 489.1989 | 9.21 | | RP | POS | | | 234586 | 239892 | 210419 | 206665 | 4.27E-01 | -0.11 | 0.64 |
| RP_NEG_484_80164a15_383 | | 484.8016 | 15.38 | | RP | NEG | | | 975715 | 223799 | 999704 | 218912 | 4.28E-01 | 0.11 | 0.68 |
| RP_NEG_526_29437a10_428 | | 526.2944 | 10.43 | | RP | NEG | | | 1862018 | 715885 | 1943658 | 783202 | 4.28E-01 | 0.11 | 0.8 |
| RP_POS_432_88565a8_926 | | 432.8857 | 8.93 | | RP | POS | | | 843674 | 117529 | 855984 | 109823 | 4.28E-01 | 0.11 | 0.58 |
| RP_POS_519_70081a10_192 | | 519.7008 | 10.19 | | RP | POS | | | 1222690 | 556510 | 1293980 | 732003 | 4.28E-01 | 0.11 | 0.64 |
| RP_NEG_170_83328a15_467 | | 170.8333 | 15.47 | | RP | NEG | | | 1.1E+07 | 1102454 | 1.1E+07 | 1312839 | 4.28E-01 | -0.11 | 0.5 |
| RP_NEG_573_12952a9_243 | | 573.1295 | 9.24 | | RP | NEG | | | 6055560 | 351377 | 6017476 | 351086 | 4.28E-01 | -0.11 | 0.96 |
| RP_POS_163_11163a13_089 | | 163.1116 | 13.09 | | RP | POS | | | 6542872 | 3210677 | 6202597 | 3078280 | 4.28E-01 | -0.11 | 0.6 |
| RP_POS_210_11116a4_605 | | 210.1112 | 4.61 | | RP | POS | | | 200344 | 97785.3 | 191022 | 74396.4 | 4.28E-01 | -0.11 | 0.62 |
| RP_POS_498_85623a9_808 | | 498.8562 | 9.81 | | RP | POS | | | 2717047 | 1214714 | 2578825 | 1321263 | 4.28E-01 | -0.11 | 0.77 |
| RP_POS_368_42496a10_339 | | 368.4250 | 10.34 | | RP | POS | | | 447110 | 217447 | 484328 | 421730 | 4.29E-01 | 0.12 | 0.54 |
| HILIC_POS_166_07234a1_461 | | 166.0723 | 1.46 | | HILIC | POS | 166.0725(100) | | 1825892 | 254093 | 1851681 | 223757 | 4.29E-01 | 0.11 | 0.46 |
| HILIC_NEG_196_90129a3_1 | | 196.9013 | 3.10 | | HILIC | NEG | | | 257491 | 70323.6 | 264392 | 57518.9 | 4.29E-01 | 0.11 | 0.62 |
| RP_POS_683_68188a11_517 | | 683.6819 | 11.52 | | RP | POS | | | 146406 | 62978 | 156329 | 110198 | 4.29E-01 | 0.11 | 0.76 |
| RP_POS_273_10922a8_612 | | 273.1092 | 8.61 | | RP | POS | 273.1087(100) | | 1568385 | 205143 | 1546926 | 192095 | 4.29E-01 | -0.11 | 0.8 |
| RP_POS_515_41425a14_356 | | 515.4143 | 14.36 | | RP | POS | | | 9.6E+08 | 2.1E+08 | 9.4E+08 | 1.8E+08 | 4.29E-01 | -0.11 | 0.81 |
| RP_POS_546_36938a14_34 | | 546.3694 | 14.34 | | RP | POS | | | 1083345 | 804373 | 1005704 | 633731 | 4.29E-01 | -0.11 | 0.99 |
| RP_POS_726_54413a13_606 | | 726.5441 | 13.61 | | RP | POS | | | 2563053 | 2284979 | 2335476 | 1936610 | 4.29E-01 | -0.11 | 0.83 |
| RP_POS_961_80359a13_495 | | 961.8036 | 13.50 | | RP | POS | | | 618417 | 521212 | 569546 | 382088 | 4.29E-01 | -0.11 | 0.7 |
| RP_POS_222_16986a6_503 | | 222.1699 | 6.50 | | RP | POS | | | 470932 | 64934.5 | 477875 | 63633.2 | 4.30E-01 | 0.11 | 0.91 |
| RP_POS_233_98206a0_546 | | 233.9821 | 0.55 | | RP | POS | | | 1132999 | 119740 | 1146714 | 132614 | 4.30E-01 | 0.11 | 0.74 |
| RP_POS_297_08261a8_977 | | 297.0826 | 8.98 | | RP | POS | | | 451000 | 115980 | 463204 | 110663 | 4.30E-01 | 0.11 | 0.59 |
| RP_POS_446_98438a9_756 | | 446.9844 | 9.76 | | RP | POS | | | 124291 | 71912 | 132514 | 79523.2 | 4.30E-01 | 0.11 | 0.5 |
| RP_POS_475_41409a13_174 | | 475.4141 | 13.17 | | RP | POS | | | 2539620 | 1038568 | 2651468 | 1032070 | 4.30E-01 | 0.11 | 0.8 |
| RP_POS_583_07642a10_278 | | 583.0764 | 10.28 | | RP | POS | | | 719458 | 387457 | 764037 | 433616 | 4.30E-01 | 0.11 | 0.7 |
| RP_POS_524_37152a10_814 | LysoPC(0:0/18:0) | 524.3715 | 10.81 | | RP | POS | 184.0743(100), 86.0974(10) | 1 | 9.9E+07 | 2.5E+07 | 1E+08 | 2.8E+07 | 4.30E-01 | 0.11 | 0.8 |
| RP_POS_273_10947a7_658 | | 273.1095 | 7.66 | | RP | POS | 273.1092(100) | | 3662686 | 301679 | 3631243 | 281442 | 4.30E-01 | -0.11 | 0.81 |
| RP_POS_177_09103a6_837 | | 177.0910 | 6.84 | | RP | POS | | | 1152418 | 368967 | 1112665 | 366770 | 4.30E-01 | -0.11 | 0.63 |
| RP_NEG_369_1745a7_488 | | 369.1745 | 7.49 | | RP | NEG | | | 1.1E+07 | 8341005 | 1.2E+07 | 5971206 | 4.31E-01 | 0.11 | 0.58 |
| RP_NEG_540_00165a10_489 | | 540.0017 | 10.49 | | RP | NEG | | | 665494 | 336984 | 700169 | 308493 | 4.31E-01 | 0.11 | 0.85 |
| RP_POS_331_21136a10_253 | | 331.2114 | 10.25 | | RP | POS | | | 1089873 | 548973 | 1146249 | 500244 | 4.31E-01 | 0.11 | 1.06 |
| RP_POS_418_3161a10_253 | | 418.3161 | 10.25 | | RP | POS | | | 1031640 | 431236 | 1076224 | 399122 | 4.31E-01 | 0.11 | 1.04 |
| HILIC_NEG_179_84026a3_233 | | 179.8403 | 3.23 | | HILIC | NEG | | | 453381 | 83171.7 | 442466 | 114439 | 4.31E-01 | -0.11 | 0.79 |
| RP_NEG_701_08514a8_487 | | 701.0851 | 8.49 | | RP | NEG | | | 330634 | 27321.1 | 327725 | 26668.7 | 4.31E-01 | -0.11 | 0.94 |
| RP_POS_149_96101a0_619 | | 149.9610 | 0.62 | | RP | POS | | | 6103413 | 1417826 | 5949188 | 1442018 | 4.31E-01 | -0.11 | 0.64 |
| RP_POS_240_00778a5_42 | | 240.0078 | 5.42 | | RP | POS | | | 929037 | 600266 | 862740 | 627034 | 4.31E-01 | -0.11 | 0.65 |
| RP_POS_247_08502a5_729 | | 247.0850 | 5.73 | | RP | POS | | | 809400 | 294025 | 777777 | 293331 | 4.31E-01 | -0.11 | 0.69 |
| RP_POS_509_84711a9_81 | | 509.8471 | 9.81 | | RP | POS | | | 2633560 | 1689163 | 2464177 | 1468106 | 4.31E-01 | -0.11 | 0.81 |
| RP_POS_780_04474a12_722 | | 780.0447 | 12.72 | | RP | POS | | | 780926 | 642907 | 850146 | 644054 | 4.32E-01 | 0.11 | 0.81 |
| RP_NEG_637_14069a9_337 | | 637.1407 | 9.34 | | RP | NEG | | | 1.5E+07 | 656594 | 1.5E+07 | 646408 | 4.32E-01 | -0.11 | 0.81 |
| RP_POS_220_16719a11_143 | | 220.1672 | 11.14 | | RP | POS | | | 2565403 | 533095 | 2509970 | 500542 | 4.32E-01 | -0.11 | 0.94 |
| RP_POS_205_12244a6_772 | | 205.1224 | 6.77 | | RP | POS | | | 864242 | 6964192 | 353063 | 790743 | 4.32E-01 | -0.13 | 0.45 |
| HILIC_POS_159_14923a1_852 | | 159.1492 | 1.85 | | HILIC | POS | | | 343652 | 704549 | 451089 | 1195397 | 4.33E-01 | 0.11 | 0.51 |
| RP_POS_384_27261a12_956 | | 384.2726 | 12.96 | | RP | POS | | | 532619 | 314954 | 569170 | 360784 | 4.33E-01 | 0.11 | 0.88 |
| RP_POS_711_66052a11_427 | | 711.6605 | 11.43 | | RP | POS | | | 124216 | 60921.3 | 130924 | 63775.3 | 4.33E-01 | 0.11 | 0.62 |
| RP_NEG_522_4787a15_499 | | 522.4787 | 15.50 | | RP | NEG | | | 378746 | 28087.1 | 375561 | 30926.5 | 4.33E-01 | -0.11 | 0.53 |
| RP_POS_140_95253a15_409 | | 140.9525 | 15.41 | | RP | POS | | | 7.2E+07 | 1.2E+07 | 7E+07 | 1.2E+07 | 4.33E-01 | -0.11 | 1.09 |
| RP_POS_165_11272a10_25 | | 165.1127 | 10.25 | | RP | POS | | | 284877 | 213029 | 262419 | 206304 | 4.33E-01 | -0.11 | 0.61 |
| RP_NEG_207_83658a15_479 | | 207.8366 | 15.48 | | RP | NEG | | | 2203551 | 149522 | 2220571 | 166722 | 4.34E-01 | 0.11 | 0.81 |
| RP_POS_201_07567a10_254 | | 201.0757 | 10.25 | | RP | POS | | | 714790 | 375979 | 753369 | 346536 | 4.34E-01 | 0.11 | 1.12 |
| RP_POS_201_09103a10_513 | | 201.0910 | 10.51 | | RP | POS | | | 4221976 | 2124547 | 4447768 | 2100463 | 4.34E-01 | 0.11 | 1.13 |
| RP_POS_872_57379a10_93 | | 872.5738 | 10.93 | | RP | POS | | | 390163 | 350136 | 427092 | 340926 | 4.34E-01 | 0.11 | 0.8 |
| HILIC_NEG_608_32019a1_252 | | 608.3202 | 1.25 | | HILIC | NEG | | | 159738 | 32523 | 156062 | 35841.2 | 4.34E-01 | -0.11 | 0.68 |
| HILIC_POS_172_13322a1_017 | | 172.1332 | 1.02 | | HILIC | POS | | | 743146 | 363863 | 703194 | 380933 | 4.34E-01 | -0.11 | 0.64 |
| HILIC_POS_387_21429a1_332 | | 387.2143 | 1.33 | | HILIC | POS | | | 312630 | 126586 | 298118 | 142954 | 4.34E-01 | -0.11 | 0.52 |
| RP_NEG_519_40613a13_172 | | 519.4061 | 13.17 | | RP | NEG | | | 2548234 | 153150 | 2531998 | 150167 | 4.34E-01 | -0.11 | 1.09 |
| RP_POS_293_06839a5_759 | | 293.0684 | 5.76 | | RP | POS | | | 1647124 | 1447043 | 1523925 | 822697 | 4.34E-01 | -0.11 | 0.72 |
| HILIC_POS_118_0864a3_048 | | 118.0864 | 3.05 | | HILIC | POS | 118.0865(100) | | 8317743 | 2073111 | 8523771 | 1796302 | 4.35E-01 | 0.11 | 0.57 |
| HILIC_POS_59_06136a4_278 | | 59.0614 | 4.28 | | HILIC | POS | | | 9744326 | 4672635 | 1E+07 | 6513464 | 4.35E-01 | 0.11 | 0.65 |
| RP_POS_299_18524a10_253 | | 299.1852 | 10.25 | | RP | POS | | | 9094105 | 4552238 | 9558717 | 4169501 | 4.35E-01 | 0.11 | 1.07 |
| RP_POS_453_34427a5_197 | | 453.3443 | 5.20 | | RP | POS | | | 567774 | 222818 | 592451 | 237818 | 4.35E-01 | 0.11 | 0.94 |
| RP_POS_489_6915a10_657 | | 489.6915 | 10.66 | | RP | POS | | | 204564 | 109791 | 220360 | 173699 | 4.35E-01 | 0.11 | 0.71 |
| RP_POS_524_37042a10_929 | LysoPC(18:0/0:0 | 524.3704 | 10.93 | | RP | POS | 184.0741(100), 104.1077(70) | 1 | 8.4E+08 | 1.9E+08 | 8.6E+08 | 2.1E+08 | 4.35E-01 | 0.11 | 0.73 |
| RP_NEG_172_99139a2_213 | | 172.9914 | 2.21 | | RP | NEG | | | 4528318 | 4161599 | 4069687 | 4403158 | 4.35E-01 | -0.11 | 0.54 |
| RP_NEG_222_11349a7_091 | | 222.1135 | 7.09 | | RP | NEG | | | 181759 | 80776.3 | 172684 | 88188.3 | 4.35E-01 | -0.11 | 0.61 |
| RP_NEG_630_30353a10_016 | | 630.3035 | 10.02 | | RP | NEG | | | 630702 | 272102 | 600629 | 289234 | 4.35E-01 | -0.11 | 0.67 |
| RP_POS_1192_32495a13_104 | | 1192.3250 | 13.10 | | RP | POS | | | 554614 | 362997 | 518335 | 318231 | 4.35E-01 | -0.11 | 0.78 |
| RP_POS_482_32376a10_23 | | 482.3238 | 10.23 | | RP | POS | | | 4.7E+07 | 1.3E+07 | 4.6E+07 | 1.3E+07 | 4.35E-01 | -0.11 | 0.72 |
| RP_POS_553_45789a11_595 | | 553.4579 | 11.60 | | RP | POS | | | 4191112 | 828366 | 4100933 | 856681 | 4.35E-01 | -0.11 | 1.03 |
| RP_POS_647_45844a13_876 | | 647.4584 | 13.88 | | RP | POS | | | 1E+08 | 3.8E+08 | 6.7E+07 | 2.6E+08 | 4.35E-01 | -0.11 | 0.61 |
| RP_NEG_1043_61108a10_293 | | 1043.6111 | 10.29 | | RP | NEG | | | 854528 | 271362 | 884611 | 291031 | 4.36E-01 | 0.11 | 0.73 |
| RP_NEG_280_97409a3_535 | | 280.9741 | 3.54 | | RP | NEG | | | 248800 | 234454 | 277766 | 299916 | 4.36E-01 | 0.11 | 0.69 |
| RP_POS_473_39944a14_221 | | 473.3994 | 14.22 | | RP | POS | | | 318498 | 274264 | 349244 | 300215 | 4.36E-01 | 0.11 | 0.75 |
| RP_POS_522_37415a9_591 | | 522.3742 | 9.59 | | RP | POS | | | 3554178 | 1573100 | 3752395 | 2069658 | 4.36E-01 | 0.11 | 1.02 |
| RP_POS_180_06558a3_537 | Hippuric acid | 180.0656 | 3.54 | | RP | POS | 105.0339(100) | 1 | 7879732 | 7760186 | 8825498 | 9714816 | 4.36E-01 | 0.11 | 0.66 |
| HILIC_NEG_176_83664a3_087 | | 176.8366 | 3.09 | | HILIC | NEG | | | 316979 | 43000.6 | 312610 | 39201.4 | 4.36E-01 | -0.11 | 0.59 |
| HILIC_POS_349_25882a1_347 | | 349.2588 | 1.35 | | HILIC | POS | | | 459310 | 185360 | 440613 | 166885 | 4.36E-01 | -0.11 | 0.56 |
| RP_NEG_552_3324a10_573 | | 552.3324 | 10.57 | | RP | NEG | | | 808839 | 306255 | 779516 | 246147 | 4.36E-01 | -0.11 | 0.59 |
| RP_NEG_554_34747a10_536 | | 554.3475 | 10.54 | | RP | NEG | | | 967229 | 295602 | 937590 | 262712 | 4.36E-01 | -0.11 | 0.67 |
| RP_POS_325_17694a9_924 | | 325.1769 | 9.92 | | RP | POS | | | 748002 | 332141 | 713252 | 321362 | 4.36E-01 | -0.11 | 0.71 |
| RP_POS_414_33652a6_628 | Petiline | 414.3365 | 6.63 | | RP | POS | 414.3339(100) | 2 | 492756 | 521622 | 439131 | 486401 | 4.36E-01 | -0.11 | 0.57 |
| RP_NEG_164_8362a15_492 | | 164.8362 | 15.49 | | RP | NEG | | | 1878711 | 72896.9 | 1886224 | 68636.5 | 4.37E-01 | 0.11 | 0.63 |
| RP_NEG_408_97125a9_53 | | 408.9713 | 9.53 | | RP | NEG | | | 798558 | 115879 | 810997 | 118081 | 4.37E-01 | 0.11 | 0.47 |
| RP_NEG_608_31842a10_359 | | 608.3184 | 10.36 | | RP | NEG | | | 8250045 | 1030792 | 8365549 | 1132078 | 4.37E-01 | 0.11 | 0.99 |
| RP_POS_213_22131a11_409 | | 213.2213 | 11.41 | | RP | POS | | | 661829 | 330146 | 698671 | 360010 | 4.37E-01 | 0.11 | 0.71 |
| RP_POS_320_8692a15_438 | | 320.8692 | 15.44 | | RP | POS | | | 583813 | 518811 | 645652 | 628655 | 4.37E-01 | 0.11 | 0.77 |
| HILIC_NEG_341_004a0_936 | | 341.0040 | 0.94 | | HILIC | NEG | | | 814843 | 82535.4 | 806402 | 76648.4 | 4.37E-01 | -0.11 | 0.72 |
| HILIC_POS_130_15923a2_992 | | 130.1592 | 2.99 | | HILIC | POS | | | 3046388 | 834035 | 2965075 | 702119 | 4.37E-01 | -0.11 | 0.81 |
| RP_POS_174_99158a0_673 | | 174.9916 | 0.67 | | RP | POS | | | 430067 | 129676 | 416872 | 119318 | 4.37E-01 | -0.11 | 1 |
| RP_POS_313_14969a9_208 | | 313.1497 | 9.21 | | RP | POS | | | 1671859 | 1010112 | 1564900 | 1003345 | 4.37E-01 | -0.11 | 1 |
| RP_POS_324_00558a15_442 | | 324.0056 | 15.44 | | RP | POS | | | 1049644 | 643241 | 988070 | 519258 | 4.37E-01 | -0.11 | 0.71 |
| RP_POS_521_88733a10_218 | | 521.8873 | 10.22 | | RP | POS | | | 3294291 | 1703916 | 3119795 | 1588471 | 4.37E-01 | -0.11 | 0.57 |
| RP_POS_641_9234a9_748 | | 641.9234 | 9.75 | | RP | POS | | | 503851 | 285622 | 475652 | 246087 | 4.37E-01 | -0.11 | 0.69 |
| RP_POS_675_81732a4_708 | | 675.8173 | 4.71 | | RP | POS | | | 702787 | 863700 | 626814 | 556453 | 4.37E-01 | -0.11 | 1 |
| HILIC_POS_159_11302a3_458 | | 159.1130 | 3.46 | | HILIC | POS | | | 105104 | 457092 | 71681.8 | 69963.3 | 4.37E-01 | -0.13 | 0.55 |
| HILIC_POS_344_22803a0_61 | | 344.2280 | 0.61 | | HILIC | POS | | | 298020 | 292955 | 343383 | 513560 | 4.38E-01 | 0.11 | 0.49 |
| RP_NEG_159_10262a6_508 | | 159.1026 | 6.51 | | RP | NEG | | | 626635 | 217717 | 650357 | 228030 | 4.38E-01 | 0.11 | 0.98 |
| RP_NEG_349_86609a15_404 | | 349.8661 | 15.40 | | RP | NEG | | | 228076 | 97831.3 | 239529 | 115642 | 4.38E-01 | 0.11 | 0.73 |
| RP_POS_442_20688a8_083 | | 442.2069 | 8.08 | | RP | POS | | | 392042 | 97988.8 | 402518 | 99401.7 | 4.38E-01 | 0.11 | 0.83 |
| HILIC_NEG_323_01312a0_935 | | 323.0131 | 0.94 | | HILIC | NEG | | | 483824 | 51323.1 | 478364 | 51605.6 | 4.38E-01 | -0.11 | 0.78 |
| RP_POS_232_0265a0_687 | | 232.0265 | 0.69 | | RP | POS | | | 451723 | 102252 | 440921 | 101347 | 4.38E-01 | -0.11 | 0.63 |
| RP_POS_487_19852a9_207 | | 487.1985 | 9.21 | | RP | POS | | | 374382 | 594013 | 318827 | 456241 | 4.38E-01 | -0.11 | 0.83 |
| RP_POS_546_79614a10_31 | | 546.7961 | 10.31 | | RP | POS | | | 1564790 | 592555 | 1500485 | 617341 | 4.38E-01 | -0.11 | 0.84 |
| HILIC_POS_161_12859a0_738 | | 161.1286 | 0.74 | | HILIC | POS | | | 597047 | 271847 | 627839 | 306178 | 4.39E-01 | 0.11 | 0.54 |
| HILIC_POS_451_23004a1_062 | | 451.2300 | 1.06 | | HILIC | POS | | | 117447 | 53367.5 | 123001 | 51605.7 | 4.39E-01 | 0.11 | 0.71 |
| RP_NEG_208_78572a15_493 | | 208.7857 | 15.49 | | RP | NEG | | | 393465 | 27442.7 | 396265 | 25540.1 | 4.39E-01 | 0.11 | 0.66 |
| HILIC_POS_342_04364a3_147 | | 342.0436 | 3.15 | | HILIC | POS | | | 409077 | 44576.8 | 403272 | 62222.9 | 4.39E-01 | -0.11 | 0.49 |
| RP_NEG_202_91844a15_399 | | 202.9184 | 15.40 | | RP | NEG | | | 1923972 | 488534 | 1873210 | 471494 | 4.39E-01 | -0.11 | 0.68 |
| RP_POS_279_64648a10_309 | | 279.6465 | 10.31 | | RP | POS | | | 4342756 | 2713697 | 4081361 | 2245648 | 4.39E-01 | -0.11 | 1.18 |
| RP_POS_378_19894a10_419 | | 378.1989 | 10.42 | | RP | POS | | | 195812 | 262573 | 168871 | 247190 | 4.39E-01 | -0.11 | 0.45 |
| RP_POS_455_96228a15_421 | | 455.9623 | 15.42 | | RP | POS | | | 524459 | 348319 | 486056 | 373701 | 4.39E-01 | -0.11 | 0.77 |
| RP_POS_793_55792a13_361 | | 793.5579 | 13.36 | | RP | POS | | | 5814514 | 3383369 | 6157292 | 3125781 | 4.40E-01 | 0.11 | 0.81 |
| HILIC_POS_146_98047a7_43 | | 146.9805 | 7.43 | | HILIC | POS | | | 858553 | 91973 | 849290 | 83742.7 | 4.40E-01 | -0.11 | 0.63 |
| RP_NEG_235_92651a13_273 | | 235.9265 | 13.27 | | RP | NEG | | | 562757 | 279442 | 532997 | 284064 | 4.40E-01 | -0.11 | 0.63 |
| HILIC_NEG_253_79381a3_342 | | 253.7938 | 3.34 | | HILIC | NEG | | | 1871058 | 521931 | 1930296 | 594409 | 4.41E-01 | 0.11 | 0.54 |
| HILIC_NEG_98_93897a3_107 | | 98.9390 | 3.11 | | HILIC | NEG | | | 903499 | 103742 | 913262 | 82127.7 | 4.41E-01 | 0.11 | 0.48 |
| RP_POS_213_01031a15_407 | | 213.0103 | 15.41 | | RP | POS | | | 208094 | 102188 | 222812 | 164798 | 4.41E-01 | 0.11 | 0.9 |
| RP_POS_412_77753a15_37 | | 412.7775 | 15.37 | | RP | POS | | | 277159 | 177887 | 296071 | 180903 | 4.41E-01 | 0.11 | 0.83 |
| RP_POS_559_05756a10_012 | | 559.0576 | 10.01 | | RP | POS | | | 630426 | 369264 | 667718 | 339950 | 4.41E-01 | 0.11 | 0.71 |
| RP_POS_637_2467a11_262 | | 637.2467 | 11.26 | | RP | POS | | | 132693 | 101769 | 143920 | 110318 | 4.41E-01 | 0.11 | 0.78 |
| RP_POS_862_62457a12_292 | | 862.6246 | 12.29 | | RP | POS | | | 773124 | 309765 | 809243 | 368043 | 4.41E-01 | 0.11 | 0.8 |
| RP_NEG_246_98108a6_542 | | 246.9811 | 6.54 | | RP | NEG | | | 855893 | 295210 | 822996 | 325823 | 4.41E-01 | -0.11 | 0.49 |
| RP_NEG_333_13495a9_813 | | 333.1350 | 9.81 | | RP | NEG | | | 49079.5 | 128787 | 38351.1 | 71282.4 | 4.41E-01 | -0.11 | 0.61 |
| RP_NEG_456_21933a11_591 | | 456.2193 | 11.59 | | RP | NEG | | | 418012 | 72110 | 410749 | 66010.1 | 4.41E-01 | -0.11 | 1.11 |
| RP_POS_531_38757a10_654 | | 531.3876 | 10.65 | | RP | POS | | | 2254837 | 570964 | 2196413 | 538799 | 4.41E-01 | -0.11 | 1.15 |
| RP_POS_540_36511a9_501 | | 540.3651 | 9.50 | | RP | POS | | | 1882628 | 846973 | 1797718 | 767908 | 4.41E-01 | -0.11 | 0.58 |
| HILIC_NEG_174_9563a7_992 | | 174.9563 | 7.99 | | HILIC | NEG | | | 1E+07 | 4253293 | 9721703 | 3840726 | 4.42E-01 | -0.1 | 0.47 |
| RP_NEG_313_23886a10_709 | | 313.2389 | 10.71 | | RP | NEG | | | 354421 | 195611 | 337109 | 133029 | 4.42E-01 | -0.11 | 0.92 |
| HILIC_POS_239_14891a0_61 | | 239.1489 | 0.61 | | HILIC | POS | | | 708319 | 325342 | 744186 | 354333 | 4.43E-01 | 0.11 | 0.52 |
| RP_POS_787_54431a13_788 | | 787.5443 | 13.79 | | RP | POS | | | 1038907 | 778362 | 1131253 | 956289 | 4.43E-01 | 0.11 | 0.98 |
| RP_POS_801_55206a11_642 | | 801.5521 | 11.64 | | RP | POS | | | 5454073 | 1953915 | 5681471 | 2328405 | 4.43E-01 | 0.11 | 0.52 |
| RP_NEG_632_81915a15_352 | | 632.8192 | 15.35 | | RP | NEG | | | 360329 | 49857.1 | 365393 | 46899.6 | 4.43E-01 | 0.1 | 0.9 |
| RP_POS_177_00702a15_385 | | 177.0070 | 15.39 | | RP | POS | | | 4067962 | 1482063 | 3901893 | 1660163 | 4.43E-01 | -0.11 | 0.54 |
| HILIC_POS_204_20033a2_859 | | 204.2003 | 2.86 | | HILIC | POS | | | 424092 | 165783 | 442377 | 181598 | 4.44E-01 | 0.11 | 0.92 |
| RP_POS_804_55048a13_121 | | 804.5505 | 13.12 | | RP | POS | | | 2E+08 | 5.3E+07 | 2E+08 | 5.4E+07 | 4.44E-01 | 0.1 | 0.66 |
| RP_POS_237_14568a7_731 | | 237.1457 | 7.73 | | RP | POS | | | 1138202 | 186903 | 1118027 | 197021 | 4.44E-01 | -0.11 | 0.74 |
| RP_POS_689_81506a11_459 | | 689.8151 | 11.46 | | RP | POS | | | 188521 | 78790.3 | 179914 | 85029.4 | 4.44E-01 | -0.11 | 0.6 |
| RP_POS_456_99643a10_085 | | 456.9964 | 10.09 | | RP | POS | | | 95585.3 | 46727.2 | 101185 | 58658.1 | 4.45E-01 | 0.11 | 0.56 |
| RP_POS_699_32098a11_521 | | 699.3210 | 11.52 | | RP | POS | | | 116511 | 58835.9 | 123567 | 73930.7 | 4.45E-01 | 0.11 | 0.67 |
| RP_NEG_174_95625a15_527 | | 174.9563 | 15.53 | | RP | NEG | | | 7173705 | 378031 | 7214090 | 393753 | 4.45E-01 | 0.1 | 0.69 |
| HILIC_POS_150_11232a3_317 | | 150.1123 | 3.32 | | HILIC | POS | 150.1126(100), 132.1026(30) | | 2629580 | 2815144 | 2386373 | 1819376 | 4.45E-01 | -0.1 | 0.79 |
| RP_NEG_220_14696a9_993 | | 220.1470 | 9.99 | | RP | NEG | | | 352642 | 37016.3 | 348718 | 38046.5 | 4.45E-01 | -0.1 | 0.74 |
| RP_POS_436_21786a6_712 | | 436.2179 | 6.71 | | RP | POS | | | 578731 | 55057.4 | 573212 | 50740.3 | 4.45E-01 | -0.1 | 1 |
| RP_POS_507_32504a10_358 | | 507.3250 | 10.36 | | RP | POS | | | 830939 | 230544 | 857607 | 275594 | 4.46E-01 | 0.11 | 0.91 |
| RP_POS_579_29266a9_513 | | 579.2927 | 9.51 | | RP | POS | | | 809901 | 699631 | 893375 | 877469 | 4.46E-01 | 0.11 | 0.91 |
| RP_POS_739_31567a11_467 | | 739.3157 | 11.47 | | RP | POS | | | 161280 | 116004 | 174975 | 142780 | 4.46E-01 | 0.11 | 0.59 |
| RP_POS_798_06207a13_444 | | 798.0621 | 13.44 | | RP | POS | | | 678065 | 643670 | 754512 | 802218 | 4.46E-01 | 0.11 | 0.59 |
| RP_POS_271_16925a9_582 | | 271.1693 | 9.58 | | RP | POS | 271.1675(100) | | 6415177 | 371161 | 6454802 | 387121 | 4.46E-01 | 0.1 | 0.63 |
| RP_NEG_314_95267a0_917 | | 314.9527 | 0.92 | | RP | NEG | | | 214805 | 42290.9 | 219266 | 43159.6 | 4.46E-01 | 0.1 | 0.47 |
| RP_POS_413_26523a11_59 | | 413.2652 | 11.59 | | RP | POS | 413.2643(100) | | 6706980 | 1426268 | 6566944 | 1266523 | 4.46E-01 | -0.1 | 0.63 |
| HILIC_POS_315_07986a7_784 | Altenuene | 315.0799 | 7.78 | | HILIC | POS | 310.0793(100) | 2 | 301075 | 220378 | 327253 | 273914 | 4.46E-01 | 0.11 | 0.51 |
| HILIC_POS_235_11873a1_709 | | 235.1187 | 1.71 | | HILIC | POS | | | 195315 | 105313 | 219343 | 298177 | 4.47E-01 | 0.12 | 0.79 |
| RP_NEG_548_29724a10_901 | | 548.2972 | 10.90 | | RP | NEG | | | 886308 | 193715 | 907487 | 211696 | 4.47E-01 | 0.1 | 0.75 |
| RP_NEG_701_15436a9_416 | | 701.1544 | 9.42 | | RP | NEG | | | 1.4E+07 | 988582 | 1.4E+07 | 977524 | 4.47E-01 | -0.1 | 0.71 |
| RP_POS_132_90262a15_395 | | 132.9026 | 15.40 | | RP | POS | | | 528694 | 225784 | 505617 | 218084 | 4.47E-01 | -0.1 | 0.62 |
| RP_POS_180_02812a15_532 | | 180.0281 | 15.53 | | RP | POS | | | 1272058 | 359942 | 1233733 | 375865 | 4.47E-01 | -0.1 | 0.81 |
| RP_POS_340_25937a6_188 | | 340.2594 | 6.19 | | RP | POS | | | 1980726 | 2120257 | 1783804 | 1680726 | 4.47E-01 | -0.1 | 0.71 |
| RP_POS_443_3335a10_649 | | 443.3335 | 10.65 | | RP | POS | | | 8461790 | 1212645 | 8335296 | 1217038 | 4.47E-01 | -0.1 | 1.11 |
| HILIC_NEG_173_10461a6_955 | Arginine | 173.1046 | 6.96 | | HILIC | NEG | 131.0831(100), 173.1043(5) | 1 | 2425452 | 481437 | 2477074 | 508891 | 4.47E-01 | 0.10 | 0.57 |

**Supplemantary table 1**: The non-targeted metabolomics results ordered by the VIP value (p value < 0.0003 are shaded dark green, p value < 0.05 are shaded light green, d values below 0 are blue and above 0 are red)

| | | | | | | | | | Cases | | Controls | | Welch test | Cohen's | |
|---|---|---|---|---|---|---|---|---|---|---|---|---|---|---|---|
| Feature_ID | Metabolite_name | Average_Mz | Average_RT_min_ | HMDB ID | RP_HILIC | POS_NEG | Cureted_MSMS | ID_level | Mean | SD | Mean2 | SD3 | p | d | VIP |
| RP_POS_745_51648a11_38 | | 745.5165 | 11.38 | | RP | POS | | | 105011 | 55581.1 | 112570 | 84625.5 | 4.48E-01 | 0.11 | 0.61 |
| HILIC_NEG_164_07185a3_908 | | 164.0719 | 3.91 | | HILIC | NEG | | | 342603 | 84671.7 | 350890 | 75393.4 | 4.48E-01 | 0.1 | 0.55 |
| HILIC_POS_363_22031a3_289 | | 363.2203 | 3.29 | | HILIC | POS | | | 19773 | 7196.51 | 20570 | 8060.28 | 4.48E-01 | 0.1 | 0.76 |
| RP_POS_223_04861a15_38 | | 223.0486 | 15.38 | | RP | POS | | | 948479 | 438653 | 994614 | 449182 | 4.48E-01 | 0.1 | 0.62 |
| RP_POS_774_96216a10_484 | | 774.9622 | 10.48 | | RP | POS | | | 378297 | 224409 | 402164 | 234628 | 4.48E-01 | 0.1 | 0.5 |
| RP_POS_780_2121a8_957 | | 780.2121 | 8.96 | | RP | POS | | | 516547 | 49242.3 | 521617 | 48564 | 4.48E-01 | 0.1 | 0.8 |
| RP_NEG_432_58133a15_488 | | 432.5813 | 15.49 | | RP | NEG | | | 675013 | 40565.5 | 670921 | 38451.5 | 4.48E-01 | -0.1 | 0.59 |
| RP_NEG_443_15622a8_069 | | 443.1562 | 8.07 | | RP | NEG | | | 809712 | 372808 | 771886 | 357716 | 4.48E-01 | -0.1 | 0.81 |
| RP_POS_355_25217a11_593 | | 355.2522 | 11.59 | | RP | POS | | | 1057276 | 716126 | 987324 | 634508 | 4.48E-01 | -0.1 | 1.18 |
| RP_POS_679_45941a9_697 | | 679.4594 | 9.70 | | RP | POS | | | 472171 | 299864 | 439112 | 332709 | 4.48E-01 | -0.1 | 0.92 |
| RP_POS_646_35529a14_343 | | 646.3553 | 14.34 | | RP | POS | | | 1437156 | 1225876 | 1337743 | 647599 | 4.48E-01 | -0.11 | 0.54 |
| HILIC_POS_76_0765a1_469 | Trimethylamine N-oxide (TMAO) | 76.0765 | 1.47 | | HILIC | POS | 59.074(100), 76.0765(50) | 1 | 1.1E+08 | 1.1E+08 | 9.8E+07 | 8.7E+07 | 4.48E-01 | -0.10 | 0.5 |
| RP_POS_217_10414a2_587 | 1,2,3,4-tetrahydro-6-methoxy-1-oxo-beta-carboline | 217.1041 | 2.59 | | RP | POS | 217.1041(100) | 2 | 550078 | 190556 | 530394 | 189653 | 4.49E-01 | -0.10 | 1.13 |
| RP_POS_130_96523a0_634 | | 130.9652 | 0.63 | | RP | POS | | | 577808 | 268946 | 606376 | 281603 | 4.49E-01 | 0.1 | 0.8 |
| RP_POS_142_06514a2_881 | | 142.0651 | 2.88 | | RP | POS | | | 581954 | 121215 | 594207 | 115669 | 4.49E-01 | 0.1 | 0.88 |
| RP_POS_255_05873a12_914 | | 255.0587 | 12.91 | | RP | POS | | | 1096042 | 688929 | 1158948 | 528775 | 4.49E-01 | 0.1 | 0.87 |
| RP_POS_551_41016a14_818 | | 551.4102 | 14.82 | | RP | POS | | | 1064757 | 958238 | 1172385 | 1101013 | 4.49E-01 | 0.1 | 0.96 |
| RP_POS_748_52209a10_202 | | 748.5221 | 10.20 | | RP | POS | | | 1646572 | 815138 | 1734765 | 879315 | 4.49E-01 | 0.1 | 0.58 |
| RP_POS_310_3107a10_97 | | 310.3107 | 10.97 | | RP | POS | | | 115485 | 61957.2 | 109739 | 49278.9 | 4.49E-01 | -0.1 | 0.72 |
| RP_POS_363_04257a0_633 | | 363.0426 | 0.63 | | RP | POS | | | 439297 | 205985 | 419390 | 179872 | 4.49E-01 | -0.1 | 0.65 |
| RP_POS_551_26947a7_071 | | 551.2695 | 7.07 | | RP | POS | | | 2.8E+07 | 2690525 | 2.8E+07 | 2502875 | 4.49E-01 | -0.1 | 1.08 |
| RP_NEG_458_14578a11_592 | | 458.1458 | 11.59 | | RP | NEG | | | 1441846 | 87048.9 | 1431637 | 107321 | 4.49E-01 | -0.11 | 0.93 |
| HILIC_POS_131_1181a3_271 | | 131.1181 | 3.27 | | HILIC | POS | | | 7797073 | 1742262 | 7983217 | 1849881 | 4.50E-01 | 0.1 | 0.8 |
| RP_POS_363_1619a10_927 | | 363.1619 | 10.93 | | RP | POS | | | 471785 | 319952 | 500264 | 231822 | 4.50E-01 | 0.1 | 0.71 |
| RP_POS_122_09671a0_7 | N,N-Dimethylaniline | 122.0967 | 0.70 | | RP | POS | 122.0966(100), 95.0501(0.5) | 2 | 677158 | 119091 | 664402 | 126888 | 4.50E-01 | -0.10 | 0.95 |
| RP_POS_145_06477a6_127 | 1-Naphthol | 145.0648 | 6.13 | | RP | POS | 117.0705(100), 145.0645(35) | 2 | 1799198 | 281036 | 1828841 | 291543 | 4.50E-01 | 0.10 | 0.75 |
| RP_POS_302_26013a15_345 | | 302.2601 | 15.35 | | RP | POS | | | 1549507 | 2824022 | 1291053 | 2184711 | 4.50E-01 | -0.1 | 0.69 |
| RP_POS_130_96317a15_381 | | 130.9632 | 15.38 | | RP | POS | | | 1212437 | 701559 | 1282428 | 657751 | 4.51E-01 | 0.1 | 0.52 |
| RP_POS_740_52209a13_05 | PE(20:4/16:0) | 740.5221 | 13.05 | | RP | POS | 599.5003(100), 313.2726(70) | 1 | 9356564 | 3737882 | 9765325 | 4143386 | 4.51E-01 | 0.10 | 0.84 |
| RP_NEG_371_15302a11_142 | Arctigenin | 371.1530 | 11.14 | | RP | NEG | 371.151(100) | 2 | 1182358 | 76149.9 | 1174010 | 84836.2 | 4.51E-01 | -0.10 | 0.79 |
| HILIC_NEG_259_8157a3_097 | | 259.8157 | 3.10 | | HILIC | NEG | | | 810644 | 61301 | 803759 | 71019.8 | 4.51E-01 | -0.1 | 0.64 |
| HILIC_POS_387_21411a4_994 | | 387.2141 | 4.99 | | HILIC | POS | | | 1006765 | 456491 | 963430 | 386042 | 4.51E-01 | -0.1 | 1.03 |
| RP_POS_374_95639a15_455 | | 374.9564 | 15.46 | | RP | POS | | | 621945 | 248119 | 597112 | 234603 | 4.51E-01 | -0.1 | 1.19 |
| HILIC_NEG_113_92633a3_144 | | 113.9263 | 3.14 | | HILIC | NEG | | | 1117908 | 120793 | 1102662 | 167154 | 4.51E-01 | -0.11 | 0.47 |
| RP_POS_407_25513a9_456 | | 407.2551 | 9.46 | | RP | POS | | | 105916 | 115728 | 122344 | 188874 | 4.52E-01 | 0.11 | 0.75 |
| HILIC_NEG_160_97604a7_406 | | 160.9760 | 7.41 | | HILIC | NEG | | | 628100 | 106867 | 639790 | 118824 | 4.52E-01 | 0.1 | 0.65 |
| RP_NEG_518_83301a15_364 | | 518.8330 | 15.36 | | RP | NEG | | | 493811 | 195789 | 515384 | 220745 | 4.52E-01 | 0.1 | 0.48 |
| RP_NEG_569_13281a8_968 | | 569.1328 | 8.97 | | RP | NEG | | | 436942 | 28602.5 | 439672 | 24672.7 | 4.52E-01 | 0.1 | 0.48 |
| RP_POS_193_05598a15_334 | | 193.0560 | 15.33 | | RP | POS | | | 269529 | 113705 | 280426 | 98711.2 | 4.52E-01 | 0.1 | 0.43 |
| RP_POS_450_31796a10_427 | | 450.3180 | 10.43 | | RP | POS | | | 2745651 | 100793 | 2756306 | 105751 | 4.52E-01 | 0.1 | 1 |
| RP_POS_622_23969a11_386 | | 622.2397 | 11.39 | | RP | POS | | | 129399 | 103700 | 139852 | 99918.6 | 4.52E-01 | 0.1 | 0.6 |
| RP_POS_822_48096a11_64 | | 822.4810 | 11.64 | | RP | POS | | | 2599369 | 1130219 | 2727011 | 1328356 | 4.52E-01 | 0.1 | 0.76 |
| RP_POS_890_52325a12_994 | | 890.5233 | 12.99 | | RP | POS | | | 712620 | 290275 | 745477 | 341499 | 4.52E-01 | 0.1 | 0.76 |
| RP_POS_228_85524a15_396 | | 228.8552 | 15.40 | | RP | POS | | | 301288 | 157409 | 286672 | 127701 | 4.52E-01 | -0.1 | 0.74 |
| RP_POS_287_64304a9_207 | | 287.6430 | 9.21 | | RP | POS | | | 438942 | 425718 | 395960 | 410638 | 4.52E-01 | -0.1 | 0.65 |
| RP_POS_385_16702a10_896 | | 385.1670 | 10.90 | | RP | POS | | | 1263127 | 701162 | 1194653 | 634518 | 4.52E-01 | -0.1 | 0.89 |
| RP_POS_497_31961a10_042 | | 497.3196 | 10.04 | | RP | POS | | | 398784 | 231711 | 375816 | 216213 | 4.52E-01 | -0.1 | 0.8 |
| RP_POS_432_28015a4_466 | | 432.2802 | 4.47 | | RP | POS | | | 464948 | 160178 | 487653 | 261812 | 4.53E-01 | 0.11 | 0.64 |
| HILIC_POS_227_07866a4_138 | | 227.0787 | 4.14 | | HILIC | POS | | | 726052 | 364318 | 765265 | 396310 | 4.53E-01 | 0.1 | 0.72 |
| RP_NEG_376_2265a9_601 | | 376.2265 | 9.60 | | RP | NEG | | | 960545 | 275647 | 986228 | 225857 | 4.53E-01 | 0.1 | 0.64 |
| RP_NEG_376_98526a8_222 | 2-Perfluorohexyl ethanoic acid | 376.9853 | 8.22 | | RP | NEG | 292.985(100), 242.986(40), 376.9844(25) | 2 | 2009376 | 444760 | 1963974 | 440115 | 4.53E-01 | -0.10 | 0.71 |
| RP_POS_255_05862a13_604 | | 255.0586 | 13.60 | | RP | POS | 255.0606(100), 137.023(20) | | 1047590 | 592066 | 985462 | 614841 | 4.53E-01 | -0.1 | 0.52 |
| RP_POS_151_14818a7_528 | | 151.1482 | 7.53 | | RP | POS | | | 389230 | 92574.7 | 379920 | 89214.5 | 4.53E-01 | -0.1 | 0.82 |
| RP_POS_642_61365a14_07 | | 642.6137 | 14.07 | | RP | POS | | | 395424 | 271484 | 367586 | 270312 | 4.53E-01 | -0.1 | 0.81 |
| HILIC_NEG_157_90086a5_932 | | 157.9009 | 5.93 | | HILIC | NEG | | | 1719906 | 544852 | 1783162 | 672205 | 4.54E-01 | 0.1 | 0.64 |
| RP_NEG_247_00102a5_961 | | 247.0010 | 5.96 | | RP | NEG | | | 1484947 | 723152 | 1563053 | 795191 | 4.54E-01 | 0.1 | 0.49 |
| RP_NEG_297_24374a10_636 | | 297.2437 | 10.64 | | RP | NEG | | | 3279488 | 660614 | 3344481 | 610889 | 4.54E-01 | 0.1 | 0.76 |
| RP_POS_203_05923a15_379 | | 203.0592 | 15.38 | | RP | POS | | | 1.1E+07 | 3202653 | 1.1E+07 | 2721617 | 4.54E-01 | 0.1 | 0.74 |
| RP_NEG_568_36121a10_931 | LysoPC 18:0 (18:0/0:0) | 568.3612 | 10.93 | | RP | NEG | 283.2642(100), 224.0691(10) | 2 | 1.6E+08 | 3.1E+07 | 1.6E+08 | 3.4E+07 | 4.54E-01 | 0.10 | 0.79 |
| RP_POS_203_12778a9_459 | | 203.1278 | 9.46 | | RP | POS | | | 2436174 | 2106026 | 2220351 | 2110764 | 4.54E-01 | -0.1 | 0.71 |
| RP_POS_494_34314a8_403 | | 494.3431 | 8.40 | | RP | POS | | | 293213 | 217819 | 272223 | 193235 | 4.54E-01 | -0.1 | 0.72 |
| RP_POS_451_33923a11_591 | | 451.3392 | 11.59 | | RP | POS | | | 422162 | 442908 | 482190 | 686113 | 4.55E-01 | 0.11 | 0.63 |
| HILIC_POS_388_31601a0_883 | | 388.3160 | 0.88 | | HILIC | POS | | | 103675 | 34333.6 | 99879.6 | 39288.1 | 4.55E-01 | -0.1 | 0.61 |
| RP_NEG_520_26581a10_469 | | 520.2658 | 10.47 | | RP | NEG | | | 444451 | 92502.7 | 434642 | 98675 | 4.55E-01 | -0.1 | 0.59 |
| RP_NEG_524_27832a10_18 | | 524.2783 | 10.18 | | RP | NEG | | | 2817905 | 809691 | 2733537 | 839992 | 4.55E-01 | -0.1 | 0.66 |
| RP_POS_504_30643a10_23 | | 504.3064 | 10.23 | | RP | POS | | | 5381048 | 1655528 | 5217837 | 1547433 | 4.55E-01 | -0.1 | 0.71 |
| RP_POS_368_21951a10_568 | | 368.2195 | 10.57 | | RP | POS | | | 207993 | 233757 | 264938 | 726695 | 4.56E-01 | 0.12 | 0.58 |
| RP_NEG_1035_66492a10_358 | | 1035.6649 | 10.36 | | RP | NEG | | | 4266304 | 1340933 | 4412969 | 1515874 | 4.56E-01 | 0.1 | 0.99 |
| RP_NEG_177_95686a15_55 | | 177.9569 | 15.55 | | RP | NEG | | | 2862631 | 582138 | 2914909 | 444956 | 4.56E-01 | 0.1 | 0.49 |
| RP_POS_195_12254a10_658 | | 195.1225 | 10.66 | | RP | POS | | | 1157314 | 311939 | 1189962 | 326476 | 4.56E-01 | 0.1 | 0.53 |
| RP_POS_548_72699a10_587 | | 548.7270 | 10.59 | | RP | POS | | | 324022 | 191001 | 345677 | 229081 | 4.56E-01 | 0.1 | 0.84 |
| RP_POS_818_56708a13_451 | | 818.5671 | 13.45 | | RP | POS | | | 3053453 | 1879385 | 3261292 | 2161591 | 4.56E-01 | 0.1 | 0.96 |
| HILIC_POS_496_34061a1_252 | LysoPC 16:0 | 496.3406 | 1.25 | | HILIC | POS | 184.0744(100), 104.1078(80) | 2 | 3.8E+08 | 7E+07 | 3.9E+08 | 7.5E+07 | 4.56E-01 | 0.10 | 0.89 |
| RP_NEG_562_3158a10_016 | | 562.3158 | 10.02 | | RP | NEG | | | 8408561 | 3536327 | 8041380 | 3651061 | 4.56E-01 | -0.1 | 0.66 |
| RP_NEG_679_15155a8_64 | | 679.1516 | 8.64 | | RP | NEG | | | 1125696 | 63603 | 1119584 | 56645.7 | 4.56E-01 | -0.1 | 0.76 |
| RP_POS_210_10983a10_25 | | 210.1098 | 10.25 | | RP | POS | | | 1372134 | 269570 | 1343915 | 282527 | 4.56E-01 | -0.1 | 0.64 |
| RP_POS_343_19418a11_593 | | 343.1942 | 11.59 | | RP | POS | | | 945803 | 225389 | 923817 | 206861 | 4.56E-01 | -0.1 | 0.81 |
| RP_POS_361_16681a13_132 | | 361.1668 | 13.13 | | RP | POS | | | 264771 | 286168 | 239011 | 220485 | 4.56E-01 | -0.1 | 0.51 |
| RP_POS_559_89032a10_54 | | 559.8903 | 10.54 | | RP | POS | | | 601785 | 429789 | 559428 | 402798 | 4.56E-01 | -0.1 | 0.85 |
| RP_NEG_309_17477a10_093 | | 309.1748 | 10.09 | | RP | NEG | 309.1752(100), 96.96(40) | | 210060 | 140731 | 233080 | 278230 | 4.57E-01 | 0.11 | 0.59 |
| HILIC_POS_375_32156a4_91 | | 375.3216 | 4.91 | | HILIC | POS | | | 207864 | 85714.2 | 219103 | 127586 | 4.57E-01 | 0.11 | 0.89 |
| RP_POS_499_27396a11_852 | | 499.2740 | 11.85 | | RP | POS | | | 1135079 | 229180 | 1157968 | 221006 | 4.57E-01 | 0.1 | 1.21 |
| RP_POS_544_71637a10_021 | | 544.7164 | 10.02 | | RP | POS | | | 618522 | 338631 | 654411 | 364068 | 4.57E-01 | 0.1 | 0.78 |
| HILIC_POS_233_18571a3_289 | | 233.1857 | 3.29 | | HILIC | POS | | | 38952.3 | 23771.6 | 36698.3 | 20652.7 | 4.57E-01 | -0.1 | 0.53 |
| RP_NEG_162_83894a11_595 | | 162.8389 | 11.60 | | RP | NEG | | | 144798 | 45355.8 | 140168 | 45596.9 | 4.57E-01 | -0.1 | 0.65 |
| RP_POS_149_99765a15_531 | | 149.9977 | 15.53 | | RP | POS | | | 2.7E+07 | 1380758 | 2.7E+07 | 1233054 | 4.57E-01 | -0.1 | 1 |
| RP_POS_627_26123a7_604 | | 627.2612 | 7.60 | | RP | POS | | | 393394 | 121427 | 381695 | 109297 | 4.57E-01 | -0.1 | 1.16 |
| HILIC_POS_133_061a6_247 | Asparagine | 133.0610 | 6.25 | | HILIC | POS | 74.0243(100), 87.056(60) | 1 | 4744101 | 1114760 | 4637388 | 987972 | 4.57E-01 | -0.10 | 0.59 |
| HILIC_NEG_257_819a3_216 | | 257.8190 | 3.22 | | HILIC | NEG | | | 2849667 | 284063 | 2881244 | 332068 | 4.58E-01 | 0.1 | 0.61 |
| RP_POS_1063_67175a10_303 | | 1063.6718 | 10.30 | | RP | POS | | | 1.3E+07 | 6989851 | 1.4E+07 | 7328440 | 4.58E-01 | 0.1 | 0.68 |
| RP_POS_485_67944a10_065 | | 485.6794 | 10.07 | | RP | POS | | | 902801 | 474602 | 950320 | 462168 | 4.58E-01 | 0.1 | 0.61 |
| RP_POS_545_70642a10_444 | | 545.7064 | 10.44 | | RP | POS | | | 538774 | 222818 | 562556 | 243279 | 4.58E-01 | 0.1 | 0.86 |
| RP_POS_594_91656a11_238 | | 594.9166 | 11.24 | | RP | POS | | | 139010 | 73522.7 | 147341 | 88625.6 | 4.58E-01 | 0.1 | 0.57 |
| RP_POS_636_44409a10_481 | | 636.4441 | 10.48 | | RP | POS | | | 241362 | 139503 | 256652 | 159189 | 4.58E-01 | 0.1 | 0.64 |
| RP_POS_651_11993a11_244 | | 651.1199 | 11.24 | | RP | POS | | | 162239 | 77257.4 | 170626 | 86735.1 | 4.58E-01 | 0.1 | 0.62 |
| RP_NEG_235_13461a8_57 | | 235.1346 | 8.57 | | RP | NEG | | | 623134 | 29871.9 | 626107 | 28905.2 | 4.59E-01 | 0.1 | 0.55 |
| RP_NEG_445_06506a0_685 | | 445.0651 | 0.69 | | RP | NEG | | | 259864 | 38098.5 | 263780 | 39106.1 | 4.59E-01 | 0.1 | 0.52 |
| RP_POS_495_68179a9_892 | | 495.6818 | 9.89 | | RP | POS | | | 604568 | 361126 | 642186 | 379829 | 4.59E-01 | 0.1 | 0.79 |
| RP_POS_546_35425a11_584 | | 546.3543 | 11.58 | | RP | POS | | | 1316878 | 343084 | 1353517 | 376945 | 4.59E-01 | 0.1 | 0.69 |
| RP_NEG_159_10274a7_023 | | 159.1027 | 7.02 | | RP | NEG | | | 689578 | 471605 | 647151 | 367486 | 4.59E-01 | -0.1 | 0.71 |
| RP_POS_329_17651a11_14 | | 329.1765 | 11.14 | | RP | POS | | | 518284 | 114603 | 502706 | 180454 | 4.59E-01 | -0.11 | 0.62 |
| HILIC_NEG_352_01123a4_11 | | 352.0112 | 4.11 | | HILIC | NEG | | | 186553 | 36088.1 | 190387 | 39416.2 | 4.60E-01 | 0.1 | 0.62 |
| RP_NEG_231_93185a13_187 | | 231.9319 | 13.19 | | RP | NEG | | | 1377897 | 595304 | 1444243 | 704921 | 4.60E-01 | 0.1 | 0.56 |
| RP_NEG_343_22821a9_925 | | 343.2282 | 9.93 | | RP | NEG | | | 197373 | 265651 | 230948 | 379503 | 4.60E-01 | 0.1 | 0.77 |
| RP_POS_399_78085a9_909 | | 399.7809 | 9.91 | | RP | POS | | | 363894 | 223028 | 391170 | 303302 | 4.60E-01 | 0.1 | 0.73 |
| RP_POS_621_4292a10_284 | | 621.4292 | 10.28 | | RP | POS | | | 1046590 | 543841 | 992089 | 535257 | 4.60E-01 | -0.1 | 0.53 |
| RP_POS_149_02362a9_523 | | 149.0236 | 9.52 | | RP | POS | 149.0231(100), 121.0287(20) | | 4E+07 | 7905675 | 4.1E+07 | 1.1E+07 | 4.61E-01 | 0.1 | 0.86 |
| HILIC_POS_219_08255a6_965 | | 219.0826 | 6.97 | | HILIC | POS | | | 242770 | 27715.5 | 245593 | 28213.7 | 4.61E-01 | 0.1 | 0.48 |
| RP_NEG_381_3381a10_785 | | 381.3381 | 10.79 | | RP | NEG | | | 157790 | 55215.4 | 163407 | 56164.7 | 4.61E-01 | 0.1 | 0.89 |
| RP_POS_237_18436a9_176 | | 237.1844 | 9.18 | | RP | POS | | | 1400615 | 88088.1 | 1409434 | 86755 | 4.61E-01 | 0.1 | 0.66 |
| RP_POS_371_21912a10_515 | | 371.2191 | 10.52 | | RP | POS | | | 6148816 | 2863352 | 6434034 | 2797824 | 4.61E-01 | 0.1 | 1.03 |
| RP_POS_438_29779a10_676 | | 438.2978 | 10.68 | | RP | POS | | | 5070265 | 1819314 | 5270144 | 2105706 | 4.61E-01 | 0.1 | 0.79 |
| RP_POS_641_39075a15_162 | | 641.3908 | 15.16 | | RP | POS | | | 1086948 | 831420 | 1175903 | 923372 | 4.61E-01 | 0.1 | 1.15 |
| RP_POS_725_55536a12_805 | | 725.5554 | 12.81 | | RP | POS | | | 5.5E+07 | 1.3E+07 | 5.7E+07 | 1.1E+07 | 4.61E-01 | 0.1 | 1.07 |
| RP_POS_808_03485a10_929 | | 808.0349 | 10.93 | | RP | POS | | | 3912953 | 1555144 | 4080094 | 1737460 | 4.61E-01 | 0.1 | 0.71 |
| RP_NEG_240_04913a0_683 | | 240.0491 | 0.68 | | RP | NEG | | | 267097 | 57640.8 | 261191 | 59373.1 | 4.61E-01 | -0.1 | 0.46 |
| RP_NEG_289_06842a0_678 | | 289.0684 | 0.68 | | RP | NEG | | | 213936 | 54981.9 | 209015 | 42986.2 | 4.61E-01 | -0.1 | 0.64 |
| RP_NEG_595_16998a11_152 | | 595.1700 | 11.15 | | RP | NEG | | | 412034 | 121296 | 399410 | 128303 | 4.61E-01 | -0.1 | 1.07 |
| RP_POS_195_13824a6_022 | | 195.1382 | 6.02 | | RP | POS | | | 966394 | 332796 | 932823 | 333316 | 4.61E-01 | -0.1 | 0.77 |
| RP_POS_231_11581a10_554 | | 231.1158 | 10.55 | | RP | POS | | | 5031464 | 1903729 | 4844072 | 1821300 | 4.61E-01 | -0.1 | 0.61 |
| RP_POS_225_10931a6_714 | | 225.1093 | 6.71 | | RP | POS | 225.1305(100) | | 680180 | 717377 | 752512 | 720269 | 4.62E-01 | 0.1 | 0.68 |
| RP_NEG_291_00781a15_474 | | 291.0078 | 15.47 | | RP | NEG | | | 1010312 | 151947 | 1025779 | 154943 | 4.62E-01 | 0.1 | 0.6 |
| RP_POS_236_11282a4_825 | | 236.1128 | 4.83 | | RP | POS | | | 9.6E+07 | 4.8E+07 | 1E+08 | 7.2E+07 | 4.62E-01 | 0.1 | 0.78 |
| RP_POS_411_76901a13_395 | | 411.7690 | 13.40 | | RP | POS | | | 2374360 | 2495841 | 2649110 | 2908760 | 4.62E-01 | 0.1 | 0.98 |
| RP_POS_417_22815a7_534 | | 417.2282 | 7.53 | | RP | POS | | | 7762942 | 816514 | 7845208 | 816724 | 4.62E-01 | 0.1 | 1.03 |
| RP_POS_599_36346a14_346 | | 599.3635 | 14.35 | | RP | POS | | | 2943041 | 1330142 | 3084753 | 1470066 | 4.62E-01 | 0.1 | 0.59 |
| RP_POS_621_60376a11_305 | | 621.6038 | 11.31 | | RP | POS | | | 289813 | 131017 | 304577 | 158713 | 4.62E-01 | 0.1 | 0.78 |
| RP_NEG_480_31018a10_901 | LysoPE 18:0 | 480.3102 | 10.90 | | RP | NEG | 283.2648(100), 480.3092(5) | 1 | 1E+07 | 2259455 | 1E+07 | 2506801 | 4.62E-01 | 0.10 | 0.82 |
| RP_POS_381_29758a11_244 | 2,3-dihydroxypropyl stearate | 381.2976 | 11.24 | | RP | POS | 381.2991(100) | 2 | 1.1E+07 | 4994522 | 1.2E+07 | 4645570 | 4.62E-01 | 0.10 | 0.73 |
| HILIC_POS_366_28439a0_847 | | 366.2844 | 0.85 | | HILIC | POS | | | 839238 | 300264 | 808389 | 311855 | 4.62E-01 | -0.1 | 0.61 |
| RP_POS_185_12828a2_758 | | 185.1283 | 2.76 | | RP | POS | | | 1751937 | 618006 | 1690780 | 598490 | 4.62E-01 | -0.1 | 0.51 |
| RP_POS_429_31775a10_529 | | 429.3178 | 10.53 | | RP | POS | | | 2251229 | 595393 | 2195254 | 520444 | 4.62E-01 | -0.1 | 0.99 |
| RP_POS_743_00726a10_51 | | 743.0073 | 10.51 | | RP | POS | | | 294871 | 183945 | 273988 | 225311 | 4.62E-01 | -0.1 | 0.6 |
| HILIC_POS_128_10707a0_618 | | 128.1071 | 0.62 | | HILIC | POS | | | 841913 | 129476 | 854963 | 130415 | 4.63E-01 | 0.1 | 0.53 |
| RP_NEG_163_8403a0_705 | | 163.8403 | 0.71 | | RP | NEG | | | 1690321 | 717858 | 1762999 | 728428 | 4.63E-01 | 0.1 | 0.62 |
| HILIC_POS_388_31616a4_917 | | 388.3162 | 4.92 | | HILIC | POS | | | 65237.4 | 23977.1 | 63040.6 | 19962.1 | 4.63E-01 | -0.1 | 0.63 |
| RP_POS_218_13821a4_892 | | 218.1382 | 4.89 | | RP | POS | | | 178777 | 92442.6 | 169473 | 92887 | 4.63E-01 | -0.1 | 0.8 |
| RP_POS_452_24792a6_733 | | 452.2479 | 6.73 | | RP | POS | | | 627383 | 50244.8 | 622619 | 44893.2 | 4.63E-01 | -0.1 | 0.73 |
| RP_NEG_528_26428a8_518 | | 528.2643 | 8.52 | | RP | NEG | | | 4080018 | 6274951 | 5391112 | 1.7E+07 | 4.64E-01 | 0.11 | 0.62 |
| RP_POS_383_23004a13_685 | | 383.2300 | 13.69 | | RP | POS | | | 228180 | 163779 | 251119 | 271752 | 4.64E-01 | 0.11 | 0.81 |
| RP_NEG_387_2869a8_955 | | 387.2869 | 8.96 | | RP | NEG | | | 296979 | 45531.1 | 301273 | 40523.4 | 4.64E-01 | 0.1 | 0.65 |
| RP_NEG_744_29462a10_361 | | 744.2946 | 10.36 | | RP | NEG | | | 706541 | 67716.9 | 713244 | 66283.5 | 4.64E-01 | 0.1 | 0.91 |
| RP_POS_143_1068a5_502 | | 143.1068 | 5.50 | | RP | POS | | | 416123 | 516858 | 469013 | 537985 | 4.64E-01 | 0.1 | 0.7 |
| RP_POS_345_15836a10_96 | | 345.1584 | 10.96 | | RP | POS | | | 263256 | 138322 | 277454 | 144638 | 4.64E-01 | 0.1 | 0.85 |
| RP_POS_482_40399a10_651 | | 482.4040 | 10.65 | | RP | POS | | | 1.5E+07 | 1172252 | 1.5E+07 | 1097270 | 4.64E-01 | 0.1 | 0.81 |
| HILIC_POS_188_06851a2_921 | 4-hydroxy-4-(pyridin-2-yl)butan-2-one | 188.0685 | 2.92 | | HILIC | POS | 188.0684(100) | 2 | 183724 | 131480 | 171819 | 106710 | 4.64E-01 | -0.10 | 1.06 |
| HILIC_NEG_200_85927a6_853 | | 200.8593 | 6.85 | | HILIC | NEG | | | 1909699 | 803652 | 1830516 | 777923 | 4.64E-01 | -0.1 | 0.54 |
| HILIC_NEG_279_01645a4_248 | | 279.0165 | 4.25 | | HILIC | NEG | | | 252169 | 104241 | 242903 | 81212 | 4.64E-01 | -0.1 | 0.59 |
| HILIC_NEG_588_89624a4_352 | | 588.8962 | 4.35 | | HILIC | NEG | | | 7524947 | 526828 | 7473514 | 501725 | 4.64E-01 | -0.1 | 0.84 |
| HILIC_POS_361_3064a3_089 | | 361.3064 | 3.09 | | HILIC | POS | | | 567084 | 907877 | 481396 | 809747 | 4.64E-01 | -0.1 | 0.82 |
| RP_POS_600_46771a10_54 | | 600.4677 | 10.54 | | RP | POS | | | 2255989 | 496049 | 2211761 | 388468 | 4.64E-01 | -0.1 | 0.71 |
| RP_POS_481_34064a9_9 | | 481.3406 | 9.90 | | RP | POS | | | 333867 | 195500 | 355631 | 235177 | 4.65E-01 | 0.1 | 0.54 |
| RP_POS_526_02551a10_339 | | 526.0255 | 10.34 | | RP | POS | | | 604693 | 307195 | 637836 | 350291 | 4.65E-01 | 0.1 | 0.67 |
| RP_POS_594_75525a11_45 | | 594.7553 | 11.45 | | RP | POS | | | 128881 | 80262.7 | 138089 | 101022 | 4.65E-01 | 0.1 | 0.81 |
| RP_POS_718_70209a11_434 | | 718.7021 | 11.43 | | RP | POS | | | 223193 | 109857 | 235267 | 129176 | 4.65E-01 | 0.1 | 0.65 |
| RP_POS_872_5379a12_881 | | 872.5379 | 12.88 | | RP | POS | | | 6214156 | 2463936 | 6447308 | 2211610 | 4.65E-01 | 0.1 | 0.62 |
| RP_NEG_526_35193a10_778 | LysoPC O-16:0 | 526.3519 | 10.78 | | RP | NEG | 466.3306(100)377.2474(20), 78.9587(40) | 1 | 6552609 | 1710087 | 6383846 | 1666221 | 4.65E-01 | -0.10 | 0.67 |
| HILIC_NEG_276_84277a7_735 | | 276.8428 | 7.74 | | HILIC | NEG | | | 1998653 | 289191 | 1969640 | 290602 | 4.65E-01 | -0.1 | 0.75 |
| RP_NEG_226_01833a4_219 | | 226.0183 | 4.22 | | RP | NEG | | | 1053276 | 1427919 | 897383 | 1662895 | 4.65E-01 | -0.1 | 1.04 |
| RP_NEG_742_5401a13_95 | | 742.5401 | 13.95 | | RP | NEG | | | 1032236 | 533629 | 979175 | 529134 | 4.65E-01 | -0.1 | 0.57 |
| RP_POS_307_91751a15_47 | | 307.9175 | 15.47 | | RP | POS | | | 119790 | 88927.9 | 111695 | 73608 | 4.65E-01 | -0.1 | 0.62 |
| HILIC_POS_184_16937a0_622 | | 184.1694 | 0.62 | | HILIC | POS | | | 1393316 | 426539 | 1435217 | 415117 | 4.66E-01 | 0.1 | 0.87 |

**Supplemantary table 1**: The non-targeted metabolomics results ordered by the VIP value (p value < 0.0003 are shaded dark green, p value < 0.05 are shaded light green, d values below 0 are blue and above 0 are red)

| | | | | | | | | | Cases | | Controls | | Welch test | Cohen's | |
|---|---|---|---|---|---|---|---|---|---|---|---|---|---|---|---|
| Feature_ID | Metabolite_name | Average_Mz | Average_RT_min_ | HMDB ID | RP_HILIC | POS_NEG | Cureted_MSMS | ID_level | Mean | SD | Mean2 | SD3 | p | d | VIP |
| RP_NEG_313_16501a10_49 | | 313.1650 | 10.49 | | RP | NEG | | | 246144 | 172272 | 265014 | 202232 | 4.66E-01 | 0.1 | 0.74 |
| HILIC_POS_202_11855a4 | | 202.1186 | 4.00 | | HILIC | POS | | | 503118 | 738749 | 442427 | 471468 | 4.66E-01 | -0.1 | 0.56 |
| HILIC_POS_319_16586a1_38 | | 319.1659 | 1.38 | | HILIC | POS | | | 550353 | 382711 | 515523 | 318117 | 4.66E-01 | -0.1 | 0.69 |
| RP_NEG_775_68054a7_729 | | 775.6805 | 7.73 | | RP | NEG | | | 847603 | 198027 | 828286 | 190309 | 4.66E-01 | -0.1 | 0.58 |
| RP_POS_144_10204a0_675 | | 144.1020 | 0.68 | | RP | POS | | | 2.6E+07 | 1E+08 | 1.7E+07 | 7E+07 | 4.66E-01 | -0.1 | 0.46 |
| RP_POS_232_15424a5_199 | | 232.1542 | 5.20 | | RP | POS | | | 251020 | 106536 | 240579 | 103246 | 4.66E-01 | -0.1 | 0.65 |
| RP_POS_366_37375a11_897 | | 366.3738 | 11.90 | | RP | POS | | | 709060 | 328488 | 676669 | 322194 | 4.66E-01 | -0.1 | 0.66 |
| RP_POS_393_33868a12_323 | | 393.3387 | 12.32 | | RP | POS | | | 459977 | 212886 | 440738 | 174008 | 4.66E-01 | -0.1 | 0.64 |
| RP_POS_299_25833a10_416 | | 299.2583 | 10.42 | | RP | POS | | | 6393390 | 2363343 | 6608463 | 1976264 | 4.67E-01 | 0.1 | 0.84 |
| RP_POS_770_56891a13_074 | | 770.5689 | 13.07 | | RP | POS | | | 7598431 | 4888652 | 8157020 | 6165001 | 4.67E-01 | 0.1 | 0.93 |
| RP_POS_209_09227a1_244 | N-Acetyl-L-phenylalanine | 209.0923 | 1.24 | | RP | POS | 94.0654(100), 146.0602(90), 192.0651(60) | 1 | 2109310 | 981046 | 2009813 | 1016760 | 4.67E-01 | -0.10 | 0.72 |
| HILIC_NEG_99_92553a5_453 | | 99.9255 | 5.45 | | HILIC | NEG | | | 3389059 | 514484 | 3331145 | 633392 | 4.67E-01 | -0.1 | 0.49 |
| RP_POS_160_07581a1_64 | | 160.0758 | 1.64 | | RP | POS | | | 2158289 | 1007937 | 2060733 | 957845 | 4.67E-01 | -0.1 | 0.59 |
| RP_POS_316_28452a10_423 | | 316.2845 | 10.42 | | RP | POS | | | 629798 | 89189.4 | 620815 | 91053.9 | 4.67E-01 | -0.1 | 1.02 |
| RP_POS_569_0708a10_291 | | 569.0708 | 10.29 | | RP | POS | | | 320997 | 185720 | 302358 | 188382 | 4.67E-01 | -0.1 | 0.65 |
| RP_NEG_144_92349a11_142 | | 144.9235 | 11.14 | | RP | NEG | | | 1492859 | 207503 | 1516377 | 258814 | 4.68E-01 | 0.1 | 0.62 |
| RP_NEG_586_81927a15_363 | | 586.8193 | 15.36 | | RP | NEG | | | 390890 | 169141 | 408193 | 178259 | 4.68E-01 | 0.1 | 0.63 |
| RP_POS_197_98491a0_699 | | 197.9849 | 0.70 | | RP | POS | | | 536490 | 363988 | 577240 | 446284 | 4.68E-01 | 0.1 | 0.63 |
| RP_POS_323_21945a8_748 | | 323.2195 | 8.75 | | RP | POS | | | 183035 | 87930.2 | 191634 | 85432 | 4.68E-01 | 0.1 | 0.64 |
| RP_POS_622_10419a10_51 | | 622.1042 | 10.51 | | RP | POS | | | 219555 | 174748 | 236988 | 176030 | 4.68E-01 | 0.1 | 0.48 |
| RP_NEG_276_98596a3_583 | | 276.9860 | 3.58 | | RP | NEG | | | 977091 | 291101 | 948777 | 279489 | 4.68E-01 | -0.1 | 0.64 |
| RP_NEG_404_15948a11_592 | | 404.1595 | 11.59 | | RP | NEG | | | 1735184 | 76108.1 | 1726996 | 87640.4 | 4.68E-01 | -0.1 | 0.71 |
| RP_POS_156_06319a11_145 | | 156.0632 | 11.15 | | RP | POS | | | 3027532 | 588064 | 2965887 | 646232 | 4.68E-01 | -0.1 | 0.54 |
| RP_POS_588_41486a9_575 | | 588.4149 | 9.58 | | RP | POS | | | 1819499 | 914021 | 1734647 | 801536 | 4.68E-01 | -0.1 | 0.62 |
| RP_POS_645_46667a10_442 | | 645.4667 | 10.44 | | RP | POS | | | 3445253 | 1384138 | 3302740 | 1479986 | 4.68E-01 | -0.1 | 0.79 |
| HILIC_POS_610_62323a10_674 | | 610.6232 | 10.67 | | HILIC | POS | | | 477649 | 79004 | 485313 | 76025.4 | 4.69E-01 | 0.1 | 0.74 |
| RP_POS_399_76947a13_784 | | 399.7695 | 13.78 | | RP | POS | | | 4596961 | 3512969 | 4968969 | 3941276 | 4.69E-01 | 0.1 | 1.09 |
| HILIC_NEG_244_84846a5_502 | | 244.8485 | 5.50 | | HILIC | NEG | | | 1071541 | 396261 | 1034007 | 363519 | 4.69E-01 | -0.1 | 0.75 |
| HILIC_NEG_257_92691a0_844 | | 257.9269 | 0.84 | | HILIC | NEG | | | 147085 | 41645.4 | 143043 | 40094.3 | 4.69E-01 | -0.1 | 0.93 |
| RP_NEG_259_15533a6_886 | | 259.1553 | 6.89 | | RP | NEG | | | 233704 | 32813.8 | 230577 | 30455.3 | 4.69E-01 | -0.1 | 1.13 |
| RP_POS_244_19061a11_113 | | 244.1906 | 11.11 | | RP | POS | | | 628106 | 880912 | 547278 | 758093 | 4.69E-01 | -0.1 | 0.59 |
| RP_POS_314_0137a15_494 | | 314.0137 | 15.49 | | RP | POS | | | 1973732 | 769961 | 2057505 | 906765 | 4.70E-01 | 0.1 | 0.65 |
| RP_POS_621_43414a11_239 | | 621.4341 | 11.24 | | RP | POS | | | 693532 | 581059 | 750357 | 570019 | 4.70E-01 | 0.1 | 0.66 |
| RP_NEG_479_37531a12_163 | | 479.3753 | 12.16 | | RP | NEG | | | 261189 | 23346.7 | 258886 | 23297.1 | 4.70E-01 | -0.1 | 0.94 |
| RP_POS_175_0963a9_459 | | 175.0963 | 9.46 | | RP | POS | | | 1563078 | 1289399 | 1434791 | 1302917 | 4.70E-01 | -0.1 | 0.7 |
| HILIC_POS_147_14919a5_37 | | 147.1492 | 5.37 | | HILIC | POS | | | 4.5E+07 | 2.2E+07 | 4.7E+07 | 2.3E+07 | 4.71E-01 | 0.1 | 0.66 |
| RP_NEG_339_8782a0_86 | | 339.8782 | 0.86 | | RP | NEG | | | 858952 | 218310 | 878352 | 176752 | 4.71E-01 | 0.1 | 0.75 |
| RP_NEG_372_90155a15_344 | | 372.9016 | 15.34 | | RP | NEG | | | 3131403 | 99978.9 | 3141989 | 113248 | 4.71E-01 | 0.1 | 0.66 |
| RP_POS_167_03389a10_252 | | 167.0339 | 10.25 | | RP | POS | | | 371158 | 226437 | 392652 | 210854 | 4.71E-01 | 0.1 | 1.13 |
| RP_POS_445_11914a11_769 | | 445.1191 | 11.77 | | RP | POS | | | 126488 | 82031 | 135029 | 90410.2 | 4.71E-01 | 0.1 | 1.21 |
| RP_POS_537_39514a15_45 | | 537.3951 | 15.45 | | RP | POS | | | 458717 | 235682 | 485074 | 291712 | 4.71E-01 | 0.1 | 0.46 |
| HILIC_NEG_289_70261a1_406 | | 289.7026 | 1.41 | | HILIC | NEG | | | 184129 | 31786.3 | 181187 | 28137.4 | 4.71E-01 | -0.1 | 0.58 |
| HILIC_POS_100_05101a1_718 | | 100.0510 | 1.72 | | HILIC | POS | | | 420648 | 221547 | 403951 | 106303 | 4.71E-01 | -0.1 | 0.39 |
| RP_POS_254_24767a10_403 | | 254.2477 | 10.40 | | RP | POS | | | 291547 | 614027 | 240602 | 415424 | 4.71E-01 | -0.1 | 0.97 |
| RP_POS_366_37289a12_153 | | 366.3729 | 12.15 | | RP | POS | | | 7065076 | 1.3E+07 | 5888534 | 1.1E+07 | 4.71E-01 | -0.1 | 0.64 |
| RP_POS_431_35889a12_109 | | 431.3589 | 12.11 | | RP | POS | | | 302931 | 124952 | 290717 | 122861 | 4.71E-01 | -0.1 | 1.27 |
| RP_POS_558_73126a10_435 | | 558.7313 | 10.44 | | RP | POS | | | 777590 | 403663 | 739541 | 370705 | 4.71E-01 | -0.1 | 0.65 |
| RP_POS_299_65378a10_937 | | 299.6538 | 10.94 | | RP | POS | | | 444867 | 230837 | 468295 | 243858 | 4.72E-01 | 0.1 | 0.71 |
| RP_POS_320_25595a9_492 | | 320.2560 | 9.49 | | RP | POS | | | 61385.3 | 85301.6 | 70389.1 | 96525.6 | 4.72E-01 | 0.1 | 1.03 |
| RP_POS_359_00656a15_498 | | 359.0066 | 15.50 | | RP | POS | | | 577334 | 154061 | 593140 | 165947 | 4.72E-01 | 0.1 | 0.66 |
| RP_POS_143_07043a9_459 | | 143.0704 | 9.46 | | RP | POS | | | 3440468 | 2766631 | 3164593 | 2830161 | 4.72E-01 | -0.1 | 0.7 |
| RP_POS_502_00739a10_058 | | 502.0074 | 10.06 | | RP | POS | | | 679797 | 364580 | 644283 | 358180 | 4.72E-01 | -0.1 | 0.98 |
| HILIC_NEG_202_92902a4_412 | | 202.9290 | 4.41 | | HILIC | NEG | | | 7386916 | 742374 | 7448859 | 517602 | 4.73E-01 | 0.1 | 0.83 |
| RP_NEG_321_13171a7_693 | | 321.1317 | 7.69 | | RP | NEG | | | 830021 | 50119.1 | 834924 | 49841.7 | 4.73E-01 | 0.1 | 0.65 |
| RP_NEG_564_34497a11_639 | | 564.3450 | 11.64 | | RP | NEG | | | 3898570 | 1047934 | 4006642 | 1142330 | 4.73E-01 | 0.1 | 0.53 |
| RP_POS_399_27399a10_517 | | 399.2740 | 10.52 | | RP | POS | | | 620744 | 436627 | 667213 | 501951 | 4.73E-01 | 0.1 | 1.16 |
| RP_POS_482_29343a9_232 | | 482.2934 | 9.23 | | RP | POS | | | 737638 | 911377 | 836352 | 1076785 | 4.73E-01 | 0.1 | 0.74 |
| HILIC_POS_331_24707a0_845 | | 331.2471 | 0.85 | | HILIC | POS | | | 893694 | 320857 | 861377 | 335826 | 4.73E-01 | -0.1 | 0.59 |
| RP_NEG_271_19205a9_228 | | 271.1921 | 9.23 | | RP | NEG | | | 1152993 | 121970 | 1141587 | 111179 | 4.73E-01 | -0.1 | 1.09 |
| RP_NEG_851_11475a9_581 | | 851.1148 | 9.58 | | RP | NEG | | | 5912729 | 363199 | 5879604 | 313754 | 4.73E-01 | -0.1 | 0.96 |
| RP_POS_145_12239a6_501 | | 145.1224 | 6.50 | | RP | POS | | | 807697 | 81133.9 | 799950 | 76892.7 | 4.73E-01 | -0.1 | 0.96 |
| RP_POS_165_0609a15_324 | | 165.0609 | 15.32 | | RP | POS | | | 1.4E+07 | 2933972 | 1.3E+07 | 2967635 | 4.73E-01 | -0.1 | 0.5 |
| RP_POS_368_21985a14_216 | | 368.2199 | 14.22 | | RP | POS | | | 163184 | 107500 | 153501 | 90518.9 | 4.73E-01 | -0.1 | 0.46 |
| RP_POS_429_37277a13_931 | | 429.3728 | 13.93 | | RP | POS | | | 3970562 | 952823 | 3878360 | 927126 | 4.73E-01 | -0.1 | 0.44 |
| RP_NEG_160_89252a14_343 | | 160.8925 | 14.34 | | RP | NEG | | | 1286532 | 575798 | 1339369 | 507211 | 4.74E-01 | 0.1 | 0.63 |
| RP_NEG_332_78976a15_38 | | 332.7898 | 15.38 | | RP | NEG | | | 3597991 | 618414 | 3657088 | 590844 | 4.74E-01 | 0.1 | 0.44 |
| RP_POS_364_2059a11_241 | | 364.2059 | 11.24 | | RP | POS | | | 244554 | 136876 | 258162 | 140691 | 4.74E-01 | 0.1 | 0.77 |
| RP_POS_628_94104a11_538 | | 628.9410 | 11.54 | | RP | POS | | | 213246 | 96293 | 224181 | 123186 | 4.74E-01 | 0.1 | 0.82 |
| HILIC_NEG_100_93348a5_397 | | 100.9335 | 5.40 | | HILIC | NEG | | | 4.8E+07 | 1.9E+07 | 4.6E+07 | 1.6E+07 | 4.74E-01 | -0.1 | 0.6 |
| HILIC_NEG_212_91113a4_917 | | 212.9111 | 4.92 | | HILIC | NEG | | | 2685336 | 1168081 | 2583525 | 917141 | 4.74E-01 | -0.1 | 0.54 |
| HILIC_POS_176_13927a3_617 | | 176.1393 | 3.62 | | HILIC | POS | | | 3104236 | 6233558 | 2438595 | 7229817 | 4.74E-01 | -0.1 | 0.55 |
| RP_POS_300_28949a11_091 | | 300.2895 | 11.09 | | RP | POS | | | 456551 | 76160.5 | 448199 | 92113.9 | 4.74E-01 | -0.1 | 0.7 |
| RP_POS_768_55298a12_796 | | 768.5530 | 12.80 | | RP | POS | | | 1.7E+07 | 5710900 | 1.6E+07 | 4995330 | 4.74E-01 | -0.1 | 0.68 |
| RP_POS_221_98396a13_192 | | 221.9840 | 13.19 | | RP | POS | | | 655553 | 348324 | 690607 | 367474 | 4.75E-01 | 0.1 | 0.72 |
| RP_POS_227_16415a8_384 | | 227.1642 | 8.38 | | RP | POS | | | 927554 | 116722 | 938400 | 106022 | 4.75E-01 | 0.1 | 0.76 |
| RP_POS_453_34204a9_157 | | 453.3420 | 9.16 | | RP | POS | | | 578140 | 344823 | 616657 | 431704 | 4.75E-01 | 0.1 | 0.76 |
| RP_POS_614_26398a11_325 | | 614.2640 | 11.33 | | RP | POS | | | 120361 | 74051.2 | 127663 | 75369.9 | 4.75E-01 | 0.1 | 0.63 |
| HILIC_NEG_163_89777a6_301 | | 163.8978 | 6.30 | | HILIC | NEG | | | 5167333 | 1916901 | 4994917 | 1626813 | 4.75E-01 | -0.1 | 0.69 |
| RP_NEG_390_76273a15_407 | | 390.7627 | 15.41 | | RP | NEG | | | 162214 | 85743 | 154702 | 68622.4 | 4.75E-01 | -0.1 | 0.64 |
| RP_POS_540_30597a10_015 | | 540.3060 | 10.02 | | RP | POS | | | 3244977 | 1554926 | 3088720 | 1637664 | 4.75E-01 | -0.1 | 0.63 |
| HILIC_NEG_351_9975a7_409 | | 351.9975 | 7.41 | | HILIC | NEG | | | 807162 | 46519 | 811650 | 45520.2 | 4.76E-01 | 0.1 | 0.72 |
| HILIC_POS_162_99458a4_805 | | 162.9946 | 4.81 | | HILIC | POS | | | 165415 | 145736 | 178504 | 123661 | 4.76E-01 | 0.1 | 0.85 |
| HILIC_POS_506_35901a1_011 | | 506.3590 | 1.01 | | HILIC | POS | | | 137591 | 75253.1 | 144498 | 66782.2 | 4.76E-01 | 0.1 | 0.77 |
| RP_NEG_242_94386a15_333 | | 242.9439 | 15.33 | | RP | NEG | | | 4675388 | 539874 | 4725478 | 491170 | 4.76E-01 | 0.1 | 0.57 |
| RP_POS_198_18532a10_649 | | 198.1853 | 10.65 | | RP | POS | | | 1.7E+07 | 5431603 | 1.8E+07 | 5019148 | 4.76E-01 | 0.1 | 0.58 |
| RP_POS_331_26312a11_038 | | 331.2631 | 11.04 | | RP | POS | | | 2001509 | 1322536 | 2123499 | 1184888 | 4.76E-01 | 0.1 | 1.09 |
| RP_POS_414_21118a7_783 | | 414.2112 | 7.78 | | RP | POS | | | 433049 | 60535.1 | 438808 | 57709.7 | 4.76E-01 | 0.1 | 0.73 |
| RP_POS_750_7226a11_536 | | 750.7226 | 11.54 | | RP | POS | | | 181132 | 89811.2 | 191679 | 121477 | 4.76E-01 | 0.1 | 0.62 |
| HILIC_POS_106_08682a5_094 | | 106.0868 | 5.09 | | HILIC | POS | 88.0761(100), 70.066(60) | | 1.7E+07 | 1.3E+07 | 1.6E+07 | 1E+07 | 4.76E-01 | -0.1 | 0.87 |
| HILIC_POS_189_07315a5_653 | | 189.0732 | 5.65 | | HILIC | POS | | | 4090462 | 359454 | 4058971 | 287874 | 4.76E-01 | -0.1 | 0.83 |
| RP_NEG_1103_65076a10_494 | | 1103.6508 | 10.49 | | RP | NEG | | | 3721573 | 323296 | 3687898 | 362766 | 4.76E-01 | -0.1 | 0.8 |
| RP_NEG_329_65689a15_497 | | 329.6569 | 15.50 | | RP | NEG | | | 354854 | 33101.8 | 351445 | 36471 | 4.76E-01 | -0.1 | 0.6 |
| RP_POS_239_1619a8_542 | | 239.1619 | 8.54 | | RP | POS | | | 8866207 | 1932516 | 8691903 | 1651778 | 4.76E-01 | -0.1 | 1.02 |
| HILIC_NEG_293_74075a3_297 | | 293.7408 | 3.30 | | HILIC | NEG | | | 125681 | 91435.7 | 134522 | 90393.5 | 4.77E-01 | 0.1 | 0.66 |
| HILIC_POS_338_34189a1_349 | | 338.3419 | 1.35 | | HILIC | POS | | | 167966 | 72587.3 | 175315 | 77861.7 | 4.77E-01 | 0.1 | 0.56 |
| RP_NEG_190_92879a13_289 | | 190.9288 | 13.29 | | RP | NEG | | | 592330 | 139170 | 605668 | 135480 | 4.77E-01 | 0.1 | 0.52 |
| RP_NEG_520_91107a11_363 | | 520.9111 | 11.36 | | RP | NEG | | | 792387 | 206785 | 814136 | 236983 | 4.77E-01 | 0.1 | 0.92 |
| RP_POS_142_08635a0_673 | | 142.0864 | 0.67 | | RP | POS | | | 468590 | 399389 | 503830 | 327347 | 4.77E-01 | 0.1 | 0.53 |
| RP_POS_267_12048a3_532 | | 267.1205 | 3.53 | | RP | POS | | | 126850 | 124162 | 140148 | 146589 | 4.77E-01 | 0.1 | 0.72 |
| HILIC_NEG_99_92571a3_161 | | 99.9257 | 3.16 | | HILIC | NEG | | | 7084561 | 1896443 | 6897372 | 1943334 | 4.77E-01 | -0.1 | 0.67 |
| RP_POS_368_28192a9_692 | | 368.2819 | 9.69 | | RP | POS | | | 77852.2 | 42876.4 | 73918.4 | 38257.6 | 4.77E-01 | -0.1 | 0.6 |
| RP_POS_604_28955a10_311 | | 604.2896 | 10.31 | | RP | POS | | | 513311 | 454087 | 473960 | 357000 | 4.77E-01 | -0.1 | 0.82 |
| HILIC_POS_421_28946a3_09 | | 421.2895 | 3.09 | | HILIC | POS | | | 72970.8 | 25944.2 | 76461.4 | 42557.2 | 4.78E-01 | 0.1 | 0.53 |
| RP_POS_395_24045a10_252 | | 395.2405 | 10.25 | | RP | POS | | | 2.7E+07 | 1.3E+07 | 2.8E+07 | 1.2E+07 | 4.78E-01 | 0.1 | 1.01 |
| HILIC_POS_106_08678a3_003 | | 106.0868 | 3.00 | | HILIC | POS | 88.0762(100), 106.0864(90), 70.066(55) | | 9911860 | 6199290 | 9340324 | 5600189 | 4.78E-01 | -0.1 | 0.84 |
| RP_POS_142_94943a15_407 | | 142.9494 | 15.41 | | RP | POS | | | 4.1E+07 | 8081537 | 4E+07 | 8535390 | 4.78E-01 | -0.1 | 1.14 |
| RP_POS_310_20108a5_748 | | 310.2011 | 5.75 | | RP | POS | | | 770686 | 403575 | 729017 | 450064 | 4.78E-01 | -0.1 | 0.63 |
| RP_POS_343_33884a12_626 | | 343.3388 | 12.63 | | RP | POS | | | 1.8E+07 | 9546528 | 1.7E+07 | 8729346 | 4.78E-01 | -0.1 | 0.81 |
| RP_POS_526_29266a10_181 | | 526.2927 | 10.18 | | RP | POS | | | 6919614 | 2046667 | 6716373 | 2135575 | 4.78E-01 | -0.1 | 0.66 |
| HILIC_NEG_112_98588a5_467 | | 112.9859 | 5.47 | | HILIC | NEG | 68.9949(100), 112.9856(5) | | 1.3E+07 | 1861850 | 1.3E+07 | 1799464 | 4.79E-01 | 0.1 | 0.7 |
| RP_NEG_255_10278a8_34 | | 255.1028 | 8.34 | | RP | NEG | | | 320001 | 568669 | 386880 | 778540 | 4.79E-01 | 0.1 | 0.95 |
| RP_POS_168_98045a15_434 | | 168.9805 | 15.43 | | RP | POS | | | 8551214 | 2016544 | 8806121 | 3057759 | 4.79E-01 | 0.1 | 0.61 |
| RP_POS_299_30527a9_272 | | 299.3053 | 9.27 | | RP | POS | | | 1287161 | 641555 | 1347539 | 608774 | 4.79E-01 | 0.1 | 0.71 |
| RP_POS_343_33899a11_255 | | 343.3390 | 11.26 | | RP | POS | | | 1.5E+07 | 9278099 | 1.6E+07 | 7537608 | 4.79E-01 | 0.1 | 0.74 |
| RP_POS_806_50732a11_642 | | 806.5073 | 11.64 | | RP | POS | | | 4632775 | 1822093 | 4822470 | 2064105 | 4.79E-01 | 0.1 | 0.68 |
| HILIC_NEG_130_99269a1_342 | | 130.9927 | 1.34 | | HILIC | NEG | | | 3342643 | 864355 | 3257704 | 888822 | 4.79E-01 | -0.1 | 0.83 |
| HILIC_NEG_207_83675a10_646 | | 207.8368 | 10.65 | | HILIC | NEG | | | 362047 | 421602 | 327764 | 285139 | 4.79E-01 | -0.1 | 0.44 |
| RP_NEG_417_2287a9_414 | | 417.2287 | 9.41 | | RP | NEG | | | 302370 | 29128.8 | 299470 | 30640.8 | 4.79E-01 | -0.1 | 0.62 |
| RP_POS_358_05011a8_096 | | 358.0501 | 8.10 | | RP | POS | | | 121842 | 53601 | 126729 | 47862.7 | 4.80E-01 | 0.1 | 0.49 |
| RP_POS_773_07831a13_785 | | 773.0783 | 13.79 | | RP | POS | | | 1543912 | 847244 | 1630921 | 942321 | 4.80E-01 | 0.1 | 0.95 |
| HILIC_NEG_209_00455a1_348 | | 209.0046 | 1.35 | | HILIC | NEG | | | 757319 | 230712 | 735614 | 219791 | 4.80E-01 | -0.1 | 0.57 |
| HILIC_POS_172_09418a4_359 | | 172.0942 | 4.36 | | HILIC | POS | | | 2505304 | 1983591 | 2324732 | 1765605 | 4.80E-01 | -0.1 | 1.03 |
| RP_NEG_198_93665a0_649 | | 198.9367 | 0.65 | | RP | NEG | | | 2110318 | 326515 | 2078808 | 325992 | 4.80E-01 | -0.1 | 0.53 |
| RP_POS_243_19521a8_3 | | 243.1952 | 8.30 | | RP | POS | | | 668078 | 85681.6 | 659657 | 88519.9 | 4.80E-01 | -0.1 | 0.76 |
| RP_POS_385_16931a9_207 | | 385.1693 | 9.21 | | RP | POS | | | 276318 | 185790 | 258069 | 191182 | 4.80E-01 | -0.1 | 0.7 |
| RP_POS_460_27551a4_801 | | 460.2755 | 4.80 | | RP | POS | | | 907503 | 115797 | 918675 | 115751 | 4.81E-01 | 0.1 | 0.7 |
| RP_POS_716_55121a10_871 | | 716.5512 | 10.87 | | RP | POS | | | 1068675 | 231707 | 1090994 | 231739 | 4.81E-01 | 0.1 | 0.79 |
| RP_POS_228_1958a11_113 | | 228.1958 | 11.11 | | RP | POS | 95.0862(100), 175.1479(50) | | 2645851 | 3586952 | 2331273 | 2964671 | 4.81E-01 | -0.1 | 0.57 |
| RP_NEG_360_65762a15_498 | | 360.6576 | 15.50 | | RP | NEG | | | 216963 | 101419 | 208255 | 79825.9 | 4.81E-01 | -0.1 | 0.48 |
| RP_POS_289_08316a7_659 | | 289.0832 | 7.66 | | RP | POS | | | 505946 | 52003.5 | 500947 | 51696.8 | 4.81E-01 | -0.1 | 1.05 |
| RP_POS_383_16653a10_895 | | 383.1665 | 10.90 | | RP | POS | | | 1027554 | 462807 | 982096 | 479337 | 4.81E-01 | -0.1 | 0.95 |
| RP_POS_623_60828a11_647 | | 623.6083 | 11.65 | | RP | POS | | | 151466 | 85801.8 | 143292 | 83760.1 | 4.81E-01 | -0.1 | 0.43 |
| RP_POS_305_24551a11_155 | | 305.2455 | 11.16 | | RP | POS | 305.1555(100) | | 357530 | 406542 | 401327 | 492976 | 4.82E-01 | 0.1 | 0.61 |
| HILIC_POS_172_09416a4_896 | | 172.0942 | 4.90 | | HILIC | POS | | | 2763728 | 2195956 | 3003395 | 2712325 | 4.82E-01 | 0.1 | 0.87 |
| RP_POS_219_08614a4_826 | | 219.0861 | 4.83 | | RP | POS | | | 600557 | 326447 | 643948 | 534208 | 4.82E-01 | 0.1 | 0.75 |
| RP_POS_975_64056a10_481 | | 975.6406 | 10.48 | | RP | POS | | | 604180 | 327009 | 637277 | 358390 | 4.82E-01 | 0.1 | 0.6 |
| RP_NEG_176_90906a11_591 | | 176.9091 | 11.59 | | RP | NEG | | | 911727 | 107131 | 902017 | 95569.6 | 4.82E-01 | -0.1 | 0.55 |
| RP_NEG_197_80814a0_729 | | 197.8081 | 0.73 | | RP | NEG | | | 3905731 | 971300 | 3794456 | 1294236 | 4.82E-01 | -0.1 | 0.68 |
| RP_POS_276_21722a9_209 | | 276.2172 | 9.21 | | RP | POS | | | 5247293 | 1843301 | 5065577 | 1927624 | 4.82E-01 | -0.1 | 0.67 |
| RP_POS_315_11133a8_844 | | 315.1113 | 8.84 | | RP | POS | | | 2182765 | 1667157 | 2030610 | 1504150 | 4.82E-01 | -0.1 | 1.23 |
| RP_POS_560_4707a11_327 | | 560.4707 | 11.33 | | RP | POS | | | 1800900 | 4366457 | 1470094 | 2383368 | 4.82E-01 | -0.1 | 0.66 |
| RP_POS_590_48108a12_486 | | 590.4811 | 12.49 | | RP | POS | | | 1270781 | 442556 | 1224599 | 510004 | 4.82E-01 | -0.1 | 0.9 |
| RP_NEG_297_15314a10_009 | Aurapten | 297.1531 | 10.01 | | RP | NEG | 297.1524(100), 183.0124(15) | 2 | 6767749 | 9448230 | 6104016 | 3487713 | 4.82E-01 | -0.10 | 0.46 |
| HILIC_POS_64_01671a4_339 | | 64.0167 | 4.34 | | HILIC | POS | | | 2.7E+08 | 3.6E+07 | 2.8E+08 | 2.5E+07 | 4.83E-01 | 0.1 | 0.67 |
| RP_POS_182_02367a15_528 | | 182.0237 | 15.53 | | RP | POS | | | 3E+07 | 1657030 | 3E+07 | 1395012 | 4.83E-01 | 0.1 | 1.03 |
| RP_POS_123_98306a15_406 | | 123.9831 | 15.41 | | RP | POS | | | 844502 | 397838 | 806937 | 385474 | 4.83E-01 | -0.1 | 0.53 |
| RP_POS_212_07423a0_632 | | 212.0742 | 0.63 | | RP | POS | | | 3708217 | 1134434 | 3597111 | 1176328 | 4.83E-01 | -0.1 | 0.47 |
| RP_POS_733_33093a4_891 | | 733.3309 | 4.89 | | RP | POS | | | 918581 | 1923732 | 781104 | 810803 | 4.83E-01 | -0.1 | 0.8 |
| RP_POS_132_10201a1_343 | L-Isoleucine | 132.1020 | 1.34 | | RP | POS | 86.0974(100) | 1 | 2E+07 | 6937974 | 1.9E+07 | 4971690 | 4.83E-01 | -0.10 | 0.61 |
| HILIC_NEG_180_9893a5_108 | | 180.9893 | 5.11 | | HILIC | NEG | | | 487688 | 253043 | 509507 | 204451 | 4.84E-01 | 0.1 | 0.41 |
| RP_NEG_1103_65198a10_358 | | 1103.6520 | 10.36 | | RP | NEG | | | 721045 | 185637 | 740032 | 208044 | 4.84E-01 | 0.1 | 0.96 |
| RP_NEG_180_86208a14_341 | | 180.8621 | 14.34 | | RP | NEG | | | 790806 | 411926 | 825590 | 315734 | 4.84E-01 | 0.1 | 0.42 |
| RP_NEG_186_04538a0_68 | | 186.0454 | 0.68 | | RP | NEG | | | 398268 | 96409.8 | 407861 | 103224 | 4.84E-01 | 0.1 | 0.81 |
| RP_NEG_342_97974a8_399 | | 342.9797 | 8.40 | | RP | NEG | | | 1297355 | 529199 | 1350013 | 567373 | 4.84E-01 | 0.1 | 0.62 |
| RP_POS_364_16934a10_714 | | 364.1693 | 10.71 | | RP | POS | | | 477821 | 370983 | 512285 | 350873 | 4.84E-01 | 0.1 | 0.94 |
| RP_POS_416_78903a13_961 | | 416.7890 | 13.96 | | RP | POS | | | 1119375 | 925007 | 1207210 | 909433 | 4.84E-01 | 0.1 | 0.76 |
| RP_POS_655_7901a10_582 | | 655.7901 | 10.58 | | RP | POS | | | 162968 | 95911.2 | 173058 | 112780 | 4.84E-01 | 0.1 | 0.48 |
| RP_POS_749_55811a12_504 | | 749.5581 | 12.50 | | RP | POS | | | 529835 | 392788 | 567082 | 385508 | 4.84E-01 | 0.1 | 0.58 |
| RP_NEG_323_22095a11_142 | | 323.2210 | 11.14 | | RP | NEG | | | 2949164 | 727999 | 2882219 | 674997 | 4.84E-01 | -0.1 | 1.3 |
| RP_POS_339_07742a8_842 | | 339.0774 | 8.84 | | RP | POS | | | 483512 | 343207 | 453070 | 294438 | 4.84E-01 | -0.1 | 1.01 |
| RP_POS_617_10004a10_375 | | 617.1000 | 10.38 | | RP | POS | | | 505277 | 280361 | 475233 | 339588 | 4.84E-01 | -0.1 | 0.82 |
| RP_POS_694_31622a11_618 | | 694.3162 | 11.62 | | RP | POS | | | 113134 | 56699.4 | 107226 | 65623.7 | 4.84E-01 | -0.1 | 0.56 |

**Supplemantary table 1**: The non-targeted metabolomics results ordered by the VIP value (p value < 0.0003 are shaded dark green, p value < 0.05 are shaded light green, d values below 0 are blue and above 0 are red)

| | | | | | | | | | Cases | | Controls | | Welch test | Cohen's | |
|---|---|---|---|---|---|---|---|---|---|---|---|---|---|---|---|
| Feature_ID | Metabolite_name | Average_Mz | Average_RT_min_ | HMDB ID | RP_HILIC | POS_NEG | Cureted_MSMS | ID_level | Mean | SD | Mean2 | SD3 | p | d | VIP |
| HILIC_NEG_131_04619a6_244 | Asparagine | 131.0462 | 6.24 | | HILIC | NEG | 114.0201(100), 70.0294(45),131.0458(30) | 1 | 1126111 | 260728 | 1102718 | 230075 | 4.84E-01 | -0.10 | 0.58 |
| HILIC_POS_169_09456a7_093 | Pyridoxamine | 169.0946 | 7.09 | | HILIC | POS | 169.0958(100) | 1 | 486819 | 167933 | 472179 | 138904 | 4.84E-01 | -0.10 | 0.53 |
| HILIC_NEG_278_01776a4 | | 278.0178 | 4.00 | | HILIC | NEG | | | 221656 | 65809.7 | 227397 | 54953.1 | 4.85E-01 | 0.1 | 0.71 |
| RP_NEG_170_83345a7_263 | | 170.8335 | 7.26 | | RP | NEG | | | 2404226 | 668781 | 2470621 | 715348 | 4.85E-01 | 0.1 | 0.66 |
| RP_NEG_357_03122a0_678 | | 357.0312 | 0.68 | | RP | NEG | | | 2091576 | 503983 | 2140076 | 511647 | 4.85E-01 | 0.1 | 0.5 |
| RP_NEG_480_31015a10_793 | LysoPE 18:0 | 480.3102 | 10.79 | | RP | NEG | 283.2637(100), 480.311(5) | 1 | 1416356 | 318876 | 1448835 | 356261 | 4.85E-01 | 0.10 | 0.8 |
| HILIC_NEG_223_00851a1_345 | | 223.0085 | 1.35 | | HILIC | NEG | | | 753582 | 230062 | 733380 | 194160 | 4.85E-01 | -0.1 | 0.96 |
| RP_POS_251_12549a4_642 | | 251.1255 | 4.64 | | RP | POS | | | 144275 | 94070.9 | 136130 | 77115.5 | 4.85E-01 | -0.1 | 1.12 |
| RP_POS_259_20502a10_065 | | 259.2050 | 10.07 | | RP | POS | | | 7358842 | 434505 | 7314103 | 494571 | 4.85E-01 | -0.1 | 0.59 |
| HILIC_POS_293_09805a7_745 | Ethylenediaminetetraacetic acid EDTA | 293.0981 | 7.75 | | HILIC | POS | 160.0613(100), 293.0962(10) | 2 | 1.6E+07 | 1372180 | 1.6E+07 | 1412709 | 4.85E-01 | -0.10 | 0.51 |
| RP_POS_130_06535a2_478 | | 130.0654 | 2.48 | | RP | POS | 130.0658(100), 102.0561(2) | | 1794318 | 593277 | 1852929 | 633340 | 4.86E-01 | 0.1 | 0.61 |
| RP_POS_153_09109a6_598 | | 153.0911 | 6.60 | | RP | POS | | | 2266029 | 257337 | 2243384 | 219776 | 4.86E-01 | -0.09 | 0.55 |
| RP_POS_207_1357a6_654 | | 207.1357 | 6.65 | | RP | POS | | | 566397 | 248410 | 542323 | 255723 | 4.86E-01 | -0.1 | 0.67 |
| RP_NEG_160_84183a11_595 | | 160.8418 | 11.60 | | RP | NEG | | | 150068 | 45920 | 154460 | 46326.2 | 4.87E-01 | 0.1 | 0.79 |
| RP_POS_266_17258a7_657 | | 266.1726 | 7.66 | | RP | POS | | | 612017 | 263466 | 638135 | 284101 | 4.87E-01 | 0.1 | 0.86 |
| RP_POS_354_08487a15_721 | | 354.0849 | 15.72 | | RP | POS | | | 4601783 | 2731346 | 4876125 | 3011789 | 4.87E-01 | 0.1 | 0.48 |
| RP_POS_703_89441a11_558 | | 703.8944 | 11.56 | | RP | POS | | | 147492 | 70010.3 | 156219 | 106729 | 4.87E-01 | 0.1 | 0.53 |
| RP_POS_716_1972a8_778 | | 716.1972 | 8.78 | | RP | POS | | | 973284 | 87791.6 | 982180 | 98055.2 | 4.87E-01 | 0.1 | 0.78 |
| RP_POS_780_21198a9_114 | | 780.2120 | 9.11 | | RP | POS | | | 714397 | 77870.8 | 721923 | 80141.3 | 4.87E-01 | 0.1 | 1.01 |
| RP_POS_166_98515a15_403 | | 166.9852 | 15.40 | | RP | POS | | | 2.2E+07 | 7123000 | 2.2E+07 | 6966123 | 4.87E-01 | 0.09 | 0.51 |
| HILIC_POS_206_11974a1_06 | | 206.1197 | 1.06 | | HILIC | POS | | | 54217.6 | 29417.6 | 51603.9 | 25777.6 | 4.87E-01 | -0.09 | 0.84 |
| RP_POS_386_2323a9_209 | | 386.2323 | 9.21 | | RP | POS | | | 2331124 | 4272394 | 1980592 | 3114402 | 4.87E-01 | -0.09 | 0.64 |
| RP_POS_297_24231a8_882 | | 297.2423 | 8.88 | | RP | POS | | | 1097043 | 171915 | 1080115 | 182724 | 4.87E-01 | -0.1 | 0.84 |
| RP_POS_327_22064a15_393 | | 327.2206 | 15.39 | | RP | POS | | | 382461 | 221955 | 364563 | 154463 | 4.87E-01 | -0.1 | 0.58 |
| RP_POS_497_315a13_232 | | 497.3150 | 13.23 | | RP | POS | | | 263841 | 180994 | 246613 | 181552 | 4.87E-01 | -0.1 | 0.59 |
| RP_POS_1184_84155a13_193 | | 1184.8416 | 13.19 | | RP | POS | | | 4582750 | 2824490 | 4854787 | 2897494 | 4.88E-01 | 0.1 | 0.72 |
| RP_POS_171_06549a3_116 | | 171.0655 | 3.12 | | RP | POS | | | 241950 | 122575 | 254238 | 135083 | 4.88E-01 | 0.1 | 0.95 |
| HILIC_POS_427_26663a0_937 | | 427.2666 | 0.94 | | HILIC | POS | | | 532084 | 99779.3 | 541381 | 96639.4 | 4.88E-01 | 0.09 | 0.63 |
| RP_NEG_723_09143a9_458 | | 723.0914 | 9.46 | | RP | NEG | | | 2982246 | 158302 | 2967510 | 152947 | 4.88E-01 | -0.09 | 0.86 |
| RP_POS_295_15164a5_599 | | 295.1516 | 5.60 | | RP | POS | | | 158698 | 117147 | 147550 | 117771 | 4.88E-01 | -0.09 | 1.08 |
| RP_POS_429_31766a10_442 | | 429.3177 | 10.44 | | RP | POS | | | 3987454 | 749245 | 3915778 | 760167 | 4.88E-01 | -0.09 | 1.14 |
| HILIC_NEG_146_96593a0_915 | | 146.9659 | 0.92 | | HILIC | NEG | | | 968037 | 266393 | 995132 | 301767 | 4.89E-01 | 0.1 | 0.58 |
| RP_NEG_168_83641a15_467 | | 168.8364 | 15.47 | | RP | NEG | | | 7221793 | 826010 | 7302076 | 865347 | 4.89E-01 | 0.09 | 0.6 |
| RP_NEG_629_11096a9_099 | | 629.1110 | 9.10 | | RP | NEG | | | 976750 | 52493.2 | 981760 | 53264.6 | 4.89E-01 | 0.09 | 0.69 |
| RP_POS_514_70508a10_486 | | 514.7051 | 10.49 | | RP | POS | | | 268027 | 203275 | 287715 | 212223 | 4.89E-01 | 0.09 | 0.57 |
| RP_POS_405_29974a9_392 | | 405.2997 | 9.39 | | RP | POS | | | 1112591 | 622771 | 1045702 | 770665 | 4.89E-01 | -0.1 | 0.74 |
| RP_POS_407_75491a13_804 | | 407.7549 | 13.80 | | RP | POS | | | 3008385 | 2241753 | 3204767 | 1931420 | 4.90E-01 | 0.09 | 0.6 |
| RP_POS_681_07794a11_425 | | 681.0779 | 11.43 | | RP | POS | | | 107331 | 62182.2 | 113098 | 60016.3 | 4.90E-01 | 0.09 | 0.57 |
| HILIC_NEG_197_04359a0_845 | | 197.0436 | 0.85 | | HILIC | NEG | | | 337769 | 83755.6 | 330119 | 78610.9 | 4.90E-01 | -0.09 | 0.78 |
| HILIC_NEG_278_83875a7_729 | | 278.8388 | 7.73 | | HILIC | NEG | | | 174532 | 26483.3 | 172025 | 26616.1 | 4.90E-01 | -0.09 | 0.66 |
| RP_NEG_608_31793a10_506 | | 608.3179 | 10.51 | | RP | NEG | | | 1.5E+07 | 2343098 | 1.5E+07 | 1954003 | 4.90E-01 | -0.09 | 0.6 |
| RP_POS_468_44089a9_974 | | 468.4409 | 9.97 | | RP | POS | | | 104542 | 168313 | 89827.4 | 144139 | 4.90E-01 | -0.09 | 0.67 |
| RP_NEG_428_96387a8_307 | | 428.9639 | 8.31 | | RP | NEG | | | 384420 | 107733 | 373367 | 124468 | 4.90E-01 | -0.1 | 0.45 |
| RP_NEG_407_18829a11_404 | | 407.1883 | 11.40 | | RP | NEG | | | 110031 | 115257 | 121905 | 134271 | 4.91E-01 | 0.1 | 0.62 |
| HILIC_NEG_162_84258a3_114 | | 162.8426 | 3.11 | | HILIC | NEG | | | 3E+07 | 3365708 | 3E+07 | 3332853 | 4.91E-01 | 0.09 | 0.45 |
| RP_POS_356_85938a15_435 | | 356.8594 | 15.44 | | RP | POS | | | 168224 | 172709 | 185595 | 193966 | 4.91E-01 | 0.09 | 0.73 |
| HILIC_NEG_452_92242a4_352 | | 452.9224 | 4.35 | | HILIC | NEG | | | 1.9E+07 | 1340037 | 1.9E+07 | 1273843 | 4.91E-01 | -0.09 | 0.87 |
| RP_NEG_527_18286a11_143 | | 527.1829 | 11.14 | | RP | NEG | | | 631932 | 113611 | 621870 | 100861 | 4.91E-01 | -0.09 | 1.23 |
| RP_POS_390_19724a15_384 | | 390.1972 | 15.38 | | RP | POS | | | 232199 | 99263.1 | 223654 | 82710.1 | 4.91E-01 | -0.09 | 0.69 |
| RP_POS_562_51874a15_186 | | 562.5187 | 15.19 | | RP | POS | | | 2.3E+07 | 2794999 | 2.3E+07 | 2909879 | 4.91E-01 | -0.09 | 0.5 |
| RP_POS_700_48828a12_328 | | 700.4883 | 12.33 | | RP | POS | | | 543127 | 609555 | 483170 | 658565 | 4.91E-01 | -0.09 | 0.47 |
| RP_POS_399_25049a9_69 | Tri(butoxyethyl)phosphate | 399.2505 | 9.69 | | RP | POS | 57.0711(100), 199.0377(98) | 2 | 1564507 | 1542548 | 1774490 | 2669054 | 4.91E-01 | 0.10 | 0.56 |
| HILIC_POS_394_31705a3_13 | | 394.3171 | 3.13 | | HILIC | POS | | | 821424 | 982477 | 931726 | 1311972 | 4.92E-01 | 0.1 | 0.61 |
| HILIC_NEG_130_99274a2_268 | | 130.9927 | 2.27 | | HILIC | NEG | | | 1517405 | 379307 | 1551516 | 348333 | 4.92E-01 | 0.09 | 0.5 |
| RP_NEG_201_88939a15_517 | | 201.8894 | 15.52 | | RP | NEG | | | 1260698 | 208337 | 1281594 | 233502 | 4.92E-01 | 0.09 | 0.59 |
| RP_NEG_254_62297a8_221 | | 254.6230 | 8.22 | | RP | NEG | | | 1797472 | 827687 | 1882676 | 968319 | 4.92E-01 | 0.09 | 0.71 |
| RP_POS_365_3024a11_242 | | 365.3024 | 11.24 | | RP | POS | | | 854223 | 675363 | 912410 | 566491 | 4.92E-01 | 0.09 | 0.6 |
| RP_POS_686_88086a11_333 | | 686.8809 | 11.33 | | RP | POS | | | 162473 | 94351.8 | 171656 | 100573 | 4.92E-01 | 0.09 | 0.62 |
| RP_POS_360_32364a11_658 | | 360.3236 | 11.66 | | RP | POS | | | 4.8E+07 | 8E+07 | 4.1E+07 | 6.6E+07 | 4.92E-01 | -0.09 | 0.61 |
| RP_NEG_385_16943a5_732 | | 385.1694 | 5.73 | | RP | NEG | | | 1119631 | 1076772 | 1210518 | 866792 | 4.93E-01 | 0.09 | 0.57 |
| RP_POS_236_07162a11_595 | | 236.0716 | 11.60 | | RP | POS | | | 1.2E+08 | 4.3E+07 | 1.2E+08 | 4.1E+07 | 4.93E-01 | 0.09 | 0.66 |
| RP_POS_420_36856a11_549 | | 420.3686 | 11.55 | | RP | POS | | | 399372 | 99641.7 | 408117 | 87631.4 | 4.93E-01 | 0.09 | 0.47 |
| RP_POS_602_75659a10_391 | | 602.7566 | 10.39 | | RP | POS | | | 580122 | 291431 | 609495 | 330993 | 4.93E-01 | 0.09 | 0.68 |
| RP_POS_280_66397a10_314 | | 280.6640 | 10.31 | | RP | POS | | | 295852 | 158111 | 281689 | 144641 | 4.93E-01 | -0.09 | 0.58 |
| RP_POS_297_24182a9_966 | | 297.2418 | 9.97 | | RP | POS | | | 1606189 | 853854 | 1521719 | 939192 | 4.93E-01 | -0.09 | 0.42 |
| RP_POS_397_30994a11_144 | | 397.3099 | 11.14 | | RP | POS | | | 1957946 | 416085 | 1917397 | 445266 | 4.93E-01 | -0.09 | 0.66 |
| RP_POS_764_04541a12_783 | | 764.0454 | 12.78 | | RP | POS | | | 1352875 | 700797 | 1290601 | 630216 | 4.93E-01 | -0.09 | 0.65 |
| RP_POS_137_09605a8_914 | | 137.0961 | 8.91 | | RP | POS | | | 1156418 | 169599 | 1175004 | 220528 | 4.94E-01 | 0.1 | 0.6 |
| RP_NEG_718_14612a9_42 | | 718.1461 | 9.42 | | RP | NEG | | | 471354 | 32250.4 | 474367 | 32163.5 | 4.94E-01 | 0.09 | 0.58 |
| RP_POS_296_20691a6_92 | | 296.2069 | 6.92 | | RP | POS | | | 186120 | 36547.4 | 189414 | 34026.3 | 4.94E-01 | 0.09 | 0.72 |
| RP_POS_346_0405a15_514 | | 346.0405 | 15.51 | | RP | POS | | | 4573663 | 1186843 | 4695773 | 1397647 | 4.94E-01 | 0.09 | 0.74 |
| RP_NEG_568_36127a10_815 | LysoPC 18:0 (18:0/0:0) | 568.3613 | 10.82 | | RP | NEG | 283.2648(100), 242.0818(2) | 1 | 2.5E+07 | 6068713 | 2.6E+07 | 6557688 | 4.94E-01 | 0.09 | 0.78 |
| HILIC_NEG_134_89478a6_136 | | 134.8948 | 6.14 | | HILIC | NEG | | | 1.7E+07 | 4629636 | 1.7E+07 | 4984484 | 4.94E-01 | -0.09 | 0.73 |
| RP_POS_281_66241a10_926 | | 281.6624 | 10.93 | | RP | POS | | | 861977 | 401521 | 826138 | 366008 | 4.94E-01 | -0.09 | 0.92 |
| RP_POS_327_25336a11_905 | | 327.2534 | 11.91 | | RP | POS | | | 1364011 | 3280765 | 1076060 | 2895858 | 4.94E-01 | -0.09 | 0.58 |
| RP_POS_469_16763a6_804 | | 469.1676 | 6.80 | | RP | POS | | | 228006 | 87102.7 | 220101 | 82249 | 4.94E-01 | -0.09 | 1.03 |
| RP_POS_495_35785a10_389 | | 495.3579 | 10.39 | | RP | POS | | | 574168 | 1281841 | 485168 | 526148 | 4.94E-01 | -0.1 | 0.51 |
| HILIC_NEG_180_97379a5_463 | | 180.9738 | 5.46 | | HILIC | NEG | | | 457895 | 35770 | 461758 | 45593.9 | 4.95E-01 | 0.09 | 0.42 |
| RP_POS_267_23181a8_428 | | 267.2318 | 8.43 | | RP | POS | | | 130928 | 56976.3 | 136716 | 65909.6 | 4.95E-01 | 0.09 | 0.67 |
| RP_POS_371_10165a9_545 | | 371.1017 | 9.55 | | RP | POS | | | 147407 | 227847 | 169348 | 241445 | 4.95E-01 | 0.09 | 0.98 |
| RP_POS_486_36826a11_137 | | 486.3683 | 11.14 | | RP | POS | | | 563135 | 982058 | 649622 | 875200 | 4.95E-01 | 0.09 | 0.78 |
| RP_POS_587_74817a10_405 | | 587.7482 | 10.41 | | RP | POS | | | 827627 | 447767 | 873051 | 517996 | 4.95E-01 | 0.09 | 0.47 |
| RP_POS_133_0347a11_194 | | 133.0347 | 11.19 | | RP | POS | | | 2359445 | 986651 | 2277992 | 761997 | 4.95E-01 | -0.09 | 0.74 |
| RP_POS_290_16006a3_908 | | 290.1601 | 3.91 | | RP | POS | | | 193878 | 145680 | 181489 | 120394 | 4.95E-01 | -0.09 | 0.66 |
| RP_POS_513_8623a9_597 | | 513.8623 | 9.60 | | RP | POS | | | 982367 | 481141 | 932832 | 568162 | 4.95E-01 | -0.09 | 0.83 |
| RP_POS_713_09253a11_295 | | 713.0925 | 11.30 | | RP | POS | | | 2170854 | 6813648 | 1657068 | 4072101 | 4.95E-01 | -0.09 | 0.68 |
| RP_NEG_286_97659a6_375 | | 286.9766 | 6.38 | | RP | NEG | | | 1750985 | 329992 | 1780804 | 312327 | 4.96E-01 | 0.09 | 0.46 |
| RP_POS_305_022a2_481 | | 305.0220 | 2.48 | | RP | POS | | | 787456 | 300197 | 817241 | 334940 | 4.96E-01 | 0.09 | 0.54 |
| HILIC_POS_226_95134a4_336 | | 226.9513 | 4.34 | | HILIC | POS | | | 4.5E+08 | 2.5E+07 | 4.5E+08 | 2.6E+07 | 4.96E-01 | -0.09 | 0.85 |
| RP_NEG_313_23932a11_081 | | 313.2393 | 11.08 | | RP | NEG | | | 1270591 | 103409 | 1260506 | 112346 | 4.96E-01 | -0.09 | 0.74 |
| RP_NEG_929_1062a9_629 | | 929.1062 | 9.63 | | RP | NEG | | | 2716815 | 167710 | 2701265 | 166379 | 4.96E-01 | -0.09 | 0.66 |
| RP_POS_229_14299a8_675 | | 229.1430 | 8.68 | | RP | POS | | | 588652 | 140786 | 575807 | 135212 | 4.96E-01 | -0.09 | 0.68 |
| RP_POS_388_81647a15_417 | | 388.8165 | 15.42 | | RP | POS | | | 186992 | 203292 | 169434 | 175385 | 4.96E-01 | -0.09 | 0.73 |
| RP_POS_460_73462a4_893 | | 460.7346 | 4.89 | | RP | POS | | | 1217668 | 1490493 | 1073648 | 1589653 | 4.96E-01 | -0.09 | 0.77 |
| RP_POS_702_99158a10_377 | | 702.9916 | 10.38 | | RP | POS | | | 634210 | 292163 | 605120 | 328133 | 4.96E-01 | -0.09 | 0.63 |
| HILIC_NEG_108_90216a3_101 | | 108.9022 | 3.10 | | HILIC | NEG | | | 902736 | 113686 | 913878 | 124820 | 4.97E-01 | 0.09 | 0.48 |
| RP_POS_1171_33276a13_183 | | 1171.3328 | 13.18 | | RP | POS | | | 1873330 | 917436 | 1957022 | 884974 | 4.97E-01 | 0.09 | 0.81 |
| RP_POS_518_32068a10_357 | | 518.3207 | 10.36 | | RP | POS | | | 8.7E+07 | 1.3E+07 | 8.9E+07 | 1.5E+07 | 4.97E-01 | 0.09 | 0.93 |
| RP_POS_622_30646a7_495 | | 622.3065 | 7.50 | | RP | POS | | | 1502773 | 138973 | 1516567 | 155714 | 4.97E-01 | 0.09 | 0.74 |
| RP_NEG_322_80472a15_377 | | 322.8047 | 15.38 | | RP | NEG | | | 4912616 | 1090077 | 4811322 | 1091543 | 4.97E-01 | -0.09 | 1.16 |
| RP_POS_658_96667a10_422 | | 658.9667 | 10.42 | | RP | POS | | | 928442 | 476511 | 882199 | 515715 | 4.97E-01 | -0.09 | 0.8 |
| RP_POS_428_89188a8_928 | TDCPP | 428.8919 | 8.93 | | RP | POS | 98.9848(100), 208.9539(25) | 2 | 533884 | 73615.6 | 540817 | 75497.2 | 4.97E-01 | 0.09 | 0.61 |
| HILIC_POS_112_05094a1_468 | | 112.0509 | 1.47 | | HILIC | POS | | | 301117 | 66552.6 | 307825 | 76822.2 | 4.98E-01 | 0.09 | 0.5 |
| RP_NEG_183_01604a0_69 | | 183.0160 | 0.69 | | RP | NEG | | | 658159 | 109421 | 668678 | 116807 | 4.98E-01 | 0.09 | 0.41 |
| RP_POS_263_14609a0_562 | | 263.1461 | 0.56 | | RP | POS | | | 423662 | 247058 | 446310 | 242046 | 4.98E-01 | 0.09 | 0.48 |
| RP_POS_277_12836a6_515 | Triethylcitrate | 277.1284 | 6.52 | | RP | POS | 157.0503(100) | 2 | 1243895 | 168418 | 1260159 | 181006 | 4.98E-01 | 0.09 | 1.06 |
| RP_POS_1013_65387a10_49 | | 1013.6539 | 10.49 | | RP | POS | | | 3.8E+07 | 3764689 | 3.7E+07 | 4173167 | 4.98E-01 | -0.09 | 0.61 |
| RP_POS_281_24741a10_471 | | 281.2474 | 10.47 | | RP | POS | | | 2884698 | 1154681 | 2779667 | 1114529 | 4.98E-01 | -0.09 | 0.58 |
| RP_POS_317_16196a10_686 | | 317.1620 | 10.69 | | RP | POS | | | 1267610 | 286149 | 1242988 | 246969 | 4.98E-01 | -0.09 | 0.79 |
| RP_POS_319_16583a3_899 | | 319.1658 | 3.90 | | RP | POS | | | 519675 | 414106 | 485687 | 321755 | 4.98E-01 | -0.09 | 0.65 |
| RP_POS_369_38364a11_657 | | 369.3836 | 11.66 | | RP | POS | | | 2840947 | 4882625 | 2426947 | 4079713 | 4.98E-01 | -0.09 | 0.63 |
| RP_POS_510_35544a10_534 | | 510.3554 | 10.53 | | RP | POS | | | 4091685 | 1318035 | 3976191 | 1180305 | 4.98E-01 | -0.09 | 0.66 |
| RP_POS_531_3241a10_311 | | 531.3241 | 10.31 | | RP | POS | | | 1614330 | 608231 | 1559289 | 580811 | 4.98E-01 | -0.09 | 0.77 |
| RP_POS_231_10167a7_271 | | 231.1017 | 7.27 | | RP | POS | | | 318065 | 83746.3 | 325954 | 86754.4 | 4.99E-01 | 0.09 | 0.62 |
| RP_POS_320_16965a4_712 | | 320.1697 | 4.71 | | RP | POS | | | 615298 | 94623.7 | 624898 | 110900 | 4.99E-01 | 0.09 | 0.75 |
| RP_POS_383_21381a11_141 | | 383.2138 | 11.14 | | RP | POS | | | 326964 | 191941 | 346157 | 219485 | 4.99E-01 | 0.09 | 0.97 |
| RP_POS_406_25031a13_209 | | 406.2503 | 13.21 | | RP | POS | | | 331225 | 263697 | 358718 | 322775 | 4.99E-01 | 0.09 | 0.71 |
| RP_POS_365_10986a8_842 | | 365.1099 | 8.84 | | RP | POS | | | 340660 | 200528 | 322194 | 199202 | 4.99E-01 | -0.09 | 1.07 |
| RP_NEG_549_29016a9_945 | | 549.2902 | 9.95 | | RP | NEG | | | 574082 | 719335 | 665847 | 1178885 | 5.00E-01 | 0.1 | 0.57 |
| HILIC_POS_356_35172a3_055 | | 356.3517 | 3.06 | | HILIC | POS | | | 2083655 | 701345 | 2146575 | 665032 | 5.00E-01 | 0.09 | 0.73 |
| RP_NEG_804_57794a13_799 | | 804.5779 | 13.80 | | RP | NEG | | | 1.2E+08 | 2.3E+07 | 1.3E+08 | 3E+07 | 5.00E-01 | 0.09 | 0.56 |
| RP_POS_401_26831a9_953 | | 401.2683 | 9.95 | | RP | POS | | | 891345 | 87839.7 | 898996 | 78562.6 | 5.00E-01 | 0.09 | 0.65 |
| RP_POS_698_0943a11_661 | | 698.0943 | 11.66 | | RP | POS | | | 129010 | 66410.4 | 135613 | 75603.2 | 5.00E-01 | 0.09 | 0.77 |
| RP_POS_759_72937a11_521 | | 759.7294 | 11.52 | | RP | POS | | | 198766 | 107292 | 210578 | 143386 | 5.00E-01 | 0.09 | 0.46 |
| RP_POS_482_32358a10_085 | LysoPC 15:0 | 482.3236 | 10.09 | | RP | POS | 184.0743(100) | 1 | 5498219 | 1510695 | 5363946 | 1407675 | 5.00E-01 | -0.09 | 0.71 |
| HILIC_POS_106_0866a4_207 | | 106.0866 | 4.21 | | HILIC | POS | 88.0762(100), 106.0863(80) | | 1.1E+07 | 4988402 | 1.1E+07 | 4533215 | 5.00E-01 | -0.09 | 0.94 |
| HILIC_NEG_138_89003a3_204 | | 138.8900 | 3.20 | | HILIC | NEG | | | 3300347 | 180563 | 3283971 | 174856 | 5.00E-01 | -0.09 | 0.54 |
| HILIC_NEG_144_92348a7_997 | | 144.9235 | 8.00 | | HILIC | NEG | | | 2.2E+07 | 9831815 | 2.1E+07 | 9842148 | 5.00E-01 | -0.09 | 0.54 |
| RP_NEG_247_85045a0_628 | | 247.8505 | 0.63 | | RP | NEG | | | 206343 | 189250 | 189310 | 180344 | 5.00E-01 | -0.09 | 0.66 |
| RP_POS_185_12834a2_379 | | 185.1283 | 2.38 | | RP | POS | | | 2637841 | 1016715 | 2544268 | 1013166 | 5.00E-01 | -0.09 | 0.51 |
| RP_POS_356_35144a10_664 | | 356.3514 | 10.66 | | RP | POS | | | 205854 | 83683.8 | 197558 | 94795.3 | 5.00E-01 | -0.09 | 0.55 |
| HILIC_POS_134_08121a2_255 | N-Methylthreonine | 134.0812 | 2.26 | | HILIC | POS | 88.0761(100), 70.0658(80) | 2 | 5850432 | 2645603 | 5613850 | 2494556 | 5.00E-01 | -0.09 | 0.56 |
| RP_POS_311_18521a8_246 | | 311.1852 | 8.25 | | RP | POS | 209.1174(100), 273.1139(58) | | 7887782 | 316897 | 7916502 | 307650 | 5.01E-01 | 0.09 | 0.66 |
| HILIC_POS_550_38739a1_116 | | 550.3874 | 1.12 | | HILIC | POS | | | 571063 | 270823 | 595329 | 257934 | 5.01E-01 | 0.09 | 0.6 |
| RP_POS_514_22357a9_524 | | 514.2236 | 9.52 | | RP | POS | | | 407902 | 144343 | 421760 | 155752 | 5.01E-01 | 0.09 | 1 |
| RP_POS_147_09155a4_804 | 5,6-Dimethylbenzimidazole | 147.0916 | 4.80 | | RP | POS | 147.0921(100) | 1 | 1045896 | 195241 | 1065410 | 225000 | 5.01E-01 | 0.09 | 0.7 |
| HILIC_POS_172_09471a6_949 | | 172.0947 | 6.95 | | HILIC | POS | | | 514905 | 192537 | 497314 | 189628 | 5.01E-01 | -0.09 | 1.08 |
| HILIC_POS_430_91336a4_337 | | 430.9134 | 4.34 | | HILIC | POS | | | 2.6E+07 | 1576587 | 2.6E+07 | 1647388 | 5.01E-01 | -0.09 | 0.82 |
| RP_NEG_429_19553a7_52 | | 429.1955 | 7.52 | | RP | NEG | | | 265463 | 204077 | 247480 | 188024 | 5.01E-01 | -0.09 | 0.52 |
| RP_NEG_765_09973a8_639 | | 765.0997 | 8.64 | | RP | NEG | | | 410046 | 29448.7 | 407459 | 26960.7 | 5.01E-01 | -0.09 | 0.89 |
| RP_POS_187_13318a10_252 | | 187.1332 | 10.25 | | RP | POS | | | 2523423 | 226895 | 2500816 | 260225 | 5.01E-01 | -0.09 | 0.52 |
| RP_POS_647_94305a10_441 | | 647.9431 | 10.44 | | RP | POS | | | 1473453 | 627700 | 1414778 | 646084 | 5.01E-01 | -0.09 | 0.65 |
| RP_POS_480_00568a0_677 | | 480.0057 | 0.68 | | RP | POS | | | 359665 | 105030 | 369136 | 101582 | 5.02E-01 | 0.09 | 0.48 |
| RP_POS_619_33429a14_348 | | 619.3343 | 14.35 | | RP | POS | | | 762354 | 532956 | 817648 | 655713 | 5.02E-01 | 0.09 | 0.73 |
| HILIC_NEG_155_86568a6_296 | | 155.8657 | 6.30 | | HILIC | NEG | | | 1358923 | 632458 | 1302679 | 596334 | 5.02E-01 | -0.09 | 0.45 |
| HILIC_NEG_315_04178a7_351 | | 315.0418 | 7.35 | | HILIC | NEG | | | 1164703 | 140004 | 1150619 | 163573 | 5.02E-01 | -0.09 | 0.65 |
| HILIC_POS_490_22733a0_812 | | 490.2273 | 0.81 | | HILIC | POS | | | 209396 | 63514.4 | 203844 | 57650 | 5.02E-01 | -0.09 | 0.69 |
| RP_NEG_305_08203a7_592 | | 305.0820 | 7.59 | | RP | NEG | | | 540704 | 30260.2 | 537749 | 33756.7 | 5.02E-01 | -0.09 | 0.54 |
| RP_POS_327_16458a9_458 | | 327.1646 | 9.46 | | RP | POS | | | 445089 | 524829 | 399681 | 467579 | 5.02E-01 | -0.09 | 0.92 |
| RP_POS_437_43454a11_676 | | 437.4345 | 11.68 | | RP | POS | | | 245109 | 52015.4 | 240585 | 46684.7 | 5.02E-01 | -0.09 | 0.82 |
| RP_NEG_293_17963a11_276 | | 293.1796 | 11.28 | | RP | NEG | 293.1806(100) | | 174217 | 139601 | 186425 | 127662 | 5.03E-01 | 0.09 | 0.45 |
| RP_NEG_196_94402a15_49 | | 196.9440 | 15.49 | | RP | NEG | | | 231296 | 50431.7 | 235750 | 47104.6 | 5.03E-01 | 0.09 | 0.75 |
| RP_POS_169_04956a10_253 | | 169.0496 | 10.25 | | RP | POS | | | 1778587 | 873187 | 1855456 | 809396 | 5.03E-01 | 0.09 | 1.06 |
| RP_POS_369_11145a6_011 | | 369.1115 | 6.01 | | RP | POS | | | 8217583 | 6573693 | 8889989 | 7925408 | 5.03E-01 | 0.09 | 0.83 |
| RP_POS_480_24329a6_835 | | 480.2433 | 6.84 | | RP | POS | | | 703828 | 57192.2 | 708677 | 49047.6 | 5.03E-01 | 0.09 | 0.78 |
| RP_NEG_218_98589a5_511 | | 218.9859 | 5.51 | | RP | NEG | | | 1830596 | 593505 | 1777874 | 559154 | 5.03E-01 | -0.09 | 0.63 |
| RP_NEG_592_26733a10_28 | | 592.2673 | 10.28 | | RP | NEG | | | 1317202 | 306134 | 1287062 | 347346 | 5.03E-01 | -0.09 | 0.63 |
| RP_POS_213_10977a11_236 | | 213.1098 | 11.24 | | RP | POS | | | 1989551 | 1056953 | 1895703 | 994963 | 5.03E-01 | -0.09 | 0.75 |
| RP_POS_299_20087a10_002 | | 299.2009 | 10.00 | | RP | POS | | | 1635609 | 277380 | 1611375 | 253602 | 5.03E-01 | -0.09 | 0.77 |
| RP_POS_379_1265a9_207 | | 379.1265 | 9.21 | | RP | POS | | | 548784 | 260229 | 523579 | 287357 | 5.03E-01 | -0.09 | 0.76 |
| RP_POS_414_30002a8_51 | | 414.3000 | 8.51 | | RP | POS | | | 1730838 | 1633644 | 1561296 | 2016812 | 5.03E-01 | -0.09 | 0.44 |
| RP_POS_629_94403a10_292 | | 629.9440 | 10.29 | | RP | POS | | | 1652346 | 795523 | 1575876 | 866177 | 5.03E-01 | -0.09 | 0.63 |

**Supplemantary table 1**: The non-targeted metabolomics results ordered by the VIP value (p value < 0.0003 are shaded dark green, p value < 0.05 are shaded light green, d values below 0 are blue and above 0 are red)

| | | | | | | | | | Cases | | Controls | | Welch test | Cohen's | |
|---|---|---|---|---|---|---|---|---|---|---|---|---|---|---|---|
| Feature_ID | Metabolite_name | Average_Mz | Average_RT_min_ | HMDB ID | RP_HILIC | POS_NEG | Cureted_MSMS | ID_level | Mean | SD | Mean2 | SD3 | p | d | VIP |
| RP_POS_670_13806a11_596 | | 670.1381 | 11.60 | | RP | POS | | | 137097 | 64962.5 | 131090 | 66187.4 | 5.03E-01 | -0.09 | 0.84 |
| HILIC_POS_261_13113a4_333 | 2-(4-oxoquinazolin-3(4H)-yl)ethyl isobutyrate | 261.1311 | 4.33 | | HILIC | POS | 261.1324(100) | 2 | 330768 | 667590 | 274431 | 566235 | 5.03E-01 | -0.09 | 0.57 |
| HILIC_POS_394_31705a4_275 | | 394.3171 | 4.28 | | HILIC | POS | | | 418834 | 172991 | 433218 | 142641 | 5.04E-01 | 0.09 | 0.6 |
| RP_POS_1018_12793a10_488 | | 1018.1279 | 10.49 | | RP | POS | | | 2706030 | 582246 | 2760409 | 606573 | 5.04E-01 | 0.09 | 0.82 |
| RP_POS_172_13319a10_556 | | 172.1332 | 10.56 | | RP | POS | | | 1946633 | 822507 | 2022248 | 832639 | 5.04E-01 | 0.09 | 0.64 |
| RP_POS_502_32843a12_866 | | 502.3284 | 12.87 | | RP | POS | | | 318871 | 518512 | 371980 | 628141 | 5.04E-01 | 0.09 | 0.79 |
| RP_POS_599_11554a9_644 | | 599.1155 | 9.64 | | RP | POS | | | 1128588 | 81089.1 | 1135827 | 77670.4 | 5.04E-01 | 0.09 | 0.7 |
| RP_POS_668_29828a11_284 | | 668.2983 | 11.28 | | RP | POS | | | 106203 | 55887.8 | 111972 | 68712.9 | 5.04E-01 | 0.09 | 0.62 |
| RP_NEG_227_20215a10_685 | Myristic acid | 227.2022 | 10.69 | | RP | NEG | 227.2009(100) | 1 | 788528 | 336442 | 759804 | 293752 | 5.04E-01 | -0.09 | 1.06 |
| RP_NEG_275_16525a9_177 | | 275.1653 | 9.18 | | RP | NEG | | | 2169937 | 84011.5 | 2162534 | 78454.8 | 5.04E-01 | -0.09 | 0.78 |
| HILIC_POS_422_30237a5_947 | | 422.3024 | 5.95 | | HILIC | POS | | | 41867.3 | 9296.29 | 42667.1 | 8281.87 | 5.05E-01 | 0.09 | 0.77 |
| RP_POS_416_3157a5_596 | | 416.3157 | 5.60 | | RP | POS | | | 340980 | 391056 | 375410 | 366011 | 5.05E-01 | 0.09 | 0.56 |
| RP_NEG_153_86868a15_495 | | 153.8687 | 15.50 | | RP | NEG | | | 1130309 | 66126.9 | 1123147 | 87511.2 | 5.05E-01 | -0.09 | 0.5 |
| RP_NEG_700_30701a10_319 | | 700.3070 | 10.32 | | RP | NEG | | | 1983998 | 199373 | 1965092 | 213719 | 5.05E-01 | -0.09 | 0.77 |
| RP_POS_452_24493a8_543 | | 452.2449 | 8.54 | | RP | POS | | | 760473 | 151817 | 747613 | 131178 | 5.05E-01 | -0.09 | 0.79 |
| RP_POS_615_45551a11_703 | | 615.4555 | 11.70 | | RP | POS | | | 513177 | 425216 | 479649 | 310750 | 5.05E-01 | -0.09 | 0.42 |
| RP_NEG_436_28357a10_675 | | 436.2836 | 10.68 | | RP | NEG | | | 1973889 | 718033 | 2044202 | 814999 | 5.06E-01 | 0.09 | 0.76 |
| RP_POS_372_87274a15_428 | | 372.8727 | 15.43 | | RP | POS | | | 159346 | 124516 | 171198 | 134757 | 5.06E-01 | 0.09 | 0.51 |
| RP_POS_415_32043a9_308 | | 415.3204 | 9.31 | | RP | POS | | | 1058191 | 331116 | 1091381 | 390976 | 5.06E-01 | 0.09 | 0.83 |
| RP_POS_457_8222a10_291 | | 457.8222 | 10.29 | | RP | POS | | | 397709 | 217438 | 422869 | 317522 | 5.06E-01 | 0.09 | 1.11 |
| RP_POS_515_6983a10_042 | | 515.6983 | 10.04 | | RP | POS | | | 273899 | 167787 | 290292 | 190148 | 5.06E-01 | 0.09 | 0.66 |
| RP_POS_156_07686a0_677 | | 156.0769 | 0.68 | | RP | POS | | | 663005 | 653675 | 598684 | 748588 | 5.06E-01 | -0.09 | 1.07 |
| RP_POS_537_39484a15_12 | | 537.3948 | 15.12 | | RP | POS | | | 1009948 | 535956 | 957538 | 607686 | 5.06E-01 | -0.09 | 0.72 |
| RP_POS_679_9754a11_404 | | 679.9754 | 11.40 | | RP | POS | | | 192972 | 128484 | 182558 | 101364 | 5.06E-01 | -0.09 | 0.81 |
| HILIC_POS_272_18497a1_035 | | 272.1850 | 1.04 | | HILIC | POS | | | 346455 | 127174 | 358482 | 137034 | 5.07E-01 | 0.09 | 0.68 |
| RP_POS_279_19296a9_161 | | 279.1930 | 9.16 | | RP | POS | | | 1322830 | 145347 | 1336255 | 149896 | 5.07E-01 | 0.09 | 1.06 |
| RP_POS_469_34586a10_716 | | 469.3459 | 10.72 | | RP | POS | | | 958239 | 379802 | 993599 | 398294 | 5.07E-01 | 0.09 | 1.14 |
| RP_POS_663_93652a9_744 | | 663.9365 | 9.74 | | RP | POS | | | 318192 | 223739 | 343561 | 319318 | 5.07E-01 | 0.09 | 0.95 |
| HILIC_NEG_198_8813a7_733 | | 198.8813 | 7.73 | | HILIC | NEG | | | 264066 | 27488.1 | 261548 | 27915.7 | 5.07E-01 | -0.09 | 0.58 |
| RP_NEG_431_17422a11_236 | | 431.1742 | 11.24 | | RP | NEG | | | 487495 | 186677 | 470618 | 185017 | 5.07E-01 | -0.09 | 0.92 |
| RP_POS_340_35724a12_088 | | 340.3572 | 12.09 | | RP | POS | | | 2789174 | 4380508 | 2425679 | 3657360 | 5.07E-01 | -0.09 | 0.61 |
| HILIC_POS_102_12829a0_811 | 1-Hexylamine | 102.1283 | 0.81 | | HILIC | POS | 102.1284(100) | 2 | 1323623 | 355657 | 1293818 | 303505 | 5.07E-01 | -0.09 | 0.54 |
| HILIC_NEG_287_70544a1_41 | | 287.7054 | 1.41 | | HILIC | NEG | | | 523390 | 84014.4 | 530741 | 78731.9 | 5.08E-01 | 0.09 | 0.44 |
| RP_NEG_312_98959a7_496 | | 312.9896 | 7.50 | | RP | NEG | | | 1310285 | 191009 | 1328030 | 200332 | 5.08E-01 | 0.09 | 0.64 |
| RP_NEG_322_76089a15_402 | | 322.7609 | 15.40 | | RP | NEG | | | 2091398 | 427380 | 2131026 | 446789 | 5.08E-01 | 0.09 | 0.76 |
| RP_POS_172_97874a15_408 | | 172.9787 | 15.41 | | RP | POS | | | 9.5E+07 | 1.9E+07 | 9.6E+07 | 2.2E+07 | 5.08E-01 | 0.09 | 0.75 |
| RP_POS_418_19315a1_91 | | 418.1932 | 1.91 | | RP | POS | | | 594815 | 401945 | 635254 | 480875 | 5.08E-01 | 0.09 | 1.17 |
| RP_POS_432_33185a10_253 | | 432.3319 | 10.25 | | RP | POS | | | 2820168 | 1238357 | 2928221 | 1153618 | 5.08E-01 | 0.09 | 0.97 |
| HILIC_POS_106_08676a0_931 | | 106.0868 | 0.93 | | HILIC | POS | 88.0762(100), 106.0863(95) | | 1.1E+07 | 4987225 | 1.1E+07 | 3246162 | 5.08E-01 | -0.09 | 1.13 |
| HILIC_POS_148_10783a2_978 | | 148.1078 | 2.98 | | HILIC | POS | | | 397973 | 190852 | 380127 | 202572 | 5.08E-01 | -0.09 | 0.77 |
| RP_NEG_1043_15283a9_458 | | 1043.1528 | 9.46 | | RP | NEG | | | 1367536 | 175980 | 1352764 | 151260 | 5.08E-01 | -0.09 | 1.05 |
| RP_NEG_286_99271a15_464 | | 286.9927 | 15.46 | | RP | NEG | | | 572358 | 92057.4 | 564166 | 89099.7 | 5.08E-01 | -0.09 | 0.6 |
| RP_NEG_594_34076a10_778 | | 594.3408 | 10.78 | | RP | NEG | | | 592818 | 153778 | 578993 | 151984 | 5.08E-01 | -0.09 | 0.63 |
| RP_POS_253_25211a10_64 | | 253.2521 | 10.64 | | RP | POS | | | 1013993 | 155260 | 1000088 | 152259 | 5.08E-01 | -0.09 | 1.08 |
| RP_POS_487_83429a9_824 | | 487.8343 | 9.82 | | RP | POS | | | 2584139 | 1469790 | 2460890 | 1259158 | 5.08E-01 | -0.09 | 0.56 |
| RP_NEG_190_92856a15_449 | | 190.9286 | 15.45 | | RP | NEG | | | 1870227 | 374834 | 1905528 | 403018 | 5.09E-01 | 0.09 | 0.87 |
| RP_POS_247_99767a0_69 | | 247.9977 | 0.69 | | RP | POS | | | 258184 | 178377 | 275380 | 200157 | 5.09E-01 | 0.09 | 0.95 |
| RP_POS_335_15094a10_713 | | 335.1509 | 10.71 | | RP | POS | | | 1800485 | 1276384 | 1914278 | 1246537 | 5.09E-01 | 0.09 | 0.92 |
| HILIC_NEG_345_88361a3_264 | | 345.8836 | 3.26 | | HILIC | NEG | | | 854784 | 210366 | 833650 | 251727 | 5.09E-01 | -0.09 | 0.76 |
| RP_NEG_203_79887a15_491 | | 203.7989 | 15.49 | | RP | NEG | | | 462136 | 26285.8 | 459635 | 28823.6 | 5.09E-01 | -0.09 | 0.45 |
| RP_NEG_243_16052a8_449 | | 243.1605 | 8.45 | | RP | NEG | | | 2087546 | 200461 | 2069757 | 193580 | 5.09E-01 | -0.09 | 1.13 |
| RP_NEG_396_97144a9_431 | | 396.9714 | 9.43 | | RP | NEG | | | 385107 | 26528.5 | 387653 | 29668.8 | 5.10E-01 | 0.09 | 0.79 |
| RP_POS_271_06323a7_678 | Genistein | 271.0632 | 7.68 | | RP | POS | 271.0616(100) | 2 | 5993753 | 4804872 | 6460895 | 5481750 | 5.10E-01 | 0.09 | 0.78 |
| RP_NEG_272_95975a15_32 | | 272.9598 | 15.32 | | RP | NEG | | | 367387 | 79824.7 | 360510 | 73083.8 | 5.10E-01 | -0.09 | 1.13 |
| RP_NEG_626_32147a10_933 | | 626.3215 | 10.93 | | RP | NEG | | | 707383 | 99245.4 | 698594 | 95890.8 | 5.10E-01 | -0.09 | 0.58 |
| RP_POS_133_10658a0_684 | | 133.1066 | 0.68 | | RP | POS | | | 1.5E+07 | 3703931 | 1.5E+07 | 2836468 | 5.10E-01 | -0.09 | 0.56 |
| RP_POS_213_1097a11_127 | | 213.1097 | 11.13 | | RP | POS | | | 2069140 | 925793 | 1989943 | 838215 | 5.10E-01 | -0.09 | 0.63 |
| RP_POS_277_14142a7_695 | | 277.1414 | 7.70 | | RP | POS | | | 1E+07 | 738352 | 1E+07 | 679927 | 5.10E-01 | -0.09 | 0.76 |
| RP_POS_281_17236a9_531 | | 281.1724 | 9.53 | | RP | POS | | | 2.1E+07 | 5688133 | 2.1E+07 | 5589390 | 5.10E-01 | -0.09 | 1.04 |
| RP_POS_581_40405a9_151 | | 581.4041 | 9.15 | | RP | POS | | | 384265 | 268735 | 360849 | 251808 | 5.10E-01 | -0.09 | 0.79 |
| HILIC_POS_133_08617a0_6 | | 133.0862 | 0.60 | | HILIC | POS | | | 672307 | 86420.4 | 680104 | 86883.1 | 5.11E-01 | 0.09 | 0.68 |
| RP_POS_185_09622a7_215 | | 185.0962 | 7.22 | | RP | POS | | | 718843 | 54276.3 | 724082 | 61502.6 | 5.11E-01 | 0.09 | 0.61 |
| RP_POS_570_35529a10_532 | | 570.3553 | 10.53 | | RP | POS | | | 3902955 | 1597829 | 4045857 | 1583779 | 5.11E-01 | 0.09 | 0.63 |
| RP_POS_251_12753a8_17 | | 251.1275 | 8.17 | | RP | POS | | | 8553260 | 518791 | 8603617 | 595771 | 5.12E-01 | 0.09 | 1.03 |
| RP_POS_637_10675a10_104 | | 637.1068 | 10.10 | | RP | POS | | | 161314 | 96202.1 | 171227 | 121120 | 5.12E-01 | 0.09 | 0.76 |
| RP_POS_510_35596a10_598 | | 510.3560 | 10.60 | | RP | POS | | | 4730085 | 1153984 | 4625315 | 1181958 | 5.12E-01 | -0.09 | 0.8 |
| RP_POS_521_41846a13_685 | | 521.4185 | 13.69 | | RP | POS | | | 6723598 | 2126604 | 6539994 | 1973537 | 5.12E-01 | -0.09 | 0.75 |
| RP_POS_369_15186a6_417 | Jasminoside B | 369.1519 | 6.42 | | RP | POS | 369.1528(100) | 2 | 1745510 | 313321 | 1719341 | 272915 | 5.13E-01 | -0.09 | 1.17 |
| HILIC_NEG_128_03539a6_135 | | 128.0354 | 6.14 | | HILIC | NEG | | | 319364 | 47517.6 | 323436 | 43710.7 | 5.13E-01 | 0.09 | 0.81 |
| RP_NEG_807_17792a8_921 | | 807.1779 | 8.92 | | RP | NEG | | | 590738 | 67473.7 | 584726 | 66972.7 | 5.13E-01 | -0.09 | 0.51 |
| RP_POS_318_19089a5_933 | | 318.1909 | 5.93 | | RP | POS | | | 167932 | 97703.9 | 159336 | 94646.6 | 5.13E-01 | -0.09 | 0.44 |
| RP_POS_536_38965a10 | | 536.3897 | 10.00 | | RP | POS | | | 9493826 | 3368765 | 9153905 | 4133395 | 5.13E-01 | -0.09 | 0.59 |
| RP_NEG_167_02087a0_915 | | 167.0209 | 0.92 | | RP | NEG | 167.0204(100), 124.0147(52) | | 7358941 | 1385730 | 7484771 | 1429419 | 5.14E-01 | 0.09 | 0.4 |
| HILIC_NEG_192_98953a1_359 | | 192.9895 | 1.36 | | HILIC | NEG | | | 517327 | 199520 | 535142 | 199378 | 5.14E-01 | 0.09 | 0.51 |
| HILIC_POS_158_964a5_464 | | 158.9640 | 5.46 | | HILIC | POS | | | 412971 | 256291 | 436436 | 268162 | 5.14E-01 | 0.09 | 0.52 |
| RP_POS_472_31152a9_613 | | 472.3115 | 9.61 | | RP | POS | | | 134691 | 119604 | 146576 | 143568 | 5.14E-01 | 0.09 | 0.93 |
| RP_POS_536_3736a10_817 | | 536.3736 | 10.82 | | RP | POS | | | 2312552 | 730501 | 2374931 | 670781 | 5.14E-01 | 0.09 | 0.6 |
| RP_POS_681_50397a10_252 | | 681.5040 | 10.25 | | RP | POS | | | 909765 | 460865 | 955529 | 552058 | 5.14E-01 | 0.09 | 0.89 |
| RP_NEG_314_7659a0_644 | | 314.7659 | 0.64 | | RP | NEG | | | 431533 | 143471 | 420617 | 100707 | 5.14E-01 | -0.09 | 0.44 |
| RP_NEG_533_4552a11_141 | | 533.4552 | 11.14 | | RP | NEG | | | 436373 | 171171 | 421853 | 155265 | 5.14E-01 | -0.09 | 1.28 |
| RP_POS_219_174a10_384 | | 219.1740 | 10.38 | | RP | POS | | | 1240109 | 367993 | 1208071 | 351657 | 5.14E-01 | -0.09 | 0.71 |
| RP_POS_482_36099a10_777 | LysoPC O-16:0 | 482.3610 | 10.78 | | RP | POS | 104.1079(100), 184.0747(25) | 1 | 2.6E+07 | 7071317 | 2.6E+07 | 7048585 | 5.14E-01 | -0.09 | 0.64 |
| HILIC_POS_142_08641a2_878 | | 142.0864 | 2.88 | | HILIC | POS | | | 285319 | 1415772 | 193824 | 490175 | 5.14E-01 | -0.1 | 0.41 |
| HILIC_NEG_200_8586a6_205 | | 200.8586 | 6.21 | | HILIC | NEG | | | 1379386 | 221743 | 1399234 | 224015 | 5.15E-01 | 0.09 | 0.67 |
| RP_POS_149_00279a0_625 | | 149.0028 | 0.63 | | RP | POS | | | 398053 | 159692 | 412608 | 166494 | 5.15E-01 | 0.09 | 0.68 |
| RP_POS_204_52904a13_104 | | 204.5290 | 13.10 | | RP | POS | | | 460181 | 224099 | 481468 | 250527 | 5.15E-01 | 0.09 | 0.66 |
| RP_POS_991_67316a10_487 | | 991.6732 | 10.49 | | RP | POS | | | 3.8E+08 | 9.1E+07 | 3.9E+08 | 9.8E+07 | 5.15E-01 | 0.09 | 0.91 |
| RP_POS_329_24753a10_896 | Docosahexaenoic acid (DHA, 22:6) | 329.2475 | 10.90 | | RP | POS | 329.248(100), 81.0705(25) | 1 | 1.2E+07 | 5661985 | 1.1E+07 | 5647717 | 5.15E-01 | -0.09 | 0.98 |
| RP_NEG_947_17737a9_704 | | 947.1774 | 9.70 | | RP | NEG | | | 2211769 | 155935 | 2198402 | 145036 | 5.15E-01 | -0.09 | 0.77 |
| HILIC_NEG_157_90083a4_315 | | 157.9008 | 4.32 | | HILIC | NEG | | | 1003661 | 250957 | 1026352 | 258923 | 5.16E-01 | 0.09 | 0.75 |
| HILIC_POS_371_23978a0_936 | | 371.2398 | 0.94 | | HILIC | POS | | | 2221748 | 179949 | 2244958 | 313403 | 5.16E-01 | 0.09 | 0.71 |
| HILIC_NEG_157_8625a3_099 | | 157.8625 | 3.10 | | HILIC | NEG | | | 1526768 | 98618.7 | 1518441 | 89194 | 5.16E-01 | -0.09 | 0.49 |
| HILIC_POS_414_33636a0_618 | | 414.3364 | 0.62 | | HILIC | POS | | | 117549 | 131794 | 106250 | 123309 | 5.16E-01 | -0.09 | 0.47 |
| RP_POS_199_16936a9_927 | | 199.1694 | 9.93 | | RP | POS | | | 8068996 | 3919405 | 7710769 | 4117915 | 5.16E-01 | -0.09 | 0.57 |
| RP_POS_397_41559a11_654 | | 397.4156 | 11.65 | | RP | POS | | | 704969 | 1123086 | 613810 | 933999 | 5.16E-01 | -0.09 | 0.57 |
| HILIC_NEG_194_90591a3_115 | | 194.9059 | 3.12 | | HILIC | NEG | | | 2098726 | 290662 | 2121731 | 230469 | 5.17E-01 | 0.09 | 0.66 |
| RP_NEG_264_00238a0_667 | | 264.0024 | 0.67 | | RP | NEG | | | 128538 | 15017.6 | 129826 | 14124.3 | 5.17E-01 | 0.09 | 0.48 |
| RP_NEG_351_01285a2_485 | | 351.0129 | 2.49 | | RP | NEG | | | 2105233 | 353062 | 2136330 | 349234 | 5.17E-01 | 0.09 | 0.6 |
| RP_POS_188_12822a4_606 | | 188.1282 | 4.61 | | RP | POS | | | 327762 | 101700 | 337686 | 119809 | 5.17E-01 | 0.09 | 0.64 |
| RP_POS_261_10989a7_239 | | 261.1099 | 7.24 | | RP | POS | | | 1439056 | 1220375 | 1330625 | 1227679 | 5.17E-01 | -0.09 | 0.83 |
| RP_POS_490_34244a9_81 | | 490.3424 | 9.81 | | RP | POS | | | 1224681 | 708034 | 1165451 | 632026 | 5.17E-01 | -0.09 | 0.63 |
| RP_POS_338_34158a11_658 | Erucamide | 338.3416 | 11.66 | | RP | POS | 338.3408(100), 83.0863(70) | 2 | 1.4E+08 | 2.3E+08 | 1.2E+08 | 1.9E+08 | 5.17E-01 | -0.09 | 0.6 |
| HILIC_NEG_132_92375a5_468 | | 132.9238 | 5.47 | | HILIC | NEG | | | 2410685 | 375644 | 2444352 | 385690 | 5.18E-01 | 0.09 | 0.58 |
| HILIC_NEG_354_99481a7_35 | | 354.9948 | 7.35 | | HILIC | NEG | | | 744437 | 35860.6 | 747621 | 36198.5 | 5.18E-01 | 0.09 | 0.61 |
| HILIC_POS_185_03255a6_136 | | 185.0326 | 6.14 | | HILIC | POS | | | 5425823 | 472342 | 5468960 | 500425 | 5.18E-01 | 0.09 | 0.76 |
| RP_NEG_184_95744a15_644 | | 184.9574 | 15.64 | | RP | NEG | | | 1404579 | 641181 | 1463051 | 678809 | 5.18E-01 | 0.09 | 0.42 |
| RP_NEG_397_22339a9_365 | | 397.2234 | 9.37 | | RP | NEG | | | 573788 | 323265 | 597637 | 213537 | 5.18E-01 | 0.09 | 0.6 |
| RP_NEG_444_1062a11_593 | | 444.1062 | 11.59 | | RP | NEG | | | 205034 | 61535.3 | 211647 | 84484.5 | 5.18E-01 | 0.09 | 0.66 |
| HILIC_POS_132_0032a4_337 | | 132.0032 | 4.34 | | HILIC | POS | | | 2.6E+07 | 1554204 | 2.6E+07 | 1681986 | 5.18E-01 | -0.09 | 1.03 |
| RP_NEG_146_90086a15_399 | | 146.9009 | 15.40 | | RP | NEG | | | 1547956 | 610638 | 1491467 | 662729 | 5.18E-01 | -0.09 | 0.54 |
| RP_POS_349_05905a9_036 | | 349.0591 | 9.04 | | RP | POS | | | 2490886 | 395307 | 2456544 | 381829 | 5.18E-01 | -0.09 | 0.66 |
| RP_POS_465_3786a10_693 | | 465.3786 | 10.69 | | RP | POS | | | 1671589 | 303399 | 1645446 | 288351 | 5.18E-01 | -0.09 | 0.93 |
| RP_POS_641_51105a15_057 | | 641.5111 | 15.06 | | RP | POS | | | 1.3E+07 | 4684423 | 1.2E+07 | 4510360 | 5.18E-01 | -0.09 | 0.74 |
| RP_POS_806_96417a10_31 | | 806.9642 | 10.31 | | RP | POS | | | 798603 | 500329 | 755940 | 467494 | 5.18E-01 | -0.09 | 0.81 |
| HILIC_NEG_215_07353a0_941 | | 215.0735 | 0.94 | | HILIC | NEG | | | 456528 | 405138 | 492129 | 402718 | 5.19E-01 | 0.09 | 0.65 |
| RP_NEG_748_52899a13_495 | | 748.5290 | 13.50 | | RP | NEG | | | 3089927 | 898359 | 3172705 | 971853 | 5.19E-01 | 0.09 | 0.74 |
| RP_POS_181_98694a0_628 | | 181.9869 | 0.63 | | RP | POS | | | 1566349 | 555737 | 1618945 | 628164 | 5.19E-01 | 0.09 | 0.53 |
| RP_POS_249_10062a15_37 | | 249.1006 | 15.37 | | RP | POS | | | 763509 | 399513 | 799464 | 413704 | 5.19E-01 | 0.09 | 0.76 |
| RP_POS_615_45929a11_144 | | 615.4593 | 11.14 | | RP | POS | | | 1229508 | 1615501 | 1349661 | 1101279 | 5.19E-01 | 0.09 | 0.43 |
| RP_POS_699_6496a11_285 | | 699.6496 | 11.29 | | RP | POS | | | 126601 | 82021.4 | 133874 | 82923 | 5.19E-01 | 0.09 | 0.47 |
| RP_POS_721_8999a11_269 | | 721.8999 | 11.27 | | RP | POS | | | 120715 | 58513.1 | 127063 | 81522.4 | 5.19E-01 | 0.09 | 1.01 |
| RP_POS_795_06818a13_247 | | 795.0682 | 13.25 | | RP | POS | | | 4809319 | 4567805 | 5222948 | 4796446 | 5.19E-01 | 0.09 | 0.97 |
| RP_POS_802_21466a9_349 | | 802.2147 | 9.35 | | RP | POS | | | 979005 | 116663 | 989256 | 115925 | 5.19E-01 | 0.09 | 0.97 |
| RP_NEG_540_33069a10_358 | LysoPC 16:0 (16:0/0:0) | 540.3307 | 10.36 | | RP | NEG | 255.2335(100) | 1 | 1.3E+08 | 2.2E+07 | 1.3E+08 | 2.4E+07 | 5.19E-01 | 0.09 | 0.94 |
| HILIC_POS_363_03564a6_309 | | 363.0356 | 6.31 | | HILIC | POS | | | 573723 | 248513 | 552230 | 239384 | 5.19E-01 | -0.09 | 0.43 |
| RP_NEG_333_65347a15_502 | | 333.6535 | 15.50 | | RP | NEG | | | 178342 | 31844.5 | 175833 | 25263.1 | 5.19E-01 | -0.09 | 0.54 |
| RP_POS_227_0787a2_484 | | 227.0787 | 2.48 | | RP | POS | | | 919281 | 335210 | 891628 | 293552 | 5.19E-01 | -0.09 | 0.47 |
| RP_NEG_271_0704a2_482 | | 271.0704 | 2.48 | | RP | NEG | | | 767853 | 137054 | 779967 | 138377 | 5.20E-01 | 0.09 | 0.51 |
| RP_POS_475_01529a10_686 | | 475.0153 | 10.69 | | RP | POS | | | 673291 | 1281547 | 817822 | 1892502 | 5.20E-01 | 0.09 | 0.75 |
| RP_POS_649_30286a10_603 | | 649.3029 | 10.60 | | RP | POS | | | 391372 | 123916 | 402382 | 126265 | 5.20E-01 | 0.09 | 0.4 |
| RP_POS_816_04486a12_766 | | 816.0449 | 12.77 | | RP | POS | | | 919265 | 635925 | 975391 | 640438 | 5.20E-01 | 0.09 | 0.81 |
| RP_NEG_524_27808a10_281 | | 524.2781 | 10.28 | | RP | NEG | | | 2.1E+07 | 5669347 | 2E+07 | 6104525 | 5.20E-01 | -0.09 | 0.62 |
| HILIC_NEG_234_81956a4_075 | | 234.8196 | 4.08 | | HILIC | NEG | | | 407121 | 88648.3 | 414141 | 71807.3 | 5.21E-01 | 0.09 | 0.75 |
| HILIC_POS_368_21985a5_66 | | 368.2199 | 5.66 | | HILIC | POS | | | 498312 | 329106 | 528774 | 361986 | 5.21E-01 | 0.09 | 0.44 |
| HILIC_POS_368_21997a7_101 | | 368.2200 | 7.10 | | HILIC | POS | | | 163608 | 31488.2 | 166351 | 31022.8 | 5.21E-01 | 0.09 | 0.6 |
| RP_POS_627_10101a10_243 | | 627.1010 | 10.24 | | RP | POS | | | 377144 | 200452 | 396084 | 227469 | 5.21E-01 | 0.09 | 0.52 |
| HILIC_POS_391_05307a3_146 | | 391.0531 | 3.15 | | HILIC | POS | | | 341928 | 54055.4 | 336750 | 62858.2 | 5.21E-01 | -0.09 | 0.89 |
| RP_POS_130_15929a3_174 | | 130.1593 | 3.17 | | RP | POS | | | 896221 | 246958 | 875714 | 221821 | 5.21E-01 | -0.09 | 0.79 |
| RP_POS_777_6936a7_731 | | 777.6936 | 7.73 | | RP | POS | | | 2370713 | 542406 | 2323293 | 537636 | 5.21E-01 | -0.09 | 0.57 |
| HILIC_POS_173_02966a4_329 | | 173.0297 | 4.33 | | HILIC | POS | | | 293705 | 176884 | 308855 | 169419 | 5.22E-01 | 0.09 | 0.52 |
| RP_NEG_245_00162a7_238 | | 245.0016 | 7.24 | | RP | NEG | | | 908611 | 101390 | 917369 | 98639.1 | 5.22E-01 | 0.09 | 0.71 |
| RP_POS_389_33682a10_249 | | 389.3368 | 10.25 | | RP | POS | | | 441866 | 148418 | 455125 | 154093 | 5.22E-01 | 0.09 | 1.05 |
| RP_POS_245_11494a7_577 | | 245.1149 | 7.58 | | RP | POS | 245.1144(100) | | 3260275 | 206165 | 3243684 | 174481 | 5.22E-01 | -0.09 | 1.01 |
| HILIC_NEG_253_79454a10_746 | | 253.7945 | 10.75 | | HILIC | NEG | | | 1008696 | 504341 | 964180 | 511575 | 5.22E-01 | -0.09 | 0.73 |
| RP_POS_201_11221a6_476 | | 201.1122 | 6.48 | | RP | POS | | | 2006017 | 1100521 | 1907860 | 1136809 | 5.22E-01 | -0.09 | 0.56 |
| RP_POS_289_63019a10_311 | | 289.6302 | 10.31 | | RP | POS | | | 2004029 | 507549 | 1960167 | 495536 | 5.22E-01 | -0.09 | 0.67 |
| RP_POS_335_07959a8_842 | | 335.0796 | 8.84 | | RP | POS | | | 504416 | 215499 | 486646 | 191930 | 5.22E-01 | -0.09 | 0.72 |
| HILIC_POS_276_14438a5_105 | Glutarylcarnitine | 276.1444 | 5.11 | | HILIC | POS | 85.02918100), 276.1437(90) | 2 | 218709 | 207192 | 237098 | 211876 | 5.22E-01 | 0.09 | 0.52 |
| RP_NEG_325_18497a10_695 | | 325.1850 | 10.70 | | RP | NEG | 325.1832(100), 183.0126(20) | | 3847920 | 2203636 | 4069471 | 2783358 | 5.23E-01 | 0.09 | 0.56 |
| RP_NEG_483_19574a10_962 | | 483.1957 | 10.96 | | RP | NEG | | | 228132 | 116212 | 237758 | 104719 | 5.23E-01 | 0.09 | 1 |
| RP_POS_373_19434a8_842 | | 373.1943 | 8.84 | | RP | POS | | | 99421.2 | 98800 | 109538 | 128363 | 5.23E-01 | 0.09 | 0.83 |
| RP_POS_551_55029a14_425 | | 551.5503 | 14.43 | | RP | POS | | | 353169 | 189452 | 369366 | 181558 | 5.23E-01 | 0.09 | 0.57 |
| RP_POS_321_315a11_656 | | 321.3150 | 11.66 | | RP | POS | | | 1977696 | 3626718 | 1688348 | 3013816 | 5.23E-01 | -0.09 | 0.6 |
| RP_POS_369_33655a12_457 | | 369.3366 | 12.46 | | RP | POS | | | 935817 | 996219 | 855803 | 840101 | 5.23E-01 | -0.09 | 0.63 |
| RP_NEG_261_85129a15_563 | | 261.8513 | 15.56 | | RP | NEG | | | 1045818 | 380826 | 1080154 | 405215 | 5.24E-01 | 0.09 | 0.43 |
| RP_POS_270_27899a10_943 | | 270.2790 | 10.94 | | RP | POS | | | 617362 | 224459 | 641152 | 307501 | 5.24E-01 | 0.09 | 0.82 |
| RP_POS_583_74628a10_277 | | 583.7463 | 10.28 | | RP | POS | | | 262149 | 178686 | 279152 | 207923 | 5.24E-01 | 0.09 | 0.52 |
| RP_POS_803_55859a13_453 | | 803.5586 | 13.45 | | RP | POS | | | 855514 | 645270 | 913764 | 687372 | 5.24E-01 | 0.09 | 1.13 |
| RP_NEG_325_18417a11_139 | | 325.1842 | 11.14 | | RP | NEG | 325.1825(100), 183.0122(10) | | 9191806 | 6049673 | 8691987 | 5448265 | 5.24E-01 | -0.09 | 0.87 |
| RP_POS_159_09193a2_884 | | 159.0919 | 2.88 | | RP | POS | | | 5844455 | 1025315 | 5932291 | 998138 | 5.25E-01 | 0.09 | 0.73 |
| RP_POS_452_39545a10_93 | | 452.3955 | 10.93 | | RP | POS | | | 7941399 | 786496 | 8006082 | 706729 | 5.25E-01 | 0.09 | 0.86 |
| RP_NEG_144_92339a13_291 | | 144.9234 | 13.29 | | RP | NEG | | | 614378 | 189158 | 595250 | 242665 | 5.25E-01 | -0.09 | 0.67 |

**Supplemantary table 1**: The non-targeted metabolomics results ordered by the VIP value (p value < 0.0003 are shaded dark green, p value < 0.05 are shaded light green, d values below 0 are blue and above 0 are red)

| | | | | | | | | | Cases | | Controls | | Welch test | Cohen's | |
|---|---|---|---|---|---|---|---|---|---|---|---|---|---|---|---|
| Feature_ID | Metabolite_name | Average_Mz | Average_RT_min_ | HMDB ID | RP_HILIC | POS_NEG | Cureted_MSMS | ID_level | Mean | SD | Mean2 | SD3 | p | d | VIP |
| RP_NEG_411_08975a0_679 | | 411.0898 | 0.68 | | RP | NEG | | | 446618 | 94796.1 | 439259 | 74936.1 | 5.25E-01 | -0.09 | 0.7 |
| RP_POS_1122_27637a9_791 | | 1122.2764 | 9.79 | | RP | POS | | | 1864294 | 255837 | 1840228 | 293760 | 5.25E-01 | -0.09 | 0.58 |
| RP_NEG_176_82623a15_474 | | 176.8262 | 15.47 | | RP | NEG | | | 718691 | 297554 | 745865 | 326532 | 5.26E-01 | 0.09 | 0.47 |
| RP_POS_198_1852a15_482 | | 198.1852 | 15.48 | | RP | POS | | | 3.1E+07 | 6127655 | 3.1E+07 | 7058504 | 5.26E-01 | 0.09 | 0.38 |
| HILIC_POS_172_09421a5_509 | | 172.0942 | 5.51 | | HILIC | POS | | | 3149019 | 2299852 | 2949028 | 2307431 | 5.26E-01 | -0.09 | 0.65 |
| RP_POS_201_12726a9_1 | | 201.1273 | 9.10 | | RP | POS | | | 908374 | 88459.8 | 900366 | 95433.3 | 5.26E-01 | -0.09 | 0.67 |
| RP_POS_304_26031a11_595 | | 304.2603 | 11.60 | | RP | POS | | | 3820271 | 801303 | 3749042 | 836096 | 5.26E-01 | -0.09 | 0.89 |
| RP_POS_722_52948a13_695 | | 722.5295 | 13.70 | | RP | POS | | | 1.1E+08 | 3.1E+07 | 1.1E+08 | 3E+07 | 5.26E-01 | -0.09 | 1.02 |
| RP_POS_765_05353a12_855 | | 765.0535 | 12.86 | | RP | POS | | | 772532 | 630543 | 722511 | 526965 | 5.26E-01 | -0.09 | 0.68 |
| HILIC_NEG_100_93351a4_428 | | 100.9335 | 4.43 | | HILIC | NEG | | | 2.2E+07 | 2756015 | 2.2E+07 | 4134669 | 5.27E-01 | 0.09 | 0.48 |
| RP_POS_256_8949a15_398 | | 256.8949 | 15.40 | | RP | POS | | | 6746993 | 1329477 | 6852801 | 1125341 | 5.27E-01 | 0.09 | 0.87 |
| RP_POS_550_38721a10_896 | | 550.3872 | 10.90 | | RP | POS | | | 1147243 | 321800 | 1176291 | 347205 | 5.27E-01 | 0.09 | 0.6 |
| HILIC_NEG_408_99136a1_271 | | 408.9914 | 1.27 | | HILIC | NEG | | | 423435 | 178991 | 407600 | 185938 | 5.27E-01 | -0.09 | 0.66 |
| RP_NEG_691_1261a9_753 | | 691.1261 | 9.75 | | RP | NEG | | | 970687 | 62190.8 | 965015 | 68149.1 | 5.27E-01 | -0.09 | 0.5 |
| RP_POS_312_32593a11_589 | | 312.3259 | 11.59 | | RP | POS | | | 1025143 | 311769 | 999602 | 280014 | 5.27E-01 | -0.09 | 0.47 |
| RP_POS_669_27063a11_374 | | 669.2706 | 11.37 | | RP | POS | | | 146234 | 89653.5 | 138578 | 87398.5 | 5.27E-01 | -0.09 | 0.7 |
| HILIC_NEG_326_98816a3_049 | | 326.9882 | 3.05 | | HILIC | NEG | | | 450293 | 75632.6 | 457222 | 83855.4 | 5.28E-01 | 0.09 | 0.76 |
| RP_NEG_594_81421a15_358 | | 594.8142 | 15.36 | | RP | NEG | | | 657615 | 66559.6 | 663191 | 62748.1 | 5.28E-01 | 0.09 | 0.64 |
| RP_POS_662_96173a11_373 | | 662.9617 | 11.37 | | RP | POS | | | 300617 | 175782 | 316609 | 192727 | 5.28E-01 | 0.09 | 0.64 |
| RP_POS_139_11177a7_327 | | 139.1118 | 7.33 | | RP | POS | | | 1132730 | 176417 | 1118055 | 164623 | 5.28E-01 | -0.09 | 0.48 |
| RP_POS_204_88904a15_396 | | 204.8890 | 15.40 | | RP | POS | | | 188745 | 104647 | 179178 | 115753 | 5.28E-01 | -0.09 | 0.76 |
| RP_POS_217_14304a9_466 | | 217.1430 | 9.47 | | RP | POS | | | 9605778 | 7472944 | 8966185 | 7339943 | 5.28E-01 | -0.09 | 0.77 |
| RP_POS_239_1617a8_599 | | 239.1617 | 8.60 | | RP | POS | | | 7633114 | 1947105 | 7474690 | 1734311 | 5.28E-01 | -0.09 | 1.04 |
| RP_POS_318_26324a9_458 | | 318.2632 | 9.46 | | RP | POS | | | 321189 | 307088 | 295628 | 286021 | 5.28E-01 | -0.09 | 0.89 |
| RP_POS_647_42328a11_824 | | 647.4233 | 11.82 | | RP | POS | | | 192839 | 121924 | 183679 | 90867.1 | 5.28E-01 | -0.09 | 0.59 |
| RP_NEG_508_79068a15_408 | | 508.7907 | 15.41 | | RP | NEG | | | 248526 | 78142.9 | 256415 | 101739 | 5.29E-01 | 0.09 | 0.59 |
| HILIC_NEG_144_92354a7_399 | | 144.9235 | 7.40 | | HILIC | NEG | | | 2.4E+07 | 7145690 | 2.3E+07 | 6869469 | 5.29E-01 | -0.09 | 0.5 |
| RP_POS_807_16931a9_539 | | 807.1693 | 9.54 | | RP | POS | | | 1128769 | 99830.7 | 1120357 | 95821.1 | 5.29E-01 | -0.09 | 0.66 |
| HILIC_NEG_99_9257a4_399 | | 99.9257 | 4.40 | | HILIC | NEG | | | 1920512 | 363026 | 1949511 | 315174 | 5.30E-01 | 0.09 | 0.78 |
| RP_POS_168_01369a0_723 | | 168.0137 | 0.72 | | RP | POS | | | 2666397 | 1316061 | 2783915 | 1408402 | 5.30E-01 | 0.09 | 0.45 |
| RP_POS_501_02423a10_054 | | 501.0242 | 10.05 | | RP | POS | | | 226735 | 120746 | 240497 | 187349 | 5.30E-01 | 0.09 | 0.51 |
| RP_NEG_541_19873a11_359 | | 541.1987 | 11.36 | | RP | NEG | | | 138893 | 40731.2 | 135443 | 39762.6 | 5.30E-01 | -0.09 | 1.21 |
| RP_POS_445_19852a8_733 | | 445.1985 | 8.73 | | RP | POS | | | 355120 | 227612 | 335199 | 235488 | 5.30E-01 | -0.09 | 0.95 |
| HILIC_POS_285_04663a3_147 | | 285.0466 | 3.15 | | HILIC | POS | | | 278390 | 118021 | 287846 | 103674 | 5.31E-01 | 0.09 | 0.63 |
| RP_POS_324_32596a11_188 | | 324.3260 | 11.19 | | RP | POS | | | 2497297 | 2682892 | 2740215 | 2951711 | 5.31E-01 | 0.09 | 0.94 |
| RP_POS_411_09424a9_612 | | 411.0942 | 9.61 | | RP | POS | | | 356750 | 65665.9 | 362611 | 70643.8 | 5.31E-01 | 0.09 | 0.8 |
| RP_POS_421_2558a10_519 | | 421.2558 | 10.52 | | RP | POS | | | 553759 | 376114 | 588971 | 437264 | 5.31E-01 | 0.09 | 1.12 |
| RP_POS_687_64508a11_392 | | 687.6451 | 11.39 | | RP | POS | | | 119109 | 72664.5 | 127398 | 113005 | 5.31E-01 | 0.09 | 0.67 |
| RP_NEG_269_21204a10_094 | | 269.2120 | 10.09 | | RP | NEG | | | 1105325 | 153412 | 1092389 | 148903 | 5.31E-01 | -0.09 | 0.95 |
| RP_POS_125_03907a15_526 | | 125.0391 | 15.53 | | RP | POS | | | 8845178 | 2092826 | 8659362 | 2225619 | 5.31E-01 | -0.09 | 0.72 |
| RP_POS_414_77203a13_004 | | 414.7720 | 13.00 | | RP | POS | | | 2803210 | 2380858 | 2624373 | 1797575 | 5.31E-01 | -0.09 | 1.09 |
| RP_POS_807_56982a13_353 | | 807.5698 | 13.35 | | RP | POS | | | 4.9E+07 | 2.2E+07 | 4.8E+07 | 2.1E+07 | 5.31E-01 | -0.09 | 0.75 |
| HILIC_POS_315_08002a7_35 | Altenuene | 315.0800 | 7.35 | | HILIC | POS | 315.0799(100) | 2 | 177169 | 35118.3 | 180590 | 43608.2 | 5.31E-01 | 0.09 | 0.61 |
| RP_POS_316_21155a2_812 | | 316.2116 | 2.81 | | RP | POS | | | 66193.8 | 76805.1 | 87361.6 | 328259 | 5.32E-01 | 0.1 | 0.74 |
| RP_NEG_549_28943a9_741 | | 549.2894 | 9.74 | | RP | NEG | | | 732068 | 763237 | 808632 | 993187 | 5.32E-01 | 0.09 | 0.62 |
| RP_POS_238_06738a14_243 | | 238.0674 | 14.24 | | RP | POS | | | 4041108 | 2449917 | 4261261 | 2681594 | 5.32E-01 | 0.09 | 0.7 |
| RP_POS_383_20386a9_574 | | 383.2039 | 9.57 | | RP | POS | 383.206(100) | | 1169703 | 296534 | 1151040 | 115885 | 5.32E-01 | -0.09 | 0.39 |
| RP_POS_599_45062a15_162 | | 599.4506 | 15.16 | | RP | POS | | | 401546 | 360787 | 371915 | 334313 | 5.32E-01 | -0.09 | 0.89 |
| RP_POS_728_52008a12_902 | | 728.5201 | 12.90 | | RP | POS | | | 4962579 | 2665188 | 4743734 | 2470372 | 5.32E-01 | -0.09 | 0.46 |
| RP_POS_834_61401a10_12 | | 834.6140 | 10.12 | | RP | POS | | | 1906871 | 865293 | 1833399 | 854079 | 5.32E-01 | -0.09 | 0.65 |
| RP_POS_882_49347a13_106 | | 882.4935 | 13.11 | | RP | POS | | | 542675 | 396030 | 512860 | 303789 | 5.32E-01 | -0.09 | 0.55 |
| HILIC_POS_249_06075a4_135 | | 249.0608 | 4.14 | | HILIC | POS | | | 1597524 | 1830122 | 1750748 | 1770193 | 5.33E-01 | 0.09 | 0.63 |
| RP_NEG_366_75391a15_434 | | 366.7539 | 15.43 | | RP | NEG | | | 1025597 | 354835 | 1058328 | 406821 | 5.33E-01 | 0.09 | 0.65 |
| RP_NEG_416_17914a0_686 | | 416.1791 | 0.69 | | RP | NEG | | | 380739 | 247980 | 403542 | 283062 | 5.33E-01 | 0.09 | 1.24 |
| RP_NEG_475_83121a15_399 | | 475.8312 | 15.40 | | RP | NEG | | | 489966 | 114270 | 499695 | 113685 | 5.33E-01 | 0.09 | 0.69 |
| RP_POS_556_38104a9_774 | | 556.3810 | 9.77 | | RP | POS | | | 885828 | 442782 | 926726 | 508785 | 5.33E-01 | 0.09 | 0.61 |
| HILIC_POS_464_38647a0_616 | | 464.3865 | 0.62 | | HILIC | POS | | | 74857.5 | 135216 | 85196.2 | 108120 | 5.33E-01 | 0.08 | 0.78 |
| RP_NEG_296_06369a0_677 | | 296.0637 | 0.68 | | RP | NEG | | | 206205 | 45003.7 | 202585 | 40102.4 | 5.33E-01 | -0.09 | 0.48 |
| RP_NEG_638_4209a11_142 | | 638.4209 | 11.14 | | RP | NEG | | | 272761 | 48225.4 | 268532 | 50774.1 | 5.33E-01 | -0.09 | 1.12 |
| RP_NEG_650_27655a10_311 | | 650.2766 | 10.31 | | RP | NEG | | | 2457425 | 571384 | 2410035 | 541521 | 5.33E-01 | -0.09 | 0.77 |
| RP_POS_159_01323a15_732 | | 159.0132 | 15.73 | | RP | POS | | | 3.4E+07 | 1.6E+07 | 3.3E+07 | 1.9E+07 | 5.33E-01 | -0.09 | 0.7 |
| RP_POS_236_0715a8_984 | | 236.0715 | 8.98 | | RP | POS | | | 7.7E+07 | 2.9E+07 | 7.4E+07 | 2.9E+07 | 5.33E-01 | -0.09 | 0.44 |
| RP_POS_393_71964a9_207 | | 393.7196 | 9.21 | | RP | POS | | | 482561 | 386447 | 447627 | 427368 | 5.33E-01 | -0.09 | 0.68 |
| HILIC_POS_149_05986a3_835 | Cinnamic acid | 149.0599 | 3.84 | | HILIC | POS | 103.0549(100), 121.065(30) | 2 | 552776 | 3798899 | 302551 | 1908139 | 5.33E-01 | -0.09 | 0.51 |
| HILIC_POS_86_09698a3_032 | | 86.0970 | 3.03 | | HILIC | POS | 86.0974(100) | | 1528831 | 555408 | 1578710 | 611841 | 5.34E-01 | 0.09 | 0.66 |
| RP_POS_131_04921a1_411 | | 131.0492 | 1.41 | | RP | POS | | | 497739 | 288153 | 524951 | 343807 | 5.34E-01 | 0.09 | 0.72 |
| RP_POS_411_21411a10_25 | | 411.2141 | 10.25 | | RP | POS | | | 1401445 | 642599 | 1452846 | 568489 | 5.34E-01 | 0.08 | 0.97 |
| RP_POS_731_74017a11_896 | | 731.7402 | 11.90 | | RP | POS | | | 145957 | 33660.4 | 148693 | 30818.7 | 5.34E-01 | 0.08 | 0.89 |
| RP_POS_614_93878a10_468 | | 614.9388 | 10.47 | | RP | POS | | | 918892 | 485553 | 871069 | 619093 | 5.34E-01 | -0.09 | 0.71 |
| RP_POS_188_07083a2_482 | | 188.0708 | 2.48 | | RP | POS | | | 1.9E+08 | 3.6E+07 | 2E+08 | 3.8E+07 | 5.35E-01 | 0.09 | 0.74 |
| HILIC_POS_173_07819a5_442 | | 173.0782 | 5.44 | | HILIC | POS | | | 374526 | 194109 | 391203 | 198142 | 5.35E-01 | 0.09 | 0.5 |
| RP_NEG_223_84885a15_484 | | 223.8489 | 15.48 | | RP | NEG | | | 217744 | 37772.8 | 220890 | 36389.3 | 5.35E-01 | 0.08 | 0.6 |
| HILIC_POS_510_35495a1_21 | LysoPC 17:0 | 510.3550 | 1.21 | | HILIC | POS | 104.1079(100), 184.0733(95) | 2 | 4227779 | 1403537 | 4107703 | 1427438 | 5.35E-01 | -0.08 | 0.83 |
| RP_NEG_327_25467a11_232 | | 327.2547 | 11.23 | | RP | NEG | | | 744184 | 1686160 | 641071 | 561437 | 5.35E-01 | -0.09 | 0.42 |
| RP_NEG_134_89478a2_892 | | 134.8948 | 2.89 | | RP | NEG | | | 2214719 | 496527 | 2258814 | 539752 | 5.36E-01 | 0.09 | 0.65 |
| RP_NEG_993_11847a9_668 | | 993.1185 | 9.67 | | RP | NEG | | | 2109342 | 214587 | 2128548 | 236940 | 5.36E-01 | 0.09 | 0.62 |
| RP_POS_331_11108a0_546 | | 331.1111 | 0.55 | | RP | POS | | | 358829 | 100426 | 369469 | 143742 | 5.36E-01 | 0.09 | 0.54 |
| RP_NEG_145_93079a15_574 | | 145.9308 | 15.57 | | RP | NEG | | | 309466 | 159337 | 323354 | 168064 | 5.36E-01 | 0.08 | 0.61 |
| HILIC_POS_235_16528a3_609 | | 235.1653 | 3.61 | | HILIC | POS | | | 3025374 | 2071146 | 2805384 | 2973334 | 5.36E-01 | -0.09 | 0.57 |
| RP_POS_184_07336a10_357 | | 184.0734 | 10.36 | | RP | POS | 184.0744(100), 86.0974(50) | | 3487612 | 710285 | 3550677 | 774950 | 5.37E-01 | 0.08 | 0.87 |
| RP_NEG_276_16089a9_767 | | 276.1609 | 9.77 | | RP | NEG | | | 928446 | 34946.6 | 931214 | 30844.9 | 5.37E-01 | 0.08 | 0.55 |
| HILIC_POS_150_1124a4_208 | | 150.1124 | 4.21 | | HILIC | POS | 150.1126(100), 132.1026(40) | | 1550039 | 1008655 | 1469371 | 907326 | 5.37E-01 | -0.08 | 0.79 |
| HILIC_NEG_200_99821a0_935 | | 200.9982 | 0.94 | | HILIC | NEG | | | 527083 | 102048 | 518964 | 90637.9 | 5.37E-01 | -0.08 | 0.67 |
| RP_NEG_691_12659a9_196 | | 691.1266 | 9.20 | | RP | NEG | | | 2695245 | 184015 | 2681160 | 150811 | 5.37E-01 | -0.08 | 0.62 |
| RP_POS_267_12067a4_083 | | 267.1207 | 4.08 | | RP | POS | | | 231037 | 187889 | 215507 | 180658 | 5.37E-01 | -0.08 | 0.99 |
| RP_POS_419_7066a7_534 | | 419.7066 | 7.53 | | RP | POS | | | 3095894 | 480962 | 3056442 | 455630 | 5.37E-01 | -0.08 | 1.06 |
| HILIC_POS_209_09193a4_112 | L-kynurenine | 209.0919 | 4.11 | | HILIC | POS | 94.0662(100), 192.0651(70) | 2 | 2220154 | 442957 | 2183872 | 419036 | 5.37E-01 | -0.08 | 0.55 |
| HILIC_POS_248_22215a0_683 | | 248.2222 | 0.68 | | HILIC | POS | | | 198034 | 361585 | 228381 | 359053 | 5.38E-01 | 0.08 | 0.52 |
| RP_POS_271_2269a8_751 | | 271.2269 | 8.75 | | RP | POS | | | 223065 | 88180.8 | 231118 | 101346 | 5.38E-01 | 0.08 | 0.76 |
| RP_POS_328_2478a7_348 | | 328.2478 | 7.35 | | RP | POS | | | 6355003 | 2560425 | 6586005 | 2881155 | 5.38E-01 | 0.08 | 0.47 |
| HILIC_NEG_112_98573a4_352 | | 112.9857 | 4.35 | | HILIC | NEG | 68.9949(100), 112.9854(4) | | 6.6E+08 | 2.7E+07 | 6.6E+08 | 2.6E+07 | 5.38E-01 | -0.08 | 0.85 |
| HILIC_POS_156_04224a5_442 | | 156.0422 | 5.44 | | HILIC | POS | | | 1065307 | 326496 | 1038794 | 304629 | 5.38E-01 | -0.08 | 0.44 |
| RP_POS_343_1427a9_459 | | 343.1427 | 9.46 | | RP | POS | | | 840978 | 1058801 | 757031 | 939318 | 5.38E-01 | -0.08 | 0.54 |
| HILIC_POS_356_00934a7_425 | | 356.0093 | 7.43 | | HILIC | POS | | | 815079 | 46848.2 | 819458 | 56180.5 | 5.39E-01 | 0.09 | 0.64 |
| RP_POS_257_11469a8_576 | | 257.1147 | 8.58 | | RP | POS | 257.1168(100) | | 885364 | 90378.7 | 892825 | 87571.6 | 5.39E-01 | 0.08 | 0.57 |
| HILIC_NEG_134_89476a0_945 | | 134.8948 | 0.95 | | HILIC | NEG | | | 3.7E+07 | 6319951 | 3.8E+07 | 6255330 | 5.39E-01 | 0.08 | 0.42 |
| RP_NEG_352_01108a2_884 | | 352.0111 | 2.88 | | RP | NEG | | | 3023113 | 450505 | 3060708 | 445083 | 5.39E-01 | 0.08 | 0.62 |
| RP_POS_290_63821a10_609 | | 290.6382 | 10.61 | | RP | POS | | | 1701251 | 411146 | 1737860 | 454706 | 5.39E-01 | 0.08 | 0.9 |
| RP_POS_331_05298a15_533 | | 331.0530 | 15.53 | | RP | POS | | | 1E+07 | 2505318 | 1E+07 | 2561053 | 5.39E-01 | 0.08 | 0.8 |
| RP_POS_900_56879a13_96 | | 900.5688 | 13.96 | | RP | POS | | | 7721160 | 2395107 | 7941498 | 2796046 | 5.39E-01 | 0.08 | 0.63 |
| RP_POS_644_49475a10_453 | | 644.4948 | 10.45 | | RP | POS | | | 2589925 | 309281 | 2563329 | 322237 | 5.39E-01 | -0.08 | 0.71 |
| RP_POS_821_37701a7_533 | | 821.3770 | 7.53 | | RP | POS | | | 580285 | 161789 | 566589 | 164118 | 5.39E-01 | -0.08 | 1.11 |
| RP_POS_129_05478a9_459 | 3-Methylglutaric acid | 129.0548 | 9.46 | | RP | POS | 55.0554(100), 83.0502(50),101.0608(40) | 1 | 2604652 | 2193670 | 2418407 | 2231618 | 5.39E-01 | -0.08 | 0.68 |
| RP_POS_343_33893a13_741 | | 343.3389 | 13.74 | | RP | POS | | | 1.3E+07 | 6944068 | 1.4E+07 | 7711516 | 5.40E-01 | 0.08 | 0.69 |
| RP_POS_604_46179a14_341 | | 604.4618 | 14.34 | | RP | POS | | | 950273 | 923737 | 1026002 | 885467 | 5.40E-01 | 0.08 | 1.09 |
| RP_POS_811_60364a13_739 | | 811.6036 | 13.74 | | RP | POS | | | 2E+07 | 1.1E+07 | 2.1E+07 | 1.1E+07 | 5.40E-01 | 0.08 | 0.92 |
| HILIC_POS_399_30716a4_283 | | 399.3072 | 4.28 | | HILIC | POS | | | 60202.2 | 61357.7 | 55983.1 | 38511.9 | 5.40E-01 | -0.08 | 0.42 |
| RP_NEG_316_94778a11_574 | | 316.9478 | 11.57 | | RP | NEG | | | 1347388 | 747864 | 1284684 | 749189 | 5.40E-01 | -0.08 | 0.94 |
| RP_POS_500_27728a10_02 | | 500.2773 | 10.02 | | RP | POS | | | 4894993 | 2910028 | 4682942 | 2161697 | 5.40E-01 | -0.08 | 0.45 |
| RP_POS_809_08484a13_782 | | 809.0848 | 13.78 | | RP | POS | | | 2991181 | 2141220 | 2842792 | 1379617 | 5.40E-01 | -0.08 | 0.72 |
| RP_POS_199_99014a15_486 | | 199.9901 | 15.49 | | RP | POS | | | 757571 | 80744.4 | 764433 | 83286.9 | 5.41E-01 | 0.08 | 0.8 |
| RP_POS_321_20944a11_237 | | 321.2094 | 11.24 | | RP | POS | | | 398626 | 287891 | 422059 | 274030 | 5.41E-01 | 0.08 | 0.97 |
| RP_POS_607_42963a10_516 | | 607.4296 | 10.52 | | RP | POS | | | 401914 | 264292 | 424087 | 266416 | 5.41E-01 | 0.08 | 0.55 |
| RP_POS_759_13342a9_483 | | 759.1334 | 9.48 | | RP | POS | | | 665336 | 143432 | 677760 | 152688 | 5.41E-01 | 0.08 | 0.71 |
| RP_POS_794_56506a13_491 | | 794.5651 | 13.49 | | RP | POS | | | 6345969 | 5099265 | 6798991 | 5677266 | 5.41E-01 | 0.08 | 0.79 |
| RP_POS_482_32443a10_902 | LysoPE 18:0 | 482.3244 | 10.90 | | RP | POS | 341.3058(100), 57.071(10) | 1 | 2.5E+07 | 5646155 | 2.5E+07 | 6541624 | 5.41E-01 | 0.08 | 0.78 |
| HILIC_NEG_259_81604a3_22 | | 259.8160 | 3.22 | | HILIC | NEG | | | 1085526 | 193109 | 1068642 | 208863 | 5.41E-01 | -0.08 | 0.5 |
| RP_POS_267_1199a3_649 | | 267.1199 | 3.65 | | RP | POS | | | 387511 | 541146 | 339217 | 606268 | 5.41E-01 | -0.08 | 0.64 |
| RP_POS_403_79449a9_636 | | 403.7945 | 9.64 | | RP | POS | | | 999194 | 589740 | 954783 | 474956 | 5.41E-01 | -0.08 | 0.65 |
| RP_POS_419_31586a11_843 | Hexadecyl ferulate | 419.3159 | 11.84 | | RP | POS | 149.0231(100), 71.0863(70) | 1 | 9226509 | 5631655 | 9702263 | 5728111 | 5.41E-01 | 0.08 | 0.55 |
| HILIC_POS_159_07634a6_381 | | 159.0763 | 6.38 | | HILIC | POS | 70.0061(100), 113.0717(60) | | 3144109 | 830264 | 3213373 | 830258 | 5.42E-01 | 0.08 | 0.52 |
| HILIC_POS_680_4801a1_352 | | 680.4801 | 1.35 | | HILIC | POS | | | 294800 | 121801 | 305042 | 123815 | 5.42E-01 | 0.08 | 0.51 |
| RP_NEG_957_20526a9_63 | | 957.2053 | 9.63 | | RP | NEG | | | 3817404 | 282154 | 3839980 | 260342 | 5.42E-01 | 0.08 | 0.93 |
| RP_POS_124_92998a15_49 | | 124.9300 | 15.49 | | RP | POS | | | 506212 | 47567.9 | 509858 | 40078.1 | 5.42E-01 | 0.08 | 0.91 |
| RP_POS_163_11157a14_251 | | 163.1116 | 14.25 | | RP | POS | | | 7053170 | 3716932 | 7350179 | 3419994 | 5.42E-01 | 0.08 | 0.41 |
| RP_POS_236_0715a11_143 | | 236.0715 | 11.14 | | RP | POS | | | 1.7E+08 | 3.8E+07 | 1.7E+08 | 3.6E+07 | 5.42E-01 | -0.08 | 0.76 |
| RP_POS_409_75412a12_732 | | 409.7541 | 12.73 | | RP | POS | | | 1853016 | 1042735 | 1767566 | 1007903 | 5.42E-01 | -0.08 | 0.54 |
| RP_NEG_541_33594a11_265 | | 541.3359 | 11.27 | | RP | NEG | | | 280612 | 555615 | 330328 | 630050 | 5.43E-01 | 0.08 | 0.46 |
| RP_POS_721_98041a10_062 | | 721.9804 | 10.06 | | RP | POS | | | 501290 | 331144 | 530702 | 370949 | 5.43E-01 | 0.08 | 0.97 |
| HILIC_NEG_188_98158a7_734 | | 188.9816 | 7.73 | | HILIC | NEG | | | 200893 | 20534.1 | 199285 | 18255.1 | 5.43E-01 | -0.08 | 0.77 |
| HILIC_NEG_343_99487a6_755 | | 343.9949 | 6.76 | | HILIC | NEG | | | 1396828 | 670545 | 1350556 | 435042 | 5.43E-01 | -0.08 | 0.5 |
| RP_NEG_318_70422a15_496 | | 318.7042 | 15.50 | | RP | NEG | | | 145639 | 43703.1 | 141426 | 55910.8 | 5.43E-01 | -0.08 | 0.75 |
| RP_NEG_529_46362a15_162 | | 529.4636 | 15.16 | | RP | NEG | | | 1902935 | 499676 | 1862256 | 480451 | 5.43E-01 | -0.08 | 0.97 |
| RP_POS_396_18991a10_936 | | 396.1899 | 10.94 | | RP | POS | | | 314711 | 147708 | 301815 | 160736 | 5.43E-01 | -0.08 | 0.51 |
| RP_POS_621_41974a9_434 | | 621.4197 | 9.43 | | RP | POS | | | 440571 | 270533 | 416940 | 294479 | 5.43E-01 | -0.08 | 0.71 |
| RP_POS_233_11497a6_598 | Dihydrokawain | 233.1150 | 6.60 | | RP | POS | 233.1136(100) | 2 | 8484223 | 2536632 | 8291208 | 2129270 | 5.44E-01 | -0.08 | 1.24 |
| RP_NEG_529_46344a13_282 | | 529.4634 | 13.28 | | RP | NEG | | | 242918 | 122875 | 252141 | 100217 | 5.44E-01 | 0.08 | 1.03 |
| RP_POS_470_2594a5_861 | | 470.2594 | 5.86 | | RP | POS | | | 338914 | 40320.9 | 342217 | 39411.4 | 5.44E-01 | 0.08 | 0.54 |
| RP_POS_621_3103a11_439 | | 621.3103 | 11.44 | | RP | POS | | | 710460 | 552043 | 759590 | 623471 | 5.44E-01 | 0.08 | 0.66 |
| RP_POS_680_87842a11_424 | | 680.8784 | 11.42 | | RP | POS | | | 148808 | 86252.1 | 156829 | 104468 | 5.44E-01 | 0.08 | 0.65 |
| RP_NEG_311_16855a11_014 | | 311.1686 | 11.01 | | RP | NEG | 311.1701(100) | | 2011161 | 1008110 | 1934512 | 843523 | 5.44E-01 | -0.08 | 0.71 |
| HILIC_NEG_227_96107a4_25 | | 227.9611 | 4.25 | | HILIC | NEG | | | 1877888 | 589619 | 1829789 | 571724 | 5.44E-01 | -0.08 | 0.66 |
| RP_NEG_157_01195a0_696 | | 157.0120 | 0.70 | | RP | NEG | | | 516902 | 91530.4 | 509426 | 88849.9 | 5.44E-01 | -0.08 | 0.73 |
| RP_NEG_610_38977a11_142 | | 610.3898 | 11.14 | | RP | NEG | | | 2257207 | 209065 | 2239261 | 222322 | 5.44E-01 | -0.08 | 0.83 |
| RP_POS_336_32593a11_357 | | 336.3259 | 11.36 | | RP | POS | | | 2778976 | 4995617 | 2403378 | 4079387 | 5.44E-01 | -0.08 | 0.57 |
| RP_POS_809_65204a14_066 | | 809.6520 | 14.07 | | RP | POS | | | 1584646 | 1343373 | 1482750 | 1120487 | 5.44E-01 | -0.08 | 0.75 |
| RP_POS_934_63879a11_433 | | 934.6388 | 11.43 | | RP | POS | | | 318247 | 472237 | 282216 | 399986 | 5.44E-01 | -0.08 | 0.68 |
| RP_NEG_745_55231a12_36 | | 745.5523 | 12.36 | | RP | NEG | | | 2.1E+07 | 3162128 | 2.2E+07 | 3283800 | 5.45E-01 | 0.08 | 0.51 |
| HILIC_NEG_136_89168a1_352 | | 136.8917 | 1.35 | | HILIC | NEG | | | 6173450 | 1663714 | 6312146 | 1682144 | 5.45E-01 | 0.08 | 0.46 |
| HILIC_NEG_99_92567a0_937 | | 99.9257 | 0.94 | | HILIC | NEG | | | 1851576 | 192589 | 1868731 | 218945 | 5.45E-01 | 0.08 | 0.58 |
| RP_POS_558_4032a9_983 | | 558.4032 | 9.98 | | RP | POS | | | 8462451 | 3264325 | 8735424 | 3322783 | 5.45E-01 | 0.08 | 0.69 |
| RP_POS_726_50421a12_434 | | 726.5042 | 12.43 | | RP | POS | | | 1083329 | 1116721 | 1185463 | 1326628 | 5.45E-01 | 0.08 | 0.7 |
| HILIC_POS_148_10803a4_853 | | 148.1080 | 4.85 | | HILIC | POS | | | 2166154 | 2625240 | 1963743 | 2281353 | 5.45E-01 | -0.08 | 0.62 |
| RP_POS_340_01974a15_438 | | 340.0197 | 15.44 | | RP | POS | | | 1328133 | 526451 | 1280301 | 617816 | 5.45E-01 | -0.08 | 0.66 |
| RP_POS_525_73999a5_485 | | 525.7400 | 5.49 | | RP | POS | | | 1887227 | 987083 | 1793104 | 1248758 | 5.45E-01 | -0.08 | 0.51 |
| RP_POS_547_38477a15_06 | | 547.3848 | 15.06 | | RP | POS | | | 4967257 | 1669753 | 5113240 | 1843364 | 5.46E-01 | 0.08 | 0.71 |
| RP_POS_675_13892a11_5 | | 675.1389 | 11.50 | | RP | POS | | | 150566 | 83187.7 | 157448 | 83304.2 | 5.46E-01 | 0.08 | 0.77 |
| RP_POS_743_99304a10_051 | | 743.9930 | 10.05 | | RP | POS | | | 316419 | 206557 | 335136 | 242403 | 5.46E-01 | 0.08 | 0.75 |
| RP_POS_774_5639a12_366 | PC 16:0_HODE | 774.5639 | 12.37 | | RP | POS | 184.0746(100) | 1 | 2287248 | 1049684 | 2217411 | 622593 | 5.46E-01 | -0.08 | 1 |
| HILIC_NEG_196_88533a7_735 | | 196.8853 | 7.74 | | HILIC | NEG | | | 6513612 | 539925 | 6465915 | 606611 | 5.46E-01 | -0.08 | 0.65 |
| HILIC_NEG_276_99146a3_05 | | 276.9915 | 3.05 | | HILIC | NEG | | | 609172 | 82326.6 | 603537 | 53458.1 | 5.46E-01 | -0.08 | 0.56 |
| HILIC_POS_355_03827a4_864 | | 355.0383 | 4.86 | | HILIC | POS | | | 124747 | 85355 | 118293 | 71382.6 | 5.46E-01 | -0.08 | 0.57 |

**Supplemantary table 1**: The non-targeted metabolomics results ordered by the VIP value (p value < 0.0003 are shaded dark green, p value < 0.05 are shaded light green, d values below 0 are blue and above 0 are red)

| | | | | | | | | | Cases | | Controls | | Welch test | Cohen's | |
|---|---|---|---|---|---|---|---|---|---|---|---|---|---|---|---|
| Feature_ID | Metabolite_name | Average_Mz | Average_RT_min_ | HMDB ID | RP_HILIC | POS_NEG | Cureted_MSMS | ID_level | Mean | SD | Mean2 | SD3 | p | d | VIP |
| RP_NEG_506_50412a15_5 | | 506.5041 | 15.50 | | RP | NEG | | | 381908 | 44577.1 | 378424 | 40058.1 | 5.46E-01 | -0.08 | 0.97 |
| RP_NEG_508_7442a4_664 | | 508.7442 | 4.66 | | RP | NEG | | | 343918 | 182950 | 328687 | 185568 | 5.46E-01 | -0.08 | 1.23 |
| RP_NEG_624_30408a10_611 | | 624.3041 | 10.61 | | RP | NEG | | | 695477 | 112503 | 685477 | 128106 | 5.46E-01 | -0.08 | 0.57 |
| RP_POS_392_33591a11_135 | | 392.3359 | 11.14 | | RP | POS | | | 233659 | 59741.1 | 228295 | 68926.4 | 5.46E-01 | -0.08 | 0.69 |
| RP_POS_613_45306a9_729 | | 613.4531 | 9.73 | | RP | POS | | | 635622 | 379903 | 603626 | 393797 | 5.46E-01 | -0.08 | 0.63 |
| RP_POS_846_52197a12_816 | | 846.5220 | 12.82 | | RP | POS | | | 5225284 | 3121833 | 4973865 | 2983169 | 5.46E-01 | -0.08 | 0.61 |
| RP_POS_518_32458a10_015 | LysoPC 18:3 | 518.3246 | 10.02 | | RP | POS | 184.0743(100), 104.1077(70) | 2 | 3E+07 | 1.3E+07 | 2.9E+07 | 1.4E+07 | 5.46E-01 | -0.08 | 0.6 |
| RP_POS_255_05869a2_538 | | 255.0587 | 2.54 | | RP | POS | 255.0567(100) | | 1150469 | 502715 | 1194355 | 555912 | 5.47E-01 | 0.08 | 0.4 |
| RP_NEG_268_98328a6_816 | | 268.9833 | 6.82 | | RP | NEG | | | 2986786 | 146105 | 2998066 | 128514 | 5.47E-01 | 0.08 | 0.63 |
| RP_NEG_397_33322a11_999 | | 397.3332 | 12.00 | | RP | NEG | | | 258692 | 37812.7 | 262152 | 45317.8 | 5.47E-01 | 0.08 | 0.95 |
| RP_POS_297_0824a10_694 | | 297.0824 | 10.69 | | RP | POS | | | 121528 | 49115.1 | 125351 | 44054.3 | 5.47E-01 | 0.08 | 0.69 |
| RP_POS_341_09888a6_225 | Agarotetrol | 341.0989 | 6.23 | | RP | POS | 341.1927(100) | 2 | 296551 | 84907 | 289849 | 78177.7 | 5.47E-01 | -0.08 | 1.05 |
| RP_POS_303_23172a10_691 | Abietic acid | 303.2317 | 10.69 | | RP | POS | 303.2323(100), 73.029(20) | 2 | 1862510 | 796956 | 1925573 | 736809 | 5.47E-01 | 0.08 | 0.94 |
| HILIC_POS_358_25937a0_818 | | 358.2594 | 0.82 | | HILIC | POS | | | 236339 | 226874 | 220217 | 164983 | 5.47E-01 | -0.08 | 0.48 |
| RP_NEG_487_11551a8_18 | | 487.1155 | 8.18 | | RP | NEG | | | 198899 | 280621 | 177151 | 248934 | 5.47E-01 | -0.08 | 0.52 |
| RP_NEG_570_76233a15_406 | | 570.7623 | 15.41 | | RP | NEG | | | 232596 | 79108.1 | 226213 | 76060.5 | 5.47E-01 | -0.08 | 0.66 |
| RP_POS_289_64911a10_93 | | 289.6491 | 10.93 | | RP | POS | | | 2101659 | 713019 | 2041539 | 742680 | 5.47E-01 | -0.08 | 0.95 |
| RP_POS_516_85492a10_022 | | 516.8549 | 10.02 | | RP | POS | | | 3262774 | 1501830 | 3138815 | 1508388 | 5.47E-01 | -0.08 | 0.56 |
| RP_POS_706_53741a12_903 | | 706.5374 | 12.90 | | RP | POS | | | 2E+07 | 9758572 | 1.9E+07 | 9551125 | 5.47E-01 | -0.08 | 0.46 |
| RP_POS_195_12312a2_586 | Octyl acetate | 195.1231 | 2.59 | | RP | POS | 195.0871(100) | 2 | 2423922 | 246124 | 2443660 | 233963 | 5.47E-01 | 0.08 | 0.49 |
| RP_POS_766_5719a13_776 | | 766.5719 | 13.78 | | RP | POS | | | 2908758 | 2749865 | 3206616 | 4224046 | 5.48E-01 | 0.09 | 0.55 |
| HILIC_POS_90_97717a5_462 | | 90.9772 | 5.46 | | HILIC | POS | | | 1458689 | 157700 | 1471827 | 161935 | 5.48E-01 | 0.08 | 0.49 |
| RP_POS_183_976a15_429 | | 183.9760 | 15.43 | | RP | POS | | | 322516 | 202116 | 337496 | 163652 | 5.48E-01 | 0.08 | 0.43 |
| RP_POS_288_28995a8_112 | | 288.2900 | 8.11 | | RP | POS | | | 1457857 | 3510700 | 1793111 | 4505707 | 5.48E-01 | 0.08 | 0.67 |
| RP_POS_478_33047a13_014 | | 478.3305 | 13.01 | | RP | POS | 478.3317(100) | | 1958354 | 1490058 | 1848219 | 1200356 | 5.48E-01 | -0.08 | 0.68 |
| HILIC_NEG_112_98566a3_98 | | 112.9857 | 3.98 | | HILIC | NEG | 68.9949(100), 112.9854(4) | | 8826439 | 3871955 | 8502166 | 3999762 | 5.48E-01 | -0.08 | 0.65 |
| RP_POS_569_4317a11_6 | | 569.4317 | 11.60 | | RP | POS | | | 766265 | 807858 | 712316 | 494703 | 5.48E-01 | -0.08 | 0.48 |
| RP_POS_765_51202a12_246 | | 765.5120 | 12.25 | | RP | POS | | | 898790 | 611156 | 852419 | 521378 | 5.48E-01 | -0.08 | 0.44 |
| RP_NEG_802_56226a13_212 | PC 34:2 (16:0_18:2) | 802.5623 | 13.21 | | RP | NEG | 279.2342(100), 255.2336(40) | 2 | 1.8E+08 | 1.7E+07 | 1.8E+08 | 1.6E+07 | 5.48E-01 | -0.08 | 0.53 |
| HILIC_POS_340_04675a3_147 | | 340.0468 | 3.15 | | HILIC | POS | | | 994408 | 141919 | 1005157 | 121388 | 5.49E-01 | 0.08 | 0.6 |
| RP_NEG_418_60574a15_492 | | 418.6057 | 15.49 | | RP | NEG | | | 759841 | 144639 | 772127 | 154299 | 5.49E-01 | 0.08 | 0.69 |
| RP_NEG_578_21942a10_59 | | 578.2194 | 10.59 | | RP | NEG | | | 404113 | 128232 | 415464 | 146707 | 5.49E-01 | 0.08 | 0.6 |
| RP_NEG_742_53937a13_205 | | 742.5394 | 13.21 | | RP | NEG | | | 698394 | 310777 | 725202 | 340448 | 5.49E-01 | 0.08 | 0.38 |
| RP_POS_438_37946a10_649 | | 438.3795 | 10.65 | | RP | POS | | | 1.6E+07 | 6837216 | 1.6E+07 | 6889618 | 5.49E-01 | 0.08 | 0.85 |
| RP_POS_772_52637a13_501 | | 772.5264 | 13.50 | | RP | POS | | | 2888464 | 4600165 | 3379776 | 6964373 | 5.49E-01 | 0.08 | 0.69 |
| RP_POS_161_98122a15_575 | | 161.9812 | 15.58 | | RP | POS | | | 4457926 | 2314858 | 4279326 | 2055584 | 5.49E-01 | -0.08 | 0.41 |
| RP_POS_275_23633a10_736 | | 275.2363 | 10.74 | | RP | POS | | | 2133570 | 476924 | 2094503 | 476227 | 5.49E-01 | -0.08 | 0.52 |
| RP_POS_229_08325a6_498 | | 229.0833 | 6.50 | | RP | POS | | | 1025474 | 162822 | 1064931 | 637511 | 5.50E-01 | 0.1 | 0.77 |
| RP_POS_273_10818a2_476 | | 273.1082 | 2.48 | | RP | POS | | | 338370 | 130210 | 351005 | 172384 | 5.50E-01 | 0.08 | 0.61 |
| RP_POS_278_15982a6_476 | | 278.1598 | 6.48 | | RP | POS | | | 2581183 | 1361425 | 2465955 | 1451279 | 5.50E-01 | -0.08 | 0.54 |
| RP_POS_343_33865a14_354 | | 343.3387 | 14.35 | | RP | POS | | | 1.4E+07 | 7271326 | 1.3E+07 | 7763709 | 5.50E-01 | -0.08 | 0.5 |
| RP_POS_364_92801a15_463 | | 364.9280 | 15.46 | | RP | POS | | | 288531 | 99805.4 | 281098 | 82684.9 | 5.50E-01 | -0.08 | 1.06 |
| RP_POS_505_39008a11_77 | | 505.3901 | 11.77 | | RP | POS | | | 505196 | 548462 | 469440 | 314687 | 5.50E-01 | -0.08 | 0.38 |
| RP_POS_280_18835a8_561 | | 280.1884 | 8.56 | | RP | POS | | | 474810 | 55531.8 | 479361 | 56154.9 | 5.51E-01 | 0.08 | 0.97 |
| RP_POS_325_23471a10_361 | | 325.2347 | 10.36 | | RP | POS | | | 1153298 | 919288 | 1233949 | 1043147 | 5.51E-01 | 0.08 | 0.63 |
| RP_POS_504_34485a13_364 | | 504.3449 | 13.36 | | RP | POS | | | 571590 | 1135329 | 668195 | 1227086 | 5.51E-01 | 0.08 | 0.62 |
| RP_POS_872_5564a13_769 | | 872.5564 | 13.77 | | RP | POS | | | 1052413 | 1144649 | 1142900 | 1079813 | 5.51E-01 | 0.08 | 1.05 |
| RP_POS_409_17572a8_802 | 2-(5-methoxy-4,8,8-trimethyl-2-oxo-2,8,9,10-tetrahydropyrano[2,3-f]chromen-3-yl)-N-(pyridin-2-yl)acetamide | 409.1757 | 8.80 | | RP | POS | 409.1745(100) | 2 | 111451 | 76046.5 | 118462 | 93386.5 | 5.51E-01 | 0.08 | 0.72 |
| RP_POS_303_09814a7_069 | | 303.0981 | 7.07 | | RP | POS | | | 252841 | 114108 | 241112 | 164478 | 5.51E-01 | -0.08 | 0.9 |
| HILIC_POS_427_26669a2_267 | | 427.2667 | 2.27 | | HILIC | POS | | | 735092 | 426745 | 771438 | 462197 | 5.52E-01 | 0.08 | 0.47 |
| RP_NEG_1171_17761a9_764 | | 1171.1776 | 9.76 | | RP | NEG | | | 4228174 | 466545 | 4265125 | 442717 | 5.52E-01 | 0.08 | 0.8 |
| RP_POS_1071_72327a10_929 | | 1071.7233 | 10.93 | | RP | POS | | | 1319289 | 728459 | 1381295 | 788378 | 5.52E-01 | 0.08 | 0.64 |
| RP_POS_274_06955a0_585 | | 274.0696 | 0.59 | | RP | POS | | | 335099 | 244844 | 355743 | 261124 | 5.52E-01 | 0.08 | 0.66 |
| RP_POS_506_3588a10_929 | | 506.3588 | 10.93 | | RP | POS | | | 911191 | 282415 | 935570 | 313396 | 5.52E-01 | 0.08 | 0.47 |
| RP_POS_549_38623a10_167 | | 549.3862 | 10.17 | | RP | POS | | | 1572168 | 653826 | 1631303 | 784565 | 5.52E-01 | 0.08 | 0.59 |
| RP_POS_721_15405a8_777 | | 721.1541 | 8.78 | | RP | POS | | | 549452 | 54894.6 | 553720 | 50343.7 | 5.52E-01 | 0.08 | 0.7 |
| HILIC_POS_371_23978a1_326 | | 371.2398 | 1.33 | | HILIC | POS | | | 3397800 | 1164941 | 3308015 | 1050951 | 5.52E-01 | -0.08 | 0.58 |
| RP_POS_244_92418a15_45 | | 244.9242 | 15.45 | | RP | POS | | | 370549 | 66990.6 | 364317 | 83848.2 | 5.52E-01 | -0.08 | 0.68 |
| RP_POS_378_24301a12_362 | | 378.2430 | 12.36 | | RP | POS | | | 1189844 | 844980 | 1123313 | 794390 | 5.52E-01 | -0.08 | 0.94 |
| RP_POS_394_21817a9_208 | | 394.2182 | 9.21 | | RP | POS | | | 804424 | 1379513 | 702971 | 1120532 | 5.52E-01 | -0.08 | 0.71 |
| RP_NEG_325_18469a11_326 | | 325.1847 | 11.33 | | RP | NEG | 325.1825(100), 183.0122(10) | | 5138352 | 4418159 | 5502126 | 4537862 | 5.53E-01 | 0.08 | 0.4 |
| HILIC_POS_267_14395a0_77 | | 267.1440 | 0.77 | | HILIC | POS | | | 1252154 | 98888.9 | 1260713 | 110544 | 5.53E-01 | 0.08 | 0.72 |
| HILIC_POS_340_24738a0_678 | | 340.2474 | 0.68 | | HILIC | POS | | | 334334 | 172962 | 348404 | 174065 | 5.53E-01 | 0.08 | 0.8 |
| RP_NEG_729_18744a9_505 | | 729.1874 | 9.51 | | RP | NEG | | | 388320 | 24539.1 | 390238 | 22813.4 | 5.53E-01 | 0.08 | 0.79 |
| RP_POS_210_11a10_578 | | 210.1100 | 10.58 | | RP | POS | | | 480527 | 188814 | 497790 | 231078 | 5.53E-01 | 0.08 | 0.48 |
| RP_POS_408_74652a12_436 | | 408.7465 | 12.44 | | RP | POS | | | 1.2E+09 | 3.2E+09 | 1.6E+09 | 4.2E+09 | 5.53E-01 | 0.08 | 0.76 |
| RP_POS_412_32669a8_754 | | 412.3267 | 8.75 | | RP | POS | | | 873831 | 490205 | 918497 | 595901 | 5.53E-01 | 0.08 | 0.74 |
| RP_POS_553_36841a14_356 | | 553.3684 | 14.36 | | RP | POS | | | 1E+08 | 3E+07 | 1.1E+08 | 3.1E+07 | 5.53E-01 | 0.08 | 0.75 |
| RP_POS_149_05969a9_79 | Cinnamic acid | 149.0597 | 9.79 | | RP | POS | 149.0231(50) | 2 | 775192 | 111159 | 784440 | 116234 | 5.53E-01 | 0.08 | 0.97 |
| RP_POS_496_34027a10_487 | LysoPC 16:0 (16:0/0:0) | 496.3403 | 10.49 | | RP | POS | 184.0743(100), 104.1078(60) | 1 | 3.1E+09 | 3.8E+08 | 3.1E+09 | 4.1E+08 | 5.53E-01 | 0.08 | 0.89 |
| RP_POS_381_2973a11_151 | | 381.2973 | 11.15 | | RP | POS | 381.2991(100) | | 1889571 | 436841 | 1855126 | 412766 | 5.53E-01 | -0.08 | 0.67 |
| RP_NEG_498_26337a10_019 | | 498.2634 | 10.02 | | RP | NEG | | | 2009149 | 1187349 | 1924919 | 890189 | 5.53E-01 | -0.08 | 0.44 |
| RP_POS_548_27472a10_281 | | 548.2747 | 10.28 | | RP | POS | | | 5270210 | 1335094 | 5155113 | 1486788 | 5.53E-01 | -0.08 | 0.62 |
| RP_POS_674_48755a10_569 | | 674.4876 | 10.57 | | RP | POS | | | 1608975 | 777284 | 1545569 | 784195 | 5.53E-01 | -0.08 | 0.81 |
| RP_POS_130_06529a2_884 | | 130.0653 | 2.88 | | RP | POS | 130.0658(100) | | 2694115 | 361736 | 2723083 | 353830 | 5.54E-01 | 0.08 | 0.56 |
| RP_POS_378_24014a9_602 | | 378.2401 | 9.60 | | RP | POS | | | 2283114 | 644914 | 2330385 | 526000 | 5.54E-01 | 0.08 | 0.61 |
| RP_POS_550_82684a10_928 | | 550.8268 | 10.93 | | RP | POS | | | 1298052 | 431263 | 1336376 | 505523 | 5.54E-01 | 0.08 | 0.91 |
| RP_POS_245_11485a8_594 | | 245.1149 | 8.59 | | RP | POS | 245.1148(100) | | 7041861 | 1162755 | 6949931 | 1110849 | 5.54E-01 | -0.08 | 1.12 |
| RP_NEG_402_88434a14_345 | | 402.8843 | 14.35 | | RP | NEG | | | 1563094 | 680525 | 1504488 | 760321 | 5.54E-01 | -0.08 | 0.78 |
| HILIC_NEG_136_89169a4_925 | | 136.8917 | 4.93 | | HILIC | NEG | | | 1.6E+07 | 6489793 | 1.7E+07 | 6670298 | 5.55E-01 | 0.08 | 0.51 |
| RP_POS_430_31622a10_268 | | 430.3162 | 10.27 | | RP | POS | | | 1118390 | 596411 | 1164879 | 558765 | 5.55E-01 | 0.08 | 0.92 |
| RP_POS_726_17188a11_626 | | 726.1719 | 11.63 | | RP | POS | | | 135951 | 115962 | 146418 | 140291 | 5.55E-01 | 0.08 | 0.79 |
| RP_POS_238_88416a15_397 | | 238.8842 | 15.40 | | RP | POS | | | 1.6E+07 | 2405387 | 1.5E+07 | 2531459 | 5.55E-01 | -0.08 | 0.78 |
| RP_POS_377_26538a9_405 | | 377.2654 | 9.41 | | RP | POS | | | 188361 | 74029.3 | 181867 | 85579.9 | 5.55E-01 | -0.08 | 0.47 |
| RP_POS_569_43298a11_237 | | 569.4330 | 11.24 | | RP | POS | | | 638802 | 1200619 | 562654 | 649152 | 5.55E-01 | -0.08 | 0.46 |
| RP_POS_132_1021a0_682 | Norleucine | 132.1021 | 0.68 | | RP | POS | 86.0976(100), 132.1029(4) | 2 | 2.2E+08 | 5.4E+07 | 2.2E+08 | 4.2E+07 | 5.55E-01 | -0.08 | 0.53 |
| RP_NEG_271_07034a2_884 | | 271.0703 | 2.88 | | RP | NEG | | | 815083 | 124152 | 825339 | 130052 | 5.56E-01 | 0.08 | 0.56 |
| HILIC_POS_130_08643a6_854 | | 130.0864 | 6.85 | | HILIC | POS | 130.1599(100), 84.0815(90) | | 516027 | 478283 | 480894 | 396429 | 5.56E-01 | -0.08 | 0.54 |
| RP_POS_311_21881a10_437 | | 311.2188 | 10.44 | | RP | POS | | | 715233 | 323891 | 691584 | 265826 | 5.56E-01 | -0.08 | 0.84 |
| RP_NEG_431_04813a0_672 | | 431.0481 | 0.67 | | RP | NEG | | | 170857 | 32561.9 | 173451 | 32077.4 | 5.57E-01 | 0.08 | 0.52 |
| RP_NEG_764_5238a13_164 | | 764.5238 | 13.16 | | RP | NEG | | | 1234485 | 418334 | 1270898 | 480768 | 5.57E-01 | 0.08 | 0.5 |
| RP_POS_227_0636a14_228 | | 227.0636 | 14.23 | | RP | POS | | | 486449 | 235802 | 507671 | 285448 | 5.57E-01 | 0.08 | 0.61 |
| RP_POS_740_52551a10_502 | | 740.5255 | 10.50 | | RP | POS | | | 981463 | 553376 | 1032399 | 695018 | 5.57E-01 | 0.08 | 0.48 |
| HILIC_POS_106_08654a3_119 | | 106.0865 | 3.12 | | HILIC | POS | 88.0762(100), 106.0863(90) | | 2E+07 | 7449684 | 1.9E+07 | 6524829 | 5.57E-01 | -0.08 | 1.17 |
| RP_NEG_384_73083a15_425 | | 384.7308 | 15.43 | | RP | NEG | | | 441629 | 170511 | 428226 | 164001 | 5.57E-01 | -0.08 | 0.53 |
| RP_NEG_468_27353a10_057 | | 468.2735 | 10.06 | | RP | NEG | | | 238429 | 85979.7 | 230673 | 104497 | 5.57E-01 | -0.08 | 0.84 |
| RP_POS_196_99599a15_4 | | 196.9960 | 15.40 | | RP | POS | | | 251510 | 258680 | 234993 | 146059 | 5.57E-01 | -0.08 | 0.49 |
| RP_POS_492_3898a11_274 | | 492.3898 | 11.27 | | RP | POS | | | 1696832 | 2483762 | 1487145 | 2710478 | 5.57E-01 | -0.08 | 0.36 |
| RP_POS_701_55902a12_361 | | 701.5590 | 12.36 | | RP | POS | | | 9.8E+07 | 2.1E+07 | 9.9E+07 | 1.8E+07 | 5.58E-01 | 0.08 | 0.79 |
| HILIC_NEG_259_0011a1_357 | | 259.0011 | 1.36 | | HILIC | NEG | | | 1417612 | 390919 | 1452081 | 460074 | 5.58E-01 | 0.08 | 0.39 |
| HILIC_POS_450_04807a3_148 | | 450.0481 | 3.15 | | HILIC | POS | | | 246166 | 49215.4 | 249747 | 40507.4 | 5.58E-01 | 0.08 | 0.43 |
| RP_NEG_566_24585a13_293 | | 566.2459 | 13.29 | | RP | NEG | | | 494045 | 80005.1 | 500143 | 72802.2 | 5.58E-01 | 0.08 | 0.62 |
| RP_POS_181_15852a8_396 | | 181.1585 | 8.40 | | RP | POS | | | 4364870 | 380388 | 4396385 | 403664 | 5.58E-01 | 0.08 | 0.6 |
| RP_NEG_1107_59436a10_491 | | 1107.5944 | 10.49 | | RP | NEG | | | 698255 | 137542 | 687761 | 125380 | 5.58E-01 | -0.08 | 0.71 |
| RP_POS_185_11514a5_73 | | 185.1151 | 5.73 | | RP | POS | | | 2E+07 | 1.2E+07 | 1.9E+07 | 1E+07 | 5.58E-01 | -0.08 | 1.23 |
| RP_NEG_465_23154a11_596 | | 465.2315 | 11.60 | | RP | NEG | | | 374600 | 97332.7 | 383270 | 117070 | 5.59E-01 | 0.08 | 0.83 |
| RP_POS_249_11166a6_707 | | 249.1117 | 6.71 | | RP | POS | | | 1245220 | 193778 | 1260572 | 190856 | 5.59E-01 | 0.08 | 0.66 |
| RP_POS_619_44073a10_662 | | 619.4407 | 10.66 | | RP | POS | | | 965857 | 338909 | 993590 | 353303 | 5.59E-01 | 0.08 | 1.04 |
| RP_POS_698_97784a10_541 | | 698.9778 | 10.54 | | RP | POS | | | 528932 | 319791 | 557010 | 376194 | 5.59E-01 | 0.08 | 0.88 |
| RP_POS_571_42792a11_727 | | 571.4279 | 11.73 | | RP | POS | | | 688277 | 442754 | 656402 | 357539 | 5.59E-01 | -0.08 | 0.36 |
| HILIC_POS_173_1286a4_106 | | 173.1286 | 4.11 | | HILIC | POS | | | 1507157 | 2939271 | 1788515 | 3960700 | 5.60E-01 | 0.08 | 0.56 |
| HILIC_POS_442_05981a3_146 | | 442.0598 | 3.15 | | HILIC | POS | | | 361354 | 70625.3 | 366396 | 56370.7 | 5.60E-01 | 0.08 | 0.62 |
| RP_NEG_893_19299a9_407 | | 893.1930 | 9.41 | | RP | NEG | | | 3682907 | 485807 | 3718520 | 410529 | 5.60E-01 | 0.08 | 0.92 |
| RP_NEG_191_8607a11_144 | | 191.8607 | 11.14 | | RP | NEG | | | 164929 | 35500.4 | 162161 | 34023.3 | 5.60E-01 | -0.08 | 0.85 |
| RP_NEG_195_8112a15_493 | | 195.8112 | 15.49 | | RP | NEG | | | 4.6E+07 | 895604 | 4.6E+07 | 947602 | 5.60E-01 | -0.08 | 0.47 |
| RP_POS_331_65341a5_59 | | 331.6534 | 5.59 | | RP | POS | | | 212572 | 101334 | 204136 | 109347 | 5.60E-01 | -0.08 | 0.76 |
| HILIC_POS_172_13318a0_63 | | 172.1332 | 0.63 | | HILIC | POS | 172.1332(100) | | 3129081 | 631007 | 3186723 | 794273 | 5.61E-01 | 0.08 | 0.52 |
| HILIC_NEG_234_82025a10_716 | | 234.8203 | 10.72 | | HILIC | NEG | | | 1747073 | 1234035 | 1840551 | 1126679 | 5.61E-01 | 0.08 | 0.45 |
| HILIC_POS_298_18604a0_768 | | 298.1860 | 0.77 | | HILIC | POS | | | 202388 | 23506.8 | 204359 | 25859.4 | 5.61E-01 | 0.08 | 0.5 |
| RP_NEG_747_56873a12_805 | | 747.5687 | 12.81 | | RP | NEG | | | 3.6E+07 | 5704702 | 3.7E+07 | 5948748 | 5.61E-01 | 0.08 | 0.99 |
| HILIC_POS_371_23972a6_941 | | 371.2397 | 6.94 | | HILIC | POS | | | 987461 | 561213 | 943539 | 545411 | 5.61E-01 | -0.08 | 0.59 |
| RP_POS_251_1256a5_227 | | 251.1256 | 5.23 | | RP | POS | | | 243171 | 200011 | 228611 | 167122 | 5.61E-01 | -0.08 | 0.81 |
| RP_POS_464_37311a11_41 | | 464.3731 | 11.41 | | RP | POS | | | 765443 | 135435 | 754831 | 132044 | 5.61E-01 | -0.08 | 0.71 |
| RP_POS_723_53925a12_361 | | 723.5393 | 12.36 | | RP | POS | | | 2.3E+07 | 5378247 | 2.3E+07 | 4669364 | 5.62E-01 | 0.08 | 0.78 |
| RP_NEG_357_28094a11_401 | | 357.2809 | 11.40 | | RP | NEG | | | 311455 | 197954 | 327945 | 215981 | 5.62E-01 | 0.08 | 0.92 |
| RP_POS_862_50555a13_113 | | 862.5056 | 13.11 | | RP | POS | | | 518036 | 255338 | 542598 | 348642 | 5.62E-01 | 0.08 | 0.53 |
| HILIC_NEG_154_99278a0_932 | | 154.9928 | 0.93 | | HILIC | NEG | | | 208139 | 27437.2 | 205896 | 28919.8 | 5.62E-01 | -0.08 | 0.67 |
| RP_NEG_240_99107a15_465 | | 240.9911 | 15.47 | | RP | NEG | | | 438542 | 76750.7 | 432130 | 84280.9 | 5.62E-01 | -0.08 | 0.6 |
| RP_POS_279_64691a10_179 | | 279.6469 | 10.18 | | RP | POS | | | 943288 | 545970 | 903206 | 467567 | 5.62E-01 | -0.08 | 1.03 |
| RP_POS_301_14072a9_44 | | 301.1407 | 9.44 | | RP | POS | | | 1.5E+07 | 1883711 | 1.5E+07 | 1833782 | 5.62E-01 | -0.08 | 0.82 |
| RP_POS_589_22504a7_07 | | 589.2250 | 7.07 | | RP | POS | | | 2154458 | 449081 | 2119513 | 434297 | 5.62E-01 | -0.08 | 1.16 |
| RP_NEG_623_20422a11_612 | | 623.2042 | 11.61 | | RP | NEG | | | 522724 | 176026 | 536055 | 161639 | 5.63E-01 | 0.08 | 0.69 |
| RP_POS_247_92902a15_385 | | 247.9290 | 15.39 | | RP | POS | | | 570270 | 273033 | 595023 | 342523 | 5.63E-01 | 0.08 | 0.47 |
| RP_POS_650_78827a11_481 | | 650.7883 | 11.48 | | RP | POS | | | 250952 | 143696 | 262722 | 153226 | 5.63E-01 | 0.08 | 0.46 |
| RP_NEG_203_88606a15_487 | | 203.8861 | 15.49 | | RP | NEG | | | 590671 | 375976 | 563713 | 308589 | 5.63E-01 | -0.08 | 0.51 |
| RP_NEG_247_17036a9_192 | | 247.1704 | 9.19 | | RP | NEG | | | 396451 | 38292 | 393543 | 35365.9 | 5.63E-01 | -0.08 | 0.54 |
| RP_POS_492_19354a11_694 | | 492.1935 | 11.69 | | RP | POS | | | 119932 | 48958.7 | 116475 | 38690 | 5.63E-01 | -0.08 | 0.69 |
| HILIC_NEG_225_06158a5_122 | | 225.0616 | 5.12 | | HILIC | NEG | | | 1.6E+07 | 4262904 | 1.6E+07 | 4893981 | 5.64E-01 | 0.08 | 0.71 |
| HILIC_NEG_272_79489a3_231 | | 272.7949 | 3.23 | | HILIC | NEG | | | 930359 | 133128 | 941113 | 138736 | 5.64E-01 | 0.08 | 0.54 |
| HILIC_POS_147_14906a5_828 | | 147.1491 | 5.83 | | HILIC | POS | | | 1E+08 | 8.3E+07 | 1.1E+08 | 8E+07 | 5.64E-01 | 0.08 | 0.54 |
| RP_NEG_340_77969a15_399 | | 340.7797 | 15.40 | | RP | NEG | | | 1736538 | 603975 | 1783068 | 578439 | 5.64E-01 | 0.08 | 0.71 |
| RP_NEG_357_0206a2_884 | | 357.0206 | 2.88 | | RP | NEG | | | 497812 | 79479.7 | 503957 | 76562.1 | 5.64E-01 | 0.08 | 0.6 |
| RP_NEG_558_33423a10_931 | | 558.3342 | 10.93 | | RP | NEG | | | 3689452 | 575846 | 3736271 | 607383 | 5.64E-01 | 0.08 | 0.65 |
| RP_NEG_627_11401a9_479 | | 627.1140 | 9.48 | | RP | NEG | | | 1481777 | 178530 | 1493995 | 131903 | 5.64E-01 | 0.08 | 0.47 |
| RP_POS_244_22705a9_402 | | 244.2271 | 9.40 | | RP | POS | | | 1059993 | 66079.6 | 1064877 | 58129.5 | 5.64E-01 | 0.08 | 0.69 |
| RP_POS_662_94818a10_265 | | 662.9482 | 10.27 | | RP | POS | | | 1881506 | 822838 | 1814933 | 862348 | 5.64E-01 | -0.08 | 0.68 |
| RP_NEG_211_91838a14_337 | | 211.9184 | 14.34 | | RP | NEG | | | 1555723 | 536715 | 1594230 | 444177 | 5.65E-01 | 0.08 | 0.38 |
| HILIC_NEG_279_01642a3_999 | | 279.0164 | 4.00 | | HILIC | NEG | | | 513652 | 185919 | 500539 | 148653 | 5.65E-01 | -0.08 | 0.51 |
| RP_POS_484_43579a12_423 | | 484.4358 | 12.42 | | RP | POS | | | 516534 | 236611 | 498501 | 222408 | 5.65E-01 | -0.08 | 0.42 |
| RP_NEG_528_82251a15_366 | | 528.8225 | 15.37 | | RP | NEG | | | 667821 | 69888.7 | 673425 | 72643.8 | 5.66E-01 | 0.08 | 0.82 |
| RP_POS_357_0083a15_496 | | 357.0083 | 15.50 | | RP | POS | | | 998326 | 153859 | 1010737 | 161791 | 5.66E-01 | 0.08 | 0.56 |
| RP_POS_388_23001a0_677 | | 388.2300 | 0.68 | | RP | POS | | | 834415 | 409451 | 868542 | 454250 | 5.66E-01 | 0.08 | 0.42 |
| RP_POS_393_22455a10_332 | | 393.2246 | 10.33 | | RP | POS | | | 1100757 | 576279 | 1145348 | 562046 | 5.66E-01 | 0.08 | 1.03 |
| RP_POS_515_0318a10_041 | | 515.0318 | 10.04 | | RP | POS | | | 1044539 | 519961 | 1086878 | 554363 | 5.66E-01 | 0.08 | 0.58 |
| RP_POS_540_44574a10_87 | | 540.4457 | 10.87 | | RP | POS | | | 7850887 | 663094 | 7900813 | 611303 | 5.66E-01 | 0.08 | 0.74 |
| RP_POS_796_68475a14_356 | | 796.6848 | 14.36 | | RP | POS | | | 1314636 | 2186865 | 1474128 | 1893382 | 5.66E-01 | 0.08 | 0.98 |
| RP_POS_323_14624a6_416 | | 323.1462 | 6.42 | | RP | POS | | | 193859 | 80559.5 | 187423 | 83150.8 | 5.66E-01 | -0.08 | 0.81 |
| RP_POS_552_38586a8_918 | | 552.3859 | 8.92 | | RP | POS | | | 382153 | 218585 | 364820 | 222771 | 5.66E-01 | -0.08 | 0.63 |
| HILIC_NEG_100_93345a5_93 | | 100.9335 | 5.93 | | HILIC | NEG | | | 2.6E+07 | 4100573 | 2.7E+07 | 4807730 | 5.67E-01 | 0.08 | 0.59 |
| HILIC_NEG_391_00143a1_363 | | 391.0014 | 1.36 | | HILIC | NEG | | | 696560 | 228899 | 714066 | 218734 | 5.67E-01 | 0.08 | 0.41 |
| RP_NEG_1091_72717a10_931 | | 1091.7272 | 10.93 | | RP | NEG | | | 5917796 | 2164843 | 6092095 | 2278096 | 5.67E-01 | 0.08 | 0.72 |

**Supplemantary table 1**: The non-targeted metabolomics results ordered by the VIP value (p value < 0.0003 are shaded dark green, p value < 0.05 are shaded light green, d values below 0 are blue and above 0 are red)

| | | | | | | | | | Cases | | Controls | | Welch test | Cohen's | |
|---|---|---|---|---|---|---|---|---|---|---|---|---|---|---|---|
| Feature_ID | Metabolite_name | Average_Mz | Average_RT_min | HMDB ID | RP_HILIC | POS_NEG | Cureted_MSMS | ID_level | Mean | SD | Mean2 | SD3 | p | d | VIP |
| RP_NEG_231_93173a14_08 | | 231.9317 | 14.08 | | RP | NEG | | | 1388806 | 865794 | 1453261 | 784853 | 5.67E-01 | 0.08 | 0.63 |
| RP_NEG_581_45563a10_961 | | 581.4556 | 10.96 | | RP | NEG | | | 108430 | 120461 | 117511 | 112105 | 5.67E-01 | 0.08 | 1.01 |
| RP_POS_201_11185a2_717 | | 201.1119 | 2.72 | | RP | POS | | | 45327.9 | 26460.5 | 47790.1 | 35071.9 | 5.67E-01 | 0.08 | 1.06 |
| RP_POS_229_17998a7_765 | | 229.1800 | 7.77 | | RP | POS | | | 545696 | 69221.1 | 551001 | 66603.6 | 5.67E-01 | 0.08 | 1.05 |
| RP_POS_310_23724a9_788 | | 310.2372 | 9.79 | | RP | POS | | | 1.1E+07 | 1274986 | 1.1E+07 | 1622635 | 5.67E-01 | 0.08 | 1.01 |
| RP_POS_509_03812a10_761 | | 509.0381 | 10.76 | | RP | POS | | | 161469 | 127253 | 172997 | 161901 | 5.67E-01 | 0.08 | 0.59 |
| HILIC_POS_553_40894a2_26 | | 553.4089 | 2.26 | | HILIC | POS | | | 286293 | 252749 | 268325 | 207456 | 5.67E-01 | -0.08 | 0.53 |
| RP_NEG_294_98297a6_491 | | 294.9830 | 6.49 | | RP | NEG | | | 4621820 | 104867 | 4611557 | 149565 | 5.67E-01 | -0.08 | 0.53 |
| RP_POS_349_18759a10_685 | | 349.1876 | 10.69 | | RP | POS | | | 1976320 | 591270 | 1925510 | 692739 | 5.67E-01 | -0.08 | 0.57 |
| RP_POS_397_36697a11_401 | | 397.3670 | 11.40 | | RP | POS | | | 435933 | 167849 | 424156 | 133731 | 5.67E-01 | -0.08 | 0.92 |
| RP_POS_640_93689a10_284 | | 640.9369 | 10.28 | | RP | POS | | | 2167572 | 1002831 | 2080336 | 1198795 | 5.67E-01 | -0.08 | 0.91 |
| HILIC_POS_174_12396a1_833 | | 174.1240 | 1.83 | | HILIC | POS | | | 1600142 | 1066031 | 1679839 | 982021 | 5.68E-01 | 0.08 | 0.49 |
| RP_NEG_481_77884a15_372 | | 481.7788 | 15.37 | | RP | NEG | | | 445385 | 171810 | 459461 | 187086 | 5.68E-01 | 0.08 | 0.45 |
| HILIC_POS_371_24002a5_654 | | 371.2400 | 5.65 | | HILIC | POS | | | 1E+07 | 1266752 | 1E+07 | 1019023 | 5.68E-01 | -0.08 | 0.76 |
| HILIC_POS_666_33301a0_82 | | 666.3330 | 0.82 | | HILIC | POS | | | 1639455 | 594532 | 1593861 | 575131 | 5.68E-01 | -0.08 | 0.49 |
| RP_NEG_445_1492a11_591 | | 445.1492 | 11.59 | | RP | NEG | | | 534262 | 53892.6 | 530083 | 53132.7 | 5.68E-01 | -0.08 | 0.71 |
| RP_NEG_615_14117a8_475 | | 615.1412 | 8.48 | | RP | NEG | | | 1308003 | 64447.5 | 1303063 | 62181.3 | 5.68E-01 | -0.08 | 0.67 |
| RP_POS_478_1781a11_029 | | 478.1781 | 11.03 | | RP | POS | | | 231953 | 44805.2 | 228378 | 46555.8 | 5.68E-01 | -0.08 | 0.7 |
| RP_POS_760_58514a14_356 | | 760.5851 | 14.36 | | RP | POS | | | 1.2E+07 | 5364762 | 1.2E+07 | 5204878 | 5.68E-01 | -0.08 | 0.76 |
| RP_NEG_165_83722a0_704 | | 165.8372 | 0.70 | | RP | NEG | | | 1662436 | 611633 | 1714274 | 707303 | 5.69E-01 | 0.08 | 0.61 |
| RP_NEG_351_01431a2_884 | | 351.0143 | 2.88 | | RP | NEG | | | 1705886 | 253391 | 1725388 | 247270 | 5.69E-01 | 0.08 | 0.61 |
| RP_POS_325_22705a5_763 | | 325.2271 | 5.76 | | RP | POS | | | 959827 | 250915 | 980054 | 266970 | 5.69E-01 | 0.08 | 0.97 |
| RP_POS_386_69699a6_644 | | 386.6970 | 6.64 | | RP | POS | | | 421878 | 85194 | 429006 | 96514.7 | 5.69E-01 | 0.08 | 0.65 |
| RP_POS_421_78293a10_956 | | 421.7829 | 10.96 | | RP | POS | | | 216432 | 114821 | 226634 | 143340 | 5.69E-01 | 0.08 | 0.81 |
| RP_NEG_307_85641a15_544 | | 307.8564 | 15.54 | | RP | NEG | | | 2208368 | 591692 | 2161091 | 619883 | 5.69E-01 | -0.08 | 0.46 |
| RP_POS_419_3508a12_228 | | 419.3508 | 12.23 | | RP | POS | | | 1584443 | 330863 | 1558711 | 329378 | 5.69E-01 | -0.08 | 0.87 |
| RP_POS_657_48529a15_017 | | 657.4853 | 15.02 | | RP | POS | | | 1312582 | 1098046 | 1229717 | 1035163 | 5.69E-01 | -0.08 | 0.77 |
| RP_NEG_416_17853a1_912 | | 416.1785 | 1.91 | | RP | NEG | | | 159728 | 104803 | 168474 | 118777 | 5.70E-01 | 0.08 | 1.22 |
| RP_POS_178_97464a15_42 | | 178.9746 | 15.42 | | RP | POS | | | 828113 | 414631 | 861907 | 451780 | 5.70E-01 | 0.08 | 0.7 |
| RP_NEG_431_22913a8_727 | | 431.2291 | 8.73 | | RP | NEG | | | 311312 | 195758 | 297440 | 162988 | 5.70E-01 | -0.08 | 0.36 |
| RP_NEG_915_1275a9_629 | | 915.1275 | 9.63 | | RP | NEG | | | 6773094 | 401052 | 6743225 | 369362 | 5.70E-01 | -0.08 | 0.92 |
| RP_POS_1067_77209a14_347 | | 1067.7721 | 14.35 | | RP | POS | | | 1879846 | 2363529 | 1726789 | 1560877 | 5.70E-01 | -0.08 | 0.77 |
| RP_POS_256_15399a5_075 | | 256.1540 | 5.08 | | RP | POS | | | 71706.3 | 32958.8 | 69219.1 | 31205.4 | 5.70E-01 | -0.08 | 1.04 |
| RP_POS_519_31921a14_111 | | 519.3192 | 14.11 | | RP | POS | | | 283260 | 185653 | 269559 | 168429 | 5.70E-01 | -0.08 | 0.36 |
| HILIC_POS_135_00279a1_328 | | 135.0028 | 1.33 | | HILIC | POS | | | 974762 | 188191 | 993582 | 280507 | 5.71E-01 | 0.08 | 0.76 |
| RP_POS_144_0808a2_482 | | 144.0808 | 2.48 | | RP | POS | | | 7780976 | 1468532 | 7895335 | 1482148 | 5.71E-01 | 0.08 | 0.67 |
| RP_POS_229_14311a5_227 | | 229.1431 | 5.23 | | RP | POS | | | 152809 | 91410.3 | 159730 | 87294.3 | 5.71E-01 | 0.08 | 0.79 |
| RP_POS_374_30518a11_195 | | 374.3052 | 11.20 | | RP | POS | | | 495628 | 350696 | 526120 | 426397 | 5.71E-01 | 0.08 | 0.83 |
| RP_POS_638_7807a11_343 | | 638.7807 | 11.34 | | RP | POS | | | 186948 | 83800.7 | 194350 | 104330 | 5.71E-01 | 0.08 | 0.87 |
| HILIC_POS_226_10503a2_853 | | 226.1050 | 2.85 | | HILIC | POS | | | 1096316 | 4638387 | 784148 | 3434564 | 5.71E-01 | -0.08 | 0.49 |
| RP_NEG_378_91904a14_34 | | 378.9190 | 14.34 | | RP | NEG | | | 1530145 | 642072 | 1480752 | 634612 | 5.71E-01 | -0.08 | 0.76 |
| RP_POS_593_91577a9_955 | | 593.9158 | 9.96 | | RP | POS | | | 1925850 | 836069 | 1847438 | 1138604 | 5.71E-01 | -0.08 | 0.85 |
| RP_POS_748_52795a13_231 | | 748.5280 | 13.23 | | RP | POS | | | 3645219 | 1712928 | 3520999 | 1503913 | 5.71E-01 | -0.08 | 0.99 |
| HILIC_POS_387_21378a4_863 | | 387.2138 | 4.86 | | HILIC | POS | | | 988693 | 512676 | 1031036 | 575138 | 5.72E-01 | 0.08 | 0.49 |
| HILIC_POS_478_29257a1_858 | | 478.2926 | 1.86 | | HILIC | POS | | | 1377532 | 610044 | 1435554 | 852943 | 5.72E-01 | 0.08 | 0.55 |
| RP_NEG_171_94678a0_86 | | 171.9468 | 0.86 | | RP | NEG | | | 2330774 | 663688 | 2378665 | 579964 | 5.72E-01 | 0.08 | 0.71 |
| RP_NEG_368_97726a8_368 | | 368.9773 | 8.37 | | RP | NEG | | | 1061264 | 641276 | 1104135 | 470086 | 5.72E-01 | 0.08 | 0.44 |
| RP_POS_243_1825a15_412 | | 243.1825 | 15.41 | | RP | POS | | | 245464 | 123145 | 254855 | 120274 | 5.72E-01 | 0.08 | 0.5 |
| RP_POS_392_19043a6_563 | | 392.1904 | 6.56 | | RP | POS | | | 1.1E+07 | 905212 | 1.1E+07 | 961311 | 5.72E-01 | 0.08 | 0.8 |
| RP_POS_490_29022a9_929 | | 490.2902 | 9.93 | | RP | POS | | | 9439083 | 4166792 | 9788599 | 4808506 | 5.72E-01 | 0.08 | 0.46 |
| RP_NEG_712_26068a10_313 | | 712.2607 | 10.31 | | RP | NEG | | | 489980 | 87295.9 | 483159 | 88933.6 | 5.72E-01 | -0.08 | 0.61 |
| RP_POS_262_03766a0_677 | | 262.0377 | 0.68 | | RP | POS | | | 391979 | 94543.9 | 385181 | 81989.4 | 5.72E-01 | -0.08 | 0.57 |
| RP_POS_575_40521a11_399 | | 575.4052 | 11.40 | | RP | POS | | | 123112 | 103434 | 114888 | 109039 | 5.72E-01 | -0.08 | 0.44 |
| RP_NEG_167_02061a0_679 | | 167.0206 | 0.68 | | RP | NEG | 167.0206(100), 124.0149(60),96.0199(10) | | 1.4E+08 | 1.7E+07 | 1.4E+08 | 1.8E+07 | 5.73E-01 | 0.08 | 0.33 |
| HILIC_POS_485_3584a1_462 | | 485.3584 | 1.46 | | HILIC | POS | | | 194290 | 503393 | 238258 | 620200 | 5.73E-01 | 0.08 | 0.53 |
| RP_POS_164_98445a11_189 | | 164.9845 | 11.19 | | RP | POS | | | 4165223 | 1678850 | 4300031 | 1805121 | 5.73E-01 | 0.08 | 0.9 |
| RP_POS_204_97923a1_386 | | 204.9792 | 1.39 | | RP | POS | | | 543250 | 575096 | 589096 | 611661 | 5.73E-01 | 0.08 | 0.91 |
| RP_POS_200_16463a7_687 | | 200.1646 | 7.69 | | RP | POS | | | 139391 | 63662.6 | 134656 | 59465.5 | 5.73E-01 | -0.08 | 0.45 |
| RP_POS_573_40625a11_657 | | 573.4063 | 11.66 | | RP | POS | | | 1698806 | 2927219 | 1488766 | 2545073 | 5.73E-01 | -0.08 | 0.62 |
| RP_POS_623_28992a7_235 | | 623.2899 | 7.24 | | RP | POS | | | 1352692 | 135252 | 1342459 | 130801 | 5.73E-01 | -0.08 | 1.01 |
| RP_POS_626_60571a11_453 | | 626.6057 | 11.45 | | RP | POS | | | 154459 | 104292 | 146919 | 92036.1 | 5.73E-01 | -0.08 | 0.86 |
| RP_NEG_498_28973a9_025 | | 498.2897 | 9.03 | | RP | NEG | 498.2869(100), 79.9577(2) | | 1.3E+07 | 2.9E+07 | 1.5E+07 | 3.6E+07 | 5.74E-01 | 0.08 | 0.6 |
| HILIC_NEG_113_92651a5_653 | | 113.9265 | 5.65 | | HILIC | NEG | | | 2351916 | 294651 | 2376279 | 334416 | 5.74E-01 | 0.08 | 0.43 |
| HILIC_NEG_144_96011a1_353 | | 144.9601 | 1.35 | | HILIC | NEG | | | 2877392 | 449939 | 2912024 | 451010 | 5.74E-01 | 0.08 | 0.83 |
| HILIC_NEG_209_83372a3_243 | | 209.8337 | 3.24 | | HILIC | NEG | | | 322730 | 93223.8 | 330210 | 100374 | 5.74E-01 | 0.08 | 0.42 |
| RP_NEG_262_992a7_346 | | 262.9920 | 7.35 | | RP | NEG | | | 1644445 | 223322 | 1663124 | 258684 | 5.74E-01 | 0.08 | 0.78 |
| RP_POS_411_23523a9_848 | | 411.2352 | 9.85 | | RP | POS | | | 1920876 | 794290 | 1984177 | 845184 | 5.74E-01 | 0.08 | 0.93 |
| RP_POS_586_26984a5_476 | | 586.2698 | 5.48 | | RP | POS | | | 279972 | 191313 | 295522 | 210608 | 5.74E-01 | 0.08 | 0.55 |
| RP_NEG_479_28357a10_887 | | 479.2836 | 10.89 | | RP | NEG | | | 326645 | 1039318 | 258200 | 741141 | 5.74E-01 | -0.08 | 0.7 |
| RP_POS_269_24731a9_408 | | 269.2473 | 9.41 | | RP | POS | | | 1907010 | 115466 | 1898809 | 98819.7 | 5.74E-01 | -0.08 | 0.6 |
| RP_POS_402_24268a11_594 | | 402.2427 | 11.59 | | RP | POS | | | 1142278 | 541560 | 1101487 | 521080 | 5.74E-01 | -0.08 | 1.09 |
| HILIC_POS_170_09239a6_279 | 1-methylhistidine | 170.0924 | 6.28 | | HILIC | POS | 124.0876(100), 170.0929(40) | 1 | 2.4E+07 | 7399294 | 2.4E+07 | 6333318 | 5.74E-01 | -0.08 | 0.5 |
| RP_POS_427_99997a15_387 | | 428.0000 | 15.39 | | RP | POS | | | 373493 | 251155 | 389846 | 174585 | 5.75E-01 | 0.08 | 0.39 |
| RP_POS_456_3299a10_09 | | 456.3299 | 10.09 | | RP | POS | | | 352606 | 222220 | 369813 | 225667 | 5.75E-01 | 0.08 | 0.9 |
| RP_POS_506_85901a15_393 | | 506.8590 | 15.39 | | RP | POS | | | 204444 | 149789 | 216141 | 154583 | 5.75E-01 | 0.08 | 0.62 |
| RP_POS_738_51361a11_516 | | 738.5136 | 11.52 | | RP | POS | | | 158525 | 75830 | 165162 | 94734.6 | 5.75E-01 | 0.08 | 0.57 |
| RP_POS_774_6001a14_349 | | 774.6001 | 14.35 | | RP | POS | | | 6685595 | 3927231 | 7022724 | 4762721 | 5.75E-01 | 0.08 | 0.66 |
| HILIC_POS_176_1026a1_362 | | 176.1026 | 1.36 | | HILIC | POS | 176.1036(100) | | 186192 | 117741 | 177333 | 113718 | 5.75E-01 | -0.08 | 0.79 |
| HILIC_NEG_347_00262a7_477 | | 347.0026 | 7.48 | | HILIC | NEG | | | 699740 | 37078.7 | 697006 | 34322.9 | 5.75E-01 | -0.08 | 0.85 |
| HILIC_POS_354_01227a7_42 | | 354.0123 | 7.42 | | HILIC | POS | | | 2011755 | 120729 | 2002475 | 120964 | 5.75E-01 | -0.08 | 1.08 |
| RP_POS_386_20938a9_008 | | 386.2094 | 9.01 | | RP | POS | | | 285334 | 68863.1 | 279999 | 70040.1 | 5.75E-01 | -0.08 | 0.43 |
| RP_NEG_258_73273a15_473 | | 258.7327 | 15.47 | | RP | NEG | | | 390533 | 100276 | 397928 | 93667.3 | 5.76E-01 | 0.08 | 0.66 |
| RP_POS_371_25818a10_131 | | 371.2582 | 10.13 | | RP | POS | | | 1933786 | 124670 | 1944424 | 150296 | 5.76E-01 | 0.08 | 0.55 |
| RP_POS_806_56885a12_785 | PC 38:4 | 806.5689 | 12.79 | | RP | POS | 184.0743(100), 86.0962(15) | 1 | 7.6E+07 | 2.9E+07 | 7.8E+07 | 3.5E+07 | 5.76E-01 | 0.08 | 0.73 |
| HILIC_NEG_168_98912a0_934 | | 168.9891 | 0.93 | | HILIC | NEG | | | 739379 | 67688.7 | 734231 | 67002.3 | 5.76E-01 | -0.08 | 0.48 |
| HILIC_NEG_180_97289a4_352 | | 180.9729 | 4.35 | | HILIC | NEG | | | 4.9E+07 | 2530914 | 4.9E+07 | 2564176 | 5.76E-01 | -0.08 | 0.86 |
| RP_POS_457_30661a9_839 | | 457.3066 | 9.84 | | RP | POS | | | 195389 | 107492 | 185785 | 138953 | 5.76E-01 | -0.08 | 0.59 |
| RP_POS_559_51569a14_049 | | 559.5157 | 14.05 | | RP | POS | | | 4.2E+07 | 4249730 | 4.2E+07 | 4155433 | 5.76E-01 | -0.08 | 0.61 |
| RP_NEG_209_15457a7_694 | | 209.1546 | 7.69 | | RP | NEG | | | 7735475 | 419888 | 7767823 | 427585 | 5.77E-01 | 0.08 | 0.63 |
| RP_NEG_232_98463a8_352 | | 232.9846 | 8.35 | | RP | NEG | | | 616277 | 77320.7 | 622638 | 88314.3 | 5.77E-01 | 0.08 | 0.43 |
| RP_POS_1195_33276a13_122 | | 1195.3328 | 13.12 | | RP | POS | | | 6233902 | 1693631 | 6361922 | 1661736 | 5.77E-01 | 0.08 | 0.6 |
| RP_POS_428_3363a10_427 | | 428.3363 | 10.43 | | RP | POS | | | 2349475 | 114930 | 2358632 | 124087 | 5.77E-01 | 0.08 | 1.02 |
| RP_POS_603_91589a10_488 | | 603.9159 | 10.49 | | RP | POS | | | 671749 | 366056 | 700592 | 388283 | 5.77E-01 | 0.08 | 0.96 |
| HILIC_NEG_384_02505a6_134 | | 384.0251 | 6.13 | | HILIC | NEG | | | 185563 | 24478.8 | 183729 | 23632.4 | 5.77E-01 | -0.08 | 0.79 |
| RP_POS_311_23682a10_895 | | 311.2368 | 10.90 | | RP | POS | | | 547144 | 256987 | 527584 | 256219 | 5.77E-01 | -0.08 | 0.91 |
| RP_POS_376_29761a11_656 | | 376.2976 | 11.66 | | RP | POS | | | 1848619 | 2649741 | 1661312 | 2274838 | 5.77E-01 | -0.08 | 0.57 |
| HILIC_POS_118_08661a7_091 | | 118.0866 | 7.09 | | HILIC | POS | | | 1.3E+07 | 3050452 | 1.3E+07 | 3021441 | 5.78E-01 | 0.08 | 0.55 |
| RP_POS_344_22772a3_994 | | 344.2277 | 3.99 | | RP | POS | | | 712448 | 249993 | 736984 | 373390 | 5.78E-01 | 0.08 | 0.57 |
| RP_POS_350_02921a4_825 | | 350.0292 | 4.83 | | RP | POS | | | 616740 | 495357 | 661195 | 649603 | 5.78E-01 | 0.08 | 0.67 |
| HILIC_POS_234_09747a3_113 | | 234.0975 | 3.11 | | HILIC | POS | | | 456739 | 165925 | 445312 | 135234 | 5.78E-01 | -0.08 | 0.65 |
| RP_POS_226_12215a7_915 | | 226.1222 | 7.92 | | RP | POS | | | 506777 | 48623.4 | 503124 | 47486 | 5.78E-01 | -0.08 | 0.62 |
| RP_POS_329_15689a5_816 | | 329.1569 | 5.82 | | RP | POS | | | 410159 | 250454 | 394320 | 164316 | 5.78E-01 | -0.08 | 0.56 |
| RP_POS_390_77237a13_214 | | 390.7724 | 13.21 | | RP | POS | | | 6440881 | 2404094 | 6275838 | 1952374 | 5.78E-01 | -0.08 | 0.82 |
| HILIC_POS_232_15408a1_557 | Butanoylcarnitine | 232.1541 | 1.56 | | HILIC | POS | 85.0292(100), 232.154(30) | 1 | 3.1E+07 | 1.5E+07 | 3E+07 | 1.1E+07 | 5.78E-01 | -0.08 | 0.57 |
| RP_POS_167_1068a7_147 | | 167.1068 | 7.15 | | RP | POS | 167.107(100) | | 2082082 | 186547 | 2098929 | 247773 | 5.79E-01 | 0.08 | 0.41 |
| RP_POS_179_14297a8_434 | | 179.1430 | 8.43 | | RP | POS | | | 4930759 | 349838 | 4957080 | 344646 | 5.79E-01 | 0.08 | 0.45 |
| RP_POS_238_0676a9_384 | | 238.0676 | 9.38 | | RP | POS | | | 2100867 | 957911 | 2173209 | 950466 | 5.79E-01 | 0.08 | 0.74 |
| RP_POS_526_28754a10_901 | | 526.2875 | 10.90 | | RP | POS | | | 405345 | 83127.2 | 412175 | 95255.3 | 5.79E-01 | 0.08 | 0.54 |
| RP_POS_800_07874a13_79 | | 800.0787 | 13.79 | | RP | POS | | | 1213810 | 748703 | 1275688 | 869218 | 5.79E-01 | 0.08 | 0.56 |
| RP_POS_734_56952a13_671 | PC 32:0 (16:0/16:0) | 734.5695 | 13.67 | | RP | POS | 184.0745(100) | 1 | 1.9E+07 | 7534376 | 1.9E+07 | 6012840 | 5.79E-01 | 0.08 | 0.69 |
| HILIC_POS_216_92239a4_295 | | 216.9224 | 4.30 | | HILIC | POS | | | 325229 | 305862 | 303623 | 265348 | 5.79E-01 | -0.08 | 0.33 |
| RP_NEG_228_98102a3_583 | | 228.9810 | 3.58 | | RP | NEG | | | 487187 | 272935 | 467961 | 235082 | 5.79E-01 | -0.08 | 0.7 |
| RP_NEG_378_91809a15_312 | | 378.9181 | 15.31 | | RP | NEG | | | 1381915 | 135290 | 1371774 | 132488 | 5.79E-01 | -0.08 | 0.55 |
| RP_NEG_622_33478a10_724 | | 622.3348 | 10.72 | | RP | NEG | | | 947138 | 234691 | 929105 | 240545 | 5.79E-01 | -0.08 | 0.8 |
| RP_POS_205_09734a2_884 | | 205.0973 | 2.88 | | RP | POS | 188.0708(100), 146.0602(80) | | 1.3E+08 | 2.3E+07 | 1.3E+08 | 2.1E+07 | 5.80E-01 | 0.08 | 0.72 |
| RP_POS_292_63812a10_599 | | 292.6381 | 10.60 | | RP | POS | | | 373156 | 202158 | 387150 | 168797 | 5.80E-01 | 0.08 | 0.58 |
| RP_POS_495_83185a9_39 | | 495.8319 | 9.39 | | RP | POS | | | 277976 | 161580 | 291340 | 188081 | 5.80E-01 | 0.08 | 0.95 |
| RP_POS_866_22668a9_588 | | 866.2267 | 9.59 | | RP | POS | | | 1765951 | 169248 | 1778263 | 156709 | 5.80E-01 | 0.08 | 0.97 |
| RP_POS_199_16924a8_599 | Lauric acid | 199.1692 | 8.60 | | RP | POS | 199.1877(100) | 2 | 4997025 | 971698 | 4925822 | 915817 | 5.80E-01 | -0.08 | 0.72 |
| HILIC_NEG_139_94466a5_652 | | 139.9447 | 5.65 | | HILIC | NEG | | | 364651 | 50097.7 | 361186 | 41786.8 | 5.80E-01 | -0.08 | 0.83 |
| HILIC_POS_162_16479a4_796 | | 162.1648 | 4.80 | | HILIC | POS | | | 6586428 | 1.9E+07 | 5197107 | 1.8E+07 | 5.80E-01 | -0.08 | 0.52 |
| RP_POS_165_11255a10_368 | | 165.1126 | 10.37 | | RP | POS | | | 355935 | 150883 | 343221 | 181528 | 5.80E-01 | -0.08 | 0.62 |
| RP_POS_287_22165a9_927 | | 287.2217 | 9.93 | | RP | POS | | | 3556641 | 1754757 | 3420815 | 1826647 | 5.80E-01 | -0.08 | 0.52 |
| HILIC_POS_349_25778a3_19 | | 349.2578 | 3.19 | | HILIC | POS | | | 230531 | 115893 | 239958 | 131793 | 5.81E-01 | 0.08 | 0.75 |
| RP_POS_262_16464a0_682 | | 262.1646 | 0.68 | | RP | POS | | | 486934 | 160083 | 499052 | 160955 | 5.81E-01 | 0.08 | 0.64 |
| RP_POS_304_26144a14_239 | | 304.2614 | 14.24 | | RP | POS | | | 733648 | 374017 | 767779 | 509252 | 5.81E-01 | 0.08 | 0.7 |
| HILIC_NEG_253_79367a3_075 | | 253.7937 | 3.08 | | HILIC | NEG | | | 275997 | 22810.1 | 274231 | 23919.6 | 5.81E-01 | -0.08 | 0.63 |
| RP_POS_209_09662a8_351 | | 209.0966 | 8.35 | | RP | POS | | | 234750 | 118689 | 225745 | 119903 | 5.81E-01 | -0.08 | 0.82 |
| RP_POS_240_15692a10_575 | | 240.1569 | 10.58 | | RP | POS | | | 842592 | 419064 | 814251 | 333475 | 5.81E-01 | -0.08 | 0.47 |
| RP_POS_540_70093a10_325 | | 540.7009 | 10.33 | | RP | POS | | | 585073 | 343568 | 560155 | 317714 | 5.81E-01 | -0.08 | 0.63 |
| RP_POS_596_60944a14_052 | | 596.6094 | 14.05 | | RP | POS | | | 1E+07 | 1583528 | 1E+07 | 1498474 | 5.81E-01 | -0.08 | 1.21 |
| RP_NEG_127_87016a15_516 | | 127.8702 | 15.52 | | RP | NEG | | | 1130699 | 249569 | 1150708 | 278857 | 5.82E-01 | 0.08 | 0.42 |
| HILIC_NEG_134_89474a3_204 | | 134.8947 | 3.20 | | HILIC | NEG | | | 4.3E+08 | 2.3E+07 | 4.3E+08 | 3.1E+07 | 5.82E-01 | -0.08 | 0.75 |
| HILIC_POS_205_06833a4_867 | D-Mannitol | 205.0683 | 4.87 | | HILIC | POS | 205.0699(100) | 2 | 2065429 | 3167566 | 1775064 | 4346204 | 5.82E-01 | -0.08 | 0.55 |
| HILIC_NEG_236_81755a7_038 | | 236.8176 | 7.04 | | HILIC | NEG | | | 271989 | 123987 | 281626 | 132202 | 5.83E-01 | 0.08 | 0.5 |
| HILIC_POS_243_18298a1_465 | | 243.1830 | 1.47 | | HILIC | POS | | | 2561420 | 4624924 | 2925717 | 5035883 | 5.83E-01 | 0.08 | 0.54 |
| RP_POS_283_26309a10_448 | | 283.2631 | 10.45 | | RP | POS | | | 2650819 | 405114 | 2686858 | 534232 | 5.83E-01 | 0.08 | 0.71 |
| RP_POS_733_71069a11_328 | | 733.7107 | 11.33 | | RP | POS | | | 175164 | 92815.1 | 181765 | 83691.3 | 5.83E-01 | 0.07 | 0.52 |
| HILIC_POS_172_09421a4_23 | | 172.0942 | 4.23 | | HILIC | POS | | | 742195 | 600187 | 700001 | 526304 | 5.83E-01 | -0.07 | 1.14 |
| HILIC_POS_375_32153a4_289 | | 375.3215 | 4.29 | | HILIC | POS | | | 248637 | 130927 | 239545 | 111896 | 5.83E-01 | -0.07 | 0.69 |
| RP_POS_642_24738a10_323 | | 642.2474 | 10.32 | | RP | POS | | | 1151714 | 275375 | 1131426 | 266335 | 5.83E-01 | -0.07 | 0.59 |
| HILIC_POS_128_06841a4_229 | | 128.0684 | 4.23 | | HILIC | POS | | | 133494 | 51359 | 129115 | 63684.8 | 5.83E-01 | -0.08 | 0.85 |
| RP_POS_284_67685a10_489 | | 284.6769 | 10.49 | | RP | POS | | | 3930848 | 1219033 | 3834803 | 1328731 | 5.83E-01 | -0.08 | 1.03 |
| RP_POS_529_70435a10_03 | | 529.7044 | 10.03 | | RP | POS | | | 960962 | 557519 | 924657 | 410281 | 5.83E-01 | -0.08 | 0.61 |
| HILIC_POS_246_17014a1_263 | Valeryl-L-carnitine | 246.1701 | 1.26 | | HILIC | POS | 85.0293(100) 246.1697(40) | 1 | 1.1E+07 | 8322039 | 1.2E+07 | 8596894 | 5.83E-01 | 0.08 | 0.67 |
| RP_POS_369_35104a13_299 | Cholesterol | 369.3510 | 13.30 | | RP | POS | 369.3574(100), 161.133(70) | 1 | 7.8E+07 | 1.2E+07 | 7.9E+07 | 1.2E+07 | 5.84E-01 | 0.07 | 1.15 |
| HILIC_POS_380_30045a3_245 | | 380.3005 | 3.25 | | HILIC | POS | | | 182138 | 92538.7 | 189581 | 104834 | 5.84E-01 | 0.08 | 0.49 |
| HILIC_POS_414_1889a5_934 | | 414.1889 | 5.93 | | HILIC | POS | | | 93098.1 | 59933.8 | 97809.6 | 65274 | 5.84E-01 | 0.08 | 0.62 |
| RP_POS_604_90619a9_954 | | 604.9062 | 9.95 | | RP | POS | | | 2230142 | 1070426 | 2318912 | 1272932 | 5.84E-01 | 0.08 | 0.73 |
| HILIC_NEG_168_83163a3_199 | | 168.8316 | 3.20 | | HILIC | NEG | | | 3142993 | 125619 | 3152137 | 119008 | 5.84E-01 | 0.07 | 0.61 |
| HILIC_NEG_196_90141a3_194 | | 196.9014 | 3.19 | | HILIC | NEG | | | 252587 | 149576 | 263847 | 151159 | 5.84E-01 | 0.07 | 0.59 |
| RP_POS_162_0761a13_065 | | 162.0761 | 13.07 | | RP | POS | | | 2614777 | 1322813 | 2706453 | 1131236 | 5.84E-01 | 0.07 | 0.62 |
| RP_POS_731_60474a13_549 | N-Stearoyl-D-sphingomyelin | 731.6047 | 13.55 | | RP | POS | 184.0743(100), 86.0958(10) | 1 | 9814789 | 4601458 | 1E+07 | 4728501 | 5.84E-01 | 0.08 | 0.8 |
| RP_NEG_642_41736a11_141 | | 642.4174 | 11.14 | | RP | NEG | | | 369761 | 44777.6 | 366548 | 41307.5 | 5.84E-01 | -0.07 | 0.84 |
| RP_POS_246_16971a4_643 | | 246.1697 | 4.64 | | RP | POS | | | 128436 | 72350.7 | 123166 | 68733.8 | 5.84E-01 | -0.07 | 1.02 |
| RP_POS_334_30786a11_59 | | 334.3079 | 11.59 | | RP | POS | | | 1242826 | 374831 | 1216211 | 338883 | 5.84E-01 | -0.07 | 0.42 |
| RP_POS_446_34656a10_766 | | 446.3466 | 10.77 | | RP | POS | | | 381537 | 51626.8 | 377649 | 52172.9 | 5.84E-01 | -0.07 | 0.62 |
| RP_POS_448_76031a13_794 | | 448.7603 | 13.79 | | RP | POS | | | 425295 | 308262 | 403410 | 277632 | 5.84E-01 | -0.07 | 0.46 |
| RP_POS_682_47461a11_477 | | 682.4746 | 11.48 | | RP | POS | | | 179735 | 114779 | 172041 | 91503.7 | 5.84E-01 | -0.07 | 0.59 |
| RP_POS_195_10191a8_496 | | 195.1019 | 8.50 | | RP | POS | | | 1.9E+07 | 801483 | 1.9E+07 | 867512 | 5.84E-01 | -0.08 | 0.79 |
| HILIC_NEG_128_0354a4_248 | Pyroglutamic acid | 128.0354 | 4.25 | | HILIC | NEG | 128.0358(100) | 1 | 9164099 | 1656894 | 9034497 | 1787724 | 5.84E-01 | -0.08 | 0.63 |
| RP_POS_297_16391a9_817 | | 297.1639 | 9.82 | | RP | POS | | | 580012 | 198240 | 595427 | 212699 | 5.85E-01 | 0.08 | 0.87 |

**Supplemantary table 1**: The non-targeted metabolomics results ordered by the VIP value (p value < 0.0003 are shaded dark green, p value < 0.05 are shaded light green, d values below 0 are blue and above 0 are red)

| | | | | | | | | | Cases | | Controls | | Welch test | Cohen's | |
|---|---|---|---|---|---|---|---|---|---|---|---|---|---|---|---|
| Feature_ID | Metabolite_name | Average_Mz | Average_RT_min_ | HMDB ID | RP_HILIC | POS_NEG | Cureted_MSMS | ID_level | Mean | SD | Mean2 | SD3 | p | d | VIP |
| RP_NEG_172_83073a11_14 | | 172.8307 | 11.14 | | RP | NEG | | | 93265.4 | 34365.1 | 95559.8 | 27219.4 | 5.85E-01 | 0.07 | 0.69 |
| RP_NEG_588_89862a11_326 | | 588.8986 | 11.33 | | RP | NEG | | | 553808 | 137987 | 563377 | 118835 | 5.85E-01 | 0.07 | 0.52 |
| RP_POS_502_83875a9_596 | | 502.8388 | 9.60 | | RP | POS | | | 1175419 | 744115 | 1228248 | 674538 | 5.85E-01 | 0.07 | 1.18 |
| RP_POS_149_02359a7_47 | | 149.0236 | 7.47 | | RP | POS | 149.023(100),121.0286(20) | | 1.2E+07 | 1628744 | 1.2E+07 | 1103591 | 5.85E-01 | -0.07 | 0.61 |
| HILIC_POS_388_31647a4_277 | | 388.3165 | 4.28 | | HILIC | POS | | | 123367 | 74462.2 | 118453 | 57283.5 | 5.85E-01 | -0.07 | 0.45 |
| RP_NEG_214_78334a14_341 | | 214.7833 | 14.34 | | RP | NEG | | | 824146 | 282141 | 802944 | 284716 | 5.85E-01 | -0.07 | 0.47 |
| RP_POS_316_28476a10_282 | | 316.2848 | 10.28 | | RP | POS | | | 1156672 | 215583 | 1140789 | 209975 | 5.85E-01 | -0.07 | 0.98 |
| RP_POS_532_33868a10_66 | | 532.3387 | 10.66 | | RP | POS | | | 6051365 | 2187609 | 5894998 | 2006404 | 5.85E-01 | -0.07 | 0.62 |
| RP_POS_526_2934a10_283 | | 526.2934 | 10.28 | | RP | POS | | | 5.3E+07 | 1.4E+07 | 5.1E+07 | 1.6E+07 | 5.85E-01 | -0.08 | 0.6 |
| RP_POS_716_52209a13_132 | | 716.5221 | 13.13 | | RP | POS | | | 4027382 | 2627489 | 4264167 | 3573159 | 5.86E-01 | 0.08 | 0.84 |
| HILIC_POS_184_07365a1_242 | | 184.0737 | 1.24 | | HILIC | POS | 184.0746(100), 86.0968(40) | | 1160148 | 342834 | 1185631 | 340978 | 5.86E-01 | 0.07 | 0.58 |
| RP_NEG_460_55887a15_495 | | 460.5589 | 15.50 | | RP | NEG | | | 617019 | 52100.1 | 621015 | 55051.7 | 5.86E-01 | 0.07 | 0.85 |
| HILIC_NEG_115_92078a3_191 | | 115.9208 | 3.19 | | HILIC | NEG | | | 2.3E+07 | 791372 | 2.3E+07 | 767389 | 5.86E-01 | -0.07 | 0.5 |
| RP_POS_319_28433a10_518 | | 319.2843 | 10.52 | | RP | POS | | | 1175819 | 192499 | 1161396 | 194877 | 5.86E-01 | -0.07 | 0.86 |
| RP_POS_508_33975a10_573 | | 508.3398 | 10.57 | | RP | POS | | | 3459686 | 1332185 | 3369091 | 1106429 | 5.86E-01 | -0.07 | 0.51 |
| RP_NEG_498_29034a9_16 | | 498.2903 | 9.16 | | RP | NEG | 498.2897(100), 124.0081(2) | | 6582313 | 9325906 | 6014680 | 5779357 | 5.86E-01 | -0.08 | 0.53 |
| RP_NEG_433_2363a10_899 | | 433.2363 | 10.90 | | RP | NEG | | | 670780 | 690681 | 727727 | 825123 | 5.87E-01 | 0.08 | 0.78 |
| RP_POS_403_23273a9_958 | | 403.2327 | 9.96 | | RP | POS | | | 2714599 | 2450379 | 2917793 | 2956017 | 5.87E-01 | 0.08 | 0.7 |
| RP_POS_196_00299a15_559 | | 196.0030 | 15.56 | | RP | POS | | | 4.8E+07 | 2.3E+07 | 5E+07 | 2.3E+07 | 5.87E-01 | 0.07 | 0.64 |
| RP_NEG_324_97302a6_951 | | 324.9730 | 6.95 | | RP | NEG | | | 446941 | 24012.9 | 445059 | 26385.4 | 5.87E-01 | -0.07 | 0.74 |
| RP_POS_304_24823a9_927 | | 304.2482 | 9.93 | | RP | POS | | | 5927417 | 2910825 | 5704962 | 3060293 | 5.87E-01 | -0.07 | 0.52 |
| RP_POS_407_24564a11_594 | | 407.2456 | 11.59 | | RP | POS | | | 1527844 | 256855 | 1510148 | 221776 | 5.87E-01 | -0.07 | 0.83 |
| RP_POS_428_83038a15_386 | | 428.8304 | 15.39 | | RP | POS | | | 753387 | 960056 | 684391 | 901963 | 5.87E-01 | -0.07 | 0.62 |
| RP_NEG_263_12912a9_386 | | 263.1291 | 9.39 | | RP | NEG | | | 150590 | 155160 | 142328 | 48955.5 | 5.87E-01 | -0.08 | 0.4 |
| RP_POS_363_21649a9_723 | | 363.2165 | 9.72 | | RP | POS | | | 839813 | 399066 | 869291 | 397257 | 5.88E-01 | 0.07 | 1.02 |
| RP_POS_543_34479a10_926 | | 543.3448 | 10.93 | | RP | POS | | | 1042511 | 478601 | 1082321 | 583320 | 5.88E-01 | 0.07 | 0.8 |
| RP_POS_257_15143a9_827 | | 257.1514 | 9.83 | | RP | POS | 257.1511(100) | | 2756455 | 271538 | 2736594 | 264421 | 5.88E-01 | -0.07 | 1.12 |
| HILIC_NEG_172_8425a3_092 | | 172.8425 | 3.09 | | HILIC | NEG | | | 1220462 | 102706 | 1212342 | 115144 | 5.88E-01 | -0.07 | 0.51 |
| HILIC_NEG_200_85924a5_306 | | 200.8592 | 5.31 | | HILIC | NEG | | | 5565468 | 2127759 | 5408199 | 2114973 | 5.88E-01 | -0.07 | 0.61 |
| RP_POS_601_435a11_217 | | 601.4350 | 11.22 | | RP | POS | | | 610613 | 598888 | 562964 | 677729 | 5.88E-01 | -0.07 | 0.73 |
| RP_POS_747_5332a10_636 | | 747.5332 | 10.64 | | RP | POS | | | 551169 | 344715 | 525879 | 338204 | 5.88E-01 | -0.07 | 0.83 |
| RP_NEG_311_16928a11_592 | | 311.1693 | 11.59 | | RP | NEG | 311.1698(100) | | 1059408 | 202606 | 1041885 | 262041 | 5.88E-01 | -0.08 | 0.4 |
| RP_POS_158_02721a4_82 | | 158.0272 | 4.82 | | RP | POS | | | 3.4E+07 | 1.1E+07 | 3.5E+07 | 1.1E+07 | 5.89E-01 | 0.07 | 0.57 |
| RP_POS_170_06017a2_883 | | 170.0602 | 2.88 | | RP | POS | | | 3752488 | 702211 | 3804027 | 693886 | 5.89E-01 | 0.07 | 0.7 |
| RP_POS_320_25824a10_692 | | 320.2582 | 10.69 | | RP | POS | | | 1210854 | 679050 | 1260457 | 663647 | 5.89E-01 | 0.07 | 0.93 |
| RP_POS_435_31216a8_601 | | 435.3122 | 8.60 | | RP | POS | | | 379843 | 248469 | 399239 | 273678 | 5.89E-01 | 0.07 | 0.58 |
| HILIC_NEG_183_93452a5_655 | | 183.9345 | 5.66 | | HILIC | NEG | | | 2828212 | 409276 | 2800441 | 344165 | 5.89E-01 | -0.07 | 0.52 |
| RP_NEG_136_89166a11_143 | | 136.8917 | 11.14 | | RP | NEG | | | 385959 | 46066.5 | 382642 | 43909.1 | 5.89E-01 | -0.07 | 0.69 |
| RP_POS_162_07619a14_206 | | 162.0762 | 14.21 | | RP | POS | | | 2921656 | 1374768 | 2823544 | 1287190 | 5.89E-01 | -0.07 | 0.76 |
| RP_POS_510_75919a4_666 | | 510.7592 | 4.67 | | RP | POS | | | 6509161 | 3699078 | 6236540 | 3675171 | 5.89E-01 | -0.07 | 1.27 |
| RP_POS_524_3736a10_329 | | 524.3736 | 10.33 | | RP | POS | | | 1159433 | 1575103 | 1059676 | 1124296 | 5.89E-01 | -0.07 | 0.38 |
| RP_POS_586_30817a10_518 | | 586.3082 | 10.52 | | RP | POS | | | 1989534 | 243416 | 1970979 | 257683 | 5.89E-01 | -0.07 | 0.62 |
| RP_POS_399_18536a9_224 | | 399.1854 | 9.22 | | RP | POS | | | 243792 | 406086 | 322344 | 1397029 | 5.90E-01 | 0.09 | 0.55 |
| RP_NEG_714_27887a10_612 | | 714.2789 | 10.61 | | RP | NEG | | | 491407 | 76687.7 | 497409 | 85430.7 | 5.90E-01 | 0.07 | 0.64 |
| RP_POS_254_85654a15_41 | | 254.8565 | 15.41 | | RP | POS | | | 331249 | 183497 | 344176 | 168678 | 5.90E-01 | 0.07 | 0.84 |
| RP_POS_533_36664a9_482 | | 533.3666 | 9.48 | | RP | POS | | | 266884 | 207207 | 282522 | 216780 | 5.90E-01 | 0.07 | 0.77 |
| RP_POS_553_87158a9_784 | | 553.8716 | 9.78 | | RP | POS | | | 1985016 | 1172256 | 2067085 | 1063686 | 5.90E-01 | 0.07 | 0.95 |
| RP_POS_564_41168a15_065 | | 564.4117 | 15.07 | | RP | POS | | | 9773595 | 2771431 | 9993028 | 3145312 | 5.90E-01 | 0.07 | 0.6 |
| RP_POS_731_41626a12_617 | | 731.4163 | 12.62 | | RP | POS | | | 513585 | 853299 | 582280 | 992757 | 5.90E-01 | 0.07 | 0.65 |
| RP_POS_569_43152a15_186 | | 569.4315 | 15.19 | | RP | POS | | | 5.6E+07 | 1.3E+07 | 5.5E+07 | 1.2E+07 | 5.90E-01 | -0.07 | 0.87 |
| RP_NEG_195_85481a11_589 | | 195.8548 | 11.59 | | RP | NEG | | | 123212 | 66594.9 | 128358 | 72726.4 | 5.91E-01 | 0.07 | 0.66 |
| RP_POS_562_32629a10_929 | | 562.3263 | 10.93 | | RP | POS | | | 6679195 | 1232997 | 6776996 | 1410089 | 5.91E-01 | 0.07 | 0.82 |
| RP_POS_570_05066a10_286 | | 570.0507 | 10.29 | | RP | POS | | | 530553 | 316139 | 553468 | 308174 | 5.91E-01 | 0.07 | 0.89 |
| RP_NEG_1121_14331a9_737 | | 1121.1433 | 9.74 | | RP | NEG | | | 1916215 | 97761.2 | 1909405 | 88176.6 | 5.91E-01 | -0.07 | 0.74 |
| RP_POS_318_90829a15_399 | | 318.9083 | 15.40 | | RP | POS | | | 1169237 | 567358 | 1127116 | 577405 | 5.91E-01 | -0.07 | 0.85 |
| RP_POS_698_28998a11_396 | | 698.2900 | 11.40 | | RP | POS | | | 253449 | 172285 | 240820 | 171568 | 5.91E-01 | -0.07 | 0.62 |
| RP_POS_269_22647a10_842 | Vitamin A acetate | 269.2265 | 10.84 | | RP | POS | 269.2253(100), 93.0705(80) | 1 | 5.7E+07 | 1E+07 | 5.6E+07 | 1.1E+07 | 5.91E-01 | -0.07 | 0.4 |
| HILIC_NEG_244_98534a5_11 | | 244.9853 | 5.11 | | HILIC | NEG | | | 617157 | 288311 | 639579 | 320361 | 5.92E-01 | 0.07 | 0.44 |
| HILIC_POS_245_98276a1_347 | | 245.9828 | 1.35 | | HILIC | POS | | | 396580 | 142801 | 406870 | 138103 | 5.92E-01 | 0.07 | 0.61 |
| RP_NEG_145_01404a0_673 | | 145.0140 | 0.67 | | RP | NEG | | | 430767 | 157441 | 442979 | 173752 | 5.92E-01 | 0.07 | 0.49 |
| RP_POS_246_16946a5_228 | | 246.1695 | 5.23 | | RP | POS | | | 289726 | 140055 | 300230 | 145959 | 5.92E-01 | 0.07 | 0.75 |
| RP_NEG_369_17419a11_292 | | 369.1742 | 11.29 | | RP | NEG | | | 393930 | 66895.7 | 389246 | 61137.6 | 5.92E-01 | -0.07 | 0.76 |
| RP_POS_501_3515a8_653 | | 501.3515 | 8.65 | | RP | POS | | | 420514 | 290528 | 400562 | 255472 | 5.92E-01 | -0.07 | 0.7 |
| RP_POS_255_11264a5_754 | | 255.1126 | 5.75 | | RP | POS | | | 2.3E+07 | 5.2E+07 | 2E+07 | 1.9E+07 | 5.92E-01 | -0.08 | 0.47 |
| RP_POS_206_08144a4_822 | Indolelactic acid | 206.0814 | 4.82 | | RP | POS | 188.0705(100), 160.0766(98) | 1 | 3992303 | 1031603 | 3924466 | 824353 | 5.92E-01 | -0.07 | 0.63 |
| HILIC_POS_142_08636a1_291 | | 142.0864 | 1.29 | | HILIC | POS | | | 1361657 | 1832803 | 1492513 | 1753552 | 5.93E-01 | 0.07 | 0.48 |
| HILIC_POS_153_09097a3_103 | | 153.0910 | 3.10 | | HILIC | POS | | | 141042 | 112485 | 149951 | 129289 | 5.93E-01 | 0.07 | 0.86 |
| RP_POS_185_15363a6_654 | | 185.1536 | 6.65 | | RP | POS | | | 551290 | 89463.8 | 558246 | 99743.2 | 5.93E-01 | 0.07 | 1.12 |
| RP_POS_235_16943a8_702 | | 235.1694 | 8.70 | | RP | POS | | | 3699200 | 243505 | 3719132 | 294803 | 5.93E-01 | 0.07 | 0.57 |
| RP_POS_522_28644a9_274 | | 522.2864 | 9.27 | | RP | POS | | | 597870 | 1194651 | 700865 | 1563836 | 5.93E-01 | 0.07 | 0.63 |
| RP_POS_607_75574a10_122 | | 607.7557 | 10.12 | | RP | POS | | | 395521 | 215408 | 412347 | 242487 | 5.93E-01 | 0.07 | 0.71 |
| HILIC_NEG_121_02942a0_933 | | 121.0294 | 0.93 | | HILIC | NEG | | | 263007 | 73152.9 | 257680 | 72597.9 | 5.93E-01 | -0.07 | 0.63 |
| HILIC_NEG_345_71832a1_406 | | 345.7183 | 1.41 | | HILIC | NEG | | | 212772 | 42697.7 | 209926 | 35538.5 | 5.93E-01 | -0.07 | 0.35 |
| RP_NEG_872_56427a13_803 | | 872.5643 | 13.80 | | RP | NEG | | | 1.3E+07 | 3551958 | 1.3E+07 | 3859461 | 5.93E-01 | -0.07 | 0.62 |
| RP_POS_297_27927a10_987 | | 297.2793 | 10.99 | | RP | POS | | | 1179524 | 141048 | 1168748 | 152699 | 5.93E-01 | -0.07 | 1 |
| RP_NEG_315_95004a0_916 | | 315.9500 | 0.92 | | RP | NEG | | | 391037 | 75696.2 | 396662 | 78474.4 | 5.94E-01 | 0.07 | 0.35 |
| RP_NEG_539_24872a7_054 | | 539.2487 | 7.05 | | RP | NEG | | | 1536891 | 685763 | 1582014 | 554996 | 5.94E-01 | 0.07 | 0.51 |
| RP_POS_320_20068a10_076 | | 320.2007 | 10.08 | | RP | POS | | | 293871 | 245560 | 311459 | 237676 | 5.94E-01 | 0.07 | 0.69 |
| RP_POS_449_15292a8_516 | | 449.1529 | 8.52 | | RP | POS | | | 1704208 | 865592 | 1767117 | 860296 | 5.94E-01 | 0.07 | 0.37 |
| RP_NEG_424_91098a15_324 | | 424.9110 | 15.32 | | RP | NEG | | | 429356 | 36027.8 | 426779 | 34762.2 | 5.94E-01 | -0.07 | 0.77 |
| RP_POS_123_08081a6_569 | | 123.0808 | 6.57 | | RP | POS | | | 2192280 | 746009 | 2135012 | 816108 | 5.94E-01 | -0.07 | 0.66 |
| HILIC_POS_264_19565a0_699 | | 264.1957 | 0.70 | | HILIC | POS | | | 2184517 | 2.4E+07 | 925787 | 9111883 | 5.94E-01 | -0.08 | 0.35 |
| RP_POS_643_28351a11_455 | | 643.2835 | 11.46 | | RP | POS | | | 706712 | 1420750 | 812810 | 1490627 | 5.95E-01 | 0.07 | 0.62 |
| HILIC_NEG_94_92501a3_198 | | 94.9250 | 3.20 | | HILIC | NEG | | | 1648975 | 219636 | 1629727 | 297460 | 5.95E-01 | -0.07 | 0.82 |
| RP_NEG_365_13992a9_974 | | 365.1399 | 9.97 | | RP | NEG | | | 121828 | 187722 | 108849 | 170371 | 5.95E-01 | -0.07 | 0.66 |
| RP_NEG_503_54669a15_496 | | 503.5467 | 15.50 | | RP | NEG | | | 606763 | 43269.3 | 603472 | 46942.7 | 5.95E-01 | -0.07 | 0.59 |
| RP_POS_201_1637a10_841 | | 201.1637 | 10.84 | | RP | POS | | | 459485 | 115159 | 451279 | 110936 | 5.95E-01 | -0.07 | 0.46 |
| RP_POS_236_07166a8_513 | | 236.0717 | 8.51 | | RP | POS | | | 7.8E+07 | 4.1E+07 | 7.5E+07 | 3.6E+07 | 5.95E-01 | -0.07 | 0.64 |
| RP_POS_369_29779a10_873 | | 369.2978 | 10.87 | | RP | POS | | | 4627826 | 885162 | 4566303 | 810750 | 5.95E-01 | -0.07 | 1.13 |
| HILIC_POS_126_0663a1_139 | 5-Methylcytosine | 126.0663 | 1.14 | | HILIC | POS | 81.0457(100), 126.066(60) | 1 | 1212818 | 852873 | 1150571 | 860222 | 5.95E-01 | -0.07 | 0.58 |
| HILIC_NEG_200_86041a5_623 | | 200.8604 | 5.62 | | HILIC | NEG | | | 2443656 | 180278 | 2465476 | 373501 | 5.96E-01 | 0.08 | 0.7 |
| HILIC_NEG_100_93363a2_276 | | 100.9336 | 2.28 | | HILIC | NEG | | | 1.9E+07 | 2972451 | 1.9E+07 | 3620087 | 5.96E-01 | 0.07 | 0.44 |
| RP_POS_1058_26501a9_917 | | 1058.2650 | 9.92 | | RP | POS | | | 1387351 | 199931 | 1402053 | 205409 | 5.96E-01 | 0.07 | 0.71 |
| RP_POS_261_22159a10_579 | | 261.2216 | 10.58 | | RP | POS | | | 3168930 | 524410 | 3207901 | 547038 | 5.96E-01 | 0.07 | 0.39 |
| HILIC_NEG_248_96037a4_352 | | 248.9604 | 4.35 | | HILIC | NEG | | | 8.5E+07 | 4867120 | 8.4E+07 | 4932833 | 5.96E-01 | -0.07 | 0.86 |
| RP_POS_167_1429a6_654 | | 167.1429 | 6.65 | | RP | POS | | | 451902 | 87044.3 | 445607 | 86441.3 | 5.96E-01 | -0.07 | 0.92 |
| RP_POS_387_16238a10_896 | | 387.1624 | 10.90 | | RP | POS | | | 464111 | 257009 | 445762 | 249364 | 5.96E-01 | -0.07 | 0.91 |
| RP_POS_470_35068a10_063 | | 470.3507 | 10.06 | | RP | POS | | | 5539322 | 2312527 | 5380032 | 2099696 | 5.96E-01 | -0.07 | 0.69 |
| RP_POS_713_29645a11_281 | | 713.2965 | 11.28 | | RP | POS | | | 151660 | 96750 | 144973 | 88058.9 | 5.96E-01 | -0.07 | 0.58 |
| HILIC_NEG_241_01045a1_265 | | 241.0105 | 1.27 | | HILIC | NEG | | | 623451 | 253155 | 642204 | 265021 | 5.97E-01 | 0.07 | 0.58 |
| RP_POS_159_96935a0_955 | | 159.9694 | 0.96 | | RP | POS | | | 4881631 | 3200849 | 5133191 | 3699213 | 5.97E-01 | 0.07 | 0.34 |
| RP_POS_236_07143a7_303 | | 236.0714 | 7.30 | | RP | POS | | | 5.2E+07 | 1.8E+07 | 5.3E+07 | 1.7E+07 | 5.97E-01 | 0.07 | 0.38 |
| RP_POS_558_29059a7_054 | | 558.2906 | 7.05 | | RP | POS | | | 741970 | 354505 | 765089 | 287271 | 5.97E-01 | 0.07 | 0.54 |
| RP_POS_577_4057a10_326 | | 577.4057 | 10.33 | | RP | POS | | | 938047 | 483809 | 981337 | 679761 | 5.97E-01 | 0.07 | 0.8 |
| RP_POS_392_80939a15_413 | | 392.8094 | 15.41 | | RP | POS | | | 599453 | 375793 | 571729 | 389075 | 5.97E-01 | -0.07 | 0.74 |
| HILIC_NEG_100_93358a7_09 | | 100.9336 | 7.09 | | HILIC | NEG | | | 1.6E+07 | 3937692 | 1.7E+07 | 3428500 | 5.98E-01 | 0.07 | 0.6 |
| RP_NEG_314_90048a15_374 | | 314.9005 | 15.37 | | RP | NEG | | | 519579 | 194607 | 533419 | 189265 | 5.98E-01 | 0.07 | 0.4 |
| RP_POS_1045_62488a10_295 | | 1045.6249 | 10.30 | | RP | POS | | | 1736073 | 768327 | 1791564 | 769998 | 5.98E-01 | 0.07 | 0.6 |
| RP_POS_589_84277a11_321 | | 589.8428 | 11.32 | | RP | POS | | | 169879 | 101332 | 177089 | 98854.9 | 5.98E-01 | 0.07 | 0.46 |
| RP_POS_284_2944a11_15 | 1-Desoxymethylsphinganine | 284.2944 | 11.15 | | RP | POS | 284.2942(100) | 2 | 2.6E+07 | 6839352 | 2.6E+07 | 7277785 | 5.98E-01 | -0.07 | 1.03 |
| HILIC_POS_205_06821a5_023 | | 205.0682 | 5.02 | | HILIC | POS | | | 451848 | 668338 | 403912 | 662766 | 5.98E-01 | -0.07 | 0.4 |
| RP_NEG_400_15549a11_142 | | 400.1555 | 11.14 | | RP | NEG | | | 855509 | 69043 | 850577 | 67913.9 | 5.98E-01 | -0.07 | 0.55 |
| RP_NEG_812_58246a13_677 | | 812.5825 | 13.68 | | RP | NEG | | | 9201772 | 2110159 | 9036628 | 2428158 | 5.98E-01 | -0.07 | 0.52 |
| RP_POS_720_58954a14_373 | | 720.5895 | 14.37 | | RP | POS | | | 229355 | 225498 | 212690 | 235828 | 5.98E-01 | -0.07 | 0.57 |
| RP_POS_237_10234a6_833 | | 237.1023 | 6.83 | | RP | POS | 194.0958(100), 237.1033(60) | | 66158 | 115140 | 60281.6 | 31431.2 | 5.98E-01 | -0.08 | 0.44 |
| RP_POS_280_13901a0_679 | | 280.1390 | 0.68 | | RP | POS | | | 606929 | 218047 | 622796 | 223276 | 5.99E-01 | 0.07 | 0.66 |
| RP_POS_314_02933a10_495 | | 314.0293 | 10.50 | | RP | POS | | | 1156082 | 592786 | 1204553 | 734505 | 5.99E-01 | 0.07 | 0.48 |
| HILIC_NEG_192_9894a0_934 | | 192.9894 | 0.93 | | HILIC | NEG | | | 627808 | 68744.7 | 622809 | 70047.2 | 5.99E-01 | -0.07 | 0.6 |
| HILIC_NEG_373_01028a0_934 | | 373.0103 | 0.93 | | HILIC | NEG | | | 572476 | 63229 | 567510 | 73516.1 | 5.99E-01 | -0.07 | 0.49 |
| RP_POS_205_12202a15_175 | | 205.1220 | 15.18 | | RP | POS | | | 592624 | 324220 | 566558 | 391604 | 5.99E-01 | -0.07 | 0.53 |
| RP_POS_770_46204a10_487 | | 770.4620 | 10.49 | | RP | POS | | | 2147453 | 430527 | 2113500 | 504518 | 5.99E-01 | -0.07 | 0.84 |
| HILIC_NEG_182_03217a2_031 | | 182.0322 | 2.03 | | HILIC | NEG | | | 218300 | 45301.9 | 221557 | 45483.1 | 6.00E-01 | 0.07 | 0.45 |
| RP_NEG_312_00223a2_162 | | 312.0022 | 2.16 | | RP | NEG | | | 966919 | 153618 | 978132 | 158754 | 6.00E-01 | 0.07 | 0.64 |
| RP_NEG_316_94803a13_899 | | 316.9480 | 13.90 | | RP | NEG | | | 1646017 | 776953 | 1699010 | 703245 | 6.00E-01 | 0.07 | 0.33 |
| RP_POS_534_29547a10_357 | | 534.2955 | 10.36 | | RP | POS | | | 3438767 | 621294 | 3485791 | 684245 | 6.00E-01 | 0.07 | 0.85 |
| HILIC_POS_64_01669a5_077 | | 64.0167 | 5.08 | | HILIC | POS | | | 9771758 | 5293461 | 9412092 | 4761468 | 6.00E-01 | -0.07 | 0.58 |
| RP_POS_179_90273a15_435 | | 179.9027 | 15.44 | | RP | POS | | | 814353 | 428876 | 783466 | 431938 | 6.00E-01 | -0.07 | 0.58 |
| RP_POS_181_03807a15_382 | | 181.0381 | 15.38 | | RP | POS | | | 6093319 | 2454960 | 5920658 | 2372690 | 6.00E-01 | -0.07 | 0.34 |
| RP_POS_275_07895a6_13 | | 275.0790 | 6.13 | | RP | POS | | | 2412627 | 6358610 | 2071785 | 2737922 | 6.00E-01 | -0.07 | 0.67 |
| RP_POS_758_56726a15_458 | PC 34:2 | 758.5673 | 15.46 | | RP | POS | 184.0746(100) | 1 | 751788 | 316723 | 774633 | 320130 | 6.00E-01 | 0.07 | 0.42 |
| HILIC_POS_232_19026a0_909 | | 232.1903 | 0.91 | | HILIC | POS | | | 234537 | 31751.8 | 236884 | 33752.3 | 6.01E-01 | 0.07 | 0.75 |
| RP_POS_153_99802a0_732 | | 153.9980 | 0.73 | | RP | POS | | | 534741 | 223812 | 550170 | 208380 | 6.01E-01 | 0.07 | 0.45 |
| RP_POS_426_97736a10_121 | | 426.9774 | 10.12 | | RP | POS | | | 168288 | 94979.2 | 175366 | 102413 | 6.01E-01 | 0.07 | 0.68 |
| HILIC_NEG_316_94724a4_352 | | 316.9472 | 4.35 | | HILIC | NEG | | | 5.6E+07 | 3473178 | 5.6E+07 | 3436763 | 6.01E-01 | -0.07 | 0.86 |
| HILIC_NEG_394_05487a6_135 | | 394.0549 | 6.14 | | HILIC | NEG | | | 193618 | 28619.3 | 191630 | 27123.8 | 6.01E-01 | -0.07 | 0.79 |
| HILIC_POS_368_22003a3_042 | | 368.2200 | 3.04 | | HILIC | POS | | | 443284 | 157527 | 432189 | 152866 | 6.01E-01 | -0.07 | 0.81 |
| RP_NEG_201_88934a14_348 | | 201.8893 | 14.35 | | RP | NEG | | | 1098114 | 447167 | 1064702 | 482890 | 6.01E-01 | -0.07 | 0.44 |
| RP_NEG_224_81195a15_45 | | 224.8120 | 15.45 | | RP | NEG | | | 707440 | 103924 | 698775 | 133867 | 6.01E-01 | -0.07 | 0.67 |
| RP_POS_153_091a10_253 | | 153.0910 | 10.25 | | RP | POS | | | 1147300 | 149379 | 1136321 | 156732 | 6.01E-01 | -0.07 | 0.64 |
| RP_POS_385_33069a12_34 | | 385.3307 | 12.34 | | RP | POS | | | 143725 | 81514.4 | 137968 | 79548.1 | 6.01E-01 | -0.07 | 0.61 |
| RP_POS_150_05853a0_679 | L-Methionine | 150.0585 | 0.68 | | RP | POS | 104.0534(100), 56.0507(70), 133.0322(60) | 2 | 2.9E+07 | 6907225 | 2.8E+07 | 6368917 | 6.01E-01 | -0.07 | 0.54 |
| RP_POS_389_33676a10_358 | | 389.3368 | 10.36 | | RP | POS | | | 504640 | 165408 | 517724 | 197751 | 6.02E-01 | 0.07 | 0.95 |
| RP_POS_560_31085a10_608 | | 560.3109 | 10.61 | | RP | POS | | | 6189444 | 1114023 | 6271626 | 1183681 | 6.02E-01 | 0.07 | 0.74 |
| RP_POS_640_94879a11_421 | | 640.9488 | 11.42 | | RP | POS | | | 276556 | 136025 | 287906 | 176520 | 6.02E-01 | 0.07 | 0.6 |
| HILIC_POS_210_10988a4_026 | | 210.1099 | 4.03 | | HILIC | POS | | | 42682.3 | 25418.7 | 40898.4 | 24632.7 | 6.02E-01 | -0.07 | 0.51 |
| RP_POS_248_97633a15_359 | | 248.9763 | 15.36 | | RP | POS | | | 339264 | 197591 | 325440 | 190587 | 6.02E-01 | -0.07 | 0.45 |
| RP_POS_603_79077a4_406 | | 603.7908 | 4.41 | | RP | POS | | | 1959242 | 2366920 | 1809849 | 1832901 | 6.02E-01 | -0.07 | 1.04 |
| RP_NEG_558_99774a0_678 | | 558.9977 | 0.68 | | RP | NEG | | | 301004 | 116373 | 309157 | 112880 | 6.03E-01 | 0.07 | 0.43 |
| RP_POS_278_24789a10_565 | | 278.2479 | 10.57 | | RP | POS | | | 230104 | 241999 | 248057 | 261087 | 6.03E-01 | 0.07 | 0.45 |
| RP_POS_505_68689a9_725 | | 505.6869 | 9.73 | | RP | POS | | | 223429 | 134318 | 235341 | 191083 | 6.03E-01 | 0.07 | 0.65 |
| RP_POS_832_58411a12_971 | | 832.5841 | 12.97 | | RP | POS | | | 4764977 | 3104073 | 5013952 | 3806766 | 6.03E-01 | 0.07 | 0.81 |
| RP_NEG_413_20074a6_872 | | 413.2007 | 6.87 | | RP | NEG | | | 564920 | 1384384 | 479905 | 1006997 | 6.03E-01 | -0.07 | 0.43 |
| RP_POS_139_05479a15_513 | | 139.0548 | 15.51 | | RP | POS | | | 1.2E+07 | 1318283 | 1.2E+07 | 1478493 | 6.03E-01 | -0.07 | 0.93 |
| RP_POS_184_54019a0_707 | | 184.5402 | 0.71 | | RP | POS | | | 1643583 | 613602 | 1600844 | 589359 | 6.03E-01 | -0.07 | 0.58 |
| RP_POS_229_14319a4_643 | | 229.1432 | 4.64 | | RP | POS | | | 106946 | 65787.6 | 102446 | 60912.9 | 6.03E-01 | -0.07 | 1.06 |
| RP_POS_304_26093a10_966 | | 304.2609 | 10.97 | | RP | POS | | | 6126278 | 2810616 | 5949849 | 2161619 | 6.03E-01 | -0.07 | 0.42 |
| RP_POS_576_39563a6_927 | | 576.3956 | 6.93 | | RP | POS | | | 413955 | 293247 | 394518 | 254641 | 6.03E-01 | -0.07 | 0.86 |
| RP_POS_602_29498a10_614 | | 602.2950 | 10.61 | | RP | POS | | | 543049 | 273114 | 524164 | 259188 | 6.03E-01 | -0.07 | 0.68 |
| RP_POS_701_40833a13_696 | | 701.4083 | 13.70 | | RP | POS | | | 8760594 | 2155762 | 8604315 | 2228128 | 6.03E-01 | -0.07 | 1.01 |
| RP_POS_320_23511a10_833 | | 320.2351 | 10.83 | | RP | POS | | | 961591 | 416433 | 1010461 | 853913 | 6.04E-01 | 0.08 | 0.72 |
| HILIC_NEG_214_07707a0_94 | | 214.0771 | 0.94 | | HILIC | NEG | | | 128816 | 117263 | 136950 | 112363 | 6.04E-01 | 0.07 | 0.64 |
| HILIC_POS_152_03177a6_136 | | 152.0318 | 6.14 | | HILIC | POS | | | 1641699 | 172172 | 1653992 | 173812 | 6.04E-01 | 0.07 | 0.73 |

**Supplemantary table 1**: The non-targeted metabolomics results ordered by the VIP value (p value < 0.0003 are shaded dark green, p value < 0.05 are shaded light green, d values below 0 are blue and above 0 are red)

| | | | | | | | | | Cases | | Controls | | Welch test | Cohen's | |
|---|---|---|---|---|---|---|---|---|---|---|---|---|---|---|---|
| Feature_ID | Metabolite_name | Average_Mz | Average_RT_min_ | HMDB ID | RP_HILIC | POS_NEG | Cureted_MSMS | ID_level | Mean | SD | Mean2 | SD3 | p | d | VIP |
| RP_POS_1148_34094a13_214 | | 1148.3409 | 13.21 | | RP | POS | | | 6924275 | 1934558 | 7059993 | 1890418 | 6.04E-01 | 0.07 | 0.71 |
| RP_NEG_287_12167a8_791 | | 287.1217 | 8.79 | | RP | NEG | | | 578494 | 1115491 | 500254 | 1095125 | 6.04E-01 | -0.07 | 0.62 |
| RP_POS_475_41553a15_185 | | 475.4155 | 15.19 | | RP | POS | | | 4E+07 | 7131814 | 3.9E+07 | 6907850 | 6.04E-01 | -0.07 | 1.02 |
| RP_POS_548_8009a10_31 | | 548.8009 | 10.31 | | RP | POS | | | 1860108 | 690644 | 1809606 | 729792 | 6.04E-01 | -0.07 | 0.68 |
| RP_POS_794_60516a14_369 | | 794.6052 | 14.37 | | RP | POS | | | 4867734 | 3571641 | 4624512 | 3294740 | 6.04E-01 | -0.07 | 0.8 |
| RP_POS_806_56586a11_594 | | 806.5659 | 11.59 | | RP | POS | | | 1642573 | 444518 | 1608828 | 499894 | 6.04E-01 | -0.07 | 0.65 |
| HILIC_NEG_132_92358a4_413 | | 132.9236 | 4.41 | | HILIC | NEG | | | 2E+07 | 1055601 | 2E+07 | 1063932 | 6.05E-01 | 0.07 | 0.72 |
| RP_POS_777_05292a12_793 | | 777.0529 | 12.79 | | RP | POS | | | 945390 | 595772 | 984789 | 522729 | 6.05E-01 | 0.07 | 1.04 |
| RP_POS_994_25153a9_739 | | 994.2515 | 9.74 | | RP | POS | | | 2076863 | 217454 | 2093192 | 241928 | 6.05E-01 | 0.07 | 0.73 |
| RP_NEG_374_13254a11_142 | | 374.1325 | 11.14 | | RP | NEG | | | 3973220 | 354580 | 3948083 | 356115 | 6.05E-01 | -0.07 | 0.69 |
| RP_POS_271_22668a9_162 | | 271.2267 | 9.16 | | RP | POS | | | 1121739 | 143482 | 1111394 | 148669 | 6.05E-01 | -0.07 | 0.84 |
| RP_POS_169_03569a0_915 | | 169.0357 | 0.92 | | RP | POS | 169.0354(100), 141.0411(30) | | 7033253 | 1383605 | 7134274 | 1473279 | 6.06E-01 | 0.07 | 0.38 |
| HILIC_POS_158_11771a2_191 | | 158.1177 | 2.19 | | HILIC | POS | | | 161894 | 134215 | 172477 | 161858 | 6.06E-01 | 0.07 | 0.64 |
| RP_NEG_315_18127a8_249 | | 315.1813 | 8.25 | | RP | NEG | | | 228123 | 23732.8 | 229836 | 24710.6 | 6.06E-01 | 0.07 | 0.56 |
| RP_NEG_394_97638a7_255 | | 394.9764 | 7.26 | | RP | NEG | | | 543554 | 66676.7 | 548742 | 78928.8 | 6.06E-01 | 0.07 | 0.41 |
| RP_POS_423_27158a10_587 | | 423.2716 | 10.59 | | RP | POS | | | 1726684 | 1004088 | 1800865 | 1091890 | 6.06E-01 | 0.07 | 1.02 |
| RP_POS_707_29535a11_382 | | 707.2954 | 11.38 | | RP | POS | | | 172859 | 117982 | 181281 | 120673 | 6.06E-01 | 0.07 | 0.72 |
| RP_POS_841_41687a12_617 | | 841.4169 | 12.62 | | RP | POS | | | 387350 | 638361 | 432617 | 644324 | 6.06E-01 | 0.07 | 0.71 |
| HILIC_NEG_315_0419a6_932 | | 315.0419 | 6.93 | | HILIC | NEG | | | 1123706 | 491092 | 1090736 | 446189 | 6.06E-01 | -0.07 | 0.61 |
| RP_NEG_300_99057a6_962 | | 300.9906 | 6.96 | | RP | NEG | | | 1073446 | 336552 | 1050349 | 319535 | 6.06E-01 | -0.07 | 0.74 |
| RP_POS_577_39337a9_453 | | 577.3934 | 9.45 | | RP | POS | | | 391647 | 379325 | 368650 | 273792 | 6.06E-01 | -0.07 | 0.82 |
| RP_POS_209_07864a2_829 | Ethyl caffeate | 209.0786 | 2.83 | | RP | POS | 209.0801(100) | 2 | 1.2E+09 | 1.7E+09 | 1.4E+09 | 1.7E+09 | 6.06E-01 | 0.07 | 0.46 |
| HILIC_NEG_163_84024a3_11 | | 163.8402 | 3.11 | | HILIC | NEG | | | 2.2E+07 | 699445 | 2.2E+07 | 644521 | 6.07E-01 | 0.07 | 0.62 |
| RP_NEG_653_30408a10_851 | | 653.3041 | 10.85 | | RP | NEG | | | 46097.7 | 25896.9 | 48153.4 | 31704.3 | 6.07E-01 | 0.07 | 0.67 |
| RP_POS_324_20172a5_448 | | 324.2017 | 5.45 | | RP | POS | | | 315384 | 102268 | 323405 | 123085 | 6.07E-01 | 0.07 | 0.85 |
| RP_POS_611_76617a11_502 | | 611.7662 | 11.50 | | RP | POS | | | 130661 | 76843.8 | 136776 | 94629.2 | 6.07E-01 | 0.07 | 0.62 |
| RP_POS_758_56921a13_213 | PC 34:2 (16:0_18:2) | 758.5692 | 13.21 | | RP | POS | 184.0747(100) | 1 | 2E+09 | 5.1E+08 | 2E+09 | 4.4E+08 | 6.07E-01 | -0.07 | 0.89 |
| RP_POS_292_17563a4_547 | | 292.1756 | 4.55 | | RP | POS | | | 230491 | 101638 | 223205 | 105381 | 6.07E-01 | -0.07 | 0.74 |
| RP_POS_623_93707a11_377 | | 623.9371 | 11.38 | | RP | POS | | | 261950 | 143281 | 274426 | 202796 | 6.08E-01 | 0.07 | 0.57 |
| RP_POS_654_45251a9_574 | | 654.4525 | 9.57 | | RP | POS | | | 445952 | 207700 | 461722 | 238585 | 6.08E-01 | 0.07 | 0.51 |
| RP_NEG_291_78537a15_483 | | 291.7854 | 15.48 | | RP | NEG | | | 307475 | 46946.6 | 304090 | 49287.7 | 6.08E-01 | -0.07 | 0.66 |
| RP_POS_149_13271a7_694 | | 149.1327 | 7.69 | | RP | POS | | | 875494 | 72754.7 | 869971 | 83420.8 | 6.08E-01 | -0.07 | 0.62 |
| RP_POS_443_33401a10_699 | | 443.3340 | 10.70 | | RP | POS | | | 9715463 | 1656957 | 9607409 | 1437883 | 6.08E-01 | -0.07 | 1 |
| RP_POS_473_34525a10_533 | | 473.3453 | 10.53 | | RP | POS | | | 1232863 | 363888 | 1208621 | 328955 | 6.08E-01 | -0.07 | 1.08 |
| RP_NEG_339_18088a7_11 | | 339.1809 | 7.11 | | RP | NEG | | | 128118 | 30130.5 | 130758 | 43098.2 | 6.09E-01 | 0.07 | 0.6 |
| RP_NEG_427_94522a0_679 | | 427.9452 | 0.68 | | RP | NEG | | | 539393 | 93420.1 | 545810 | 90350.4 | 6.09E-01 | 0.07 | 0.46 |
| RP_NEG_862_39722a5_622 | | 862.3972 | 5.62 | | RP | NEG | | | 1295902 | 571390 | 1252260 | 664193 | 6.09E-01 | -0.07 | 0.83 |
| RP_POS_186_14906a15_428 | | 186.1491 | 15.43 | | RP | POS | | | 654475 | 255352 | 637054 | 244069 | 6.09E-01 | -0.07 | 0.5 |
| RP_POS_278_15976a6_42 | | 278.1598 | 6.42 | | RP | POS | | | 2782740 | 2260410 | 2624586 | 2260385 | 6.09E-01 | -0.07 | 0.49 |
| RP_POS_318_26364a9_929 | | 318.2636 | 9.93 | | RP | POS | | | 906363 | 439770 | 875200 | 450754 | 6.09E-01 | -0.07 | 0.52 |
| RP_POS_546_86389a9_585 | | 546.8639 | 9.59 | | RP | POS | | | 694362 | 363684 | 666912 | 415038 | 6.09E-01 | -0.07 | 0.56 |
| RP_POS_639_44653a9_751 | | 639.4465 | 9.75 | | RP | POS | | | 2335740 | 1074482 | 2254080 | 1239013 | 6.09E-01 | -0.07 | 0.7 |
| RP_POS_658_44928a10_09 | | 658.4493 | 10.09 | | RP | POS | | | 750371 | 400050 | 723454 | 371362 | 6.09E-01 | -0.07 | 0.78 |
| HILIC_POS_217_08144a6_583 | | 217.0814 | 6.58 | | HILIC | POS | | | 6994183 | 1115094 | 7069333 | 1044164 | 6.10E-01 | 0.07 | 0.64 |
| HILIC_POS_256_10974a0_615 | | 256.1097 | 0.62 | | HILIC | POS | | | 8837729 | 1.2E+07 | 9518790 | 6880216 | 6.10E-01 | 0.07 | 0.52 |
| RP_NEG_840_57678a13_482 | | 840.5768 | 13.48 | | RP | NEG | | | 2012301 | 718600 | 2062800 | 729611 | 6.10E-01 | 0.07 | 0.96 |
| RP_POS_1069_71631a10_929 | | 1069.7163 | 10.93 | | RP | POS | | | 7617014 | 4079753 | 7910622 | 4308928 | 6.10E-01 | 0.07 | 0.64 |
| RP_POS_176_07405a2_918 | | 176.0741 | 2.92 | | RP | POS | | | 4170486 | 3449764 | 4518298 | 5991396 | 6.10E-01 | 0.07 | 0.85 |
| RP_POS_273_18442a10_268 | | 273.1844 | 10.27 | | RP | POS | | | 594151 | 82140.2 | 600457 | 96741.9 | 6.10E-01 | 0.07 | 0.7 |
| RP_POS_550_38672a11_008 | | 550.3867 | 11.01 | | RP | POS | | | 9080020 | 2525591 | 9262062 | 2679018 | 6.10E-01 | 0.07 | 0.63 |
| RP_POS_551_33008a10_932 | | 551.3301 | 10.93 | | RP | POS | | | 3064813 | 940306 | 3134696 | 1051968 | 6.10E-01 | 0.07 | 0.82 |
| RP_POS_653_12604a11_558 | | 653.1260 | 11.56 | | RP | POS | | | 176964 | 87421.1 | 184001 | 111081 | 6.10E-01 | 0.07 | 0.65 |
| RP_POS_699_9657a10_075 | | 699.9657 | 10.08 | | RP | POS | | | 813452 | 423476 | 845188 | 480235 | 6.10E-01 | 0.07 | 0.9 |
| HILIC_POS_184_05775a4_443 | | 184.0578 | 4.44 | | HILIC | POS | | | 647690 | 273265 | 626394 | 329276 | 6.10E-01 | -0.07 | 0.68 |
| RP_NEG_440_88818a15_336 | | 440.8882 | 15.34 | | RP | NEG | | | 2366406 | 81769.4 | 2360570 | 85220.8 | 6.10E-01 | -0.07 | 0.66 |
| RP_POS_242_87975a15_398 | | 242.8798 | 15.40 | | RP | POS | | | 5422272 | 1240040 | 5341332 | 1089385 | 6.10E-01 | -0.07 | 0.74 |
| RP_POS_317_11465a15_413 | | 317.1147 | 15.41 | | RP | POS | | | 803900 | 234644 | 785397 | 288600 | 6.10E-01 | -0.07 | 0.86 |
| RP_POS_778_53503a12_723 | | 778.5350 | 12.72 | | RP | POS | | | 2E+07 | 1.2E+07 | 1.9E+07 | 1.1E+07 | 6.10E-01 | -0.07 | 0.63 |
| RP_POS_131_04945a2_158 | trans-Cinnamic acid | 131.0495 | 2.16 | | RP | POS | 103.0549(100) | 2 | 561524 | 102420 | 568761 | 104780 | 6.10E-01 | 0.07 | 0.67 |
| HILIC_POS_257_21078a3_143 | | 257.2108 | 3.14 | | HILIC | POS | | | 209432 | 69755.6 | 214143 | 65870.5 | 6.11E-01 | 0.07 | 0.35 |
| RP_NEG_384_93658a14_347 | | 384.9366 | 14.35 | | RP | NEG | | | 2841282 | 1424205 | 2942327 | 1477355 | 6.11E-01 | 0.07 | 0.55 |
| RP_POS_397_27066a11_241 | | 397.2707 | 11.24 | | RP | POS | | | 456592 | 209746 | 469999 | 176653 | 6.11E-01 | 0.07 | 0.65 |
| RP_POS_425_21463a9_958 | | 425.2146 | 9.96 | | RP | POS | | | 2893004 | 2425328 | 3079127 | 2864976 | 6.11E-01 | 0.07 | 0.72 |
| HILIC_POS_365_06735a4_851 | | 365.0674 | 4.85 | | HILIC | POS | | | 316924 | 262761 | 300610 | 207824 | 6.11E-01 | -0.07 | 0.67 |
| RP_NEG_979_13959a9_848 | | 979.1396 | 9.85 | | RP | NEG | | | 3876985 | 227696 | 3861819 | 209469 | 6.11E-01 | -0.07 | 0.96 |
| RP_POS_255_2317a10_254 | | 255.2317 | 10.25 | | RP | POS | | | 1370996 | 160258 | 1359877 | 159649 | 6.11E-01 | -0.07 | 0.65 |
| RP_POS_426_32324a10_121 | | 426.3232 | 10.12 | | RP | POS | | | 1194435 | 534252 | 1154934 | 594242 | 6.11E-01 | -0.07 | 0.7 |
| RP_NEG_572_04333a0_679 | | 572.0433 | 0.68 | | RP | NEG | | | 414371 | 89176.7 | 421118 | 103617 | 6.12E-01 | 0.07 | 0.46 |
| RP_POS_418_7485a13_119 | | 418.7485 | 13.12 | | RP | POS | | | 4990613 | 2325907 | 5152861 | 2347385 | 6.12E-01 | 0.07 | 0.91 |
| RP_POS_553_31232a10_606 | | 553.3123 | 10.61 | | RP | POS | | | 289915 | 186640 | 302262 | 170722 | 6.12E-01 | 0.07 | 0.58 |
| RP_POS_593_08954a10_529 | | 593.0895 | 10.53 | | RP | POS | | | 183247 | 85110.7 | 189413 | 91902.5 | 6.12E-01 | 0.07 | 0.41 |
| RP_POS_132_1021a1_438 | | 132.1021 | 1.44 | | RP | POS | 86.0974(100), 132.1026(2) | | 4.3E+07 | 1.4E+07 | 4.2E+07 | 1E+07 | 6.12E-01 | -0.07 | 0.53 |
| HILIC_NEG_176_83672a3_226 | | 176.8367 | 3.23 | | HILIC | NEG | | | 636465 | 267961 | 617764 | 270993 | 6.12E-01 | -0.07 | 0.78 |
| HILIC_POS_150_1125a5_795 | | 150.1125 | 5.80 | | HILIC | POS | | | 4678273 | 1672578 | 4561317 | 1693015 | 6.12E-01 | -0.07 | 0.83 |
| RP_POS_436_77771a13_773 | | 436.7777 | 13.77 | | RP | POS | | | 1163108 | 788583 | 1113252 | 655747 | 6.12E-01 | -0.07 | 0.53 |
| RP_POS_809_5285a13_113 | | 809.5285 | 13.11 | | RP | POS | | | 2016204 | 1160446 | 1941403 | 1004809 | 6.12E-01 | -0.07 | 0.7 |
| HILIC_NEG_144_92355a1_502 | | 144.9236 | 1.50 | | HILIC | NEG | | | 1.3E+07 | 7047660 | 1.3E+07 | 6304929 | 6.13E-01 | 0.07 | 0.58 |
| HILIC_NEG_200_85922a4_296 | | 200.8592 | 4.30 | | HILIC | NEG | | | 1527456 | 141662 | 1536948 | 133185 | 6.13E-01 | 0.07 | 0.84 |
| RP_NEG_735_07483a9_195 | | 735.0748 | 9.20 | | RP | NEG | | | 377978 | 27340.6 | 379765 | 24520.5 | 6.13E-01 | 0.07 | 0.73 |
| RP_POS_453_23892a7_635 | | 453.2389 | 7.64 | | RP | POS | | | 503854 | 174091 | 517268 | 208774 | 6.13E-01 | 0.07 | 0.69 |
| HILIC_NEG_334_05865a4_852 | | 334.0587 | 4.85 | | HILIC | NEG | | | 88176.4 | 56938.2 | 84618.7 | 46267.1 | 6.13E-01 | -0.07 | 0.65 |
| HILIC_POS_172_09427a5_26 | | 172.0943 | 5.26 | | HILIC | POS | | | 3494470 | 2043077 | 3369272 | 1585642 | 6.13E-01 | -0.07 | 0.86 |
| RP_POS_288_13614a10_607 | | 288.1361 | 10.61 | | RP | POS | | | 740739 | 308153 | 720554 | 276956 | 6.13E-01 | -0.07 | 0.9 |
| RP_POS_456_35321a8_771 | | 456.3532 | 8.77 | | RP | POS | | | 1170373 | 597771 | 1123007 | 748925 | 6.13E-01 | -0.07 | 0.81 |
| RP_POS_219_04684a6_648 | Diphenylsulfone | 219.0468 | 6.65 | | RP | POS | 141.0011(100), 219.0466(85) | 2 | 2087178 | 424326 | 2115629 | 401000 | 6.13E-01 | 0.07 | 0.45 |
| RP_NEG_525_224a11_091 | | 525.2240 | 11.09 | | RP | NEG | | | 50517.1 | 103775 | 63068.5 | 227602 | 6.14E-01 | 0.08 | 0.39 |
| RP_POS_221_15324a7_885 | | 221.1532 | 7.89 | | RP | POS | 221.1528(100) | | 3179787 | 259752 | 3196430 | 224630 | 6.14E-01 | 0.07 | 0.36 |
| HILIC_NEG_301_72702a3_3 | | 301.7270 | 3.30 | | HILIC | NEG | | | 189544 | 120665 | 198188 | 129140 | 6.14E-01 | 0.07 | 0.59 |
| RP_NEG_407_17282a2_885 | | 407.1728 | 2.89 | | RP | NEG | | | 226458 | 75797.3 | 231655 | 75073.8 | 6.14E-01 | 0.07 | 0.63 |
| RP_POS_241_177a7_502 | | 241.1770 | 7.50 | | RP | POS | | | 1488971 | 460362 | 1520976 | 466585 | 6.14E-01 | 0.07 | 0.53 |
| RP_POS_316_21173a3_216 | | 316.2117 | 3.22 | | RP | POS | | | 161517 | 272788 | 187362 | 442257 | 6.14E-01 | 0.07 | 0.86 |
| HILIC_POS_294_93915a4_337 | | 294.9392 | 4.34 | | HILIC | POS | | | 2.9E+07 | 1682899 | 2.9E+07 | 1733232 | 6.14E-01 | -0.07 | 0.76 |
| RP_NEG_430_11295a11_141 | | 430.1130 | 11.14 | | RP | NEG | | | 428857 | 42302.8 | 426053 | 39178.2 | 6.14E-01 | -0.07 | 0.79 |
| HILIC_NEG_476_27786a1_877 | LysoPE 18:2 | 476.2779 | 1.88 | | HILIC | NEG | 279.2339(100), 476.2752(5) | 2 | 492500 | 214107 | 508746 | 251923 | 6.14E-01 | 0.07 | 0.72 |
| HILIC_NEG_200_85928a4_21 | | 200.8593 | 4.21 | | HILIC | NEG | | | 2209553 | 878170 | 2269947 | 875800 | 6.15E-01 | 0.07 | 0.62 |
| RP_POS_231_116a15_691 | | 231.1160 | 15.69 | | RP | POS | | | 4875381 | 2128899 | 5048816 | 2809416 | 6.15E-01 | 0.07 | 0.45 |
| RP_POS_698_48828a11_393 | | 698.4883 | 11.39 | | RP | POS | | | 304830 | 190674 | 321896 | 287814 | 6.15E-01 | 0.07 | 0.56 |
| RP_POS_897_47565a12_617 | | 897.4757 | 12.62 | | RP | POS | | | 403817 | 673610 | 450433 | 681319 | 6.15E-01 | 0.07 | 0.7 |
| HILIC_POS_150_11237a5_048 | | 150.1124 | 5.05 | | HILIC | POS | 150.1126(100), 132.1027(40) | | 3749822 | 2354008 | 3584737 | 2436952 | 6.15E-01 | -0.07 | 0.71 |
| HILIC_POS_499_05951a7_668 | | 499.0595 | 7.67 | | HILIC | POS | | | 462775 | 27205.7 | 460844 | 28773.2 | 6.15E-01 | -0.07 | 0.42 |
| RP_NEG_316_9483a10_434 | | 316.9483 | 10.43 | | RP | NEG | | | 485746 | 195771 | 471375 | 219189 | 6.15E-01 | -0.07 | 0.42 |
| HILIC_NEG_216_85353a7_743 | | 216.8535 | 7.74 | | HILIC | NEG | | | 563411 | 128013 | 572315 | 131158 | 6.16E-01 | 0.07 | 0.45 |
| HILIC_POS_334_08167a5_474 | | 334.0817 | 5.47 | | HILIC | POS | | | 342297 | 175385 | 353471 | 151966 | 6.16E-01 | 0.07 | 0.42 |
| RP_POS_212_07426a15_539 | | 212.0743 | 15.54 | | RP | POS | | | 4148640 | 2109958 | 4298777 | 2250055 | 6.16E-01 | 0.07 | 0.53 |
| RP_POS_777_54437a10_33 | | 777.5444 | 10.33 | | RP | POS | | | 873582 | 528509 | 908723 | 498790 | 6.16E-01 | 0.07 | 0.82 |
| HILIC_POS_228_19598a1_021 | | 228.1960 | 1.02 | | HILIC | POS | | | 713993 | 326981 | 691966 | 316830 | 6.16E-01 | -0.07 | 0.49 |
| RP_NEG_231_9151a0_682 | | 231.9151 | 0.68 | | RP | NEG | | | 428030 | 66000.2 | 424004 | 51689.5 | 6.16E-01 | -0.07 | 0.7 |
| HILIC_POS_480_34592a1_066 | LysoPC 15:1 | 480.3459 | 1.07 | | HILIC | POS | 480.3423(100), 104.1079(90) | 2 | 1737271 | 679048 | 1783484 | 669161 | 6.16E-01 | 0.07 | 0.53 |
| HILIC_POS_245_09969a2_26 | | 245.0997 | 2.26 | | HILIC | POS | 245.099(100) | | 575345 | 342206 | 597205 | 299877 | 6.17E-01 | 0.07 | 0.47 |
| HILIC_POS_289_12552a0_771 | | 289.1255 | 0.77 | | HILIC | POS | | | 266748 | 28960 | 268691 | 27955.7 | 6.17E-01 | 0.07 | 0.47 |
| HILIC_POS_586_27118a0_764 | | 586.2712 | 0.76 | | HILIC | POS | | | 40383.9 | 44476.2 | 44587.2 | 72715 | 6.17E-01 | 0.07 | 0.48 |
| RP_NEG_156_92979a15_394 | | 156.9298 | 15.39 | | RP | NEG | | | 2631612 | 581351 | 2670168 | 547799 | 6.17E-01 | 0.07 | 0.54 |
| RP_POS_284_16971a3_582 | | 284.1697 | 3.58 | | RP | POS | | | 306709 | 38016.7 | 309209 | 35150.2 | 6.17E-01 | 0.07 | 0.95 |
| RP_POS_591_84174a11_393 | | 591.8417 | 11.39 | | RP | POS | | | 123606 | 53659.2 | 127783 | 66758.3 | 6.17E-01 | 0.07 | 0.59 |
| RP_POS_593_4422a11_716 | | 593.4422 | 11.72 | | RP | POS | | | 564639 | 418878 | 539289 | 324090 | 6.17E-01 | -0.07 | 0.32 |
| RP_NEG_203_08315a2_884 | | 203.0832 | 2.88 | | RP | NEG | 116.05(100), 203.0829(65) | | 1.8E+07 | 3088728 | 1.8E+07 | 3052318 | 6.18E-01 | 0.07 | 0.6 |
| RP_POS_575_88489a9_775 | | 575.8849 | 9.78 | | RP | POS | | | 1572426 | 903290 | 1507475 | 990351 | 6.18E-01 | -0.07 | 0.87 |
| RP_POS_688_39355a11_143 | | 688.3936 | 11.14 | | RP | POS | | | 488910 | 181542 | 476498 | 182083 | 6.18E-01 | -0.07 | 0.68 |
| HILIC_POS_59_06135a3_199 | | 59.0614 | 3.20 | | HILIC | POS | | | 7821697 | 2786811 | 8015936 | 2914360 | 6.19E-01 | 0.07 | 0.35 |
| RP_NEG_576_20386a10_301 | | 576.2039 | 10.30 | | RP | NEG | | | 443461 | 133704 | 453333 | 153974 | 6.19E-01 | 0.07 | 0.72 |
| RP_POS_306_09479a15_54 | | 306.0948 | 15.54 | | RP | POS | | | 1174715 | 532323 | 1211464 | 545740 | 6.19E-01 | 0.07 | 0.48 |
| HILIC_NEG_112_98567a0_782 | | 112.9857 | 0.78 | | HILIC | NEG | 68.9949(100), 112.9855(2) | | 7328878 | 4523000 | 7045642 | 3836142 | 6.19E-01 | -0.07 | 0.63 |
| HILIC_NEG_381_00635a6_685 | | 381.0064 | 6.69 | | HILIC | NEG | | | 1101756 | 158439 | 1091349 | 147932 | 6.19E-01 | -0.07 | 0.42 |
| RP_POS_261_13327a6_594 | | 261.1333 | 6.59 | | RP | POS | | | 649572 | 819185 | 594880 | 793087 | 6.19E-01 | -0.07 | 0.52 |
| RP_POS_315_15668a8_67 | | 315.1567 | 8.67 | | RP | POS | | | 2943150 | 169131 | 2932748 | 137440 | 6.19E-01 | -0.07 | 0.42 |
| RP_POS_614_33972a10_934 | | 614.3397 | 10.93 | | RP | POS | | | 3446580 | 581920 | 3404698 | 643490 | 6.19E-01 | -0.07 | 0.51 |
| RP_NEG_164_07147a1_412 | | 164.0715 | 1.41 | | RP | NEG | 147.0452(100), 164.0709(50), 72.0085(35) | | 9368801 | 2368605 | 9524072 | 2226276 | 6.20E-01 | 0.07 | 0.53 |
| HILIC_POS_387_21381a4_606 | | 387.2138 | 4.61 | | HILIC | POS | | | 419118 | 259835 | 438334 | 301863 | 6.20E-01 | 0.07 | 0.56 |
| RP_NEG_207_13882a6_886 | | 207.1388 | 6.89 | | RP | NEG | | | 336656 | 103485 | 343932 | 110345 | 6.20E-01 | 0.07 | 0.53 |
| RP_NEG_806_50189a10_419 | | 806.5019 | 10.42 | | RP | NEG | | | 273880 | 295036 | 291425 | 223118 | 6.20E-01 | 0.07 | 0.36 |
| RP_POS_341_35278a10_833 | | 341.3528 | 10.83 | | RP | POS | | | 906311 | 637207 | 951657 | 694211 | 6.20E-01 | 0.07 | 1.29 |
| RP_POS_638_56964a11_896 | | 638.5696 | 11.90 | | RP | POS | | | 1743557 | 3587614 | 1995927 | 3839039 | 6.20E-01 | 0.07 | 0.8 |
| RP_NEG_255_82214a0_639 | | 255.8221 | 0.64 | | RP | NEG | | | 670343 | 272791 | 654981 | 178061 | 6.20E-01 | -0.07 | 0.53 |
| RP_NEG_787_10187a9_525 | | 787.1019 | 9.53 | | RP | NEG | | | 4878777 | 286232 | 4861004 | 239271 | 6.20E-01 | -0.07 | 1.07 |
| HILIC_NEG_176_99847a5_112 | FT-PFCA | 176.9985 | 5.11 | | HILIC | NEG | 92.9956(100), 176.9988(5) | 2 | 1431001 | 678049 | 1481056 | 786482 | 6.20E-01 | 0.07 | 0.38 |
| HILIC_NEG_590_34729a1_163 | | 590.3473 | 1.16 | | HILIC | NEG | | | 183443 | 70368.2 | 188394 | 75410.7 | 6.21E-01 | 0.07 | 0.73 |
| RP_NEG_395_24451a8_637 | | 395.2445 | 8.64 | | RP | NEG | | | 231984 | 36572 | 234416 | 35469.4 | 6.21E-01 | 0.07 | 0.7 |
| RP_POS_170_11665a10_833 | | 170.1167 | 10.83 | | RP | POS | | | 1023432 | 550342 | 1060638 | 551081 | 6.21E-01 | 0.07 | 0.95 |
| RP_POS_493_35342a9_378 | | 493.3534 | 9.38 | | RP | POS | | | 1319838 | 682532 | 1368996 | 762067 | 6.21E-01 | 0.07 | 0.78 |
| RP_POS_502_29056a10_594 | | 502.2906 | 10.59 | | RP | POS | | | 5450791 | 1650119 | 5572877 | 1926199 | 6.21E-01 | 0.07 | 0.6 |
| RP_NEG_129_05592a3_583 | Ketoleucine | 129.0559 | 3.58 | | RP | NEG | 129.0557(100) | 2 | 2.7E+07 | 7998680 | 2.6E+07 | 7208470 | 6.21E-01 | -0.07 | 0.77 |
| RP_NEG_293_1767a8_305 | | 293.1767 | 8.31 | | RP | NEG | | | 1823438 | 299904 | 1804068 | 274352 | 6.21E-01 | -0.07 | 0.81 |
| RP_POS_276_11893a0_575 | | 276.1189 | 0.58 | | RP | POS | | | 2859357 | 757599 | 2810670 | 685367 | 6.21E-01 | -0.07 | 0.56 |
| RP_POS_349_06363a15_537 | | 349.0636 | 15.54 | | RP | POS | | | 1547130 | 641782 | 1506191 | 571336 | 6.21E-01 | -0.07 | 0.77 |
| RP_POS_679_14331a9_416 | | 679.1433 | 9.42 | | RP | POS | | | 481963 | 55002.2 | 478498 | 47871.5 | 6.21E-01 | -0.07 | 0.9 |
| HILIC_NEG_157_03674a1_884 | Allatoin | 157.0367 | 1.88 | | HILIC | NEG | 97.004(100), 114.0296(70) | 1 | 610159 | 368482 | 632914 | 307750 | 6.21E-01 | 0.07 | 0.53 |
| RP_NEG_636_34882a10_933 | | 636.3488 | 10.93 | | RP | NEG | | | 1.3E+07 | 1888104 | 1.3E+07 | 2010915 | 6.22E-01 | 0.07 | 0.67 |
| RP_NEG_723_08704a9_337 | | 723.0870 | 9.34 | | RP | NEG | | | 3492067 | 319675 | 3512334 | 283710 | 6.22E-01 | 0.07 | 0.54 |
| RP_POS_631_43945a11_364 | | 631.4395 | 11.36 | | RP | POS | | | 297549 | 147580 | 307742 | 154120 | 6.22E-01 | 0.07 | 0.56 |
| HILIC_POS_133_03197a4_63 | | 133.0320 | 4.63 | | HILIC | POS | | | 3602857 | 1230271 | 3523184 | 1140057 | 6.22E-01 | -0.07 | 0.57 |
| RP_NEG_463_14612a11_592 | | 463.1461 | 11.59 | | RP | NEG | | | 844804 | 52679.8 | 840893 | 62195.5 | 6.22E-01 | -0.07 | 0.75 |
| RP_NEG_701_15454a9_747 | | 701.1545 | 9.75 | | RP | NEG | | | 5392019 | 224684 | 5377031 | 220677 | 6.22E-01 | -0.07 | 0.48 |
| RP_POS_177_14847a6_468 | | 177.1485 | 6.47 | | RP | POS | | | 1164571 | 109263 | 1157065 | 112950 | 6.22E-01 | -0.07 | 0.87 |
| RP_POS_367_20325a10_893 | | 367.2033 | 10.89 | | RP | POS | | | 430505 | 247862 | 414080 | 239452 | 6.22E-01 | -0.07 | 0.89 |
| RP_POS_384_08231a6_124 | Pantoprazole | 384.0823 | 6.12 | | RP | POS | 138.0547(100), 200.0386(65) | 2 | 3.1E+07 | 2.1E+07 | 3E+07 | 2E+07 | 6.22E-01 | -0.07 | 0.6 |
| HILIC_POS_245_09946a4_415 | | 245.0995 | 4.42 | | HILIC | POS | 245.0945(100) | | 460807 | 247337 | 479887 | 311830 | 6.23E-01 | 0.07 | 0.55 |
| HILIC_NEG_173_92607a7_397 | | 173.9261 | 7.40 | | HILIC | NEG | | | 1139617 | 557984 | 1180786 | 654727 | 6.23E-01 | 0.07 | 0.39 |
| RP_NEG_565_99005a10_607 | | 565.9901 | 10.61 | | RP | NEG | | | 208747 | 140813 | 218102 | 137664 | 6.23E-01 | 0.07 | 0.77 |
| RP_POS_188_07071a2_883 | | 188.0707 | 2.88 | | RP | POS | | | 1.9E+08 | 3.4E+07 | 1.9E+08 | 3.2E+07 | 6.23E-01 | 0.07 | 0.72 |
| RP_POS_147_04416a0_679 | | 147.0442 | 0.68 | | RP | POS | 119.0492(100), 147.0448(38) | | 4328094 | 1255823 | 4248444 | 1123462 | 6.23E-01 | -0.07 | 0.55 |
| HILIC_NEG_176_998a1_348 | | 176.9980 | 1.35 | | HILIC | NEG | | | 1060735 | 310382 | 1039874 | 309909 | 6.23E-01 | -0.07 | 0.37 |
| HILIC_NEG_243_96796a1_442 | | 243.9680 | 1.44 | | HILIC | NEG | | | 5739150 | 1160718 | 5667910 | 965933 | 6.23E-01 | -0.07 | 0.79 |

**Supplemantary table 1**: The non-targeted metabolomics results ordered by the VIP value (p value < 0.0003 are shaded dark green, p value < 0.05 are shaded light green, d values below 0 are blue and above 0 are red)

| | | | | | | | | | Cases | | Controls | | Welch test | Cohen's | |
|---|---|---|---|---|---|---|---|---|---|---|---|---|---|---|---|
| Feature_ID | Metabolite_name | Average_Mz | Average_RT_min_ | HMDB ID | RP_HILIC | POS_NEG | Cureted_MSMS | ID_level | Mean | SD | Mean2 | SD3 | p | d | VIP |
| HILIC_POS_387_21402a5_785 | | 387.2140 | 5.79 | | HILIC | POS | | | 1654836 | 1098615 | 1585125 | 980915 | 6.23E-01 | -0.07 | 0.65 |
| RP_NEG_242_98582a6_588 | | 242.9858 | 6.59 | | RP | NEG | | | 3703688 | 625933 | 3663708 | 567119 | 6.23E-01 | -0.07 | 0.6 |
| RP_NEG_379_15628a10_388 | | 379.1563 | 10.39 | | RP | NEG | | | 311376 | 162616 | 299410 | 190239 | 6.23E-01 | -0.07 | 0.34 |
| RP_NEG_601_44379a11_142 | | 601.4438 | 11.14 | | RP | NEG | | | 387199 | 126808 | 379027 | 116680 | 6.23E-01 | -0.07 | 1.26 |
| RP_POS_136_59586a11_656 | | 136.5959 | 11.66 | | RP | POS | | | 2169345 | 3876451 | 1930544 | 3248160 | 6.23E-01 | -0.07 | 0.59 |
| RP_POS_236_07153a10_971 | | 236.0715 | 10.97 | | RP | POS | | | 9E+07 | 2.3E+07 | 8.8E+07 | 2.3E+07 | 6.23E-01 | -0.07 | 0.42 |
| RP_POS_388_79013a9_909 | | 388.7901 | 9.91 | | RP | POS | | | 382337 | 231971 | 365701 | 259509 | 6.23E-01 | -0.07 | 0.62 |
| RP_POS_414_18503a10_893 | | 414.1850 | 10.89 | | RP | POS | | | 325827 | 168353 | 314531 | 168139 | 6.23E-01 | -0.07 | 0.86 |
| RP_NEG_204_06651a4_821 | 3-Indolelactic acid | 204.0665 | 4.82 | | RP | NEG | 204.0667(100), 158.0606(80) | 2 | 3769718 | 965987 | 3710875 | 792618 | 6.23E-01 | -0.07 | 0.6 |
| RP_NEG_171_10281a6_083 | | 171.1028 | 6.08 | | RP | NEG | | | 510297 | 79482.5 | 515917 | 87170.9 | 6.24E-01 | 0.07 | 0.66 |
| RP_NEG_542_79877a15_389 | | 542.7988 | 15.39 | | RP | NEG | | | 639936 | 186796 | 652540 | 189207 | 6.24E-01 | 0.07 | 0.54 |
| RP_POS_175_11157a7_723 | | 175.1116 | 7.72 | | RP | POS | | | 1761802 | 164038 | 1772464 | 154369 | 6.24E-01 | 0.07 | 0.88 |
| RP_POS_590_41064a7_36 | | 590.4106 | 7.36 | | RP | POS | | | 595809 | 419883 | 624612 | 438389 | 6.24E-01 | 0.07 | 1.01 |
| RP_POS_596_26392a10_356 | | 596.2639 | 10.36 | | RP | POS | | | 826025 | 149880 | 836132 | 151794 | 6.24E-01 | 0.07 | 0.75 |
| RP_POS_652_18835a8_903 | | 652.1884 | 8.90 | | RP | POS | | | 899829 | 92773.5 | 905728 | 83522.1 | 6.24E-01 | 0.07 | 0.86 |
| RP_POS_1168_3075a13_202 | | 1168.3075 | 13.20 | | RP | POS | | | 8.4E+07 | 2.2E+08 | 7E+07 | 1.7E+08 | 6.24E-01 | -0.07 | 0.7 |
| RP_POS_167_97141a0_632 | | 167.9714 | 0.63 | | RP | POS | | | 996470 | 461477 | 966080 | 445754 | 6.24E-01 | -0.07 | 0.51 |
| RP_POS_475_33707a10_042 | | 475.3371 | 10.04 | | RP | POS | | | 1198333 | 594623 | 1157759 | 614458 | 6.24E-01 | -0.07 | 0.81 |
| RP_POS_871_18213a9_408 | | 871.1821 | 9.41 | | RP | POS | | | 744243 | 95489.4 | 737749 | 97805 | 6.24E-01 | -0.07 | 0.78 |
| RP_POS_120_08111a1_411 | | 120.0811 | 1.41 | | RP | POS | 120.0809(100) | | 1.5E+07 | 4486929 | 1.5E+07 | 4326661 | 6.25E-01 | 0.07 | 0.55 |
| HILIC_NEG_231_93176a3_101 | | 231.9318 | 3.10 | | HILIC | NEG | | | 522715 | 98958 | 528776 | 82957.3 | 6.25E-01 | 0.07 | 0.5 |
| HILIC_NEG_312_83633a4_059 | | 312.8363 | 4.06 | | HILIC | NEG | | | 862214 | 207022 | 876366 | 215165 | 6.25E-01 | 0.07 | 0.47 |
| RP_NEG_154_93269a15_393 | | 154.9327 | 15.39 | | RP | NEG | | | 4833492 | 821407 | 4779595 | 794510 | 6.25E-01 | -0.07 | 0.63 |
| RP_NEG_665_17334a9_541 | | 665.1733 | 9.54 | | RP | NEG | | | 683435 | 38032.3 | 680920 | 37320.5 | 6.25E-01 | -0.07 | 0.64 |
| RP_POS_181_12215a6_569 | | 181.1222 | 6.57 | | RP | POS | | | 702496 | 185598 | 688683 | 222837 | 6.25E-01 | -0.07 | 0.54 |
| RP_POS_408_32538a12_323 | | 408.3254 | 12.32 | | RP | POS | | | 164521 | 75204.2 | 159669 | 70362.4 | 6.25E-01 | -0.07 | 0.71 |
| RP_POS_485_3555a10_53 | | 485.3555 | 10.53 | | RP | POS | | | 236586 | 133685 | 228251 | 116695 | 6.25E-01 | -0.07 | 0.47 |
| RP_POS_528_39117a10_4 | | 528.3912 | 10.40 | | RP | POS | | | 2918056 | 1402984 | 2830151 | 1233895 | 6.25E-01 | -0.07 | 0.71 |
| RP_POS_570_30945a6_838 | | 570.3095 | 6.84 | | RP | POS | | | 1316038 | 1206044 | 1246669 | 871362 | 6.25E-01 | -0.07 | 0.54 |
| RP_POS_700_5271a13_464 | | 700.5271 | 13.46 | | RP | POS | | | 1305082 | 629876 | 1263668 | 611552 | 6.25E-01 | -0.07 | 0.8 |
| HILIC_POS_346_22192a0_747 | | 346.2219 | 0.75 | | HILIC | POS | | | 544795 | 916958 | 608939 | 998550 | 6.26E-01 | 0.07 | 0.53 |
| RP_NEG_221_94724a14_343 | | 221.9472 | 14.34 | | RP | NEG | | | 3054246 | 1413292 | 3145054 | 1317104 | 6.26E-01 | 0.07 | 0.42 |
| RP_NEG_501_08624a8_832 | | 501.0862 | 8.83 | | RP | NEG | | | 414208 | 30043.4 | 416225 | 30364.3 | 6.26E-01 | 0.07 | 0.95 |
| RP_POS_170_06029a2_482 | | 170.0603 | 2.48 | | RP | POS | | | 3738682 | 827716 | 3791592 | 765812 | 6.26E-01 | 0.07 | 0.65 |
| RP_POS_551_03882a10_165 | | 551.0388 | 10.17 | | RP | POS | | | 552437 | 260932 | 574388 | 377247 | 6.26E-01 | 0.07 | 0.46 |
| RP_NEG_350_98703a7_25 | | 350.9870 | 7.25 | | RP | NEG | | | 1061880 | 261819 | 1045010 | 246036 | 6.26E-01 | -0.07 | 0.91 |
| RP_NEG_483_77829a15_378 | | 483.7783 | 15.38 | | RP | NEG | | | 292873 | 136432 | 283858 | 134424 | 6.26E-01 | -0.07 | 0.71 |
| RP_POS_357_2193a8_842 | | 357.2193 | 8.84 | | RP | POS | | | 332934 | 503800 | 307220 | 244705 | 6.26E-01 | -0.07 | 0.54 |
| RP_POS_396_2645a12_812 | | 396.2645 | 12.81 | | RP | POS | | | 137240 | 97939.8 | 130871 | 93660.6 | 6.26E-01 | -0.07 | 0.61 |
| RP_POS_823_14294a9_539 | | 823.1429 | 9.54 | | RP | POS | | | 829985 | 98349.2 | 823991 | 82208.6 | 6.26E-01 | -0.07 | 0.66 |
| HILIC_POS_155_10648a1_254 | | 155.1065 | 1.25 | | HILIC | POS | | | 180612 | 332954 | 203438 | 352228 | 6.27E-01 | 0.07 | 0.75 |
| RP_NEG_193_92699a0_643 | | 193.9270 | 0.64 | | RP | NEG | | | 949475 | 348934 | 973000 | 358572 | 6.27E-01 | 0.07 | 0.49 |
| RP_NEG_313_00024a2_162 | | 313.0002 | 2.16 | | RP | NEG | | | 2055693 | 343355 | 2078765 | 349878 | 6.27E-01 | 0.07 | 0.63 |
| RP_NEG_693_12506a9_522 | | 693.1251 | 9.52 | | RP | NEG | | | 669165 | 45183.1 | 672389 | 51109.4 | 6.27E-01 | 0.07 | 0.76 |
| RP_POS_281_13583a3_451 | | 281.1358 | 3.45 | | RP | POS | | | 3107824 | 3066599 | 3469736 | 6839462 | 6.27E-01 | 0.07 | 0.51 |
| RP_POS_346_11621a15_323 | | 346.1162 | 15.32 | | RP | POS | | | 3866362 | 757041 | 3921393 | 882292 | 6.27E-01 | 0.07 | 0.38 |
| HILIC_NEG_200_85918a5_497 | | 200.8592 | 5.50 | | HILIC | NEG | | | 8063042 | 1624132 | 7953761 | 1657284 | 6.27E-01 | -0.07 | 0.76 |
| HILIC_POS_217_99323a4_833 | | 217.9932 | 4.83 | | HILIC | POS | | | 390940 | 151719 | 380975 | 148749 | 6.27E-01 | -0.07 | 0.65 |
| RP_POS_316_32126a8_77 | | 316.3213 | 8.77 | | RP | POS | | | 497258 | 712486 | 544676 | 717566 | 6.28E-01 | 0.07 | 0.45 |
| RP_POS_554_39429a10_304 | | 554.3943 | 10.30 | | RP | POS | | | 262815 | 154734 | 275814 | 225397 | 6.28E-01 | 0.07 | 0.65 |
| RP_POS_583_38788a14_354 | | 583.3879 | 14.35 | | RP | POS | | | 4881020 | 3694431 | 5173911 | 4940281 | 6.28E-01 | 0.07 | 0.79 |
| RP_POS_584_47144a10_924 | | 584.4714 | 10.92 | | RP | POS | | | 4909355 | 523687 | 4940537 | 419581 | 6.28E-01 | 0.07 | 0.78 |
| RP_POS_595_84698a11_537 | | 595.8470 | 11.54 | | RP | POS | | | 117576 | 53782 | 121594 | 65813.9 | 6.28E-01 | 0.07 | 0.9 |
| HILIC_NEG_384_93552a4_352 | | 384.9355 | 4.35 | | HILIC | NEG | | | 2.8E+07 | 1877977 | 2.8E+07 | 1786295 | 6.28E-01 | -0.07 | 0.86 |
| HILIC_NEG_80_97431a3_164 | | 80.9743 | 3.16 | | HILIC | NEG | | | 7123819 | 316494 | 7103421 | 300079 | 6.28E-01 | -0.07 | 0.64 |
| RP_POS_192_06526a1_913 | | 192.0653 | 1.91 | | RP | POS | | | 494327 | 105836 | 487168 | 109830 | 6.28E-01 | -0.07 | 0.56 |
| RP_POS_589_26556a9_209 | | 589.2656 | 9.21 | | RP | POS | | | 545728 | 642471 | 505599 | 570921 | 6.28E-01 | -0.07 | 0.63 |
| RP_POS_618_92084a10_306 | | 618.9208 | 10.31 | | RP | POS | | | 2035255 | 1035919 | 1957496 | 1276618 | 6.28E-01 | -0.07 | 0.82 |
| HILIC_NEG_278_01758a4_256 | | 278.0176 | 4.26 | | HILIC | NEG | | | 121320 | 34131 | 119296 | 27396.3 | 6.29E-01 | -0.07 | 0.53 |
| RP_POS_193_15913a8_009 | | 193.1591 | 8.01 | | RP | POS | | | 1938888 | 244171 | 1922231 | 258402 | 6.29E-01 | -0.07 | 0.64 |
| RP_POS_360_04797a8_096 | | 360.0480 | 8.10 | | RP | POS | | | 137968 | 125535 | 131907 | 46514.7 | 6.29E-01 | -0.07 | 0.37 |
| HILIC_NEG_144_92355a5_655 | | 144.9236 | 5.66 | | HILIC | NEG | | | 4.1E+07 | 8367085 | 4.2E+07 | 8481136 | 6.30E-01 | 0.07 | 0.34 |
| HILIC_NEG_82_97145a3_165 | | 82.9715 | 3.17 | | HILIC | NEG | | | 2849456 | 136042 | 2840891 | 124617 | 6.30E-01 | -0.07 | 0.64 |
| HILIC_POS_201_06099a4_826 | | 201.0610 | 4.83 | | HILIC | POS | | | 329868 | 159450 | 320249 | 133424 | 6.30E-01 | -0.07 | 0.77 |
| RP_POS_288_9209a15_398 | | 288.9209 | 15.40 | | RP | POS | | | 4538149 | 1242101 | 4456877 | 1225995 | 6.30E-01 | -0.07 | 0.65 |
| RP_POS_298_04147a15_508 | | 298.0415 | 15.51 | | RP | POS | | | 5341676 | 5937491 | 5021720 | 3690245 | 6.30E-01 | -0.07 | 0.77 |
| RP_POS_780_55334a12_529 | PC 36:5 (18:2_18:3) | 780.5533 | 12.53 | | RP | POS | 184.0744(100), 780.555(2) | 1 | 1.4E+07 | 7709833 | 1.3E+07 | 7671837 | 6.30E-01 | -0.07 | 0.65 |
| RP_POS_287_08865a4_37 | 5,7,4'-Trihydroxy-8-methylflavanone | 287.0887 | 4.37 | | RP | POS | 287.0884(100 | 2 | 339760 | 661050 | 306368 | 323721 | 6.30E-01 | -0.07 | 0.62 |
| HILIC_NEG_155_8656a7_744 | | 155.8656 | 7.74 | | HILIC | NEG | | | 1596512 | 635310 | 1638536 | 641981 | 6.31E-01 | 0.07 | 0.5 |
| RP_POS_419_31567a9_897 | | 419.3157 | 9.90 | | RP | POS | | | 2231020 | 1261357 | 2314004 | 1264678 | 6.31E-01 | 0.07 | 0.46 |
| RP_POS_581_38727a10_156 | | 581.3873 | 10.16 | | RP | POS | | | 244018 | 155948 | 255318 | 184252 | 6.31E-01 | 0.07 | 0.92 |
| RP_POS_592_46979a12_222 | | 592.4698 | 12.22 | | RP | POS | | | 533175 | 340879 | 555295 | 332883 | 6.31E-01 | 0.07 | 0.41 |
| RP_POS_261_10962a7_316 | | 261.1096 | 7.32 | | RP | POS | | | 2351518 | 1893601 | 2223853 | 1985222 | 6.31E-01 | -0.07 | 0.81 |
| RP_POS_319_15668a10_107 | | 319.1567 | 10.11 | | RP | POS | | | 303499 | 213244 | 288759 | 233488 | 6.31E-01 | -0.07 | 0.56 |
| RP_POS_363_19452a7_359 | | 363.1945 | 7.36 | | RP | POS | | | 644834 | 598581 | 610759 | 438766 | 6.31E-01 | -0.07 | 0.53 |
| RP_POS_808_58527a13_437 | | 808.5853 | 13.44 | | RP | POS | | | 1.3E+08 | 5.5E+07 | 1.3E+08 | 4.4E+07 | 6.31E-01 | -0.07 | 0.74 |
| RP_POS_391_28366a11_59 | | 391.2837 | 11.59 | | RP | POS | 149.023(100),71.0862(15) | | 6641630 | 2839113 | 6823482 | 2720876 | 6.32E-01 | 0.07 | 0.45 |
| RP_POS_191_02248a15_372 | | 191.0225 | 15.37 | | RP | POS | | | 3317361 | 1141819 | 3390117 | 1084210 | 6.32E-01 | 0.07 | 0.74 |
| RP_POS_348_26666a11_232 | | 348.2667 | 11.23 | | RP | POS | | | 7.05E+16 | 1.38E+17 | 8.05E+16 | 1.63E+17 | 6.32E-01 | 0.07 | 0.63 |
| RP_POS_900_56976a13_668 | | 900.5698 | 13.67 | | RP | POS | | | 880822 | 616932 | 923308 | 675158 | 6.32E-01 | 0.07 | 0.34 |
| HILIC_NEG_405_01743a1_241 | | 405.0174 | 1.24 | | HILIC | NEG | | | 348291 | 136591 | 339296 | 138027 | 6.32E-01 | -0.07 | 0.5 |
| RP_POS_257_0134a0_682 | | 257.0134 | 0.68 | | RP | POS | | | 546607 | 112334 | 539839 | 94789.4 | 6.32E-01 | -0.07 | 0.58 |
| RP_POS_369_1059a9_456 | | 369.1059 | 9.46 | | RP | POS | | | 199672 | 158547 | 188118 | 189957 | 6.32E-01 | -0.07 | 0.68 |
| RP_POS_615_92786a9_944 | | 615.9279 | 9.94 | | RP | POS | | | 1493149 | 736342 | 1445094 | 731351 | 6.32E-01 | -0.07 | 0.58 |
| RP_POS_130_05016a1_037 | L-5-Oxoproline | 130.0502 | 1.04 | | RP | POS | 84.0449(100), 130.0502(40) | 2 | 1375450 | 250122 | 1357609 | 289485 | 6.32E-01 | -0.07 | 0.45 |
| HILIC_NEG_291_0079a1_229 | | 291.0079 | 1.23 | | HILIC | NEG | | | 974085 | 353488 | 996830 | 343202 | 6.33E-01 | 0.07 | 0.35 |
| HILIC_POS_176_0657a3_911 | | 176.0657 | 3.91 | | HILIC | POS | | | 1394841 | 1127585 | 1465992 | 1055766 | 6.33E-01 | 0.07 | 0.68 |
| RP_NEG_324_99026a15_459 | | 324.9903 | 15.46 | | RP | NEG | | | 824459 | 119924 | 832440 | 124319 | 6.33E-01 | 0.07 | 0.45 |
| RP_NEG_957_20605a9_874 | | 957.2061 | 9.87 | | RP | NEG | | | 4747321 | 262315 | 4762933 | 217670 | 6.33E-01 | 0.07 | 0.86 |
| RP_POS_408_3681a10_931 | | 408.3681 | 10.93 | | RP | POS | | | 6697519 | 575010 | 6735800 | 596142 | 6.33E-01 | 0.07 | 0.78 |
| RP_POS_465_67355a10_402 | | 465.6736 | 10.40 | | RP | POS | | | 231893 | 141398 | 242201 | 170828 | 6.33E-01 | 0.07 | 0.64 |
| HILIC_NEG_199_80498a3_09 | | 199.8050 | 3.09 | | HILIC | NEG | | | 1E+07 | 901391 | 9974818 | 1045568 | 6.33E-01 | -0.07 | 0.6 |
| HILIC_NEG_212_91119a4_298 | | 212.9112 | 4.30 | | HILIC | NEG | | | 1590162 | 99019.1 | 1583806 | 95929 | 6.33E-01 | -0.07 | 0.59 |
| RP_NEG_173_90926a14_346 | | 173.9093 | 14.35 | | RP | NEG | | | 1058844 | 619614 | 1020171 | 567642 | 6.33E-01 | -0.07 | 0.47 |
| HILIC_POS_132_10208a1_322 | Isoleucine | 132.1021 | 1.32 | | HILIC | POS | 132.1027(100) | 2 | 539389 | 135279 | 547894 | 125777 | 6.33E-01 | 0.07 | 0.71 |
| RP_POS_572_36896a11_009 | | 572.3690 | 11.01 | | RP | POS | | | 1570888 | 517476 | 1604654 | 518746 | 6.34E-01 | 0.07 | 0.71 |
| HILIC_POS_154_02669a4_89 | | 154.0267 | 4.89 | | HILIC | POS | | | 216725 | 128455 | 208786 | 116431 | 6.34E-01 | -0.06 | 0.61 |
| RP_NEG_330_98019a8_017 | | 330.9802 | 8.02 | | RP | NEG | | | 1590928 | 431691 | 1564411 | 385818 | 6.34E-01 | -0.06 | 0.45 |
| RP_POS_259_13766a8_843 | | 259.1377 | 8.84 | | RP | POS | | | 205950 | 190949 | 194250 | 169547 | 6.34E-01 | -0.06 | 0.96 |
| RP_POS_820_52448a12_874 | | 820.5245 | 12.87 | | RP | POS | | | 4382480 | 2584986 | 4227387 | 2195090 | 6.34E-01 | -0.06 | 0.66 |
| RP_NEG_554_34814a10_724 | | 554.3481 | 10.72 | | RP | NEG | | | 1E+07 | 2548271 | 1E+07 | 2589359 | 6.34E-01 | -0.07 | 0.78 |
| RP_POS_636_43555a10_101 | | 636.4356 | 10.10 | | RP | POS | | | 1048550 | 572192 | 1011599 | 563161 | 6.34E-01 | -0.07 | 0.54 |
| HILIC_POS_146_05989a4_099 | Indole-3-carboxyaldehyde | 146.0599 | 4.10 | | HILIC | POS | 118.0658(100), 146.0602(60) | 2 | 2389016 | 5390926 | 2769075 | 6176550 | 6.34E-01 | 0.07 | 0.68 |
| RP_NEG_139_26991a0_678 | | 139.2699 | 0.68 | | RP | NEG | | | 282274 | 73377.6 | 287094 | 74885.3 | 6.35E-01 | 0.07 | 0.33 |
| RP_NEG_636_34967a10_816 | | 636.3497 | 10.82 | | RP | NEG | | | 2454508 | 547638 | 2492051 | 602206 | 6.35E-01 | 0.07 | 0.7 |
| RP_POS_184_07327a10_477 | | 184.0733 | 10.48 | | RP | POS | | | 2785903 | 491631 | 2818603 | 513122 | 6.35E-01 | 0.07 | 0.76 |
| RP_POS_403_78116a13_468 | | 403.7812 | 13.47 | | RP | POS | | | 2031766 | 1931263 | 2163333 | 2105801 | 6.35E-01 | 0.07 | 0.83 |
| RP_POS_478_32278a10_379 | | 478.3228 | 10.38 | | RP | POS | | | 439755 | 471407 | 471297 | 498006 | 6.35E-01 | 0.07 | 0.41 |
| RP_POS_585_34015a15_059 | | 585.3402 | 15.06 | | RP | POS | | | 4090080 | 1341495 | 4183175 | 1508422 | 6.35E-01 | 0.07 | 0.88 |
| HILIC_NEG_155_86589a3_214 | | 155.8659 | 3.21 | | HILIC | NEG | | | 1.9E+07 | 1494977 | 1.9E+07 | 1111508 | 6.35E-01 | 0.06 | 0.42 |
| RP_NEG_267_14407a9_816 | | 267.1441 | 9.82 | | RP | NEG | | | 88380 | 632828 | 126152 | 535839 | 6.35E-01 | 0.06 | 0.35 |
| RP_POS_562_32635a10_81 | | 562.3264 | 10.81 | | RP | POS | | | 382418 | 231787 | 396528 | 204228 | 6.35E-01 | 0.06 | 0.66 |
| HILIC_NEG_226_99525a1_232 | | 226.9953 | 1.23 | | HILIC | NEG | | | 1645932 | 578731 | 1608989 | 559899 | 6.35E-01 | -0.06 | 0.62 |
| RP_POS_261_22137a10_236 | | 261.2214 | 10.24 | | RP | POS | | | 3139477 | 262010 | 3123862 | 221125 | 6.35E-01 | -0.06 | 0.4 |
| RP_POS_514_39478a9_137 | | 514.3948 | 9.14 | | RP | POS | | | 2495177 | 1469571 | 2405205 | 1314270 | 6.35E-01 | -0.06 | 0.6 |
| RP_POS_520_0354a10_191 | | 520.0354 | 10.19 | | RP | POS | | | 996142 | 576550 | 1040667 | 769374 | 6.36E-01 | 0.07 | 0.54 |
| RP_NEG_465_80353a15_386 | | 465.8035 | 15.39 | | RP | NEG | | | 445267 | 173856 | 456628 | 177061 | 6.36E-01 | 0.06 | 0.44 |
| RP_POS_406_74768a13_214 | | 406.7477 | 13.21 | | RP | POS | | | 6648038 | 3539408 | 6858086 | 2975714 | 6.36E-01 | 0.06 | 0.91 |
| RP_NEG_241_14471a8_71 | | 241.1447 | 8.71 | | RP | NEG | | | 452568 | 99071.9 | 446008 | 103418 | 6.36E-01 | -0.06 | 0.54 |
| RP_POS_290_63913a10_011 | | 290.6391 | 10.01 | | RP | POS | | | 275828 | 182339 | 264995 | 153920 | 6.36E-01 | -0.06 | 0.75 |
| HILIC_POS_104_10733a1_257 | | 104.1073 | 1.26 | | HILIC | POS | 104.1079(100), 60.082(20) | | 1311125 | 588650 | 1271303 | 635240 | 6.36E-01 | -0.07 | 0.47 |
| HILIC_POS_673_52881a0_884 | | 673.5288 | 0.88 | | HILIC | POS | | | 240400 | 118726 | 248467 | 129792 | 6.37E-01 | 0.06 | 0.57 |
| RP_POS_412_30884a9_641 | | 412.3088 | 9.64 | | RP | POS | | | 2084766 | 1098591 | 2149564 | 918809 | 6.37E-01 | 0.06 | 0.85 |
| RP_NEG_242_98633a6_077 | | 242.9863 | 6.08 | | RP | NEG | | | 2058246 | 311738 | 2038574 | 298751 | 6.37E-01 | -0.06 | 0.4 |
| RP_POS_135_03203a15_327 | | 135.0320 | 15.33 | | RP | POS | | | 1.3E+07 | 3500000 | 1.3E+07 | 3619086 | 6.37E-01 | -0.06 | 0.41 |
| RP_POS_330_89822a15_41 | | 330.8982 | 15.41 | | RP | POS | | | 4923504 | 1881376 | 4818674 | 1369049 | 6.37E-01 | -0.06 | 0.88 |
| RP_POS_146_0601a2_883 | | 146.0601 | 2.88 | | RP | POS | 146.0819(100), 118.0864(80) | | 1.5E+07 | 2730559 | 1.5E+07 | 2630184 | 6.38E-01 | 0.06 | 0.73 |
| HILIC_NEG_183_93456a4_861 | | 183.9346 | 4.86 | | HILIC | NEG | | | 2748558 | 941148 | 2813288 | 1058802 | 6.38E-01 | 0.06 | 0.58 |
| RP_NEG_127_87019a14_334 | | 127.8702 | 14.33 | | RP | NEG | | | 512679 | 226377 | 526916 | 216473 | 6.38E-01 | 0.06 | 0.43 |
| RP_NEG_855_66119a14_94 | | 855.6612 | 14.94 | | RP | NEG | | | 2420683 | 1305504 | 2510192 | 1458912 | 6.38E-01 | 0.06 | 0.62 |
| HILIC_POS_118_08652a1_083 | | 118.0865 | 1.08 | | HILIC | POS | 118.0863(100) | | 3590057 | 1912203 | 3483273 | 1412236 | 6.38E-01 | -0.06 | 0.35 |
| HILIC_POS_398_215a5_934 | | 398.2150 | 5.93 | | HILIC | POS | | | 124600 | 101454 | 118303 | 94778.3 | 6.38E-01 | -0.06 | 0.58 |
| RP_POS_295_22421a10_107 | | 295.2242 | 10.11 | | RP | POS | | | 1912614 | 237388 | 1897941 | 220426 | 6.38E-01 | -0.06 | 0.88 |
| RP_POS_335_21951a8_481 | | 335.2195 | 8.48 | | RP | POS | | | 645495 | 226671 | 631569 | 206990 | 6.38E-01 | -0.06 | 0.53 |
| HILIC_NEG_293_04926a5_119 | | 293.0493 | 5.12 | | HILIC | NEG | | | 8324135 | 3012416 | 8529910 | 3364872 | 6.39E-01 | 0.06 | 0.49 |
| RP_NEG_357_89801a15_402 | | 357.8980 | 15.40 | | RP | NEG | | | 448672 | 128722 | 457250 | 138042 | 6.39E-01 | 0.06 | 0.62 |
| RP_NEG_870_5484a13_221 | | 870.5484 | 13.22 | | RP | NEG | | | 1.1E+07 | 4945312 | 1.2E+07 | 5017771 | 6.39E-01 | 0.06 | 0.3 |
| RP_POS_605_38159a14_348 | | 605.3816 | 14.35 | | RP | POS | | | 2306119 | 1079135 | 2377047 | 1130091 | 6.39E-01 | 0.06 | 0.79 |
| RP_POS_217_17941a8_601 | | 217.1794 | 8.60 | | RP | POS | | | 545447 | 133429 | 537203 | 124429 | 6.39E-01 | -0.06 | 0.7 |
| RP_POS_271_03217a15_419 | | 271.0322 | 15.42 | | RP | POS | | | 1102444 | 402803 | 1079472 | 315588 | 6.39E-01 | -0.06 | 0.45 |
| RP_POS_426_83084a15_399 | | 426.8308 | 15.40 | | RP | POS | | | 709085 | 250349 | 691572 | 290239 | 6.39E-01 | -0.06 | 0.73 |
| HILIC_POS_204_09808a1_353 | 1h-indole-3-butanoic acid | 204.0981 | 1.35 | | HILIC | POS | 204.0979(100) | 2 | 236147 | 135152 | 227750 | 127372 | 6.39E-01 | -0.06 | 0.39 |
| HILIC_NEG_268_98666a4_251 | | 268.9867 | 4.25 | | HILIC | NEG | | | 681190 | 276407 | 697828 | 245898 | 6.40E-01 | 0.06 | 0.6 |
| RP_NEG_382_73248a15_424 | | 382.7325 | 15.42 | | RP | NEG | | | 929148 | 317338 | 950130 | 337022 | 6.40E-01 | 0.06 | 0.57 |
| RP_POS_172_09454a15_254 | | 172.0945 | 15.25 | | RP | POS | | | 1271557 | 624723 | 1310789 | 602806 | 6.40E-01 | 0.06 | 0.31 |
| HILIC_POS_342_18329a0_814 | | 342.1833 | 0.81 | | HILIC | POS | | | 1584516 | 584500 | 1548424 | 546351 | 6.40E-01 | -0.06 | 0.53 |
| RP_NEG_338_78229a15_441 | | 338.7823 | 15.44 | | RP | NEG | | | 3757588 | 1437502 | 3664153 | 1483739 | 6.40E-01 | -0.06 | 0.49 |
| RP_POS_282_90463a15_349 | | 282.9046 | 15.35 | | RP | POS | | | 1335658 | 425897 | 1308018 | 438145 | 6.40E-01 | -0.06 | 0.62 |
| RP_POS_136_07561a1_225 | Octopamine | 136.0756 | 1.23 | | RP | POS | 91.0546(100), 136.0759(95) | 2 | 2623344 | 861505 | 2573499 | 700626 | 6.40E-01 | -0.06 | 0.5 |
| RP_NEG_1159_71497a10_932 | | 1159.7150 | 10.93 | | RP | NEG | | | 949469 | 297381 | 969370 | 323374 | 6.41E-01 | 0.06 | 0.67 |
| RP_POS_321_2399a10_777 | | 321.2399 | 10.78 | | RP | POS | | | 3762521 | 656146 | 3805375 | 686460 | 6.41E-01 | 0.06 | 0.94 |
| RP_POS_327_33731a9_652 | | 327.3373 | 9.65 | | RP | POS | | | 1093566 | 155429 | 1102482 | 125331 | 6.41E-01 | 0.06 | 0.56 |
| RP_POS_457_32346a8_624 | | 457.3235 | 8.62 | | RP | POS | | | 410388 | 255204 | 427617 | 282231 | 6.41E-01 | 0.06 | 0.53 |
| RP_POS_338_34137a11_049 | | 338.3414 | 11.05 | | RP | POS | 338.3405(100) | | 7142734 | 459909 | 7112694 | 480897 | 6.41E-01 | -0.06 | 0.68 |
| HILIC_NEG_294_98279a0_934 | | 294.9828 | 0.93 | | HILIC | NEG | | | 657652 | 74267 | 653000 | 71759.4 | 6.41E-01 | -0.06 | 0.49 |
| RP_POS_361_30655a10_366 | | 361.3066 | 10.37 | | RP | POS | | | 2278628 | 429507 | 2251123 | 432496 | 6.41E-01 | -0.06 | 0.95 |
| RP_POS_275_21179a4_732 | | 275.2118 | 4.73 | | RP | POS | | | 1.8E+07 | 3519503 | 1.7E+07 | 5071257 | 6.41E-01 | -0.07 | 0.68 |
| HILIC_NEG_568_36243a1_17 | LysoPC 18:0 (18:0/0:0) | 568.3624 | 1.17 | | HILIC | NEG | 283.2652(100) | 2 | 1824497 | 499805 | 1856913 | 514558 | 6.41E-01 | 0.06 | 0.66 |
| RP_POS_160_07582a2_884 | N-Acetylindole | 160.0758 | 2.88 | | RP | POS | 160.0963(100), 118.0658(60) | 2 | 1196754 | 223684 | 1210700 | 214334 | 6.41E-01 | 0.06 | 0.65 |
| RP_POS_123_04431a0_68 | 4-Hydroxybenzaldehyde | 123.0443 | 0.68 | | RP | POS | 95.0496(100), 123.0443(50) | 1 | 2503640 | 737124 | 2459694 | 647481 | 6.41E-01 | -0.06 | 0.52 |
| HILIC_POS_468_30917a1_349 | LysoPC 14:0 | 468.3092 | 1.35 | | HILIC | POS | 184.0745(100), 104.1078(75) | 2 | 9638346 | 3909282 | 9892563 | 4051092 | 6.41E-01 | 0.06 | 0.46 |
| RP_POS_342_15576a2_104 | | 342.1558 | 2.10 | | RP | POS | | | 627370 | 313981 | 653553 | 478327 | 6.42E-01 | 0.07 | 0.68 |

**Supplemantary table 1**: The non-targeted metabolomics results ordered by the VIP value (p value < 0.0003 are shaded dark green, p value < 0.05 are shaded light green, d values below 0 are blue and above 0 are red)

| | | | | | | | | | Cases | | Controls | | Welch test | Cohen's | |
|---|---|---|---|---|---|---|---|---|---|---|---|---|---|---|---|
| Feature_ID | Metabolite_name | Average_Mz | Average_RT_min_ | HMDB ID | RP_HILIC | POS_NEG | Cureted_MSMS | ID_level | Mean | SD | Mean2 | SD3 | p | d | VIP |
| HILIC_POS_116_07101a7_154 | | 116.0710 | 7.15 | | HILIC | POS | 70.0661(100), 116.0713(55) | | 7101062 | 1717487 | 7213626 | 1812312 | 6.42E-01 | 0.06 | 0.49 |
| HILIC_POS_203_05246a5_105 | | 203.0525 | 5.11 | | HILIC | POS | 203.0537(100) | | 5.1E+07 | 2.2E+07 | 5.3E+07 | 2.5E+07 | 6.42E-01 | 0.06 | 0.63 |
| RP_NEG_559_99561a0_678 | | 559.9956 | 0.68 | | RP | NEG | | | 487582 | 161161 | 498043 | 167702 | 6.42E-01 | 0.06 | 0.32 |
| RP_POS_298_31018a11_295 | | 298.3102 | 11.30 | | RP | POS | | | 324864 | 90260.8 | 331325 | 110133 | 6.42E-01 | 0.06 | 0.67 |
| RP_NEG_308_99857a15_473 | | 308.9986 | 15.47 | | RP | NEG | | | 1870768 | 169106 | 1859905 | 172420 | 6.42E-01 | -0.06 | 0.38 |
| RP_POS_133_03172a0_679 | | 133.0317 | 0.68 | | RP | POS | | | 4346109 | 1026026 | 4282052 | 990580 | 6.42E-01 | -0.06 | 0.53 |
| RP_POS_395_27634a10_822 | | 395.2763 | 10.82 | | RP | POS | | | 743921 | 767707 | 701529 | 566822 | 6.42E-01 | -0.06 | 0.59 |
| RP_POS_152_07068a2_508 | Acetaminophen | 152.0707 | 2.51 | | RP | POS | 152.07(100), 110.0607(70) | 1 | 1.9E+07 | 8.7E+07 | 2.6E+07 | 1.1E+08 | 6.42E-01 | 0.06 | 0.73 |
| RP_NEG_476_27875a10_182 | LysoPE 18:2 | 476.2788 | 10.18 | | RP | NEG | 279.2339(100),476.2809(5) | 1 | 4005888 | 1637719 | 4124078 | 2030493 | 6.42E-01 | 0.06 | 0.69 |
| HILIC_POS_150_05814a4_63 | L-Methionine | 150.0581 | 4.63 | | HILIC | POS | 104.0535(100), 133.0324(60) | 1 | 1.8E+07 | 5752583 | 1.7E+07 | 5369191 | 6.42E-01 | -0.06 | 0.56 |
| RP_NEG_415_2168a8_343 | | 415.2168 | 8.34 | | RP | NEG | | | 833357 | 503274 | 864380 | 478487 | 6.43E-01 | 0.06 | 0.47 |
| RP_POS_400_21667a4_992 | | 400.2167 | 4.99 | | RP | POS | | | 89136.5 | 36501 | 91732.9 | 44451 | 6.43E-01 | 0.06 | 0.67 |
| RP_POS_1019_63257a10_488 | | 1019.6326 | 10.49 | | RP | POS | | | 4978020 | 936497 | 4913202 | 1087442 | 6.43E-01 | -0.06 | 0.77 |
| RP_POS_333_24182a10_783 | | 333.2418 | 10.78 | | RP | POS | | | 292880 | 565883 | 256849 | 569001 | 6.43E-01 | -0.06 | 0.47 |
| RP_POS_394_91318a15_382 | | 394.9132 | 15.38 | | RP | POS | | | 268230 | 117269 | 261432 | 97864.8 | 6.43E-01 | -0.06 | 0.55 |
| RP_POS_401_76492a12_738 | | 401.7649 | 12.74 | | RP | POS | | | 1222285 | 958262 | 1159645 | 1014447 | 6.43E-01 | -0.06 | 0.64 |
| RP_POS_483_37601a11_782 | | 483.3760 | 11.78 | | RP | POS | | | 426541 | 437425 | 402444 | 325045 | 6.43E-01 | -0.06 | 0.51 |
| RP_POS_541_89264a10_37 | | 541.8926 | 10.37 | | RP | POS | | | 1542175 | 695037 | 1498836 | 675515 | 6.43E-01 | -0.06 | 0.6 |
| RP_POS_548_27435a10_181 | | 548.2744 | 10.18 | | RP | POS | | | 768653 | 236626 | 752869 | 259218 | 6.43E-01 | -0.06 | 0.56 |
| HILIC_POS_221_16968a1_673 | | 221.1697 | 1.67 | | HILIC | POS | | | 218015 | 358604 | 240632 | 357557 | 6.44E-01 | 0.06 | 0.5 |
| HILIC_POS_59_06134a0_749 | | 59.0613 | 0.75 | | HILIC | POS | | | 5876628 | 3028522 | 6086379 | 3543730 | 6.44E-01 | 0.06 | 0.6 |
| RP_NEG_1043_15259a9_873 | | 1043.1526 | 9.87 | | RP | NEG | | | 3478186 | 209865 | 3491284 | 204716 | 6.44E-01 | 0.06 | 0.96 |
| RP_POS_559_0672a10_434 | | 559.0672 | 10.43 | | RP | POS | | | 299996 | 188102 | 311801 | 185207 | 6.44E-01 | 0.06 | 0.76 |
| RP_POS_631_77441a10_363 | | 631.7744 | 10.36 | | RP | POS | | | 337023 | 244756 | 352938 | 257278 | 6.44E-01 | 0.06 | 0.54 |
| RP_POS_313_14319a9_475 | Benzyl butyl phthalate | 313.1432 | 9.48 | | RP | POS | 91.0546(100), 149.023(40) | 2 | 1040262 | 170677 | 1030143 | 150988 | 6.44E-01 | -0.06 | 0.97 |
| HILIC_POS_159_11272a4_821 | | 159.1127 | 4.82 | | HILIC | POS | | | 399161 | 879099 | 349639 | 695038 | 6.44E-01 | -0.06 | 0.53 |
| RP_NEG_216_78081a14_343 | | 216.7808 | 14.34 | | RP | NEG | | | 754153 | 330040 | 734561 | 292752 | 6.44E-01 | -0.06 | 0.45 |
| RP_POS_334_17734a4_176 | | 334.1773 | 4.18 | | RP | POS | | | 48949 | 66818.3 | 44763 | 65696.3 | 6.44E-01 | -0.06 | 0.66 |
| HILIC_POS_169_05826a6_136 | | 169.0583 | 6.14 | | HILIC | POS | | | 2.7E+07 | 2682798 | 2.7E+07 | 2744814 | 6.45E-01 | 0.06 | 0.81 |
| RP_NEG_275_16513a9_065 | | 275.1651 | 9.07 | | RP | NEG | | | 1192594 | 47009.1 | 1195441 | 43640.5 | 6.45E-01 | 0.06 | 0.81 |
| RP_POS_647_9552a11_662 | | 647.9552 | 11.66 | | RP | POS | | | 164955 | 77398.8 | 170780 | 103684 | 6.45E-01 | 0.06 | 0.65 |
| RP_POS_898_51581a13_709 | | 898.5158 | 13.71 | | RP | POS | | | 743495 | 364135 | 767585 | 396667 | 6.45E-01 | 0.06 | 0.97 |
| RP_NEG_158_97874a15_319 | | 158.9787 | 15.32 | | RP | NEG | | | 7603741 | 573307 | 7571484 | 452484 | 6.45E-01 | -0.06 | 0.9 |
| RP_NEG_455_60254a15_497 | | 455.6025 | 15.50 | | RP | NEG | | | 940003 | 63660.1 | 935789 | 69333.9 | 6.45E-01 | -0.06 | 0.34 |
| RP_POS_143_95905a0_621 | | 143.9591 | 0.62 | | RP | POS | | | 776275 | 277851 | 758698 | 279308 | 6.45E-01 | -0.06 | 0.64 |
| RP_POS_229_21616a11_665 | | 229.2162 | 11.67 | | RP | POS | | | 265082 | 133339 | 256483 | 139188 | 6.45E-01 | -0.06 | 0.86 |
| RP_POS_344_66089a5_649 | | 344.6609 | 5.65 | | RP | POS | | | 1991239 | 1327896 | 1913421 | 1152545 | 6.45E-01 | -0.06 | 1.09 |
| RP_POS_387_1319a8_842 | | 387.1319 | 8.84 | | RP | POS | | | 219697 | 107214 | 212788 | 111928 | 6.45E-01 | -0.06 | 0.58 |
| RP_POS_533_37982a10_373 | | 533.3798 | 10.37 | | RP | POS | | | 761348 | 396379 | 734397 | 453183 | 6.45E-01 | -0.06 | 0.85 |
| RP_POS_450_99796a10_425 | | 450.9980 | 10.43 | | RP | POS | | | 140247 | 69114.4 | 146318 | 114608 | 6.46E-01 | 0.07 | 0.41 |
| RP_NEG_551_12744a8_368 | | 551.1274 | 8.37 | | RP | NEG | | | 862059 | 36525.4 | 864327 | 35748.9 | 6.46E-01 | 0.06 | 0.47 |
| RP_POS_1002_65741a10_356 | | 1002.6574 | 10.36 | | RP | POS | | | 577486 | 668169 | 620716 | 704953 | 6.46E-01 | 0.06 | 0.57 |
| RP_POS_237_14801a9_79 | | 237.1480 | 9.79 | | RP | POS | | | 1218344 | 173195 | 1231235 | 228379 | 6.46E-01 | 0.06 | 0.66 |
| HILIC_NEG_134_89479a1_48 | | 134.8948 | 1.48 | | HILIC | NEG | | | 2.7E+07 | 6164573 | 2.7E+07 | 6373122 | 6.46E-01 | -0.06 | 0.32 |
| HILIC_NEG_197_90666a0_897 | | 197.9067 | 0.90 | | HILIC | NEG | | | 153732 | 35906.1 | 151276 | 41598.3 | 6.46E-01 | -0.06 | 0.5 |
| RP_NEG_344_81592a15_396 | | 344.8159 | 15.40 | | RP | NEG | | | 2627541 | 566394 | 2592838 | 539290 | 6.46E-01 | -0.06 | 0.52 |
| RP_POS_285_24225a9_677 | | 285.2423 | 9.68 | | RP | POS | | | 1495122 | 181276 | 1483786 | 179974 | 6.46E-01 | -0.06 | 0.66 |
| RP_POS_303_14328a6_227 | | 303.1433 | 6.23 | | RP | POS | | | 1533279 | 149507 | 1523769 | 152703 | 6.46E-01 | -0.06 | 0.85 |
| RP_POS_505_30252a13_224 | | 505.3025 | 13.22 | | RP | POS | | | 403914 | 190794 | 392590 | 170355 | 6.46E-01 | -0.06 | 0.5 |
| RP_POS_708_51007a13_696 | | 708.5101 | 13.70 | | RP | POS | | | 8908760 | 2156544 | 8775992 | 2078468 | 6.46E-01 | -0.06 | 0.88 |
| RP_POS_386_19949a10_834 | | 386.1995 | 10.83 | | RP | POS | | | 557099 | 287758 | 577736 | 360699 | 6.47E-01 | 0.06 | 0.97 |
| RP_POS_419_35074a13_171 | | 419.3507 | 13.17 | | RP | POS | | | 741425 | 358626 | 763410 | 345405 | 6.47E-01 | 0.06 | 0.85 |
| HILIC_POS_147_0439a5_117 | | 147.0439 | 5.12 | | HILIC | POS | | | 234574 | 85242 | 229653 | 72500 | 6.47E-01 | -0.06 | 0.56 |
| RP_NEG_595_28986a11_389 | | 595.2899 | 11.39 | | RP | NEG | | | 159737 | 219260 | 147033 | 188245 | 6.47E-01 | -0.06 | 0.5 |
| RP_NEG_915_12775a9_538 | | 915.1278 | 9.54 | | RP | NEG | | | 4783194 | 291520 | 4766665 | 238116 | 6.47E-01 | -0.06 | 1.1 |
| RP_POS_189_09109a7_709 | | 189.0911 | 7.71 | | RP | POS | | | 2835246 | 3271066 | 2629603 | 3297819 | 6.47E-01 | -0.06 | 0.51 |
| RP_POS_266_13855a7_708 | | 266.1386 | 7.71 | | RP | POS | | | 1389401 | 2000722 | 1263749 | 2008385 | 6.47E-01 | -0.06 | 0.59 |
| RP_POS_402_0108a0_678 | | 402.0108 | 0.68 | | RP | POS | | | 1095776 | 199334 | 1083936 | 180067 | 6.47E-01 | -0.06 | 0.45 |
| RP_POS_454_82892a9_841 | | 454.8289 | 9.84 | | RP | POS | | | 2308536 | 1077955 | 2240115 | 1107721 | 6.47E-01 | -0.06 | 0.52 |
| RP_POS_556_89661a10_171 | | 556.8966 | 10.17 | | RP | POS | | | 2976880 | 1304023 | 2898054 | 1218743 | 6.47E-01 | -0.06 | 0.8 |
| RP_NEG_624_33978a8_632 | | 624.3398 | 8.63 | | RP | NEG | | | 750551 | 1908877 | 660549 | 853330 | 6.47E-01 | -0.07 | 0.39 |
| HILIC_POS_118_0864a7_75 | | 118.0864 | 7.75 | | HILIC | POS | 140.069(100), 74.0974(40) | | 1.9E+07 | 5680859 | 1.9E+07 | 5753116 | 6.48E-01 | 0.06 | 0.46 |
| HILIC_NEG_270_00513a0_939 | | 270.0051 | 0.94 | | HILIC | NEG | | | 65667.6 | 51668.5 | 68736.9 | 46894.8 | 6.48E-01 | 0.06 | 0.67 |
| RP_NEG_598_29083a10_518 | | 598.2908 | 10.52 | | RP | NEG | | | 712118 | 58579.5 | 716160 | 69535.1 | 6.48E-01 | 0.06 | 0.4 |
| RP_NEG_626_27716a10_359 | | 626.2772 | 10.36 | | RP | NEG | | | 1811561 | 297477 | 1830657 | 313199 | 6.48E-01 | 0.06 | 0.97 |
| RP_NEG_691_12646a9_525 | | 691.1265 | 9.53 | | RP | NEG | | | 2111179 | 118509 | 2118165 | 105740 | 6.48E-01 | 0.06 | 0.66 |
| RP_POS_282_16974a7_659 | | 282.1697 | 7.66 | | RP | POS | | | 149568 | 79287.9 | 154087 | 66134.6 | 6.48E-01 | 0.06 | 0.82 |
| RP_POS_423_7692a13_419 | | 423.7692 | 13.42 | | RP | POS | | | 1161242 | 899577 | 1214684 | 816283 | 6.48E-01 | 0.06 | 0.52 |
| HILIC_NEG_182_98698a6_135 | | 182.9870 | 6.14 | | HILIC | NEG | | | 836809 | 280351 | 819593 | 271361 | 6.48E-01 | -0.06 | 0.86 |
| HILIC_POS_612_32275a1_592 | | 612.3228 | 1.59 | | HILIC | POS | | | 459144 | 225603 | 445745 | 204336 | 6.48E-01 | -0.06 | 0.7 |
| RP_NEG_217_86526a15_572 | | 217.8653 | 15.57 | | RP | NEG | | | 620207 | 315830 | 602088 | 267165 | 6.48E-01 | -0.06 | 0.41 |
| RP_NEG_225_79858a15_499 | | 225.7986 | 15.50 | | RP | NEG | | | 285468 | 39529.7 | 282917 | 41953.6 | 6.48E-01 | -0.06 | 0.67 |
| RP_NEG_431_19589a11_605 | | 431.1959 | 11.61 | | RP | NEG | | | 1504612 | 377267 | 1480061 | 406117 | 6.48E-01 | -0.06 | 0.67 |
| RP_NEG_509_28943a11_701 | | 509.2894 | 11.70 | | RP | NEG | | | 161561 | 123616 | 154523 | 102442 | 6.48E-01 | -0.06 | 0.73 |
| RP_POS_278_15994a6_595 | | 278.1599 | 6.60 | | RP | POS | | | 2596599 | 2504244 | 2441432 | 2474952 | 6.48E-01 | -0.06 | 0.47 |
| RP_POS_304_26099a11_236 | | 304.2610 | 11.24 | | RP | POS | | | 2099594 | 1266790 | 2026690 | 1075608 | 6.48E-01 | -0.06 | 0.71 |
| RP_POS_361_22183a9_576 | | 361.2218 | 9.58 | | RP | POS | | | 585558 | 159532 | 578018 | 73271.6 | 6.48E-01 | -0.06 | 0.29 |
| RP_POS_447_31033a9_332 | | 447.3103 | 9.33 | | RP | POS | | | 596865 | 340821 | 576180 | 323524 | 6.48E-01 | -0.06 | 0.58 |
| RP_POS_643_45013a11_496 | | 643.4501 | 11.50 | | RP | POS | | | 161666 | 86232.9 | 156671 | 74433.9 | 6.48E-01 | -0.06 | 0.53 |
| HILIC_POS_176_07411a1_047 | | 176.0741 | 1.05 | | HILIC | POS | | | 3682306 | 3237604 | 3954557 | 5133270 | 6.49E-01 | 0.07 | 0.83 |
| RP_NEG_510_09592a2_885 | | 510.0959 | 2.89 | | RP | NEG | | | 321780 | 92814.2 | 327485 | 90818.3 | 6.49E-01 | 0.06 | 0.58 |
| RP_POS_238_0672a11_586 | | 238.0672 | 11.59 | | RP | POS | | | 4294582 | 2055209 | 4408609 | 1621587 | 6.49E-01 | 0.06 | 0.65 |
| RP_POS_704_52191a12_435 | | 704.5219 | 12.44 | | RP | POS | | | 6941762 | 5276388 | 7278876 | 5540679 | 6.49E-01 | 0.06 | 0.39 |
| RP_POS_213_14603a7_42 | | 213.1460 | 7.42 | | RP | POS | | | 4123377 | 1303618 | 4046964 | 1158702 | 6.49E-01 | -0.06 | 1.23 |
| RP_POS_334_18613a6_575 | | 334.1861 | 6.58 | | RP | POS | | | 255743 | 82305.1 | 250349 | 90363.9 | 6.49E-01 | -0.06 | 0.44 |
| RP_POS_591_26251a10_45 | | 591.2625 | 10.45 | | RP | POS | | | 246107 | 105561 | 240295 | 81443.8 | 6.49E-01 | -0.06 | 0.48 |
| RP_POS_390_22333a6_783 | | 390.2233 | 6.78 | | RP | POS | | | 1743249 | 153200 | 1753200 | 166386 | 6.50E-01 | 0.06 | 0.79 |
| HILIC_POS_151_1442a1_467 | | 151.1442 | 1.47 | | HILIC | POS | | | 2.3E+07 | 7E+07 | 1.9E+07 | 5.6E+07 | 6.50E-01 | -0.06 | 0.35 |
| RP_NEG_259_00095a15_477 | | 259.0010 | 15.48 | | RP | NEG | | | 1543321 | 281724 | 1526525 | 260712 | 6.50E-01 | -0.06 | 0.9 |
| RP_NEG_679_18848a9_7 | | 679.1885 | 9.70 | | RP | NEG | | | 395981 | 40419.4 | 393501 | 39544.1 | 6.50E-01 | -0.06 | 0.59 |
| RP_NEG_819_15222a9_347 | | 819.1522 | 9.35 | | RP | NEG | | | 1427149 | 139234 | 1418353 | 143761 | 6.50E-01 | -0.06 | 0.77 |
| RP_POS_1156_32288a13_201 | | 1156.3229 | 13.20 | | RP | POS | | | 1390186 | 366868 | 1365435 | 424348 | 6.50E-01 | -0.06 | 0.78 |
| RP_POS_155_10651a10_361 | | 155.1065 | 10.36 | | RP | POS | | | 1879793 | 304672 | 1860638 | 312780 | 6.50E-01 | -0.06 | 0.51 |
| RP_POS_386_21906a11_145 | | 386.2191 | 11.15 | | RP | POS | | | 494093 | 306317 | 473652 | 346866 | 6.50E-01 | -0.06 | 0.54 |
| RP_POS_712_49274a9_901 | | 712.4927 | 9.90 | | RP | POS | | | 1243637 | 620526 | 1206311 | 585500 | 6.50E-01 | -0.06 | 0.62 |
| RP_NEG_265_14783a9_881 | | 265.1478 | 9.88 | | RP | NEG | 265.1465(100), 96.9601(65) | | 4481034 | 7159073 | 4080317 | 5843510 | 6.51E-01 | -0.06 | 0.46 |
| HILIC_NEG_114_02452a1_354 | | 114.0245 | 1.35 | | HILIC | NEG | | | 265520 | 86048.7 | 259948 | 93545.1 | 6.51E-01 | -0.06 | 0.62 |
| RP_NEG_266_10385a8_955 | | 266.1039 | 8.96 | | RP | NEG | | | 2017003 | 70866.9 | 2013055 | 57227.5 | 6.51E-01 | -0.06 | 0.47 |
| RP_NEG_475_16275a10_894 | | 475.1628 | 10.89 | | RP | NEG | | | 318660 | 141091 | 310008 | 138561 | 6.51E-01 | -0.06 | 0.93 |
| RP_POS_339_28683a11_157 | | 339.2868 | 11.16 | | RP | POS | | | 1841113 | 928553 | 1795577 | 521229 | 6.51E-01 | -0.06 | 0.41 |
| RP_POS_355_09058a9_207 | | 355.0906 | 9.21 | | RP | POS | | | 508332 | 339427 | 487769 | 325426 | 6.51E-01 | -0.06 | 0.6 |
| RP_POS_498_28827a8_464 | | 498.2883 | 8.46 | | RP | POS | | | 435282 | 738113 | 393528 | 615558 | 6.51E-01 | -0.06 | 0.33 |
| RP_POS_651_95886a10_27 | | 651.9589 | 10.27 | | RP | POS | | | 1312175 | 643166 | 1264793 | 855151 | 6.51E-01 | -0.06 | 0.72 |
| RP_POS_414_18964a9_587 | | 414.1896 | 9.59 | | RP | POS | | | 895498 | 962529 | 995491 | 2013260 | 6.52E-01 | 0.07 | 0.66 |
| RP_POS_517_34259a10_836 | | 517.3426 | 10.84 | | RP | POS | | | 1520227 | 745370 | 1616345 | 2004328 | 6.52E-01 | 0.07 | 0.59 |
| HILIC_NEG_130_92883a4_404 | | 130.9288 | 4.40 | | HILIC | NEG | | | 815339 | 197225 | 827074 | 184312 | 6.52E-01 | 0.06 | 0.88 |
| RP_NEG_146_92348a0_647 | | 146.9235 | 0.65 | | RP | NEG | | | 587642 | 224455 | 601819 | 234154 | 6.52E-01 | 0.06 | 0.65 |
| RP_POS_304_93338a15_398 | | 304.9334 | 15.40 | | RP | POS | | | 4169026 | 1157306 | 4234760 | 984583 | 6.52E-01 | 0.06 | 0.74 |
| HILIC_NEG_118_89951a3_135 | | 118.8995 | 3.14 | | HILIC | NEG | | | 486152 | 149655 | 477027 | 146576 | 6.52E-01 | -0.06 | 0.53 |
| RP_POS_1010_64484a10_485 | | 1010.6448 | 10.49 | | RP | POS | | | 789806 | 468607 | 764035 | 368179 | 6.52E-01 | -0.06 | 0.57 |
| RP_POS_163_97652a0_62 | | 163.9765 | 0.62 | | RP | POS | | | 8327853 | 1672580 | 8213115 | 2001460 | 6.52E-01 | -0.06 | 0.44 |
| HILIC_POS_394_31531a7_781 | | 394.3153 | 7.78 | | HILIC | POS | | | 1677429 | 530175 | 1709167 | 503138 | 6.53E-01 | 0.06 | 0.39 |
| RP_POS_433_78293a10_925 | | 433.7829 | 10.93 | | RP | POS | | | 207463 | 119728 | 215823 | 148494 | 6.53E-01 | 0.06 | 0.62 |
| RP_POS_491_32779a10_713 | | 491.3278 | 10.71 | | RP | POS | | | 1148339 | 407532 | 1175334 | 464445 | 6.53E-01 | 0.06 | 1.05 |
| HILIC_NEG_203_79877a3_231 | | 203.7988 | 3.23 | | HILIC | NEG | | | 224263 | 111673 | 217496 | 108528 | 6.53E-01 | -0.06 | 0.81 |
| RP_NEG_198_93665a15_769 | | 198.9367 | 15.77 | | RP | NEG | | | 1.1E+07 | 7037167 | 1E+07 | 7208779 | 6.53E-01 | -0.06 | 0.46 |
| RP_POS_361_30624a10_253 | | 361.3062 | 10.25 | | RP | POS | | | 1825168 | 216991 | 1812021 | 211668 | 6.53E-01 | -0.06 | 0.88 |
| RP_POS_388_35468a12_068 | | 388.3547 | 12.07 | | RP | POS | | | 3655170 | 6757185 | 3269014 | 5830592 | 6.53E-01 | -0.06 | 0.55 |
| RP_POS_430_27536a11_599 | | 430.2754 | 11.60 | | RP | POS | | | 640023 | 129621 | 632587 | 113409 | 6.53E-01 | -0.06 | 0.58 |
| RP_POS_183_08037a8_082 | Benzophenone | 183.0804 | 8.08 | | RP | POS | 105.0338(100), 183.081(20) | 2 | 2047686 | 603548 | 2010258 | 612149 | 6.53E-01 | -0.06 | 0.57 |
| RP_NEG_411_65305a15_495 | | 411.6531 | 15.50 | | RP | NEG | | | 869074 | 67377.2 | 873494 | 75994.4 | 6.54E-01 | 0.06 | 0.49 |
| RP_NEG_435_21014a11_588 | | 435.2101 | 11.59 | | RP | NEG | | | 82993.6 | 56712.1 | 86458.1 | 56300.7 | 6.54E-01 | 0.06 | 0.53 |
| RP_POS_144_08078a2_883 | | 144.0808 | 2.88 | | RP | POS | | | 7642493 | 1443096 | 7728443 | 1368817 | 6.54E-01 | 0.06 | 0.71 |
| RP_POS_209_03352a15_381 | | 209.0335 | 15.38 | | RP | POS | | | 904390 | 397368 | 928660 | 394974 | 6.54E-01 | 0.06 | 0.42 |
| RP_POS_290_15997a4_298 | | 290.1600 | 4.30 | | RP | POS | | | 198278 | 88452.7 | 204228 | 103985 | 6.54E-01 | 0.06 | 0.6 |
| RP_POS_318_24026a9_27 | | 318.2403 | 9.27 | | RP | POS | | | 170313 | 181214 | 182865 | 222771 | 6.54E-01 | 0.06 | 0.97 |
| RP_NEG_387_18521a6_827 | | 387.1852 | 6.83 | | RP | NEG | 387.1831(100) | | 818227 | 588042 | 781928 | 596919 | 6.54E-01 | -0.06 | 0.37 |
| RP_NEG_755_13879a9_791 | | 755.1388 | 9.79 | | RP | NEG | | | 1271389 | 61520.4 | 1267456 | 66298.3 | 6.54E-01 | -0.06 | 0.71 |
| RP_POS_277_17993a8_304 | | 277.1799 | 8.30 | | RP | POS | | | 1526759 | 254002 | 1512332 | 218790 | 6.54E-01 | -0.06 | 0.84 |
| RP_POS_313_16238a5_358 | | 313.1624 | 5.36 | | RP | POS | | | 378411 | 308760 | 360263 | 285649 | 6.54E-01 | -0.06 | 0.98 |
| RP_POS_463_34225a9_841 | | 463.3423 | 9.84 | | RP | POS | | | 6580648 | 2498583 | 6405296 | 3131519 | 6.54E-01 | -0.06 | 0.78 |
| RP_POS_568_87787a9_574 | | 568.8779 | 9.57 | | RP | POS | | | 520679 | 335458 | 541582 | 347172 | 6.55E-01 | 0.06 | 0.75 |
| RP_POS_323_25549a11_075 | | 323.2555 | 11.08 | | RP | POS | | | 1813685 | 500953 | 1781113 | 558202 | 6.55E-01 | -0.06 | 0.9 |
| RP_POS_328_23035a10_634 | | 328.2304 | 10.63 | | RP | POS | | | 5307224 | 1474819 | 5214256 | 1563354 | 6.55E-01 | -0.06 | 0.42 |
| RP_POS_424_36346a10_526 | | 424.3635 | 10.53 | | RP | POS | | | 3551324 | 645809 | 3513425 | 597106 | 6.55E-01 | -0.06 | 0.85 |
| RP_POS_758_56732a11_143 | | 758.5673 | 11.14 | | RP | POS | | | 2152136 | 480284 | 2123213 | 468421 | 6.55E-01 | -0.06 | 0.56 |
| RP_POS_156_95262a15_489 | | 156.9526 | 15.49 | | RP | POS | | | 1401821 | 91441 | 1407301 | 88708.1 | 6.56E-01 | 0.06 | 0.96 |
| RP_POS_330_99765a0_674 | | 330.9977 | 0.67 | | RP | POS | | | 632358 | 117887 | 639174 | 106554 | 6.56E-01 | 0.06 | 0.48 |
| RP_POS_539_04822a10_321 | | 539.0482 | 10.32 | | RP | POS | | | 811455 | 465915 | 848010 | 694438 | 6.56E-01 | 0.06 | 0.4 |
| RP_POS_739_51343a12_356 | | 739.5134 | 12.36 | | RP | POS | | | 476298 | 300050 | 494162 | 286998 | 6.56E-01 | 0.06 | 0.88 |
| RP_NEG_1021_21777a9_504 | | 1021.2178 | 9.50 | | RP | NEG | | | 1224530 | 136681 | 1216401 | 130574 | 6.56E-01 | -0.06 | 0.88 |
| RP_NEG_283_22778a9_844 | | 283.2278 | 9.84 | | RP | NEG | | | 402688 | 84515.9 | 397507 | 85303.7 | 6.56E-01 | -0.06 | 0.99 |
| RP_POS_384_34766a11_049 | | 384.3477 | 11.05 | | RP | POS | | | 5077081 | 258824 | 5059000 | 326133 | 6.56E-01 | -0.06 | 0.63 |
| RP_POS_510_35458a10_659 | | 510.3546 | 10.66 | | RP | POS | | | 3.2E+07 | 1.1E+07 | 3.2E+07 | 9586026 | 6.56E-01 | -0.06 | 0.63 |
| HILIC_POS_418_19409a6_507 | | 418.1941 | 6.51 | | HILIC | POS | | | 68643.7 | 45839.5 | 71539.3 | 49106.7 | 6.57E-01 | 0.06 | 1.14 |
| RP_NEG_514_27411a13_287 | | 514.2741 | 13.29 | | RP | NEG | | | 946452 | 185808 | 958037 | 195097 | 6.57E-01 | 0.06 | 0.96 |
| RP_POS_270_91025a15_397 | | 270.9103 | 15.40 | | RP | POS | | | 2.8E+07 | 6890511 | 2.8E+07 | 6190939 | 6.57E-01 | 0.06 | 1.03 |
| RP_POS_475_67593a10_233 | | 475.6759 | 10.23 | | RP | POS | | | 584050 | 343425 | 604242 | 323794 | 6.57E-01 | 0.06 | 0.58 |
| RP_POS_432_31064a8_515 | | 432.3106 | 8.52 | | RP | POS | 414.3015(100), 339.2697(40) | | 1921349 | 1583055 | 1825839 | 1563360 | 6.57E-01 | -0.06 | 0.34 |
| HILIC_NEG_194_90588a3_196 | | 194.9059 | 3.20 | | HILIC | NEG | | | 1781633 | 101547 | 1775622 | 96477.3 | 6.57E-01 | -0.06 | 0.45 |
| RP_POS_213_10986a15_195 | | 213.1099 | 15.20 | | RP | POS | | | 2642580 | 1112045 | 2570182 | 1257032 | 6.57E-01 | -0.06 | 0.62 |
| RP_POS_250_21396a10_169 | | 250.2140 | 10.17 | | RP | POS | | | 1111721 | 217332 | 1097906 | 236211 | 6.57E-01 | -0.06 | 0.67 |
| RP_POS_379_28955a12_798 | | 379.2896 | 12.80 | | RP | POS | | | 1078749 | 867155 | 1030325 | 734977 | 6.57E-01 | -0.06 | 0.78 |
| RP_POS_411_24521a11_591 | | 411.2452 | 11.59 | | RP | POS | | | 933844 | 221097 | 917774 | 296236 | 6.57E-01 | -0.06 | 1 |
| RP_NEG_230_98631a7_573 | | 230.9863 | 7.57 | | RP | NEG | | | 2053715 | 201129 | 2066824 | 228692 | 6.58E-01 | 0.06 | 0.52 |
| RP_NEG_562_29596a13_281 | | 562.2960 | 13.28 | | RP | NEG | | | 424528 | 92871.7 | 430197 | 94515.4 | 6.58E-01 | 0.06 | 1.02 |
| RP_POS_172_13319a10_362 | | 172.1332 | 10.36 | | RP | POS | | | 1.7E+07 | 1519575 | 1.7E+07 | 1548574 | 6.58E-01 | 0.06 | 0.89 |
| RP_POS_672_07184a11_449 | | 672.0718 | 11.45 | | RP | POS | | | 126750 | 67291.3 | 131482 | 86132.3 | 6.58E-01 | 0.06 | 0.68 |
| HILIC_NEG_195_81094a3_089 | | 195.8109 | 3.09 | | HILIC | NEG | | | 1.9E+07 | 1641897 | 1.9E+07 | 1935146 | 6.58E-01 | -0.06 | 0.64 |
| HILIC_NEG_270_79727a3_252 | | 270.7973 | 3.25 | | HILIC | NEG | | | 1424556 | 212715 | 1412195 | 196835 | 6.58E-01 | -0.06 | 0.42 |
| HILIC_POS_116_10733a0_849 | | 116.1073 | 0.85 | | HILIC | POS | | | 1365650 | 404032 | 1339674 | 447985 | 6.58E-01 | -0.06 | 0.43 |
| RP_NEG_664_32104a5_651 | | 664.3210 | 5.65 | | RP | NEG | | | 566031 | 178539 | 555803 | 160611 | 6.58E-01 | -0.06 | 0.68 |

**Supplemantary table 1**: The non-targeted metabolomics results ordered by the VIP value (p value < 0.0003 are shaded dark green, p value < 0.05 are shaded light green, d values below 0 are blue and above 0 are red)

| | | | | | | | | | Cases | | Controls | | Welch test | Cohen's | |
|---|---|---|---|---|---|---|---|---|---|---|---|---|---|---|---|
| Feature_ID | Metabolite_name | Average_Mz | Average_RT_min_ | HMDB ID | RP_HILIC | POS_NEG | Cureted_MSMS | ID_level | Mean | SD | Mean2 | SD3 | p | d | VIP |
| RP_POS_518_42371a13_81 | | 518.4237 | 13.81 | | RP | POS | | | 2453298 | 859214 | 2393926 | 1074306 | 6.58E-01 | -0.06 | 0.53 |
| RP_POS_844_53528a13_784 | | 844.5353 | 13.78 | | RP | POS | | | 668178 | 386587 | 647258 | 307295 | 6.58E-01 | -0.06 | 0.94 |
| RP_POS_205_09744a2_482 | Tryptophan | 205.0974 | 2.48 | | RP | POS | 188.0708(100), 146.0602(60) | 1 | 1.3E+08 | 2.5E+07 | 1.3E+08 | 2.4E+07 | 6.58E-01 | 0.06 | 0.68 |
| RP_POS_187_07159a2_505 | | 187.0716 | 2.51 | | RP | POS | | | 1387096 | 679212 | 1430603 | 753380 | 6.59E-01 | 0.06 | 0.65 |
| RP_POS_214_17976a6_871 | | 214.1798 | 6.87 | | RP | POS | | | 583677 | 70845 | 588490 | 86446.8 | 6.59E-01 | 0.06 | 0.59 |
| RP_POS_276_22983a10_343 | | 276.2298 | 10.34 | | RP | POS | | | 72874.3 | 113131 | 80169.8 | 126804 | 6.59E-01 | 0.06 | 0.97 |
| RP_POS_327_21631a10_598 | | 327.2163 | 10.60 | | RP | POS | | | 912050 | 606534 | 950176 | 651203 | 6.59E-01 | 0.06 | 1.26 |
| RP_POS_391_71027a14_35 | | 391.7103 | 14.35 | | RP | POS | | | 186655 | 171750 | 195838 | 133270 | 6.59E-01 | 0.06 | 0.61 |
| RP_POS_283_65964a10_487 | | 283.6596 | 10.49 | | RP | POS | | | 270528 | 342034 | 250577 | 321142 | 6.59E-01 | -0.06 | 0.37 |
| RP_POS_383_29526a11_593 | | 383.2953 | 11.59 | | RP | POS | | | 1213262 | 180883 | 1202016 | 190837 | 6.59E-01 | -0.06 | 0.76 |
| RP_POS_497_25876a6_047 | | 497.2588 | 6.05 | | RP | POS | | | 2325483 | 316145 | 2305813 | 334877 | 6.59E-01 | -0.06 | 1.17 |
| RP_POS_575_41327a10_666 | | 575.4133 | 10.67 | | RP | POS | | | 1666835 | 604043 | 1631753 | 560784 | 6.59E-01 | -0.06 | 0.96 |
| RP_POS_737_50446a9_942 | | 737.5045 | 9.94 | | RP | POS | | | 260298 | 157340 | 250475 | 167196 | 6.59E-01 | -0.06 | 0.9 |
| HILIC_NEG_234_81947a4_762 | | 234.8195 | 4.76 | | HILIC | NEG | | | 461161 | 214727 | 473615 | 200200 | 6.60E-01 | 0.06 | 0.65 |
| RP_POS_146_99637a15_385 | | 146.9964 | 15.39 | | RP | POS | | | 812748 | 438947 | 836837 | 365044 | 6.60E-01 | 0.06 | 0.36 |
| RP_POS_345_13162a15_417 | | 345.1316 | 15.42 | | RP | POS | | | 109650 | 32119.8 | 111726 | 36336.1 | 6.60E-01 | 0.06 | 0.9 |
| RP_NEG_520_91113a11_193 | | 520.9111 | 11.19 | | RP | NEG | | | 707787 | 139043 | 699151 | 147523 | 6.60E-01 | -0.06 | 0.63 |
| HILIC_NEG_204_06654a0_706 | Indolelactic acid | 204.0665 | 0.71 | | HILIC | NEG | 117.9284(100), 158.0625(80) | 2 | 347023 | 91662.4 | 342015 | 75536.6 | 6.60E-01 | -0.06 | 0.65 |
| RP_NEG_458_83835a15_367 | | 458.8384 | 15.37 | | RP | NEG | | | 1636522 | 154978 | 1644767 | 121061 | 6.61E-01 | 0.06 | 0.7 |
| RP_POS_236_07162a0_616 | | 236.0716 | 0.62 | | RP | POS | | | 3.6E+07 | 3331479 | 3.6E+07 | 3141247 | 6.61E-01 | 0.06 | 0.87 |
| HILIC_POS_123_04052a7_411 | | 123.0405 | 7.41 | | HILIC | POS | 123.0999(70) | | 321658 | 97538.5 | 315640 | 102554 | 6.61E-01 | -0.06 | 0.68 |
| RP_POS_136_07565a0_68 | | 136.0757 | 0.68 | | RP | POS | 91.0548(100), 136.0763(85) | | 1.7E+07 | 4979906 | 1.7E+07 | 4448943 | 6.61E-01 | -0.06 | 0.53 |
| RP_POS_359_11511a9_458 | | 359.1151 | 9.46 | | RP | POS | | | 960116 | 977184 | 902973 | 934441 | 6.61E-01 | -0.06 | 0.85 |
| RP_POS_170_1176a10_36 | | 170.1176 | 10.36 | | RP | POS | | | 1623935 | 345443 | 1643356 | 306624 | 6.62E-01 | 0.06 | 0.77 |
| RP_POS_193_15883a9_276 | | 193.1588 | 9.28 | | RP | POS | | | 2339192 | 740332 | 2384914 | 784157 | 6.62E-01 | 0.06 | 0.88 |
| RP_POS_510_35471a10_722 | LysoPC 17:0 | 510.3547 | 10.72 | | RP | POS | 184.0743(100), 104.1077(60), 510.3586(30) | 1 | 4E+07 | 1E+07 | 3.9E+07 | 1E+07 | 6.62E-01 | -0.06 | 0.78 |
| RP_NEG_920_56458a13_253 | | 920.5646 | 13.25 | | RP | NEG | | | 2113287 | 1118970 | 2044410 | 1181114 | 6.62E-01 | -0.06 | 0.66 |
| RP_POS_345_17636a11_235 | | 345.1764 | 11.24 | | RP | POS | | | 341206 | 224059 | 329657 | 162766 | 6.62E-01 | -0.06 | 0.67 |
| RP_POS_375_16422a6_562 | | 375.1642 | 6.56 | | RP | POS | | | 474771 | 47115.7 | 472035 | 44539.2 | 6.62E-01 | -0.06 | 0.61 |
| RP_POS_930_23944a9_708 | | 930.2394 | 9.71 | | RP | POS | | | 1940317 | 190761 | 1928467 | 204391 | 6.62E-01 | -0.06 | 0.78 |
| RP_POS_237_1095a6_05 | Pestalotin | 237.1095 | 6.05 | | RP | POS | 237.1115(100) | 2 | 407799 | 251744 | 423571 | 273001 | 6.62E-01 | 0.06 | 0.71 |
| HILIC_NEG_134_89478a4_933 | | 134.8948 | 4.93 | | HILIC | NEG | | | 6E+07 | 2.2E+07 | 6.2E+07 | 2E+07 | 6.63E-01 | 0.06 | 0.53 |
| RP_NEG_612_26105a10_359 | | 612.2611 | 10.36 | | RP | NEG | | | 2101939 | 291448 | 2120338 | 322082 | 6.63E-01 | 0.06 | 0.91 |
| RP_POS_665_51105a14_839 | | 665.5111 | 14.84 | | RP | POS | | | 1569739 | 1247078 | 1635795 | 975088 | 6.63E-01 | 0.06 | 0.66 |
| HILIC_POS_149_02689a6_089 | | 149.0269 | 6.09 | | HILIC | POS | | | 298375 | 117059 | 289298 | 176335 | 6.63E-01 | -0.06 | 0.86 |
| RP_NEG_304_91357a15_354 | | 304.9136 | 15.35 | | RP | NEG | | | 1.5E+07 | 488175 | 1.5E+07 | 990135 | 6.63E-01 | -0.06 | 0.86 |
| RP_NEG_772_32562a10_939 | | 772.3256 | 10.94 | | RP | NEG | | | 1171158 | 298349 | 1152218 | 333567 | 6.63E-01 | -0.06 | 0.64 |
| HILIC_POS_356_35153a1_365 | | 356.3515 | 1.37 | | HILIC | POS | | | 1212803 | 564301 | 1245382 | 533136 | 6.64E-01 | 0.06 | 0.99 |
| RP_NEG_594_37854a11_01 | | 594.3785 | 11.01 | | RP | NEG | | | 2417491 | 677096 | 2458243 | 694118 | 6.64E-01 | 0.06 | 0.63 |
| RP_POS_265_93918a15_362 | | 265.9392 | 15.36 | | RP | POS | | | 482922 | 256903 | 498452 | 264806 | 6.64E-01 | 0.06 | 0.66 |
| RP_POS_333_29913a10_684 | | 333.2991 | 10.68 | | RP | POS | | | 1038780 | 177016 | 1049862 | 193892 | 6.64E-01 | 0.06 | 0.53 |
| RP_POS_390_98386a0_678 | | 390.9839 | 0.68 | | RP | POS | | | 1294967 | 397297 | 1317431 | 361321 | 6.64E-01 | 0.06 | 0.4 |
| RP_POS_766_5733a14_234 | | 766.5733 | 14.23 | | RP | POS | | | 784980 | 661740 | 827754 | 764426 | 6.64E-01 | 0.06 | 0.49 |
| RP_NEG_228_91624a0_683 | | 228.9162 | 0.68 | | RP | NEG | | | 387531 | 58379 | 384324 | 49892.2 | 6.64E-01 | -0.06 | 0.83 |
| RP_POS_229_18018a8_302 | | 229.1802 | 8.30 | | RP | POS | | | 536408 | 69286.4 | 532155 | 73508 | 6.64E-01 | -0.06 | 0.75 |
| RP_POS_519_23999a6_046 | | 519.2400 | 6.05 | | RP | POS | | | 2719000 | 803334 | 2670049 | 841153 | 6.64E-01 | -0.06 | 1.22 |
| RP_POS_637_49359a10_374 | | 637.4936 | 10.37 | | RP | POS | | | 2780695 | 309474 | 2763185 | 282049 | 6.64E-01 | -0.06 | 0.61 |
| RP_NEG_476_2785a10_303 | LysoPE 18:2 | 476.2785 | 10.30 | | RP | NEG | 279.2336(100), 196.0369(20) | 2 | 2.6E+07 | 9458668 | 2.7E+07 | 1.2E+07 | 6.64E-01 | 0.06 | 0.68 |
| RP_NEG_546_28174a10_591 | | 546.2817 | 10.59 | | RP | NEG | | | 1173165 | 351747 | 1195281 | 390750 | 6.65E-01 | 0.06 | 0.52 |
| RP_POS_236_07153a9_87 | | 236.0715 | 9.87 | | RP | POS | | | 7.3E+07 | 2.8E+07 | 7.4E+07 | 3E+07 | 6.65E-01 | 0.06 | 0.54 |
| RP_POS_265_25247a10_615 | | 265.2525 | 10.62 | | RP | POS | | | 1858756 | 465091 | 1887104 | 491274 | 6.65E-01 | 0.06 | 0.82 |
| RP_POS_312_88681a15_412 | | 312.8868 | 15.41 | | RP | POS | | | 1727550 | 571969 | 1763816 | 645866 | 6.65E-01 | 0.06 | 0.91 |
| RP_POS_326_34119a11_781 | | 326.3412 | 11.78 | | RP | POS | | | 456597 | 80143.4 | 461241 | 76972 | 6.65E-01 | 0.06 | 0.59 |
| RP_POS_500_37894a8_8 | | 500.3789 | 8.80 | | RP | POS | | | 832267 | 440762 | 867984 | 712193 | 6.65E-01 | 0.06 | 0.69 |
| RP_NEG_450_84335a15_368 | | 450.8434 | 15.37 | | RP | NEG | | | 372162 | 102436 | 365202 | 128613 | 6.65E-01 | -0.06 | 0.65 |
| RP_POS_229_89313a15_378 | | 229.8931 | 15.38 | | RP | POS | | | 457388 | 188549 | 446682 | 174394 | 6.65E-01 | -0.06 | 0.51 |
| RP_POS_271_11496a4_37 | | 271.1150 | 4.37 | | RP | POS | | | 442168 | 1067653 | 381182 | 994858 | 6.65E-01 | -0.06 | 0.47 |
| RP_POS_378_21149a8_077 | | 378.2115 | 8.08 | | RP | POS | | | 310340 | 95089.6 | 304191 | 110493 | 6.65E-01 | -0.06 | 0.51 |
| RP_POS_711_50616a10_379 | | 711.5062 | 10.38 | | RP | POS | | | 2949113 | 1409703 | 2871497 | 1223650 | 6.65E-01 | -0.06 | 0.77 |
| RP_POS_738_51782a13_693 | | 738.5178 | 13.69 | | RP | POS | | | 477537 | 302241 | 459428 | 308305 | 6.65E-01 | -0.06 | 0.74 |
| RP_NEG_654_13367a9_339 | | 654.1337 | 9.34 | | RP | NEG | | | 382369 | 41288.8 | 384733 | 38898.6 | 6.66E-01 | 0.06 | 0.4 |
| RP_NEG_654_30945a10_931 | | 654.3095 | 10.93 | | RP | NEG | | | 2786962 | 476719 | 2816483 | 519159 | 6.66E-01 | 0.06 | 0.8 |
| RP_POS_286_95947a15_443 | | 286.9595 | 15.44 | | RP | POS | | | 372845 | 136787 | 383525 | 211274 | 6.66E-01 | 0.06 | 0.36 |
| RP_POS_489_42703a11_6 | | 489.4270 | 11.60 | | RP | POS | | | 535795 | 123992 | 543500 | 136089 | 6.66E-01 | 0.06 | 0.68 |
| HILIC_NEG_291_00818a5_117 | | 291.0082 | 5.12 | | HILIC | NEG | | | 547439 | 237425 | 534093 | 216238 | 6.66E-01 | -0.06 | 0.3 |
| HILIC_POS_566_88879a4_338 | | 566.8888 | 4.34 | | HILIC | POS | | | 7434669 | 458260 | 7406731 | 485576 | 6.66E-01 | -0.06 | 0.74 |
| RP_NEG_1173_76331a12_545 | | 1173.7633 | 12.55 | | RP | NEG | | | 309153 | 519457 | 280737 | 447540 | 6.66E-01 | -0.06 | 0.45 |
| RP_NEG_385_16965a7_158 | | 385.1697 | 7.16 | | RP | NEG | | | 712372 | 597024 | 679865 | 509355 | 6.66E-01 | -0.06 | 0.42 |
| RP_POS_301_65182a10_492 | | 301.6518 | 10.49 | | RP | POS | | | 1493832 | 604404 | 1459786 | 550932 | 6.66E-01 | -0.06 | 0.6 |
| RP_POS_328_3212a11_107 | | 328.3212 | 11.11 | | RP | POS | | | 489238 | 101525 | 483471 | 94446.5 | 6.66E-01 | -0.06 | 0.63 |
| RP_POS_394_34659a12_328 | | 394.3466 | 12.33 | | RP | POS | | | 8367052 | 2260334 | 8242652 | 1971708 | 6.66E-01 | -0.06 | 1.19 |
| RP_POS_453_16119a12_375 | | 453.1612 | 12.38 | | RP | POS | | | 425940 | 198707 | 414435 | 191644 | 6.66E-01 | -0.06 | 0.97 |
| HILIC_POS_179_07913a5_118 | | 179.0791 | 5.12 | | HILIC | POS | | | 2520348 | 6393406 | 3180725 | 1.4E+07 | 6.67E-01 | 0.06 | 0.81 |
| HILIC_POS_252_04576a2_98 | | 252.0458 | 2.98 | | HILIC | POS | | | 779076 | 145290 | 786854 | 120033 | 6.67E-01 | 0.06 | 0.66 |
| HILIC_POS_362_01968a5_148 | | 362.0197 | 5.15 | | HILIC | POS | | | 110041 | 24248.6 | 111438 | 23356.4 | 6.67E-01 | 0.06 | 0.52 |
| RP_NEG_180_86206a15_37 | | 180.8621 | 15.37 | | RP | NEG | | | 3233502 | 415708 | 3259833 | 473675 | 6.67E-01 | 0.06 | 0.66 |
| RP_POS_422_3269a9_021 | | 422.3269 | 9.02 | | RP | POS | | | 826102 | 346968 | 845673 | 319466 | 6.67E-01 | 0.06 | 0.59 |
| RP_POS_650_62225a11_478 | | 650.6223 | 11.48 | | RP | POS | | | 216465 | 103796 | 223382 | 128034 | 6.67E-01 | 0.06 | 0.59 |
| RP_POS_994_25116a9_898 | | 994.2512 | 9.90 | | RP | POS | | | 1507984 | 155092 | 1517828 | 176736 | 6.67E-01 | 0.06 | 0.77 |
| HILIC_NEG_226_86201a0_849 | | 226.8620 | 0.85 | | HILIC | NEG | | | 157903 | 17740.6 | 156874 | 17315.3 | 6.67E-01 | -0.06 | 0.55 |
| RP_NEG_428_0517a0_679 | | 428.0517 | 0.68 | | RP | NEG | | | 267920 | 77215.7 | 263824 | 62501.4 | 6.67E-01 | -0.06 | 0.35 |
| RP_POS_338_21744a6_378 | | 338.2174 | 6.38 | | RP | POS | | | 1532498 | 96190.5 | 1526966 | 92065.2 | 6.67E-01 | -0.06 | 0.83 |
| HILIC_POS_176_06564a4_113 | Gentiannine | 176.0656 | 4.11 | | HILIC | POS | 176.1036(100) | 2 | 1151820 | 702470 | 1199040 | 879443 | 6.67E-01 | 0.06 | 0.64 |
| HILIC_NEG_136_89172a2_272 | | 136.8917 | 2.27 | | HILIC | NEG | | | 1.3E+07 | 1936625 | 1.4E+07 | 2371156 | 6.68E-01 | 0.06 | 0.6 |
| HILIC_NEG_343_88663a3_257 | | 343.8866 | 3.26 | | HILIC | NEG | | | 1349799 | 262652 | 1366242 | 294128 | 6.68E-01 | 0.06 | 0.59 |
| HILIC_NEG_396_89148a4_435 | | 396.8915 | 4.44 | | HILIC | NEG | | | 177847 | 92891.6 | 183603 | 102313 | 6.68E-01 | 0.06 | 1 |
| RP_POS_588_08032a10_405 | | 588.0803 | 10.41 | | RP | POS | | | 717482 | 331640 | 740033 | 423609 | 6.68E-01 | 0.06 | 0.49 |
| RP_POS_733_51758a10_362 | | 733.5176 | 10.36 | | RP | POS | | | 2249835 | 1049308 | 2314533 | 1149327 | 6.68E-01 | 0.06 | 0.39 |
| RP_NEG_201_03838a0_685 | | 201.0384 | 0.69 | | RP | NEG | | | 1758115 | 395073 | 1736795 | 334681 | 6.68E-01 | -0.06 | 0.65 |
| RP_NEG_372_70264a15_456 | | 372.7026 | 15.46 | | RP | NEG | | | 1641276 | 152761 | 1632129 | 158258 | 6.68E-01 | -0.06 | 0.68 |
| RP_POS_466_24539a6_42 | | 466.2454 | 6.42 | | RP | POS | | | 458638 | 172492 | 449223 | 149523 | 6.68E-01 | -0.06 | 0.68 |
| RP_POS_544_45782a13_074 | | 544.4578 | 13.07 | | RP | POS | | | 8376501 | 1.9E+07 | 7385271 | 1.5E+07 | 6.68E-01 | -0.06 | 0.5 |
| RP_POS_746_60321a14_525 | | 746.6032 | 14.53 | | RP | POS | | | 3661350 | 2473331 | 3527283 | 2118563 | 6.68E-01 | -0.06 | 0.52 |
| RP_POS_478_29294a10_182 | | 478.2929 | 10.18 | | RP | POS | 337.2749(100) | | 9712184 | 3992375 | 9977761 | 4971426 | 6.69E-01 | 0.06 | 0.68 |
| RP_POS_161_96983a0_623 | | 161.9698 | 0.62 | | RP | POS | | | 611927 | 223097 | 625022 | 224679 | 6.69E-01 | 0.06 | 0.59 |
| RP_POS_196_16948a0_646 | | 196.1695 | 0.65 | | RP | POS | | | 3905415 | 1754795 | 4012118 | 1881892 | 6.69E-01 | 0.06 | 0.54 |
| RP_POS_463_31915a12_439 | | 463.3192 | 12.44 | | RP | POS | | | 148682 | 106624 | 154943 | 107545 | 6.69E-01 | 0.06 | 1.06 |
| RP_POS_476_32108a11_028 | | 476.3211 | 11.03 | | RP | POS | | | 348468 | 45564.7 | 351292 | 50614.2 | 6.69E-01 | 0.06 | 0.63 |
| RP_NEG_340_05112a0_674 | | 340.0511 | 0.67 | | RP | NEG | | | 513041 | 127855 | 506126 | 109502 | 6.69E-01 | -0.06 | 0.51 |
| RP_POS_329_1568a5_235 | | 329.1568 | 5.24 | | RP | POS | | | 271895 | 137095 | 264773 | 107600 | 6.69E-01 | -0.06 | 1 |
| HILIC_NEG_423_00732a1_274 | | 423.0073 | 1.27 | | HILIC | NEG | | | 426173 | 178873 | 436749 | 183821 | 6.70E-01 | 0.06 | 0.55 |
| HILIC_POS_226_02431a3_963 | | 226.0243 | 3.96 | | HILIC | POS | | | 218796 | 92466.7 | 224270 | 94944.9 | 6.70E-01 | 0.06 | 0.66 |
| RP_NEG_341_98279a2_885 | | 341.9828 | 2.89 | | RP | NEG | | | 464071 | 79155.5 | 468669 | 78728.8 | 6.70E-01 | 0.06 | 0.52 |
| HILIC_NEG_136_89169a4_271 | | 136.8917 | 4.27 | | HILIC | NEG | | | 5147544 | 1868304 | 5036473 | 1931863 | 6.70E-01 | -0.06 | 0.58 |
| HILIC_POS_148_10791a4_188 | | 148.1079 | 4.19 | | HILIC | POS | | | 312053 | 182499 | 302584 | 142901 | 6.70E-01 | -0.06 | 0.6 |
| RP_NEG_285_12622a8_476 | | 285.1262 | 8.48 | | RP | NEG | | | 4124691 | 140012 | 4116829 | 130580 | 6.70E-01 | -0.06 | 0.35 |
| RP_POS_351_22928a10_896 | | 351.2293 | 10.90 | | RP | POS | | | 2.2E+07 | 1.1E+07 | 2.2E+07 | 1.1E+07 | 6.70E-01 | -0.06 | 0.9 |
| RP_POS_810_59998a14_357 | PC 38:4 (18:0_20:4) | 810.6000 | 14.36 | | RP | POS | 184.0745(100), 86.0961(20) | 1 | 6524252 | 3535027 | 6735232 | 3681525 | 6.70E-01 | 0.06 | 0.61 |
| RP_POS_208_13356a0_699 | Benzoylcholine | 208.1336 | 0.70 | | RP | POS | 149.0593(100), 208.1341(65) | 2 | 1139343 | 64010.6 | 1135779 | 58566.3 | 6.70E-01 | -0.06 | 0.47 |
| RP_POS_225_09039a9_575 | | 225.0904 | 9.58 | | RP | POS | peak 100 missing | | 688063 | 231088 | 701840 | 242584 | 6.71E-01 | 0.06 | 0.7 |
| RP_NEG_171_94682a0_702 | | 171.9468 | 0.70 | | RP | NEG | | | 1873414 | 320353 | 1891311 | 297809 | 6.71E-01 | 0.06 | 0.69 |
| RP_POS_134_99644a15_378 | | 134.9964 | 15.38 | | RP | POS | | | 7804838 | 2722013 | 7969377 | 2917467 | 6.71E-01 | 0.06 | 0.41 |
| RP_NEG_449_15149a8_617 | | 449.1515 | 8.62 | | RP | NEG | | | 173035 | 144796 | 164583 | 146405 | 6.71E-01 | -0.06 | 0.49 |
| RP_NEG_650_27716a10_18 | | 650.2772 | 10.18 | | RP | NEG | | | 493819 | 178834 | 483426 | 178693 | 6.71E-01 | -0.06 | 0.68 |
| RP_POS_318_11957a0_679 | | 318.1196 | 0.68 | | RP | POS | | | 2407281 | 1153643 | 2470191 | 1023880 | 6.72E-01 | 0.06 | 0.5 |
| RP_POS_416_77982a13_778 | | 416.7798 | 13.78 | | RP | POS | | | 601772 | 538172 | 632321 | 519278 | 6.72E-01 | 0.06 | 0.49 |
| HILIC_NEG_355_01974a0_933 | | 355.0197 | 0.93 | | HILIC | NEG | | | 337674 | 38442.6 | 335377 | 40706.3 | 6.72E-01 | -0.06 | 0.54 |
| RP_NEG_280_9826a8_171 | | 280.9826 | 8.17 | | RP | NEG | | | 2341479 | 278640 | 2325792 | 264172 | 6.72E-01 | -0.06 | 0.6 |
| RP_NEG_283_11044a8_53 | | 283.1104 | 8.53 | | RP | NEG | | | 343496 | 151918 | 334592 | 155381 | 6.72E-01 | -0.06 | 0.44 |
| RP_NEG_446_10318a11_594 | | 446.1032 | 11.59 | | RP | NEG | | | 144228 | 34469 | 142256 | 33708.4 | 6.72E-01 | -0.06 | 0.45 |
| RP_NEG_979_14001a9_668 | | 979.1400 | 9.67 | | RP | NEG | | | 4798914 | 305714 | 4782289 | 270858 | 6.72E-01 | -0.06 | 1.07 |
| RP_POS_234_13393a4_703 | | 234.1339 | 4.70 | | RP | POS | | | 367159 | 141021 | 358878 | 144530 | 6.72E-01 | -0.06 | 0.44 |
| RP_POS_288_63358a10_311 | | 288.6336 | 10.31 | | RP | POS | | | 5924605 | 1557807 | 5839114 | 1403164 | 6.72E-01 | -0.06 | 0.78 |
| RP_POS_582_41669a10_662 | | 582.4167 | 10.66 | | RP | POS | | | 263337 | 183299 | 252613 | 187204 | 6.72E-01 | -0.06 | 0.61 |
| RP_POS_454_29303a10_475 | LysoPE 16:0 | 454.2930 | 10.48 | | RP | POS | 313.2726(100),57.071(10) | 1 | 3E+07 | 6637793 | 2.9E+07 | 7230058 | 6.72E-01 | -0.06 | 0.58 |
| HILIC_POS_106_08675a1_361 | | 106.0868 | 1.36 | | HILIC | POS | 88.0763(100), 106.0564(75) | | 1.3E+07 | 6574355 | 1.3E+07 | 6820261 | 6.73E-01 | 0.06 | 0.83 |
| RP_NEG_722_29559a10_934 | | 722.2956 | 10.93 | | RP | NEG | | | 605321 | 95249.3 | 611177 | 106159 | 6.73E-01 | 0.06 | 0.73 |
| RP_POS_265_00055a0_586 | | 265.0006 | 0.59 | | RP | POS | | | 5377500 | 1096234 | 5443470 | 1180300 | 6.73E-01 | 0.06 | 0.45 |
| RP_POS_860_50909a13_785 | | 860.5091 | 13.79 | | RP | POS | | | 399279 | 347505 | 417968 | 303310 | 6.73E-01 | 0.06 | 0.74 |
| RP_NEG_180_98935a15_476 | | 180.9894 | 15.48 | | RP | NEG | | | 2990027 | 237816 | 2976036 | 246510 | 6.73E-01 | -0.06 | 0.48 |
| RP_NEG_413_65076a15_496 | | 413.6508 | 15.50 | | RP | NEG | | | 945990 | 69315.3 | 950492 | 85300.1 | 6.74E-01 | 0.06 | 0.63 |
| RP_NEG_300_93625a0_68 | | 300.9363 | 0.68 | | RP | NEG | | | 405916 | 87896.2 | 401367 | 70699.7 | 6.74E-01 | -0.06 | 0.72 |
| RP_POS_137_04582a0_679 | | 137.0458 | 0.68 | | RP | POS | 137.0467(100), 110.0356(3) | | 5.1E+07 | 2.6E+07 | 5.2E+07 | 2.3E+07 | 6.75E-01 | 0.06 | 0.58 |
| HILIC_POS_156_13847a0_787 | | 156.1385 | 0.79 | | HILIC | POS | | | 672117 | 174738 | 681639 | 158412 | 6.75E-01 | 0.06 | 0.49 |
| RP_NEG_230_01305a1_393 | | 230.0131 | 1.39 | | RP | NEG | | | 3112421 | 1.4E+07 | 3978896 | 1.6E+07 | 6.75E-01 | 0.06 | 0.69 |
| RP_POS_147_04419a9_175 | | 147.0442 | 9.18 | | RP | POS | | | 577029 | 45986.8 | 579603 | 44044.4 | 6.75E-01 | 0.06 | 0.58 |
| RP_POS_261_22147a10_443 | | 261.2215 | 10.44 | | RP | POS | | | 3482893 | 1637418 | 3578265 | 1680502 | 6.75E-01 | 0.06 | 0.3 |
| RP_POS_271_19016a7_148 | | 271.1902 | 7.15 | | RP | POS | | | 202068 | 109828 | 208563 | 116047 | 6.75E-01 | 0.06 | 0.69 |
| RP_POS_573_07318a10_42 | | 573.0732 | 10.42 | | RP | POS | | | 846448 | 425988 | 870693 | 420080 | 6.75E-01 | 0.06 | 0.73 |
| RP_POS_663_06885a11_475 | | 663.0689 | 11.48 | | RP | POS | | | 145707 | 72421.8 | 149979 | 76062.8 | 6.75E-01 | 0.06 | 0.62 |
| RP_POS_744_45618a11_593 | | 744.4562 | 11.59 | | RP | POS | | | 1659620 | 1924690 | 1792063 | 2589950 | 6.75E-01 | 0.06 | 0.8 |
| HILIC_NEG_144_92355a4_638 | | 144.9236 | 4.64 | | HILIC | NEG | | | 4.2E+07 | 2.2E+07 | 4.1E+07 | 2.2E+07 | 6.75E-01 | -0.06 | 0.54 |
| HILIC_NEG_247_89268a0_898 | | 247.8927 | 0.90 | | HILIC | NEG | | | 1414106 | 324517 | 1394253 | 364453 | 6.75E-01 | -0.06 | 0.52 |
| RP_NEG_695_50598a15_265 | | 695.5060 | 15.27 | | RP | NEG | | | 1523540 | 375722 | 1502609 | 355067 | 6.75E-01 | -0.06 | 0.51 |
| RP_POS_151_14821a7_695 | | 151.1482 | 7.70 | | RP | POS | | | 1457645 | 104278 | 1451594 | 106553 | 6.75E-01 | -0.06 | 0.48 |
| RP_POS_177_08705a0_544 | | 177.0871 | 0.54 | | RP | POS | | | 3148311 | 914286 | 3095797 | 919749 | 6.75E-01 | -0.06 | 0.76 |
| RP_POS_423_76944a13_24 | | 423.7694 | 13.24 | | RP | POS | | | 1610638 | 922048 | 1560808 | 819027 | 6.75E-01 | -0.06 | 0.67 |
| RP_POS_628_93774a11_288 | | 628.9377 | 11.29 | | RP | POS | | | 266474 | 152623 | 258773 | 116893 | 6.75E-01 | -0.06 | 0.69 |
| HILIC_NEG_115_02097a1_345 | | 115.0210 | 1.35 | | HILIC | NEG | | | 1403196 | 336117 | 1423360 | 367088 | 6.76E-01 | 0.06 | 0.7 |
| RP_NEG_335_04944a0_678 | | 335.0494 | 0.68 | | RP | NEG | | | 3.8E+07 | 9352932 | 3.8E+07 | 9733442 | 6.76E-01 | 0.06 | 0.27 |
| RP_POS_563_46454a14_247 | | 563.4645 | 14.25 | | RP | POS | | | 900325 | 511590 | 931519 | 572926 | 6.76E-01 | 0.06 | 0.32 |
| HILIC_POS_161_12814a1_363 | | 161.1281 | 1.36 | | HILIC | POS | | | 801011 | 375403 | 779257 | 385904 | 6.76E-01 | -0.06 | 0.39 |
| HILIC_POS_582_5929a10_65 | | 582.5929 | 10.65 | | HILIC | POS | | | 493002 | 84050.1 | 488495 | 74187.7 | 6.76E-01 | -0.06 | 0.53 |
| RP_NEG_376_98578a7_366 | | 376.9858 | 7.37 | | RP | NEG | | | 2073021 | 62025.4 | 2069093 | 73848.5 | 6.76E-01 | -0.06 | 0.46 |
| RP_POS_192_10201a4_377 | | 192.1020 | 4.38 | | RP | POS | | | 5123302 | 6028772 | 4776988 | 6090112 | 6.76E-01 | -0.06 | 0.49 |
| RP_POS_301_14078a13_295 | | 301.1408 | 13.30 | | RP | POS | | | 1212165 | 865036 | 1163188 | 851904 | 6.76E-01 | -0.06 | 0.98 |
| RP_POS_524_37122a11_141 | | 524.3712 | 11.14 | | RP | POS | | | 1.2E+07 | 3229691 | 1.2E+07 | 3430998 | 6.76E-01 | -0.06 | 0.6 |
| RP_POS_672_25317a10_536 | | 672.2532 | 10.54 | | RP | POS | | | 448270 | 181863 | 438953 | 145457 | 6.76E-01 | -0.06 | 0.47 |
| HILIC_NEG_230_01279a0_628 | | 230.0128 | 0.63 | | HILIC | NEG | | | 453139 | 1848219 | 565847 | 2080773 | 6.77E-01 | 0.06 | 0.69 |
| RP_NEG_331_95352a2_886 | | 331.9535 | 2.89 | | RP | NEG | | | 198870 | 52364.8 | 201846 | 52006.6 | 6.77E-01 | 0.06 | 0.55 |
| RP_POS_250_12827a4_825 | | 250.1283 | 4.83 | | RP | POS | | | 708097 | 276360 | 725399 | 324583 | 6.77E-01 | 0.06 | 0.83 |
| RP_POS_274_18723a0_576 | | 274.1872 | 0.58 | | RP | POS | | | 1947674 | 759823 | 1994969 | 886566 | 6.77E-01 | 0.06 | 0.78 |

**Supplemantary table 1**: The non-targeted metabolomics results ordered by the VIP value (p value < 0.0003 are shaded dark green, p value < 0.05 are shaded light green, d values below 0 are blue and above 0 are red)

| | | | | | | | | | Cases | | Controls | | Welch test | Cohen's | |
|---|---|---|---|---|---|---|---|---|---|---|---|---|---|---|---|
| Feature_ID | Metabolite_name | Average_Mz | Average_RT_min_ | HMDB ID | RP_HILIC | POS_NEG | Cureted_MSMS | ID_level | Mean | SD | Mean2 | SD3 | p | d | VIP |
| RP_POS_997_62469a10_306 | | 997.6247 | 10.31 | | RP | POS | | | 5138124 | 3234283 | 5348875 | 4049904 | 6.77E-01 | 0.06 | 0.69 |
| HILIC_NEG_144_92352a3_106 | | 144.9235 | 3.11 | | HILIC | NEG | | | 1.4E+07 | 1779455 | 1.4E+07 | 1748918 | 6.77E-01 | -0.06 | 0.37 |
| HILIC_NEG_234_81963a3_349 | | 234.8196 | 3.35 | | HILIC | NEG | | | 3890998 | 1859622 | 3779675 | 2030715 | 6.77E-01 | -0.06 | 0.65 |
| HILIC_NEG_524_27942a1_723 | | 524.2794 | 1.72 | | HILIC | NEG | | | 292131 | 90493.4 | 286748 | 97914.9 | 6.77E-01 | -0.06 | 0.48 |
| RP_POS_229_14117a6_118 | | 229.1412 | 6.12 | | RP | POS | | | 396177 | 160900 | 387147 | 156249 | 6.77E-01 | -0.06 | 0.64 |
| RP_POS_551_42413a11_657 | | 551.4241 | 11.66 | | RP | POS | | | 2084115 | 4006171 | 1865767 | 3679632 | 6.77E-01 | -0.06 | 0.57 |
| RP_POS_277_09451a5_759 | Dihydromethysticin | 277.0945 | 5.76 | | RP | POS | 277.0957(100) | 2 | 4409369 | 9355638 | 4008932 | 4144025 | 6.77E-01 | -0.06 | 0.64 |
| RP_POS_347_22079a7_05 | | 347.2208 | 7.05 | | RP | POS | | | 533134 | 254533 | 546337 | 211703 | 6.78E-01 | 0.06 | 0.45 |
| RP_POS_798_59943a13_75 | | 798.5994 | 13.75 | | RP | POS | | | 5008859 | 2881782 | 5182245 | 3195742 | 6.78E-01 | 0.06 | 1 |
| RP_POS_890_54083a13_944 | | 890.5408 | 13.94 | | RP | POS | | | 1568067 | 1056438 | 1630292 | 1130434 | 6.78E-01 | 0.06 | 0.63 |
| HILIC_NEG_129_05585a0_838 | | 129.0559 | 0.84 | | HILIC | NEG | 129.0557(100) | | 4.5E+07 | 1.3E+07 | 4.4E+07 | 1.2E+07 | 6.78E-01 | -0.06 | 0.74 |
| HILIC_NEG_152_88649a3_203 | | 152.8865 | 3.20 | | HILIC | NEG | | | 329525 | 128033 | 321989 | 136849 | 6.78E-01 | -0.06 | 0.61 |
| RP_NEG_338_8681a15_395 | | 338.8681 | 15.40 | | RP | NEG | | | 2500117 | 494864 | 2471795 | 503339 | 6.78E-01 | -0.06 | 0.97 |
| RP_POS_155_10649a10_252 | | 155.1065 | 10.25 | | RP | POS | | | 1736289 | 162425 | 1726557 | 179116 | 6.78E-01 | -0.06 | 0.66 |
| RP_POS_352_24521a10_253 | | 352.2452 | 10.25 | | RP | POS | | | 1.4E+07 | 1304431 | 1.4E+07 | 1408344 | 6.78E-01 | -0.06 | 0.73 |
| RP_POS_423_75311a13_46 | | 423.7531 | 13.46 | | RP | POS | | | 435894 | 344246 | 415479 | 372292 | 6.78E-01 | -0.06 | 0.71 |
| HILIC_NEG_312_83636a5_379 | | 312.8364 | 5.38 | | HILIC | NEG | | | 1542050 | 394713 | 1564507 | 399607 | 6.79E-01 | 0.06 | 0.62 |
| RP_POS_173_09631a7_322 | | 173.0963 | 7.32 | | RP | POS | | | 927195 | 303870 | 946238 | 362123 | 6.79E-01 | 0.06 | 0.45 |
| RP_POS_251_95061a15_548 | | 251.9506 | 15.55 | | RP | POS | | | 455164 | 199385 | 466646 | 205821 | 6.79E-01 | 0.06 | 0.41 |
| RP_POS_524_16248a8_598 | | 524.1625 | 8.60 | | RP | POS | | | 599260 | 58160.4 | 602580 | 59045.3 | 6.79E-01 | 0.06 | 0.82 |
| RP_POS_576_37378a10_275 | | 576.3738 | 10.28 | | RP | POS | | | 402523 | 174048 | 413253 | 202038 | 6.79E-01 | 0.06 | 0.89 |
| HILIC_POS_147_14934a4_809 | | 147.1493 | 4.81 | | HILIC | POS | | | 2.5E+07 | 7480594 | 2.5E+07 | 9801304 | 6.79E-01 | -0.06 | 0.34 |
| RP_NEG_226_96605a14_345 | | 226.9661 | 14.35 | | RP | NEG | | | 1298732 | 563501 | 1266858 | 563012 | 6.79E-01 | -0.06 | 0.7 |
| RP_NEG_966_56787a14_098 | | 966.5679 | 14.10 | | RP | NEG | | | 4013861 | 1541422 | 3926334 | 1554064 | 6.79E-01 | -0.06 | 0.67 |
| RP_POS_155_10683a6_268 | | 155.1068 | 6.27 | | RP | POS | | | 1358389 | 502606 | 1330367 | 488697 | 6.79E-01 | -0.06 | 0.58 |
| RP_POS_504_34332a10_778 | | 504.3433 | 10.78 | | RP | POS | | | 3806920 | 1124768 | 3741291 | 1187469 | 6.79E-01 | -0.06 | 0.49 |
| HILIC_POS_518_32153a1_124 | | 518.3215 | 1.12 | | HILIC | POS | | | 1402799 | 465402 | 1429416 | 476144 | 6.80E-01 | 0.06 | 0.66 |
| RP_POS_295_18866a7_724 | | 295.1887 | 7.72 | | RP | POS | | | 240521 | 98577.4 | 247262 | 134936 | 6.80E-01 | 0.06 | 0.7 |
| RP_POS_808_58453a13_257 | | 808.5845 | 13.26 | | RP | POS | | | 2.2E+08 | 5E+07 | 2.3E+08 | 5.3E+07 | 6.80E-01 | 0.06 | 0.67 |
| HILIC_POS_217_15428a5_471 | | 217.1543 | 5.47 | | HILIC | POS | | | 434056 | 771371 | 392888 | 693105 | 6.80E-01 | -0.06 | 0.51 |
| RP_POS_245_0746a9_211 | | 245.0746 | 9.21 | | RP | POS | | | 268534 | 210552 | 256783 | 205950 | 6.80E-01 | -0.06 | 0.83 |
| RP_POS_476_27722a10_017 | | 476.2772 | 10.02 | | RP | POS | | | 4130292 | 2596626 | 3988634 | 2433615 | 6.80E-01 | -0.06 | 0.52 |
| RP_POS_468_30853a9_929 | LysoPC 14:0 (14:0/0:0) | 468.3085 | 9.93 | | RP | POS | 184.0742(100), 104.1077(70) | 1 | 8.4E+07 | 3.2E+07 | 8.6E+07 | 3.5E+07 | 6.80E-01 | 0.06 | 0.4 |
| HILIC_NEG_134_89476a5_322 | | 134.8948 | 5.32 | | HILIC | NEG | | | 2.5E+07 | 1.1E+07 | 2.6E+07 | 1.2E+07 | 6.81E-01 | 0.06 | 0.43 |
| RP_NEG_338_78192a15_374 | | 338.7819 | 15.37 | | RP | NEG | | | 4182494 | 624201 | 4221466 | 744694 | 6.81E-01 | 0.06 | 0.28 |
| HILIC_NEG_294_98303a1_313 | | 294.9830 | 1.31 | | HILIC | NEG | | | 523163 | 255433 | 509524 | 231670 | 6.81E-01 | -0.06 | 0.38 |
| HILIC_POS_188_03531a6_115 | | 188.0353 | 6.12 | | HILIC | POS | | | 382162 | 304446 | 363579 | 351604 | 6.81E-01 | -0.06 | 0.83 |
| RP_NEG_614_3468a10_437 | | 614.3468 | 10.44 | | RP | NEG | | | 3155439 | 1018555 | 3094486 | 1139262 | 6.81E-01 | -0.06 | 0.57 |
| RP_POS_279_95496a15_527 | | 279.9550 | 15.53 | | RP | POS | | | 699685 | 158755 | 690640 | 162378 | 6.81E-01 | -0.06 | 0.75 |
| RP_POS_250_13174a8_842 | | 250.1317 | 8.84 | | RP | POS | | | 187222 | 257157 | 201093 | 238602 | 6.82E-01 | 0.06 | 0.42 |
| RP_POS_464_43024a11_951 | | 464.4302 | 11.95 | | RP | POS | | | 133024 | 63636.3 | 136969 | 75698.1 | 6.82E-01 | 0.06 | 0.47 |
| HILIC_NEG_520_90887a4_352 | | 520.9089 | 4.35 | | HILIC | NEG | | | 1.2E+07 | 835851 | 1.2E+07 | 840901 | 6.82E-01 | -0.06 | 0.82 |
| HILIC_POS_171_14925a3_069 | | 171.1493 | 3.07 | | HILIC | POS | | | 4220060 | 2038995 | 4105396 | 2055105 | 6.82E-01 | -0.06 | 0.59 |
| RP_POS_283_1153a6_565 | | 283.1153 | 6.57 | | RP | POS | | | 3345022 | 4271497 | 3112222 | 4064173 | 6.82E-01 | -0.06 | 0.47 |
| RP_POS_345_25711a10_635 | | 345.2571 | 10.64 | | RP | POS | | | 1E+07 | 2735599 | 1E+07 | 3077880 | 6.82E-01 | -0.06 | 0.38 |
| RP_POS_386_25421a8_06 | | 386.2542 | 8.06 | | RP | POS | | | 203510 | 67307.1 | 199795 | 65476.6 | 6.82E-01 | -0.06 | 0.44 |
| RP_POS_465_81931a9_841 | | 465.8193 | 9.84 | | RP | POS | | | 2235062 | 1184310 | 2168278 | 1201638 | 6.82E-01 | -0.06 | 0.69 |
| RP_NEG_598_51587a14_038 | | 598.5159 | 14.04 | | RP | NEG | | | 598225 | 119507 | 604448 | 104249 | 6.83E-01 | 0.06 | 0.42 |
| RP_POS_464_38647a11_138 | | 464.3865 | 11.14 | | RP | POS | | | 885906 | 1677774 | 971613 | 1405666 | 6.83E-01 | 0.06 | 0.88 |
| RP_POS_593_74915a10_138 | | 593.7492 | 10.14 | | RP | POS | | | 209202 | 142478 | 217368 | 149348 | 6.83E-01 | 0.06 | 0.3 |
| RP_POS_721_50256a11_49 | | 721.5026 | 11.49 | | RP | POS | | | 324427 | 219142 | 337275 | 238590 | 6.83E-01 | 0.06 | 0.36 |
| RP_POS_748_54242a9_396 | | 748.5424 | 9.40 | | RP | POS | | | 1933946 | 977391 | 1993912 | 1147850 | 6.83E-01 | 0.06 | 0.42 |
| RP_POS_335_12509a9_472 | | 335.1251 | 9.47 | | RP | POS | | | 1041997 | 162441 | 1032325 | 181929 | 6.83E-01 | -0.06 | 0.51 |
| RP_POS_344_10202a9_179 | | 344.1020 | 9.18 | | RP | POS | | | 425537 | 304018 | 407463 | 339292 | 6.83E-01 | -0.06 | 0.6 |
| RP_POS_570_4574a10_652 | | 570.4574 | 10.65 | | RP | POS | | | 9784146 | 779920 | 9741086 | 762660 | 6.83E-01 | -0.06 | 1.09 |
| RP_POS_677_47504a11_584 | | 677.4750 | 11.58 | | RP | POS | | | 210734 | 119803 | 203806 | 127411 | 6.83E-01 | -0.06 | 0.41 |
| RP_POS_804_55066a11_594 | | 804.5507 | 11.59 | | RP | POS | | | 1526125 | 320384 | 1508355 | 316885 | 6.83E-01 | -0.06 | 0.56 |
| HILIC_POS_189_07314a2_259 | | 189.0731 | 2.26 | | HILIC | POS | | | 1725903 | 425918 | 1751164 | 475923 | 6.84E-01 | 0.06 | 0.39 |
| HILIC_POS_542_32281a1_204 | | 542.3228 | 1.20 | | HILIC | POS | | | 3749674 | 1687173 | 3665269 | 1359620 | 6.84E-01 | -0.06 | 0.55 |
| RP_NEG_524_33618a10_693 | | 524.3362 | 10.69 | | RP | NEG | | | 6916324 | 1802111 | 6820159 | 1659019 | 6.84E-01 | -0.06 | 0.64 |
| RP_POS_255_23172a9_982 | | 255.2317 | 9.98 | | RP | POS | | | 1414018 | 187256 | 1404504 | 155782 | 6.84E-01 | -0.06 | 0.62 |
| RP_POS_387_27853a11_591 | | 387.2785 | 11.59 | | RP | POS | | | 479721 | 79671.3 | 475681 | 65990.2 | 6.84E-01 | -0.06 | 0.72 |
| RP_POS_410_80386a9_88 | | 410.8039 | 9.88 | | RP | POS | | | 982273 | 513189 | 950397 | 619026 | 6.84E-01 | -0.06 | 0.61 |
| RP_POS_511_68701a10_357 | | 511.6870 | 10.36 | | RP | POS | | | 443277 | 234554 | 427955 | 304974 | 6.84E-01 | -0.06 | 0.36 |
| RP_POS_522_28619a9_132 | | 522.2862 | 9.13 | | RP | POS | 148.0049(100) 486.2679(60) | | 1011706 | 3634422 | 1212476 | 3603113 | 6.85E-01 | 0.06 | 0.51 |
| HILIC_NEG_128_9285a4_261 | | 128.9285 | 4.26 | | HILIC | NEG | | | 785195 | 275994 | 800485 | 274655 | 6.85E-01 | 0.06 | 0.45 |
| RP_NEG_180_06679a1_225 | | 180.0668 | 1.23 | | RP | NEG | 180.0662(100), 163.0399(80), 119.0501(48) | | 1998249 | 692341 | 1962712 | 592568 | 6.85E-01 | -0.06 | 0.49 |
| HILIC_NEG_157_90103a3_586 | | 157.9010 | 3.59 | | HILIC | NEG | | | 1994525 | 1002488 | 1939608 | 981671 | 6.85E-01 | -0.06 | 0.43 |
| RP_NEG_173_92593a15_559 | | 173.9259 | 15.56 | | RP | NEG | | | 1048177 | 348472 | 1029055 | 341438 | 6.85E-01 | -0.06 | 0.33 |
| RP_NEG_214_78333a11_588 | | 214.7833 | 11.59 | | RP | NEG | | | 159363 | 44537.3 | 156789 | 47853.3 | 6.85E-01 | -0.06 | 0.49 |
| RP_POS_203_15038a0_57 | | 203.1504 | 0.57 | | RP | POS | | | 5822357 | 842804 | 5774112 | 890696 | 6.85E-01 | -0.06 | 0.46 |
| RP_POS_237_1843a7_68 | | 237.1843 | 7.68 | | RP | POS | | | 653862 | 211237 | 642795 | 188880 | 6.85E-01 | -0.06 | 0.7 |
| RP_POS_417_81107a10_111 | | 417.8111 | 10.11 | | RP | POS | | | 533068 | 276722 | 549910 | 326982 | 6.86E-01 | 0.06 | 0.67 |
| RP_POS_727_57581a12_498 | | 727.5758 | 12.50 | | RP | POS | | | 4249371 | 1846205 | 4356720 | 2015245 | 6.86E-01 | 0.06 | 0.52 |
| RP_POS_182_08138a1_225 | | 182.0814 | 1.23 | | RP | POS | 136.0758(100), 159.0139(55) | | 1.2E+07 | 4380046 | 1.2E+07 | 3650575 | 6.86E-01 | -0.06 | 0.51 |
| HILIC_NEG_153_86836a3_098 | | 153.8684 | 3.10 | | HILIC | NEG | | | 1.8E+07 | 1048527 | 1.8E+07 | 974706 | 6.86E-01 | -0.06 | 0.47 |
| RP_POS_315_17395a10_154 | | 315.1740 | 10.15 | | RP | POS | | | 1.3E+07 | 4146259 | 1.3E+07 | 3915015 | 6.86E-01 | -0.06 | 0.54 |
| RP_POS_320_16943a5_434 | | 320.1694 | 5.43 | | RP | POS | | | 443725 | 59780.2 | 440455 | 58425.9 | 6.86E-01 | -0.06 | 0.55 |
| RP_POS_366_2316a10_833 | | 366.2316 | 10.83 | | RP | POS | | | 345608 | 249927 | 332764 | 216980 | 6.86E-01 | -0.06 | 0.99 |
| HILIC_POS_125_00003a1_25 | | 125.0000 | 1.25 | | HILIC | POS | | | 333891 | 262996 | 349391 | 296001 | 6.87E-01 | 0.06 | 0.31 |
| RP_POS_181_12256a6_039 | | 181.1226 | 6.04 | | RP | POS | | | 891600 | 236181 | 905104 | 252980 | 6.87E-01 | 0.06 | 0.79 |
| RP_POS_345_13025a4_336 | | 345.1303 | 4.34 | | RP | POS | | | 692289 | 503704 | 720155 | 507691 | 6.87E-01 | 0.06 | 0.56 |
| RP_POS_612_55615a11_408 | | 612.5562 | 11.41 | | RP | POS | | | 284300 | 469315 | 315114 | 624069 | 6.87E-01 | 0.06 | 0.44 |
| RP_POS_619_29095a12_615 | | 619.2910 | 12.62 | | RP | POS | | | 307477 | 522579 | 337279 | 555526 | 6.87E-01 | 0.06 | 0.62 |
| HILIC_NEG_315_04193a7_093 | | 315.0419 | 7.09 | | HILIC | NEG | | | 1317752 | 159487 | 1308884 | 162167 | 6.87E-01 | -0.06 | 0.56 |
| HILIC_NEG_197_8078a3_244 | | 197.8078 | 3.24 | | HILIC | NEG | | | 4.7E+07 | 1.6E+07 | 4.8E+07 | 1.8E+07 | 6.88E-01 | 0.06 | 0.53 |
| RP_POS_233_09531a3_496 | | 233.0953 | 3.50 | | RP | POS | | | 1395005 | 1381413 | 1489105 | 1945854 | 6.88E-01 | 0.06 | 0.9 |
| RP_POS_384_98434a15_449 | | 384.9843 | 15.45 | | RP | POS | | | 542713 | 168874 | 552990 | 201138 | 6.88E-01 | 0.06 | 0.7 |
| RP_POS_462_14594a11_771 | | 462.1459 | 11.77 | | RP | POS | | | 374904 | 205185 | 387703 | 254394 | 6.88E-01 | 0.06 | 1.03 |
| HILIC_POS_131_11803a4_963 | | 131.1180 | 4.96 | | HILIC | POS | | | 1374007 | 768769 | 1412777 | 647524 | 6.88E-01 | 0.05 | 0.34 |
| RP_POS_181_04922a7_471 | | 181.0492 | 7.47 | | RP | POS | | | 1135262 | 158157 | 1142720 | 113930 | 6.88E-01 | 0.05 | 0.57 |
| RP_POS_536_36597a11_852 | | 536.3660 | 11.85 | | RP | POS | | | 986036 | 186727 | 996112 | 181021 | 6.88E-01 | 0.05 | 1.13 |
| RP_POS_681_50427a10_363 | | 681.5043 | 10.36 | | RP | POS | | | 1133841 | 606374 | 1166424 | 580758 | 6.88E-01 | 0.05 | 0.71 |
| RP_POS_182_08124a0_68 | | 182.0812 | 0.68 | | RP | POS | 136.0763(100), 165.0548(45) | | 8.1E+07 | 2.3E+07 | 8E+07 | 2.1E+07 | 6.88E-01 | -0.05 | 0.51 |
| RP_NEG_218_98923a15_487 | | 218.9892 | 15.49 | | RP | NEG | | | 1721887 | 486206 | 1697373 | 409875 | 6.88E-01 | -0.05 | 0.67 |
| RP_POS_256_15924a9_208 | | 256.1592 | 9.21 | | RP | POS | | | 421083 | 447588 | 395364 | 484795 | 6.88E-01 | -0.06 | 0.86 |
| RP_POS_147_11267a15_451 | Lysine | 147.1127 | 15.45 | | RP | POS | 130.0867(100), 84.9607(35) | 2 | 336811 | 76400.1 | 340864 | 71683.5 | 6.88E-01 | 0.05 | 0.59 |
| RP_POS_512_32178a9_374 | | 512.3218 | 9.37 | | RP | POS | | | 398763 | 431707 | 426349 | 556985 | 6.89E-01 | 0.06 | 0.68 |
| RP_NEG_757_1355a9_577 | | 757.1355 | 9.58 | | RP | NEG | | | 763206 | 64645.6 | 766814 | 66978.2 | 6.89E-01 | 0.05 | 0.51 |
| RP_POS_227_12732a3_92 | | 227.1273 | 3.92 | | RP | POS | | | 153683 | 84105.8 | 158269 | 83265.5 | 6.89E-01 | 0.05 | 0.68 |
| RP_POS_565_4115a10_17 | | 565.4115 | 10.17 | | RP | POS | | | 8579632 | 3372360 | 8767941 | 3506615 | 6.89E-01 | 0.05 | 0.74 |
| RP_POS_746_56403a10_667 | | 746.5640 | 10.67 | | RP | POS | | | 1083460 | 275804 | 1096752 | 210872 | 6.89E-01 | 0.05 | 0.59 |
| HILIC_POS_362_92651a4_337 | | 362.9265 | 4.34 | | HILIC | POS | | | 3.6E+07 | 2156537 | 3.6E+07 | 2243771 | 6.89E-01 | -0.05 | 0.75 |
| RP_NEG_183_00468a5_452 | | 183.0047 | 5.45 | | RP | NEG | | | 126186 | 51199.5 | 123303 | 53942.5 | 6.89E-01 | -0.05 | 0.41 |
| RP_POS_312_15924a8_991 | | 312.1592 | 8.99 | | RP | POS | | | 1524479 | 2131726 | 1425362 | 1482827 | 6.89E-01 | -0.05 | 0.34 |
| RP_POS_842_52332a13_207 | | 842.5233 | 13.21 | | RP | POS | | | 973048 | 434206 | 947731 | 485229 | 6.89E-01 | -0.06 | 0.84 |
| HILIC_NEG_100_9335a1_069 | | 100.9335 | 1.07 | | HILIC | NEG | | | 1.3E+07 | 2954051 | 1.3E+07 | 2706207 | 6.90E-01 | 0.05 | 0.72 |
| HILIC_NEG_132_90031a1_344 | | 132.9003 | 1.34 | | HILIC | NEG | | | 826473 | 293912 | 842735 | 300846 | 6.90E-01 | 0.05 | 0.7 |
| RP_NEG_247_74924a15_494 | | 247.7492 | 15.49 | | RP | NEG | | | 346796 | 25351.2 | 348219 | 26704.7 | 6.90E-01 | 0.05 | 0.67 |
| RP_NEG_452_27759a9_93 | | 452.2776 | 9.93 | | RP | NEG | | | 991440 | 364462 | 1012475 | 403190 | 6.90E-01 | 0.05 | 0.45 |
| RP_POS_1111_67102a10_292 | | 1111.6710 | 10.29 | | RP | POS | | | 634529 | 572765 | 662078 | 438094 | 6.90E-01 | 0.05 | 0.52 |
| RP_POS_467_00143a9_91 | | 467.0014 | 9.91 | | RP | POS | | | 95338.2 | 47469.6 | 97952.1 | 48438.6 | 6.90E-01 | 0.05 | 0.4 |
| RP_POS_898_55353a13_419 | | 898.5535 | 13.42 | | RP | POS | | | 1597022 | 1254359 | 1671541 | 1455432 | 6.90E-01 | 0.05 | 0.51 |
| HILIC_NEG_112_98564a3_064 | | 112.9856 | 3.06 | | HILIC | NEG | 68.9949(100), 112.9854(4) | | 9564706 | 385048 | 9543218 | 400801 | 6.90E-01 | -0.05 | 0.74 |
| RP_POS_1066_6283a10_308 | | 1066.6283 | 10.31 | | RP | POS | | | 609160 | 585886 | 579840 | 492747 | 6.90E-01 | -0.05 | 0.64 |
| RP_POS_162_95882a15_422 | | 162.9588 | 15.42 | | RP | POS | | | 2615095 | 1225118 | 2545380 | 1319166 | 6.90E-01 | -0.05 | 0.6 |
| RP_POS_446_32584a10_263 | | 446.3258 | 10.26 | | RP | POS | | | 155246 | 81865 | 150557 | 89324.5 | 6.90E-01 | -0.05 | 0.49 |
| RP_POS_428_78888a13_795 | | 428.7889 | 13.80 | | RP | POS | | | 146458 | 101168 | 140121 | 127386 | 6.90E-01 | -0.06 | 0.67 |
| RP_POS_142_1591a8_986 | | 142.1591 | 8.99 | | RP | POS | | | 684055 | 150564 | 691994 | 141905 | 6.91E-01 | 0.05 | 0.49 |
| RP_POS_537_39606a14_355 | | 537.3961 | 14.36 | | RP | POS | | | 1.1E+09 | 2.8E+08 | 1.1E+09 | 2.6E+08 | 6.91E-01 | 0.05 | 0.91 |
| RP_POS_622_3067a7_605 | | 622.3067 | 7.61 | | RP | POS | | | 1702283 | 163160 | 1711064 | 159672 | 6.91E-01 | 0.05 | 0.73 |
| HILIC_NEG_216_90984a7_733 | | 216.9098 | 7.73 | | HILIC | NEG | | | 246587 | 18648 | 245489 | 21361.2 | 6.91E-01 | -0.05 | 0.45 |
| HILIC_POS_253_11584a4_867 | | 253.1158 | 4.87 | | HILIC | POS | | | 322386 | 394591 | 301152 | 386474 | 6.91E-01 | -0.05 | 0.48 |
| RP_NEG_241_9742a0_685 | | 241.9742 | 0.69 | | RP | NEG | | | 346617 | 75965.5 | 342739 | 67439.7 | 6.91E-01 | -0.05 | 0.84 |
| RP_NEG_637_1413a9_099 | | 637.1413 | 9.10 | | RP | NEG | | | 2.1E+07 | 1006433 | 2.1E+07 | 1019214 | 6.91E-01 | -0.05 | 0.78 |
| RP_POS_155_02969a15_461 | | 155.0297 | 15.46 | | RP | POS | | | 2441203 | 749721 | 2403603 | 636764 | 6.91E-01 | -0.05 | 0.6 |
| RP_POS_267_64688a10_486 | | 267.6469 | 10.49 | | RP | POS | | | 3330431 | 2153806 | 3215250 | 2092397 | 6.91E-01 | -0.05 | 1.01 |
| RP_POS_276_14447a6_127 | | 276.1445 | 6.13 | | RP | POS | | | 336728 | 267208 | 322812 | 246879 | 6.91E-01 | -0.05 | 0.53 |
| RP_POS_646_47485a9_118 | | 646.4749 | 9.12 | | RP | POS | | | 1861175 | 1472717 | 1792076 | 1066919 | 6.91E-01 | -0.05 | 0.87 |
| HILIC_POS_328_24811a0_704 | | 328.2481 | 0.70 | | HILIC | POS | | | 3112257 | 1346691 | 3188275 | 1449065 | 6.92E-01 | 0.05 | 0.38 |
| RP_NEG_308_99765a6_359 | | 308.9977 | 6.36 | | RP | NEG | | | 3398105 | 508257 | 3428035 | 587235 | 6.92E-01 | 0.05 | 0.33 |
| RP_NEG_345_16849a9_112 | | 345.1685 | 9.11 | | RP | NEG | | | 281111 | 22649 | 282329 | 22304.9 | 6.92E-01 | 0.05 | 0.72 |
| RP_NEG_558_21381a13_3 | | 558.2138 | 13.30 | | RP | NEG | | | 139542 | 59888.6 | 142616 | 54138.1 | 6.92E-01 | 0.05 | 0.58 |
| RP_POS_149_09598a8_18 | | 149.0960 | 8.18 | | RP | POS | | | 6612508 | 536761 | 6640932 | 513230 | 6.92E-01 | 0.05 | 0.78 |
| RP_POS_258_16956a5_327 | | 258.1696 | 5.33 | | RP | POS | | | 77911.9 | 42187.9 | 80026.7 | 36218 | 6.92E-01 | 0.05 | 0.88 |
| RP_POS_816_42206a7_534 | | 816.4221 | 7.53 | | RP | POS | | | 2532631 | 202869 | 2543685 | 204628 | 6.92E-01 | 0.05 | 0.63 |
| HILIC_NEG_266_94608a4_826 | | 266.9461 | 4.83 | | HILIC | NEG | | | 754081 | 212088 | 742917 | 201027 | 6.92E-01 | -0.05 | 0.32 |
| RP_NEG_318_98065a7_628 | | 318.9807 | 7.63 | | RP | NEG | | | 3850481 | 245265 | 3838005 | 216533 | 6.92E-01 | -0.05 | 0.64 |
| RP_POS_409_27188a15_414 | | 409.2719 | 15.41 | | RP | POS | | | 723905 | 244379 | 710508 | 250055 | 6.92E-01 | -0.05 | 0.79 |
| RP_POS_648_45496a11_405 | | 648.4550 | 11.41 | | RP | POS | | | 211432 | 187701 | 202198 | 154112 | 6.92E-01 | -0.05 | 0.48 |
| RP_POS_890_48517a13_115 | | 890.4852 | 13.12 | | RP | POS | | | 777722 | 284214 | 795767 | 370814 | 6.93E-01 | 0.06 | 0.39 |
| HILIC_POS_312_02008a7_422 | | 312.0201 | 7.42 | | HILIC | POS | | | 177293 | 13914 | 178016 | 12925.4 | 6.93E-01 | 0.05 | 0.52 |
| RP_NEG_437_28909a11_745 | | 437.2891 | 11.75 | | RP | NEG | | | 175842 | 20478.1 | 176925 | 19658.2 | 6.93E-01 | 0.05 | 1.25 |
| RP_POS_235_08534a15_376 | | 235.0853 | 15.38 | | RP | POS | | | 5566354 | 2002477 | 5676731 | 2076138 | 6.93E-01 | 0.05 | 0.5 |
| RP_POS_284_12802a8_678 | | 284.1280 | 8.68 | | RP | POS | | | 583163 | 673711 | 620974 | 722506 | 6.93E-01 | 0.05 | 0.37 |
| HILIC_POS_375_32162a7_028 | | 375.3216 | 7.03 | | HILIC | POS | | | 522370 | 425996 | 504246 | 234220 | 6.93E-01 | -0.05 | 0.75 |
| RP_NEG_1107_16455a9_896 | | 1107.1646 | 9.90 | | RP | NEG | | | 3043967 | 217558 | 3032062 | 223543 | 6.93E-01 | -0.05 | 0.7 |
| RP_POS_163_13274a6_503 | | 163.1327 | 6.50 | | RP | POS | | | 3069289 | 693258 | 3026112 | 880115 | 6.93E-01 | -0.05 | 0.88 |
| RP_POS_271_65842a10_181 | | 271.6584 | 10.18 | | RP | POS | | | 635715 | 330024 | 618867 | 297087 | 6.93E-01 | -0.05 | 0.93 |
| RP_POS_274_16473a4_716 | | 274.1647 | 4.72 | | RP | POS | | | 452861 | 251908 | 439575 | 240863 | 6.93E-01 | -0.05 | 0.62 |
| RP_POS_457_35089a10_874 | | 457.3509 | 10.87 | | RP | POS | | | 6090282 | 832634 | 6043056 | 908958 | 6.93E-01 | -0.05 | 1.05 |
| RP_POS_146_06012a4_263 | | 146.0601 | 4.26 | | RP | POS | 146.0601(100), 118.0864(40) | | 590431 | 1897572 | 511505 | 945483 | 6.93E-01 | -0.06 | 0.53 |
| HILIC_NEG_131_91554a3_154 | | 131.9155 | 3.15 | | HILIC | NEG | | | 514154 | 188320 | 524554 | 197116 | 6.94E-01 | 0.05 | 0.35 |
| HILIC_NEG_242_05202a5_116 | | 242.0520 | 5.12 | | HILIC | NEG | | | 1317785 | 744253 | 1354999 | 642671 | 6.94E-01 | 0.05 | 0.46 |
| HILIC_NEG_352_01462a6_685 | | 352.0146 | 6.69 | | HILIC | NEG | | | 233941 | 28772.3 | 235518 | 29657.8 | 6.94E-01 | 0.05 | 0.36 |
| RP_POS_265_17987a8_503 | | 265.1799 | 8.50 | | RP | POS | | | 3065029 | 222352 | 3077153 | 228148 | 6.94E-01 | 0.05 | 0.47 |
| RP_POS_504_306a10_902 | | 504.3060 | 10.90 | | RP | POS | | | 3937040 | 825532 | 3986115 | 977310 | 6.94E-01 | 0.05 | 0.73 |
| RP_POS_734_50781a9_894 | | 734.5078 | 9.89 | | RP | POS | | | 663684 | 307858 | 682263 | 374447 | 6.94E-01 | 0.05 | 0.68 |
| HILIC_NEG_245_89465a0_898 | | 245.8947 | 0.90 | | HILIC | NEG | | | 1.1E+07 | 2506185 | 1.1E+07 | 2827693 | 6.94E-01 | -0.05 | 0.51 |
| RP_NEG_170_83302a15_329 | | 170.8330 | 15.33 | | RP | NEG | | | 4535179 | 368734 | 4514168 | 409116 | 6.94E-01 | -0.05 | 0.88 |
| RP_POS_267_12054a5_049 | | 267.1205 | 5.05 | | RP | POS | | | 190711 | 128142 | 183374 | 142606 | 6.94E-01 | -0.05 | 1.08 |
| RP_POS_312_21671a6_41 | | 312.2167 | 6.41 | | RP | POS | | | 728942 | 338552 | 748252 | 377396 | 6.95E-01 | 0.05 | 0.46 |

**Supplemantary table 1**: The non-targeted metabolomics results ordered by the VIP value (p value < 0.0003 are shaded dark green, p value < 0.05 are shaded light green, d values below 0 are blue and above 0 are red)

| | | | | | | | | | Cases | | Controls | | Welch test | Cohen's | |
|---|---|---|---|---|---|---|---|---|---|---|---|---|---|---|---|
| Feature_ID | Metabolite_name | Average_Mz | Average_RT_min_ | HMDB ID | RP_HILIC | POS_NEG | Cureted_MSMS | ID_level | Mean | SD | Mean2 | SD3 | p | d | VIP |
| RP_POS_425_79276a10_924 | | 425.7928 | 10.92 | | RP | POS | | | 233929 | 218806 | 245162 | 201407 | 6.95E-01 | 0.05 | 0.81 |
| RP_POS_546_45465a14_356 | | 546.4547 | 14.36 | | RP | POS | | | 4.7E+07 | 1.6E+07 | 4.8E+07 | 1.4E+07 | 6.95E-01 | 0.05 | 0.48 |
| RP_POS_624_63965a15_303 | | 624.6397 | 15.30 | | RP | POS | | | 3473797 | 1693508 | 3560480 | 1549376 | 6.95E-01 | 0.05 | 0.65 |
| RP_POS_858_5976a14_114 | | 858.5976 | 14.11 | | RP | POS | | | 5954880 | 4457637 | 6193312 | 4431821 | 6.95E-01 | 0.05 | 0.87 |
| HILIC_POS_90_09191a1_662 | | 90.0919 | 1.66 | | HILIC | POS | | | 789116 | 257993 | 776906 | 198678 | 6.95E-01 | -0.05 | 0.72 |
| RP_NEG_166_92424a15_468 | | 166.9242 | 15.47 | | RP | NEG | | | 435777 | 79152.2 | 431492 | 80555.4 | 6.95E-01 | -0.05 | 0.38 |
| RP_POS_515_31256a10_487 | | 515.3126 | 10.49 | | RP | POS | | | 4911871 | 2003733 | 4802918 | 2049853 | 6.95E-01 | -0.05 | 1.1 |
| HILIC_NEG_136_89169a6_136 | | 136.8917 | 6.14 | | HILIC | NEG | | | 5020944 | 1204046 | 5091258 | 1406987 | 6.96E-01 | 0.05 | 0.56 |
| RP_POS_585_07837a11_394 | | 585.0784 | 11.39 | | RP | POS | | | 97928.9 | 38296.7 | 100239 | 47017.1 | 6.96E-01 | 0.05 | 0.65 |
| RP_NEG_224_97995a0_678 | | 224.9800 | 0.68 | | RP | NEG | | | 208512 | 25418.4 | 207097 | 27418.2 | 6.96E-01 | -0.05 | 0.62 |
| RP_POS_232_92831a15_369 | | 232.9283 | 15.37 | | RP | POS | | | 5122658 | 1317927 | 5056451 | 1167283 | 6.96E-01 | -0.05 | 0.61 |
| RP_POS_451_99182a9_923 | | 451.9918 | 9.92 | | RP | POS | | | 153984 | 102681 | 148411 | 105391 | 6.96E-01 | -0.05 | 0.57 |
| RP_POS_744_92114a11_645 | | 744.9211 | 11.65 | | RP | POS | | | 108116 | 64012 | 111135 | 49512 | 6.97E-01 | 0.05 | 0.74 |
| HILIC_NEG_185_96265a0_937 | | 185.9627 | 0.94 | | HILIC | NEG | | | 1723354 | 493347 | 1698197 | 454183 | 6.97E-01 | -0.05 | 0.4 |
| RP_NEG_466_55023a15_494 | | 466.5502 | 15.49 | | RP | NEG | | | 258744 | 31990.4 | 257196 | 26458 | 6.97E-01 | -0.05 | 0.58 |
| RP_NEG_750_52887a12_904 | | 750.5289 | 12.90 | | RP | NEG | | | 3027626 | 1307350 | 2956779 | 1351534 | 6.97E-01 | -0.05 | 0.36 |
| RP_POS_553_06549a10_702 | | 553.0655 | 10.70 | | RP | POS | | | 199110 | 121526 | 207672 | 188896 | 6.98E-01 | 0.06 | 0.59 |
| RP_NEG_720_27838a10_614 | | 720.2784 | 10.61 | | RP | NEG | | | 562687 | 92982.9 | 567621 | 93181.6 | 6.98E-01 | 0.05 | 0.73 |
| RP_NEG_770_31085a10_616 | | 770.3109 | 10.62 | | RP | NEG | | | 1051178 | 209754 | 1061792 | 191292 | 6.98E-01 | 0.05 | 0.61 |
| RP_POS_715_57605a12_65 | | 715.5761 | 12.65 | | RP | POS | | | 1640965 | 782482 | 1686411 | 916356 | 6.98E-01 | 0.05 | 0.58 |
| HILIC_NEG_234_81973a4_916 | | 234.8197 | 4.92 | | HILIC | NEG | | | 371660 | 104748 | 365992 | 108730 | 6.98E-01 | -0.05 | 0.69 |
| RP_NEG_260_73077a15_471 | | 260.7308 | 15.47 | | RP | NEG | | | 441727 | 228735 | 431632 | 150716 | 6.98E-01 | -0.05 | 0.5 |
| RP_POS_318_1904a5_695 | | 318.1904 | 5.70 | | RP | POS | | | 251395 | 128620 | 243948 | 149889 | 6.98E-01 | -0.05 | 0.5 |
| RP_POS_337_15625a10_154 | | 337.1563 | 10.15 | | RP | POS | | | 4954465 | 1594887 | 4871362 | 1538062 | 6.98E-01 | -0.05 | 0.58 |
| HILIC_NEG_183_93407a1_263 | | 183.9341 | 1.26 | | HILIC | NEG | | | 1739568 | 750994 | 1779233 | 750398 | 6.99E-01 | 0.05 | 0.39 |
| RP_NEG_251_78716a15_497 | | 251.7872 | 15.50 | | RP | NEG | | | 636798 | 47615.8 | 639289 | 46623.5 | 6.99E-01 | 0.05 | 0.57 |
| RP_NEG_532_29907a10_359 | | 532.2991 | 10.36 | | RP | NEG | | | 951523 | 140141 | 959267 | 151914 | 6.99E-01 | 0.05 | 0.91 |
| RP_POS_146_11763a0_556 | | 146.1176 | 0.56 | | RP | POS | | | 8392226 | 1453134 | 8475954 | 1685983 | 6.99E-01 | 0.05 | 0.52 |
| RP_POS_350_89221a15_337 | | 350.8922 | 15.34 | | RP | POS | | | 301416 | 223585 | 313077 | 218085 | 6.99E-01 | 0.05 | 0.83 |
| HILIC_NEG_259_00113a4_244 | | 259.0011 | 4.24 | | HILIC | NEG | | | 578650 | 233989 | 566384 | 230526 | 6.99E-01 | -0.05 | 0.7 |
| HILIC_NEG_312_8363a4_47 | | 312.8363 | 4.47 | | HILIC | NEG | | | 2205075 | 859193 | 2158093 | 914905 | 6.99E-01 | -0.05 | 0.54 |
| RP_NEG_230_98637a6_076 | | 230.9864 | 6.08 | | RP | NEG | | | 2599312 | 868281 | 2555992 | 773532 | 6.99E-01 | -0.05 | 0.49 |
| RP_NEG_273_82697a15_51 | | 273.8270 | 15.51 | | RP | NEG | | | 546045 | 50121.6 | 543464 | 47640.6 | 6.99E-01 | -0.05 | 0.45 |
| RP_POS_211_0946a6_717 | | 211.0946 | 6.72 | | RP | POS | | | 750167 | 446940 | 725311 | 490169 | 6.99E-01 | -0.05 | 0.76 |
| RP_POS_308_86795a15_402 | | 308.8680 | 15.40 | | RP | POS | | | 175678 | 116270 | 169881 | 103473 | 6.99E-01 | -0.05 | 0.46 |
| RP_POS_551_41034a15_001 | | 551.4103 | 15.00 | | RP | POS | | | 6338394 | 2134439 | 6215117 | 2482604 | 6.99E-01 | -0.05 | 0.61 |
| RP_POS_563_37732a9_137 | | 563.3773 | 9.14 | | RP | POS | | | 193695 | 149618 | 185778 | 149679 | 6.99E-01 | -0.05 | 0.72 |
| HILIC_POS_106_08674a1_837 | | 106.0867 | 1.84 | | HILIC | POS | 88.0761(100), 106.0872(90) | | 2E+07 | 1E+07 | 2E+07 | 8837652 | 7.00E-01 | 0.05 | 0.78 |
| HILIC_POS_352_24615a5_933 | | 352.2462 | 5.93 | | HILIC | POS | | | 139614 | 89875.9 | 145589 | 129741 | 7.00E-01 | 0.05 | 0.92 |
| RP_POS_652_95544a11_556 | | 652.9554 | 11.56 | | RP | POS | | | 188154 | 91211.1 | 193737 | 116912 | 7.00E-01 | 0.05 | 0.63 |
| RP_POS_137_09613a10_36 | | 137.0961 | 10.36 | | RP | POS | | | 1251707 | 239144 | 1239243 | 235003 | 7.00E-01 | -0.05 | 0.49 |
| RP_POS_165_05481a0_68 | | 165.0548 | 0.68 | | RP | POS | | | 1.5E+07 | 4316061 | 1.5E+07 | 3904863 | 7.00E-01 | -0.05 | 0.5 |
| RP_POS_346_21552a10_155 | | 346.2155 | 10.16 | | RP | POS | | | 495432 | 167472 | 486918 | 156425 | 7.00E-01 | -0.05 | 0.58 |
| RP_POS_455_66919a10_564 | | 455.6692 | 10.56 | | RP | POS | | | 168786 | 86609.2 | 164193 | 87533.3 | 7.00E-01 | -0.05 | 0.46 |
| RP_POS_728_58582a11_858 | | 728.5858 | 11.86 | | RP | POS | | | 245982 | 266076 | 234683 | 157873 | 7.00E-01 | -0.05 | 0.33 |
| HILIC_NEG_142_92921a5_65 | | 142.9292 | 5.65 | | HILIC | NEG | | | 1740273 | 269996 | 1725375 | 294591 | 7.01E-01 | -0.05 | 0.55 |
| RP_NEG_154_89828a15_49 | | 154.8983 | 15.49 | | RP | NEG | | | 504295 | 28263.2 | 502843 | 27056.5 | 7.01E-01 | -0.05 | 0.62 |
| RP_NEG_316_70758a15_497 | | 316.7076 | 15.50 | | RP | NEG | | | 254756 | 49341.4 | 252092 | 51871.6 | 7.01E-01 | -0.05 | 0.58 |
| RP_NEG_376_12888a11_142 | | 376.1289 | 11.14 | | RP | NEG | | | 1267370 | 119156 | 1261054 | 121091 | 7.01E-01 | -0.05 | 0.6 |
| RP_POS_191_16396a7_42 | | 191.1640 | 7.42 | | RP | POS | | | 3047259 | 129733 | 3039750 | 153581 | 7.01E-01 | -0.05 | 0.46 |
| RP_POS_261_10034a15_39 | | 261.1003 | 15.39 | | RP | POS | | | 398978 | 193049 | 388817 | 193838 | 7.01E-01 | -0.05 | 0.44 |
| RP_POS_313_23657a8_48 | | 313.2366 | 8.48 | | RP | POS | | | 661572 | 217988 | 650709 | 196391 | 7.01E-01 | -0.05 | 0.49 |
| RP_POS_451_16446a8_573 | | 451.1645 | 8.57 | | RP | POS | | | 105763 | 108406 | 99954.8 | 112305 | 7.01E-01 | -0.05 | 0.37 |
| RP_NEG_239_06718a8_297 | Dinoseb | 239.0672 | 8.30 | | RP | NEG | 239.0666(100) | 2 | 123812 | 44422.4 | 121734 | 35118.1 | 7.01E-01 | -0.05 | 0.53 |
| HILIC_NEG_129_93625a6_293 | | 129.9363 | 6.29 | | HILIC | NEG | | | 2515744 | 1120024 | 2577744 | 1237450 | 7.02E-01 | 0.05 | 0.45 |
| RP_NEG_203_88664a14_335 | | 203.8866 | 14.34 | | RP | NEG | | | 654462 | 242917 | 667966 | 270342 | 7.02E-01 | 0.05 | 0.56 |
| RP_POS_489_29575a12_147 | | 489.2958 | 12.15 | | RP | POS | | | 527097 | 166193 | 536408 | 187688 | 7.02E-01 | 0.05 | 0.66 |
| RP_POS_822_53998a13_39 | | 822.5400 | 13.39 | | RP | POS | | | 7004495 | 4712861 | 7247062 | 4555225 | 7.02E-01 | 0.05 | 0.72 |
| HILIC_NEG_205_83961a3_246 | | 205.8396 | 3.25 | | HILIC | NEG | | | 1221959 | 275479 | 1206952 | 295490 | 7.02E-01 | -0.05 | 0.64 |
| RP_NEG_302_99142a15_505 | | 302.9914 | 15.51 | | RP | NEG | | | 795895 | 75379.3 | 791507 | 90440.7 | 7.02E-01 | -0.05 | 0.45 |
| RP_POS_445_36835a13_149 | | 445.3684 | 13.15 | | RP | POS | | | 2832617 | 733627 | 2795736 | 681372 | 7.02E-01 | -0.05 | 0.34 |
| HILIC_POS_142_1225a0_966 | | 142.1225 | 0.97 | | HILIC | POS | | | 896690 | 289591 | 911268 | 270247 | 7.03E-01 | 0.05 | 0.74 |
| RP_POS_553_45862a14_233 | | 553.4586 | 14.23 | | RP | POS | | | 1589665 | 946495 | 1637022 | 877014 | 7.03E-01 | 0.05 | 0.38 |
| RP_NEG_1151_65234a10_312 | | 1151.6523 | 10.31 | | RP | NEG | | | 1198023 | 398377 | 1176879 | 412202 | 7.03E-01 | -0.05 | 0.66 |
| RP_NEG_136_87241a14_338 | | 136.8724 | 14.34 | | RP | NEG | | | 859300 | 403974 | 840657 | 312806 | 7.03E-01 | -0.05 | 0.23 |
| RP_POS_556_27441a11_147 | | 556.2744 | 11.15 | | RP | POS | | | 320194 | 194538 | 310102 | 193294 | 7.03E-01 | -0.05 | 0.64 |
| HILIC_POS_104_07114a4_671 | N,N-Dimethylglycine | 104.0711 | 4.67 | | HILIC | POS | 58.0663(100), 104.0716(25) | 2 | 4361249 | 3016215 | 4231979 | 1909677 | 7.03E-01 | -0.05 | 0.45 |
| RP_NEG_512_29974a9_93 | | 512.2997 | 9.93 | | RP | NEG | | | 2.4E+07 | 8610539 | 2.4E+07 | 9335684 | 7.04E-01 | 0.05 | 0.45 |
| RP_POS_225_1481a8_102 | | 225.1481 | 8.10 | | RP | POS | | | 2448104 | 1653490 | 2529582 | 1494257 | 7.04E-01 | 0.05 | 0.52 |
| RP_POS_254_04335a15_418 | | 254.0434 | 15.42 | | RP | POS | | | 2230559 | 678704 | 2264445 | 630837 | 7.04E-01 | 0.05 | 0.61 |
| RP_POS_833_65027a14_94 | | 833.6503 | 14.94 | | RP | POS | | | 2047212 | 2119774 | 2168645 | 2507322 | 7.04E-01 | 0.05 | 0.52 |
| RP_POS_561_23535a8_842 | | 561.2354 | 8.84 | | RP | POS | | | 199146 | 119025 | 192836 | 123837 | 7.04E-01 | -0.05 | 0.79 |
| RP_POS_894_51715a14_093 | | 894.5172 | 14.09 | | RP | POS | | | 1.6E+07 | 2.8E+07 | 1.4E+07 | 2.9E+07 | 7.04E-01 | -0.05 | 0.74 |
| HILIC_POS_277_10443a2_974 | | 277.1044 | 2.97 | | HILIC | POS | | | 329129 | 177361 | 338283 | 176925 | 7.05E-01 | 0.05 | 0.66 |
| RP_NEG_359_97528a0_681 | | 359.9753 | 0.68 | | RP | NEG | | | 303864 | 76959.4 | 307613 | 68407.1 | 7.05E-01 | 0.05 | 0.34 |
| RP_NEG_580_28754a9_93 | | 580.2875 | 9.93 | | RP | NEG | | | 1690134 | 598895 | 1722353 | 640933 | 7.05E-01 | 0.05 | 0.46 |
| RP_POS_289_15051a0_68 | | 289.1505 | 0.68 | | RP | POS | | | 664628 | 590914 | 699010 | 718723 | 7.05E-01 | 0.05 | 1.29 |
| RP_POS_412_77744a14_081 | | 412.7774 | 14.08 | | RP | POS | | | 5282710 | 9275912 | 5771929 | 9593863 | 7.05E-01 | 0.05 | 0.54 |
| RP_POS_731_17267a11_514 | | 731.1727 | 11.51 | | RP | POS | | | 533534 | 993098 | 589624 | 1156212 | 7.05E-01 | 0.05 | 0.62 |
| RP_POS_227_12785a6_445 | | 227.1279 | 6.45 | | RP | POS | | | 841913 | 241439 | 828942 | 257237 | 7.05E-01 | -0.05 | 0.73 |
| RP_POS_524_85162a9_588 | | 524.8516 | 9.59 | | RP | POS | | | 912218 | 536756 | 886362 | 464443 | 7.05E-01 | -0.05 | 0.47 |
| RP_POS_517_30511a13_116 | | 517.3051 | 13.12 | | RP | POS | | | 460070 | 256666 | 472062 | 209006 | 7.06E-01 | 0.05 | 0.68 |
| RP_POS_748_58112a13_934 | | 748.5811 | 13.93 | | RP | POS | | | 283910 | 356763 | 300567 | 292393 | 7.06E-01 | 0.05 | 0.8 |
| HILIC_NEG_249_89148a0_898 | | 249.8915 | 0.90 | | HILIC | NEG | | | 337188 | 77038.8 | 332928 | 87027 | 7.06E-01 | -0.05 | 0.49 |
| RP_NEG_235_92657a15_615 | | 235.9266 | 15.62 | | RP | NEG | | | 9543638 | 4810096 | 9280539 | 5324003 | 7.06E-01 | -0.05 | 0.45 |
| RP_POS_165_05492a1_225 | | 165.0549 | 1.23 | | RP | POS | | | 2157598 | 768769 | 2121091 | 650228 | 7.06E-01 | -0.05 | 0.5 |
| RP_POS_255_05859a10_566 | | 255.0586 | 10.57 | | RP | POS | | | 1243151 | 324582 | 1226931 | 304983 | 7.06E-01 | -0.05 | 0.5 |
| HILIC_NEG_235_82018a5_869 | | 235.8202 | 5.87 | | HILIC | NEG | | | 212204 | 83232.7 | 216509 | 83964.7 | 7.07E-01 | 0.05 | 0.35 |
| HILIC_POS_228_19595a1_349 | | 228.1960 | 1.35 | | HILIC | POS | | | 700085 | 263049 | 713169 | 247270 | 7.07E-01 | 0.05 | 0.46 |
| RP_NEG_301_16599a8_582 | | 301.1660 | 8.58 | | RP | NEG | | | 565522 | 246652 | 578096 | 242527 | 7.07E-01 | 0.05 | 0.95 |
| RP_POS_430_88705a8_926 | | 430.8871 | 8.93 | | RP | POS | | | 1091188 | 133943 | 1098209 | 138428 | 7.07E-01 | 0.05 | 0.53 |
| RP_POS_442_2998a10_089 | | 442.2998 | 10.09 | | RP | POS | | | 167569 | 96438.4 | 173001 | 112735 | 7.07E-01 | 0.05 | 0.81 |
| RP_POS_443_39127a11_344 | | 443.3913 | 11.34 | | RP | POS | | | 558744 | 552020 | 589579 | 636606 | 7.07E-01 | 0.05 | 0.86 |
| RP_POS_607_39008a13_696 | | 607.3901 | 13.70 | | RP | POS | | | 554688 | 370848 | 574860 | 409227 | 7.07E-01 | 0.05 | 0.81 |
| RP_POS_637_04889a11_259 | | 637.0489 | 11.26 | | RP | POS | | | 126078 | 69908 | 130109 | 84801.9 | 7.07E-01 | 0.05 | 0.4 |
| RP_NEG_280_98309a7_342 | | 280.9831 | 7.34 | | RP | NEG | | | 2946617 | 520713 | 2920016 | 515405 | 7.07E-01 | -0.05 | 0.55 |
| RP_NEG_299_73068a15_495 | | 299.7307 | 15.50 | | RP | NEG | | | 729714 | 42969.9 | 727473 | 44073.5 | 7.07E-01 | -0.05 | 0.78 |
| RP_POS_267_12018a4_149 | | 267.1202 | 4.15 | | RP | POS | | | 205223 | 173070 | 196714 | 158999 | 7.07E-01 | -0.05 | 1.11 |
| RP_POS_165_09088a11_222 | Phenethylacetate | 165.0909 | 11.22 | | RP | POS | 165.0906(100), 104.9634(30) | 2 | 1549292 | 486462 | 1524901 | 463810 | 7.07E-01 | -0.05 | 0.57 |
| HILIC_POS_106_08658a5_325 | | 106.0866 | 5.33 | | HILIC | POS | 88.0761(100), 70.066(60) | | 2.4E+07 | 2E+07 | 2.5E+07 | 2.3E+07 | 7.08E-01 | 0.05 | 0.53 |
| HILIC_NEG_136_89172a5_653 | | 136.8917 | 5.65 | | HILIC | NEG | | | 4841758 | 1567416 | 4924402 | 1655483 | 7.08E-01 | 0.05 | 0.41 |
| HILIC_NEG_352_99716a7_352 | | 352.9972 | 7.35 | | HILIC | NEG | | | 1464613 | 66730 | 1468027 | 66406.2 | 7.08E-01 | 0.05 | 0.51 |
| RP_NEG_502_27094a9_93 | | 502.2709 | 9.93 | | RP | NEG | | | 728828 | 262397 | 742914 | 285860 | 7.08E-01 | 0.05 | 0.4 |
| RP_POS_138_0914a3_602 | | 138.0914 | 3.60 | | RP | POS | | | 69225.2 | 47442.3 | 71754.8 | 50838.5 | 7.08E-01 | 0.05 | 0.92 |
| RP_NEG_271_01526a0_669 | | 271.0153 | 0.67 | | RP | NEG | | | 252546 | 85510.3 | 248502 | 72726.6 | 7.08E-01 | -0.05 | 0.32 |
| RP_POS_169_08601a2_998 | | 169.0860 | 3.00 | | RP | POS | | | 441583 | 226810 | 430597 | 203263 | 7.08E-01 | -0.05 | 0.84 |
| RP_POS_346_17935a10_684 | | 346.1794 | 10.68 | | RP | POS | | | 524502 | 125273 | 518199 | 120803 | 7.08E-01 | -0.05 | 0.88 |
| RP_POS_350_22964a10_252 | | 350.2296 | 10.25 | | RP | POS | | | 625635 | 110657 | 619358 | 131967 | 7.08E-01 | -0.05 | 0.55 |
| RP_POS_372_12369a9_207 | | 372.1237 | 9.21 | | RP | POS | | | 465534 | 348023 | 447221 | 364681 | 7.08E-01 | -0.05 | 0.8 |
| RP_POS_496_62576a10_484 | | 496.6258 | 10.48 | | RP | POS | | | 551233 | 439017 | 531734 | 323276 | 7.08E-01 | -0.05 | 0.54 |
| HILIC_NEG_255_8223a3_215 | | 255.8223 | 3.22 | | HILIC | NEG | | | 2679072 | 237247 | 2691465 | 248135 | 7.09E-01 | 0.05 | 0.5 |
| RP_POS_501_37564a10_872 | | 501.3756 | 10.87 | | RP | POS | | | 2863978 | 558702 | 2891686 | 529789 | 7.09E-01 | 0.05 | 1.22 |
| RP_POS_535_35492a10_929 | | 535.3549 | 10.93 | | RP | POS | | | 1713546 | 515673 | 1741763 | 581072 | 7.09E-01 | 0.05 | 0.83 |
| RP_POS_569_36688a15_067 | | 569.3669 | 15.07 | | RP | POS | | | 2.5E+07 | 7033328 | 2.5E+07 | 7881951 | 7.09E-01 | 0.05 | 0.77 |
| HILIC_NEG_119_9077a6_292 | | 119.9077 | 6.29 | | HILIC | NEG | | | 2248542 | 980194 | 2198451 | 985543 | 7.09E-01 | -0.05 | 0.42 |
| HILIC_NEG_248_85771a4_352 | | 248.8577 | 4.35 | | HILIC | NEG | | | 351103 | 84302 | 346891 | 80921.8 | 7.09E-01 | -0.05 | 0.29 |
| HILIC_POS_176_00871a4_856 | | 176.0087 | 4.86 | | HILIC | POS | | | 291368 | 127978 | 284803 | 129192 | 7.09E-01 | -0.05 | 0.59 |
| HILIC_POS_245_18585a1_944 | | 245.1859 | 1.94 | | HILIC | POS | | | 304806 | 261606 | 287985 | 377974 | 7.09E-01 | -0.05 | 0.57 |
| RP_POS_449_32651a9_379 | | 449.3265 | 9.38 | | RP | POS | | | 1852103 | 1043832 | 1799528 | 1016575 | 7.09E-01 | -0.05 | 0.81 |
| RP_NEG_170_83331a11_141 | | 170.8333 | 11.14 | | RP | NEG | | | 209824 | 51546.4 | 212373 | 48910.2 | 7.10E-01 | 0.05 | 0.76 |
| RP_POS_235_16916a9_092 | | 235.1692 | 9.09 | | RP | POS | | | 4.6E+07 | 1764882 | 4.6E+07 | 1741164 | 7.10E-01 | 0.05 | 1.07 |
| RP_POS_299_22156a9_463 | | 299.2216 | 9.46 | | RP | POS | | | 994646 | 357665 | 1013970 | 399007 | 7.10E-01 | 0.05 | 0.86 |
| RP_POS_329_21048a10_057 | | 329.2105 | 10.06 | | RP | POS | | | 734629 | 81126.8 | 738984 | 89159 | 7.10E-01 | 0.05 | 0.47 |
| RP_POS_449_15289a0_629 | | 449.1529 | 0.63 | | RP | POS | | | 1315377 | 193342 | 1324978 | 184394 | 7.10E-01 | 0.05 | 0.56 |
| RP_POS_704_49731a10_235 | | 704.4973 | 10.24 | | RP | POS | | | 3447222 | 1516704 | 3538652 | 2000077 | 7.10E-01 | 0.05 | 0.76 |
| RP_NEG_291_15988a8_669 | | 291.1599 | 8.67 | | RP | NEG | | | 1641354 | 87675.7 | 1636810 | 90722.1 | 7.10E-01 | -0.05 | 0.36 |
| RP_POS_167_14316a7_695 | | 167.1432 | 7.70 | | RP | POS | | | 1085680 | 82353.8 | 1081428 | 84584.6 | 7.10E-01 | -0.05 | 0.62 |
| RP_POS_382_29938a10_662 | | 382.2994 | 10.66 | | RP | POS | | | 145848 | 96522.1 | 141474 | 76018.5 | 7.10E-01 | -0.05 | 0.62 |
| RP_POS_411_75201a13_79 | | 411.7520 | 13.79 | | RP | POS | | | 638540 | 432911 | 614574 | 501756 | 7.10E-01 | -0.05 | 0.96 |
| RP_POS_651_44238a9_916 | | 651.4424 | 9.92 | | RP | POS | | | 539677 | 289981 | 523260 | 348973 | 7.10E-01 | -0.05 | 0.62 |
| RP_POS_670_60736a14_048 | | 670.6074 | 14.05 | | RP | POS | | | 642854 | 326511 | 627276 | 289321 | 7.10E-01 | -0.05 | 0.62 |
| RP_NEG_294_98279a6_068 | | 294.9828 | 6.07 | | RP | NEG | | | 2008563 | 649855 | 2041547 | 650162 | 7.11E-01 | 0.05 | 0.48 |
| RP_POS_240_88046a15_398 | | 240.8805 | 15.40 | | RP | POS | | | 8636729 | 1530882 | 8708072 | 1296200 | 7.11E-01 | 0.05 | 0.83 |
| RP_POS_273_17957a10_689 | | 273.1796 | 10.69 | | RP | POS | | | 152480 | 115219 | 157797 | 95523.6 | 7.11E-01 | 0.05 | 0.52 |
| RP_POS_389_25311a9_85 | | 389.2531 | 9.85 | | RP | POS | | | 624857 | 288667 | 640062 | 310390 | 7.11E-01 | 0.05 | 0.91 |
| RP_POS_613_47992a13_912 | | 613.4799 | 13.91 | | RP | POS | | | 967086 | 746008 | 1012731 | 1010840 | 7.11E-01 | 0.05 | 0.73 |
| RP_POS_797_59015a14_095 | | 797.5902 | 14.10 | | RP | POS | | | 1E+07 | 1.3E+07 | 1.1E+07 | 1.4E+07 | 7.11E-01 | 0.05 | 0.83 |
| HILIC_NEG_164_03865a6_09 | | 164.0387 | 6.09 | | HILIC | NEG | | | 254050 | 124604 | 247442 | 135460 | 7.11E-01 | -0.05 | 0.75 |
| RP_NEG_192_98962a6_399 | | 192.9896 | 6.40 | | RP | NEG | | | 2190929 | 474906 | 2167586 | 449150 | 7.11E-01 | -0.05 | 0.36 |
| RP_NEG_348_99075a8_756 | | 348.9908 | 8.76 | | RP | NEG | | | 829546 | 91957.6 | 825041 | 86471.9 | 7.11E-01 | -0.05 | 0.26 |
| RP_NEG_601_16296a9_469 | | 601.1630 | 9.47 | | RP | NEG | | | 690690 | 42106.5 | 688599 | 40675 | 7.11E-01 | -0.05 | 0.83 |
| RP_NEG_740_52551a13_26 | | 740.5255 | 13.26 | | RP | NEG | | | 698843 | 338514 | 680395 | 385515 | 7.11E-01 | -0.05 | 0.41 |
| RP_POS_271_22668a9_935 | | 271.2267 | 9.94 | | RP | POS | | | 1017093 | 128883 | 1010747 | 122220 | 7.11E-01 | -0.05 | 0.88 |
| RP_POS_328_23264a4_89 | | 328.2326 | 4.89 | | RP | POS | | | 380170 | 220617 | 369869 | 186892 | 7.11E-01 | -0.05 | 0.51 |
| RP_POS_478_97968a10_23 | | 478.9797 | 10.23 | | RP | POS | | | 210236 | 102127 | 205464 | 86868.6 | 7.11E-01 | -0.05 | 0.47 |
| RP_POS_568_07416a10_679 | | 568.0742 | 10.68 | | RP | POS | | | 223199 | 119176 | 216412 | 145072 | 7.11E-01 | -0.05 | 0.64 |
| HILIC_POS_257_1467a3_619 | | 257.1467 | 3.62 | | HILIC | POS | | | 7757267 | 6.2E+07 | 1.1E+07 | 5.9E+07 | 7.12E-01 | 0.05 | 0.46 |
| RP_NEG_277_82156a15_504 | | 277.8216 | 15.50 | | RP | NEG | | | 152433 | 28197 | 153739 | 23702.8 | 7.12E-01 | 0.05 | 0.53 |
| RP_NEG_599_32019a12_938 | | 599.3202 | 12.94 | | RP | NEG | | | 1.8E+07 | 1.7E+07 | 1.9E+07 | 1.8E+07 | 7.12E-01 | 0.05 | 0.69 |
| RP_NEG_766_53973a13_83 | | 766.5397 | 13.83 | | RP | NEG | | | 2197574 | 799401 | 2234631 | 672881 | 7.12E-01 | 0.05 | 0.63 |
| RP_POS_303_18011a8_585 | | 303.1801 | 8.59 | | RP | POS | | | 1041301 | 464139 | 1064471 | 453615 | 7.12E-01 | 0.05 | 1 |
| HILIC_POS_150_11237a4_804 | | 150.1124 | 4.80 | | HILIC | POS | 150.1128(100), 132.1028(35) | | 2310336 | 1315979 | 2249353 | 1108385 | 7.12E-01 | -0.05 | 1.08 |
| HILIC_POS_144_98193a3_187 | | 144.9819 | 3.19 | | HILIC | POS | | | 360229 | 221646 | 349565 | 202010 | 7.12E-01 | -0.05 | 0.28 |
| RP_NEG_385_63263a15_5 | | 385.6326 | 15.50 | | RP | NEG | | | 249721 | 31183.2 | 248005 | 36122 | 7.12E-01 | -0.05 | 0.64 |
| RP_POS_592_33734a10_436 | | 592.3373 | 10.44 | | RP | POS | | | 1602386 | 563091 | 1572083 | 629877 | 7.12E-01 | -0.05 | 0.52 |
| HILIC_NEG_283_71011a1_406 | | 283.7101 | 1.41 | | HILIC | NEG | | | 247224 | 41132.6 | 249230 | 38690.1 | 7.13E-01 | 0.05 | 0.42 |
| HILIC_POS_191_00824a0_846 | | 191.0082 | 0.85 | | HILIC | POS | | | 223177 | 78778.4 | 226993 | 73336.2 | 7.13E-01 | 0.05 | 0.71 |
| RP_NEG_302_97211a2_163 | | 302.9721 | 2.16 | | RP | NEG | | | 324440 | 66177.1 | 327811 | 67604.4 | 7.13E-01 | 0.05 | 0.56 |
| RP_NEG_473_34833a11_264 | | 473.3483 | 11.26 | | RP | NEG | | | 1772358 | 4994376 | 2032660 | 5319565 | 7.13E-01 | 0.05 | 0.32 |
| RP_NEG_473_83411a15_399 | | 473.8341 | 15.40 | | RP | NEG | | | 304300 | 81091.7 | 308770 | 95116.4 | 7.13E-01 | 0.05 | 0.36 |
| RP_POS_132_08115a2_884 | | 132.0812 | 2.88 | | RP | POS | | | 2275911 | 405722 | 2296205 | 402100 | 7.13E-01 | 0.05 | 0.64 |
| RP_POS_222_18307a9_447 | | 222.1831 | 9.45 | | RP | POS | | | 1209372 | 509583 | 1235559 | 529180 | 7.13E-01 | 0.05 | 0.67 |

**Supplemantary table 1**: The non-targeted metabolomics results ordered by the VIP value (p value < 0.0003 are shaded dark green, p value < 0.05 are shaded light green, d values below 0 are blue and above 0 are red)

| | | | | | | | | | Cases | | Controls | | Welch test | Cohen's | |
|---|---|---|---|---|---|---|---|---|---|---|---|---|---|---|---|
| Feature_ID | Metabolite_name | Average_Mz | Average_RT_min_ | HMDB ID | RP_HILIC | POS_NEG | Cureted_MSMS | ID_level | Mean | SD | Mean2 | SD3 | p | d | VIP |
| RP_POS_291_19513a9_918 | | 291.1951 | 9.92 | | RP | POS | | | 785305 | 115974 | 791192 | 117883 | 7.13E-01 | 0.05 | 0.41 |
| RP_POS_375_16824a9_456 | | 375.1682 | 9.46 | | RP | POS | | | 176847 | 151386 | 185852 | 199730 | 7.13E-01 | 0.05 | 0.65 |
| RP_POS_575_49213a14_048 | | 575.4921 | 14.05 | | RP | POS | | | 4654955 | 1849938 | 4755892 | 2134564 | 7.13E-01 | 0.05 | 0.75 |
| HILIC_NEG_183_93454a6_112 | | 183.9345 | 6.11 | | HILIC | NEG | | | 950655 | 324088 | 934920 | 303833 | 7.13E-01 | -0.05 | 0.62 |
| RP_POS_258_89163a15_399 | | 258.8916 | 15.40 | | RP | POS | | | 3735748 | 821051 | 3694029 | 835138 | 7.13E-01 | -0.05 | 0.47 |
| RP_POS_334_02505a0_63 | | 334.0251 | 0.63 | | RP | POS | | | 392442 | 193175 | 382871 | 188460 | 7.13E-01 | -0.05 | 0.99 |
| RP_POS_350_98621a8_091 | | 350.9862 | 8.09 | | RP | POS | | | 1093855 | 253284 | 1080914 | 260111 | 7.13E-01 | -0.05 | 0.39 |
| RP_POS_432_28012a4_612 | | 432.2801 | 4.61 | | RP | POS | | | 807436 | 208089 | 821276 | 322352 | 7.14E-01 | 0.05 | 0.53 |
| RP_POS_607_09283a11_341 | | 607.0928 | 11.34 | | RP | POS | | | 118135 | 64941.9 | 121745 | 77553.1 | 7.14E-01 | 0.05 | 0.77 |
| HILIC_NEG_100_93343a5_657 | | 100.9334 | 5.66 | | HILIC | NEG | | | 3.7E+07 | 6033096 | 3.7E+07 | 6443713 | 7.14E-01 | -0.05 | 0.44 |
| RP_NEG_138_86906a15_418 | | 138.8691 | 15.42 | | RP | NEG | | | 626055 | 235372 | 613994 | 244982 | 7.14E-01 | -0.05 | 0.56 |
| RP_NEG_626_27667a10_492 | | 626.2767 | 10.49 | | RP | NEG | | | 5521704 | 757738 | 5481004 | 852802 | 7.14E-01 | -0.05 | 0.91 |
| RP_POS_471_6723a10_081 | | 471.6723 | 10.08 | | RP | POS | | | 152446 | 90100.5 | 147280 | 112803 | 7.14E-01 | -0.05 | 0.37 |
| RP_POS_744_55426a13_947 | | 744.5543 | 13.95 | | RP | POS | | | 3422877 | 4690486 | 3195912 | 4379605 | 7.14E-01 | -0.05 | 0.77 |
| RP_POS_816_5509a13_075 | | 816.5509 | 13.08 | | RP | POS | | | 4192965 | 2007136 | 4081674 | 2392307 | 7.14E-01 | -0.05 | 0.69 |
| RP_POS_169_0358a0_678 | Uric acid | 169.0358 | 0.68 | | RP | POS | 169.0352(100), 141.0412(20) | 2 | 7.6E+07 | 1.7E+07 | 7.7E+07 | 1.5E+07 | 7.14E-01 | 0.05 | 0.39 |
| HILIC_POS_202_15488a3_432 | | 202.1549 | 3.43 | | HILIC | POS | | | 1233486 | 2380769 | 1445997 | 5338891 | 7.15E-01 | 0.06 | 0.76 |
| RP_POS_235_15376a6_724 | | 235.1538 | 6.72 | | RP | POS | | | 551793 | 55439.7 | 554665 | 59220.7 | 7.15E-01 | 0.05 | 0.72 |
| HILIC_POS_136_07562a5_12 | | 136.0756 | 5.12 | | HILIC | POS | | | 1874469 | 646666 | 1844780 | 548514 | 7.15E-01 | -0.05 | 0.53 |
| HILIC_NEG_165_83742a0_941 | | 165.8374 | 0.94 | | HILIC | NEG | | | 582140 | 70321.2 | 578402 | 78373.5 | 7.15E-01 | -0.05 | 0.29 |
| HILIC_NEG_212_91115a4_096 | | 212.9112 | 4.10 | | HILIC | NEG | | | 2650516 | 628248 | 2620825 | 565942 | 7.15E-01 | -0.05 | 0.38 |
| HILIC_NEG_215_99699a4_852 | | 215.9970 | 4.85 | | HILIC | NEG | | | 201649 | 123397 | 196441 | 84648.2 | 7.15E-01 | -0.05 | 0.62 |
| HILIC_POS_228_02216a4_853 | | 228.0222 | 4.85 | | HILIC | POS | | | 809951 | 331723 | 794265 | 299054 | 7.15E-01 | -0.05 | 0.62 |
| HILIC_POS_366_28418a3_126 | | 366.2842 | 3.13 | | HILIC | POS | | | 638057 | 95751.4 | 633068 | 103463 | 7.15E-01 | -0.05 | 0.33 |
| RP_POS_162_076a0_645 | | 162.0760 | 0.65 | | RP | POS | | | 5898625 | 2149163 | 5792332 | 2113214 | 7.15E-01 | -0.05 | 0.55 |
| RP_POS_229_15849a8_98 | | 229.1585 | 8.98 | | RP | POS | | | 984762 | 81922.9 | 980816 | 76733.1 | 7.15E-01 | -0.05 | 0.79 |
| RP_POS_289_04962a2_885 | | 289.0496 | 2.89 | | RP | POS | | | 192912 | 66853.6 | 189391 | 73450.3 | 7.15E-01 | -0.05 | 0.53 |
| RP_POS_533_32562a8_458 | | 533.3256 | 8.46 | | RP | POS | | | 603461 | 995135 | 557381 | 856761 | 7.15E-01 | -0.05 | 0.47 |
| RP_POS_237_22092a10_107 | | 237.2209 | 10.11 | | RP | POS | | | 1048430 | 143208 | 1055606 | 145177 | 7.16E-01 | 0.05 | 0.81 |
| RP_POS_781_55261a13_21 | | 781.5526 | 13.21 | | RP | POS | | | 8.8E+07 | 3E+07 | 9E+07 | 2.8E+07 | 7.16E-01 | 0.05 | 0.7 |
| HILIC_NEG_118_92428a3_061 | | 118.9243 | 3.06 | | HILIC | NEG | | | 803882 | 162588 | 795544 | 171470 | 7.16E-01 | -0.05 | 0.42 |
| HILIC_POS_371_24002a3_881 | | 371.2400 | 3.88 | | HILIC | POS | | | 4875717 | 6096155 | 4591497 | 5377838 | 7.16E-01 | -0.05 | 0.36 |
| RP_NEG_215_86826a14_33 | | 215.8683 | 14.33 | | RP | NEG | | | 333576 | 150187 | 326031 | 152725 | 7.16E-01 | -0.05 | 0.49 |
| RP_NEG_328_88028a15_374 | | 328.8803 | 15.37 | | RP | NEG | | | 9158176 | 1286271 | 9092062 | 1364667 | 7.16E-01 | -0.05 | 0.54 |
| RP_POS_371_26398a12_794 | | 371.2640 | 12.79 | | RP | POS | | | 1737633 | 1864492 | 1657386 | 1370138 | 7.16E-01 | -0.05 | 1.09 |
| RP_POS_374_36285a9_684 | | 374.3629 | 9.68 | | RP | POS | | | 2099877 | 4020493 | 1932929 | 2677037 | 7.16E-01 | -0.05 | 0.36 |
| RP_POS_420_25259a11_591 | | 420.2526 | 11.59 | | RP | POS | | | 589002 | 310768 | 573547 | 310776 | 7.16E-01 | -0.05 | 0.98 |
| RP_POS_200_07408a6_17 | N-Ethyl-o-toluenesulfonamide | 200.0741 | 6.17 | | RP | POS | 91.0546(100), 155.0167(85),119.0612(70) | 2 | 316861 | 201054 | 307805 | 163875 | 7.16E-01 | -0.05 | 0.68 |
| RP_POS_165_03117a1_792 | 5-(hydroxymethyl)pyrimidine-2,4-diol | 165.0312 | 1.79 | | RP | POS | 165.0305(100) | 2 | 3079141 | 757674 | 3116469 | 743202 | 7.16E-01 | 0.05 | 0.55 |
| RP_POS_205_0285a0_681 | | 205.0285 | 0.68 | | RP | POS | | | 493621 | 137971 | 500139 | 125839 | 7.17E-01 | 0.05 | 0.69 |
| RP_POS_478_38916a11_841 | | 478.3892 | 11.84 | | RP | POS | | | 808055 | 752520 | 842070 | 623968 | 7.17E-01 | 0.05 | 0.65 |
| RP_POS_451_96664a15_403 | | 451.9666 | 15.40 | | RP | POS | | | 1853868 | 607569 | 1823478 | 617707 | 7.17E-01 | -0.05 | 0.73 |
| RP_POS_701_50519a9_673 | | 701.5052 | 9.67 | | RP | POS | | | 678018 | 375014 | 658255 | 418397 | 7.17E-01 | -0.05 | 0.99 |
| RP_POS_790_5351a13_833 | | 790.5351 | 13.83 | | RP | POS | | | 1541113 | 1118610 | 1490634 | 927894 | 7.17E-01 | -0.05 | 0.82 |
| HILIC_POS_188_071a4_108 | Atrazine-desethyl | 188.0710 | 4.11 | | HILIC | POS | 146.0604(100) | 2 | 4.1E+07 | 7713070 | 4.2E+07 | 7602366 | 7.17E-01 | 0.05 | 0.68 |
| RP_NEG_326_08826a1_849 | | 326.0883 | 1.85 | | RP | NEG | 150.0597(100), 113.0244(95) | | 2508559 | 3643519 | 2750256 | 5730769 | 7.18E-01 | 0.05 | 0.6 |
| RP_NEG_757_13544a9_277 | | 757.1354 | 9.28 | | RP | NEG | | | 654517 | 56361.5 | 657093 | 48175.8 | 7.18E-01 | 0.05 | 0.83 |
| RP_POS_398_31122a8_327 | | 398.3112 | 8.33 | | RP | POS | | | 543239 | 438052 | 561673 | 309185 | 7.18E-01 | 0.05 | 0.38 |
| HILIC_POS_242_01894a5_123 | | 242.0189 | 5.12 | | HILIC | POS | | | 242694 | 69136.6 | 239367 | 65764.8 | 7.18E-01 | -0.05 | 0.51 |
| RP_NEG_298_99332a7_797 | | 298.9933 | 7.80 | | RP | NEG | | | 983946 | 183796 | 975504 | 158684 | 7.18E-01 | -0.05 | 0.42 |
| RP_NEG_380_97751a8_594 | | 380.9775 | 8.59 | | RP | NEG | | | 1387232 | 621787 | 1419901 | 698238 | 7.19E-01 | 0.05 | 0.36 |
| RP_NEG_395_24219a11_23 | | 395.2422 | 11.23 | | RP | NEG | | | 22326.3 | 93958.5 | 26863.3 | 90835.6 | 7.19E-01 | 0.05 | 0.31 |
| RP_POS_796_52417a13_296 | | 796.5242 | 13.30 | | RP | POS | | | 1.2E+07 | 4029408 | 1.2E+07 | 3781628 | 7.19E-01 | 0.05 | 0.59 |
| HILIC_NEG_243_89915a0_898 | | 243.8992 | 0.90 | | HILIC | NEG | | | 2.9E+07 | 6491258 | 2.9E+07 | 7299242 | 7.19E-01 | -0.05 | 0.51 |
| HILIC_NEG_332_82718a5_352 | | 332.8272 | 5.35 | | HILIC | NEG | | | 320550 | 66605.6 | 317409 | 61261.6 | 7.19E-01 | -0.05 | 0.59 |
| RP_NEG_215_86879a0_636 | | 215.8688 | 0.64 | | RP | NEG | | | 370492 | 174464 | 363256 | 119290 | 7.19E-01 | -0.05 | 0.31 |
| RP_POS_823_14301a9_346 | | 823.1430 | 9.35 | | RP | POS | | | 796844 | 107596 | 792287 | 77939.5 | 7.19E-01 | -0.05 | 0.76 |
| RP_POS_195_13782a8_047 | | 195.1378 | 8.05 | | RP | POS | | | 1400674 | 216772 | 1410974 | 204794 | 7.20E-01 | 0.05 | 0.53 |
| RP_POS_765_98761a10_498 | | 765.9876 | 10.50 | | RP | POS | | | 6147506 | 1367990 | 6209281 | 1161790 | 7.20E-01 | 0.05 | 0.32 |
| RP_POS_217_1586a10_065 | | 217.1586 | 10.07 | | RP | POS | 217.1588(100) | | 3989577 | 235522 | 3977941 | 238501 | 7.20E-01 | -0.05 | 0.94 |
| HILIC_POS_371_24017a4_501 | | 371.2402 | 4.50 | | HILIC | POS | | | 4847811 | 2614354 | 4725546 | 2384378 | 7.20E-01 | -0.05 | 0.36 |
| RP_POS_412_86356a15_367 | | 412.8636 | 15.37 | | RP | POS | | | 339314 | 149607 | 332756 | 118771 | 7.20E-01 | -0.05 | 0.68 |
| HILIC_POS_478_29263a1_638 | | 478.2926 | 1.64 | | HILIC | POS | | | 156988 | 73407 | 161340 | 100503 | 7.21E-01 | 0.05 | 0.61 |
| RP_NEG_598_29077a10_359 | | 598.2908 | 10.36 | | RP | NEG | | | 532845 | 56042.5 | 535851 | 65965.2 | 7.21E-01 | 0.05 | 0.82 |
| RP_POS_500_68964a10_053 | | 500.6896 | 10.05 | | RP | POS | | | 769497 | 398015 | 789565 | 421777 | 7.21E-01 | 0.05 | 0.46 |
| HILIC_POS_144_10187a4_1 | | 144.1019 | 4.10 | | HILIC | POS | 144.1026(100), 98.0969(30) | | 7118137 | 6205928 | 6854336 | 4599287 | 7.21E-01 | -0.05 | 0.55 |
| RP_NEG_287_12314a7_691 | | 287.1231 | 7.69 | | RP | NEG | | | 310202 | 187003 | 299319 | 249749 | 7.21E-01 | -0.05 | 0.47 |
| RP_POS_341_23221a7_931 | | 341.2322 | 7.93 | | RP | POS | | | 394102 | 112145 | 388025 | 134160 | 7.21E-01 | -0.05 | 0.63 |
| RP_POS_401_26395a11_591 | | 401.2640 | 11.59 | | RP | POS | | | 103138 | 140430 | 96660.9 | 125677 | 7.21E-01 | -0.05 | 0.45 |
| RP_POS_584_43457a10_643 | | 584.4346 | 10.64 | | RP | POS | | | 614862 | 140107 | 608294 | 129469 | 7.21E-01 | -0.05 | 0.49 |
| RP_POS_636_77948a10_483 | | 636.7795 | 10.48 | | RP | POS | | | 235030 | 134459 | 228348 | 138991 | 7.21E-01 | -0.05 | 0.98 |
| RP_NEG_431_31662a9_306 | | 431.3166 | 9.31 | | RP | NEG | | | 353492 | 114371 | 359548 | 132050 | 7.22E-01 | 0.05 | 0.85 |
| RP_POS_478_29297a10_304 | | 478.2930 | 10.30 | | RP | POS | | | 6.7E+07 | 2.4E+07 | 6.8E+07 | 3E+07 | 7.22E-01 | 0.05 | 0.67 |
| RP_POS_700_57233a12_534 | | 700.5723 | 12.53 | | RP | POS | | | 393725 | 212598 | 403841 | 204237 | 7.22E-01 | 0.05 | 0.86 |
| RP_POS_768_53223a13_704 | | 768.5322 | 13.70 | | RP | POS | | | 1756531 | 881917 | 1799142 | 872792 | 7.22E-01 | 0.05 | 0.89 |
| HILIC_NEG_308_99866a1_361 | | 308.9987 | 1.36 | | HILIC | NEG | | | 1335724 | 404296 | 1316418 | 391342 | 7.22E-01 | -0.05 | 0.37 |
| RP_NEG_262_99289a15_465 | | 262.9929 | 15.47 | | RP | NEG | | | 1132165 | 108580 | 1126696 | 115236 | 7.22E-01 | -0.05 | 0.43 |
| RP_POS_236_07158a13_89 | | 236.0716 | 13.89 | | RP | POS | | | 1.3E+08 | 5.3E+07 | 1.3E+08 | 5.4E+07 | 7.22E-01 | -0.05 | 0.82 |
| RP_POS_575_49152a15_406 | | 575.4915 | 15.41 | | RP | POS | | | 454926 | 161087 | 447262 | 154374 | 7.22E-01 | -0.05 | 0.79 |
| RP_POS_599_75726a11_358 | | 599.7573 | 11.36 | | RP | POS | | | 129153 | 69923.5 | 125980 | 61147.3 | 7.22E-01 | -0.05 | 0.47 |
| HILIC_POS_227_11337a6_774 | L-Carnosine | 227.1134 | 6.77 | | HILIC | POS | 110.0715(100), 227.114(20) | 1 | 425199 | 342518 | 410030 | 285306 | 7.23E-01 | -0.05 | 0.98 |
| RP_NEG_351_9975a15_521 | | 351.9975 | 15.52 | | RP | NEG | | | 582059 | 85838.4 | 586284 | 88346.7 | 7.23E-01 | 0.05 | 0.56 |
| RP_POS_602_42993a9_953 | | 602.4299 | 9.95 | | RP | POS | | | 7080827 | 2978308 | 7223094 | 2899310 | 7.23E-01 | 0.05 | 0.75 |
| RP_POS_790_53766a13_062 | | 790.5377 | 13.06 | | RP | POS | | | 3074861 | 1920379 | 2983207 | 1870721 | 7.23E-01 | -0.05 | 0.84 |
| RP_POS_455_71564a7_631 | | 455.7156 | 7.63 | | RP | POS | | | 261726 | 78887.6 | 265750 | 86874.6 | 7.24E-01 | 0.05 | 1.03 |
| RP_POS_766_57428a13_435 | | 766.5743 | 13.44 | | RP | POS | | | 5.4E+07 | 1.2E+07 | 5.5E+07 | 1.3E+07 | 7.24E-01 | 0.05 | 0.61 |
| RP_NEG_130_94434a10_402 | | 130.9443 | 10.40 | | RP | NEG | | | 2823140 | 1152290 | 2769764 | 1060737 | 7.24E-01 | -0.05 | 0.29 |
| RP_NEG_474_26328a10_016 | | 474.2633 | 10.02 | | RP | NEG | | | 1699478 | 1068243 | 1649469 | 1005077 | 7.24E-01 | -0.05 | 0.5 |
| RP_NEG_915_60059a11_43 | | 915.6006 | 11.43 | | RP | NEG | | | 101843 | 145089 | 95520.6 | 117483 | 7.24E-01 | -0.05 | 0.62 |
| RP_POS_203_12801a7_655 | | 203.1280 | 7.66 | | RP | POS | | | 1009790 | 175966 | 1002047 | 145445 | 7.24E-01 | -0.05 | 0.7 |
| RP_POS_666_96155a10_092 | | 666.9616 | 10.09 | | RP | POS | | | 1084004 | 523449 | 1059047 | 509554 | 7.24E-01 | -0.05 | 0.46 |
| RP_POS_247_01514a15_331 | | 247.0151 | 15.33 | | RP | POS | | | 200452 | 119325 | 205947 | 109681 | 7.25E-01 | 0.05 | 1.02 |
| RP_POS_301_25247a9_245 | | 301.2525 | 9.25 | | RP | POS | | | 435241 | 356914 | 460502 | 633632 | 7.25E-01 | 0.05 | 0.6 |
| RP_POS_196_16992a15_512 | | 196.1699 | 15.51 | | RP | POS | | | 5796646 | 2252042 | 5688016 | 2257656 | 7.25E-01 | -0.05 | 0.56 |
| RP_POS_275_07977a15_377 | | 275.0798 | 15.38 | | RP | POS | | | 354957 | 190379 | 345139 | 214949 | 7.25E-01 | -0.05 | 0.76 |
| RP_POS_343_33899a11_598 | | 343.3390 | 11.60 | | RP | POS | | | 1.6E+07 | 5172030 | 1.5E+07 | 5834888 | 7.25E-01 | -0.05 | 0.64 |
| RP_POS_697_48999a10_077 | | 697.4900 | 10.08 | | RP | POS | | | 3341065 | 1691104 | 3264806 | 1493305 | 7.25E-01 | -0.05 | 0.59 |
| RP_NEG_227_90462a0_728 | | 227.9046 | 0.73 | | RP | NEG | | | 186753 | 81074.9 | 190732 | 84575.5 | 7.26E-01 | 0.05 | 0.26 |
| RP_POS_315_25235a10_634 | | 315.2524 | 10.63 | | RP | POS | | | 2439494 | 205765 | 2449208 | 200478 | 7.26E-01 | 0.05 | 0.88 |
| HILIC_NEG_117_91898a0_933 | | 117.9190 | 0.93 | | HILIC | NEG | | | 887638 | 101651 | 883192 | 84346.1 | 7.26E-01 | -0.05 | 0.57 |
| HILIC_NEG_157_86272a3_213 | | 157.8627 | 3.21 | | HILIC | NEG | | | 2733812 | 244856 | 2721925 | 250324 | 7.26E-01 | -0.05 | 0.45 |
| RP_NEG_393_17123a11_471 | | 393.1712 | 11.47 | | RP | NEG | | | 147567 | 97307.3 | 142922 | 96433.3 | 7.26E-01 | -0.05 | 0.37 |
| RP_NEG_505_54333a15_493 | | 505.5433 | 15.49 | | RP | NEG | | | 309416 | 28632.4 | 307963 | 31752.7 | 7.26E-01 | -0.05 | 0.27 |
| RP_POS_195_10194a7_649 | | 195.1019 | 7.65 | | RP | POS | | | 6370789 | 237413 | 6358459 | 273451 | 7.26E-01 | -0.05 | 0.66 |
| RP_POS_318_04773a15_385 | | 318.0477 | 15.39 | | RP | POS | | | 242739 | 120404 | 237140 | 113675 | 7.26E-01 | -0.05 | 0.42 |
| RP_POS_446_25751a12_997 | | 446.2575 | 13.00 | | RP | POS | | | 177110 | 125543 | 171087 | 125739 | 7.26E-01 | -0.05 | 0.96 |
| RP_POS_536_16589a12_249 | | 536.1659 | 12.25 | | RP | POS | | | 1629328 | 739165 | 1594039 | 732404 | 7.26E-01 | -0.05 | 1.09 |
| HILIC_NEG_115_92085a5_794 | | 115.9209 | 5.79 | | HILIC | NEG | | | 4.6E+07 | 6663759 | 4.7E+07 | 5427648 | 7.27E-01 | 0.05 | 0.44 |
| HILIC_NEG_193_81564a3_219 | | 193.8156 | 3.22 | | HILIC | NEG | | | 1413405 | 814133 | 1450125 | 729191 | 7.27E-01 | 0.05 | 0.29 |
| RP_NEG_253_145a7_694 | | 253.1450 | 7.69 | | RP | NEG | | | 8634761 | 467168 | 8657221 | 471778 | 7.27E-01 | 0.05 | 0.52 |
| RP_POS_497_3447a11_278 | | 497.3447 | 11.28 | | RP | POS | | | 1110006 | 2742470 | 1236104 | 2559222 | 7.27E-01 | 0.05 | 0.62 |
| HILIC_POS_177_07463a1_224 | | 177.0746 | 1.22 | | HILIC | POS | | | 359556 | 158509 | 351999 | 158498 | 7.27E-01 | -0.05 | 0.53 |
| RP_POS_158_02711a0_638 | | 158.0271 | 0.64 | | RP | POS | | | 6E+07 | 1.6E+07 | 5.9E+07 | 1.6E+07 | 7.27E-01 | -0.05 | 0.52 |
| RP_POS_523_71301a10_738 | | 523.7130 | 10.74 | | RP | POS | | | 157548 | 77809.1 | 153391 | 94200.4 | 7.27E-01 | -0.05 | 0.75 |
| RP_POS_783_06854a13_351 | | 783.0685 | 13.35 | | RP | POS | | | 3686630 | 3101255 | 3531300 | 3380789 | 7.27E-01 | -0.05 | 0.75 |
| RP_POS_166_08649a2_162 | Phenylalanine | 166.0865 | 2.16 | | RP | POS | 120.0809(100), 166.0871(5) | 1 | 1.1E+08 | 1.9E+07 | 1.1E+08 | 1.9E+07 | 7.27E-01 | 0.05 | 0.64 |
| HILIC_POS_203_15005a6_459 | N,N-Dimethylarginine | 203.1501 | 6.46 | | HILIC | POS | 203.1501(100), 116.0714(60) | 2 | 5352211 | 1169750 | 5398328 | 749753 | 7.27E-01 | 0.05 | 0.52 |
| HILIC_NEG_157_90083a2_279 | | 157.9008 | 2.28 | | HILIC | NEG | | | 3619834 | 1651940 | 3699107 | 1679195 | 7.28E-01 | 0.05 | 0.41 |
| HILIC_NEG_512_3006a1_351 | | 512.3006 | 1.35 | | HILIC | NEG | | | 296749 | 124122 | 302806 | 129702 | 7.28E-01 | 0.05 | 0.4 |
| RP_POS_605_42523a10_507 | | 605.4252 | 10.51 | | RP | POS | | | 388536 | 288485 | 405784 | 413993 | 7.28E-01 | 0.05 | 0.48 |
| RP_POS_615_49542a14_801 | | 615.4954 | 14.80 | | RP | POS | | | 4473014 | 2427938 | 4593704 | 2629196 | 7.28E-01 | 0.05 | 0.48 |
| HILIC_NEG_328_07852a1_444 | | 328.0785 | 1.44 | | HILIC | NEG | | | 111913 | 36556.7 | 110365 | 28840.4 | 7.28E-01 | -0.05 | 0.79 |
| RP_NEG_243_06223a1_25 | | 243.0622 | 1.25 | | RP | NEG | | | 414215 | 127576 | 408893 | 96406.9 | 7.28E-01 | -0.05 | 0.68 |
| RP_POS_154_99033a0_708 | | 154.9903 | 0.71 | | RP | POS | | | 1849311 | 527472 | 1824889 | 501388 | 7.28E-01 | -0.05 | 0.63 |
| RP_POS_215_12724a4_899 | | 215.1272 | 4.90 | | RP | POS | | | 242158 | 73305.1 | 238621 | 75356.9 | 7.28E-01 | -0.05 | 1.05 |
| RP_POS_285_24176a9_558 | | 285.2418 | 9.56 | | RP | POS | | | 6990687 | 845648 | 6951579 | 799257 | 7.28E-01 | -0.05 | 1.05 |
| RP_POS_383_28296a12_108 | | 383.2830 | 12.11 | | RP | POS | | | 199250 | 83620.8 | 195085 | 91052.9 | 7.28E-01 | -0.05 | 1.19 |
| RP_POS_690_49835a9_126 | | 690.4984 | 9.13 | | RP | POS | | | 1109141 | 649926 | 1079520 | 600862 | 7.28E-01 | -0.05 | 1.05 |
| RP_POS_730_30945a11_501 | | 730.3095 | 11.50 | | RP | POS | | | 147153 | 86020.4 | 142980 | 89294.1 | 7.28E-01 | -0.05 | 0.53 |
| RP_POS_480_34555a10_692 | LysoPC 15:1 | 480.3456 | 10.69 | | RP | POS | 104.1077(100),480.3415(80) | 1 | 3E+07 | 8208683 | 3E+07 | 7672261 | 7.28E-01 | -0.05 | 0.61 |
| HILIC_NEG_212_91104a1_308 | | 212.9110 | 1.31 | | HILIC | NEG | | | 1620460 | 358376 | 1637695 | 368102 | 7.29E-01 | 0.05 | 0.6 |
| HILIC_POS_356_27872a0_633 | | 356.2787 | 0.63 | | HILIC | POS | | | 1004608 | 505598 | 1028560 | 507203 | 7.29E-01 | 0.05 | 0.44 |
| HILIC_POS_62_06096a3_767 | | 62.0610 | 3.77 | | HILIC | POS | | | 6218184 | 1314972 | 6275403 | 1106716 | 7.29E-01 | 0.05 | 0.43 |
| RP_NEG_399_22116a8_69 | | 399.2212 | 8.69 | | RP | NEG | | | 3.9E+07 | 7.2E+07 | 4.5E+07 | 1.4E+08 | 7.29E-01 | 0.05 | 0.62 |
| RP_POS_175_02554a15_688 | | 175.0255 | 15.69 | | RP | POS | | | 2424439 | 1607048 | 2497028 | 1464009 | 7.29E-01 | 0.05 | 0.45 |
| RP_POS_219_10747a15_33 | | 219.1075 | 15.33 | | RP | POS | | | 1286757 | 313175 | 1301031 | 290852 | 7.29E-01 | 0.05 | 0.28 |
| RP_POS_301_14062a9_523 | | 301.1406 | 9.52 | | RP | POS | | | 3.3E+07 | 9070637 | 3.3E+07 | 9975325 | 7.29E-01 | 0.05 | 1.03 |
| RP_POS_582_74945a10_66 | | 582.7495 | 10.66 | | RP | POS | | | 229744 | 148055 | 237269 | 167777 | 7.29E-01 | 0.05 | 0.59 |
| HILIC_NEG_245_07822a1_55 | | 245.0782 | 1.55 | | HILIC | NEG | | | 108085 | 24414.4 | 107119 | 16193.5 | 7.29E-01 | -0.05 | 0.33 |
| HILIC_NEG_498_28992a1_386 | | 498.2899 | 1.39 | | HILIC | NEG | | | 198065 | 325201 | 185709 | 189308 | 7.29E-01 | -0.05 | 0.49 |
| HILIC_POS_235_09248a7_747 | | 235.0925 | 7.75 | | HILIC | POS | | | 547929 | 43946.5 | 545977 | 38824.2 | 7.29E-01 | -0.05 | 0.65 |
| RP_POS_149_01222a15_375 | | 149.0122 | 15.38 | | RP | POS | | | 9.1E+07 | 3.2E+07 | 9E+07 | 3E+07 | 7.29E-01 | -0.05 | 0.27 |
| RP_NEG_164_07198a2_162 | Phenylalanine | 164.0720 | 2.16 | | RP | NEG | 174.0452(100), 164.0709(50), 72.0086(35) | 1 | 5077578 | 886754 | 5119563 | 885951 | 7.29E-01 | 0.05 | 0.58 |
| RP_NEG_297_15341a9_828 | Aurapten | 297.1534 | 9.83 | | RP | NEG | 297.1524(100), 183.0123(20) | 2 | 6027541 | 2897571 | 6160503 | 2734520 | 7.29E-01 | 0.05 | 0.33 |
| RP_NEG_391_00073a0_679 | | 391.0007 | 0.68 | | RP | NEG | | | 619897 | 117537 | 625297 | 111328 | 7.30E-01 | 0.05 | 0.56 |
| RP_POS_330_26367a7_704 | | 330.2637 | 7.70 | | RP | POS | | | 1025192 | 715163 | 1057408 | 651912 | 7.30E-01 | 0.05 | 0.46 |
| RP_POS_506_31088a13_792 | | 506.3109 | 13.79 | | RP | POS | | | 366704 | 161396 | 374942 | 185474 | 7.30E-01 | 0.05 | 0.76 |
| HILIC_POS_209_15366a1_376 | | 209.1537 | 1.38 | | HILIC | POS | | | 112918 | 58572.4 | 109998 | 64544.7 | 7.30E-01 | -0.05 | 0.78 |
| RP_POS_652_66974a12_954 | | 652.6697 | 12.95 | | RP | POS | | | 181645 | 176277 | 173794 | 157111 | 7.30E-01 | -0.05 | 0.9 |
| RP_POS_468_30844a9_769 | LysoPC 14:0 (0:0/14:0) | 468.3084 | 9.77 | | RP | POS | 184.0744(100) | 1 | 9718159 | 3647978 | 9897676 | 3929293 | 7.30E-01 | 0.05 | 0.38 |
| RP_NEG_498_1803a12_655 | | 498.1803 | 12.66 | | RP | NEG | | | 325168 | 175607 | 333398 | 174240 | 7.31E-01 | 0.05 | 0.42 |
| RP_POS_156_06326a15_234 | | 156.0633 | 15.23 | | RP | POS | | | 2282690 | 1731890 | 2362335 | 1663940 | 7.31E-01 | 0.05 | 0.38 |
| RP_POS_598_41888a10_263 | | 598.4189 | 10.26 | | RP | POS | | | 283368 | 212875 | 293894 | 232136 | 7.31E-01 | 0.05 | 0.42 |
| RP_POS_657_66266a11_309 | | 657.6627 | 11.31 | | RP | POS | | | 121744 | 59871.8 | 124761 | 67483 | 7.31E-01 | 0.05 | 0.53 |
| HILIC_POS_195_12288a1_34 | | 195.1229 | 1.34 | | HILIC | POS | | | 593344 | 337241 | 579562 | 249652 | 7.31E-01 | -0.05 | 0.52 |
| HILIC_POS_387_21411a1_111 | | 387.2141 | 1.11 | | HILIC | POS | | | 334466 | 169842 | 326221 | 179889 | 7.31E-01 | -0.05 | 0.63 |
| RP_NEG_191_86032a11_593 | | 191.8603 | 11.59 | | RP | NEG | | | 255956 | 62578.3 | 252960 | 64679.6 | 7.31E-01 | -0.05 | 0.65 |
| RP_NEG_230_98634a6_49 | | 230.9863 | 6.49 | | RP | NEG | | | 5727628 | 109630 | 5722195 | 120083 | 7.31E-01 | -0.05 | 0.35 |
| RP_NEG_333_09351a0_681 | | 333.0935 | 0.68 | | RP | NEG | | | 2054230 | 658706 | 2027027 | 503277 | 7.31E-01 | -0.05 | 0.81 |
| RP_NEG_619_28986a10_775 | | 619.2899 | 10.78 | | RP | NEG | | | 599226 | 1006529 | 557578 | 769189 | 7.31E-01 | -0.05 | 0.25 |
| RP_NEG_636_26141a10_181 | | 636.2614 | 10.18 | | RP | NEG | | | 657024 | 214226 | 646716 | 224192 | 7.31E-01 | -0.05 | 0.66 |
| RP_NEG_979_13989a9_587 | | 979.1399 | 9.59 | | RP | NEG | | | 3591561 | 241466 | 3580785 | 217803 | 7.31E-01 | -0.05 | 1 |

**Supplemantary table 1**: The non-targeted metabolomics results ordered by the VIP value (p value < 0.0003 are shaded dark green, p value < 0.05 are shaded light green, d values below 0 are blue and above 0 are red)

| | | | | | | | | | Cases | | Controls | | Welch test | Cohen's | |
|---|---|---|---|---|---|---|---|---|---|---|---|---|---|---|---|
| Feature_ID | Metabolite_name | Average_Mz | Average_RT_min_ | HMDB ID | RP_HILIC | POS_NEG | Cureted_MSMS | ID_level | Mean | SD | Mean2 | SD3 | p | d | VIP |
| RP_POS_148_00195a15_542 | | 148.0020 | 15.54 | | RP | POS | | | 1009025 | 370932 | 991502 | 374290 | 7.31E-01 | -0.05 | 0.32 |
| RP_POS_163_0755a7_312 | | 163.0755 | 7.31 | | RP | POS | | | 1765835 | 916476 | 1720730 | 995646 | 7.31E-01 | -0.05 | 0.6 |
| RP_POS_570_35547a10_436 | | 570.3555 | 10.44 | | RP | POS | | | 1.2E+07 | 4027148 | 1.2E+07 | 4453906 | 7.31E-01 | -0.05 | 0.48 |
| RP_POS_618_47583a13_551 | | 618.4758 | 13.55 | | RP | POS | | | 739227 | 277223 | 724688 | 334884 | 7.31E-01 | -0.05 | 0.29 |
| RP_POS_273_10904a7_304 | Angolensin (r) | 273.1090 | 7.30 | | RP | POS | 273.1078(100) | 2 | 605405 | 70117 | 602348 | 60588.3 | 7.31E-01 | -0.05 | 0.51 |
| HILIC_POS_110_00918a7_422 | | 110.0092 | 7.42 | | HILIC | POS | | | 616950 | 145244 | 623862 | 149116 | 7.32E-01 | 0.05 | 0.44 |
| HILIC_POS_198_05264a4_581 | | 198.0526 | 4.58 | | HILIC | POS | | | 725379 | 1920605 | 903548 | 4858308 | 7.32E-01 | 0.05 | 0.96 |
| RP_POS_355_01089a15_498 | | 355.0109 | 15.50 | | RP | POS | | | 1800429 | 233787 | 1812104 | 261247 | 7.32E-01 | 0.05 | 0.66 |
| RP_POS_454_38977a9_937 | | 454.3898 | 9.94 | | RP | POS | | | 484665 | 356769 | 501926 | 377480 | 7.32E-01 | 0.05 | 0.61 |
| RP_POS_476_30688a4_862 | | 476.3069 | 4.86 | | RP | POS | | | 670500 | 161468 | 679591 | 217786 | 7.32E-01 | 0.05 | 0.67 |
| RP_NEG_679_15131a8_719 | | 679.1513 | 8.72 | | RP | NEG | | | 777494 | 40524.5 | 775620 | 39511 | 7.32E-01 | -0.05 | 0.46 |
| RP_POS_236_07132a10_243 | | 236.0713 | 10.24 | | RP | POS | | | 7.2E+07 | 1.5E+07 | 7.1E+07 | 1.7E+07 | 7.32E-01 | -0.05 | 0.46 |
| RP_POS_378_24219a9_208 | | 378.2422 | 9.21 | | RP | POS | | | 727614 | 1755985 | 658698 | 1174164 | 7.32E-01 | -0.05 | 0.57 |
| HILIC_NEG_113_9263a4_859 | | 113.9263 | 4.86 | | HILIC | NEG | | | 2899180 | 1119144 | 2949901 | 1057020 | 7.33E-01 | 0.05 | 0.55 |
| RP_POS_272_6655a10_608 | | 272.6655 | 10.61 | | RP | POS | | | 1199718 | 446262 | 1220800 | 457905 | 7.33E-01 | 0.05 | 1.09 |
| HILIC_NEG_228_98148a0_84 | | 228.9815 | 0.84 | | HILIC | NEG | | | 5588719 | 2074056 | 5498106 | 1821372 | 7.33E-01 | -0.05 | 0.81 |
| RP_NEG_284_84982a15_409 | | 284.8498 | 15.41 | | RP | NEG | | | 706286 | 259330 | 694557 | 245088 | 7.33E-01 | -0.05 | 0.67 |
| RP_NEG_691_12622a9_416 | | 691.1262 | 9.42 | | RP | NEG | | | 2370416 | 173417 | 2362798 | 153885 | 7.33E-01 | -0.05 | 0.59 |
| RP_POS_561_81805a10_928 | | 561.8181 | 10.93 | | RP | POS | | | 440883 | 199592 | 431816 | 189472 | 7.33E-01 | -0.05 | 0.59 |
| HILIC_POS_205_09698a4_109 | Tryptophan | 205.0970 | 4.11 | | HILIC | POS | 188.0708(100), 146.0602(60) | 1 | 7E+07 | 1.3E+07 | 7.1E+07 | 1.3E+07 | 7.33E-01 | 0.05 | 0.68 |
| RP_NEG_255_07928a8_254 | | 255.0793 | 8.25 | | RP | NEG | | | 454812 | 27130.2 | 456059 | 26509.6 | 7.34E-01 | 0.05 | 0.96 |
| RP_POS_628_26489a6_09 | | 628.2649 | 6.09 | | RP | POS | | | 223949 | 114368 | 229951 | 140469 | 7.34E-01 | 0.05 | 0.41 |
| HILIC_POS_160_06013a7_746 | | 160.0601 | 7.75 | | HILIC | POS | | | 595935 | 50565.9 | 593520 | 52939.4 | 7.34E-01 | -0.05 | 0.46 |
| HILIC_POS_165_05489a5_12 | | 165.0549 | 5.12 | | HILIC | POS | | | 2133959 | 753253 | 2101517 | 646866 | 7.34E-01 | -0.05 | 0.53 |
| HILIC_POS_371_23965a0_686 | | 371.2397 | 0.69 | | HILIC | POS | | | 1680748 | 452329 | 1658651 | 493921 | 7.34E-01 | -0.05 | 0.45 |
| RP_POS_389_31091a11_593 | | 389.3109 | 11.59 | | RP | POS | | | 1349067 | 340791 | 1333941 | 313166 | 7.34E-01 | -0.05 | 1.02 |
| RP_POS_533_50165a10_716 | | 533.5017 | 10.72 | | RP | POS | | | 269702 | 260790 | 258809 | 209353 | 7.34E-01 | -0.05 | 1 |
| HILIC_NEG_153_86868a7_745 | | 153.8687 | 7.75 | | HILIC | NEG | | | 2473144 | 940320 | 2515583 | 898628 | 7.35E-01 | 0.05 | 0.49 |
| HILIC_POS_406_32968a5_674 | | 406.3297 | 5.67 | | HILIC | POS | | | 42712.4 | 36778.3 | 44440.4 | 37849.3 | 7.35E-01 | 0.05 | 0.42 |
| RP_NEG_269_1311a8_772 | | 269.1311 | 8.77 | | RP | NEG | | | 499443 | 1170797 | 551647 | 1088471 | 7.35E-01 | 0.05 | 0.48 |
| RP_NEG_318_00952a2_162 | | 318.0095 | 2.16 | | RP | NEG | | | 315706 | 54907.4 | 318269 | 55804.8 | 7.35E-01 | 0.05 | 0.55 |
| RP_POS_313_16144a3_754 | | 313.1614 | 3.75 | | RP | POS | | | 577002 | 169861 | 586176 | 218682 | 7.35E-01 | 0.05 | 1.03 |
| RP_POS_448_29123a9_628 | | 448.2912 | 9.63 | | RP | POS | | | 304177 | 137320 | 310813 | 148287 | 7.35E-01 | 0.05 | 1.02 |
| RP_POS_485_36014a12_439 | | 485.3601 | 12.44 | | RP | POS | | | 1303749 | 264384 | 1315186 | 231216 | 7.35E-01 | 0.05 | 0.6 |
| RP_POS_574_38092a9_97 | | 574.3809 | 9.97 | | RP | POS | | | 195935 | 112113 | 190413 | 125406 | 7.35E-01 | -0.05 | 0.56 |
| RP_POS_568_82648a11_299 | | 568.8265 | 11.30 | | RP | POS | | | 106946 | 39309.5 | 109467 | 64880.2 | 7.36E-01 | 0.05 | 0.54 |
| RP_NEG_216_85394a15_5 | | 216.8539 | 15.50 | | RP | NEG | | | 322564 | 79811.5 | 318642 | 89296.3 | 7.36E-01 | -0.05 | 0.68 |
| RP_POS_199_1304a6_464 | | 199.1304 | 6.46 | | RP | POS | | | 1393345 | 508389 | 1370947 | 464521 | 7.36E-01 | -0.05 | 1.04 |
| RP_POS_368_22037a15_412 | | 368.2204 | 15.41 | | RP | POS | | | 1279948 | 464069 | 1258918 | 449654 | 7.36E-01 | -0.05 | 0.37 |
| RP_POS_769_55798a13_214 | | 769.5580 | 13.21 | | RP | POS | | | 2.7E+07 | 8482381 | 2.7E+07 | 8537317 | 7.36E-01 | -0.05 | 0.84 |
| HILIC_NEG_183_93454a2_272 | | 183.9345 | 2.27 | | HILIC | NEG | | | 3405458 | 508931 | 3430220 | 562371 | 7.37E-01 | 0.05 | 0.44 |
| HILIC_POS_391_06714a6_308 | | 391.0671 | 6.31 | | HILIC | POS | | | 341980 | 141995 | 349327 | 173510 | 7.37E-01 | 0.05 | 0.32 |
| RP_POS_254_93288a15_446 | | 254.9329 | 15.45 | | RP | POS | | | 481078 | 313622 | 497671 | 397512 | 7.37E-01 | 0.05 | 0.6 |
| RP_POS_458_85629a15_374 | | 458.8563 | 15.37 | | RP | POS | | | 109133 | 68760.2 | 112167 | 63516.1 | 7.37E-01 | 0.05 | 0.5 |
| HILIC_NEG_117_919a4_329 | | 117.9190 | 4.33 | | HILIC | NEG | | | 538871 | 382426 | 523318 | 296300 | 7.37E-01 | -0.05 | 0.42 |
| RP_POS_147_05013a15_335 | | 147.0501 | 15.34 | | RP | POS | | | 237124 | 109495 | 232400 | 97225.6 | 7.37E-01 | -0.05 | 0.41 |
| RP_POS_501_33267a9_805 | | 501.3327 | 9.81 | | RP | POS | | | 227575 | 162877 | 220067 | 163463 | 7.37E-01 | -0.05 | 0.7 |
| RP_POS_626_43927a10_535 | | 626.4393 | 10.54 | | RP | POS | | | 221821 | 505914 | 202444 | 336435 | 7.37E-01 | -0.05 | 0.43 |
| HILIC_POS_123_04425a5_105 | Benzoic acid | 123.0443 | 5.11 | | HILIC | POS | 95.0501(100), 123.044(40) | 2 | 426557 | 198319 | 417827 | 183130 | 7.37E-01 | -0.05 | 0.44 |
| RP_NEG_316_94797a14_337 | | 316.9480 | 14.34 | | RP | NEG | | | 4756359 | 1392424 | 4817515 | 1292377 | 7.38E-01 | 0.05 | 0.3 |
| RP_POS_282_8584a15_397 | | 282.8584 | 15.40 | | RP | POS | | | 216694 | 116818 | 221649 | 100261 | 7.38E-01 | 0.05 | 0.79 |
| RP_POS_520_86804a9_796 | | 520.8680 | 9.80 | | RP | POS | | | 2411122 | 1124269 | 2461336 | 1071148 | 7.38E-01 | 0.05 | 0.54 |
| RP_NEG_155_9518a0_683 | | 155.9518 | 0.68 | | RP | NEG | | | 505450 | 94858.9 | 501398 | 82789.9 | 7.38E-01 | -0.05 | 0.69 |
| RP_NEG_273_17075a7_462 | | 273.1708 | 7.46 | | RP | NEG | | | 265464 | 34467.2 | 263973 | 30893.5 | 7.38E-01 | -0.05 | 0.96 |
| RP_NEG_343_6803a15_496 | | 343.6803 | 15.50 | | RP | NEG | | | 594662 | 47654.8 | 592404 | 50904.7 | 7.38E-01 | -0.05 | 0.95 |
| RP_POS_188_07069a4_822 | | 188.0707 | 4.82 | | RP | POS | | | 664734 | 176582 | 657508 | 140775 | 7.38E-01 | -0.05 | 0.63 |
| RP_POS_556_44165a10_538 | | 556.4417 | 10.54 | | RP | POS | | | 3233651 | 659816 | 3206219 | 543427 | 7.38E-01 | -0.05 | 0.52 |
| HILIC_NEG_134_8947a4_063 | | 134.8947 | 4.06 | | HILIC | NEG | | | 2.1E+07 | 7300743 | 2.1E+07 | 7693627 | 7.39E-01 | 0.05 | 0.46 |
| RP_NEG_367_24753a10_678 | | 367.2475 | 10.68 | | RP | NEG | | | 211318 | 254195 | 200312 | 230822 | 7.39E-01 | -0.05 | 0.67 |
| RP_NEG_829_18011a9_538 | | 829.1801 | 9.54 | | RP | NEG | | | 8761744 | 462858 | 8741077 | 446888 | 7.39E-01 | -0.05 | 0.9 |
| RP_POS_341_1568a5_401 | | 341.1568 | 5.40 | | RP | POS | | | 292574 | 133974 | 286389 | 137698 | 7.39E-01 | -0.05 | 1.01 |
| RP_POS_457_30692a12_226 | | 457.3069 | 12.23 | | RP | POS | | | 228485 | 108937 | 223572 | 106786 | 7.39E-01 | -0.05 | 0.64 |
| RP_NEG_268_97849a0_684 | | 268.9785 | 0.68 | | RP | NEG | | | 388139 | 95859.3 | 392317 | 89037.1 | 7.40E-01 | 0.05 | 0.64 |
| RP_NEG_409_23651a10_959 | | 409.2365 | 10.96 | | RP | NEG | | | 131097 | 183412 | 138540 | 145696 | 7.40E-01 | 0.05 | 0.72 |
| RP_POS_210_10995a11_596 | | 210.1100 | 11.60 | | RP | POS | | | 2941409 | 434527 | 2959317 | 358519 | 7.40E-01 | 0.05 | 0.52 |
| RP_POS_291_19498a9_057 | | 291.1950 | 9.06 | | RP | POS | | | 3095378 | 351940 | 3111320 | 350244 | 7.40E-01 | 0.05 | 0.47 |
| RP_POS_930_23938a9_875 | | 930.2394 | 9.88 | | RP | POS | | | 1380483 | 174320 | 1388577 | 182046 | 7.40E-01 | 0.05 | 0.68 |
| RP_NEG_130_94438a12_65 | | 130.9444 | 12.65 | | RP | NEG | | | 3046214 | 1479073 | 2980286 | 1433034 | 7.40E-01 | -0.05 | 0.84 |
| RP_POS_167_07004a9_115 | | 167.0700 | 9.12 | | RP | POS | | | 1276877 | 218717 | 1267102 | 212572 | 7.40E-01 | -0.05 | 0.36 |
| RP_POS_278_24524a11_599 | | 278.2452 | 11.60 | | RP | POS | | | 464325 | 129656 | 457688 | 159207 | 7.40E-01 | -0.05 | 0.41 |
| RP_POS_285_63876a10_492 | | 285.6388 | 10.49 | | RP | POS | | | 991613 | 548084 | 967192 | 528665 | 7.40E-01 | -0.05 | 0.53 |
| RP_POS_391_17218a7_439 | | 391.1722 | 7.44 | | RP | POS | | | 238713 | 80541.7 | 235179 | 75657.8 | 7.40E-01 | -0.05 | 0.97 |
| RP_POS_604_42285a11_351 | | 604.4229 | 11.35 | | RP | POS | | | 165592 | 157636 | 159347 | 117962 | 7.40E-01 | -0.05 | 0.48 |
| RP_POS_704_51514a9_404 | | 704.5151 | 9.40 | | RP | POS | | | 2881936 | 1462118 | 2813275 | 1557172 | 7.40E-01 | -0.05 | 0.6 |
| HILIC_POS_139_05026a0_695 | | 139.0503 | 0.70 | | HILIC | POS | 121.0401(100), 139.0759(40) | | 881695 | 1319264 | 935508 | 1066663 | 7.41E-01 | 0.05 | 0.35 |
| HILIC_POS_212_99982a4_849 | | 212.9998 | 4.85 | | HILIC | POS | | | 965895 | 155182 | 972542 | 139544 | 7.41E-01 | 0.05 | 0.24 |
| RP_POS_825_54242a12_996 | | 825.5424 | 13.00 | | RP | POS | | | 1104526 | 713384 | 1147605 | 1118210 | 7.41E-01 | 0.05 | 0.73 |
| RP_POS_185_11729a9_463 | | 185.1173 | 9.46 | | RP | POS | | | 9957779 | 7570193 | 9604880 | 8014909 | 7.41E-01 | -0.05 | 0.7 |
| HILIC_NEG_165_83743a6_983 | | 165.8374 | 6.98 | | HILIC | NEG | | | 296166 | 101592 | 300937 | 109449 | 7.42E-01 | 0.05 | 0.43 |
| RP_NEG_248_93602a0_679 | | 248.9360 | 0.68 | | RP | NEG | | | 967116 | 102027 | 971875 | 108956 | 7.42E-01 | 0.05 | 0.33 |
| RP_POS_1019_60712a10_307 | | 1019.6071 | 10.31 | | RP | POS | | | 1373638 | 671530 | 1408377 | 844641 | 7.42E-01 | 0.05 | 0.66 |
| RP_POS_689_48749a11_425 | | 689.4875 | 11.43 | | RP | POS | | | 151759 | 99946.7 | 157920 | 161660 | 7.42E-01 | 0.05 | 0.64 |
| RP_NEG_336_97318a8_188 | | 336.9732 | 8.19 | | RP | NEG | | | 1229710 | 368127 | 1214383 | 316111 | 7.42E-01 | -0.04 | 0.42 |
| RP_POS_181_55092a15_417 | | 181.5509 | 15.42 | | RP | POS | | | 196357 | 83639.6 | 192681 | 80096.4 | 7.42E-01 | -0.04 | 0.63 |
| RP_POS_316_12088a12_142 | | 316.1209 | 12.14 | | RP | POS | | | 1557475 | 4293230 | 1374197 | 3886338 | 7.42E-01 | -0.04 | 0.6 |
| RP_POS_343_104a9_207 | | 343.1040 | 9.21 | | RP | POS | | | 239672 | 150264 | 233103 | 142025 | 7.42E-01 | -0.04 | 0.53 |
| RP_POS_374_18735a10_355 | | 374.1874 | 10.36 | | RP | POS | | | 197057 | 121273 | 191972 | 105221 | 7.42E-01 | -0.04 | 0.74 |
| RP_POS_482_32425a10_147 | | 482.3243 | 10.15 | | RP | POS | | | 4030757 | 1608820 | 3961476 | 1481538 | 7.42E-01 | -0.04 | 0.51 |
| RP_POS_553_72479a10_305 | | 553.7248 | 10.31 | | RP | POS | | | 754800 | 448983 | 735611 | 406957 | 7.42E-01 | -0.04 | 0.59 |
| RP_POS_235_09642a7_721 | | 235.0964 | 7.72 | | RP | POS | | | 2397876 | 177834 | 2389992 | 172250 | 7.42E-01 | -0.05 | 0.55 |
| RP_POS_271_22693a9_559 | | 271.2269 | 9.56 | | RP | POS | | | 3643762 | 421272 | 3624546 | 431630 | 7.42E-01 | -0.05 | 0.8 |
| RP_POS_537_87781a10_57 | | 537.8778 | 10.57 | | RP | POS | | | 363857 | 185085 | 372840 | 212250 | 7.43E-01 | 0.05 | 0.99 |
| HILIC_NEG_174_86081a3_156 | | 174.8608 | 3.16 | | HILIC | NEG | | | 719309 | 141379 | 725096 | 117870 | 7.43E-01 | 0.04 | 0.64 |
| RP_NEG_323_10703a6_014 | | 323.1070 | 6.01 | | RP | NEG | | | 3083320 | 1739206 | 3161696 | 1753235 | 7.43E-01 | 0.04 | 0.72 |
| RP_POS_366_92484a15_463 | | 366.9248 | 15.46 | | RP | POS | | | 293343 | 56503.4 | 295952 | 59677.3 | 7.43E-01 | 0.04 | 0.47 |
| RP_POS_435_3074a11_593 | | 435.3074 | 11.59 | | RP | POS | | | 696117 | 87819.5 | 699990 | 85111.9 | 7.43E-01 | 0.04 | 0.62 |
| HILIC_POS_199_99046a4_339 | | 199.9905 | 4.34 | | HILIC | POS | | | 2916116 | 808063 | 2882027 | 719252 | 7.43E-01 | -0.04 | 0.84 |
| RP_POS_207_15913a6_119 | | 207.1591 | 6.12 | | RP | POS | | | 1134112 | 366767 | 1118615 | 325504 | 7.43E-01 | -0.04 | 0.64 |
| RP_POS_271_09406a7_707 | | 271.0941 | 7.71 | | RP | POS | | | 7551065 | 1.2E+07 | 7006878 | 1.2E+07 | 7.43E-01 | -0.04 | 0.53 |
| RP_POS_533_71606a10_598 | | 533.7161 | 10.60 | | RP | POS | | | 345732 | 262119 | 334889 | 223560 | 7.43E-01 | -0.04 | 0.47 |
| HILIC_POS_414_18893a3_038 | | 414.1889 | 3.04 | | HILIC | POS | | | 135707 | 59418.6 | 132865 | 66598.2 | 7.43E-01 | -0.05 | 0.62 |
| RP_POS_237_10236a5_759 | Carbamazepine | 237.1024 | 5.76 | | RP | POS | 194.0955(100), 237.1032(50) | 2 | 3.2E+08 | 1.2E+08 | 3.2E+08 | 5.2E+07 | 7.43E-01 | -0.05 | 0.48 |
| RP_POS_236_07126a15_485 | | 236.0713 | 15.49 | | RP | POS | | | 1.9E+07 | 7003092 | 2E+07 | 8440573 | 7.44E-01 | 0.05 | 0.42 |
| RP_NEG_213_91524a15_538 | | 213.9152 | 15.54 | | RP | NEG | | | 1030952 | 405438 | 1047927 | 356351 | 7.44E-01 | 0.04 | 0.9 |
| RP_NEG_940_55176a13_811 | | 940.5518 | 13.81 | | RP | NEG | | | 3410788 | 1418183 | 3473888 | 1403887 | 7.44E-01 | 0.04 | 0.38 |
| RP_POS_376_15399a9_458 | | 376.1540 | 9.46 | | RP | POS | | | 482526 | 570658 | 456640 | 586943 | 7.44E-01 | -0.04 | 0.52 |
| RP_POS_604_56738a15_195 | | 604.5674 | 15.20 | | RP | POS | | | 179171 | 176277 | 171127 | 182846 | 7.44E-01 | -0.04 | 0.69 |
| RP_POS_688_51801a9_975 | | 688.5180 | 9.98 | | RP | POS | | | 2459676 | 1275315 | 2403503 | 1247148 | 7.44E-01 | -0.04 | 0.8 |
| HILIC_NEG_117_92866a3_149 | | 117.9287 | 3.15 | | HILIC | NEG | | | 917577 | 161461 | 909513 | 194680 | 7.44E-01 | -0.05 | 0.36 |
| HILIC_NEG_212_91115a4_196 | | 212.9112 | 4.20 | | HILIC | NEG | | | 2180671 | 643730 | 2213165 | 798486 | 7.45E-01 | 0.05 | 0.34 |
| RP_POS_443_69962a5_622 | | 443.6996 | 5.62 | | RP | POS | | | 429187 | 233440 | 442249 | 336463 | 7.45E-01 | 0.05 | 0.65 |
| RP_NEG_371_17014a7_49 | | 371.1701 | 7.49 | | RP | NEG | | | 438016 | 395624 | 453443 | 298228 | 7.45E-01 | 0.04 | 0.53 |
| RP_POS_288_64053a10_608 | | 288.6405 | 10.61 | | RP | POS | | | 4578780 | 1318436 | 4637487 | 1318670 | 7.45E-01 | 0.04 | 1.02 |
| RP_POS_576_46619a14_356 | | 576.4662 | 14.36 | | RP | POS | | | 2.4E+07 | 4787300 | 2.4E+07 | 4320005 | 7.45E-01 | 0.04 | 0.6 |
| HILIC_NEG_100_93346a3_11 | | 100.9335 | 3.11 | | HILIC | NEG | | | 2E+07 | 634176 | 2E+07 | 657943 | 7.45E-01 | -0.04 | 1.02 |
| HILIC_NEG_373_01041a0_591 | | 373.0104 | 0.59 | | HILIC | NEG | | | 731318 | 291374 | 719021 | 262430 | 7.45E-01 | -0.04 | 0.4 |
| HILIC_POS_160_03439a4_86 | | 160.0344 | 4.86 | | HILIC | POS | | | 1.1E+07 | 4912060 | 1.1E+07 | 4483156 | 7.45E-01 | -0.04 | 0.66 |
| RP_NEG_327_18094a10_881 | | 327.1809 | 10.88 | | RP | NEG | | | 187207 | 148947 | 181078 | 127612 | 7.45E-01 | -0.04 | 0.27 |
| RP_NEG_346_81342a15_396 | | 346.8134 | 15.40 | | RP | NEG | | | 2330345 | 506161 | 2307542 | 517731 | 7.45E-01 | -0.04 | 0.87 |
| RP_POS_627_15295a11_447 | | 627.1530 | 11.45 | | RP | POS | | | 109550 | 54320 | 112433 | 73150.1 | 7.46E-01 | 0.05 | 0.6 |
| HILIC_POS_226_95122a5_289 | | 226.9512 | 5.29 | | HILIC | POS | | | 897194 | 371188 | 914290 | 396994 | 7.46E-01 | 0.04 | 0.38 |
| RP_NEG_161_89398a0_64 | | 161.8940 | 0.64 | | RP | NEG | | | 259562 | 113281 | 264605 | 114517 | 7.46E-01 | 0.04 | 0.39 |
| RP_NEG_300_87006a15_402 | | 300.8701 | 15.40 | | RP | NEG | | | 134962 | 69436.1 | 137809 | 59422.5 | 7.46E-01 | 0.04 | 0.76 |
| RP_NEG_637_14081a9_459 | | 637.1408 | 9.46 | | RP | NEG | | | 8998673 | 447242 | 9017744 | 417390 | 7.46E-01 | 0.04 | 0.81 |
| RP_POS_337_06396a0_677 | | 337.0640 | 0.68 | | RP | POS | | | 1060808 | 398087 | 1077640 | 363187 | 7.46E-01 | 0.04 | 0.37 |
| RP_POS_353_31638a9_821 | | 353.3164 | 9.82 | | RP | POS | | | 221574 | 64931.1 | 224258 | 56732.5 | 7.46E-01 | 0.04 | 0.51 |
| HILIC_POS_131_11807a3_176 | | 131.1181 | 3.18 | | HILIC | POS | | | 4108477 | 1211274 | 4056694 | 1128501 | 7.46E-01 | -0.04 | 0.31 |
| RP_POS_233_15314a8_55 | | 233.1531 | 8.55 | | RP | POS | | | 3681973 | 171747 | 3673822 | 193157 | 7.46E-01 | -0.04 | 0.72 |
| RP_POS_700_28772a10_949 | | 700.2877 | 10.95 | | RP | POS | | | 925711 | 348497 | 910121 | 355197 | 7.46E-01 | -0.04 | 0.51 |
| RP_POS_714_50665a12_724 | | 714.5067 | 12.72 | | RP | POS | | | 667076 | 798295 | 630343 | 854450 | 7.46E-01 | -0.04 | 0.57 |
| RP_POS_555_41144a9_445 | | 555.4114 | 9.45 | | RP | POS | | | 579492 | 311259 | 596945 | 454867 | 7.47E-01 | 0.05 | 1.01 |
| RP_POS_140_06816a0_667 | | 140.0682 | 0.67 | | RP | POS | 140.0789(100) | | 492993 | 105815 | 497197 | 85769.3 | 7.47E-01 | 0.04 | 0.59 |
| HILIC_NEG_115_92075a5_654 | | 115.9208 | 5.65 | | HILIC | NEG | | | 5.1E+07 | 8449951 | 5.1E+07 | 8548825 | 7.47E-01 | 0.04 | 0.41 |
| RP_POS_183_10164a9_58 | | 183.1016 | 9.58 | | RP | POS | | | 1347902 | 550577 | 1372269 | 552132 | 7.47E-01 | 0.04 | 0.94 |
| RP_POS_586_48718a14_341 | | 586.4872 | 14.34 | | RP | POS | | | 512117 | 490682 | 534044 | 501011 | 7.47E-01 | 0.04 | 0.69 |
| RP_POS_252_97255a0_72 | | 252.9726 | 0.72 | | RP | POS | | | 694013 | 419596 | 677418 | 334704 | 7.47E-01 | -0.04 | 0.72 |
| RP_POS_394_35226a10_644 | | 394.3523 | 10.64 | | RP | POS | | | 8839677 | 690580 | 8810817 | 622388 | 7.47E-01 | -0.04 | 0.71 |
| RP_POS_435_25116a10_951 | | 435.2512 | 10.95 | | RP | POS | | | 94359.5 | 132662 | 88789.1 | 120503 | 7.47E-01 | -0.04 | 0.88 |
| RP_NEG_227_90352a15_61 | | 227.9035 | 15.61 | | RP | NEG | | | 1218434 | 375272 | 1234528 | 359050 | 7.48E-01 | 0.04 | 0.35 |
| RP_POS_275_633a10_488 | | 275.6330 | 10.49 | | RP | POS | | | 4777752 | 1315143 | 4833343 | 1222450 | 7.48E-01 | 0.04 | 0.89 |
| HILIC_POS_166_05305a6_093 | Methioninesulfoxide | 166.0531 | 6.09 | | HILIC | POS | 74.0243(100), 102.0551(20) | 2 | 3586996 | 1829900 | 3501880 | 2017907 | 7.48E-01 | -0.04 | 0.79 |
| RP_POS_248_90018a15_386 | | 248.9002 | 15.39 | | RP | POS | | | 788110 | 391761 | 770630 | 404281 | 7.48E-01 | -0.04 | 0.59 |
| RP_POS_433_2915a11_236 | | 433.2915 | 11.24 | | RP | POS | | | 796941 | 366699 | 782494 | 294080 | 7.48E-01 | -0.04 | 0.6 |
| HILIC_POS_184_0943a4_725 | | 184.0943 | 4.73 | | HILIC | POS | | | 1382870 | 8198526 | 1895559 | 1.4E+07 | 7.49E-01 | 0.05 | 0.77 |
| RP_NEG_384_93668a11_112 | | 384.9367 | 11.11 | | RP | NEG | | | 925860 | 386923 | 944352 | 452025 | 7.49E-01 | 0.04 | 0.63 |
| RP_POS_193_1588a8_453 | | 193.1588 | 8.45 | | RP | POS | | | 2115934 | 169728 | 2123763 | 185967 | 7.49E-01 | 0.04 | 0.36 |
| RP_POS_317_30487a11_159 | | 317.3049 | 11.16 | | RP | POS | | | 402038 | 168039 | 408741 | 140033 | 7.49E-01 | 0.04 | 0.29 |
| HILIC_NEG_165_89461a6_299 | | 165.8946 | 6.30 | | HILIC | NEG | | | 1201992 | 492566 | 1179538 | 529542 | 7.49E-01 | -0.04 | 0.67 |
| RP_NEG_376_99399a0_67 | | 376.9940 | 0.67 | | RP | NEG | | | 235335 | 40015.1 | 233653 | 37087.9 | 7.49E-01 | -0.04 | 0.53 |
| RP_POS_129_12741a8_542 | | 129.1274 | 8.54 | | RP | POS | | | 988669 | 183132 | 980999 | 168591 | 7.49E-01 | -0.04 | 0.67 |
| RP_POS_477_35864a10_267 | | 477.3586 | 10.27 | | RP | POS | | | 2674410 | 1291088 | 2618847 | 1252049 | 7.49E-01 | -0.04 | 0.47 |
| RP_POS_565_42603a11_336 | | 565.4260 | 11.34 | | RP | POS | | | 3390000 | 3453937 | 3245634 | 3157502 | 7.49E-01 | -0.04 | 0.7 |
| RP_POS_569_36389a12_482 | | 569.3639 | 12.48 | | RP | POS | | | 1463788 | 632876 | 1435147 | 671714 | 7.49E-01 | -0.04 | 0.9 |
| HILIC_POS_501_05417a3_147 | | 501.0542 | 3.15 | | HILIC | POS | | | 508442 | 56204.5 | 505366 | 80383.3 | 7.49E-01 | -0.05 | 0.48 |
| HILIC_NEG_117_05592a0_732 | 2-Hydroxyvaleric acid | 117.0559 | 0.73 | | HILIC | NEG | 71.0497(100), 117.0553(90) | 2 | 6187189 | 1648825 | 6104508 | 2068110 | 7.49E-01 | -0.04 | 0.31 |
| RP_NEG_701_15424a9_527 | | 701.1542 | 9.53 | | RP | NEG | | | 1.1E+07 | 499485 | 1.1E+07 | 478897 | 7.50E-01 | 0.04 | 0.8 |
| RP_POS_645_4563a11_55 | | 645.4563 | 11.55 | | RP | POS | | | 172724 | 120630 | 177867 | 115734 | 7.50E-01 | 0.04 | 0.72 |
| RP_POS_730_7121a11_492 | | 730.7121 | 11.49 | | RP | POS | | | 149952 | 100116 | 155446 | 144068 | 7.50E-01 | 0.04 | 0.71 |
| RP_POS_807_16919a9_348 | | 807.1692 | 9.35 | | RP | POS | | | 801782 | 98008.9 | 805967 | 94403.4 | 7.50E-01 | 0.04 | 0.85 |
| RP_NEG_137_02437a5_551 | P-Hydroxybenzoic acid | 137.0244 | 5.55 | | RP | NEG | 93.0339(100), 137.0238(40) | 2 | 410587 | 249099 | 421116 | 234565 | 7.50E-01 | 0.04 | 0.93 |
| RP_NEG_234_91397a0_683 | | 234.9140 | 0.68 | | RP | NEG | | | 669732 | 93168.5 | 665962 | 80832.2 | 7.50E-01 | -0.04 | 0.73 |
| RP_POS_238_06754a9_115 | | 238.0675 | 9.12 | | RP | POS | | | 1892316 | 837573 | 1856989 | 789295 | 7.50E-01 | -0.04 | 0.27 |

**Supplemantary table 1**: The non-targeted metabolomics results ordered by the VIP value (p value < 0.0003 are shaded dark green, p value < 0.05 are shaded light green, d values below 0 are blue and above 0 are red)

| | | | | | | | | | Cases | | Controls | | Welch test | Cohen's | |
|---|---|---|---|---|---|---|---|---|---|---|---|---|---|---|---|
| Feature_ID | Metabolite_name | Average_Mz | Average_RT_min_ | HMDB ID | RP_HILIC | POS_NEG | Cureted_MSMS | ID_level | Mean | SD | Mean2 | SD3 | p | d | VIP |
| RP_NEG_328_75229a15_453 | | 328.7523 | 15.45 | | RP | NEG | | | 4086819 | 6258905 | 4364677 | 6541126 | 7.51E-01 | 0.04 | 0.6 |
| RP_NEG_584_22968a9_93 | | 584.2297 | 9.93 | | RP | NEG | | | 409671 | 149783 | 416546 | 165254 | 7.51E-01 | 0.04 | 0.54 |
| RP_POS_132_95854a15_509 | | 132.9585 | 15.51 | | RP | POS | | | 2.9E+07 | 5504169 | 2.9E+07 | 5599772 | 7.51E-01 | 0.04 | 0.64 |
| HILIC_NEG_113_92628a4_249 | | 113.9263 | 4.25 | | HILIC | NEG | | | 1789635 | 633110 | 1763140 | 590188 | 7.51E-01 | -0.04 | 0.47 |
| HILIC_POS_440_18951a0_928 | | 440.1895 | 0.93 | | HILIC | POS | | | 428995 | 115012 | 423962 | 116812 | 7.51E-01 | -0.04 | 0.58 |
| RP_NEG_279_01596a1_438 | | 279.0160 | 1.44 | | RP | NEG | | | 1338855 | 386221 | 1323907 | 304184 | 7.51E-01 | -0.04 | 0.39 |
| RP_NEG_957_20593a9_701 | | 957.2059 | 9.70 | | RP | NEG | | | 9885902 | 540047 | 9863630 | 487701 | 7.51E-01 | -0.04 | 0.7 |
| RP_POS_558_29407a10_313 | | 558.2941 | 10.31 | | RP | POS | | | 6022961 | 1029398 | 5977487 | 1061474 | 7.51E-01 | -0.04 | 0.59 |
| RP_POS_611_59796a11_497 | | 611.5980 | 11.50 | | RP | POS | | | 136200 | 82434.2 | 132952 | 67804.8 | 7.51E-01 | -0.04 | 0.45 |
| HILIC_POS_132_10188a4_219 | Norleucine | 132.1019 | 4.22 | | HILIC | POS | 86.0975(100), 132.1028(5) | 2 | 1.2E+08 | 5.9E+07 | 1.1E+08 | 4.7E+07 | 7.51E-01 | -0.04 | 0.43 |
| HILIC_NEG_312_83633a4_29 | | 312.8363 | 4.29 | | HILIC | NEG | | | 781753 | 92128 | 785430 | 78677.2 | 7.52E-01 | 0.04 | 0.57 |
| RP_POS_564_39813a10_154 | | 564.3981 | 10.15 | | RP | POS | | | 365128 | 213140 | 376113 | 284176 | 7.52E-01 | 0.04 | 0.67 |
| RP_NEG_704_33594a10_816 | | 704.3359 | 10.82 | | RP | NEG | | | 795980 | 177822 | 804244 | 203722 | 7.53E-01 | 0.04 | 0.66 |
| RP_POS_173_15379a8_074 | | 173.1538 | 8.07 | | RP | POS | | | 685513 | 63912.4 | 688107 | 56956.3 | 7.53E-01 | 0.04 | 0.89 |
| RP_POS_209_11784a4_59 | | 209.1178 | 4.59 | | RP | POS | | | 110796 | 74070.4 | 114006 | 74890.2 | 7.53E-01 | 0.04 | 0.54 |
| HILIC_NEG_295_73547a3_075 | | 295.7355 | 3.08 | | HILIC | NEG | | | 77239.4 | 13133 | 76676.8 | 13003.7 | 7.53E-01 | -0.04 | 0.54 |
| RP_POS_203_09155a6_515 | | 203.0916 | 6.52 | | RP | POS | | | 346226 | 68651 | 343160 | 73612 | 7.53E-01 | -0.04 | 0.7 |
| RP_NEG_160_84213a11_143 | | 160.8421 | 11.14 | | RP | NEG | | | 102059 | 28856.4 | 103260 | 27348.7 | 7.54E-01 | 0.04 | 0.55 |
| RP_NEG_648_27557a9_929 | | 648.2756 | 9.93 | | RP | NEG | | | 425836 | 147416 | 432450 | 159952 | 7.54E-01 | 0.04 | 0.45 |
| RP_POS_788_5564a13_432 | | 788.5564 | 13.43 | | RP | POS | | | 1.4E+07 | 2890936 | 1.4E+07 | 3561478 | 7.54E-01 | 0.04 | 0.56 |
| HILIC_NEG_212_91112a5_353 | | 212.9111 | 5.35 | | HILIC | NEG | | | 2702650 | 472895 | 2681913 | 494266 | 7.54E-01 | -0.04 | 0.52 |
| RP_NEG_224_99588a6_645 | | 224.9959 | 6.65 | | RP | NEG | | | 877912 | 78114.7 | 874734 | 70622.1 | 7.54E-01 | -0.04 | 0.44 |
| RP_NEG_297_73383a15_493 | | 297.7338 | 15.49 | | RP | NEG | | | 1562869 | 79802.4 | 1559715 | 68044.8 | 7.54E-01 | -0.04 | 0.79 |
| RP_POS_357_27411a12_123 | | 357.2741 | 12.12 | | RP | POS | | | 239870 | 107753 | 235446 | 98992.7 | 7.54E-01 | -0.04 | 0.54 |
| HILIC_POS_171_07611a6_138 | | 171.0761 | 6.14 | | HILIC | POS | | | 2609882 | 416848 | 2626835 | 379543 | 7.55E-01 | 0.04 | 0.89 |
| RP_POS_195_13805a6_474 | | 195.1381 | 6.47 | | RP | POS | | | 2229096 | 891598 | 2265579 | 819463 | 7.55E-01 | 0.04 | 0.64 |
| RP_POS_208_15491a6_5 | | 208.1549 | 6.50 | | RP | POS | | | 1.8E+07 | 1699252 | 1.9E+07 | 1860936 | 7.55E-01 | 0.04 | 1.02 |
| RP_NEG_299_14929a9_832 | | 299.1493 | 9.83 | | RP | NEG | | | 137817 | 102148 | 132821 | 128468 | 7.55E-01 | -0.04 | 0.49 |
| RP_POS_602_44696a9_118 | | 602.4470 | 9.12 | | RP | POS | | | 2055378 | 1363817 | 2000121 | 1233296 | 7.55E-01 | -0.04 | 0.73 |
| RP_POS_649_44775a10_48 | | 649.4478 | 10.48 | | RP | POS | | | 471932 | 243071 | 462531 | 199961 | 7.55E-01 | -0.04 | 0.47 |
| RP_POS_245_18578a4_062 | Leucylleucine | 245.1858 | 4.06 | | RP | POS | 86.0974(100)245.1866(10) | 2 | 1536336 | 969892 | 1474132 | 1763510 | 7.55E-01 | -0.05 | 0.4 |
| HILIC_NEG_112_98567a2_273 | | 112.9857 | 2.27 | | HILIC | NEG | 68.9949(100), 112.9854(4) | | 1.4E+07 | 2879388 | 1.4E+07 | 2891112 | 7.56E-01 | 0.04 | 0.55 |
| HILIC_POS_234_09685a1_39 | | 234.0969 | 1.39 | | HILIC | POS | | | 583013 | 334124 | 594216 | 184922 | 7.56E-01 | 0.04 | 0.45 |
| HILIC_POS_637_42834a1_211 | | 637.4283 | 1.21 | | HILIC | POS | | | 147605 | 64022.9 | 150258 | 60990.8 | 7.56E-01 | 0.04 | 0.55 |
| RP_NEG_1067_61084a10_287 | | 1067.6108 | 10.29 | | RP | NEG | | | 414439 | 176762 | 421863 | 173122 | 7.56E-01 | 0.04 | 0.6 |
| RP_NEG_194_08263a8_476 | | 194.0826 | 8.48 | | RP | NEG | | | 1140607 | 80411.2 | 1144694 | 108025 | 7.56E-01 | 0.04 | 0.4 |
| RP_POS_383_36639a13_666 | | 383.3664 | 13.67 | | RP | POS | | | 314443 | 298947 | 326590 | 275067 | 7.56E-01 | 0.04 | 0.8 |
| RP_POS_598_2663a10_478 | | 598.2663 | 10.48 | | RP | POS | | | 1350777 | 760395 | 1384808 | 831593 | 7.56E-01 | 0.04 | 0.45 |
| HILIC_NEG_118_99263a0_935 | | 118.9926 | 0.94 | | HILIC | NEG | | | 950922 | 95306.9 | 946981 | 90353.2 | 7.56E-01 | -0.04 | 0.51 |
| HILIC_NEG_205_83989a10_652 | | 205.8399 | 10.65 | | HILIC | NEG | | | 398048 | 417616 | 381420 | 366685 | 7.56E-01 | -0.04 | 0.24 |
| RP_NEG_830_59314a14_098 | PC 36:2 (18:0/18:2) | 830.5931 | 14.10 | | RP | NEG | 279.2341(100) | 2 | 9.7E+07 | 1.8E+07 | 9.8E+07 | 1.9E+07 | 7.56E-01 | 0.04 | 0.47 |
| HILIC_POS_633_39728a1_224 | | 633.3973 | 1.22 | | HILIC | POS | | | 1612225 | 605098 | 1586173 | 625985 | 7.57E-01 | -0.04 | 0.63 |
| RP_NEG_239_12947a8_207 | | 239.1295 | 8.21 | | RP | NEG | | | 542031 | 42432.3 | 540386 | 35489.6 | 7.57E-01 | -0.04 | 0.45 |
| RP_POS_609_42932a11_414 | | 609.4293 | 11.41 | | RP | POS | | | 152454 | 78728.6 | 148890 | 88659.5 | 7.57E-01 | -0.04 | 0.44 |
| RP_POS_641_73431a6_089 | | 641.7343 | 6.09 | | RP | POS | | | 486997 | 187703 | 478230 | 222418 | 7.57E-01 | -0.04 | 0.52 |
| RP_POS_255_05846a10_428 | | 255.0585 | 10.43 | | RP | POS | | | 1301055 | 509116 | 1324779 | 603050 | 7.58E-01 | 0.04 | 0.68 |
| RP_POS_272_90707a15_397 | | 272.9071 | 15.40 | | RP | POS | | | 1.5E+07 | 3971818 | 1.6E+07 | 3734476 | 7.58E-01 | 0.04 | 0.98 |
| RP_POS_531_47589a15_179 | | 531.4759 | 15.18 | | RP | POS | | | 5300956 | 3691206 | 5453110 | 3543650 | 7.58E-01 | 0.04 | 0.69 |
| RP_POS_537_36066a10_197 | | 537.3607 | 10.20 | | RP | POS | | | 308614 | 180541 | 316670 | 199514 | 7.58E-01 | 0.04 | 0.64 |
| HILIC_POS_103_0547a3_91 | | 103.0547 | 3.91 | | HILIC | POS | 103.0549(100) | | 441900 | 773226 | 408320 | 817655 | 7.58E-01 | -0.04 | 0.59 |
| RP_NEG_341_19632a11_426 | | 341.1963 | 11.43 | | RP | NEG | | | 57643.2 | 54178.7 | 55439.8 | 50483 | 7.58E-01 | -0.04 | 0.54 |
| RP_NEG_362_75989a15_416 | | 362.7599 | 15.42 | | RP | NEG | | | 319565 | 149824 | 313190 | 152145 | 7.58E-01 | -0.04 | 0.41 |
| RP_NEG_862_59894a13_64 | | 862.5989 | 13.64 | | RP | NEG | | | 1974328 | 2120909 | 1880551 | 2305612 | 7.58E-01 | -0.04 | 0.47 |
| RP_POS_240_15698a15_293 | | 240.1570 | 15.29 | | RP | POS | | | 2283716 | 948663 | 2241109 | 1064993 | 7.58E-01 | -0.04 | 0.82 |
| RP_POS_630_74432a6_088 | | 630.7443 | 6.09 | | RP | POS | | | 2446110 | 931618 | 2404362 | 1041653 | 7.58E-01 | -0.04 | 0.56 |
| HILIC_POS_284_18567a1_028 | | 284.1857 | 1.03 | | HILIC | POS | | | 362344 | 175494 | 370089 | 191753 | 7.59E-01 | 0.04 | 0.24 |
| RP_NEG_310_85733a15_407 | | 310.8573 | 15.41 | | RP | NEG | | | 171701 | 103687 | 176610 | 127284 | 7.59E-01 | 0.04 | 0.64 |
| RP_NEG_369_17477a8_215 | | 369.1748 | 8.22 | | RP | NEG | | | 167323 | 274699 | 178587 | 263993 | 7.59E-01 | 0.04 | 0.61 |
| RP_POS_352_3562a11_643 | | 352.3562 | 11.64 | | RP | POS | | | 636533 | 204041 | 644252 | 164819 | 7.59E-01 | 0.04 | 0.62 |
| RP_POS_513_37042a13_165 | | 513.3704 | 13.17 | | RP | POS | | | 162879 | 109603 | 167642 | 116452 | 7.59E-01 | 0.04 | 0.36 |
| HILIC_NEG_155_86572a3_098 | | 155.8657 | 3.10 | | HILIC | NEG | | | 1.1E+07 | 584193 | 1.1E+07 | 617701 | 7.59E-01 | -0.04 | 0.51 |
| RP_NEG_463_20953a10_894 | | 463.2095 | 10.89 | | RP | NEG | | | 610414 | 259953 | 599340 | 266955 | 7.59E-01 | -0.04 | 0.89 |
| RP_POS_147_04408a1_225 | | 147.0441 | 1.23 | | RP | POS | | | 595190 | 215709 | 587075 | 172061 | 7.59E-01 | -0.04 | 0.49 |
| RP_POS_179_10692a7_843 | | 179.1069 | 7.84 | | RP | POS | | | 4477496 | 333107 | 4462508 | 377432 | 7.59E-01 | -0.04 | 0.48 |
| RP_POS_189_09105a7_492 | | 189.0911 | 7.49 | | RP | POS | | | 834571 | 982698 | 793504 | 974993 | 7.59E-01 | -0.04 | 0.46 |
| RP_POS_337_27112a11_062 | | 337.2711 | 11.06 | | RP | POS | | | 1546837 | 884733 | 1513110 | 726677 | 7.59E-01 | -0.04 | 0.45 |
| HILIC_POS_165_1122a0_556 | | 165.1122 | 0.56 | | HILIC | POS | | | 1236344 | 560793 | 1259646 | 556965 | 7.60E-01 | 0.04 | 0.49 |
| HILIC_POS_352_24496a4_377 | | 352.2450 | 4.38 | | HILIC | POS | | | 253991 | 152083 | 260638 | 164833 | 7.60E-01 | 0.04 | 0.49 |
| RP_NEG_160_84178a15_492 | | 160.8418 | 15.49 | | RP | NEG | | | 5860371 | 140822 | 5866810 | 164022 | 7.60E-01 | 0.04 | 0.29 |
| RP_NEG_327_18082a11_66 | | 327.1808 | 11.66 | | RP | NEG | | | 59353.4 | 40745.8 | 61279.5 | 50111.5 | 7.60E-01 | 0.04 | 0.37 |
| RP_POS_218_01111a0_667 | | 218.0111 | 0.67 | | RP | POS | | | 306029 | 62548.7 | 308477 | 55015.5 | 7.60E-01 | 0.04 | 0.42 |
| HILIC_NEG_218_84953a5_845 | | 218.8495 | 5.85 | | HILIC | NEG | | | 189381 | 82406.7 | 186334 | 63718.3 | 7.60E-01 | -0.04 | 0.44 |
| HILIC_POS_375_32169a0_899 | | 375.3217 | 0.90 | | HILIC | POS | | | 283094 | 296104 | 271231 | 273137 | 7.60E-01 | -0.04 | 0.65 |
| RP_NEG_168_83611a15_331 | | 168.8361 | 15.33 | | RP | NEG | | | 2892476 | 375531 | 2908215 | 381032 | 7.61E-01 | 0.04 | 0.57 |
| RP_NEG_590_11951a8_981 | | 590.1195 | 8.98 | | RP | NEG | | | 503235 | 34266.1 | 501861 | 32051.9 | 7.61E-01 | -0.04 | 0.78 |
| RP_POS_1015_66998a10_461 | | 1015.6700 | 10.46 | | RP | POS | | | 3600478 | 4650950 | 3420035 | 4059288 | 7.61E-01 | -0.04 | 0.61 |
| RP_POS_684_97113a11_325 | | 684.9711 | 11.33 | | RP | POS | | | 145656 | 74868.8 | 142393 | 81127.6 | 7.61E-01 | -0.04 | 0.63 |
| RP_POS_478_32986a13_803 | | 478.3299 | 13.80 | | RP | POS | 478.3317(100) | | 1457299 | 2011280 | 1540945 | 2027349 | 7.62E-01 | 0.04 | 0.66 |
| RP_NEG_477_82883a15_393 | | 477.8288 | 15.39 | | RP | NEG | | | 484038 | 103167 | 488229 | 99780.6 | 7.62E-01 | 0.04 | 0.46 |
| RP_NEG_230_9969a3_771 | | 230.9969 | 3.77 | | RP | NEG | | | 363839 | 681715 | 331752 | 845149 | 7.62E-01 | -0.04 | 0.49 |
| RP_NEG_397_22614a10_473 | | 397.2261 | 10.47 | | RP | NEG | | | 34377.4 | 46094.8 | 32754.1 | 32239.4 | 7.62E-01 | -0.04 | 0.51 |
| RP_POS_172_16963a7_776 | | 172.1696 | 7.78 | | RP | POS | | | 5.5E+07 | 4.2E+07 | 5.3E+07 | 3.8E+07 | 7.62E-01 | -0.04 | 0.36 |
| HILIC_POS_176_103a6_381 | Citrulline | 176.1030 | 6.38 | | HILIC | POS | 159.077(100), 113.0717(60) 70.0661(50) | 1 | 7118049 | 1809422 | 7191635 | 1749461 | 7.62E-01 | 0.04 | 0.42 |
| RP_POS_772_58472a13_492 | PC 35:2 (18:2_17:0) | 772.5847 | 13.49 | | RP | POS | 184.0745(100), 772.5852(10) | 1 | 2.1E+07 | 6214449 | 2.1E+07 | 6310293 | 7.62E-01 | -0.04 | 0.41 |
| RP_POS_441_98856a10_11 | | 441.9886 | 10.11 | | RP | POS | | | 107762 | 52569.9 | 110148 | 62118.9 | 7.63E-01 | 0.04 | 0.54 |
| HILIC_POS_142_1226a1_354 | | 142.1226 | 1.35 | | HILIC | POS | | | 148519 | 79766.3 | 145490 | 67521.6 | 7.63E-01 | -0.04 | 0.33 |
| HILIC_POS_116_07089a3_086 | | 116.0709 | 3.09 | | HILIC | POS | 116.0714(100) | | 2989563 | 1271771 | 3043216 | 1333961 | 7.64E-01 | 0.04 | 0.46 |
| HILIC_NEG_236_79959a3_101 | | 236.7996 | 3.10 | | HILIC | NEG | | | 176019 | 50289.4 | 177989 | 45883.1 | 7.64E-01 | 0.04 | 0.6 |
| RP_POS_227_12798a5_043 | | 227.1280 | 5.04 | | RP | POS | | | 263342 | 149177 | 269571 | 153654 | 7.64E-01 | 0.04 | 0.67 |
| RP_POS_321_24054a11_601 | | 321.2405 | 11.60 | | RP | POS | | | 903139 | 176662 | 911338 | 217026 | 7.64E-01 | 0.04 | 0.63 |
| RP_POS_511_02811a9_888 | | 511.0281 | 9.89 | | RP | POS | | | 139292 | 89327.5 | 143154 | 97870.3 | 7.64E-01 | 0.04 | 0.27 |
| HILIC_POS_195_12259a0_611 | | 195.1226 | 0.61 | | HILIC | POS | | | 1.9E+07 | 2240655 | 1.8E+07 | 2011366 | 7.64E-01 | -0.04 | 0.52 |
| HILIC_POS_394_31543a7_033 | | 394.3154 | 7.03 | | HILIC | POS | | | 593053 | 248702 | 582657 | 256486 | 7.64E-01 | -0.04 | 0.32 |
| HILIC_POS_414_18906a2_955 | | 414.1891 | 2.96 | | HILIC | POS | | | 101046 | 43994.9 | 99116.4 | 49501.8 | 7.64E-01 | -0.04 | 0.63 |
| RP_POS_185_11507a15_202 | | 185.1151 | 15.20 | | RP | POS | | | 2848676 | 1422987 | 2793793 | 1253969 | 7.64E-01 | -0.04 | 0.39 |
| RP_POS_284_33099a9_206 | | 284.3310 | 9.21 | | RP | POS | | | 1659911 | 918415 | 1620465 | 998131 | 7.64E-01 | -0.04 | 0.42 |
| RP_POS_585_91772a10_32 | | 585.9177 | 10.32 | | RP | POS | | | 1766559 | 909482 | 1728962 | 923973 | 7.64E-01 | -0.04 | 0.53 |
| HILIC_NEG_353_99622a7_413 | | 353.9962 | 7.41 | | HILIC | NEG | | | 336138 | 21001.3 | 337003 | 21299.2 | 7.65E-01 | 0.04 | 0.6 |
| HILIC_POS_310_02225a7_419 | | 310.0223 | 7.42 | | HILIC | POS | | | 434312 | 25510.1 | 435385 | 26767.3 | 7.65E-01 | 0.04 | 0.66 |
| HILIC_POS_387_21375a0_705 | | 387.2138 | 0.71 | | HILIC | POS | | | 195969 | 72837.2 | 199103 | 80032.6 | 7.65E-01 | 0.04 | 0.92 |
| RP_NEG_615_14081a8_563 | | 615.1408 | 8.56 | | RP | NEG | | | 801997 | 39426.8 | 803496 | 34210.5 | 7.65E-01 | 0.04 | 0.48 |
| RP_NEG_810_56635a13_432 | | 810.5664 | 13.43 | | RP | NEG | | | 6601888 | 1303033 | 6663357 | 1655865 | 7.65E-01 | 0.04 | 0.56 |
| HILIC_POS_352_24634a0_937 | | 352.2463 | 0.94 | | HILIC | POS | | | 77531.4 | 43097.5 | 75845.3 | 39530.5 | 7.65E-01 | -0.04 | 0.53 |
| RP_NEG_320_88663a15_374 | | 320.8866 | 15.37 | | RP | NEG | | | 2158420 | 779071 | 2127252 | 746464 | 7.65E-01 | -0.04 | 0.45 |
| RP_NEG_947_6109a10_478 | | 947.6109 | 10.48 | | RP | NEG | | | 1570971 | 396735 | 1553734 | 440207 | 7.65E-01 | -0.04 | 0.62 |
| RP_POS_242_24771a10_415 | | 242.2477 | 10.42 | | RP | POS | | | 224158 | 167603 | 215926 | 226721 | 7.65E-01 | -0.04 | 1.02 |
| RP_POS_266_13852a7_492 | | 266.1385 | 7.49 | | RP | POS | | | 477615 | 611633 | 452788 | 605397 | 7.65E-01 | -0.04 | 0.51 |
| RP_POS_120_08105a2_162 | | 120.0811 | 2.16 | | RP | POS | 120.0808(100), 103.0549(20) | | 8260222 | 1559197 | 8322674 | 1515098 | 7.66E-01 | 0.04 | 0.64 |
| RP_NEG_193_85744a14_332 | | 193.8574 | 14.33 | | RP | NEG | | | 139559 | 88958 | 143257 | 92402 | 7.66E-01 | 0.04 | 0.36 |
| RP_NEG_292_9838a7_529 | | 292.9838 | 7.53 | | RP | NEG | | | 1515665 | 463023 | 1536150 | 533997 | 7.66E-01 | 0.04 | 0.52 |
| RP_NEG_353_20059a10_491 | | 353.2006 | 10.49 | | RP | NEG | | | 146987 | 198236 | 154749 | 184881 | 7.66E-01 | 0.04 | 0.26 |
| RP_POS_307_22437a9_84 | | 307.2244 | 9.84 | | RP | POS | | | 1807499 | 372122 | 1821736 | 328426 | 7.66E-01 | 0.04 | 0.94 |
| RP_POS_585_45038a13_647 | | 585.4504 | 13.65 | | RP | POS | | | 413941 | 347140 | 428930 | 384281 | 7.66E-01 | 0.04 | 0.69 |
| RP_POS_702_5368a10_658 | | 702.5368 | 10.66 | | RP | POS | | | 2317764 | 276436 | 2328846 | 268795 | 7.66E-01 | 0.04 | 0.82 |
| RP_NEG_563_10059a8_98 | | 563.1006 | 8.98 | | RP | NEG | | | 2801181 | 141656 | 2795590 | 134034 | 7.66E-01 | -0.04 | 0.65 |
| RP_POS_478_36703a11_335 | | 478.3670 | 11.34 | | RP | POS | | | 246236 | 144684 | 241259 | 99557.7 | 7.66E-01 | -0.04 | 0.61 |
| RP_POS_563_22821a12_617 | | 563.2282 | 12.62 | | RP | POS | | | 605653 | 937721 | 568369 | 897594 | 7.66E-01 | -0.04 | 0.54 |
| RP_POS_183_07806a5_748 | Triethylphosphate | 183.0781 | 5.75 | | RP | POS | 98.9894(100), 127.0163(40) | 2 | 576499 | 214995 | 585208 | 213099 | 7.66E-01 | 0.04 | 0.64 |
| RP_POS_153_07011a15_491 | | 153.0701 | 15.49 | | RP | POS | | | 3692151 | 957751 | 3728069 | 820624 | 7.67E-01 | 0.04 | 0.38 |
| RP_POS_239_23676a10_493 | | 239.2368 | 10.49 | | RP | POS | | | 304248 | 181471 | 311337 | 169826 | 7.67E-01 | 0.04 | 0.5 |
| RP_POS_588_50024a14_352 | | 588.5002 | 14.35 | | RP | POS | | | 2955518 | 1164546 | 3004472 | 1241407 | 7.67E-01 | 0.04 | 0.37 |
| HILIC_POS_634_87457a4_338 | | 634.8746 | 4.34 | | HILIC | POS | | | 4816746 | 281558 | 4804777 | 307631 | 7.67E-01 | -0.04 | 0.74 |
| RP_POS_306_2764a11_149 | | 306.2764 | 11.15 | | RP | POS | | | 1.5E+07 | 4071738 | 1.5E+07 | 4493292 | 7.67E-01 | -0.04 | 0.99 |
| RP_POS_369_26096a10_669 | | 369.2610 | 10.67 | | RP | POS | | | 682773 | 132134 | 677445 | 130590 | 7.67E-01 | -0.04 | 0.92 |
| RP_POS_285_13068a5_048 | | 285.1307 | 5.05 | | RP | POS | 285.1286(100) | | 156352 | 97519.5 | 160506 | 107187 | 7.68E-01 | 0.04 | 0.98 |
| RP_NEG_278_98727a7_445 | | 278.9873 | 7.45 | | RP | NEG | | | 839675 | 293573 | 851914 | 312103 | 7.68E-01 | 0.04 | 0.67 |
| RP_POS_197_11757a6_851 | | 197.1176 | 6.85 | | RP | POS | | | 1700244 | 187722 | 1707767 | 185926 | 7.68E-01 | 0.04 | 0.57 |
| RP_POS_593_59772a12_944 | | 593.5977 | 12.94 | | RP | POS | | | 4952136 | 8123902 | 5259804 | 7212741 | 7.68E-01 | 0.04 | 0.64 |
| HILIC_NEG_134_89481a0_708 | | 134.8948 | 0.71 | | HILIC | NEG | | | 2E+07 | 4576104 | 2E+07 | 5225767 | 7.68E-01 | -0.04 | 0.92 |
| HILIC_NEG_98_93906a4_258 | | 98.9391 | 4.26 | | HILIC | NEG | | | 1373763 | 421602 | 1357467 | 388720 | 7.68E-01 | -0.04 | 0.57 |
| RP_NEG_376_98566a7_532 | | 376.9857 | 7.53 | | RP | NEG | | | 2694976 | 320251 | 2707781 | 318847 | 7.69E-01 | 0.04 | 0.44 |
| RP_POS_342_01642a15_45 | | 342.0164 | 15.45 | | RP | POS | | | 623342 | 417045 | 642344 | 515406 | 7.69E-01 | 0.04 | 0.39 |
| HILIC_NEG_200_8588a1_329 | | 200.8588 | 1.33 | | HILIC | NEG | | | 374012 | 127497 | 368998 | 122408 | 7.69E-01 | -0.04 | 0.41 |
| RP_NEG_193_85739a11_594 | | 193.8574 | 11.59 | | RP | NEG | | | 218740 | 67666 | 216344 | 52207 | 7.69E-01 | -0.04 | 0.63 |
| RP_NEG_295_73642a15_493 | | 295.7364 | 15.49 | | RP | NEG | | | 1297236 | 84956.4 | 1294356 | 58284.1 | 7.69E-01 | -0.04 | 0.49 |
| RP_NEG_365_14008a9_832 | | 365.1401 | 9.83 | | RP | NEG | | | 168921 | 140118 | 163103 | 149308 | 7.69E-01 | -0.04 | 0.54 |
| HILIC_POS_86_09712a4_219 | 2-Methylpyrrolidine | 86.0971 | 4.22 | | HILIC | POS | 86.0975(100), 69.071(60) | 2 | 2.6E+07 | 1.1E+07 | 2.6E+07 | 8236466 | 7.69E-01 | -0.04 | 0.39 |
| RP_POS_282_27939a11_139 | | 282.2794 | 11.14 | | RP | POS | 282.2775(100), 69.071(60) | | 9014548 | 3775759 | 9196836 | 5144548 | 7.70E-01 | 0.04 | 0.83 |
| RP_POS_312_02838a10_489 | | 312.0284 | 10.49 | | RP | POS | | | 255855 | 159034 | 262119 | 154658 | 7.70E-01 | 0.04 | 0.4 |
| RP_NEG_617_73663a6_088 | | 617.7366 | 6.09 | | RP | NEG | | | 1.2E+07 | 3938931 | 1.2E+07 | 4648157 | 7.70E-01 | -0.04 | 0.55 |
| RP_POS_380_2551a10_029 | | 380.2551 | 10.03 | | RP | POS | | | 5397811 | 2033365 | 5316822 | 2013396 | 7.70E-01 | -0.04 | 0.59 |
| RP_POS_391_77991a13_802 | | 391.7799 | 13.80 | | RP | POS | | | 2867774 | 1785759 | 2799366 | 1639404 | 7.70E-01 | -0.04 | 0.62 |
| RP_POS_705_48712a9_746 | | 705.4871 | 9.75 | | RP | POS | | | 504708 | 267664 | 493669 | 282603 | 7.70E-01 | -0.04 | 0.38 |
| RP_POS_794_56885a12_946 | | 794.5689 | 12.95 | | RP | POS | | | 6171663 | 3227602 | 6055370 | 2612684 | 7.70E-01 | -0.04 | 0.79 |
| HILIC_POS_553_40607a5_82 | | 553.4061 | 5.82 | | HILIC | POS | | | 247317 | 194978 | 254760 | 179315 | 7.71E-01 | 0.04 | 0.42 |
| RP_POS_156_06311a9_328 | | 156.0631 | 9.33 | | RP | POS | | | 1008102 | 441805 | 1025664 | 441605 | 7.71E-01 | 0.04 | 0.57 |
| RP_POS_166_98495a15_471 | | 166.9850 | 15.47 | | RP | POS | | | 1.7E+07 | 4908895 | 1.7E+07 | 5124853 | 7.71E-01 | 0.04 | 0.7 |
| RP_POS_460_27957a10_676 | | 460.2796 | 10.68 | | RP | POS | | | 659688 | 274453 | 671136 | 299478 | 7.71E-01 | 0.04 | 0.69 |
| RP_POS_500_27515a10_305 | | 500.2752 | 10.31 | | RP | POS | | | 5946565 | 1792575 | 6027968 | 2229651 | 7.71E-01 | 0.04 | 0.64 |
| RP_POS_738_20337a9_279 | | 738.2034 | 9.28 | | RP | POS | | | 626845 | 65226.5 | 629534 | 69668 | 7.71E-01 | 0.04 | 0.96 |
| RP_NEG_306_93506a0_679 | | 306.9351 | 0.68 | | RP | NEG | | | 702866 | 134319 | 697809 | 120865 | 7.71E-01 | -0.04 | 0.78 |
| HILIC_POS_182_08136a5_12 | Tyrosine | 182.0814 | 5.12 | | HILIC | POS | 136.076(100), 165.0545(40) | 1 | 1.3E+07 | 4654364 | 1.3E+07 | 4054649 | 7.71E-01 | -0.04 | 0.53 |
| HILIC_POS_106_08675a4_544 | | 106.0868 | 4.54 | | HILIC | POS | 88.0761(100), 70.066(60) | | 1.1E+07 | 1.1E+07 | 1.2E+07 | 9372677 | 7.72E-01 | 0.04 | 1.02 |
| HILIC_NEG_297_73294a3_074 | | 297.7329 | 3.07 | | HILIC | NEG | | | 90764.8 | 14015.4 | 91316.3 | 13817.9 | 7.72E-01 | 0.04 | 0.51 |
| RP_NEG_316_94821a11_188 | | 316.9482 | 11.19 | | RP | NEG | | | 1313794 | 415583 | 1331448 | 470275 | 7.72E-01 | 0.04 | 1.15 |
| RP_NEG_358_99451a6_936 | | 358.9945 | 6.94 | | RP | NEG | | | 2437332 | 975828 | 2477750 | 1058855 | 7.72E-01 | 0.04 | 0.47 |
| HILIC_POS_112_89604a3_066 | | 112.8960 | 3.07 | | HILIC | POS | | | 406416 | 175691 | 398622 | 212826 | 7.72E-01 | -0.04 | 0.82 |
| HILIC_POS_315_17822a1_585 | | 315.1782 | 1.59 | | HILIC | POS | | | 508048 | 255545 | 498418 | 231335 | 7.72E-01 | -0.04 | 0.65 |
| HILIC_POS_59_06134a3_551 | | 59.0613 | 3.55 | | HILIC | POS | | | 7757934 | 5632018 | 7542451 | 5289507 | 7.72E-01 | -0.04 | 0.26 |
| RP_NEG_278_01859a1_438 | | 278.0186 | 1.44 | | RP | NEG | | | 518255 | 138954 | 513282 | 113017 | 7.72E-01 | -0.04 | 0.41 |
| RP_NEG_375_1319a10_684 | | 375.1319 | 10.68 | | RP | NEG | | | 448388 | 78419.4 | 445366 | 74327.3 | 7.72E-01 | -0.04 | 0.79 |
| HILIC_POS_211_13268a0_708 | | 211.1327 | 0.71 | | HILIC | POS | | | 777658 | 424575 | 797720 | 569478 | 7.73E-01 | 0.04 | 0.68 |

**Supplemantary table 1**: The non-targeted metabolomics results ordered by the VIP value (p value < 0.0003 are shaded dark green, p value < 0.05 are shaded light green, d values below 0 are blue and above 0 are red)

| | | | | | | | | | Cases | | Controls | | Welch test | Cohen's | |
|---|---|---|---|---|---|---|---|---|---|---|---|---|---|---|---|
| Feature_ID | Metabolite_name | Average_Mz | Average_RT_min_ | HMDB ID | RP_HILIC | POS_NEG | Cureted_MSMS | ID_level | Mean | SD | Mean2 | SD3 | p | d | VIP |
| RP_POS_610_30847a10_319 | | 610.3085 | 10.32 | | RP | POS | | | 1272829 | 214553 | 1281440 | 222329 | 7.73E-01 | 0.04 | 0.45 |
| RP_POS_678_47308a11_125 | | 678.4731 | 11.13 | | RP | POS | | | 304411 | 432561 | 320961 | 408259 | 7.73E-01 | 0.04 | 0.98 |
| RP_NEG_755_13885a9_582 | | 755.1389 | 9.58 | | RP | NEG | | | 2384225 | 164116 | 2377753 | 164063 | 7.73E-01 | -0.04 | 0.49 |
| RP_POS_424_77832a13_941 | | 424.7783 | 13.94 | | RP | POS | | | 2771424 | 2234576 | 2690195 | 1890143 | 7.73E-01 | -0.04 | 0.86 |
| RP_POS_625_38947a13_551 | | 625.3895 | 13.55 | | RP | POS | | | 3161392 | 1158017 | 3115131 | 1180764 | 7.73E-01 | -0.04 | 0.67 |
| RP_POS_174_56111a15_434 | | 174.5611 | 15.43 | | RP | POS | | | 250231 | 134819 | 255559 | 136890 | 7.74E-01 | 0.04 | 0.48 |
| RP_POS_364_19571a6_417 | | 364.1957 | 6.42 | | RP | POS | | | 7063906 | 482657 | 7083238 | 498491 | 7.74E-01 | 0.04 | 0.81 |
| RP_POS_383_14066a10_163 | | 383.1407 | 10.16 | | RP | POS | | | 948389 | 1705720 | 1023375 | 2060429 | 7.74E-01 | 0.04 | 0.22 |
| HILIC_NEG_353_01138a6_685 | | 353.0114 | 6.69 | | HILIC | NEG | | | 3104140 | 422594 | 3088512 | 375944 | 7.74E-01 | -0.04 | 0.56 |
| RP_NEG_829_1803a9_819 | | 829.1803 | 9.82 | | RP | NEG | | | 6691248 | 326642 | 6679434 | 277894 | 7.74E-01 | -0.04 | 0.54 |
| RP_POS_472_98953a10_08 | | 472.9895 | 10.08 | | RP | POS | | | 388977 | 208682 | 381113 | 193271 | 7.74E-01 | -0.04 | 0.55 |
| RP_POS_582_40576a11_268 | | 582.4058 | 11.27 | | RP | POS | | | 298526 | 296045 | 287098 | 287783 | 7.74E-01 | -0.04 | 0.56 |
| RP_POS_801_98712a10_312 | | 801.9871 | 10.31 | | RP | POS | | | 2261463 | 736109 | 2231295 | 792939 | 7.74E-01 | -0.04 | 0.56 |
| RP_POS_205_14392a6_994 | | 205.1439 | 6.99 | | RP | POS | | | 1094930 | 98693.5 | 1098787 | 99080.7 | 7.75E-01 | 0.04 | 0.6 |
| RP_POS_209_15398a8_999 | | 209.1540 | 9.00 | | RP | POS | | | 2249486 | 185415 | 2257156 | 204208 | 7.75E-01 | 0.04 | 0.3 |
| RP_POS_356_27875a8_224 | | 356.2788 | 8.22 | | RP | POS | | | 2324648 | 1262381 | 2372596 | 1197654 | 7.75E-01 | 0.04 | 0.43 |
| RP_POS_394_24295a7_933 | | 394.2430 | 7.93 | | RP | POS | | | 340864 | 50778.4 | 343007 | 58006.3 | 7.75E-01 | 0.04 | 0.83 |
| RP_POS_167_07051a8_496 | | 167.0705 | 8.50 | | RP | POS | | | 8860243 | 320417 | 8848219 | 297043 | 7.75E-01 | -0.04 | 0.92 |
| RP_POS_338_18085a5_277 | | 338.1809 | 5.28 | | RP | POS | | | 451023 | 50701 | 449195 | 43222.8 | 7.75E-01 | -0.04 | 0.81 |
| RP_POS_696_47406a12_019 | | 696.4741 | 12.02 | | RP | POS | | | 176977 | 111142 | 172594 | 112915 | 7.75E-01 | -0.04 | 0.41 |
| RP_POS_744_5907a14_348 | | 744.5907 | 14.35 | | RP | POS | | | 2115133 | 1476477 | 2059513 | 1378467 | 7.75E-01 | -0.04 | 0.45 |
| RP_POS_431_64908a10_28 | | 431.6491 | 10.28 | | RP | POS | | | 306503 | 487662 | 326876 | 550207 | 7.76E-01 | 0.04 | 0.61 |
| RP_POS_722_10114a11_266 | | 722.1011 | 11.27 | | RP | POS | | | 219102 | 103155 | 223353 | 114204 | 7.76E-01 | 0.04 | 0.59 |
| RP_POS_315_2269a11_368 | | 315.2269 | 11.37 | | RP | POS | 315.2279(100) | | 200834 | 248742 | 191447 | 233903 | 7.76E-01 | -0.04 | 0.58 |
| RP_POS_226_21655a10_832 | | 226.2166 | 10.83 | | RP | POS | | | 618235 | 325735 | 605997 | 303951 | 7.76E-01 | -0.04 | 0.93 |
| RP_POS_592_37793a10_15 | | 592.3779 | 10.15 | | RP | POS | | | 382959 | 204691 | 375016 | 203771 | 7.76E-01 | -0.04 | 0.51 |
| RP_NEG_129_05591a3_315 | | 129.0559 | 3.32 | | RP | NEG | | | 1E+07 | 2927612 | 1.1E+07 | 2627553 | 7.77E-01 | 0.04 | 0.74 |
| HILIC_NEG_115_92074a10_597 | | 115.9207 | 10.60 | | HILIC | NEG | | | 5E+07 | 1.9E+07 | 5E+07 | 1.8E+07 | 7.77E-01 | -0.04 | 0.72 |
| RP_POS_141_05006a15_467 | | 141.0501 | 15.47 | | RP | POS | | | 1975060 | 417315 | 1959115 | 405705 | 7.77E-01 | -0.04 | 0.47 |
| RP_POS_503_65912a10_046 | | 503.6591 | 10.05 | | RP | POS | | | 160386 | 74280 | 157389 | 80025.2 | 7.77E-01 | -0.04 | 0.53 |
| RP_POS_568_57825a14_049 | | 568.5783 | 14.05 | | RP | POS | | | 3426384 | 299180 | 3413848 | 343416 | 7.77E-01 | -0.04 | 0.94 |
| RP_POS_594_37732a10_51 | | 594.3773 | 10.51 | | RP | POS | | | 42187.9 | 38243.7 | 40796.4 | 33724.4 | 7.77E-01 | -0.04 | 0.92 |
| RP_POS_608_91974a9_76 | | 608.9197 | 9.76 | | RP | POS | | | 713270 | 395194 | 699322 | 326436 | 7.77E-01 | -0.04 | 0.95 |
| HILIC_POS_524_3714a1_168 | LysoPC 18:0 (0:0/18:0) | 524.3714 | 1.17 | | HILIC | POS | 184.0746(100), 104.1079(75) | 2 | 5E+07 | 1.6E+07 | 5.1E+07 | 1.4E+07 | 7.77E-01 | 0.04 | 0.51 |
| RP_POS_263_20056a9_275 | | 263.2006 | 9.28 | | RP | POS | 263.2022(100), 93.0704(95) | | 3452180 | 3537470 | 3616357 | 4779049 | 7.78E-01 | 0.04 | 0.59 |
| RP_POS_408_36804a10_873 | | 408.3680 | 10.87 | | RP | POS | | | 6755920 | 561003 | 6777664 | 566843 | 7.78E-01 | 0.04 | 0.81 |
| RP_POS_590_89032a9_578 | | 590.8903 | 9.58 | | RP | POS | | | 348119 | 215277 | 356860 | 236014 | 7.78E-01 | 0.04 | 0.85 |
| HILIC_NEG_145_0621a6_136 | Glutamine | 145.0621 | 6.14 | | HILIC | NEG | 115.9203(100), 127.0521(35) | 2 | 1.5E+07 | 2551993 | 1.5E+07 | 2425622 | 7.78E-01 | -0.04 | 0.81 |
| HILIC_POS_161_23116a4_811 | | 161.2312 | 4.81 | | HILIC | POS | | | 123735 | 74954.4 | 120840 | 75123.9 | 7.78E-01 | -0.04 | 0.7 |
| RP_POS_227_1273a3_646 | | 227.1273 | 3.65 | | RP | POS | | | 132971 | 87508.4 | 129979 | 68332.4 | 7.78E-01 | -0.04 | 0.82 |
| RP_POS_387_19247a8_803 | | 387.1925 | 8.80 | | RP | POS | | | 524041 | 287548 | 511842 | 339664 | 7.78E-01 | -0.04 | 0.46 |
| HILIC_POS_140_14349a0_707 | | 140.1435 | 0.71 | | HILIC | POS | | | 2639848 | 785817 | 2669795 | 776743 | 7.79E-01 | 0.04 | 0.55 |
| RP_NEG_398_21152a11_59 | | 398.2115 | 11.59 | | RP | NEG | | | 359337 | 64618.8 | 361772 | 62587 | 7.79E-01 | 0.04 | 0.58 |
| HILIC_NEG_266_91592a0_901 | | 266.9159 | 0.90 | | HILIC | NEG | | | 649348 | 97244.5 | 645228 | 115422 | 7.79E-01 | -0.04 | 0.34 |
| RP_NEG_269_10391a5_402 | | 269.1039 | 5.40 | | RP | NEG | | | 544364 | 22536.2 | 543383 | 27781.9 | 7.79E-01 | -0.04 | 0.51 |
| RP_NEG_395_21997a10_894 | | 395.2200 | 10.89 | | RP | NEG | | | 2209295 | 1042518 | 2169108 | 1051346 | 7.79E-01 | -0.04 | 0.9 |
| RP_POS_352_24548a11_237 | | 352.2455 | 11.24 | | RP | POS | | | 1389340 | 627182 | 1366931 | 542792 | 7.79E-01 | -0.04 | 0.73 |
| HILIC_POS_138_09154a0_725 | | 138.0915 | 0.73 | | HILIC | POS | | | 1848779 | 688722 | 1875614 | 715538 | 7.80E-01 | 0.04 | 0.29 |
| RP_NEG_246_98158a6_822 | | 246.9816 | 6.82 | | RP | NEG | | | 4125716 | 2280411 | 4215139 | 2392765 | 7.80E-01 | 0.04 | 0.55 |
| RP_POS_210_10992a10_369 | | 210.1099 | 10.37 | | RP | POS | | | 1191825 | 501620 | 1212223 | 559031 | 7.80E-01 | 0.04 | 0.63 |
| RP_POS_272_22226a9_556 | | 272.2223 | 9.56 | | RP | POS | | | 892326 | 70460.3 | 895071 | 72843.6 | 7.80E-01 | 0.04 | 0.74 |
| RP_POS_756_92694a11_432 | | 756.9269 | 11.43 | | RP | POS | | | 173086 | 110927 | 177683 | 127389 | 7.80E-01 | 0.04 | 0.53 |
| HILIC_POS_406_32953a0_944 | | 406.3295 | 0.94 | | HILIC | POS | | | 48169 | 34993.5 | 46910 | 31013.8 | 7.80E-01 | -0.04 | 0.65 |
| RP_POS_282_27875a12_981 | | 282.2788 | 12.98 | | RP | POS | 282.814(100), 83.0863(70) | | 2650736 | 1941009 | 2728922 | 2148746 | 7.81E-01 | 0.04 | 0.44 |
| HILIC_POS_121_03993a0_696 | | 121.0399 | 0.70 | | HILIC | POS | | | 430376 | 709550 | 454166 | 546026 | 7.81E-01 | 0.04 | 0.32 |
| RP_NEG_358_99573a7_286 | | 358.9957 | 7.29 | | RP | NEG | | | 1924208 | 526858 | 1942870 | 457869 | 7.81E-01 | 0.04 | 0.51 |
| RP_NEG_479_28345a10_448 | | 479.2835 | 10.45 | | RP | NEG | | | 981132 | 4592600 | 1144247 | 4012186 | 7.81E-01 | 0.04 | 0.47 |
| RP_NEG_172_83073a14_339 | | 172.8307 | 14.34 | | RP | NEG | | | 1135958 | 521926 | 1116964 | 481461 | 7.81E-01 | -0.04 | 0.54 |
| RP_POS_470_3681a9_154 | | 470.3681 | 9.15 | | RP | POS | | | 2130550 | 1103022 | 2088737 | 1100339 | 7.81E-01 | -0.04 | 0.75 |
| RP_NEG_234_97577a0_689 | | 234.9758 | 0.69 | | RP | NEG | | | 751606 | 222483 | 759394 | 190604 | 7.82E-01 | 0.04 | 0.62 |
| RP_NEG_331_65567a15_497 | | 331.6557 | 15.50 | | RP | NEG | | | 408181 | 38816.7 | 409811 | 46367 | 7.82E-01 | 0.04 | 0.38 |
| HILIC_NEG_333_09378a1_447 | | 333.0938 | 1.45 | | HILIC | NEG | | | 325848 | 111372 | 322086 | 88271.1 | 7.82E-01 | -0.04 | 0.82 |
| HILIC_POS_387_21378a5_918 | | 387.2138 | 5.92 | | HILIC | POS | | | 1288659 | 375572 | 1273547 | 419069 | 7.82E-01 | -0.04 | 0.58 |
| RP_NEG_729_18781a9_601 | | 729.1878 | 9.60 | | RP | NEG | | | 550816 | 32659.5 | 552023 | 31627.2 | 7.83E-01 | 0.04 | 0.56 |
| RP_POS_262_63614a9_926 | | 262.6361 | 9.93 | | RP | POS | | | 236487 | 137830 | 241797 | 143448 | 7.83E-01 | 0.04 | 0.39 |
| RP_POS_277_18356a14_339 | | 277.1836 | 14.34 | | RP | POS | | | 326490 | 204045 | 334980 | 241302 | 7.83E-01 | 0.04 | 0.66 |
| RP_POS_385_17087a14_327 | | 385.1709 | 14.33 | | RP | POS | | | 285318 | 193966 | 291591 | 139189 | 7.83E-01 | 0.04 | 0.56 |
| HILIC_NEG_174_99876a0_933 | | 174.9988 | 0.93 | | HILIC | NEG | | | 311082 | 38341.3 | 309552 | 42425.5 | 7.83E-01 | -0.04 | 0.61 |
| RP_NEG_271_22849a10_106 | | 271.2285 | 10.11 | | RP | NEG | | | 2057750 | 234368 | 2049010 | 229955 | 7.83E-01 | -0.04 | 0.95 |
| RP_NEG_423_10345a7_674 | | 423.1035 | 7.67 | | RP | NEG | | | 302625 | 22270.1 | 301791 | 21941.3 | 7.83E-01 | -0.04 | 0.65 |
| RP_NEG_452_27945a10_476 | | 452.2795 | 10.48 | | RP | NEG | | | 1.3E+07 | 3024109 | 1.3E+07 | 3407184 | 7.83E-01 | -0.04 | 0.57 |
| RP_NEG_504_79568a15_409 | | 504.7957 | 15.41 | | RP | NEG | | | 244940 | 54309.2 | 242682 | 64160.8 | 7.83E-01 | -0.04 | 0.3 |
| RP_POS_202_97079a15_425 | | 202.9708 | 15.43 | | RP | POS | | | 627533 | 305996 | 615459 | 333524 | 7.83E-01 | -0.04 | 0.8 |
| RP_POS_489_69116a11_636 | | 489.6912 | 11.64 | | RP | POS | | | 2549651 | 8355148 | 2281647 | 5847685 | 7.83E-01 | -0.04 | 0.57 |
| HILIC_POS_170_09238a6_874 | 3-Methylhistidine | 170.0924 | 6.87 | | HILIC | POS | 170.0929(100), 109.0762(40) | 2 | 9040821 | 1.6E+07 | 9569787 | 1.2E+07 | 7.83E-01 | 0.04 | 0.44 |
| RP_POS_758_56915a14_359 | PC 34:2 | 758.5692 | 14.36 | | RP | POS | 184.0746(1000) | 1 | 6504205 | 4087789 | 6366366 | 3266177 | 7.83E-01 | -0.04 | 0.35 |
| HILIC_NEG_201_12476a4_682 | | 201.1248 | 4.68 | | HILIC | NEG | | | 137184 | 185705 | 143609 | 158272 | 7.84E-01 | 0.04 | 0.37 |
| RP_NEG_353_05188a0_671 | | 353.0519 | 0.67 | | RP | NEG | | | 220580 | 50489 | 222354 | 44475.9 | 7.84E-01 | 0.04 | 0.4 |
| RP_POS_302_24536a10_547 | | 302.2454 | 10.55 | | RP | POS | | | 4637905 | 2168248 | 4751866 | 3616648 | 7.84E-01 | 0.04 | 0.88 |
| RP_POS_388_35431a11_805 | | 388.3543 | 11.81 | | RP | POS | | | 4051595 | 6358650 | 4326274 | 8065133 | 7.84E-01 | 0.04 | 0.29 |
| RP_POS_871_1817a9_588 | | 871.1817 | 9.59 | | RP | POS | | | 1184447 | 99810.8 | 1188050 | 92745.2 | 7.84E-01 | 0.04 | 0.58 |
| RP_NEG_201_14975a8_462 | | 201.1498 | 8.46 | | RP | NEG | | | 425280 | 228147 | 415442 | 288658 | 7.84E-01 | -0.04 | 0.57 |
| RP_NEG_237_11311a7_15 | | 237.1131 | 7.15 | | RP | NEG | | | 248875 | 12024.8 | 248371 | 14614.9 | 7.84E-01 | -0.04 | 0.3 |
| RP_NEG_688_26062a10_501 | | 688.2606 | 10.50 | | RP | NEG | | | 680779 | 171447 | 674787 | 149444 | 7.84E-01 | -0.04 | 0.56 |
| RP_NEG_729_18756a9_798 | | 729.1876 | 9.80 | | RP | NEG | | | 567871 | 30837.1 | 566746 | 29370.6 | 7.84E-01 | -0.04 | 0.56 |
| RP_POS_227_20036a8_302 | | 227.2004 | 8.30 | | RP | POS | | | 2127185 | 152019 | 2120986 | 175508 | 7.84E-01 | -0.04 | 0.6 |
| RP_POS_529_38007a10_461 | | 529.3801 | 10.46 | | RP | POS | | | 501760 | 218560 | 492566 | 265225 | 7.84E-01 | -0.04 | 0.49 |
| HILIC_POS_128_01936a7_421 | | 128.0194 | 7.42 | | HILIC | POS | | | 233532 | 98119.3 | 237129 | 95374.6 | 7.85E-01 | 0.04 | 0.49 |
| RP_POS_696_49969a10_543 | | 696.4997 | 10.54 | | RP | POS | | | 1560388 | 690221 | 1589399 | 842262 | 7.85E-01 | 0.04 | 0.53 |
| RP_POS_847_57819a10_356 | | 847.5782 | 10.36 | | RP | POS | | | 292746 | 127526 | 297324 | 118765 | 7.85E-01 | 0.04 | 0.56 |
| RP_POS_264_26871a10_025 | | 264.2687 | 10.03 | | RP | POS | | | 381033 | 165847 | 374975 | 159336 | 7.85E-01 | -0.04 | 0.6 |
| RP_POS_553_38989a12_488 | | 553.3899 | 12.49 | | RP | POS | | | 1.1E+07 | 3258812 | 1.1E+07 | 3554670 | 7.85E-01 | -0.04 | 0.61 |
| RP_POS_578_07764a10_543 | | 578.0776 | 10.54 | | RP | POS | | | 410749 | 244433 | 400717 | 286956 | 7.85E-01 | -0.04 | 0.55 |
| RP_POS_301_14075a11_142 | | 301.1408 | 11.14 | | RP | POS | | | 1804923 | 715980 | 1833480 | 811493 | 7.86E-01 | 0.04 | 0.89 |
| RP_POS_632_10608a10_364 | | 632.1061 | 10.36 | | RP | POS | | | 349485 | 196903 | 357289 | 221017 | 7.86E-01 | 0.04 | 0.68 |
| RP_POS_682_48352a10_25 | | 682.4835 | 10.25 | | RP | POS | | | 3496258 | 1502136 | 3557813 | 1781110 | 7.86E-01 | 0.04 | 0.9 |
| RP_NEG_186_11394a4_715 | | 186.1139 | 4.72 | | RP | NEG | | | 587473 | 140444 | 582223 | 142569 | 7.86E-01 | -0.04 | 0.81 |
| RP_NEG_428_97723a9_361 | | 428.9772 | 9.36 | | RP | NEG | | | 871589 | 66904.1 | 868605 | 90165.1 | 7.86E-01 | -0.04 | 0.27 |
| RP_POS_300_21677a5_908 | | 300.2168 | 5.91 | | RP | POS | | | 369400 | 274989 | 359152 | 277247 | 7.86E-01 | -0.04 | 0.34 |
| RP_POS_758_56873a11_592 | PC 34:2 | 758.5687 | 11.59 | | RP | POS | 184.0743(100) | 1 | 1755652 | 829833 | 1727189 | 705525 | 7.86E-01 | -0.04 | 0.32 |
| HILIC_NEG_185_96281a0_704 | | 185.9628 | 0.70 | | HILIC | NEG | | | 121244 | 56048.2 | 123411 | 60914.1 | 7.87E-01 | 0.04 | 0.96 |
| RP_POS_200_98943a2_736 | | 200.9894 | 2.74 | | RP | POS | | | 645159 | 187343 | 652067 | 186461 | 7.87E-01 | 0.04 | 0.61 |
| HILIC_NEG_144_99173a5_119 | | 144.9917 | 5.12 | | HILIC | NEG | | | 3147324 | 1494566 | 3088034 | 1686532 | 7.87E-01 | -0.04 | 0.68 |
| RP_POS_191_1797a8_881 | | 191.1797 | 8.88 | | RP | POS | | | 761005 | 374554 | 747142 | 375823 | 7.87E-01 | -0.04 | 0.58 |
| RP_POS_510_75916a4_496 | | 510.7592 | 4.50 | | RP | POS | | | 6183806 | 3681909 | 6045160 | 3815057 | 7.87E-01 | -0.04 | 1.26 |
| HILIC_POS_148_04248a5_747 | | 148.0425 | 5.75 | | HILIC | POS | | | 258475 | 193958 | 265788 | 203423 | 7.88E-01 | 0.04 | 0.82 |
| RP_NEG_414_13651a10_683 | | 414.1365 | 10.68 | | RP | NEG | | | 273211 | 37622.7 | 274658 | 40752.9 | 7.88E-01 | 0.04 | 0.41 |
| RP_POS_151_045a15_333 | | 151.0450 | 15.33 | | RP | POS | | | 3982462 | 913632 | 4017307 | 972093 | 7.88E-01 | 0.04 | 0.46 |
| RP_POS_464_21191a6_805 | | 464.2119 | 6.81 | | RP | POS | | | 1073574 | 100837 | 1077147 | 93961.3 | 7.88E-01 | 0.04 | 0.86 |
| RP_POS_497_39578a13_174 | | 497.3958 | 13.17 | | RP | POS | | | 2983602 | 863829 | 3015858 | 889118 | 7.88E-01 | 0.04 | 0.65 |
| RP_POS_595_58478a11_593 | | 595.5848 | 11.59 | | RP | POS | | | 714879 | 324791 | 726940 | 331164 | 7.88E-01 | 0.04 | 0.91 |
| RP_POS_844_52484a12_778 | | 844.5248 | 12.78 | | RP | POS | | | 1384737 | 860168 | 1415258 | 806352 | 7.88E-01 | 0.04 | 0.77 |
| RP_NEG_374_78583a15_412 | | 374.7858 | 15.41 | | RP | NEG | | | 604274 | 238065 | 595541 | 238071 | 7.88E-01 | -0.04 | 0.67 |
| RP_POS_367_31802a11_595 | | 367.3180 | 11.60 | | RP | POS | | | 822669 | 455436 | 808335 | 324601 | 7.88E-01 | -0.04 | 0.42 |
| RP_POS_261_14459a3_393 | gamma-Glutamylleucine | 261.1446 | 3.39 | | RP | POS | 86.0973(100), 132.1026(60) | 2 | 587234 | 254423 | 595309 | 185405 | 7.88E-01 | 0.04 | 0.48 |
| RP_NEG_447_13568a8_515 | | 447.1357 | 8.52 | | RP | NEG | | | 2272179 | 274423 | 2286020 | 446271 | 7.89E-01 | 0.04 | 0.78 |
| RP_POS_300_07498a15_331 | | 300.0750 | 15.33 | | RP | POS | | | 803271 | 235630 | 812772 | 279004 | 7.89E-01 | 0.04 | 0.24 |
| RP_POS_380_27917a10_518 | | 380.2792 | 10.52 | | RP | POS | | | 424045 | 295704 | 434673 | 287192 | 7.89E-01 | 0.04 | 0.8 |
| RP_POS_643_52661a11_952 | | 643.5266 | 11.95 | | RP | POS | | | 4628736 | 4827553 | 4787386 | 3850839 | 7.89E-01 | 0.04 | 0.79 |
| HILIC_NEG_200_85924a4_048 | | 200.8592 | 4.05 | | HILIC | NEG | | | 1749700 | 304330 | 1738224 | 322079 | 7.89E-01 | -0.04 | 0.63 |
| HILIC_POS_475_13361a4_157 | | 475.1336 | 4.16 | | HILIC | POS | | | 110456 | 23696.6 | 109519 | 27125.6 | 7.89E-01 | -0.04 | 0.44 |
| RP_NEG_368_97922a6_983 | | 368.9792 | 6.98 | | RP | NEG | | | 739074 | 194158 | 732227 | 180402 | 7.89E-01 | -0.04 | 0.59 |
| RP_POS_400_8685a15_393 | | 400.8685 | 15.39 | | RP | POS | | | 550129 | 265315 | 541328 | 217694 | 7.89E-01 | -0.04 | 0.36 |
| RP_POS_687_47015a10_239 | | 687.4702 | 10.24 | | RP | POS | | | 466484 | 268894 | 456041 | 298875 | 7.89E-01 | -0.04 | 0.62 |
| RP_POS_295_18781a9_891 | Cinchonidine | 295.1878 | 9.89 | | RP | POS | 295.187688100) | 2 | 3633500 | 1429485 | 3587847 | 1071863 | 7.89E-01 | -0.04 | 0.68 |
| RP_POS_279_15887a9_44 | | 279.1589 | 9.44 | | RP | POS | 149.023(100), 57.0707(18) | | 1.7E+07 | 1246393 | 1.7E+07 | 1319357 | 7.90E-01 | 0.04 | 0.71 |
| RP_NEG_661_25446a5_79 | | 661.2545 | 5.79 | | RP | NEG | | | 1880067 | 822125 | 1909188 | 782579 | 7.90E-01 | 0.04 | 0.49 |
| RP_POS_476_33676a9_733 | | 476.3368 | 9.73 | | RP | POS | | | 229921 | 123674 | 234840 | 143877 | 7.90E-01 | 0.04 | 0.87 |
| HILIC_NEG_452_27844a1_92 | | 452.2784 | 1.92 | | HILIC | NEG | | | 271723 | 76703.2 | 268776 | 84405.2 | 7.90E-01 | -0.04 | 0.62 |
| RP_NEG_323_04199a0_688 | | 323.0420 | 0.69 | | RP | NEG | | | 449119 | 83467.5 | 446197 | 77360.1 | 7.90E-01 | -0.04 | 0.59 |
| RP_POS_129_07007a7_707 | | 129.0701 | 7.71 | | RP | POS | | | 1.5E+07 | 2.3E+07 | 1.4E+07 | 2.4E+07 | 7.90E-01 | -0.04 | 0.51 |
| RP_POS_149_11745a4_073 | | 149.1175 | 4.07 | | RP | POS | | | 892861 | 598601 | 872556 | 519102 | 7.90E-01 | -0.04 | 0.42 |
| RP_POS_298_69226a10_93 | | 298.6923 | 10.93 | | RP | POS | | | 1241076 | 502684 | 1221583 | 562731 | 7.90E-01 | -0.04 | 1 |
| RP_POS_370_25644a12_353 | | 370.2564 | 12.35 | | RP | POS | | | 1109501 | 866548 | 1080518 | 734563 | 7.90E-01 | -0.04 | 1.06 |
| RP_POS_231_11589a0_875 | | 231.1159 | 0.88 | | RP | POS | | | 1.2E+07 | 6532265 | 1.2E+07 | 6941843 | 7.91E-01 | 0.04 | 0.69 |
| RP_POS_544_29749a6_16 | | 544.2975 | 6.16 | | RP | POS | | | 716509 | 75759.2 | 719151 | 70460.4 | 7.91E-01 | 0.04 | 0.69 |
| RP_NEG_320_80807a15_389 | | 320.8081 | 15.39 | | RP | NEG | | | 4780560 | 1320944 | 4731208 | 1393026 | 7.91E-01 | -0.04 | 0.65 |
| RP_POS_445_36655a12_439 | | 445.3666 | 12.44 | | RP | POS | | | 4737286 | 798069 | 4711411 | 633952 | 7.91E-01 | -0.04 | 0.4 |
| RP_POS_560_0473a10_435 | | 560.0473 | 10.44 | | RP | POS | | | 635868 | 323006 | 623893 | 335800 | 7.91E-01 | -0.04 | 0.35 |
| RP_POS_844_52411a13_008 | | 844.5241 | 13.01 | | RP | POS | | | 1E+07 | 5154024 | 1E+07 | 4425494 | 7.91E-01 | -0.04 | 0.83 |
| RP_POS_240_09959a10_358 | Securinine | 240.0996 | 10.36 | | RP | POS | 240.0996(100) | 2 | 1861891 | 367517 | 1876286 | 423377 | 7.92E-01 | 0.04 | 1.04 |
| RP_POS_1074_15466a10_306 | | 1074.1547 | 10.31 | | RP | POS | | | 451047 | 228931 | 459538 | 241352 | 7.92E-01 | 0.04 | 0.53 |
| RP_POS_280_65515a10_607 | | 280.6552 | 10.61 | | RP | POS | | | 2385023 | 1135208 | 2429045 | 1285074 | 7.92E-01 | 0.04 | 1.19 |
| RP_POS_306_93198a15_406 | | 306.9320 | 15.41 | | RP | POS | | | 2704199 | 803274 | 2734993 | 892592 | 7.92E-01 | 0.04 | 0.58 |
| RP_POS_441_65411a10_105 | | 441.6541 | 10.11 | | RP | POS | | | 176628 | 127528 | 181305 | 131546 | 7.92E-01 | 0.04 | 0.41 |
| RP_POS_605_27734a10_31 | | 605.2773 | 10.31 | | RP | POS | | | 834362 | 261066 | 825023 | 258129 | 7.92E-01 | -0.04 | 0.54 |
| HILIC_NEG_243_0621a1_452 | Uridine | 243.0621 | 1.45 | | HILIC | NEG | 110.0247(100), 200.0575(30) | 2 | 1406405 | 414628 | 1393280 | 316645 | 7.92E-01 | -0.04 | 0.75 |
| RP_NEG_301_21762a10_69 | | 301.2176 | 10.69 | | RP | NEG | 301.2161(100) | | 2473832 | 1392366 | 2522321 | 1321845 | 7.93E-01 | 0.04 | 0.92 |
| HILIC_POS_176_10312a3_019 | | 176.1031 | 3.02 | | HILIC | POS | 176.1036(100) | | 170431 | 129069 | 174931 | 122023 | 7.93E-01 | 0.04 | 1.03 |
| RP_POS_478_33026a13_214 | | 478.3303 | 13.21 | | RP | POS | 478.3311(100) | | 3037260 | 2126808 | 3116640 | 2275293 | 7.93E-01 | 0.04 | 0.76 |
| HILIC_POS_635_41412a1_22 | | 635.4141 | 1.22 | | HILIC | POS | | | 507949 | 186525 | 514472 | 178015 | 7.93E-01 | 0.04 | 0.62 |
| RP_POS_143_01424a14_351 | | 143.0142 | 14.35 | | RP | POS | | | 2986276 | 1317092 | 3028550 | 1051708 | 7.93E-01 | 0.04 | 0.6 |
| RP_POS_129_07007a7_491 | | 129.0701 | 7.49 | | RP | POS | | | 5467387 | 9895843 | 5113108 | 9877388 | 7.93E-01 | -0.04 | 0.51 |
| RP_POS_264_84726a15_443 | | 264.8473 | 15.44 | | RP | POS | | | 273727 | 153326 | 268296 | 149892 | 7.93E-01 | -0.04 | 0.94 |
| RP_POS_363_11121a9_456 | | 363.1112 | 9.46 | | RP | POS | | | 357438 | 349662 | 344038 | 391370 | 7.93E-01 | -0.04 | 0.72 |
| RP_POS_496_05084a10_483 | | 496.0508 | 10.48 | | RP | POS | | | 567563 | 437025 | 552743 | 390928 | 7.93E-01 | -0.04 | 0.53 |
| RP_POS_896_53741a12_924 | | 896.5374 | 12.92 | | RP | POS | | | 2763114 | 4022389 | 2629562 | 3446393 | 7.93E-01 | -0.04 | 0.56 |
| RP_NEG_274_10461a0_574 | | 274.1046 | 0.57 | | RP | NEG | | | 813488 | 181149 | 820019 | 184436 | 7.94E-01 | 0.04 | 0.36 |
| RP_NEG_765_16577a9_582 | | 765.1658 | 9.58 | | RP | NEG | | | 1.1E+07 | 536209 | 1.1E+07 | 517131 | 7.94E-01 | 0.04 | 0.68 |
| RP_POS_159_11679a6_052 | | 159.1168 | 6.05 | | RP | POS | | | 649698 | 169575 | 655972 | 180462 | 7.94E-01 | 0.04 | 0.55 |
| RP_POS_338_34128a10_667 | | 338.3413 | 10.67 | | RP | POS | | | 983265 | 292008 | 994210 | 318172 | 7.94E-01 | 0.04 | 0.43 |
| RP_POS_382_23956a12_356 | | 382.2396 | 12.36 | | RP | POS | | | 120487 | 78101.1 | 123523 | 90486 | 7.94E-01 | 0.04 | 0.68 |

**Supplemantary table 1**: The non-targeted metabolomics results ordered by the VIP value (p value < 0.0003 are shaded dark green, p value < 0.05 are shaded light green, d values below 0 are blue and above 0 are red)

| | | | | | | | | | Cases | | Controls | | Welch test | Cohen's | |
|---|---|---|---|---|---|---|---|---|---|---|---|---|---|---|---|
| Feature_ID | Metabolite_name | Average_Mz | Average_RT_min_ | HMDB ID | RP_HILIC | POS_NEG | Cureted_MSMS | ID_level | Mean | SD | Mean2 | SD3 | p | d | VIP |
| RP_POS_607_0929a10_516 | | 607.0929 | 10.52 | | RP | POS | | | 331335 | 240492 | 339626 | 224249 | 7.94E-01 | 0.04 | 0.3 |
| RP_POS_271_11511a4_254 | | 271.1151 | 4.25 | | RP | POS | | | 213681 | 468729 | 198208 | 402267 | 7.94E-01 | -0.04 | 0.41 |
| RP_POS_790_57166a13_673 | | 790.5717 | 13.67 | | RP | POS | | | 1.4E+07 | 5843467 | 1.4E+07 | 5781676 | 7.94E-01 | -0.04 | 0.61 |
| HILIC_NEG_121_90455a6_296 | | 121.9046 | 6.30 | | HILIC | NEG | | | 554692 | 272925 | 564945 | 302365 | 7.95E-01 | 0.04 | 0.36 |
| HILIC_NEG_285_70825a1_409 | | 285.7083 | 1.41 | | HILIC | NEG | | | 617197 | 101759 | 620597 | 90016.4 | 7.95E-01 | 0.04 | 0.35 |
| RP_POS_164_98453a15_365 | | 164.9845 | 15.37 | | RP | POS | | | 2.3E+07 | 1.3E+07 | 2.3E+07 | 1.1E+07 | 7.95E-01 | 0.04 | 0.35 |
| RP_POS_507_01419a10_204 | | 507.0142 | 10.20 | | RP | POS | | | 684271 | 364907 | 697616 | 385678 | 7.95E-01 | 0.04 | 0.47 |
| RP_POS_534_05298a10_598 | | 534.0530 | 10.60 | | RP | POS | | | 300346 | 228738 | 293115 | 180066 | 7.95E-01 | -0.04 | 0.5 |
| RP_POS_644_26245a10_613 | | 644.2625 | 10.61 | | RP | POS | | | 2207239 | 448745 | 2190854 | 469723 | 7.95E-01 | -0.04 | 0.41 |
| RP_POS_327_07761a9_036 | Triphenylphosphate | 327.0776 | 9.04 | | RP | POS | 327.0776(100), 233.0356(25) | 2 | 1.5E+07 | 2125496 | 1.5E+07 | 1438618 | 7.95E-01 | 0.04 | 0.46 |
| RP_POS_287_23648a10_901 | | 287.2365 | 10.90 | | RP | POS | 287.236(100), 81.0705(45) | | 1055993 | 534333 | 1072947 | 429416 | 7.96E-01 | 0.04 | 0.41 |
| RP_NEG_369_22879a7_817 | | 369.2288 | 7.82 | | RP | NEG | | | 488412 | 549737 | 511990 | 754494 | 7.96E-01 | 0.04 | 0.48 |
| RP_NEG_423_23972a8_842 | | 423.2397 | 8.84 | | RP | NEG | | | 207287 | 17116.1 | 207953 | 20143.2 | 7.96E-01 | 0.04 | 0.47 |
| RP_POS_173_96158a0_6 | | 173.9616 | 0.60 | | RP | POS | | | 892710 | 218000 | 900476 | 221224 | 7.96E-01 | 0.04 | 0.4 |
| RP_POS_828_55109a11_593 | | 828.5511 | 11.59 | | RP | POS | | | 848461 | 275060 | 857470 | 237181 | 7.96E-01 | 0.04 | 0.39 |
| RP_POS_174_99231a8_091 | | 174.9923 | 8.09 | | RP | POS | | | 204747 | 99994.3 | 201386 | 90970.7 | 7.96E-01 | -0.04 | 0.48 |
| RP_POS_576_53467a15_178 | | 576.5347 | 15.18 | | RP | POS | | | 5400141 | 1498419 | 5349274 | 1389613 | 7.96E-01 | -0.04 | 0.4 |
| RP_POS_577_13251a9_644 | | 577.1325 | 9.64 | | RP | POS | | | 1676838 | 230912 | 1669371 | 192719 | 7.96E-01 | -0.04 | 1.14 |
| RP_POS_449_39627a14_597 | | 449.3963 | 14.60 | | RP | POS | | | 310324 | 204666 | 319879 | 317062 | 7.97E-01 | 0.04 | 0.47 |
| HILIC_POS_446_05383a3_147 | | 446.0538 | 3.15 | | HILIC | POS | | | 1101510 | 312200 | 1111796 | 276046 | 7.97E-01 | 0.03 | 0.74 |
| RP_POS_425_44559a11_897 | | 425.4456 | 11.90 | | RP | POS | | | 4103214 | 2216650 | 4184989 | 2424217 | 7.98E-01 | 0.04 | 0.59 |
| RP_POS_504_34406a10_61 | | 504.3441 | 10.61 | | RP | POS | | | 891414 | 333721 | 902837 | 318733 | 7.98E-01 | 0.04 | 0.52 |
| RP_POS_526_43201a10_652 | | 526.4320 | 10.65 | | RP | POS | | | 1.4E+07 | 870325 | 1.4E+07 | 987070 | 7.98E-01 | 0.04 | 0.88 |
| RP_POS_695_11664a9_415 | | 695.1166 | 9.42 | | RP | POS | | | 468754 | 54340 | 470618 | 52119.9 | 7.98E-01 | 0.04 | 0.49 |
| HILIC_NEG_202_92909a3_163 | | 202.9291 | 3.16 | | HILIC | NEG | | | 4411505 | 482191 | 4428197 | 473123 | 7.98E-01 | 0.03 | 0.39 |
| HILIC_NEG_271_07086a4_097 | | 271.0709 | 4.10 | | HILIC | NEG | | | 116709 | 24229.9 | 117532 | 22939.1 | 7.98E-01 | 0.03 | 0.57 |
| RP_POS_172_97716a0_911 | | 172.9772 | 0.91 | | RP | POS | | | 2.4E+07 | 1.2E+07 | 2.3E+07 | 1.2E+07 | 7.98E-01 | -0.03 | 0.23 |
| RP_POS_475_41565a14_532 | | 475.4157 | 14.53 | | RP | POS | | | 806114 | 474685 | 790426 | 423081 | 7.98E-01 | -0.03 | 0.35 |
| RP_POS_209_09229a1_915 | | 209.0923 | 1.92 | | RP | POS | 94.0654(100), 146.0601(70) | | 1853453 | 358667 | 1840610 | 372735 | 7.98E-01 | -0.04 | 0.47 |
| RP_NEG_199_80519a15_493 | | 199.8052 | 15.49 | | RP | NEG | | | 2.8E+07 | 543317 | 2.8E+07 | 580072 | 7.98E-01 | -0.04 | 0.35 |
| RP_POS_764_55859a12_986 | | 764.5586 | 12.99 | | RP | POS | | | 4706895 | 4304161 | 4509128 | 6563496 | 7.98E-01 | -0.04 | 0.65 |
| RP_NEG_216_78081a11_586 | | 216.7808 | 11.59 | | RP | NEG | | | 153421 | 46294.4 | 155154 | 52496.6 | 7.99E-01 | 0.04 | 0.81 |
| RP_POS_227_17493a3_259 | | 227.1749 | 3.26 | | RP | POS | | | 384834 | 147914 | 390643 | 181132 | 7.99E-01 | 0.04 | 0.35 |
| RP_POS_241_06786a4_826 | | 241.0679 | 4.83 | | RP | POS | | | 3.3E+07 | 1.8E+07 | 3.4E+07 | 2.2E+07 | 7.99E-01 | 0.04 | 0.66 |
| RP_POS_490_35269a10_219 | | 490.3527 | 10.22 | | RP | POS | | | 980529 | 440564 | 998880 | 588330 | 7.99E-01 | 0.04 | 0.61 |
| RP_NEG_349_01288a2_885 | | 349.0129 | 2.89 | | RP | NEG | | | 577938 | 82500.6 | 580830 | 83742.5 | 7.99E-01 | 0.03 | 0.53 |
| RP_POS_618_24658a10_36 | | 618.2466 | 10.36 | | RP | POS | | | 1473172 | 150745 | 1478573 | 158407 | 7.99E-01 | 0.03 | 0.25 |
| RP_NEG_327_23309a10_895 | Docosahexanoic acid | 327.2331 | 10.90 | | RP | NEG | 327.2336(100), 283.2428(90), 59.0132(70) | 1 | 2.5E+07 | 1.3E+07 | 2.5E+07 | 1.3E+07 | 7.99E-01 | -0.03 | 0.91 |
| RP_POS_360_16418a5_517 | | 360.1642 | 5.52 | | RP | POS | | | 357370 | 187214 | 351145 | 171019 | 7.99E-01 | -0.03 | 1.02 |
| HILIC_POS_518_32202a1_264 | LysoPC 18:3 | 518.3220 | 1.26 | | HILIC | POS | 104.1078(100), 184.0743(75) | 2 | 9178531 | 1602980 | 9233397 | 1552237 | 7.99E-01 | 0.03 | 0.54 |
| RP_POS_781_5285a9_908 | | 781.5285 | 9.91 | | RP | POS | | | 312783 | 217353 | 321773 | 290189 | 8.00E-01 | 0.04 | 0.98 |
| RP_POS_227_07932a2_883 | | 227.0793 | 2.88 | | RP | POS | | | 1315828 | 305782 | 1325589 | 260610 | 8.00E-01 | 0.03 | 0.41 |
| RP_POS_357_27859a9_099 | | 357.2786 | 9.10 | | RP | POS | | | 580957 | 1012586 | 615303 | 971382 | 8.00E-01 | 0.03 | 0.58 |
| HILIC_POS_394_31546a0_893 | | 394.3155 | 0.89 | | HILIC | POS | | | 225577 | 51278.4 | 223990 | 40711.3 | 8.00E-01 | -0.03 | 0.67 |
| RP_NEG_554_30176a10_311 | | 554.3018 | 10.31 | | RP | NEG | | | 4474622 | 866389 | 4442920 | 957055 | 8.00E-01 | -0.03 | 0.61 |
| RP_POS_401_34113a11_417 | 7a-Hydroxy-4-cholesten-3-one | 401.3411 | 11.42 | | RP | POS | 401.3431(100), 97.0654(80) | 1 | 2273087 | 1844838 | 2211726 | 1709678 | 8.00E-01 | -0.03 | 0.56 |
| RP_NEG_328_16251a9_829 | | 328.1625 | 9.83 | | RP | NEG | | | 381050 | 913075 | 410681 | 814704 | 8.01E-01 | 0.03 | 0.48 |
| RP_POS_163_11156a10_511 | | 163.1116 | 10.51 | | RP | POS | | | 1726425 | 992708 | 1759044 | 902999 | 8.01E-01 | 0.03 | 0.62 |
| RP_POS_326_00333a15_451 | | 326.0033 | 15.45 | | RP | POS | | | 441984 | 207634 | 449396 | 220324 | 8.01E-01 | 0.03 | 0.31 |
| RP_POS_742_53851a13_262 | | 742.5385 | 13.26 | | RP | POS | | | 2469885 | 1533629 | 2525879 | 1694252 | 8.01E-01 | 0.03 | 0.71 |
| RP_POS_228_23218a10_17 | Myristamide | 228.2322 | 10.17 | | RP | POS | 228.2316(100) | 1 | 1.3E+07 | 1465463 | 1.3E+07 | 1760061 | 8.01E-01 | -0.03 | 0.8 |
| RP_NEG_309_87701a7_6 | | 309.8770 | 7.60 | | RP | NEG | | | 299861 | 407963 | 286250 | 382525 | 8.01E-01 | -0.03 | 0.67 |
| RP_NEG_493_15402a6_981 | | 493.1540 | 6.98 | | RP | NEG | | | 496082 | 156898 | 490178 | 182356 | 8.01E-01 | -0.03 | 0.4 |
| RP_NEG_469_798a15_384 | | 469.7980 | 15.38 | | RP | NEG | | | 284452 | 114115 | 288431 | 117475 | 8.02E-01 | 0.03 | 0.32 |
| RP_POS_259_20575a10_712 | | 259.2058 | 10.71 | | RP | POS | | | 7078935 | 757328 | 7104635 | 739584 | 8.02E-01 | 0.03 | 0.39 |
| RP_POS_603_52179a15_278 | | 603.5218 | 15.28 | | RP | POS | | | 4214755 | 2897049 | 4292872 | 1577297 | 8.02E-01 | 0.03 | 0.25 |
| HILIC_POS_376_9928a7_352 | | 376.9928 | 7.35 | | HILIC | POS | | | 177440 | 13012.9 | 176996 | 12874 | 8.02E-01 | -0.03 | 0.39 |
| RP_NEG_893_19318a9_587 | | 893.1932 | 9.59 | | RP | NEG | | | 5933060 | 359686 | 5921287 | 328621 | 8.02E-01 | -0.03 | 0.81 |
| HILIC_POS_139_05026a1_048 | Urocanic acid | 139.0503 | 1.05 | | HILIC | POS | 139.0511(100), 121.0396(60) | 1 | 1716658 | 926502 | 1682531 | 1052674 | 8.02E-01 | -0.03 | 0.3 |
| RP_POS_495_36066a10_835 | | 495.3607 | 10.84 | | RP | POS | | | 2973693 | 817107 | 3020986 | 1723048 | 8.03E-01 | 0.04 | 0.66 |
| HILIC_POS_157_09731a4_858 | | 157.0973 | 4.86 | | HILIC | POS | | | 2299238 | 922359 | 2331405 | 959523 | 8.03E-01 | 0.03 | 0.63 |
| HILIC_POS_226_95103a5_46 | | 226.9510 | 5.46 | | HILIC | POS | | | 1915495 | 393793 | 1928096 | 346673 | 8.03E-01 | 0.03 | 0.33 |
| RP_NEG_256_98544a7_366 | | 256.9854 | 7.37 | | RP | NEG | | | 5588835 | 230313 | 5596455 | 218250 | 8.03E-01 | 0.03 | 0.33 |
| RP_POS_488_9837a10_07 | | 488.9837 | 10.07 | | RP | POS | | | 179432 | 102639 | 183107 | 112090 | 8.03E-01 | 0.03 | 0.39 |
| RP_POS_519_04108a10_618 | | 519.0411 | 10.62 | | RP | POS | | | 235892 | 152642 | 241389 | 168177 | 8.03E-01 | 0.03 | 0.38 |
| RP_POS_550_26459a11_24 | | 550.2646 | 11.24 | | RP | POS | | | 351745 | 219236 | 358930 | 203324 | 8.03E-01 | 0.03 | 0.76 |
| RP_NEG_128_03546a1_039 | | 128.0355 | 1.04 | | RP | NEG | | | 685447 | 110156 | 681332 | 128854 | 8.03E-01 | -0.03 | 0.5 |
| RP_NEG_145_89331a10_893 | | 145.8933 | 10.89 | | RP | NEG | | | 172611 | 91697.5 | 169470 | 92691 | 8.03E-01 | -0.03 | 0.91 |
| RP_NEG_243_19678a9_008 | | 243.1968 | 9.01 | | RP | NEG | | | 333861 | 63693.2 | 331719 | 62116.5 | 8.03E-01 | -0.03 | 0.58 |
| RP_POS_429_3725a11_222 | | 429.3725 | 11.22 | | RP | POS | | | 1370401 | 522688 | 1352984 | 500124 | 8.03E-01 | -0.03 | 0.6 |
| RP_POS_720_55389a13_098 | | 720.5539 | 13.10 | | RP | POS | | | 1049594 | 1094124 | 1011018 | 1162039 | 8.03E-01 | -0.03 | 0.61 |
| RP_POS_333_15408a6_343 | | 333.1541 | 6.34 | | RP | POS | | | 384050 | 346333 | 408684 | 930709 | 8.04E-01 | 0.04 | 0.86 |
| HILIC_NEG_230_98576a0_925 | | 230.9858 | 0.93 | | HILIC | NEG | | | 416246 | 54750 | 418112 | 55002.8 | 8.04E-01 | 0.03 | 0.6 |
| RP_NEG_230_98613a15_472 | | 230.9861 | 15.47 | | RP | NEG | | | 1899818 | 148458 | 1904883 | 149748 | 8.04E-01 | 0.03 | 0.54 |
| HILIC_POS_160_10805a2_562 | | 160.1081 | 2.56 | | HILIC | POS | | | 1393798 | 1852626 | 1319303 | 2456102 | 8.04E-01 | -0.03 | 0.77 |
| HILIC_POS_499_05734a3_146 | | 499.0573 | 3.15 | | HILIC | POS | | | 695591 | 103972 | 691966 | 109056 | 8.04E-01 | -0.03 | 0.34 |
| RP_POS_329_15668a4_337 | | 329.1567 | 4.34 | | RP | POS | | | 174988 | 94469.9 | 171393 | 114932 | 8.04E-01 | -0.03 | 0.75 |
| HILIC_NEG_234_8196a0_975 | | 234.8196 | 0.98 | | HILIC | NEG | | | 425023 | 203505 | 431879 | 203062 | 8.05E-01 | 0.03 | 0.41 |
| RP_POS_607_93042a10_305 | | 607.9304 | 10.31 | | RP | POS | | | 1745367 | 835170 | 1774955 | 911338 | 8.05E-01 | 0.03 | 0.61 |
| RP_POS_259_22662a10_388 | | 259.2266 | 10.39 | | RP | POS | | | 362662 | 105023 | 359128 | 104230 | 8.05E-01 | -0.03 | 0.25 |
| RP_POS_266_15933a4_538 | | 266.1593 | 4.54 | | RP | POS | | | 1468191 | 92799.1 | 1464655 | 113868 | 8.05E-01 | -0.03 | 0.64 |
| RP_POS_313_16248a5_732 | | 313.1625 | 5.73 | | RP | POS | | | 407673 | 342278 | 397068 | 289440 | 8.05E-01 | -0.03 | 0.8 |
| RP_POS_805_55396a12_864 | | 805.5540 | 12.86 | | RP | POS | | | 3.9E+07 | 4.8E+07 | 3.8E+07 | 5.1E+07 | 8.05E-01 | -0.03 | 0.64 |
| RP_NEG_1131_66345a10_295 | | 1131.6635 | 10.30 | | RP | NEG | | | 869290 | 327718 | 880134 | 317792 | 8.06E-01 | 0.03 | 0.53 |
| RP_NEG_279_1969a9_274 | | 279.1969 | 9.27 | | RP | NEG | | | 806477 | 882604 | 841747 | 1171974 | 8.06E-01 | 0.03 | 0.58 |
| RP_NEG_324_75818a15_455 | | 324.7582 | 15.46 | | RP | NEG | | | 5277997 | 9610415 | 5600998 | 9647678 | 8.06E-01 | 0.03 | 0.58 |
| RP_POS_392_70084a6_781 | | 392.7008 | 6.78 | | RP | POS | | | 544622 | 148340 | 549510 | 142729 | 8.06E-01 | 0.03 | 1.11 |
| RP_POS_559_32404a9_526 | | 559.3240 | 9.53 | | RP | POS | | | 834822 | 481199 | 852316 | 550204 | 8.06E-01 | 0.03 | 0.93 |
| RP_NEG_514_28479a7_352 | | 514.2848 | 7.35 | | RP | NEG | 514.285(100) | | 1038777 | 2182217 | 976500 | 1518663 | 8.06E-01 | -0.03 | 0.86 |
| RP_POS_325_20047a8_793 | | 325.2005 | 8.79 | | RP | POS | | | 1455584 | 205745 | 1448707 | 204453 | 8.06E-01 | -0.03 | 0.43 |
| RP_POS_187_13271a10_361 | | 187.1327 | 10.36 | | RP | POS | | | 3072419 | 404113 | 3058981 | 401395 | 8.07E-01 | -0.03 | 0.6 |
| RP_POS_227_1274a3_837 | | 227.1274 | 3.84 | | RP | POS | | | 185585 | 96351 | 182470 | 90233.2 | 8.07E-01 | -0.03 | 0.71 |
| RP_POS_391_16422a9_005 | | 391.1642 | 9.01 | | RP | POS | | | 562551 | 94692.2 | 559425 | 92996.3 | 8.07E-01 | -0.03 | 0.63 |
| RP_POS_515_34991a10_218 | | 515.3499 | 10.22 | | RP | POS | | | 263827 | 162095 | 258111 | 178882 | 8.07E-01 | -0.03 | 0.9 |
| RP_POS_710_10016a11_452 | | 710.1002 | 11.45 | | RP | POS | | | 294515 | 187170 | 288502 | 174093 | 8.07E-01 | -0.03 | 0.48 |
| RP_NEG_301_20239a8_398 | | 301.2024 | 8.40 | | RP | NEG | | | 264135 | 31599 | 265178 | 31308.9 | 8.08E-01 | 0.03 | 0.88 |
| RP_POS_213_11247a3_547 | | 213.1125 | 3.55 | | RP | POS | | | 98868.1 | 80585.5 | 101239 | 62462.5 | 8.08E-01 | 0.03 | 0.63 |
| RP_POS_338_23187a9_438 | | 338.2319 | 9.44 | | RP | POS | | | 755167 | 135166 | 759506 | 126034 | 8.08E-01 | 0.03 | 0.8 |
| RP_POS_546_35406a11_144 | | 546.3541 | 11.14 | | RP | POS | | | 4506695 | 1039824 | 4543405 | 1156546 | 8.08E-01 | 0.03 | 0.57 |
| RP_POS_636_95294a10_441 | | 636.9529 | 10.44 | | RP | POS | | | 909563 | 370455 | 922486 | 404814 | 8.08E-01 | 0.03 | 0.4 |
| HILIC_NEG_114_94918a3_158 | | 114.9492 | 3.16 | | HILIC | NEG | | | 646235 | 89043.7 | 643355 | 84895.8 | 8.08E-01 | -0.03 | 0.34 |
| HILIC_POS_131_08148a5_626 | | 131.0815 | 5.63 | | HILIC | POS | | | 454342 | 1085152 | 422122 | 854925 | 8.08E-01 | -0.03 | 0.42 |
| RP_NEG_273_05325a3_764 | | 273.0533 | 3.76 | | RP | NEG | | | 533829 | 103242 | 530564 | 94161.8 | 8.08E-01 | -0.03 | 0.54 |
| RP_POS_128_95079a0_719 | | 128.9508 | 0.72 | | RP | POS | | | 2563635 | 651777 | 2539256 | 794072 | 8.08E-01 | -0.03 | 0.55 |
| RP_POS_184_16982a10_835 | | 184.1698 | 10.84 | | RP | POS | | | 149230 | 119048 | 145238 | 121370 | 8.08E-01 | -0.03 | 0.8 |
| RP_POS_206_11769a7_708 | | 206.1177 | 7.71 | | RP | POS | | | 1158177 | 2101375 | 1087812 | 2123237 | 8.08E-01 | -0.03 | 0.53 |
| RP_POS_332_20255a7_997 | | 332.2026 | 8.00 | | RP | POS | | | 265289 | 36131.2 | 264016 | 40165 | 8.08E-01 | -0.03 | 0.55 |
| RP_POS_646_78223a10_353 | | 646.7822 | 10.35 | | RP | POS | | | 269418 | 183020 | 263514 | 173211 | 8.08E-01 | -0.03 | 0.43 |
| RP_NEG_172_82983a15_33 | | 172.8298 | 15.33 | | RP | NEG | | | 2158691 | 236724 | 2165656 | 187079 | 8.09E-01 | 0.03 | 0.51 |
| RP_POS_281_24741a10_925 | | 281.2474 | 10.93 | | RP | POS | | | 2193178 | 1160552 | 2232156 | 1190070 | 8.09E-01 | 0.03 | 0.64 |
| RP_POS_331_00247a8_092 | | 331.0025 | 8.09 | | RP | POS | | | 1988572 | 492232 | 2002618 | 359818 | 8.09E-01 | 0.03 | 0.56 |
| RP_POS_680_13843a11_408 | | 680.1384 | 11.41 | | RP | POS | | | 215924 | 113027 | 220132 | 138213 | 8.09E-01 | 0.03 | 0.55 |
| HILIC_NEG_172_00723a4_849 | | 172.0072 | 4.85 | | HILIC | NEG | | | 159666 | 145409 | 155713 | 91956.2 | 8.09E-01 | -0.03 | 0.49 |
| HILIC_NEG_480_30875a1_247 | | 480.3088 | 1.25 | | HILIC | NEG | | | 140640 | 31237.7 | 139559 | 34067.2 | 8.09E-01 | -0.03 | 0.69 |
| RP_NEG_225_89706a15_596 | | 225.8971 | 15.60 | | RP | NEG | | | 1421887 | 700053 | 1397370 | 777014 | 8.09E-01 | -0.03 | 0.39 |
| RP_NEG_302_93546a0_679 | | 302.9355 | 0.68 | | RP | NEG | | | 252916 | 58628.2 | 251125 | 50405.2 | 8.09E-01 | -0.03 | 0.79 |
| RP_POS_287_14899a4_597 | | 287.1490 | 4.60 | | RP | POS | | | 1206176 | 256947 | 1197345 | 275070 | 8.09E-01 | -0.03 | 0.5 |
| RP_POS_493_65414a10_218 | | 493.6541 | 10.22 | | RP | POS | | | 211693 | 98057.5 | 207726 | 136220 | 8.09E-01 | -0.03 | 0.68 |
| RP_POS_660_55579a14_836 | | 660.5558 | 14.84 | | RP | POS | | | 620821 | 638014 | 601778 | 517949 | 8.09E-01 | -0.03 | 0.36 |
| RP_POS_673_9715a10_251 | | 673.9715 | 10.25 | | RP | POS | | | 1030765 | 472804 | 1049314 | 629196 | 8.10E-01 | 0.03 | 0.75 |
| HILIC_NEG_144_92355a5_796 | | 144.9236 | 5.80 | | HILIC | NEG | | | 3.8E+07 | 6865308 | 3.8E+07 | 6260450 | 8.10E-01 | -0.03 | 0.41 |
| RP_NEG_336_78464a15_372 | | 336.7846 | 15.37 | | RP | NEG | | | 6755653 | 1432865 | 6706521 | 1549371 | 8.10E-01 | -0.03 | 0.69 |
| RP_POS_279_23187a9_94 | | 279.2319 | 9.94 | | RP | POS | | | 1497431 | 511446 | 1481232 | 478301 | 8.10E-01 | -0.03 | 0.55 |
| RP_POS_738_50421a13_127 | | 738.5042 | 13.13 | | RP | POS | | | 709701 | 670552 | 684867 | 816845 | 8.10E-01 | -0.03 | 0.64 |
| HILIC_POS_151_04784a6_136 | | 151.0478 | 6.14 | | HILIC | POS | | | 2323113 | 241531 | 2331026 | 242120 | 8.11E-01 | 0.03 | 0.7 |
| RP_POS_529_42834a14_992 | | 529.4283 | 14.99 | | RP | POS | | | 1519457 | 1564985 | 1573428 | 1719712 | 8.11E-01 | 0.03 | 0.52 |
| RP_NEG_176_90907a14_349 | | 176.9091 | 14.35 | | RP | NEG | | | 1840834 | 875833 | 1813447 | 807989 | 8.11E-01 | -0.03 | 0.5 |
| RP_POS_188_12825a10_362 | | 188.1283 | 10.36 | | RP | POS | | | 4302032 | 478267 | 4285888 | 505514 | 8.11E-01 | -0.03 | 0.68 |
| RP_POS_243_11607a15_535 | | 243.1161 | 15.54 | | RP | POS | | | 311579 | 207074 | 304996 | 195358 | 8.11E-01 | -0.03 | 0.5 |
| RP_POS_345_18423a1_438 | | 345.1842 | 1.44 | | RP | POS | | | 1782975 | 659923 | 1763881 | 510668 | 8.11E-01 | -0.03 | 0.38 |
| RP_POS_245_13135a10_43 | | 245.1314 | 10.43 | | RP | POS | | | 939355 | 358332 | 951081 | 361950 | 8.12E-01 | 0.03 | 0.58 |
| RP_POS_283_71765a5_754 | | 283.7177 | 5.75 | | RP | POS | | | 462126 | 123062 | 466299 | 132565 | 8.12E-01 | 0.03 | 0.52 |
| HILIC_NEG_315_94867a4_828 | | 315.9487 | 4.83 | | HILIC | NEG | | | 107618 | 17895 | 107082 | 15284.6 | 8.12E-01 | -0.03 | 0.27 |
| RP_NEG_765_16571a9_277 | | 765.1657 | 9.28 | | RP | NEG | | | 9831767 | 816813 | 9806227 | 757807 | 8.12E-01 | -0.03 | 1.06 |
| RP_POS_191_14279a9_695 | | 191.1428 | 9.70 | | RP | POS | | | 2096688 | 274855 | 2088199 | 249518 | 8.12E-01 | -0.03 | 0.56 |
| RP_POS_152_07066a2_25 | | 152.0707 | 2.25 | | RP | POS | 152.07(100), 110.0607(85) | | 1.1E+07 | 6E+07 | 1.3E+07 | 6.1E+07 | 8.13E-01 | 0.03 | 0.62 |
| HILIC_POS_156_0419a3_687 | | 156.0419 | 3.69 | | HILIC | POS | | | 1835679 | 2405802 | 1919563 | 2742903 | 8.13E-01 | 0.03 | 0.27 |
| HILIC_POS_526_29193a1_704 | | 526.2919 | 1.70 | | HILIC | POS | | | 809078 | 317270 | 819014 | 297640 | 8.13E-01 | 0.03 | 0.49 |
| RP_NEG_384_93304a0_679 | | 384.9330 | 0.68 | | RP | NEG | | | 297107 | 50675.1 | 298686 | 47023 | 8.13E-01 | 0.03 | 0.37 |
| RP_NEG_650_35059a13_175 | | 650.3506 | 13.18 | | RP | NEG | | | 421089 | 128142 | 425532 | 144062 | 8.13E-01 | 0.03 | 0.39 |
| HILIC_POS_118_08656a4_11 | | 118.0866 | 4.11 | | HILIC | POS | 118.0866(100) | | 1.2E+07 | 7205871 | 1.2E+07 | 5993725 | 8.13E-01 | -0.03 | 0.27 |
| RP_NEG_1043_1521a9_63 | | 1043.1521 | 9.63 | | RP | NEG | | | 2631315 | 209235 | 2624636 | 204920 | 8.13E-01 | -0.03 | 0.98 |
| RP_POS_162_07623a15_479 | | 162.0762 | 15.48 | | RP | POS | | | 7615272 | 1505859 | 7564956 | 1590547 | 8.13E-01 | -0.03 | 0.72 |
| RP_POS_474_2276a8_841 | | 474.2276 | 8.84 | | RP | POS | | | 224498 | 163699 | 219044 | 172844 | 8.13E-01 | -0.03 | 0.68 |
| HILIC_POS_371_23987a5_107 | | 371.2399 | 5.11 | | HILIC | POS | | | 8452613 | 4037591 | 8573949 | 3529519 | 8.14E-01 | 0.03 | 0.48 |
| HILIC_POS_553_40649a4_867 | | 553.4065 | 4.87 | | HILIC | POS | | | 386071 | 178374 | 391941 | 185401 | 8.14E-01 | 0.03 | 0.55 |
| RP_POS_213_10988a14_236 | | 213.1099 | 14.24 | | RP | POS | | | 2270018 | 1028870 | 2313314 | 1569398 | 8.14E-01 | 0.03 | 0.74 |
| RP_POS_624_27997a10_611 | | 624.2800 | 10.61 | | RP | POS | | | 1096094 | 218875 | 1103287 | 227858 | 8.14E-01 | 0.03 | 0.53 |
| HILIC_NEG_192_98903a3_052 | | 192.9890 | 3.05 | | HILIC | NEG | | | 205185 | 29636.9 | 204264 | 27662 | 8.14E-01 | -0.03 | 0.46 |
| RP_POS_177_0546a7_47 | | 177.0546 | 7.47 | | RP | POS | | | 7191108 | 998930 | 7163732 | 703800 | 8.14E-01 | -0.03 | 0.52 |
| RP_POS_514_29395a11_147 | | 514.2940 | 11.15 | | RP | POS | | | 151913 | 76332.1 | 154391 | 78111.3 | 8.15E-01 | 0.03 | 0.34 |
| RP_POS_542_42615a9_811 | | 542.4262 | 9.81 | | RP | POS | | | 1034123 | 483045 | 1053457 | 689481 | 8.15E-01 | 0.03 | 1.03 |
| RP_POS_564_6828a11_378 | | 564.6828 | 11.38 | | RP | POS | | | 90765.2 | 39337 | 92214.6 | 49944.9 | 8.15E-01 | 0.03 | 0.74 |
| HILIC_NEG_146_9214a0_937 | | 146.9214 | 0.94 | | HILIC | NEG | | | 708337 | 87461.4 | 705305 | 100492 | 8.15E-01 | -0.03 | 0.48 |
| RP_NEG_501_54852a15_497 | | 501.5485 | 15.50 | | RP | NEG | | | 704520 | 51260.6 | 702956 | 46649.3 | 8.15E-01 | -0.03 | 0.36 |
| RP_POS_128_0623a7_707 | | 128.0623 | 7.71 | | RP | POS | | | 1570451 | 2629789 | 1485329 | 2688698 | 8.15E-01 | -0.03 | 0.52 |
| RP_POS_222_18303a9_267 | | 222.1830 | 9.27 | | RP | POS | | | 776613 | 181089 | 771150 | 161307 | 8.15E-01 | -0.03 | 1.14 |
| RP_POS_267_12021a3_836 | | 267.1202 | 3.84 | | RP | POS | | | 199968 | 354296 | 189453 | 306963 | 8.15E-01 | -0.03 | 0.43 |
| RP_POS_326_24509a9_356 | | 326.2451 | 9.36 | | RP | POS | | | 229630 | 75071.4 | 227075 | 83472.1 | 8.15E-01 | -0.03 | 0.86 |
| RP_POS_633_45319a10_878 | | 633.4532 | 10.88 | | RP | POS | | | 516424 | 219963 | 509465 | 216056 | 8.15E-01 | -0.03 | 0.75 |
| RP_NEG_1035_66492a10_489 | | 1035.6649 | 10.49 | | RP | NEG | | | 5E+07 | 7876656 | 5E+07 | 8989671 | 8.16E-01 | 0.03 | 0.91 |
| RP_NEG_310_00189a2_163 | | 310.0019 | 2.16 | | RP | NEG | | | 445851 | 74181.3 | 448220 | 74887.1 | 8.16E-01 | 0.03 | 0.57 |

**Supplemantary table 1**: The non-targeted metabolomics results ordered by the VIP value (p value < 0.0003 are shaded dark green, p value < 0.05 are shaded light green, d values below 0 are blue and above 0 are red)

| | | | | | | | | | Cases | | Controls | | Welch test | Cohen's | |
|---|---|---|---|---|---|---|---|---|---|---|---|---|---|---|---|
| Feature_ID | Metabolite_name | Average_Mz | Average_RT_min_ | HMDB ID | RP_HILIC | POS_NEG | Cureted_MSMS | ID_level | Mean | SD | Mean2 | SD3 | p | d | VIP |
| RP_NEG_580_23602a10_899 | | 580.2360 | 10.90 | | RP | NEG | | | 290202 | 66775.9 | 292425 | 72694.2 | 8.16E-01 | 0.03 | 0.68 |
| RP_NEG_896_56464a13_463 | | 896.5646 | 13.46 | | RP | NEG | | | 1.2E+07 | 2571308 | 1.2E+07 | 2610311 | 8.16E-01 | 0.03 | 0.48 |
| RP_POS_155_1432a8_087 | | 155.1432 | 8.09 | | RP | POS | | | 290312 | 76399.3 | 292859 | 82831.5 | 8.16E-01 | 0.03 | 0.68 |
| RP_POS_186_9929a1_675 | | 186.9929 | 1.68 | | RP | POS | | | 766803 | 200386 | 773282 | 207196 | 8.16E-01 | 0.03 | 0.59 |
| RP_POS_478_34915a9_607 | | 478.3492 | 9.61 | | RP | POS | | | 4272784 | 1993562 | 4340951 | 2258260 | 8.16E-01 | 0.03 | 0.55 |
| RP_POS_340_21161a4_885 | | 340.2116 | 4.89 | | RP | POS | 340.2142(100) | | 595713 | 42030.1 | 594407 | 40252.9 | 8.16E-01 | -0.03 | 0.26 |
| HILIC_POS_387_21387a2_913 | | 387.2139 | 2.91 | | HILIC | POS | | | 558063 | 416186 | 545843 | 354810 | 8.16E-01 | -0.03 | 0.59 |
| RP_NEG_328_99506a1_227 | | 328.9951 | 1.23 | | RP | NEG | | | 466746 | 159050 | 462008 | 139593 | 8.16E-01 | -0.03 | 0.45 |
| RP_POS_186_14909a10_252 | | 186.1491 | 10.25 | | RP | POS | | | 2578534 | 304947 | 2568838 | 304949 | 8.16E-01 | -0.03 | 0.58 |
| RP_POS_315_66696a10_936 | | 315.6670 | 10.94 | | RP | POS | | | 518908 | 240654 | 511386 | 233822 | 8.16E-01 | -0.03 | 0.76 |
| RP_POS_400_29056a6_359 | | 400.2906 | 6.36 | | RP | POS | | | 932887 | 427881 | 919871 | 390811 | 8.16E-01 | -0.03 | 0.47 |
| HILIC_NEG_195_81107a3_247 | | 195.8111 | 3.25 | | HILIC | NEG | | | 3.7E+07 | 1.3E+07 | 3.7E+07 | 1.4E+07 | 8.17E-01 | 0.03 | 0.42 |
| RP_NEG_544_26495a10_303 | | 544.2650 | 10.30 | | RP | NEG | | | 1190977 | 310366 | 1201874 | 370376 | 8.17E-01 | 0.03 | 0.68 |
| RP_POS_194_89449a15_404 | | 194.8945 | 15.40 | | RP | POS | | | 147873 | 81109.7 | 150576 | 88564.5 | 8.17E-01 | 0.03 | 0.4 |
| RP_POS_329_00516a8_091 | | 329.0052 | 8.09 | | RP | POS | | | 7000467 | 1462007 | 7037571 | 858114 | 8.17E-01 | 0.03 | 0.73 |
| RP_POS_560_50311a15_175 | | 560.5031 | 15.18 | | RP | POS | | | 2115834 | 883183 | 2142952 | 831818 | 8.17E-01 | 0.03 | 0.37 |
| RP_POS_586_4361a10_668 | | 586.4361 | 10.67 | | RP | POS | | | 538934 | 339030 | 549377 | 323790 | 8.17E-01 | 0.03 | 0.74 |
| HILIC_NEG_219_87561a0_898 | | 219.8756 | 0.90 | | HILIC | NEG | | | 262839 | 31525.5 | 261760 | 36132.9 | 8.17E-01 | -0.03 | 0.45 |
| RP_NEG_236_10545a8_303 | | 236.1055 | 8.30 | | RP | NEG | | | 316027 | 54535 | 314372 | 50175.3 | 8.17E-01 | -0.03 | 0.77 |
| RP_POS_262_04977a5_734 | | 262.0498 | 5.73 | | RP | POS | | | 186911 | 75497 | 184563 | 73380.8 | 8.17E-01 | -0.03 | 0.54 |
| RP_POS_709_32788a11_579 | | 709.3279 | 11.58 | | RP | POS | | | 115764 | 56812.7 | 113841 | 64134.5 | 8.17E-01 | -0.03 | 0.62 |
| HILIC_NEG_87_00842a1_744 | | 87.0084 | 1.74 | | HILIC | NEG | | | 4355212 | 3655038 | 4472144 | 3761082 | 8.18E-01 | 0.03 | 0.51 |
| RP_NEG_124_01524a0_678 | | 124.0152 | 0.68 | | RP | NEG | | | 364636 | 49538.9 | 366298 | 55491.5 | 8.18E-01 | 0.03 | 0.23 |
| RP_POS_249_02806a0_586 | | 249.0281 | 0.59 | | RP | POS | | | 1120633 | 337736 | 1130805 | 310046 | 8.18E-01 | 0.03 | 0.95 |
| RP_POS_756_72321a11_435 | | 756.7232 | 11.44 | | RP | POS | | | 164508 | 89203.2 | 167080 | 75083.3 | 8.18E-01 | 0.03 | 0.48 |
| RP_POS_228_19574a7_748 | | 228.1957 | 7.75 | | RP | POS | 95.0861(100), 109.102(55), 175.1474(45) | | 1136738 | 507219 | 1122356 | 412211 | 8.18E-01 | -0.03 | 0.44 |
| RP_NEG_263_77252a15_486 | | 263.7725 | 15.49 | | RP | NEG | | | 188959 | 34726 | 187903 | 32590 | 8.18E-01 | -0.03 | 0.23 |
| RP_NEG_750_54529a14_242 | | 750.5453 | 14.24 | | RP | NEG | | | 1303983 | 483881 | 1287171 | 572092 | 8.18E-01 | -0.03 | 0.69 |
| RP_NEG_793_48108a11_593 | | 793.4811 | 11.59 | | RP | NEG | | | 367973 | 150041 | 363363 | 143455 | 8.18E-01 | -0.03 | 1.1 |
| RP_NEG_896_56451a13_332 | | 896.5645 | 13.33 | | RP | NEG | | | 5135093 | 2514286 | 5047989 | 2975344 | 8.18E-01 | -0.03 | 0.8 |
| RP_POS_221_11714a9_071 | | 221.1171 | 9.07 | | RP | POS | | | 1864238 | 142000 | 1859715 | 145343 | 8.18E-01 | -0.03 | 0.56 |
| RP_POS_413_32297a10_873 | | 413.3230 | 10.87 | | RP | POS | | | 5059868 | 846255 | 5032944 | 859254 | 8.18E-01 | -0.03 | 1.11 |
| RP_NEG_556_33197a9_708 | | 556.3320 | 9.71 | | RP | NEG | | | 277064 | 362635 | 287392 | 301669 | 8.19E-01 | 0.03 | 0.52 |
| RP_NEG_659_07758a9_244 | | 659.0776 | 9.24 | | RP | NEG | | | 598681 | 50403.4 | 600181 | 45472 | 8.19E-01 | 0.03 | 0.71 |
| RP_POS_175_10774a0_578 | | 175.1077 | 0.58 | | RP | POS | | | 1410966 | 623674 | 1430579 | 627973 | 8.19E-01 | 0.03 | 0.5 |
| RP_POS_176_97424a15_408 | | 176.9742 | 15.41 | | RP | POS | | | 3.5E+07 | 8061642 | 3.6E+07 | 9659539 | 8.19E-01 | 0.03 | 0.46 |
| RP_POS_229_10701a8_585 | | 229.1070 | 8.59 | | RP | POS | | | 394127 | 171202 | 399365 | 163971 | 8.19E-01 | 0.03 | 0.98 |
| RP_POS_569_43451a11_142 | | 569.4345 | 11.14 | | RP | POS | | | 2301060 | 5322269 | 2490086 | 6604259 | 8.19E-01 | 0.03 | 0.6 |
| RP_POS_768_55316a13_836 | | 768.5532 | 13.84 | | RP | POS | | | 6355963 | 3034901 | 6442340 | 2510517 | 8.19E-01 | 0.03 | 0.77 |
| HILIC_POS_297_07956a4_855 | | 297.0796 | 4.86 | | HILIC | POS | | | 902152 | 575830 | 884783 | 535042 | 8.19E-01 | -0.03 | 0.66 |
| RP_POS_770_60516a13_948 | | 770.6052 | 13.95 | | RP | POS | | | 8128073 | 2807965 | 8038761 | 2901009 | 8.19E-01 | -0.03 | 0.46 |
| HILIC_POS_283_0498a3_146 | | 283.0498 | 3.15 | | HILIC | POS | | | 193678 | 50382.2 | 195120 | 42648.7 | 8.20E-01 | 0.03 | 0.84 |
| RP_POS_198_1852a10_563 | | 198.1852 | 10.56 | | RP | POS | | | 1.7E+07 | 7203562 | 1.7E+07 | 7544267 | 8.20E-01 | 0.03 | 0.36 |
| RP_POS_201_14827a7_141 | | 201.1483 | 7.14 | | RP | POS | | | 473658 | 103614 | 476963 | 107931 | 8.20E-01 | 0.03 | 0.82 |
| RP_POS_282_06406a15_33 | | 282.0641 | 15.33 | | RP | POS | | | 597223 | 155540 | 602530 | 182597 | 8.20E-01 | 0.03 | 0.21 |
| HILIC_NEG_134_89473a3_108 | | 134.8947 | 3.11 | | HILIC | NEG | | | 3.1E+08 | 7691191 | 3.1E+08 | 7153522 | 8.20E-01 | -0.03 | 0.71 |
| RP_NEG_283_24332a10_894 | | 283.2433 | 10.89 | | RP | NEG | | | 241529 | 127404 | 237552 | 128376 | 8.20E-01 | -0.03 | 0.9 |
| RP_NEG_295_18964a10_684 | | 295.1896 | 10.68 | | RP | NEG | | | 301401 | 115061 | 298028 | 102572 | 8.20E-01 | -0.03 | 0.98 |
| RP_NEG_704_336a10_935 | | 704.3360 | 10.94 | | RP | NEG | | | 3438897 | 563611 | 3420601 | 606821 | 8.20E-01 | -0.03 | 0.63 |
| RP_NEG_851_1145a9_785 | | 851.1145 | 9.79 | | RP | NEG | | | 3643648 | 188087 | 3638138 | 167288 | 8.20E-01 | -0.03 | 0.85 |
| RP_POS_184_06039a0_6 | | 184.0604 | 0.60 | | RP | POS | | | 342002 | 352688 | 332724 | 243790 | 8.20E-01 | -0.03 | 0.38 |
| RP_POS_619_75214a6_088 | | 619.7521 | 6.09 | | RP | POS | | | 3E+07 | 1.1E+07 | 3E+07 | 1.3E+07 | 8.20E-01 | -0.03 | 0.46 |
| HILIC_POS_276_11908a7_267 | Glu-Gln | 276.1191 | 7.27 | | HILIC | POS | 130.05(100) | 2 | 206874 | 44109.3 | 208324 | 48737.2 | 8.20E-01 | 0.03 | 0.49 |
| RP_NEG_164_92699a15_424 | | 164.9270 | 15.42 | | RP | NEG | | | 768325 | 642069 | 787778 | 614225 | 8.21E-01 | 0.03 | 0.41 |
| RP_NEG_204_98952a15_474 | | 204.9895 | 15.47 | | RP | NEG | | | 1605081 | 209015 | 1611475 | 204291 | 8.21E-01 | 0.03 | 0.5 |
| RP_POS_120_10825a6_983 | | 120.1083 | 6.98 | | RP | POS | | | 210484 | 91635.4 | 213753 | 116147 | 8.21E-01 | 0.03 | 0.45 |
| RP_POS_358_36771a9_675 | | 358.3677 | 9.68 | | RP | POS | | | 263811 | 569720 | 283200 | 672096 | 8.21E-01 | 0.03 | 0.31 |
| RP_POS_480_2384a9_209 | | 480.2384 | 9.21 | | RP | POS | | | 1293968 | 479929 | 1309775 | 534082 | 8.21E-01 | 0.03 | 0.58 |
| RP_POS_596_26489a10_493 | | 596.2649 | 10.49 | | RP | POS | | | 891198 | 484593 | 907055 | 534628 | 8.21E-01 | 0.03 | 0.33 |
| RP_NEG_493_2814a9_371 | | 493.2814 | 9.37 | | RP | NEG | | | 524103 | 507985 | 505549 | 668563 | 8.21E-01 | -0.03 | 0.55 |
| RP_POS_428_33701a11_235 | | 428.3370 | 11.24 | | RP | POS | | | 186594 | 95969.4 | 183809 | 85197.1 | 8.21E-01 | -0.03 | 0.54 |
| RP_POS_469_36505a12_338 | | 469.3651 | 12.34 | | RP | POS | | | 1576983 | 781838 | 1557044 | 495251 | 8.21E-01 | -0.03 | 0.71 |
| HILIC_NEG_68_99507a0_573 | | 68.9951 | 0.57 | | HILIC | NEG | | | 1432937 | 252096 | 1425013 | 262728 | 8.22E-01 | -0.03 | 0.54 |
| RP_POS_1061_65393a10_31 | | 1061.6539 | 10.31 | | RP | POS | | | 9056891 | 3627562 | 8942205 | 3798487 | 8.22E-01 | -0.03 | 0.64 |
| RP_POS_611_92822a11_257 | | 611.9282 | 11.26 | | RP | POS | | | 163972 | 84793 | 161334 | 86300.5 | 8.22E-01 | -0.03 | 0.42 |
| RP_POS_572_43695a9_454 | | 572.4370 | 9.45 | | RP | POS | | | 3925387 | 1957057 | 3989490 | 2207978 | 8.23E-01 | 0.03 | 0.56 |
| HILIC_NEG_134_89491a7_382 | | 134.8949 | 7.38 | | HILIC | NEG | | | 1E+07 | 1360685 | 1E+07 | 1205786 | 8.23E-01 | -0.03 | 0.24 |
| HILIC_NEG_312_83606a7_011 | | 312.8361 | 7.01 | | HILIC | NEG | | | 393529 | 131438 | 388798 | 172512 | 8.23E-01 | -0.03 | 0.49 |
| RP_NEG_296_97778a7_52 | | 296.9778 | 7.52 | | RP | NEG | | | 1202111 | 174121 | 1196684 | 180882 | 8.23E-01 | -0.03 | 0.6 |
| RP_POS_259_19037a9_531 | | 259.1904 | 9.53 | | RP | POS | | | 2.2E+07 | 4868890 | 2.2E+07 | 5623555 | 8.23E-01 | -0.03 | 0.55 |
| RP_POS_461_36243a11_798 | | 461.3624 | 11.80 | | RP | POS | | | 269161 | 228003 | 263321 | 152134 | 8.23E-01 | -0.03 | 0.33 |
| RP_POS_627_26141a7_494 | | 627.2614 | 7.49 | | RP | POS | | | 327705 | 145554 | 323435 | 134423 | 8.23E-01 | -0.03 | 1.17 |
| RP_POS_886_51959a14_083 | | 886.5196 | 14.08 | | RP | POS | | | 174639 | 201379 | 169759 | 112964 | 8.23E-01 | -0.03 | 0.47 |
| HILIC_POS_123_05558a0_607 | Nicotinamide | 123.0556 | 0.61 | | HILIC | POS | 123.0557(100), 80.0501(10) | 1 | 833780 | 791370 | 815459 | 362795 | 8.23E-01 | -0.03 | 0.41 |
| HILIC_POS_173_07812a4_365 | | 173.0781 | 4.37 | | HILIC | POS | | | 1068038 | 283941 | 1077111 | 308672 | 8.24E-01 | 0.03 | 0.52 |
| RP_POS_257_04193a4_825 | | 257.0419 | 4.83 | | RP | POS | | | 2192362 | 1158339 | 2231254 | 1377770 | 8.24E-01 | 0.03 | 0.71 |
| RP_POS_578_42651a15_06 | | 578.4265 | 15.06 | | RP | POS | | | 1012965 | 592361 | 1036052 | 873444 | 8.24E-01 | 0.03 | 0.56 |
| RP_POS_784_10187a14_351 | | 784.1019 | 14.35 | | RP | POS | | | 682626 | 737391 | 700478 | 421485 | 8.24E-01 | 0.03 | 0.52 |
| RP_POS_864_4093a5_624 | | 864.4093 | 5.62 | | RP | POS | | | 2761578 | 1189936 | 2807180 | 1723577 | 8.24E-01 | 0.03 | 0.61 |
| HILIC_NEG_153_86868a6_879 | | 153.8687 | 6.88 | | HILIC | NEG | | | 869778 | 482475 | 856069 | 420737 | 8.24E-01 | -0.03 | 0.3 |
| RP_POS_336_20148a5_389 | | 336.2015 | 5.39 | | RP | POS | | | 510918 | 195289 | 505257 | 178590 | 8.24E-01 | -0.03 | 0.73 |
| RP_POS_629_4021a11_824 | | 629.4021 | 11.82 | | RP | POS | | | 602944 | 478183 | 588808 | 451913 | 8.24E-01 | -0.03 | 0.5 |
| RP_POS_709_47003a9_408 | | 709.4700 | 9.41 | | RP | POS | | | 215006 | 132542 | 210640 | 152376 | 8.24E-01 | -0.03 | 0.72 |
| HILIC_POS_188_12819a3_107 | | 188.1282 | 3.11 | | HILIC | POS | | | 228245 | 118764 | 231883 | 121954 | 8.25E-01 | 0.03 | 0.36 |
| RP_NEG_144_90388a15_402 | | 144.9039 | 15.40 | | RP | NEG | | | 1566537 | 648080 | 1585518 | 610251 | 8.25E-01 | 0.03 | 0.64 |
| RP_NEG_369_20508a10_689 | | 369.2051 | 10.69 | | RP | NEG | | | 234214 | 135839 | 238186 | 127664 | 8.25E-01 | 0.03 | 0.92 |
| RP_POS_325_21368a10_69 | | 325.2137 | 10.69 | | RP | POS | | | 2412726 | 1318644 | 2451192 | 1237266 | 8.25E-01 | 0.03 | 0.88 |
| RP_POS_394_40454a12_481 | | 394.4045 | 12.48 | | RP | POS | | | 2282384 | 997594 | 2311028 | 902909 | 8.25E-01 | 0.03 | 0.71 |
| RP_POS_564_42865a12_442 | | 564.4287 | 12.44 | | RP | POS | | | 437504 | 187353 | 443223 | 190012 | 8.25E-01 | 0.03 | 0.41 |
| RP_NEG_1083_66455a10_31 | | 1083.6646 | 10.31 | | RP | NEG | | | 1E+07 | 4869416 | 1E+07 | 4981344 | 8.25E-01 | -0.03 | 0.63 |
| RP_POS_530_37268a8_912 | | 530.3727 | 8.91 | | RP | POS | | | 474891 | 269788 | 466706 | 271143 | 8.25E-01 | -0.03 | 0.6 |
| RP_POS_1173_84985a13_238 | | 1173.8499 | 13.24 | | RP | POS | | | 2304441 | 2292224 | 2370975 | 2144234 | 8.26E-01 | 0.03 | 0.77 |
| RP_POS_436_37811a12_23 | | 436.3781 | 12.23 | | RP | POS | | | 1564431 | 259732 | 1572335 | 265695 | 8.26E-01 | 0.03 | 0.63 |
| HILIC_NEG_343_88641a3_09 | | 343.8864 | 3.09 | | HILIC | NEG | | | 168625 | 28199 | 167759 | 29283 | 8.26E-01 | -0.03 | 0.87 |
| HILIC_POS_558_29443a1_221 | | 558.2944 | 1.22 | | HILIC | POS | | | 234024 | 316560 | 224587 | 313236 | 8.26E-01 | -0.03 | 0.24 |
| RP_POS_253_63113a9_926 | | 253.6311 | 9.93 | | RP | POS | | | 340466 | 178603 | 334687 | 202907 | 8.26E-01 | -0.03 | 0.84 |
| RP_NEG_223_09799a6_977 | Olivetolcarboxylic acid | 223.0980 | 6.98 | | RP | NEG | 223.0966(100), 179.0959(10) | 2 | 1259035 | 77459.5 | 1256352 | 98334 | 8.26E-01 | -0.03 | 0.42 |
| HILIC_NEG_138_89001a3_108 | | 138.8900 | 3.11 | | HILIC | NEG | | | 2467136 | 104542 | 2470158 | 98671 | 8.27E-01 | 0.03 | 0.7 |
| RP_POS_207_00156a15_406 | | 207.0016 | 15.41 | | RP | POS | | | 572092 | 265136 | 579269 | 216356 | 8.27E-01 | 0.03 | 0.35 |
| HILIC_NEG_231_93185a5_654 | | 231.9319 | 5.65 | | HILIC | NEG | | | 515553 | 119949 | 512305 | 98010.7 | 8.27E-01 | -0.03 | 0.47 |
| RP_NEG_283_17068a10_001 | | 283.1707 | 10.00 | | RP | NEG | | | 436921 | 379832 | 426232 | 337443 | 8.27E-01 | -0.03 | 0.73 |
| RP_NEG_743_16321a8_775 | | 743.1632 | 8.78 | | RP | NEG | | | 822241 | 62444.3 | 820543 | 51731.1 | 8.27E-01 | -0.03 | 0.82 |
| RP_POS_311_29422a10_203 | | 311.2942 | 10.20 | | RP | POS | | | 654669 | 95653.1 | 651633 | 105986 | 8.27E-01 | -0.03 | 0.42 |
| RP_POS_591_69867a11_398 | | 591.6987 | 11.40 | | RP | POS | | | 158017 | 96022.3 | 155105 | 98897.8 | 8.27E-01 | -0.03 | 0.34 |
| RP_POS_253_10699a7_331 | Trinexapac-ethyl | 253.1070 | 7.33 | | RP | POS | 179.1074(100), 253.1049(40) | 2 | 1980696 | 137102 | 1976227 | 159696 | 8.27E-01 | -0.03 | 0.63 |
| HILIC_POS_185_95972a5_384 | | 185.9597 | 5.38 | | HILIC | POS | | | 195630 | 103373 | 198781 | 108849 | 8.28E-01 | 0.03 | 0.78 |
| RP_NEG_852_5766a13_247 | | 852.5766 | 13.25 | | RP | NEG | | | 1.9E+07 | 3730493 | 1.9E+07 | 3759236 | 8.28E-01 | 0.03 | 0.65 |
| RP_POS_252_04556a15_526 | | 252.0456 | 15.53 | | RP | POS | | | 6358323 | 1157616 | 6391951 | 1107789 | 8.28E-01 | 0.03 | 0.68 |
| RP_POS_330_30075a9_569 | | 330.3008 | 9.57 | | RP | POS | | | 228142 | 86916.9 | 230541 | 74895.9 | 8.28E-01 | 0.03 | 0.37 |
| RP_POS_650_43799a10_774 | | 650.4380 | 10.77 | | RP | POS | | | 495371 | 293853 | 504706 | 328912 | 8.28E-01 | 0.03 | 0.87 |
| RP_NEG_203_0145a0_686 | | 203.0145 | 0.69 | | RP | NEG | | | 313903 | 68971.4 | 311985 | 60866.4 | 8.28E-01 | -0.03 | 0.64 |
| RP_POS_362_27206a10_878 | | 362.2721 | 10.88 | | RP | POS | | | 537419 | 158120 | 532697 | 159510 | 8.28E-01 | -0.03 | 0.31 |
| RP_POS_553_45801a15_186 | | 553.4580 | 15.19 | | RP | POS | | | 2E+08 | 3.7E+07 | 2E+08 | 3.5E+07 | 8.28E-01 | -0.03 | 1.09 |
| HILIC_NEG_236_81673a4_197 | | 236.8167 | 4.20 | | HILIC | NEG | | | 198693 | 98159.1 | 201661 | 103005 | 8.29E-01 | 0.03 | 0.29 |
| RP_NEG_219_95174a15_559 | | 219.9517 | 15.56 | | RP | NEG | | | 1495106 | 292987 | 1503336 | 265624 | 8.29E-01 | 0.03 | 0.33 |
| RP_NEG_251_0927a6_491 | | 251.0927 | 6.49 | | RP | NEG | | | 302923 | 16867.1 | 303473 | 19998.3 | 8.29E-01 | 0.03 | 0.29 |
| RP_NEG_990_56775a13_948 | | 990.5678 | 13.95 | | RP | NEG | | | 1996967 | 843827 | 2021532 | 820692 | 8.29E-01 | 0.03 | 0.48 |
| RP_POS_249_13293a4_538 | | 249.1329 | 4.54 | | RP | POS | | | 1746892 | 119824 | 1750651 | 132910 | 8.29E-01 | 0.03 | 0.72 |
| RP_POS_537_37836a9_143 | | 537.3784 | 9.14 | | RP | POS | | | 334493 | 207595 | 340637 | 208880 | 8.29E-01 | 0.03 | 0.74 |
| RP_POS_631_45044a10_118 | | 631.4504 | 10.12 | | RP | POS | | | 7287507 | 2896835 | 7380952 | 3363437 | 8.29E-01 | 0.03 | 0.69 |
| HILIC_NEG_252_95609a4_866 | | 252.9561 | 4.87 | | HILIC | NEG | | | 654971 | 231568 | 648526 | 207036 | 8.29E-01 | -0.03 | 0.62 |
| HILIC_NEG_272_95938a7_41 | | 272.9594 | 7.41 | | HILIC | NEG | | | 92640.5 | 37257.7 | 91567.5 | 35358 | 8.29E-01 | -0.03 | 0.8 |
| HILIC_POS_189_07315a7_104 | | 189.0732 | 7.10 | | HILIC | POS | | | 501250 | 117430 | 497881 | 111225 | 8.29E-01 | -0.03 | 0.49 |
| HILIC_POS_158_08389a4_042 | 3-Methylcrotonylglycine | 158.0839 | 4.04 | | HILIC | POS | 158.0846(100), 53.0396(90) | 2 | 423446 | 1631313 | 377755 | 1467791 | 8.29E-01 | -0.03 | 0.55 |
| RP_NEG_512_29993a9_77 | | 512.2999 | 9.77 | | RP | NEG | | | 3080754 | 1126388 | 3115174 | 1211098 | 8.30E-01 | 0.03 | 0.41 |
| RP_POS_701_09125a11_477 | | 701.0913 | 11.48 | | RP | POS | | | 216930 | 134143 | 220601 | 116147 | 8.30E-01 | 0.03 | 0.46 |
| RP_POS_170_11751a10_252 | | 170.1175 | 10.25 | | RP | POS | | | 2246912 | 212054 | 2240293 | 237348 | 8.30E-01 | -0.03 | 0.59 |
| RP_NEG_564_33112a10_31 | LysoPC 18:2 | 564.3311 | 10.31 | | RP | NEG | 279.2339(100), 504.3123(18) | 1 | 2.2E+08 | 5.8E+07 | 2.1E+08 | 6E+07 | 8.30E-01 | -0.03 | 0.63 |
| HILIC_NEG_292_94287a3_916 | | 292.9429 | 3.92 | | HILIC | NEG | | | 146106 | 48598.7 | 147643 | 55909.2 | 8.31E-01 | 0.03 | 0.5 |
| HILIC_POS_211_13266a3_03 | | 211.1327 | 3.03 | | HILIC | POS | | | 674967 | 309923 | 684288 | 328827 | 8.31E-01 | 0.03 | 0.68 |
| RP_POS_303_19287a9_275 | | 303.1929 | 9.28 | | RP | POS | | | 1386414 | 1195001 | 1427782 | 1580022 | 8.31E-01 | 0.03 | 0.59 |
| RP_POS_434_25687a13_111 | | 434.2569 | 13.11 | | RP | POS | | | 250573 | 169568 | 255807 | 188236 | 8.31E-01 | 0.03 | 0.56 |
| HILIC_POS_171_14922a1_372 | | 171.1492 | 1.37 | | HILIC | POS | | | 3060228 | 1306726 | 3019736 | 1448365 | 8.31E-01 | -0.03 | 0.51 |
| RP_POS_189_04315a15_376 | | 189.0432 | 15.38 | | RP | POS | | | 2638441 | 945246 | 2610864 | 943153 | 8.31E-01 | -0.03 | 0.42 |
| RP_POS_531_40771a12_488 | | 531.4077 | 12.49 | | RP | POS | | | 1.1E+07 | 3260213 | 1.1E+07 | 3409000 | 8.31E-01 | -0.03 | 0.37 |
| HILIC_POS_172_13312a1_361 | | 172.1331 | 1.36 | | HILIC | POS | | | 734664 | 342328 | 744727 | 348936 | 8.32E-01 | 0.03 | 0.52 |
| RP_NEG_328_16226a9_662 | | 328.1623 | 9.66 | | RP | NEG | | | 125842 | 193737 | 130939 | 159550 | 8.32E-01 | 0.03 | 0.47 |
| RP_POS_303_88373a0_799 | | 303.8837 | 0.80 | | RP | POS | | | 471774 | 229612 | 478189 | 214465 | 8.32E-01 | 0.03 | 0.22 |
| RP_POS_544_43964a14_356 | | 544.4396 | 14.36 | | RP | POS | | | 3932978 | 1017177 | 3962427 | 1011171 | 8.32E-01 | 0.03 | 0.51 |
| RP_POS_654_33026a10_568 | | 654.3303 | 10.57 | | RP | POS | | | 6924646 | 2384249 | 7029695 | 4388437 | 8.32E-01 | 0.03 | 0.62 |
| RP_POS_682_36438a10_569 | | 682.3644 | 10.57 | | RP | POS | | | 232930 | 117362 | 237719 | 197103 | 8.32E-01 | 0.03 | 0.53 |
| HILIC_NEG_213_88008a3_148 | | 213.8801 | 3.15 | | HILIC | NEG | | | 170797 | 57723.8 | 169175 | 54589 | 8.32E-01 | -0.03 | 0.42 |
| RP_NEG_765_16559a9_482 | | 765.1656 | 9.48 | | RP | NEG | | | 1.2E+07 | 701312 | 1.2E+07 | 645857 | 8.32E-01 | -0.03 | 0.64 |
| RP_POS_338_19885a10_835 | | 338.1989 | 10.84 | | RP | POS | | | 889192 | 288352 | 880307 | 320873 | 8.32E-01 | -0.03 | 0.57 |
| RP_POS_457_27112a11_843 | | 457.2711 | 11.84 | | RP | POS | | | 191850 | 120775 | 188324 | 122804 | 8.32E-01 | -0.03 | 0.35 |
| HILIC_NEG_100_93348a4_861 | | 100.9335 | 4.86 | | HILIC | NEG | | | 6.5E+07 | 1.2E+07 | 6.5E+07 | 1.1E+07 | 8.33E-01 | 0.03 | 0.55 |
| HILIC_POS_344_01379a6_31 | | 344.0138 | 6.31 | | HILIC | POS | | | 800532 | 296682 | 809697 | 333650 | 8.33E-01 | 0.03 | 0.38 |
| HILIC_POS_415_28265a2_924 | | 415.2827 | 2.92 | | HILIC | POS | | | 108313 | 45009.1 | 109625 | 46125.4 | 8.33E-01 | 0.03 | 0.45 |
| RP_NEG_856_60895a14_447 | | 856.6090 | 14.45 | | RP | NEG | | | 1.4E+07 | 5442246 | 1.4E+07 | 5896578 | 8.33E-01 | 0.03 | 0.52 |
| RP_POS_311_1459a6_079 | | 311.1459 | 6.08 | | RP | POS | | | 208292 | 108328 | 211322 | 101870 | 8.33E-01 | 0.03 | 0.98 |
| RP_NEG_277_98444a3_316 | | 277.9844 | 3.32 | | RP | NEG | | | 1073455 | 291776 | 1065488 | 261004 | 8.33E-01 | -0.03 | 0.76 |
| RP_NEG_469_33261a10_269 | 18b-glycyrrhetinic acid | 469.3326 | 10.27 | | RP | NEG | 469.3357(100) | 2 | 1655781 | 2517986 | 1581826 | 2618262 | 8.33E-01 | -0.03 | 0.17 |
| RP_NEG_191_86032a14_337 | | 191.8603 | 14.34 | | RP | NEG | | | 175923 | 99323.3 | 178784 | 99777.6 | 8.34E-01 | 0.03 | 0.28 |
| HILIC_NEG_165_8373a3_114 | | 165.8373 | 3.11 | | HILIC | NEG | | | 2.1E+07 | 626630 | 2.1E+07 | 629713 | 8.34E-01 | -0.03 | 0.8 |
| HILIC_POS_459_30823a2_924 | | 459.3082 | 2.92 | | HILIC | POS | | | 106292 | 51741.9 | 104871 | 47451.5 | 8.34E-01 | -0.03 | 0.59 |
| RP_POS_625_41205a10_115 | | 625.4121 | 10.12 | | RP | POS | | | 188514 | 98392.5 | 185526 | 108613 | 8.34E-01 | -0.03 | 0.82 |
| RP_POS_790_53601a12_792 | | 790.5360 | 12.79 | | RP | POS | | | 6590272 | 3580915 | 6488468 | 3512091 | 8.34E-01 | -0.03 | 0.57 |
| HILIC_POS_331_24716a1_35 | | 331.2472 | 1.35 | | HILIC | POS | | | 891645 | 337641 | 901925 | 380101 | 8.35E-01 | 0.03 | 0.51 |
| HILIC_POS_371_24002a5_928 | | 371.2400 | 5.93 | | HILIC | POS | | | 5854638 | 774256 | 5876760 | 775783 | 8.35E-01 | 0.03 | 0.47 |
| RP_POS_267_15662a8_078 | | 267.1566 | 8.08 | | RP | POS | | | 523057 | 240474 | 530206 | 258600 | 8.35E-01 | 0.03 | 0.5 |
| HILIC_NEG_158_93925a5_465 | | 158.9393 | 5.47 | | HILIC | NEG | | | 413923 | 50250.9 | 412410 | 55288.2 | 8.35E-01 | -0.03 | 0.9 |
| RP_NEG_227_12917a8_106 | | 227.1292 | 8.11 | | RP | NEG | | | 225528 | 95550.9 | 222895 | 90008.4 | 8.35E-01 | -0.03 | 0.35 |

**Supplemantary table 1**: The non-targeted metabolomics results ordered by the VIP value (p value < 0.0003 are shaded dark green, p value < 0.05 are shaded light green, d values below 0 are blue and above 0 are red)

| | | | | | | | | | Cases | | Controls | | Welch test | Cohen's | |
|---|---|---|---|---|---|---|---|---|---|---|---|---|---|---|---|
| Feature_ID | Metabolite_name | Average_Mz | Average_RT_min_ | HMDB ID | RP_HILIC | POS_NEG | Cureted_MSMS | ID_level | Mean | SD | Mean2 | SD3 | p | d | VIP |
| RP_NEG_531_19525a10_893 | | 531.1953 | 10.89 | | RP | NEG | | | 234426 | 97699.8 | 231573 | 101787 | 8.35E-01 | -0.03 | 0.9 |
| HILIC_NEG_100_93361a7_388 | | 100.9336 | 7.39 | | HILIC | NEG | | | 1.9E+07 | 5172066 | 1.9E+07 | 5440737 | 8.36E-01 | 0.03 | 0.52 |
| HILIC_NEG_341_00409a1_361 | | 341.0041 | 1.36 | | HILIC | NEG | | | 823943 | 257250 | 831481 | 273006 | 8.36E-01 | 0.03 | 0.35 |
| RP_NEG_417_64307a15_493 | | 417.6431 | 15.49 | | RP | NEG | | | 225778 | 50927.9 | 227179 | 48153.6 | 8.36E-01 | 0.03 | 0.56 |
| RP_NEG_528_2641a7_292 | | 528.2641 | 7.29 | | RP | NEG | | | 869057 | 2454802 | 818480 | 872639 | 8.36E-01 | -0.03 | 0.81 |
| HILIC_NEG_397_88928a4_472 | | 397.8893 | 4.47 | | HILIC | NEG | | | 889940 | 311672 | 898821 | 318589 | 8.37E-01 | 0.03 | 0.35 |
| RP_NEG_193_0873a7_647 | | 193.0873 | 7.65 | | RP | NEG | | | 1.5E+07 | 404433 | 1.5E+07 | 414108 | 8.37E-01 | 0.03 | 0.59 |
| RP_POS_313_23712a10_718 | | 313.2371 | 10.72 | | RP | POS | | | 1534990 | 835869 | 1557371 | 763264 | 8.37E-01 | 0.03 | 0.9 |
| RP_POS_408_33148a10_659 | | 408.3315 | 10.66 | | RP | POS | | | 769675 | 113880 | 772940 | 118270 | 8.37E-01 | 0.03 | 0.57 |
| HILIC_POS_152_05424a4_604 | | 152.0542 | 4.60 | | HILIC | POS | | | 673000 | 262499 | 665897 | 245074 | 8.37E-01 | -0.03 | 0.54 |
| RP_NEG_243_96843a0_685 | | 243.9684 | 0.69 | | RP | NEG | | | 7197996 | 1483240 | 7159050 | 1298646 | 8.37E-01 | -0.03 | 0.77 |
| RP_POS_655_62231a11_386 | | 655.6223 | 11.39 | | RP | POS | | | 178312 | 153131 | 174384 | 126449 | 8.37E-01 | -0.03 | 0.8 |
| RP_NEG_279_23337a10_31 | | 279.2334 | 10.31 | | RP | NEG | | | 289475 | 93784.8 | 292251 | 103516 | 8.38E-01 | 0.03 | 0.66 |
| RP_POS_451_65704a9_924 | | 451.6570 | 9.92 | | RP | POS | | | 231180 | 156972 | 235490 | 152012 | 8.38E-01 | 0.03 | 0.46 |
| RP_POS_286_20123a5_398 | | 286.2012 | 5.40 | | RP | POS | 286.2011(100), 85.0291(100) | | 2571186 | 923828 | 2542370 | 1116938 | 8.38E-01 | -0.03 | 0.35 |
| RP_NEG_608_34479a9_118 | | 608.3448 | 9.12 | | RP | NEG | | | 255448 | 241130 | 249094 | 213927 | 8.38E-01 | -0.03 | 0.53 |
| RP_POS_375_32306a10_717 | | 375.3231 | 10.72 | | RP | POS | | | 154017 | 175939 | 148405 | 220442 | 8.38E-01 | -0.03 | 0.68 |
| RP_POS_482_29343a9_113 | | 482.2934 | 9.11 | | RP | POS | | | 868433 | 1560744 | 819464 | 1889440 | 8.38E-01 | -0.03 | 0.54 |
| RP_POS_548_50226a14_252 | | 548.5023 | 14.25 | | RP | POS | | | 2010368 | 927528 | 1984951 | 891088 | 8.38E-01 | -0.03 | 0.34 |
| RP_POS_564_89594a9_775 | | 564.8959 | 9.78 | | RP | POS | | | 1441967 | 708520 | 1423269 | 636129 | 8.38E-01 | -0.03 | 0.81 |
| RP_POS_655_94a10_102 | | 655.9400 | 10.10 | | RP | POS | | | 1947719 | 868590 | 1920054 | 1082384 | 8.38E-01 | -0.03 | 0.62 |
| HILIC_POS_286_20139a0_938 | Octenoyl-L-carnitine | 286.2014 | 0.94 | | HILIC | POS | 85.0291(100), 286.2001(40) | 2 | 1.3E+07 | 6001864 | 1.3E+07 | 6733864 | 8.38E-01 | 0.03 | 0.23 |
| HILIC_POS_232_12891a5_796 | | 232.1289 | 5.80 | | HILIC | POS | | | 127729 | 115556 | 131241 | 135430 | 8.39E-01 | 0.03 | 1.23 |
| RP_NEG_331_19189a6_981 | | 331.1919 | 6.98 | | RP | NEG | | | 586952 | 204857 | 593379 | 252519 | 8.39E-01 | 0.03 | 0.39 |
| RP_POS_215_16357a7_138 | | 215.1636 | 7.14 | | RP | POS | | | 405203 | 47818.1 | 406599 | 52156.1 | 8.39E-01 | 0.03 | 0.76 |
| RP_POS_598_28143a6_983 | | 598.2814 | 6.98 | | RP | POS | | | 797978 | 232693 | 805163 | 279186 | 8.39E-01 | 0.03 | 0.39 |
| HILIC_NEG_115_92095a7_995 | | 115.9210 | 8.00 | | HILIC | NEG | | | 3E+07 | 1.8E+07 | 3E+07 | 1.7E+07 | 8.39E-01 | -0.03 | 0.8 |
| HILIC_POS_287_04407a3_147 | | 287.0441 | 3.15 | | HILIC | POS | | | 218794 | 169969 | 214124 | 167480 | 8.39E-01 | -0.03 | 0.69 |
| HILIC_POS_58_06614a1_468 | | 58.0661 | 1.47 | | HILIC | POS | | | 3315082 | 1747453 | 3266028 | 1772868 | 8.39E-01 | -0.03 | 0.43 |
| RP_POS_568_29565a5_651 | | 568.2957 | 5.65 | | RP | POS | | | 395648 | 168141 | 391371 | 140234 | 8.39E-01 | -0.03 | 0.64 |
| RP_NEG_181_08694a6_202 | | 181.0869 | 6.20 | | RP | NEG | | | 1760540 | 90525.5 | 1762900 | 80651.6 | 8.40E-01 | 0.03 | 1.01 |
| RP_NEG_248_96086a14_338 | | 248.9609 | 14.34 | | RP | NEG | | | 5489004 | 1245362 | 5525124 | 1356488 | 8.40E-01 | 0.03 | 0.44 |
| RP_NEG_428_18591a11_143 | | 428.1859 | 11.14 | | RP | NEG | | | 260693 | 37772.5 | 259643 | 38099.2 | 8.40E-01 | -0.03 | 1.13 |
| RP_NEG_664_4223a10_722 | | 664.4223 | 10.72 | | RP | NEG | | | 28234.3 | 47532.6 | 27127.4 | 32783 | 8.40E-01 | -0.03 | 0.67 |
| RP_POS_153_94798a15_483 | | 153.9480 | 15.48 | | RP | POS | | | 537724 | 94720 | 534690 | 121669 | 8.40E-01 | -0.03 | 0.55 |
| RP_POS_402_77322a13_118 | | 402.7732 | 13.12 | | RP | POS | | | 5022519 | 3857630 | 4914535 | 3934587 | 8.40E-01 | -0.03 | 0.82 |
| RP_POS_540_44568a10_926 | | 540.4457 | 10.93 | | RP | POS | | | 7030707 | 599711 | 7013056 | 668767 | 8.40E-01 | -0.03 | 0.47 |
| RP_POS_543_44391a11_337 | | 543.4439 | 11.34 | | RP | POS | | | 806631 | 1405471 | 772505 | 1079415 | 8.40E-01 | -0.03 | 0.44 |
| RP_POS_175_11166a8_085 | | 175.1117 | 8.09 | | RP | POS | | | 5290828 | 435222 | 5303040 | 452082 | 8.41E-01 | 0.03 | 0.44 |
| RP_POS_247_0849a15_382 | | 247.0849 | 15.38 | | RP | POS | | | 697285 | 318891 | 706848 | 370443 | 8.41E-01 | 0.03 | 0.44 |
| RP_POS_791_03656a12_73 | | 791.0366 | 12.73 | | RP | POS | | | 1760089 | 1015750 | 1788287 | 1036523 | 8.41E-01 | 0.03 | 0.46 |
| RP_POS_182_05794a1_789 | | 182.0579 | 1.79 | | RP | POS | | | 654206 | 164074 | 649787 | 157861 | 8.41E-01 | -0.03 | 0.47 |
| RP_POS_252_04482a9_337 | | 252.0448 | 9.34 | | RP | POS | | | 1589624 | 585025 | 1572331 | 667265 | 8.41E-01 | -0.03 | 0.24 |
| RP_POS_430_3891a9_691 | | 430.3891 | 9.69 | | RP | POS | | | 1274642 | 3623806 | 1199803 | 1637655 | 8.41E-01 | -0.03 | 0.28 |
| RP_POS_628_49957a10_867 | | 628.4996 | 10.87 | | RP | POS | | | 3441984 | 320751 | 3432479 | 365666 | 8.41E-01 | -0.03 | 0.93 |
| RP_POS_363_21658a6_982 | Hydrocortisone | 363.2166 | 6.98 | | RP | POS | 363.2177(100), 121.0654(95) | 2 | 2.9E+07 | 9483398 | 2.9E+07 | 1.2E+07 | 8.41E-01 | 0.03 | 0.42 |
| HILIC_NEG_115_92082a4_254 | | 115.9208 | 4.25 | | HILIC | NEG | | | 5E+07 | 1.2E+07 | 5E+07 | 1.1E+07 | 8.42E-01 | 0.03 | 0.76 |
| RP_NEG_1185_15503a9_765 | | 1185.1550 | 9.77 | | RP | NEG | | | 1608373 | 217894 | 1614104 | 202706 | 8.42E-01 | 0.03 | 0.32 |
| RP_POS_159_98201a15_411 | | 159.9820 | 15.41 | | RP | POS | | | 2596902 | 1102633 | 2625956 | 1038654 | 8.42E-01 | 0.03 | 0.6 |
| RP_POS_231_07838a8_469 | | 231.0784 | 8.47 | | RP | POS | | | 934324 | 771698 | 957099 | 885515 | 8.42E-01 | 0.03 | 0.4 |
| RP_POS_508_85696a10_43 | | 508.8570 | 10.43 | | RP | POS | | | 801048 | 454151 | 814267 | 512305 | 8.42E-01 | 0.03 | 0.78 |
| RP_POS_703_57666a12_807 | | 703.5767 | 12.81 | | RP | POS | | | 2E+08 | 4.8E+07 | 2.1E+08 | 4.1E+07 | 8.42E-01 | 0.03 | 1.06 |
| RP_POS_431_1777a12_373 | | 431.1777 | 12.37 | | RP | POS | | | 1.3E+07 | 3305098 | 1.3E+07 | 3337448 | 8.42E-01 | -0.03 | 0.6 |
| RP_POS_609_41559a13_555 | | 609.4156 | 13.56 | | RP | POS | | | 1.6E+07 | 5582361 | 1.5E+07 | 5501476 | 8.42E-01 | -0.03 | 0.51 |
| HILIC_NEG_201_03802a1_34 | | 201.0380 | 1.34 | | HILIC | NEG | | | 1293736 | 327894 | 1302294 | 305386 | 8.43E-01 | 0.03 | 0.64 |
| RP_NEG_159_89404a14_342 | | 159.8940 | 14.34 | | RP | NEG | | | 1363153 | 527897 | 1376977 | 494578 | 8.43E-01 | 0.03 | 0.42 |
| RP_NEG_292_98322a7_223 | | 292.9832 | 7.22 | | RP | NEG | | | 1605328 | 358082 | 1615160 | 368985 | 8.43E-01 | 0.03 | 0.67 |
| RP_NEG_506_32541a10_976 | | 506.3254 | 10.98 | | RP | NEG | | | 280877 | 83119.8 | 283145 | 84715.5 | 8.43E-01 | 0.03 | 0.5 |
| RP_NEG_929_1059a9_348 | | 929.1059 | 9.35 | | RP | NEG | | | 1287544 | 98700.7 | 1290202 | 98150.1 | 8.43E-01 | 0.03 | 1 |
| RP_POS_265_02295a0_548 | | 265.0230 | 0.55 | | RP | POS | | | 890857 | 315525 | 899883 | 347296 | 8.43E-01 | 0.03 | 1.12 |
| RP_POS_327_1734a3_26 | | 327.1734 | 3.26 | | RP | POS | | | 745685 | 349082 | 754558 | 307955 | 8.43E-01 | 0.03 | 0.66 |
| RP_NEG_249_11311a8_789 | | 249.1131 | 8.79 | | RP | NEG | | | 232815 | 36395.2 | 231883 | 32497.9 | 8.43E-01 | -0.03 | 0.65 |
| RP_NEG_473_28265a12_385 | | 473.2827 | 12.39 | | RP | NEG | | | 121814 | 82610.8 | 119799 | 66918 | 8.43E-01 | -0.03 | 0.46 |
| RP_NEG_637_1424a9_699 | | 637.1424 | 9.70 | | RP | NEG | | | 2878041 | 247496 | 2871534 | 232799 | 8.43E-01 | -0.03 | 0.33 |
| HILIC_NEG_200_85916a6_463 | | 200.8592 | 6.46 | | HILIC | NEG | | | 1605277 | 818531 | 1628766 | 915970 | 8.44E-01 | 0.03 | 0.47 |
| RP_POS_227_12732a3_535 | | 227.1273 | 3.54 | | RP | POS | | | 130500 | 87449.1 | 132868 | 88962 | 8.44E-01 | 0.03 | 0.94 |
| RP_POS_325_16159a8_584 | | 325.1616 | 8.58 | | RP | POS | | | 1313685 | 465003 | 1326308 | 475268 | 8.44E-01 | 0.03 | 0.97 |
| RP_NEG_1175_77917a12_606 | | 1175.7792 | 12.61 | | RP | NEG | | | 2619376 | 3533424 | 2526828 | 3350781 | 8.44E-01 | -0.03 | 0.5 |
| RP_NEG_243_96761a0_862 | | 243.9676 | 0.86 | | RP | NEG | | | 1663392 | 1061743 | 1636561 | 934105 | 8.44E-01 | -0.03 | 0.73 |
| RP_POS_249_18262a8_299 | | 249.1826 | 8.30 | | RP | POS | | | 738611 | 285672 | 731140 | 269750 | 8.44E-01 | -0.03 | 0.87 |
| RP_POS_386_15146a9_035 | | 386.1515 | 9.04 | | RP | POS | | | 632683 | 112953 | 629079 | 149097 | 8.44E-01 | -0.03 | 0.93 |
| RP_POS_441_33295a12_229 | | 441.3330 | 12.23 | | RP | POS | | | 2180368 | 506266 | 2167354 | 461645 | 8.44E-01 | -0.03 | 1.03 |
| HILIC_NEG_190_85938a0_919 | | 190.8594 | 0.92 | | HILIC | NEG | | | 298420 | 69512.4 | 300304 | 71183 | 8.45E-01 | 0.03 | 0.24 |
| HILIC_POS_265_03476a4_158 | | 265.0348 | 4.16 | | HILIC | POS | | | 161118 | 57576.1 | 162690 | 59735.6 | 8.45E-01 | 0.03 | 0.4 |
| RP_POS_459_14719a4_825 | | 459.1472 | 4.83 | | RP | POS | | | 536468 | 818213 | 559474 | 895779 | 8.45E-01 | 0.03 | 0.69 |
| HILIC_NEG_289_06778a1_452 | | 289.0678 | 1.45 | | HILIC | NEG | | | 497065 | 148041 | 493563 | 113827 | 8.45E-01 | -0.03 | 0.73 |
| RP_POS_345_17505a9_208 | | 345.1751 | 9.21 | | RP | POS | | | 550594 | 302115 | 539612 | 483922 | 8.45E-01 | -0.03 | 0.93 |
| RP_POS_387_25275a8_452 | | 387.2528 | 8.45 | | RP | POS | | | 859642 | 620532 | 845204 | 459595 | 8.45E-01 | -0.03 | 0.72 |
| RP_POS_801_03827a13_201 | | 801.0383 | 13.20 | | RP | POS | | | 1270095 | 812645 | 1249994 | 692898 | 8.45E-01 | -0.03 | 0.7 |
| HILIC_POS_144_10202a2_888 | | 144.1020 | 2.89 | | HILIC | POS | 144.1026(100), 98.096(5) | | 771834 | 713832 | 791354 | 755864 | 8.46E-01 | 0.03 | 0.66 |
| RP_NEG_643_29077a10_755 | | 643.2908 | 10.76 | | RP | NEG | | | 77704.5 | 64991.4 | 79299.5 | 55368.6 | 8.46E-01 | 0.03 | 0.32 |
| RP_POS_557_88116a14_344 | | 557.8812 | 14.34 | | RP | POS | | | 3.4E+07 | 9.7E+07 | 3.6E+07 | 5.4E+07 | 8.46E-01 | 0.03 | 0.82 |
| RP_POS_784_58124a14_36 | | 784.5812 | 14.36 | | RP | POS | | | 2432443 | 1602286 | 2470821 | 1292367 | 8.46E-01 | 0.03 | 0.74 |
| HILIC_POS_150_11235a6_161 | | 150.1124 | 6.16 | | HILIC | POS | 150.1125(100), 132.1046(40) | | 5475954 | 1922749 | 5425092 | 1915886 | 8.46E-01 | -0.03 | 0.65 |
| HILIC_POS_162_05835a4_763 | | 162.0584 | 4.76 | | HILIC | POS | | | 2348572 | 5115790 | 2211281 | 5216111 | 8.46E-01 | -0.03 | 0.56 |
| RP_NEG_289_10602a7_576 | | 289.1060 | 7.58 | | RP | NEG | | | 498200 | 27712.2 | 497461 | 28074.3 | 8.46E-01 | -0.03 | 0.55 |
| HILIC_NEG_257_81863a3_098 | | 257.8186 | 3.10 | | HILIC | NEG | | | 2398454 | 165540 | 2403039 | 179554 | 8.47E-01 | 0.03 | 0.74 |
| RP_POS_254_17506a8_42 | | 254.1751 | 8.42 | | RP | POS | | | 1401045 | 170343 | 1405785 | 186379 | 8.47E-01 | 0.03 | 0.99 |
| RP_POS_320_11008a15_519 | | 320.1101 | 15.52 | | RP | POS | | | 1286284 | 388494 | 1296243 | 369055 | 8.47E-01 | 0.03 | 0.76 |
| RP_POS_746_50958a13_359 | | 746.5096 | 13.36 | | RP | POS | | | 1425712 | 680522 | 1407956 | 668781 | 8.47E-01 | -0.03 | 0.56 |
| RP_POS_782_56897a14_364 | PC 36:4 (18:2/18:2) | 782.5690 | 14.36 | | RP | POS | 184.0744(100), 782.5809(2) | 1 | 3022419 | 2202266 | 3078654 | 2078534 | 8.47E-01 | 0.03 | 0.51 |
| HILIC_POS_166_08614a3_908 | | 166.0861 | 3.91 | | HILIC | POS | 120.0807(100), 166.0871(10) | | 1.9E+08 | 3.9E+07 | 1.9E+08 | 3.9E+07 | 8.48E-01 | 0.03 | 0.52 |
| HILIC_POS_386_17862a2_434 | | 386.1786 | 2.43 | | HILIC | POS | | | 188934 | 91715.9 | 191544 | 106271 | 8.48E-01 | 0.03 | 0.54 |
| RP_NEG_1149_24304a10_064 | | 1149.2430 | 10.06 | | RP | NEG | | | 2267043 | 1332109 | 2298399 | 1070209 | 8.48E-01 | 0.03 | 0.49 |
| RP_POS_431_26385a9_629 | | 431.2639 | 9.63 | | RP | POS | | | 479150 | 213984 | 485018 | 231408 | 8.48E-01 | 0.03 | 1.01 |
| HILIC_POS_70_06599a4_858 | | 70.0660 | 4.86 | | HILIC | POS | | | 3E+07 | 9092388 | 2.9E+07 | 8478310 | 8.48E-01 | -0.03 | 0.68 |
| RP_NEG_168_83649a11_139 | | 168.8365 | 11.14 | | RP | NEG | | | 130266 | 41666.3 | 129123 | 45244.3 | 8.48E-01 | -0.03 | 0.72 |
| RP_NEG_378_24176a10_028 | | 378.2418 | 10.03 | | RP | NEG | | | 2142544 | 822283 | 2121159 | 810041 | 8.48E-01 | -0.03 | 0.59 |
| RP_POS_290_91821a15_406 | | 290.9182 | 15.41 | | RP | POS | | | 2161877 | 642824 | 2144469 | 681912 | 8.48E-01 | -0.03 | 0.66 |
| HILIC_POS_132_0657a7_746 | 5-Aminolevulinic acid | 132.0657 | 7.75 | | HILIC | POS | 114.0553(100), 86.06(70) | 2 | 733753 | 81422.3 | 736052 | 92766.1 | 8.48E-01 | 0.03 | 0.4 |
| HILIC_NEG_142_94437a1_743 | | 142.9444 | 1.74 | | HILIC | NEG | | | 622998 | 653196 | 639837 | 637744 | 8.49E-01 | 0.03 | 0.47 |
| RP_POS_255_23166a10_518 | | 255.2317 | 10.52 | | RP | POS | | | 586377 | 740309 | 606945 | 830136 | 8.49E-01 | 0.03 | 0.52 |
| RP_POS_416_3645a12_743 | | 416.3645 | 12.74 | | RP | POS | | | 718008 | 439640 | 727830 | 317586 | 8.49E-01 | 0.03 | 0.6 |
| RP_POS_449_3606a12_308 | | 449.3606 | 12.31 | | RP | POS | | | 1357724 | 163065 | 1362052 | 168196 | 8.49E-01 | 0.03 | 0.71 |
| HILIC_NEG_200_85922a7_019 | | 200.8592 | 7.02 | | HILIC | NEG | | | 1900415 | 831259 | 1879471 | 779356 | 8.49E-01 | -0.03 | 0.7 |
| RP_NEG_334_78592a15_372 | | 334.7859 | 15.37 | | RP | NEG | | | 6721731 | 918171 | 6696611 | 1000308 | 8.49E-01 | -0.03 | 0.8 |
| RP_POS_502_3273a10_69 | | 502.3273 | 10.69 | | RP | POS | | | 5512199 | 1598258 | 5471340 | 1539350 | 8.49E-01 | -0.03 | 0.56 |
| RP_POS_588_17487a8_766 | | 588.1749 | 8.77 | | RP | POS | | | 862333 | 78901.6 | 860357 | 73372.8 | 8.49E-01 | -0.03 | 0.98 |
| RP_POS_364_25653a12_509 | | 364.2565 | 12.51 | | RP | POS | | | 372158 | 236717 | 365732 | 259021 | 8.50E-01 | -0.03 | 0.42 |
| RP_POS_457_98053a10_097 | | 457.9805 | 10.10 | | RP | POS | | | 398524 | 242503 | 392791 | 203026 | 8.50E-01 | -0.03 | 0.58 |
| RP_POS_540_42566a12_093 | | 540.4257 | 12.09 | | RP | POS | | | 556001 | 347753 | 546576 | 377926 | 8.50E-01 | -0.03 | 0.55 |
| RP_POS_666_33282a5_653 | | 666.3328 | 5.65 | | RP | POS | | | 1886702 | 729548 | 1868852 | 659262 | 8.50E-01 | -0.03 | 0.76 |
| HILIC_NEG_186_93405a1_743 | | 186.9341 | 1.74 | | HILIC | NEG | | | 2001322 | 2076602 | 2054263 | 2039405 | 8.51E-01 | 0.03 | 0.47 |
| RP_NEG_174_95627a13_29 | | 174.9563 | 13.29 | | RP | NEG | | | 1977995 | 498098 | 1990462 | 476105 | 8.51E-01 | 0.03 | 0.48 |
| RP_NEG_380_73422a15_429 | | 380.7342 | 15.43 | | RP | NEG | | | 1332410 | 459594 | 1344382 | 470479 | 8.51E-01 | 0.03 | 0.79 |
| RP_POS_268_26349a10_603 | | 268.2635 | 10.60 | | RP | POS | | | 112216 | 171271 | 116777 | 182385 | 8.51E-01 | 0.03 | 0.92 |
| RP_POS_366_26367a10_515 | | 366.2637 | 10.52 | | RP | POS | | | 1413909 | 798243 | 1433314 | 714293 | 8.51E-01 | 0.03 | 0.82 |
| HILIC_POS_306_66458a1_589 | | 306.6646 | 1.59 | | HILIC | POS | | | 150212 | 94440.3 | 147963 | 81490.6 | 8.51E-01 | -0.03 | 0.68 |
| RP_NEG_421_22815a11_509 | | 421.2282 | 11.51 | | RP | NEG | | | 59978.7 | 34845.5 | 59052.3 | 37234.9 | 8.51E-01 | -0.03 | 0.43 |
| RP_NEG_628_72894a6_088 | | 628.7289 | 6.09 | | RP | NEG | | | 797633 | 257038 | 790395 | 300490 | 8.51E-01 | -0.03 | 0.51 |
| RP_POS_172_97719a0_693 | | 172.9772 | 0.69 | | RP | POS | | | 8172311 | 5359292 | 8045014 | 4575500 | 8.51E-01 | -0.03 | 0.54 |
| RP_NEG_354_84494a15_399 | | 354.8449 | 15.40 | | RP | NEG | | | 950937 | 206548 | 956517 | 228798 | 8.52E-01 | 0.03 | 0.73 |
| RP_POS_198_99219a2_743 | | 198.9922 | 2.74 | | RP | POS | | | 2054496 | 540257 | 2068049 | 524154 | 8.52E-01 | 0.03 | 0.68 |
| RP_POS_417_36844a11_181 | | 417.3684 | 11.18 | | RP | POS | | | 197458 | 167894 | 206124 | 435524 | 8.52E-01 | 0.03 | 0.54 |
| RP_POS_443_80811a9_86 | | 443.8081 | 9.86 | | RP | POS | | | 1685958 | 781705 | 1707610 | 902998 | 8.52E-01 | 0.03 | 0.49 |
| RP_POS_681_07294a11_182 | | 681.0729 | 11.18 | | RP | POS | | | 141096 | 64103.1 | 142890 | 75292.7 | 8.52E-01 | 0.03 | 0.47 |
| RP_POS_930_23889a9_63 | | 930.2389 | 9.63 | | RP | POS | | | 1601160 | 166467 | 1605601 | 179375 | 8.52E-01 | 0.03 | 0.9 |
| HILIC_NEG_260_95578a3_987 | | 260.9558 | 3.99 | | HILIC | NEG | | | 150349 | 68127.7 | 148764 | 56926.6 | 8.52E-01 | -0.03 | 0.46 |
| RP_POS_491_34558a9_729 | | 491.3456 | 9.73 | | RP | POS | | | 180803 | 110975 | 177228 | 160009 | 8.52E-01 | -0.03 | 0.48 |
| RP_POS_577_5174a11_507 | | 577.5174 | 11.51 | | RP | POS | | | 346464 | 379473 | 336937 | 367211 | 8.52E-01 | -0.03 | 0.35 |
| RP_POS_732_54688a10_458 | | 732.5469 | 10.46 | | RP | POS | | | 680181 | 176045 | 675578 | 183213 | 8.52E-01 | -0.03 | 0.43 |
| RP_POS_816_64081a11_828 | | 816.6408 | 11.83 | | RP | POS | | | 384989 | 429238 | 375391 | 326834 | 8.52E-01 | -0.03 | 0.39 |
| RP_NEG_1059_66504a10_352 | | 1059.6650 | 10.35 | | RP | NEG | | | 1362758 | 516190 | 1375989 | 526993 | 8.53E-01 | 0.03 | 0.68 |
| RP_POS_285_25757a8_104 | | 285.2576 | 8.10 | | RP | POS | | | 377828 | 401038 | 388432 | 431491 | 8.53E-01 | 0.03 | 0.8 |
| RP_POS_674_97345a11_511 | | 674.9735 | 11.51 | | RP | POS | | | 207063 | 135783 | 210685 | 148323 | 8.53E-01 | 0.03 | 0.58 |
| RP_NEG_148_8999a15_405 | | 148.8999 | 15.41 | | RP | NEG | | | 457618 | 191005 | 452384 | 219316 | 8.53E-01 | -0.03 | 0.34 |
| RP_NEG_459_59714a15_497 | | 459.5971 | 15.50 | | RP | NEG | | | 905744 | 72152.3 | 903924 | 71530.6 | 8.53E-01 | -0.03 | 0.49 |
| RP_NEG_309_17462a10_278 | | 309.1746 | 10.28 | | RP | NEG | 309.1757(100), 96.9602(45) | | 628624 | 512210 | 641115 | 481078 | 8.54E-01 | 0.03 | 0.62 |
| HILIC_NEG_234_82031a4_283 | | 234.8203 | 4.28 | | HILIC | NEG | | | 339481 | 55829.8 | 340852 | 53502.7 | 8.54E-01 | 0.03 | 0.65 |
| HILIC_POS_178_08611a4_746 | | 178.0861 | 4.75 | | HILIC | POS | | | 674320 | 2971305 | 751503 | 3165587 | 8.54E-01 | 0.03 | 0.32 |
| RP_NEG_457_23666a10_697 | | 457.2367 | 10.70 | | RP | NEG | | | 455946 | 1166824 | 488757 | 1406070 | 8.54E-01 | 0.03 | 0.21 |
| RP_POS_357_68604a5_52 | | 357.6860 | 5.52 | | RP | POS | | | 1175274 | 208823 | 1180541 | 210290 | 8.54E-01 | 0.03 | 0.74 |
| RP_NEG_151_0401a5 | | 151.0401 | 5.00 | | RP | NEG | | | 531799 | 1144213 | 506727 | 854337 | 8.54E-01 | -0.03 | 0.34 |
| RP_POS_134_95872a15_464 | | 134.9587 | 15.46 | | RP | POS | | | 9766710 | 3076908 | 9691724 | 2888861 | 8.54E-01 | -0.03 | 0.31 |
| RP_POS_312_16479a4_312 | | 312.1648 | 4.31 | | RP | POS | | | 104244 | 39803.1 | 103146 | 46661.4 | 8.54E-01 | -0.03 | 0.4 |
| RP_POS_223_10715a2_816 | | 223.1072 | 2.82 | | RP | POS | | | 721257 | 295433 | 728405 | 276024 | 8.55E-01 | 0.03 | 0.47 |
| HILIC_POS_174_12378a1_885 | | 174.1238 | 1.89 | | HILIC | POS | | | 1458560 | 1025383 | 1482074 | 868112 | 8.55E-01 | 0.02 | 0.48 |
| RP_POS_527_40173a11_756 | | 527.4017 | 11.76 | | RP | POS | | | 526827 | 352843 | 534662 | 274131 | 8.55E-01 | 0.02 | 0.18 |
| RP_POS_233_98232a0_68 | | 233.9823 | 0.68 | | RP | POS | | | 2656511 | 615007 | 2642259 | 531476 | 8.55E-01 | -0.02 | 0.67 |
| HILIC_NEG_194_98898a1_351 | | 194.9890 | 1.35 | | HILIC | NEG | | | 1906451 | 564880 | 1889547 | 758197 | 8.55E-01 | -0.03 | 0.18 |
| RP_POS_587_43372a13_55 | | 587.4337 | 13.55 | | RP | POS | | | 1291547 | 507443 | 1277857 | 583026 | 8.55E-01 | -0.03 | 0.54 |
| RP_POS_631_40265a15_167 | | 631.4027 | 15.17 | | RP | POS | | | 917393 | 754764 | 896383 | 902503 | 8.55E-01 | -0.03 | 0.77 |
| RP_POS_694_49542a13_696 | | 694.4954 | 13.70 | | RP | POS | | | 1.3E+07 | 3124844 | 1.3E+07 | 3190487 | 8.55E-01 | -0.03 | 0.89 |
| RP_POS_530_36969a11_126 | | 530.3697 | 11.13 | | RP | POS | | | 124701 | 260532 | 132372 | 346390 | 8.56E-01 | 0.03 | 0.58 |
| RP_POS_282_27966a15_281 | | 282.2797 | 15.28 | | RP | POS | 282.2813(100), 69.0709(70) | | 2918079 | 1261884 | 2949633 | 1276121 | 8.56E-01 | 0.02 | 0.4 |
| HILIC_NEG_313_0007a3_933 | | 313.0007 | 3.93 | | HILIC | NEG | | | 175462 | 74865.7 | 177306 | 73579.7 | 8.56E-01 | 0.02 | 0.36 |
| RP_NEG_339_19937a13_16 | | 339.1994 | 13.16 | | RP | NEG | | | 841175 | 703160 | 855804 | 468956 | 8.56E-01 | 0.02 | 0.21 |
| RP_POS_320_17017a6_227 | | 320.1702 | 6.23 | | RP | POS | | | 2.5E+07 | 2647912 | 2.5E+07 | 2696400 | 8.56E-01 | 0.02 | 0.89 |
| RP_POS_300_91492a15_354 | | 300.9149 | 15.35 | | RP | POS | | | 140902 | 67802.1 | 139396 | 53746.9 | 8.56E-01 | -0.02 | 0.42 |
| HILIC_POS_218_09241a4_08 | | 218.0924 | 4.08 | | HILIC | POS | | | 299194 | 266474 | 305632 | 256788 | 8.57E-01 | 0.02 | 0.5 |
| RP_NEG_199_03789a0_585 | | 199.0379 | 0.59 | | RP | NEG | | | 4043464 | 1042164 | 4072024 | 1254526 | 8.57E-01 | 0.02 | 0.26 |
| RP_POS_422_25778a13_214 | | 422.2578 | 13.21 | | RP | POS | | | 431283 | 336190 | 438926 | 286347 | 8.57E-01 | 0.02 | 0.6 |

**Supplemantary table 1**: The non-targeted metabolomics results ordered by the VIP value (p value < 0.0003 are shaded dark green, p value < 0.05 are shaded light green, d values below 0 are blue and above 0 are red)

| | | | | | | | | | Cases | | Controls | | Welch test | Cohen's | |
|---|---|---|---|---|---|---|---|---|---|---|---|---|---|---|---|
| Feature_ID | Metabolite_name | Average_Mz | Average_RT_min_ | HMDB ID | RP_HILIC | POS_NEG | Cureted_MSMS | ID_level | Mean | SD | Mean2 | SD3 | p | d | VIP |
| RP_POS_473_8175a9_389 | | 473.8175 | 9.39 | | RP | POS | | | 339260 | 181289 | 343988 | 200053 | 8.57E-01 | 0.02 | 0.65 |
| RP_POS_330_06879a15_379 | | 330.0688 | 15.38 | | RP | POS | | | 2.7E+07 | 8726929 | 2.7E+07 | 8780410 | 8.57E-01 | -0.02 | 0.55 |
| RP_POS_548_50537a15_185 | | 548.5054 | 15.19 | | RP | POS | | | 2E+08 | 2.4E+07 | 2E+08 | 2.2E+07 | 8.57E-01 | -0.02 | 0.83 |
| RP_POS_449_15274a9_345 | | 449.1527 | 9.35 | | RP | POS | | | 1264933 | 482823 | 1250585 | 657759 | 8.57E-01 | -0.03 | 0.36 |
| RP_POS_223_10719a2_362 | | 223.1072 | 2.36 | | RP | POS | | | 797372 | 399131 | 807075 | 395762 | 8.58E-01 | 0.02 | 0.53 |
| RP_POS_294_64404a10_494 | | 294.6440 | 10.49 | | RP | POS | | | 993313 | 517376 | 1004933 | 437509 | 8.58E-01 | 0.02 | 0.71 |
| RP_POS_300_19907a8_01 | | 300.1991 | 8.01 | | RP | POS | | | 908122 | 3312069 | 988645 | 3295104 | 8.58E-01 | 0.02 | 0.38 |
| HILIC_NEG_162_83929a3_186 | | 162.8393 | 3.19 | | HILIC | NEG | | | 3.2E+07 | 3113069 | 3.2E+07 | 3052142 | 8.58E-01 | -0.02 | 0.19 |
| RP_NEG_445_10431a8_647 | | 445.1043 | 8.65 | | RP | NEG | | | 993695 | 81213.3 | 991712 | 81106.1 | 8.58E-01 | -0.02 | 0.81 |
| RP_NEG_463_34439a12_227 | | 463.3444 | 12.23 | | RP | NEG | | | 141809 | 49419.8 | 140498 | 56945.3 | 8.58E-01 | -0.02 | 0.63 |
| RP_POS_808_57532a12_785 | | 808.5753 | 12.79 | | RP | POS | | | 9881847 | 4122962 | 9789844 | 3423882 | 8.58E-01 | -0.02 | 0.53 |
| RP_POS_141_00066a0_634 | Phosphonoacetic acid | 141.0007 | 0.63 | | RP | POS | 95.0502(100), 53.0396(85), 105.0452(78) | 2 | 2.3E+07 | 5166755 | 2.3E+07 | 6232064 | 8.58E-01 | 0.02 | 0.63 |
| HILIC_POS_211_13286a1_352 | | 211.1329 | 1.35 | | HILIC | POS | | | 796574 | 492254 | 808137 | 464687 | 8.59E-01 | 0.02 | 0.45 |
| RP_NEG_816_53436a13_118 | | 816.5344 | 13.12 | | RP | NEG | | | 3019089 | 615609 | 3034071 | 620750 | 8.59E-01 | 0.02 | 0.7 |
| RP_POS_226_08905a15_51 | | 226.0891 | 15.51 | | RP | POS | | | 758820 | 267832 | 765862 | 307192 | 8.59E-01 | 0.02 | 0.22 |
| RP_POS_344_01529a15_432 | | 344.0153 | 15.43 | | RP | POS | | | 324511 | 203129 | 330048 | 247489 | 8.59E-01 | 0.02 | 0.95 |
| HILIC_NEG_161_91351a0_902 | | 161.9135 | 0.90 | | HILIC | NEG | | | 1155820 | 146280 | 1151670 | 189133 | 8.59E-01 | -0.02 | 0.57 |
| RP_POS_194_15163a7_774 | | 194.1516 | 7.77 | | RP | POS | | | 2308483 | 1414059 | 2276497 | 1228707 | 8.59E-01 | -0.02 | 0.48 |
| HILIC_POS_189_08714a6_579 | | 189.0871 | 6.58 | | HILIC | POS | | | 496908 | 180052 | 501345 | 187359 | 8.60E-01 | 0.02 | 0.44 |
| RP_POS_311_08881a3_984 | | 311.0888 | 3.98 | | RP | POS | | | 988447 | 559947 | 1001532 | 528947 | 8.60E-01 | 0.02 | 1.05 |
| RP_POS_445_36728a11_218 | | 445.3673 | 11.22 | | RP | POS | | | 506906 | 163561 | 510999 | 174257 | 8.60E-01 | 0.02 | 0.61 |
| HILIC_NEG_136_89169a1_571 | | 136.8917 | 1.57 | | HILIC | NEG | | | 4585099 | 3166294 | 4511044 | 2982492 | 8.60E-01 | -0.02 | 0.39 |
| HILIC_POS_253_16438a0_554 | | 253.1644 | 0.55 | | HILIC | POS | | | 1007758 | 341015 | 1000006 | 303570 | 8.60E-01 | -0.02 | 0.57 |
| RP_NEG_205_83961a14_347 | | 205.8396 | 14.35 | | RP | NEG | | | 265010 | 169954 | 261038 | 160148 | 8.60E-01 | -0.02 | 0.32 |
| RP_POS_480_82553a9_605 | | 480.8255 | 9.61 | | RP | POS | | | 1350042 | 802587 | 1332180 | 680277 | 8.60E-01 | -0.02 | 0.5 |
| HILIC_NEG_273_01666a5_115 | | 273.0167 | 5.12 | | HILIC | NEG | | | 217605 | 105421 | 220103 | 103475 | 8.61E-01 | 0.02 | 0.42 |
| RP_POS_131_10667a4_075 | | 131.1067 | 4.08 | | RP | POS | | | 80246 | 63625.5 | 81906.9 | 73751.2 | 8.61E-01 | 0.02 | 0.32 |
| RP_POS_541_29053a7_837 | | 541.2905 | 7.84 | | RP | POS | | | 545506 | 100735 | 548086 | 113649 | 8.61E-01 | 0.02 | 0.64 |
| HILIC_POS_120_06575a1_018 | | 120.0658 | 1.02 | | HILIC | POS | 120.0657(100) | | 1.1E+07 | 5608293 | 1E+07 | 6691538 | 8.61E-01 | -0.02 | 0.52 |
| HILIC_POS_255_09732a4_539 | | 255.0973 | 4.54 | | HILIC | POS | | | 72069.6 | 81218.4 | 69787.2 | 105587 | 8.61E-01 | -0.02 | 0.42 |
| RP_POS_181_98746a15_602 | | 181.9875 | 15.60 | | RP | POS | | | 6753290 | 3675759 | 6667499 | 3499984 | 8.61E-01 | -0.02 | 0.74 |
| RP_POS_462_65341a10_239 | | 462.6534 | 10.24 | | RP | POS | | | 405253 | 192541 | 400580 | 198466 | 8.61E-01 | -0.02 | 0.34 |
| RP_NEG_213_02292a5_479 | | 213.0229 | 5.48 | | RP | NEG | | | 1E+07 | 1.1E+07 | 1E+07 | 1.5E+07 | 8.62E-01 | 0.02 | 0.18 |
| RP_NEG_215_01852a5_478 | | 215.0185 | 5.48 | | RP | NEG | | | 471062 | 497331 | 484946 | 645454 | 8.62E-01 | 0.02 | 0.19 |
| RP_NEG_306_98294a6_39 | | 306.9829 | 6.39 | | RP | NEG | | | 868706 | 139445 | 872338 | 163082 | 8.62E-01 | 0.02 | 0.43 |
| RP_POS_231_11581a14_215 | | 231.1158 | 14.22 | | RP | POS | | | 3292267 | 1899925 | 3334937 | 1689655 | 8.62E-01 | 0.02 | 0.34 |
| RP_POS_806_58307a9_645 | | 806.5831 | 9.65 | | RP | POS | | | 2564898 | 1534767 | 2601711 | 1562190 | 8.62E-01 | 0.02 | 0.26 |
| RP_NEG_957_20587a9_458 | | 957.2059 | 9.46 | | RP | NEG | | | 2186809 | 265125 | 2180656 | 254074 | 8.62E-01 | -0.02 | 0.88 |
| RP_POS_217_07443a15_382 | | 217.0744 | 15.38 | | RP | POS | | | 1662663 | 725663 | 1645029 | 759167 | 8.62E-01 | -0.02 | 0.56 |
| RP_POS_365_26575a11_62 | | 365.2658 | 11.62 | | RP | POS | | | 94498.5 | 65056.5 | 92996 | 61269.2 | 8.62E-01 | -0.02 | 0.52 |
| RP_POS_485_36011a11_957 | | 485.3601 | 11.96 | | RP | POS | | | 555535 | 504528 | 542781 | 561174 | 8.62E-01 | -0.02 | 0.89 |
| RP_POS_570_35583a10_193 | | 570.3558 | 10.19 | | RP | POS | | | 1652192 | 723925 | 1634374 | 774621 | 8.62E-01 | -0.02 | 0.53 |
| RP_NEG_203_08247a2_482 | Tryptophan | 203.0825 | 2.48 | | RP | NEG | 116.0506(100), 203.0836(60), 142.0664(30) | 1 | 1.9E+07 | 3710654 | 1.9E+07 | 3405106 | 8.62E-01 | 0.02 | 0.47 |
| HILIC_NEG_160_84227a3_17 | | 160.8423 | 3.17 | | HILIC | NEG | | | 1.2E+07 | 3577503 | 1.2E+07 | 2636923 | 8.63E-01 | 0.02 | 0.44 |
| HILIC_NEG_244_98738a6_134 | | 244.9874 | 6.13 | | HILIC | NEG | | | 606425 | 138766 | 609603 | 130306 | 8.63E-01 | 0.02 | 0.99 |
| HILIC_POS_100_07616a0_563 | | 100.0762 | 0.56 | | HILIC | POS | | | 6028543 | 8370742 | 6228146 | 8553037 | 8.63E-01 | 0.02 | 0.46 |
| RP_NEG_421_22638a10_828 | | 421.2264 | 10.83 | | RP | NEG | | | 236847 | 268423 | 243113 | 263928 | 8.63E-01 | 0.02 | 0.41 |
| RP_POS_462_30603a9_958 | | 462.3060 | 9.96 | | RP | POS | | | 303102 | 248080 | 309773 | 309687 | 8.63E-01 | 0.02 | 0.61 |
| RP_NEG_540_33105a15_475 | | 540.3311 | 15.48 | | RP | NEG | | | 257166 | 298712 | 264805 | 349182 | 8.64E-01 | 0.02 | 0.62 |
| RP_POS_215_1403a8_917 | | 215.1403 | 8.92 | | RP | POS | | | 623476 | 107638 | 626517 | 146240 | 8.64E-01 | 0.02 | 0.93 |
| HILIC_NEG_136_89169a5_373 | | 136.8917 | 5.37 | | HILIC | NEG | | | 8044167 | 2789190 | 7977478 | 2881061 | 8.64E-01 | -0.02 | 0.58 |
| HILIC_NEG_157_90079a1_325 | | 157.9008 | 1.33 | | HILIC | NEG | | | 1072452 | 461574 | 1062184 | 420237 | 8.64E-01 | -0.02 | 0.42 |
| RP_NEG_339_19968a11_594 | | 339.1997 | 11.59 | | RP | NEG | | | 4796147 | 3651122 | 4709020 | 3763862 | 8.64E-01 | -0.02 | 0.79 |
| RP_POS_419_35226a15_185 | | 419.3523 | 15.19 | | RP | POS | | | 1.5E+07 | 2495601 | 1.5E+07 | 2302051 | 8.64E-01 | -0.02 | 1.04 |
| RP_POS_750_54291a13_686 | | 750.5429 | 13.69 | | RP | POS | | | 1640720 | 1861503 | 1579188 | 3138064 | 8.64E-01 | -0.02 | 0.66 |
| HILIC_POS_361_3064a0_898 | | 361.3064 | 0.90 | | HILIC | POS | | | 288304 | 102302 | 290593 | 95416.4 | 8.65E-01 | 0.02 | 0.85 |
| RP_NEG_185_07817a8_888 | | 185.0782 | 8.89 | | RP | NEG | | | 349289 | 21088.8 | 349797 | 22597.1 | 8.65E-01 | 0.02 | 0.39 |
| RP_NEG_995_61169a10_306 | | 995.6117 | 10.31 | | RP | NEG | | | 2312804 | 1164984 | 2342778 | 1378131 | 8.65E-01 | 0.02 | 0.64 |
| RP_POS_581_27856a10_396 | | 581.2786 | 10.40 | | RP | POS | | | 1003953 | 449077 | 1014139 | 427621 | 8.65E-01 | 0.02 | 0.57 |
| HILIC_NEG_247_98534a6_134 | | 247.9853 | 6.13 | | HILIC | NEG | | | 134247 | 35468.8 | 133402 | 37014.4 | 8.65E-01 | -0.02 | 0.25 |
| RP_NEG_155_0715a6_752 | | 155.0715 | 6.75 | | RP | NEG | | | 361385 | 217360 | 356588 | 196102 | 8.65E-01 | -0.02 | 0.42 |
| RP_POS_334_29538a10_232 | | 334.2954 | 10.23 | | RP | POS | | | 435843 | 110784 | 433413 | 99488.1 | 8.65E-01 | -0.02 | 0.98 |
| RP_POS_362_26831a12_356 | | 362.2683 | 12.36 | | RP | POS | | | 938040 | 582192 | 925620 | 489034 | 8.65E-01 | -0.02 | 0.99 |
| RP_POS_471_33957a9_381 | | 471.3396 | 9.38 | | RP | POS | | | 1474224 | 681319 | 1457434 | 752589 | 8.65E-01 | -0.02 | 0.59 |
| RP_POS_493_32889a10_27 | | 493.3289 | 10.27 | | RP | POS | 493.3312(100) | | 1113042 | 1858252 | 1155244 | 1807266 | 8.66E-01 | 0.02 | 0.12 |
| RP_NEG_174_82954a11_587 | | 174.8295 | 11.59 | | RP | NEG | | | 121471 | 64587.4 | 122918 | 61252.2 | 8.66E-01 | 0.02 | 0.53 |
| RP_NEG_457_59921a15_497 | | 457.5992 | 15.50 | | RP | NEG | | | 1214898 | 76412 | 1216840 | 90473.4 | 8.66E-01 | 0.02 | 0.57 |
| RP_POS_131_96149a15_383 | | 131.9615 | 15.38 | | RP | POS | | | 4360608 | 1463529 | 4398623 | 1784942 | 8.66E-01 | 0.02 | 0.23 |
| RP_NEG_355_22806a9_58 | | 355.2281 | 9.58 | | RP | NEG | | | 427067 | 230810 | 421388 | 259789 | 8.66E-01 | -0.02 | 0.76 |
| RP_POS_202_14351a5_904 | | 202.1435 | 5.90 | | RP | POS | | | 402488 | 330815 | 395949 | 237337 | 8.66E-01 | -0.02 | 0.61 |
| RP_POS_330_29953a10_161 | | 330.2995 | 10.16 | | RP | POS | | | 1091994 | 227375 | 1086756 | 227622 | 8.66E-01 | -0.02 | 0.83 |
| RP_POS_543_90063a10_194 | | 543.9006 | 10.19 | | RP | POS | | | 3997304 | 1625233 | 3958851 | 1706472 | 8.66E-01 | -0.02 | 0.72 |
| RP_POS_563_38983a9_971 | | 563.3898 | 9.97 | | RP | POS | | | 1460375 | 651394 | 1443201 | 811798 | 8.66E-01 | -0.02 | 0.46 |
| RP_NEG_325_18463a15_457 | | 325.1846 | 15.46 | | RP | NEG | 325.1826(100), 183.0122(10) | | 752891 | 107787 | 755232 | 97582.4 | 8.67E-01 | 0.02 | 0.34 |
| HILIC_NEG_218_98833a0_936 | | 218.9883 | 0.94 | | HILIC | NEG | | | 702697 | 255973 | 708963 | 288501 | 8.67E-01 | 0.02 | 0.64 |
| RP_NEG_1085_23096a9_764 | | 1085.2310 | 9.76 | | RP | NEG | | | 4834580 | 337781 | 4841900 | 302410 | 8.67E-01 | 0.02 | 0.75 |
| RP_POS_289_95609a15_623 | | 289.9561 | 15.62 | | RP | POS | | | 315622 | 136356 | 319019 | 157740 | 8.67E-01 | 0.02 | 0.95 |
| RP_POS_365_23212a6_675 | | 365.2321 | 6.68 | | RP | POS | | | 738054 | 272469 | 744192 | 262687 | 8.67E-01 | 0.02 | 0.56 |
| RP_POS_770_60504a14_068 | | 770.6050 | 14.07 | | RP | POS | | | 6426723 | 3995913 | 6513084 | 3577992 | 8.67E-01 | 0.02 | 0.81 |
| HILIC_NEG_115_92075a5_443 | | 115.9208 | 5.44 | | HILIC | NEG | | | 5.9E+07 | 1.2E+07 | 5.9E+07 | 1.2E+07 | 8.67E-01 | -0.02 | 0.25 |
| RP_POS_229_17998a7_528 | | 229.1800 | 7.53 | | RP | POS | | | 188428 | 71179.3 | 186646 | 83331.1 | 8.67E-01 | -0.02 | 0.67 |
| RP_POS_492_00214a10_218 | | 492.0021 | 10.22 | | RP | POS | | | 792509 | 428074 | 783349 | 374176 | 8.67E-01 | -0.02 | 0.64 |
| RP_NEG_133_06592a5_478 | | 133.0659 | 5.48 | | RP | NEG | | | 323556 | 360306 | 333287 | 475416 | 8.68E-01 | 0.02 | 0.18 |
| RP_NEG_381_23141a11_76 | | 381.2314 | 11.76 | | RP | NEG | | | 7098.81 | 8992.12 | 7319.98 | 10319.5 | 8.68E-01 | 0.02 | 0.31 |
| RP_POS_388_34198a11_456 | | 388.3420 | 11.46 | | RP | POS | | | 670286 | 139278 | 673215 | 119545 | 8.68E-01 | 0.02 | 0.67 |
| RP_POS_568_2962a7_071 | | 568.2962 | 7.07 | | RP | POS | | | 2E+08 | 1.4E+07 | 2E+08 | 1.3E+07 | 8.68E-01 | 0.02 | 0.85 |
| RP_POS_727_7063a11_413 | | 727.7063 | 11.41 | | RP | POS | | | 2094711 | 4812395 | 2211841 | 5460026 | 8.68E-01 | 0.02 | 0.73 |
| HILIC_POS_175_0341a0_839 | | 175.0341 | 0.84 | | HILIC | POS | | | 747640 | 226748 | 742339 | 237837 | 8.68E-01 | -0.02 | 0.79 |
| HILIC_NEG_115_92069a1_067 | | 115.9207 | 1.07 | | HILIC | NEG | | | 2.6E+07 | 5595072 | 2.5E+07 | 6124002 | 8.68E-01 | -0.02 | 0.32 |
| RP_NEG_200_91965a15_399 | | 200.9197 | 15.40 | | RP | NEG | | | 2935944 | 598049 | 2922734 | 568595 | 8.68E-01 | -0.02 | 0.33 |
| RP_POS_532_44043a14_356 | | 532.4404 | 14.36 | | RP | POS | | | 1.2E+09 | 2.2E+08 | 1.2E+09 | 2.1E+08 | 8.68E-01 | -0.02 | 0.51 |
| HILIC_POS_184_16942a0_762 | | 184.1694 | 0.76 | | HILIC | POS | | | 8750145 | 1265191 | 8775964 | 1035958 | 8.69E-01 | 0.02 | 0.64 |
| RP_NEG_416_07462a11_144 | | 416.0746 | 11.14 | | RP | NEG | | | 152229 | 31003.4 | 152946 | 32585.6 | 8.69E-01 | 0.02 | 0.56 |
| RP_POS_371_32578a9_731 | | 371.3258 | 9.73 | | RP | POS | | | 974177 | 259107 | 980524 | 298786 | 8.69E-01 | 0.02 | 0.63 |
| RP_POS_569_3855a12_45 | | 569.3855 | 12.45 | | RP | POS | | | 775709 | 243241 | 781774 | 288484 | 8.69E-01 | 0.02 | 0.75 |
| HILIC_NEG_199_80505a3_242 | | 199.8051 | 3.24 | | HILIC | NEG | | | 2.3E+07 | 9188934 | 2.2E+07 | 1E+07 | 8.69E-01 | -0.02 | 0.58 |
| RP_NEG_353_20047a10_673 | | 353.2005 | 10.67 | | RP | NEG | | | 163787 | 298562 | 156667 | 328142 | 8.69E-01 | -0.02 | 0.48 |
| RP_POS_209_19014a8_883 | | 209.1901 | 8.88 | | RP | POS | | | 703073 | 350790 | 695264 | 340112 | 8.69E-01 | -0.02 | 0.54 |
| RP_POS_685_4361a13_696 | | 685.4361 | 13.70 | | RP | POS | | | 6.3E+07 | 1.8E+07 | 6.2E+07 | 1.9E+07 | 8.69E-01 | -0.02 | 0.95 |
| RP_POS_774_54132a14_241 | | 774.5413 | 14.24 | | RP | POS | | | 972901 | 749329 | 956272 | 732034 | 8.69E-01 | -0.02 | 0.8 |
| HILIC_POS_200_06837a4_728 | | 200.0684 | 4.73 | | HILIC | POS | | | 1164094 | 1992224 | 1218358 | 2738490 | 8.70E-01 | 0.02 | 0.47 |
| HILIC_NEG_112_98586a5_121 | | 112.9859 | 5.12 | | HILIC | NEG | 68.9949(100), 112.9856(5) | | 5.6E+07 | 1.8E+07 | 5.6E+07 | 1.7E+07 | 8.70E-01 | 0.02 | 0.39 |
| HILIC_POS_140_06828a2_277 | | 140.0683 | 2.28 | | HILIC | POS | | | 199286 | 126823 | 202312 | 141269 | 8.70E-01 | 0.02 | 0.92 |
| HILIC_NEG_540_3316a1_25 | LysoPC 16:0 | 540.3316 | 1.25 | | HILIC | NEG | 225.2334(100) | 2 | 1.1E+07 | 2006003 | 1.1E+07 | 2133899 | 8.70E-01 | 0.02 | 0.83 |
| RP_POS_236_07159a9_51 | | 236.0716 | 9.51 | | RP | POS | | | 6.9E+07 | 1.1E+07 | 6.9E+07 | 1.2E+07 | 8.70E-01 | -0.02 | 0.29 |
| RP_POS_243_23206a11_664 | | 243.2321 | 11.66 | | RP | POS | | | 241998 | 134612 | 238919 | 140599 | 8.70E-01 | -0.02 | 0.86 |
| RP_POS_341_30466a11_246 | | 341.3047 | 11.25 | | RP | POS | | | 1479511 | 529871 | 1468664 | 444333 | 8.70E-01 | -0.02 | 0.46 |
| RP_POS_534_71234a10_176 | | 534.7123 | 10.18 | | RP | POS | | | 963173 | 454265 | 952732 | 476410 | 8.70E-01 | -0.02 | 0.65 |
| RP_POS_637_94208a9_929 | | 637.9421 | 9.93 | | RP | POS | | | 938377 | 420197 | 928734 | 440213 | 8.70E-01 | -0.02 | 0.53 |
| HILIC_POS_100_07624a1_027 | 2-Piperidone | 100.0762 | 1.03 | | HILIC | POS | 100.0766(100) | 2 | 1724116 | 1648461 | 1684913 | 1841750 | 8.70E-01 | -0.02 | 0.48 |
| HILIC_NEG_174_8392a3_092 | | 174.8392 | 3.09 | | HILIC | NEG | | | 1123019 | 97237.6 | 1125162 | 95831.2 | 8.71E-01 | 0.02 | 0.46 |
| RP_POS_552_19812a11_317 | | 552.1981 | 11.32 | | RP | POS | | | 228183 | 93656.2 | 230062 | 75820 | 8.71E-01 | 0.02 | 0.29 |
| RP_POS_654_45502a11_479 | | 654.4550 | 11.48 | | RP | POS | | | 100295 | 73951.1 | 101930 | 73429.6 | 8.71E-01 | 0.02 | 0.51 |
| RP_NEG_1057_13208a9_708 | | 1057.1321 | 9.71 | | RP | NEG | | | 2195983 | 131118 | 2193324 | 108487 | 8.71E-01 | -0.02 | 0.81 |
| RP_NEG_221_94638a13_807 | | 221.9464 | 13.81 | | RP | NEG | | | 1968450 | 961727 | 1947897 | 895351 | 8.71E-01 | -0.02 | 0.51 |
| RP_POS_1164_3092a13_2 | | 1164.3092 | 13.20 | | RP | POS | | | 583729 | 470846 | 573943 | 411782 | 8.71E-01 | -0.02 | 0.37 |
| RP_POS_225_09087a8_419 | | 225.0909 | 8.42 | | RP | POS | | | 387738 | 414993 | 378666 | 404964 | 8.71E-01 | -0.02 | 0.5 |
| RP_POS_359_35153a11_973 | | 359.3515 | 11.97 | | RP | POS | | | 171647 | 70100.6 | 170031 | 74762.9 | 8.71E-01 | -0.02 | 0.41 |
| RP_NEG_262_99289a7_029 | | 262.9929 | 7.03 | | RP | NEG | | | 2480158 | 320897 | 2486879 | 292205 | 8.72E-01 | 0.02 | 0.75 |
| RP_POS_211_11218a8_84 | | 211.1122 | 8.84 | | RP | POS | | | 2481004 | 1168626 | 2507404 | 1222340 | 8.72E-01 | 0.02 | 0.5 |
| RP_NEG_326_98911a7_241 | | 326.9891 | 7.24 | | RP | NEG | 242.9874(100), 326.988(10) | | 3028624 | 1343704 | 2999136 | 1337338 | 8.72E-01 | -0.02 | 0.6 |
| HILIC_NEG_173_92589a3_149 | | 173.9259 | 3.15 | | HILIC | NEG | | | 480359 | 105224 | 477581 | 140742 | 8.72E-01 | -0.02 | 0.55 |
| HILIC_POS_197_08951a5_792 | | 197.0895 | 5.79 | | HILIC | POS | | | 174077 | 118666 | 171601 | 107422 | 8.72E-01 | -0.02 | 0.64 |
| RP_NEG_262_99222a7_589 | | 262.9922 | 7.59 | | RP | NEG | | | 1127147 | 137288 | 1124240 | 126878 | 8.72E-01 | -0.02 | 0.29 |
| RP_NEG_308_90762a15_361 | | 308.9076 | 15.36 | | RP | NEG | | | 530306 | 155000 | 526748 | 166002 | 8.72E-01 | -0.02 | 0.37 |
| RP_NEG_338_98782a7_677 | | 338.9878 | 7.68 | | RP | NEG | | | 1711488 | 782001 | 1694253 | 783370 | 8.72E-01 | -0.02 | 0.32 |
| RP_NEG_339_19983a12_415 | | 339.1998 | 12.42 | | RP | NEG | | | 2649552 | 3004295 | 2582482 | 3093210 | 8.72E-01 | -0.02 | 0.41 |
| RP_NEG_487_11569a8_014 | | 487.1157 | 8.01 | | RP | NEG | | | 905639 | 59801.1 | 904408 | 52541.2 | 8.72E-01 | -0.02 | 0.87 |
| RP_NEG_609_55798a15_28 | | 609.5580 | 15.28 | | RP | NEG | | | 6894914 | 555307 | 6883290 | 505475 | 8.72E-01 | -0.02 | 0.52 |
| RP_POS_592_25085a11_375 | | 592.2509 | 11.38 | | RP | POS | | | 113106 | 92107.2 | 111334 | 69283 | 8.72E-01 | -0.02 | 0.52 |
| RP_POS_652_45233a11_398 | | 652.4523 | 11.40 | | RP | POS | | | 184930 | 130923 | 182166 | 120357 | 8.72E-01 | -0.02 | 0.31 |
| HILIC_POS_342_01501a5_581 | | 342.0150 | 5.58 | | HILIC | POS | | | 103997 | 46702.9 | 105093 | 52645.8 | 8.73E-01 | 0.02 | 0.4 |
| HILIC_POS_395_04813a3_147 | | 395.0481 | 3.15 | | HILIC | POS | | | 162749 | 55988.8 | 163915 | 50909.4 | 8.73E-01 | 0.02 | 0.76 |
| RP_POS_696_98456a11_441 | | 696.9846 | 11.44 | | RP | POS | | | 250461 | 217709 | 255333 | 227411 | 8.73E-01 | 0.02 | 0.62 |
| HILIC_POS_147_14923a5_602 | | 147.1492 | 5.60 | | HILIC | POS | | | 5.1E+07 | 2.2E+07 | 5E+07 | 2.4E+07 | 8.73E-01 | -0.02 | 0.74 |
| RP_NEG_1021_21796a9_897 | | 1021.2180 | 9.90 | | RP | NEG | | | 3996622 | 265044 | 3991254 | 227622 | 8.73E-01 | -0.02 | 0.74 |
| RP_NEG_550_24609a8_533 | | 550.2461 | 8.53 | | RP | NEG | | | 302377 | 289100 | 296296 | 268745 | 8.73E-01 | -0.02 | 0.26 |
| RP_POS_675_35229a12_617 | | 675.3523 | 12.62 | | RP | POS | | | 305203 | 597697 | 292409 | 577145 | 8.73E-01 | -0.02 | 0.56 |
| HILIC_POS_119_0493a5_097 | | 119.0493 | 5.10 | | HILIC | POS | | | 117174 | 51922.6 | 118223 | 45509.3 | 8.74E-01 | 0.02 | 0.62 |
| RP_NEG_180_97318a14_348 | | 180.9732 | 14.35 | | RP | NEG | | | 1286221 | 692381 | 1301279 | 700563 | 8.74E-01 | 0.02 | 0.47 |
| RP_NEG_640_25708a10_499 | | 640.2571 | 10.50 | | RP | NEG | | | 467408 | 167679 | 471233 | 183747 | 8.74E-01 | 0.02 | 0.79 |
| RP_POS_279_12018a4_747 | | 279.1202 | 4.75 | | RP | POS | | | 226665 | 258628 | 232700 | 293219 | 8.74E-01 | 0.02 | 0.53 |
| RP_POS_466_17453a8_531 | | 466.1745 | 8.53 | | RP | POS | | | 493425 | 193451 | 497760 | 205188 | 8.74E-01 | 0.02 | 0.51 |
| RP_POS_620_49133a14_359 | | 620.4913 | 14.36 | | RP | POS | | | 2317651 | 1013775 | 2340358 | 1072623 | 8.74E-01 | 0.02 | 0.59 |
| HILIC_NEG_358_99576a0_935 | | 358.9958 | 0.94 | | HILIC | NEG | | | 860559 | 92096.8 | 858602 | 88605.6 | 8.74E-01 | -0.02 | 0.72 |
| HILIC_POS_356_27951a0_558 | | 356.2795 | 0.56 | | HILIC | POS | | | 831601 | 320193 | 824617 | 323296 | 8.74E-01 | -0.02 | 0.71 |
| RP_NEG_225_8978a13_275 | | 225.8978 | 13.28 | | RP | NEG | | | 135212 | 100766 | 132831 | 116233 | 8.74E-01 | -0.02 | 0.5 |
| RP_NEG_429_1796a11_239 | | 429.1796 | 11.24 | | RP | NEG | | | 1363097 | 407928 | 1354342 | 397590 | 8.74E-01 | -0.02 | 0.82 |
| RP_POS_451_30203a10_796 | | 451.3020 | 10.80 | | RP | POS | | | 460384 | 543205 | 449720 | 443085 | 8.74E-01 | -0.02 | 0.71 |
| HILIC_NEG_144_92357a4_266 | | 144.9236 | 4.27 | | HILIC | NEG | | | 3.6E+07 | 1E+07 | 3.6E+07 | 1.2E+07 | 8.75E-01 | 0.02 | 0.54 |
| RP_NEG_347_87003a15_401 | | 347.8700 | 15.40 | | RP | NEG | | | 216757 | 111801 | 218971 | 94965.8 | 8.75E-01 | 0.02 | 0.3 |
| RP_POS_542_32178a10_312 | | 542.3218 | 10.31 | | RP | POS | | | 1.4E+08 | 2.4E+07 | 1.4E+08 | 2.7E+07 | 8.75E-01 | 0.02 | 0.54 |
| RP_POS_195_1382a5_97 | | 195.1382 | 5.97 | | RP | POS | | | 755790 | 122188 | 753217 | 116654 | 8.75E-01 | -0.02 | 0.84 |
| RP_POS_665_29761a11_437 | | 665.2976 | 11.44 | | RP | POS | | | 136978 | 81137.6 | 135124 | 90021.1 | 8.75E-01 | -0.02 | 0.58 |
| RP_POS_163_03891a7_471 | | 163.0389 | 7.47 | | RP | POS | 163.0393(100) | | 1333558 | 183086 | 1336956 | 135388 | 8.76E-01 | 0.02 | 0.59 |
| HILIC_POS_503_05246a3_147 | | 503.0525 | 3.15 | | HILIC | POS | | | 232344 | 41479 | 233190 | 37893.2 | 8.76E-01 | 0.02 | 0.4 |
| RP_NEG_480_30984a10_489 | | 480.3098 | 10.49 | | RP | NEG | | | 1.9E+07 | 1970223 | 1.9E+07 | 2247747 | 8.76E-01 | 0.02 | 0.88 |
| RP_POS_313_27347a11_19 | | 313.2735 | 11.19 | | RP | POS | | | 468822 | 382959 | 460859 | 365652 | 8.76E-01 | -0.02 | 0.31 |
| RP_POS_393_29761a11_78 | | 393.2976 | 11.78 | | RP | POS | | | 4564893 | 1227271 | 4539434 | 1170056 | 8.76E-01 | -0.02 | 0.57 |
| RP_POS_603_08984a10_39 | | 603.0898 | 10.39 | | RP | POS | | | 273898 | 218987 | 269352 | 207497 | 8.76E-01 | -0.02 | 0.47 |
| RP_POS_758_57367a13_718 | PC 34:2 (16:0_18:2) | 758.5737 | 13.72 | | RP | POS | 184.0745(100) | 1 | 1.6E+07 | 9766412 | 1.6E+07 | 9311065 | 8.76E-01 | -0.02 | 0.67 |
| RP_NEG_327_25421a10_688 | | 327.2542 | 10.69 | | RP | NEG | | | 208087 | 62318.1 | 209442 | 65658.9 | 8.77E-01 | 0.02 | 0.69 |

**Supplemantary table 1**: The non-targeted metabolomics results ordered by the VIP value (p value < 0.0003 are shaded dark green, p value < 0.05 are shaded light green, d values below 0 are blue and above 0 are red)

| | | | | | | | | | Cases | | Controls | | Welch test | Cohen's | |
|---|---|---|---|---|---|---|---|---|---|---|---|---|---|---|---|
| Feature_ID | Metabolite_name | Average_Mz | Average_RT_min_ | HMDB ID | RP_HILIC | POS_NEG | Cureted_MSMS | ID_level | Mean | SD | Mean2 | SD3 | p | d | VIP |
| RP_POS_199_00806a0_694 | | 199.0081 | 0.69 | | RP | POS | | | 715788 | 85623.8 | 717601 | 85356.6 | 8.77E-01 | 0.02 | 0.46 |
| RP_POS_232_12904a0_681 | | 232.1290 | 0.68 | | RP | POS | | | 416284 | 387976 | 425277 | 456221 | 8.77E-01 | 0.02 | 1.24 |
| RP_NEG_409_65601a15_493 | | 409.6560 | 15.49 | | RP | NEG | | | 369138 | 37751.2 | 368260 | 44293.1 | 8.77E-01 | -0.02 | 0.65 |
| RP_POS_152_01637a5_791 | 2-Hydroxybenzothiazole | 152.0164 | 5.79 | | RP | POS | 152.0164(100) | 2 | 1.2E+07 | 5340335 | 1.2E+07 | 4285511 | 8.77E-01 | 0.02 | 0.54 |
| RP_POS_1109_65259a10_294 | | 1109.6526 | 10.29 | | RP | POS | | | 574503 | 243582 | 579803 | 260291 | 8.78E-01 | 0.02 | 0.39 |
| RP_POS_515_31281a10_354 | | 515.3128 | 10.35 | | RP | POS | | | 582964 | 275283 | 589679 | 352192 | 8.78E-01 | 0.02 | 1.07 |
| RP_POS_545_37756a8_693 | | 545.3776 | 8.69 | | RP | POS | | | 324780 | 217446 | 329169 | 200508 | 8.78E-01 | 0.02 | 0.35 |
| RP_POS_582_74268a11_314 | | 582.7427 | 11.31 | | RP | POS | | | 101083 | 50646.5 | 102287 | 62274.7 | 8.78E-01 | 0.02 | 0.45 |
| RP_POS_597_09039a10_645 | | 597.0904 | 10.65 | | RP | POS | | | 266533 | 161103 | 270098 | 175865 | 8.78E-01 | 0.02 | 0.5 |
| RP_POS_617_56598a11_592 | | 617.5660 | 11.59 | | RP | POS | | | 1280564 | 713772 | 1297126 | 843011 | 8.78E-01 | 0.02 | 0.9 |
| HILIC_NEG_200_99745a6_135 | | 200.9975 | 6.14 | | HILIC | NEG | | | 857849 | 237996 | 852993 | 225857 | 8.78E-01 | -0.02 | 0.84 |
| HILIC_NEG_261_89243a3_097 | | 261.8924 | 3.10 | | HILIC | NEG | | | 114468 | 44572.4 | 113565 | 41791.6 | 8.78E-01 | -0.02 | 0.5 |
| HILIC_POS_384_34644a3_12 | | 384.3464 | 3.12 | | HILIC | POS | | | 84928.6 | 61280.6 | 83723.9 | 54263.9 | 8.78E-01 | -0.02 | 0.31 |
| RP_POS_205_14343a7_272 | | 205.1434 | 7.27 | | RP | POS | | | 3060680 | 295868 | 3054719 | 275129 | 8.78E-01 | -0.02 | 1.03 |
| RP_POS_549_8902a9_983 | | 549.8902 | 9.98 | | RP | POS | | | 3011301 | 1369465 | 2979837 | 1600751 | 8.78E-01 | -0.02 | 0.85 |
| RP_NEG_150_00179a5_79 | | 150.0018 | 5.79 | | RP | NEG | 150.0011(100) | | 2390944 | 1056614 | 2410614 | 838250 | 8.79E-01 | 0.02 | 0.54 |
| RP_POS_1096_04492a14_051 | | 1096.0449 | 14.05 | | RP | POS | | | 1718109 | 348087 | 1724724 | 287136 | 8.79E-01 | 0.02 | 0.9 |
| RP_POS_416_37296a12_342 | | 416.3730 | 12.34 | | RP | POS | | | 1307382 | 186596 | 1311269 | 187932 | 8.79E-01 | 0.02 | 0.62 |
| RP_POS_539_70807a9_873 | | 539.7081 | 9.87 | | RP | POS | | | 380757 | 193397 | 384754 | 190502 | 8.79E-01 | 0.02 | 0.66 |
| RP_POS_694_9859a11_378 | | 694.9859 | 11.38 | | RP | POS | | | 200469 | 169907 | 204448 | 206686 | 8.79E-01 | 0.02 | 0.72 |
| RP_NEG_160_93552a11_139 | | 160.9355 | 11.14 | | RP | NEG | | | 274145 | 29454.6 | 273437 | 37386.5 | 8.79E-01 | -0.02 | 0.36 |
| RP_NEG_310_00177a1_429 | | 310.0018 | 1.43 | | RP | NEG | | | 767591 | 311906 | 760987 | 321710 | 8.79E-01 | -0.02 | 0.28 |
| RP_NEG_819_15222a9_538 | | 819.1522 | 9.54 | | RP | NEG | | | 1786732 | 110152 | 1784480 | 106806 | 8.79E-01 | -0.02 | 0.72 |
| RP_POS_421_32846a12_341 | | 421.3285 | 12.34 | | RP | POS | | | 1656430 | 365162 | 1649391 | 313055 | 8.79E-01 | -0.02 | 0.88 |
| HILIC_POS_447_34384a4_874 | | 447.3438 | 4.87 | | HILIC | POS | | | 16530.3 | 6668.73 | 16661.6 | 6050.16 | 8.80E-01 | 0.02 | 0.26 |
| HILIC_POS_497_05637a7_67 | | 497.0564 | 7.67 | | HILIC | POS | | | 198194 | 11899.6 | 198470 | 14466.7 | 8.80E-01 | 0.02 | 0.28 |
| RP_POS_197_0787a6_15 | | 197.0787 | 6.15 | | RP | POS | | | 577140 | 199289 | 580970 | 172195 | 8.80E-01 | 0.02 | 0.79 |
| RP_POS_278_63922a10_013 | | 278.6392 | 10.01 | | RP | POS | | | 216612 | 168436 | 220164 | 174109 | 8.80E-01 | 0.02 | 0.55 |
| RP_POS_648_48773a14_342 | | 648.4877 | 14.34 | | RP | POS | | | 1227982 | 556777 | 1240349 | 634234 | 8.80E-01 | 0.02 | 0.23 |
| HILIC_POS_157_09715a5_118 | | 157.0972 | 5.12 | | HILIC | POS | | | 3037718 | 5833715 | 2920083 | 5591515 | 8.80E-01 | -0.02 | 0.78 |
| HILIC_POS_188_09167a5_942 | | 188.0917 | 5.94 | | HILIC | POS | | | 347164 | 203117 | 342721 | 225049 | 8.80E-01 | -0.02 | 0.81 |
| RP_POS_236_07152a13_075 | | 236.0715 | 13.08 | | RP | POS | | | 9.3E+07 | 3.2E+07 | 9.2E+07 | 2.7E+07 | 8.80E-01 | -0.02 | 0.52 |
| RP_POS_332_26144a9_172 | | 332.2614 | 9.17 | | RP | POS | | | 484648 | 707605 | 470086 | 706122 | 8.80E-01 | -0.02 | 0.38 |
| RP_POS_181_0498a9_523 | | 181.0498 | 9.52 | | RP | POS | | | 4780045 | 1016450 | 4806027 | 1451011 | 8.81E-01 | 0.02 | 0.72 |
| RP_POS_500_2749a10_179 | | 500.2749 | 10.18 | | RP | POS | | | 1102113 | 465853 | 1112791 | 563777 | 8.81E-01 | 0.02 | 0.63 |
| RP_POS_692_14886a11_544 | | 692.1489 | 11.54 | | RP | POS | | | 221481 | 94172 | 223916 | 136911 | 8.81E-01 | 0.02 | 0.46 |
| RP_POS_796_52673a13_686 | | 796.5267 | 13.69 | | RP | POS | | | 2806913 | 3388508 | 2877241 | 3450529 | 8.81E-01 | 0.02 | 0.75 |
| RP_POS_1199_86816a13_791 | | 1199.8682 | 13.79 | | RP | POS | | | 772927 | 553559 | 762010 | 514937 | 8.81E-01 | -0.02 | 0.37 |
| RP_POS_147_08057a7_3 | Benzylidenacetone | 147.0806 | 7.30 | | RP | POS | 147.0802(100), 129.0697(90) | 2 | 427832 | 94341.9 | 425994 | 85219.6 | 8.81E-01 | -0.02 | 0.61 |
| HILIC_NEG_200_85857a5_414 | | 200.8586 | 5.41 | | HILIC | NEG | | | 5001044 | 2039550 | 5041292 | 1946329 | 8.82E-01 | 0.02 | 0.23 |
| RP_NEG_136_87204a15_454 | | 136.8720 | 15.45 | | RP | NEG | | | 1512553 | 229960 | 1518377 | 327297 | 8.82E-01 | 0.02 | 0.58 |
| RP_POS_311_03564a15_485 | | 311.0356 | 15.49 | | RP | POS | | | 77672.1 | 26145.6 | 78241.2 | 29601.6 | 8.82E-01 | 0.02 | 0.67 |
| RP_POS_328_90042a15_408 | | 328.9004 | 15.41 | | RP | POS | | | 6028444 | 1861644 | 6060689 | 1324278 | 8.82E-01 | 0.02 | 0.95 |
| HILIC_POS_226_04506a5_12 | | 226.0451 | 5.12 | | HILIC | POS | | | 1376118 | 344149 | 1369488 | 312914 | 8.82E-01 | -0.02 | 0.5 |
| RP_POS_259_20511a10_659 | | 259.2051 | 10.66 | | RP | POS | | | 5373324 | 1240285 | 5348358 | 1227527 | 8.82E-01 | -0.02 | 0.29 |
| RP_POS_518_32019a11_594 | | 518.3202 | 11.59 | | RP | POS | | | 1429317 | 320610 | 1422743 | 326586 | 8.82E-01 | -0.02 | 0.64 |
| HILIC_POS_424_34116a0_605 | | 424.3412 | 0.61 | | HILIC | POS | | | 333160 | 120243 | 335384 | 101847 | 8.83E-01 | 0.02 | 0.67 |
| RP_POS_189_12708a7_29 | | 189.1271 | 7.29 | | RP | POS | | | 620482 | 155675 | 623476 | 142651 | 8.83E-01 | 0.02 | 0.56 |
| RP_POS_330_29962a9_673 | | 330.2996 | 9.67 | | RP | POS | | | 276919 | 115020 | 279087 | 100506 | 8.83E-01 | 0.02 | 0.56 |
| RP_NEG_257_92645a3_581 | | 257.9265 | 3.58 | | RP | NEG | | | 444931 | 199435 | 441167 | 177084 | 8.83E-01 | -0.02 | 0.57 |
| RP_NEG_418_0719a11_144 | | 418.0719 | 11.14 | | RP | NEG | | | 114197 | 31991.1 | 113484 | 38079.3 | 8.83E-01 | -0.02 | 0.52 |
| RP_POS_189_15981a15_447 | | 189.1598 | 15.45 | | RP | POS | | | 595740 | 179008 | 591932 | 197809 | 8.83E-01 | -0.02 | 0.79 |
| RP_POS_201_12726a8_354 | | 201.1273 | 8.35 | | RP | POS | | | 2529720 | 126229 | 2527066 | 136956 | 8.83E-01 | -0.02 | 0.76 |
| RP_POS_638_72687a6_088 | | 638.7269 | 6.09 | | RP | POS | | | 1015532 | 425911 | 1005964 | 510869 | 8.83E-01 | -0.02 | 0.69 |
| RP_POS_793_55847a13_113 | | 793.5585 | 13.11 | | RP | POS | | | 2.8E+07 | 4.7E+07 | 2.7E+07 | 3.9E+07 | 8.83E-01 | -0.02 | 0.63 |
| RP_POS_939_59528a11_433 | | 939.5953 | 11.43 | | RP | POS | | | 360328 | 487908 | 351392 | 406649 | 8.83E-01 | -0.02 | 0.51 |
| HILIC_NEG_315_0416a6_126 | | 315.0416 | 6.13 | | HILIC | NEG | | | 2272851 | 577363 | 2284186 | 564235 | 8.84E-01 | 0.02 | 0.35 |
| RP_POS_1184_3407a13_274 | | 1184.3407 | 13.27 | | RP | POS | | | 4462014 | 1615953 | 4494263 | 1606319 | 8.84E-01 | 0.02 | 0.65 |
| RP_POS_227_12485a6_991 | | 227.1249 | 6.99 | | RP | POS | | | 797322 | 197154 | 792898 | 239001 | 8.84E-01 | -0.02 | 0.88 |
| RP_POS_295_19424a11_14 | | 295.1942 | 11.14 | | RP | POS | | | 412299 | 207761 | 407641 | 254781 | 8.84E-01 | -0.02 | 0.56 |
| HILIC_POS_106_08678a2_247 | | 106.0868 | 2.25 | | HILIC | POS | 88.0762(100), 106.0863(70) | | 1.5E+07 | 7703498 | 1.5E+07 | 7277991 | 8.85E-01 | 0.02 | 1.03 |
| RP_POS_478_32718a10_484 | | 478.3272 | 10.48 | | RP | POS | 478.3316(100) | | 3291221 | 807212 | 3307301 | 815056 | 8.85E-01 | 0.02 | 0.3 |
| RP_NEG_362_6557a15_498 | | 362.6557 | 15.50 | | RP | NEG | | | 159834 | 62216.5 | 161042 | 60014.3 | 8.85E-01 | 0.02 | 0.56 |
| RP_POS_350_03683a15_462 | | 350.0368 | 15.46 | | RP | POS | | | 968179 | 260240 | 973177 | 246221 | 8.85E-01 | 0.02 | 0.5 |
| RP_POS_588_17468a8_478 | | 588.1747 | 8.48 | | RP | POS | | | 1118644 | 106830 | 1120762 | 106480 | 8.85E-01 | 0.02 | 1.01 |
| RP_POS_862_51074a12_876 | | 862.5107 | 12.88 | | RP | POS | | | 962817 | 770134 | 978000 | 762001 | 8.85E-01 | 0.02 | 0.53 |
| HILIC_NEG_201_80214a3_238 | | 201.8021 | 3.24 | | HILIC | NEG | | | 4320269 | 1676770 | 4285158 | 1855072 | 8.85E-01 | -0.02 | 0.39 |
| RP_POS_234_89574a15_461 | | 234.8957 | 15.46 | | RP | POS | | | 390307 | 51865.2 | 389377 | 42175.4 | 8.85E-01 | -0.02 | 0.68 |
| RP_POS_644_49512a10_016 | | 644.4951 | 10.02 | | RP | POS | | | 1283445 | 645598 | 1270131 | 698164 | 8.85E-01 | -0.02 | 0.77 |
| RP_NEG_178_97778a15_546 | Difluoro(perfluoromethoxy)acetic acid | 178.9778 | 15.55 | | RP | NEG | 134.9883(100), 178.9779(15) | 2 | 8218332 | 1179736 | 8241878 | 1195571 | 8.85E-01 | 0.02 | 0.57 |
| RP_POS_256_26324a10_714 | Palmitic acid | 256.2632 | 10.71 | | RP | POS | 256.2624(100), 88.0762(10) | 1 | 5.4E+07 | 1.8E+07 | 5.4E+07 | 2E+07 | 8.86E-01 | 0.02 | 1.11 |
| HILIC_POS_243_05257a4_112 | | 243.0526 | 4.11 | | HILIC | POS | | | 1230595 | 563178 | 1241628 | 560397 | 8.86E-01 | 0.02 | 0.55 |
| RP_NEG_496_80225a15_413 | | 496.8023 | 15.41 | | RP | NEG | | | 273253 | 46002.9 | 274246 | 53968.1 | 8.86E-01 | 0.02 | 0.57 |
| RP_POS_268_96021a0_682 | | 268.9602 | 0.68 | | RP | POS | | | 796740 | 159510 | 799786 | 152085 | 8.86E-01 | 0.02 | 0.51 |
| RP_POS_581_40393a9_393 | | 581.4039 | 9.39 | | RP | POS | | | 398061 | 210686 | 402495 | 238797 | 8.86E-01 | 0.02 | 0.83 |
| RP_POS_811_66797a14_95 | | 811.6680 | 14.95 | | RP | POS | | | 1.2E+07 | 6599429 | 1.2E+07 | 5581171 | 8.86E-01 | 0.02 | 0.63 |
| HILIC_NEG_264_00256a4_858 | | 264.0026 | 4.86 | | HILIC | NEG | | | 188769 | 34552.2 | 188188 | 24701.9 | 8.86E-01 | -0.02 | 0.27 |
| HILIC_NEG_97_93107a3_154 | | 97.9311 | 3.15 | | HILIC | NEG | | | 264272 | 98974.6 | 262399 | 93257.2 | 8.86E-01 | -0.02 | 0.21 |
| RP_NEG_179_07169a6_899 | | 179.0717 | 6.90 | | RP | NEG | | | 416132 | 26306.8 | 415581 | 29629.3 | 8.86E-01 | -0.02 | 0.51 |
| RP_NEG_262_02304a0_67 | | 262.0230 | 0.67 | | RP | NEG | | | 248152 | 46367.8 | 247166 | 53508.9 | 8.86E-01 | -0.02 | 0.49 |
| RP_NEG_290_84189a15_412 | | 290.8419 | 15.41 | | RP | NEG | | | 442200 | 170862 | 438824 | 174619 | 8.86E-01 | -0.02 | 0.71 |
| RP_POS_593_13165a8_478 | | 593.1317 | 8.48 | | RP | POS | | | 506235 | 43412.2 | 505376 | 43803.9 | 8.86E-01 | -0.02 | 0.52 |
| RP_POS_203_12788a10_362 | | 203.1279 | 10.36 | | RP | POS | | | 467989 | 193030 | 471784 | 197140 | 8.87E-01 | 0.02 | 0.5 |
| RP_POS_338_23209a9_521 | | 338.2321 | 9.52 | | RP | POS | | | 1349266 | 463776 | 1358825 | 514417 | 8.87E-01 | 0.02 | 1.02 |
| RP_POS_509_69891a10_348 | | 509.6989 | 10.35 | | RP | POS | | | 781089 | 514337 | 791092 | 517616 | 8.87E-01 | 0.02 | 0.69 |
| RP_POS_653_46161a10_102 | | 653.4616 | 10.10 | | RP | POS | | | 6107695 | 2325774 | 6160188 | 2991063 | 8.87E-01 | 0.02 | 0.51 |
| HILIC_POS_162_05019a4_845 | | 162.0502 | 4.85 | | HILIC | POS | | | 2105869 | 1329358 | 2081816 | 1163176 | 8.87E-01 | -0.02 | 0.57 |
| HILIC_POS_387_21396a5_652 | | 387.2140 | 5.65 | | HILIC | POS | | | 1733564 | 453346 | 1724958 | 431827 | 8.87E-01 | -0.02 | 0.47 |
| RP_NEG_249_07675a7_881 | | 249.0768 | 7.88 | | RP | NEG | | | 518687 | 28138.1 | 518144 | 27851.8 | 8.87E-01 | -0.02 | 0.41 |
| RP_NEG_261_77484a15_487 | | 261.7748 | 15.49 | | RP | NEG | | | 203573 | 37705.1 | 202803 | 41073.5 | 8.87E-01 | -0.02 | 0.49 |
| RP_NEG_673_06226a9_098 | | 673.0623 | 9.10 | | RP | NEG | | | 441178 | 34652.1 | 440542 | 30991.3 | 8.87E-01 | -0.02 | 0.45 |
| RP_POS_175_1078a0_654 | | 175.1078 | 0.65 | | RP | POS | | | 1023767 | 1239534 | 999343 | 1271228 | 8.87E-01 | -0.02 | 0.79 |
| RP_POS_201_6813a10_833 | | 201.6813 | 10.83 | | RP | POS | | | 228760 | 97816.6 | 226842 | 99367.4 | 8.87E-01 | -0.02 | 0.91 |
| RP_POS_236_07147a12_945 | | 236.0715 | 12.95 | | RP | POS | | | 1.1E+08 | 5.1E+07 | 1.1E+08 | 4.2E+07 | 8.87E-01 | -0.02 | 0.65 |
| RP_POS_338_34131a11_41 | | 338.3413 | 11.41 | | RP | POS | | | 1235988 | 537872 | 1225946 | 495014 | 8.87E-01 | -0.02 | 0.45 |
| RP_POS_256_09244a3_176 | Amfenac | 256.0924 | 3.18 | | RP | POS | 256.091(100), 238.0832(25) | 2 | 179454 | 91270.3 | 181100 | 78102.3 | 8.87E-01 | 0.02 | 0.39 |
| RP_NEG_573_13025a8_98 | | 573.1303 | 8.98 | | RP | NEG | | | 2.2E+07 | 1125068 | 2.2E+07 | 1083203 | 8.88E-01 | 0.02 | 0.81 |
| RP_POS_303_0386a4_824 | | 303.0386 | 4.82 | | RP | POS | | | 973435 | 766031 | 988586 | 809693 | 8.88E-01 | 0.02 | 0.65 |
| RP_POS_464_28287a9_222 | | 464.2829 | 9.22 | | RP | POS | 464.288(100),126.0229(60) | | 1206316 | 1687373 | 1175763 | 1496355 | 8.88E-01 | -0.02 | 0.5 |
| RP_POS_257_17465a9_58 | | 257.1747 | 9.58 | | RP | POS | | | 3715971 | 2418257 | 3669714 | 2383282 | 8.88E-01 | -0.02 | 0.98 |
| HILIC_NEG_183_93459a1_772 | | 183.9346 | 1.77 | | HILIC | NEG | | | 2165665 | 1083394 | 2143931 | 1189491 | 8.89E-01 | -0.02 | 0.43 |
| RP_NEG_384_93674a11_194 | | 384.9367 | 11.19 | | RP | NEG | | | 1039191 | 286469 | 1033699 | 288700 | 8.89E-01 | -0.02 | 0.37 |
| RP_POS_251_04642a10_289 | | 251.0464 | 10.29 | | RP | POS | | | 408399 | 97333.9 | 406572 | 94136.2 | 8.89E-01 | -0.02 | 0.47 |
| RP_POS_625_32745a14_343 | | 625.3275 | 14.34 | | RP | POS | | | 2252725 | 1044104 | 2233951 | 931586 | 8.89E-01 | -0.02 | 0.49 |
| RP_NEG_201_03493a0_585 | | 201.0349 | 0.59 | | RP | NEG | | | 1328468 | 349140 | 1335739 | 414008 | 8.90E-01 | 0.02 | 0.25 |
| RP_NEG_221_09325a2_818 | | 221.0933 | 2.82 | | RP | NEG | | | 199595 | 77082.9 | 201003 | 72561.9 | 8.90E-01 | 0.02 | 0.51 |
| RP_POS_558_29541a10_178 | | 558.2954 | 10.18 | | RP | POS | | | 748232 | 462684 | 757212 | 483465 | 8.90E-01 | 0.02 | 0.62 |
| RP_POS_205_08621a9_44 | | 205.0862 | 9.44 | | RP | POS | | | 2044685 | 199833 | 2040998 | 189394 | 8.90E-01 | -0.02 | 0.83 |
| RP_POS_426_32541a10_661 | | 426.3254 | 10.66 | | RP | POS | | | 198022 | 139843 | 195621 | 114109 | 8.90E-01 | -0.02 | 0.6 |
| RP_POS_706_974a10_235 | | 706.9740 | 10.24 | | RP | POS | | | 1062024 | 512618 | 1052547 | 491108 | 8.90E-01 | -0.02 | 0.87 |
| HILIC_POS_120_0811a3_908 | | 120.0811 | 3.91 | | HILIC | POS | 120.0807(100), 103.0543(20) | | 4.4E+07 | 9610305 | 4.4E+07 | 9485976 | 8.91E-01 | 0.02 | 0.51 |
| HILIC_NEG_115_92081a5_928 | | 115.9208 | 5.93 | | HILIC | NEG | | | 4.3E+07 | 8170088 | 4.3E+07 | 7876288 | 8.91E-01 | 0.02 | 0.48 |
| HILIC_NEG_317_8512a3_247 | | 317.8512 | 3.25 | | HILIC | NEG | | | 480579 | 70003.3 | 481896 | 70132.7 | 8.91E-01 | 0.02 | 0.73 |
| HILIC_POS_368_21985a5_933 | | 368.2199 | 5.93 | | HILIC | POS | | | 351184 | 220882 | 356145 | 296409 | 8.91E-01 | 0.02 | 0.89 |
| RP_POS_1039_67236a10_308 | | 1039.6724 | 10.31 | | RP | POS | | | 4.6E+07 | 3.1E+07 | 4.6E+07 | 3.5E+07 | 8.91E-01 | 0.02 | 0.61 |
| RP_POS_223_09621a7_47 | | 223.0962 | 7.47 | | RP | POS | | | 4507893 | 649872 | 4518038 | 427527 | 8.91E-01 | 0.02 | 0.55 |
| RP_POS_376_95453a15_455 | | 376.9545 | 15.46 | | RP | POS | | | 620091 | 177217 | 623668 | 200641 | 8.91E-01 | 0.02 | 0.97 |
| RP_NEG_399_18408a11_592 | | 399.1841 | 11.59 | | RP | NEG | | | 2047444 | 108540 | 2045303 | 119642 | 8.91E-01 | -0.02 | 0.87 |
| RP_POS_134_95857a0_635 | | 134.9586 | 0.64 | | RP | POS | | | 612858 | 325481 | 606398 | 362133 | 8.91E-01 | -0.02 | 0.71 |
| RP_POS_174_14883a0_681 | | 174.1488 | 0.68 | | RP | POS | | | 2785611 | 642047 | 2774113 | 589234 | 8.91E-01 | -0.02 | 0.56 |
| RP_POS_569_43231a13_287 | | 569.4323 | 13.29 | | RP | POS | | | 233094 | 113558 | 231103 | 99022.5 | 8.91E-01 | -0.02 | 0.65 |
| RP_POS_766_54303a15_178 | | 766.5430 | 15.18 | | RP | POS | | | 2498571 | 1572779 | 2470494 | 1423493 | 8.91E-01 | -0.02 | 0.85 |
| HILIC_NEG_144_92361a3_205 | | 144.9236 | 3.21 | | HILIC | NEG | | | 1.2E+07 | 372261 | 1.2E+07 | 383702 | 8.92E-01 | 0.02 | 0.25 |
| RP_NEG_314_98907a7_661 | | 314.9891 | 7.66 | | RP | NEG | | | 6223779 | 347818 | 6230576 | 382306 | 8.92E-01 | 0.02 | 0.23 |
| RP_POS_340_24863a7_553 | | 340.2486 | 7.55 | | RP | POS | | | 268648 | 164274 | 271641 | 159137 | 8.92E-01 | 0.02 | 0.81 |
| HILIC_POS_106_08673a1_462 | | 106.0867 | 1.46 | | HILIC | POS | | | 9052565 | 4259263 | 8947918 | 6574934 | 8.92E-01 | -0.02 | 0.91 |
| RP_NEG_437_16211a7_49 | | 437.1621 | 7.49 | | RP | NEG | | | 412308 | 515140 | 404718 | 291264 | 8.92E-01 | -0.02 | 0.57 |
| RP_POS_259_20547a10_176 | | 259.2055 | 10.18 | | RP | POS | | | 3744223 | 228558 | 3739606 | 266496 | 8.92E-01 | -0.02 | 0.3 |
| RP_POS_399_30844a10_695 | | 399.3084 | 10.70 | | RP | POS | | | 7693329 | 1192349 | 7672338 | 1085787 | 8.92E-01 | -0.02 | 1.14 |
| RP_NEG_206_78903a15_494 | | 206.7890 | 15.49 | | RP | NEG | | | 866089 | 36045.4 | 866817 | 42390.1 | 8.93E-01 | 0.02 | 0.52 |
| RP_NEG_379_15689a10_54 | | 379.1569 | 10.54 | | RP | NEG | | | 215387 | 136242 | 218438 | 187396 | 8.93E-01 | 0.02 | 0.3 |
| RP_NEG_722_51483a13_353 | | 722.5148 | 13.35 | | RP | NEG | | | 1851358 | 561430 | 1862400 | 628129 | 8.93E-01 | 0.02 | 0.72 |
| RP_POS_311_25778a10_676 | | 311.2578 | 10.68 | | RP | POS | | | 2487811 | 461222 | 2498547 | 670870 | 8.93E-01 | 0.02 | 0.67 |
| RP_POS_209_07864a3_363 | | 209.0786 | 3.36 | | RP | POS | 209.0768(100) | | 508644 | 251974 | 503918 | 262531 | 8.93E-01 | -0.02 | 1.1 |
| RP_NEG_938_53619a13_238 | | 938.5362 | 13.24 | | RP | NEG | | | 2134267 | 578771 | 2121956 | 734355 | 8.93E-01 | -0.02 | 0.3 |
| RP_POS_335_21915a10_718 | | 335.2192 | 10.72 | | RP | POS | | | 1867454 | 935460 | 1851594 | 802789 | 8.93E-01 | -0.02 | 0.84 |
| RP_POS_707_49023a10_673 | | 707.4902 | 10.67 | | RP | POS | | | 341753 | 124679 | 339143 | 155208 | 8.93E-01 | -0.02 | 0.6 |
| RP_POS_740_52173a12_803 | | 740.5217 | 12.80 | | RP | POS | | | 629091 | 785808 | 614360 | 812062 | 8.93E-01 | -0.02 | 0.47 |
| HILIC_NEG_137_02422a0_605 | Salicylic acid | 137.0242 | 0.61 | | HILIC | NEG | 93.034(100), 137.024(30) | 1 | 538247 | 432146 | 545564 | 364172 | 8.93E-01 | 0.02 | 0.79 |
| RP_POS_195_12265a2_061 | Octyl acetate | 195.1227 | 2.06 | | RP | POS | 195.0871(100) | 2 | 6219713 | 739444 | 6234087 | 818431 | 8.93E-01 | 0.02 | 0.55 |
| RP_NEG_524_47522a15_499 | | 524.4752 | 15.50 | | RP | NEG | | | 325609 | 33525 | 326175 | 29032.2 | 8.94E-01 | 0.02 | 0.64 |
| RP_POS_238_06674a10_537 | | 238.0667 | 10.54 | | RP | POS | | | 3705099 | 1201167 | 3727888 | 1284137 | 8.94E-01 | 0.02 | 0.39 |
| RP_POS_524_27063a6_968 | | 524.2706 | 6.97 | | RP | POS | | | 2553774 | 184182 | 2556888 | 158922 | 8.94E-01 | 0.02 | 0.88 |
| RP_POS_616_44287a10_304 | | 616.4429 | 10.30 | | RP | POS | | | 5815171 | 2524298 | 5860341 | 2420929 | 8.94E-01 | 0.02 | 0.65 |
| RP_NEG_452_92398a11_094 | | 452.9240 | 11.09 | | RP | NEG | | | 956399 | 393558 | 949611 | 350758 | 8.94E-01 | -0.02 | 0.72 |
| RP_NEG_555_21466a11_594 | | 555.2147 | 11.59 | | RP | NEG | | | 839957 | 179314 | 836322 | 216075 | 8.94E-01 | -0.02 | 0.39 |
| RP_POS_287_63293a10_181 | | 287.6329 | 10.18 | | RP | POS | | | 783717 | 327399 | 777779 | 322484 | 8.94E-01 | -0.02 | 0.79 |
| RP_POS_582_50897a13_704 | | 582.5090 | 13.70 | | RP | POS | | | 288501 | 362862 | 282660 | 279851 | 8.94E-01 | -0.02 | 0.37 |
| RP_NEG_336_78503a15_438 | | 336.7850 | 15.44 | | RP | NEG | | | 5662806 | 1595128 | 5693180 | 1764132 | 8.95E-01 | 0.02 | 0.58 |
| RP_NEG_381_93344a0_677 | | 381.9334 | 0.68 | | RP | NEG | | | 193771 | 34290.1 | 194409 | 36010.5 | 8.95E-01 | 0.02 | 0.22 |
| RP_POS_217_1217a8_245 | | 217.1217 | 8.25 | | RP | POS | | | 4103341 | 192396 | 4106868 | 198777 | 8.95E-01 | 0.02 | 0.86 |
| RP_POS_227_12791a5_976 | | 227.1279 | 5.98 | | RP | POS | | | 421623 | 197989 | 425000 | 176200 | 8.95E-01 | 0.02 | 0.53 |
| RP_POS_472_34772a7_534 | | 472.3477 | 7.53 | | RP | POS | | | 474546 | 326936 | 480451 | 327481 | 8.95E-01 | 0.02 | 0.9 |
| RP_NEG_162_83899a15_492 | | 162.8390 | 15.49 | | RP | NEG | | | 5801954 | 141177 | 5799175 | 163461 | 8.95E-01 | -0.02 | 0.45 |
| RP_NEG_356_84247a15_401 | | 356.8425 | 15.40 | | RP | NEG | | | 808757 | 205618 | 805001 | 209461 | 8.95E-01 | -0.02 | 0.62 |
| RP_NEG_632_31873a10_181 | | 632.3187 | 10.18 | | RP | NEG | | | 2636007 | 818531 | 2620633 | 872379 | 8.95E-01 | -0.02 | 0.61 |
| RP_POS_173_98489a1_675 | | 173.9849 | 1.68 | | RP | POS | | | 2529888 | 944587 | 2513422 | 891320 | 8.95E-01 | -0.02 | 0.89 |
| RP_POS_184_99544a15_432 | | 184.9954 | 15.43 | | RP | POS | | | 482262 | 273395 | 477646 | 238648 | 8.95E-01 | -0.02 | 0.37 |
| RP_POS_236_07143a15_266 | | 236.0714 | 15.27 | | RP | POS | | | 2.1E+08 | 1E+08 | 2.1E+08 | 7.9E+07 | 8.95E-01 | -0.02 | 0.49 |
| RP_POS_358_0293a15_434 | | 358.0293 | 15.43 | | RP | POS | | | 2527150 | 912346 | 2509253 | 1055562 | 8.95E-01 | -0.02 | 0.41 |
| RP_POS_492_40271a11_706 | | 492.4027 | 11.71 | | RP | POS | | | 504002 | 84924.7 | 502509 | 81357.7 | 8.95E-01 | -0.02 | 0.76 |
| HILIC_NEG_174_08839a6_378 | Citrulline | 174.0884 | 6.38 | | HILIC | NEG | 131.0829(100), 132.085(2) | 1 | 427382 | 110946 | 425384 | 111478 | 8.95E-01 | -0.02 | 0.39 |

**Supplemantary table 1**: The non-targeted metabolomics results ordered by the VIP value (p value < 0.0003 are shaded dark green, p value < 0.05 are shaded light green, d values below 0 are blue and above 0 are red)

| | | | | | | | | | Cases | | Controls | | Welch test | Cohen's | |
|---|---|---|---|---|---|---|---|---|---|---|---|---|---|---|---|
| Feature_ID | Metabolite_name | Average_Mz | Average_RT_min_ | HMDB ID | RP_HILIC | POS_NEG | Cureted_MSMS | ID_level | Mean | SD | Mean2 | SD3 | p | d | VIP |
| RP_POS_327_07742a13_697 | | 327.0774 | 13.70 | | RP | POS | 327.0771(100), 233.0355(25) | | 1031019 | 590943 | 1042169 | 652975 | 8.96E-01 | 0.02 | 0.54 |
| HILIC_NEG_234_82047a6_91 | | 234.8205 | 6.91 | | HILIC | NEG | | | 829418 | 367172 | 836015 | 369822 | 8.96E-01 | 0.02 | 0.44 |
| RP_NEG_412_19214a11_592 | | 412.1921 | 11.59 | | RP | NEG | | | 1609435 | 287567 | 1614569 | 288238 | 8.96E-01 | 0.02 | 0.66 |
| RP_NEG_464_31509a11_063 | | 464.3151 | 11.06 | | RP | NEG | | | 1189483 | 413329 | 1197616 | 484298 | 8.96E-01 | 0.02 | 0.64 |
| RP_POS_252_1076a0_545 | | 252.1076 | 0.55 | | RP | POS | | | 650195 | 243443 | 654463 | 234202 | 8.96E-01 | 0.02 | 0.62 |
| RP_POS_659_9552a9_922 | | 659.9552 | 9.92 | | RP | POS | | | 664566 | 343136 | 671017 | 374169 | 8.96E-01 | 0.02 | 0.69 |
| HILIC_NEG_132_92348a3_17 | | 132.9235 | 3.17 | | HILIC | NEG | | | 1.5E+07 | 912996 | 1.5E+07 | 865861 | 8.96E-01 | -0.02 | 0.82 |
| HILIC_POS_277_09827a4_35 | | 277.0983 | 4.35 | | HILIC | POS | | | 334009 | 483377 | 323376 | 671092 | 8.96E-01 | -0.02 | 0.35 |
| RP_POS_207_96626a0_67 | | 207.9663 | 0.67 | | RP | POS | | | 279507 | 65933.7 | 278379 | 61033.1 | 8.96E-01 | -0.02 | 0.62 |
| RP_POS_726_51074a10_219 | | 726.5107 | 10.22 | | RP | POS | | | 2340513 | 1010434 | 2321167 | 1140135 | 8.96E-01 | -0.02 | 0.73 |
| RP_POS_424_34106a9_271 | Linoleyl carnitine | 424.3411 | 9.27 | | RP | POS | 85.0293(100), 424.3443(60) | 2 | 6593284 | 2168713 | 6629432 | 1884487 | 8.96E-01 | 0.02 | 0.65 |
| RP_NEG_407_20786a6_981 | Cortisol | 407.2079 | 6.98 | | RP | NEG | 331.193(100), 297.1474(25), 315.1596(9) | 2 | 8039647 | 2768899 | 8095847 | 3411116 | 8.96E-01 | 0.02 | 0.39 |
| RP_NEG_865_09283a9_583 | | 865.0928 | 9.58 | | RP | NEG | | | 1950345 | 192057 | 1953897 | 207509 | 8.97E-01 | 0.02 | 0.26 |
| RP_POS_233_07693a2_227 | | 233.0769 | 2.23 | | RP | POS | | | 4404455 | 910662 | 4422665 | 1123413 | 8.97E-01 | 0.02 | 0.59 |
| RP_NEG_309_85675a15_593 | | 309.8568 | 15.59 | | RP | NEG | | | 842139 | 405598 | 835028 | 398732 | 8.97E-01 | -0.02 | 0.38 |
| RP_POS_377_05954a15_559 | | 377.0595 | 15.56 | | RP | POS | | | 4525038 | 2198317 | 4486293 | 2188258 | 8.97E-01 | -0.02 | 0.59 |
| RP_POS_418_20648a4_193 | | 418.2065 | 4.19 | | RP | POS | | | 562441 | 239409 | 558266 | 234441 | 8.97E-01 | -0.02 | 0.7 |
| RP_POS_461_99481a9_74 | | 461.9948 | 9.74 | | RP | POS | | | 128405 | 76752.3 | 127109 | 69580 | 8.97E-01 | -0.02 | 0.4 |
| RP_POS_485_02048a10_53 | | 485.0205 | 10.53 | | RP | POS | | | 272103 | 157716 | 269421 | 146818 | 8.97E-01 | -0.02 | 0.71 |
| RP_POS_589_41571a11_553 | | 589.4157 | 11.55 | | RP | POS | | | 715374 | 603801 | 703932 | 678233 | 8.97E-01 | -0.02 | 0.56 |
| RP_POS_724_52722a13_362 | | 724.5272 | 13.36 | | RP | POS | | | 6293313 | 2146104 | 6255446 | 2151871 | 8.97E-01 | -0.02 | 0.74 |
| HILIC_NEG_144_92352a5_358 | | 144.9235 | 5.36 | | HILIC | NEG | | | 5.2E+07 | 1.1E+07 | 5.2E+07 | 9480515 | 8.98E-01 | 0.02 | 0.39 |
| RP_NEG_404_00308a0_682 | | 404.0031 | 0.68 | | RP | NEG | | | 265747 | 80541.2 | 267029 | 65468.1 | 8.98E-01 | 0.02 | 0.25 |
| RP_NEG_893_1933a9_848 | | 893.1933 | 9.85 | | RP | NEG | | | 5816431 | 335840 | 5821961 | 297869 | 8.98E-01 | 0.02 | 0.57 |
| RP_POS_633_44275a11_43 | | 633.4428 | 11.43 | | RP | POS | | | 183825 | 105145 | 185444 | 80839.6 | 8.98E-01 | 0.02 | 0.24 |
| RP_POS_824_52179a13_689 | | 824.5218 | 13.69 | | RP | POS | | | 1082400 | 549989 | 1093098 | 655386 | 8.98E-01 | 0.02 | 0.84 |
| RP_NEG_326_98907a6_598 | | 326.9891 | 6.60 | | RP | NEG | 242.9873(100), 326.988(10) | | 2160893 | 92618.2 | 2159288 | 91299.8 | 8.98E-01 | -0.02 | 0.24 |
| RP_NEG_476_77078a15_392 | | 476.7708 | 15.39 | | RP | NEG | | | 398503 | 123710 | 396259 | 130773 | 8.98E-01 | -0.02 | 0.83 |
| RP_POS_172_13348a10_253 | | 172.1335 | 10.25 | | RP | POS | | | 1.3E+07 | 936027 | 1.3E+07 | 1122307 | 8.98E-01 | -0.02 | 0.46 |
| RP_POS_252_18047a6_724 | | 252.1805 | 6.72 | | RP | POS | | | 804806 | 85949.7 | 803326 | 83536 | 8.98E-01 | -0.02 | 0.97 |
| RP_POS_264_19574a4_124 | | 264.1957 | 4.12 | | RP | POS | | | 2E+07 | 2.1E+07 | 2E+07 | 1.2E+07 | 8.98E-01 | -0.02 | 0.43 |
| RP_POS_287_63284a10_31 | | 287.6328 | 10.31 | | RP | POS | | | 4790013 | 1529282 | 4764506 | 1379770 | 8.98E-01 | -0.02 | 0.93 |
| RP_POS_297_18475a11_105 | | 297.1848 | 11.11 | | RP | POS | | | 574690 | 534248 | 565035 | 564348 | 8.98E-01 | -0.02 | 0.85 |
| RP_POS_796_58386a13_38 | | 796.5839 | 13.38 | | RP | POS | | | 1.4E+07 | 3740506 | 1.4E+07 | 3592114 | 8.98E-01 | -0.02 | 0.51 |
| HILIC_NEG_226_99539a5_118 | | 226.9954 | 5.12 | | HILIC | NEG | 142.9925(100), 226.9965(30) | | 1217574 | 617395 | 1228429 | 627533 | 8.99E-01 | 0.02 | 0.44 |
| RP_NEG_693_12482a9_417 | | 693.1248 | 9.42 | | RP | NEG | | | 742392 | 53820.3 | 743331 | 53990.2 | 8.99E-01 | 0.02 | 0.44 |
| RP_POS_219_17409a15_175 | | 219.1741 | 15.18 | | RP | POS | | | 447950 | 333966 | 453284 | 284066 | 8.99E-01 | 0.02 | 0.51 |
| RP_POS_337_1669a13_225 | | 337.1669 | 13.23 | | RP | POS | | | 461813 | 382832 | 469066 | 448543 | 8.99E-01 | 0.02 | 0.87 |
| RP_POS_491_84717a9_598 | | 491.8472 | 9.60 | | RP | POS | | | 1301335 | 661470 | 1314568 | 835054 | 8.99E-01 | 0.02 | 0.66 |
| RP_POS_501_39706a13_808 | | 501.3971 | 13.81 | | RP | POS | | | 1908708 | 1453184 | 1939518 | 2007182 | 8.99E-01 | 0.02 | 0.49 |
| HILIC_NEG_335_04984a4_822 | | 335.0498 | 4.82 | | HILIC | NEG | | | 445211 | 184969 | 442056 | 178863 | 8.99E-01 | -0.02 | 0.33 |
| HILIC_NEG_357_05551a1_449 | | 357.0555 | 1.45 | | HILIC | NEG | | | 352639 | 105232 | 350966 | 88374.4 | 8.99E-01 | -0.02 | 0.75 |
| RP_NEG_414_61227a15_493 | | 414.6123 | 15.49 | | RP | NEG | | | 300886 | 77167.5 | 299646 | 65966.4 | 8.99E-01 | -0.02 | 0.54 |
| RP_POS_129_05479a9_533 | | 129.0548 | 9.53 | | RP | POS | | | 890251 | 370035 | 883400 | 416789 | 8.99E-01 | -0.02 | 0.61 |
| RP_POS_245_22598a10_513 | | 245.2260 | 10.51 | | RP | POS | | | 653631 | 235961 | 648616 | 326465 | 8.99E-01 | -0.02 | 0.9 |
| RP_POS_436_7608a13_947 | | 436.7608 | 13.95 | | RP | POS | | | 531469 | 366841 | 525529 | 319727 | 8.99E-01 | -0.02 | 0.47 |
| HILIC_POS_244_07924a5_088 | | 244.0792 | 5.09 | | HILIC | POS | | | 1794725 | 917916 | 1811265 | 999272 | 9.00E-01 | 0.02 | 0.55 |
| RP_NEG_466_29321a10_701 | | 466.2932 | 10.70 | | RP | NEG | | | 313985 | 103663 | 315849 | 112325 | 9.00E-01 | 0.02 | 0.77 |
| RP_NEG_544_75757a15_38 | | 544.7576 | 15.38 | | RP | NEG | | | 581825 | 108677 | 583728 | 111678 | 9.00E-01 | 0.02 | 0.36 |
| RP_POS_408_71475a7_534 | | 408.7148 | 7.53 | | RP | POS | | | 304495 | 52430.5 | 305361 | 48802.4 | 9.00E-01 | 0.02 | 0.42 |
| RP_NEG_268_90698a15_389 | | 268.9070 | 15.39 | | RP | NEG | | | 456631 | 177841 | 453446 | 192761 | 9.00E-01 | -0.02 | 0.4 |
| RP_NEG_436_57663a15_491 | | 436.5766 | 15.49 | | RP | NEG | | | 479929 | 53067.5 | 479103 | 43242.6 | 9.00E-01 | -0.02 | 0.22 |
| RP_POS_239_12779a7_333 | | 239.1278 | 7.33 | | RP | POS | | | 1873955 | 1133811 | 1852940 | 1297708 | 9.00E-01 | -0.02 | 0.6 |
| RP_POS_146_06027a2_482 | Indole-3-carboxyaldehyde | 146.0603 | 2.48 | | RP | POS | 118.0657(100), 146.0602(50) | 2 | 1.5E+07 | 2989344 | 1.5E+07 | 2689814 | 9.00E-01 | -0.02 | 0.61 |
| RP_POS_353_15088a9_556 | 2-(4-(3,4-dihydro-2H-benzo[b][1,4]dioxepin-7-yl)-3-methyl-1H-pyrazol-5-yl)-5-methoxyphenol | 353.1509 | 9.56 | | RP | POS | 353.1507(100) | 2 | 968037 | 355596 | 962050 | 339340 | 9.00E-01 | -0.02 | 0.34 |
| RP_POS_201_08865a5_729 | | 201.0887 | 5.73 | | RP | POS | | | 485157 | 274011 | 480509 | 272540 | 9.01E-01 | -0.02 | 1.02 |
| RP_POS_358_25864a7_096 | | 358.2586 | 7.10 | | RP | POS | | | 575830 | 485407 | 568389 | 395905 | 9.01E-01 | -0.02 | 0.2 |
| RP_POS_378_19299a5_941 | | 378.1930 | 5.94 | | RP | POS | | | 325385 | 148218 | 323025 | 130439 | 9.01E-01 | -0.02 | 0.62 |
| RP_POS_427_35666a12_651 | | 427.3567 | 12.65 | | RP | POS | | | 238201 | 148221 | 235848 | 130870 | 9.01E-01 | -0.02 | 0.42 |
| RP_POS_299_08911a4_257 | | 299.0891 | 4.26 | | RP | POS | 299.1479(100) | | 508932 | 163854 | 511262 | 111436 | 9.02E-01 | 0.02 | 0.62 |
| RP_NEG_221_85178a15_495 | | 221.8518 | 15.50 | | RP | NEG | | | 232721 | 74169.7 | 233925 | 68812.7 | 9.02E-01 | 0.02 | 0.52 |
| RP_NEG_726_45551a10_775 | | 726.4555 | 10.78 | | RP | NEG | | | 104171 | 66534.9 | 105306 | 68378 | 9.02E-01 | 0.02 | 0.9 |
| RP_POS_473_32626a9_333 | | 473.3263 | 9.33 | | RP | POS | | | 1286324 | 2029732 | 1324167 | 2418586 | 9.02E-01 | 0.02 | 0.71 |
| HILIC_POS_172_1331a3_253 | | 172.1331 | 3.25 | | HILIC | POS | | | 377944 | 214590 | 374320 | 214124 | 9.02E-01 | -0.02 | 0.32 |
| RP_NEG_881_52612a12_357 | | 881.5261 | 12.36 | | RP | NEG | | | 1276391 | 204309 | 1272764 | 226230 | 9.02E-01 | -0.02 | 0.46 |
| RP_POS_225_11211a5_345 | | 225.1121 | 5.35 | | RP | POS | | | 946855 | 154514 | 944463 | 131485 | 9.02E-01 | -0.02 | 0.32 |
| RP_POS_265_17752a8_761 | | 265.1775 | 8.76 | | RP | POS | | | 1226027 | 172756 | 1223235 | 161295 | 9.02E-01 | -0.02 | 0.86 |
| RP_POS_377_20343a11_237 | | 377.2034 | 11.24 | | RP | POS | | | 280273 | 176327 | 277693 | 129957 | 9.02E-01 | -0.02 | 0.89 |
| RP_POS_518_40997a14_356 | | 518.4100 | 14.36 | | RP | POS | | | 1.1E+07 | 4456881 | 1.1E+07 | 3755116 | 9.02E-01 | -0.02 | 0.41 |
| HILIC_POS_311_18555a1_36 | | 311.1856 | 1.36 | | HILIC | POS | | | 99504.3 | 44078.7 | 98787.7 | 42230.6 | 9.03E-01 | -0.02 | 0.46 |
| RP_NEG_326_00497a0_688 | | 326.0050 | 0.69 | | RP | NEG | | | 279931 | 110629 | 278180 | 100128 | 9.03E-01 | -0.02 | 0.58 |
| RP_NEG_627_11462a9_337 | | 627.1146 | 9.34 | | RP | NEG | | | 2156836 | 107432 | 2154943 | 117509 | 9.03E-01 | -0.02 | 0.49 |
| RP_POS_193_15904a7_684 | | 193.1590 | 7.68 | | RP | POS | | | 2558160 | 136928 | 2555757 | 150415 | 9.03E-01 | -0.02 | 0.37 |
| RP_POS_207_0177a15_392 | | 207.0177 | 15.39 | | RP | POS | | | 605281 | 333461 | 600145 | 285870 | 9.03E-01 | -0.02 | 0.51 |
| RP_POS_429_37262a12_339 | | 429.3726 | 12.34 | | RP | POS | | | 3687312 | 1466579 | 3667871 | 846912 | 9.03E-01 | -0.02 | 0.45 |
| HILIC_POS_280_98688a4_844 | | 280.9869 | 4.84 | | HILIC | POS | | | 99008.5 | 38067.1 | 99647 | 39457 | 9.04E-01 | 0.02 | 0.44 |
| RP_POS_267_64655a10_356 | | 267.6466 | 10.36 | | RP | POS | | | 1714861 | 857688 | 1729664 | 925549 | 9.04E-01 | 0.02 | 1.2 |
| RP_POS_317_11469a9_522 | | 317.1147 | 9.52 | | RP | POS | | | 1638192 | 525947 | 1647252 | 564481 | 9.04E-01 | 0.02 | 1.04 |
| RP_POS_371_10037a10_251 | | 371.1004 | 10.25 | | RP | POS | | | 644629 | 333203 | 650555 | 379712 | 9.04E-01 | 0.02 | 0.56 |
| RP_POS_663_26727a5_79 | | 663.2673 | 5.79 | | RP | POS | | | 3616504 | 2036207 | 3646874 | 1648055 | 9.04E-01 | 0.02 | 0.6 |
| RP_NEG_207_103a6_823 | | 207.1030 | 6.82 | | RP | NEG | | | 739307 | 33589.5 | 738737 | 35347.7 | 9.04E-01 | -0.02 | 0.4 |
| RP_POS_339_23166a9_583 | | 339.2317 | 9.58 | | RP | POS | | | 1026906 | 582372 | 1016714 | 649386 | 9.04E-01 | -0.02 | 0.74 |
| HILIC_NEG_134_89473a7_747 | | 134.8947 | 7.75 | | HILIC | NEG | | | 2.4E+07 | 6252316 | 2.4E+07 | 5550499 | 9.05E-01 | 0.02 | 0.43 |
| RP_POS_1199_77368a12_616 | | 1199.7737 | 12.62 | | RP | POS | | | 5988749 | 7482199 | 6115412 | 8065067 | 9.05E-01 | 0.02 | 0.66 |
| RP_POS_374_77417a9_4 | | 374.7742 | 9.40 | | RP | POS | | | 268582 | 164021 | 271722 | 213764 | 9.05E-01 | 0.02 | 0.68 |
| HILIC_NEG_312_83606a4_897 | | 312.8361 | 4.90 | | HILIC | NEG | | | 1561068 | 340846 | 1555636 | 327086 | 9.05E-01 | -0.02 | 0.58 |
| RP_NEG_299_14905a9_97 | | 299.1491 | 9.97 | | RP | NEG | | | 126544 | 242807 | 123264 | 158230 | 9.05E-01 | -0.02 | 0.21 |
| RP_POS_236_07155a10_324 | | 236.0716 | 10.32 | | RP | POS | | | 4.1E+07 | 2E+07 | 4.1E+07 | 1.9E+07 | 9.05E-01 | -0.02 | 0.41 |
| RP_POS_785_35223a12_616 | | 785.3522 | 12.62 | | RP | POS | | | 454007 | 888670 | 440807 | 731173 | 9.05E-01 | -0.02 | 0.52 |
| RP_NEG_915_12732a9_818 | | 915.1273 | 9.82 | | RP | NEG | | | 3986962 | 212764 | 3990218 | 189798 | 9.06E-01 | 0.02 | 0.85 |
| RP_POS_227_17561a15_465 | | 227.1756 | 15.47 | | RP | POS | | | 226883 | 144480 | 229087 | 130712 | 9.06E-01 | 0.02 | 0.74 |
| RP_POS_630_46246a10_616 | | 630.4625 | 10.62 | | RP | POS | | | 1125098 | 626370 | 1136963 | 814092 | 9.06E-01 | 0.02 | 0.75 |
| HILIC_NEG_259_00113a5_114 | | 259.0011 | 5.11 | | HILIC | NEG | | | 1014069 | 497062 | 1006200 | 477879 | 9.06E-01 | -0.02 | 0.3 |
| HILIC_POS_248_14915a4_158 | | 248.1492 | 4.16 | | HILIC | POS | | | 1958315 | 1.1E+07 | 1813386 | 6549118 | 9.06E-01 | -0.02 | 0.81 |
| RP_NEG_355_28616a11_632 | | 355.2862 | 11.63 | | RP | NEG | | | 208584 | 440062 | 201822 | 400476 | 9.06E-01 | -0.02 | 0.29 |
| RP_POS_390_81418a15_434 | | 390.8142 | 15.43 | | RP | POS | | | 217842 | 118084 | 215883 | 125025 | 9.06E-01 | -0.02 | 0.53 |
| HILIC_NEG_564_33173a1_224 | LysoPC 18:2 | 564.3317 | 1.22 | | HILIC | NEG | 279.2341(100), 504.3128(20) | 2 | 2724039 | 929292 | 2708400 | 1008831 | 9.06E-01 | -0.02 | 0.55 |
| RP_POS_506_42038a12_317 | | 506.4204 | 12.32 | | RP | POS | | | 481215 | 293363 | 486101 | 316213 | 9.07E-01 | 0.02 | 0.86 |
| RP_POS_512_33423a9_954 | | 512.3342 | 9.95 | | RP | POS | | | 1173779 | 402366 | 1180816 | 467323 | 9.07E-01 | 0.02 | 1.05 |
| RP_NEG_311_16852a10_321 | | 311.1685 | 10.32 | | RP | NEG | 311.1699(100) | | 1.1E+07 | 9555952 | 1.1E+07 | 1.1E+07 | 9.07E-01 | -0.02 | 0.76 |
| HILIC_NEG_244_9854a0_935 | | 244.9854 | 0.94 | | HILIC | NEG | | | 1159771 | 96846 | 1158130 | 108104 | 9.07E-01 | -0.02 | 0.45 |
| HILIC_POS_427_26648a5_926 | | 427.2665 | 5.93 | | HILIC | POS | | | 486114 | 167381 | 483596 | 149081 | 9.07E-01 | -0.02 | 0.64 |
| RP_NEG_1149_2428a9_789 | | 1149.2428 | 9.79 | | RP | NEG | | | 4507414 | 494521 | 4499121 | 536055 | 9.07E-01 | -0.02 | 0.43 |
| RP_NEG_449_15143a8_359 | | 449.1514 | 8.36 | | RP | NEG | | | 100651 | 76432.6 | 99378.2 | 81745.7 | 9.07E-01 | -0.02 | 0.56 |
| RP_POS_473_34518a10_447 | | 473.3452 | 10.45 | | RP | POS | | | 2253326 | 524746 | 2244691 | 555236 | 9.07E-01 | -0.02 | 0.97 |
| RP_POS_846_54053a13_429 | | 846.5405 | 13.43 | | RP | POS | | | 1522208 | 1456465 | 1496022 | 1769484 | 9.07E-01 | -0.02 | 0.36 |
| HILIC_POS_132_10207a3_339 | Alanine betaine | 132.1021 | 3.34 | | HILIC | POS | 132.1028(100), 58.074(10) | 2 | 2999018 | 7567578 | 3150762 | 1.1E+07 | 9.07E-01 | 0.02 | 0.41 |
| HILIC_POS_179_00484a3_654 | | 179.0048 | 3.65 | | HILIC | POS | | | 1859625 | 3360112 | 1925171 | 4687667 | 9.08E-01 | 0.02 | 0.36 |
| RP_POS_263_90164a15_539 | | 263.9016 | 15.54 | | RP | POS | | | 284115 | 129118 | 286184 | 132682 | 9.08E-01 | 0.02 | 0.53 |
| RP_POS_322_18005a6_033 | | 322.1801 | 6.03 | | RP | POS | | | 227191 | 222015 | 230675 | 220941 | 9.08E-01 | 0.02 | 0.75 |
| RP_POS_414_30008a7_286 | | 414.3001 | 7.29 | | RP | POS | | | 538369 | 1492519 | 557257 | 852857 | 9.08E-01 | 0.02 | 0.71 |
| HILIC_POS_101_0715a6_137 | | 101.0715 | 6.14 | | HILIC | POS | | | 1201073 | 178731 | 1198356 | 165334 | 9.08E-01 | -0.02 | 0.75 |
| HILIC_POS_352_24615a5_08 | | 352.2462 | 5.08 | | HILIC | POS | | | 229210 | 159992 | 226902 | 133964 | 9.08E-01 | -0.02 | 0.69 |
| RP_POS_539_29803a10_784 | | 539.2980 | 10.78 | | RP | POS | | | 366197 | 387692 | 360767 | 297986 | 9.08E-01 | -0.02 | 0.89 |
| RP_POS_149_01199a8_614 | | 149.0120 | 8.61 | | RP | POS | 149.023(100), 121.0287(25) | | 3212875 | 829445 | 3224756 | 691178 | 9.09E-01 | 0.02 | 0.76 |
| RP_POS_402_35788a9_314 | | 402.3579 | 9.31 | | RP | POS | 283.2647(100) | | 1661303 | 3508450 | 1729967 | 5047342 | 9.09E-01 | 0.02 | 0.69 |
| HILIC_NEG_279_03967a1_455 | | 279.0397 | 1.46 | | HILIC | NEG | | | 298684 | 82089.3 | 299805 | 61398.7 | 9.09E-01 | 0.02 | 0.61 |
| RP_NEG_397_24515a11_593 | | 397.2452 | 11.59 | | RP | NEG | | | 557860 | 172463 | 560329 | 143493 | 9.09E-01 | 0.02 | 0.42 |
| RP_POS_462_82755a9_387 | | 462.8276 | 9.39 | | RP | POS | | | 469028 | 244489 | 472854 | 243742 | 9.09E-01 | 0.02 | 0.81 |
| HILIC_POS_520_33972a1_219 | LysoPC(0:0/18:2) | 520.3397 | 1.22 | | HILIC | POS | 184.0743(100) | 3 | 7.9E+07 | 3.3E+07 | 7.8E+07 | 3.3E+07 | 9.09E-01 | -0.02 | 0.64 |
| RP_POS_144_0656a1_84 | | 144.0656 | 1.84 | | RP | POS | | | 1070697 | 435559 | 1063614 | 468652 | 9.09E-01 | -0.02 | 0.28 |
| RP_POS_154_01237a5_79 | | 154.0124 | 5.79 | | RP | POS | | | 500551 | 217875 | 497481 | 176829 | 9.09E-01 | -0.02 | 0.56 |
| RP_POS_282_27917a9_848 | | 282.2792 | 9.85 | | RP | POS | | | 1090873 | 170023 | 1088055 | 188787 | 9.09E-01 | -0.02 | 0.53 |
| RP_POS_300_03802a15_363 | | 300.0380 | 15.36 | | RP | POS | | | 467074 | 217998 | 463643 | 222728 | 9.09E-01 | -0.02 | 0.56 |
| HILIC_NEG_132_90024a0_708 | | 132.9002 | 0.71 | | HILIC | NEG | | | 788532 | 220904 | 791944 | 220934 | 9.10E-01 | 0.02 | 0.54 |
| HILIC_POS_258_16965a1_266 | | 258.1697 | 1.27 | | HILIC | POS | | | 262654 | 119412 | 264611 | 131231 | 9.10E-01 | 0.02 | 0.88 |
| RP_NEG_393_17145a10_72 | | 393.1715 | 10.72 | | RP | NEG | | | 177280 | 101515 | 178960 | 113698 | 9.10E-01 | 0.02 | 0.26 |
| RP_NEG_540_66107a10_489 | | 540.6611 | 10.49 | | RP | NEG | | | 666597 | 237383 | 670532 | 270500 | 9.10E-01 | 0.02 | 0.57 |
| RP_POS_186_14914a6_27 | | 186.1491 | 6.27 | | RP | POS | | | 2921162 | 443976 | 2927999 | 436547 | 9.10E-01 | 0.02 | 0.84 |
| HILIC_POS_179_04057a3_147 | | 179.0406 | 3.15 | | HILIC | POS | | | 374856 | 109667 | 373044 | 122863 | 9.10E-01 | -0.02 | 0.45 |
| RP_NEG_134_89476a2_164 | | 134.8948 | 2.16 | | RP | NEG | | | 2231826 | 300635 | 2226639 | 362241 | 9.10E-01 | -0.02 | 0.47 |
| RP_NEG_504_30991a10_31 | | 504.3099 | 10.31 | | RP | NEG | | | 6603085 | 1808144 | 6574221 | 1900633 | 9.10E-01 | -0.02 | 0.62 |
| RP_NEG_681_29718a10_567 | | 681.2972 | 10.57 | | RP | NEG | | | 1657169 | 601998 | 1643947 | 1019470 | 9.10E-01 | -0.02 | 0.73 |
| RP_POS_153_0051a15_392 | | 153.0051 | 15.39 | | RP | POS | | | 1855336 | 868912 | 1842202 | 832834 | 9.10E-01 | -0.02 | 0.37 |
| RP_POS_306_87198a15_367 | | 306.8720 | 15.37 | | RP | POS | | | 479731 | 206847 | 476298 | 234398 | 9.10E-01 | -0.02 | 0.81 |
| RP_POS_582_59259a14_047 | | 582.5926 | 14.05 | | RP | POS | | | 2391809 | 208163 | 2388292 | 240496 | 9.10E-01 | -0.02 | 0.68 |
| HILIC_POS_150_11237a0_739 | | 150.1124 | 0.74 | | HILIC | POS | 150.1128(100) | | 1770920 | 857840 | 1785486 | 1034437 | 9.11E-01 | 0.02 | 0.66 |
| RP_POS_307_22424a8_423 | | 307.2242 | 8.42 | | RP | POS | | | 171542 | 90484.3 | 172942 | 93577.3 | 9.11E-01 | 0.02 | 0.73 |
| RP_NEG_142_99239a15_487 | | 142.9924 | 15.49 | | RP | NEG | | | 455544 | 106566 | 454017 | 93777.1 | 9.11E-01 | -0.02 | 0.49 |
| RP_POS_541_37604a9_986 | | 541.3760 | 9.99 | | RP | POS | | | 1492580 | 891002 | 1478789 | 920734 | 9.11E-01 | -0.02 | 0.58 |
| RP_POS_820_59851a9_885 | | 820.5985 | 9.89 | | RP | POS | | | 3012428 | 1501562 | 2989412 | 1518635 | 9.11E-01 | -0.02 | 0.95 |
| RP_POS_464_28305a9_096 | | 464.2831 | 9.10 | | RP | POS | 464.2785(100), 126.0229(60) | | 1703084 | 8402219 | 1839788 | 9564004 | 9.12E-01 | 0.02 | 0.4 |
| RP_NEG_230_01306a1_956 | | 230.0131 | 1.96 | | RP | NEG | | | 2599484 | 7066241 | 2713608 | 7967117 | 9.12E-01 | 0.02 | 0.51 |
| RP_NEG_703_1189a9_097 | | 703.1189 | 9.10 | | RP | NEG | | | 294894 | 24506.1 | 295321 | 30885.3 | 9.12E-01 | 0.02 | 0.66 |
| RP_POS_159_09157a2_483 | | 159.0916 | 2.48 | | RP | POS | | | 5787026 | 1160579 | 5804523 | 1144021 | 9.12E-01 | 0.02 | 0.52 |
| RP_POS_466_32907a11_064 | | 466.3291 | 11.06 | | RP | POS | | | 3008202 | 1108636 | 3026099 | 1254025 | 9.12E-01 | 0.02 | 0.7 |
| RP_POS_536_03107a10_176 | | 536.0311 | 10.18 | | RP | POS | | | 745823 | 320411 | 750975 | 358794 | 9.12E-01 | 0.02 | 0.35 |
| RP_POS_637_30548a10_568 | | 637.3055 | 10.57 | | RP | POS | | | 1.2E+07 | 4175563 | 1.2E+07 | 7508122 | 9.12E-01 | 0.02 | 0.67 |
| RP_NEG_556_29865a10_311 | | 556.2987 | 10.31 | | RP | NEG | | | 1279891 | 251652 | 1283922 | 284845 | 9.13E-01 | 0.02 | 0.61 |
| HILIC_POS_140_06825a4_864 | | 140.0683 | 4.86 | | HILIC | POS | 140.0692(100), 70.0974(50) | | 2516974 | 786083 | 2528760 | 788407 | 9.13E-01 | 0.01 | 0.66 |
| RP_NEG_206_97244a15_536 | | 206.9724 | 15.54 | | RP | NEG | | | 2752964 | 555608 | 2760953 | 513196 | 9.13E-01 | 0.01 | 0.33 |
| HILIC_NEG_127_05141a6_135 | | 127.0514 | 6.14 | | HILIC | NEG | | | 600818 | 104955 | 599274 | 101399 | 9.13E-01 | -0.01 | 0.82 |
| RP_NEG_427_11481a11_141 | | 427.1148 | 11.14 | | RP | NEG | | | 360801 | 36534.8 | 360271 | 34294.9 | 9.13E-01 | -0.01 | 0.65 |
| RP_POS_774_60046a14_094 | | 774.6005 | 14.09 | | RP | POS | | | 3225494 | 2931838 | 3185633 | 2427090 | 9.13E-01 | -0.01 | 0.65 |
| RP_NEG_218_77814a15_47 | | 218.7781 | 15.47 | | RP | NEG | | | 1243338 | 76657.4 | 1241924 | 107856 | 9.13E-01 | -0.02 | 0.47 |
| RP_POS_780_55237a13_224 | | 780.5524 | 13.22 | | RP | POS | | | 1.9E+08 | 5.8E+07 | 1.9E+08 | 6.1E+07 | 9.13E-01 | -0.02 | 0.42 |
| RP_POS_597_44873a11_595 | | 597.4487 | 11.60 | | RP | POS | | | 889636 | 1367607 | 905247 | 705367 | 9.14E-01 | 0.02 | 0.36 |
| HILIC_POS_366_2843a1_349 | | 366.2843 | 1.35 | | HILIC | POS | | | 797760 | 347634 | 802885 | 343696 | 9.14E-01 | 0.01 | 0.31 |
| RP_NEG_401_03839a0_683 | | 401.0384 | 0.68 | | RP | NEG | | | 389310 | 58435.2 | 390120 | 51865.3 | 9.14E-01 | 0.01 | 0.38 |
| RP_NEG_893_19299a9_675 | | 893.1930 | 9.68 | | RP | NEG | | | 1.2E+07 | 600078 | 1.2E+07 | 566275 | 9.14E-01 | 0.01 | 0.74 |
| RP_POS_331_32111a11_464 | | 331.3211 | 11.46 | | RP | POS | | | 258663 | 70790.6 | 259672 | 66907.9 | 9.14E-01 | 0.01 | 0.53 |

**Supplemantary table 1**: The non-targeted metabolomics results ordered by the VIP value (p value < 0.0003 are shaded dark green, p value < 0.05 are shaded light green, d values below 0 are blue and above 0 are red)

| | | | | | | | | | Cases | | Controls | | Welch test | Cohen's | |
|---|---|---|---|---|---|---|---|---|---|---|---|---|---|---|---|
| Feature_ID | Metabolite_name | Average_Mz | Average_RT_min_ | HMDB ID | RP_HILIC | POS_NEG | Cureted_MSMS | ID_level | Mean | SD | Mean2 | SD3 | p | d | VIP |
| RP_POS_772_6134a11_843 | | 772.6134 | 11.84 | | RP | POS | | | 265966 | 250253 | 269480 | 229713 | 9.14E-01 | 0.01 | 0.33 |
| RP_NEG_152_97858a15_559 | | 152.9786 | 15.56 | | RP | NEG | | | 1080910 | 288314 | 1076577 | 295699 | 9.14E-01 | -0.01 | 0.61 |
| RP_NEG_248_96094a11_161 | | 248.9609 | 11.16 | | RP | NEG | | | 1643032 | 869602 | 1630637 | 810350 | 9.14E-01 | -0.01 | 0.4 |
| RP_NEG_255_19696a9_16 | | 255.1970 | 9.16 | | RP | NEG | | | 208044 | 30716.5 | 207555 | 35028.5 | 9.14E-01 | -0.01 | 1.01 |
| RP_NEG_283_70993a15_495 | | 283.7099 | 15.50 | | RP | NEG | | | 1504329 | 62006.7 | 1503401 | 63873 | 9.14E-01 | -0.01 | 0.41 |
| RP_POS_329_1571a5_53 | | 329.1571 | 5.53 | | RP | POS | | | 309554 | 165957 | 307053 | 170624 | 9.14E-01 | -0.01 | 0.88 |
| RP_POS_638_57111a11_977 | | 638.5711 | 11.98 | | RP | POS | | | 1694558 | 2405282 | 1662889 | 1884958 | 9.14E-01 | -0.01 | 0.57 |
| HILIC_POS_159_09175a4_109 | 1-Benzylimidazole | 159.0918 | 4.11 | | HILIC | POS | 159.0921(100) | 2 | 5734060 | 1014143 | 5719028 | 1025324 | 9.14E-01 | -0.01 | 0.72 |
| RP_NEG_599_52838a15_277 | | 599.5284 | 15.28 | | RP | NEG | | | 1791397 | 344859 | 1796268 | 324720 | 9.15E-01 | 0.01 | 0.45 |
| RP_POS_192_97954a15_394 | | 192.9795 | 15.39 | | RP | POS | | | 405082 | 226621 | 408116 | 188809 | 9.15E-01 | 0.01 | 0.37 |
| RP_POS_540_39026a14_353 | | 540.3903 | 14.35 | | RP | POS | | | 1E+07 | 4368051 | 1E+07 | 3641470 | 9.15E-01 | 0.01 | 0.68 |
| RP_POS_546_03131a10_024 | | 546.0313 | 10.02 | | RP | POS | | | 423810 | 243639 | 427266 | 228540 | 9.15E-01 | 0.01 | 0.53 |
| RP_POS_575_05756a10_42 | | 575.0576 | 10.42 | | RP | POS | | | 550886 | 225820 | 554575 | 273427 | 9.15E-01 | 0.01 | 0.49 |
| RP_NEG_1085_23096a9_917 | | 1085.2310 | 9.92 | | RP | NEG | | | 3349172 | 229089 | 3345805 | 231122 | 9.15E-01 | -0.01 | 0.45 |
| RP_NEG_363_17679a10_683 | | 363.1768 | 10.68 | | RP | NEG | | | 123242 | 44898.8 | 122623 | 40586.3 | 9.15E-01 | -0.01 | 1 |
| RP_NEG_391_28616a9_019 | | 391.2862 | 9.02 | | RP | NEG | | | 337347 | 437023 | 331087 | 422516 | 9.15E-01 | -0.01 | 0.29 |
| RP_POS_271_20569a10_692 | | 271.2057 | 10.69 | | RP | POS | | | 1525627 | 498099 | 1518119 | 523992 | 9.15E-01 | -0.01 | 0.54 |
| RP_POS_318_11752a12_142 | | 318.1175 | 12.14 | | RP | POS | | | 522029 | 1256832 | 503644 | 1252640 | 9.15E-01 | -0.01 | 0.65 |
| RP_POS_471_34714a10_27 | Glycyrrhetic acid | 471.3471 | 10.27 | | RP | POS | 471.3434(100) | 2 | 9748451 | 1.5E+07 | 9974445 | 1.6E+07 | 9.15E-01 | 0.01 | 0.12 |
| HILIC_POS_106_08685a4_8 | | 106.0869 | 4.80 | | HILIC | POS | 88.0761(100), 70.066(60) | | 9646738 | 3806166 | 9701021 | 3726564 | 9.16E-01 | 0.01 | 1.03 |
| RP_NEG_309_02582a0_67 | | 309.0258 | 0.67 | | RP | NEG | | | 195671 | 39266.7 | 196249 | 40766.9 | 9.16E-01 | 0.01 | 0.27 |
| RP_POS_560_8811a9_983 | | 560.8811 | 9.98 | | RP | POS | | | 3080219 | 1716565 | 3105015 | 1735785 | 9.16E-01 | 0.01 | 0.52 |
| HILIC_NEG_183_93459a4_241 | | 183.9346 | 4.24 | | HILIC | NEG | | | 3853939 | 902204 | 3838659 | 1180124 | 9.16E-01 | -0.01 | 0.28 |
| RP_NEG_325_31165a12_408 | | 325.3117 | 12.41 | | RP | NEG | | | 77489.5 | 29320.7 | 77062 | 29684.1 | 9.16E-01 | -0.01 | 0.72 |
| RP_POS_151_94971a15_424 | | 151.9497 | 15.42 | | RP | POS | | | 392362 | 299181 | 388213 | 278190 | 9.16E-01 | -0.01 | 0.57 |
| RP_POS_237_22156a9_645 | | 237.2216 | 9.65 | | RP | POS | | | 1598151 | 200629 | 1595216 | 205445 | 9.16E-01 | -0.01 | 0.82 |
| RP_POS_607_49768a11_593 | | 607.4977 | 11.59 | | RP | POS | | | 1233605 | 693044 | 1223500 | 701128 | 9.16E-01 | -0.01 | 1.13 |
| RP_POS_617_33673a14_353 | | 617.3367 | 14.35 | | RP | POS | | | 3605463 | 2534824 | 3563798 | 3151844 | 9.16E-01 | -0.01 | 0.91 |
| HILIC_NEG_118_92418a5_118 | | 118.9242 | 5.12 | | HILIC | NEG | | | 3256574 | 1083142 | 3272087 | 1098592 | 9.17E-01 | 0.01 | 0.17 |
| HILIC_NEG_200_85922a5_847 | | 200.8592 | 5.85 | | HILIC | NEG | | | 2839932 | 1163496 | 2856076 | 1107739 | 9.17E-01 | 0.01 | 0.29 |
| RP_POS_465_23346a4_98 | | 465.2335 | 4.98 | | RP | POS | | | 495196 | 360568 | 500432 | 369761 | 9.17E-01 | 0.01 | 0.99 |
| RP_POS_520_6969a9_721 | | 520.6969 | 9.72 | | RP | POS | | | 132857 | 83376.4 | 134072 | 87296.5 | 9.17E-01 | 0.01 | 0.38 |
| RP_POS_566_39886a9_58 | | 566.3989 | 9.58 | | RP | POS | | | 2273709 | 1252147 | 2291015 | 1170682 | 9.17E-01 | 0.01 | 1.05 |
| RP_POS_819_5722a13_784 | | 819.5722 | 13.78 | | RP | POS | | | 8469697 | 2E+07 | 8776089 | 2.3E+07 | 9.17E-01 | 0.01 | 0.49 |
| RP_POS_898_55359a13_279 | | 898.5536 | 13.28 | | RP | POS | | | 2190654 | 4372089 | 2251091 | 4167256 | 9.17E-01 | 0.01 | 0.53 |
| HILIC_NEG_234_82027a6_453 | | 234.8203 | 6.45 | | HILIC | NEG | | | 935132 | 426821 | 929700 | 336261 | 9.17E-01 | -0.01 | 0.31 |
| RP_NEG_369_17447a7_933 | | 369.1745 | 7.93 | | RP | NEG | | | 4.5E+07 | 1.6E+08 | 4.3E+07 | 9.5E+07 | 9.17E-01 | -0.01 | 0.46 |
| RP_POS_322_00781a15_432 | | 322.0078 | 15.43 | | RP | POS | | | 2040883 | 1205427 | 2022165 | 1396463 | 9.17E-01 | -0.01 | 0.66 |
| RP_POS_385_19843a9_721 | | 385.1984 | 9.72 | | RP | POS | | | 879985 | 461779 | 873886 | 397610 | 9.17E-01 | -0.01 | 0.94 |
| HILIC_NEG_262_98447a4_867 | | 262.9845 | 4.87 | | HILIC | NEG | | | 471710 | 174107 | 474058 | 160137 | 9.18E-01 | 0.01 | 0.62 |
| HILIC_POS_104_05339a4_639 | | 104.0534 | 4.64 | | HILIC | POS | | | 1573048 | 4339450 | 1632875 | 4200436 | 9.18E-01 | 0.01 | 0.43 |
| HILIC_POS_331_24719a3_992 | | 331.2472 | 3.99 | | HILIC | POS | | | 263627 | 165500 | 265848 | 150731 | 9.18E-01 | 0.01 | 0.27 |
| HILIC_POS_466_24609a0_783 | | 466.2461 | 0.78 | | HILIC | POS | | | 196876 | 86695.1 | 198097 | 87748.3 | 9.18E-01 | 0.01 | 0.42 |
| RP_NEG_400_86307a15_369 | | 400.8631 | 15.37 | | RP | NEG | | | 806205 | 134785 | 808136 | 137829 | 9.18E-01 | 0.01 | 0.34 |
| RP_NEG_883_16504a9_673 | | 883.1650 | 9.67 | | RP | NEG | | | 2557677 | 150462 | 2559600 | 123592 | 9.18E-01 | 0.01 | 0.73 |
| RP_POS_239_10667a8_023 | | 239.1067 | 8.02 | | RP | POS | | | 629014 | 269523 | 632843 | 274876 | 9.18E-01 | 0.01 | 0.76 |
| RP_POS_353_35245a11_159 | | 353.3525 | 11.16 | | RP | POS | | | 5.45E+23 | 5.73E+23 | 5.53E+23 | 5.59E+23 | 9.18E-01 | 0.01 | 1.17 |
| RP_POS_557_41241a10_541 | | 557.4124 | 10.54 | | RP | POS | | | 1267816 | 641157 | 1277518 | 726253 | 9.18E-01 | 0.01 | 0.79 |
| RP_NEG_231_96753a0_685 | | 231.9675 | 0.69 | | RP | NEG | | | 475342 | 121137 | 473734 | 108986 | 9.18E-01 | -0.01 | 0.79 |
| RP_POS_1039_6676a10_501 | | 1039.6676 | 10.50 | | RP | POS | | | 1513118 | 517750 | 1505539 | 558210 | 9.18E-01 | -0.01 | 0.39 |
| RP_POS_346_27399a10_896 | | 346.2740 | 10.90 | | RP | POS | | | 2.2E+07 | 1.1E+07 | 2.2E+07 | 1.2E+07 | 9.18E-01 | -0.01 | 0.85 |
| RP_POS_271_13272a8_746 | Parthenolide | 271.1327 | 8.75 | | RP | POS | | | 883144 | 66402.4 | 884045 | 63823.9 | 9.19E-01 | 0.01 | 0.25 |
| HILIC_NEG_144_92355a4_113 | | 144.9236 | 4.11 | | HILIC | NEG | | | 2.4E+07 | 8068196 | 2.4E+07 | 8246332 | 9.19E-01 | 0.01 | 0.32 |
| HILIC_NEG_144_92355a5_456 | | 144.9236 | 5.46 | | HILIC | NEG | | | 3.6E+07 | 1.5E+07 | 3.6E+07 | 1.6E+07 | 9.19E-01 | 0.01 | 0.84 |
| HILIC_POS_490_29141a1_349 | | 490.2914 | 1.35 | | HILIC | POS | | | 380785 | 174578 | 383290 | 185514 | 9.19E-01 | 0.01 | 0.27 |
| RP_NEG_551_12744a8_272 | | 551.1274 | 8.27 | | RP | NEG | | | 1249855 | 75019.7 | 1250858 | 69346.6 | 9.19E-01 | 0.01 | 0.97 |
| RP_POS_222_16988a7_276 | | 222.1699 | 7.28 | | RP | POS | | | 514939 | 75691 | 515978 | 73844.7 | 9.19E-01 | 0.01 | 0.85 |
| RP_POS_274_12186a0_626 | | 274.1219 | 0.63 | | RP | POS | | | 4636847 | 529877 | 4644665 | 594311 | 9.19E-01 | 0.01 | 0.58 |
| RP_POS_362_09421a15_394 | | 362.0942 | 15.39 | | RP | POS | | | 1087862 | 536344 | 1095447 | 548362 | 9.19E-01 | 0.01 | 0.45 |
| RP_POS_763_52759a10_039 | | 763.5276 | 10.04 | | RP | POS | | | 754127 | 375181 | 759183 | 352291 | 9.19E-01 | 0.01 | 0.59 |
| HILIC_POS_174_14909a0_717 | | 174.1491 | 0.72 | | HILIC | POS | | | 7477804 | 1822704 | 7452555 | 1805698 | 9.19E-01 | -0.01 | 0.51 |
| RP_NEG_186_11336a11_591 | | 186.1134 | 11.59 | | RP | NEG | | | 569768 | 75899.3 | 568663 | 81925.1 | 9.19E-01 | -0.01 | 0.88 |
| RP_NEG_342_8627a15_395 | | 342.8627 | 15.40 | | RP | NEG | | | 2513454 | 479638 | 2506872 | 469116 | 9.19E-01 | -0.01 | 0.94 |
| RP_NEG_504_30988a10_181 | | 504.3099 | 10.18 | | RP | NEG | | | 946193 | 340960 | 941304 | 362541 | 9.19E-01 | -0.01 | 0.6 |
| RP_POS_1194_81946a12_616 | | 1194.8195 | 12.62 | | RP | POS | | | 1.1E+07 | 1.6E+07 | 1.1E+07 | 1.6E+07 | 9.19E-01 | -0.01 | 0.62 |
| RP_POS_464_19159a6_469 | | 464.1916 | 6.47 | | RP | POS | | | 953553 | 1138473 | 936407 | 1313120 | 9.19E-01 | -0.01 | 0.34 |
| HILIC_POS_368_21988a1_333 | | 368.2199 | 1.33 | | HILIC | POS | | | 59390.4 | 22602.1 | 59738.5 | 27413.5 | 9.20E-01 | 0.01 | 0.48 |
| HILIC_POS_434_12558a4_873 | | 434.1256 | 4.87 | | HILIC | POS | | | 160716 | 130862 | 162506 | 128754 | 9.20E-01 | 0.01 | 0.5 |
| RP_NEG_197_80794a15_493 | | 197.8079 | 15.49 | | RP | NEG | | | 5.9E+07 | 1103436 | 5.9E+07 | 1260336 | 9.20E-01 | 0.01 | 0.33 |
| RP_POS_278_24567a10_714 | | 278.2457 | 10.71 | | RP | POS | | | 9334474 | 3403802 | 9382892 | 3649278 | 9.20E-01 | 0.01 | 1.12 |
| RP_POS_590_48193a14_347 | | 590.4819 | 14.35 | | RP | POS | | | 1932411 | 1536247 | 1954706 | 1683329 | 9.20E-01 | 0.01 | 0.54 |
| RP_POS_646_20636a6_088 | | 646.2064 | 6.09 | | RP | POS | | | 543744 | 228125 | 547188 | 267468 | 9.20E-01 | 0.01 | 0.48 |
| RP_POS_780_58966a13_711 | | 780.5897 | 13.71 | | RP | POS | | | 2141127 | 1566269 | 2163226 | 1639863 | 9.20E-01 | 0.01 | 0.8 |
| RP_NEG_144_9234a14_348 | | 144.9234 | 14.35 | | RP | NEG | | | 839647 | 378866 | 834390 | 389793 | 9.20E-01 | -0.01 | 0.59 |
| RP_NEG_499_55179a15_494 | | 499.5518 | 15.49 | | RP | NEG | | | 467630 | 45632.6 | 466998 | 45909.9 | 9.20E-01 | -0.01 | 0.65 |
| RP_POS_266_15985a4_973 | | 266.1599 | 4.97 | | RP | POS | | | 1.6E+08 | 5.9E+08 | 1.5E+08 | 2.7E+08 | 9.20E-01 | -0.01 | 0.48 |
| RP_POS_608_42462a11_505 | | 608.4246 | 11.51 | | RP | POS | | | 140885 | 83558.6 | 139415 | 124052 | 9.20E-01 | -0.01 | 0.28 |
| HILIC_NEG_184_99159a0_845 | | 184.9916 | 0.85 | | HILIC | NEG | | | 391033 | 132113 | 389365 | 113292 | 9.21E-01 | -0.01 | 0.7 |
| HILIC_NEG_199_80544a0_896 | | 199.8054 | 0.90 | | HILIC | NEG | | | 100827 | 18966.9 | 100565 | 19804.4 | 9.21E-01 | -0.01 | 0.85 |
| RP_NEG_244_98534a15_476 | | 244.9853 | 15.48 | | RP | NEG | | | 2056714 | 218734 | 2053768 | 215040 | 9.21E-01 | -0.01 | 0.41 |
| RP_NEG_273_05325a3_966 | | 273.0533 | 3.97 | | RP | NEG | | | 823814 | 147341 | 821854 | 143047 | 9.21E-01 | -0.01 | 0.63 |
| RP_POS_141_091a3_047 | | 141.0910 | 3.05 | | RP | POS | | | 293382 | 121269 | 291623 | 137688 | 9.21E-01 | -0.01 | 0.5 |
| RP_POS_144_94801a15_407 | | 144.9480 | 15.41 | | RP | POS | | | 2.7E+07 | 7522779 | 2.7E+07 | 6684061 | 9.21E-01 | -0.01 | 1.04 |
| RP_POS_253_21602a9_821 | | 253.2160 | 9.82 | | RP | POS | | | 1281557 | 151601 | 1279559 | 144245 | 9.21E-01 | -0.01 | 0.63 |
| RP_POS_260_63892a10_226 | | 260.6389 | 10.23 | | RP | POS | | | 146927 | 71464 | 145955 | 72651.8 | 9.21E-01 | -0.01 | 0.73 |
| HILIC_NEG_116_92846a5_155 | | 116.9285 | 5.16 | | HILIC | NEG | | | 5329208 | 2319745 | 5358298 | 2024437 | 9.22E-01 | 0.01 | 0.54 |
| HILIC_POS_380_3009a1_322 | | 380.3009 | 1.32 | | HILIC | POS | | | 209966 | 78555.9 | 211086 | 86975.6 | 9.22E-01 | 0.01 | 0.23 |
| RP_NEG_266_91058a15_388 | | 266.9106 | 15.39 | | RP | NEG | | | 966054 | 315448 | 970201 | 307095 | 9.22E-01 | 0.01 | 0.41 |
| RP_POS_235_09645a6_397 | | 235.0965 | 6.40 | | RP | POS | | | 326441 | 76777 | 327506 | 81479.8 | 9.22E-01 | 0.01 | 0.58 |
| RP_POS_640_31769a7_235 | | 640.3177 | 7.24 | | RP | POS | | | 1E+07 | 680363 | 1E+07 | 631268 | 9.22E-01 | 0.01 | 0.91 |
| HILIC_NEG_167_02126a4_824 | | 167.0213 | 4.82 | | HILIC | NEG | 167.0206(100), 124.0149(55) | | 1E+07 | 2015616 | 1E+07 | 1981168 | 9.22E-01 | -0.01 | 0.3 |
| RP_POS_166_98489a11_193 | | 166.9849 | 11.19 | | RP | POS | | | 7428482 | 2935716 | 7391564 | 2606366 | 9.22E-01 | -0.01 | 0.35 |
| RP_POS_659_28705a10_569 | | 659.2871 | 10.57 | | RP | POS | | | 5554930 | 1920816 | 5518519 | 3247009 | 9.22E-01 | -0.01 | 0.72 |
| RP_POS_820_58392a13_236 | | 820.5839 | 13.24 | | RP | POS | | | 5860463 | 6607134 | 5765454 | 7534973 | 9.22E-01 | -0.01 | 0.68 |
| HILIC_NEG_187_04236a1_345 | 1-Hydroxy-2-naphthoic acid | 187.0424 | 1.35 | | HILIC | NEG | 187.0421(100) | 2 | 1E+07 | 3569312 | 1E+07 | 3127615 | 9.22E-01 | 0.01 | 0.79 |
| RP_POS_166_98488a10_415 | | 166.9849 | 10.42 | | RP | POS | | | 5437036 | 2577553 | 5470329 | 2450515 | 9.23E-01 | 0.01 | 0.63 |
| RP_POS_314_0148a0_643 | | 314.0148 | 0.64 | | RP | POS | | | 2345708 | 1015031 | 2357115 | 717745 | 9.23E-01 | 0.01 | 0.41 |
| RP_POS_316_05283a0_638 | | 316.0528 | 0.64 | | RP | POS | | | 369559 | 192790 | 366629 | 242453 | 9.23E-01 | -0.01 | 0.8 |
| HILIC_POS_224_12794a4_788 | 4-Hydroxybenzoylcholine | 224.1279 | 4.79 | | HILIC | POS | 224.1279(100), 165.0541(40) | 2 | 750736 | 215154 | 753742 | 236657 | 9.23E-01 | 0.01 | 0.26 |
| HILIC_NEG_136_89168a0_945 | | 136.8917 | 0.95 | | HILIC | NEG | | | 1.1E+07 | 2175549 | 1.1E+07 | 2018606 | 9.24E-01 | 0.01 | 0.29 |
| HILIC_POS_371_24139a4_885 | | 371.2414 | 4.89 | | HILIC | POS | | | 7264477 | 3079263 | 7302995 | 2864356 | 9.24E-01 | 0.01 | 0.57 |
| RP_NEG_391_00143a7_257 | | 391.0014 | 7.26 | | RP | NEG | | | 3526210 | 1047373 | 3540049 | 1085154 | 9.24E-01 | 0.01 | 0.42 |
| RP_POS_201_11194a4_9 | | 201.1119 | 4.90 | | RP | POS | | | 148679 | 83759.1 | 149751 | 80417.4 | 9.24E-01 | 0.01 | 0.61 |
| RP_POS_294_16943a7_025 | | 294.1694 | 7.03 | | RP | POS | | | 647484 | 58517.4 | 648296 | 65330.8 | 9.24E-01 | 0.01 | 1.01 |
| RP_POS_436_7999a9_625 | | 436.7999 | 9.63 | | RP | POS | | | 1094985 | 581092 | 1102636 | 589482 | 9.24E-01 | 0.01 | 0.59 |
| HILIC_NEG_115_92081a4_099 | | 115.9208 | 4.10 | | HILIC | NEG | | | 3.8E+07 | 1E+07 | 3.8E+07 | 8162569 | 9.24E-01 | -0.01 | 0.3 |
| HILIC_POS_138_05254a4_858 | | 138.0525 | 4.86 | | HILIC | POS | | | 1636646 | 916301 | 1625225 | 833741 | 9.24E-01 | -0.01 | 0.63 |
| HILIC_POS_289_1507a5_987 | | 289.1507 | 5.99 | | HILIC | POS | | | 96583.1 | 82086 | 95464.7 | 89273 | 9.24E-01 | -0.01 | 1.35 |
| RP_NEG_350_98727a7_457 | | 350.9873 | 7.46 | | RP | NEG | | | 1222569 | 384988 | 1217510 | 393240 | 9.24E-01 | -0.01 | 0.59 |
| RP_NEG_563_1004a9_243 | | 563.1004 | 9.24 | | RP | NEG | | | 755210 | 49410.7 | 754552 | 51174.1 | 9.24E-01 | -0.01 | 0.51 |
| RP_NEG_964_552a13_466 | | 964.5520 | 13.47 | | RP | NEG | | | 3155153 | 2059738 | 3183160 | 2291954 | 9.25E-01 | 0.01 | 0.54 |
| RP_POS_630_47754a9_73 | | 630.4775 | 9.73 | | RP | POS | | | 3712549 | 1592466 | 3732341 | 1488876 | 9.25E-01 | 0.01 | 0.7 |
| RP_NEG_437_2916a9_261 | | 437.2916 | 9.26 | | RP | NEG | 391.2837(100) | | 437522 | 594530 | 431331 | 355560 | 9.25E-01 | -0.01 | 0.74 |
| RP_NEG_193_81577a15_493 | | 193.8158 | 15.49 | | RP | NEG | | | 2720705 | 79488 | 2719669 | 82342.8 | 9.25E-01 | -0.01 | 0.38 |
| RP_POS_236_07153a0_557 | | 236.0715 | 0.56 | | RP | POS | | | 2.1E+07 | 2080785 | 2.1E+07 | 4982539 | 9.25E-01 | -0.01 | 0.47 |
| RP_POS_275_08017a6_498 | | 275.0802 | 6.50 | | RP | POS | | | 748826 | 380999 | 744104 | 354254 | 9.25E-01 | -0.01 | 1.2 |
| RP_POS_342_18298a5_65 | | 342.1830 | 5.65 | | RP | POS | | | 7958562 | 2841773 | 7923482 | 2629176 | 9.25E-01 | -0.01 | 0.63 |
| RP_POS_446_315a9_844 | | 446.3150 | 9.84 | | RP | POS | | | 796560 | 352190 | 791486 | 429315 | 9.25E-01 | -0.01 | 0.87 |
| RP_POS_601_5199a15_048 | | 601.5199 | 15.05 | | RP | POS | | | 1288517 | 1348374 | 1270875 | 1399659 | 9.25E-01 | -0.01 | 0.67 |
| HILIC_POS_187_03036a6_144 | | 187.0304 | 6.14 | | HILIC | POS | | | 373156 | 35631.8 | 373635 | 39723.1 | 9.26E-01 | 0.01 | 0.42 |
| RP_POS_144_17474a3_269 | | 144.1747 | 3.27 | | RP | POS | | | 714957 | 261318 | 718049 | 228549 | 9.26E-01 | 0.01 | 0.56 |
| RP_POS_351_10095a7_089 | | 351.1010 | 7.09 | | RP | POS | | | 3878762 | 2273314 | 3912324 | 2941885 | 9.26E-01 | 0.01 | 0.37 |
| RP_NEG_216_78024a15_327 | | 216.7802 | 15.33 | | RP | NEG | | | 1197132 | 110854 | 1195610 | 125833 | 9.26E-01 | -0.01 | 0.4 |
| RP_NEG_421_22729a11_113 | | 421.2273 | 11.11 | | RP | NEG | | | 129237 | 89701.1 | 128030 | 99136.4 | 9.26E-01 | -0.01 | 0.27 |
| RP_POS_271_09399a7_491 | | 271.0940 | 7.49 | | RP | POS | | | 5070319 | 1.1E+07 | 4930124 | 1.1E+07 | 9.26E-01 | -0.01 | 0.49 |
| RP_POS_331_18826a9_053 | | 331.1883 | 9.05 | | RP | POS | | | 1199604 | 120389 | 1198002 | 131240 | 9.26E-01 | -0.01 | 0.44 |
| RP_POS_724_70325a11_339 | | 724.7033 | 11.34 | | RP | POS | | | 285011 | 143452 | 283297 | 128122 | 9.26E-01 | -0.01 | 0.45 |
| RP_NEG_564_33099a10_181 | LysoPC 18:2 | 564.3310 | 10.18 | | RP | NEG | 279.2336(100), 504.312(15) | 1 | 3.6E+07 | 1.2E+07 | 3.6E+07 | 1.3E+07 | 9.26E-01 | -0.01 | 0.61 |
| RP_POS_359_31616a11_245 | 1-Monostearin | 359.3162 | 11.25 | | RP | POS | 57.071(100), 341.3055(75) | 2 | 2743643 | 938955 | 2754463 | 773431 | 9.26E-01 | 0.01 | 0.42 |
| RP_POS_511_38428a9_483 | | 511.3843 | 9.48 | | RP | POS | | | 395124 | 291942 | 398282 | 209453 | 9.27E-01 | 0.01 | 0.53 |
| HILIC_NEG_212_99638a0_934 | | 212.9964 | 0.93 | | HILIC | NEG | | | 333117 | 35684.8 | 332634 | 40445.7 | 9.27E-01 | -0.01 | 0.74 |
| HILIC_POS_170_11757a0_558 | | 170.1176 | 0.56 | | HILIC | POS | | | 998878 | 553279 | 992473 | 467188 | 9.27E-01 | -0.01 | 0.97 |
| RP_POS_252_04549a15_423 | | 252.0455 | 15.42 | | RP | POS | | | 3.2E+07 | 4832142 | 3.2E+07 | 4627279 | 9.28E-01 | 0.01 | 0.62 |
| RP_NEG_462_17773a6_469 | | 462.1777 | 6.47 | | RP | NEG | | | 372009 | 442474 | 366191 | 494058 | 9.28E-01 | -0.01 | 0.34 |
| RP_POS_543_72009a10_449 | | 543.7201 | 10.45 | | RP | POS | | | 849071 | 394849 | 844527 | 344855 | 9.28E-01 | -0.01 | 0.54 |
| RP_POS_717_59045a13_002 | | 717.5905 | 13.00 | | RP | POS | | | 612182 | 451986 | 606733 | 426640 | 9.28E-01 | -0.01 | 0.7 |
| RP_NEG_313_004a0_682 | | 313.0040 | 0.68 | | RP | NEG | | | 461092 | 146131 | 462788 | 133966 | 9.29E-01 | 0.01 | 0.63 |
| RP_POS_236_07144a14_255 | | 236.0714 | 14.26 | | RP | POS | | | 1.5E+08 | 6.7E+07 | 1.5E+08 | 7.1E+07 | 9.29E-01 | 0.01 | 0.25 |
| RP_POS_251_00017a8_09 | | 251.0002 | 8.09 | | RP | POS | | | 956160 | 272631 | 959342 | 254534 | 9.29E-01 | 0.01 | 0.58 |
| RP_POS_556_44281a10_457 | | 556.4428 | 10.46 | | RP | POS | | | 6078478 | 571584 | 6085319 | 549238 | 9.29E-01 | 0.01 | 0.68 |
| RP_POS_782_56873a13_119 | PC(18:2/18:2) | 782.5687 | 13.12 | | RP | POS | 184.0745(100), 782.5599(2) | 1 | 1.1E+09 | 2.9E+08 | 1.1E+09 | 2.3E+08 | 9.29E-01 | -0.01 | 0.56 |
| HILIC_POS_161_12856a3_067 | | 161.1286 | 3.07 | | HILIC | POS | | | 1149077 | 391898 | 1153963 | 420654 | 9.30E-01 | 0.01 | 0.5 |
| HILIC_POS_263_08688a7_749 | | 263.0869 | 7.75 | | HILIC | POS | | | 685190 | 60156.4 | 686089 | 85341.3 | 9.30E-01 | 0.01 | 0.33 |
| RP_NEG_325_04779a0_667 | | 325.0478 | 0.67 | | RP | NEG | | | 150085 | 45444.5 | 150567 | 34594.8 | 9.30E-01 | 0.01 | 0.35 |
| RP_NEG_326_75583a15_453 | | 326.7558 | 15.45 | | RP | NEG | | | 5280376 | 1698636 | 5300867 | 1729786 | 9.30E-01 | 0.01 | 0.38 |
| RP_POS_294_15488a6_515 | | 294.1549 | 6.52 | | RP | POS | | | 188025 | 71777.6 | 188863 | 67449.8 | 9.30E-01 | 0.01 | 0.55 |
| RP_POS_635_43207a9_73 | | 635.4321 | 9.73 | | RP | POS | | | 462751 | 285303 | 466755 | 368954 | 9.30E-01 | 0.01 | 0.9 |
| HILIC_NEG_87_00843a1_342 | | 87.0084 | 1.34 | | HILIC | NEG | | | 520147 | 179280 | 518126 | 158679 | 9.30E-01 | -0.01 | 0.67 |
| RP_POS_327_00766a8_091 | Tris(1-chloro-2-propyl)phosphate | 327.0077 | 8.09 | | RP | POS | 98.985(100) | 2 | 7198670 | 1380848 | 7184477 | 989047 | 9.30E-01 | -0.01 | 0.73 |
| HILIC_NEG_183_9344a0_941 | | 183.9344 | 0.94 | | HILIC | NEG | | | 4206889 | 358620 | 4211174 | 362002 | 9.31E-01 | 0.01 | 0.91 |
| HILIC_NEG_265_00009a4_863 | | 265.0001 | 4.86 | | HILIC | NEG | | | 838096 | 242461 | 840566 | 176025 | 9.31E-01 | 0.01 | 0.38 |
| HILIC_POS_72_08165a4_856 | | 72.0817 | 4.86 | | HILIC | POS | | | 3.5E+07 | 8406602 | 3.5E+07 | 6113198 | 9.31E-01 | 0.01 | 0.49 |
| RP_NEG_201_80229a15_493 | | 201.8023 | 15.49 | | RP | NEG | | | 5946550 | 146901 | 5948245 | 140984 | 9.31E-01 | 0.01 | 0.33 |
| RP_POS_200_97366a15_423 | | 200.9737 | 15.42 | | RP | POS | | | 1457509 | 572463 | 1449848 | 701956 | 9.31E-01 | -0.01 | 0.46 |
| RP_POS_421_73761a12_735 | | 421.7376 | 12.74 | | RP | POS | | | 548173 | 367445 | 543024 | 479821 | 9.31E-01 | -0.01 | 0.53 |
| RP_POS_520_3399a10_308 | LysoPC 18:2 (0:0/18:2) | 520.3399 | 10.31 | | RP | POS | 184.0743(100) | 1 | 1.1E+09 | 3.2E+08 | 1.1E+09 | 3.6E+08 | 9.31E-01 | 0.01 | 0.58 |
| RP_POS_233_06927a15_366 | | 233.0693 | 15.37 | | RP | POS | | | 525574 | 381466 | 529978 | 370103 | 9.32E-01 | 0.01 | 0.85 |
| RP_POS_273_22122a7_961 | | 273.2212 | 7.96 | | RP | POS | | | 266697 | 255965 | 269598 | 242440 | 9.32E-01 | 0.01 | 0.6 |
| RP_POS_530_37311a10_035 | | 530.3731 | 10.04 | | RP | POS | | | 259243 | 148899 | 261091 | 167326 | 9.32E-01 | 0.01 | 0.54 |
| RP_POS_256_29922a8_663 | | 256.2992 | 8.66 | | RP | POS | | | 319688 | 242723 | 316783 | 256982 | 9.32E-01 | -0.01 | 0.4 |
| RP_POS_368_84195a15_395 | | 368.8420 | 15.40 | | RP | POS | | | 536009 | 312680 | 532858 | 226256 | 9.32E-01 | -0.01 | 0.39 |
| RP_POS_820_52411a13_124 | | 820.5241 | 13.12 | | RP | POS | | | 1E+07 | 3391409 | 1E+07 | 3885555 | 9.32E-01 | -0.01 | 0.32 |

**Supplemantary table 1**: The non-targeted metabolomics results ordered by the VIP value (p value < 0.0003 are shaded dark green, p value < 0.05 are shaded light green, d values below 0 are blue and above 0 are red)

| | | | | | | | | | Cases | | Controls | | Welch test | Cohen's | |
|---|---|---|---|---|---|---|---|---|---|---|---|---|---|---|---|
| Feature_ID | Metabolite_name | Average_Mz | Average_RT_min_ | HMDB ID | RP_HILIC | POS_NEG | Cureted_MSMS | ID_level | Mean | SD | Mean2 | SD3 | p | d | VIP |
| RP_POS_285_07886a7_985 | Diazepam | 285.0789 | 7.99 | | RP | POS | 285.0778(100),154.0418(10) | 2 | 1968135 | 3305210 | 2008635 | 3585433 | 9.32E-01 | 0.01 | 0.73 |
| HILIC_POS_273_26468a6_958 | | 273.2647 | 6.96 | | HILIC | POS | | | 96738.4 | 44708.1 | 97243.4 | 43483.9 | 9.33E-01 | 0.01 | 0.62 |
| RP_POS_132_08092a2_482 | | 132.0809 | 2.48 | | RP | POS | | | 2242707 | 511660 | 2248953 | 573192 | 9.33E-01 | 0.01 | 0.48 |
| RP_POS_175_14832a8_922 | | 175.1483 | 8.92 | | RP | POS | | | 2891388 | 263469 | 2894318 | 248702 | 9.33E-01 | 0.01 | 0.2 |
| RP_POS_254_97151a0_585 | | 254.9715 | 0.59 | | RP | POS | | | 1115392 | 172141 | 1117484 | 190270 | 9.33E-01 | 0.01 | 0.35 |
| RP_POS_285_8858a15_387 | | 285.8858 | 15.39 | | RP | POS | | | 193783 | 135696 | 195383 | 141686 | 9.33E-01 | 0.01 | 0.71 |
| RP_POS_549_7218a10_169 | | 549.7218 | 10.17 | | RP | POS | | | 339114 | 196587 | 341845 | 265369 | 9.33E-01 | 0.01 | 0.35 |
| HILIC_NEG_147_95769a1_35 | | 147.9577 | 1.35 | | HILIC | NEG | | | 238221 | 75512.2 | 237358 | 75251.7 | 9.33E-01 | -0.01 | 0.62 |
| HILIC_POS_208_15437a0_802 | | 208.1544 | 0.80 | | HILIC | POS | | | 1820941 | 930239 | 1807397 | 1344805 | 9.33E-01 | -0.01 | 0.81 |
| RP_NEG_502_86084a15_348 | | 502.8608 | 15.35 | | RP | NEG | | | 1577675 | 93651 | 1576546 | 101718 | 9.33E-01 | -0.01 | 0.85 |
| RP_POS_1069_62488a10_29 | | 1069.6249 | 10.29 | | RP | POS | | | 654101 | 365147 | 649941 | 359923 | 9.33E-01 | -0.01 | 0.54 |
| RP_POS_272_14886a4_42 | | 272.1489 | 4.42 | | RP | POS | | | 103223 | 67261.9 | 102417 | 71709.8 | 9.33E-01 | -0.01 | 0.92 |
| RP_POS_163_03886a9_44 | | 163.0389 | 9.44 | | RP | POS | 163.0393(100) | | 2176881 | 176253 | 2178857 | 174089 | 9.34E-01 | 0.01 | 0.76 |
| HILIC_POS_315_12772a6_137 | | 315.1277 | 6.14 | | HILIC | POS | | | 374013 | 35716.6 | 374555 | 56250.8 | 9.34E-01 | 0.01 | 0.62 |
| RP_NEG_757_13544a9_484 | | 757.1354 | 9.48 | | RP | NEG | | | 674837 | 55875.3 | 675421 | 48229.9 | 9.34E-01 | 0.01 | 0.56 |
| RP_NEG_894_54877a13_121 | | 894.5488 | 13.12 | | RP | NEG | | | 1.2E+07 | 2209120 | 1.2E+07 | 2254387 | 9.34E-01 | 0.01 | 0.16 |
| RP_POS_230_98251a0_696 | | 230.9825 | 0.70 | | RP | POS | | | 519541 | 97639.7 | 520637 | 97269.1 | 9.34E-01 | 0.01 | 0.56 |
| HILIC_NEG_323_0134a0_591 | | 323.0134 | 0.59 | | HILIC | NEG | | | 635159 | 252646 | 632428 | 231452 | 9.34E-01 | -0.01 | 0.39 |
| RP_POS_251_12788a8_633 | | 251.1279 | 8.63 | | RP | POS | | | 2021141 | 303794 | 2017864 | 280246 | 9.34E-01 | -0.01 | 0.43 |
| RP_POS_194_11513a10_252 | | 194.1151 | 10.25 | | RP | POS | 194.155(100) | | 1777821 | 207060 | 1780163 | 215144 | 9.35E-01 | 0.01 | 0.58 |
| HILIC_POS_480_35315a4_035 | | 480.3532 | 4.04 | | HILIC | POS | | | 130499 | 87855.7 | 131422 | 78569.7 | 9.35E-01 | 0.01 | 0.61 |
| RP_NEG_326_98938a7_028 | | 326.9894 | 7.03 | | RP | NEG | | | 2399773 | 628100 | 2392623 | 653482 | 9.35E-01 | -0.01 | 0.62 |
| RP_POS_163_11166a13_244 | | 163.1117 | 13.24 | | RP | POS | | | 6214554 | 3366972 | 6175690 | 3574343 | 9.35E-01 | -0.01 | 0.36 |
| RP_POS_471_00507a10_076 | | 471.0051 | 10.08 | | RP | POS | | | 705091 | 430895 | 700805 | 334159 | 9.35E-01 | -0.01 | 0.45 |
| RP_POS_532_33728a10_722 | | 532.3373 | 10.72 | | RP | POS | | | 6578546 | 1803454 | 6557736 | 1926605 | 9.35E-01 | -0.01 | 0.8 |
| RP_POS_607_2912a10_606 | | 607.2912 | 10.61 | | RP | POS | | | 1172912 | 232712 | 1170229 | 244368 | 9.35E-01 | -0.01 | 0.52 |
| RP_POS_642_20862a11_294 | | 642.2086 | 11.29 | | RP | POS | | | 673350 | 189199 | 671380 | 168012 | 9.35E-01 | -0.01 | 0.4 |
| HILIC_NEG_89_02396a1_346 | Lactic acid | 89.0240 | 1.35 | | HILIC | NEG | 89.0244(100), 71.0128(9) | 1 | 3.9E+08 | 1.1E+08 | 3.9E+08 | 9.3E+07 | 9.35E-01 | 0.01 | 0.65 |
| HILIC_POS_217_10437a4_403 | 1,2,3,4-tetrahydro-6-methoxy-1-oxo-beta-carboline | 217.1044 | 4.40 | | HILIC | POS | 217.1044(100) | 2 | 509468 | 331152 | 513781 | 423798 | 9.35E-01 | 0.01 | 0.32 |
| RP_POS_275_0672a3_956 | | 275.0672 | 3.96 | | RP | POS | | | 263263 | 64170.4 | 263995 | 67796.8 | 9.36E-01 | 0.01 | 0.61 |
| RP_POS_663_45404a13_695 | CE(16:0) | 663.4540 | 13.70 | | RP | POS | 495.27(100), 439.2003(65) | 2 | 1.6E+08 | 3.6E+07 | 1.6E+08 | 4E+07 | 9.36E-01 | -0.01 | 0.86 |
| RP_NEG_434_87201a15_355 | | 434.8720 | 15.36 | | RP | NEG | | | 1996686 | 119453 | 1995394 | 116492 | 9.36E-01 | -0.01 | 0.87 |
| RP_POS_295_22385a9_645 | | 295.2239 | 9.65 | | RP | POS | | | 3664373 | 437248 | 3659633 | 421992 | 9.36E-01 | -0.01 | 0.9 |
| RP_POS_500_02942a10_517 | | 500.0294 | 10.52 | | RP | POS | | | 228866 | 140737 | 227252 | 151556 | 9.36E-01 | -0.01 | 0.39 |
| HILIC_POS_520_34039a1_111 | LysoPC 18:2 (0:0/18:2) | 520.3404 | 1.11 | | HILIC | POS | 184.0743(100) | 2 | 1.1E+07 | 9294366 | 1.1E+07 | 1.3E+07 | 9.36E-01 | 0.01 | 0.55 |
| HILIC_POS_118_08654a4_856 | | 118.0865 | 4.86 | | HILIC | POS | 118.0866(100) | | 1.3E+08 | 2.8E+07 | 1.3E+08 | 2.1E+07 | 9.37E-01 | 0.01 | 0.53 |
| HILIC_POS_123_08078a1_357 | | 123.0808 | 1.36 | | HILIC | POS | | | 132149 | 102955 | 131157 | 79978.7 | 9.37E-01 | -0.01 | 0.91 |
| HILIC_POS_208_03976a7_993 | | 208.0398 | 7.99 | | HILIC | POS | | | 1487248 | 602584 | 1480637 | 626750 | 9.37E-01 | -0.01 | 0.51 |
| RP_POS_358_11832a9_456 | | 358.1183 | 9.46 | | RP | POS | | | 130385 | 131434 | 128712 | 172351 | 9.37E-01 | -0.01 | 0.53 |
| RP_POS_514_37158a10_512 | | 514.3716 | 10.51 | | RP | POS | | | 293104 | 222326 | 290800 | 204258 | 9.37E-01 | -0.01 | 0.36 |
| HILIC_NEG_326_98935a1_366 | | 326.9894 | 1.37 | | HILIC | NEG | 242.9877(100), 326.988(10) | | 886249 | 352656 | 889795 | 320426 | 9.38E-01 | 0.01 | 0.45 |
| HILIC_NEG_254_97223a4_863 | | 254.9722 | 4.86 | | HILIC | NEG | | | 583081 | 151872 | 584495 | 112450 | 9.38E-01 | 0.01 | 0.43 |
| RP_POS_1037_65295a10_352 | | 1037.6530 | 10.35 | | RP | POS | | | 963022 | 569638 | 968953 | 545953 | 9.38E-01 | 0.01 | 0.69 |
| RP_POS_390_19824a10_845 | | 390.1982 | 10.85 | | RP | POS | | | 525009 | 414681 | 529259 | 385253 | 9.38E-01 | 0.01 | 0.54 |
| RP_POS_653_62195a11_315 | | 653.6220 | 11.32 | | RP | POS | | | 205271 | 95244.4 | 206429 | 118349 | 9.38E-01 | 0.01 | 0.56 |
| RP_POS_558_27496a11_148 | | 558.2750 | 11.15 | | RP | POS | | | 782323 | 235027 | 779747 | 246508 | 9.38E-01 | -0.01 | 0.59 |
| RP_POS_559_375a10_17 | | 559.3750 | 10.17 | | RP | POS | | | 247860 | 147791 | 246331 | 138631 | 9.38E-01 | -0.01 | 0.35 |
| RP_POS_794_50952a12_808 | | 794.5095 | 12.81 | | RP | POS | | | 2891373 | 2286808 | 2867445 | 2201459 | 9.38E-01 | -0.01 | 0.67 |
| HILIC_NEG_144_92337a1_067 | | 144.9234 | 1.07 | | HILIC | NEG | | | 2E+07 | 3652746 | 2E+07 | 4127656 | 9.39E-01 | 0.01 | 0.5 |
| HILIC_POS_232_02747a0_628 | | 232.0275 | 0.63 | | HILIC | POS | | | 891608 | 2184262 | 915507 | 2386071 | 9.39E-01 | 0.01 | 0.52 |
| RP_NEG_130_94437a11_142 | | 130.9444 | 11.14 | | RP | NEG | | | 7796088 | 919629 | 7805802 | 942926 | 9.39E-01 | 0.01 | 0.36 |
| RP_NEG_134_89481a15_496 | | 134.8948 | 15.50 | | RP | NEG | | | 1316459 | 257291 | 1319795 | 360741 | 9.39E-01 | 0.01 | 0.48 |
| RP_NEG_207_93095a15_648 | | 207.9310 | 15.65 | | RP | NEG | | | 5348570 | 2510567 | 5377665 | 2967503 | 9.39E-01 | 0.01 | 0.48 |
| RP_NEG_840_57648a13_365 | | 840.5765 | 13.37 | | RP | NEG | | | 1593160 | 483639 | 1598131 | 467668 | 9.39E-01 | 0.01 | 0.36 |
| RP_POS_307_18842a8_295 | | 307.1884 | 8.30 | | RP | POS | | | 684707 | 215143 | 686855 | 195513 | 9.39E-01 | 0.01 | 0.55 |
| RP_POS_471_33917a10_071 | | 471.3392 | 10.07 | | RP | POS | | | 350118 | 231951 | 352552 | 231296 | 9.39E-01 | 0.01 | 0.57 |
| RP_POS_743_15698a9_483 | | 743.1570 | 9.48 | | RP | POS | | | 859831 | 119396 | 861091 | 120962 | 9.39E-01 | 0.01 | 0.47 |
| RP_NEG_193_85739a11_144 | | 193.8574 | 11.14 | | RP | NEG | | | 136459 | 32991.8 | 136088 | 37247.8 | 9.39E-01 | -0.01 | 0.53 |
| RP_POS_426_34167a9_183 | | 426.3417 | 9.18 | | RP | POS | | | 733569 | 467747 | 728890 | 432493 | 9.39E-01 | -0.01 | 0.5 |
| RP_POS_393_29721a11_12 | | 393.2972 | 11.12 | | RP | POS | | | 1618043 | 1101982 | 1629203 | 1081979 | 9.40E-01 | 0.01 | 0.46 |
| RP_POS_623_33075a14_344 | | 623.3308 | 14.34 | | RP | POS | | | 3343217 | 1909969 | 3367240 | 2618599 | 9.40E-01 | 0.01 | 1.19 |
| RP_POS_725_10522a11_34 | | 725.1052 | 11.34 | | RP | POS | | | 287263 | 225079 | 289759 | 258973 | 9.40E-01 | 0.01 | 0.3 |
| HILIC_POS_130_08629a3_939 | | 130.0863 | 3.94 | | HILIC | POS | 84.0817(1009, 130.1589(60) | | 9026742 | 1.7E+07 | 8867669 | 1.4E+07 | 9.40E-01 | -0.01 | 0.76 |
| RP_NEG_409_22369a6_672 | | 409.2237 | 6.67 | | RP | NEG | | | 207088 | 78817.5 | 206305 | 74314.6 | 9.40E-01 | -0.01 | 0.63 |
| RP_POS_331_16849a0_67 | | 331.1685 | 0.67 | | RP | POS | | | 3319711 | 438288 | 3315641 | 359145 | 9.40E-01 | -0.01 | 0.47 |
| RP_NEG_421_22742a10_639 | | 421.2274 | 10.64 | | RP | NEG | | | 256227 | 412593 | 260587 | 441093 | 9.41E-01 | 0.01 | 0.27 |
| RP_POS_214_91777a15_365 | | 214.9178 | 15.37 | | RP | POS | | | 2E+07 | 2692885 | 2E+07 | 3141721 | 9.41E-01 | 0.01 | 0.38 |
| HILIC_POS_190_14352a0_765 | | 190.1435 | 0.77 | | HILIC | POS | | | 1588708 | 491083 | 1583626 | 510069 | 9.41E-01 | -0.01 | 0.79 |
| RP_NEG_765_16559a9_785 | | 765.1656 | 9.79 | | RP | NEG | | | 6799316 | 276766 | 6796708 | 241576 | 9.41E-01 | -0.01 | 0.58 |
| RP_POS_247_22693a9_741 | | 247.2269 | 9.74 | | RP | POS | | | 208280 | 134007 | 206915 | 135197 | 9.41E-01 | -0.01 | 0.39 |
| RP_POS_282_62411a10_278 | | 282.6241 | 10.28 | | RP | POS | | | 171219 | 88982.9 | 170268 | 96907.6 | 9.41E-01 | -0.01 | 0.68 |
| RP_POS_313_27356a10_652 | | 313.2736 | 10.65 | | RP | POS | | | 1748310 | 596440 | 1742408 | 572830 | 9.41E-01 | -0.01 | 0.41 |
| RP_POS_401_37775a13_288 | | 401.3778 | 13.29 | | RP | POS | | | 668395 | 338021 | 665029 | 328436 | 9.41E-01 | -0.01 | 0.8 |
| RP_POS_741_52881a12_799 | | 741.5288 | 12.80 | | RP | POS | | | 3137790 | 2334677 | 3115725 | 2018075 | 9.41E-01 | -0.01 | 1.17 |
| HILIC_POS_104_07103a5_349 | 2-Aminoisobutyric acid | 104.0710 | 5.35 | | HILIC | POS | 58.0663(100), 104.1079(20) | 2 | 1595069 | 2193305 | 1617443 | 2250368 | 9.41E-01 | 0.01 | 0.6 |
| HILIC_POS_130_15903a4_039 | | 130.1590 | 4.04 | | HILIC | POS | | | 1841028 | 1021678 | 1851023 | 996102 | 9.42E-01 | 0.01 | 0.4 |
| RP_NEG_190_8912a15_403 | | 190.8912 | 15.40 | | RP | NEG | | | 2324052 | 833524 | 2332109 | 789885 | 9.42E-01 | 0.01 | 0.52 |
| RP_NEG_333_95023a2_885 | | 333.9502 | 2.89 | | RP | NEG | | | 122215 | 32050.1 | 122535 | 32072.8 | 9.42E-01 | 0.01 | 0.49 |
| RP_POS_318_30051a10_252 | | 318.3005 | 10.25 | | RP | POS | | | 291843 | 86989.5 | 292687 | 83403.7 | 9.42E-01 | 0.01 | 0.37 |
| HILIC_NEG_115_92079a1_301 | | 115.9208 | 1.30 | | HILIC | NEG | | | 1.6E+07 | 2696140 | 1.6E+07 | 2380801 | 9.42E-01 | -0.01 | 0.81 |
| RP_NEG_226_80962a15_431 | | 226.8096 | 15.43 | | RP | NEG | | | 489767 | 149808 | 488282 | 147553 | 9.42E-01 | -0.01 | 0.38 |
| RP_POS_285_13071a5_197 | | 285.1307 | 5.20 | | RP | POS | | | 153816 | 80438.8 | 153063 | 71718.2 | 9.42E-01 | -0.01 | 0.89 |
| RP_POS_756_55121a13_661 | | 756.5512 | 13.66 | | RP | POS | | | 4821392 | 5122967 | 4771239 | 5009889 | 9.42E-01 | -0.01 | 0.58 |
| HILIC_NEG_102_95685a4_362 | | 102.9569 | 4.36 | | HILIC | NEG | | | 1113837 | 396975 | 1117796 | 406193 | 9.43E-01 | 0.01 | 0.57 |
| HILIC_NEG_355_00732a6_686 | | 355.0073 | 6.69 | | HILIC | NEG | | | 356836 | 34784.3 | 357185 | 36094.6 | 9.43E-01 | 0.01 | 0.39 |
| HILIC_NEG_690_88116a4_353 | | 690.8812 | 4.35 | | HILIC | NEG | | | 204973 | 63257.2 | 205604 | 65393.4 | 9.43E-01 | 0.01 | 0.49 |
| RP_POS_223_09364a4_894 | | 223.0936 | 4.89 | | RP | POS | | | 139874 | 114491 | 140997 | 113676 | 9.43E-01 | 0.01 | 0.99 |
| RP_POS_444_76343a13_789 | | 444.7634 | 13.79 | | RP | POS | | | 304148 | 423461 | 307918 | 345242 | 9.43E-01 | 0.01 | 0.59 |
| HILIC_NEG_299_72995a3_322 | | 299.7300 | 3.32 | | HILIC | NEG | | | 1003593 | 70326.6 | 1002836 | 82118 | 9.43E-01 | -0.01 | 0.43 |
| RP_NEG_322_84738a15_379 | | 322.8474 | 15.38 | | RP | NEG | | | 854996 | 331736 | 851848 | 317116 | 9.43E-01 | -0.01 | 0.45 |
| RP_NEG_657_33282a9_825 | | 657.3328 | 9.83 | | RP | NEG | | | 151376 | 484016 | 147088 | 403444 | 9.43E-01 | -0.01 | 0.58 |
| RP_POS_632_45636a8_84 | | 632.4564 | 8.84 | | RP | POS | | | 433335 | 243704 | 430345 | 347612 | 9.43E-01 | -0.01 | 0.95 |
| RP_POS_230_24744a8_125 | | 230.2474 | 8.13 | | RP | POS | | | 359355 | 588315 | 365533 | 678414 | 9.44E-01 | 0.01 | 0.23 |
| RP_POS_721_7052a11_52 | | 721.7052 | 11.52 | | RP | POS | | | 235708 | 127388 | 237125 | 163578 | 9.44E-01 | 0.01 | 0.51 |
| HILIC_NEG_195_81079a0_896 | | 195.8108 | 0.90 | | HILIC | NEG | | | 171828 | 30706.9 | 171525 | 32077.5 | 9.44E-01 | -0.01 | 0.9 |
| HILIC_NEG_162_98271a1_353 | | 162.9827 | 1.35 | | HILIC | NEG | 118.9923(100), 162.999(30) | | 1462662 | 378991 | 1466964 | 513926 | 9.45E-01 | 0.01 | 0.47 |
| HILIC_NEG_256_96869a4_863 | | 256.9687 | 4.86 | | HILIC | NEG | | | 180493 | 48662.9 | 180891 | 35840.5 | 9.45E-01 | 0.01 | 0.44 |
| RP_NEG_215_86836a15_576 | | 215.8684 | 15.58 | | RP | NEG | | | 961304 | 576779 | 966208 | 459432 | 9.45E-01 | 0.01 | 0.44 |
| RP_NEG_250_93463a15_662 | | 250.9346 | 15.66 | | RP | NEG | | | 745831 | 351047 | 749116 | 347817 | 9.45E-01 | 0.01 | 0.33 |
| RP_POS_253_14091a4_647 | | 253.1409 | 4.65 | | RP | POS | | | 387484 | 155156 | 388840 | 133130 | 9.45E-01 | 0.01 | 0.61 |
| RP_POS_680_47888a13_696 | | 680.4789 | 13.70 | | RP | POS | | | 1.9E+07 | 3986550 | 1.9E+07 | 4608552 | 9.45E-01 | 0.01 | 0.86 |
| RP_POS_686_68103a11_335 | | 686.6810 | 11.34 | | RP | POS | | | 144239 | 66448.4 | 144875 | 69305.2 | 9.45E-01 | 0.01 | 0.48 |
| RP_POS_694_64746a11_38 | | 694.6475 | 11.38 | | RP | POS | | | 232352 | 211435 | 234339 | 210257 | 9.45E-01 | 0.01 | 0.7 |
| HILIC_NEG_369_00507a6_234 | | 369.0051 | 6.23 | | HILIC | NEG | | | 740159 | 153746 | 738694 | 155958 | 9.45E-01 | -0.01 | 0.31 |
| RP_NEG_552_2677a13_296 | | 552.2677 | 13.30 | | RP | NEG | | | 266598 | 49936.5 | 266161 | 42841.6 | 9.45E-01 | -0.01 | 0.65 |
| RP_POS_227_02567a15_392 | | 227.0257 | 15.39 | | RP | POS | | | 260270 | 362681 | 256507 | 423723 | 9.45E-01 | -0.01 | 0.51 |
| RP_POS_343_1908a5_942 | | 343.1908 | 5.94 | | RP | POS | | | 604998 | 173344 | 603557 | 132732 | 9.45E-01 | -0.01 | 0.45 |
| RP_POS_351_18411a10_687 | | 351.1841 | 10.69 | | RP | POS | | | 554987 | 137946 | 553744 | 123976 | 9.45E-01 | -0.01 | 0.56 |
| RP_NEG_679_15094a8_901 | | 679.1509 | 8.90 | | RP | NEG | | | 715919 | 40148.8 | 716297 | 41040.6 | 9.46E-01 | 0.01 | 0.57 |
| RP_POS_1029_62573a10_486 | | 1029.6257 | 10.49 | | RP | POS | | | 611529 | 409292 | 615022 | 344564 | 9.46E-01 | 0.01 | 0.33 |
| RP_POS_205_1588a9_702 | | 205.1588 | 9.70 | | RP | POS | | | 1.3E+07 | 261514 | 1.3E+07 | 291303 | 9.46E-01 | 0.01 | 0.53 |
| RP_POS_337_01404a15_485 | | 337.0140 | 15.49 | | RP | POS | | | 332535 | 71564.3 | 333192 | 70886.3 | 9.46E-01 | 0.01 | 0.85 |
| RP_POS_476_79846a12_368 | | 476.7985 | 12.37 | | RP | POS | | | 8938085 | 2.9E+07 | 9179128 | 2.2E+07 | 9.46E-01 | 0.01 | 0.62 |
| HILIC_NEG_144_92355a6_167 | | 144.9236 | 6.17 | | HILIC | NEG | | | 2.9E+07 | 5865643 | 2.9E+07 | 7710933 | 9.46E-01 | -0.01 | 0.23 |
| RP_NEG_332_78961a15_438 | | 332.7896 | 15.44 | | RP | NEG | | | 3150600 | 830092 | 3143472 | 728496 | 9.46E-01 | -0.01 | 0.24 |
| RP_NEG_368_75204a15_429 | | 368.7520 | 15.43 | | RP | NEG | | | 709196 | 255864 | 706907 | 239908 | 9.46E-01 | -0.01 | 0.26 |
| RP_POS_419_23627a10_883 | | 419.2363 | 10.88 | | RP | POS | | | 708042 | 214354 | 706068 | 214156 | 9.46E-01 | -0.01 | 0.48 |
| RP_POS_429_79099a9_386 | | 429.7910 | 9.39 | | RP | POS | | | 432588 | 231884 | 429768 | 355503 | 9.46E-01 | -0.01 | 1.01 |
| RP_POS_516_46222a13_684 | | 516.4622 | 13.68 | | RP | POS | | | 4293771 | 1195015 | 4282252 | 1298125 | 9.46E-01 | -0.01 | 0.29 |
| RP_POS_669_07007a11_364 | | 669.0701 | 11.36 | | RP | POS | | | 136441 | 65816.7 | 135855 | 62032.2 | 9.46E-01 | -0.01 | 0.74 |
| HILIC_POS_161_12859a6_812 | L-beta-Homolysine | 161.1286 | 6.81 | | HILIC | POS | 84.0814(100) | 2 | 5647659 | 2877592 | 5618926 | 3240358 | 9.46E-01 | -0.01 | 0.28 |
| HILIC_NEG_291_00787a1_359 | | 291.0079 | 1.36 | | HILIC | NEG | | | 954009 | 267449 | 956433 | 268030 | 9.47E-01 | 0.01 | 0.24 |
| RP_NEG_146_99742a15_365 | | 146.9974 | 15.37 | | RP | NEG | | | 1027985 | 414698 | 1031933 | 456041 | 9.47E-01 | 0.01 | 0.28 |
| RP_POS_169_08597a4_082 | | 169.0860 | 4.08 | | RP | POS | | | 799930 | 310871 | 802938 | 352105 | 9.47E-01 | 0.01 | 0.66 |
| RP_POS_179_10648a6_831 | | 179.1065 | 6.83 | | RP | POS | | | 3460855 | 331851 | 3463711 | 297486 | 9.47E-01 | 0.01 | 0.17 |
| RP_POS_205_09184a15_334 | | 205.0918 | 15.33 | | RP | POS | | | 609802 | 185827 | 611475 | 181428 | 9.47E-01 | 0.01 | 0.51 |
| RP_POS_369_18231a9_006 | | 369.1823 | 9.01 | | RP | POS | | | 1415853 | 228651 | 1417907 | 227049 | 9.47E-01 | 0.01 | 0.34 |
| RP_POS_478_33157a11_097 | | 478.3316 | 11.10 | | RP | POS | | | 1019606 | 1034727 | 1029064 | 1029050 | 9.47E-01 | 0.01 | 0.88 |
| RP_POS_599_43561a9_435 | | 599.4356 | 9.44 | | RP | POS | | | 577300 | 344610 | 580719 | 398582 | 9.47E-01 | 0.01 | 0.92 |
| RP_NEG_313_16376a10_195 | | 313.1638 | 10.20 | | RP | NEG | | | 177181 | 103133 | 176273 | 96529.9 | 9.47E-01 | -0.01 | 0.42 |
| RP_NEG_356_97971a8_551 | | 356.9797 | 8.55 | | RP | NEG | | | 2092525 | 737540 | 2085811 | 736653 | 9.47E-01 | -0.01 | 0.55 |
| RP_POS_221_09189a0_692 | | 221.0919 | 0.69 | | RP | POS | | | 533133 | 382050 | 530339 | 230059 | 9.47E-01 | -0.01 | 0.5 |
| RP_POS_429_16769a11_598 | | 429.1677 | 11.60 | | RP | POS | | | 443504 | 88770.8 | 442733 | 81264.7 | 9.47E-01 | -0.01 | 0.82 |
| RP_POS_530_35504a10 | | 530.3550 | 10.00 | | RP | POS | | | 285989 | 182458 | 284224 | 201561 | 9.47E-01 | -0.01 | 0.79 |
| HILIC_POS_116_07111a4_858 | Proline | 116.0711 | 4.86 | | HILIC | POS | 70.0662100, 116.0715(45) | 1 | 5.1E+08 | 1.6E+08 | 5.1E+08 | 1.4E+08 | 9.47E-01 | -0.01 | 0.68 |
| RP_NEG_236_91653a0_683 | | 236.9165 | 0.68 | | RP | NEG | | | 257431 | 46601.3 | 257836 | 44440.5 | 9.48E-01 | 0.01 | 0.71 |
| RP_POS_295_22437a10_278 | | 295.2244 | 10.28 | | RP | POS | | | 737476 | 438724 | 741451 | 454910 | 9.48E-01 | 0.01 | 0.6 |
| RP_POS_318_21857a10_543 | | 318.2186 | 10.54 | | RP | POS | | | 251110 | 153610 | 252456 | 149497 | 9.48E-01 | 0.01 | 1.02 |
| RP_POS_615_33795a14_356 | | 615.3380 | 14.36 | | RP | POS | | | 6674659 | 3406842 | 6704633 | 3276364 | 9.48E-01 | 0.01 | 0.9 |
| HILIC_POS_150_11229a3_121 | | 150.1123 | 3.12 | | HILIC | POS | 150.1126(100) | | 5059976 | 2500268 | 5037152 | 2641807 | 9.48E-01 | -0.01 | 1.16 |
| RP_NEG_235_97406a0_689 | | 235.9741 | 0.69 | | RP | NEG | | | 1242648 | 370829 | 1239561 | 317703 | 9.48E-01 | -0.01 | 0.66 |
| RP_NEG_344_97922a7_466 | | 344.9792 | 7.47 | | RP | NEG | | | 1515778 | 618437 | 1510354 | 592758 | 9.48E-01 | -0.01 | 0.28 |
| RP_NEG_356_13055a10_687 | | 356.1306 | 10.69 | | RP | NEG | | | 420532 | 47237.1 | 420136 | 41606.1 | 9.48E-01 | -0.01 | 0.48 |
| RP_NEG_645_69702a6_086 | | 645.6970 | 6.09 | | RP | NEG | | | 275546 | 89103.1 | 274666 | 105777 | 9.48E-01 | -0.01 | 0.53 |
| RP_POS_1186_29053a9_813 | | 1186.2905 | 9.81 | | RP | POS | | | 1536319 | 256488 | 1534042 | 257724 | 9.48E-01 | -0.01 | 0.42 |
| RP_POS_872_36926a10_569 | | 872.3693 | 10.57 | | RP | POS | | | 465357 | 163887 | 463320 | 272120 | 9.48E-01 | -0.01 | 0.64 |
| HILIC_POS_273_10992a3_023 | | 273.1099 | 3.02 | | HILIC | POS | 273.1868(100) | | 189120 | 138986 | 190295 | 131396 | 9.49E-01 | 0.01 | 0.34 |
| HILIC_NEG_163_84039a0_94 | | 163.8404 | 0.94 | | HILIC | NEG | | | 617406 | 83917.7 | 618134 | 83820.4 | 9.49E-01 | 0.01 | 0.28 |
| HILIC_NEG_302_9711a3_93 | | 302.9711 | 3.93 | | HILIC | NEG | | | 193886 | 143610 | 195222 | 162709 | 9.49E-01 | 0.01 | 0.66 |
| HILIC_POS_330_26422a0_641 | | 330.2642 | 0.64 | | HILIC | POS | | | 659682 | 390123 | 663129 | 395089 | 9.49E-01 | 0.01 | 0.4 |
| HILIC_POS_444_0567a3_146 | | 444.0567 | 3.15 | | HILIC | POS | | | 956979 | 137367 | 958059 | 111173 | 9.49E-01 | 0.01 | 0.35 |
| RP_POS_198_18517a0_633 | | 198.1852 | 0.63 | | RP | POS | | | 2.1E+07 | 4044513 | 2.1E+07 | 4580722 | 9.49E-01 | 0.01 | 0.64 |
| RP_POS_213_12341a0_679 | | 213.1234 | 0.68 | | RP | POS | | | 364718 | 323605 | 367369 | 285224 | 9.49E-01 | 0.01 | 0.98 |
| RP_POS_376_90103a15_374 | | 376.9010 | 15.37 | | RP | POS | | | 1571700 | 951635 | 1579928 | 944643 | 9.49E-01 | 0.01 | 0.38 |
| RP_POS_397_31055a11_596 | | 397.3106 | 11.60 | | RP | POS | | | 2498601 | 505662 | 2503101 | 526387 | 9.49E-01 | 0.01 | 0.64 |
| RP_POS_405_73984a12_812 | | 405.7398 | 12.81 | | RP | POS | | | 398772 | 430942 | 402558 | 428825 | 9.49E-01 | 0.01 | 0.6 |
| RP_POS_791_53821a13_81 | | 791.5382 | 13.81 | | RP | POS | | | 999628 | 1267909 | 1009456 | 981279 | 9.49E-01 | 0.01 | 0.63 |
| RP_POS_949_62555a10_48 | | 949.6256 | 10.48 | | RP | POS | | | 5060696 | 1503185 | 5074630 | 1691062 | 9.49E-01 | 0.01 | 0.59 |
| RP_POS_453_32361a10_067 | | 453.3236 | 10.07 | | RP | POS | | | 814446 | 408376 | 810366 | 516936 | 9.49E-01 | -0.01 | 1.11 |
| RP_POS_758_00946a10_348 | | 758.0095 | 10.35 | | RP | POS | | | 475869 | 341175 | 472748 | 371313 | 9.49E-01 | -0.01 | 0.67 |
| HILIC_POS_344_6611a0_81 | | 344.6611 | 0.81 | | HILIC | POS | | | 130301 | 57525.8 | 130826 | 63529.9 | 9.50E-01 | 0.01 | 0.53 |
| RP_NEG_694_26398a10_505 | | 694.2640 | 10.51 | | RP | NEG | | | 551145 | 65315.7 | 551672 | 57587.1 | 9.50E-01 | 0.01 | 0.4 |

**Supplemantary table 1**: The non-targeted metabolomics results ordered by the VIP value (p value < 0.0003 are shaded dark green, p value < 0.05 are shaded light green, d values below 0 are blue and above 0 are red)

| | | | | | | | | | Cases | | Controls | | Welch test | Cohen's | |
|---|---|---|---|---|---|---|---|---|---|---|---|---|---|---|---|
| Feature_ID | Metabolite_name | Average_Mz | Average_RT_min_ | HMDB ID | RP_HILIC | POS_NEG | Cureted_MSMS | ID_level | Mean | SD | Mean2 | SD3 | p | d | VIP |
| RP_POS_569_43243a12_666 | | 569.4324 | 12.67 | | RP | POS | | | 307678 | 195803 | 309200 | 161137 | 9.50E-01 | 0.01 | 0.84 |
| RP_POS_592_4173a11_382 | | 592.4173 | 11.38 | | RP | POS | | | 131458 | 65592.3 | 132029 | 67598.4 | 9.50E-01 | 0.01 | 0.57 |
| RP_POS_864_52301a13_466 | | 864.5230 | 13.47 | | RP | POS | | | 1368159 | 794515 | 1374816 | 749089 | 9.50E-01 | 0.01 | 0.37 |
| HILIC_NEG_153_86853a3_214 | | 153.8685 | 3.21 | | HILIC | NEG | | | 2.9E+07 | 2620835 | 2.9E+07 | 2212518 | 9.50E-01 | -0.01 | 0.44 |
| HILIC_POS_274_18729a6_679 | | 274.1873 | 6.68 | | HILIC | POS | | | 1311787 | 532326 | 1307003 | 582087 | 9.50E-01 | -0.01 | 0.75 |
| RP_NEG_237_76492a15_495 | | 237.7649 | 15.50 | | RP | NEG | | | 676814 | 33645.2 | 676521 | 34800.1 | 9.50E-01 | -0.01 | 0.21 |
| RP_POS_179_14297a8_211 | | 179.1430 | 8.21 | | RP | POS | | | 3134646 | 155098 | 3133362 | 147512 | 9.50E-01 | -0.01 | 0.58 |
| RP_POS_595_70612a11_539 | | 595.7061 | 11.54 | | RP | POS | | | 164410 | 97681.8 | 163625 | 87257.8 | 9.50E-01 | -0.01 | 0.63 |
| HILIC_NEG_136_89163a4_059 | | 136.8916 | 4.06 | | HILIC | NEG | | | 6455391 | 2217869 | 6475357 | 2491604 | 9.51E-01 | 0.01 | 0.49 |
| HILIC_POS_375_32184a3_092 | | 375.3218 | 3.09 | | HILIC | POS | | | 920349 | 789636 | 926924 | 788810 | 9.51E-01 | 0.01 | 0.52 |
| RP_NEG_327_12723a10_685 | | 327.1272 | 10.69 | | RP | NEG | | | 1125582 | 375027 | 1128431 | 300526 | 9.51E-01 | 0.01 | 0.64 |
| RP_NEG_332_83408a15_396 | | 332.8341 | 15.40 | | RP | NEG | | | 6175934 | 1033985 | 6184271 | 943133 | 9.51E-01 | 0.01 | 0.78 |
| RP_POS_646_45551a9_93 | | 646.4555 | 9.93 | | RP | POS | | | 4348440 | 1961187 | 4366722 | 2352542 | 9.51E-01 | 0.01 | 0.69 |
| HILIC_POS_210_05032a3_901 | | 210.0503 | 3.90 | | HILIC | POS | | | 2394262 | 578996 | 2389043 | 665939 | 9.51E-01 | -0.01 | 0.6 |
| HILIC_POS_84_04513a6_137 | | 84.0451 | 6.14 | | HILIC | POS | | | 2049933 | 338306 | 2047232 | 307595 | 9.51E-01 | -0.01 | 0.8 |
| RP_POS_366_23062a15_358 | | 366.2306 | 15.36 | | RP | POS | | | 2.1E+08 | 7E+08 | 2E+08 | 5.3E+08 | 9.51E-01 | -0.01 | 0.48 |
| HILIC_POS_157_07629a4_059 | 2,2-Dipyridyl | 157.0763 | 4.06 | | HILIC | POS | 157.0767(100) | 2 | 601903 | 272766 | 604115 | 261670 | 9.52E-01 | 0.01 | 0.97 |
| RP_POS_253_21291a10_425 | | 253.2129 | 10.43 | | RP | POS | | | 971582 | 354651 | 974487 | 348549 | 9.52E-01 | 0.01 | 0.8 |
| RP_POS_275_0083a0_687 | | 275.0083 | 0.69 | | RP | POS | | | 361745 | 110901 | 362571 | 88468.9 | 9.52E-01 | 0.01 | 0.48 |
| RP_POS_678_07367a11_349 | | 678.0737 | 11.35 | | RP | POS | | | 165505 | 104321 | 166458 | 124143 | 9.52E-01 | 0.01 | 0.28 |
| RP_POS_689_51093a14_6 | | 689.5109 | 14.60 | | RP | POS | | | 948331 | 1450558 | 964009 | 2188800 | 9.52E-01 | 0.01 | 0.45 |
| HILIC_POS_388_22943a6_899 | | 388.2294 | 6.90 | | HILIC | POS | | | 240662 | 113517 | 239712 | 115214 | 9.52E-01 | -0.01 | 0.57 |
| RP_POS_233_0769a2_509 | | 233.0769 | 2.51 | | RP | POS | | | 1826295 | 755466 | 1819512 | 879537 | 9.52E-01 | -0.01 | 0.6 |
| RP_POS_255_23166a10_107 | | 255.2317 | 10.11 | | RP | POS | | | 2062600 | 251819 | 2060496 | 260812 | 9.52E-01 | -0.01 | 0.71 |
| RP_POS_367_10974a9_456 | | 367.1097 | 9.46 | | RP | POS | | | 1676091 | 3909877 | 1646964 | 3117572 | 9.52E-01 | -0.01 | 0.63 |
| RP_POS_422_18298a7_533 | | 422.1830 | 7.53 | | RP | POS | | | 442389 | 162629 | 441154 | 137214 | 9.52E-01 | -0.01 | 0.63 |
| RP_POS_610_18256a12_753 | | 610.1826 | 12.75 | | RP | POS | | | 1201079 | 398591 | 1197764 | 407169 | 9.52E-01 | -0.01 | 0.92 |
| RP_POS_319_22485a9_965 | Andrograpanin | 319.2249 | 9.97 | | RP | POS | 319.2261(100) | 2 | 1124195 | 643029 | 1118778 | 666801 | 9.52E-01 | -0.01 | 0.48 |
| HILIC_POS_350_01505a7_403 | | 350.0151 | 7.40 | | HILIC | POS | | | 429462 | 28752.3 | 429709 | 31766.8 | 9.53E-01 | 0.01 | 0.52 |
| RP_NEG_429_24985a9_627 | | 429.2499 | 9.63 | | RP | NEG | | | 318544 | 139683 | 319721 | 153752 | 9.53E-01 | 0.01 | 0.98 |
| RP_NEG_459_29135a11_094 | | 459.2914 | 11.09 | | RP | NEG | | | 2856211 | 2942228 | 2879480 | 2823370 | 9.53E-01 | 0.01 | 0.9 |
| RP_POS_255_23131a9_645 | | 255.2313 | 9.65 | | RP | POS | | | 3456946 | 393866 | 3460206 | 407996 | 9.53E-01 | 0.01 | 0.77 |
| RP_POS_587_46417a13_704 | | 587.4642 | 13.70 | | RP | POS | | | 637919 | 683459 | 643217 | 642879 | 9.53E-01 | 0.01 | 0.37 |
| HILIC_NEG_400_88898a1_342 | | 400.8890 | 1.34 | | HILIC | NEG | | | 158487 | 38334.3 | 158174 | 39971.6 | 9.53E-01 | -0.01 | 0.56 |
| RP_NEG_144_92354a10_43 | | 144.9235 | 10.43 | | RP | NEG | | | 809336 | 314019 | 806909 | 292963 | 9.53E-01 | -0.01 | 0.34 |
| RP_NEG_162_83887a11_143 | | 162.8389 | 11.14 | | RP | NEG | | | 97764.4 | 22966.7 | 97531 | 33411.7 | 9.53E-01 | -0.01 | 0.55 |
| RP_POS_200_2009a9_448 | | 200.2009 | 9.45 | | RP | POS | | | 2.3E+07 | 2541190 | 2.3E+07 | 2972777 | 9.53E-01 | -0.01 | 0.6 |
| RP_POS_319_01117a4_824 | | 319.0112 | 4.82 | | RP | POS | | | 491885 | 391957 | 488572 | 429506 | 9.53E-01 | -0.01 | 0.74 |
| HILIC_NEG_192_90021a3_151 | | 192.9002 | 3.15 | | HILIC | NEG | | | 199084 | 59140.8 | 198613 | 60343.8 | 9.54E-01 | -0.01 | 0.34 |
| HILIC_POS_650_43799a0_652 | | 650.4380 | 0.65 | | HILIC | POS | | | 66656.5 | 52478.1 | 66261.8 | 48175.5 | 9.54E-01 | -0.01 | 0.56 |
| RP_NEG_136_89162a11_59 | | 136.8916 | 11.59 | | RP | NEG | | | 391667 | 141289 | 390517 | 149469 | 9.54E-01 | -0.01 | 0.34 |
| RP_NEG_296_9985a6_74 | | 296.9985 | 6.74 | | RP | NEG | | | 4319959 | 531475 | 4316231 | 412861 | 9.54E-01 | -0.01 | 0.56 |
| RP_POS_169_09718a2_232 | | 169.0972 | 2.23 | | RP | POS | | | 402201 | 659215 | 398452 | 210656 | 9.54E-01 | -0.01 | 0.28 |
| RP_POS_486_34814a10_067 | | 486.3481 | 10.07 | | RP | POS | | | 219475 | 115770 | 218528 | 123124 | 9.54E-01 | -0.01 | 0.46 |
| RP_POS_499_43561a13_682 | | 499.4356 | 13.68 | | RP | POS | | | 2744303 | 806601 | 2737586 | 874375 | 9.54E-01 | -0.01 | 0.38 |
| HILIC_NEG_100_93344a1_366 | | 100.9334 | 1.37 | | HILIC | NEG | | | 9266769 | 3215380 | 9292559 | 3420260 | 9.55E-01 | 0.01 | 0.58 |
| HILIC_NEG_234_81978a6_1 | | 234.8198 | 6.10 | | HILIC | NEG | | | 856635 | 426230 | 859959 | 437108 | 9.55E-01 | 0.01 | 0.3 |
| RP_POS_406_20279a7_241 | | 406.2028 | 7.24 | | RP | POS | | | 1.3E+07 | 960826 | 1.3E+07 | 965677 | 9.55E-01 | 0.01 | 0.66 |
| RP_POS_836_61548a14_117 | | 836.6155 | 14.12 | | RP | POS | | | 2.3E+07 | 9207329 | 2.3E+07 | 8887507 | 9.55E-01 | -0.01 | 0.67 |
| HILIC_POS_375_32205a10_567 | | 375.3221 | 10.57 | | HILIC | POS | | | 668741 | 2077714 | 683089 | 1719262 | 9.56E-01 | 0.01 | 0.3 |
| HILIC_NEG_110_89923a3_102 | | 110.8992 | 3.10 | | HILIC | NEG | | | 786900 | 111354 | 787751 | 114561 | 9.56E-01 | 0.01 | 0.42 |
| HILIC_NEG_115_92078a4_408 | | 115.9208 | 4.41 | | HILIC | NEG | | | 2.9E+07 | 9653844 | 2.9E+07 | 9337955 | 9.56E-01 | 0.01 | 0.29 |
| HILIC_POS_351_57819a1_009 | | 351.5782 | 1.01 | | HILIC | POS | | | 90922 | 104017 | 91717 | 105383 | 9.56E-01 | 0.01 | 0.37 |
| RP_POS_200_20076a9_268 | | 200.2008 | 9.27 | | RP | POS | | | 1.7E+07 | 427375 | 1.7E+07 | 590962 | 9.56E-01 | 0.01 | 0.53 |
| RP_POS_584_474a10_867 | | 584.4740 | 10.87 | | RP | POS | | | 5331193 | 457056 | 5334533 | 426191 | 9.56E-01 | 0.01 | 0.69 |
| RP_POS_624_2973a10_925 | | 624.2973 | 10.93 | | RP | POS | | | 694454 | 290809 | 696821 | 327030 | 9.56E-01 | 0.01 | 0.87 |
| RP_POS_675_26074a10_567 | | 675.2607 | 10.57 | | RP | POS | | | 397331 | 161776 | 398999 | 261007 | 9.56E-01 | 0.01 | 0.88 |
| RP_POS_813_52319a13_112 | | 813.5232 | 13.11 | | RP | POS | | | 568074 | 477388 | 572736 | 711574 | 9.56E-01 | 0.01 | 0.64 |
| RP_POS_464_33258a9_175 | | 464.3326 | 9.18 | | RP | POS | | | 367224 | 183028 | 365729 | 212109 | 9.56E-01 | -0.01 | 0.36 |
| RP_POS_193_15906a6_967 | | 193.1591 | 6.97 | | RP | POS | | | 3649514 | 135271 | 3650500 | 133113 | 9.57E-01 | 0.01 | 0.76 |
| RP_POS_319_15765a10_688 | | 319.1577 | 10.69 | | RP | POS | | | 433899 | 114579 | 434707 | 104263 | 9.57E-01 | 0.01 | 0.61 |
| RP_POS_542_37213a14_358 | | 542.3721 | 14.36 | | RP | POS | | | 3754608 | 3724756 | 3782496 | 3752045 | 9.57E-01 | 0.01 | 0.82 |
| RP_NEG_318_81046a15_391 | | 318.8105 | 15.39 | | RP | NEG | | | 2518175 | 883235 | 2511194 | 1009980 | 9.57E-01 | -0.01 | 0.29 |
| RP_NEG_509_11652a9_128 | | 509.1165 | 9.13 | | RP | NEG | | | 640031 | 64771.9 | 639564 | 63039.1 | 9.57E-01 | -0.01 | 0.84 |
| RP_POS_373_22537a11_14 | | 373.2254 | 11.14 | | RP | POS | | | 512087 | 165756 | 510959 | 139181 | 9.57E-01 | -0.01 | 0.77 |
| RP_POS_447_31912a9_759 | | 447.3191 | 9.76 | | RP | POS | | | 108981 | 104188 | 108180 | 110470 | 9.57E-01 | -0.01 | 0.57 |
| RP_POS_301_17676a9_113 | Adrenosterone | 301.1768 | 9.11 | | RP | POS | 301.1798(100) | 2 | 7182205 | 321444 | 7184345 | 269939 | 9.58E-01 | 0.01 | 0.89 |
| HILIC_NEG_116_92848a7_74 | | 116.9285 | 7.74 | | HILIC | NEG | | | 5370083 | 2009646 | 5383308 | 1686454 | 9.58E-01 | 0.01 | 0.56 |
| RP_NEG_239_90286a15_569 | | 239.9029 | 15.57 | | RP | NEG | | | 1254218 | 390203 | 1256907 | 355515 | 9.58E-01 | 0.01 | 0.4 |
| RP_POS_190_9637a0_672 | | 190.9637 | 0.67 | | RP | POS | | | 963904 | 162382 | 964938 | 126403 | 9.58E-01 | 0.01 | 0.66 |
| RP_POS_806_56628a13_467 | | 806.5663 | 13.47 | | RP | POS | | | 1.9E+08 | 6.7E+07 | 1.9E+08 | 7.1E+07 | 9.58E-01 | 0.01 | 0.76 |
| HILIC_NEG_197_80804a0_896 | | 197.8080 | 0.90 | | HILIC | NEG | | | 221159 | 39044.1 | 220861 | 42957 | 9.58E-01 | -0.01 | 0.88 |
| HILIC_POS_106_08675a5_785 | | 106.0868 | 5.79 | | HILIC | POS | | | 1.6E+07 | 6658480 | 1.6E+07 | 6482384 | 9.58E-01 | -0.01 | 0.51 |
| RP_NEG_339_21606a9_645 | | 339.2161 | 9.65 | | RP | NEG | | | 265890 | 37022.1 | 265632 | 35447.6 | 9.58E-01 | -0.01 | 0.78 |
| RP_NEG_537_14893a9_381 | | 537.1489 | 9.38 | | RP | NEG | | | 363470 | 27356.6 | 363269 | 28122 | 9.58E-01 | -0.01 | 0.56 |
| HILIC_POS_212_14902a0_61 | Terbutylazine-2-hydroxy | 212.1490 | 0.61 | | HILIC | POS | 212.1452(100) | 2 | 1137560 | 141849 | 1136582 | 131385 | 9.58E-01 | -0.01 | 0.41 |
| HILIC_POS_132_98734a1_754 | | 132.9873 | 1.75 | | HILIC | POS | | | 489762 | 323633 | 492085 | 342814 | 9.59E-01 | 0.01 | 0.52 |
| HILIC_POS_346_00925a6_311 | | 346.0093 | 6.31 | | HILIC | POS | | | 2.1E+07 | 6792268 | 2.1E+07 | 6924813 | 9.59E-01 | 0.01 | 0.4 |
| HILIC_POS_450_34204a3_293 | | 450.3420 | 3.29 | | HILIC | POS | | | 172110 | 97947.8 | 171463 | 87901.4 | 9.59E-01 | -0.01 | 0.5 |
| RP_NEG_398_86459a15_364 | | 398.8646 | 15.36 | | RP | NEG | | | 1224060 | 171502 | 1222817 | 181782 | 9.59E-01 | -0.01 | 0.38 |
| RP_POS_516_68311a10_043 | | 516.6831 | 10.04 | | RP | POS | | | 594883 | 288013 | 592570 | 358070 | 9.59E-01 | -0.01 | 0.56 |
| RP_POS_250_06464a3_01 | Sulfapyridine | 250.0646 | 3.01 | | RP | POS | 156.012(100), 250.0655(70) | 2 | 2.3E+08 | 6E+07 | 2.3E+08 | 5.6E+07 | 9.59E-01 | -0.01 | 0.55 |
| RP_POS_282_27896a13_131 | | 282.2790 | 13.13 | | RP | POS | 282.2777(100), 83.0864(70) | | 1609506 | 882046 | 1616344 | 1106031 | 9.60E-01 | 0.01 | 0.89 |
| RP_NEG_270_01044a0_666 | | 270.0104 | 0.67 | | RP | NEG | | | 144339 | 29768.6 | 144524 | 24105.3 | 9.60E-01 | 0.01 | 0.26 |
| RP_NEG_313_00018a1_406 | | 313.0002 | 1.41 | | RP | NEG | | | 4584163 | 1122585 | 4592230 | 1218466 | 9.60E-01 | 0.01 | 0.37 |
| HILIC_NEG_231_93204a3_206 | | 231.9320 | 3.21 | | HILIC | NEG | | | 499564 | 195758 | 498107 | 225375 | 9.60E-01 | -0.01 | 0.59 |
| HILIC_NEG_294_98306a0_591 | | 294.9831 | 0.59 | | HILIC | NEG | | | 599899 | 148118 | 598866 | 151461 | 9.60E-01 | -0.01 | 0.36 |
| RP_POS_614_48358a10_7 | | 614.4836 | 10.70 | | RP | POS | | | 7178041 | 680096 | 7173160 | 739831 | 9.60E-01 | -0.01 | 0.76 |
| RP_POS_726_55859a12_8 | | 726.5586 | 12.80 | | RP | POS | | | 2.5E+07 | 1.6E+07 | 2.5E+07 | 1.4E+07 | 9.60E-01 | -0.01 | 1.07 |
| HILIC_POS_118_08652a6_129 | | 118.0865 | 6.13 | | HILIC | POS | 140.069(100), 74.0974(40) | | 1.1E+07 | 4036122 | 1.1E+07 | 4267206 | 9.61E-01 | 0.01 | 0.26 |
| HILIC_NEG_328_81033a2_933 | | 328.8103 | 2.93 | | HILIC | NEG | | | 249225 | 42995.2 | 249505 | 40959.4 | 9.61E-01 | 0.01 | 0.52 |
| HILIC_POS_243_18295a1_599 | | 243.1830 | 1.60 | | HILIC | POS | | | 1131038 | 1163401 | 1139276 | 1281026 | 9.61E-01 | 0.01 | 0.4 |
| RP_NEG_371_19058a8_179 | | 371.1906 | 8.18 | | RP | NEG | | | 261586 | 131491 | 262376 | 106461 | 9.61E-01 | 0.01 | 0.29 |
| RP_POS_191_0858a9_463 | | 191.0858 | 9.46 | | RP | POS | 191.0864(100) | | 5915385 | 1725245 | 5904802 | 1444159 | 9.61E-01 | -0.01 | 0.45 |
| HILIC_NEG_196_02287a4_237 | | 196.0229 | 4.24 | | HILIC | NEG | | | 210137 | 35120.6 | 209892 | 37625.8 | 9.61E-01 | -0.01 | 0.59 |
| RP_NEG_185_87532a15_499 | | 185.8753 | 15.50 | | RP | NEG | | | 205799 | 49174.6 | 205466 | 49213.4 | 9.61E-01 | -0.01 | 0.49 |
| RP_POS_504_70163a10_639 | | 504.7016 | 10.64 | | RP | POS | | | 182361 | 114973 | 181562 | 122229 | 9.61E-01 | -0.01 | 0.39 |
| RP_POS_612_32263a7_166 | | 612.3226 | 7.17 | | RP | POS | | | 1335081 | 104976 | 1334399 | 100529 | 9.61E-01 | -0.01 | 0.83 |
| HILIC_NEG_291_00784a0_935 | | 291.0078 | 0.94 | | HILIC | NEG | | | 900508 | 84426.2 | 901072 | 89194.2 | 9.62E-01 | 0.01 | 0.62 |
| HILIC_POS_207_11269a3_835 | | 207.1127 | 3.84 | | HILIC | POS | | | 196534 | 233442 | 198016 | 223361 | 9.62E-01 | 0.01 | 0.57 |
| RP_NEG_266_94595a0_68 | | 266.9460 | 0.68 | | RP | NEG | | | 7981443 | 898866 | 7987361 | 900021 | 9.62E-01 | 0.01 | 0.19 |
| RP_NEG_336_97311a6_781 | | 336.9731 | 6.78 | | RP | NEG | | | 502779 | 70105 | 503217 | 65658.8 | 9.62E-01 | 0.01 | 0.42 |
| RP_POS_288_17993a4_539 | | 288.1799 | 4.54 | | RP | POS | | | 70942 | 28327.1 | 71136.1 | 31595.9 | 9.62E-01 | 0.01 | 1 |
| RP_POS_302_12689a15_401 | | 302.1269 | 15.40 | | RP | POS | | | 239529 | 106422 | 240199 | 98889.9 | 9.62E-01 | 0.01 | 0.31 |
| RP_POS_475_29797a11_921 | | 475.2980 | 11.92 | | RP | POS | | | 167015 | 141890 | 167957 | 146258 | 9.62E-01 | 0.01 | 0.44 |
| RP_POS_295_11179a7_069 | | 295.1118 | 7.07 | | RP | POS | | | 512949 | 240296 | 511463 | 221577 | 9.62E-01 | -0.01 | 1.12 |
| RP_POS_505_86432a10_019 | | 505.8643 | 10.02 | | RP | POS | | | 3270980 | 1537494 | 3260848 | 1607761 | 9.62E-01 | -0.01 | 0.97 |
| RP_POS_620_52435a15_167 | | 620.5244 | 15.17 | | RP | POS | | | 296933 | 210345 | 295552 | 209975 | 9.62E-01 | -0.01 | 0.63 |
| HILIC_NEG_157_0118a1_34 | | 157.0118 | 1.34 | | HILIC | NEG | | | 1825601 | 342678 | 1827720 | 328591 | 9.63E-01 | 0.01 | 0.78 |
| HILIC_POS_375_32162a4_078 | | 375.3216 | 4.08 | | HILIC | POS | | | 281024 | 108015 | 281830 | 138404 | 9.63E-01 | 0.01 | 0.6 |
| RP_NEG_282_98248a7_515 | | 282.9825 | 7.52 | | RP | NEG | | | 3374089 | 1135096 | 3381538 | 1234130 | 9.63E-01 | 0.01 | 0.51 |
| RP_NEG_607_05042a8_98 | | 607.0504 | 8.98 | | RP | NEG | | | 270363 | 22750.5 | 270512 | 24419.1 | 9.63E-01 | 0.01 | 0.58 |
| RP_POS_529_42908a14_35 | | 529.4291 | 14.35 | | RP | POS | | | 997538 | 842095 | 1002588 | 742790 | 9.63E-01 | 0.01 | 0.52 |
| RP_POS_729_29126a12_616 | | 729.2913 | 12.62 | | RP | POS | | | 575531 | 1062492 | 582824 | 1211501 | 9.63E-01 | 0.01 | 0.57 |
| HILIC_POS_173_12839a3_911 | | 173.1284 | 3.91 | | HILIC | POS | | | 828533 | 3781011 | 804036 | 3995325 | 9.63E-01 | -0.01 | 0.66 |
| RP_NEG_434_57861a15_488 | | 434.5786 | 15.49 | | RP | NEG | | | 744568 | 49392.4 | 744262 | 47189.7 | 9.63E-01 | -0.01 | 0.37 |
| RP_POS_585_06299a10_27 | | 585.0630 | 10.27 | | RP | POS | | | 471731 | 214635 | 470203 | 262313 | 9.63E-01 | -0.01 | 0.5 |
| RP_NEG_399_17471a6_98 | | 399.1747 | 6.98 | | RP | NEG | | | 199757 | 72086.9 | 200259 | 87555.4 | 9.64E-01 | 0.01 | 0.38 |
| RP_POS_643_39148a15_163 | | 643.3915 | 15.16 | | RP | POS | | | 620747 | 283888 | 622426 | 257728 | 9.64E-01 | 0.01 | 0.85 |
| RP_POS_246_944a15_365 | | 246.9440 | 15.37 | | RP | POS | | | 1.8E+07 | 4730947 | 1.7E+07 | 4493820 | 9.64E-01 | -0.01 | 0.46 |
| RP_POS_385_19925a6_982 | | 385.1993 | 6.98 | | RP | POS | | | 3343676 | 1011507 | 3336903 | 1189936 | 9.64E-01 | -0.01 | 0.41 |
| RP_POS_621_76904a11_301 | | 621.7690 | 11.30 | | RP | POS | | | 185145 | 92864 | 184511 | 111556 | 9.64E-01 | -0.01 | 0.74 |
| RP_POS_656_12695a10_582 | | 656.1270 | 10.58 | | RP | POS | | | 178408 | 105204 | 177704 | 119285 | 9.64E-01 | -0.01 | 0.49 |
| RP_POS_762_97888a10_486 | | 762.9789 | 10.49 | | RP | POS | | | 1659782 | 763125 | 1655139 | 729097 | 9.64E-01 | -0.01 | 0.82 |
| HILIC_NEG_186_04614a1_344 | | 186.0461 | 1.34 | | HILIC | NEG | | | 2409961 | 842851 | 2414666 | 733251 | 9.65E-01 | 0.01 | 0.79 |
| HILIC_POS_538_388a1_127 | | 538.3880 | 1.13 | | HILIC | POS | | | 150782 | 69497.4 | 151203 | 69426.5 | 9.65E-01 | 0.01 | 0.47 |
| RP_POS_464_33295a8_884 | | 464.3330 | 8.88 | | RP | POS | | | 304241 | 209581 | 305456 | 194652 | 9.65E-01 | 0.01 | 0.46 |
| RP_POS_553_45795a14_528 | | 553.4580 | 14.53 | | RP | POS | | | 4140887 | 2338433 | 4154666 | 2319423 | 9.65E-01 | 0.01 | 0.77 |
| RP_NEG_442_16736a11_142 | | 442.1674 | 11.14 | | RP | NEG | | | 1319613 | 104791 | 1319011 | 97721.1 | 9.65E-01 | -0.01 | 0.64 |
| RP_NEG_829_1803a9_63 | | 829.1803 | 9.63 | | RP | NEG | | | 1E+07 | 523333 | 1E+07 | 473292 | 9.65E-01 | -0.01 | 0.53 |
| RP_POS_432_70825a5_624 | | 432.7083 | 5.62 | | RP | POS | | | 2027849 | 922996 | 2021696 | 1126076 | 9.65E-01 | -0.01 | 0.48 |
| RP_POS_595_42236a9_766 | | 595.4224 | 9.77 | | RP | POS | | | 4420023 | 1868332 | 4407656 | 2168919 | 9.65E-01 | -0.01 | 0.74 |
| RP_NEG_532_2998a10_491 | | 532.2998 | 10.49 | | RP | NEG | | | 2608096 | 350184 | 2605906 | 403970 | 9.66E-01 | -0.01 | 0.42 |
| RP_POS_292_17542a7_149 | | 292.1754 | 7.15 | | RP | POS | | | 360081 | 221264 | 358741 | 241122 | 9.66E-01 | -0.01 | 0.41 |
| RP_POS_345_03311a0_645 | | 345.0331 | 0.65 | | RP | POS | | | 1537140 | 740420 | 1532639 | 796838 | 9.66E-01 | -0.01 | 0.69 |
| RP_POS_770_53613a10_189 | | 770.5361 | 10.19 | | RP | POS | | | 1131435 | 572735 | 1128227 | 519756 | 9.66E-01 | -0.01 | 0.8 |
| RP_NEG_172_83023a11_591 | | 172.8302 | 11.59 | | RP | NEG | | | 240373 | 69430.5 | 239927 | 84778 | 9.67E-01 | -0.01 | 0.73 |
| HILIC_POS_284_1701a0_77 | | 284.1701 | 0.77 | | HILIC | POS | | | 1877600 | 156392 | 1878512 | 170871 | 9.68E-01 | 0.01 | 0.48 |
| RP_NEG_162_98268a15_499 | | 162.9827 | 15.50 | | RP | NEG | | | 1190832 | 316266 | 1192457 | 273836 | 9.68E-01 | 0.01 | 0.49 |
| RP_POS_339_32635a11_786 | | 339.3264 | 11.79 | | RP | POS | | | 468668 | 145792 | 469470 | 149665 | 9.68E-01 | 0.01 | 0.51 |
| RP_POS_454_29294a10_353 | | 454.2929 | 10.35 | | RP | POS | | | 5007976 | 1285067 | 5015660 | 1473798 | 9.68E-01 | 0.01 | 0.54 |
| RP_POS_415_2406a11_59 | | 415.2406 | 11.59 | | RP | POS | | | 220679 | 89795.5 | 220188 | 90343 | 9.68E-01 | -0.01 | 0.89 |
| RP_POS_610_54022a14_755 | | 610.5402 | 14.76 | | RP | POS | | | 1870692 | 1541672 | 1863195 | 1220555 | 9.68E-01 | -0.01 | 0.5 |
| HILIC_POS_351_24393a1_011 | | 351.2439 | 1.01 | | HILIC | POS | | | 147596 | 143061 | 148377 | 149316 | 9.69E-01 | 0.01 | 0.4 |
| HILIC_POS_361_02029a5_476 | | 361.0203 | 5.48 | | HILIC | POS | | | 308134 | 111828 | 308773 | 129038 | 9.69E-01 | 0.01 | 0.41 |
| RP_POS_348_10126a3_589 | | 348.1013 | 3.59 | | RP | POS | | | 2342495 | 2995427 | 2358853 | 3121727 | 9.69E-01 | 0.01 | 0.82 |
| RP_POS_459_24991a10_611 | | 459.2499 | 10.61 | | RP | POS | | | 509383 | 1047419 | 515622 | 1247615 | 9.69E-01 | 0.01 | 0.52 |
| RP_POS_714_16003a11_488 | | 714.1600 | 11.49 | | RP | POS | | | 171538 | 142167 | 172324 | 151140 | 9.69E-01 | 0.01 | 0.62 |
| RP_NEG_125_87312a14_339 | | 125.8731 | 14.34 | | RP | NEG | | | 752864 | 275904 | 754273 | 271638 | 9.70E-01 | 0.01 | 0.26 |
| RP_NEG_343_99503a15_495 | | 343.9950 | 15.50 | | RP | NEG | | | 1395623 | 105032 | 1396151 | 101536 | 9.70E-01 | 0.01 | 0.71 |
| RP_POS_133_03461a10_521 | | 133.0346 | 10.52 | | RP | POS | | | 2336228 | 1067155 | 2341817 | 1092023 | 9.70E-01 | 0.01 | 0.65 |
| RP_POS_346_04034a15_455 | | 346.0403 | 15.46 | | RP | POS | | | 5310671 | 1417884 | 5318745 | 1646712 | 9.70E-01 | 0.01 | 0.63 |
| RP_NEG_627_11456a9_099 | | 627.1146 | 9.10 | | RP | NEG | | | 3200095 | 160312 | 3199253 | 166243 | 9.70E-01 | -0.01 | 0.62 |
| RP_POS_222_99428a15_401 | | 222.9943 | 15.40 | | RP | POS | | | 319047 | 172606 | 318197 | 161613 | 9.70E-01 | -0.01 | 0.49 |
| RP_POS_263_23691a10_535 | | 263.2369 | 10.54 | | RP | POS | | | 4104703 | 1433707 | 4096987 | 1520263 | 9.70E-01 | -0.01 | 0.89 |
| RP_POS_515_36731a9_379 | | 515.3673 | 9.38 | | RP | POS | | | 1042073 | 552610 | 1039282 | 527045 | 9.70E-01 | -0.01 | 0.51 |
| RP_POS_725_51837a10_658 | | 725.5184 | 10.66 | | RP | POS | | | 622738 | 299952 | 620872 | 398912 | 9.70E-01 | -0.01 | 0.74 |
| HILIC_POS_247_09273a7_744 | | 247.0927 | 7.74 | | HILIC | POS | | | 805800 | 51822.1 | 806073 | 57860.6 | 9.71E-01 | 0 | 0.68 |
| RP_NEG_181_91875a0_7 | | 181.9188 | 0.70 | | RP | NEG | | | 675460 | 137805 | 676130 | 134021 | 9.71E-01 | 0 | 0.65 |
| RP_NEG_883_16473a9_407 | | 883.1647 | 9.41 | | RP | NEG | | | 876716 | 103536 | 877218 | 98469.8 | 9.71E-01 | 0 | 0.77 |
| RP_POS_267_15671a8_449 | | 267.1567 | 8.45 | | RP | POS | | | 1398108 | 185845 | 1397261 | 159271 | 9.71E-01 | 0 | 0.89 |
| RP_POS_517_37054a10_525 | | 517.3705 | 10.53 | | RP | POS | | | 653154 | 336682 | 651589 | 295535 | 9.71E-01 | 0 | 0.66 |

**Supplemantary table 1**: The non-targeted metabolomics results ordered by the VIP value (p value < 0.0003 are shaded dark green, p value < 0.05 are shaded light green, d values below 0 are blue and above 0 are red)

| | | | | | | | | | Cases | | Controls | | Welch test | Cohen's | |
|---|---|---|---|---|---|---|---|---|---|---|---|---|---|---|---|
| Feature_ID | Metabolite_name | Average_Mz | Average_RT_min_ | HMDB ID | RP_HILIC | POS_NEG | Cureted_MSMS | ID_level | Mean | SD | Mean2 | SD3 | p | d | VIP |
| HILIC_NEG_197_80829a10_942 | | 197.8083 | 10.94 | | HILIC | NEG | | | 2.1E+07 | 1.1E+07 | 2.1E+07 | 1.1E+07 | 9.71E-01 | -0.01 | 0.46 |
| RP_NEG_390_98102a8_845 | | 390.9810 | 8.85 | | RP | NEG | | | 1281173 | 450714 | 1278952 | 431901 | 9.71E-01 | -0.01 | 0.48 |
| HILIC_POS_175_11867a7_093 | | 175.1187 | 7.09 | | HILIC | POS | 175.1193(100), 70.0659(75), 116.071(55) | | 1579981 | 425506 | 1582059 | 444583 | 9.72E-01 | 0 | 0.49 |
| HILIC_NEG_100_93359a5_794 | | 100.9336 | 5.79 | | HILIC | NEG | | | 3E+07 | 6481825 | 3E+07 | 5251345 | 9.72E-01 | 0 | 0.37 |
| HILIC_NEG_171_88551a0_9 | | 171.8855 | 0.90 | | HILIC | NEG | | | 228036 | 25130 | 228161 | 25965.7 | 9.72E-01 | 0 | 0.5 |
| RP_NEG_331_95181a6_811 | | 331.9518 | 6.81 | | RP | NEG | | | 380293 | 75811.3 | 380652 | 74933.5 | 9.72E-01 | 0 | 0.46 |
| RP_NEG_356_98029a7_095 | | 356.9803 | 7.10 | | RP | NEG | | | 637003 | 79326.9 | 636642 | 72381.1 | 9.72E-01 | 0 | 0.5 |
| RP_POS_240_15887a7_577 | | 240.1589 | 7.58 | | RP | POS | | | 3756873 | 425292 | 3754971 | 365158 | 9.72E-01 | 0 | 0.64 |
| RP_POS_280_86136a15_402 | | 280.8614 | 15.40 | | RP | POS | | | 234849 | 127005 | 235510 | 143284 | 9.72E-01 | 0 | 0.55 |
| RP_POS_289_15433a9_355 | | 289.1543 | 9.36 | | RP | POS | | | 457850 | 118413 | 457266 | 128385 | 9.72E-01 | 0 | 0.86 |
| RP_POS_295_22635a7_905 | | 295.2264 | 7.91 | | RP | POS | | | 152825 | 63140.8 | 152426 | 98052.1 | 9.72E-01 | 0 | 0.35 |
| RP_POS_381_21552a11_594 | | 381.2155 | 11.59 | | RP | POS | | | 249178 | 141700 | 248461 | 155546 | 9.72E-01 | 0 | 0.93 |
| RP_POS_559_51965a10_546 | | 559.5197 | 10.55 | | RP | POS | | | 114694 | 105377 | 115179 | 95974.5 | 9.72E-01 | 0 | 1 |
| HILIC_NEG_358_99603a1_289 | | 358.9960 | 1.29 | | HILIC | NEG | | | 707160 | 317732 | 705728 | 296618 | 9.73E-01 | 0 | 0.48 |
| RP_NEG_316_94788a13_281 | | 316.9479 | 13.28 | | RP | NEG | | | 973212 | 289626 | 974472 | 264808 | 9.73E-01 | 0 | 0.53 |
| RP_POS_204_12308a3_374 | | 204.1231 | 3.37 | | RP | POS | | | 298147 | 181055 | 298970 | 181091 | 9.73E-01 | 0 | 0.66 |
| RP_POS_522_79688a10_488 | | 522.7969 | 10.49 | | RP | POS | | | 4753051 | 756397 | 4749812 | 655337 | 9.73E-01 | 0 | 0.85 |
| RP_POS_751_00049a10_197 | | 751.0005 | 10.20 | | RP | POS | | | 444035 | 230969 | 445333 | 313849 | 9.73E-01 | 0 | 0.46 |
| RP_POS_816_58734a13_792 | | 816.5873 | 13.79 | | RP | POS | | | 1.3E+07 | 6097196 | 1.3E+07 | 6017370 | 9.73E-01 | 0 | 0.28 |
| RP_POS_197_07875a3_633 | | 197.0788 | 3.63 | | RP | POS | | | 72667.9 | 44970.8 | 72877.2 | 48786.8 | 9.74E-01 | 0 | 0.92 |
| RP_NEG_452_27783a10_353 | LysoPE 16:0 | 452.2778 | 10.35 | | RP | NEG | 255.2328(100), 452.283(2) | 2 | 2138712 | 551759 | 2141319 | 601289 | 9.74E-01 | 0.00 | 0.55 |
| HILIC_POS_106_08682a3_301 | | 106.0868 | 3.30 | | HILIC | POS | 88.0763(100), 106.0864(90) | | 1.1E+07 | 7114190 | 1E+07 | 6360907 | 9.75E-01 | 0 | 0.85 |
| HILIC_NEG_166_83476a3_112 | | 166.8348 | 3.11 | | HILIC | NEG | | | 6901442 | 267886 | 6902502 | 236757 | 9.75E-01 | 0 | 0.76 |
| HILIC_NEG_202_92931a5_468 | | 202.9293 | 5.47 | | HILIC | NEG | | | 805762 | 91910.4 | 806156 | 92457.1 | 9.75E-01 | 0 | 0.59 |
| HILIC_POS_233_07878a2_981 | | 233.0788 | 2.98 | | HILIC | POS | | | 781399 | 305371 | 782705 | 300824 | 9.75E-01 | 0 | 0.45 |
| RP_NEG_473_39972a13_173 | | 473.3997 | 13.17 | | RP | NEG | | | 4143520 | 302030 | 4142235 | 295081 | 9.75E-01 | 0 | 1.18 |
| RP_POS_355_65237a5_649 | | 355.6524 | 5.65 | | RP | POS | | | 283620 | 174276 | 284258 | 119788 | 9.75E-01 | 0 | 0.52 |
| RP_POS_371_23975a9_635 | | 371.2398 | 9.64 | | RP | POS | | | 726767 | 129205 | 726228 | 125801 | 9.75E-01 | 0 | 0.76 |
| RP_POS_245_11488a7_801 | | 245.1149 | 7.80 | | RP | POS | 245.1142(100) | | 882508 | 110658 | 882978 | 121097 | 9.76E-01 | 0 | 0.89 |
| HILIC_POS_150_11267a0_893 | | 150.1127 | 0.89 | | HILIC | POS | 150.1127(100), 132.1046(45) | | 2839870 | 1467767 | 2845436 | 1246279 | 9.76E-01 | 0 | 1.2 |
| HILIC_NEG_315_04202a5_869 | | 315.0420 | 5.87 | | HILIC | NEG | | | 1778049 | 1276817 | 1783333 | 1301202 | 9.76E-01 | 0 | 0.43 |
| HILIC_POS_443_3342a4_281 | | 443.3342 | 4.28 | | HILIC | POS | | | 64186.4 | 48882 | 64375.7 | 42469.1 | 9.76E-01 | 0 | 0.31 |
| RP_NEG_589_37354a12_054 | | 589.3735 | 12.05 | | RP | NEG | | | 153650 | 20681.9 | 153566 | 20472 | 9.76E-01 | 0 | 0.93 |
| RP_POS_372_95865a15_454 | | 372.9587 | 15.45 | | RP | POS | | | 424437 | 159079 | 425079 | 148089 | 9.76E-01 | 0 | 1.14 |
| RP_POS_436_21753a6_785 | | 436.2175 | 6.79 | | RP | POS | | | 898494 | 77207.7 | 898186 | 72692.3 | 9.76E-01 | 0 | 0.81 |
| HILIC_NEG_183_06633a3_048 | | 183.0663 | 3.05 | | HILIC | NEG | | | 841749 | 73380 | 841447 | 78000.3 | 9.77E-01 | 0 | 0.5 |
| HILIC_POS_178_10741a0_778 | | 178.1074 | 0.78 | | HILIC | POS | | | 523088 | 251809 | 522070 | 258761 | 9.77E-01 | 0 | 0.61 |
| RP_NEG_349_01266a2_483 | | 349.0127 | 2.48 | | RP | NEG | | | 736479 | 131489 | 737002 | 135567 | 9.77E-01 | 0 | 0.45 |
| RP_NEG_456_16107a10_896 | | 456.1611 | 10.90 | | RP | NEG | | | 157456 | 62621.3 | 157207 | 65541.8 | 9.77E-01 | 0 | 0.98 |
| RP_POS_173_07846a0_705 | | 173.0785 | 0.71 | | RP | POS | | | 688344 | 156002 | 687749 | 140643 | 9.77E-01 | 0 | 0.77 |
| RP_POS_542_32147a10_18 | | 542.3215 | 10.18 | | RP | POS | | | 2.1E+07 | 9466874 | 2.1E+07 | 9801908 | 9.77E-01 | 0 | 0.58 |
| HILIC_NEG_132_92339a4_332 | | 132.9234 | 4.33 | | HILIC | NEG | | | 5392593 | 441875 | 5394192 | 412797 | 9.78E-01 | 0 | 0.14 |
| HILIC_POS_190_08643a3_865 | | 190.0864 | 3.87 | | HILIC | POS | | | 151104 | 414397 | 152652 | 398950 | 9.78E-01 | 0 | 0.47 |
| HILIC_POS_368_22a4_528 | | 368.2200 | 4.53 | | HILIC | POS | | | 206339 | 166236 | 206912 | 144859 | 9.78E-01 | 0 | 0.97 |
| RP_NEG_265_00067a0_678 | | 265.0007 | 0.68 | | RP | NEG | | | 496402 | 94727.6 | 496748 | 85389.1 | 9.78E-01 | 0 | 0.26 |
| RP_NEG_367_06805a0_688 | | 367.0681 | 0.69 | | RP | NEG | | | 468712 | 94415.4 | 468370 | 89073.1 | 9.78E-01 | 0 | 0.58 |
| RP_POS_273_20581a9_891 | | 273.2058 | 9.89 | | RP | POS | | | 2236658 | 880669 | 2239742 | 746257 | 9.78E-01 | 0 | 0.74 |
| RP_POS_283_11493a4_262 | | 283.1149 | 4.26 | | RP | POS | | | 184564 | 107495 | 184149 | 108444 | 9.78E-01 | 0 | 1 |
| RP_POS_518_21649a9_207 | | 518.2165 | 9.21 | | RP | POS | | | 385028 | 422028 | 383339 | 479245 | 9.78E-01 | 0 | 0.54 |
| RP_POS_604_46051a13_554 | | 604.4605 | 13.55 | | RP | POS | | | 9613515 | 3250048 | 9601442 | 3279231 | 9.78E-01 | 0 | 0.3 |
| HILIC_NEG_203_08298a4_107 | Tryptophan | 203.0830 | 4.11 | | HILIC | NEG | 116.0506(100), 203.0834(70) | 1 | 285185 | 61105.3 | 284959 | 59624.8 | 9.78E-01 | 0.00 | 0.67 |
| RP_POS_271_09436a6_706 | Alpinetin | 271.0944 | 6.71 | | RP | POS | 271.0948(100) | 2 | 523321 | 181231 | 522654 | 185703 | 9.79E-01 | 0.00 | 1.07 |
| RP_NEG_321_21024a12_527 | | 321.2102 | 12.53 | | RP | NEG | | | 20401.8 | 92916.2 | 20088.9 | 82189.5 | 9.79E-01 | 0 | 0.39 |
| RP_NEG_554_30255a10_181 | | 554.3026 | 10.18 | | RP | NEG | | | 1026002 | 337819 | 1024716 | 361383 | 9.79E-01 | 0 | 0.57 |
| RP_POS_306_2399a10_363 | | 306.2399 | 10.36 | | RP | POS | | | 1076281 | 269975 | 1077330 | 302945 | 9.79E-01 | 0 | 0.62 |
| RP_POS_494_84079a10_041 | | 494.8408 | 10.04 | | RP | POS | | | 2720229 | 1327659 | 2724931 | 1274862 | 9.79E-01 | 0 | 0.78 |
| RP_POS_680_97687a10_403 | | 680.9769 | 10.40 | | RP | POS | | | 835039 | 471521 | 836829 | 508996 | 9.79E-01 | 0 | 0.83 |
| RP_NEG_209_00444a15_488 | | 209.0044 | 15.49 | | RP | NEG | | | 634054 | 145338 | 634481 | 110800 | 9.80E-01 | 0 | 0.43 |
| RP_NEG_394_97647a8_096 | | 394.9765 | 8.10 | | RP | NEG | | | 1037997 | 463764 | 1036463 | 419298 | 9.80E-01 | 0 | 0.52 |
| RP_POS_153_0546a4_231 | Vanillin | 153.0546 | 4.23 | | RP | POS | 125.0596(100), 93.0338(98), 111.044(80) | 2 | 895352 | 154324 | 894812 | 158341 | 9.80E-01 | 0.00 | 0.5 |
| RP_NEG_325_18448a11_589 | | 325.1845 | 11.59 | | RP | NEG | 325.1829(100), 183.0125(15) | | 4518779 | 2048090 | 4525501 | 1983484 | 9.81E-01 | 0 | 0.64 |
| HILIC_POS_150_11238a5_634 | | 150.1124 | 5.63 | | HILIC | POS | 150.1126(100) | | 3259890 | 1866664 | 3266158 | 1997239 | 9.81E-01 | 0 | 0.58 |
| HILIC_NEG_348_99942a7_475 | | 348.9994 | 7.48 | | HILIC | NEG | | | 208770 | 16415.9 | 208717 | 16721.6 | 9.81E-01 | 0 | 0.94 |
| RP_NEG_318_04575a0_679 | | 318.0458 | 0.68 | | RP | NEG | | | 315113 | 55210.8 | 315314 | 63864.1 | 9.81E-01 | 0 | 0.44 |
| RP_POS_535_43317a13_295 | | 535.4332 | 13.30 | | RP | POS | | | 401504 | 441248 | 402875 | 391031 | 9.81E-01 | 0 | 0.35 |
| RP_POS_633_92609a10_118 | | 633.9261 | 10.12 | | RP | POS | | | 2441772 | 1256438 | 2445846 | 1228388 | 9.81E-01 | 0 | 0.62 |
| RP_POS_696_37872a10_568 | | 696.3787 | 10.57 | | RP | POS | | | 1161015 | 495462 | 1163229 | 779799 | 9.81E-01 | 0 | 0.84 |
| HILIC_POS_130_04996a6_137 | | 130.0500 | 6.14 | | HILIC | POS | 84.045(100), 130.0503(30) | | 7.1E+07 | 1.2E+07 | 7.1E+07 | 1.1E+07 | 9.82E-01 | 0 | 0.78 |
| RP_NEG_200_85863a11_592 | | 200.8586 | 11.59 | | RP | NEG | | | 479837 | 141004 | 480291 | 158481 | 9.82E-01 | 0 | 0.4 |
| RP_NEG_312_0022a1_41 | | 312.0022 | 1.41 | | RP | NEG | | | 2002412 | 525419 | 2004200 | 602567 | 9.82E-01 | 0 | 0.53 |
| RP_NEG_453_60522a15_493 | | 453.6052 | 15.49 | | RP | NEG | | | 331184 | 34086.7 | 331068 | 41830.9 | 9.82E-01 | 0 | 0.65 |
| RP_POS_156_06329a14_248 | | 156.0633 | 14.25 | | RP | POS | | | 2047618 | 917314 | 2050685 | 1103702 | 9.82E-01 | 0 | 0.45 |
| RP_POS_487_36111a10_651 | | 487.3611 | 10.65 | | RP | POS | | | 4102825 | 779406 | 4105335 | 803504 | 9.82E-01 | 0 | 1.09 |
| RP_POS_496_33981a11_144 | | 496.3398 | 11.14 | | RP | POS | | | 3561836 | 844051 | 3564464 | 862180 | 9.82E-01 | 0 | 0.66 |
| RP_POS_553_45905a11_236 | | 553.4591 | 11.24 | | RP | POS | | | 1433474 | 713343 | 1431427 | 619733 | 9.82E-01 | 0 | 0.78 |
| RP_POS_611_76697a10_63 | | 611.7670 | 10.63 | | RP | POS | | | 254299 | 184142 | 254904 | 198118 | 9.82E-01 | 0 | 0.59 |
| RP_NEG_540_33026a10_489 | LysoPC 16:0 (16:0/0:0) | 540.3303 | 10.49 | | RP | NEG | 255.233(100) | 1 | 5.4E+08 | 5.3E+07 | 5.4E+08 | 6E+07 | 9.83E-01 | 0.00 | 0.86 |
| HILIC_NEG_341_99936a6_307 | | 341.9994 | 6.31 | | HILIC | NEG | | | 884795 | 339192 | 883798 | 352096 | 9.83E-01 | 0 | 0.88 |
| RP_NEG_572_43158a14_146 | | 572.4316 | 14.15 | | RP | NEG | | | 116632 | 55484.4 | 116458 | 62732.7 | 9.83E-01 | 0 | 0.52 |
| RP_POS_136_0733a3_262 | | 136.0733 | 3.26 | | RP | POS | | | 594078 | 451904 | 595387 | 439374 | 9.83E-01 | 0 | 1.03 |
| RP_POS_162_07608a9_354 | | 162.0761 | 9.35 | | RP | POS | | | 4063701 | 2125713 | 4057923 | 1762429 | 9.83E-01 | 0 | 0.38 |
| RP_POS_315_16193a10_68 | | 315.1619 | 10.68 | | RP | POS | | | 1050529 | 333209 | 1049599 | 316693 | 9.83E-01 | 0 | 0.61 |
| RP_POS_594_13025a11_244 | | 594.1303 | 11.24 | | RP | POS | | | 93701.9 | 42070.8 | 93578 | 45109.9 | 9.83E-01 | 0 | 0.31 |
| HILIC_POS_126_06649a1_885 | 5-Methylcytosine | 126.0665 | 1.89 | | HILIC | POS | 126.0659(100), 109.0347(10) | 1 | 750399 | 1307176 | 746893 | 1141011 | 9.83E-01 | 0.00 | 0.32 |
| HILIC_NEG_172_84262a3_228 | | 172.8426 | 3.23 | | HILIC | NEG | | | 2119355 | 707149 | 2121199 | 680331 | 9.84E-01 | 0 | 0.35 |
| HILIC_NEG_554_34521a1_203 | | 554.3452 | 1.20 | | HILIC | NEG | | | 157547 | 51318.5 | 157678 | 45389.7 | 9.84E-01 | 0 | 0.3 |
| RP_NEG_276_03873a0_69 | | 276.0387 | 0.69 | | RP | NEG | | | 503308 | 89499.7 | 503543 | 77918.1 | 9.84E-01 | 0 | 0.39 |
| RP_NEG_397_17947a6_981 | | 397.1795 | 6.98 | | RP | NEG | | | 672654 | 237625 | 673378 | 282713 | 9.84E-01 | 0 | 0.39 |
| RP_POS_170_11748a5_906 | | 170.1175 | 5.91 | | RP | POS | | | 353546 | 155002 | 353957 | 153636 | 9.84E-01 | 0 | 0.51 |
| RP_POS_347_18719a10_686 | | 347.1872 | 10.69 | | RP | POS | | | 1163286 | 349494 | 1162260 | 395976 | 9.84E-01 | 0 | 0.56 |
| RP_POS_552_36694a10_106 | | 552.3669 | 10.11 | | RP | POS | | | 146336 | 205319 | 146987 | 264899 | 9.84E-01 | 0 | 0.48 |
| RP_POS_659_52142a10_251 | | 659.5214 | 10.25 | | RP | POS | | | 470469 | 250001 | 469839 | 201569 | 9.84E-01 | 0 | 0.64 |
| RP_POS_777_54218a13_201 | | 777.5422 | 13.20 | | RP | POS | | | 2578696 | 1414365 | 2574520 | 1686138 | 9.84E-01 | 0 | 1.02 |
| RP_POS_848_55573a13_951 | | 848.5557 | 13.95 | | RP | POS | | | 7573364 | 7422305 | 7551825 | 8254946 | 9.84E-01 | 0 | 0.87 |
| HILIC_POS_150_11234a7_744 | | 150.1123 | 7.74 | | HILIC | POS | 150.1128(100),132.1028(40) | | 8527730 | 2431202 | 8521348 | 2416983 | 9.85E-01 | 0 | 0.98 |
| HILIC_POS_355_01086a7_359 | | 355.0109 | 7.36 | | HILIC | POS | | | 3559063 | 163880 | 3559519 | 184930 | 9.85E-01 | 0 | 0.31 |
| RP_NEG_378_7359a15_432 | | 378.7359 | 15.43 | | RP | NEG | | | 1119988 | 336295 | 1119124 | 330115 | 9.85E-01 | 0 | 0.75 |
| RP_POS_167_07011a8_594 | | 167.0701 | 8.59 | | RP | POS | | | 4641327 | 456863 | 4642505 | 476889 | 9.85E-01 | 0 | 0.28 |
| RP_POS_428_30304a8_322 | | 428.3030 | 8.32 | | RP | POS | | | 414596 | 252876 | 413915 | 278763 | 9.85E-01 | 0 | 0.83 |
| RP_NEG_293_1767a8_367 | | 293.1767 | 8.37 | | RP | NEG | | | 1388729 | 76378 | 1388892 | 61600.4 | 9.86E-01 | 0 | 0.33 |
| RP_NEG_376_78403a15_411 | | 376.7840 | 15.41 | | RP | NEG | | | 613906 | 308846 | 614605 | 268234 | 9.86E-01 | 0 | 0.68 |
| RP_NEG_993_11841a9_407 | | 993.1184 | 9.41 | | RP | NEG | | | 986204 | 110850 | 985957 | 102226 | 9.86E-01 | 0 | 0.64 |
| RP_POS_276_21716a8_851 | | 276.2172 | 8.85 | | RP | POS | | | 742974 | 222848 | 743472 | 185433 | 9.86E-01 | 0 | 0.51 |
| RP_NEG_714_50842a13_127 | L-alpha-Phosphatidylethanolamine | 714.5084 | 13.13 | | RP | NEG | 279.2332(100), 255.2329(35), 714.5129(20) | 2 | 1186003 | 484316 | 1184670 | 583084 | 9.86E-01 | 0.00 | 0.45 |
| HILIC_NEG_208_96597a4_86 | | 208.9660 | 4.86 | | HILIC | NEG | | | 165044 | 48699.9 | 164944 | 39254.9 | 9.87E-01 | 0 | 0.57 |
| RP_NEG_170_83344a11_592 | | 170.8334 | 11.59 | | RP | NEG | | | 538713 | 88789.2 | 538916 | 93660.3 | 9.87E-01 | 0 | 0.8 |
| RP_POS_249_18246a8_881 | | 249.1825 | 8.88 | | RP | POS | | | 1054603 | 435894 | 1053633 | 420179 | 9.87E-01 | 0 | 0.65 |
| RP_POS_329_21454a14_356 | | 329.2145 | 14.36 | | RP | POS | | | 8353437 | 1841297 | 8357442 | 1700521 | 9.87E-01 | 0 | 0.72 |
| RP_POS_395_38858a11_216 | | 395.3886 | 11.22 | | RP | POS | | | 360853 | 108678 | 361124 | 139412 | 9.87E-01 | 0 | 0.66 |
| RP_POS_483_33505a9_597 | | 483.3351 | 9.60 | | RP | POS | | | 548981 | 279154 | 549556 | 244867 | 9.87E-01 | 0 | 0.64 |
| HILIC_NEG_151_87405a3_095 | | 151.8741 | 3.10 | | HILIC | NEG | | | 837085 | 60962.7 | 836973 | 51445.6 | 9.88E-01 | 0 | 0.7 |
| RP_NEG_224_99561a15_471 | | 224.9956 | 15.47 | | RP | NEG | | | 772025 | 98246.9 | 771816 | 105742 | 9.88E-01 | 0 | 0.3 |
| RP_NEG_316_98413a8_409 | | 316.9841 | 8.41 | | RP | NEG | | | 938844 | 73263.1 | 938985 | 63678.4 | 9.88E-01 | 0 | 0.26 |
| RP_POS_249_18484a8_409 | | 249.1848 | 8.41 | | RP | POS | | | 1.4E+07 | 606442 | 1.4E+07 | 607716 | 9.88E-01 | 0 | 0.82 |
| RP_POS_293_17258a7_149 | | 293.1726 | 7.15 | | RP | POS | | | 221372 | 127784 | 221637 | 134439 | 9.88E-01 | 0 | 0.87 |
| RP_POS_300_86917a15_411 | | 300.8692 | 15.41 | | RP | POS | | | 1106539 | 455168 | 1107494 | 466306 | 9.88E-01 | 0 | 0.75 |
| RP_POS_343_07254a8_842 | | 343.0725 | 8.84 | | RP | POS | | | 163030 | 75051 | 162873 | 81592.1 | 9.88E-01 | 0 | 0.66 |
| RP_POS_350_25336a10_832 | | 350.2534 | 10.83 | | RP | POS | | | 384011 | 155137 | 384330 | 158677 | 9.88E-01 | 0 | 0.94 |
| RP_POS_778_53754a12_316 | | 778.5375 | 12.32 | | RP | POS | | | 3073661 | 2188826 | 3077565 | 1718462 | 9.88E-01 | 0 | 0.45 |
| RP_POS_864_62506a9_86 | | 864.6251 | 9.86 | | RP | POS | | | 1982465 | 928730 | 1980499 | 911117 | 9.88E-01 | 0 | 0.42 |
| HILIC_NEG_186_10364a4_854 | | 186.1036 | 4.85 | | HILIC | NEG | | | 1484720 | 617875 | 1483493 | 710201 | 9.89E-01 | 0 | 0.42 |
| HILIC_NEG_261_81339a3_216 | | 261.8134 | 3.22 | | HILIC | NEG | | | 131512 | 40974.5 | 131596 | 46601.2 | 9.89E-01 | 0 | 0.15 |
| RP_NEG_573_04291a0_679 | | 573.0429 | 0.68 | | RP | NEG | | | 477568 | 109742 | 477773 | 112609 | 9.89E-01 | 0 | 0.4 |
| RP_POS_295_19012a8_369 | | 295.1901 | 8.37 | | RP | POS | | | 2699271 | 133641 | 2698997 | 147208 | 9.89E-01 | 0 | 0.43 |
| HILIC_NEG_131_08273a6_378 | Ornithine | 131.0827 | 6.38 | | HILIC | NEG | 131.0829(100), 100.9252(30) | 2 | 260848 | 66357.8 | 260724 | 67419.8 | 9.89E-01 | 0.00 | 0.4 |
| RP_NEG_186_99898a15_479 | | 186.9990 | 15.48 | | RP | NEG | | | 512750 | 112161 | 512538 | 121760 | 9.90E-01 | 0 | 0.4 |
| RP_NEG_195_13898a8_208 | | 195.1390 | 8.21 | | RP | NEG | | | 334490 | 29789.9 | 334534 | 23888.8 | 9.90E-01 | 0 | 0.57 |
| RP_POS_321_2399a10_291 | | 321.2399 | 10.29 | | RP | POS | | | 5528445 | 1143442 | 5526502 | 1188712 | 9.90E-01 | 0 | 1.03 |
| RP_POS_352_24518a14_237 | | 352.2452 | 14.24 | | RP | POS | | | 809390 | 414202 | 810102 | 436283 | 9.90E-01 | 0 | 0.53 |
| RP_POS_389_97498a0_678 | | 389.9750 | 0.68 | | RP | POS | | | 343940 | 110795 | 344116 | 95790.7 | 9.90E-01 | 0 | 0.27 |
| RP_POS_468_30875a10_701 | | 468.3088 | 10.70 | | RP | POS | | | 821112 | 274906 | 820627 | 295082 | 9.90E-01 | 0 | 0.83 |
| RP_POS_538_35052a9_193 | | 538.3505 | 9.19 | | RP | POS | | | 572717 | 264360 | 572291 | 216175 | 9.90E-01 | 0 | 0.43 |
| RP_POS_586_90796a9_767 | | 586.9080 | 9.77 | | RP | POS | | | 1068638 | 510546 | 1069657 | 667463 | 9.90E-01 | 0 | 0.76 |
| HILIC_POS_224_12796a5_465 | | 224.1280 | 5.47 | | HILIC | POS | 224.128(100), 165.0545(40) | | 3683761 | 729257 | 3682490 | 845298 | 9.91E-01 | 0 | 0.21 |
| HILIC_NEG_129_03664a0_937 | | 129.0366 | 0.94 | | HILIC | NEG | | | 317495 | 139956 | 317712 | 137927 | 9.91E-01 | 0 | 0.41 |
| HILIC_NEG_136_89174a3_204 | | 136.8917 | 3.20 | | HILIC | NEG | | | 1.4E+08 | 7449252 | 1.4E+08 | 8685223 | 9.91E-01 | 0 | 0.52 |
| HILIC_NEG_221_01247a1_343 | | 221.0125 | 1.34 | | HILIC | NEG | | | 1015898 | 385850 | 1016451 | 333506 | 9.91E-01 | 0 | 0.73 |
| RP_NEG_431_18521a8_802 | | 431.1852 | 8.80 | | RP | NEG | | | 132690 | 87186 | 132839 | 102490 | 9.91E-01 | 0 | 0.45 |
| RP_NEG_452_92407a12_169 | | 452.9241 | 12.17 | | RP | NEG | | | 1760533 | 861907 | 1761740 | 778877 | 9.91E-01 | 0 | 0.58 |
| RP_POS_1083_92896a15_163 | | 1083.9290 | 15.16 | | RP | POS | | | 497123 | 258122 | 496691 | 283073 | 9.91E-01 | 0 | 0.71 |
| RP_POS_314_88477a15_412 | | 314.8848 | 15.41 | | RP | POS | | | 1895789 | 777725 | 1894619 | 717155 | 9.91E-01 | 0 | 0.87 |
| RP_POS_317_11459a9_437 | | 317.1146 | 9.44 | | RP | POS | | | 800713 | 133771 | 800504 | 135273 | 9.91E-01 | 0 | 0.52 |
| RP_POS_328_28464a10_755 | | 328.2846 | 10.76 | | RP | POS | | | 480137 | 60292.2 | 480223 | 54898.8 | 9.91E-01 | 0 | 0.61 |
| RP_POS_444_31635a6_541 | | 444.3164 | 6.54 | | RP | POS | | | 818684 | 734832 | 817621 | 594351 | 9.91E-01 | 0 | 0.64 |
| RP_POS_518_3219a11_143 | | 518.3219 | 11.14 | | RP | POS | | | 1509227 | 294567 | 1509759 | 378305 | 9.91E-01 | 0 | 0.74 |
| RP_POS_668_34821a10_57 | | 668.3482 | 10.57 | | RP | POS | | | 700179 | 273899 | 699574 | 443317 | 9.91E-01 | 0 | 0.55 |
| HILIC_POS_147_07632a6_138 | Cholic acid (dimer) | 147.0763 | 6.14 | | HILIC | POS | 84.0452(100), 130.0506(80) | 1 | 1.3E+08 | 2.1E+07 | 1.3E+08 | 1.9E+07 | 9.91E-01 | 0.00 | 0.75 |
| HILIC_NEG_397_88922a5_46 | | 397.8892 | 5.46 | | HILIC | NEG | | | 463372 | 55956.3 | 463454 | 60408.1 | 9.92E-01 | 0 | 0.69 |
| RP_NEG_188_94972a0_681 | | 188.9497 | 0.68 | | RP | NEG | | | 1238259 | 123956 | 1238449 | 138495 | 9.92E-01 | 0 | 0.69 |
| RP_NEG_191_94772a0_86 | | 191.9477 | 0.86 | | RP | NEG | | | 2213301 | 550198 | 2213972 | 492478 | 9.92E-01 | 0 | 0.79 |
| RP_NEG_339_68634a15_497 | | 339.6863 | 15.50 | | RP | NEG | | | 518241 | 40675.3 | 518296 | 38698.9 | 9.92E-01 | 0 | 0.69 |
| RP_POS_310_23535a9_325 | | 310.2354 | 9.33 | | RP | POS | | | 2933158 | 227388 | 2933465 | 218557 | 9.92E-01 | 0 | 0.61 |
| RP_POS_434_32086a9_647 | | 434.3209 | 9.65 | | RP | POS | | | 3268768 | 1480209 | 3266594 | 1743579 | 9.92E-01 | 0 | 0.51 |
| RP_POS_574_06696a10_003 | | 574.0670 | 10.00 | | RP | POS | | | 398442 | 226255 | 398097 | 248294 | 9.92E-01 | 0 | 0.4 |
| HILIC_NEG_198_90463a0_898 | | 198.9046 | 0.90 | | HILIC | NEG | | | 961358 | 252662 | 960969 | 341056 | 9.93E-01 | 0 | 0.4 |
| HILIC_NEG_268_91125a0_902 | | 268.9113 | 0.90 | | HILIC | NEG | | | 244539 | 37443.9 | 244590 | 44377.7 | 9.93E-01 | 0 | 0.3 |
| HILIC_NEG_341_00394a5_106 | | 341.0039 | 5.11 | | HILIC | NEG | | | 408797 | 208355 | 409037 | 178519 | 9.93E-01 | 0 | 0.59 |
| RP_NEG_464_55191a15_495 | | 464.5519 | 15.50 | | RP | NEG | | | 592322 | 49914.4 | 592382 | 50288.5 | 9.93E-01 | 0 | 0.83 |
| RP_NEG_702_32312a10_612 | | 702.3231 | 10.61 | | RP | NEG | | | 3154264 | 381383 | 3154722 | 433466 | 9.93E-01 | 0 | 0.65 |
| RP_POS_432_81729a9_86 | | 432.8173 | 9.86 | | RP | POS | | | 1804543 | 933355 | 1803390 | 993981 | 9.93E-01 | 0 | 0.56 |
| HILIC_POS_146_11754a2_216 | Acetylcholine | 146.1175 | 2.22 | | HILIC | POS | 146.117(100), 87.0448(5) | 2 | 738291 | 907794 | 739395 | 1039723 | 9.93E-01 | 0.00 | 0.74 |
| RP_NEG_267_14426a9_656 | | 267.1443 | 9.66 | | RP | NEG | | | 18512.5 | 27019.8 | 18484.5 | 30834.1 | 9.94E-01 | 0 | 0.24 |

**Supplemantary table 1**: The non-targeted metabolomics results ordered by the VIP value (p value < 0.0003 are shaded dark green, p value < 0.05 are shaded light green, d values below 0 are blue and above 0 are red)

| | | | | | | | | | Cases | | Controls | | Welch test | Cohen's | |
|---|---|---|---|---|---|---|---|---|---|---|---|---|---|---|---|
| Feature_ID | Metabolite_name | Average_Mz | Average_RT_min_ | HMDB ID | RP_HILIC | POS_NEG | Cureted_MSMS | ID_level | Mean | SD | Mean2 | SD3 | p | d | VIP |
| RP_NEG_372_99069a8_657 | | 372.9907 | 8.66 | | RP | NEG | | | 808285 | 215815 | 808519 | 230817 | 9.94E-01 | 0 | 0.4 |
| RP_NEG_481_28131a9_218 | | 481.2813 | 9.22 | | RP | NEG | | | 696026 | 1065953 | 694600 | 1575227 | 9.94E-01 | 0 | 0.3 |
| RP_POS_356_27982a9_685 | | 356.2798 | 9.69 | | RP | POS | | | 363762 | 225141 | 364000 | 235118 | 9.94E-01 | 0 | 0.8 |
| RP_POS_563_06964a10_559 | | 563.0696 | 10.56 | | RP | POS | | | 479165 | 257378 | 478918 | 264670 | 9.94E-01 | 0 | 0.56 |
| RP_POS_604_31122a10_931 | | 604.3112 | 10.93 | | RP | POS | | | 858401 | 434549 | 858896 | 495006 | 9.94E-01 | 0 | 0.46 |
| RP_POS_752_55829a14_25 | | 752.5583 | 14.25 | | RP | POS | | | 3914964 | 1983045 | 3912926 | 1846878 | 9.94E-01 | 0 | 0.82 |
| HILIC_NEG_146_96123a7_998 | | 146.9612 | 8.00 | | HILIC | NEG | | | 392420 | 169481 | 392597 | 219714 | 9.95E-01 | 0 | 0.33 |
| HILIC_POS_141_06596a5_243 | | 141.0660 | 5.24 | | HILIC | POS | | | 1727263 | 2958655 | 1729537 | 2753774 | 9.95E-01 | 0 | 0.5 |
| HILIC_POS_209_13841a0_546 | | 209.1384 | 0.55 | | HILIC | POS | | | 4734351 | 3387069 | 4737107 | 3300450 | 9.95E-01 | 0 | 0.46 |
| RP_NEG_330_83612a15_396 | | 330.8361 | 15.40 | | RP | NEG | | | 7173342 | 963861 | 7172521 | 938613 | 9.95E-01 | 0 | 0.7 |
| RP_POS_152_96948a0_632 | | 152.9695 | 0.63 | | RP | POS | | | 501496 | 229365 | 501690 | 232030 | 9.95E-01 | 0 | 0.62 |
| RP_POS_474_32663a6_182 | | 474.3266 | 6.18 | | RP | POS | | | 594232 | 390734 | 594533 | 366755 | 9.95E-01 | 0 | 0.85 |
| RP_POS_476_84222a9_823 | | 476.8422 | 9.82 | | RP | POS | | | 2574849 | 1255973 | 2575934 | 1237406 | 9.95E-01 | 0 | 0.59 |
| RP_POS_586_30878a10_358 | | 586.3088 | 10.36 | | RP | POS | | | 1480855 | 194042 | 1481018 | 211811 | 9.95E-01 | 0 | 0.81 |
| RP_POS_149_02338a9_44 | | 149.0234 | 9.44 | | RP | POS | 149.023(100), 121.0286(12) | | 1.6E+07 | 1132484 | 1.6E+07 | 1181468 | 9.96E-01 | 0 | 0.75 |
| HILIC_NEG_213_04977a6_135 | | 213.0498 | 6.14 | | HILIC | NEG | | | 308091 | 31116.7 | 308068 | 32964 | 9.96E-01 | 0 | 0.79 |
| HILIC_NEG_242_92738a4_864 | | 242.9274 | 4.86 | | HILIC | NEG | | | 207389 | 58119.4 | 207428 | 54036.9 | 9.96E-01 | 0 | 0.65 |
| HILIC_NEG_84_93823a3_135 | | 84.9382 | 3.14 | | HILIC | NEG | | | 254263 | 65659.8 | 254213 | 72346.3 | 9.96E-01 | 0 | 0.43 |
| HILIC_POS_148_04265a1_834 | | 148.0427 | 1.83 | | HILIC | POS | | | 595952 | 488574 | 596311 | 537454 | 9.96E-01 | 0 | 0.58 |
| RP_NEG_280_98334a7_568 | | 280.9833 | 7.57 | | RP | NEG | | | 1829128 | 281404 | 1829353 | 343640 | 9.96E-01 | 0 | 0.55 |
| RP_NEG_360_99051a8_449 | | 360.9905 | 8.45 | | RP | NEG | | | 1180501 | 174556 | 1180637 | 181965 | 9.96E-01 | 0 | 0.21 |
| RP_POS_279_159a7_384 | | 279.1590 | 7.38 | | RP | POS | | | 1187306 | 432334 | 1187032 | 437517 | 9.96E-01 | 0 | 0.45 |
| RP_POS_282_20386a7_747 | | 282.2039 | 7.75 | | RP | POS | | | 657414 | 301971 | 657239 | 260921 | 9.96E-01 | 0 | 0.43 |
| RP_POS_373_21109a10_895 | | 373.2111 | 10.90 | | RP | POS | | | 973940 | 493544 | 973615 | 506866 | 9.96E-01 | 0 | 0.83 |
| RP_POS_510_03323a10_349 | | 510.0332 | 10.35 | | RP | POS | | | 626927 | 444479 | 626601 | 423383 | 9.96E-01 | 0 | 0.47 |
| RP_POS_530_03961a10_031 | | 530.0396 | 10.03 | | RP | POS | | | 743223 | 346427 | 742983 | 400721 | 9.96E-01 | 0 | 0.45 |
| RP_POS_743_15631a9_279 | | 743.1563 | 9.28 | | RP | POS | | | 672638 | 69561.4 | 672689 | 68312.8 | 9.96E-01 | 0 | 0.6 |
| RP_POS_394_25766a6_981 | | 394.2577 | 6.98 | | RP | POS | | | 226574 | 131871 | 226489 | 152129 | 9.97E-01 | 0 | 0.55 |
| RP_POS_468_32983a9_823 | | 468.3298 | 9.82 | | RP | POS | | | 984526 | 481512 | 984252 | 509476 | 9.97E-01 | 0 | 0.53 |
| RP_POS_506_02203a9_723 | | 506.0220 | 9.72 | | RP | POS | | | 157563 | 97818.7 | 157516 | 106414 | 9.97E-01 | 0 | 0.45 |
| RP_POS_785_52795a13_208 | | 785.5280 | 13.21 | | RP | POS | | | 3252441 | 1501986 | 3251858 | 1214175 | 9.97E-01 | 0 | 0.42 |
| RP_POS_500_30386a9_124 | | 500.3039 | 9.12 | | RP | POS | 464.2791(100), 126.0228(40) | | 864386 | 1716247 | 865001 | 1592787 | 9.98E-01 | 0 | 0.39 |
| RP_POS_221_15327a9_92 | | 221.1533 | 9.92 | | RP | POS | 221.1532(100) | | 1626260 | 211222 | 1626184 | 217476 | 9.98E-01 | 0 | 0.33 |
| RP_NEG_498_29047a9_342 | | 498.2905 | 9.34 | | RP | NEG | 498.2872(100), 79.957(2) | | 7247239 | 1.9E+07 | 7254676 | 1.6E+07 | 9.98E-01 | 0 | 0.64 |
| HILIC_NEG_134_89478a4_763 | | 134.8948 | 4.76 | | HILIC | NEG | | | 2.6E+07 | 1.3E+07 | 2.6E+07 | 1.1E+07 | 9.98E-01 | 0 | 0.43 |
| HILIC_NEG_268_98666a3_995 | | 268.9867 | 4.00 | | HILIC | NEG | | | 916377 | 554625 | 916571 | 528727 | 9.98E-01 | 0 | 0.32 |
| HILIC_POS_366_2847a3_221 | | 366.2847 | 3.22 | | HILIC | POS | | | 469896 | 136233 | 469932 | 126452 | 9.98E-01 | 0 | 0.39 |
| RP_NEG_671_06287a9_098 | | 671.0629 | 9.10 | | RP | NEG | | | 408029 | 26973.1 | 408038 | 28068 | 9.98E-01 | 0 | 0.63 |
| RP_POS_520_33997a10_18 | LysoPC 18:2 (0:0/18:2) | 520.3400 | 10.18 | | RP | POS | 184.0743(100), 86.0974(12) | 1 | 1.4E+08 | 5.6E+07 | 1.4E+08 | 5.9E+07 | 9.98E-01 | 0.00 | 0.58 |
| RP_POS_163_97652a15_571 | | 163.9765 | 15.57 | | RP | POS | | | 2E+08 | 7.4E+07 | 2E+08 | 7.2E+07 | 9.99E-01 | 0 | 0.7 |
| RP_POS_289_90549a15_546 | | 289.9055 | 15.55 | | RP | POS | | | 504961 | 197395 | 504917 | 209971 | 9.99E-01 | 0 | 0.31 |
| RP_POS_489_35226a10_379 | | 489.3523 | 10.38 | | RP | POS | | | 225700 | 182328 | 225668 | 180521 | 9.99E-01 | 0 | 0.37 |
| RP_POS_870_52277a12_74 | | 870.5228 | 12.74 | | RP | POS | | | 7383226 | 2530566 | 7382963 | 2497850 | 9.99E-01 | 0 | 0.73 |
| HILIC_NEG_211_87418a3_135 | | 211.8742 | 3.14 | | HILIC | NEG | | | 235108 | 57691.5 | 235110 | 67989.2 | 1.00E+00 | 0 | 0.2 |
| RP_POS_183_13799a6_719 | | 183.1380 | 6.72 | | RP | POS | | | 680543 | 77899.4 | 680546 | 81465.8 | 1.00E+00 | 0 | 0.19 |
| RP_POS_397_30322a9_914 | | 397.3032 | 9.91 | | RP | POS | | | 504840 | 365746 | 504813 | 390780 | 1.00E+00 | 0 | 0.6 |
